# Investigating 24-Hour Urine Expected Ranges of Non-Kidney Stone Formers: an EAU Endourology Research Group Systematic Review and Meta-Analysis

**DOI:** 10.64898/2026.09.17.26363294

**Authors:** Kimberley A. Noble, Anastasia Glubb, Parag Roy, Jacqueline Howard, Lazaros Tzelves, Steffi Yuen, Paul Cook, Bhaskar K. Somani, Daniel G. Fuster, Pietro Manuel Ferraro, Gary C. Curhan, Sarah Howles, John A. Sayer, Oisín N. Kavanagh, Robert Geraghty

**Author notes:** Joint first author (Kimberley A. Noble, Anastasia Glubb). Joint senior author (Oisín N. Kavanagh, Robert Geraghty).

## Abstract

**Background and Objective:** There is substantial variation in reference ranges for 24-hour urine collections, leading to diagnostic uncertainty. We aimed to curate robust evidence for expected ranges of 24-hour urine collections in the non-kidney stone forming population.

**Methods:** We performed a systematic review (PROSPERO ID: CRD42024590784) of healthy adults without kidney stone disease undergoing 24-hour urine collections. Each component was meta-analysed in R, stratified by age, sex and ancestry, meta-regression and sensitivity analyses for study design, collection completeness and bias. An explicit eight-branch decision framework selected the recommended analysis per component. Expected ranges were derived from the pooled SD (95% interval); evidence was quantified by GRADE.

**Key Findings and Limitations:** Seventy-two articles were included (10,284 participants). Expected ranges (mmol / 24 h unless stated): volume 1.0–2.5L, calcium 1.9–6.2, oxalate 0.2–0.6 (men) / 0.2–0.4 (women), urate 2.0–5.1(men)/2.3–3.5(women), pH 5.6–6.3, citrate 1.2–4.5, creatinine 12.2– 16.3(men)/7.2–12.6(women), phosphate 19–26, sodium 98–253, potassium 28–77, magnesium 3.1–4.8, ammonium 28–46(men)/20–40(women), chloride 84–218, sulphate 17–26(men)/7–30(women), urea 234–445. Principal limitations were: high between-study heterogeneity (I^2^>95% for most components) and assay/laboratory variability (most marked for citrate and potassium).

**Conclusions and Clinical Implications:** This is the first study to attempt a robust characterization of 24-hour urine expected ranges using meta-analysis. Six of the derived expected ranges (pH, oxalate, magnesium, sulphate, urate, creatinine) were broadly concordant with existing reference ranges, providing an evidence base for those specific components.

**Take Home Message:** This study establishes expected 24-hour urine values in a non-stone-forming population and provides empirical support for the derivation of reference ranges for several common urine analytes. I t also provides a repository of data for further analyses.

**Patient summary:** The existing 24-hour urine reference ranges are poorly characterised. These are important as they are used by clinicians to diagnose and guide treatment for conditions predisposing to kidney stone formation. We have robustly determined expected ranges for people without kidney stones after reviewing existing studies in more than 10,000 people. For several urine components the available data remain limited, and our ranges should be revisited as more evidence accumulates.

## INTRODUCTION

Twenty-four hour urine collections are often used to diagnose underlying metabolic abnormalities in patients with kidney stone disease^1^. These can guide patient management, including initiation of preventative therapies and response to dietary and drug interventions. However, there is significant global variation in the definition of 24-hour urine reference ranges^1–4^. Determinants of existing reference ranges are infrequently cited^3^, and where references do exist, they are often from small studies from 30–40 years ago^5^. There is also evidence of variation between sexes^6^, ancestries^7^ and ages^8^, which are often not accounted for in published reference ranges.

There are broadly two ways to derive a reference range for biochemical values. The first is a statistical (diagnostic) approach, which utilises a healthy population, presumes a normal distribution of the component and takes a confidence interval of the values from that population^2,9^. The second is a clinically informed (therapeutic) approach, utilising studies where a particular benefit threshold is demonstrated, for example a 24-hour urine volume >2.5 L reduces risk of subsequent stone formation^10^.

Given the ambiguity surrounding 24-hour urine reference ranges, we aimed to utilise the statistical (diagnostic) approach to derive standardised expected ranges for healthy adults without kidney stone disease using a systematic review and meta-analysis of studies reporting 24-hour urine values in these populations. We examined ammonium, calcium, chloride, citrate, creatinine, magnesium, oxalate, pH, phosphate, potassium, sodium, sulphate, urate, urea and volume. Our secondary aim was to develop an online application for our results to be explored and integrated into laboratory reporting and clinical use.

## METHODS

### Search strategy

A systematic review of the literature was performed in accordance with the Preferred Reporting Items for Systematic Reviews and Meta-Analyses (PRISMA) statement [see **supplementary table** 1]^11^. Google Scholar, Ovid MEDLINE, PubMed, Scopus and Web of Science databases were searched for eligible studies. Terms used included ‘human urine’ or ‘urine composition’ and ‘24-hour’ or ‘24-h’ or ‘24 hour’. The search was performed by a professional librarian (JH). The review was registered on PROSPERO (ID: CRD42024590784). We also examined individual papers and guidelines, e.g. European Association of Urology (EAU) and American Urological Association (AUA) guidelines. The search criteria [see **Appendix 1**] and results are available [see data availability / **Appendices 2–3**].

### Eligibility criteria

Studies included randomised controlled trials (RCTs) and observational cohort studies. We excluded case reports and opinion papers. Participants included all adults ≥18 years old without diagnosed kidney stone disease. Participants were included based on “no diagnosed kidney stone disease” as reported by the original authors. We included studies that described the measurement of the following 24-hour urine components: ammonium, calcium, chloride, citrate, creatinine, magnesium, oxalate, pH, phosphate, potassium, sodium, sulphate, urate, urea and urine volume.

### Data extraction

Two reviewers (AG and KN) extracted data independently, with any conflict resolved by the senior authors (RG and OK). Data were extracted into an Excel spreadsheet (see data availability). Corresponding authors of papers with missing or ambiguous data (e.g. unreported standard deviations, unclear cohort sizes) were contacted by email for clarification or supplementary data. If median/interquartile range was reported rather than mean/standard deviation, we then converted from the former to the latter^12^. For RCTs, the pre-intervention (baseline) values from each arm were extracted.

### Data analysis

We used R (R Foundation for Statistical Computing, Vienna, Austria) to perform a meta-analysis of means (metamean) with the package ‘meta’. We generated meta-analysed expected ranges based on the assumption of normal distribution, apart from volume where inspection of the per-study distribution suggested a right-skew and we therefore performed a log-transformation; other components did not show a comparable skew on visual inspection of forest plots and were retained on the natural scale. The expected range was calculated according to existing standards for reporting reference ranges (<5% / >95% for normal distribution)^9^. A random-effects model was used when substantial heterogeneity was suspected, while a fixed-effect model was applied when there was minimal heterogeneity. Heterogeneity was evaluated using I^2^, τ^2^, and Cochran’s Q statistics^13^. Publication bias was assessed statistically with Egger’s and Begg’s tests, and visually with Funnel/Baujat plots.

We performed sub-analyses for age, sex, ancestry, and country of study (used as a proxy for laboratory assay type, since direct assay types were rarely reported). Sensitivity analyses were performed for study design (RCT only) and risk of bias (low only and low/moderate only). Differences between sub-analyses were tested with Wald tests.

Where there were sufficient data, we performed meta-regression using age, sex, ancestry, study size and overall risk of bias as covariates. Some studies contributed multiple cohorts, therefore the number of contributing cohorts (k) per component is therefore greater than the number of unique studies.

We applied an explicit eight-branch decision framework (Figure 1; full specification in Appendix 4 section 4) to select the recommended analysis per component. Branches evaluated, in order: data sufficiency; robust sex, age, ancestry, and assay/country differences; meta-regression effectiveness; low risk-of-bias sub-analysis; and overall pooled as default. Where the framework selected an assay-stratified branch the result is reported as the overall pooled estimate. This was due to the uncertainty in assay ascertainment, which limited our ability to be more specific. In some cases, we have utilised country as a proxy for assay (given the difference in practice between countries). This framework prioritised data sufficiency and robust stratum differences before falling back on overall pooled analyses, ensuring transparent and reproducible analysis selection.

### Bias analysis

We used the Cochrane risk of bias tools for both observational studies (exposures)^14^ and randomised trials^15^. We summarised bias into an overall category. This was based on: any domain being high risk then overall was assigned high risk; if any two domains were moderate then overall was assigned moderate; otherwise overall was assigned to low risk.

### GRADE analysis

We utilised the GRADE methodology to determine the level of evidence for each meta-analysis-derived 24-hour urine reference range15. Within this expected-range context, GRADE ratings reflect certainty that further research will change the estimate. High indicates very high confidence in the estimated range; Moderate indicates further research is likely to have an important impact on confidence and may change the estimate; Low and Very Low indicate further research is very likely to change the estimate. We emphasise that a Low or Very Low rating reflects between-study heterogeneity and limited stratified data rather than an incorrect pooled estimate; these ranges should be interpreted alongside the caveats described per component (Table 1 footnote).

### Reference ranges

Clinically available reference ranges for 24-hour urine biochemistry were used as a comparison to our calculated expected ranges, including: the European Association of Urology Guidelines^1^, Mayo Clinic Laboratories^16^, Litholink^17^, Hong Kong [see **Appendix 4**] and two NHS trusts in the UK (York^18^ and Bristol^19^).

### Code/Data availability

The statistical code, search results and data used for these analyses are provided in **Appendix 4**. A public repository and shiny application will be released at the time of publication (URLs withheld pending peer review).

## RESULTS

### Search Results

The literature search identified 527 records, of which 72 studies were included after screening [see **Figure 2** and Supplementary Table 1]. These studies comprised 10,284 participants with a mean age of 47.7 years (±10 years). Sex distribution included 3,245 men and 6,134 women; several studies did not report sex-specific data.

Six studies reported data stratified by ancestry (White European, Black/Afro-Caribbean and East Asian)^6,20–24^, while 15 studies examined specific age groups beyond the standard ≥18 years category^6,24–33^. Eleven studies were randomised controlled trials; for these, baseline (pre-intervention) values from each arm were extracted^21,32,34–42^.

Per-component sample sizes are reported in Table 1, ranging from the smallest (urea, k=13 cohorts, n=3,876) to the largest (calcium, k=100 cohorts, n=10,284). Studies originated from multiple countries, with the largest contributions from the USA, UK and Italy.

### Bias Analysis

Overall, 34 studies were at high risk of bias, 22 were at moderate risk and the remainder were at low risk [see **Appendix 4 section 8**].

### Meta-Analysis Results

#### Overall

Across the 15 components, the framework selected sex-stratified analysis for five (ammonium, creatinine, oxalate, sulphate, urate), meta-regression for six (calcium, chloride, magnesium, pH, phosphate, sodium), and the unadjusted overall pooled estimate for two (urea, volume). Assay variability was detected in two (citrate, potassium) and these are reported as the overall pooled estimate. Overall results are detailed in **Table 1. Table 2** compares the calculated expected range of non-kidney stone forming populations to existing reference ranges. All analyses associated with each 24-hour urine component are detailed in **Appendix 4, section 6**. We briefly summarise the results below; a detailed explanation of per-component analysis selection and GRADE rating is provided in **Appendix 4**, section 9.

#### Ammonium

Forty-five cohorts contributed (n=4,841). Sex differences on meta-regression (p=0.035) supported sex-stratified analysis: 28–46 mmol/24 h for men (n=535) and 20–40 mmol/24 h for women (n=3,857), both of Moderate GRADE rating. The men’s range is concordant with EAU (<50) and Mayo (15–56); the women’s range is lower than published comparators [see Table 2; Appendix 4 sections 6.12 & 9.14].

#### Calcium

One hundred cohorts contributed (n=10,284). Meta-regression explained 29% of the heterogeneity (R^2^=29.4%, above the 10% threshold) and thus was the recommended analysis (n=5,418), yielding 1.9– 6.2 mmol/24 h. The GRADE rating was Low, reflecting residual heterogeneity. The range lies between the NHS (2.5–7.5)/NDH (2–7.4) and Mayo (<5)/Litholink [see Table 2; Appendix 4 sections 6.2 & 9.4].

#### Chloride

Fifty-one cohorts contributed (n=3,534). Meta-regression explained ~30% of the heterogeneity (R^2^=29.7%) and was therefore the recommended analysis (n=3,173), yielding 84–218 mmol/24 h, with a Moderate GRADE rating. Existing reference ranges vary widely (Mayo 34–286; NHS/NDH 110–250; Litholink 50–120) and our range falls between these [see Table 2; Appendix 4 sections 6.13 & 9.15].

#### Citrate

Ninety-seven cohorts contributed (n=9,458). The framework detected significant between-assay/country variability (minimum Wald p<0.001, 44.8% relative shift) and therefore we report the overall pooled estimate of 1.2–4.5 mmol/24 h. The GRADE rating was Very Low. Our range is concordant with NHS (0.6–4.8) and Mayo (1.3–6.2) [see Table 2; Appendix 4 sections 6.7 & 9.9].

#### Creatinine

Seventy-four cohorts contributed (n=9,058). Highly significant sex differences on meta-regression (p<0.001) supported sex-stratified analysis: 12.2–16.3 mmol/24 h for men (n=760, GRADE Very Low) and 7.2–12.6 mmol/24 h for women (n=4,217, GRADE Low). Both ranges fall within published comparators (EAU, NHS, Mayo, NDH) and are broadly concordant [see Table 2; Appendix 4 sections 6.4 & 9.6].

#### Magnesium

Eighty-seven cohorts contributed (n=9,513). Meta-regression explained ~17% of the heterogeneity (R^2^=17.3%) and thus was the recommended analysis (n=4,724), yielding 3.1–4.8 mmol/24 h with a Moderate GRADE rating. The range seems to sit within the middle of the existing reference ranges: EAU (>3), NHS (2.4–6.5), Mayo (2.1–4.3) and Litholink (2.6–7.1) [see Table 2; Appendix 4 sections 6.11 & 9.13].

#### Oxalate

Ninety-two cohorts contributed (n=9,725). Sex differences on meta-regression (p=0.012) supported sex-stratified analysis: 0.2–0.6 mmol/24 h for men (n=860) and 0.2–0.4 mmol/24 h for women (n=4,177), both had a Very Low GRADE rating. The men’s upper limit slightly exceeds the EAU/NHS/Mayo cut-off (<0.5); the women’s range matches Litholink (<0.4) [see Table 2; Appendix 4 sections 6.5 & 9.7].

#### pH

Ninety-five cohorts contributed (n=9,226). Meta-regression explained ~12% of the heterogeneity (R^2^=11.6%) and therefore was the recommended analysis (n=4,379), yielding 5.6–6.3, with a Moderate GRADE rating. The range is concordant with Litholink (5.7–6.3) and falls within the broader EAU (5.5–7) and Mayo/NDH (4.5–8) ranges [see Table 2; Appendix 4 sections 6.6 & 9.8].

#### Phosphate

Fifty-six cohorts contributed (n=8,104). Meta-regression explained ~77% of the heterogeneity (R^2^=76.8%) with significant small-study bias on Egger’s test (p=0.001). Meta-regression was the recommended analysis (n=3,974), yielding 19–26 mmol/24h, with a Low GRADE rating. This range is narrower than all published comparators, which span <35 (EAU) to 7.3–58 (Mayo) [see Table 2; Appendix 4 sections 6.8 & 9.10].

#### Potassium

Ninety-three cohorts contributed (n=9,418). The framework detected significant between-assay/country variability (minimum Wald p=0.010, 44.9% relative shift). We therefore report the overall pooled estimate of 28–77 mmol/24 h. This estimate has a Very Low GRADE rating. The range is concordant with the lower portion of NHS (25–125) and Mayo (16–105) [see Table 2; Appendix 4 sections 6.10 & 9.12].

#### Sodium

Ninety-four cohorts contributed (n=9,120). Meta-regression explained ~12% of the heterogeneity (R^2^=11.5%), with the moderator-adjusted estimate differing substantially from the unadjusted overall. The meta-regression was the recommended analysis (n=3,945), yielding 98–253 mmol/24 h, with a Moderate GRADE rating. The range is concordant with the upper limits of NHS (40–250) and NDH (40–220) but exceeds Litholink (50–120) [see Table 2; Appendix 4 sections 6.9 & 9.11].

#### Sulphate

Forty-one cohorts contributed (n=4,857). There were highly significant sex differences on meta-regression (p<0.001) supporting sex-stratified analysis: 17–26 mmol/24 h for men (n=642) and 7–30 mmol/24 h for women (n=3,794), both had a Moderate GRADE rating. Sex-specific comparators were not available in existing reference ranges, however our ranges fall within Mayo’s overall (7–47) [see Table 2; Appendix 4 sections 6.14 & 9.16].

#### Urate

Ninety-three cohorts contributed (n=9,544). Significant sex differences were detected on meta-regression (p=0.042) supporting sex-stratified analysis: 2.0–5.1 mmol/24 h for men (n=860) and 2.3–3.5 mmol/24 h for women (n=4,177), both with a Very Low GRADE rating. Both sex-specific ranges are broadly concordant with published comparators (EAU <5/<4, Mayo, Litholink) [see Table 2; Appendix 4 sections 6.3 & 9.5].

#### Urea

Thirteen cohorts contributed (n=3,876). Meta-regression was uninterpretable given the small dataset, therefore the overall pooled analysis was used, yielding 234–445 mmol/24 h, with a Low GRADE rating. This range is broadly concordant with NHS (250–570), Litholink (249.9–535.5) and Mayo (124.9–749.7) reference ranges [see Table 2; Appendix 4 sections 6.15 & 9.17].

#### Volume

Eighty-nine cohorts contributed (n=9,144). Meta-regression explained little heterogeneity (R^2^=7.6%, below the 10% threshold) and no stratum effect was significant, therefore the overall pooled analysis was used, yielding 1.0–2.5 L/24 h, with a High GRADE rating. The lower limit exceeds the EAU minimum (>0.5) and the upper limit matches Litholink’s therapeutic threshold (>2.5 L) [see Table 2; Appendix 4 sections 6.1 & 9.3].

### DISCUSSION

This is the first meta-analysis attempting to define expected ranges of 24-hour urine biochemistry values in adult non-kidney-stone formers. Our analyses were constructed to stratify results by age, sex and ancestry. We also performed sensitivity analyses based on study design, collection completeness, risk of bias and country (used as a proxy for assay type), and applied a pre-specified decision framework [see **Figure 1**] to select the recommended analysis per component. Six of the derived expected ranges (pH, oxalate, magnesium, sulphate, urate, creatinine) were broadly concordant with previously published reference ranges [see **Table 2**], thus providing an evidence base for these specific urine components.

As with any review, we were limited by the data available, and this forms the major limitation of our study. However, we performed extensive statistical analyses for heterogeneity and publication bias, along with sensitivity and bias analyses. These allow us to give an accurate GRADE rating for each component, aiding the robustness and interpretability of our results.

Ancestry-stratified analyses were possible for six studies only; all per-ancestry sub-analyses were underpowered, with most strata containing one to three studies. Ancestry-related differences are therefore not reliably characterised in this work and represent a clear gap for future research. For citrate and potassium specifically, our analyses detected significant between-assay/country differences (minimum Wald p<0.001 and p=0.010 respectively); standardisation of laboratory methods for these two analytes would materially reduce this between-source variability.

We are careful to describe our findings as expected ranges, not reference ranges. This is because the distribution of a urine component in a healthy population does not necessarily infer risk. For example, the reference range for 24-hour urine volume is defined by the EAU as >0.5L^1^, whilst Litholink (the most widely utilised laboratory in the US) defines it as >2.5L^17^. This demonstrates the difference between diagnostic and therapeutic thresholds. Lower 24-hour urine volumes increase the risk of stone formation^6,10^, and may explain why an individual has formed one. However, the goal of kidney stone management is not just diagnosis but also recurrence prevention. Borghi et al.^10^ demonstrated by RCT that higher urine volumes (>2.5L) significantly reduced subsequent recurrences in calcium stone formers, hence the rationale behind Litholink’s reference range. This is reflected in our data, with the upper limit being 2.5L/24h in a non-stone forming population, again reinforcing the point that the 24-hour urine volume stone formers should aim for to prevent kidney stone disease recurrence, is greater than the upper limit of the non-stone-forming distribution observed in our data^10,17^.

These data highlight the lack of reference range standardisation for 24-hour urine tests. This is partly because reference ranges have been derived through different (diagnostic vs therapeutic) approaches and from varying populations. For 24-hour urine values, both approaches have been utilised in existing ranges resulting in differing reference ranges [see **table 2**]. This demonstrates the uncertainty in whether these are derived from unaffected patient populations or treatment thresholds. In our data, these components were graded Low or Very Low for certainty of evidence. This suggests that the current 24-hour urine reference ranges, if based on similar data, are of similarly limited certainty. These are particularly important components given their pathophysiological relevance to kidney stone disease. For example, there are clear data that suggest increasing urine calcium increases the risk of forming kidney stones^6^, with a continuous, dose-response relationship that does not respect a single cut-off threshold^43^. We acknowledge that no single component determines kidney stone risk in isolation, and that any threshold-based approach trades sensitivity for specificity. In other words, if a high threshold is set, we risk not including people where that particular component is driving their disease and therefore not treating them effectively. For example, for 24-hour urine calcium, given that relative reduction, rather than absolute, is associated with decreased risk of recurrence^44^, there is an argument that the upper limit should be set at a lower level to capture all those at risk of recurrence.

Several factors contribute to the high between-study heterogeneity observed across 24-hour urine components. Differences in collection completeness, assays, background diet, hydration status, and inclusion criteria for ‘healthy’ all plausibly contribute and are not fully captured in the studies we have examined.

Future studies should aim to replicate these results to improve robustness, and whilst our framework adjusts for the dominant moderators, the residual unexplained variability across most components warrants harmonisation of collection and assay standards in future primary studies. Until such harmonisation is achieved, future studies including 24-hour urine results should report assay type, collection completeness and demographic strata consistently, enabling future meta-analyses to model these moderators directly.

Twenty-four-hour urine reference range derivation for patients with kidney stones is complex. As we stated earlier, the other method of determining reference ranges is by identifying therapeutic thresholds. Further work is needed to collate the evidence for these thresholds, which can then be integrated and contextualised with the data detailed in this paper to derive reference ranges for 24-hour urine collections in patients with kidney stones.

## CONCLUSION

This study establishes expected 24-hour urine values in a non-stone-forming population and provides empirical support for published reference ranges for several common urine analytes. Following a pre-specified decision framework, the expected ranges we derived are concordant with published reference ranges for pH, magnesium, sulphate, oxalate, urate and creatinine, providing an evidence base for those components. Further work is needed to delineate treatment thresholds for the 24-hour urine components detailed here, to allow for reference range determination.

## Data Availability

All data produced are available online at https://github.com/rg2u17/24_hour_urine and https://endourology.shinyapps.io/24_hr_ranges/

https://github.com/rg2u17/24_hour_urine

## Conflicts of Interest

RG, BS and LT are members of the EAU guidelines panel on Urolithiasis. JAS has previously been a consultant for the EAU guidelines panel on Urolithiasis.

## Author Contributions

Conceptualization: RG, OK; Data Curation: JH, AG, PR, KN, RG; Formal Analysis: RG; Writing — Original Draft: KN, AG, RG, OK; Writing — Review and Editing: JH, LT, SY, PC, BKS, DGF, PMF, GC, SH, JAS, ON, RG

## Acknowledgements

We are grateful to Dr Sara Best and Dr John Asplin for their valuable insights.

## Financial Disclosures

RG is funded by NIHR as Doctoral Fellowship (NIHR304667). KN would like to thank the Barbour Foundation for funding her PhD Studentship. This study was funded, in part, by an Engineering and Physical Sciences Research Council grant (EP/Y014596/1) and by Kidney Research UK, through a donation made by the Thompson Family Trust in memory of David Thompson (NEPH_ST_001_20240827). JAS is funded by LifeArc, Medical Research Council (MR/Y007808/1), Kidney Research UK (Paed_RP_001_20180925, RP_007_20210729), the Northern Counties Kidney Research Fund (20/01) and the European Union’s Horizon Europe research and innovation programme and from UKRI under grant agreement No: 101080717 (TheRaCil).

