## Supplementary material for "Investigating 24-Hour Urine Expected Ranges of Non-Kidney Stone Formers: an EAU Endourology Research Group Systematic Review and Meta-Analysis": Tables and Figures

#### Table 1

| **Meta-Analysis Type** | **Patients, n** | **95% CI** | **Q statistic** | **Q p-value** | **GRADE Rating** |
| --- | --- | --- | --- | --- | --- |
| **Ammonium (mmol/24hr)** | | | | | |
| Male | 535 | 28 – 46 | 267.48 | <0.001 | Moderate ⊕⊕⊕◯ |
| Female | 3857 | 20 – 40 | 1303.78 | <0.001 | Moderate ⊕⊕⊕◯ |
| **Calcium (mmol/24hr)** | | | | | |
| Meta-Regression | 5418 | 1.9 – 6.2 | 2744.79 | <0.001 | Low ⊕⊕◯◯ |
| **Chloride (mmol/24hr)** | | | | | |
| Meta-Regression | 3173 | 84 – 218 | 553.32 | <0.001 | Moderate ⊕⊕⊕◯ |
| **Citrate (mmol/24hr)*** | | | | | |
| Overall | 9458 | 1.2 – 4.5 | 16945.00 | <0.001 | Very Low ⊕◯◯◯ |
| **Creatinine (mmol/24hr)** | | | | | |
| Male | 760 | 12.2 – 16.3 | 196.17 | <0.001 | Very Low ⊕◯◯◯ |
| Female | 4217 | 7.2 – 12.6 | 1625.32 | <0.001 | Low ⊕⊕◯◯ |
| **Magnesium (mmol/24hr)** | | | | | |
| Meta-Regression | 4724 | 3.1 – 4.8 | 306.00 | <0.001 | Moderate ⊕⊕⊕◯ |
| **Oxalate (mmol/24hr)** | | | | | |
| Male | 860 | 0.2 – 0.6 | 3373.74 | <0.001 | Very Low ⊕◯◯◯ |
| Female | 4177 | 0.2 – 0.4 | 439.70 | <0.001 | Very Low ⊕◯◯◯ |
| **pH** | | | | | |
| Meta-Regression | 4379 | 5.6 – 6.3 | 324.21 | <0.001 | Moderate ⊕⊕⊕◯ |
| **Phosphate (mmol/24hr)** | | | | | |
| Meta-Regression | 3974 | 19 – 26 | 183.33 | <0.001 | Low ⊕⊕◯◯ |
| **Potassium (mmol/24hr)*** | | | | | |
| Overall | 9418 | 28 – 77 | 7644.37 | <0.001 | Very Low ⊕◯◯◯ |
| **Sodium (mmol/24hr)** | | | | | |
| Meta-Regression | 3945 | 98 – 253 | 857.39 | <0.001 | Moderate ⊕⊕⊕◯ |
| **Sulphate (mmol/24hr)** | | | | | |
| Male | 642 | 17 – 26 | 588.12 | <0.001 | Moderate ⊕⊕⊕◯ |
| Female | 3794 | 7 – 30 | 7067.01 | <0.001 | Moderate ⊕⊕⊕◯ |
| **Urate (mmol/24hr)** | | | | | |
| Male | 860 | 2 – 5.1 | 1118.46 | <0.001 | Very Low ⊕◯◯◯ |
| Female | 4177 | 2.3 – 3.5 | 1324.26 | <0.001 | Very Low ⊕◯◯◯ |
| **Urea (mmol/24hr)** | | | | | |
| Overall | 3876 | 234 – 445 | 3061.65 | <0.001 | Low ⊕⊕◯◯ |
| **Volume (L/24hr)** | | | | | |
| Overall | 9144 | 1 – 2.5 | 9158.78 | <0.001 | High ⊕⊕⊕⊕ |

**Table 1.** Summary of most robust meta-analysis results. *95% CI: Expected range presuming component is normally distributed (save for volume where we assumed a right skew), Q p-value = Cochran's Q heterogeneity test; RoB = Risk of Bias. *=varies significantly by assay proxy*

#### Table 2

| **Sex** | **Existing Reference Ranges** | | | | | **Expected Range**  **in this Study** | **Expected Range**  **GRADE Rating** |
| --- | --- | --- | --- | --- | --- | --- | --- |
|  | EAU | NHS | Mayo Clinic | Litholink | NDH |  |  |
| **Ammonium (Mmol / 24hrs)** | | | | | | | |
| **Overall** | < 50 | - | 15 - 56 | 18 - 56 | - | **-** | - |
| **Men** | - | - | - | - | - | **28 – 46** | Moderate ⊕⊕⊕◯ |
| **Women** | - | - | - | - | - | **20 – 40** | Moderate ⊕⊕⊕◯ |
| **Calcium (Mmol / 24hrs)** | | | | | | | |
| **Overall** | < 8 | 2.5 - 7.5 | < 5 | - | 2 - 7.4 | **1.9 – 6.2** | Low ⊕⊕◯◯ |
| **Men** | - | - | - | 1 - 6.2 | - | **-** | - |
| **Women** | - | - | - | 0.7 - 5 | - | **-** | - |
| **Chloride (Mmol / 24hrs)** | | | | | | | |
| **Overall** | - | 110 - 250 | 34 - 286 | 50 - 120 | 110 - 250 | **84 – 218** | Moderate ⊕⊕⊕◯ |
| **Citrate (Mmol / 24hrs)*** | | | | | | | |
| **Overall** | - | 0.6 - 4.8 | 1.3 - 6.2 | - | - | **1.2 – 4.5** | Very Low ⊕◯◯◯ |
| **Men** | > 1.7 | - | - | > 2.1 | - | **-** | - |
| **Women** | > 1.9 | - | - | > 2.8 | - | **-** | - |
| **Creatinine (Mmol / 24hrs)** | | | | | | | |
| **Men** | 13 - 18 | 9 - 19 | 8.2 - 26.1 | - | 7.1 - 17.7 | **12.2 – 16.3** | Very Low ⊕◯◯◯ |
| **Women** | 7 - 13 | 6 - 13 | 5.3 - 15.8 | - | 5.3 - 15.9 | **7.2 – 12.6** | Low ⊕⊕◯◯ |
| **Magnesium (Mmol / 24hrs)** | | | | | | | |
| **Overall** | > 3 | 2.4 - 6.5 | 2.1 - 4.3 | 2.6 - 7.1 | - | **3.1 – 4.8** | Moderate ⊕⊕⊕◯ |
| **Oxalate (Mmol / 24hrs)** | | | | | | | |
| **Overall** | < 0.5 | 0.1 - 0.5 | 0.1 - 0.5 | - | - | **-** | - |
| **Men** | - | - | - | < 0.5 | - | **0.2 – 0.6** | Very Low ⊕◯◯◯ |
| **Women** | - | - | - | < 0.4 | - | **0.2 – 0.4** | Very Low ⊕◯◯◯ |
| **pH** | | | | | | | |
| **Overall** | 5.5 - 7 | - | 4.5 - 8 | 5.7 - 6.3 | 4.5 - 8 | **5.6 – 6.3** | Moderate ⊕⊕⊕◯ |
| **Phosphate (Mmol / 24hrs)** | | | | | | | |
| **Overall** | < 35 | 15 - 50 | 7.3 - 58 | 19.4 - 38.7 | 12.9 - 42 | **19 – 26** | Low ⊕⊕◯◯ |
| **Potassium (Mmol / 24hrs)*** | | | | | | | |
| **Overall** | - | 25 - 125 | 16 - 105 | 40 - 100 | 25 - 125 | **28 – 77** | Very Low ⊕◯◯◯ |
| **Sodium (Mmol / 24hrs)** | | | | | | | |
| **Overall** | - | 40 - 250 | 22 - 328 | 50 - 120 | 40 - 220 | **98 – 253** | Moderate ⊕⊕⊕◯ |
| **Sulphate (Mmol / 24hrs)** | | | | | | | |
| **Overall** | - | - | 7 - 47 | 12.5 - 28 | - | **-** | - |
| **Men** | - | - | - | - | - | **17 – 26** | Moderate ⊕⊕⊕◯ |
| **Women** | - | - | - | - | - | **7 – 30** | Moderate ⊕⊕⊕◯ |
| **Urate (Mmol / 24hrs)** | | | | | | | |
| **Overall** | - | 1.5 - 4.5 | - | - | 1.5 - 4.5 | **-** | - |
| **Men** | < 5 | - | 1.2 - 5.9 | 2.7 - 4.8 | - | **2 – 5.1** | Very Low ⊕◯◯◯ |
| **Women** | < 4 | - | 1.5 - 4.5 | 2.2 - 4.5 | - | **2.3 – 3.5** | Very Low ⊕◯◯◯ |
| **Urea (Mmol / 24hrs)** | | | | | | | |
| **Overall** | - | 250 - 570 | 124.9 - 749.7 | 249.9 - 535.5 | 250 - 4.5 | **234 – 445** | Low ⊕⊕◯◯ |
| **Volume (L / 24hrs)** | | | | | | | |
| **Overall** | > 0.5 | < 3 | - | > 2.5 | - | **1 – 2.5** | High ⊕⊕⊕⊕ |

**Table 2.** Comparison of meta-analysed expected ranges to existing reference ranges. **=varies significantly by assay proxy*

### Figures

#### Figure 1

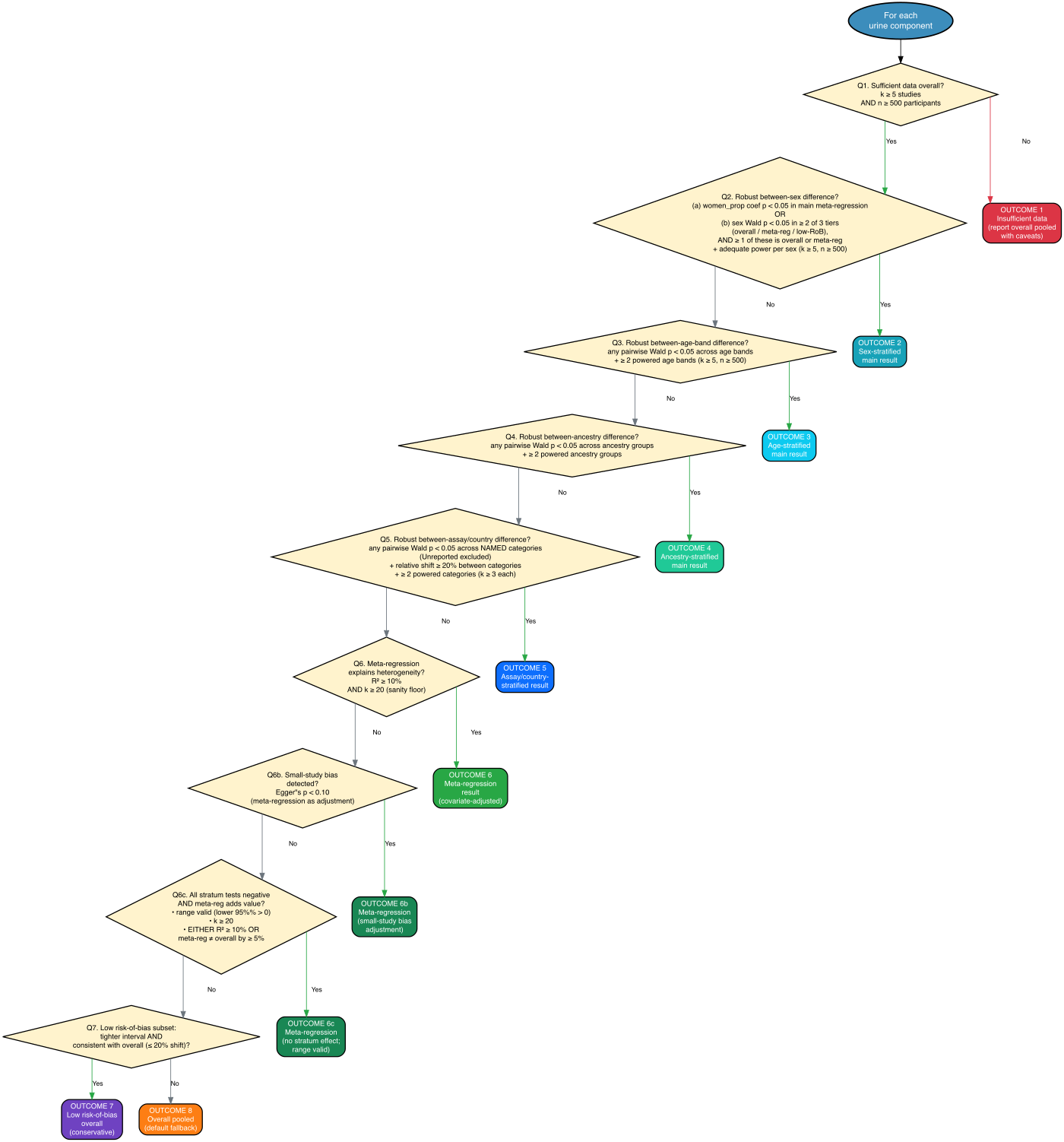

**Figure 1.** Decision framework used to select the recommended analysis per 24-hour urine component. Each component was evaluated against eight ordered branches (Q1 data sufficiency → Q2 sex → Q3 age → Q4 ancestry → Q5 assay/country → Q6 meta-regression effective → Q6b meta-regression with Egger's override → Q6c meta-regression with stratum-effect override → Q7 low risk-of-bias → Q8 overall pooled). The first branch whose evidence criteria were met determined the recommended analysis. Thresholds: k≥5, n≥500; R²≥10% with k≥20 for Q6; Wald p<0.05; Egger's p<0.10; consistency ≤20% (Q/df ratio ≤150%).

#### Figure 2

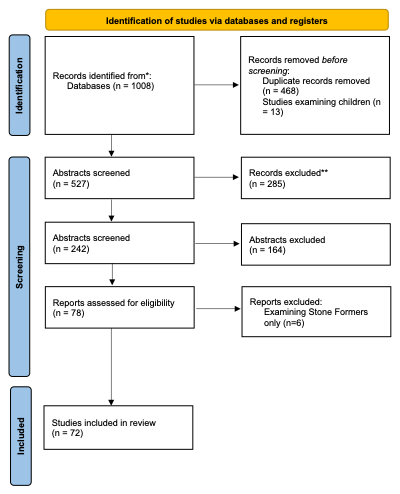

**Figure 2**. PRISMA flow diagram.

### Supplementary Tables

#### Supplementary Table 1

| **Section and Topic** | **Item #** | **Checklist item** | **Location where item is reported** |
| --- | --- | --- | --- |
| **TITLE** | | |  |
| Title | 1 | Identify the report as a systematic review. | See Title |
| **ABSTRACT** | | |  |
| Abstract | 2 | See the PRISMA 2020 for Abstracts checklist. | See Abstract |
| **INTRODUCTION** | | |  |
| Rationale | 3 | Describe the rationale for the review in the context of existing knowledge. | See Introduction |
| Objectives | 4 | Provide an explicit statement of the objective(s) or question(s) the review addresses. | See Introduction / PICO in methods |
| **METHODS** | | |  |
| Eligibility criteria | 5 | Specify the inclusion and exclusion criteria for the review and how studies were grouped for the syntheses. | See Methods / PROSPERO |
| Information sources | 6 | Specify all databases, registers, websites, organisations, reference lists and other sources searched or consulted to identify studies. Specify the date when each source was last searched or consulted. | See Methods / PROSPERO |
| Search strategy | 7 | Present the full search strategies for all databases, registers and websites, including any filters and limits used. | See Methods / PROSPERO |
| Selection process | 8 | Specify the methods used to decide whether a study met the inclusion criteria of the review, including how many reviewers screened each record and each report retrieved, whether they worked independently, and if applicable, details of automation tools used in the process. | See Methods / PROSPERO |
| Data collection process | 9 | Specify the methods used to collect data from reports, including how many reviewers collected data from each report, whether they worked independently, any processes for obtaining or confirming data from study investigators, and if applicable, details of automation tools used in the process. | See Methods / PROSPERO |
| Data items | 10a | List and define all outcomes for which data were sought. Specify whether all results that were compatible with each outcome domain in each study were sought (e.g. for all measures, time points, analyses), and if not, the methods used to decide which results to collect. | See Methods / PROSPERO |
|  | 10b | List and define all other variables for which data were sought (e.g. participant and intervention characteristics, funding sources). Describe any assumptions made about any missing or unclear information. | See Methods / PROSPERO |
| Study risk of bias assessment | 11 | Specify the methods used to assess risk of bias in the included studies, including details of the tool(s) used, how many reviewers assessed each study and whether they worked independently, and if applicable, details of automation tools used in the process. | See Methods / PROSPERO |
| Effect measures | 12 | Specify for each outcome the effect measure(s) (e.g. risk ratio, mean difference) used in the synthesis or presentation of results. | See Methods / PROSPERO |
| Synthesis methods | 13a | Describe the processes used to decide which studies were eligible for each synthesis (e.g. tabulating the study intervention characteristics and comparing against the planned groups for each synthesis (item #5)). | See Methods / PROSPERO |
|  | 13b | Describe any methods required to prepare the data for presentation or synthesis, such as handling of missing summary statistics, or data conversions. | See Methods / PROSPERO |
|  | 13c | Describe any methods used to tabulate or visually display results of individual studies and syntheses. | See Methods / PROSPERO |
|  | 13d | Describe any methods used to synthesize results and provide a rationale for the choice(s). If meta-analysis was performed, describe the model(s), method(s) to identify the presence and extent of statistical heterogeneity, and software package(s) used. | See Methods / PROSPERO / appendix 2 |
|  | 13e | Describe any methods used to explore possible causes of heterogeneity among study results (e.g. subgroup analysis, meta-regression). | See Methods / PROSPERO |
|  | 13f | Describe any sensitivity analyses conducted to assess robustness of the synthesized results. | See Methods / PROSPERO |
| Reporting bias assessment | 14 | Describe any methods used to assess risk of bias due to missing results in a synthesis (arising from reporting biases). | See Methods / PROSPERO |
| Certainty assessment | 15 | Describe any methods used to assess certainty (or confidence) in the body of evidence for an outcome. | See Methods / PROSPERO |
| **RESULTS** | | |  |
| Study selection | 16a | Describe the results of the search and selection process, from the number of records identified in the search to the number of studies included in the review, ideally using a flow diagram. | See results |
|  | 16b | Cite studies that might appear to meet the inclusion criteria, but which were excluded, and explain why they were excluded. | See results |
| Study characteristics | 17 | Cite each included study and present its characteristics. | See results |
| Risk of bias in studies | 18 | Present assessments of risk of bias for each included study. | See results |
| Results of individual studies | 19 | For all outcomes, present, for each study: (a) summary statistics for each group (where appropriate) and (b) an effect estimate and its precision (e.g. confidence/credible interval), ideally using structured tables or plots. | See results |
| Results of syntheses | 20a | For each synthesis, briefly summarise the characteristics and risk of bias among contributing studies. | See results |
|  | 20b | Present results of all statistical syntheses conducted. If meta-analysis was done, present for each the summary estimate and its precision (e.g. confidence/credible interval) and measures of statistical heterogeneity. If comparing groups, describe the direction of the effect. | See results |
|  | 20c | Present results of all investigations of possible causes of heterogeneity among study results. | See results |
|  | 20d | Present results of all sensitivity analyses conducted to assess the robustness of the synthesized results. | See results |
| Reporting biases | 21 | Present assessments of risk of bias due to missing results (arising from reporting biases) for each synthesis assessed. | See results |
| Certainty of evidence | 22 | Present assessments of certainty (or confidence) in the body of evidence for each outcome assessed. | See results |
| **DISCUSSION** | | |  |
| Discussion | 23a | Provide a general interpretation of the results in the context of other evidence. | See Discussion |
|  | 23b | Discuss any limitations of the evidence included in the review. | See Discussion |
|  | 23c | Discuss any limitations of the review processes used. | See Discussion |
|  | 23d | Discuss implications of the results for practice, policy, and future research. | See Discussion |
| **OTHER INFORMATION** | | |  |
| Registration and protocol | 24a | Provide registration information for the review, including register name and registration number, or state that the review was not registered. | See Methods / PROSPERO |
|  | 24b | Indicate where the review protocol can be accessed, or state that a protocol was not prepared. | See Methods / PROSPERO / Appendix 3 |
|  | 24c | Describe and explain any amendments to information provided at registration or in the protocol. | N/A |
| Support | 25 | Describe sources of financial or non-financial support for the review, and the role of the funders or sponsors in the review. | Supporting information |
| Competing interests | 26 | Declare any competing interests of review authors. | Supporting information |
| Availability of data, code and other materials | 27 | Report which of the following are publicly available and where they can be found: template data collection forms; data extracted from included studies; data used for all analyses; analytic code; any other materials used in the review. | Code freely available as appendix 2. Data clearly detailed in Results |

#### Supplementary Table 2

| Study | Age ± SD (years) | Total, n | Men,n | Women, n | Race | Age Band | Study Type |
| --- | --- | --- | --- | --- | --- | --- | --- |
| Goldschmied, 1975^1^ | - | 55 | 28 | 27 | mixed | 18+ | Observational Cohort |
| Welshman, 1975^2^ | - | 89 | 89 | 0 | mixed | 18+ | Observational Cohort |
| Welshman, 1976^3^ | - | 158 | 73 | 85 | mixed | 18+ | Observational Cohort |
| Marya, 1979^4^ | - | 50 | NA | NA | mixed | 18+ | Observational Cohort |
| Stern, 1980^5^ | - | 9 | NA | NA | mixed | 18+ | Observational Cohort |
| Hesse et al. 1986^6^ | - | 25 | 0 | 25 | mixed | 20-30 | Observational Cohort |
|  | - | 25 | 0 | 25 | mixed | 30-40 | Observational Cohort |
|  | - | 25 | 0 | 25 | mixed | 40-50 | Observational Cohort |
|  | - | 25 | 0 | 25 | mixed | 50-60 | Observational Cohort |
|  | - | 25 | 0 | 25 | mixed | 60+ | Observational Cohort |
|  | - | 25 | 25 | 0 | mixed | 20-30 | Observational Cohort |
|  | - | 25 | 25 | 0 | mixed | 30-40 | Observational Cohort |
|  | - | 25 | 25 | 0 | mixed | 40-50 | Observational Cohort |
|  | - | 25 | 25 | 0 | mixed | 50-60 | Observational Cohort |
|  | - | 25 | 25 | 0 | white | 60+ | Observational Cohort |
| Bingham, 1988^7^ | - | 8 | 5 | 3 | mixed | 18+ | Observational Cohort |
| Michelacci, 1989^8^ | - | 23 | 11 | 12 | mixed | 18+ | Observational Cohort |
| Bataille, 1991^9^ | 35±5 | 62 | 41 | 21 | mixed | 30-40 | Observational Cohort |
| Wabner & Pak 1992^10^ | 38±8.6 | 12 | 0 | 12 | mixed | 18+ | RCT |
| Erwin et al. 1994^11^ | 40±1.5 | 22 | 16 | 6 | mixed | 40-50 | Observational Cohort |
| Buchholz et al. 1996^12^ | 34±10.8 | 37 | 37 | 0 | mixed | 18+ | Observational Cohort |
| Christie et al. 1996^13^ | 39±3 | 17 | 7 | 10 | mixed | 30-40 | Observational Cohort |
| Lemann et al. 1996^14^ | 43.9±10.2 | 94 | 44 | 50 | mixed | 18+ | Observational Cohort |
| Rodgers, 1997^15^ | - | 40 | 20 | 20 | mixed | 18+ | Observational Cohort |
| Hess et al. 1999^16^ | 39±1.3 | 107 | 107 | 0 | mixed | 30-40 | Observational Cohort |
| Matin and Streem et al. 2000^17^ | - | 26 | 9 | 17 | mixed | 18+ | Observational Cohort |
| Holmes et al. 2001^18^ | 29±4 | 12 | 6 | 6 | mixed | 20-30 | Observational Cohort |
| Miyake et al. 2001^19^ | 31.5±4.4 | 11 | 11 | 0 | mixed | 30-40 | Observational Cohort |
| Terris et al. 2001^20^ | 30.2±NA | 5 | 3 | 2 | mixed | 20-30 | Observational Cohort |
| Rodgers and Lewandowski, 2002^21^ | - | 10 | 10 | 0 | black | 18+ | RCT |
|  | - | 10 | 10 | 0 | white | 18+ | RCT |
| Kinder et al. 2002^22^ | - | 111 | 0 | 111 | mixed | 18+ | Observational Cohort |
|  | - | 66 | 66 | 0 | mixed | 18+ | Observational Cohort |
| Allie and Rodgers, 2003^23^ | - | 8 | 8 | 0 | mixed | 20-30 | RCT |
| Baxmann et al. 2003^24^ | 37±16 | 20 | 8 | 12 | mixed | 18+ | RCT |
| Huang et al. 2003^25^ | 50.8±1.8 | 32 | 27 | 5 | mixed | 50-60 | Observational Cohort |
| Kuo et al. 2003^26^ | - | 4 | NA | NA | mixed | 18+ | Observational Cohort |
| Ogawa et al. 2003^27^ | 54.1±16.2 | 22 | 6 | 16 | mixed | 18+ | Observational Cohort |
| Traxer et al. 2003^28^ | 38.1±11.3 | 12 | 6 | 6 | mixed | 18+ | RCT |
| Abate et al. 2004^29^ | 31±11 | 55 | 33 | 22 | mixed | 18+ | Observational Cohort |
| Chai and Liebman 2004^30^ | - | 9 | 5 | 4 | mixed | 18+ | RCT |
| Stitchantrakul et al. 2004^31^ | 22.7±1.9 | 38 | 38 | 0 | mixed | 20-30 | RCT |
| Defoor et al. 2005^32^ | - | 168 | 97 | 71 | mixed | 18+ | Observational Cohort |
| Gettman et al. 2005^33^ | 34.8±9.6 | 12 | 6 | 6 | mixed | 18+ | Observational Cohort |
| Shah et al. 2005^34^ | 30±NA | 101 | 54 | 47 | mixed | 18+ | Observational Cohort |
|  | 30±5 | 61 | 12 | 49 | mixed | 30-40 | Observational Cohort |
| Cameron et al. 2006^35^ | 49±8 | 59 | 24 | 35 | mixed | 18+ | Observational Cohort |
|  | 52±8 | 24 | 10 | 14 | mixed | 18+ | Observational Cohort |
| Matsumoto et al. 2006^36^ | - | 10 | 5 | 5 | mixed | 18+ | RCT |
| Pais et al. 2007^37^ | 43±10 | 61 | 12 | 49 | mixed | 18+ | Observational Cohort |
| Stitchantrakul et al. 2007^38^ | 42.6±1.9 | 34 | 14 | 20 | mixed | 40-50 | Observational Cohort |
| Ciacci et al. 2008^39^ | 35.8±NA | 45 | 9 | 36 | mixed | 18+ | Observational Cohort |
| Baxmann et al. 2008^40^ | 36.6±13.6 | 170 | 78 | 92 | mixed | 18+ | Observational Cohort |
| Rodgers et al. 2009^41^ | 50±10 | 18 | 16 | 2 | white | 18+ | Observational Cohort |
|  | 50±10 | 17 | 10 | 7 | black | 18+ | Observational Cohort |
|  | 50±10 | 37 | 19 | 18 | white | 18+ | Observational Cohort |
| Sweeney et al. 2009^42^ | 36.3±10.6 | 14 | NA | NA | mixed | 18+ | RCT |
| Maalouf et al. 2010^43^ | 52±10 | 16 | 8 | 8 | mixed | 18+ | Observational Cohort |
|  | 54±11 | 9 | 5 | 4 | mixed | 18+ | Observational Cohort |
| Semins et al. 2010^44^ | - | 96 | 96 | 0 | mixed | 18+ | Observational Cohort |
|  | - | 72 | 0 | 72 | mixed | 18+ | Observational Cohort |
| Duffey et al. 2011^45^ | 46±10.4 | 51 | 16 | 35 | mixed | 18+ | Observational Cohort |
| Parvin et al. 2011^46^ | 38.36±6.907 | 109 | 109 | 0 | mixed | 18+ | Observational Cohort |
| Arrabal-Polo et al. 2012^47^ | 49.53±10.14 | 60 | NA | NA | mixed | 18+ | Observational Cohort |
| Goodarzi et al. 2012^48^ | 39.9±12.6 | 27 | 18 | 9 | mixed | 18+ | Observational Cohort |
| Herrel et al. 2012^49^ | 38.7±NA | 10 | NA | NA | mixed | 18+ | RCT |
| Hong et al. 2012^50^ | 50.2±10.8 | 30 | 12 | 18 | mixed | 18+ | Observational Cohort |
| Sorensen et al. 2012^51^ | 58±12 | 50 | 13 | 37 | mixed | 18+ | Observational Cohort |
| Nouvenne et al. 2013^52^ | 46±6 | 39 | 0 | 39 | mixed | 18+ | Observational Cohort |
| Elkoushy et al. 2014^53^ | - | 204 | 57 | 147 | mixed | 18+ | Observational Cohort |
| Lange et al. 2014^54^ | 25.3±2.7 | 15 | 8 | 7 | mixed | 18+ | RCT |
| Pigna et al. 2014^55^ | 52.1±13.6 | 21 | 21 | 0 | mixed | 18+ | Observational Cohort |
| Ma et al. 2014^56^ | 54.2±2.4 | 29 | NA | NA | mixed | 18+ | Observational Cohort |
| Doenyas-Barak et al. 2015^57^ | 48.7±12.5 | 22 | 22 | 0 | mixed | 18+ | Observational Cohort |
|  | 44.7±15.3 | 26 | 0 | 26 | mixed | 18+ | Observational Cohort |
| Grases et al. 2015^58^ | - | 75 | 42 | 33 | mixed | 18+ | Observational Cohort |
| Arrabal-Polo et al. 2015^59^ | 51.81±10.32 | 61 | 31 | 30 | mixed | 18+ | Observational Cohort |
| Perinpam et al. 2015^60^ | 65.4±9 | 709 | 293 | 416 | mixed | 18+ | Observational Cohort |
| Rodgers et al. 2015^61^ | - | 15 | 15 | 0 | mixed | 18+ | Observational Cohort |
| Rodríguez-Rodríguez et al. 2015^62^ | 35.7±11.2 | 418 | 196 | 222 | white | 18+ | Observational Cohort |
| Doizi et al. 2016^63^ | 57.6±9.7 | 9 (Obese) | 5 | 4 | mixed | 18+ | Observational Cohort |
|  | 54.3±9.5 | 12 (Lean) | 6 | 6 | mixed | 18+ | Observational Cohort |
| Arora et al. 2017^64^ | 49.4±14.9 | 20 | 10 | 10 | mixed | 18+ | Observational Cohort |
| Deng et al. 2017^65^ | 48.86±16.25 | 376 | 188 | 188 | mixed | 18+ | Observational Cohort |
|  | 53.96±19.28 | 24 | 12 | 12 | mixed | 18+ | Observational Cohort |
|  | 52.14±14.25 | 149 | 64 | 85 | mixed | 18+ | Observational Cohort |
|  | 50.03±12.84 | 35 | 14 | 21 | mixed | 18+ | Observational Cohort |
| Athanasatouet al. 2018^66^ | - | 163 | NA | NA | white | 18+ | Observational Cohort |
| Mai et al. 2019^67^ | 39.98±8.43 | 160 | 0 | 160 | mixed | 18+ | Observational Cohort |
| Baric et al. 2019^68^ | 21±2 | 53 | 25 | 28 | mixed | 20-30 | Observational Cohort |
| Thongprayoon et al. 2022^69^ | 45.33±18.59 | 440 | 215 | 225 | mixed | 18+ | Observational Cohort |
| Pieters et al. 2022^70^ | 54±12 | 287 | 112 | 175 | mixed | 18+ | Observational Cohort |
| Cao et al. 2023^71^ | 49.08±12.7 | 40 | 28 | 12 | east asian | 18+ | Observational Cohort |
| *HPFS^72,73^ | 48.67±0.52 | 6 | 6 | 0 | white | 40-50 | Observational Cohort |
| *NHS I ^72,73^ | 47.09±2.29 | 524 | 0 | 524 | white | 40-50 | Observational Cohort |
| *NHS II ^72,73^ | 46.86±2.03 | 29 | 0 | 29 | black | 40-50 | Observational Cohort |
| *HPFS^72,73^ | 55.63±3.02 | 79 | 79 | 0 | white | 50-60 | Observational Cohort |
| *NHS I ^72,73^ | 54.25±2.69 | 2409 | 0 | 2409 | white | 50-60 | Observational Cohort |
| *NHS II ^72,73^ | 54.92±2.65 | 131 | 0 | 131 | black | 50-60 | Observational Cohort |
| *HPFS^72,73^ | 64.06±2.79 | 215 | 215 | 0 | white | 60+ | Observational Cohort |
| *NHS I ^72,73^ | 64.59±2.95 | 231 | 0 | 231 | white | 60+ | Observational Cohort |
| *NHS II ^72,73^ | 64.86±3.02 | 93 | 0 | 93 | black | 60+ | Observational Cohort |
| *HPFS^72,73^ | 72.3±1.68 | 89 | 89 | 0 | white | 60+ | Observational Cohort |
| *NHS I ^72,73^ | 72.36±1.54 | 96 | 0 | 96 | white | 60+ | Observational Cohort |
| *NHS II ^72,73^ | 71.73±1.34 | 51 | 0 | 51 | black | 60+ | Observational Cohort |

**Supplementary Table 1.** Summary of Papers included in Meta-Analysis. * = summary data from Healthcare Professionals Follow-up study and the Nurses Health Study I/II was kindly provided by Professor G. Curhan amalgamating the two references studies.

#### Supplementary Table 3

| Study | Urine Component | | | | | | | | | | | | | | |
| --- | --- | --- | --- | --- | --- | --- | --- | --- | --- | --- | --- | --- | --- | --- | --- |
|  | Volume | pH | Creatinine | Calcium | Urate | Oxalate | Citrate | Phosphate | Sodium | Potassium | Magnesium | Ammonium | Chloride | Sulphate | Urea |
| Goldschmied, 1975^1^ | No | No | No | Yes | No | No | No | No | No | No | No | No | No | No | No |
| Welshman, 1975^2^ | No | No | No | Yes | No | No | No | No | Yes | Yes | Yes | No | No | No | No |
| Welshman, 1976^3^ | No | No | No | No | No | No | Yes | No | No | No | No | No | No | No | No |
| Marya, 1979^4^ | No | Yes | No | No | No | No | No | No | No | No | No | No | No | No | No |
| Stern, 1980^5^ | Yes | Yes | No | No | Yes | No | No | No | No | No | No | Yes | No | No | No |
| Hesse et al. 1986^6^ | Yes | Yes | Yes | Yes | Yes | Yes | Yes | No | Yes | Yes | Yes | Yes | Yes | Yes | No |
| Bingham, 1988^7^ | No | No | Yes | No | No | No | No | No | Yes | Yes | No | Yes | No | No | Yes |
| Michelacci, 1989^8^ | No | No | No | No | No | No | No | No | No | No | No | No | No | No | No |
| Bataille, 1991^9^ | No | No | No | No | No | No | No | No | No | No | No | No | No | No | No |
| Wabner & Pak 1992^10^ | Yes | Yes | No | Yes | Yes | Yes | Yes | No | Yes | Yes | Yes | Yes | No | No | No |
| Erwin et al. 1994^11^ | Yes | Yes | No | Yes | Yes | Yes | Yes | No | No | No | No | No | No | No | No |
| Buchholz et al. 1996^12^ | No | Yes | No | Yes | Yes | Yes | No | No | No | No | No | No | No | No | No |
| Christie et al. 1996^13^ | Yes | Yes | No | Yes | Yes | Yes | No | No | Yes | Yes | No | No | No | No | No |
| Lemann et al. 1996^14^ | Yes | Yes | No | Yes | No | Yes | Yes | Yes | Yes | Yes | Yes | Yes | No | Yes | No |
| Rodgers, 1997^15^ | Yes | No | No | Yes | No | Yes | Yes | No | No | No | Yes | No | No | No | No |
| Hess et al. 1999^16^ | Yes | Yes | No | Yes | Yes | Yes | Yes | Yes | Yes | No | Yes | No | No | Yes | Yes |
| Matin and Streem et al. 2000^17^ | Yes | No | No | Yes | Yes | Yes | Yes | No | Yes | No | No | No | No | No | No |
| Holmes et al. 2001^18^ | Yes | No | No | No | No | No | No | No | No | No | No | No | No | No | No |
| Miyake et al. 2001^19^ | Yes | Yes | Yes | Yes | Yes | Yes | Yes | No | No | No | Yes | No | No | No | No |
| Terris et al. 2001^20^ | No | No | No | No | No | No | No | No | No | No | Yes | No | No | No | No |
| Rodgers and Lewandowski, 2002^21^ | No | No | No | No | No | No | No | No | No | No | No | No | No | No | No |
| Kinder et al. 2002^22^ | No | Yes | No | Yes | No | No | No | No | No | No | No | No | No | No | No |
| Allie and Rodgers, 2003^23^ | No | Yes | Yes | Yes | Yes | Yes | Yes | Yes | Yes | Yes | Yes | No | Yes | No | No |
| Baxmann et al. 2003^24^ | Yes | No | Yes | Yes | Yes | Yes | Yes | No | Yes | Yes | Yes | No | No | No | Yes |
| Huang et al. 2003^25^ | No | Yes | No | No | No | No | No | No | No | No | Yes | No | No | No | No |
| Kuo et al. 2003^26^ | Yes | Yes | No | Yes | No | No | No | No | No | No | No | No | No | No | No |
| Ogawa et al. 2003^27^ | No | No | No | Yes | No | Yes | Yes | No | No | No | Yes | No | No | No | No |
| Traxer et al. 2003^28^ | Yes | Yes | Yes | Yes | Yes | No | Yes | Yes | Yes | Yes | Yes | Yes | Yes | Yes | No |
| Abate et al. 2004^29^ | Yes | Yes | Yes | No | No | No | Yes | No | Yes | Yes | No | Yes | Yes | Yes | No |
| Chai and Liebman 2004^30^ | No | No | No | No | No | No | No | No | No | No | No | No | No | No | No |
| Stitchantrakul et al. 2004^31^ | Yes | No | Yes | Yes | Yes | Yes | Yes | Yes | Yes | Yes | Yes | No | Yes | No | Yes |
| Defoor et al. 2005^32^ | No | Yes | No | No | No | No | No | No | No | No | No | No | No | No | No |
| Gettman et al. 2005^33^ | Yes | Yes | Yes | Yes | Yes | Yes | Yes | Yes | Yes | No | No | No | No | No | No |
| Shah et al. 2005^34^ | No | No | No | No | No | No | Yes | No | No | No | No | No | No | No | No |
| Cameron et al. 2006^35^ | Yes | Yes | No | No | Yes | No | Yes | No | No | Yes | No | Yes | No | Yes | No |
| Matsumoto et al. 2006^36^ | Yes | Yes | Yes | Yes | Yes | Yes | Yes | Yes | Yes | Yes | Yes | No | No | Yes | No |
| Pais et al. 2007^37^ | No | No | No | No | No | No | No | No | No | No | No | No | No | No | No |
| Stitchantrakul et al. 2007^38^ | Yes | No | Yes | Yes | Yes | Yes | Yes | Yes | Yes | Yes | Yes | No | Yes | No | Yes |
| Ciacci et al. 2008^39^ | No | No | No | No | Yes | No | Yes | No | No | No | No | No | No | No | No |
| Baxmann et al. 2008^40^ | No | No | Yes | No | No | No | No | No | No | No | No | No | No | No | No |
| Rodgers et al. 2009^41^ | No | Yes | No | Yes | Yes | Yes | Yes | No | Yes | Yes | Yes | No | No | No | No |
| Sweeney et al. 2009^42^ | Yes | Yes | Yes | Yes | Yes | Yes | Yes | No | Yes | Yes | Yes | Yes | Yes | Yes | No |
| Maalouf et al. 2010^43^ | Yes | Yes | Yes | Yes | Yes | Yes | Yes | Yes | Yes | Yes | Yes | Yes | Yes | Yes | No |
| Semins et al. 2010^44^ | No | No | No | No | No | No | No | No | No | No | No | No | No | No | No |
| Duffey et al. 2011^45^ | Yes | Yes | No | Yes | No | Yes | Yes | No | Yes | No | Yes | No | No | No | No |
| Parvin et al. 2011^46^ | Yes | Yes | Yes | Yes | Yes | Yes | Yes | Yes | Yes | Yes | Yes | No | Yes | No | Yes |
| Arrabal-Polo et al. 2012^47^ | Yes | Yes | No | Yes | Yes | Yes | No | No | Yes | Yes | Yes | No | No | No | No |
| Goodarzi et al. 2012^48^ | Yes | Yes | Yes | No | Yes | No | Yes | No | No | No | No | No | No | No | No |
| Herrel et al. 2012^49^ | Yes | Yes | Yes | Yes | Yes | Yes | Yes | Yes | Yes | Yes | Yes | No | No | No | No |
| Hong et al. 2012^50^ | Yes | Yes | No | Yes | Yes | Yes | Yes | Yes | Yes | Yes | Yes | No | No | No | No |
| Sorensen et al. 2012^51^ | Yes | Yes | No | Yes | Yes | Yes | Yes | Yes | Yes | No | No | No | No | No | No |
| Nouvenne et al. 2013^52^ | Yes | Yes | Yes | Yes | Yes | Yes | Yes | Yes | Yes | Yes | Yes | Yes | Yes | Yes | Yes |
| Elkoushy et al. 2014^53^ | Yes | No | No | Yes | No | Yes | Yes | Yes | No | No | No | No | No | No | No |
| Lange et al. 2014^54^ | Yes | No | Yes | Yes | Yes | Yes | Yes | Yes | Yes | Yes | Yes | No | No | No | No |
| Pigna et al. 2014^55^ | Yes | Yes | Yes | Yes | Yes | Yes | Yes | Yes | Yes | Yes | No | Yes | No | Yes | No |
| Ma et al. 2014^56^ | No | Yes | No | No | No | Yes | No | No | No | No | No | No | No | No | No |
| Doenyas-Barak et al. 2015^57^ | No | No | Yes | No | No | No | No | No | Yes | Yes | No | No | Yes | No | No |
| Grases et al. 2015^58^ | Yes | No | No | Yes | Yes | Yes | Yes | Yes | No | No | Yes | No | No | No | No |
| Arrabal-Polo et al. 2015^59^ | No | No | No | Yes | Yes | Yes | Yes | No | No | No | No | No | No | No | No |
| Perinpam et al. 2015^60^ | No | No | No | Yes | Yes | Yes | No | No | No | No | Yes | No | No | No | No |
| Rodgers et al. 2015^61^ | Yes | Yes | Yes | Yes | Yes | Yes | Yes | No | Yes | Yes | Yes | No | Yes | No | No |
| Rodríguez-Rodríguez et al. 2015^62^ | No | No | Yes | No | No | No | No | No | No | Yes | No | No | No | No | No |
| Doizi et al. 2016^63^ | Yes | Yes | No | Yes | Yes | Yes | Yes | No | Yes | Yes | Yes | Yes | Yes | No | No |
| Arora et al. 2017^64^ | Yes | Yes | No | Yes | Yes | Yes | Yes | Yes | Yes | Yes | Yes | Yes | Yes | Yes | No |
| Deng et al. 2017^65^ | Yes | Yes | Yes | Yes | Yes | Yes | Yes | Yes | Yes | Yes | Yes | No | Yes | No | No |
| Athanasatouet al. 2018^66^ | Yes | No | Yes | No | No | No | No | No | Yes | Yes | No | No | No | No | No |
| Mai et al. 2019^67^ | Yes | Yes | Yes | Yes | Yes | Yes | Yes | Yes | Yes | Yes | Yes | No | Yes | No | No |
| Baric et al. 2019^68^ | Yes | No | No | No | No | No | No | No | Yes | Yes | No | No | No | No | Yes |
| Thongprayoon et al. 2022^69^ | Yes | Yes | Yes | Yes | Yes | Yes | Yes | Yes | Yes | Yes | Yes | No | No | No | No |
| Pieters et al. 2022^70^ | No | No | Yes | No | No | No | No | No | No | No | No | No | No | No | No |
| Cao et al. 2023^71^ | No | No | No | Yes | No | No | No | No | Yes | No | Yes | No | No | No | No |
| *HPFS^72,73^ | Yes | Yes | Yes | Yes | Yes | Yes | Yes | Yes | Yes | Yes | Yes | Yes | No | Yes | No |
| *NHS I ^72,73^ | Yes | Yes | Yes | Yes | Yes | Yes | Yes | Yes | Yes | Yes | Yes | Yes | No | Yes | No |
| *NHS II ^72,73^ | Yes | Yes | Yes | Yes | Yes | Yes | Yes | Yes | Yes | Yes | Yes | Yes | No | Yes | No |

**Supplementary Table 2.** Summary of availability of data per urine component and paper.
