## Appendix 3 for "Investigating 24-Hour Urine Expected Ranges of Non-Kidney Stone Formers: an EAU Endourology Research Group Systematic Review and Meta-Analysis"

Evidence Search Results from your Knowledge & Library Service

1. **Preventive Pharmacological Therapy and Risk of Recurrent Urinary Stone Disease.**

**Item Type:** Journal Article

**Authors:** Arivoli, Kumaran;Valicevic, Autumn N.;Oerline, Mary K.;Hsi, Ryan S.;Patel, Sanjeevkumar R.;Hollingsworth, John M. and Shahinian, Vahakn B.

**Publication Date:** May 01 ,2024

**Journal:** Clinical Journal of the American Society of Nephrology: CJASN 19(5), pp. 565–572

**Abstract: BACKGROUND**: Urinary stone disease is a prevalent condition associated with a high recurrence risk. Preventive pharmacological therapy has been proposed to reduce recurrent stone episodes. However, limited evidence exists regarding its effectiveness, contributing to its underutilization by physicians. This study aimed to evaluate the association between preventive pharmacological therapy (thiazide diuretics, alkali therapy, and uric acid-lowering medications) and clinically significant urinary stone disease recurrence. **METHODS**: Using data from the Veterans Health Administration, adults with an index episode of urinary stone disease from 2012 through 2019 and at least one urinary abnormality (hypercalciuria, hypocitraturia, or hyperuricosuria) on 24-hour urine collection were included. The primary outcome was a composite variable representing recurrent stone events that resulted in emergency department visits, hospitalizations, or surgery for urinary stone disease. Cox proportional hazards regression was performed to estimate the association between preventive pharmacological therapy use and recurrent urinary stone disease while adjusting for relevant baseline patient characteristics. **RESULTS**: Among the cohort of patients with urinary abnormalities ( n =5637), treatment with preventive pharmacological therapy was associated with a significant 19% lower risk of recurrent urinary stone disease during the 12-36-month period after the initial urine collection (hazard ratio, 0.81; 95% confidence interval, 0.65 to 1.00; P = 0.0496). However, the effectiveness of preventive pharmacological therapy diminished over longer follow-up periods (12-48 and 12-60 months after the urine collection) and did not reach statistical significance. When examining specific urinary abnormalities, only alkali therapy for hypocitraturia was associated with a significant 26% lower recurrence risk within the 12-36-month timeframe (hazard ratio, 0.74; 95% confidence interval, 0.56 to 0.97; P = 0.03). **CONCLUSIONS**: When considering all urinary abnormalities together, this study demonstrates that the use of preventive pharmacological therapy is associated with a lower risk of clinically significant recurrent episodes of urinary stone disease in the 12-36 month timeframe after urine collection, although only the association with the use of alkali therapy for hypocitraturia was significant when individual abnormalities were examined. Copyright © 2024 by the American Society of Nephrology.

**URL:** <https://ovidsp.ovid.com/ovidweb.cgi?T=JS&CSC=Y&NEWS=N&PAGE=fulltext&D=med25&DO=10.2215%2fCJN.0000000000000428> <https://libkey.io/libraries/2592/openurl?genre=article&sid=OVID:medline&id=pmid:38345854&id=doi:10.2215%2FCJN.0000000000000428&issn=1555-9041&isbn=&volume=19&issue=5&spage=565&pages=565-572&date=2024&title=Clinical+Journal+of+The+American+Society+of+Nephrology%3A+CJASN&atitle=Preventive+Pharmacological+Therapy+and+Risk+of+Recurrent+Urinary+Stone+Disease.&aulast=Arivoli&pid=%3Cauthor%3EArivoli+K%3BValicevic+AN%3BOerline+MK%3BHsi+RS%3BPatel+SR%3BHollingsworth+JM%3BShahinian+VB%3C%2Fauthor%3E%3CAN%3E38345854%3C%2FAN%3E%3CDT%3EJournal+Article%3C%2FDT%3E>

2. **Predicting urinary stone recurrence: a joint model analysis of repeated 24-hour urine collections from the MSTONE database.**

**Item Type:** Journal Article

**Authors:** Kong, Zifang;Johnson, Brett A.;Maalouf, Naim M.;Nakada, Stephen Y.;Tracy, Chad R.;Steinberg, Ryan L.;Miller, Nicole;Antonelli, Jodi A.;Lotan, Yair;Pearle, Margaret S. and Liu, Yu-Lun

**Publication Date:** Nov 01 ,2024

**Journal:** Urolithiasis 52(1), pp. 156

**Abstract:** To address the limitations in existing urinary stone recurrence (USR) models, including failure to account for changes in 24-hour urine (24U) parameters over time and ignoring multiplicity of stone recurrences, we presented a novel statistical method to jointly model temporal trends in 24U parameters and multiple recurrent stone events. The MSTONE database spanning May 2001 to April 2015 was analyzed. A joint recurrent model was employed, combining a linear mixed-effects model for longitudinal 24U parameters and a recurrent event model with a dynamic first-order Autoregressive (AR(1)) structure. A mixture cure component was included to handle patient heterogeneity. Comparisons were made with existing methods, multivariable Cox regression and conditional Prentice-Williams-Peterson regression, both applied to established nomograms. Among 396 patients (median follow-up of 2.93 years; IQR, 1.53-4.36 years), 34.6% remained free of stone recurrence throughout the study period, 30.0% experienced a single recurrence, and 35.4% had multiple recurrences. The joint recurrent model with a mixture cure component identified significant associations between 24U parameters - including urine pH (adjusted HR = 1.991; 95% CI 1.490-2.660; p < 0.001), total volume (adjusted HR = 0.700; 95% CI 0.501-0.977; p = 0.036), potassium (adjusted HR = 0.983; 95% CI 0.974-0.991; p < 0.001), uric acid (adjusted HR = 1.528; 95% CI 1.105-2.113, p = 0.010), calcium (adjusted HR = 1.164; 95% CI 1.052-1.289; p = 0.003), and citrate (adjusted HR = 0.796; 95% CI 0.706-0.897; p < 0.001), and USR, achieving better predictive performance compared to existing methods. 24U parameters play an important role in prevention of USR, and therefore, patients with a history of stones are recommended to closely monitor for future recurrence by regularly conducting 24U tests. Copyright © 2024. The Author(s).

**URL:** <https://ovidsp.ovid.com/ovidweb.cgi?T=JS&CSC=Y&NEWS=N&PAGE=fulltext&D=medl&DO=10.1007%2fs00240-024-01653-5> <https://libkey.io/libraries/2592/openurl?genre=article&sid=OVID:medline&id=pmid:39485566&id=doi:10.1007%2Fs00240-024-01653-5&issn=2194-7228&isbn=&volume=52&issue=1&spage=156&pages=156&date=2024&title=Urolithiasis&atitle=Predicting+urinary+stone+recurrence%3A+a+joint+model+analysis+of+repeated+24-hour+urine+collections+from+the+MSTONE+database.&aulast=Kong&pid=%3Cauthor%3EKong+Z%3BJohnson+BA%3BMaalouf+NM%3BNakada+SY%3BTracy+CR%3BSteinberg+RL%3BMiller+N%3BAntonelli+JA%3BLotan+Y%3BPearle+MS%3BLiu+YL%3C%2Fauthor%3E%3CAN%3E39485566%3C%2FAN%3E%3CDT%3EJournal+Article%3C%2FDT%3E>

3. **Understanding the Barriers to Preventive Pharmacological Therapy Use in Older Patients With Urinary Stone Disease.**

**Item Type:** Journal Article

**Authors:** Krampe, Noah;Oerline, Mary K.;Hsi, Ryan S.;Crivelli, Joseph J.;Asplin, John R.;Shahinian, Vahakn B. and Hollingsworth, John M.

**Publication Date:** 2024

**Journal:** Urology Practice 11(1), pp. 218–225

**Abstract: INTRODUCTION**: Despite compelling clinical trial evidence and professional society guideline recommendations, prescription rates of preventative pharmacological therapy (PPT) for urinary stone disease are low. We sought to understand how patient- and clinician-level factors contribute to the decision to prescribe PPT after an index stone event. **METHODS**: We identified Medicare beneficiaries with urinary stone disease who had a 24-hour urine collection processed by a central laboratory. Among the subset with a urine chemistry abnormality (ie, hypercalciuria, hypocitraturia, hyperuricosuria, or low urine pH), we determined whether PPT was prescribed within 6 months of their collection. After assigning patients to the clinicians who ordered their collection, we fit multilevel models to determine how much of the variation in PPT prescription was attributable to patient vs clinician factors. **RESULTS**: Of the 11,563 patients meeting inclusion criteria, 33.6% were prescribed PPT. There was nearly sevenfold variation between the treating clinician with the lowest prescription rate (11%) and the one with the highest (75%). Nineteen percent of this variation was attributable to clinician factors. After accounting for measured patient differences and clinician volume, patients had twice the odds of being prescribed PPT if they were treated by a nephrologist (odds ratio [OR], 2.15; 95% CI, 1.79-2.57) or a primary care physician (OR, 1.78; 95% CI, 1.22-2.58) compared to being treated by a urologist. **CONCLUSIONS**: These findings suggest that the type of clinician whom a patient sees for his stone care determines, to a large extent, whether PPT will be prescribed.

**URL:** <https://ovidsp.ovid.com/ovidweb.cgi?T=JS&CSC=Y&NEWS=N&PAGE=fulltext&D=med25&DO=10.1097%2fUPJ.0000000000000480> <https://libkey.io/libraries/2592/openurl?genre=article&sid=OVID:medline&id=pmid:37903744&id=doi:10.1097%2FUPJ.0000000000000480&issn=2352-0779&isbn=&volume=11&issue=1&spage=218&pages=218-225&date=2024&title=Urology+Practice&atitle=Understanding+the+Barriers+to+Preventive+Pharmacological+Therapy+Use+in+Older+Patients+With+Urinary+Stone+Disease.&aulast=Krampe&pid=%3Cauthor%3EKrampe+N%3BOerline+MK%3BHsi+RS%3BCrivelli+JJ%3BAsplin+JR%3BShahinian+VB%3BHollingsworth+JM%3C%2Fauthor%3E%3CAN%3E37903744%3C%2FAN%3E%3CDT%3EJournal+Article%3C%2FDT%3E>

4. **Do We Really Need Stone Analysis and 24-Hour Urine Testing? No.**

**Item Type:** Journal Article

**Authors:** Modai, Jonathan and Scotland, Kymora B.

**Publication Date:** Se ,2024

**Journal:** Journal of Endourology 38(9), pp. 898–900

**URL:** <https://ovidsp.ovid.com/ovidweb.cgi?T=JS&CSC=Y&NEWS=N&PAGE=fulltext&D=medl&DO=10.1089%2fend.2024.0270> <https://libkey.io/libraries/2592/openurl?genre=article&sid=OVID:medline&id=pmid:38874954&id=doi:10.1089%2Fend.2024.0270&issn=0892-7790&isbn=&volume=38&issue=9&spage=898&pages=898-900&date=2024&title=Journal+of+Endourology&atitle=Do+We+Really+Need+Stone+Analysis+and+24-Hour+Urine+Testing%3F+No.&aulast=Modai&pid=%3Cauthor%3EModai+J%3BScotland+KB%3C%2Fauthor%3E%3CAN%3E38874954%3C%2FAN%3E%3CDT%3EJournal+Article%3C%2FDT%3E>

5. **The Yield of Genetic Testing in Management of Nephrolithiasis.**

**Item Type:** Journal Article

**Authors:** Pearce, Robert J.;Sui, Wilson;Yang, Heiko;Chi, Thomas and Stoller, Marshall

**Publication Date:** No ,2024

**Journal:** Urology 193, pp. 27–34

**Abstract: OBJECTIVE**: To describe the yield and indications for performing genetic testing in patients with nephrolithiasis. Genetic testing for nephrolithiasis is becoming more accessible and rapid due to technologic advances. This study aimed to determine the diagnostic yield of genetic testing in a cohort of high-risk stone formers and to identify 24-hour urine characteristics to prompt genetic screening. **MATERIALS AND METHODS**: We retrospectively identified patients who underwent genetic testing for nephrolithiasis from 2020 to 22 at a single institution using a custom PerkinElmer genomics panel for nephrolithiasis. We compared characteristics of patients with and without genetic abnormalities. We used receiver operator characteristic (ROC) analysis to identify candidate thresholds for genetic testing. **RESULTS**: Fourteen of 36 patients (39%) who underwent genetic testing had identifiable mutations. Five patients (14%) had known pathogenic mutations, including genes for primary hyperoxaluria (PH2 and PH3), cystinuria, and enamel-renal syndrome. The remaining mutations were variants of uncertain significance. Of the 14 patients with identified mutations, only 6 had concordant 24-hour urine abnormalities, including 3/5 with known pathogenic mutations. In patients with urine oxalate >= 40 mg/day, 3/29 (10.3%) had PH2 or PH3. ROC analysis showed that an oxalate threshold of >= 80 mg/day may have promising screening characteristics. **CONCLUSION**: Genetic testing for nephrolithiasis remains controversial due to unknown yield and the time and energy required to discuss results with patients. This preliminary report describes the yield and identifies clinical factors and a potential cut-off that may assist clinicians in deciding when genetic profiling should be pursued. Copyright © 2024. Published by Elsevier Inc.

**URL:** <https://ovidsp.ovid.com/ovidweb.cgi?T=JS&CSC=Y&NEWS=N&PAGE=fulltext&D=medl&DO=10.1016%2fj.urology.2024.07.002> <https://libkey.io/libraries/2592/openurl?genre=article&sid=OVID:medline&id=pmid:38992507&id=doi:10.1016%2Fj.urology.2024.07.002&issn=0090-4295&isbn=&volume=193&issue=&spage=27&pages=27-34&date=2024&title=Urology&atitle=The+Yield+of+Genetic+Testing+in+Management+of+Nephrolithiasis.&aulast=Pearce&pid=%3Cauthor%3EPearce+RJ%3BSui+W%3BYang+H%3BChi+T%3BStoller+M%3C%2Fauthor%3E%3CAN%3E38992507%3C%2FAN%3E%3CDT%3EJournal+Article%3C%2FDT%3E>

6. **Is the "Pandemic Diet" Nonlithogenic?.**

**Item Type:** Journal Article

**Authors:** Shkolnik, Brian;Zhao, Kelley;Lu, Jennifer Y.;Kothari, Pankti;Herfel, Scott;Schulsinger, Hailey and Schulsinger, David

**Publication Date:** Ju ,2024

**Journal:** Journal of Endourology 38(7), pp. 697–700

**Abstract:** Introduction and

**URL:** <https://ovidsp.ovid.com/ovidweb.cgi?T=JS&CSC=Y&NEWS=N&PAGE=fulltext&D=medl&DO=10.1089%2fend.2023.0535> <https://libkey.io/libraries/2592/openurl?genre=article&sid=OVID:medline&id=pmid:38623784&id=doi:10.1089%2Fend.2023.0535&issn=0892-7790&isbn=&volume=38&issue=7&spage=697&pages=697-700&date=2024&title=Journal+of+Endourology&atitle=Is+the+%22Pandemic+Diet%22+Nonlithogenic%3F.&aulast=Shkolnik&pid=%3Cauthor%3EShkolnik+B%3BZhao+K%3BLu+JY%3BKothari+P%3BHerfel+S%3BSchulsinger+H%3BSchulsinger+D%3C%2Fauthor%3E%3CAN%3E38623784%3C%2FAN%3E%3CDT%3EJournal+Article%3C%2FDT%3E>

7. **Associations Between Provider Specialty and Use of Follow-up Testing Among Patients on Preventive Pharmacological Therapy for Urinary Stone Disease.**

**Item Type:** Journal Article

**Authors:** Crivelli, Joseph J.;Yan, Phyllis L.;Shahinian, Vahakn;Hsi, Ryan S. and Hollingsworth, John M.

**Publication Date:** 2023

**Journal:** Urology Practice 10(4), pp. 400–406

**Abstract: INTRODUCTION**: The AUA Medical Management of Kidney Stones guideline outlines recommendations on follow-up testing for patients prescribed preventive pharmacological therapy. We evaluated adherence to these recommendations by provider specialty. **METHODS**: Using claims data from working-age adults with urinary stone disease (2008-2019), we identified patients prescribed a preventive pharmacological therapy agent (a thiazide diuretic, alkali citrate therapy, allopurinol, or a combination thereof) and the specialty of the prescribing physician (urology, nephrology, and general practice). Next, we identified patients who completed a 24-hour urine collection prior to their prescription fill. We then measured adherence to 3 recommendations outlined in the AUA guideline. Finally, we fit multivariable logistic regression models evaluating associations between prescribing provider specialty and adherence to recommended follow-up testing. **RESULTS**: Among 2,600 patients meeting study criteria, 1,523 (59%) adhered to >=1 follow-up testing recommendation, with a significant increase over the study period. Nephrologists had higher odds of adherence to >=1 follow-up test compared to urologists (odds ratio, 1.52; 95% confidence interval, 1.19-1.94; P : Among 2,600 patients meeting study criteria, 1,523 (59%) adhered to >=1 follow-up testing recommendation, with a significant increase over the study period. Nephrologists had higher odds of adherence to >=1 follow-up test compared to urologists (odds ratio, 1.52; 95% confidence interval, 1.19-1.94; P **CONCLUSIONS**: Following initiation of preventive pharmacological therapy, adherence to guideline-recommended follow-up testing was low overall. There exist meaningful specialty-specific differences in the use of this testing.

**URL:** <https://ovidsp.ovid.com/ovidweb.cgi?T=JS&CSC=Y&NEWS=N&PAGE=fulltext&D=med24&DO=10.1097%2fUPJ.0000000000000414> <https://libkey.io/libraries/2592/openurl?genre=article&sid=OVID:medline&id=pmid:37341368&id=doi:10.1097%2FUPJ.0000000000000414&issn=2352-0779&isbn=&volume=10&issue=4&spage=400&pages=400-406&date=2023&title=Urology+Practice&atitle=Associations+Between+Provider+Specialty+and+Use+of+Follow-up+Testing+Among+Patients+on+Preventive+Pharmacological+Therapy+for+Urinary+Stone+Disease.&aulast=Crivelli&pid=%3Cauthor%3ECrivelli+JJ%3BYan+PL%3BShahinian+V%3BHsi+RS%3BHollingsworth+JM%3C%2Fauthor%3E%3CAN%3E37341368%3C%2FAN%3E%3CDT%3EJournal+Article%3C%2FDT%3E>

8. **Breaking the Cycle of Recurrent Calcium Stone Disease**

**Item Type:** Journal Article

**Authors:** Malieckal, Deepa A.;Ganesan, Calyani;Mendez, Denisse Arellano and Pao, Alan C.

**Publication Date:** 2023

**Journal:** Advances in Kidney Disease and Health 30(2), pp. 164–176

**Abstract:** Calcium stones are common and recurrent in nature, yet few therapeutic tools are available for secondary prevention. Personalized approaches for stone prevention have been informed by 24-hour urine testing to guide dietary and medical interventions. However, current evidence is conflicting about whether an approach guided by 24-hour urine testing is more effective than a generic one. The available medications for stone prevention, namely thiazide diuretics, alkali, and allopurinol, are not always prescribed consistently, dosed correctly, or tolerated well by patients. New treatments on the horizon hold the promise of preventing calcium oxalate stones by degrading oxalate in the gut, reprogramming the gut microbiome to reduce oxalate absorption, or knocking down expression of enzymes involved in hepatic oxalate production. New treatments are also needed to target Randall's plaque, the root cause of calcium stone formation. Copyright © 2022 National Kidney Foundation, Inc. Published by Elsevier Inc. All rights reserved.

**URL:** <https://ovidsp.ovid.com/ovidweb.cgi?T=JS&CSC=Y&NEWS=N&PAGE=fulltext&D=med23&DO=10.1053%2fj.akdh.2022.12.004> <https://libkey.io/libraries/2592/openurl?genre=article&sid=OVID:medline&id=pmid:36868731&id=doi:10.1053%2Fj.akdh.2022.12.004&issn=2949-8139&isbn=&volume=30&issue=2&spage=164&pages=164-176&date=2023&title=Advances+in+Kidney+Disease+and+Health&atitle=Breaking+the+Cycle+of+Recurrent+Calcium+Stone+Disease.&aulast=Malieckal&pid=%3Cauthor%3EMalieckal+DA%3BGanesan+C%3BMendez+DA%3BPao+AC%3C%2Fauthor%3E%3CAN%3E36868731%3C%2FAN%3E%3CDT%3EJournal+Article%3C%2FDT%3E>

9. **Serum and 24-hour urinary tests cost-effectiveness in stone formers.**

**Item Type:** Journal Article

**Authors:** Mohammadi, Abdolreza;Farabi, Hiro;Baghdadabad, Leila Zareian;Narouie, Behzad;Reis, Leonardo Oliveira and Aghamir, Seyed Mohammad Kazem

**Publication Date:** Aug 27 ,2023

**Journal:** BMC Urology 23(1), pp. 141

**Abstract: OBJECTIVE**: To assess the routine serum and 24-hour urine tests proficiency in diagnosing the baseline metabolic abnormality of kidney stone formers. **METHODS**: This study analyzes the routine serum and 24-hour urine tests proficiency in diagnosing the baseline metabolic abnormality of kidney stone formers. The sensitivity and specificity, false positive, and negative results of the tests are extracted from diagnostic kits used in the laboratories of the target community. To accurately infer the results, a simulation based on 1000 people was used through 22 standard laboratory tests (Additional File 2), including calcium, oxalate, phosphate, uric acid, sulphate, potassium, sodium, citrate, and magnesium in 24-hour urine; and calcium, creatinine, Vit D, uric acid, and intact parathyroid hormone (PTH) in serum. The incremental cost-effectiveness ratio (ICER) was calculated and compared for each diagnostic test versus other diagnostic tests according to the incremental cost required for correct diagnoses of stone causes. **RESULTS**: Urinary uric acid, citrate, and serum potassium constitute the cost-effectiveness boundary curve in this study. This means that other diagnostic tests are not cost-effective compared to these three tests in terms of indexing at least one item of cost and effectiveness. The ICER index for each correct diagnosis with the urinary uric acid test was $ 1.25 per diagnosis, the most cost-effective test compared to serum potassium and urinary citrate. **CONCLUSION**: The simplified blood and 24-hour urine metabolic evaluation, including urinary uric acid, citrate, and serum potassium, constitute the cost-effectiveness boundary curve. The most cost-effective test was urinary uric acid measurement. Copyright © 2023. BioMed Central Ltd., part of Springer Nature.

**URL:** <https://ovidsp.ovid.com/ovidweb.cgi?T=JS&CSC=Y&NEWS=N&PAGE=fulltext&D=med24&DO=10.1186%2fs12894-023-01310-w> <https://libkey.io/libraries/2592/openurl?genre=article&sid=OVID:medline&id=pmid:37635222&id=doi:10.1186%2Fs12894-023-01310-w&issn=1471-2490&isbn=&volume=23&issue=1&spage=141&pages=141&date=2023&title=BMC+Urology&atitle=Serum+and+24-hour+urinary+tests+cost-effectiveness+in+stone+formers.&aulast=Mohammadi&pid=%3Cauthor%3EMohammadi+A%3BFarabi+H%3BBaghdadabad+LZ%3BNarouie+B%3BReis+LO%3BAghamir+SMK%3C%2Fauthor%3E%3CAN%3E37635222%3C%2FAN%3E%3CDT%3EJournal+Article%3C%2FDT%3E>

10. **Attitudes of urologists on metabolic evaluation for urolithiasis: outcomes of a global survey from 57 countries.**

**Item Type:** Journal Article

**Authors:** Karagoz, Mehmet Ali;Guven, Selcuk;Tefik, Tzevat;Gokce, Mehmet Ilker;Kiremit, Murat Can;Atar, Feyzi Arda;Ibis, Muhammed Arif;Yitgin, Yasin;Boyuk, Abubekir;Verep, Samed and Sarica, Kemal

**Publication Date:** De ,2022

**Journal:** Urolithiasis 50(6), pp. 711–720

**Abstract:** Although stone disease is an important health problem with high incidence and recurrence rates, it is a preventable disease. Attitudes and practices of urologists regarding the prevention of recurrence continue to be a subject of debate. In this context, an online survey study was conducted involving 305 urologists from 57 different countries. The first 7 questions collected demographic data about the urologists and the remaining 23 questions were about the recurrence and metabolic evaluation, medical treatment, and follow-up of urinary stone disease. Most urologists (85.2%) thought that metabolic examination was important. Approximately one-third of the participants (34.1%) performed 24-hour urine analysis and stone analysis was ordered by 87.5% of the urologists. Metabolic analysis was performed for all patients by 14.7% of the participants. For pediatric patients this rate was 68.5%, and for adults with recurrence the rate was 81.6%. Reasons cited by the urologists for not performing metabolic analysis included not feeling confident doing so (18.3%), having limited facilities in their hospital (26.5%), having an excessive daily workload (31.8%), patient-related factors (27.5%), and referring patients to other departments for metabolic evaluation (20.9%). Although majority of the responding urologists do consider the metabolic analysis as vital important, they seemed not to be willing to perform these tests with the same degree of enthusiasm in their daily practice. Our results show that urologists need support in performing and interpreting 24-hour urine analysis, improving their knowledge levels, and communicating with patients. Urology residency training should focus more on the prevention of urinary stone recurrence in addition to the surgical training. Copyright © 2022. The Author(s), under exclusive licence to Springer-Verlag GmbH Germany, part of Springer Nature.

**URL:** <https://ovidsp.ovid.com/ovidweb.cgi?T=JS&CSC=Y&NEWS=N&PAGE=fulltext&D=med22&DO=10.1007%2fs00240-022-01362-x> <https://libkey.io/libraries/2592/openurl?genre=article&sid=OVID:medline&id=pmid:36169664&id=doi:10.1007%2Fs00240-022-01362-x&issn=2194-7228&isbn=&volume=50&issue=6&spage=711&pages=711-720&date=2022&title=Urolithiasis&atitle=Attitudes+of+urologists+on+metabolic+evaluation+for+urolithiasis%3A+outcomes+of+a+global+survey+from+57+countries.&aulast=Karagoz&pid=%3Cauthor%3EKaragoz+MA%3BGuven+S%3BTefik+T%3BGokce+MI%3BKiremit+MC%3BAtar+FA%3BIbis+MA%3BYitgin+Y%3BBoyuk+A%3BVerep+S%3BSarica+K%3C%2Fauthor%3E%3CAN%3E36169664%3C%2FAN%3E%3CDT%3EJournal+Article%3C%2FDT%3E>

11. **Twenty-four-hour Urine Testing and Urinary Stone Disease Recurrence in Veterans.**

**Item Type:** Journal Article

**Authors:** Song, Shen;Thomas, I-Chun;Ganesan, Calyani;Velaer, Kyla N.;Chertow, Glenn M.;Pao, Alan C. and Leppert, John T.

**Publication Date:** 2022

**Journal:** Urology 159, pp. 33–40

**Abstract: OBJECTIVE**: To determine whether 24-hour urine testing in Veterans with USD (urinary stone disease) reduces or delays urinary stone recurrence. **METHODS**: Cohort study of national health record data from Veterans Health Administration from 2007 through 2013. We utilized a study population of 130,129 Veterans with USD based on diagnostic or procedural codes and excluded those with USD claims in the 2 years before cohort entry. We then created a propensity-score matched cohort of 14,854 Veterans based on completion of 24-hour urine testing within 6 months of stone diagnosis. Primary outcome was time-to-next clinically significant stone event, defined as an emergency department visit, inpatient admission related to a urinary stone, or urologic stone procedure with 5-year follow up. **RESULTS**: Of 14,854 Veterans in the propensity-score matched cohort, 8560 (57.6%) experienced a recurrent USD event. Completion of 24-hour urine testing was associated with a higher risk of developing a second stone event (hazard ratio [HR] 1.17, 95% confidence interval [95% CI] 1.12-1.22). Among Veterans with known recurrent disease, we examined time to a third stone event. In this cohort of 4736 patients, completion of 24-hour urine testing was not associated with a higher risk of developing a third stone event (HR 1.06, 95% CI 0.99-1.12). **CONCLUSION**: Completion of 24-hour urine testing was not associated with a reduction in urinary stone recurrence. These findings challenge the validity of a longstanding recommendation in general medicine, nephrology, and urology practice. Copyright © 2021 Elsevier Inc. All rights reserved.

**URL:** <https://ovidsp.ovid.com/ovidweb.cgi?T=JS&CSC=Y&NEWS=N&PAGE=fulltext&D=med21&DO=10.1016%2fj.urology.2021.10.005> <https://libkey.io/libraries/2592/openurl?genre=article&sid=OVID:medline&id=pmid:34688771&id=doi:10.1016%2Fj.urology.2021.10.005&issn=0090-4295&isbn=&volume=159&issue=&spage=33&pages=33-40&date=2022&title=Urology&atitle=Twenty-four-hour+Urine+Testing+and+Urinary+Stone+Disease+Recurrence+in+Veterans.&aulast=Song&pid=%3Cauthor%3ESong+S%3BThomas+IC%3BGanesan+C%3BVelaer+KN%3BChertow+GM%3BPao+AC%3BLeppert+JT%3C%2Fauthor%3E%3CAN%3E34688771%3C%2FAN%3E%3CDT%3EJournal+Article%3C%2FDT%3E>

12. **Nuclear Magnetic Resonance Metabolomic Profiling and Urine Chemistries in Incident Kidney Stone Formers Compared with Controls.**

**Item Type:** Journal Article

**Authors:** Thongprayoon, Charat;Vuckovic, Ivan;Vaughan, Lisa E.;Macura, Slobodan;Larson, Nicholas B.;D'Costa, Matthew R.;Lieske, John C.;Rule, Andrew D. and Denic, Aleksandar

**Publication Date:** No ,2022

**Journal:** Journal of the American Society of Nephrology 33(11), pp. 2071–2086

**Abstract: BACKGROUND**: The urine metabolites and chemistries that contribute to kidney stone formation are not fully understood. This study examined differences between the urine metabolic and chemistries profiles of first-time stone formers and controls. **METHODS**: High-resolution ^1^H-nuclear magnetic resonance (NMR) spectroscopy-based metabolomic analysis was performed in 24-hour urine samples from a prospective cohort of 418 first-time symptomatic kidney stone formers and 440 controls. In total, 48 NMR-quantified metabolites in addition to 12 standard urine chemistries were assayed. Analysis of covariance was used to determine the association of stone former status with urine metabolites or chemistries after adjusting for age and sex and correcting for the false discovery rate. Gradient-boosted machine methods with nested cross-validation were applied to predict stone former status. **RESULTS**: Among the standard urine chemistries, stone formers had lower urine oxalate and potassium and higher urine calcium, phosphate, and creatinine. Among NMR urine metabolites, stone formers had lower hippuric acid, trigonelline, 2-furoylglycine, imidazole, and citrate and higher creatine and alanine. A cross-validated model using urine chemistries, age, and sex yielded a mean AUC of 0.76 (95% CI, 0.73 to 0.79). A cross-validated model using urine chemistries, NMR-quantified metabolites, age, and sex did not meaningfully improve the discrimination (mean AUC, 0.78; 95% CI, 0.75 to 0.81). In this combined model, among the top ten discriminating features, four were urine chemistries and six NMR-quantified metabolites. **CONCLUSIONS**: Although NMR-quantified metabolites did not improve discrimination, several urine metabolic profiles were identified that may improve understanding of kidney stone pathogenesis. Copyright © 2022 by the American Society of Nephrology.

**URL:** <https://ovidsp.ovid.com/ovidweb.cgi?T=JS&CSC=Y&NEWS=N&PAGE=fulltext&D=med22&DO=10.1681%2fASN.2022040416> <https://libkey.io/libraries/2592/openurl?genre=article&sid=OVID:medline&id=pmid:36316097&id=doi:10.1681%2FASN.2022040416&issn=1046-6673&isbn=&volume=33&issue=11&spage=2071&pages=2071-2086&date=2022&title=Journal+of+the+American+Society+of+Nephrology&atitle=Nuclear+Magnetic+Resonance+Metabolomic+Profiling+and+Urine+Chemistries+in+Incident+Kidney+Stone+Formers+Compared+with+Controls.&aulast=Thongprayoon&pid=%3Cauthor%3EThongprayoon+C%3BVuckovic+I%3BVaughan+LE%3BMacura+S%3BLarson+NB%3BD%27Costa+MR%3BLieske+JC%3BRule+AD%3BDenic+A%3C%2Fauthor%3E%3CAN%3E36316097%3C%2FAN%3E%3CDT%3EJournal+Article%3C%2FDT%3E>

13. **Stone Morphology Distinguishes Two Pathways of Idiopathic Calcium Oxalate Stone Pathogenesis.**

**Item Type:** Journal Article

**Authors:** Williams, James C. Jr;Al-Awadi, Haider;Muthenini, Manognya;Bledsoe, Sharon B.;El-Achkar, Tarek;Evan, Andrew P.;Coe, Fredric;Lingeman, James E. and Worcester, Elaine M.

**Publication Date:** 2022

**Journal:** Journal of Endourology 36(5), pp. 694–702

**Abstract: Introduction:** About 1 in 11 Americans will experience a kidney stone, but underlying causes remain obscure. The objective of the present study was to separate idiopathic calcium oxalate stone formers by whether or not they showed positive evidence of forming a stone on Randall's plaque (RP). **Materials and Methods:** In patients undergoing either percutaneous or ureteroscopic procedures for kidney stone removal, all stone material was extracted and analyzed using micro-CT imaging to identify those attached to RP. Twenty-four-hour urine samples were collected weeks after the stone removal procedure and patients were off of medications that would affect urine composition. The endoscopic video was analyzed for papillary pathology (RP, pitting, plugging, dilated ducts, and loss of papillary shape) by an observer blinded to the data on stone type. The percent papillary area occupied by RP and ductal plugging was quantified using image analysis software. **Results:** Patients having even one stone on RP (N = 36) did not differ from non-RP patients (N = 37) in age, sex, BMI, or other clinical characteristics. Compared with the non-RP group, RP stone formers had more numerous, but smaller, stones, more abundant papillary RP formation, and fewer ductal plugs, both by quantitative measurement of surface area (on average, three times more plaque area, but only 41% as much plug area as in non-RP patients) and by semiquantitative visual grading. Serum and blood values did not differ between RP and non-RP stone formers by any measure. **Conclusions:** Growth of many small stones on plaque seems the pathogenetic scheme for the RP stone-forming phenotype, whereas the non-RP phenotype stone pathogenesis pathway is less obvious. Higher papillary plugging in non-RP patients suggests that plugs play a role in stone formation and that these patients have a greater degree of papillary damage. Underlying mechanisms that create these distinctive phenotypes are presently unknown.

**URL:** <https://ovidsp.ovid.com/ovidweb.cgi?T=JS&CSC=Y&NEWS=N&PAGE=fulltext&D=med21&DO=10.1089%2fend.2021.0685> <https://libkey.io/libraries/2592/openurl?genre=article&sid=OVID:medline&id=pmid:34915736&id=doi:10.1089%2Fend.2021.0685&issn=0892-7790&isbn=&volume=36&issue=5&spage=694&pages=694-702&date=2022&title=Journal+of+Endourology&atitle=Stone+Morphology+Distinguishes+Two+Pathways+of+Idiopathic+Calcium+Oxalate+Stone+Pathogenesis.&aulast=Williams&pid=%3Cauthor%3EWilliams+JC+Jr%3BAl-Awadi+H%3BMuthenini+M%3BBledsoe+SB%3BEl-Achkar+T%3BEvan+AP%3BCoe+F%3BLingeman+JE%3BWorcester+EM%3C%2Fauthor%3E%3CAN%3E34915736%3C%2FAN%3E%3CDT%3EJournal+Article%3C%2FDT%3E>

14. **Variation in Urinary Stone Parameters Throughout the Day and the Effect of Increased Fluid and Citrate Supplementation.**

**Item Type:** Journal Article

**Authors:** Bhatt, Vikas;White, Mark D.;Listman, James;Feustel, Paul J.;Howe, Adam and Kogan, Barry A.

**Publication Date:** 2021

**Journal:** Journal of Endourology 35(10), pp. 1548–1554

**Abstract: Background:** Because 24-hour urine collections are cumbersome, many studies have evaluated the use of spot urine samples as a substitute, mostly finding poor concordance between the two. Daily variation in stone parameters probably contributes to the lack of concordance, but specific variation in various stone parameters is not well delineated. The variations likely lead to peaks and troughs, which can increase the risk of stone formation. **Methods:** We prospectively recruited 20 nonstone-forming patients, recording their total fluid intake over 24 hours and collecting voids at first morning, 9 to 10 A.M., 1 to 2 P.M., and 4 to 5 P.M. for evaluation of pH, specific gravity, calcium, citrate, and creatinine. Participants were then asked to double their fluid intake and take a daily True Lemon supplement over the course of the next 3 days. Urine was recollected postintervention. **Results:** Baseline [citrate]/[creatinine] increased throughout the day such that the 5 P.M. level was significantly higher compared with first void (0.58 vs 0.42, p = 0.027); [calcium]/[creatinine] daily variation was not statistically significant, but showed a distinct pattern that was present in both sets of collections. Daily [calcium]/[citrate] variation was significantly (p = 0.004) and consistently highest in the early morning on both day 1 (0.43) and day 4 (0.45). There was no significant variation in specific gravity and pH. Increasing fluid intake and citrate supplementation increase the daily variation in pH and [citrate]/[creatinine], but did not increase the values compared with their respective preintervention void times. There was also no detectable postintervention effect on [Ca]/[creatinine] or specific gravity. **Conclusions:** Urinary citrate concentration follows a circadian pattern, while urinary calcium has a diurnal excretion pattern. [Calcium]:[citrate] is highest in the early morning, indicating a high-risk time of day for stone formation. Spot urine samples identify a key time of day, which 24-hour urine collections may miss, for clinical monitoring.

**URL:** <https://ovidsp.ovid.com/ovidweb.cgi?T=JS&CSC=Y&NEWS=N&PAGE=fulltext&D=med19&DO=10.1089%2fend.2020.1212> <https://libkey.io/libraries/2592/openurl?genre=article&sid=OVID:medline&id=pmid:33637013&id=doi:10.1089%2Fend.2020.1212&issn=0892-7790&isbn=&volume=35&issue=10&spage=1548&pages=1548-1554&date=2021&title=Journal+of+Endourology&atitle=Variation+in+Urinary+Stone+Parameters+Throughout+the+Day+and+the+Effect+of+Increased+Fluid+and+Citrate+Supplementation.&aulast=Bhatt&pid=%3Cauthor%3EBhatt+V%3BWhite+MD%3BListman+J%3BFeustel+PJ%3BHowe+A%3BKogan+BA%3C%2Fauthor%3E%3CAN%3E33637013%3C%2FAN%3E%3CDT%3EJournal+Article%3C%2FDT%3E>

15. **Nutritional status assessed by the Controlling Nutritional Status (CONUT) score as a predictor of recurrence of urolithiasis.**

**Item Type:** Journal Article

**Authors:** Lee, Hee Youn;Kang, Ho Won;Kim, Kyeong;Ha, Yun-Sok;Kim, Won Tae;Kim, Yong-June;Yun, Seok Joong;Kim, Wun-Jae and Lee, Sang-Cheol

**Publication Date:** 2021

**Journal:** Investigative and Clinical Urology 62(5), pp. 553–559

**Abstract: PURPOSE**: We aimed to determine the influence of nutritional status on urinary metabolic abnormalities and stone recurrence in patients with urolithiasis. **MATERIALS AND METHODS**: We analyzed data for 464 stone-formers and 464 propensity-score-matched control patients that had been collected between 2003 and 2015. Nutritional status was evaluated by use of the Controlling Nutritional Status (CONUT) score, and patients were placed into two CONUT score categories (0-1 and >=2). Serum and 24-hour urinary metabolites were evaluated in 464 stone-formers. Kaplan-Meier and multivariate Cox regression analyses were performed to assess the influence of nutritional status on stone recurrence. Stone recurrence was defined as radiographic appearance of new stones during the follow-up period. **RESULTS**: Stone-formers showed a higher prevalence of poor nutrition (CONUT score >=2) than did the propensity-score-matched control patients (p: Stone-formers showed a higher prevalence of poor nutrition (CONUT score >=2) than did the propensity-score-matched control patients (p: Stone-formers showed a higher prevalence of poor nutrition (CONUT score >=2) than did the propensity-score-matched control patients (p**CONCLUSIONS**: The CONUT score, an easily measured immunonutritional biomarker, is independently associated with a higher risk for stone recurrence in patients with urolithiasis. This implies that not only dietary excess, but also undernourished status, may be associated with aberrations in urine physicochemistry and stone recurrence. Copyright © The Korean Urological Association, 2021.

**URL:** <https://ovidsp.ovid.com/ovidweb.cgi?T=JS&CSC=Y&NEWS=N&PAGE=fulltext&D=med20&DO=10.4111%2ficu.20210031> <https://libkey.io/libraries/2592/openurl?genre=article&sid=OVID:medline&id=pmid:34387033&id=doi:10.4111%2Ficu.20210031&issn=2466-0493&isbn=&volume=62&issue=5&spage=553&pages=553-559&date=2021&title=Investigative+And+Clinical+Urology&atitle=Nutritional+status+assessed+by+the+Controlling+Nutritional+Status+%28CONUT%29+score+as+a+predictor+of+recurrence+of+urolithiasis.&aulast=Lee&pid=%3Cauthor%3ELee+HY%3BKang+HW%3BKim+K%3BHa+YS%3BKim+WT%3BKim+YJ%3BYun+SJ%3BKim+WJ%3BLee+SC%3C%2Fauthor%3E%3CAN%3E34387033%3C%2FAN%3E%3CDT%3EJournal+Article%3C%2FDT%3E>

16. **Re: Twenty-Four Hour Urine Testing and Prescriptions for Urinary Stone Disease-Related Medications in Veterans.**

**Item Type:** Journal Article

**Authors:** Assimos, Dean G.

**Publication Date:** 2020

**Journal:** Journal of Urology 203(6), pp. 1056–1057

**URL:** <https://ovidsp.ovid.com/ovidweb.cgi?T=JS&CSC=Y&NEWS=N&PAGE=fulltext&D=med17&DO=10.1097%2fJU.0000000000000997.01> <https://libkey.io/libraries/2592/openurl?genre=article&sid=OVID:medline&id=pmid:32155362&id=doi:10.1097%2FJU.0000000000000997.01&issn=0022-5347&isbn=&volume=203&issue=6&spage=1056&pages=1056-1057&date=2020&title=Journal+of+Urology&atitle=Re%3A+Twenty-Four+Hour+Urine+Testing+and+Prescriptions+for+Urinary+Stone+Disease-Related+Medications+in+Veterans.&aulast=Assimos&pid=%3Cauthor%3EAssimos+DG%3C%2Fauthor%3E%3CAN%3E32155362%3C%2FAN%3E%3CDT%3EJournal+Article%3C%2FDT%3E>

17. **The Impact of Alternative Alkalinizing Agents on 24-Hour Urine Parameters.**

**Item Type:** Journal Article

**Authors:** Boydston, Kohldon;Terry, Russell;Winship, Brenton;Davis, Leah;Yttri, Sarah;Carlos, Evan;Scales, Charles;Lipkin, Michael and Preminger, Glenn M.

**Publication Date:** 2020

**Journal:** Urology 142, pp. 55–59

**Abstract: OBJECTIVES**: To determine if alternative alkalinizing agents lead to similar changes in 24-hour urine pH and citrate compared to potassium citrate (KCIT). Many stone formers cannot tolerate KCIT due to side effects or cost. In these patients, we have prescribed potassium bicarbonate or sodium bicarbonate as alternative alkali (AA), though their efficacy is unclear. **METHODS**: We performed a retrospective cohort study of adult stone formers seen from 2000 to 2018 with 24-hour urine analyses. Two analyses were performed. The first evaluated the alkalinizing and citraturic effects in patients with baseline low urine pH or hypocitraturia off of any alkalinizing medications, who were subsequently treated with either KCIT or AA. The second analysis compared the pH and citrate in patients changing from KCIT to an AA. Reasons for switching were abstracted by chart review and cost savings percentages were calculated using GoodRx medication prices. **RESULTS**: When starting alkali therapy, the median increase in pH from baseline was 0.64 for KCIT and 0.51 for AA (P=.077), and the median increase in citrate from baseline was 231 mg for KCIT and 171 mg for AA (P=.109). When switching alkali therapy, median pH and citrate did not significantly change. Hyperkalemia (24%), GI upset (19%), and cost (17%) were the most common reasons cited for switching to an AA. AA represented a savings of 86%-92% compared to KCIT. **CONCLUSION**: Alternative alkali appear to offer comparable improvements in 24-hour urine parameters and significant cost-savings compared to KCIT. Copyright © 2020 Elsevier Inc. All rights reserved.

**URL:** <https://ovidsp.ovid.com/ovidweb.cgi?T=JS&CSC=Y&NEWS=N&PAGE=fulltext&D=med17&DO=10.1016%2fj.urology.2020.04.047> <https://libkey.io/libraries/2592/openurl?genre=article&sid=OVID:medline&id=pmid:32330528&id=doi:10.1016%2Fj.urology.2020.04.047&issn=0090-4295&isbn=&volume=142&issue=&spage=55&pages=55-59&date=2020&title=Urology&atitle=The+Impact+of+Alternative+Alkalinizing+Agents+on+24-Hour+Urine+Parameters.&aulast=Boydston&pid=%3Cauthor%3EBoydston+K%3BTerry+R%3BWinship+B%3BDavis+L%3BYttri+S%3BCarlos+E%3BScales+C%3BLipkin+M%3BPreminger+GM%3C%2Fauthor%3E%3CAN%3E32330528%3C%2FAN%3E%3CDT%3EJournal+Article%3C%2FDT%3E>

18. **SLIPS-LAB-A bioinspired bioanalysis system for metabolic evaluation of urinary stone disease.**

**Item Type:** Journal Article

**Authors:** Li, Hui;Shkolyar, Eugene;Wang, Jing;Conti, Simon;Pao, Alan C.;Liao, Joseph C.;Wong, Tak-Sing and Wong, Pak Kin

**Publication Date:** 2020

**Journal:** Science Advances 6(21), pp. eaba8535

**Abstract:** Urinary stone disease is among the most common medical conditions. Standard evaluation of urinary stone disease involves a metabolic workup of stone formers based on measurement of minerals and solutes excreted in 24-hour urine samples. Nevertheless, 24-hour urine testing is slow, expensive, and inconvenient for patients, which has hindered widespread adoption in clinical practice. Here, we demonstrate SLIPS-LAB (Slippery Liquid-Infused Porous Surface Laboratory), a droplet-based bioanalysis system, for rapid measurement of urinary stone-associated analytes. The ultra-repellent and antifouling properties of SLIPS, which is a biologically inspired surface technology, allow autonomous liquid handling and manipulation of physiological samples without complicated sample preparation procedures and supporting equipment. We pilot a study that examines key urinary analytes in clinical samples from patients with urinary stone. The simplicity and speed of SLIPS-LAB hold the potential to provide actionable diagnostic information for patients with urinary stone disease and rapid feedback for responses to dietary and pharmacologic treatments. Copyright © 2020 The Authors, some rights reserved; exclusive licensee American Association for the Advancement of Science. No claim to original U.S. Government Works. Distributed under a Creative Commons Attribution NonCommercial License 4.0 (CC BY-NC).

**URL:** <https://ovidsp.ovid.com/ovidweb.cgi?T=JS&CSC=Y&NEWS=N&PAGE=fulltext&D=med18&DO=10.1126%2fsciadv.aba8535> <https://libkey.io/libraries/2592/openurl?genre=article&sid=OVID:medline&id=pmid:32494753&id=doi:10.1126%2Fsciadv.aba8535&issn=2375-2548&isbn=&volume=6&issue=21&spage=eaba8535&pages=eaba8535&date=2020&title=Science+Advances&atitle=SLIPS-LAB-A+bioinspired+bioanalysis+system+for+metabolic+evaluation+of+urinary+stone+disease.&aulast=Li&pid=%3Cauthor%3ELi+H%3BShkolyar+E%3BWang+J%3BConti+S%3BPao+AC%3BLiao+JC%3BWong+TS%3BWong+PK%3C%2Fauthor%3E%3CAN%3E32494753%3C%2FAN%3E%3CDT%3EJournal+Article%3C%2FDT%3E>

19. **Pharmacological Dilutional Therapy Using the Vasopressin Antagonist Tolvaptan for Young Patients With Cystinuria: A Pilot Investigation.**

**Item Type:** Journal Article

**Authors:** Nelson, Caleb P.;Kurtz, Michael P.;Venna, Alyssia;Cilento, Bartley G. Jr and Baum, Michelle A.

**Publication Date:** 2020

**Journal:** Urology 144, pp. 65–70

**Abstract: OBJECTIVE**: To perform a pilot study of short-term safety, tolerability, and impact on urinary stone risk parameters of the vasopressin V2-receptor antagonist tolvaptan (which increases urinary excretion of free water) among adolescents and young adults with cystinuria. **MATERIALS AND METHODS**: We enrolled cystinuria patients age 12-25 years. Subjects were treated for 4 days at low-dose tolvaptan (0.3 mg/kg/day, maximum 30 mg) and 4 days at high dose (0.6 mg/kg/day, maximum 60 mg). Twenty-four-hour urine collections were done at baseline, day 3-4 of the dosing period, day 7-8 of the dosing period, and 3-6 days after washout. Primary outcome was cystine capacity (mg/L, target capacity > 0). Secondary outcomes included other urinary/serum parameters, tolerability, and thirst response. **RESULTS**: Two females (17, 23 years) and 2 males (13, 24 years) were enrolled. Cystine capacity respectively went from baseline of -312, -82, -353, and -628 mg/L to 97, 111, 75, and -3 mg/L on high dose (Figure 1). Twenty-four-hour volume went from 1.96, 3.0, 2.1, and 0.91 L to 11.74, 6.5, 9.9, and 2.8 L on high dose (Figure 2). There were no abnormalities in serum electrolytes or liver enzymes. Subjects did experience extreme thirst (9/10 on visual scale), but none discontinued treatment or reduced dose. **CONCLUSION**: Dilutional therapy with tolvaptan increased both cystine capacity and urinary volumes. This treatment approach has the potential to reduce recurrence of stones in this population. Further investigation should study longer term effects and safety, and determine optimal dosing to improve tolerability. Copyright © 2020 Elsevier Inc. All rights reserved.

**URL:** <https://ovidsp.ovid.com/ovidweb.cgi?T=JS&CSC=Y&NEWS=N&PAGE=fulltext&D=med18&DO=10.1016%2fj.urology.2020.07.002> <https://libkey.io/libraries/2592/openurl?genre=article&sid=OVID:medline&id=pmid:32683063&id=doi:10.1016%2Fj.urology.2020.07.002&issn=0090-4295&isbn=&volume=144&issue=&spage=65&pages=65-70&date=2020&title=Urology&atitle=Pharmacological+Dilutional+Therapy+Using+the+Vasopressin+Antagonist+Tolvaptan+for+Young+Patients+With+Cystinuria%3A+A+Pilot+Investigation.&aulast=Nelson&pid=%3Cauthor%3ENelson+CP%3BKurtz+MP%3BVenna+A%3BCilento+BG+Jr%3BBaum+MA%3C%2Fauthor%3E%3CAN%3E32683063%3C%2FAN%3E%3CDT%3EJournal+Article%3C%2FDT%3E>

20. **Initial collection of an inadequate 24-hour urine sample in children does not predict subsequent inadequate collections.**

**Item Type:** Journal Article

**Authors:** Chan, K. H.;Moser, E. A.;Whittam, B. M.;Misseri, R.;Cain, M. P.;Krambeck, A. and Schwaderer, A.

**Publication Date:** Fe ,2019

**Journal:** Journal of Pediatric Urology 15(1), pp. 74.e1–74.e7

**Abstract: INTRODUCTION**: Approximately half of adult stone formers submit specimens that are either under or over collections as determined by 24-h creatinine/kg. Previously identified predictors of inadequate collection in adults include female sex, older age, higher body mass index (BMI), vitamin D supplementation, and weekday collection. **OBJECTIVE**: The objective of this study is to determine risk factors for inadequate 24-h urinary specimen collection in the pediatric population. **STUDY DESIGN**: A retrospective analysis of all children (: A retrospective analysis of all children (**RESULTS**: Of 367 patients, 80 had an adequate collection (21.9%): median age, 13 years (interquartile range, 8-16); 61.1% female; 93.5% white; 19.5% obese; and 13.0% overweight. No parental or child factors were associated with inadequate collection (Summary Table). Of inadequate collections, more than 80% were over collections. In the 175 patients with more than one 24-h urinary specimen collection, the effect of an initial inadequate collection on subsequent inadequate collections was not significant after controlling for the correlation of samples from the same patient (p = 0.8). **DISCUSSION**: Any parental or child factors associated with the collection of inadequate 24-h urine specimens in children were not found. An initial inadequate collection does not predict subsequent inadequate collections. It was surprising that >80% of the inadequate collections were over collections rather than under collections. Possible explanations are that children collected urine samples for longer than the 24-h period or that stone-forming children produce more creatinine per 24-h period than healthy children due to hyperfiltration. **CONCLUSION**: Inadequate collections are very common, and the risk factors for them are unclear. A repeat collection would be suggested if the first is inadequate. Further studies must be planned to explore barriers to accurate specimen collection using qualitative research methodology. Copyright © 2018 Journal of Pediatric Urology Company. Published by Elsevier Ltd. All rights reserved.

**URL:** <https://ovidsp.ovid.com/ovidweb.cgi?T=JS&CSC=Y&NEWS=N&PAGE=fulltext&D=med16&DO=10.1016%2fj.jpurol.2018.10.019> <https://libkey.io/libraries/2592/openurl?genre=article&sid=OVID:medline&id=pmid:30467015&id=doi:10.1016%2Fj.jpurol.2018.10.019&issn=1477-5131&isbn=&volume=15&issue=1&spage=74.e1&pages=74.e1-74.e7&date=2019&title=Journal+of+pediatric+urology&atitle=Initial+collection+of+an+inadequate+24-hour+urine+sample+in+children+does+not+predict+subsequent+inadequate+collections.&aulast=Chan&pid=%3Cauthor%3EChan+KH%3BMoser+EA%3BWhittam+BM%3BMisseri+R%3BCain+MP%3BKrambeck+A%3BSchwaderer+A%3C%2Fauthor%3E%3CAN%3E30467015%3C%2FAN%3E%3CDT%3EJournal+Article%3C%2FDT%3E>

21. **Urinary sulphated glycosaminoglycan insufficiency and chondroitin sulphate supplement in urolithiasis.**

**Item Type:** Journal Article

**Authors:** Dissayabutra, Thasinas;Kalpongnukul, Nuttiya;Chindaphan, Kanokporn;Srisa-Art, Monpicha;Ungjaroenwathana, Wattanachai;Kaewwongse, Maroot;Iampenkhae, Kroonpong and Tosukhowong, Piyaratana

**Publication Date:** 2019

**Journal:** PLoS ONE [Electronic Resource] 14(3), pp. e0213180

**Abstract:** Familial members of urolithiasis have high risk for stone development. We observed the low sulphated glycosaminoglycan (GAG) excretion in urolithiasis patients and their descendants. In this study, we investigated urinary excretion of sulphated GAG, chondroitin sulphate (CS), heparan sulphate (HS) and hyaluronic acid (HA) in urolithiasis and their children, and explored the effect of CS and HA supplement in urolithic hyperoxaluric rats. The 24-hour urines were collected from urolithiasis patients (28) and their children (40), as well as healthy controls (45) and their children (33) to measure urinary sulphated GAG, CS, HS and HA excretion rate. Our result showed that urinary sulphated GAG and CS were diminished in both urolithiasis patients and their children, while decreased HS and increased HA were observed only in urolithiasis patients. Percentage of HS per sulphated GAG increased in both urolithiasis patients and their children. In hyperoxaluric rats induced by ethylene glycol and vitamin D, we found that CS supplement could prevent stone formation, while HA supplement had no effect on stone formation. Our study revealed that decreased urinary GAG and CS excretion are common in familial members of urolithiasis patients, and CS supplement might be beneficial in calcium oxalate urolithiasis prophylaxis for hyperoxaluric patients.

**URL:** <https://ovidsp.ovid.com/ovidweb.cgi?T=JS&CSC=Y&NEWS=N&PAGE=fulltext&D=med16&DO=10.1371%2fjournal.pone.0213180> [https://libkey.io/libraries/2592/openurl?genre=article&sid=OVID:medline&id=pmid:30845174&id=doi:10.1371%2Fjournal.pone.0213180&issn=1932-6203&isbn=&volume=14&issue=3&spage=e0213180&pages=e0213180&date=2019&title=PLoS+ONE+%5BElectronic+Resource%5D&atitle=Urinary+sulphated+glycosaminoglycan+insufficiency+and+chondroitin+sulphate+supplement+in+urolithiasis.&aulast=Dissayabutra&pid=%3Cauthor%3EDissayabutra+T%3BKalpongnukul+N%3BChindaphan+K%3BSrisa-Art+M%3BUngjaroenwathana+W%3BKaewwongse+M%3BIampenkhae+K%3BTosukhowong+P%3C%2Fauthor%3E%3CAN%3E30845174%3C%2FAN%3E%3CDT%3EJournal+Article%3C%2FDT%3E](https://libkey.io/libraries/2592/openurl?genre=article&sid=OVID:medline&id=pmid:30845174&id=doi:10.1371%2Fjournal.pone.0213180&issn=1932-6203&isbn=&volume=14&issue=3&spage=e0213180&pages=e0213180&date=2019&title=PLoS+ONE+%5BElectronic+Resource%5D&atitle=Urinary+sulfated+glycosaminoglycan+insufficiency+and+chondroitin+sulfate+supplement+in+urolithiasis.&aulast=Dissayabutra&pid=%3Cauthor%3EDissayabutra+T%3BKalpongnukul+N%3BChindaphan+K%3BSrisa-Art+M%3BUngjaroenwathana+W%3BKaewwongse+M%3BIampenkhae+K%3BTosukhowong+P%3C%2Fauthor%3E%3CAN%3E30845174%3C%2FAN%3E%3CDT%3EJournal+Article%3C%2FDT%3E)

22. **Patients with primary hyperoxaluria type 2 have significant morbidity and require careful follow-up.**

**Item Type:** Journal Article

**Authors:** Garrelfs, Sander F.;Rumsby, Gill;Peters-Sengers, Hessel;Erger, Florian;Groothoff, Jaap W.;Beck, Bodo B.;Oosterveld, Michiel J. S.;Pelle, Alessandra;Neuhaus, Thomas;Adams, Brigitte;Cochat, Pierre;Salido, Eduardo;Lipkin, Graham W.;Hoppe, Bernd and Hulton, Sally-Anne

**Publication Date:** 2019

**Journal:** Kidney International 96(6), pp. 1389–1399

**Abstract:** Primary hyperoxaluria type 2 is a rare inherited disorder of glyoxylate metabolism causing nephrocalcinosis, renal stone formation and ultimately kidney failure. Previously, primary hyperoxaluria type 2 was considered to have a more favorable prognosis than primary hyperoxaluria type 1, but earlier reports are limited by low patient numbers and short follow up periods. Here we report on the clinical, genetic, and biochemical findings from the largest cohort of patients with primary hyperoxaluria type 2, obtained by a retrospective record review of genetically confirmed cases in the OxalEurope registry, a dataset containing 101 patients from eleven countries. Median follow up was 12.4 years. Median ages at first symptom and diagnosis for index cases were 3.2 years and 8.0 years, respectively. Urolithiasis was the most common presenting feature (82.8% of patients). Genetic analysis revealed 18 novel mutations in the GRHPR gene. Of 238 spot-urine analyses, 23 (9.7%) were within the normal range for oxalate as compared to less than 4% of 24-hour urine collections. Median intra-individual variation of 24-hour oxalate excretion was substantial (34.1%). At time of review, 12 patients were lost to follow-up; 45 of the remaining 89 patients experienced chronic kidney disease stage 2 or greater and 22 patients had reached stage 5. Median renal survival was 43.3 years, including 15 kidney transplantations in 11 patients (1 combined with liver transplantation). Renal outcome did not correlate with genotype, biochemical parameters or initially present nephrocalcinosis. Thus, primary hyperoxaluria type 2 is a disease with significant morbidity. Accurate diagnosis by 24-hour urine analysis and genetic testing are required with careful follow-up. Copyright © 2019 International Society of Nephrology. All rights reserved.

**URL:** <https://ovidsp.ovid.com/ovidweb.cgi?T=JS&CSC=Y&NEWS=N&PAGE=fulltext&D=med16&DO=10.1016%2fj.kint.2019.08.018> <https://libkey.io/libraries/2592/openurl?genre=article&sid=OVID:medline&id=pmid:31685312&id=doi:10.1016%2Fj.kint.2019.08.018&issn=0085-2538&isbn=&volume=96&issue=6&spage=1389&pages=1389-1399&date=2019&title=Kidney+International&atitle=Patients+with+primary+hyperoxaluria+type+2+have+significant+morbidity+and+require+careful+follow-up.&aulast=Garrelfs&pid=%3Cauthor%3EGarrelfs+SF%3BRumsby+G%3BPeters-Sengers+H%3BErger+F%3BGroothoff+JW%3BBeck+BB%3BOosterveld+MJS%3BPelle+A%3BNeuhaus+T%3BAdams+B%3BCochat+P%3BSalido+E%3BLipkin+GW%3BHoppe+B%3BHulton+SA%3C%2Fauthor%3E%3CAN%3E31685312%3C%2FAN%3E%3CDT%3EJournal+Article%3C%2FDT%3E>

23. **Twenty-four-hour urine osmolality as a representative index of adequate hydration and a predictor of recurrence in patients with urolithiasis.**

**Item Type:** Journal Article

**Authors:** Kang, Ho Won;Seo, Sung Pil;Ha, Yun-Sok;Kim, Won Tae;Kim, Yong-June;Yun, Seok-Joong;Kim, Wun-Jae and Lee, Sang-Cheol

**Publication Date:** Ju ,2019

**Journal:** International Urology & Nephrology 51(7), pp. 1129–1135

**Abstract: PURPOSE**: To determine the value of 24-h urine osmolality (UOsm) as a representative index of adequate hydration and predictor of stone recurrence in patients with urolithiasis. **METHODS**: Medical records of consecutive patients presenting with renal or ureteric stones between 1994 and 2017 were retrospectively reviewed. Patients were grouped according to the results of 24-h UOsm (low : Medical records of consecutive patients presenting with renal or ureteric stones between 1994 and 2017 were retrospectively reviewed. Patients were grouped according to the results of 24-h UOsm (low _2_O, high > 564 mOsm/kg H_2_O). Metabolic parameters and risk of stone recurrence were compared between the two groups. **RESULTS**: The low urine concentration group were more likely to be older, to be female, and to have a lower body mass index and higher glomerular filtration rate than the high concentration group (each P : The low urine concentration group were more likely to be older, to be female, and to have a lower body mass index and higher glomerular filtration rate than the high concentration group (each P : The low urine concentration group were more likely to be older, to be female, and to have a lower body mass index and higher glomerular filtration rate than the high concentration group (each P **CONCLUSIONS**: UOsm is a promising approach to assessing hydration and predicting stone recurrence in patients with urolithiasis. Maintaining UOsm : UOsm is a promising approach to assessing hydration and predicting stone recurrence in patients with urolithiasis. Maintaining UOsm _2_O may reduce the risk of stone recurrence.

**URL:** <https://ovidsp.ovid.com/ovidweb.cgi?T=JS&CSC=Y&NEWS=N&PAGE=fulltext&D=med16&DO=10.1007%2fs11255-019-02108-2> <https://libkey.io/libraries/2592/openurl?genre=article&sid=OVID:medline&id=pmid:31089946&id=doi:10.1007%2Fs11255-019-02108-2&issn=0301-1623&isbn=&volume=51&issue=7&spage=1129&pages=1129-1135&date=2019&title=International+Urology+%26+Nephrology&atitle=Twenty-four-hour+urine+osmolality+as+a+representative+index+of+adequate+hydration+and+a+predictor+of+recurrence+in+patients+with+urolithiasis.&aulast=Kang&pid=%3Cauthor%3EKang+HW%3BSeo+SP%3BHa+YS%3BKim+WT%3BKim+YJ%3BYun+SJ%3BKim+WJ%3BLee+SC%3C%2Fauthor%3E%3CAN%3E31089946%3C%2FAN%3E%3CDT%3EJournal+Article%3C%2FDT%3E>

24. **Evaluation and Comparison of Metabolic Disorders between Patients with Unilateral and Bilateral Staghorn Renal Stones.**

**Item Type:** Journal Article

**Authors:** Mohammadi Sichani, Mehrdad;Jafarpisheh, Amir and Ghoreifi, Alireza

**Publication Date:** 2019

**Journal:** Urology Journal 16(3), pp. 242–245

**Abstract: PURPOSE**: Metabolic disorders are common in patients with staghorn renal stones. Aim of this study was to evaluate and compare the metabolic disorders in patients with unilateral and bilateral staghorn stones. **MATERIALS AND METHODS**: In this cross sectional study, 78 patients who underwent percutaneous nephrolithotomy (PCNL) for staghorn renal stones were included. The urine volume, the level of calcium, oxalate, uric acid, phos-phate, sodium, citrate, creatinine, and cystine from 24 hour urine collection as well as the serum levels of calcium, phosphorus, magnesium, creatinine, blood urea nitrogen (BUN), parathyroid hormone (PTH) and uric acid were recorded and compared among the two groups with unilateral and bilateral renal stones. **RESULTS**: 56 patients (71.8%) had unilateral and 22 (28.2%) had bilateral renal stones. At least one abnormal meta-bolic factor was found in 32 (57.1%) and 15 (68.2%) patients with unilateral and bilateral renal stones, respectively (P = .044). Cystine urine levels and serum levels of BUN were higher in cases with bilateral compared to unilateral renal stones (36.4% vs. 12.5%, P = .025 and 27.3% vs. 1.8%, P = .002, respectively). **CONCLUSION**: Metabolic factors are strongly correlated with the formation of staghorn renal stones specially bilat-eral ones. In our study among different metabolic factors, cystine urine levels and serum levels of BUN were sig-nificantly higher in patients with bilateral renal stones. Proper metabolic assessments are recommended in patients with staghorn urolithiasis.

**URL:** <https://ovidsp.ovid.com/ovidweb.cgi?T=JS&CSC=Y&NEWS=N&PAGE=fulltext&D=med16&DO=10.22037%2fuj.v0i0.4316> <https://libkey.io/libraries/2592/openurl?genre=article&sid=OVID:medline&id=pmid:30206923&id=doi:10.22037%2Fuj.v0i0.4316&issn=1735-1308&isbn=&volume=16&issue=3&spage=242&pages=242-245&date=2019&title=Urology+Journal&atitle=Evaluation+and+Comparison+of+Metabolic+Disorders+between+Patients+with+Unilateral+and+Bilateral+Staghorn+Renal+Stones.&aulast=Mohammadi+Sichani&pid=%3Cauthor%3EMohammadi+Sichani+M%3BJafarpisheh+A%3BGhoreifi+A%3C%2Fauthor%3E%3CAN%3E30206923%3C%2FAN%3E%3CDT%3EComparative+Study%3C%2FDT%3E>

25. **Renal Disorders in Pulmonary Sarcoidosis Patients.**

**Item Type:** Journal Article

**Authors:** Yassari, Fatemeh;Kiani, Arda;Taghavi, Kimia;Abdi, Ebrahim;Emami, Habib;Seifi, Sharareh and Abedini, Atefeh

**Publication Date:** Ja ,2019

**Journal:** Iranian Journal of Kidney Diseases 13(1), pp. 21–26

**Abstract: INTRODUCTION**: The frequency of kidney disorders varies in pulmonary sarcoidosis patients. Since the prevalence of kidney disorders among Iranian sarcoidosis patients is uncertain, this study aimed to evaluate kidney disorders and associated manifestations in Iranian pulmonary sarcoidosis patients. **MATERIALS AND METHODS**: One hundred patients with confirmed granuloma as pulmonary sarcoidosis were studied for renal disorders. Size of urinary tract and the presence of renal stones were checked via clinical examination and urinary organ ultrasonography. Patients' 24-hour urine sample was examined for pH, calcium, protein (over 250 mg) and creatinine (over 1.4 mg). **RESULTS**: Thirty-three percent of the patients expressed renal disorders simultaneously.Uric acid in pulmonary sarcoidosis patients could be correlated with the probability of developing renal stone. In addition, 1,25-dihydroxyvitamin D levels above 30 ng/mL and uric acid levels above 7 mg/dL in urine were directly correlated with renal disorders in sarcoidosis patients. **CONCLUSIONS**: Urinalysis is an easy and reliable method for assessing renal disorders in sarcoidosis patients. The current study proposes inclusion of urinalysis in routine checkups of sarcoidosis individuals.

**URL:** <https://ovidsp.ovid.com/ovidweb.cgi?T=JS&CSC=Y&NEWS=N&PAGE=fulltext&D=med16&AN=30851715> <https://libkey.io/libraries/2592/openurl?genre=article&sid=OVID:medline&id=pmid:30851715&id=doi:&issn=1735-8582&isbn=&volume=13&issue=1&spage=21&pages=21-26&date=2019&title=Iranian+journal+of+Kidney+Diseases&atitle=Renal+Disorders+in+Pulmonary+Sarcoidosis+Patients.&aulast=Yassari&pid=%3Cauthor%3EYassari+F%3BKiani+A%3BTaghavi+K%3BAbdi+E%3BEmami+H%3BSeifi+S%3BAbedini+A%3C%2Fauthor%3E%3CAN%3E30851715%3C%2FAN%3E%3CDT%3EJournal+Article%3C%2FDT%3E>

26. **Claudin-14 Gene Polymorphisms and Urine Calcium Excretion.**

**Item Type:** Journal Article

**Authors:** Arcidiacono, Teresa;Simonini, Marco;Lanzani, Chiara;Citterio, Lorena;Salvi, Erika;Barlassina, Cristina;Spotti, Donatella;Cusi, Daniele;Manunta, Paolo and Vezzoli, Giuseppe

**Publication Date:** 2018

**Journal:** Clinical Journal of the American Society of Nephrology: CJASN 13(10), pp. 1542–1549

**Abstract: BACKGROUND AND OBJECTIVES**: Claudin-16 and -19 are proteins forming pores for the paracellular reabsorption of divalent cations in the ascending limb of Henle loop; conversely, claudin-14 decreases ion permeability of these pores. Single-nucleotide polymorphisms in gene coding for claudin-14 were associated with kidney stones and calcium excretion. This study aimed to explore the association of claudin-14, claudin-16, and claudin-19 single-nucleotide polymorphisms with calcium excretion. **DESIGN, SETTING, PARTICIPANTS, & MEASUREMENTS**: We performed a retrospective observational study of 393 patients with hypertension who were naive to antihypertensive drugs, in whom we measured 24-hour urine calcium excretion; history of kidney stones was ascertained by interview; 370 of these patients underwent an intravenous 0.9% sodium chloride infusion (2 L in 2 hours) to evaluate the response of calcium excretion in three different 2-hour urine samples collected before, during, and after saline infusion. Genotypes of claudin-14, claudin-16, and claudin-19 were obtained from data of a previous genome-wide association study in the same patients. **RESULTS**: Thirty-one single-nucleotide polymorphisms of the 3' region of the claudin-14 gene were significantly associated with 24-hour calcium excretion and calcium excretion after saline infusion. The most significant associated single-nucleotide polymorphism was rs219755 (24-hour calcium excretion in GG, 225+/-124 mg/24 hours; 24-hour calcium excretion in GA, 194+/-100 mg/24 hours; 24-hour calcium excretion in AA, 124+/-73 mg/24 hours; P: Thirty-one single-nucleotide polymorphisms of the 3' region of the claudin-14 gene were significantly associated with 24-hour calcium excretion and calcium excretion after saline infusion. The most significant associated single-nucleotide polymorphism was rs219755 (24-hour calcium excretion in GG, 225+/-124 mg/24 hours; 24-hour calcium excretion in GA, 194+/-100 mg/24 hours; 24-hour calcium excretion in AA, 124+/-73 mg/24 hours; P**CONCLUSIONS**: Claudin-14 genotype at the 3' region is associated with calcium excretion in 24-hour urine and after the calciuretic stimulus of saline infusion. Copyright © 2018 by the American Society of Nephrology.

**URL:** <https://ovidsp.ovid.com/ovidweb.cgi?T=JS&CSC=Y&NEWS=N&PAGE=fulltext&D=med15&DO=10.2215%2fCJN.01770218> <https://libkey.io/libraries/2592/openurl?genre=article&sid=OVID:medline&id=pmid:30232134&id=doi:10.2215%2FCJN.01770218&issn=1555-9041&isbn=&volume=13&issue=10&spage=1542&pages=1542-1549&date=2018&title=Clinical+Journal+of+The+American+Society+of+Nephrology%3A+CJASN&atitle=Claudin-14+Gene+Polymorphisms+and+Urine+Calcium+Excretion.&aulast=Arcidiacono&pid=%3Cauthor%3EArcidiacono+T%3BSimonini+M%3BLanzani+C%3BCitterio+L%3BSalvi+E%3BBarlassina+C%3BSpotti+D%3BCusi+D%3BManunta+P%3BVezzoli+G%3C%2Fauthor%3E%3CAN%3E30232134%3C%2FAN%3E%3CDT%3EJournal+Article%3C%2FDT%3E>

27. **Impact of Potassium Citrate vs Citric Acid on Urinary Stone Risk in Calcium Phosphate Stone Formers.**

**Item Type:** Journal Article

**Authors:** Doizi, Steeve;Poindexter, John R.;Pearle, Margaret S.;Blanco, Francisco;Moe, Orson W.;Sakhaee, Khashayar and Maalouf, Naim M.

**Publication Date:** 2018

**Journal:** Journal of Urology 200(6), pp. 1278–1284

**Abstract: PURPOSE**: To our knowledge no medication has been shown to be effective for preventing recurrent calcium phosphate urinary stones. Potassium citrate may protect against calcium phosphate stones by enhancing urine citrate excretion and lowering urine calcium but it raises urine pH, which increases calcium phosphate saturation and may negate the beneficial effects. Citric acid can potentially raise urine citrate but not pH and, thus, it may be a useful countermeasure against calcium phosphate stones. We assessed whether these 2 agents could significantly alter urine composition and reduce calcium phosphate saturation. **MATERIALS AND METHODS**: In a crossover metabolic study 13 recurrent calcium phosphate stone formers without hypercalciuria were evaluated at the end of 3, 1-week study phases during which they consumed a fixed metabolic diet and received assigned study medications, including citric acid 30 mEq twice daily, potassium citrate 20 mEq twice daily or matching placebo. We collected 24-hour urine specimens to perform urine chemistry studies and calculate calcium phosphate saturation indexes. **RESULTS**: Urine parameters did not significantly differ between the citric acid and placebo phases. Potassium citrate significantly increased urine pH, potassium and citrate compared to citric acid and placebo (p : Urine parameters did not significantly differ between the citric acid and placebo phases. Potassium citrate significantly increased urine pH, potassium and citrate compared to citric acid and placebo (p **CONCLUSIONS**: Citric acid at a dose of 60 mEq per day did not significantly alter urine composition in calcium phosphate stone formers. The long-term impact of potassium citrate on calcium phosphate stone recurrence needs to be studied further. Copyright © 2018 American Urological Association Education and Research, Inc. Published by Elsevier Inc. All rights reserved.

**URL:** <https://ovidsp.ovid.com/ovidweb.cgi?T=JS&CSC=Y&NEWS=N&PAGE=fulltext&D=med15&DO=10.1016%2fj.juro.2018.07.039> <https://libkey.io/libraries/2592/openurl?genre=article&sid=OVID:medline&id=pmid:30036516&id=doi:10.1016%2Fj.juro.2018.07.039&issn=0022-5347&isbn=&volume=200&issue=6&spage=1278&pages=1278-1284&date=2018&title=Journal+of+Urology&atitle=Impact+of+Potassium+Citrate+vs+Citric+Acid+on+Urinary+Stone+Risk+in+Calcium+Phosphate+Stone+Formers.&aulast=Doizi&pid=%3Cauthor%3EDoizi+S%3BPoindexter+JR%3BPearle+MS%3BBlanco+F%3BMoe+OW%3BSakhaee+K%3BMaalouf+NM%3C%2Fauthor%3E%3CAN%3E30036516%3C%2FAN%3E%3CDT%3EJournal+Article%3C%2FDT%3E>

28. **A multiregional Italian cohort of 24-hour urine metabolic evaluation in renal stone formers.**

**Item Type:** Journal Article

**Authors:** Esperto, Francesco;Marangella, Martino;Trinchieri, Alberto;Petrarulo, Michele and Miano, Roberto

**Publication Date:** Fe ,2018

**Journal:** Minerva Urologica E Nefrologica 70(1), pp. 87–94

**Abstract: BACKGROUND**: Nephrolithiasis is a common condition with several studies documenting an increased prevalence over the past four decades. EAU and AUA guidelines recommend 24-hour urine metabolic evaluation in high-risk stone formers. Aim of this study is to retrospectively evaluate the first three years of experience with LithoTest R (Biohealth Italia Srl, Turin, Italy) through the analysis of demographic, clinical and biochemical data collected from a large cohort of patients with kidney stones. **METHODS**: We retrospectively analyzed data from the LithoCenter database, including data from outpatient consultations, between January 2007 and December 2009 from all over Italy. LithoTest R was performed through a 24-hour urine collection and included measurements of urine volume and pH, 24-hour excretion of creatinine as well as main cations and anions, including calcium, magnesium sodium potassium, ammonium, uric acid, oxalate, citrate, phosphate, inorganic sulphate and chloride. Urine state of saturation for calcium oxalate (betaCaOx), calcium hydrogen phosphate or brushite (betabsh) and uric acid (betaUA) were also calculated by means of the computer program LithoRisk. Brand's test for cystinuria was also carried out. Statistical analysis was performed using the S-PSS software v. 22.0. **RESULTS**: The number of patients with data available for analysis was 435, of whom 236 were male (54%) and 199 female (46%). Complete 24-hour urine measurements were available for all 435 patients. Compared to men, women had significantly lower values for creatinine, urate, oxalate, phosphate, sodium, potassium, magnesium and chloride excretion, whereas 24-hour pH and citrate excretion were higher. No significant differences were found for the other examined variables. betaCaOx and betaUA were significantly higher in men than women, whereas no significant difference was found for betabsh. There was a direct relationship between calcium and sodium urine excretion. Excessive sodium excretion was recorded in 191 patients (44%) and low urine volumes in 201 (46.2%). Hyperoxaluria was observed in 118 patients (27.3%), hypercalciuria in 115 (26.6%), hyperuricosuria in 153 (35.4%), hypomagnesuria in 96 (22.2%), and hypocitraturia in 134 patients (31%). Hyperexcretion of sodium, hypocitraturia and hyperoxaluria were most frequent in males. betaCaOx was significantly higher in the setting of hypercalciuria, hypocitraturia, hyperoxaluria and urine pH below 5.5. **CONCLUSIONS**: Our findings in a large cohort of high-risk stone-forming patients show significant differences in urinary metabolic profiles between men and women. Carrying on the collection and analysis of data by LithoTest R from 2009 to 2015 and matching urinary and dietary data could eventually improve our understanding on the metabolic profile of stone-formers in Italy.

**URL:** <https://ovidsp.ovid.com/ovidweb.cgi?T=JS&CSC=Y&NEWS=N&PAGE=fulltext&D=med15&DO=10.23736%2fS0393-2249.17.02961-7> <https://libkey.io/libraries/2592/openurl?genre=article&sid=OVID:medline&id=pmid:28882031&id=doi:10.23736%2FS0393-2249.17.02961-7&issn=0393-2249&isbn=&volume=70&issue=1&spage=87&pages=87-94&date=2018&title=Minerva+Urologica+e+Nefrologica&atitle=A+multiregional+Italian+cohort+of+24-hour+urine+metabolic+evaluation+in+renal+stone+formers.&aulast=Esperto&pid=%3Cauthor%3EEsperto+F%3BMarangella+M%3BTrinchieri+A%3BPetrarulo+M%3BMiano+R%3C%2Fauthor%3E%3CAN%3E28882031%3C%2FAN%3E%3CDT%3EJournal+Article%3C%2FDT%3E>

29. **Short-Term Changes in Urinary Relative Supersaturation Predict Recurrence of Kidney Stones: A Tool to Guide Preventive Measures in Urolithiasis.**

**Item Type:** Journal Article

**Authors:** Ferraro, Pietro Manuel;Ticinesi, Andrea;Meschi, Tiziana;Rodgers, Allen;Di Maio, Federica;Fulignati, Pierluigi;Borghi, Loris and Gambaro, Giovanni

**Publication Date:** 2018

**Journal:** Journal of Urology 200(5), pp. 1082–1087

**Abstract: PURPOSE**: Kidney stone disease is characterized by a relatively high rate of recurrence. In our study we analyzed the association between relative supersaturation and the risk of stone recurrence. Additionally, we examined the association between the risk of recurrence and changes in relative supersaturation and urinary composition after 1 week of medical treatment. **MATERIALS AND METHODS**: We performed a post hoc analysis of data from a previously published randomized controlled trial comparing the effect of 2 diets in 120 men with recurrent calcium oxalate stones and hypercalciuria. Baseline and followup 24-hour urine parameters were used to calculate the relative supersaturation of calcium oxalate, calcium phosphate and uric acid using the EQUIL2, JESS and LithoRisk computer programs. Cox models were used to calculate the estimated association between each baseline relative supersaturation, and 1-week changes and the risk of recurrence during followup. **RESULTS**: During a 5-year followup 35 patients (34%) experienced recurrence. A reduction in calcium oxalate relative supersaturation at 1 week was significantly associated with a lower risk of recurrence using the EQUIL2 calculation (for every 10% reduction from baseline HR 0.92, 95% CI 0.86-1.00, p = 0.044). However, there was no association for relative supersaturation calculated by other methods or for the relative supersaturation of other salts. Changes in the 24-hour urine excretion of citrate, potassium and magnesium were significantly associated with a risk of recurrence. **CONCLUSIONS**: In recurrent stone formers with hypercalciuria baseline values and changes in the relative supersaturation of calcium oxalate may be associated with the risk of recurrence. Changes in urinary citrate, potassium and magnesium following dietary intervention may also be predictive. Copyright © 2018 American Urological Association Education and Research, Inc. Published by Elsevier Inc. All rights reserved.

**URL:** <https://ovidsp.ovid.com/ovidweb.cgi?T=JS&CSC=Y&NEWS=N&PAGE=fulltext&D=med15&DO=10.1016%2fj.juro.2018.06.029> <https://libkey.io/libraries/2592/openurl?genre=article&sid=OVID:medline&id=pmid:29940247&id=doi:10.1016%2Fj.juro.2018.06.029&issn=0022-5347&isbn=&volume=200&issue=5&spage=1082&pages=1082-1087&date=2018&title=Journal+of+Urology&atitle=Short-Term+Changes+in+Urinary+Relative+Supersaturation+Predict+Recurrence+of+Kidney+Stones%3A+A+Tool+to+Guide+Preventive+Measures+in+Urolithiasis.&aulast=Ferraro&pid=%3Cauthor%3EFerraro+PM%3BTicinesi+A%3BMeschi+T%3BRodgers+A%3BDi+Maio+F%3BFulignati+P%3BBorghi+L%3BGambaro+G%3C%2Fauthor%3E%3CAN%3E29940247%3C%2FAN%3E%3CDT%3EEvaluation+Study%3C%2FDT%3E>

30. **Study of Urine Composition of Patients With Recurrent Nephrolithiasis in Lorestan, Iran.**

**Item Type:** Journal Article

**Authors:** Hadian, Babak;Zafar-Mohtashami, Azita and Ghorbani, Fershteh

**Publication Date:** 2018

**Journal:** Iranian Journal of Kidney Diseases 12(1), pp. 22–26

**Abstract: INTRODUCTION**: Nephrolithiasis is one of the most common urinary tract diseases. After the first episode of urinary calculus, the risk of recurrence is nearly 40% to 50% at 5 years. Nephrolithiasis is a systemic disease that is associated with some metabolic disorders. This study aimed to provide a picture of the frequency of metabolic abnormalities in patients with nephrolithiasis from west part of Iran. **MATERIALS AND METHODS**: Patients with recurrent urinary tract calculi referred to the Nephrology-Urology Clinics in Khorramabad city were recruited. After collection of demographic data of all the patients, 24-hour urine and blood samples were taken to measure biochemical factors. **RESULTS**: Of the 232 participants, 125 were males and 107 were females. Hyperoxaluria was seen in 93 (40.1%) of the participants, hypercalciuria in 55 (23.7%), hypocitraturia in 58 (25%), and hyperuricosuria in 33 (14.9%). Hyperoxaluria in the males was significantly more frequent than in the female patients. There were no significant differences between the two groups in other urinary metabolic disorders. **CONCLUSIONS**: Patients with nephrolithiasis from Lorestan province may have metabolic characteristics varying from those of regions; ethnicity may be a possible reason. Variation of dietary regimens, such as the amount of meat or vegetable in the diet that can change oxalate, calcium, or citrate of urine, might have influenced the results. Time of sampling is another factor. Population-specific studies are helpful to health care providers for preventive planning for nephrolithiasis.

**URL:** <https://ovidsp.ovid.com/ovidweb.cgi?T=JS&CSC=Y&NEWS=N&PAGE=fulltext&D=med15&AN=29421773> <https://libkey.io/libraries/2592/openurl?genre=article&sid=OVID:medline&id=pmid:29421773&id=doi:&issn=1735-8582&isbn=&volume=12&issue=1&spage=22&pages=22-26&date=2018&title=Iranian+journal+of+Kidney+Diseases&atitle=Study+of+Urine+Composition+of+Patients+With+Recurrent+Nephrolithiasis+in+Lorestan%2C+Iran.&aulast=Hadian&pid=%3Cauthor%3EHadian+B%3BZafar-Mohtashami+A%3BGhorbani+F%3C%2Fauthor%3E%3CAN%3E29421773%3C%2FAN%3E%3CDT%3EJournal+Article%3C%2FDT%3E>

31. **Urine kidney injury markers do not increase following gastric bypass: a multi-center cross-sectional study.**

**Item Type:** Journal Article

**Authors:** Hinck, Bryan D.;Miyaoka, Ricardo;Lingeman, James E.;Assimos, Dean G.;Matlaga, Brian R.;Pramanik, Rocky;Asplin, John;Cohen, Benjamin and Monga, Manoj

**Publication Date:** 2018

**Journal:** Canadian Journal of Urology 25(1), pp. 9199–9204

**Abstract: INTRODUCTION**: To determine if markers of kidney injury correlate with urinary oxalate excretion. If so, such biomarkers might be early predictors of oxalate nephropathy. Gastric bypass surgery for obesity is known to be associated with postoperative hyperoxaluria, which can lead to urolithiasis and kidney damage. **MATERIALS AND METHODS**: Patients were recruited from four large academic centers > 6 months following completion of gastric bypass surgery. Patients provided a spot urine sample for analysis of three markers of kidney injury: 8-iso-Prostaglandin F2 alpha, N-acetyl- beta -D-Glucosaminidase, and Neutrophil gelatinase-associated lipocalin. Patients also provided 24 hour urine samples for stone risk analysis. **RESULTS**: A total of 46 study patients provided samples, the average age was 48.4 +/- 11.3. There were 40 women and 6 men. There was no difference in the level of any of the three inflammatory markers between the study group and the reference range generated from healthy non-hyperoxaluric subjects. Neither oxalate excretion nor supersaturation of calcium oxalate correlated with any of the injury markers. There was no difference noted between those with hyperoxaluria (n = 17) and those with normoxaluria (n = 29) with respect to any of the injury markers. **CONCLUSIONS**: Though hyperoxaluria was common after bypass surgery, markers of kidney injury were not elevated after surgery. No correlation was found between urine oxalate excretion and any of the injury markers.

**URL:** <https://ovidsp.ovid.com/ovidweb.cgi?T=JS&CSC=Y&NEWS=N&PAGE=fulltext&D=med15&AN=29524975> <https://libkey.io/libraries/2592/openurl?genre=article&sid=OVID:medline&id=pmid:29524975&id=doi:&issn=1195-9479&isbn=&volume=25&issue=1&spage=9199&pages=9199-9204&date=2018&title=Canadian+Journal+of+Urology&atitle=Urine+kidney+injury+markers+do+not+increase+following+gastric+bypass%3A+a+multi-center+cross-sectional+study.&aulast=Hinck&pid=%3Cauthor%3EHinck+BD%3BMiyaoka+R%3BLingeman+JE%3BAssimos+DG%3BMatlaga+BR%3BPramanik+R%3BAsplin+J%3BCohen+B%3BMonga+M%3C%2Fauthor%3E%3CAN%3E29524975%3C%2FAN%3E%3CDT%3EComparative+Study%3C%2FDT%3E>

32. **Etiopathogenesis of Nephrolithiasis in Ulcerative Colitis Patients with the Ileal Pouch Anal Anastomosis.**

**Item Type:** Journal Article

**Authors:** Arora, Zubin;Mukewar, Saurabh;Lopez, Rocio;Camino, Debra;Shen, Bo and Hall, Phillip

**Publication Date:** 2017

**Journal:** Inflammatory Bowel Diseases 23(5), pp. 840–846

**Abstract: BACKGROUND**: Our previous study showed that nephrolithiasis is a common complication in ulcerative colitis patients after ileal pouch anal anastomosis (IPAA). However, the pathogenesis of nephrolithiasis in IPAA patients has not been studied. The aim of this study was to compare urine and serum metabolic compositions in IPAA patients with nephrolithiasis and controls with IPAA and no nephrolithiasis. **METHODS**: Using cross-sectional study design, serum and 24-hour urine metabolic compositions were compared between IPAA patients with nephrolithiasis (the study group) and those without (the control group). Urinary supersaturation of calcium oxalate, calcium phosphate, and uric acid was calculated. **RESULTS**: A total of 40 patients were enrolled in the study. There were no significant differences in serum electrolytes, vitamin D, parathyroid hormone, and kidney function tests between the study (n = 20) and control groups (n = 20). Patients in the study group were found to have a significantly higher 24-hour urine supersaturation of calcium oxalate (8.8 versus 5.0, P = 0.037) and calcium phosphate (0.61 versus 0.27, P = 0.028) as compared with controls. Nineteen (95%) patients in the study group were symptomatic due to nephrolithiasis with several requiring procedural intervention for treatment, including ureteroscopy in 3 (15%) patients, lithotripsy in 5 (25%) patients, and percutaneous surgery in 1 (5%) patient. **CONCLUSIONS**: Ulcerative colitis-IPAA patients are at risk for the development of calcium oxalate and calcium phosphate stones. Nephrolithiasis is symptomatic in a majority of the patients and frequently requires procedural intervention for treatment.

**URL:** <https://ovidsp.ovid.com/ovidweb.cgi?T=JS&CSC=Y&NEWS=N&PAGE=fulltext&D=med14&DO=10.1097%2fMIB.0000000000001070> <https://libkey.io/libraries/2592/openurl?genre=article&sid=OVID:medline&id=pmid:28301430&id=doi:10.1097%2FMIB.0000000000001070&issn=1078-0998&isbn=&volume=23&issue=5&spage=840&pages=840-846&date=2017&title=Inflammatory+Bowel+Diseases&atitle=Etiopathogenesis+of+Nephrolithiasis+in+Ulcerative+Colitis+Patients+with+the+Ileal+Pouch+Anal+Anastomosis.&aulast=Arora&pid=%3Cauthor%3EArora+Z%3BMukewar+S%3BLopez+R%3BCamino+D%3BShen+B%3BHall+P%3C%2Fauthor%3E%3CAN%3E28301430%3C%2FAN%3E%3CDT%3EJournal+Article%3C%2FDT%3E>

33. **Influence of weight status on 24-hour urine composition in adults without urolithiasis: A nationwide study based on a Chinese Han population.**

**Item Type:** Journal Article

**Authors:** Deng, Tuo;Mai, Zanlin;Cai, Chao;Duan, Xiaolu;Zhu, Wei;Zhang, Tao;Wu, Wenqi and Zeng, Guohua

**Publication Date:** 2017

**Journal:** PLoS ONE [Electronic Resource] 12(9), pp. e0184655

**Abstract: OBJECTIVES**: This study sought to explore the influence of different body weight statuses on 24-hour urine compositions in adults without urolithiasis based on a nationwide study of a Chinese Han population. **MATERIAL AND METHODS**: Twenty-four-hour urine samples from 584 Chinese Han adults without urolithiasis in six cities were analyzed. The participants were divided into four body weight status types according to their body mass indices (BMIs) according to WHO guidelines. The baseline characteristics and 24-hour urine compositions of the standard weight group were compared with those of the underweight, overweight and obese groups. The influences of different body weight statuses on the 24-hour urine compositions were explored using univariate and multivariate logistic regressions. **RESULTS**: The numbers of participants in the underweight, standard weight, overweight and obese status groups were 24, 376, 149 and 35, respectively. The overweight and obese groups suffered significantly higher risks of hypertension and diabetes mellitus than the standard weight group. In the univariate analyses, compared with the standard weight group, the overweight group had significantly higher levels of urine citrate (mean difference [MD] = 0.51 mmol, 95% confidence interval [CI]: 0.15-0.87, P = 0.001), potassium (MD = 6.63 mmol, 95% CI: 1.13-12.14, P = 0.01) and magnesium (MD = 0.38 mmol, 95% CI: 0.08-0.69, P = 0.014). Significant increases in urine citrate (MD = 0.85 mmol, 95% CI: 0.01-1.68, P = 0.046), magnesium (MD = 0.69 mmol, 95% CI: 0.13-1.25, P = 0.016) and phosphate (MD = 2.28 mmol, 95% CI: 0.03-4.54, P = 0.047) were found in the obese group. No significant differences were detected between the standard weight and underweight groups. In the multivariate logistic regression analyses, we only observed significantly higher levels of urine potassium (odds ratio [OR] = 1.02, 95% CI: 1.00-1.04, P = 0.03) in the overweight group and phosphate (OR = 1.32, 95% CI: 1.05-1.66, P = 0.018) in the obese group when compared with the standard weight group. **CONCLUSIONS**: Nonstone-forming adults with overweight or obese statuses were at higher risks of hypertension and diabetes mellitus. Obese nonstone-formers might have a greater risk of urinary stone formation due to increased urinary phosphate excretion. Additionally, underweight status had no influence on 24-hour urine composition.

**URL:** <https://ovidsp.ovid.com/ovidweb.cgi?T=JS&CSC=Y&NEWS=N&PAGE=fulltext&D=med14&DO=10.1371%2fjournal.pone.0184655> <https://libkey.io/libraries/2592/openurl?genre=article&sid=OVID:medline&id=pmid:28886192&id=doi:10.1371%2Fjournal.pone.0184655&issn=1932-6203&isbn=&volume=12&issue=9&spage=e0184655&pages=e0184655&date=2017&title=PLoS+ONE+%5BElectronic+Resource%5D&atitle=Influence+of+weight+status+on+24-hour+urine+composition+in+adults+without+urolithiasis%3A+A+nationwide+study+based+on+a+Chinese+Han+population.&aulast=Deng&pid=%3Cauthor%3EDeng+T%3BMai+Z%3BCai+C%3BDuan+X%3BZhu+W%3BZhang+T%3BWu+W%3BZeng+G%3C%2Fauthor%3E%3CAN%3E28886192%3C%2FAN%3E%3CDT%3EJournal+Article%3C%2FDT%3E>

34. **Analyte variations in consecutive 24-hour urine collections in children.**

**Item Type:** Journal Article

**Authors:** Ellison, Jonathan S.;Hollingsworth, John M.;Langman, Craig B.;Asplin, John R.;Schwaderer, Andrew L.;Yan, Phyllis;Bierlein, Maggie;Barraza, Mark A.;Defoor, William R.;Figueroa, T. Ernesto;Jackson, Elizabeth C.;Jayanthi, Venkata R.;Johnson, Emilie K.;Joseph, David B. and Shnorhavorian, Margarett

**Publication Date:** De ,2017

**Journal:** Journal of Pediatric Urology 13(6), pp. 632.e1–632.e7

**Abstract: PURPOSE**: The metabolic evaluation of children with nephrolithiasis begins with a 24-h urine collection. For adults, the diagnostic yield increases with consecutive collections; however, little is known regarding the variability of multiple 24-h studies in the pediatric population. We sought to evaluate the variability of consecutive 24-h urine collection in children through a multi-institutional study hypothesizing that compared with a single collection, consecutive 24-h urine collections would reveal a greater degree of clinically useful information in the evaluation of children at risk for nephrolithiasis. **MATERIALS AND METHODS**: Including data from six institutions, we identified children less than 18 years of age considered at risk for recurrent nephrolithiasis, undergoing metabolic evaluation. We evaluated a subset of patients performing two collections with urine creatinine varying by 10% or less during a 7-day period. Discordance between repeat collections based on normative urine chemistry values was evaluated. **RESULTS**: A total of 733 children met inclusion criteria, and in over a third both urine calcium and urine volume differed by 30% or more between samples. Urine oxalate demonstrated greater variation between collections in children =30% and 50%, respectively, are shown in the table. **CONCLUSIONS**: Our analysis indicates that stone risk in as many as one in four children may be misclassified if normative values of only a single 24-h urine are used. In light of these findings, repeat 24-h urine collections prior to targeted intervention to modify stone risk are advised to increase diagnostic yield in children at risk for nephrolithiasis. Copyright © 2017 Journal of Pediatric Urology Company. Published by Elsevier Ltd. All rights reserved.

**URL:** <https://ovidsp.ovid.com/ovidweb.cgi?T=JS&CSC=Y&NEWS=N&PAGE=fulltext&D=med14&DO=10.1016%2fj.jpurol.2017.06.014> <https://libkey.io/libraries/2592/openurl?genre=article&sid=OVID:medline&id=pmid:28739373&id=doi:10.1016%2Fj.jpurol.2017.06.014&issn=1477-5131&isbn=&volume=13&issue=6&spage=632.e1&pages=632.e1-632.e7&date=2017&title=Journal+of+pediatric+urology&atitle=Analyte+variations+in+consecutive+24-hour+urine+collections+in+children.&aulast=Ellison&pid=%3Cauthor%3EEllison+JS%3BHollingsworth+JM%3BLangman+CB%3BAsplin+JR%3BSchwaderer+AL%3BYan+P%3BBierlein+M%3BBarraza+MA%3BDefoor+WR%3BFigueroa+TE%3BJackson+EC%3BJayanthi+VR%3BJohnson+EK%3BJoseph+DB%3BShnorhavorian+M%3C%2Fauthor%3E%3CAN%3E28739373%3C%2FAN%3E%3CDT%3EJournal+Article%3C%2FDT%3E>

35. **Can a Simplified 12-Hour Nighttime Urine Collection Predict Urinary Stone Risk?.**

**Item Type:** Journal Article

**Authors:** Hinck, Bryan D.;Ganesan, Vishnuvardhan;Tarplin, Sarah;Asplin, John;Granja, Ignacio;Calle, Juan;Sivalingam, Sri and Monga, Manoj

**Publication Date:** Oc ,2017

**Journal:** Urology 108, pp. 40–45

**Abstract: OBJECTIVE**: To determine if there is correlation between nighttime 12-hour and traditional 24-hour urine collection in regard to chemistry values and the supersaturations of calcium oxalate, calcium phosphate, and uric acid for the metabolic evaluation of nephrolithiasis. **MATERIALS AND METHODS**: Ninety-five patients were prospectively enrolled from 2013 to 2015. Patients >18 years of age who presented to a tertiary stone clinic and who would normally be counseled for 24-hour urine collection were eligible for the study. Participants completed 24-hour urine collections twice, with each divided into 2 separate 12-hour collections. Day-time collection began after the first morning void and continued for 12 hours. The night collection proceeded for the next 12 hours through the first morning void. **RESULTS**: Forty-nine 24-hour samples from 35 patients met inclusion criteria and were included in the analysis. Overall, there was strong correlation between the night 12-hour and the 24-hour urine collections with R^2^ ranging from 0.76 for pH to 0.96 for Citrate. In our analysis of variability, the nighttime 12-hour collection differed from the 24-hour collection by 30% in 1-9 patients (2.0%-18.4%) based on individual chemistry value. Diagnosis of underlying metabolic abnormalities was concordant in 92% of patients. **CONCLUSION**: A 12-hour nighttime collection has strong correlation with 24-hour urine collection. As such, simplifying the metabolic evaluation to a 12-hour overnight collection may be feasible-improving compliance and decreasing patient burden. Copyright © 2017 Elsevier Inc. All rights reserved.

**URL:** <https://ovidsp.ovid.com/ovidweb.cgi?T=JS&CSC=Y&NEWS=N&PAGE=fulltext&D=med14&DO=10.1016%2fj.urology.2017.06.028> <https://libkey.io/libraries/2592/openurl?genre=article&sid=OVID:medline&id=pmid:28705575&id=doi:10.1016%2Fj.urology.2017.06.028&issn=0090-4295&isbn=&volume=108&issue=&spage=40&pages=40-45&date=2017&title=Urology&atitle=Can+a+Simplified+12-Hour+Nighttime+Urine+Collection+Predict+Urinary+Stone+Risk%3F.&aulast=Hinck&pid=%3Cauthor%3EHinck+BD%3BGanesan+V%3BTarplin+S%3BAsplin+J%3BGranja+I%3BCalle+J%3BSivalingam+S%3BMonga+M%3C%2Fauthor%3E%3CAN%3E28705575%3C%2FAN%3E%3CDT%3EJournal+Article%3C%2FDT%3E>

36. **Medical and dietary interventions for preventing recurrent urinary stones in children**

**Item Type:** Journal Article

**Authors:** Kern, Adam;Grimsby, Gwen;Mayo, Helen and Baker, Linda A.

**Publication Date:** Nov 09 ,2017

**Journal:** Cochrane Database of Systematic Reviews 11, pp. CD011252

**Abstract: BACKGROUND**: Nephrolithiasis, or urinary stone disease, in children causes significant morbidity, and is increasing in prevalence in the North American population. Therefore, medical and dietary interventions (MDI) for recurrent urinary stones in children are poised to gain increasing importance in the clinical armamentarium. **OBJECTIVES**: To assess the effects of medical and dietary interventions (MDI) for the prevention of idiopathic urinary stones in children aged from one to 18 years. **SEARCH METHODS**: We searched multiple databases using search terms relevant to this review, including studies identified from the Cochrane Central Register of Controlled Trials (CENTRAL, 2017, Issue 1), MEDLINE OvidSP (1946 to 14 February 2017), Embase OvidSP (1980 to 14 February 2017), International Clinical Trials Register (ICTRP) Search Portal and ClinicalTrials.gov. Additionally, we handsearched renal-related journals and the proceedings of major renal conferences, and reviewed weekly current awareness alerts for selected renal journals. The date of the last search was 14 February 2017. There were no language restrictions. **SELECTION CRITERIA**: Randomized controlled trials of at least one year of MDI versus control for prevention of recurrent idiopathic (non-syndromic) nephrolithiasis in children. **DATA COLLECTION AND ANALYSIS**: We used standard methodologic procedures expected by Cochrane. Titles and abstracts were identified by search criteria and then screened for relevance, and then data extraction and risk of bias assessment were carried out. We assessed the quality of evidence using GRADE. **MAIN RESULTS**: The search identified one study of 125 children (72 boys and 53 girls) with calcium-containing idiopathic nephrolithiasis and normal renal morphology following initial treatment with shockwave lithotripsy (SWL). Patients were randomized to oral potassium citrate 1 mEq/kg per day for 12 months versus no specific medication or preventive measure with results reported for a total of 96 patients (48 per group). This included children who were stone-free (n = 52) or had residual stone fragments (n = 44) following SWL. Primary outcomes:Medical therapy may lower rates of stone recurrence with a risk ratio (RR) of 0.19 (95% confidence interval (CI) 0.06 to 0.60; low quality evidence). This corresponds to 270 fewer stone recurrences per 1000 (133 fewer to 313 fewer) children. We downgraded the quality of evidence by two levels for very serious study limitations related to unclear allocation concealment (selection bias) and a high risk of performance, detection and attrition bias. While the data for adverse events were incomplete, they reported that six of 48 (12.5%) children receiving potassium citrate left the trial because of adverse effects. This corresponds to a RR of 13.0 (95% CI 0.75 to 224.53; very low quality evidence); an absolute effect size estimate could not be generated. We downgraded the quality of evidence for study limitations and imprecision. We found no information on retreatment rates. **SECONDARY OUTCOMES**: We found no evidence on serum electrolytes, 24-hour urine collection parameters or time to new stone formation. We were unable to perform any preplanned secondary analyses. **AUTHORS' CONCLUSIONS**: Oral potassium citrate supplementation may reduce recurrent calcium urinary stone formation in children following SWL; however, our confidence in this finding is limited. A substantial number of children stopped the medication due to adverse events. There is no trial evidence on retreatment rates. There is a critical need for additional well-designed trials in children with nephrolithiasis.

**URL:** <https://ovidsp.ovid.com/ovidweb.cgi?T=JS&CSC=Y&NEWS=N&PAGE=fulltext&D=med14&DO=10.1002%2f14651858.CD011252.pub2> <https://libkey.io/libraries/2592/openurl?genre=article&sid=OVID:medline&id=pmid:29117629&id=doi:10.1002%2F14651858.CD011252.pub2&issn=1361-6137&isbn=&volume=11&issue=11&spage=CD011252&pages=CD011252&date=2017&title=Cochrane+Database+of+Systematic+Reviews&atitle=Medical+and+dietary+interventions+for+preventing+recurrent+urinary+stones+in+children.&aulast=Kern&pid=%3Cauthor%3EKern+A%3BGrimsby+G%3BMayo+H%3BBaker+LA%3C%2Fauthor%3E%3CAN%3E29117629%3C%2FAN%3E%3CDT%3EJournal+Article%3C%2FDT%3E>

37. **Integrative Analysis of miRNA and mRNA Expression Profiles in Calcium Oxalate Nephrolithiasis Rat Model.**

**Item Type:** Journal Article

**Authors:** Lan, Chuangxin;Chen, Dong;Liang, Xiongfa;Huang, Jian;Zeng, Tao;Duan, Xiaolu;Chen, Kang;Lai, Yongchang;Yang, Dong;Li, Shujue;Jiang, Chonghe and Wu, Wenqi

**Publication Date:** 2017

**Journal:** BioMed Research International 2017, pp. 8306736

**Abstract:** The microRNA (miRNA) expression profiles and their biological functions in calcium oxalate nephrolithiasis remain unclear. In this study, we investigate the miRNA and mRNA expression profiles of kidney tissues in calcium oxalate stone rats. 16 Sprague Dawley rats were divided into control group and stone-forming group. 24-hour urine samples and kidney tissues were collected for biochemical and histological determination after 4 weeks. MiRNA and mRNA microarray were applied to evaluate the miRNA and mRNA expression profiles. To validate the microarray results, the quantitative real-time PCR (qRT-PCR) was performed. A total of 38 miRNAs and 2728 mRNAs were significantly and differentially expressed in kidney tissues of stone-forming group versus control group. Gene Ontology (GO) analysis revealed that most of the target genes were enriched in terms of oxidation reduction, ion transport, inflammatory response, and response to wounding. Kyoto Encyclopedia of Genes and Genomes (KEGG) pathway analysis of these targets highlights their critical role in cytokine-cytokine receptor interaction, gap junction, and chemokine signaling pathway. Furthermore, the reliability of the microarray-based results was confirmed by using qRT-PCR determination. The miRNA and mRNA expressions in calcium oxalate stone rat kidneys might provide a basis for further research on urolithiasis mechanism.

**URL:** <https://ovidsp.ovid.com/ovidweb.cgi?T=JS&CSC=Y&NEWS=N&PAGE=fulltext&D=med14&DO=10.1155%2f2017%2f8306736> <https://libkey.io/libraries/2592/openurl?genre=article&sid=OVID:medline&id=pmid:29392139&id=doi:10.1155%2F2017%2F8306736&issn=2314-6141&isbn=&volume=2017&issue=&spage=8306736&pages=8306736&date=2017&title=BioMed+Research+International&atitle=Integrative+Analysis+of+miRNA+and+mRNA+Expression+Profiles+in+Calcium+Oxalate+Nephrolithiasis+Rat+Model.&aulast=Lan&pid=%3Cauthor%3ELan+C%3BChen+D%3BLiang+X%3BHuang+J%3BZeng+T%3BDuan+X%3BChen+K%3BLai+Y%3BYang+D%3BLi+S%3BJiang+C%3BWu+W%3C%2Fauthor%3E%3CAN%3E29392139%3C%2FAN%3E%3CDT%3EJournal+Article%3C%2FDT%3E>

38. **Age, Body Mass Index, and Gender Predict 24-Hour Urine Parameters in Recurrent Idiopathic Calcium Oxalate Stone Formers.**

**Item Type:** Journal Article

**Authors:** Otto, Brandon J.;Bozorgmehri, Shahab;Kuo, Jennifer;Canales, Muna;Bird, Vincent G. and Canales, Benjamin

**Publication Date:** De ,2017

**Journal:** Journal of Endourology 31(12), pp. 1335–1341

**Abstract: INTRODUCTION AND OBJECTIVE**: Despite guidelines, routine 24-hour urine testing is completed in : Despite guidelines, routine 24-hour urine testing is completed in **METHODS**: We performed a retrospective study of 392 consecutive patients from 2007 to 2014 with >=2 lifetime stone episodes, >70% calcium oxalate by mineral analysis, and >=1 24-hour urine collection. We compared mean 24-hour urine values by age in decades. We used logistic regression and receiver operating characteristic (ROC) curve analysis to assess the predictive ability of age, gender, body mass index (BMI), and comorbidities to detect abnormal 24-hour urine parameters. **RESULTS**: The mean age of the cohort was 51 +/- 16 years. Older age was associated with greater urinary oxalate (p-trend : The mean age of the cohort was 51 +/- 16 years. Older age was associated with greater urinary oxalate (p-trend : The mean age of the cohort was 51 +/- 16 years. Older age was associated with greater urinary oxalate (p-trend **CONCLUSIONS**: In our recurrent calcium oxalate cohort, age significantly impacted urinary calcium, oxalate, citrate, and pH. Along with gender and BMI, age can be used to predict key 24-hour urine stone risk results. These data lay the foundation for a risk prediction tool, which could be a surrogate for 24-hour urine results in recurrent stone formers, who are unwilling or unable to complete metabolic testing. Further validation of these findings is needed in other stone populations.

**URL:** <https://ovidsp.ovid.com/ovidweb.cgi?T=JS&CSC=Y&NEWS=N&PAGE=fulltext&D=med14&DO=10.1089%2fend.2017.0352> <https://libkey.io/libraries/2592/openurl?genre=article&sid=OVID:medline&id=pmid:29084490&id=doi:10.1089%2Fend.2017.0352&issn=0892-7790&isbn=&volume=31&issue=12&spage=1335&pages=1335-1341&date=2017&title=Journal+of+Endourology&atitle=Age%2C+Body+Mass+Index%2C+and+Gender+Predict+24-Hour+Urine+Parameters+in+Recurrent+Idiopathic+Calcium+Oxalate+Stone+Formers.&aulast=Otto&pid=%3Cauthor%3EOtto+BJ%3BBozorgmehri+S%3BKuo+J%3BCanales+M%3BBird+VG%3BCanales+B%3C%2Fauthor%3E%3CAN%3E29084490%3C%2FAN%3E%3CDT%3EJournal+Article%3C%2FDT%3E>

39. **Vascular Disease and Kidney Stones: Abdominal Aortic Calcifications Are Associated with Low Urine pH and Hypocitraturia.**

**Item Type:** Journal Article

**Authors:** Patel, Nishant D.;Ward, Ryan D.;Calle, Juan;Remer, Erick M. and Monga, Manoj

**Publication Date:** 2017

**Journal:** Journal of Endourology 31(9), pp. 956–961

**Abstract: OBJECTIVE**: Vascular calcifications are associated with nephrolithiasis. Although studies have demonstrated correlations with vascular disease and calcium stones in kidney stone formers (KSF), an etiologic link has remained elusive. As a noncontrast CT (NCCT) scan is typically part of a stone evaluation, our objective was to evaluate the association of NCCT-based assessment of abdominal aortic calcifications (AACs) with 24-hour urine parameters and stone composition. **MATERIALS AND METHODS**: Ninety-seven KSF were included with CT imaging and 24-hour urine studies. For each patient, semi-automated CT software was utilized to provide an AAC Agatston score from the celiac axis to the aortic bifurcation. Univariate analysis was performed to compare patients with or without AAC. Multivariate logistic regression was performed to assess for variables associated with 24-hour urine parameters and stone composition. **RESULTS**: The presence of AAC was associated with hypertension, diabetes, peripheral vascular disease, and coronary artery disease. Patients with any AAC showed lower 24-hour urine citrate (399 vs 593 mg/day, p : The presence of AAC was associated with hypertension, diabetes, peripheral vascular disease, and coronary artery disease. Patients with any AAC showed lower 24-hour urine citrate (399 vs 593 mg/day, p : The presence of AAC was associated with hypertension, diabetes, peripheral vascular disease, and coronary artery disease. Patients with any AAC showed lower 24-hour urine citrate (399 vs 593 mg/day, p : The presence of AAC was associated with hypertension, diabetes, peripheral vascular disease, and coronary artery disease. Patients with any AAC showed lower 24-hour urine citrate (399 vs 593 mg/day, p **CONCLUSIONS**: NCCT-based diagnosis of AAC is associated with low urine pH, hypocitraturia, and uric acid stone formation. The presence of AAC could be considered an additional prognosticator for the utility of alkalinization therapy.

**URL:** <https://ovidsp.ovid.com/ovidweb.cgi?T=JS&CSC=Y&NEWS=N&PAGE=fulltext&D=med14&DO=10.1089%2fend.2017.0350> <https://libkey.io/libraries/2592/openurl?genre=article&sid=OVID:medline&id=pmid:28605936&id=doi:10.1089%2Fend.2017.0350&issn=0892-7790&isbn=&volume=31&issue=9&spage=956&pages=956-961&date=2017&title=Journal+of+Endourology&atitle=Vascular+Disease+and+Kidney+Stones%3A+Abdominal+Aortic+Calcifications+Are+Associated+with+Low+Urine+pH+and+Hypocitraturia.&aulast=Patel&pid=%3Cauthor%3EPatel+ND%3BWard+RD%3BCalle+J%3BRemer+EM%3BMonga+M%3C%2Fauthor%3E%3CAN%3E28605936%3C%2FAN%3E%3CDT%3EJournal+Article%3C%2FDT%3E>

40. **Association of urinary citrate excretion, pH, and net gastrointestinal alkali absorption with diet, diuretic use, and blood glucose concentration.**

**Item Type:** Journal Article

**Authors:** Perinpam, Majuran;Ware, Erin B.;Smith, Jennifer A.;Turner, Stephen T.;Kardia, Sharon L. R. and Lieske, John C.

**Publication Date:** Oc ,2017

**Journal:** Physiological Reports 5(19)
**Abstract:** Urinary citrate (Ucit) protects against urinary stone formation. Acid base status and diet influence Ucit. However, the effect of demographics, diet, and glucose metabolism on Ucit excretion, urinary pH (U-pH) and net gastrointestinal alkali absorption (NAA) are not known. Twenty-four hour urine samples, blood glucose, creatinine, and cystatin C were obtained from non-Hispanic white sibships in Rochester, MN (n = 446; 64.5 +/- 9 years; 58% female). Diet was assessed by a food frequency questionnaire. The impact of blood glucose, demographics and dietary elements on Ucit excretion, U-pH, and NAA were evaluated in bivariate and multivariable models and interaction models that included age, sex, and weight. NAA significantly associated with Ucit and U-pH In multivariate models Ucit increased with age, weight, eGFR_Cys_, and blood glucose, but decreased with loop diuretic and thiazide use. U-pH decreased with serum creatinine, blood glucose, and dietary protein but increased with dietary potassium. NAA was higher in males and increased with age, weight, eGFR_Cys_ and dietary potassium. Significant interactions were observed for Ucit excretion with age and blood glucose, weight and eGFR_Cys,_ and sex and thiazide use. Blood glucose had a significant and independent effect on U-pH and also Ucit. This study provides the first evidence that blood glucose could influence urinary stone risk independent of urinary pH, potentially providing new insight into the association of obesity and urinary stone disease. Copyright © 2017 The Authors. Physiological Reports published by Wiley Periodicals, Inc. on behalf of The Physiological Society and the American Physiological Society.

**URL:** <https://ovidsp.ovid.com/ovidweb.cgi?T=JS&CSC=Y&NEWS=N&PAGE=fulltext&D=med14&DO=10.14814%2fphy2.13411> <https://libkey.io/libraries/2592/openurl?genre=article&sid=OVID:medline&id=pmid:29038354&id=doi:10.14814%2Fphy2.13411&issn=2051-817X&isbn=&volume=5&issue=19&spage=&pages=&date=2017&title=Physiological+Reports&atitle=Association+of+urinary+citrate+excretion%2C+pH%2C+and+net+gastrointestinal+alkali+absorption+with+diet%2C+diuretic+use%2C+and+blood+glucose+concentration.&aulast=Perinpam&pid=%3Cauthor%3EPerinpam+M%3BWare+EB%3BSmith+JA%3BTurner+ST%3BKardia+SLR%3BLieske+JC%3C%2Fauthor%3E%3CAN%3E29038354%3C%2FAN%3E%3CDT%3EJournal+Article%3C%2FDT%3E>

41. **24 Hour urine metabolic differences between solitary and multiple stone formers: Results of the Collaboration on Urolithiasis in Pediatrics (CUP) working group.**

**Item Type:** Journal Article

**Authors:** Saitz, Theodore R.;Mongoue-Tchokote, Solange;Sharadin, Cynthia;Giel, Dana W.;Corbett, Sean;Kovacevic, Larisa and Bayne, Aaron P.

**Publication Date:** Oc ,2017

**Journal:** Journal of Pediatric Urology 13(5), pp. 506.e1–506.e5

**Abstract: INTRODUCTION**: Specific factors associated with the risk of developing pediatric urinary stone disease remain unclear, especially those that may be associated with recurrent stone disease. **OBJECTIVE**: We compared the results of 24-h urine collections in children with a solitary stone episode to those with multiple stone episodes to determine if there is a difference that may be associated with multiple stone formation in children. **STUDY DESIGN**: A multi-institutional retrospective analysis was completed to assess 24-h urinary metabolic profiles in children with urolithiasis aged 2-18 years old. Differences in mean urine collections between the two groups were assessed using chi-square tests to test the associations among gender, stone type, and multiple stone status, as well as multivariate analyses using general linear models. **RESULTS**: We analyzed 142 solitary stone patients and 136 multiple stone patients from four centers were included. Multiple stone patients were older than solitary stone patients (mean 13.4 +/- 3.6 years vs. 12 +/- 3.9 years, p = 0.002). Females were more likely to have multiple stones (58% vs. 39%, p = 0.002). BMI was not associated with multiple stones (p = 0.8467). Multiple stone formers had lower urine volumes, although this did not reach statistical significance when compared with solitary stone formation (20.4 mL/kg/day +/- 11.5 vs. 22.9 +/- 13.0, p = 0.0880). Higher values for super-saturation of calcium oxalate were associated with multiple stone disease in univariate (p = 0.0485) and multivariate analysis (p = 0.0469) (Figure). Centers located in the Southeast of the United States saw a higher proportion of children with multiple stones (Tennessee 62.7%, Virginia 44.4%, Oregon 31.6%, Michigan 27.3%, p : We analyzed 142 solitary stone patients and 136 multiple stone patients from four centers were included. Multiple stone patients were older than solitary stone patients (mean 13.4 +/- 3.6 years vs. 12 +/- 3.9 years, p = 0.002). Females were more likely to have multiple stones (58% vs. 39%, p = 0.002). BMI was not associated with multiple stones (p = 0.8467). Multiple stone formers had lower urine volumes, although this did not reach statistical significance when compared with solitary stone formation (20.4 mL/kg/day +/- 11.5 vs. 22.9 +/- 13.0, p = 0.0880). Higher values for super-saturation of calcium oxalate were associated with multiple stone disease in univariate (p = 0.0485) and multivariate analysis (p = 0.0469) (Figure). Centers located in the Southeast of the United States saw a higher proportion of children with multiple stones (Tennessee 62.7%, Virginia 44.4%, Oregon 31.6%, Michigan 27.3%, p **DISCUSSION**: In a large multi-institutional retrospective analysis we found that multiple stone disease was associated with higher super-saturations of calcium oxalate. Many urinary parameters changed with patient age, highlighting that the values should be interpreted with respect to patient age. The inability to comment on follow-up because of the nature of our dataset is a limitation of this study. **CONCLUSION**: Multiple stone disease in children is associated with higher super-saturation calcium oxalate, while lower urinary volume may also be associated with multiple stones; however, further study is required. Early metabolic evaluation may help risk stratify children likely to form multiple stones. Copyright © 2017 Journal of Pediatric Urology Company. Published by Elsevier Ltd. All rights reserved.

**URL:** <https://ovidsp.ovid.com/ovidweb.cgi?T=JS&CSC=Y&NEWS=N&PAGE=fulltext&D=med14&DO=10.1016%2fj.jpurol.2017.03.015> <https://libkey.io/libraries/2592/openurl?genre=article&sid=OVID:medline&id=pmid:28526618&id=doi:10.1016%2Fj.jpurol.2017.03.015&issn=1477-5131&isbn=&volume=13&issue=5&spage=506.e1&pages=506.e1-506.e5&date=2017&title=Journal+of+pediatric+urology&atitle=24+Hour+urine+metabolic+differences+between+solitary+and+multiple+stone+formers%3A+Results+of+the+Collaboration+on+Urolithiasis+in+Pediatrics+%28CUP%29+working+group.&aulast=Saitz&pid=%3Cauthor%3ESaitz+TR%3BMongoue-Tchokote+S%3BSharadin+C%3BGiel+DW%3BCorbett+S%3BKovacevic+L%3BBayne+AP%3C%2Fauthor%3E%3CAN%3E28526618%3C%2FAN%3E%3CDT%3EComparative+Study%3C%2FDT%3E>

42. **Metabolic investigation in patients with nephrolithiasis.**

**Item Type:** Journal Article

**Authors:** Santos, Francilayne Moretto Dos;Peres, Aline Krampe;Mandotti, Michel Roberto and Peres, Luis Alberto Batista

**Publication Date:** 2017

**Journal:** Einstein 15(4), pp. 452–456

**Abstract: OBJECTIVE**: To evaluate the prevalence of metabolic disorders associated with nephrolithiasis in a female population. **METHODS**: A retrospective study on 1,737 patients with evidence of recent formation of renal stones, being 54% females. The laboratory investigation consisted of at least two samples of blood and 24-hour urine to assess calcium, uric acid, citrate and creatinine levels, qualitative cystinuria, urinary pH following fasting and 12-hour water restriction, urine culture, serum creatinine and parathyroid hormone. **RESULTS**: The most frequent alterations were hypercalciuria (40.9%), urinary tract infection (23.2%), hypocitraturia (22.4%), low urinary volume (20.5%) and hyperuricosuria (16%). **CONCLUSION**: The most frequent metabolic alterations in females were hypocitraturia, urinary tract infection, low urinary volume and hyperuricosuria.

**URL:** <https://ovidsp.ovid.com/ovidweb.cgi?T=JS&CSC=Y&NEWS=N&PAGE=fulltext&D=med14&DO=10.1590%2fS1679-45082017AO4029> <https://libkey.io/libraries/2592/openurl?genre=article&sid=OVID:medline&id=pmid:29267427&id=doi:10.1590%2FS1679-45082017AO4029&issn=1679-4508&isbn=&volume=15&issue=4&spage=452&pages=452-456&date=2017&title=Einstein&atitle=Metabolic+investigation+in+patients+with+nephrolithiasis.&aulast=Santos&pid=%3Cauthor%3ESantos+FMD%3BPeres+AK%3BMandotti+MR%3BPeres+LAB%3C%2Fauthor%3E%3CAN%3E29267427%3C%2FAN%3E%3CDT%3EJournal+Article%3C%2FDT%3E>

43. **Physicochemical factors of the urine of children with hypercalciuria**

**Item Type:** Journal Article

**Authors:** Ubetagoyena-Arrieta, Mercedes;Martinez Saenz de Jubera, Jorge;Corera-Casty, Maria Nieves;Gonzalez Perez-Yarza, Eduardo and Garcia-Albizua, Irati

**Publication Date:** Ap ,2017

**Journal:** Archivos Espanoles De Urologia 70(3), pp. 342–348

**Abstract: OBJECTIVE**: Hypercalciuria is a common lithogenic risk factor. The aim of this study was, first, to study the characteristics of urine biochemical factors of children with hypercalciuria (HC) and compare them with those of children without hypercalciuria. Second, to analyze the differences between children with HC and lithiasis (HCL) and children with HC and no lithiasis (HCNL). **METHODS**: The sample was composed by 111 cases with HC, divided into 2 groups: HCNL group, consisting of 93 cases with no personal history of kidney stones, and HCL group, 18 cases with personal history of kidney stones. As a cohort control group, 113 healthy children were used. Creatinine, urea, sodium, potassium, chlorine, uric acid, calcium, phosphorus, magnesium and osmolality: blood and 24-hour urine following parameters were determined. Oxalate and citrate were determined in urine. **RESULTS**: The mean values of natriuresis, uricosuria, phosphaturia, magnesuria, citraturia, calcium oxalate and calcium phosphate saturation were higher in HCNL than in control group. The HCL group had phosphaturia, calcium oxalate and calcium phosphate saturations more elevated compared with the control group. There were no significant differences in urinary excretion of various parameters when the groups HCL and HCNL were compared. **CONCLUSIONS**: Our results show that children with hypercalciuria without lithiasis associated show an increase in natriuresis, phosphaturia, uricosuria, magnesuria and citraturia. We found no differences between these urinary abnormalities when compared hypercalciuric children without lithiasis with those with hypercalciuria and urolithiasis.

**URL:** <https://ovidsp.ovid.com/ovidweb.cgi?T=JS&CSC=Y&NEWS=N&PAGE=fulltext&D=med14&AN=28422036> <https://libkey.io/libraries/2592/openurl?genre=article&sid=OVID:medline&id=pmid:28422036&id=doi:&issn=0004-0614&isbn=&volume=70&issue=3&spage=342&pages=342-348&date=2017&title=Archivos+Espanoles+de+Urologia&atitle=Caracteristicas+de+los+factores+fisicoquimicos+que+componen+la+orina+de+los+ninos+con+hipercalciuria.&aulast=Ubetagoyena-Arrieta&pid=%3Cauthor%3EUbetagoyena-Arrieta+M%3BMartinez+Saenz+de+Jubera+J%3BCorera-Casty+MN%3BGonzalez+Perez-Yarza+E%3BGarcia-Albizua+I%3C%2Fauthor%3E%3CAN%3E28422036%3C%2FAN%3E%3CDT%3EJournal+Article%3C%2FDT%3E>

44. **Use of urinary concentrations in mg/dl in relation to absolute values in 24-hour samples for the evaluation of lithogenic factors in stone forming patients.**

**Item Type:** Journal Article

**Authors:** Arrabal-Polo, M. A.;Cano-Garcia, M. C.;Hidalgo-Agullo, G. and Roletto-Salmo, L.

**Publication Date:** 2016

**Journal:** Archivos Espanoles De Urologia 69(2), pp. 53–58

**Abstract: OBJECTIVE**: The aim of this study is to analyze urine concentrations (mg/dl) of different lithogenic factors in a sample of 24 h as a predictor of these changes rather than absolute values depend on the volume of diuresis. **METHODS**: A total of 131 patients from the North Almeria Health Management Area (Spain) with urinary calstone disease in whom a metabolic study was indicated were included from June 2014 to May 2015. The concentrations of calcium, oxalate, uric acid, citrate and magnesium were measured in the urine, and the calcium/citrate ratio was calculated. The classifications used were: hypercalciuria (>260mg/24h), hyperuricosuria (>750mg/24h), hyperoxaluria (>40mg/24h), hypocitraturia (: A total of 131 patients from the North Almeria Health Management Area (Spain) with urinary calstone disease in whom a metabolic study was indicated were included from June 2014 to May 2015. The concentrations of calcium, oxalate, uric acid, citrate and magnesium were measured in the urine, and the calcium/citrate ratio was calculated. The classifications used were: hypercalciuria (>260mg/24h), hyperuricosuria (>750mg/24h), hyperoxaluria (>40mg/24h), hypocitraturia (: A total of 131 patients from the North Almeria Health Management Area (Spain) with urinary calstone disease in whom a metabolic study was indicated were included from June 2014 to May 2015. The concentrations of calcium, oxalate, uric acid, citrate and magnesium were measured in the urine, and the calcium/citrate ratio was calculated. The classifications used were: hypercalciuria (>260mg/24h), hyperuricosuria (>750mg/24h), hyperoxaluria (>40mg/24h), hypocitraturia (**RESULTS**: A cut-off point of 12.55mg/dl, with a sensitivity of 90% and a specificity of 85% and a relative risk (RR) of 51.2 (13.9-188.4), was estimated for urinary calcium. For oxalate the cut-off point was 1.86mg/dl, with a sensitivity of 91% and a specificity of 84% with an estimated RR of 67.2 (8.3-540.6). As regards the uric acid concentration in urine, a cut-off point of 31.2mg/dl was estimated, with a sensitivity of 85% and a specificity of 70% and a RR of 12 (3.8-37.6). For citrate the cut-off point was 18.8mg/dl, with a sensitivity and specificity of 82% and 74%, respectively, with a RR of 13.7 (4.4- 42.6). The cut-off point for magnesium was 2.26mg/dl with a sensitivity of 95% and specificity of 78%, with a RR of 67.6 (11.4-398.3). **CONCLUSION**: The determination of urine concentrations, instead of absolute values, depends to a large extent on urine output, appears to be useful when estimating classic metabolic alterations and should be taken into account in the evaluation of patients with urinary stone disease.

**URL:** <https://ovidsp.ovid.com/ovidweb.cgi?T=JS&CSC=Y&NEWS=N&PAGE=fulltext&D=med13&AN=26959969> <https://libkey.io/libraries/2592/openurl?genre=article&sid=OVID:medline&id=pmid:26959969&id=doi:&issn=0004-0614&isbn=&volume=69&issue=2&spage=53&pages=53-8&date=2016&title=Archivos+Espanoles+de+Urologia&atitle=Uso+de+las+concentraciones+urinarias+en+mg%2Fdl+en+relacion+a+los+valores+absolutos+en+24+h+para+la+evaluacion+de+factores+litogenicos+en+pacientes+litiasicos.&aulast=Arrabal-Polo&pid=%3Cauthor%3EArrabal-Polo+MA%3BCano-Garcia+MC%3BHidalgo-Agullo+G%3BRoletto-Salmo+L%3C%2Fauthor%3E%3CAN%3E26959969%3C%2FAN%3E%3CDT%3EJournal+Article%3C%2FDT%3E>

45. **Assessment of Urine Proteomics in Type 1 Primary Hyperoxaluria.**

**Item Type:** Journal Article

**Authors:** Brooks, Ellen R.;Hoppe, Bernd;Milliner, Dawn S.;Salido, Eduardo;Rim, John;Krevitt, Leah M.;Olson, Julie B.;Price, Heather E.;Vural, Gulsah and Langman, Craig B.

**Publication Date:** 2016

**Journal:** American Journal of Nephrology 43(4), pp. 293–303

**Abstract: BACKGROUND**: Primary hyperoxaluria type 1 (PH1) and idiopathic hypercalciuria (IHC) are stone-forming diseases that may result in the formation of calcium (Ca) oxalate (Ox) stones, nephrocalcinosis, and progressive chronic kidney disease (CKD). Poorer clinical outcome in PH1 is segregated by the highest urine (Ur)-Ox (UrOx), while IHC outcomes are not predictable by UrCa. We hypothesized that differences would be found in selected Ur-protein (PRO) patterns in PH1 and IHC, compared to healthy intra-familial sibling controls (C) of PH1 patients. We also hypothesized that the PRO patterns associated with higher UrOx levels would reflect injury, inflammation, biomineralization, and abnormal tissue repair processes in PH1. **METHODS**: Twenty four-hour Ur samples were obtained from 3 cohorts: PH1 (n = 47); IHC (n = 35) and C (n = 13) and were analyzed using targeted platform-based multi-analyte profile immunoassays and for UrOx and UrCa by biochemical measurements. **RESULTS**: Known stone matrix constituents, osteopontin, calbindin, and vitronectin were lowest in PH1 (C > IHC > PH1; p C (p = 0.03 to p IHC (p IHC (p **CONCLUSION**: PH1 Ur-PROs reflected overt inflammation, chemotaxis, oxidative stress, growth factors (including EGF), and pro-angiogenic and calcification regulation/inhibition compared to the C and IHC cohorts. Many of the up- and downregulated PH1-PROs found in this study are also found in CKD, acute kidney injury, stone formers, and/or stone matrices. Further data analyses may provide evidence for PH1 unique PROs or demonstrate a poorer clinical outcome. Copyright © 2016 S. Karger AG, Basel.

**URL:** <https://ovidsp.ovid.com/ovidweb.cgi?T=JS&CSC=Y&NEWS=N&PAGE=fulltext&D=med13&DO=10.1159%2f000445448> <https://libkey.io/libraries/2592/openurl?genre=article&sid=OVID:medline&id=pmid:27161247&id=doi:10.1159%2F000445448&issn=0250-8095&isbn=&volume=43&issue=4&spage=293&pages=293-303&date=2016&title=American+Journal+of+Nephrology&atitle=Assessment+of+Urine+Proteomics+in+Type+1+Primary+Hyperoxaluria.&aulast=Brooks&pid=%3Cauthor%3EBrooks+ER%3BHoppe+B%3BMilliner+DS%3BSalido+E%3BRim+J%3BKrevitt+LM%3BOlson+JB%3BPrice+HE%3BVural+G%3BLangman+CB%3C%2Fauthor%3E%3CAN%3E27161247%3C%2FAN%3E%3CDT%3EComparative+Study%3C%2FDT%3E>

46. **Prospective evaluation of urinary metabolic indices in severely obese adolescents after weight loss surgery.**

**Item Type:** Journal Article

**Authors:** DeFoor, W. Robert;Asplin, John R.;Kollar, Linda;Jackson, Elizabeth;Jenkins, Todd;Schulte, Marion and Inge, Thomas

**Publication Date:** Fe ,2016

**Journal:** Surgery for Obesity & Related Diseases 12(2), pp. 363–367

**Abstract: BACKGROUND**: Observational studies in obese adults have found abnormal urinary metabolic indices that predispose to nephrolithiasis. Few studies have been performed in severely obese adolescents. **OBJECTIVES**: To assess urinary stone risk factors in severely obese adolescents and in those undergoing 2 types of weight loss surgery. **SETTING**: Children's hospital, United States. **METHODS**: A prospective cross-sectional study was performed to assess urinary metabolic profiles in severely obese adolescents who either have not undergone any gastrointestinal surgery or who have undergone Roux-en-Y gastric bypass (RYGB) or vertical sleeve gastrectomy (SG). Twenty-four-hour urine collections were performed at home and evaluated at a central laboratory. Established normal reference ranges for adults were used in the analysis. A linear regression analysis was performed assessing the relationship of the study group with each of the outcomes. **RESULTS**: A total of 55 samples were analyzed from 14 severely obese adolescents and from 17 severely obese adolescents after bariatric surgery (RYGB, 10; SG, 7). Median body mass index was similar between the RYGB and SG groups. The median 24-hour excretion of oxalate was significantly elevated in the RYGB group. Calcium and uric acid excretion and the median supersaturation of calcium oxalate, calcium phosphate, and uric acid were similar among all groups. **CONCLUSIONS**: Elevated excretion of oxalate in the urine of severely obese adolescents and in those who have undergone RYGB may portend increased risk for kidney stone formation. Larger longitudinal studies are needed to verify these findings and to determine the clinical risk of developing stone disease in these patient populations. Copyright © 2016 American Society for Metabolic and Bariatric Surgery. Published by Elsevier Inc. All rights reserved.

**URL:** <https://ovidsp.ovid.com/ovidweb.cgi?T=JS&CSC=Y&NEWS=N&PAGE=fulltext&D=med13&DO=10.1016%2fj.soard.2015.03.013> <https://libkey.io/libraries/2592/openurl?genre=article&sid=OVID:medline&id=pmid:26077697&id=doi:10.1016%2Fj.soard.2015.03.013&issn=1550-7289&isbn=&volume=12&issue=2&spage=363&pages=363-7&date=2016&title=Surgery+for+Obesity+%26+Related+Diseases&atitle=Prospective+evaluation+of+urinary+metabolic+indices+in+severely+obese+adolescents+after+weight+loss+surgery.&aulast=DeFoor&pid=%3Cauthor%3EDeFoor+WR%3BAsplin+JR%3BKollar+L%3BJackson+E%3BJenkins+T%3BSchulte+M%3BInge+T%3C%2Fauthor%3E%3CAN%3E26077697%3C%2FAN%3E%3CDT%3EJournal+Article%3C%2FDT%3E>

47. **Contemporary Attitudes and Practice Patterns of North American Urologists in Investigating Stone-Forming Patients-A Survey of Endourological Society Members.**

**Item Type:** Journal Article

**Authors:** McGuire, Barry B.;Matulewicz, Richard S.;Zuccarino-Crowe, Rian;Nadler, Robert B. and Perry, Kent T.

**Publication Date:** Ap ,2016

**Journal:** Journal of Endourology 30(4), pp. 460–464

**Abstract: INTRODUCTION**: Recent evidence would suggest a low rate of metabolic assessment in stone formers, even in those deemed as high risk. We wished to assess the attitudes and practice patterns of metabolic work up in North American members of the Endourological Society as part of the management of stone-forming patients. **METHODS**: A 12-question online multiple-choice questionnaire (using Survey Monkey( R)) was distributed to all members of the Endourological Society through e-mail. Descriptive analyses were performed. **RESULTS**: A total of 124 North American members of the Endourological Society responded (90% endourologists, 65% fellowship trained). Ninety-seven percent perform metabolic assessments without referring to a consultant. Eighty-three percent use a commercial analysis company and 17% request serum or urine parameters individually. Ninety-seven percent believe that 24-48-hour urine collection is a better way of assessing patients for metabolic abnormalities than a "basic analysis." Many respondents (37%) would be more likely to metabolically assess if results were easier to interpret, and 35% would like assistance/advice in the interpretation of results. At initial investigation of a first-time stone former, 87% of respondents use serum chemistry, 48% use 24-hour urine, 26% use 48-hour urine (two consecutive 24-hour urine collections), 54% send stone for analysis, and 7% do not investigate. On recurrent stone formers, 69% use serum chemistry, 73% use 24/48-hour urine, and 23% send stone for analysis. On routine follow-up, 36% check serum chemistry, 55% use 24-hour urine, 2% use 48-hour urine, and 29% do not metabolically evaluate. The majority agree that pharmacologic therapy plays a strong role in preventing recurrence (90%). After initiating pharmacologic therapy, 59% reassess using serum chemistry and 84% and 7% use 24/48-hour urine collection, respectively. Physicians re-evaluate patients after 1 month (7%), 1-2 months (10%), 2-4 months (44%), 4-6 months (30%), or after 6-12 months (7%). **CONCLUSION**: This snapshot assessment of Endourological Society members' practices in the metabolic investigation of stone-forming patients demonstrates wide testing variations. Many physicians expressed interest in assistance/advice in the interpretation of the metabolic assessment results.

**URL:** <https://ovidsp.ovid.com/ovidweb.cgi?T=JS&CSC=Y&NEWS=N&PAGE=fulltext&D=med13&DO=10.1089%2fend.2015.0207> <https://libkey.io/libraries/2592/openurl?genre=article&sid=OVID:medline&id=pmid:26738410&id=doi:10.1089%2Fend.2015.0207&issn=0892-7790&isbn=&volume=30&issue=4&spage=460&pages=460-4&date=2016&title=Journal+of+Endourology&atitle=Contemporary+Attitudes+and+Practice+Patterns+of+North+American+Urologists+in+Investigating+Stone-Forming+Patients-A+Survey+of+Endourological+Society+Members.&aulast=McGuire&pid=%3Cauthor%3EMcGuire+BB%3BMatulewicz+RS%3BZuccarino-Crowe+R%3BNadler+RB%3BPerry+KT%3C%2Fauthor%3E%3CAN%3E26738410%3C%2FAN%3E%3CDT%3EJournal+Article%3C%2FDT%3E>

48. **Dipstick Spot urine pH does not accurately represent 24 hour urine PH measured by an electrode.**

**Item Type:** Journal Article

**Authors:** Omar, Mohamed;Sarkissian, Carl;Jianbo, Li;Calle, Juan and Monga, Manoj

**Publication Date:** 2016

**Journal:** International Braz J Urol 42(3), pp. 546–549

**Abstract: OBJECTIVES**: To determine whether spot urine pH measured by dipstick is an accurate representation of 24 hours urine pH measured by an electrode. **MATERIALS AND METHODS**: We retrospectively reviewed urine pH results of patients who presented to the urology stone clinic. For each patient we recorded the most recente pH result measured by dipstick from a spot urine sample that preceded the result of a 24-hour urine pH measured by the use of a pH electrode. Patients were excluded if there was a change in medications or dietary recommendations or if the two samples were more than 4 months apart. A difference of more than 0.5 pH was considered na inaccurate result. **RESULTS**: A total 600 patients were retrospectively reviewed for the pH results. The mean difference in pH between spot urine value and the 24 hours collection values was 0.52+/-0.45 pH. Higher pH was associated with lower accuracy (p: A total 600 patients were retrospectively reviewed for the pH results. The mean difference in pH between spot urine value and the 24 hours collection values was 0.52+/-0.45 pH. Higher pH was associated with lower accuracy (p6.5. Samples taken more than 75 days apart had only 49% the accuracy of more recent samples (p6.5. Samples taken more than 75 days apart had only 49% the accuracy of more recent samples (p6.5. Samples taken more than 75 days apart had only 49% the accuracy of more recent samples (p**CONCLUSIONS**: Spot urine pH by dipstick is not an accurate method for evaluation of the patients with urolithiasis. Patients with alkaline urine are more prone to error with reliance on spot urine pH.

**URL:** <https://ovidsp.ovid.com/ovidweb.cgi?T=JS&CSC=Y&NEWS=N&PAGE=fulltext&D=med13&DO=10.1590%2fS1677-5538.IBJU.2015.0071> <https://libkey.io/libraries/2592/openurl?genre=article&sid=OVID:medline&id=pmid:27286119&id=doi:10.1590%2FS1677-5538.IBJU.2015.0071&issn=1677-5538&isbn=&volume=42&issue=3&spage=546&pages=546-9&date=2016&title=International+Braz+J+Urol&atitle=Dipstick+Spot+urine+pH+does+not+accurately+represent+24+hour+urine+PH+measured+by+an+electrode.&aulast=Omar&pid=%3Cauthor%3EOmar+M%3BSarkissian+C%3BJianbo+L%3BCalle+J%3BMonga+M%3C%2Fauthor%3E%3CAN%3E27286119%3C%2FAN%3E%3CDT%3EJournal+Article%3C%2FDT%3E>

49. **Re: Underuse of 24-hour urine collection among children with incident urinary stones: a quality-of-care concern?.**

**Item Type:** Journal Article

**Authors:** Canning, Douglas A.

**Publication Date:** Ma ,2015

**Journal:** Journal of Urology 193(3), pp. 990

**URL:** <https://ovidsp.ovid.com/ovidweb.cgi?T=JS&CSC=Y&NEWS=N&PAGE=fulltext&D=med12&DO=10.1016%2fj.juro.2014.12.034> <https://libkey.io/libraries/2592/openurl?genre=article&sid=OVID:medline&id=pmid:25765427&id=doi:10.1016%2Fj.juro.2014.12.034&issn=0022-5347&isbn=&volume=193&issue=3&spage=990&pages=990&date=2015&title=Journal+of+Urology&atitle=Re%3A+Underuse+of+24-hour+urine+collection+among+children+with+incident+urinary+stones%3A+a+quality-of-care+concern%3F.&aulast=Canning&pid=%3Cauthor%3ECanning+DA%3C%2Fauthor%3E%3CAN%3E25765427%3C%2FAN%3E%3CDT%3EEditorial%3C%2FDT%3E>

50. **Provider variation in the quality of metabolic stone management.**

**Item Type:** Journal Article

**Authors:** Dauw, Casey A.;Alruwaily, Abdulrahman F.;Bierlein, Maggie J.;Asplin, John R.;Ghani, Khurshid R.;Wolf, J. Stuart Jr and Hollingsworth, John M.

**Publication Date:** Ma ,2015

**Journal:** Journal of Urology 193(3), pp. 885–890

**Abstract: PURPOSE**: Urinary stone disease is a chronic condition for which secondary prevention (dietary and medical therapy guided by 24-hour urine collection results) has an important role. Assessing the response to these interventions with followup testing is recommended and yet to our knowledge provider compliance with these guidelines is unknown. **MATERIALS AND METHODS**: Using Litholink R files from 1995 to 2013 we identified adults with urinary stone disease who underwent metabolic evaluation and the providers who ordered the evaluation. By focusing on patients with an abnormality on the initial collection we determined the proportion who underwent a followup test within 6 months of the initial test. Multilevel modeling was done to quantify variation in followup testing among providers after accounting for various patient and provider factors. **RESULTS**: A total of 208,125 patients had an abnormality on the initial collection, of whom only 33,413 (16.1%) performed a repeat collection within 6 months. While most variation in followup testing was attributable to the patient, the provider contribution was nontrivial (18.0%). The specialty of the ordering provider was important. Patients who saw a urologist had 24% lower odds of repeat testing compared to those who saw a primary care physician (OR 0.76, 95% CI 0.67-0.86, p : A total of 208,125 patients had an abnormality on the initial collection, of whom only 33,413 (16.1%) performed a repeat collection within 6 months. While most variation in followup testing was attributable to the patient, the provider contribution was nontrivial (18.0%). The specialty of the ordering provider was important. Patients who saw a urologist had 24% lower odds of repeat testing compared to those who saw a primary care physician (OR 0.76, 95% CI 0.67-0.86, p **CONCLUSIONS**: Followup testing is uncommon in patients with an abnormal initial 24-hour urine collection. Given the observed provider variation, efforts to educate providers on the value of followup testing are likely to have salutary effects on patients with metabolic stone disease. Copyright © 2015 American Urological Association Education and Research, Inc. Published by Elsevier Inc. All rights reserved.

**URL:** <https://ovidsp.ovid.com/ovidweb.cgi?T=JS&CSC=Y&NEWS=N&PAGE=fulltext&D=med12&DO=10.1016%2fj.juro.2014.09.111> <https://libkey.io/libraries/2592/openurl?genre=article&sid=OVID:medline&id=pmid:25286012&id=doi:10.1016%2Fj.juro.2014.09.111&issn=0022-5347&isbn=&volume=193&issue=3&spage=885&pages=885-90&date=2015&title=Journal+of+Urology&atitle=Provider+variation+in+the+quality+of+metabolic+stone+management.&aulast=Dauw&pid=%3Cauthor%3EDauw+CA%3BAlruwaily+AF%3BBierlein+MJ%3BAsplin+JR%3BGhani+KR%3BWolf+JS+Jr%3BHollingsworth+JM%3C%2Fauthor%3E%3CAN%3E25286012%3C%2FAN%3E%3CDT%3EComparative+Study%3C%2FDT%3E>

51. **Daily sodium and potassium excretion can be estimated by scheduled spot urine collections.**

**Item Type:** Journal Article

**Authors:** Doenyas-Barak, Keren;Beberashvili, Ilia;Bar-Chaim, Adina;Averbukh, Zhan;Vogel, Ofir and Efrati, Shai

**Publication Date:** 2015

**Journal:** Nephron 130(1), pp. 35–40

**Abstract: AIM**: The evaluation of sodium and potassium intake is part of the optimal management of hypertension, metabolic syndrome, renal stones, and other conditions. To date, no convenient method for its evaluation exists, as the gold standard method of 24-hour urine collection is cumbersome and often incorrectly performed, and methods that use spot or shorter collections are not accurate enough to replace the gold standard. The aim of this study was to evaluate the correlation and agreement between a new method that uses multiple-scheduled spot urine collection and the gold standard method of 24-hour urine collection. **METHODS**: The urine sodium or potassium to creatinine ratios were determined for four scheduled spot urine samples. The mean ratios of the four spot samples and the ratios of each of the single spot samples were corrected for estimated creatinine excretion and compared to the gold standard. **RESULTS**: A significant linear correlation was demonstrated between the 24-hour urinary solute excretions and estimated excretion evaluated by any of the scheduled spot urine samples. The correlation of the mean of the four spots was better than for any of the single spots. Bland-Altman plots showed that the differences between these measurements were within the limits of agreement. **CONCLUSION**: Four scheduled spot urine samples can be used as a convenient method for estimation of 24-hour sodium or potassium excretion. Copyright © 2015 S. Karger AG, Basel.

**URL:** <https://ovidsp.ovid.com/ovidweb.cgi?T=JS&CSC=Y&NEWS=N&PAGE=fulltext&D=med12&DO=10.1159%2f000430105> <https://libkey.io/libraries/2592/openurl?genre=article&sid=OVID:medline&id=pmid:25998865&id=doi:10.1159%2F000430105&issn=1660-8151&isbn=&volume=130&issue=1&spage=35&pages=35-40&date=2015&title=Nephron&atitle=Daily+sodium+and+potassium+excretion+can+be+estimated+by+scheduled+spot+urine+collections.&aulast=Doenyas-Barak&pid=%3Cauthor%3EDoenyas-Barak+K%3BBeberashvili+I%3BBar-Chaim+A%3BAverbukh+Z%3BVogel+O%3BEfrati+S%3C%2Fauthor%3E%3CAN%3E25998865%3C%2FAN%3E%3CDT%3EJournal+Article%3C%2FDT%3E>

52. **Factors Associated With the Lower Prevalence of Nephrolithiasis in Children Compared With Adults.**

**Item Type:** Journal Article

**Authors:** Grases, Felix;Rodriguez, Adrian;Costa-Bauza, Antonia;Saez-Torres, Concepcion;Rodrigo, Dolores;Gomez, Cristina;Mir-Perello, Concepcion and Frontera, Guiem

**Publication Date:** Se ,2015

**Journal:** Urology 86(3), pp. 587–592

**Abstract: OBJECTIVE**: To determine the reasons behind the lower prevalence of kidney stones in children by assessing urinary lithogenic parameters in healthy children, healthy adults, and 3 groups of stone-former patients. **METHODS**: The study subjects included 75 healthy adults, 105 healthy children, 62 patients with previous calcium oxalate monohydrate papillary stones, 120 patients with previous calcium oxalate monohydrate unattached stones, and 248 patients with previous calcium oxalate dihydrate stones. Twenty-four-hour urine samples were collected, and the urinary lithogenic parameters were measured. **RESULTS**: Calcium, magnesium, and phosphorous concentration differed significantly between healthy children and adults. Except citrate, all solute/creatinine ratios differed between healthy children and adults. However, these differences were much more important in the cases of calcium and magnesium. The calcium/creatinine ratio was 2-fold lower, whereas the magnesium/creatinine ratio was 2-fold higher, in healthy children than that in healthy adults (P : Calcium, magnesium, and phosphorous concentration differed significantly between healthy children and adults. Except citrate, all solute/creatinine ratios differed between healthy children and adults. However, these differences were much more important in the cases of calcium and magnesium. The calcium/creatinine ratio was 2-fold lower, whereas the magnesium/creatinine ratio was 2-fold higher, in healthy children than that in healthy adults (P **CONCLUSION**: Ratios of calcium and magnesium to creatinine, as well as morphoanatomic factors and lifestyle habits, may explain the lower prevalence of nephrolithiasis in children than those in adults. Copyright © 2015 Elsevier Inc. All rights reserved.

**URL:** <https://ovidsp.ovid.com/ovidweb.cgi?T=JS&CSC=Y&NEWS=N&PAGE=fulltext&D=med12&DO=10.1016%2fj.urology.2015.06.028> <https://libkey.io/libraries/2592/openurl?genre=article&sid=OVID:medline&id=pmid:26144339&id=doi:10.1016%2Fj.urology.2015.06.028&issn=0090-4295&isbn=&volume=86&issue=3&spage=587&pages=587-92&date=2015&title=Urology&atitle=Factors+Associated+With+the+Lower+Prevalence+of+Nephrolithiasis+in+Children+Compared+With+Adults.&aulast=Grases&pid=%3Cauthor%3EGrases+F%3BRodriguez+A%3BCosta-Bauza+A%3BSaez-Torres+C%3BRodrigo+D%3BGomez+C%3BMir-Perello+C%3BFrontera+G%3C%2Fauthor%3E%3CAN%3E26144339%3C%2FAN%3E%3CDT%3EComparative+Study%3C%2FDT%3E>

53. **Does socioeconomic status, age, or gender influence appointment attendance and completion of 24-hour urine collections?.**

**Item Type:** Journal Article

**Authors:** Sninsky, Brian C.;Nakada, Stephen Y. and Penniston, Kristina L.

**Publication Date:** Ma ,2015

**Journal:** Urology 85(3), pp. 568–573

**Abstract: OBJECTIVE**: To examine the association of age, gender, and socioeconomic status with appointment attendance and completion of 24-hour urine collections to identify patient characteristics associated with barriers to treatment. **METHODS**: A retrospective review was performed under institutional review board approval for new patients scheduled at a single metabolic stone clinic over a 1.5-year period. Postal codes were used to identify poverty and education levels using US Census Bureau data. We evaluated 3 years of adherence based on "no shows," percentage of stone clinic appointments attended, and percentage of 24-hour urine collections completed. Quintile analyses were used to further explore nonlinear data. **RESULTS**: A total of 447 patients were included (246 men and 201 women). Mean age was 49 +/- 14.5 years. Younger age, areas of higher poverty, and areas of lower education were all weakly to moderately associated with increased "no show" rates and decreased clinic attendance. Men tended to not show for appointments more frequently than women (P = .043). When grouped by age groups, younger age was strongly correlated with decreased 24-hour urine collection completion (R = 0.75). When divided into age quintiles, relationships between "no shows" and clinic attendance were strengthened as they were when patients were divided into poverty quintiles. **CONCLUSION**: It is critical for providers to be aware of health disparities in medical stone management. Low education, high poverty, and younger age are associated with suboptimal evaluation follow-up. Of note, the lowest adherence was in younger patients, a population that requires the most urgent workup. Copyright © 2015 Elsevier Inc. All rights reserved.

**URL:** <https://ovidsp.ovid.com/ovidweb.cgi?T=JS&CSC=Y&NEWS=N&PAGE=fulltext&D=med12&DO=10.1016%2fj.urology.2014.10.043> <https://libkey.io/libraries/2592/openurl?genre=article&sid=OVID:medline&id=pmid:25733267&id=doi:10.1016%2Fj.urology.2014.10.043&issn=0090-4295&isbn=&volume=85&issue=3&spage=568&pages=568-73&date=2015&title=Urology&atitle=Does+socioeconomic+status%2C+age%2C+or+gender+influence+appointment+attendance+and+completion+of+24-hour+urine+collections%3F.&aulast=Sninsky&pid=%3Cauthor%3ESninsky+BC%3BNakada+SY%3BPenniston+KL%3C%2Fauthor%3E%3CAN%3E25733267%3C%2FAN%3E%3CDT%3EJournal+Article%3C%2FDT%3E>

54. **Collection and storage of urine specimens for measurement of urolithiasis risk factors.**

**Item Type:** Journal Article

**Authors:** Wu, Wenqi;Yang, Dong;Tiselius, Hans-Goran;Ou, Lili;Mai, Zanlin;Chen, Kang;Zhu, Hanliang;Xu, Shaohong;Zhao, Zhijian and Zeng, Guohua

**Publication Date:** Fe ,2015

**Journal:** Urology 85(2), pp. 299–303

**Abstract: OBJECTIVE**: To evaluate how different methods for storage and preservation of urine samples affected the outcome of analysis of risk factors for stone formation. **METHODS**: Spot urine samples were collected from 21 healthy volunteers. Each fresh urine sample was divided into ten 10-mL aliquots: 2 without preservative, 2 with thymol, 2 with toluene, 2 with hydrochloric acid (HCl), and 2 with sodium azide. One sample of each pair was stored at 4 degreeC and the other at room temperature. The concentrations of calcium, magnesium, sodium, phosphate, urate, oxalate, citrate, and pH in each urine sample were analyzed immediately after collection (0 hour) and after 24 and 48 hours. **RESULTS**: There were no significant differences in calcium, oxalate, magnesium, phosphate, sodium, urate or pH (without acidification) between samples with different preservation methods (P >.05). Urinary citrate, however, was significantly lower in the urine collected with HCl than when other preservatives were used, both at room temperature and at 4 degreeC. Urine pH was significantly higher after 48 hours than after 24 hours, whether the samples were stored at room temperature or at 4 degreeC. **CONCLUSION**: Antibacterial preservatives (eg, thymol or toluene) can be recommended as preservatives for 24-hour urine collections. Ideally, the samples should be stored at 4 degreeC. When HCl is used as a preservative, it seems essential to neutralize the samples before analysis. This is particularly obvious with the chromatographic method used for analysis of citrate that was used in this study. Copyright © 2015 Elsevier Inc. All rights reserved.

**URL:** <https://ovidsp.ovid.com/ovidweb.cgi?T=JS&CSC=Y&NEWS=N&PAGE=fulltext&D=med12&DO=10.1016%2fj.urology.2014.10.030> <https://libkey.io/libraries/2592/openurl?genre=article&sid=OVID:medline&id=pmid:25623670&id=doi:10.1016%2Fj.urology.2014.10.030&issn=0090-4295&isbn=&volume=85&issue=2&spage=299&pages=299-303&date=2015&title=Urology&atitle=Collection+and+storage+of+urine+specimens+for+measurement+of+urolithiasis+risk+factors.&aulast=Wu&pid=%3Cauthor%3EWu+W%3BYang+D%3BTiselius+HG%3BOu+L%3BMai+Z%3BChen+K%3BZhu+H%3BXu+S%3BZhao+Z%3BZeng+G%3C%2Fauthor%3E%3CAN%3E25623670%3C%2FAN%3E%3CDT%3EComparative+Study%3C%2FDT%3E>

55. **Predisposing factors for infantile urinary calculus in south-west of Iran.**

**Item Type:** Journal Article

**Authors:** Alemzadeh-Ansari, Mohammad Hasan;Valavi, Ehsan and Ahmadzadeh, Ali

**Publication Date:** Ja ,2014

**Journal:** Iranian Journal of Kidney Diseases 8(1), pp. 53–57

**Abstract: INTRODUCTION**: Urinary calculi in infants are relatively infrequent, but their incidence has increased in the recent decades. The aim of this study was to investigate the clinical presentation, metabolic risk factors, and urinary tract abnormalities in infants suffering from kidney calculus. **MATERIALS AND METHODS**: A total of 152 infants were admitted between 2009 and 2012 with ultrasonography-proven urolithiasis. A Foley catheter was fixed and 24-hour urine samples were analyzed for calcium, citrate, oxalate, uric acid, and magnesium. For detecting cystinuria, qualitative measurement of urinary cystine was done by nitroprusside test. Urinary tract structural abnormalities were also evaluated. **RESULTS**: The mean age at the diagnosis of kidney calculus was 5.46 months (range, 15 days to 12 months). The most common clinical findings were restlessness and urinary tract infection. A family history of calculi was found in 67.1% of the patients and 68.4% were born to consanguineous marriages. Metabolic abnormalities and urinary tract abnormalities were found in 96.1% and 15.1% of children, respectively. Urinary tract abnormalities were more common in girls. The most common metabolic risk factors were hypercalciuria (79.6%) and hypocitraturia (40.9%). Hyperoxaluria and hypomagnesuria were found in about 28% of patients, both of which were associated with bilateral urolithiasis. **CONCLUSIONS**: These findings show that urinary metabolic abnormalities are very common in infants with urolithiasis. Appropriate evaluation of urinary metabolic parameters can lead us to proper diagnosis and treatment.

**URL:** <https://ovidsp.ovid.com/ovidweb.cgi?T=JS&CSC=Y&NEWS=N&PAGE=fulltext&D=med11&AN=24413722> <https://libkey.io/libraries/2592/openurl?genre=article&sid=OVID:medline&id=pmid:24413722&id=doi:&issn=1735-8582&isbn=&volume=8&issue=1&spage=53&pages=53-7&date=2014&title=Iranian+journal+of+Kidney+Diseases&atitle=Predisposing+factors+for+infantile+urinary+calculus+in+south-west+of+Iran.&aulast=Alemzadeh-Ansari&pid=%3Cauthor%3EAlemzadeh-Ansari+MH%3BValavi+E%3BAhmadzadeh+A%3C%2Fauthor%3E%3CAN%3E24413722%3C%2FAN%3E%3CDT%3EJournal+Article%3C%2FDT%3E>

56. **Determinants of urolithiasis before and after parathyroidectomy in patients with primary hyperparathyroidism.**

**Item Type:** Journal Article

**Authors:** Elkoushy, Mohamed A.;Yu, Alice X.;Tabah, Roger;Payne, Richard J.;Dragomir, Alice and Andonian, Sero

**Publication Date:** Ju ,2014

**Journal:** Urology 84(1), pp. 22–26

**Abstract: OBJECTIVE**: To assess the determinants of urolithiasis in patients with primary hyperparathyroidism (PHPT) before and after parathyroidectomy (PTX). **METHODS**: Institutional Research Ethics approval was obtained. A retrospective review was performed for patients presenting with PHPT to the stone, surgical oncology, and otolaryngology clinics at 2 tertiary-care centers from January 2006 to November 2011. Demographic, clinical, and surgical data were collected together with 24-hour urine collections before and after PTX. **RESULTS**: Of 332 patients undergoing PTX, 255 (68.2% female patients) had PHPT. Mean age was 60.3 years (range, 18-91). Before PTX, renal calcification was detected in 51 (20%) patients, nephrolithiasis in 48 (18.8%), and nephrocalcinosis in 3 (1.2%) patients. Compared with PHPT patients without stones, PHPT patients with stones were significantly younger (56.4 vs 61.3 years, P=.02), less likely to be female (54.9% vs 71.9%, P=.03), and had significantly lower levels of vitamin D (19.7 vs 23.5 ng/mL, P=.03). Nine patients (3.5%) developed stones after PTX and were found to have significantly higher post-PTX total serum calcium levels when compared with those without stones. Although hypercalciuria was detected in 62% of pre-PTX stone formers, none of those who tested had post-PTX hypercalciuria (P: Of 332 patients undergoing PTX, 255 (68.2% female patients) had PHPT. Mean age was 60.3 years (range, 18-91). Before PTX, renal calcification was detected in 51 (20%) patients, nephrolithiasis in 48 (18.8%), and nephrocalcinosis in 3 (1.2%) patients. Compared with PHPT patients without stones, PHPT patients with stones were significantly younger (56.4 vs 61.3 years, P=.02), less likely to be female (54.9% vs 71.9%, P=.03), and had significantly lower levels of vitamin D (19.7 vs 23.5 ng/mL, P=.03). Nine patients (3.5%) developed stones after PTX and were found to have significantly higher post-PTX total serum calcium levels when compared with those without stones. Although hypercalciuria was detected in 62% of pre-PTX stone formers, none of those who tested had post-PTX hypercalciuria (P**CONCLUSION**: Pre-PTX urolithiasis was associated with younger age, male gender, and lower levels of vitamin D, whereas post-PTX urolithiasis was independently predicted by male gender and hypercalcemia. Copyright © 2014 Elsevier Inc. All rights reserved.

**URL:** <https://ovidsp.ovid.com/ovidweb.cgi?T=JS&CSC=Y&NEWS=N&PAGE=fulltext&D=med11&DO=10.1016%2fj.urology.2014.01.016> <https://libkey.io/libraries/2592/openurl?genre=article&sid=OVID:medline&id=pmid:24703459&id=doi:10.1016%2Fj.urology.2014.01.016&issn=0090-4295&isbn=&volume=84&issue=1&spage=22&pages=22-6&date=2014&title=Urology&atitle=Determinants+of+urolithiasis+before+and+after+parathyroidectomy+in+patients+with+primary+hyperparathyroidism.&aulast=Elkoushy&pid=%3Cauthor%3EElkoushy+MA%3BYu+AX%3BTabah+R%3BPayne+RJ%3BDragomir+A%3BAndonian+S%3C%2Fauthor%3E%3CAN%3E24703459%3C%2FAN%3E%3CDT%3EJournal+Article%3C%2FDT%3E>

57. **Underuse of 24-hour urine collection among children with incident urinary stones: a quality-of-care concern?.**

**Item Type:** Journal Article

**Authors:** Ellison, Jonathan S.;Kaufman, Samuel R.;Kraft, Kate H.;Wolf, J. Stuart Jr;Hollenbeck, Brent K. and Hollingsworth, John M.

**Publication Date:** Au ,2014

**Journal:** Urology 84(2), pp. 457–461

**Abstract: OBJECTIVE**: To determine the prevalence of 24-hour urine collection among first-time pediatric stone formers and investigate factors associated with its use. **MATERIALS AND METHODS**: Using data from the MarketScan Commercial Claims and Encounters Database (2002-2006), children presenting to the emergency department with an incident episode of upper urinary tract stone disease were identified through a diagnosis code-based algorithm. The number of patients performing a 24-hour urine collection within 6 months of this encounter was calculated. Temporal trends in the prevalence of use were assessed. We fitted multivariate logistic regression models to examine factors associated with testing. **RESULTS**: In total, 1848 children presented with an incident upper tract stone, of which 12.0% submitted a 24-hour urine collection. This percentage remained stable over the study period. Testing was more common among younger patients (P : In total, 1848 children presented with an incident upper tract stone, of which 12.0% submitted a 24-hour urine collection. This percentage remained stable over the study period. Testing was more common among younger patients (P : In total, 1848 children presented with an incident upper tract stone, of which 12.0% submitted a 24-hour urine collection. This percentage remained stable over the study period. Testing was more common among younger patients (P : In total, 1848 children presented with an incident upper tract stone, of which 12.0% submitted a 24-hour urine collection. This percentage remained stable over the study period. Testing was more common among younger patients (P **CONCLUSION**: Despite rates of stone recurrence, 24-hour urine collection appears to be underused among children. Efforts to increase its use are therefore likely to benefit pediatric patients with urinary stone disease. Copyright Published by Elsevier Inc.

**URL:** <https://ovidsp.ovid.com/ovidweb.cgi?T=JS&CSC=Y&NEWS=N&PAGE=fulltext&D=med11&DO=10.1016%2fj.urology.2014.04.035> <https://libkey.io/libraries/2592/openurl?genre=article&sid=OVID:medline&id=pmid:24958480&id=doi:10.1016%2Fj.urology.2014.04.035&issn=0090-4295&isbn=&volume=84&issue=2&spage=457&pages=457-61&date=2014&title=Urology&atitle=Underuse+of+24-hour+urine+collection+among+children+with+incident+urinary+stones%3A+a+quality-of-care+concern%3F.&aulast=Ellison&pid=%3Cauthor%3EEllison+JS%3BKaufman+SR%3BKraft+KH%3BWolf+JS+Jr%3BHollenbeck+BK%3BHollingsworth+JM%3C%2Fauthor%3E%3CAN%3E24958480%3C%2FAN%3E%3CDT%3EJournal+Article%3C%2FDT%3E>

58. **Comparison of the metabolic profile of mixed calcium oxalate/uric acid stone formers to that of pure calcium oxalate and pure uric acid stone formers.**

**Item Type:** Journal Article

**Authors:** Friedlander, Justin I.;Moreira, Daniel M.;Hartman, Christopher;Elsamra, Sammy E.;Smith, Arthur D. and Okeke, Zeph

**Publication Date:** Au ,2014a

**Journal:** Urology 84(2), pp. 289–294

**Abstract: OBJECTIVE**: To compare the metabolic profile of patients who form mixed calcium oxalate (CaOx)/uric acid (UA) stones to those of pure CaOx and pure UA stone formers. **METHODS**: We performed a retrospective review of 232 patients, with both stone composition analysis and 24-hour urine collection, seen between March 2002 and April 2012. Analysis of 24-hour urine constituents across the 3 stone groups (pure UA, pure CaOx, and mixed CaOx/UA) was performed using univariate analysis of variance and multivariate linear regression models adjusting for clinical and demographic factors and 24-hour urine collection elements. **RESULTS**: A total of 27 patients (11.6%) had mixed CaOx/UA, 122 (52.6%) had pure CaOx, and 83 (35.8%) had pure UA calculi. Univariate analysis demonstrated significant differences between mixed CaOx/UA patients and pure CaOx patients for urine pH (mixed, 5.63 +/- 0.49 vs pure, CaOx 5.93 +/- 0.51; P = .009) and supersaturation (SS) UA (mixed, 1.84 +/- 1.09 vs pure, CaOx 1.26 +/- 0.93; P = .01), and a significant difference between mixed CaOx/UA patients and pure UA patients for SS CaOx (mixed, 7.18 +/- 4.23 vs pure, UA 4.90 +/- 2.96; P = .005). Multivariate analysis demonstrated that mixed CaOx/UA patients had no significant difference in SS CaOx as compared with pure CaOx patients (difference, -0.27; P = .66), whereas at the same time had no significant difference in SS UA as compared with pure UA patients (-0.07; P = .69). **CONCLUSION**: The metabolic profile of patients who form mixed CaOx/UA stones demonstrates abnormalities that promote both CaOx and UA stone formation. Dietary and medical management for this group of patients should address treatment of both defects. Copyright © 2014 Elsevier Inc. All rights reserved.

**URL:** <https://ovidsp.ovid.com/ovidweb.cgi?T=JS&CSC=Y&NEWS=N&PAGE=fulltext&D=med11&DO=10.1016%2fj.urology.2014.04.019> <https://libkey.io/libraries/2592/openurl?genre=article&sid=OVID:medline&id=pmid:24929942&id=doi:10.1016%2Fj.urology.2014.04.019&issn=0090-4295&isbn=&volume=84&issue=2&spage=289&pages=289-94&date=2014&title=Urology&atitle=Comparison+of+the+metabolic+profile+of+mixed+calcium+oxalate%2Furic+acid+stone+formers+to+that+of+pure+calcium+oxalate+and+pure+uric+acid+stone+formers.&aulast=Friedlander&pid=%3Cauthor%3EFriedlander+JI%3BMoreira+DM%3BHartman+C%3BElsamra+SE%3BSmith+AD%3BOkeke+Z%3C%2Fauthor%3E%3CAN%3E24929942%3C%2FAN%3E%3CDT%3EComparative+Study%3C%2FDT%3E>

59. **Age-related changes in 24-hour urine composition must be considered in the medical management of nephrolithiasis.**

**Item Type:** Journal Article

**Authors:** Friedlander, Justin I.;Moreira, Daniel M.;Hartman, Christopher;Elsamra, Sammy E.;Smith, Arthur D. and Okeke, Zeph

**Publication Date:** Ju ,2014b

**Journal:** Journal of Endourology 28(7), pp. 871–876

**Abstract: BACKGROUND AND PURPOSE**: Over the past 50 years, there has been an upward shift in the age of peak incidence of stone disease, yet less is known regarding how the urinary biochemical profile changes with aging. Therefore, we sought to examine the relationship between age and 24-hour urine composition. **METHODS**: We retrospectively reviewed a database of our tertiary care stone clinic patients seen from March 2002 to February 2012. Analysis of pretreatment 24-hour urine collections across age groups was performed using univariate analysis of variance and multivariate linear regression models adjusting for clinical and demographic factors and 24-hour urine parameters. **RESULTS**: A total of 1115 patients were broken down into age groups consisting of =65 years (356; 31.9%). Univariate analysis found significant ascending trends with aging for mean body mass index, while mean urine pH, 24-hour calcium, uric acid (UA), ammonium, creatinine, and supersaturation (SS) of calcium oxalate (CaOx) and calcium phosphate (CaP) decreased with age (all P for trend =65 years (356; 31.9%). Univariate analysis found significant ascending trends with aging for mean body mass index, while mean urine pH, 24-hour calcium, uric acid (UA), ammonium, creatinine, and supersaturation (SS) of calcium oxalate (CaOx) and calcium phosphate (CaP) decreased with age (all P for trend =65 years (356; 31.9%). Univariate analysis found significant ascending trends with aging for mean body mass index, while mean urine pH, 24-hour calcium, uric acid (UA), ammonium, creatinine, and supersaturation (SS) of calcium oxalate (CaOx) and calcium phosphate (CaP) decreased with age (all P for trend **CONCLUSIONS**: There are numerous age-related changes in the metabolic profile as seen on 24-hour urine collection. This highlights the importance of evaluating stone-forming patients of all ages with 24-hour urine collections because both the type and degree of metabolic abnormality may change with age.

**URL:** <https://ovidsp.ovid.com/ovidweb.cgi?T=JS&CSC=Y&NEWS=N&PAGE=fulltext&D=med11&DO=10.1089%2fend.2014.0002> <https://libkey.io/libraries/2592/openurl?genre=article&sid=OVID:medline&id=pmid:24571654&id=doi:10.1089%2Fend.2014.0002&issn=0892-7790&isbn=&volume=28&issue=7&spage=871&pages=871-6&date=2014&title=Journal+of+Endourology&atitle=Age-related+changes+in+24-hour+urine+composition+must+be+considered+in+the+medical+management+of+nephrolithiasis.&aulast=Friedlander&pid=%3Cauthor%3EFriedlander+JI%3BMoreira+DM%3BHartman+C%3BElsamra+SE%3BSmith+AD%3BOkeke+Z%3C%2Fauthor%3E%3CAN%3E24571654%3C%2FAN%3E%3CDT%3EJournal+Article%3C%2FDT%3E>

60. **Prospective evaluation of 24-hour urine profiles following bariatric surgery in a modern comprehensive care bariatric clinic.**

**Item Type:** Journal Article

**Authors:** Hutchinson, Ryan;Parker, Alexander S.;Arnold, Michelle;Bowers, Steven P.;Haley, William E.;Diehl, Nancy;Stone, Ronald;Lynch, Scott A.;Smith, C. Daniel and Thiel, David D.

**Publication Date:** Ma ,2014

**Journal:** Clinical Nephrology 81(5), pp. 331–337

**Abstract: AIMS**: To prospectively examine the effect of modern bariatric surgery on 24-hour urine parameters in a comprehensive care bariatric practice (CCBP). **MATERIALS AND METHODS**: 47 consecutive patients in our CCBP underwent serum and 24-hour urine analysis pre-operatively, and 30 returned at 12 months for repeat testing. Paired comparisons for serum metabolite and 24-hour urine measures were performed using a Wilcoxon signed-rank test for continuous variables and McNemar's test for categorical variables. Statistical tests were two-sided, with threshold of significance set at p = 0.05. **RESULTS**: All 30 patients with pre-operative and 12-month follow-up analysis were free of stone events. 20 (67%) had Roux-en-Y gastric bypass (RYGB), 6 (20%) had laparoscopic gastric banding (LGB), and 4 (13%)h ad laparoscopic sleeve gastrectomy (LSG). 24-hour urinary parameters were available for 27 patients. Median urine oxalate (mmol) was 0.29 pre-operatively and 0.21 at 12 months (p = 0.048). Median urine calcium (mg) was 143 pre-operatively and 180 at 12 months (p = 0.11). Median citrate excretion was 527 pre-operatively and 782 at 12 months (p = 0.22). Median serum creatinine was 0.7 pre-operatively and 0.8 at 12 months (p : All 30 patients with pre-operative and 12-month follow-up analysis were free of stone events. 20 (67%) had Roux-en-Y gastric bypass (RYGB), 6 (20%) had laparoscopic gastric banding (LGB), and 4 (13%)h ad laparoscopic sleeve gastrectomy (LSG). 24-hour urinary parameters were available for 27 patients. Median urine oxalate (mmol) was 0.29 pre-operatively and 0.21 at 12 months (p = 0.048). Median urine calcium (mg) was 143 pre-operatively and 180 at 12 months (p = 0.11). Median citrate excretion was 527 pre-operatively and 782 at 12 months (p = 0.22). Median serum creatinine was 0.7 pre-operatively and 0.8 at 12 months (p **CONCLUSIONS**: Modern bariatric surgery (RYGB, LGB, and LSG) as part of a CCBP can still demonstrate alterations of select urinary parameters (particularly oxalate and citrate) in select patients associated with an increased risk of urolithiasis at 1 year follow-up.

**URL:** <https://ovidsp.ovid.com/ovidweb.cgi?T=JS&CSC=Y&NEWS=N&PAGE=fulltext&D=med11&DO=10.5414%2fCN108154> <https://libkey.io/libraries/2592/openurl?genre=article&sid=OVID:medline&id=pmid:24495903&id=doi:10.5414%2FCN108154&issn=0301-0430&isbn=&volume=81&issue=5&spage=331&pages=331-7&date=2014&title=Clinical+Nephrology&atitle=Prospective+evaluation+of+24-hour+urine+profiles+following+bariatric+surgery+in+a+modern+comprehensive+care+bariatric+clinic.&aulast=Hutchinson&pid=%3Cauthor%3EHutchinson+R%3BParker+AS%3BArnold+M%3BBowers+SP%3BHaley+WE%3BDiehl+N%3BStone+R%3BLynch+SA%3BSmith+CD%3BThiel+DD%3C%2Fauthor%3E%3CAN%3E24495903%3C%2FAN%3E%3CDT%3EJournal+Article%3C%2FDT%3E>

61. **Hypertriglyceridemia is associated with increased risk for stone recurrence in patients with urolithiasis.**

**Item Type:** Journal Article

**Authors:** Kang, Ho Won;Seo, Sung Pil;Kim, Won Tae;Kim, Yong-June;Yun, Seok-Joong;Lee, Sang-Cheol and Kim, Wun-Jae

**Publication Date:** Oc ,2014

**Journal:** Urology 84(4), pp. 766–771

**Abstract: OBJECTIVE**: To assess the influence of dyslipidemia on urinary lithogenic metabolites and stone recurrence in stone formers. **MATERIALS AND METHODS**: We retrospectively selected 321 patients with urolithiasis who had been followed up for >24 months between 2004 and 2009. Fasting blood samples were taken, and serum lipid profiles were measured. All subjects also underwent 24-hour urinary metabolic evaluation and stone analysis. The radiographic appearance of new stones was defined as stone recurrence. **RESULTS**: There was no significant correlation between lipid profiles and 24-hour urine metabolites (all P >.05). Stone formers with hypertriglyceridemia had significantly higher urinary calcium, sodium, uric acid, magnesium, and potassium excretions. Only in a subgroup of uric acid stone, hypertriglyceridemia was significantly associated with decreased urinary pH. Those with low high-density lipoprotein (HDL) cholesterolemia had higher urinary sodium, magnesium, and potassium excretions, whereas those with high low-density lipoprotein (LDL) cholesterolemia had lower urinary sodium excretion. Stone analysis revealed that uric acid stones were more commonly found in patients with hypertriglyceridemia and low-HDL cholesterolemia. After a median follow-up of 35.0 months, 109 patients (34% of cohort) had stone recurrence. Stone recurrence was more common in the hypertriglyceridemia group compared with the normal triglyceridemia group (45.9% vs 29.7%; P = .005). The multivariate Cox regression model revealed that hypertriglyceridemia is associated independently with stone recurrence (hazard ratio, 1.857; 95% confidence interval, 1.211-2.847; P = .005). Kaplan-Meier curves showed similar results. **CONCLUSION**: Our study showed that serum lipid profile is associated with urine metabolic alterations. More importantly, hypertriglyceridemia is independently associated with increased risk for stone recurrence in patients with urolithiasis. Copyright © 2014 Elsevier Inc. All rights reserved.

**URL:** <https://ovidsp.ovid.com/ovidweb.cgi?T=JS&CSC=Y&NEWS=N&PAGE=fulltext&D=med11&DO=10.1016%2fj.urology.2014.06.013> <https://libkey.io/libraries/2592/openurl?genre=article&sid=OVID:medline&id=pmid:25106943&id=doi:10.1016%2Fj.urology.2014.06.013&issn=0090-4295&isbn=&volume=84&issue=4&spage=766&pages=766-71&date=2014&title=Urology&atitle=Hypertriglyceridemia+is+associated+with+increased+risk+for+stone+recurrence+in+patients+with+urolithiasis.&aulast=Kang&pid=%3Cauthor%3EKang+HW%3BSeo+SP%3BKim+WT%3BKim+YJ%3BYun+SJ%3BLee+SC%3BKim+WJ%3C%2Fauthor%3E%3CAN%3E25106943%3C%2FAN%3E%3CDT%3EJournal+Article%3C%2FDT%3E>

62. **Fish oil supplementation and urinary oxalate excretion in normal subjects on a low-oxalate diet.**

**Item Type:** Journal Article

**Authors:** Lange, Jessica N.;Mufarrij, Patrick W.;Easter, Linda;Knight, John;Holmes, Ross P. and Assimos, Dean G.

**Publication Date:** Oc ,2014

**Journal:** Urology 84(4), pp. 779–781

**Abstract: OBJECTIVE**: To determine if fish oil supplementation reduces endogenous oxalate synthesis in healthy subjects. **MATERIALS AND METHODS**: Fifteen healthy non-stone-forming adults participated in this study. Subjects first abstained from using vitamins, medications, or foods enriched in omega-3 fatty acids for 30 days. Next, they collected two 24-hour urine specimens while consuming a self-selected diet. Subjects consumed an extremely low-oxalate and normal-calcium diet for 5 days and collected 24-hour urine specimens on the last 3 days of this diet. Next, the subjects took 2 fish oil capsules containing 650-mg eicosapentaenoic acid and 450-mg docosahexaenoic acid twice daily for 30 days. They consumed a self-selected diet on days 1-25 and the controlled diet on days 26-30. Twenty-four-hour urine samples were collected on days 28-30. Excretion levels of urinary analytes including oxalate and glycolate were analyzed. **RESULTS**: Although there was a significant reduction in urinary oxalate, magnesium, and potassium excretions and an increase in uric acid excretion during the controlled dietary phases compared with the self-selected diet, there were no significant differences in their excretion during controlled diet phases with and without fish oil supplementation. **CONCLUSION**: These results suggest that fish oil supplementation does not reduce endogenous oxalate synthesis or urinary oxalate excretion in normal adults during periods of extremely low oxalate intake. However, these results do not challenge the previously described reduction in urinary oxalate excretion demonstrated in normal subjects consuming a moderate amount of oxalate in conjunction with fish oil. Copyright © 2014 Elsevier Inc. All rights reserved.

**URL:** <https://ovidsp.ovid.com/ovidweb.cgi?T=JS&CSC=Y&NEWS=N&PAGE=fulltext&D=med11&DO=10.1016%2fj.urology.2014.04.052> <https://libkey.io/libraries/2592/openurl?genre=article&sid=OVID:medline&id=pmid:25102784&id=doi:10.1016%2Fj.urology.2014.04.052&issn=0090-4295&isbn=&volume=84&issue=4&spage=779&pages=779-81&date=2014&title=Urology&atitle=Fish+oil+supplementation+and+urinary+oxalate+excretion+in+normal+subjects+on+a+low-oxalate+diet.&aulast=Lange&pid=%3Cauthor%3ELange+JN%3BMufarrij+PW%3BEaster+L%3BKnight+J%3BHolmes+RP%3BAssimos+DG%3C%2Fauthor%3E%3CAN%3E25102784%3C%2FAN%3E%3CDT%3EJournal+Article%3C%2FDT%3E>

63. **Body fat content and distribution and urinary risk factors for nephrolithiasis.**

**Item Type:** Journal Article

**Authors:** Pigna, Federica;Sakhaee, Khashayar;Adams-Huet, Beverley and Maalouf, Naim M.

**Publication Date:** Ja ,2014

**Journal:** Clinical Journal of the American Society of Nephrology: CJASN 9(1), pp. 159–165

**Abstract: BACKGROUND AND OBJECTIVES**: Obesity is associated with a higher risk of nephrolithiasis. However, it is not known whether higher body fat mass or abnormal fat distribution influences stone risk independent of dietary factors. **DESIGN, SETTING, PARTICIPANTS, & MEASUREMENTS**: In this cross-sectional study, non-stone-forming men with no known kidney disease and with a wide range of body weight collected a 24-hour urine specimen while consuming a fixed metabolic diet. They underwent dual-energy x-ray absorptiometry to assess body composition and fat distribution. Urinary risk factors for nephrolithiasis and urine saturation with respect to calcium oxalate and uric acid (assessed as supersaturation index [SI]) were correlated with various measures of adiposity. **RESULTS**: Study participants included 21 men with a mean age of 52.1 years, mean weight of 91.1 kg, and mean total fat mass of 24.3 kg. Twenty-four-hour urine pH and SI uric acid were more closely correlated with fat mass than with lean mass or total body weight. Both 24-hour urine pH and SI uric acid were also significantly correlated with truncal fat mass but not with leg fat mass. Moreover, there was a significant negative correlation between truncal/leg fat mass and NH4(+)/net acid excretion ratio (R=-0.62; P=0.009). However, there was no significant association between SI calcium oxalate and body weight, lean mass, fat mass, trunk fat mass, or leg fat mass. **CONCLUSIONS**: The association between 24-hour urine pH and SI uric acid and various measures of adiposity suggest that total body fat and trunk fat are more strongly associated with risk factors for uric acid stone formation than are total body weight and lean body mass. Under a controlled metabolic diet, adiposity is not associated with risk factors for calcium oxalate stones. Further studies are needed to confirm these findings in larger populations that include women and patients who form stones.

**URL:** <https://ovidsp.ovid.com/ovidweb.cgi?T=JS&CSC=Y&NEWS=N&PAGE=fulltext&D=med11&DO=10.2215%2fCJN.06180613> <https://libkey.io/libraries/2592/openurl?genre=article&sid=OVID:medline&id=pmid:24202136&id=doi:10.2215%2FCJN.06180613&issn=1555-9041&isbn=&volume=9&issue=1&spage=159&pages=159-65&date=2014&title=Clinical+Journal+of+The+American+Society+of+Nephrology%3A+CJASN&atitle=Body+fat+content+and+distribution+and+urinary+risk+factors+for+nephrolithiasis.&aulast=Pigna&pid=%3Cauthor%3EPigna+F%3BSakhaee+K%3BAdams-Huet+B%3BMaalouf+NM%3C%2Fauthor%3E%3CAN%3E24202136%3C%2FAN%3E%3CDT%3EJournal+Article%3C%2FDT%3E>

64. **Type-2 diabetes and kidney stones: impact of diabetes medications and glycemic control.**

**Item Type:** Journal Article

**Authors:** Torricelli, Fabio C. M.;De, Shubha;Gebreselassie, Surafel;Li, Ina;Sarkissian, Carl and Monga, Manoj

**Publication Date:** Se ,2014

**Journal:** Urology 84(3), pp. 544–548

**Abstract: OBJECTIVE**: To evaluate the impact of diabetic medications and glycemic control on the urine pH, 24-hour urine stone risk profile, and stone composition. **PATIENTS AND METHODS**: We retrospectively reviewed our database searching for type-2 diabetic patients with kidney stones from July 2002 to January 2013. Patients were divided in 2 groups according to their diabetic medications: insulin vs oral antihyperglycemics. Patients were compared based on their urine collections and stone composition. A linear regression was done to assess which variables could predict a low urine pH. In a subgroup analysis, patients on thiazolidinediones (ie, pioglitazone) were compared with patients on other oral antihyperglycemics. **RESULTS**: We analyzed 1831 type-2 diabetic patients with stone disease; 375 (20.5%) were included in the insulin group and 1456 (79.5%) in the antihyperglycemics group. Linear regression revealed male gender (P = .011) and insulin therapy (P : We analyzed 1831 type-2 diabetic patients with stone disease; 375 (20.5%) were included in the insulin group and 1456 (79.5%) in the antihyperglycemics group. Linear regression revealed male gender (P = .011) and insulin therapy (P : We analyzed 1831 type-2 diabetic patients with stone disease; 375 (20.5%) were included in the insulin group and 1456 (79.5%) in the antihyperglycemics group. Linear regression revealed male gender (P = .011) and insulin therapy (P : We analyzed 1831 type-2 diabetic patients with stone disease; 375 (20.5%) were included in the insulin group and 1456 (79.5%) in the antihyperglycemics group. Linear regression revealed male gender (P = .011) and insulin therapy (P **CONCLUSION**: Urine pH is inversely related to HbA1c level. Insulin therapy is associated with higher urine pH than oral antihyperglycemic agents despite higher HbA1c suggesting that insulin may modify urine pH independent of glycemic control. Copyright © 2014 Elsevier Inc. All rights reserved.

**URL:** <https://ovidsp.ovid.com/ovidweb.cgi?T=JS&CSC=Y&NEWS=N&PAGE=fulltext&D=med11&DO=10.1016%2fj.urology.2014.02.074> <https://libkey.io/libraries/2592/openurl?genre=article&sid=OVID:medline&id=pmid:25168531&id=doi:10.1016%2Fj.urology.2014.02.074&issn=0090-4295&isbn=&volume=84&issue=3&spage=544&pages=544-8&date=2014&title=Urology&atitle=Type-2+diabetes+and+kidney+stones%3A+impact+of+diabetes+medications+and+glycemic+control.&aulast=Torricelli&pid=%3Cauthor%3ETorricelli+FC%3BDe+S%3BGebreselassie+S%3BLi+I%3BSarkissian+C%3BMonga+M%3C%2Fauthor%3E%3CAN%3E25168531%3C%2FAN%3E%3CDT%3EJournal+Article%3C%2FDT%3E>

65. **Use of the probability of stone formation (PSF) score to assess stone forming risk and treatment response in a cohort of Brazilian stone formers.**

**Item Type:** Journal Article

**Authors:** Turney, Benjamin;Robertson, William;Wiseman, Oliver;Amaro, Carmen Regina P. R.;Leitao, Victor A.;Silva, Isabela Leme da and Amaro, Joao Luiz

**Publication Date:** 2014

**Journal:** International Braz J Urol 40(4), pp. 507–512

**Abstract: INTRODUCTION**: The aim was to confirm that PSF (probability of stone formation) changed appropriately following medical therapy on recurrent stone formers. **MATERIALS AND METHODS**: Data were collected on 26 Brazilian stone-formers. A baseline 24-hour urine collection was performed prior to treatment. Details of the medical treatment initiated for stone-disease were recorded. A PSF calculation was performed on the 24 hour urine sample using the 7 urinary parameters required: voided volume, oxalate, calcium, urate, pH, citrate and magnesium. A repeat 24-hour urine sample was performed for PSF calculation after treatment. Comparison was made between the PSF scores before and during treatment. **RESULTS**: At baseline, 20 of the 26 patients (77%) had a high PSF score (> 0.5). Of the 26 patients, 17 (65%) showed an overall reduction in their PSF profiles with a medical treatment regimen. Eleven patients (42%) changed from a high risk (PSF > 0.5) to a low risk (PSF 0.5) during both assessments. **CONCLUSIONS**: The PSF score reduced following medical treatment in the majority of patients in this cohort.

**URL:** <https://ovidsp.ovid.com/ovidweb.cgi?T=JS&CSC=Y&NEWS=N&PAGE=fulltext&D=med11&DO=10.1590%2fS1677-5538.IBJU.2014.04.09> <https://libkey.io/libraries/2592/openurl?genre=article&sid=OVID:medline&id=pmid:25251955&id=doi:10.1590%2FS1677-5538.IBJU.2014.04.09&issn=1677-5538&isbn=&volume=40&issue=4&spage=507&pages=507-12&date=2014&title=International+Braz+J+Urol&atitle=Use+of+the+probability+of+stone+formation+%28PSF%29+score+to+assess+stone+forming+risk+and+treatment+response+in+a+cohort+of+Brazilian+stone+formers.&aulast=Turney&pid=%3Cauthor%3ETurney+B%3BRobertson+W%3BWiseman+O%3BAmaro+CR%3BLeitao+VA%3BSilva+IL%3BAmaro+JL%3C%2Fauthor%3E%3CAN%3E25251955%3C%2FAN%3E%3CDT%3EJournal+Article%3C%2FDT%3E>

66. **The characteristics of the stone and urine composition in Chinese stone formers: primary report of a single-center results.**

**Item Type:** Journal Article

**Authors:** Wu, Wenqi;Yang, Dong;Tiselius, Hans-Goran;Ou, Lili;Liang, Yeping;Zhu, Hanliang;Li, Shujue and Zeng, Guohua

**Publication Date:** Ap ,2014

**Journal:** Urology 83(4), pp. 732–737

**Abstract: OBJECTIVE**: To assess urine composition in Chinese patients with urolithiasis. **METHODS**: Five hundred seven Chinese patients with urolithiasis from our center in southern China were included in this study. Analysis of stone composition was performed using infrared spectrometry. From all patients, 24-hour urine samples were collected for analysis of urinary variables. Some ion activity product risk indices were also calculated. **RESULTS**: The major stone constituents in the 507 analyzed stones were as follows: calcium oxalate (78.3%), infection stone components (14.6%), uric acid (3.6%), and calcium phosphate (3.4%). Only 1 stone was composed of cystine (0.2%). Of all patients, 504 (99.4%) had 1 or several urinary metabolic abnormalities. Hypocitraturia was recorded in 93.9%, high sodium excretion in 58.6%, small urine volume in 45.6%, hyperoxaluria in 31.0%, hypercalciuria in 26.0%, hyperuricosuria in 19.3%, and hyperphosphaturia in 2.8%. Moreover, high sodium excretion was more frequent in men than women (59.2% vs 49.3%, P = .027), whereas hypercalciuria was more common in women (34.5% vs 20.4%, P : The major stone constituents in the 507 analyzed stones were as follows: calcium oxalate (78.3%), infection stone components (14.6%), uric acid (3.6%), and calcium phosphate (3.4%). Only 1 stone was composed of cystine (0.2%). Of all patients, 504 (99.4%) had 1 or several urinary metabolic abnormalities. Hypocitraturia was recorded in 93.9%, high sodium excretion in 58.6%, small urine volume in 45.6%, hyperoxaluria in 31.0%, hypercalciuria in 26.0%, hyperuricosuria in 19.3%, and hyperphosphaturia in 2.8%. Moreover, high sodium excretion was more frequent in men than women (59.2% vs 49.3%, P = .027), whereas hypercalciuria was more common in women (34.5% vs 20.4%, P : The major stone constituents in the 507 analyzed stones were as follows: calcium oxalate (78.3%), infection stone components (14.6%), uric acid (3.6%), and calcium phosphate (3.4%). Only 1 stone was composed of cystine (0.2%). Of all patients, 504 (99.4%) had 1 or several urinary metabolic abnormalities. Hypocitraturia was recorded in 93.9%, high sodium excretion in 58.6%, small urine volume in 45.6%, hyperoxaluria in 31.0%, hypercalciuria in 26.0%, hyperuricosuria in 19.3%, and hyperphosphaturia in 2.8%. Moreover, high sodium excretion was more frequent in men than women (59.2% vs 49.3%, P = .027), whereas hypercalciuria was more common in women (34.5% vs 20.4%, P : The major stone constituents in the 507 analyzed stones were as follows: calcium oxalate (78.3%), infection stone components (14.6%), uric acid (3.6%), and calcium phosphate (3.4%). Only 1 stone was composed of cystine (0.2%). Of all patients, 504 (99.4%) had 1 or several urinary metabolic abnormalities. Hypocitraturia was recorded in 93.9%, high sodium excretion in 58.6%, small urine volume in 45.6%, hyperoxaluria in 31.0%, hypercalciuria in 26.0%, hyperuricosuria in 19.3%, and hyperphosphaturia in 2.8%. Moreover, high sodium excretion was more frequent in men than women (59.2% vs 49.3%, P = .027), whereas hypercalciuria was more common in women (34.5% vs 20.4%, P **CONCLUSION**: The results of urinary risk factors for stone formation in this study might serve as a basis for design of recurrence prevention. It is of interest to note that some of the demonstrated abnormalities differ from that in reports from other countries. Copyright © 2014 Elsevier Inc. All rights reserved.

**URL:** <https://ovidsp.ovid.com/ovidweb.cgi?T=JS&CSC=Y&NEWS=N&PAGE=fulltext&D=med11&DO=10.1016%2fj.urology.2013.11.012> <https://libkey.io/libraries/2592/openurl?genre=article&sid=OVID:medline&id=pmid:24485999&id=doi:10.1016%2Fj.urology.2013.11.012&issn=0090-4295&isbn=&volume=83&issue=4&spage=732&pages=732-7&date=2014&title=Urology&atitle=The+characteristics+of+the+stone+and+urine+composition+in+Chinese+stone+formers%3A+primary+report+of+a+single-center+results.&aulast=Wu&pid=%3Cauthor%3EWu+W%3BYang+D%3BTiselius+HG%3BOu+L%3BLiang+Y%3BZhu+H%3BLi+S%3BZeng+G%3C%2Fauthor%3E%3CAN%3E24485999%3C%2FAN%3E%3CDT%3EJournal+Article%3C%2FDT%3E>

67. **Variation in urinary stone composition between adult Uyghur and Han patients with urolithiasis in Xinjiang, China.**

**Item Type:** Journal Article

**Authors:** Wumaner, Aikebaier;Keremu, Aziguli;Wumaier, Dilixiati and Wang, Qing

**Publication Date:** Oc ,2014

**Journal:** Urology 84(4), pp. 772–778

**Abstract: OBJECTIVE**: To analyze variations in urinary stone composition between Uyghur and Han patients with urolithiasis in Xinjiang, China, and to explore the possible factors associated with urinary stone composition in Uyghur and Han patients. **MATERIALS AND METHODS**: We analyzed the components of urinary stones in 317 adults (152 Uyghur and 165 Han) with urolithiasis admitted to our hospital from March 2009 to June 2011. Urinary stones were collected by endoscopic or open surgical procedures and analyzed by infrared spectroscopy. Also analyzed were clinical data, including patient's age, sex, geographic distribution, blood tests, and urine tests. Logistic regression was performed to analyze the association between stone components and the clinical characteristics of the patients. **RESULTS**: There were significant differences between Uyghur and Han patients in age, geographic distribution, serum concentrations of uric acid, potassium, and phosphorus, urine magnesium concentrations, 24-hour volumes of urine, and in stone components of calcium oxalate monohydrate, calcium oxalate dihydrate plus carbapatite, and uric acid. Differences in sex, family history, complications, sites of calculi, or other blood and urine test results were not significant. Logistic regression analysis revealed that geographic location, ethnicity, blood potassium concentration, and urine volume were significantly correlated with the calcium oxalate dihydrate plus carbapatite component of urinary stones. **CONCLUSION**: Urinary stone composition differs between Uyghur and Han patients with urolithiasis, possibly as a result of geographic distribution. Copyright © 2014 Elsevier Inc. All rights reserved.

**URL:** <https://ovidsp.ovid.com/ovidweb.cgi?T=JS&CSC=Y&NEWS=N&PAGE=fulltext&D=med11&DO=10.1016%2fj.urology.2014.04.056> <https://libkey.io/libraries/2592/openurl?genre=article&sid=OVID:medline&id=pmid:25109561&id=doi:10.1016%2Fj.urology.2014.04.056&issn=0090-4295&isbn=&volume=84&issue=4&spage=772&pages=772-8&date=2014&title=Urology&atitle=Variation+in+urinary+stone+composition+between+adult+Uyghur+and+Han+patients+with+urolithiasis+in+Xinjiang%2C+China.&aulast=Wumaner&pid=%3Cauthor%3EWumaner+A%3BKeremu+A%3BWumaier+D%3BWang+Q%3C%2Fauthor%3E%3CAN%3E25109561%3C%2FAN%3E%3CDT%3EComparative+Study%3C%2FDT%3E>

68. **Initial presentation in psychiatry emergency room led to diagnosis of many urinary bladder stones in a male patient.**

**Item Type:** Journal Article

**Authors:** El-Hennawy, Adel S.;Nagaraja, Aarathi and Mahmood, Aza K.

**Publication Date:** Ja ,2013

**Journal:** American Journal of Therapeutics 20(1), pp. 107–110

**Abstract:** The first case of man who presented to psychiatry emergency room for evaluation of abnormal behavior because of urinary stones was reported. Careful evaluation of patient led to a diagnosis of 37 urinary bladder stones in an Egyptian man with obstructive uropathy and metabolic defects in the form of hyperoxaluria and hypocitraturia. Knowledge of the differential diagnosis of metabolic defects can lead to successful outcome in preventing reformation of urinary tract stones after surgery. A 61-year-old Egyptian man presented to psychiatry emergency room because he was found lying on floor in bathroom to urinate by his wife who thought her husband needed psychiatric evaluation. Patient gave history of frequent urination and dysuria on and off for 3 years. In the last 3 months before his presentation to emergency room, he got into a habit of lying down on his left side when he went to bathroom to urinate because it was easier for him to pass urine. Renal consultation requested because of presence of red blood cells in urinalysis. Computed tomography of the abdomen and pelvis showed bilateral hydronephrosis and multiple bladder stones. Twenty-four-hour urine collection showed low urinary citrate and high oxalate. Patient underwent open vesicolithotomy and removal of 36 stones. Stone analysis showed 75% uric acid and 25% calcium oxalate. Patient did very well after surgery, and 1 month later, he underwent transuretheral resection of prostate without any complications. Now patient has no difficulty passing urine and he has no recent attack of urinary tract infection. Knowledge of the differential diagnosis of metabolic defects in men with urinary bladder stones would hopefully provide clinicians with the proper diagnostic tools to more specifically treat such patients with improved success in preventing reformation of urinary tract stones after surgery.

**URL:** <https://ovidsp.ovid.com/ovidweb.cgi?T=JS&CSC=Y&NEWS=N&PAGE=fulltext&D=med10&DO=10.1097%2fMJT.0b013e3182541ca2> <https://libkey.io/libraries/2592/openurl?genre=article&sid=OVID:medline&id=pmid:23299232&id=doi:10.1097%2FMJT.0b013e3182541ca2&issn=1075-2765&isbn=&volume=20&issue=1&spage=107&pages=107-10&date=2013&title=American+Journal+of+Therapeutics&atitle=Initial+presentation+in+psychiatry+emergency+room+led+to+diagnosis+of+many+urinary+bladder+stones+in+a+male+patient.&aulast=El-Hennawy&pid=%3Cauthor%3EEl-Hennawy+AS%3BNagaraja+A%3BMahmood+AK%3C%2Fauthor%3E%3CAN%3E23299232%3C%2FAN%3E%3CDT%3ECase+Reports%3C%2FDT%3E>

69. **24-hour urine collection in the metabolic evaluation of stone formers: is one study adequate?.**

**Item Type:** Journal Article

**Authors:** Healy, Kelly A.;Hubosky, Scott G. and Bagley, Demetrius H.

**Publication Date:** Ma ,2013

**Journal:** Journal of Endourology 27(3), pp. 374–378

**Abstract: UNLABELLED**: Abstract Purpose: To determine whether one vs two 24-hour urine collections is optimal in the metabolic evaluation of nephrolithiasis. **METHODS**: We retrospectively reviewed all 24-hour urine collections performed at our tertiary stone clinic from July 1997 to February 2012. We identified patients with two 24-hour urine collections performed : We retrospectively reviewed all 24-hour urine collections performed at our tertiary stone clinic from July 1997 to February 2012. We identified patients with two 24-hour urine collections performed **RESULTS**: A total of 813 subjects submitted two 24-hour urine collections 0.05), (range 0.061-0.9983). Pearson correlation demonstrated a high degree of correlation between two 24-hour urines for all variables (r=0.66-0.95, each P0.05), (range 0.061-0.9983). Pearson correlation demonstrated a high degree of correlation between two 24-hour urines for all variables (r=0.66-0.95, each P**CONCLUSIONS**: A single 24-hour urine collection may have changed clinical decision making in up to 45% of patients. Therefore, we recommend two collections to optimize the diagnostic yield and appropriately target stone prevention strategies.

**URL:** <https://ovidsp.ovid.com/ovidweb.cgi?T=JS&CSC=Y&NEWS=N&PAGE=fulltext&D=med10&DO=10.1089%2fend.2012.0216> <https://libkey.io/libraries/2592/openurl?genre=article&sid=OVID:medline&id=pmid:22967013&id=doi:10.1089%2Fend.2012.0216&issn=0892-7790&isbn=&volume=27&issue=3&spage=374&pages=374-8&date=2013&title=Journal+of+Endourology&atitle=24-hour+urine+collection+in+the+metabolic+evaluation+of+stone+formers%3A+is+one+study+adequate%3F.&aulast=Healy&pid=%3Cauthor%3EHealy+KA%3BHubosky+SG%3BBagley+DH%3C%2Fauthor%3E%3CAN%3E22967013%3C%2FAN%3E%3CDT%3EJournal+Article%3C%2FDT%3E>

70. **Twenty-four-hour urine constituents in stone formers: a study from the northeast part of Peninsular Malaysia.**

**Item Type:** Journal Article

**Authors:** Hussein, N. S.;Sadiq, S. M.;Kamaliah, M. D.;Norakmal, A. W. and Gohar, M. N.

**Publication Date:** Ma ,2013

**Journal:** Saudi Journal of Kidney Diseases & Transplantation 24(3), pp. 630–637

**Abstract:** Urolithiasis is a common disease with increasing incidence and prevalence world-wide, probably more common in industrialized countries. The metabolic evaluation of 24-h urine collection has been considered as part of the management of urinary stone patients. The aim of this study was to evaluate the 24-h urine constituents in stone formers and its relation to demographic data in the northeast part of Peninsular Malaysia. One hundred and six patients were recruited in this study from two hospitals in the same geographical region; 96 patients fulfilled the inclusion criteria and an informed consent was obtained from all subjects. The 24-h urine was collected in sterile bottles with a preservative agent and calcium, oxalate, citrate, uric acid, magnesium and phosphate were tested using commercial kits on a Roche Hitachi 912 chemistry analyzer. The age (mean +/- SD) of 96 patients was 56.45 +/- 13.43 years and 82.3% of the patients were male while 17.7% were female. The 24-h urine abnormalities were hypercalciuria (14.5%), hyperoxaluria (61.4%), hypocitraturia (57.2%), hyperuricouria (19.7%), hypomagnesuria (59.3%) and hyperphosphaturia (12.5%). Hyperoxaluria (61.4%) was the most common abnormality detected during the analysis of 24-h urine constituents in contradiction to industrial countries, where hypercalciuria was the most common finding. The high frequencies of hypomagnesuria and hypocitraturia reflect the important role of magnesium and citrate in stone formation and their prophylactic role in the treatment of urinary stone disease in the given population.

**URL:** <https://ovidsp.ovid.com/ovidweb.cgi?T=JS&CSC=Y&NEWS=N&PAGE=fulltext&D=med10&DO=10.4103%2f1319-2442.111090> <https://libkey.io/libraries/2592/openurl?genre=article&sid=OVID:medline&id=pmid:23640651&id=doi:10.4103%2F1319-2442.111090&issn=1319-2442&isbn=&volume=24&issue=3&spage=630&pages=630-7&date=2013&title=Saudi+Journal+of+Kidney+Diseases+%26+Transplantation&atitle=Twenty-four-hour+urine+constituents+in+stone+formers%3A+a+study+from+the+northeast+part+of+Peninsular+Malaysia.&aulast=Hussein&pid=%3Cauthor%3EHussein+NS%3BSadiq+SM%3BKamaliah+MD%3BNorakmal+AW%3BGohar+MN%3C%2Fauthor%3E%3CAN%3E23640651%3C%2FAN%3E%3CDT%3EJournal+Article%3C%2FDT%3E>

71. **Differences in 24-hour urine composition between apatite and brushite stone formers.**

**Item Type:** Journal Article

**Authors:** Moreira, Daniel M.;Friedlander, Justin I.;Hartman, Christopher;Elsamra, Sammy E.;Smith, Arthur D. and Okeke, Zeph

**Publication Date:** Oc ,2013

**Journal:** Urology 82(4), pp. 768–772

**Abstract: OBJECTIVE**: To determine the differences in 24-hour urine composition between apatite and brushite stone formers. **METHODS**: We conducted a retrospective review of 110 calcium phosphate (CaP) stone formers with both stone composition analysis and 24-hour urinalysis available. Analysis of 24-hour urine constituents between apatite and brushite stone formers was performed using univariable t test and multivariable linear regression models, adjusting for clinical and demographic factors. **RESULTS**: A total of 97 patients (88%) had predominantly apatite stones and 13 patients (12%) had predominantly brushite stones. In univariable analysis, brushite stone formers had significantly higher mean 24-hour urinary calcium excretion (apatite = 204.8 +/- 103.5 mg vs brushite = 329.7 +/- 136.6 mg, P = .007), higher mean super saturation (SS) CaP (apatite = 1.423 +/- 0.867 vs brushite = 2.576 +/- 0.171, P = .004) and lower mean SS uric acid (apatite = 0.688 +/- 0.796 vs brushite = 0.345 +/- 0.190, P : A total of 97 patients (88%) had predominantly apatite stones and 13 patients (12%) had predominantly brushite stones. In univariable analysis, brushite stone formers had significantly higher mean 24-hour urinary calcium excretion (apatite = 204.8 +/- 103.5 mg vs brushite = 329.7 +/- 136.6 mg, P = .007), higher mean super saturation (SS) CaP (apatite = 1.423 +/- 0.867 vs brushite = 2.576 +/- 0.171, P = .004) and lower mean SS uric acid (apatite = 0.688 +/- 0.796 vs brushite = 0.345 +/- 0.190, P : A total of 97 patients (88%) had predominantly apatite stones and 13 patients (12%) had predominantly brushite stones. In univariable analysis, brushite stone formers had significantly higher mean 24-hour urinary calcium excretion (apatite = 204.8 +/- 103.5 mg vs brushite = 329.7 +/- 136.6 mg, P = .007), higher mean super saturation (SS) CaP (apatite = 1.423 +/- 0.867 vs brushite = 2.576 +/- 0.171, P = .004) and lower mean SS uric acid (apatite = 0.688 +/- 0.796 vs brushite = 0.345 +/- 0.190, P : A total of 97 patients (88%) had predominantly apatite stones and 13 patients (12%) had predominantly brushite stones. In univariable analysis, brushite stone formers had significantly higher mean 24-hour urinary calcium excretion (apatite = 204.8 +/- 103.5 mg vs brushite = 329.7 +/- 136.6 mg, P = .007), higher mean super saturation (SS) CaP (apatite = 1.423 +/- 0.867 vs brushite = 2.576 +/- 0.171, P = .004) and lower mean SS uric acid (apatite = 0.688 +/- 0.796 vs brushite = 0.345 +/- 0.190, P **CONCLUSION**: In a cohort of CaP stone formers, brushite stones were associated with higher urinary calcium excretion and higher urinary SS CaP when compared to apatite stones. Aggressive measures to reduce urinary calcium may be particularly helpful to prevent brushite stone formation. Copyright © 2013 Elsevier Inc. All rights reserved.

**URL:** <https://ovidsp.ovid.com/ovidweb.cgi?T=JS&CSC=Y&NEWS=N&PAGE=fulltext&D=med10&DO=10.1016%2fj.urology.2013.04.025> <https://libkey.io/libraries/2592/openurl?genre=article&sid=OVID:medline&id=pmid:23791217&id=doi:10.1016%2Fj.urology.2013.04.025&issn=0090-4295&isbn=&volume=82&issue=4&spage=768&pages=768-72&date=2013&title=Urology&atitle=Differences+in+24-hour+urine+composition+between+apatite+and+brushite+stone+formers.&aulast=Moreira&pid=%3Cauthor%3EMoreira+DM%3BFriedlander+JI%3BHartman+C%3BElsamra+SE%3BSmith+AD%3BOkeke+Z%3C%2Fauthor%3E%3CAN%3E23791217%3C%2FAN%3E%3CDT%3EComparative+Study%3C%2FDT%3E>

72. **Influence of lean and fat mass on bone mineral density and on urinary stone risk factors in healthy women.**

**Item Type:** Journal Article

**Authors:** Nouvenne, Antonio;Ticinesi, Andrea;Guerra, Angela;Folesani, Giuseppina;Allegri, Franca;Pinelli, Silvana;Baroni, Paolo;Pedrazzoni, Mario;Lippi, Giuseppe;Terranegra, Annalisa;Dogliotti, Elena;Soldati, Laura;Borghi, Loris and Meschi, Tiziana

**Publication Date:** Oct 07 ,2013

**Journal:** Journal of Translational Medicine 11, pp. 248

**Abstract: BACKGROUND**: The role of body composition (lean mass and fat mass) on urine chemistries and bone quality is still debated. Our aim was therefore to determine the effect of lean mass and fat mass on urine composition and bone mineral density (BMD) in a cohort of healthy females. **MATERIALS AND METHODS**: 78 female volunteers (mean age 46 +/- 6 years) were enrolled at the Stone Clinic of Parma University Hospital and subdued to 24-hour urine collection for lithogenic risk profile, DEXA, and 3-day dietary diary. We defined two mathematical indexes derived from body composition measurement (index of lean mass-ILM, and index of fat mass-IFM) and the cohort was split using the median value of each index, obtaining groups differing only for lean or fat mass. We then analyzed differences in urine composition, dietary intakes and BMD. **RESULTS**: The women with high values of ILM had significantly higher excretion of creatinine (991 +/- 194 vs 1138 +/- 191 mg/day, p = 0.001), potassium (47 +/- 13 vs 60 +/- 18 mEq/day, p : The women with high values of ILM had significantly higher excretion of creatinine (991 +/- 194 vs 1138 +/- 191 mg/day, p = 0.001), potassium (47 +/- 13 vs 60 +/- 18 mEq/day, p : The women with high values of ILM had significantly higher excretion of creatinine (991 +/- 194 vs 1138 +/- 191 mg/day, p = 0.001), potassium (47 +/- 13 vs 60 +/- 18 mEq/day, p : The women with high values of ILM had significantly higher excretion of creatinine (991 +/- 194 vs 1138 +/- 191 mg/day, p = 0.001), potassium (47 +/- 13 vs 60 +/- 18 mEq/day, p **CONCLUSIONS**: Lean mass seems to significantly influence urine composition both in terms of lithogenesis promoters and inhibitors, while fat mass does not. Lean mass influences bone quality only in limb skeleton, while fat mass influences bone quality only in axial sites.

**URL:** <https://ovidsp.ovid.com/ovidweb.cgi?T=JS&CSC=Y&NEWS=N&PAGE=fulltext&D=med10&DO=10.1186%2f1479-5876-11-248> <https://libkey.io/libraries/2592/openurl?genre=article&sid=OVID:medline&id=pmid:24099643&id=doi:10.1186%2F1479-5876-11-248&issn=1479-5876&isbn=&volume=11&issue=&spage=248&pages=248&date=2013&title=Journal+of+Translational+Medicine&atitle=Influence+of+lean+and+fat+mass+on+bone+mineral+density+and+on+urinary+stone+risk+factors+in+healthy+women.&aulast=Nouvenne&pid=%3Cauthor%3ENouvenne+A%3BTicinesi+A%3BGuerra+A%3BFolesani+G%3BAllegri+F%3BPinelli+S%3BBaroni+P%3BPedrazzoni+M%3BLippi+G%3BTerranegra+A%3BDogliotti+E%3BSoldati+L%3BBorghi+L%3BMeschi+T%3C%2Fauthor%3E%3CAN%3E24099643%3C%2FAN%3E%3CDT%3EJournal+Article%3C%2FDT%3E>

73. **Biochemical determinants of severe lithogenic activity in patients with idiopathic calcium nephrolithiasis.**

**Item Type:** Journal Article

**Authors:** Arrabal-Polo, Miguel Angel;Arrabal-Martin, Miguel;de Haro-Munoz, Tomas;Poyatos-Andujar, Antonio;Palaeo-Yago, Francisco and Zuluaga-Gomez, Armando

**Publication Date:** Ja ,2012

**Journal:** Urology 79(1), pp. 48–54

**Abstract: OBJECTIVE**: To analyze the biochemical alterations in plasma and the urine determinants of severe lithogenic activity in patients with idiopathic calcium nephrolithiasis. **METHODS**: We performed a cross-sectional study of 120 patients divided into 2 groups: group 1, 60 patients without nephrolithiasis; and group 2, 60 patients with severe and/or recurrent calcium nephrolithiasis. In all patients, a study of renal function, calcium metabolism, and bone remodeling markers, and a study of the lithogenic factors were performed in urine after fasting and in 24-hour urine samples. **RESULTS**: We observed greater values for phosphorus in group 1 than in group 2 (P=.03). Also, we found greater values for intact parathyroid hormone (P=.01), osteocalcin (P=.000), and beta-crosslaps (P=.000) in group 2 than in group 1. In the 24-hour urine samples, significant differences were found between groups 1 and 2 in calciuria (11.7 vs 17.4 mg/dL; P=.000), citraturia (50.6 vs 33.5 mg/dL; P=.002), calcium/creatinine quotient (0.14 vs 0.20; P=.001), calcium/citrate quotient (0.05 vs 0.13; P=.04), and calcium/creatinine quotient after fasting (0.09 vs 0.16; P=.000). **CONCLUSION**: We consider the determinants of severe and/or recurrent calcium lithiasis to be hypercalciuria and hypocitraturia and a calcium/citrate quotient>0.06. As risk markers we can consider phosphatemia35, alkaline phosphatase>80 U/L, intact parathyroid hormone>60 pg/mL, osteocalcin>16 ng/mL, beta-crosslaps>0.400 ng/mL, and beta-crosslaps/osteocalcin quotient>0.028. Copyright © 2012 Elsevier Inc. All rights reserved.

**URL:** <https://ovidsp.ovid.com/ovidweb.cgi?T=JS&CSC=Y&NEWS=N&PAGE=fulltext&D=med9&DO=10.1016%2fj.urology.2011.07.1382> <https://libkey.io/libraries/2592/openurl?genre=article&sid=OVID:medline&id=pmid:21908029&id=doi:10.1016%2Fj.urology.2011.07.1382&issn=0090-4295&isbn=&volume=79&issue=1&spage=48&pages=48-54&date=2012&title=Urology&atitle=Biochemical+determinants+of+severe+lithogenic+activity+in+patients+with+idiopathic+calcium+nephrolithiasis.&aulast=Arrabal-Polo&pid=%3Cauthor%3EArrabal-Polo+MA%3BArrabal-Martin+M%3Bde+Haro-Munoz+T%3BPoyatos-Andujar+A%3BPalaeo-Yago+F%3BZuluaga-Gomez+A%3C%2Fauthor%3E%3CAN%3E21908029%3C%2FAN%3E%3CDT%3EComparative+Study%3C%2FDT%3E>

74. **Prevalence and metabolic abnormalities of vitamin D-inadequate patients presenting with urolithiasis to a tertiary stone clinic.**

**Item Type:** Journal Article

**Authors:** Elkoushy, Mohamed A.;Sabbagh, Ramsey;Unikowsky, Bernard and Andonian, Sero

**Publication Date:** Ap ,2012

**Journal:** Urology 79(4), pp. 781–785

**Abstract: OBJECTIVE**: To assess the prevalence and metabolic abnormalities of patients with inadequate vitamin D (VD) presenting with urolithiasis to a tertiary stone clinic in North America. **MATERIAL AND METHODS**: A retrospective review of consecutive patients presenting from August 2009 to January 2010 was performed. Demographic and clinical data were collected together with metabolic stone workup, including 2 24-hour urine collections and serum 25-hydroxy VD [25(OH)D]. VD inadequacy (VDI) included VD deficiency and VD insufficiency defined as 25-(OH) VD levels : A retrospective review of consecutive patients presenting from August 2009 to January 2010 was performed. Demographic and clinical data were collected together with metabolic stone workup, including 2 24-hour urine collections and serum 25-hydroxy VD [25(OH)D]. VD inadequacy (VDI) included VD deficiency and VD insufficiency defined as 25-(OH) VD levels **RESULTS**: Of 101 patients, 81 (80.2%) were found to have VDI: 34 (33.7%) were deficient and 47 (46.5%) had insufficient VD. Mean age was 50.4 +/- 15.8 years and the mean body mass index was 28.7 +/- 5.8 kg/m(2). Forty-two percent were smokers, 51% were recurrent stone formers, and 54% had positive family history of urolithiasis. Hyperparathyroidism was detected in 25.9% of patients, of which 91% of them were secondary to VDI. Hypocalcemia and hyperuricemia were found in 36% and 11% of patients, respectively. Of 81 VDI patients, 69 (85.2%) had complete 24-hour urine collections, of which 92.7% had at least one abnormality. However, only 40% of patients with normal VD had metabolic abnormalities (P : Of 101 patients, 81 (80.2%) were found to have VDI: 34 (33.7%) were deficient and 47 (46.5%) had insufficient VD. Mean age was 50.4 +/- 15.8 years and the mean body mass index was 28.7 +/- 5.8 kg/m(2). Forty-two percent were smokers, 51% were recurrent stone formers, and 54% had positive family history of urolithiasis. Hyperparathyroidism was detected in 25.9% of patients, of which 91% of them were secondary to VDI. Hypocalcemia and hyperuricemia were found in 36% and 11% of patients, respectively. Of 81 VDI patients, 69 (85.2%) had complete 24-hour urine collections, of which 92.7% had at least one abnormality. However, only 40% of patients with normal VD had metabolic abnormalities (P **CONCLUSION**: In this preliminary study, patients presenting with urolithiasis were found to have a high prevalence of inadequate VD associated with abnormalities on metabolic stone work-up. Copyright © 2012 Elsevier Inc. All rights reserved.

**URL:** <https://ovidsp.ovid.com/ovidweb.cgi?T=JS&CSC=Y&NEWS=N&PAGE=fulltext&D=med9&DO=10.1016%2fj.urology.2011.09.004> <https://libkey.io/libraries/2592/openurl?genre=article&sid=OVID:medline&id=pmid:22035763&id=doi:10.1016%2Fj.urology.2011.09.004&issn=0090-4295&isbn=&volume=79&issue=4&spage=781&pages=781-5&date=2012&title=Urology&atitle=Prevalence+and+metabolic+abnormalities+of+vitamin+D-inadequate+patients+presenting+with+urolithiasis+to+a+tertiary+stone+clinic.&aulast=Elkoushy&pid=%3Cauthor%3EElkoushy+MA%3BSabbagh+R%3BUnikowsky+B%3BAndonian+S%3C%2Fauthor%3E%3CAN%3E22035763%3C%2FAN%3E%3CDT%3EJournal+Article%3C%2FDT%3E>

75. **Metabolic assessment of elderly men with urolithiasis.**

**Item Type:** Journal Article

**Authors:** Freitas Junior, Celso Heitor;Mazzucchi, Eduardo;Danilovic, Alexandre;Brito, Artur Henrique and Srougi, Miguel

**Publication Date:** 2012

**Journal:** Clinics (Sao Paulo, Brazil) 67(5), pp. 457–461

**Abstract: OBJECTIVE**: To assess the presence of metabolic disorders in elderly men with urolithiasis. **METHODS**: We performed a case-control study. The inclusion criteria were as follows: (1) men older than 60 years of age and either (2) antecedent renal colic or an incidental diagnosis of urinary lithiasis after age 60 (case arm) or (3) no antecedent renal colic or incidental diagnosis of urolithiasis (control arm). Each individual underwent an interview, and those who were selected underwent all clinical protocol examinations: serum levels of total and ionized calcium, uric acid, phosphorus, glucose, urea, creatinine and parathyroid hormone, urine culture, and analysis of 24-hour urine samples (levels of calcium, citrate, creatinine, uric acid and sodium, pH and urine volume). Each case arm patient underwent two complete metabolic urinary investigations, whereas each control arm individual underwent one examination. ClinicalTrials.gov: NCT01246531. **RESULTS**: A total of 51 subjects completed the clinical investigation: 25 in the case arm and 26 in the control arm. In total, 56% of the case arm patients had hypocitraturia (vs. 15.4% in the control arm; p = 0.002). Hypernatriuria was detected in 64% of the case arm patients and in 30.8% of the controls (p = 0.017). **CONCLUSION**: Hypocitraturia and hypernatriuria are the main metabolic disorders in elderly men with urolithiasis.

**URL:** <https://ovidsp.ovid.com/ovidweb.cgi?T=JS&CSC=Y&NEWS=N&PAGE=fulltext&D=med9&DO=10.6061%2fclinics%2f2012%2805%2909> <https://libkey.io/libraries/2592/openurl?genre=article&sid=OVID:medline&id=pmid:22666789&id=doi:10.6061%2Fclinics%2F2012%2805%2909&issn=1807-5932&isbn=&volume=67&issue=5&spage=457&pages=457-61&date=2012&title=Clinics+%28Sao+Paulo%2C+Brazil%29&atitle=Metabolic+assessment+of+elderly+men+with+urolithiasis.&aulast=Freitas+Junior&pid=%3Cauthor%3EFreitas+Junior+CH%3BMazzucchi+E%3BDanilovic+A%3BBrito+AH%3BSrougi+M%3C%2Fauthor%3E%3CAN%3E22666789%3C%2FAN%3E%3CDT%3EJournal+Article%3C%2FDT%3E>

76. **Comparison of 24-hour urinary citrate excretion in stone formers and healthy volunteers.**

**Item Type:** Journal Article

**Authors:** Goodarzi, Mohammad Taghi;Forouzanfar, Farzad;Moaddab, Amir Hossein;Karimian, Mohammad and Sabzevar, Niloofar Kazemi

**Publication Date:** No ,2012

**Journal:** Saudi Journal of Kidney Diseases & Transplantation 23(6), pp. 1227–1231

**Abstract:** Low urinary citrate excretion is a risk factor in stone formers (SF). This study aimed to measure the urinary citrate excretion in SF and healthy volunteers at our center from 12 June 2008 to 20 August 2009. There were 28 SF patients (18 males and ten females) and 27 (18 males and nine females) age-matched healthy adult volunteers who participated in this study. Both groups had a similar living environment, extrinsic factors, diet and genetic descent. After collecting 24-h urine, citrate was measured using an enzymatic kit. Routine urinalysis and 24-h creatinine and uric acid were also performed. There was a significant difference in urinary citrate excretion level among SF (mean 310, SD 260 mg/L) and normal volunteer subjects (mean 800, SD 300 mg/L). By applying the previously defined normal values (320 mg/24 h) of urinary citrate in the local population, 43% of the SF in our study group was hypocitric, and none among the controls. We conclude that prevalence of hypocitraturia in stone formers was higher than that in healthy volunteers in our population.

**URL:** <https://ovidsp.ovid.com/ovidweb.cgi?T=JS&CSC=Y&NEWS=N&PAGE=fulltext&D=med9&DO=10.4103%2f1319-2442.103564> <https://libkey.io/libraries/2592/openurl?genre=article&sid=OVID:medline&id=pmid:23168853&id=doi:10.4103%2F1319-2442.103564&issn=1319-2442&isbn=&volume=23&issue=6&spage=1227&pages=1227-31&date=2012&title=Saudi+Journal+of+Kidney+Diseases+%26+Transplantation&atitle=Comparison+of+24-hour+urinary+citrate+excretion+in+stone+formers+and+healthy+volunteers.&aulast=Goodarzi&pid=%3Cauthor%3EGoodarzi+MT%3BForouzanfar+F%3BMoaddab+AH%3BKarimian+M%3BSabzevar+NK%3C%2Fauthor%3E%3CAN%3E23168853%3C%2FAN%3E%3CDT%3EJournal+Article%3C%2FDT%3E>

77. **Urinary stone risk and cola consumption.**

**Item Type:** Journal Article

**Authors:** Herrel, Lindsey;Pattaras, John;Solomon, Tania and Ogan, Kenneth

**Publication Date:** No ,2012

**Journal:** Urology 80(5), pp. 990–994

**Abstract: OBJECTIVE**: To evaluate the effect of cola consumption on urinary stone risk factors in a controlled metabolic environment with a prospective cross-over study. **METHODS**: Thirteen participants (10 normal and 3 calcium oxalate stone-formers) underwent 2 6-day phases on a controlled metabolic diet. During phase 1, subjects ingested 1 liter of cola daily, followed by a 3-week washout period. During phase 2, subjects ingested 1 liter of deionized water daily. Twenty-four-hour urine collections and serum metabolic panels were obtained for stone risk factors at the end of each phase. Urine and serum results of the cola phase were compared with those of the water phase. **RESULT**: Normal subjects and stone-formers were combined for analysis after determining no significant difference between the 2 groups. No significant differences were found among cola and water treatment groups for normal subjects, stone-formers, or the pooled group. **CONCLUSION**: Cola exerts no detectable change in urinary risk factors associated with calcium oxalate stone formation. Cola consumption may not increase stone risk and may be an acceptable alternative source of daily fluid for patients unwilling to increase water consumption. Copyright © 2012 Elsevier Inc. All rights reserved.

**URL:** <https://ovidsp.ovid.com/ovidweb.cgi?T=JS&CSC=Y&NEWS=N&PAGE=fulltext&D=med9&DO=10.1016%2fj.urology.2012.07.003> <https://libkey.io/libraries/2592/openurl?genre=article&sid=OVID:medline&id=pmid:23017784&id=doi:10.1016%2Fj.urology.2012.07.003&issn=0090-4295&isbn=&volume=80&issue=5&spage=990&pages=990-4&date=2012&title=Urology&atitle=Urinary+stone+risk+and+cola+consumption.&aulast=Herrel&pid=%3Cauthor%3EHerrel+L%3BPattaras+J%3BSolomon+T%3BOgan+K%3C%2Fauthor%3E%3CAN%3E23017784%3C%2FAN%3E%3CDT%3EComparative+Study%3C%2FDT%3E>

78. **Urinary metabolic evaluation of stone formers-a Malaysian perspective.**

**Item Type:** Journal Article

**Authors:** Hong, Yet Hoi;Dublin, Norman;Razack, Azad Hassan;Mohd, Mustafa Ali and Husain, Ruby

**Publication Date:** Se ,2012

**Journal:** Urology 80(3), pp. 529–534

**Abstract: OBJECTIVE**: To investigate the urinary metabolic excretion pattern among local stone formers given the great differences in the intrinsic and extrinsic risk factors as well as the urinary metabolic excretions compared with other populations. **METHODS**: Thirty urinary stone formers out of an initial 62 recruited provided a complete 24-hour urine sample for metabolic evaluation. Student's t-test and Pearson correlation test were used for the statistical analysis. **RESULTS**: Urinary volume (1719 +/- 712 vs 1215 +/- 575, P : Urinary volume (1719 +/- 712 vs 1215 +/- 575, P : Urinary volume (1719 +/- 712 vs 1215 +/- 575, P **CONCLUSION**: Elevated urinary oxalate level was the most important urinary risk factor among the local stone formers. A low urinary citrate excretion appeared to be a general phenomenon among the studied cohorts. Copyright © 2012 Elsevier Inc. All rights reserved.

**URL:** <https://ovidsp.ovid.com/ovidweb.cgi?T=JS&CSC=Y&NEWS=N&PAGE=fulltext&D=med9&DO=10.1016%2fj.urology.2012.02.053> <https://libkey.io/libraries/2592/openurl?genre=article&sid=OVID:medline&id=pmid:22578827&id=doi:10.1016%2Fj.urology.2012.02.053&issn=0090-4295&isbn=&volume=80&issue=3&spage=529&pages=529-34&date=2012&title=Urology&atitle=Urinary+metabolic+evaluation+of+stone+formers-a+Malaysian+perspective.&aulast=Hong&pid=%3Cauthor%3EHong+YH%3BDublin+N%3BRazack+AH%3BMohd+MA%3BHusain+R%3C%2Fauthor%3E%3CAN%3E22578827%3C%2FAN%3E%3CDT%3EJournal+Article%3C%2FDT%3E>

79. **Effects of drinking water quality on urinary parameters in men with and without urinary tract stones.**

**Item Type:** Journal Article

**Authors:** Mirzazadeh, Majid;Nouran, Mina Ghadimi;Richards, Kyle A. and Zare, Mariam

**Publication Date:** Ma ,2012

**Journal:** Urology 79(3), pp. 501–507

**Abstract: OBJECTIVE**: There is some controversy regarding the impact of water hardness on stone risk. Our study addresses this issue in a controlled setting. **MATERIAL AND METHODS**: Fifteen stone-former (SF) and 14 non-stone-former (NSF) males participated in this study in 3 intervals of 2 days each. Subjects collected a 24-hour urine sample while consuming a self-selected diet. They were then administered controlled diets. During the first 2 days, patients consumed water of minimal hardness (WMH), followed by tap water (TW) with moderate hardness on days 3 and 4, and mineral water (MW) on the final 2 days. Calcium (Ca), phosphorus, uric acid, oxalate, citrate, magnesium (Mg), sodium, potassium, and creatinine (Cr) content were measured in 24-hour urine samples on days 2, 4, and 6. **RESULTS**: Differences in water hardness and analytes were statistically significant among the different water types (P : Differences in water hardness and analytes were statistically significant among the different water types (P : Differences in water hardness and analytes were statistically significant among the different water types (P : Differences in water hardness and analytes were statistically significant among the different water types (P : Differences in water hardness and analytes were statistically significant among the different water types (P : Differences in water hardness and analytes were statistically significant among the different water types (P : Differences in water hardness and analytes were statistically significant among the different water types (P : Differences in water hardness and analytes were statistically significant among the different water types (P **CONCLUSIONS**: NSF had significantly higher urinary Mg/g Cr excretion rate before intervention than SF (P < .01). Increasing drinking water hardness while controlling for all other factors increased Ca/g Cr ratio in SF, rendering them at least theoretically more inclined to stone formation. Copyright A© 2012 Elsevier Inc. All rights reserved.

**URL:** <https://ovidsp.ovid.com/ovidweb.cgi?T=JS&CSC=Y&NEWS=N&PAGE=fulltext&D=med9&DO=10.1016%2fj.urology.2011.10.025> <https://libkey.io/libraries/2592/openurl?genre=article&sid=OVID:medline&id=pmid:22173182&id=doi:10.1016%2Fj.urology.2011.10.025&issn=0090-4295&isbn=&volume=79&issue=3&spage=501&pages=501-7&date=2012&title=Urology&atitle=Effects+of+drinking+water+quality+on+urinary+parameters+in+men+with+and+without+urinary+tract+stones.&aulast=Mirzazadeh&pid=%3Cauthor%3EMirzazadeh+M%3BNouran+MG%3BRichards+KA%3BZare+M%3C%2Fauthor%3E%3CAN%3E22173182%3C%2FAN%3E%3CDT%3EComparative+Study%3C%2FDT%3E>

80. **Omega-3 fatty acids eicosapentaenoic acid and docosahexaenoic acid in the management of hypercalciuric stone formers.**

**Item Type:** Journal Article

**Authors:** Ortiz-Alvarado, Omar;Miyaoka, Ricardo;Kriedberg, Carly;Leavitt, David A.;Moeding, Angela;Stessman, Michelle and Monga, Manoj

**Publication Date:** Fe ,2012

**Journal:** Urology 79(2), pp. 282–286

**Abstract: OBJECTIVE**: To investigate the use of fish oil in the dietary management of hypercalciuric stone formers. Prostaglandins have been linked to urinary calcium excretion, suggesting a role for omega-3 fatty acids in the treatment of hypercalciuric urolithiasis. **METHODS**: We retrospectively studied a cohort of patients treated at our stone clinics from July 2007 to February 2009. Patients' urinary risk factors for stone disease were evaluated with pre- and post-intervention 24-hour urine collections. All patients received empiric dietary recommendations for intake of fluids, sodium, protein, and citric juices. All subjects with hypercalciuria (urinary calcium>250 mg/d for males or >200 mg/d for females) on at least two 24-hour urine collections were counseled to supplement their diet with fish oil (1200 mg/d). **RESULTS**: Twenty-nine patients were followed for 9.86+/-8.96 months. The mean age was 43.38+/-13.78 years. Urinary calcium levels decreased in 52% of patients, with 24% converting to normocalciuria. The average urinary calcium (mg/d) decreased significantly from baseline (329.27+/-96.23 to 247.47+/-84.53, P: Twenty-nine patients were followed for 9.86+/-8.96 months. The mean age was 43.38+/-13.78 years. Urinary calcium levels decreased in 52% of patients, with 24% converting to normocalciuria. The average urinary calcium (mg/d) decreased significantly from baseline (329.27+/-96.23 to 247.47+/-84.53, P**CONCLUSION**: Omega-3 fatty acids combined with empiric dietary counseling results in a measurable decrease in urinary calcium and oxalate excretion and an increase in urinary citrate in hypercalciuric stone formers. Copyright © 2012 Elsevier Inc. All rights reserved.

**URL:** <https://ovidsp.ovid.com/ovidweb.cgi?T=JS&CSC=Y&NEWS=N&PAGE=fulltext&D=med9&DO=10.1016%2fj.urology.2011.08.022> <https://libkey.io/libraries/2592/openurl?genre=article&sid=OVID:medline&id=pmid:22000931&id=doi:10.1016%2Fj.urology.2011.08.022&issn=0090-4295&isbn=&volume=79&issue=2&spage=282&pages=282-6&date=2012&title=Urology&atitle=Omega-3+fatty+acids+eicosapentaenoic+acid+and+docosahexaenoic+acid+in+the+management+of+hypercalciuric+stone+formers.&aulast=Ortiz-Alvarado&pid=%3Cauthor%3EOrtiz-Alvarado+O%3BMiyaoka+R%3BKriedberg+C%3BLeavitt+DA%3BMoeding+A%3BStessman+M%3BMonga+M%3C%2Fauthor%3E%3CAN%3E22000931%3C%2FAN%3E%3CDT%3EJournal+Article%3C%2FDT%3E>

81. **Prevalence of nephrolithiasis in human immunodeficiency virus infected patients on the highly active antiretroviral therapy.**

**Item Type:** Journal Article

**Authors:** Raheem, Omer A.;Mirheydar, Hossein S.;Palazzi, Kerrin;Chenoweth, Marianne;Lakin, Charles and Sur, Roger L.

**Publication Date:** Au ,2012

**Journal:** Journal of Endourology 26(8), pp. 1095–1098

**Abstract: BACKGROUND AND PURPOSE**: Protease inhibitors, specifically indinavir, have historically been implicated as a cause of nephrolithiasis in the human immunodeficiency virus (HIV) infected patients. There is a paucity of data, however, on stone disease with nonindinavir etiologies since the introduction of highly active antiretroviral therapy (HAART). We sought to describe the prevalence of nephrolithiasis in the HIV population since the use of HAART. **PATIENTS AND METHODS**: We retrospectively reviewed HIV-positive patients currently receiving HAART treatment in whom image proven kidney and/or ureteral urolithiasis developed, between 1998 and 2010. A detailed analysis of patients' current treatment, surgical intervention, and metabolic studies was performed. **RESULTS**: A total of 436 HIV-positive patients were included and 46 (11%) patients had nephrolithiasis. Each patient included in this study was receiving nonindinavir-based antiretroviral therapy. There were 41 men of whom 36 were Caucasian. Eleven (24%) patients underwent 24-hour urine collections with 11 metabolic abnormalities identified. Stone analysis was available for seven patients (four calcium oxalate monohydrate, one cystine, one uric acid, and one atazanavir). **CONCLUSIONS**: We report the largest series of nephrolithiasis in an HIV population since the introduction of HAART and highlight not only the similar prevalence of nephrolithiasis to the non-HIV population but also the lack of consistent comprehensive metabolic evaluations in HIV patients with recurrent nephrolithiasis.

**URL:** <https://ovidsp.ovid.com/ovidweb.cgi?T=JS&CSC=Y&NEWS=N&PAGE=fulltext&D=med9&DO=10.1089%2fend.2011.0639> <https://libkey.io/libraries/2592/openurl?genre=article&sid=OVID:medline&id=pmid:22429050&id=doi:10.1089%2Fend.2011.0639&issn=0892-7790&isbn=&volume=26&issue=8&spage=1095&pages=1095-8&date=2012&title=Journal+of+Endourology&atitle=Prevalence+of+nephrolithiasis+in+human+immunodeficiency+virus+infected+patients+on+the+highly+active+antiretroviral+therapy.&aulast=Raheem&pid=%3Cauthor%3ERaheem+OA%3BMirheydar+HS%3BPalazzi+K%3BChenoweth+M%3BLakin+C%3BSur+RL%3C%2Fauthor%3E%3CAN%3E22429050%3C%2FAN%3E%3CDT%3EJournal+Article%3C%2FDT%3E>

82. **Twenty-four-hour urinary trace element excretion: reference intervals and interpretive issues.**

**Item Type:** Journal Article

**Authors:** Sieniawska, Christine E.;Jung, Lynne C.;Olufadi, Rasaq and Walker, Valerie

**Publication Date:** Ju ,2012

**Journal:** Annals of Clinical Biochemistry 49(Pt 4), pp. 341–351

**Abstract: BACKGROUND**: Introduction of inductively coupled plasma mass spectrometry (ICP-MS) into clinical laboratories has led to an increasing application of analyses to risk assessment for toxicity from environmental exposure to trace elements, and in occupational monitoring. Interpretation of results from random urine samples may be problematic and measurement of excretion over 24 h is sometimes preferable. Recent reference data are sparse. **METHODS**: Twenty-four-hour urine samples from 111 healthy adults from the renal stones clinic in Southampton, UK, were analysed for 31 trace elements using ICP-MS and for zinc using atomic absorption spectroscopy. Non-parametric 0.95 coverage intervals were determined for trace element excretion per 24 h and as a ratio to creatinine, for the full study cohort and separately for men (n=77) and women (n=34). **RESULTS**: Beryllium was undetectable in 95% of samples, bismuth in 87% and uranium in 75%. In comparison with published ranges, reference intervals for this cohort were higher for molybdenum, tin and vanadium, and for arsenic due to inclusion of fish arsenicals. Aluminium, chromium, iron, lead and mercury were lower. In our cohort, 24-h excretion of 17 elements was significantly higher in men than in women. However, when expressed as trace element to creatinine ratios, the situation reversed strikingly. Because of their lower creatinine excretion, ratios for 18 elements were significantly higher for women. **CONCLUSIONS**: New adult reference intervals were obtained for 24-h urine trace element excretion. Trace element:creatinine ratios must be used cautiously, with separate ranges for men and women.

**URL:** <https://ovidsp.ovid.com/ovidweb.cgi?T=JS&CSC=Y&NEWS=N&PAGE=fulltext&D=med9&DO=10.1258%2facb.2011.011179> <https://libkey.io/libraries/2592/openurl?genre=article&sid=OVID:medline&id=pmid:22362086&id=doi:10.1258%2Facb.2011.011179&issn=0004-5632&isbn=&volume=49&issue=4&spage=341&pages=341-51&date=2012&title=Annals+of+Clinical+Biochemistry&atitle=Twenty-four-hour+urinary+trace+element+excretion%3A+reference+intervals+and+interpretive+issues.&aulast=Sieniawska&pid=%3Cauthor%3ESieniawska+CE%3BJung+LC%3BOlufadi+R%3BWalker+V%3C%2Fauthor%3E%3CAN%3E22362086%3C%2FAN%3E%3CDT%3EJournal+Article%3C%2FDT%3E>

83. **Differences in metabolic urinary abnormalities in stone forming and nonstone forming patients with primary hyperparathyroidism.**

**Item Type:** Journal Article

**Authors:** Sorensen, Mathew D.;Duh, Quan-Yang;Grogan, Raymon H.;Tran, Thanh C. and Stoller, Marshall L.

**Publication Date:** Ma ,2012

**Journal:** Surgery 151(3), pp. 477–483

**Abstract: BACKGROUND**: Hyperparathyroidism is associated with hypercalciuria and nephrolithiasis. Urine calcium excretion decreases after parathyroidectomy, but whether there is a differential decrease between stone and nonstone formers remains controversial. We evaluated differences between stone formers and non-stone formers in serum and urinary parameters before and after parathyroidectomy. **METHODS**: 90 patients, 40 with and 50 without a history of nephrolithiasis underwent parathyroidectomy for hyperparathyroidism; 24-hour urine samples were collected before and after parathyroidectomy. Overall, 92% of patients provided samples before parathyroidectomy and 63% after parathyroidectomy. Preoperative, postoperative and changes in urinary parameters were evaluated. **RESULTS**: Preoperative hypercalciuria was present in ~65% in both groups (P = .68). Parathyroidectomy decreased serum levels of calcium, parathyroid hormone, and urinary calcium, but there were no differences between stone formers and nonstone formers. Stone formers were 12-fold (P = .001) more likely to resolve an increase in supersaturation of calcium oxalate (SSCaOx), and after adjustment for age, sex, and BMI were 46-fold (P = .002) more likely to resolve an increase in SSCaOx. After parathyroidectomy, the rate of stone recurrence was 23% and male sex (aOR 20, P = .032) and increasing BMI (aOR 1.23, P = .038) were the only independent predictors of stone recurrence after adjusting for age. No other factor evaluated preoperatively, postoperatively, or the change after parathyroidectomy differentiated stone and nonstone formers or predicted stone recurrence. **CONCLUSION**: Metabolic evaluation did not differentiate stone formers from nonstone formers reliably. Stone formers were more likely to resolve an increase in SSCaOx after parathyroidectomy. Male sex and increasing BMI were independently associated with stone recurrence after parathyroidectomy. Copyright A© 2012 Mosby, Inc. All rights reserved.

**URL:** <https://ovidsp.ovid.com/ovidweb.cgi?T=JS&CSC=Y&NEWS=N&PAGE=fulltext&D=med9&DO=10.1016%2fj.surg.2011.07.039> <https://libkey.io/libraries/2592/openurl?genre=article&sid=OVID:medline&id=pmid:21893327&id=doi:10.1016%2Fj.surg.2011.07.039&issn=0039-6060&isbn=&volume=151&issue=3&spage=477&pages=477-83&date=2012&title=Surgery&atitle=Differences+in+metabolic+urinary+abnormalities+in+stone+forming+and+nonstone+forming+patients+with+primary+hyperparathyroidism.&aulast=Sorensen&pid=%3Cauthor%3ESorensen+MD%3BDuh+QY%3BGrogan+RH%3BTran+TC%3BStoller+ML%3C%2Fauthor%3E%3CAN%3E21893327%3C%2FAN%3E%3CDT%3EJournal+Article%3C%2FDT%3E>

84. **Persistence of hypercalciuria after successful surgical treatment for primary hyperparathyroidism.**

**Item Type:** Journal Article

**Authors:** Spivacow, Francisco Rodolfo;Negri, Armando Luis;del Valle, Elisa Elena;Fradinger, Erich;Martinez, Carolina and Polonsky, Ana

**Publication Date:** Ju ,2012

**Journal:** International Urology & Nephrology 44(3), pp. 857–863

**Abstract: UNLABELLED**: Primary hyperparathyroidism (PHPT) causes hypercalciuria and stone disease in a subset of patients. Hypercalciuria typically normalizes after surgery, although the risk of stone formation may persist up to 10 years. There are few reports in the literature that show persistent hypercalciuria despite normalization of serum calcium after parathyroid surgery. We retrospectively analyzed 111 patients with PHPT from the osteoporosis, and stone clinics seen between 1999 and 2006. We selected only patients who had a complete metabolic profile that included 24-hour collections before and at least 3 months after parathyroidectomy. We excluded patients who had creatinine clearance : Primary hyperparathyroidism (PHPT) causes hypercalciuria and stone disease in a subset of patients. Hypercalciuria typically normalizes after surgery, although the risk of stone formation may persist up to 10 years. There are few reports in the literature that show persistent hypercalciuria despite normalization of serum calcium after parathyroid surgery. We retrospectively analyzed 111 patients with PHPT from the osteoporosis, and stone clinics seen between 1999 and 2006. We selected only patients who had a complete metabolic profile that included 24-hour collections before and at least 3 months after parathyroidectomy. We excluded patients who had creatinine clearance **CONCLUSIONS**: Persistently increased fractional excretion of calcium could explain the sustained increased risk of stone disease in patients with PHPT for many years after successful parathyroidectomy.

**URL:** <https://ovidsp.ovid.com/ovidweb.cgi?T=JS&CSC=Y&NEWS=N&PAGE=fulltext&D=med9&DO=10.1007%2fs11255-011-9953-6> <https://libkey.io/libraries/2592/openurl?genre=article&sid=OVID:medline&id=pmid:21505756&id=doi:10.1007%2Fs11255-011-9953-6&issn=0301-1623&isbn=&volume=44&issue=3&spage=857&pages=857-63&date=2012&title=International+Urology+%26+Nephrology&atitle=Persistence+of+hypercalciuria+after+successful+surgical+treatment+for+primary+hyperparathyroidism.&aulast=Spivacow&pid=%3Cauthor%3ESpivacow+FR%3BNegri+AL%3Bdel+Valle+EE%3BFradinger+E%3BMartinez+C%3BPolonsky+A%3C%2Fauthor%3E%3CAN%3E21505756%3C%2FAN%3E%3CDT%3EJournal+Article%3C%2FDT%3E>

85. **The role of salt abuse on risk for hypercalciuria.**

**Item Type:** Journal Article

**Authors:** Damasio, Patricia C. G.;Amaro, Carmen R. P. R.;Cunha, Natalia B.;Pichutte, Ana C.;Goldberg, Jose;Padovani, Carlos R. and Amaro, Joao L.

**Publication Date:** Jan 06 ,2011

**Journal:** Nutrition Journal 10, pp. 3

**Abstract: BACKGROUND**: Elevated sodium excretion in urine resulting from excessive sodium intake can lead to hypercalciuria and contribute to the formation of urinary stones. The aim of this study was to evaluate salt intake in patients with urinary lithiasis and idiopathic hypercalciuria (IH). **METHODS**: Between August 2007 and June 2008, 105 lithiasic patients were distributed into 2 groups: Group 1 (n = 55): patients with IH (urinary calcium excretion > 250 mg in women and 300 mg in men with normal serum calcium); Group 2 (n = 50): normocalciuric patients (NC). Inclusion criteria were: age over 18 years, normal renal function (creatinine clearance >= 60 ml/min), absent proteinuria and negative urinary culture. Pregnant women, patients with intestinal pathologies, chronic diarrhea or using corticoids were excluded. The protocol of metabolic investigation was based on non-consecutive collection of two 24-hour samples for dosages of: calcium, sodium, uric acid, citrate, oxalate, magnesium and urinary volume. Food intake was evaluated by the three-day dietary record quantitative method, and the Body Mass Index (BMI) was calculated and classified according to the World Health Organization (WHO). Sodium intake was evaluated based on 24-hour urinary sodium excretion. **RESULTS**: The distribution in both groups as regards mean age (42.11 +/- 10.61 vs. 46.14 +/- 11.52), weight (77.14 +/- 16.03 vs. 75.99 +/- 15.80), height (1.64 +/- 0.10 vs. 1.64 +/- plusorminus 0.08) and BMI (28.78 +/- 5.81 vs. 28.07 +/- 5.27) was homogeneous. Urinary excretion of calcium (433.33 +/- 141.92 vs. 188.93 +/- 53.09), sodium (280.08 +/- 100.94 vs. 200.44.93 +/- 65.81), uric acid (880.63 +/- 281.50 vs. 646.74 +/- 182.76) and magnesium (88.78 +/- 37.53 vs. 64.34 +/- 31.84) was significantly higher in the IH group (p : The distribution in both groups as regards mean age (42.11 +/- 10.61 vs. 46.14 +/- 11.52), weight (77.14 +/- 16.03 vs. 75.99 +/- 15.80), height (1.64 +/- 0.10 vs. 1.64 +/- plusorminus 0.08) and BMI (28.78 +/- 5.81 vs. 28.07 +/- 5.27) was homogeneous. Urinary excretion of calcium (433.33 +/- 141.92 vs. 188.93 +/- 53.09), sodium (280.08 +/- 100.94 vs. 200.44.93 +/- 65.81), uric acid (880.63 +/- 281.50 vs. 646.74 +/- 182.76) and magnesium (88.78 +/- 37.53 vs. 64.34 +/- 31.84) was significantly higher in the IH group (p **CONCLUSIONS**: This study showed that salt intake was higher in patients with IH as compared to NC.

**URL:** <https://ovidsp.ovid.com/ovidweb.cgi?T=JS&CSC=Y&NEWS=N&PAGE=fulltext&D=med8&DO=10.1186%2f1475-2891-10-3> <https://libkey.io/libraries/2592/openurl?genre=article&sid=OVID:medline&id=pmid:21211048&id=doi:10.1186%2F1475-2891-10-3&issn=1475-2891&isbn=&volume=10&issue=1&spage=3&pages=3&date=2011&title=Nutrition+Journal&atitle=The+role+of+salt+abuse+on+risk+for+hypercalciuria.&aulast=Damasio&pid=%3Cauthor%3EDamasio+PC%3BAmaro+CR%3BCunha+NB%3BPichutte+AC%3BGoldberg+J%3BPadovani+CR%3BAmaro+JL%3C%2Fauthor%3E%3CAN%3E21211048%3C%2FAN%3E%3CDT%3EJournal+Article%3C%2FDT%3E>

86. **Oxalobacter colonization in the morbidly obese and correlation with urinary stone risk.**

**Item Type:** Journal Article

**Authors:** Duffey, Branden G.;Miyaoka, Ricardo;Holmes, Ross;Assimos, Dean;Hinck, Bryan;Korman, Emily;Kieley, Fred;Ikramuddin, Sayeed;Kellogg, Todd;Moeding, Angela and Monga, Manoj

**Publication Date:** Se ,2011

**Journal:** Urology 78(3), pp. 531–534

**Abstract: OBJECTIVE**: To establish the baseline preoperative prevalence of Oxalobacter formigenes (OF) colonization in a cohort of obese patients scheduled for Roux-en-Y gastric bypass (RYGB) and determine the effect of OF colonization on urinary oxalate excretion. It has been proposed that loss of OF colonization after RYGB may contribute to the development of hyperoxaluria. **METHODS**: Adult patients scheduled to undergo RYGB were requested to provide a stool specimen and 24-hour urine collection before surgery. OF colonization status was determined by the calcium precipitation test. The 24-hour urine specimens were analyzed by the Litholink Corporation (Chicago, IL). **RESULTS**: Of the 51 patients submitting initial stool specimens, only 8 (16%) tested positive for OF, whereas 43 (84%) were negative. Patients colonized with OF were older than uncolonized subjects (52.9+/-6.8 vs 46.0+/-10.4 years, P=.03). Urinary oxalate was not significantly different between these groups (P=.14). **CONCLUSION**: OF colonization is uncommon in morbidly obese patients (16%) before surgery. Because hyperoxaluria develops in more than 50% of patients after RYGB, it is unlikely that loss of OF colonization is the primary cause. Copyright © 2011 Elsevier Inc. All rights reserved.

**URL:** <https://ovidsp.ovid.com/ovidweb.cgi?T=JS&CSC=Y&NEWS=N&PAGE=fulltext&D=med8&DO=10.1016%2fj.urology.2011.01.011> <https://libkey.io/libraries/2592/openurl?genre=article&sid=OVID:medline&id=pmid:21529902&id=doi:10.1016%2Fj.urology.2011.01.011&issn=0090-4295&isbn=&volume=78&issue=3&spage=531&pages=531-4&date=2011&title=Urology&atitle=Oxalobacter+colonization+in+the+morbidly+obese+and+correlation+with+urinary+stone+risk.&aulast=Duffey&pid=%3Cauthor%3EDuffey+BG%3BMiyaoka+R%3BHolmes+R%3BAssimos+D%3BHinck+B%3BKorman+E%3BKieley+F%3BIkramuddin+S%3BKellogg+T%3BMoeding+A%3BMonga+M%3C%2Fauthor%3E%3CAN%3E21529902%3C%2FAN%3E%3CDT%3EJournal+Article%3C%2FDT%3E>

87. **Effect of renal function on urinary mineral excretion and stone composition.**

**Item Type:** Journal Article

**Authors:** Kadlec, Adam O.;Greco, Kristin A.;Fridirici, Zachary C.;Gerber, Daniel and Turk, Thomas M. T.

**Publication Date:** Oc ,2011

**Journal:** Urology 78(4), pp. 744–747

**Abstract: OBJECTIVE**: To determine the effect of renal function on urinary mineral stone excretion and composition of kidney stones in patients undergoing urologic intervention for nephrolithiasis. **METHODS**: We performed a retrospective analysis of 158 patients with 193 kidney stones who underwent endourologic intervention for nephrolithiasis. The patients were grouped by the estimated glomerular filtration rate (eGFR). The kidney stone composition and results of the 24-hour urinalysis were reported for each group. Statistical analysis was performed using the chi-square test, Kruskal-Wallis test, and analysis of variance. **RESULTS**: The kidney stone composition profile differed significantly between the groups, with uric acid stones associated with a lower eGFR and calcium phosphate stones associated with a greater eGFR. A lower eGFR correlated with lower urine pH, lower calcium excretion, and greater oxalate excretion. Excretion of uric acid and the body mass index did not differ between groups. **CONCLUSION**: Different degrees of renal function correlated with certain types of kidney stones. The urine pH was lower in patients with impaired renal function. Renal function might be an underappreciated factor that affects kidney stone composition and urinary mineral excretion. Copyright © 2011 Elsevier Inc. All rights reserved.

**URL:** <https://ovidsp.ovid.com/ovidweb.cgi?T=JS&CSC=Y&NEWS=N&PAGE=fulltext&D=med8&DO=10.1016%2fj.urology.2011.04.007> <https://libkey.io/libraries/2592/openurl?genre=article&sid=OVID:medline&id=pmid:21664652&id=doi:10.1016%2Fj.urology.2011.04.007&issn=0090-4295&isbn=&volume=78&issue=4&spage=744&pages=744-7&date=2011&title=Urology&atitle=Effect+of+renal+function+on+urinary+mineral+excretion+and+stone+composition.&aulast=Kadlec&pid=%3Cauthor%3EKadlec+AO%3BGreco+KA%3BFridirici+ZC%3BGerber+D%3BTurk+TM%3C%2Fauthor%3E%3CAN%3E21664652%3C%2FAN%3E%3CDT%3EJournal+Article%3C%2FDT%3E>

88. **Oxalate and sucralose absorption in idiopathic calcium oxalate stone formers.**

**Item Type:** Journal Article

**Authors:** Knight, John;Jiang, Juquan;Wood, Kyle D.;Holmes, Ross P. and Assimos, Dean G.

**Publication Date:** Au ,2011

**Journal:** Urology 78(2), pp. 475.e9–475.e13

**Abstract: OBJECTIVES**: To better understand intestinal oxalate transport by correlating oxalate and sucralose absorption in idiopathic calcium oxalate stone formers. Oxalate has been hypothesized to undergo absorption in the large and small intestine by both paracellular and transepithelial transport. Sucralose is a chlorinated sugar that is absorbed by paracellular mechanisms. **METHODS**: Idiopathic calcium oxalate stone formers were recruited to provide urine specimens on both a self-selected diet and after a meal containing 90 mg of (13)C(2-)oxalate and 5 g of sucralose, and a stool sample for determination of Oxalobacter formigenes colonization. The 24-hour urine collections were fractionated into the first 6 hours and the subsequent 18 hours. Sucralose and oxalate excretion were measured during these periods and used to estimate absorption. **RESULTS**: Thirty-eight subjects were evaluated. The majority of both the (13)C(2-)oxalate and sucralose absorption occurred within the 0-6-hour collection. The (13)C(2-)oxalate and sucralose absorptions were significantly correlated at the 0-6 hour, the 6-24 hour, and the total 24-hour time periods (P 15% absorption) also absorbed significantly more sucralose during the 0-6 hour and whole 24-hour time points (P 15% absorption) also absorbed significantly more sucralose during the 0-6 hour and whole 24-hour time points (P **CONCLUSIONS**: The results suggest that most oxalate is absorbed in the proximal portion of the gastrointestinal tract and that paracellular transport is involved. Augmented paracellular transport, as evidenced by increased sucralose absorption, may also influence oxalate absorption. Copyright © 2011 Elsevier Inc. All rights reserved.

**URL:** <https://ovidsp.ovid.com/ovidweb.cgi?T=JS&CSC=Y&NEWS=N&PAGE=fulltext&D=med8&DO=10.1016%2fj.urology.2011.04.008> <https://libkey.io/libraries/2592/openurl?genre=article&sid=OVID:medline&id=pmid:21676449&id=doi:10.1016%2Fj.urology.2011.04.008&issn=0090-4295&isbn=&volume=78&issue=2&spage=475.e9&pages=475.e9-475.e13&date=2011&title=Urology&atitle=Oxalate+and+sucralose+absorption+in+idiopathic+calcium+oxalate+stone+formers.&aulast=Knight&pid=%3Cauthor%3EKnight+J%3BJiang+J%3BWood+KD%3BHolmes+RP%3BAssimos+DG%3C%2Fauthor%3E%3CAN%3E21676449%3C%2FAN%3E%3CDT%3EComparative+Study%3C%2FDT%3E>

89. **Determinants of osteopenia in male renal-stone-disease patients with idiopathic hypercalciuria.**

**Item Type:** Journal Article

**Authors:** Letavernier, Emmanuel;Traxer, Olivier;Daudon, Michel;Tligui, Mohammed;Hubert-Brierre, Jerome;Guerrot, Dominique;Sebag, Aline;Baud, Laurent and Haymann, Jean-Philippe

**Publication Date:** Ma ,2011

**Journal:** Clinical Journal of the American Society of Nephrology: CJASN 6(5), pp. 1149–1154

**Abstract: BACKGROUND AND OBJECTIVES**: Bone demineralization is frequent in renal-stone formers with hypercalciuria. Although this pathologic link has been recognized for decades, the underlying mechanisms and risk factors associated with osteopenia/osteoporosis in this population remain partially understood. **DESIGN, SETTING, PARTICIPANTS, & MEASUREMENTS**: This study retrospectively analyzed determinants of low bone mineral density (BMD) in 65 idiopathic hypercalciuric male renal-stone formers. Clinical and biologic evaluation included BMD measurement, bone-remodeling markers, analysis of calcium metabolism with oral calcium load test, and dietary inquiry. **RESULTS**: Patients with osteopenia (n=23, 35% of the population) presented significantly higher fasting calciuria as compared with normal bone density patients (n=42) (calcium/creatinine ratio was 0.32 versus 0.24 mmol/mmol; P=0.006). Analysis of the whole population revealed a negative association between fasting hypercalciuria and BMD (P = 0.003), independent of confounding variables including body-mass index and tobacco consumption. The fasting calcium/creatinine ratio above 0.25 mmol/mmol was associated with a 3.8-fold increase in the risk of low BMD. **CONCLUSION**: In our study, fasting hypercalciuria after a 2-day calcium-restricted diet appears as the only biologic factor associated with low BMD, suggesting a bone-calcium efflux. Our results support the view of a parathyroid-independent pathologic process that remains to be identified. Hypercalciuric patients with low BMD do not excrete more calcium in 24-hour urine samples than patients without low BMD. Copyright © 2011 by the American Society of Nephrology

**URL:** <https://ovidsp.ovid.com/ovidweb.cgi?T=JS&CSC=Y&NEWS=N&PAGE=fulltext&D=med8&DO=10.2215%2fCJN.10191110> <https://libkey.io/libraries/2592/openurl?genre=article&sid=OVID:medline&id=pmid:21441130&id=doi:10.2215%2FCJN.10191110&issn=1555-9041&isbn=&volume=6&issue=5&spage=1149&pages=1149-54&date=2011&title=Clinical+Journal+of+The+American+Society+of+Nephrology%3A+CJASN&atitle=Determinants+of+osteopenia+in+male+renal-stone-disease+patients+with+idiopathic+hypercalciuria.&aulast=Letavernier&pid=%3Cauthor%3ELetavernier+E%3BTraxer+O%3BDaudon+M%3BTligui+M%3BHubert-Brierre+J%3BGuerrot+D%3BSebag+A%3BBaud+L%3BHaymann+JP%3C%2Fauthor%3E%3CAN%3E21441130%3C%2FAN%3E%3CDT%3EJournal+Article%3C%2FDT%3E>

90. **Detection of absorptive hypercalciuria type I without the oral calcium load test.**

**Item Type:** Journal Article

**Authors:** Pak, Charles Y. C.;Sakhaee, Khashayar and Pearle, Margaret S.

**Publication Date:** Ma ,2011

**Journal:** Journal of Urology 185(3), pp. 915–919

**Abstract: PURPOSE**: We retrospectively analyzed the validity of a simple method of detecting absorptive hypercalciuria type I, a common stone forming condition with hypercalciuria that is believed to be due to high intestinal calcium absorption. The method is based on urinary calcium derived from 24-hour urine collections while on random and restricted diets rather than on a calciuric response to an oral calcium load. **MATERIALS AND METHODS**: A group of 916 well characterized patients with idiopathic calcium oxalate urolithiasis comprised the study group. We also analyzed a subgroup of 695 patients, excluding 221 with dietary abuse, defined as urinary sodium greater than 150 mEq daily and/or sulphate greater than 35 mmol daily, to eliminate potential confounding dietary factors affecting the diagnosis. In each group absorptive hypercalciuria type I was detected by the old criteria, requiring an exaggerated calciuric response to an oral calcium load test, and by the new criteria, based on 24-hour urinary calcium 200 mg or greater daily while on random and restricted diets. **RESULTS**: Using the old criteria as the gold standard the positive and negative predictive values, sensitivity and specificity of the new criteria were 80.1%, 95.9%, 90.8% and 90.5%, respectively. When excluding patients with dietary abuse the values were 85.9%, 97.2%, 92.4% and 94.5%, respectively. **CONCLUSIONS**: Absorptive hypercalciuria type I may be reliably detected by a simple method based on high 24-hour urinary calcium while on random and restricted diets, especially when excluding patients with evidence of dietary abuse during the restricted diet. Copyright © 2011 American Urological Association Education and Research, Inc. Published by Elsevier Inc. All rights reserved.

**URL:** <https://ovidsp.ovid.com/ovidweb.cgi?T=JS&CSC=Y&NEWS=N&PAGE=fulltext&D=med8&DO=10.1016%2fj.juro.2010.10.067> <https://libkey.io/libraries/2592/openurl?genre=article&sid=OVID:medline&id=pmid:21251672&id=doi:10.1016%2Fj.juro.2010.10.067&issn=0022-5347&isbn=&volume=185&issue=3&spage=915&pages=915-9&date=2011&title=Journal+of+Urology&atitle=Detection+of+absorptive+hypercalciuria+type+I+without+the+oral+calcium+load+test.&aulast=Pak&pid=%3Cauthor%3EPak+CY%3BSakhaee+K%3BPearle+MS%3C%2Fauthor%3E%3CAN%3E21251672%3C%2FAN%3E%3CDT%3EJournal+Article%3C%2FDT%3E>

91. **The most important metabolic risk factors in recurrent urinary stone formers.**

**Item Type:** Journal Article

**Authors:** Parvin, Mahmoud;Shakhssalim, Nasser;Basiri, Abbas;Miladipour, Amir Hossein;Golestan, Banafsheh;Mohammadi Torbati, Peyman;Azadvari, Mohaddeseh and Eftekhari, Sanaz

**Publication Date:** 2011

**Journal:** Urology Journal 8(2), pp. 99–106

**Abstract: PURPOSE**: To evaluate different urinary factors contributing to idiopathic calcium stone disease for determining appropriate medical treatments. **MATERIALS AND METHODS**: Two 24-hour urine samples were collected from 106 male recurrent idiopathic calcium stone formers and another 109 randomly selected men as the control group matching for age. **RESULTS**: Cases had significantly higher mean urine oxalate, calcium, uric acid, and chloride in comparison with the healthy controls (P : Cases had significantly higher mean urine oxalate, calcium, uric acid, and chloride in comparison with the healthy controls (P **CONCLUSION**: Oxalate seems to play the most important role as urinary stone risk factor in our population followed by calcium and uric acid. In addition to the risk factors, it seems that supersaturation as the sum of all risk factors probably has a high predictive value.

**URL:** <https://ovidsp.ovid.com/ovidweb.cgi?T=JS&CSC=Y&NEWS=N&PAGE=fulltext&D=med8&AN=21656467> <https://libkey.io/libraries/2592/openurl?genre=article&sid=OVID:medline&id=pmid:21656467&id=doi:&issn=1735-1308&isbn=&volume=8&issue=2&spage=99&pages=99-106&date=2011&title=Urology+Journal&atitle=The+most+important+metabolic+risk+factors+in+recurrent+urinary+stone+formers.&aulast=Parvin&pid=%3Cauthor%3EParvin+M%3BShakhssalim+N%3BBasiri+A%3BMiladipour+AH%3BGolestan+B%3BMohammadi+Torbati+P%3BAzadvari+M%3BEftekhari+S%3C%2Fauthor%3E%3CAN%3E21656467%3C%2FAN%3E%3CDT%3EJournal+Article%3C%2FDT%3E>

92. **Investigation of nephrolithiasis in the West of Parana.**

**Item Type:** Journal Article

**Authors:** Peres, Luis Alberto Batista;de Almeida, Leandro Pelegrini;Bolson, Luana Bertinato;Brites, Mariana de Freitas;David, Juliano Maximiano and Tazima, Leandro

**Publication Date:** 2011

**Journal:** Jornal Brasileiro De Nefrologia 33(2), pp. 160–165

**Abstract: INTRODUCTION**: Nephrolithiasis is com>mon and has a high rate of recurrence. **OBJECTIVES**: To assess the prevalence of the main metabolic and anatomical changes and the chemical analysis of stone found in patients with nephrolithiasis in the West region of Parana. **METHODS**: Retrospective study with 681 adult patients with nephrolithiasis. The laboratory investigation included at least two samples of 24-hour urine test with doses of calcium, uric acid, citrate, oxalate, sodium and creatinine; qualitative cystinuria, urinary pH following 12-hour fast and water restriction, urine culture and chemical analysis, when the stones were available. Renal imaging techniques included at least renal ultrasound and excretory urogram. **RESULTS**: The metabolic changes most frequently found were: hypercalciuria (51.8%), hyperuricosuria (27.6%), and hypocitraturia (23.5%). Chemical analysis of stones showed calcium oxa late in 85.7% of the cases. The most frequently anatomical changes were renal cyst, duplicated ureter, and ureteropelvic junction obstruction. **CONCLUSIONS**: This paper served as a base for knowing the characteristics of patients with nephrolithiasis in the West area of Parana.

**URL:** <https://ovidsp.ovid.com/ovidweb.cgi?T=JS&CSC=Y&NEWS=N&PAGE=fulltext&D=med8&AN=21789428> <https://libkey.io/libraries/2592/openurl?genre=article&sid=OVID:medline&id=pmid:21789428&id=doi:&issn=0101-2800&isbn=&volume=33&issue=2&spage=160&pages=160-5&date=2011&title=Jornal+Brasileiro+de+Nefrologia&atitle=Investigation+of+nephrolithiasis+in+the+West+of+Parana.&aulast=Peres&pid=%3Cauthor%3EPeres+LA%3Bde+Almeida+LP%3BBolson+LB%3BBrites+Mde+F%3BDavid+JM%3BTazima+L%3C%2Fauthor%3E%3CAN%3E21789428%3C%2FAN%3E%3CDT%3EJournal+Article%3C%2FDT%3E>

93. **Adequacy of a single 24-hour urine collection for metabolic evaluation of recurrent nephrolithiasis.**

**Item Type:** Journal Article

**Authors:** Castle, Scott M.;Cooperberg, Matthew R.;Sadetsky, Natalia;Eisner, Brian H. and Stoller, Marshall L.

**Publication Date:** Au ,2010

**Journal:** Journal of Urology 184(2), pp. 579–583

**Abstract: PURPOSE**: There is much debate about whether 1 or 2, 24-hour urinalyses are adequate for metabolic evaluation of stone formers. We determined whether repeat 24-hour urine collection provides information similar to that of the initial 24-hour urine collection and whether repeat collection is necessary. **MATERIALS AND METHODS**: We analyzed 2, 24-hour urine collections in 777 patients obtained from 2001 to 2005. Samples were collected 3 days or less apart before pharmacological intervention and analyzed elsewhere for routine stone risk profiles of urine calcium, oxalate, citrate, uric acid, sodium, potassium, magnesium, phosphorus, ammonium, chloride, urea nitrogen and creatinine. **RESULTS**: No parameters showed a statistically significant difference between 24-hour urine samples 1 and 2 when mean values were compared (pairwise t test each p >0.05, range 0.06 to 0.87). Using Pearson's correlation all parameters showed positive correlation coefficients (r = 0.68 to 0.89, each p 0.05) while for the remaining 6 p value was 0.05) while for the remaining 6 p value was **CONCLUSIONS**: One 24-hour urine sample is sufficient for metabolic evaluation of recurrent stone disease. There is no significant difference in 12 urinary parameters between 24-hour urine samples collected within 3 days of each other. This information is useful to providers and may decrease patient inconvenience and the overall cost of metabolic stone evaluation. Copyright (c) 2010 American Urological Association Education and Research, Inc. Published by Elsevier Inc. All rights reserved.

**URL:** <https://ovidsp.ovid.com/ovidweb.cgi?T=JS&CSC=Y&NEWS=N&PAGE=fulltext&D=med8&DO=10.1016%2fj.juro.2010.03.129> <https://libkey.io/libraries/2592/openurl?genre=article&sid=OVID:medline&id=pmid:20639021&id=doi:10.1016%2Fj.juro.2010.03.129&issn=0022-5347&isbn=&volume=184&issue=2&spage=579&pages=579-83&date=2010&title=Journal+of+Urology&atitle=Adequacy+of+a+single+24-hour+urine+collection+for+metabolic+evaluation+of+recurrent+nephrolithiasis.&aulast=Castle&pid=%3Cauthor%3ECastle+SM%3BCooperberg+MR%3BSadetsky+N%3BEisner+BH%3BStoller+ML%3C%2Fauthor%3E%3CAN%3E20639021%3C%2FAN%3E%3CDT%3EJournal+Article%3C%2FDT%3E>

94. **Urinary lithiasis and idiopathic hypercalciuria: the importance of dietary intake evaluation.**

**Item Type:** Journal Article

**Authors:** Damasio, Patricia C. G.;Amaro, Carmen R. P.;Berto, Silvia J. P.;Cunha, Natalia B.;Pichutte, Ana C.;Padovani, Carlos R. and Amaro, Joao L.

**Publication Date:** 2010

**Journal:** International Braz J Urol 36(5), pp. 557–562

**Abstract: PURPOSE**: To evaluate food intake of patients with urinary lithiasis and idiopathic hypercalciuria (IH). **MATERIALS AND METHODS**: Between August 2007 and June 2008, 105 patients with lithiasis were distributed into 2 groups: Group 1 (n = 55)--patients with IH (urinary calcium excretion > 250 mg in women and 300 mg in men with normal serum calcium); Group 2 (n = 50)--normocalciuria (NC) patients. Inclusion criteria were: age over 18, normal renal function (creatinine clearance >= 60 mL/min), absent proteinuria and negative urinary culture. Pregnant women, patients with some intestinal pathology, chronic diarrhea or using corticoids were excluded. The protocol of metabolic investigation was based on non-consecutive collection of two 24-hour samples for dosages of: calcium, sodium, uric acid, citrate, oxalate, magnesium and urinary volume. Food intake was evaluated through the quantitative method of Dietary Register of three days. **RESULTS**: Urinary excretion of calcium (433.33 +/- 141.92 vs. 188.93 +/- 53.09), sodium (280.08 +/- 100.94 vs. 200.44.93 +/- 65.81), uric acid (880.63 +/- 281.50 vs. 646.74 +/- 182.76) and magnesium (88.78 +/- 37.53 vs. 64.34 +/- 31.84) was significantly higher in the IH group in comparison to the NC group (p : Urinary excretion of calcium (433.33 +/- 141.92 vs. 188.93 +/- 53.09), sodium (280.08 +/- 100.94 vs. 200.44.93 +/- 65.81), uric acid (880.63 +/- 281.50 vs. 646.74 +/- 182.76) and magnesium (88.78 +/- 37.53 vs. 64.34 +/- 31.84) was significantly higher in the IH group in comparison to the NC group (p **CONCLUSION**: In our study, no difference was observed in the food intake of patients with urinary lithiasis and IH or NC.

**URL:** <https://ovidsp.ovid.com/ovidweb.cgi?T=JS&CSC=Y&NEWS=N&PAGE=fulltext&D=med8&DO=10.1590%2fs1677-55382010000500005> <https://libkey.io/libraries/2592/openurl?genre=article&sid=OVID:medline&id=pmid:21044372&id=doi:10.1590%2Fs1677-55382010000500005&issn=1677-5538&isbn=&volume=36&issue=5&spage=557&pages=557-62&date=2010&title=International+Braz+J+Urol&atitle=Urinary+lithiasis+and+idiopathic+hypercalciuria%3A+the+importance+of+dietary+intake+evaluation.&aulast=Damasio&pid=%3Cauthor%3EDamasio+PC%3BAmaro+CR%3BBerto+SJ%3BCunha+NB%3BPichutte+AC%3BPadovani+CR%3BAmaro+JL%3C%2Fauthor%3E%3CAN%3E21044372%3C%2FAN%3E%3CDT%3EComparative+Study%3C%2FDT%3E>

95. **Relationship between body mass index and quantitative 24-hour urine chemistries in patients with nephrolithiasis.**

**Item Type:** Journal Article

**Authors:** Eisner, Brian H.;Eisenberg, Michael L. and Stoller, Marshall L.

**Publication Date:** Ju ,2010

**Journal:** Urology 75(6), pp. 1289–1293

**Abstract: OBJECTIVES**: To examine the relationship between body mass index and 24-hour urine constituents in a population of stone-forming patients. **METHODS**: A total of 880 patients who presented to a metabolic stone clinic for initial evaluation were analyzed. Patients were stratified by gender and divided into quartiles of body mass index. Associations between body mass index (BMI) and urine parameters were explored using bivariate and multivariate linear regression. **RESULTS**: On bivariate analysis, increasing body mass index was associated with a significant increase in sodium, calcium, citrate, uric acid, magnesium, calcium oxalate, uric acid, and a decrease in pH in men. In women, it was associated with a significant increase in sodium, uric acid, oxalate, uric acid, and decreasing pH. On multivariate analysis, BMI was associated only with increases in sodium and calcium oxalate and decrease in pH in men. In women, multivariate analysis demonstrated positive association between BMI and urine sodium, creatinine, and phosphate and a negative relationship with urine citrate and sulphate. **CONCLUSIONS**: Increasing body mass index was related to several risk factors for urinary stone disease in this study, including increasing urine sodium and decreasing pH in men and increasing urine uric acid, sodium, and decreasing urine citrate in women. Just as general recommendations for patients with nephrolithiasis include high voided volumes, low dietary sodium, and low animal protein intake, perhaps weight reduction should be included as part of the counseling of stone-formers to optimize 24-hour urine parameters. Copyright (c) 2010 Elsevier Inc. All rights reserved.

**URL:** <https://ovidsp.ovid.com/ovidweb.cgi?T=JS&CSC=Y&NEWS=N&PAGE=fulltext&D=med8&DO=10.1016%2fj.urology.2009.09.024> <https://libkey.io/libraries/2592/openurl?genre=article&sid=OVID:medline&id=pmid:20018350&id=doi:10.1016%2Fj.urology.2009.09.024&issn=0090-4295&isbn=&volume=75&issue=6&spage=1289&pages=1289-93&date=2010&title=Urology&atitle=Relationship+between+body+mass+index+and+quantitative+24-hour+urine+chemistries+in+patients+with+nephrolithiasis.&aulast=Eisner&pid=%3Cauthor%3EEisner+BH%3BEisenberg+ML%3BStoller+ML%3C%2Fauthor%3E%3CAN%3E20018350%3C%2FAN%3E%3CDT%3EComparative+Study%3C%2FDT%3E>

96. **Twenty-four hour and spot urine metabolic evaluations: correlations versus agreements.**

**Item Type:** Journal Article

**Authors:** Hong, Yet Hoi;Dublin, Norman;Razack, Azad Hassan;Mohd, Mustafa Ali and Husain, Ruby

**Publication Date:** Ju ,2010

**Journal:** Urology 75(6), pp. 1294–1298

**Abstract: OBJECTIVES**: To investigate the correlations and agreements between the solute/creatinine ratios from the 24-hour and early morning spot urine samples for metabolic evaluation in stone-formers given the various pitfalls with the 24-hour urinary metabolic evaluation in stone-formers. **METHODS**: 30 urinary stone-formers out of an initial 62 recruited provided a complete 24-hour urine and early morning spot urine samples for metabolic evaluation. Pearson correlation and Bland and Altman Test were used to assess the correlations and agreements. **RESULTS**: Significant correlations were established between the 24-hour urinary solute excretions and the corresponding early morning spot urine solute/creatinine ratios for calcium, magnesium, urate, potassium, oxalate, citrate, and the Differential Gibb's free energy value of calcium oxalate DG(CaOx) values. However, all these solute/creatinine measurements between the 24-hour and early morning spot urine samples were judged to be not within the acceptable limits based on the estimated "limit of agreement" by the Bland and Altman Test of Agreement. Diurnal circadian rhythm and postprandial excretion surge are thought to be responsible for the disagreements. **CONCLUSIONS**: Thus, the early morning spot urine is not suitable to be used interchangeably to replace the 24-hour urine collection in the evaluation of urinary metabolic abnormalities in stone-formers. A good correlation does not translate to an agreement between the 2 measurements. Copyright (c) 2010 Elsevier Inc. All rights reserved.

**URL:** <https://ovidsp.ovid.com/ovidweb.cgi?T=JS&CSC=Y&NEWS=N&PAGE=fulltext&D=med8&DO=10.1016%2fj.urology.2009.08.061> <https://libkey.io/libraries/2592/openurl?genre=article&sid=OVID:medline&id=pmid:19914693&id=doi:10.1016%2Fj.urology.2009.08.061&issn=0090-4295&isbn=&volume=75&issue=6&spage=1294&pages=1294-8&date=2010&title=Urology&atitle=Twenty-four+hour+and+spot+urine+metabolic+evaluations%3A+correlations+versus+agreements.&aulast=Hong&pid=%3Cauthor%3EHong+YH%3BDublin+N%3BRazack+AH%3BMohd+MA%3BHusain+R%3C%2Fauthor%3E%3CAN%3E19914693%3C%2FAN%3E%3CDT%3EComparative+Study%3C%2FDT%3E>

97. **Metabolic evaluation in stone disease metabolic differences between the pediatric and adult patients with stone disease.**

**Item Type:** Journal Article

**Authors:** Karabacak, Osman Raif;Ipek, Bulent;Ozturk, Ufuk;Demirel, Fuat;Saltas, Hakan and Altug, Ugur

**Publication Date:** Ju ,2010

**Journal:** Urology 76(1), pp. 238–241

**Abstract: OBJECTIVE**: To determine the metabolic risk factors in children and adults with stone disease and any differences between the metabolic risk factors of children and adult patients with stone disease. **METHODS**: Between 1999 and 2007, 285 adults with recurrent stone disease and 71 children with primary or recurrent urinary stone disease underwent metabolic evaluation in our clinic. The evaluation included blood chemistry studies and 24-hour urine collection in the pediatric and adult groups. **RESULTS**: The mean age of the children was 9.35 years (range: 1-14) and 42.1 years (range: 14-71) in the adult patients. Metabolic risk factors were demonstrated in 90.53% (n = 258) of the adult and 88.73% (n = 63) of the pediatric patients. Although hypercalciuria (50.5%, n = 144) was the most common metabolic risk factor in the adult group, hypocitraturia was the most common metabolic risk factor in the pediatric group (57.74%, n = 41). Furthermore, in the adult group, 136 patients had more than 1 risk factor, and in the pediatric group, 24 patients had more than 1 risk factor. **CONCLUSION**: Although hypocitraturia is the most common risk factor in pediatric urolithiasis patients and hypercalciuria is the most common risk factor in adult patients, all patients with stone formation should be given a limited metabolic evaluation because such patients may also have a metabolic abnormality. Copyright 2010 Elsevier Inc. All rights reserved.

**URL:** <https://ovidsp.ovid.com/ovidweb.cgi?T=JS&CSC=Y&NEWS=N&PAGE=fulltext&D=med8&DO=10.1016%2fj.urology.2010.01.036> <https://libkey.io/libraries/2592/openurl?genre=article&sid=OVID:medline&id=pmid:20350756&id=doi:10.1016%2Fj.urology.2010.01.036&issn=0090-4295&isbn=&volume=76&issue=1&spage=238&pages=238-41&date=2010&title=Urology&atitle=Metabolic+evaluation+in+stone+disease+metabolic+differences+between+the+pediatric+and+adult+patients+with+stone+disease.&aulast=Karabacak&pid=%3Cauthor%3EKarabacak+OR%3BIpek+B%3BOzturk+U%3BDemirel+F%3BSaltas+H%3BAltug+U%3C%2Fauthor%3E%3CAN%3E20350756%3C%2FAN%3E%3CDT%3EComparative+Study%3C%2FDT%3E>

98. **Metabolic basis for low urine pH in type 2 diabetes.**

**Item Type:** Journal Article

**Authors:** Maalouf, Naim M.;Cameron, Mary Ann;Moe, Orson W. and Sakhaee, Khashayar

**Publication Date:** Ju ,2010

**Journal:** Clinical Journal of the American Society of Nephrology: CJASN 5(7), pp. 1277–1281

**Abstract: BACKGROUND AND OBJECTIVES**: Type 2 diabetes is associated with excessively low urine pH, which increases the risk for uric acid nephrolithiasis. This study was conducted to assess the metabolic basis responsible for the excessive urinary acidity of individuals with type 2 diabetes. **DESIGN, SETTING, PARTICIPANTS, & MEASUREMENTS**: Nine non-stone-forming patients who had type 2 diabetes and low urine pH and 16 age- and body mass index-matched non-stone-forming volunteers without type 2 diabetes were maintained on a constant metabolic diet for 7 days, and 24-hour urine was collected on the last 2 days of the diet. **RESULTS**: Urine dietary markers (potassium, sulphate, phosphorus, and urea nitrogen) were not different between the two groups. Patients with type 2 diabetes exhibited a significantly lower 24-hour urine pH (5.45+/-0.27 versus 5.90+/-0.42; P: Urine dietary markers (potassium, sulphate, phosphorus, and urea nitrogen) were not different between the two groups. Patients with type 2 diabetes exhibited a significantly lower 24-hour urine pH (5.45+/-0.27 versus 5.90+/-0.42; P: Urine dietary markers (potassium, sulphate, phosphorus, and urea nitrogen) were not different between the two groups. Patients with type 2 diabetes exhibited a significantly lower 24-hour urine pH (5.45+/-0.27 versus 5.90+/-0.42; P: Urine dietary markers (potassium, sulphate, phosphorus, and urea nitrogen) were not different between the two groups. Patients with type 2 diabetes exhibited a significantly lower 24-hour urine pH (5.45+/-0.27 versus 5.90+/-0.42; P**CONCLUSIONS**: The overly acidic urine in patients with type 2 diabetes persists after controlling for dietary factors, body size, and age. The lower pH is due to a combination of greater NAE and lower use of ammonia buffers in patients with diabetes, which predisposes them to uric acid urolithiasis.

**URL:** <https://ovidsp.ovid.com/ovidweb.cgi?T=JS&CSC=Y&NEWS=N&PAGE=fulltext&D=med8&DO=10.2215%2fCJN.08331109> <https://libkey.io/libraries/2592/openurl?genre=article&sid=OVID:medline&id=pmid:20413437&id=doi:10.2215%2FCJN.08331109&issn=1555-9041&isbn=&volume=5&issue=7&spage=1277&pages=1277-81&date=2010&title=Clinical+Journal+of+The+American+Society+of+Nephrology%3A+CJASN&atitle=Metabolic+basis+for+low+urine+pH+in+type+2+diabetes.&aulast=Maalouf&pid=%3Cauthor%3EMaalouf+NM%3BCameron+MA%3BMoe+OW%3BSakhaee+K%3C%2Fauthor%3E%3CAN%3E20413437%3C%2FAN%3E%3CDT%3EControlled+Clinical+Trial%3C%2FDT%3E>

99. **Bonn Risk Index based micromethod for assessing risk of urinary calcium oxalate stone formation.**

**Item Type:** Journal Article

**Authors:** Porowski, T.;Mrozek, P.;Sidun, J.;Zoch-Zwierz, W.;Konstantynowicz, J.;Kirejczyk, J. K.;Motkowski, R. and Laube, N.

**Publication Date:** Ma ,2010

**Journal:** Journal of Urology 183(3), pp. 1157–1162

**Abstract: PURPOSE**: The Bonn Risk Index has been used to evaluate the risk of urinary calcium oxalate stone formation. According to the original method, risk should be determined based on a 200 ml urine sample taken from a 24-hour collection. We evaluated whether the Bonn Risk Index can also be effectively determined in small urine samples. **MATERIALS AND METHODS**: We studied 190 children and adolescents with nocturia and calcium oxalate urolithiasis. Initially Bonn Risk Index was determined according to the original method of Laube. Subsequently Bonn Risk Index was calculated using a computer program controlling a specially designed system to define the time point of induced crystallization based on consecutive urine samples of 1.5, 2.0 and 3.0 ml. **RESULTS**: No significant differences were found in Bonn Risk Index between values obtained from 200 ml samples and those based on the micromethod with urine samples of 2 and 3 ml. **CONCLUSIONS**: Assessment of risk of urinary calcium oxalate stone formation with Bonn Risk Index in small urine volumes, based on prototype equipment controlled by specialized computer software, is comparable to the original method. This finding facilitates the procedure and improves Bonn Risk Index determination in children. Copyright 2010 American Urological Association Education and Research, Inc. Published by Elsevier Inc. All rights reserved.

**URL:** <https://ovidsp.ovid.com/ovidweb.cgi?T=JS&CSC=Y&NEWS=N&PAGE=fulltext&D=med8&DO=10.1016%2fj.juro.2009.11.054> <https://libkey.io/libraries/2592/openurl?genre=article&sid=OVID:medline&id=pmid:20096860&id=doi:10.1016%2Fj.juro.2009.11.054&issn=0022-5347&isbn=&volume=183&issue=3&spage=1157&pages=1157-62&date=2010&title=Journal+of+Urology&atitle=Bonn+Risk+Index+based+micromethod+for+assessing+risk+of+urinary+calcium+oxalate+stone+formation.&aulast=Porowski&pid=%3Cauthor%3EPorowski+T%3BMrozek+P%3BSidun+J%3BZoch-Zwierz+W%3BKonstantynowicz+J%3BKirejczyk+JK%3BMotkowski+R%3BLaube+N%3C%2Fauthor%3E%3CAN%3E20096860%3C%2FAN%3E%3CDT%3EComparative+Study%3C%2FDT%3E>

100. **Stone forming risk factors in patients with type Ia glycogen storage disease.**

**Item Type:** Journal Article

**Authors:** Scales, Charles D. Jr;Chandrashekar, Aravind S.;Robinson, Marnie R.;Cantor, David A.;Sullivan, Jennifer;Haleblian, George E.;Leitao, Victor A.;Sur, Roger L.;Borawski, Kristy M.;Koeberl, Dwight;Kishnani, Priya S. and Preminger, Glenn M.

**Publication Date:** Ma ,2010

**Journal:** Journal of Urology 183(3), pp. 1022–1025

**Abstract: PURPOSE**: Patients with type Ia glycogen storage disease have an increased recurrent nephrolithiasis rate. We identified stone forming risk factors in patients with type Ia glycogen storage disease vs those in stone formers without the disease. **MATERIALS AND METHODS**: Patients with type Ia glycogen storage disease were prospectively enrolled from our metabolic clinic. Patient 24-hour urine parameters were compared to those in age and gender matched stone forming controls. **RESULTS**: We collected 24-hour urine samples from 13 patients with type Ia glycogen storage disease. Average +/- SD age was 27.0 +/- 13.0 years and 6 patients (46%) were male. Compared to age and gender matched hypocitraturic, stone forming controls patients had profound hypocitraturia (urinary citrate 70 vs 344 mg daily, p = 0.009). When comparing creatinine adjusted urinary values, patients had profound hypocitraturia (0.119 vs 0.291 mg/mg creatinine, p = 0.005) and higher oxalate (0.026 vs 0.021 mg/mg creatinine, p = 0.038) vs other stone formers. **CONCLUSIONS**: Patients with type Ia glycogen storage disease have profound hypocitraturia, as evidenced by 24-hour urine collections, even compared to other stone formers. This may be related to a recurrent nephrolithiasis rate greater than in the overall population. These findings may be used to support different treatment modalities, timing and/or doses to prevent urinary lithiasis in patients with type Ia glycogen storage disease. Copyright 2010 American Urological Association Education and Research, Inc. Published by Elsevier Inc. All rights reserved.

**URL:** <https://ovidsp.ovid.com/ovidweb.cgi?T=JS&CSC=Y&NEWS=N&PAGE=fulltext&D=med8&DO=10.1016%2fj.juro.2009.11.040> <https://libkey.io/libraries/2592/openurl?genre=article&sid=OVID:medline&id=pmid:20092831&id=doi:10.1016%2Fj.juro.2009.11.040&issn=0022-5347&isbn=&volume=183&issue=3&spage=1022&pages=1022-5&date=2010&title=Journal+of+Urology&atitle=Stone+forming+risk+factors+in+patients+with+type+Ia+glycogen+storage+disease.&aulast=Scales&pid=%3Cauthor%3EScales+CD+Jr%3BChandrashekar+AS%3BRobinson+MR%3BCantor+DA%3BSullivan+J%3BHaleblian+GE%3BLeitao+VA%3BSur+RL%3BBorawski+KM%3BKoeberl+D%3BKishnani+PS%3BPreminger+GM%3C%2Fauthor%3E%3CAN%3E20092831%3C%2FAN%3E%3CDT%3EJournal+Article%3C%2FDT%3E>

101. **Effect of dietary changes on urinary oxalate excretion and calcium oxalate supersaturation in patients with hyperoxaluric stone formation.**

**Item Type:** Journal Article

**Authors:** Penniston, Kristina L. and Nakada, Stephen Y.

**Publication Date:** Ma ,2009

**Journal:** Urology 73(3), pp. 484–489

**Abstract: OBJECTIVES**: To test the hypothesis that patients with hyperoxaluria, who modified their dietary calcium intake, would reduce their urinary oxalate excretion without raising their urinary calcium excretion. Diet is a major factor in idiopathic calcium oxalate urolithiasis, yet controversy exists regarding the ideal clinical recommendations. Approximately 20% of patients with calcium oxalate stone formation have hyperoxaluria (> or = 45 mg oxalate/d). Calcium supplements to bind dietary oxalate have been suggested, but clinical evidence of this therapy is lacking. **METHODS**: Of 144 adult patients with stone formation seen by a registered dietitian from September 2006 to September 2007, 26 (18%) had hyperoxaluria on > or = 1 24-hour urinalyses. Of those with > or = 2 complete 24-hour collections and whose hyperoxaluria was observed before their last visit with the registered dietitian, 22 patients were identified. The patients were retrospectively separated into 2 groups according to whether they had been advised dietary changes alone (diet group, n = 10) or calcium citrate with meals, in addition to the dietary changes (supplement group, n = 12). The mean follow-up time was 317 and 266 days for the diet and supplement groups, respectively. Statistical comparisons within and between groups were made for urinary risk factors. **RESULTS**: Urinary oxalate excretion decreased from 56 to 43 mg/d and from 60 to 46 mg/d in the diet and supplement groups, respectively (P = .003 and P = .038, respectively). Calcium oxalate supersaturation decreased from 3.48 to 1.83 and from 2.37 to 1.52 in the diet and supplement groups, respectively (P = .043 and P = .002, respectively). Urinary calcium excretion did not change in either group. **CONCLUSIONS**: Gastrointestinal binding of oxalate by calcium is an effective clinical strategy for hyperoxaluria, whether mediated by calcium citrate with meals or by inclusion of calcium-containing foods with meals.

**URL:** <https://ovidsp.ovid.com/ovidweb.cgi?T=JS&CSC=Y&NEWS=N&PAGE=fulltext&D=med7&DO=10.1016%2fj.urology.2008.10.035> <https://libkey.io/libraries/2592/openurl?genre=article&sid=OVID:medline&id=pmid:19118878&id=doi:10.1016%2Fj.urology.2008.10.035&issn=0090-4295&isbn=&volume=73&issue=3&spage=484&pages=484-9&date=2009&title=Urology&atitle=Effect+of+dietary+changes+on+urinary+oxalate+excretion+and+calcium+oxalate+supersaturation+in+patients+with+hyperoxaluric+stone+formation.&aulast=Penniston&pid=%3Cauthor%3EPenniston+KL%3BNakada+SY%3C%2Fauthor%3E%3CAN%3E19118878%3C%2FAN%3E%3CDT%3EJournal+Article%3C%2FDT%3E>

102. **Risk factors for renal calcium stone formation in South African and European young adults.**

**Item Type:** Journal Article

**Authors:** Rodgers, Allen;Allie-Hamdulay, Shameez;Pinnock, Diane;Baretta, Gretchen and Trinchieri, Alberto

**Publication Date:** Se ,2009

**Journal:** Archivio Italiano Di Urologia, Andrologia 81(3), pp. 171–174

**Abstract: OBJECTIVES**: The different susceptibility to renal stone disease of white and black people has been previously explained in terms of intrinsic (genetics) and extrinsic (diet, lifestyle) factors. However, in South Africa, the absence of stone disease in the black population has not yet been fully explained by either of these. The aim of the present study was to identify potential differences between black and white subjects in South Africa and white subjects in Europe with respect to their relative dietary and urinary risk factors for renal stone formation. **MATERIALS AND METHODS**: A total of 72 healthy subjects (45 males and 27 females, age range 21-30 years) with no previous history of renal stone disease or specific diseases predisposing to renal stone formation were recruited in South Africa (SA) and in Italy (IT). They were divided in three groups: South African blacks (SA-B), South African whites (SA-W) and Italian whites (IT-W). Each participant provided a 24-hour dietary record and 24-hour urine sample taken over the same period. Nutrients and calories were calculated by means of food composition tables using a computerised procedure. Urinary concentrations of potassium, sodium, calcium, phosphate, oxalate, urate, citrate, magnesium, and creatinine, together with the pH and urinary volumes, were measured. **RESULTS**: The mean carbohydrate intake was significantly higher in SA-B (293+90 g/day) than in both SA-W (194+74, p = 0.002) and IT-W (212 +/- 81; p = 0.000). Daily magnesium intake was higher in SA-B (290+124 mg/day) than in IT-W (176+73 mg/day, p = 0.002). The mean daily urinary excretion of calcium was significantly (p = 0.029) lower in SA-B (3.07 +/- 1.68 mmol/day) with respect to SA-W (4.65 +/- 2.44 mmol/day) and IT-W (4.51 +/- 1.89 mmol/day) whereas mean daily urinary excretion of citrate was significantly (P = 0.012) higher in SA-B (3.36 +/- 1.4 mmol/day) than in SA-W (3.09 +/- 1.45 mmol/day) and IT-W (2.36 +/- 0.98 mmol/day). **CONCLUSION**: Although the carbohydrate intake and the percent of energy from carbohydrate of black subjects in this study were higher with respect to white controls, we were not able to show any other relevant difference of the known dietary stone risk patterns between black and white subjects. On the other hand the urinary patterns of black controls seem to be more favourable in term of risk for stone formation than those of white controls showing a lower calcium excretion and a higher citrate excretion in the urine. Our result of higher carbohydrate intake in black subjects is counter-intuitive as it suggests a higher risk of stone formation in this group. This puzzling result may have arisen because our subjects were recruited from the urban population rather than from rural areas, suggesting that western diets and lifestyles may ultimately change the stone incidence profile in the black population.

**URL:** <https://ovidsp.ovid.com/ovidweb.cgi?T=JS&CSC=Y&NEWS=N&PAGE=fulltext&D=med7&AN=19911680> <https://libkey.io/libraries/2592/openurl?genre=article&sid=OVID:medline&id=pmid:19911680&id=doi:&issn=1124-3562&isbn=&volume=81&issue=3&spage=171&pages=171-4&date=2009&title=Archivio+Italiano+di+Urologia%2C+Andrologia&atitle=Risk+factors+for+renal+calcium+stone+formation+in+South+African+and+European+young+adults.&aulast=Rodgers&pid=%3Cauthor%3ERodgers+A%3BAllie-Hamdulay+S%3BPinnock+D%3BBaretta+G%3BTrinchieri+A%3C%2Fauthor%3E%3CAN%3E19911680%3C%2FAN%3E%3CDT%3EComparative+Study%3C%2FDT%3E>

103. **Role of overweight status on stone-forming risk factors in children: a prospective study.**

**Item Type:** Journal Article

**Authors:** Sarica, Kemal;Eryildirim, Bilal;Yencilek, Faruk and Kuyumcuoglu, Ugur

**Publication Date:** Ma ,2009

**Journal:** Urology 73(5), pp. 1003–1007

**Abstract: OBJECTIVES**: To evaluate the possible role of being overweight on stone-forming risk factors in children. **METHODS**: A total of 94 children (43 boys and 51 girls, male/female ratio 1:1.8) who were taking no medication or dietary modifications before treatment were included in the study. After a detailed stone disease history, the systolic and diastolic blood pressures were precisely measured and recorded for all patients. The body mass index, 24-hour urine values, and serum stone-forming risk parameters were evaluated in 44 overweight (17 boys and 27 girls; group 1) and 50 normal (26 boys and 24 girls; group 2) children. The results of each group were compared using the Wilcoxon rank sum test. **RESULTS**: The evaluation of the stone-forming risk factors in both groups revealed that the overweight status might be responsible for the increased excretion of these substances in such children. Most of the children in group 1 demonstrated hypocitraturia and hyperoxaluria (9/44, 20.5%) compared with the patients in group 2. Although the mean urinary oxalate level was 0.74 +/- 0.81 mg/kg/24 h for boys and 0.69 +/- 0.72 mg/kg/24 h for girls in group 1, relatively lower values were noted in group 2 (0.42 +/- 0.52 and 0.45 +/- 0.57 mg/kg/24 h for the boys and girls, respectively). Similarly, the children in group 1 had elevated mean urinary calcium and lower citrate excretion compared with the group 2 patients. **CONCLUSIONS**: Overweight status in children might be associated with an elevated risk of stone formation in both sexes owing to the alterations in urine composition. Obese children could be more prone to stone formation, and they should be evaluated and followed up for this aspect.

**URL:** <https://ovidsp.ovid.com/ovidweb.cgi?T=JS&CSC=Y&NEWS=N&PAGE=fulltext&D=med7&DO=10.1016%2fj.urology.2008.11.038> <https://libkey.io/libraries/2592/openurl?genre=article&sid=OVID:medline&id=pmid:19193407&id=doi:10.1016%2Fj.urology.2008.11.038&issn=0090-4295&isbn=&volume=73&issue=5&spage=1003&pages=1003-7&date=2009&title=Urology&atitle=Role+of+overweight+status+on+stone-forming+risk+factors+in+children%3A+a+prospective+study.&aulast=Sarica&pid=%3Cauthor%3ESarica+K%3BEryildirim+B%3BYencilek+F%3BKuyumcuoglu+U%3C%2Fauthor%3E%3CAN%3E19193407%3C%2FAN%3E%3CDT%3EComparative+Study%3C%2FDT%3E>

104. **Metabolic and physico-chemical urolithiasis parameters in the first morning urine.**

**Item Type:** Journal Article

**Authors:** Seric, Vatroslav;Dutour-Sikiric, Maja;Mihaljevic, Ivan;Tucak-Zoric, Sandra;Bilic-Curcic, Ines and Babic-Ivancic, Vesna

**Publication Date:** De ,2009

**Journal:** Collegium Antropologicum 33(Suppl 2), pp. 85–92

**Abstract:** The 24-hour urine is golden standard for metabolic assessment of stone formers. However, due to the difficulties in collecting almost 1/3 of the samples can not be used for the analysis. Therefore, we analyzed first morning urine and calculated different risk indexes in order to asses possibility of using it in determining urolithiasis risk. Subjects were divided into 4 groups: male patients (n = 31, age 18-64), female patients (n = 31, age 25-63), male controls (n = 16, age 25-64) and female controls (n = 19, age 21-65). First morning urine pH, concentrations of calcium, magnesium, phosphate, sodium, potassium, chloride, citrate, urate, oxalate, creatinine and glycosaminoglycans were determined. Based on them, ionic concentrations and activity products of calcium oxalate and phosphate were calculated by EQUIL 2. In addition, different risk indices were calculated. The results showed that both patients and control groups had metabolic disorders, but the frequencies of occurrence were statistically independent. Significant difference in concentration of urinary constituents between corresponding patient and control groups was observed only for glycosaminoglycans in female subjects. Ca/Cit ratio and Baggio index could differentiate between both corresponding patients and control groups. The results indicate that interplay between stone formation inhibitors and promotors is responsible for urinary stone formation and that the first morning urine could be used in assessing urolithiasis risk and its prevention.

**URL:** <https://ovidsp.ovid.com/ovidweb.cgi?T=JS&CSC=Y&NEWS=N&PAGE=fulltext&D=med7&AN=20120404> <https://libkey.io/libraries/2592/openurl?genre=article&sid=OVID:medline&id=pmid:20120404&id=doi:&issn=0350-6134&isbn=&volume=33&issue=2&spage=85&pages=85-92&date=2009&title=Collegium+Antropologicum&atitle=Metabolic+and+physico-chemical+urolithiasis+parameters+in+the+first+morning+urine.&aulast=Seric&pid=%3Cauthor%3ESeric+V%3BDutour-Sikiric+M%3BMihaljevic+I%3BTucak-Zoric+S%3BBilic-Curcic+I%3BBabic-Ivancic+V%3C%2Fauthor%3E%3CAN%3E20120404%3C%2FAN%3E%3CDT%3EJournal+Article%3C%2FDT%3E>

105. **Effect of carbohydrate-electrolyte sports beverages on urinary stone risk factors.**

**Item Type:** Journal Article

**Authors:** Sweeney, Danielle D.;Tomaszewski, Jeffrey J.;Ricchiuti, Daniel D. and Averch, Timothy D.

**Publication Date:** Se ,2009

**Journal:** Journal of Urology 182(3), pp. 992–997

**Abstract: PURPOSE**: We evaluated the effects of consuming carbohydrate-electrolyte sports beverages (Gatorade((R))) on urinary stone risk factors. **MATERIALS AND METHODS**: Twelve normal subjects (5 men, 7 women) and 12 hypercalciuric stone formers (2 men, 10 women) participated in a 4-week prospective, crossover study consisting of 3 study phases. In phase 1 subjects were placed on a monitored stone prevention diet that was continued throughout the study. In phase 2 subjects ingested 2 l Gatorade daily followed by a 7-day washout period. In phase 3 subjects ingested 2 l water daily. On the final day of phases 1, 2 and 3 a 24-hour urine collection and blood sample were analyzed for stone risk factors. Effects of group and phase were tested using repeated measures ANOVA and paired t tests. **RESULTS**: Changes in urinary risk factors after Gatorade consumption revealed no statistically significant difference between normal subjects and stone formers. However, intrasubject variation occurred in both groups. Gatorade consumption in both groups increased urinary pH (p = 0.006), urinary chloride (p = 0.044) and urinary sodium (p = 0.008), and decreased urinary potassium (p = 0.035) and urinary uric acid (p = 0.019) in a statistically significant manner. In response to Gatorade consumption urinary volume, calcium and citrate were unchanged compared to water consumption and baseline. **CONCLUSIONS**: Gatorade increased mean urinary sodium and chloride levels compared to water and baseline. However, the results were within normal urinary parameters. The change did not appear to be clinically significant as urinary calcium was unchanged. Overall consumption of Gatorade does not increase or decrease urinary stone risk factors.

**URL:** <https://ovidsp.ovid.com/ovidweb.cgi?T=JS&CSC=Y&NEWS=N&PAGE=fulltext&D=med7&DO=10.1016%2fj.juro.2009.05.020> <https://libkey.io/libraries/2592/openurl?genre=article&sid=OVID:medline&id=pmid:19616798&id=doi:10.1016%2Fj.juro.2009.05.020&issn=0022-5347&isbn=&volume=182&issue=3&spage=992&pages=992-7&date=2009&title=Journal+of+Urology&atitle=Effect+of+carbohydrate-electrolyte+sports+beverages+on+urinary+stone+risk+factors.&aulast=Sweeney&pid=%3Cauthor%3ESweeney+DD%3BTomaszewski+JJ%3BRicchiuti+DD%3BAverch+TD%3C%2Fauthor%3E%3CAN%3E19616798%3C%2FAN%3E%3CDT%3EComparative+Study%3C%2FDT%3E>

106. **Dissolution of radiolucent renal stones by oral alkalinization with potassium citrate/potassium bicarbonate.**

**Item Type:** Journal Article

**Authors:** Trinchieri, Alberto;Esposito, Nicola and Castelnuovo, Chiara

**Publication Date:** Se ,2009

**Journal:** Archivio Italiano Di Urologia, Andrologia 81(3), pp. 188–191

**Abstract: AIM**: Uric acid stone disease is dependent on three pathogenetic factors: acid urine pH, low urine volume, and hyperuricosuria. However, the most important factor for uric acid stone formation is persistently acidic urine that represents a prerequisite for uric acid stone formation and growth. Urinary alkalization with alkali administration has been advocated for dissolution of stones on the basis of estabilished clinical experience. The aim of this study was to evaluate the clinical efficacy of therapy with potassium citrate/potassium bicarbonate for dissolution of radiolucent stones. **PATIENTS AND METHODS**: A total of 8 patients with radiolucent stones (: A total of 8 patients with radiolucent stones (**RESULTS**: During the first period of treatment stone burden remained unchanged in all patients. On the contrary after 6 weeks of potassium citrate/bicarbonate treatment, complete stone dissolution was found in three of the patients. In the other five cases a partial dissolution was observed and in two of them complete dissolution of the stone was achieved after prolongation of the treatment for 4 and 6 month respectively. Mean urinary volumes were unchanged during all the two study periods. Mean urinary pH was significantly higher during the potassium citrate/bicarbonate treatment period in comparison to the first study period (morning 6.60 +/- 1.06 vs 5.53 +/- 0.51, p = 0.030; afternoon 6.53 +/- 0.70 vs 5.63 +/- 0.41, p = 0.007; night 6.57 +/- 0.51 vs 5.98 +/- 0.80, p = 0.092). Tolerance of the drug was good, and no serious effects were observed sufficient to interrupt treatment. None of the patients required subsequent interventions for stone treatment. **CONCLUSION**: Urinary alkalization with potassium citrate/bicarbonate is a well tolerated and highly effective treatment, resulting in dissolution of nonobstructing uric acid stones.

**URL:** <https://ovidsp.ovid.com/ovidweb.cgi?T=JS&CSC=Y&NEWS=N&PAGE=fulltext&D=med7&AN=19911683> <https://libkey.io/libraries/2592/openurl?genre=article&sid=OVID:medline&id=pmid:19911683&id=doi:&issn=1124-3562&isbn=&volume=81&issue=3&spage=188&pages=188-91&date=2009&title=Archivio+Italiano+di+Urologia%2C+Andrologia&atitle=Dissolution+of+radiolucent+renal+stones+by+oral+alkalinization+with+potassium+citrate%2Fpotassium+bicarbonate.&aulast=Trinchieri&pid=%3Cauthor%3ETrinchieri+A%3BEsposito+N%3BCastelnuovo+C%3C%2Fauthor%3E%3CAN%3E19911683%3C%2FAN%3E%3CDT%3EJournal+Article%3C%2FDT%3E>

107. **Effect of potassium citrate therapy on the risk of renal stone formation during spaceflight.**

**Item Type:** Journal Article

**Authors:** Whitson, Peggy A.;Pietrzyk, Robert A.;Jones, Jeffrey A.;Nelman-Gonzalez, Mayra;Hudson, Edgar K. and Sams, Clarence F.

**Publication Date:** No ,2009

**Journal:** Journal of Urology 182(5), pp. 2490–2496

**Abstract: PURPOSE**: Exposure to microgravity affects human physiology and results in changes in urinary chemical composition during and after spaceflight, favoring an increased risk of renal stones. We assessed the efficacy of potassium citrate to decrease the stone risk during and after spaceflight. **MATERIALS AND METHODS**: The study was done in 30 long duration spaceflight crew members to the space stations Mir and International Space Station. Before, during and after spaceflight 24-hour urine samples were collected to assess the renal stone risk. Potassium citrate (20 mEq) was ingested daily by International Space Station crew members in a double-blind, placebo controlled study. Mir crew members performed the identical protocol but did not ingest medication. **RESULTS**: Potassium citrate treated crew members had decreased urinary calcium excretion and maintained the calcium oxalate supersaturation risk at preflight levels compared to that in controls. Increased urinary pH in the treatment group decreased the risk of uric acid stones. **CONCLUSIONS**: Results from this investigation suggest that supplementation with potassium citrate may decrease the risk of renal stone formation during and immediately after spaceflight.

**URL:** <https://ovidsp.ovid.com/ovidweb.cgi?T=JS&CSC=Y&NEWS=N&PAGE=fulltext&D=med7&DO=10.1016%2fj.juro.2009.07.010> <https://libkey.io/libraries/2592/openurl?genre=article&sid=OVID:medline&id=pmid:19765769&id=doi:10.1016%2Fj.juro.2009.07.010&issn=0022-5347&isbn=&volume=182&issue=5&spage=2490&pages=2490-6&date=2009&title=Journal+of+Urology&atitle=Effect+of+potassium+citrate+therapy+on+the+risk+of+renal+stone+formation+during+spaceflight.&aulast=Whitson&pid=%3Cauthor%3EWhitson+PA%3BPietrzyk+RA%3BJones+JA%3BNelman-Gonzalez+M%3BHudson+EK%3BSams+CF%3C%2Fauthor%3E%3CAN%3E19765769%3C%2FAN%3E%3CDT%3EJournal+Article%3C%2FDT%3E>

108. **Urinary stone disease in adults with celiac disease: prevalence, incidence and urinary determinants.**

**Item Type:** Journal Article

**Authors:** Ciacci, Carolina;Spagnuolo, Giuliano;Tortora, Raffaella;Bucci, Cristina;Franzese, Domenica;Zingone, Fabiana and Cirillo, Massimo

**Publication Date:** Se ,2008

**Journal:** Journal of Urology 180(3), pp. 974–979

**Abstract: PURPOSE**: Intestinal diseases may cause urinary stone disease via hyperoxaluria or diarrhea induced hyperconcentrated acidic urine. Data are missing on urinary stone disease in celiac disease, a common malabsorptive disorder. In this study we analyzed urinary stone disease and urine composition in adults with celiac disease. **MATERIALS AND METHODS**: Study patients were 18 years or older, untreated, and newly diagnosed with celiac disease by serum markers and jejunal biopsy. Clinical presentation of celiac disease was assessed focusing on 5 disorders of diarrhea, and deficiency of calorie (low body mass index or weight loss), lipid (low prothrombin time or low serum lipids), iron (low hemoglobin or low serum ferritin) and calcium (low serum calcium or low bone densitometry). Urinary stone disease history was assessed by questionnaire (imaging, stone excretion, stone disruption/removal). Urinary variables were measured in a 24-hour collection in a subgroup of patients. **RESULTS**: Under untreated conditions (baseline) urinary stone disease was independent of celiac disease presentation and more prevalent in patients with celiac disease than in a population sample used as a control (608 and 3,540, 7.9% and 5.0%, sex and age adjusted odds ratio 4.0, 95% CI 2.7-5.9). Excluding from analysis individuals with baseline urinary stone disease, the incidence of urinary stone disease history was not significantly different between the treated celiac disease (gluten-free diet) and control population (458 and 3,003, 2.4% vs 3.9%). The urine of untreated patients with celiac disease differed from that of healthy volunteers with 120% higher oxalate and 43% lower calcium (in 45 and 45, p : Under untreated conditions (baseline) urinary stone disease was independent of celiac disease presentation and more prevalent in patients with celiac disease than in a population sample used as a control (608 and 3,540, 7.9% and 5.0%, sex and age adjusted odds ratio 4.0, 95% CI 2.7-5.9). Excluding from analysis individuals with baseline urinary stone disease, the incidence of urinary stone disease history was not significantly different between the treated celiac disease (gluten-free diet) and control population (458 and 3,003, 2.4% vs 3.9%). The urine of untreated patients with celiac disease differed from that of healthy volunteers with 120% higher oxalate and 43% lower calcium (in 45 and 45, p : Under untreated conditions (baseline) urinary stone disease was independent of celiac disease presentation and more prevalent in patients with celiac disease than in a population sample used as a control (608 and 3,540, 7.9% and 5.0%, sex and age adjusted odds ratio 4.0, 95% CI 2.7-5.9). Excluding from analysis individuals with baseline urinary stone disease, the incidence of urinary stone disease history was not significantly different between the treated celiac disease (gluten-free diet) and control population (458 and 3,003, 2.4% vs 3.9%). The urine of untreated patients with celiac disease differed from that of healthy volunteers with 120% higher oxalate and 43% lower calcium (in 45 and 45, p **CONCLUSIONS**: Urinary stone disease risk is high in untreated patients with celiac disease independent of overt malabsorption. Hyperoxaluria is likely the underlying disorder. A gluten-free diet reduces urinary stone disease risk and oxaluria.

**URL:** <https://ovidsp.ovid.com/ovidweb.cgi?T=JS&CSC=Y&NEWS=N&PAGE=fulltext&D=med7&DO=10.1016%2fj.juro.2008.05.007> <https://libkey.io/libraries/2592/openurl?genre=article&sid=OVID:medline&id=pmid:18639267&id=doi:10.1016%2Fj.juro.2008.05.007&issn=0022-5347&isbn=&volume=180&issue=3&spage=974&pages=974-9&date=2008&title=Journal+of+Urology&atitle=Urinary+stone+disease+in+adults+with+celiac+disease%3A+prevalence%2C+incidence+and+urinary+determinants.&aulast=Ciacci&pid=%3Cauthor%3ECiacci+C%3BSpagnuolo+G%3BTortora+R%3BBucci+C%3BFranzese+D%3BZingone+F%3BCirillo+M%3C%2Fauthor%3E%3CAN%3E18639267%3C%2FAN%3E%3CDT%3EJournal+Article%3C%2FDT%3E>

109. **Lithogenic risk factors in the morbidly obese population.**

**Item Type:** Journal Article

**Authors:** Duffey, Branden G.;Pedro, Renato N.;Kriedberg, Carly;Weiland, Derek;Melquist, Jonathan;Ikramuddin, Sayeed;Kellogg, Todd;Makhlouf, Antoine A. and Monga, Manoj

**Publication Date:** Ap ,2008

**Journal:** Journal of Urology 179(4), pp. 1401–1406

**Abstract: PURPOSE**: To our knowledge baseline lithogenic risk factors in the morbidly obese population are currently unknown. Prior studies evaluated known stone formers and correlated risk with increasing body mass index. We describe risk factors for urinary stone formation in a group of unselected morbidly obese patients. **MATERIALS AND METHODS**: Patients scheduled for gastric bypass provided a 24-hour urine collection before surgery. Patient demographics, medications and supplement consumption were recorded. A dietary intake diary was converted into daily kcal, Ca, Na and protein consumption. Differences between groups based on gender, history of diabetes or nephrolithiasis, diuretic use and Ca supplementation were evaluated. Correlation of stone risk parameters with body mass index was evaluated. **RESULTS**: A total of 45 patients provided samples for analysis. Mean +/- SD body mass index was 49.5 +/- 9.1 kg/m(2) and mean age was 47.0 +/- 10.5 years. Overall 97.8% of patients had at least 1 lithogenic risk factor identified. Low urinary volume was the most common abnormality, affecting 71.1% of patients. Male patients excreted significantly more Ox (p = 0.0014), Na (p = 0.020), PO(4) (p = 0.0083) and SO(4) (p = 0.0014) than females. Patients with a history of nephrolithiasis excreted significantly more oxalate (p = 0.018) and had higher relative Na urate supersaturation (p = 0.00093) than nonstone formers. Hydrochlorothiazide use was associated with significantly increased Na urate relative supersaturation (p = 0.0097). Increasing body mass index was inversely associated with Mg (r = -0.38, p = 0.01) and brushite (r = -0.30, p = 0.04). **CONCLUSIONS**: Of our cohort of morbidly obese patients 98% had at least 1 lithogenic risk factor identified on 24-hour urine collection. This study identified a high urinary stone risk in the morbidly obese and suggests possible avenues for dietary and/or pharmacological preventive measures. Future studies will determine how bariatric surgery alters these risk factors.

**URL:** <https://ovidsp.ovid.com/ovidweb.cgi?T=JS&CSC=Y&NEWS=N&PAGE=fulltext&D=med7&DO=10.1016%2fj.juro.2007.11.072> <https://libkey.io/libraries/2592/openurl?genre=article&sid=OVID:medline&id=pmid:18289566&id=doi:10.1016%2Fj.juro.2007.11.072&issn=0022-5347&isbn=&volume=179&issue=4&spage=1401&pages=1401-6&date=2008&title=Journal+of+Urology&atitle=Lithogenic+risk+factors+in+the+morbidly+obese+population.&aulast=Duffey&pid=%3Cauthor%3EDuffey+BG%3BPedro+RN%3BKriedberg+C%3BWeiland+D%3BMelquist+J%3BIkramuddin+S%3BKellogg+T%3BMakhlouf+AA%3BMonga+M%3C%2Fauthor%3E%3CAN%3E18289566%3C%2FAN%3E%3CDT%3EJournal+Article%3C%2FDT%3E>

110. **Roux-en-Y gastric bypass is associated with early increased risk factors for development of calcium oxalate nephrolithiasis.**

**Item Type:** Journal Article

**Authors:** Duffey, Branden G.;Pedro, Renato N.;Makhlouf, Antoine;Kriedberg, Carly;Stessman, Michelle;Hinck, Bryan;Ikramuddin, Sayeed;Kellogg, Todd;Slusarek, Bridget and Monga, Manoj

**Publication Date:** Ju ,2008

**Journal:** Journal of the American College of Surgeons 206(6), pp. 1145–1153

**Abstract: BACKGROUND**: Patients treated for obesity with jejunoileal bypass (JIB) experienced a marked increased risk of hyperoxaluria, nephrolithiasis, and oxalate nephropathy developing. Jejunoileal bypass has been abandoned and replaced with other options, including Roux-en-Y gastric bypass (RYGB). Changes in urinary lithogenic risk factors after RYGB are currently unknown. Our purpose was to determine whether RYGB is associated with elevated risk of developing calcium oxalate stone formation through increased urinary oxalate excretion and relative supersaturation of calcium oxalate. **STUDY DESIGN**: A prospective longitudinal cohort study of 24 morbidly obese adults (9 men and 15 women) recruited from a university-based bariatric surgery clinic scheduled to undergo RYGB between December 2005 and April 2007. Patients provided 24-hour urine collections for analysis 7 days before and 90 days after operation. Primary outcomes were changes in 24-hour urinary oxalate excretion and relative supersaturation of calcium oxalate from baseline to 3 months post-RYGB. **RESULTS**: Compared with their baseline, patients undergoing RYGB had increased urinary oxalate excretion (31 +/- 10 mg/d versus 41 +/- 18 mg/d; p = 0.026) and relative supersaturation of calcium oxalate (1.73 +/- 0.81 versus 3.47 +/- 2.59; p = 0.030) 3 months post-RYGB in six patients (25%). De novo hyperoxaluria developed. There were no preoperative patient characteristics predictive of development of de novo hyperoxaluria or the magnitude of change of daily oxalate excretion. **CONCLUSIONS**: This prospective study indicates that RYGB is associated with an earlier increase in urinary oxalate excretion and relative supersaturation of calcium oxalate than previously reported. Additional studies are needed to determine longterm post-RYGB changes in urinary oxalate excretion and identify patients that might be at risk for hyperoxaluria developing.

**URL:** <https://ovidsp.ovid.com/ovidweb.cgi?T=JS&CSC=Y&NEWS=N&PAGE=fulltext&D=med7&DO=10.1016%2fj.jamcollsurg.2008.01.015> <https://libkey.io/libraries/2592/openurl?genre=article&sid=OVID:medline&id=pmid:18501812&id=doi:10.1016%2Fj.jamcollsurg.2008.01.015&issn=1072-7515&isbn=&volume=206&issue=6&spage=1145&pages=1145-53&date=2008&title=Journal+of+the+American+College+of+Surgeons&atitle=Roux-en-Y+gastric+bypass+is+associated+with+early+increased+risk+factors+for+development+of+calcium+oxalate+nephrolithiasis.&aulast=Duffey&pid=%3Cauthor%3EDuffey+BG%3BPedro+RN%3BMakhlouf+A%3BKriedberg+C%3BStessman+M%3BHinck+B%3BIkramuddin+S%3BKellogg+T%3BSlusarek+B%3BMonga+M%3C%2Fauthor%3E%3CAN%3E18501812%3C%2FAN%3E%3CDT%3EClinical+Trial%3C%2FDT%3E>

111. **Impact of obesity in patients with urolithiasis and its prognostic usefulness in stone recurrence.**

**Item Type:** Journal Article

**Authors:** Lee, Sang-Cheol;Kim, Yong-June;Kim, Tae-Hwan;Yun, Seok-Joong;Lee, Nam Kyu and Kim, Wun-Jae

**Publication Date:** Fe ,2008

**Journal:** Journal of Urology 179(2), pp. 570–574

**Abstract: PURPOSE**: Recent studies have suggested an increased prevalence of urolithiasis and recurrence associated with obesity. We assessed the influence of obesity on stone risk factors as well as on stone recurrence. **MATERIALS AND METHODS**: A database of patient history, body mass index, and serum and urine chemistry was analyzed for 704 consecutive stone formers (467 first time stone formers and 247 recurrent stone formers). Obesity was defined as body mass index greater than 25 kg/m(2). The effect of obesity on stone risk factors and recurrence were stratified according to stone episodes. Of these, 163 (23.2%) patients who had been followed for more than 36 months (median 54, range 5 to 148) were included in recurrence analysis. **RESULTS**: Obesity was significantly associated with stone episodes (p = 0.043). Obese stone formers excreted increased amounts of sodium, calcium, uric acid and citrate, while the urinary pH in a 24-hour urine sample was decreased compared to nonobese stone formers (p : Obesity was significantly associated with stone episodes (p = 0.043). Obese stone formers excreted increased amounts of sodium, calcium, uric acid and citrate, while the urinary pH in a 24-hour urine sample was decreased compared to nonobese stone formers (p **CONCLUSIONS**: This study reveals that obesity is associated with metabolic alterations and urinary stone recurrence. Weight control may be considered one of the preventive modalities against recurrent stone formation, especially in first time stone formers.

**URL:** <https://ovidsp.ovid.com/ovidweb.cgi?T=JS&CSC=Y&NEWS=N&PAGE=fulltext&D=med7&DO=10.1016%2fj.juro.2007.09.040> <https://libkey.io/libraries/2592/openurl?genre=article&sid=OVID:medline&id=pmid:18078957&id=doi:10.1016%2Fj.juro.2007.09.040&issn=0022-5347&isbn=&volume=179&issue=2&spage=570&pages=570-4&date=2008&title=Journal+of+Urology&atitle=Impact+of+obesity+in+patients+with+urolithiasis+and+its+prognostic+usefulness+in+stone+recurrence.&aulast=Lee&pid=%3Cauthor%3ELee+SC%3BKim+YJ%3BKim+TH%3BYun+SJ%3BLee+NK%3BKim+WJ%3C%2Fauthor%3E%3CAN%3E18078957%3C%2FAN%3E%3CDT%3EJournal+Article%3C%2FDT%3E>

112. **Induction of progressive profound hypocitraturia with increasing doses of topiramate.**

**Item Type:** Journal Article

**Authors:** Warner, Brad W.;LaGrange, Chad A.;Tucker, Tarvez;Bensalem-Owen, Meriem and Pais, Vernon M. Jr

**Publication Date:** 2008

**Journal:** Urology 72(1), pp. 29–32

**Abstract: OBJECTIVES**: To provide prospective, longitudinal evidence of the effects of topiramate, an antiepileptic medication prescribed for migraine headaches, on stone-risk factors, specifically as pertaining to dosing and rapidity of onset. **METHODS**: Patients scheduled to begin topiramate therapy were recruited to participate in the study. Enrolled subjects collected a pretreatment 24-hour urine specimen with subsequent 24-hour urine specimens collected 5 days after beginning topiramate and after each dose escalation. **RESULTS**: Six subjects enrolled in the study, 4 of whom completed two additional urine collections after initiating topiramate therapy. The pretreatment urine collections of the 4 subjects with additional samples revealed the following mean (range) values: urine volume 1550 (1300 to 1900) mL, pH 6.75 (6 to 7), creatinine 1436.3 (1196 to 1590) mg/day, calcium 305.8 (209 to 423) mg/day, and citrate 606.8 (290 to 860) mg/day. Five days after initiation of topiramate, mean calcium decreased to 211.5 mg/day (31% decrease), and mean citrate decreased to 398 (99 to 804) mg/day, an average decrease of 39.8% (6.5% to 65.9%) per patient. After a dose escalation, calcium increased to 286.8 mg/day, but citrate decreased further to 209 (119 to 353) mg/day, an average decrease of 65.1% (57.9% to 71.7%) per patient from pretreatment levels. **CONCLUSIONS**: Topiramate therapy induces a profound decrease in urinary citrate levels, equivalent to the levels seen in distal renal tubular acidosis. Citrate levels decrease quickly after the start of topiramate therapy and continue to decrease with escalating doses.

**URL:** <https://ovidsp.ovid.com/ovidweb.cgi?T=JS&CSC=Y&NEWS=N&PAGE=fulltext&D=med7&DO=10.1016%2fj.urology.2008.01.042> <https://libkey.io/libraries/2592/openurl?genre=article&sid=OVID:medline&id=pmid:18436283&id=doi:10.1016%2Fj.urology.2008.01.042&issn=0090-4295&isbn=&volume=72&issue=1&spage=29&pages=29-32%3B+discussion+32-3&date=2008&title=Urology&atitle=Induction+of+progressive+profound+hypocitraturia+with+increasing+doses+of+topiramate.&aulast=Warner&pid=%3Cauthor%3EWarner+BW%3BLaGrange+CA%3BTucker+T%3BBensalem-Owen+M%3BPais+VM+Jr%3C%2Fauthor%3E%3CAN%3E18436283%3C%2FAN%3E%3CDT%3EJournal+Article%3C%2FDT%3E>

113. **Effect of dietary control of urinary uric acid excretion in calcium oxalate stone formers and non-stone-forming controls.**

**Item Type:** Journal Article

**Authors:** Pais, Vernon M. Jr;Holmes, Ross P. and Assimos, Dean G.

**Publication Date:** Fe ,2007

**Journal:** Journal of Endourology 21(2), pp. 232–235

**Abstract: BACKGROUND AND PURPOSE**: Hyperuricosuria is a well-recognized risk factor for calcium oxalate urolithiasis. Some studies have demonstrated elevated urinary uric acid excretion in stone formers compared with non-stone-forming controls; nevertheless, these studies were limited by patient consumption of self-selected diets. With the recognition that dietary differences may induce variations in urinary uric acid excretion, we evaluated excretion of this compound in stone formers and controls consuming a standardized diet. **SUBJECTS AND METHODS**: A standardized formula diet was administered to 65 calcium oxalate stone formers and 61 age-matched non-stone-forming controls. During the 3 days of dietary intervention, 24-hour urine collections were obtained. Mean urinary uric acid excretion indexed to urinary creatinine was calculated for each subject, and the results in the two groups were compared. **RESULTS**: Stone-forming subjects did not have an elevation in urinary uric excretion compared with control subjects, with mean indexed urinary uric acid excretions of 337 +/- 64 mg/g of creatinine and 379 +/- 76 mg/g of creatinine, respectively. **CONCLUSIONS**: With dietary standardization, there was no observed increase in urinary uric acid excretion in our sampled populations. These findings emphasize the role of dietary factors in urinary uric acid excretion and highlight the potential value of dietary interventions.

**URL:** <https://ovidsp.ovid.com/ovidweb.cgi?T=JS&CSC=Y&NEWS=N&PAGE=fulltext&D=med6&DO=10.1089%2fend.2007.2218> <https://libkey.io/libraries/2592/openurl?genre=article&sid=OVID:medline&id=pmid:17338626&id=doi:10.1089%2Fend.2007.2218&issn=0892-7790&isbn=&volume=21&issue=2&spage=232&pages=232-5&date=2007&title=Journal+of+Endourology&atitle=Effect+of+dietary+control+of+urinary+uric+acid+excretion+in+calcium+oxalate+stone+formers+and+non-stone-forming+controls.&aulast=Pais&pid=%3Cauthor%3EPais+VM+Jr%3BHolmes+RP%3BAssimos+DG%3C%2Fauthor%3E%3CAN%3E17338626%3C%2FAN%3E%3CDT%3EClinical+Trial%3C%2FDT%3E>

114. **Urinary risk factors for recurrent calcium stone formation in Thai stone formers.**

**Item Type:** Journal Article

**Authors:** Stitchantrakul, Wasana;Kochakarn, Wachira;Ruangraksa, Chatuporn and Domrongkitchaiporn, Somnuek

**Publication Date:** Ap ,2007

**Journal:** Journal of the Medical Association of Thailand 90(4), pp. 688–698

**Abstract: OBJECTIVE**: To survey the urinary risk factors associated with recurrent calcium stone and the contribution of renal tubular acidosis to the prevalence of recurrent calcium stone formation in Thai recurrent stone formers. **MATERIAL AND METHOD**: There were 86 consecutive recurrent calcium stone formers. Three-day dietary record, serum biochemical parameters, first morning urine pH, and two 24-hour urine collections were obtainedfrom each subject. Urinary risk factors for calcium stone formation were determined from the average of the 2-day urine collection. Normal controls were 34 subjects matched for aged, sex, and weight, and without a history of renal stone formation. **RESULTS**: Seven patients (8.1%) were diagnosed as incomplete renal tubular acidosis (iRTA). Among the 79 idiopathic calcium stone formers (ISF), 69.6%, 15.2%, 10.1%, 7.2% and 1.3% of patients were hypocitraturia, hypercalciuria, low urinary volume, hyperuricosuria and hyperoxaluria, respectively. The common combinations of risk factors were hypocitraturia plus low urine output (8.9%) or plus hypercalciuria (7.6%). There were significant differences between ISF and normal controls in urinary oxalate excretion (0.16 +/- 0.01 vs 0.12 +/- 0.01, p : Seven patients (8.1%) were diagnosed as incomplete renal tubular acidosis (iRTA). Among the 79 idiopathic calcium stone formers (ISF), 69.6%, 15.2%, 10.1%, 7.2% and 1.3% of patients were hypocitraturia, hypercalciuria, low urinary volume, hyperuricosuria and hyperoxaluria, respectively. The common combinations of risk factors were hypocitraturia plus low urine output (8.9%) or plus hypercalciuria (7.6%). There were significant differences between ISF and normal controls in urinary oxalate excretion (0.16 +/- 0.01 vs 0.12 +/- 0.01, p : Seven patients (8.1%) were diagnosed as incomplete renal tubular acidosis (iRTA). Among the 79 idiopathic calcium stone formers (ISF), 69.6%, 15.2%, 10.1%, 7.2% and 1.3% of patients were hypocitraturia, hypercalciuria, low urinary volume, hyperuricosuria and hyperoxaluria, respectively. The common combinations of risk factors were hypocitraturia plus low urine output (8.9%) or plus hypercalciuria (7.6%). There were significant differences between ISF and normal controls in urinary oxalate excretion (0.16 +/- 0.01 vs 0.12 +/- 0.01, p : Seven patients (8.1%) were diagnosed as incomplete renal tubular acidosis (iRTA). Among the 79 idiopathic calcium stone formers (ISF), 69.6%, 15.2%, 10.1%, 7.2% and 1.3% of patients were hypocitraturia, hypercalciuria, low urinary volume, hyperuricosuria and hyperoxaluria, respectively. The common combinations of risk factors were hypocitraturia plus low urine output (8.9%) or plus hypercalciuria (7.6%). There were significant differences between ISF and normal controls in urinary oxalate excretion (0.16 +/- 0.01 vs 0.12 +/- 0.01, p : Seven patients (8.1%) were diagnosed as incomplete renal tubular acidosis (iRTA). Among the 79 idiopathic calcium stone formers (ISF), 69.6%, 15.2%, 10.1%, 7.2% and 1.3% of patients were hypocitraturia, hypercalciuria, low urinary volume, hyperuricosuria and hyperoxaluria, respectively. The common combinations of risk factors were hypocitraturia plus low urine output (8.9%) or plus hypercalciuria (7.6%). There were significant differences between ISF and normal controls in urinary oxalate excretion (0.16 +/- 0.01 vs 0.12 +/- 0.01, p : Seven patients (8.1%) were diagnosed as incomplete renal tubular acidosis (iRTA). Among the 79 idiopathic calcium stone formers (ISF), 69.6%, 15.2%, 10.1%, 7.2% and 1.3% of patients were hypocitraturia, hypercalciuria, low urinary volume, hyperuricosuria and hyperoxaluria, respectively. The common combinations of risk factors were hypocitraturia plus low urine output (8.9%) or plus hypercalciuria (7.6%). There were significant differences between ISF and normal controls in urinary oxalate excretion (0.16 +/- 0.01 vs 0.12 +/- 0.01, p : Seven patients (8.1%) were diagnosed as incomplete renal tubular acidosis (iRTA). Among the 79 idiopathic calcium stone formers (ISF), 69.6%, 15.2%, 10.1%, 7.2% and 1.3% of patients were hypocitraturia, hypercalciuria, low urinary volume, hyperuricosuria and hyperoxaluria, respectively. The common combinations of risk factors were hypocitraturia plus low urine output (8.9%) or plus hypercalciuria (7.6%). There were significant differences between ISF and normal controls in urinary oxalate excretion (0.16 +/- 0.01 vs 0.12 +/- 0.01, p : Seven patients (8.1%) were diagnosed as incomplete renal tubular acidosis (iRTA). Among the 79 idiopathic calcium stone formers (ISF), 69.6%, 15.2%, 10.1%, 7.2% and 1.3% of patients were hypocitraturia, hypercalciuria, low urinary volume, hyperuricosuria and hyperoxaluria, respectively. The common combinations of risk factors were hypocitraturia plus low urine output (8.9%) or plus hypercalciuria (7.6%). There were significant differences between ISF and normal controls in urinary oxalate excretion (0.16 +/- 0.01 vs 0.12 +/- 0.01, p **CONCLUSIONS**: Hypocitraturia was the most common urinary risk factor found in Thai recurrent idiopathic calcium stone formers followed by hypercalciuria and low urinary volume. Almost one-fourth of the stone formers had multiple risk factors. Hypocitraturia might result from low potassium and low alkaline intake. iRTA was common among recurrent calcium stone formers. Determination of morning urine pH should be a part of the investigations for urinary risk factors to avoid overlooking the diagnosis of iRTA.

**URL:** <https://ovidsp.ovid.com/ovidweb.cgi?T=JS&CSC=Y&NEWS=N&PAGE=fulltext&D=med6&AN=17487123> <https://libkey.io/libraries/2592/openurl?genre=article&sid=OVID:medline&id=pmid:17487123&id=doi:&issn=0125-2208&isbn=&volume=90&issue=4&spage=688&pages=688-98&date=2007&title=Journal+of+the+Medical+Association+of+Thailand&atitle=Urinary+risk+factors+for+recurrent+calcium+stone+formation+in+Thai+stone+formers.&aulast=Stitchantrakul&pid=%3Cauthor%3EStitchantrakul+W%3BKochakarn+W%3BRuangraksa+C%3BDomrongkitchaiporn+S%3C%2Fauthor%3E%3CAN%3E17487123%3C%2FAN%3E%3CDT%3EJournal+Article%3C%2FDT%3E>

115. **Urinary citrate levels do not correlate with urinary pH in patients with urinary stone formation.**

**Item Type:** Journal Article

**Authors:** Whitson, Jared M.;Cooperberg, Matthew R.;Stackhouse, G. Bennett and Stoller, Marshall L.

**Publication Date:** Oc ,2007

**Journal:** Urology 70(4), pp. 634–637

**Abstract: OBJECTIVES**: Urinary excretion of citrate is dependent on glomerular filtration, tubular reabsorption, and excretion. Acid base status is thought to play a significant role in urinary citrate excretion. It has been assumed that increased urinary citrate will increase urinary pH. The aim of this study was to confirm the association of increased urinary citrate levels with increased urinary pH. **METHODS**: The 24-hour urine collections of all patients with stones referred to our clinic in the past 4 years were reviewed. The samples were collected and analyzed for routine stone risk profiles by a commercial laboratory (Litholink, Chicago, Ill). The Student t test and analysis of variance were used to compare the mean values as applicable. Pearson's correlations were also calculated for each variable. **RESULTS**: A total of 572 patients had at least one 24-hour urine sample from the past 4 years. The mean urinary citrate was 305 mg/day. The mean urinary pH of all patients was 6.14. Statistical evaluation of all patients showed no correlation between urinary citrate and pH (r = -0.04, P = 0.36). In a subset of patients with urinary potassium greater than 100 mEq/day (n = 100), urinary citrate and urinary pH were both increased; however, there was still no correlation between the two (r = 0.011, P = 0.806). **CONCLUSIONS**: Despite the current dogma that increasing urinary citrate increases urinary pH, in a cohort of patients with urinary stone formation who provided 24-hour urine specimens, no correlation was found between urinary citrate and urinary pH levels.

**URL:** <https://ovidsp.ovid.com/ovidweb.cgi?T=JS&CSC=Y&NEWS=N&PAGE=fulltext&D=med6&DO=10.1016%2fj.urology.2007.04.052> <https://libkey.io/libraries/2592/openurl?genre=article&sid=OVID:medline&id=pmid:17991528&id=doi:10.1016%2Fj.urology.2007.04.052&issn=0090-4295&isbn=&volume=70&issue=4&spage=634&pages=634-7&date=2007&title=Urology&atitle=Urinary+citrate+levels+do+not+correlate+with+urinary+pH+in+patients+with+urinary+stone+formation.&aulast=Whitson&pid=%3Cauthor%3EWhitson+JM%3BCooperberg+MR%3BStackhouse+GB%3BStoller+ML%3C%2Fauthor%3E%3CAN%3E17991528%3C%2FAN%3E%3CDT%3EJournal+Article%3C%2FDT%3E>

116. **Urine composition in type 2 diabetes: predisposition to uric acid nephrolithiasis.**

**Item Type:** Journal Article

**Authors:** Cameron, Mary Ann;Maalouf, Naim M.;Adams-Huet, Beverley;Moe, Orson W. and Sakhaee, Khashayar

**Publication Date:** Ma ,2006

**Journal:** Journal of the American Society of Nephrology 17(5), pp. 1422–1428

**Abstract:** Type 2 diabetes is a risk factor for nephrolithiasis in general and has been associated with uric acid stones in particular. The purpose of this study was to identify the metabolic features that place patients with type 2 diabetes at increased risk for uric acid nephrolithiasis. Three groups of individuals were recruited for this outpatient study: patients who have type 2 diabetes and are not stone formers (n = 24), patients who do not have diabetes and are uric acid stone formers (UASF; n = 8), and normal volunteers (NV; n = 59). Participants provided a fasting blood sample and a single 24-h urine collection for stone risk analysis. Twenty-four-hour urine volume and total uric acid did not differ among the three groups. Patients with type 2 diabetes and UASF had lower 24-h urine pH than NV. Urine pH inversely correlated with both body weight and 24-h urine sulphate in all groups. Urine pH remained significantly lower in patients with type 2 diabetes and UASF than NV after adjustment for weight and urine sulphate (P < 0.01). For a given urine sulphate, urine net acid excretion tended to be higher in patients with type 2 diabetes versus NV. With increasing urine sulphate, NV and patients with type 2 diabetes had a similar rise in urine ammonium, whereas in UASF, ammonium excretion remained unchanged. The main risk factor for uric acid nephrolithiasis in patients with type 2 diabetes is a low urine pH. Higher body mass and increased acid intake can contribute to but cannot entirely account for the lower urine pH in patients with type 2 diabetes.

**URL:** <https://ovidsp.ovid.com/ovidweb.cgi?T=JS&CSC=Y&NEWS=N&PAGE=fulltext&D=med6&DO=10.1681%2fASN.2005121246> <https://libkey.io/libraries/2592/openurl?genre=article&sid=OVID:medline&id=pmid:16597681&id=doi:10.1681%2FASN.2005121246&issn=1046-6673&isbn=&volume=17&issue=5&spage=1422&pages=1422-8&date=2006&title=Journal+of+the+American+Society+of+Nephrology&atitle=Urine+composition+in+type+2+diabetes%3A+predisposition+to+uric+acid+nephrolithiasis.&aulast=Cameron&pid=%3Cauthor%3ECameron+MA%3BMaalouf+NM%3BAdams-Huet+B%3BMoe+OW%3BSakhaee+K%3C%2Fauthor%3E%3CAN%3E16597681%3C%2FAN%3E%3CDT%3EJournal+Article%3C%2FDT%3E>

117. **Effect of high and low calcium diets on stone forming risk during liberal oxalate intake.**

**Item Type:** Journal Article

**Authors:** Matsumoto, Edward D.;Heller, Howard J.;Adams-Huet, Beverley;Brinkley, Linda J.;Pak, Charles Y. C. and Pearle, Margaret S.

**Publication Date:** Ju ,2006

**Journal:** Journal of Urology 176(1), pp. 132–136

**Abstract: PURPOSE**: Recent studies suggest that a high calcium diet protects against calcium oxalate stone formation. We compared the effect of high and low calcium diets on urinary saturation of calcium oxalate during liberal oxalate intake. **MATERIALS AND METHODS**: A total of 10 healthy subjects (5 male, 5 female) participated in a 2-phase, randomized, crossover study comparing high (1,000 mg daily) and low (400 mg daily) calcium intake on a liberal oxalate diet (200 mg daily). During each phase subjects adhered to an instructed diet for 3 days followed by a controlled, metabolic diet for 4 days. Blood and 24-hour urine specimens collected on the last 2 days of each phase were analyzed for serum biochemistry studies and stone risk factors, respectively. **RESULTS**: Urinary calcium was higher (mean +/- SD 171 +/- 64 vs 124 +/- 49 mg daily, p = 0.002) and oxalate was lower (25 +/- 4.8 vs 27 +/- 4 mg daily, p = 0.02) on the high vs low calcium diet. Overall, the urinary relative saturation ratio of calcium oxalate was higher on the high compared with the low calcium diet (3.3 vs 2.5, p : Urinary calcium was higher (mean +/- SD 171 +/- 64 vs 124 +/- 49 mg daily, p = 0.002) and oxalate was lower (25 +/- 4.8 vs 27 +/- 4 mg daily, p = 0.02) on the high vs low calcium diet. Overall, the urinary relative saturation ratio of calcium oxalate was higher on the high compared with the low calcium diet (3.3 vs 2.5, p **CONCLUSIONS**: In normal subjects urinary saturation of calcium oxalate was higher on a high calcium diet than a low calcium diet during liberal oxalate intake because the decrease in urinary oxalate did not overcome the effect of increased calcium. A high calcium diet during liberal oxalate intake may pose an increased risk of calcium oxalate stone formation.

**URL:** <https://ovidsp.ovid.com/ovidweb.cgi?T=JS&CSC=Y&NEWS=N&PAGE=fulltext&D=med6&DO=10.1016%2fS0022-5347%2806%2900565-9> <https://libkey.io/libraries/2592/openurl?genre=article&sid=OVID:medline&id=pmid:16753387&id=doi:10.1016%2FS0022-5347%2806%2900565-9&issn=0022-5347&isbn=&volume=176&issue=1&spage=132&pages=132-6&date=2006&title=Journal+of+Urology&atitle=Effect+of+high+and+low+calcium+diets+on+stone+forming+risk+during+liberal+oxalate+intake.&aulast=Matsumoto&pid=%3Cauthor%3EMatsumoto+ED%3BHeller+HJ%3BAdams-Huet+B%3BBrinkley+LJ%3BPak+CY%3BPearle+MS%3C%2Fauthor%3E%3CAN%3E16753387%3C%2FAN%3E%3CDT%3EComparative+Study%3C%2FDT%3E>

118. **Comparative value of orange juice versus lemonade in reducing stone-forming risk.**

**Item Type:** Journal Article

**Authors:** Odvina, Clarita V.

**Publication Date:** No ,2006

**Journal:** Clinical Journal of the American Society of Nephrology: CJASN 1(6), pp. 1269–1274

**Abstract:** Foods that are high in citrate content generally are assumed to deliver alkali load when consumed irrespective of the accompanying cation. The object of this randomized, crossover study was to compare the effects of orange juice with those of lemonade on acid-base profile and urinary stone risks under controlled metabolic conditions. Thirteen volunteers (nine healthy subjects and four stone formers) sequentially received distilled water, orange juice, or lemonade while on constant metabolic diet. Twenty-four-hour urine samples were collected for acid-base parameters and stone risk analysis. Orange juice but not lemonade provided alkali as evidenced by higher net gastrointestinal alkali absorption and higher urinary pH and citrate compared with control. Urinary calcium was not significantly different, but urinary oxalate was higher during the orange juice phase. The calculated supersaturation of calcium oxalate was lower in the orange juice phase compared with control. Calculated undissociated uric acid was lower in the orange juice phase compared with both control and lemonade phases. The calculated supersaturation of brushite was significantly higher in the orange juice phase compared with both control and lemonade phases. Despite comparable citrate content, this study showed that orange juice has greater alkalinizing and citraturic effects than lemonade. Consumption of orange juice was associated with lower calculated calcium oxalate supersaturation and lower calculated undissociated uric acid. This short-term study suggests that orange juice consumption could result in biochemical modification of stone risk factors; however, additional studies are needed to evaluate its role in long-term prevention of recurrent nephrolithiasis.

**URL:** <https://ovidsp.ovid.com/ovidweb.cgi?T=JS&CSC=Y&NEWS=N&PAGE=fulltext&D=med6&DO=10.2215%2fCJN.00800306> <https://libkey.io/libraries/2592/openurl?genre=article&sid=OVID:medline&id=pmid:17699358&id=doi:10.2215%2FCJN.00800306&issn=1555-9041&isbn=&volume=1&issue=6&spage=1269&pages=1269-74&date=2006&title=Clinical+Journal+of+The+American+Society+of+Nephrology%3A+CJASN&atitle=Comparative+value+of+orange+juice+versus+lemonade+in+reducing+stone-forming+risk.&aulast=Odvina&pid=%3Cauthor%3EOdvina+CV%3C%2Fauthor%3E%3CAN%3E17699358%3C%2FAN%3E%3CDT%3EJournal+Article%3C%2FDT%3E>

119. **Medical management of common urinary calculi**

**Item Type:** Journal Article

**Authors:** Pietrow, Paul K. and Karellas, Michael E.

**Publication Date:** Jul 01 ,2006

**Journal:** American Family Physician 74(1), pp. 86–94

**Abstract:** Nephrolithiasis is a common condition affecting nearly 5 percent of U.S. men and women during their lifetimes. Recurrent calculi can be prevented in most patients by the use of a simplified evaluation, reasonable dietary and fluid recommendations, and directed pharmacologic intervention. Serum studies and 24-hour urine collections are the mainstays of metabolic investigation and usually are warranted in patients with recurrent calculi. Although some stones are the result of inherited conditions, most result from a complex interaction between diet, fluid habits, and genetic predisposition. Calcium-sparing diuretics such as thiazides often are used to treat hypercalciuria. Citrate medications increase levels of this naturally occurring stone inhibitor. Allopurinol can be helpful in patients with hyperuricosuria, and urease inhibitors can help break the cycle of infectious calculi. Aggressive fluid intake and moderated intake of salt, calcium, and meat are recommended for most patients. [References: 30]

**URL:** <https://ovidsp.ovid.com/ovidweb.cgi?T=JS&CSC=Y&NEWS=N&PAGE=fulltext&D=med6&AN=16848382> <https://libkey.io/libraries/2592/openurl?genre=article&sid=OVID:medline&id=pmid:16848382&id=doi:&issn=0002-838X&isbn=&volume=74&issue=1&spage=86&pages=86-94&date=2006&title=American+Family+Physician&atitle=Medical+management+of+common+urinary+calculi.&aulast=Pietrow&pid=%3Cauthor%3EPietrow+PK%3BKarellas+ME%3C%2Fauthor%3E%3CAN%3E16848382%3C%2FAN%3E%3CDT%3EJournal+Article%3C%2FDT%3E>

120. **Intestinal oxalate absorption is higher in idiopathic calcium oxalate stone formers than in healthy controls: measurements with the [(13)C2]oxalate absorption test.**

**Item Type:** Journal Article

**Authors:** Voss, Susanne;Hesse, Albrecht;Zimmermann, Diana J.;Sauerbruch, Tilman and von Unruh, Gerd E.

**Publication Date:** Ma ,2006

**Journal:** Journal of Urology 175(5), pp. 1711–1715

**Abstract: PURPOSE**: We assessed the importance of oxalate hyperabsorption for idiopathic calcium oxalate urolithiasis, oxalate absorption in healthy volunteers and recurrent calcium oxalate stone formers was compared. **MATERIALS AND METHODS**: The [(13)C2]oxalate absorption test, a standardized, radioactivity-free test, was performed. On 2 days 24-hour urine was collected and an identical standard diet containing 800 mg Ca daily was maintained. On the morning of day 2 a capsule containing 0.37 mmol sodium [(13)C2]oxalate was ingested. A total of 120 healthy volunteers (60 women and 60 men) and 120 patients (30 women and 90 men) with idiopathic CaOx urolithiasis (60% or greater CaOx) were tested. **RESULTS**: Mean intestinal oxalate absorption in the volunteers was 8.0 +/- 4.4%, and in the patients was 10.2 +/- 5.2% (p : Mean intestinal oxalate absorption in the volunteers was 8.0 +/- 4.4%, and in the patients was 10.2 +/- 5.2% (p : Mean intestinal oxalate absorption in the volunteers was 8.0 +/- 4.4%, and in the patients was 10.2 +/- 5.2% (p : Mean intestinal oxalate absorption in the volunteers was 8.0 +/- 4.4%, and in the patients was 10.2 +/- 5.2% (p **CONCLUSIONS**: In high oxalate absorbers dietary oxalate has a significant role in oxalate excretion and, therefore, increases the risk of calcium oxalate stone formation.

**URL:** <https://ovidsp.ovid.com/ovidweb.cgi?T=JS&CSC=Y&NEWS=N&PAGE=fulltext&D=med6&DO=10.1016%2fS0022-5347%2805%2901001-3> <https://libkey.io/libraries/2592/openurl?genre=article&sid=OVID:medline&id=pmid:16600737&id=doi:10.1016%2FS0022-5347%2805%2901001-3&issn=0022-5347&isbn=&volume=175&issue=5&spage=1711&pages=1711-5&date=2006&title=Journal+of+Urology&atitle=Intestinal+oxalate+absorption+is+higher+in+idiopathic+calcium+oxalate+stone+formers+than+in+healthy+controls%3A+measurements+with+the+%5B%2813%29C2%5Doxalate+absorption+test.&aulast=Voss&pid=%3Cauthor%3EVoss+S%3BHesse+A%3BZimmermann+DJ%3BSauerbruch+T%3Bvon+Unruh+GE%3C%2Fauthor%3E%3CAN%3E16600737%3C%2FAN%3E%3CDT%3EJournal+Article%3C%2FDT%3E>

121. **Metabolic assessment in patients with urinary lithiasis.**

**Item Type:** Journal Article

**Authors:** Amaro, Carmen R.;Goldberg, Jose;Amaro, Joao L. and Padovani, Carlos R.

**Publication Date:** 2005

**Journal:** International Braz J Urol 31(1), pp. 29–33

**Abstract: INTRODUCTION**: Metabolic investigation in patients with urinary lithiasis is very important for preventing recurrence of disease. The objective of this work was to diagnose and to determine the prevalence of metabolic disorders, to assess the quality of the water consumed and volume of diuresis as potential risk factors for this pathology. **PATIENTS AND METHODS**: We studied 182 patients older than 12 years. We included patients with history and/or imaging tests confirming at least 2 stones, with creatinine clearance > or = 60 mL/min and negative urine culture. The protocol consisted in the collection of 2, 24-hour urine samples, for dosing Ca, P, uric acid, Na, K, Mg, Ox and Ci, glycemia and serum levels of Ca, P, Uric acid, Na, K, Cl, Mg, U and Cr, urinary pH and urinary acidification test. **RESULTS**: 158 patients fulfilled the inclusion criteria. Among these, 151 (95.5%) presented metabolic changes, with 94 (62.2%) presenting isolated metabolic change and 57 (37.8%) had mixed changes. The main disorders detected were hypercalciuria (74%), hypocitraturia (37.3%), hyperoxaluria (24.1%), hypomagnesuria (21%), hyperuricosuria (20.2%), primary hyperparathyroidism (1.8%), secondary hyperparathyroidism (0.6%) and renal tubular acidosis (0.6). **CONCLUSION**: Metabolic change was diagnosed in 95.5% of patients. These results warrant the metabolic study and follow-up in patients with recurrent lithiasis in order to decrease the recurrence rate through specific treatments, modification in alimentary and behavioral habits.

**URL:** <https://ovidsp.ovid.com/ovidweb.cgi?T=JS&CSC=Y&NEWS=N&PAGE=fulltext&D=med6&DO=10.1590%2fs1677-55382005000100006> <https://libkey.io/libraries/2592/openurl?genre=article&sid=OVID:medline&id=pmid:15763005&id=doi:10.1590%2Fs1677-55382005000100006&issn=1677-5538&isbn=&volume=31&issue=1&spage=29&pages=29-33&date=2005&title=International+Braz+J+Urol&atitle=Metabolic+assessment+in+patients+with+urinary+lithiasis.&aulast=Amaro&pid=%3Cauthor%3EAmaro+CR%3BGoldberg+J%3BAmaro+JL%3BPadovani+CR%3C%2Fauthor%3E%3CAN%3E15763005%3C%2FAN%3E%3CDT%3EJournal+Article%3C%2FDT%3E>

122. **Results of a prospective trial to compare normal urine supersaturation in children and adults.**

**Item Type:** Journal Article

**Authors:** Defoor, William;Asplin, John;Jackson, Elizabeth;Jackson, Chad;Reddy, Pramod;Sheldon, Curtis and Minevich, Eugene

**Publication Date:** Oc ,2005

**Journal:** Journal of Urology 174(4 Pt 2), pp. 1708–1710

**Abstract: PURPOSE**: Urinary stone disease is relatively rare in children but is often due to metabolic abnormalities. Urinary supersaturation (SS) indices have been suggested as more precise predictors of stone recurrence. We previously reported urinary metabolic evaluations in stone-forming children using adult references. We now assess normal SS values in children. **MATERIAL AND METHODS**: A prospective trial was performed to assess urinary metabolic profiles in children with no personal or family history of urinary calculi. The 24-hour urine collections were performed and evaluated at an outside central laboratory. Urine chemistries were adjusted for urine creatinine. Urine pH and SS for calcium oxalate, calcium phosphate and uric acid were also assessed. The data were compared to a cohort of normal adults. **RESULTS**: A total of 168 samples from 168 adults (58% males) and 51 samples from 30 children (59% males) were evaluated. The mean age of the children was 8.8 years (range 5 to 18). Calcium oxalate SS was similar between children and adults, and urine pH was the major cause of differences seen in the calcium phosphate and uric acid SS indices. Children had a much higher excretion of calcium, oxalate and citrate when adjusted for creatinine. **CONCLUSIONS**: There are significant differences in the normal ranges of urine chemistries in children and adults. An understanding of normal supersaturation values may allow metabolic abnormalities in stone-forming children to be more precisely diagnosed and treated.

**URL:** <https://ovidsp.ovid.com/ovidweb.cgi?T=JS&CSC=Y&NEWS=N&PAGE=fulltext&D=med6&DO=10.1097%2f01.ju.0000175998.64711.45> <https://libkey.io/libraries/2592/openurl?genre=article&sid=OVID:medline&id=pmid:16148687&id=doi:10.1097%2F01.ju.0000175998.64711.45&issn=0022-5347&isbn=&volume=174&issue=4&spage=1708&pages=1708-10&date=2005&title=Journal+of+Urology&atitle=Results+of+a+prospective+trial+to+compare+normal+urine+supersaturation+in+children+and+adults.&aulast=Defoor&pid=%3Cauthor%3EDefoor+W%3BAsplin+J%3BJackson+E%3BJackson+C%3BReddy+P%3BSheldon+C%3BMinevich+E%3C%2Fauthor%3E%3CAN%3E16148687%3C%2FAN%3E%3CDT%3EClinical+Trial%3C%2FDT%3E>

123. **Effect of cranberry juice consumption on urinary stone risk factors.**

**Item Type:** Journal Article

**Authors:** Gettman, Matthew T.;Ogan, Kenneth;Brinkley, Linda J.;Adams-Huet, Beverley;Pak, Charles Y. C. and Pearle, Margaret S.

**Publication Date:** 2005

**Journal:** Journal of Urology 174(2), pp. 590–594

**Abstract: PURPOSE**: We evaluated the effect of cranberry juice on urinary stone risk factors. **MATERIALS AND METHODS**: A total of 12 normal subjects and 12 calcium oxalate stone formers underwent 2, 7-day phases of study in random order while on a controlled metabolic diet. Subjects ingested 1 l of cranberry juice (CBJ) daily in 1 phase and 1 l of deionized water in the other phase. On the last 2 days of each phase 2, 24-hour urine collections and blood samples were obtained for stone risk factors and serum chemistries. **RESULTS**: No significant differences were found between normal subjects and stone formers in response to CBJ and, therefore, the groups were combined. CBJ significantly increased urinary calcium (from 154 to 177 mg per day, p =0.0008) and urinary oxalate (from 26.4 to 29.2 mg per day, p =0.04), thereby increasing urinary saturation of calcium oxalate by 18%. Urinary citrate was unchanged and urinary magnesium increased slightly. Urinary pH decreased (from 5.97 to 5.67, p =0.0005), and urinary ammonium, titratable acidity and net acid excretion increased during CBJ ingestion. Urinary uric acid decreased (from 544 to 442 mg per day, p : No significant differences were found between normal subjects and stone formers in response to CBJ and, therefore, the groups were combined. CBJ significantly increased urinary calcium (from 154 to 177 mg per day, p =0.0008) and urinary oxalate (from 26.4 to 29.2 mg per day, p =0.04), thereby increasing urinary saturation of calcium oxalate by 18%. Urinary citrate was unchanged and urinary magnesium increased slightly. Urinary pH decreased (from 5.97 to 5.67, p =0.0005), and urinary ammonium, titratable acidity and net acid excretion increased during CBJ ingestion. Urinary uric acid decreased (from 544 to 442 mg per day, p **CONCLUSIONS**: CBJ exerts a mixed effect on urinary stone forming propensity. It reduces urinary pH likely by providing an acid load and decreases urinary uric acid perhaps by retarding urate synthesis. Overall CBJ increases the risk of calcium oxalate and uric acid stone formation but decreases the risk of brushite stones.

**URL:** <https://ovidsp.ovid.com/ovidweb.cgi?T=JS&CSC=Y&NEWS=N&PAGE=fulltext&D=med6&DO=10.1097%2f01.ju.0000165168.68054.f8> <https://libkey.io/libraries/2592/openurl?genre=article&sid=OVID:medline&id=pmid:16006907&id=doi:10.1097%2F01.ju.0000165168.68054.f8&issn=0022-5347&isbn=&volume=174&issue=2&spage=590&pages=590-4%3B+quiz+801&date=2005&title=Journal+of+Urology&atitle=Effect+of+cranberry+juice+consumption+on+urinary+stone+risk+factors.&aulast=Gettman&pid=%3Cauthor%3EGettman+MT%3BOgan+K%3BBrinkley+LJ%3BAdams-Huet+B%3BPak+CY%3BPearle+MS%3C%2Fauthor%3E%3CAN%3E16006907%3C%2FAN%3E%3CDT%3EClinical+Trial%3C%2FDT%3E>

124. **Increased frequency of ultrasonographic findings suggestive of renal stones in patients with ankylosing spondylitis.**

**Item Type:** Journal Article

**Authors:** Korkmaz, C.;Ozcan, A. and Akcar, N.

**Publication Date:** 2005

**Journal:** Clinical & Experimental Rheumatology 23(3), pp. 389–392

**Abstract: OBJECTIVE**: To determine the frequency of hypercalciuria and renal stones in ankylosing spondylitis (AS) sufferers. **METHODS**: This study involved 83 consecutive AS patients (21 female, 62 male; mean age 36.7 yr), 72 consecutive Behcet's disease (BD) patients (29 female, 43 male; mean age 37.7 yr) as disease control and 92 healthy control (HC) (26 female, 66 male; mean age 32.9 yr.) Twenty-four hour urine analyses for urinary calcium and uric acid levels were performed in each patient. Likewise, blood samples for erythrocyte sedimentation rate (ESR), C-reactive protein (CRP), parathyroid hormone (PTH), calcium and uric acid evaluation were taken. Renal ultrasonography to evaluate the presence of renal stone was performed in patients with AS, as well as in the BD patients and HC individuals after a fasting period of 12 hours. **RESULTS**: 20 of the 80 (25%) patients with AS were diagnosed with renal stones. Only 4/72 (5.5%) BD patients, and 3/98 (3.3%) HC individuals had renal stones by ultrasonography. AS patients had a significantly higher frequency of renal stones compared with BD (p : 20 of the 80 (25%) patients with AS were diagnosed with renal stones. Only 4/72 (5.5%) BD patients, and 3/98 (3.3%) HC individuals had renal stones by ultrasonography. AS patients had a significantly higher frequency of renal stones compared with BD (p : 20 of the 80 (25%) patients with AS were diagnosed with renal stones. Only 4/72 (5.5%) BD patients, and 3/98 (3.3%) HC individuals had renal stones by ultrasonography. AS patients had a significantly higher frequency of renal stones compared with BD (p : 20 of the 80 (25%) patients with AS were diagnosed with renal stones. Only 4/72 (5.5%) BD patients, and 3/98 (3.3%) HC individuals had renal stones by ultrasonography. AS patients had a significantly higher frequency of renal stones compared with BD (p **CONCLUSION**: Renal stone prevalence was found to increase in AS patients. The likelihood of renal stone formation was also found to increase with the extension of the disease duration of AS sufferers.

**URL:** <https://ovidsp.ovid.com/ovidweb.cgi?T=JS&CSC=Y&NEWS=N&PAGE=fulltext&D=med6&AN=15971429> <https://libkey.io/libraries/2592/openurl?genre=article&sid=OVID:medline&id=pmid:15971429&id=doi:&issn=0392-856X&isbn=&volume=23&issue=3&spage=389&pages=389-92&date=2005&title=Clinical+%26+Experimental+Rheumatology&atitle=Increased+frequency+of+ultrasonographic+findings+suggestive+of+renal+stones+in+patients+with+ankylosing+spondylitis.&aulast=Korkmaz&pid=%3Cauthor%3EKorkmaz+C%3BOzcan+A%3BAkcar+N%3C%2Fauthor%3E%3CAN%3E15971429%3C%2FAN%3E%3CDT%3EJournal+Article%3C%2FDT%3E>

125. **Use of a probiotic to decrease enteric hyperoxaluria.**

**Item Type:** Journal Article

**Authors:** Lieske, John C.;Goldfarb, David S.;De Simone, Claudio and Regnier, Cynthia

**Publication Date:** Se ,2005

**Journal:** Kidney International 68(3), pp. 1244–1249

**Abstract: BACKGROUND**: Patients with inflammatory bowel disease have a 10- to 100-fold increased risk of nephrolithiasis, with enteric hyperoxaluria being the major risk factor for these and other patients with fat malabsorptive states. Endogenous components of the intestinal microflora can potentially limit dietary oxalate absorption. **METHODS**: Ten patients were studied with chronic fat malabsorption, calcium oxalate stones, and hyperoxaluria thought to be caused by jejunoileal bypass (1) and Roux-en-Y gastric bypass surgery for obesity (4), dumping syndrome secondary to gastrectomy (2), celiac sprue (1), chronic pancreatitis (1), and ulcerative colitis in remission (1). For 3 months, patients received increasing doses of a lactic acid bacteria mixture (Oxadrop), VSL Pharmaceuticals), followed by a washout month. Twenty-four-hour urine collections were performed at baseline and after each month. **RESULTS**: Mean urinary oxalate excretion fell by 19% after 1 month (1 dose per day, P : Mean urinary oxalate excretion fell by 19% after 1 month (1 dose per day, P : Mean urinary oxalate excretion fell by 19% after 1 month (1 dose per day, P **CONCLUSION**: Manipulation of gastrointestinal (GI) flora can influence urinary oxalate excretion to reduce urinary supersaturation levels. These changes could have a salutary effect on stone formation rates. Further studies will be needed to establish the optimal dosing regimen.

**URL:** <https://ovidsp.ovid.com/ovidweb.cgi?T=JS&CSC=Y&NEWS=N&PAGE=fulltext&D=med6&DO=10.1111%2fj.1523-1755.2005.00520.x> <https://libkey.io/libraries/2592/openurl?genre=article&sid=OVID:medline&id=pmid:16105057&id=doi:10.1111%2Fj.1523-1755.2005.00520.x&issn=0085-2538&isbn=&volume=68&issue=3&spage=1244&pages=1244-9&date=2005&title=Kidney+International&atitle=Use+of+a+probiotic+to+decrease+enteric+hyperoxaluria.&aulast=Lieske&pid=%3Cauthor%3ELieske+JC%3BGoldfarb+DS%3BDe+Simone+C%3BRegnier+C%3C%2Fauthor%3E%3CAN%3E16105057%3C%2FAN%3E%3CDT%3EClinical+Trial%3C%2FDT%3E>

126. **Segregation of urine calcium excretion in families ascertained for nephrolithiasis: evidence for a major gene.**

**Item Type:** Journal Article

**Authors:** Loredo-Osti, J. C.;Roslin, Nicole M.;Tessier, Josee;Fujiwara, T. Mary;Morgan, Kenneth and Bonnardeaux, Alain

**Publication Date:** Se ,2005

**Journal:** Kidney International 68(3), pp. 966–971

**Abstract: BACKGROUND**: The quantitative genetics of urine calcium excretion has not been established. It is a trait of interest because hypercalciuria is commonly found in subjects with nephrolithiasis. The aim of this study was to model the segregation of this trait in a sample of French-Canadian families ascertained through a stone former. **METHODS**: Major gene, polygenic, and mixed models were fit to 24-hour urine calcium excretion from 567 individuals in 221 nuclear families, while simultaneously taking into account gender, age at examination, body mass index (BMI), and the use of thiazide drugs. The nuclear families were extracted from 154 pedigrees, some of which were four generations, with at least two siblings with a history of calcium stones. **RESULTS**: All the proposed genetic models fit the data significantly better than the null model. The most parsimonious model was the mixed codominant/polygenic model but it was statistically indistinguishable from the single-gene codominant model. In both of these models the heritability attributable to the major gene was estimated to be 0.58. **CONCLUSION**: Our results suggest that a major gene with a relatively large effect on variation in urine calcium excretion is segregating in French-Canadian families with stone formers. This implies that the power of quantitative trait segregation analysis of urine calcium excretion may be increased in these families, and results indicate that it should be feasible to genetically map the quantitative trait locus.

**URL:** <https://ovidsp.ovid.com/ovidweb.cgi?T=JS&CSC=Y&NEWS=N&PAGE=fulltext&D=med6&DO=10.1111%2fj.1523-1755.2005.00490.x> <https://libkey.io/libraries/2592/openurl?genre=article&sid=OVID:medline&id=pmid:16105027&id=doi:10.1111%2Fj.1523-1755.2005.00490.x&issn=0085-2538&isbn=&volume=68&issue=3&spage=966&pages=966-71&date=2005&title=Kidney+International&atitle=Segregation+of+urine+calcium+excretion+in+families+ascertained+for+nephrolithiasis%3A+evidence+for+a+major+gene.&aulast=Loredo-Osti&pid=%3Cauthor%3ELoredo-Osti+JC%3BRoslin+NM%3BTessier+J%3BFujiwara+TM%3BMorgan+K%3BBonnardeaux+A%3C%2Fauthor%3E%3CAN%3E16105027%3C%2FAN%3E%3CDT%3EJournal+Article%3C%2FDT%3E>

127. **Enteric hyperoxaluria, nephrolithiasis, and oxalate nephropathy: potentially serious and unappreciated complications of Roux-en-Y gastric bypass.**

**Item Type:** Journal Article

**Authors:** Nelson, Wayne K.;Houghton, Scott G.;Milliner, Dawn S.;Lieske, John C. and Sarr, Michael G.

**Publication Date:** 2005

**Journal:** Surgery for Obesity & Related Diseases 1(5), pp. 481–485

**Abstract: BACKGROUND**: Neither the presence nor prevalence of enteric hyperoxaluria has been recognized after Roux-en-Y gastric bypass (RYGBP). We have noted a high rate of oxalate nephrolithiasis and even 2 patients with oxalate nephropathy in this patient population postoperatively. Our aim was to determine the frequency of the occurrence and effects of enteric hyperoxaluria after RYGBP. **METHODS**: Retrospective review of all patients at our institution diagnosed with calcium oxalate nephrolithiasis or oxalate nephropathy after standard (n = 14) or distal (n = 9) RYGBP. The mean postoperative follow-up was 55 months. **RESULTS**: A total of 23 patients (14 men and 9 women; mean age 45 years; mean preoperative body mass index 55 kg/m(2)) developed enteric hyperoxaluria after RYGBP, defined by the presence of oxalate nephropathy (n = 2) or calcium oxalate nephrolithiasis (n = 21) and increased 24-hour excretion of urinary oxalate and/or calcium oxalate supersaturation. Enteric hyperoxaluria was recognized after a mean weight loss of 46 kg at 29 months (range 2-85) after RYGBP. Two patients developed renal failure and required chronic hemodialysis. Of the 21 patients with nephrolithiasis, 14 had no history of nephrolithiasis preoperatively, and 19 of 21 required lithotripsy or other intervention. Of the 23 patients, 20 tested had increased oxalate excretion, and 14 of 15 tested had high urine calcium oxalate supersaturation. **CONCLUSION**: Enteric hyperoxaluria, nephrolithiasis, and oxalate nephropathy must be considered with the other risks of RYGBP. Efforts should be made to identify factors that predispose patients to developing hyperoxaluria.

**URL:** <https://ovidsp.ovid.com/ovidweb.cgi?T=JS&CSC=Y&NEWS=N&PAGE=fulltext&D=med6&DO=10.1016%2fj.soard.2005.07.002> <https://libkey.io/libraries/2592/openurl?genre=article&sid=OVID:medline&id=pmid:16925274&id=doi:10.1016%2Fj.soard.2005.07.002&issn=1550-7289&isbn=&volume=1&issue=5&spage=481&pages=481-5&date=2005&title=Surgery+for+Obesity+%26+Related+Diseases&atitle=Enteric+hyperoxaluria%2C+nephrolithiasis%2C+and+oxalate+nephropathy%3A+potentially+serious+and+unappreciated+complications+of+Roux-en-Y+gastric+bypass.&aulast=Nelson&pid=%3Cauthor%3ENelson+WK%3BHoughton+SG%3BMilliner+DS%3BLieske+JC%3BSarr+MG%3C%2Fauthor%3E%3CAN%3E16925274%3C%2FAN%3E%3CDT%3EJournal+Article%3C%2FDT%3E>

128. **Effect of dietary modification on urinary stone risk factors.**

**Item Type:** Journal Article

**Authors:** Pak, Charles Y. C.;Odvina, Clarita V.;Pearle, Margaret S.;Sakhaee, Khashayar;Peterson, Roy D.;Poindexter, John R. and Brinkley, Linda J.

**Publication Date:** No ,2005

**Journal:** Kidney International 68(5), pp. 2264–2273

**Abstract: BACKGROUND**: This study was undertaken to ascertain the effect of dietary modification on urinary stone risks, and to determine whether the response depends on the prevailing urinary calcium. **METHODS**: A retrospective data analysis was conducted from our stone registry involving 951 patients with calcareous stones undergoing ambulatory evaluation, whereby 24-hour urine samples were collected during random diet and after dietary modification composed of restriction of calcium, oxalate, sodium, and meat products. Samples were analyzed for stone risk factors. Urinary calcium was also obtained after overnight fast and following a 1 g-calcium load. Changes produced by dietary modification from the random diet were evaluated in 356 patients with moderate-severe hypercalciuria (>6.88 mmol/day, group I), 243 patients with mild hypercalciuria (5.00-6.88 mmol/day, group II), and 352 with normocalciuria (: A retrospective data analysis was conducted from our stone registry involving 951 patients with calcareous stones undergoing ambulatory evaluation, whereby 24-hour urine samples were collected during random diet and after dietary modification composed of restriction of calcium, oxalate, sodium, and meat products. Samples were analyzed for stone risk factors. Urinary calcium was also obtained after overnight fast and following a 1 g-calcium load. Changes produced by dietary modification from the random diet were evaluated in 356 patients with moderate-severe hypercalciuria (>6.88 mmol/day, group I), 243 patients with mild hypercalciuria (5.00-6.88 mmol/day, group II), and 352 with normocalciuria (**RESULTS**: Urinary calcium postcalcium load and the percentage of patients with absorptive hypercalciuria type I were highest in group I, intermediate in group II, and lowest in group III. During dietary modification, urinary calcium declined by 29% in group I, 19% in group II, and 10% in group III. Urinary oxalate did not change. Urinary saturation of calcium oxalate declined by only 12% in group I, 6% in group II, and nonsignificantly in group III, owing to various physicochemical changes in urinary biochemistry, which attenuated the effect of the decline in urinary calcium. Urinary saturation of brushite declined in all 3 groups due to the fall in urinary calcium, phosphorus, and pH. This reduction was more marked in the hypercalciuric groups than in the normocalciuric group. Urinary saturation of monosodium urate also decreased from a decline in urinary sodium and uric acid. **CONCLUSION**: Secondary rise in urinary oxalate occurring from calcium restriction can be avoided by concurrent dietary oxalate restriction. Dietary modification (restriction of dietary calcium, oxalate, sodium, and meat products) is more useful in reducing urinary saturation of calcium oxalate among patients with hypercalciuria than among those with normocalciuria.

**URL:** <https://ovidsp.ovid.com/ovidweb.cgi?T=JS&CSC=Y&NEWS=N&PAGE=fulltext&D=med6&DO=10.1111%2fj.1523-1755.2005.00685.x> <https://libkey.io/libraries/2592/openurl?genre=article&sid=OVID:medline&id=pmid:16221228&id=doi:10.1111%2Fj.1523-1755.2005.00685.x&issn=0085-2538&isbn=&volume=68&issue=5&spage=2264&pages=2264-73&date=2005&title=Kidney+International&atitle=Effect+of+dietary+modification+on+urinary+stone+risk+factors.&aulast=Pak&pid=%3Cauthor%3EPak+CY%3BOdvina+CV%3BPearle+MS%3BSakhaee+K%3BPeterson+RD%3BPoindexter+JR%3BBrinkley+LJ%3C%2Fauthor%3E%3CAN%3E16221228%3C%2FAN%3E%3CDT%3EJournal+Article%3C%2FDT%3E>

129. **Genetic and dietary factors in urinary citrate excretion.**

**Item Type:** Journal Article

**Authors:** Shah, Ojas;Assimos, Dean G. and Holmes, Ross P.

**Publication Date:** Ma ,2005

**Journal:** Journal of Endourology 19(2), pp. 177–182

**Abstract: BACKGROUND AND PURPOSE**: Hypocitraturia, an important risk factor for calcium oxalate nephrolithiasis, is the result of numerous factors. We studied citrate excretion by patients with and without stones consuming normal and controlled formula diets. **SUBJECTS AND METHODS**: Subjects with and without a history of calcium oxalate stones (N = 101 per group) provided two or three 24-hour urine specimens during consumption of self-selected diets. Data also were collected on subsets of subjects consuming formula (Ensure) diets. Citrate was determined using the citrate lyase method of Petrarulo and associates, and values for multiple specimens were averaged. The data were adjusted for creatinine excretion and examined on a per-day basis. **RESULTS**: The mean citrate excretion of the non-stone formers was slightly but not significantly higher than that of the stone formers (442 +/- 217 versus 378 +/- 153 mg/g of creatinine). All statistical analyses revealed highly significant differences between, but not within, individuals, a result compatible with a genetic influence. In the normal population, 5% of subjects had a citrate excretion : The mean citrate excretion of the non-stone formers was slightly but not significantly higher than that of the stone formers (442 +/- 217 versus 378 +/- 153 mg/g of creatinine). All statistical analyses revealed highly significant differences between, but not within, individuals, a result compatible with a genetic influence. In the normal population, 5% of subjects had a citrate excretion : The mean citrate excretion of the non-stone formers was slightly but not significantly higher than that of the stone formers (442 +/- 217 versus 378 +/- 153 mg/g of creatinine). All statistical analyses revealed highly significant differences between, but not within, individuals, a result compatible with a genetic influence. In the normal population, 5% of subjects had a citrate excretion **CONCLUSION**: These findings imply a genetic influence on citrate excretion, as has already been demonstrated for calcium excretion. Further studies of genetic influences on calcium oxalate stone formation are warranted.

**URL:** <https://ovidsp.ovid.com/ovidweb.cgi?T=JS&CSC=Y&NEWS=N&PAGE=fulltext&D=med6&DO=10.1089%2fend.2005.19.177> <https://libkey.io/libraries/2592/openurl?genre=article&sid=OVID:medline&id=pmid:15798414&id=doi:10.1089%2Fend.2005.19.177&issn=0892-7790&isbn=&volume=19&issue=2&spage=177&pages=177-82&date=2005&title=Journal+of+Endourology&atitle=Genetic+and+dietary+factors+in+urinary+citrate+excretion.&aulast=Shah&pid=%3Cauthor%3EShah+O%3BAssimos+DG%3BHolmes+RP%3C%2Fauthor%3E%3CAN%3E15798414%3C%2FAN%3E%3CDT%3EJournal+Article%3C%2FDT%3E>

130. **The efficacy of dietary intervention on urinary risk factors for stone formation in recurrent calcium oxalate stone patients.**

**Item Type:** Journal Article

**Authors:** Siener, Roswitha;Schade, Natalie;Nicolay, Claudia;von Unruh, Gerd E. and Hesse, Albrecht

**Publication Date:** Ma ,2005

**Journal:** Journal of Urology 173(5), pp. 1601–1605

**Abstract: PURPOSE**: Nutrition is suggested to be the major environmental risk factor in idiopathic calcium oxalate stone disease. The study was designed to evaluate the effect of dietary intervention on urinary risk factors for recurrence in calcium oxalate stone formers. **MATERIALS AND METHODS**: A total of 76 men and 31 women with idiopathic calcium oxalate stone disease collected 24-hour urine on their habitual, self-selected diets and after 7 days on a balanced standardized diet according to the recommendations for calcium oxalate stone formers. **RESULTS**: On the usual diet, a urine volume of less than 2.0 l per 24 hours was present in 57.9%, hypercalciuria in 25.2%, hypomagnesuria in 18.7%, hyperoxaluria in 14.0%, hyperuricosuria in 41.3% and hypocitraturia in 57.0% of patients. The frequency of metabolic abnormalities and the risk of calcium oxalate stone formation decreased significantly on the ingestion of the balanced diet, due to the significant increase in urinary volume, pH and citrate excretion and the significant decrease in urinary calcium and uric acid excretion. No change occurred in urinary oxalate and magnesium excretion. **CONCLUSIONS**: The evaluation of urinary risk profiles of the patients on their usual dietary habits revealed a high risk for calcium oxalate stone formation. A low fluid intake and an increased intake of protein and alcohol were identified as the most important dietary risk factors. The shift to a nutritionally balanced diet according to the recommendations for calcium oxalate stone formers significantly reduced the stone forming potential.

**URL:** <https://ovidsp.ovid.com/ovidweb.cgi?T=JS&CSC=Y&NEWS=N&PAGE=fulltext&D=med6&DO=10.1097%2f01.ju.0000154626.16349.d3> <https://libkey.io/libraries/2592/openurl?genre=article&sid=OVID:medline&id=pmid:15821507&id=doi:10.1097%2F01.ju.0000154626.16349.d3&issn=0022-5347&isbn=&volume=173&issue=5&spage=1601&pages=1601-5&date=2005&title=Journal+of+Urology&atitle=The+efficacy+of+dietary+intervention+on+urinary+risk+factors+for+stone+formation+in+recurrent+calcium+oxalate+stone+patients.&aulast=Siener&pid=%3Cauthor%3ESiener+R%3BSchade+N%3BNicolay+C%3Bvon+Unruh+GE%3BHesse+A%3C%2Fauthor%3E%3CAN%3E15821507%3C%2FAN%3E%3CDT%3EJournal+Article%3C%2FDT%3E>

131. **Pediatric stone disease: an evolving experience.**

**Item Type:** Journal Article

**Authors:** Sternberg, Kevan;Greenfield, Saul P.;Williot, Pierre and Wan, Julian

**Publication Date:** 2005

**Journal:** Journal of Urology 174(4 Pt 2), pp. 1711–1714

**Abstract: PURPOSE**: The presentation and management of pediatric stone disease have changed due to early identification and treatment of congenital urological conditions, as well as technological advances. Therefore, we reviewed our 12-year experience. **MATERIALS AND METHODS**: A total of 123 patients with 158 stones presented from 1991 to 2003. The 46 boys and 77 girls were 8 months to 25 years old, including 5 boys (4%) younger than 3 years, 46 (37%) 4 to 12 years old and 72 (59%) older than 13 years. A 24-hour urine collection was done in 50 patients (41%) and serum chemistry studies were performed in 66 (54%). **RESULTS**: The seasons of presentation were fall in 41% of cases, summer in 24%, spring in 22% and winter in 13%. Of the patients 94 (76%) had loin pain, 10 (8%) had urinary tract infections, 13 (11%) had a history urinary tract infection, 18 (15%) had gross hematuria, 14 (11%) had structural urological abnormalities and 7 (6%) had neurogenic bladder. Metabolic abnormalities were uncommon and included hypercalciuria in 12%, hyperoxaluria in 2% and cystinuria in 2%. A total of 57 patients (46%) passed the stones and 34 (28%) underwent extracorporeal shock wave lithotripsy, of whom 24 (71%) became stone-free. Ureteroscopy was performed in 10 patients (8%) 6 to 19 years old, percutaneous nephrostolithotomy was done in 4 (3%) and 4 (3%) underwent open surgery. Stone analysis showed calcium based in 88% of the cases, struvite in 7% and cystine in 5%. **CONCLUSIONS**: The majority of patients had no congenital abnormalities. Early diagnosis of urological abnormalities and urinary infection, and appropriate management of neurogenic bladder may have reduced the incidence in those groups. Most stones are calcium based but occur in the absence of metabolic disturbances. More patients presented in the fall, perhaps reflecting the increased concentration of urine in the summer. Half of the patients passed the stones and shock wave lithotripsy was curative in most others. Ureteroscopy, percutaneous nephrostolithotomy and open surgery were rarely required.

**URL:** <https://ovidsp.ovid.com/ovidweb.cgi?T=JS&CSC=Y&NEWS=N&PAGE=fulltext&D=med6&DO=10.1097%2f01.ju.0000179537.36472.59> <https://libkey.io/libraries/2592/openurl?genre=article&sid=OVID:medline&id=pmid:16148688&id=doi:10.1097%2F01.ju.0000179537.36472.59&issn=0022-5347&isbn=&volume=174&issue=4&spage=1711&pages=1711-4%3B+discussion+1714&date=2005&title=Journal+of+Urology&atitle=Pediatric+stone+disease%3A+an+evolving+experience.&aulast=Sternberg&pid=%3Cauthor%3ESternberg+K%3BGreenfield+SP%3BWilliot+P%3BWan+J%3C%2Fauthor%3E%3CAN%3E16148688%3C%2FAN%3E%3CDT%3EJournal+Article%3C%2FDT%3E>

132. **Is 24-hour urinary calcium a surrogate marker for dietary calcium intake?.**

**Item Type:** Journal Article

**Authors:** Toren, Paul J. and Norman, Richard W.

**Publication Date:** Ma ,2005

**Journal:** Urology 65(3), pp. 459–462

**Abstract: OBJECTIVES**: To determine whether the 24-hour urinary excretion of calcium was a reliable surrogate marker for dietary calcium intake. Although dietary calcium intake has been negatively correlated with the risk of recurrent calcium-based stones, detailed dietary histories are not routinely evaluated in most patients with recurrent stone formation. **METHODS**: The dietary records and corresponding 24-hour urine collections of 68 randomly selected women with a history of calcium-based renal stones and two or more outpatient clinic visits were studied. Subjects were excluded if they had conditions or took medications affecting calcium absorption or excretion. Multivariate regression analysis was performed on the most recent set of data per patient with 24-hour urinary calcium as the dependent variable. Independent variables included age, weight, and dietary calcium, sodium, potassium, magnesium, fiber, and animal protein. Regression analysis was performed on the differences between the first and last visits for dietary and urinary data. Using tertiles, the positive predictive value of 24-hour urinary calcium for the respective dietary intake tertiles was calculated. **RESULTS**: The regression model on absolute values showed all dietary parameters to have squared partial correlation coefficients of less than 0.3 (P = 0.015, R2 = 0.264). In the second regression analysis, the model did not significantly explain the variance (P = 0.656). The positive predictive value of a mean 24-hour urinary calcium level less than 3.75 mmol/L for calcium intake less than 585 mg/day was 45%. **CONCLUSIONS**: The results of the present study revealed that the 24-hour urinary calcium cannot be used as a surrogate marker for dietary intake of calcium. A detailed dietary history is needed for all women with recurrent stone formation for proper assessment and potential modification of calcium intake to reduce recurrences.

**URL:** <https://ovidsp.ovid.com/ovidweb.cgi?T=JS&CSC=Y&NEWS=N&PAGE=fulltext&D=med6&DO=10.1016%2fj.urology.2004.10.025> <https://libkey.io/libraries/2592/openurl?genre=article&sid=OVID:medline&id=pmid:15780355&id=doi:10.1016%2Fj.urology.2004.10.025&issn=0090-4295&isbn=&volume=65&issue=3&spage=459&pages=459-62&date=2005&title=Urology&atitle=Is+24-hour+urinary+calcium+a+surrogate+marker+for+dietary+calcium+intake%3F.&aulast=Toren&pid=%3Cauthor%3EToren+PJ%3BNorman+RW%3C%2Fauthor%3E%3CAN%3E15780355%3C%2FAN%3E%3CDT%3EJournal+Article%3C%2FDT%3E>

133. **The metabolic syndrome and uric acid nephrolithiasis: novel features of renal manifestation of insulin resistance.**

**Item Type:** Journal Article

**Authors:** Abate, Nicola;Chandalia, Manisha;Cabo-Chan, Alberto V. Jr;Moe, Orson W. and Sakhaee, Khashayar

**Publication Date:** Fe ,2004

**Journal:** Kidney International 65(2), pp. 386–392

**Abstract: BACKGROUND**: Uric acid nephrolithiasis primarily results from low urinary pH, which increases the concentration of the insoluble undissociated uric acid, causing formation of both uric acid and mixed uric acid/calcium oxalate stones. These patients have recently been described as exhibiting features of insulin resistance. This study was designed to evaluate if insulin resistance is associated with excessively low urinary pH in overtly healthy volunteers (non-stone formers) and if insulin resistance may explain the excessively low urinary pH in patients with uric acid nephrolithiasis. **METHODS**: Fifty-five healthy volunteers (non stone-formers) with a large range of body mass index and 13 patients with recurrent uric acid nephrolithiasis underwent hyperinsulinemic euglycemic clamp, 24-hour urinary studies, and anthropometric measurements of adiposity. A subgroup of 35 non-stone formers had 2-hour timed urinary collection before and during the hyperinsulinemic phase of the clamp studies. **RESULTS**: For the non-stone former population, low insulin sensitivity measured as glucose disposal rate significantly correlated with low 24-hour urinary pH (r= 0. 35; P= 0.01). In addition to the previously described acidic urine pH and hypouricosuria, patients with recurrent uric acid nephrolithiasis were found to be severely insulin resistant (glucose disposal rate: uric acid stone-formers vs. normals; 4.1 +/- 1.3 vs. 6.9 +/- 2.1 mg/min/kg of lean body mass, P= 0.008). Acute hyperinsulinemia was associated with higher urinary pH (6.1 +/- 0.7 at baseline to 6.8 +/- 0.7 during hyperinsulinemia; P : For the non-stone former population, low insulin sensitivity measured as glucose disposal rate significantly correlated with low 24-hour urinary pH (r= 0. 35; P= 0.01). In addition to the previously described acidic urine pH and hypouricosuria, patients with recurrent uric acid nephrolithiasis were found to be severely insulin resistant (glucose disposal rate: uric acid stone-formers vs. normals; 4.1 +/- 1.3 vs. 6.9 +/- 2.1 mg/min/kg of lean body mass, P= 0.008). Acute hyperinsulinemia was associated with higher urinary pH (6.1 +/- 0.7 at baseline to 6.8 +/- 0.7 during hyperinsulinemia; P : For the non-stone former population, low insulin sensitivity measured as glucose disposal rate significantly correlated with low 24-hour urinary pH (r= 0. 35; P= 0.01). In addition to the previously described acidic urine pH and hypouricosuria, patients with recurrent uric acid nephrolithiasis were found to be severely insulin resistant (glucose disposal rate: uric acid stone-formers vs. normals; 4.1 +/- 1.3 vs. 6.9 +/- 2.1 mg/min/kg of lean body mass, P= 0.008). Acute hyperinsulinemia was associated with higher urinary pH (6.1 +/- 0.7 at baseline to 6.8 +/- 0.7 during hyperinsulinemia; P **CONCLUSION**: We conclude that one renal manifestation of insulin resistance may be low urinary ammonium and pH. This defect can result in increased risk of uric acid precipitation despite normouricosuria.

**URL:** <https://ovidsp.ovid.com/ovidweb.cgi?T=JS&CSC=Y&NEWS=N&PAGE=fulltext&D=med5&DO=10.1111%2fj.1523-1755.2004.00386.x> <https://libkey.io/libraries/2592/openurl?genre=article&sid=OVID:medline&id=pmid:14717908&id=doi:10.1111%2Fj.1523-1755.2004.00386.x&issn=0085-2538&isbn=&volume=65&issue=2&spage=386&pages=386-92&date=2004&title=Kidney+International&atitle=The+metabolic+syndrome+and+uric+acid+nephrolithiasis%3A+novel+features+of+renal+manifestation+of+insulin+resistance.&aulast=Abate&pid=%3Cauthor%3EAbate+N%3BChandalia+M%3BCabo-Chan+AV+Jr%3BMoe+OW%3BSakhaee+K%3C%2Fauthor%3E%3CAN%3E14717908%3C%2FAN%3E%3CDT%3EJournal+Article%3C%2FDT%3E>

134. **Assessment of oxalate absorption from almonds and black beans with and without the use of an extrinsic label.**

**Item Type:** Journal Article

**Authors:** Chai, Weiwen and Liebman, Michael

**Publication Date:** Se ,2004

**Journal:** Journal of Urology 172(3), pp. 953–957

**Abstract: PURPOSE**: Oxalate bioavailability is an important determinant of whether the consumption of a particular food is a high risk in individuals predisposed to kidney stones. We estimated and compared oxalate absorption from a high oxalate containing legume (black beans) and a high oxalate containing nut (almonds). We also compared an isotope method using extrinsically labeled oxalate and an oxalate load method to assess oxalate absorption. **MATERIALS AND METHODS**: Six male and 5 female subjects participated in the 4 oxalate load tests, namely almonds, almonds with 20 mg C2-oxalic acid, black beans and black beans with 20 mg C2-oxalic acid. Each treatment provided a total of 120 mg oxalate, after which timed urine samples were collected for the analysis of oxalate, calcium and creatinine. **RESULTS**: Average oxalate absorption from the 2 almond treatments (5.9%) using the oxalate load method was significantly higher than that from the 2 black bean treatments (1.8%) during the 24-hour post-oxalate load collection period. In contrast, C2-oxalic acid absorption from the almond (7.9%) and black bean (8.6%) treatments did not significantly differ. **CONCLUSIONS**: The higher oxalate absorption from almonds than from black beans suggests that the relative amount of soluble and insoluble oxalate in food has an important role in the determination of oxalate absorption. Since extrinsically provided C2-oxalate and oxalate naturally occurring in the high oxalate test foods appeared to be differentially absorbed, the data do not support the use of extrinsically labeled oxalate to assess food oxalate absorption.

**URL:** <https://ovidsp.ovid.com/ovidweb.cgi?T=JS&CSC=Y&NEWS=N&PAGE=fulltext&D=med5&DO=10.1097%2f01.ju.0000135918.00761.8a> <https://libkey.io/libraries/2592/openurl?genre=article&sid=OVID:medline&id=pmid:15311007&id=doi:10.1097%2F01.ju.0000135918.00761.8a&issn=0022-5347&isbn=&volume=172&issue=3&spage=953&pages=953-7&date=2004&title=Journal+of+Urology&atitle=Assessment+of+oxalate+absorption+from+almonds+and+black+beans+with+and+without+the+use+of+an+extrinsic+label.&aulast=Chai&pid=%3Cauthor%3EChai+W%3BLiebman+M%3C%2Fauthor%3E%3CAN%3E15311007%3C%2FAN%3E%3CDT%3EJournal+Article%3C%2FDT%3E>

135. **Determination of the calcium oxalate crystallization risk from urine samples: the BONN Risk Index in comparison to other risk formulas.**

**Item Type:** Journal Article

**Authors:** Laube, Norbert;Hergarten, Stefan;Hoppe, Bernd;Schmidt, Matthias and Hesse, Albrecht

**Publication Date:** Ju ,2004

**Journal:** Journal of Urology 172(1), pp. 355–359

**Abstract: PURPOSE**: Regular risk evaluation and risk monitoring during stone therapy are recommended measures to ensure reduction of recurrence of crystal formation. This strategy optimizes the patient treatment by a more individual approach and decreases expensive over treatment. We evaluated the BONN Risk Index (BRI) through data actualization and evaluation refinement. The BRI was compared with the most common methods of risk evaluation, namely calculation of relative urinary calcium oxalate (CaOx) supersaturation (RS) and of the urine activity product (AP)CaOx index to estimate the urine AP with respect to CaOx. **MATERIALS AND METHODS**: A total of 201, 12 and 24-hour urine samples were collected from 95 healthy volunteers and from 106 CaOx stone formers. Crystallization experiments following the BRI method were performed. RS and APCaOx were calculated from urinalysis. Data were indexed and individually grouped into 8 classes, and frequency distributions were plotted. A calculation scheme for the BRI based estimation of the statistical probability of a clinically healthy person being a (still nondetected) CaOx stone former is provided. **RESULTS**: Logarithmically arranged BRI groups from healthy subject and patient data showed Gaussian frequency distributions. Compared with RS and APCaOx BRI allowed optimum distinction between healthy subjects and stone formers. The healthy subject probability of already being a CaOx stone former strongly increased with increasing BRI. **CONCLUSIONS**: The BRI for evaluating CaOx crystallization risk allows reliable distinction between healthy subjects and CaOx stone formers. Although RS and APCaOx require much more analytical efforts for determination, their results do not show higher reliability.

**URL:** <https://ovidsp.ovid.com/ovidweb.cgi?T=JS&CSC=Y&NEWS=N&PAGE=fulltext&D=med5&DO=10.1097%2f01.ju.0000123822.20291.4d> <https://libkey.io/libraries/2592/openurl?genre=article&sid=OVID:medline&id=pmid:15201810&id=doi:10.1097%2F01.ju.0000123822.20291.4d&issn=0022-5347&isbn=&volume=172&issue=1&spage=355&pages=355-9&date=2004&title=Journal+of+Urology&atitle=Determination+of+the+calcium+oxalate+crystallization+risk+from+urine+samples%3A+the+BONN+Risk+Index+in+comparison+to+other+risk+formulas.&aulast=Laube&pid=%3Cauthor%3ELaube+N%3BHergarten+S%3BHoppe+B%3BSchmidt+M%3BHesse+A%3C%2Fauthor%3E%3CAN%3E15201810%3C%2FAN%3E%3CDT%3EComparative+Study%3C%2FDT%3E>

136. **Renal response to lithogenic and anti-lithogenic supplement challenges in a stone-free population group.**

**Item Type:** Journal Article

**Authors:** Lewandowski, Sonja and Rodgers, Allen L.

**Publication Date:** Ju ,2004

**Journal:** Journal of Renal Nutrition 14(3), pp. 170–179

**Abstract: OBJECTIVE**: In South Africa, urolithiasis is extremely rare in the black population, but is common in the white population. The objective of this study was to investigate the individual effects of 5 different dietary and supplemental challenges (high dietary calcium, calcium supplement, vitamin B6 supplement, L-glutamine supplement, and L-cysteine supplement) on the urinary risk factors for calcium oxalate urolithiasis in subjects from both race groups. **DESIGN**: Complete Latin Square design. **SETTING**: University research laboratory. **SUBJECTS**: Subjects were recruited from the student cohort of the University of Cape Town (10 male subjects from each race group). Selection criteria were no history of renal or metabolic diseases, and no chronic or acute medication. Subjects served as their own controls. **INTERVENTION**: After 7 days on a self-selected standardized diet, a 24-hour baseline urine sample was collected. A second 24-hour urine sample was collected after 5 days on the prescribed dietary or supplemental challenge. These were analyzed for biochemical and physicochemical risk factors. Additionally, 24-hour dietary recall questionnaires were recorded at baseline and after the 5-day test period, and were analyzed using a food analysis program. Statistical analysis of variance was performed on all of the data. **MAIN OUTCOME MEASURES**: Urine composition, relative supersaturation of urinary salts, calcium oxalate metastable limit, and Tiselius risk index. **RESULTS**: None of the protocols altered any of the urinary biochemical or physicochemical risk factors in black subjects. In white subjects, the calcium diet significantly increased urinary potassium (P =.0001) and decreased the relative supersaturation of brushite (P =.035); the calcium supplement significantly decreased the Tiselius risk index (P =.014); vitamin B6 supplement significantly decreased urinary calcium (P =.016), urinary phosphate (P =.027), and the relative supersaturation of brushite (P =.004); L-glutamine supplement significantly decreased relative supersaturation of calcium oxalate (P =.01); L-cystine supplement significantly decreased urinary calcium (P =.031) and the Tiselius risk index (P =.013). **CONCLUSIONS**: Because none of the challenges had an effect on the urinary risk factors in black subjects, it is speculated that a renal or gastrointestinal homeostatic adjustment occurs in this group, thereby keeping urinary concentration of substances in balance.

**URL:** <https://ovidsp.ovid.com/ovidweb.cgi?T=JS&CSC=Y&NEWS=N&PAGE=fulltext&D=med5&DO=10.1053%2fj.jrn.2004.04.007> <https://libkey.io/libraries/2592/openurl?genre=article&sid=OVID:medline&id=pmid:15232796&id=doi:10.1053%2Fj.jrn.2004.04.007&issn=1051-2276&isbn=&volume=14&issue=3&spage=170&pages=170-9&date=2004&title=Journal+of+Renal+Nutrition&atitle=Renal+response+to+lithogenic+and+anti-lithogenic+supplement+challenges+in+a+stone-free+population+group.&aulast=Lewandowski&pid=%3Cauthor%3ELewandowski+S%3BRodgers+AL%3C%2Fauthor%3E%3CAN%3E15232796%3C%2FAN%3E%3CDT%3EClinical+Trial%3C%2FDT%3E>

137. **Association of urinary pH with body weight in nephrolithiasis.**

**Item Type:** Journal Article

**Authors:** Maalouf, Naim M.;Sakhaee, Khashayar;Parks, Joan H.;Coe, Frederic L.;Adams-Huet, Beverley and Pak, Charles Y. C.

**Publication Date:** Ap ,2004

**Journal:** Kidney International 65(4), pp. 1422–1425

**Abstract: BACKGROUND**: The prevalence of kidney stone disease in the United States is progressively increasing, paralleling the growing rate of obesity. Uric acid nephrolithiasis, a condition associated with a low urinary pH, has been linked to obesity and insulin resistance. Based on these observations, we hypothesized that urinary pH may be inversely associated to body weight in nephrolithiasis. **METHODS**: Data were retrieved from 4883 patients with nephrolithiasis who underwent ambulatory evaluation at two established stone clinics in Dallas and Chicago. The patients collected 24-hour urine samples on an outpatient basis, while avoiding any drug that could alter urinary pH. Patients were divided in increasing sextiles of body weight, and urinary pH was adjusted for urinary creatinine and for age. **RESULTS**: Urinary pH had a strong, graded inverse association with body weight. Urinary creatinine and age were both found to be significant covariates of urinary pH, while gender was not a significant independent variable after adjustment for urinary creatinine. Mean 24-hour urinary pH, adjusted for age and urinary creatinine, were 6.09, 6.04, 6.01, 5.99, 5.97, and 5.91 for sextiles of body weight in increasing order from Dallas (P for linear trend : Urinary pH had a strong, graded inverse association with body weight. Urinary creatinine and age were both found to be significant covariates of urinary pH, while gender was not a significant independent variable after adjustment for urinary creatinine. Mean 24-hour urinary pH, adjusted for age and urinary creatinine, were 6.09, 6.04, 6.01, 5.99, 5.97, and 5.91 for sextiles of body weight in increasing order from Dallas (P for linear trend : Urinary pH had a strong, graded inverse association with body weight. Urinary creatinine and age were both found to be significant covariates of urinary pH, while gender was not a significant independent variable after adjustment for urinary creatinine. Mean 24-hour urinary pH, adjusted for age and urinary creatinine, were 6.09, 6.04, 6.01, 5.99, 5.97, and 5.91 for sextiles of body weight in increasing order from Dallas (P for linear trend **CONCLUSION**: We conclude that urinary pH is inversely related to body weight among patients with stones. The results confirm the previously proposed scheme that obesity may sometimes cause uric acid nephrolithiasis by producing excessively acid urine due to insulin resistance.

**URL:** <https://ovidsp.ovid.com/ovidweb.cgi?T=JS&CSC=Y&NEWS=N&PAGE=fulltext&D=med5&DO=10.1111%2fj.1523-1755.2004.00522.x> <https://libkey.io/libraries/2592/openurl?genre=article&sid=OVID:medline&id=pmid:15086484&id=doi:10.1111%2Fj.1523-1755.2004.00522.x&issn=0085-2538&isbn=&volume=65&issue=4&spage=1422&pages=1422-5&date=2004&title=Kidney+International&atitle=Association+of+urinary+pH+with+body+weight+in+nephrolithiasis.&aulast=Maalouf&pid=%3Cauthor%3EMaalouf+NM%3BSakhaee+K%3BParks+JH%3BCoe+FL%3BAdams-Huet+B%3BPak+CY%3C%2Fauthor%3E%3CAN%3E15086484%3C%2FAN%3E%3CDT%3EJournal+Article%3C%2FDT%3E>

138. **Screening for biochemical abnormalities in urolithiasis patients.**

**Item Type:** Journal Article

**Authors:** Orakzai, Nasir;Hanbury, Damian C. and Farrington, Kenneth

**Publication Date:** 2004

**Journal:** Journal of Ayub Medical College, Abbottabad: JAMC 16(2), pp. 60–63

**Abstract: BACKGROUND**: The significance of biochemical screening in stone formers has been a debated topic. This study was conducted to investigate the frequency of biochemical abnormalities in our urolithiasis patients and to compare the abnormality between the first time and recurrent stone formers so that this information would help in assessing the value of biochemical screening in our practice. **METHODS**: Over a twenty-one month period, new and recurrent stone disease patients had one random blood specimen and two random 24-hour urine collections analysed for biochemical abnormalities. Serum was checked for calcium, urate, phosphate and creatinine. The urines were measured for volumes, calcium, oxalate, urate, citrate, cystine and pH. **RESULTS**: Out of total of 113 patients, 83 (73%) had some urinary or blood abnormality. Highest number of abnormalities were in urine. Low volume 33 (39.76%), hypercalciuria 33 (39.76%) and hyperoxaluria 20 (24.1%) were the main urinary abnormalities. Elevated serum creatinine in 10 (12.05%) was commonest blood abnormality. Females had significantly higher frequencies of low urinary volume (48% vs 21%, p=0.001), hyperoxaluria (38% vs 11%, p=0.002) and hypocitraturia (37% vs 0%, p: Out of total of 113 patients, 83 (73%) had some urinary or blood abnormality. Highest number of abnormalities were in urine. Low volume 33 (39.76%), hypercalciuria 33 (39.76%) and hyperoxaluria 20 (24.1%) were the main urinary abnormalities. Elevated serum creatinine in 10 (12.05%) was commonest blood abnormality. Females had significantly higher frequencies of low urinary volume (48% vs 21%, p=0.001), hyperoxaluria (38% vs 11%, p=0.002) and hypocitraturia (37% vs 0%, p**CONCLUSION**: A high frequency of urinary biochemical abnormality and equal abnormality frequencies among first time and recurrent stone formers highlights the significance of biochemical screening even in cases of initial stone presentation. We feel such diagnostic evaluation would help in providing precise treatment and efficient prophylaxis.

**URL:** <https://ovidsp.ovid.com/ovidweb.cgi?T=JS&CSC=Y&NEWS=N&PAGE=fulltext&D=med5&AN=15455621> <https://libkey.io/libraries/2592/openurl?genre=article&sid=OVID:medline&id=pmid:15455621&id=doi:&issn=1025-9589&isbn=&volume=16&issue=2&spage=60&pages=60-3&date=2004&title=Journal+of+Ayub+Medical+College%2C+Abbottabad%3A+JAMC&atitle=Screening+for+biochemical+abnormalities+in+urolithiasis+patients.&aulast=Orakzai&pid=%3Cauthor%3EOrakzai+N%3BHanbury+DC%3BFarrington+K%3C%2Fauthor%3E%3CAN%3E15455621%3C%2FAN%3E%3CDT%3EJournal+Article%3C%2FDT%3E>

139. **Metabolic abnormalities associated with renal calculi in patients with horseshoe kidneys.**

**Item Type:** Journal Article

**Authors:** Raj, Ganesh V.;Auge, Brian K.;Assimos, Dean and Preminger, Glenn M.

**Publication Date:** Ma ,2004

**Journal:** Journal of Endourology 18(2), pp. 157–161

**Abstract: BACKGROUND AND PURPOSE**: Horseshoe kidneys are a complex anatomic variant of fused kidneys, with a 20% reported incidence of associated calculi. Anatomic causes such as high insertion of the ureter on the renal pelvis and obstruction of the ureteropelvic junction are thought to contribute to stone formation via impaired drainage, with urinary stasis, and an increased incidence of infection. In this multi-institutional study, we evaluated whether metabolic factors contributed to stone development in patients with horseshoe kidneys. **PATIENTS AND METHODS**: A retrospective review of 37 patients with horseshoe kidneys was performed to determine if these patients had metabolic derangements that might have contributed to calculus formation. Stone compositions as well as 24-hour urine collections were examined. Specific data points of interest were total urine volume; urine pH; urine concentrations of calcium, sodium, uric acid, oxalate, and citrate; and number of abnormalities per patient per 24-hour urine collection. These data were compared with those of a group of 13 patients with stones in caliceal diverticula as well as 24 age-, race-, and sex-matched controls with stones in anatomically normal kidneys. **RESULTS**: Eleven (9 men and 2 women) of the 37 patients (30%) with renal calculi in horseshoe kidneys had complete metabolic evaluations available for review. All patients were noted to have at least one abnormality, with an average of 2.68 abnormalities per 24-hour urine collection (range 1-4). One patient had primary hyperparathyroidism and underwent a parathyroidectomy. Low urine volumes were noted in eight patients on at least one of the two specimens (range 350-1640 mL/day). Hypercalciuria, hyperoxaluria, hyperuricosuria, and hypocitraturia were noted in seven, three, six, and six patients, respectively. No patients were found to have gouty diathesis or developed cystine stones. Comparative metabolic analyses of patients with renal calculi in caliceal diverticula or normal kidneys revealed a distinct profile in patients with horseshoe kidneys, with a higher incidence of hypocitraturia. **CONCLUSIONS**: All patients with renal calculi in horseshoe kidneys were noted to have metabolic abnormalities predisposing to stone formation. In this initial series of 11 patients, hypovolemia, hypercalcuria and hypocitraturia were most common metabolic defects. These findings suggest that metabolic derangements play a role in stone formation in patients with a horseshoe kidney. Patients with calculi in anatomically abnormal kidneys should be considered for a metabolic evaluation to identify their stone-forming risk factors in order to initiate preventative selective medical therapy and reduce the risk of recurrent calculus formation.

**URL:** <https://ovidsp.ovid.com/ovidweb.cgi?T=JS&CSC=Y&NEWS=N&PAGE=fulltext&D=med5&DO=10.1089%2f089277904322959798> <https://libkey.io/libraries/2592/openurl?genre=article&sid=OVID:medline&id=pmid:15072623&id=doi:10.1089%2F089277904322959798&issn=0892-7790&isbn=&volume=18&issue=2&spage=157&pages=157-61&date=2004&title=Journal+of+Endourology&atitle=Metabolic+abnormalities+associated+with+renal+calculi+in+patients+with+horseshoe+kidneys.&aulast=Raj&pid=%3Cauthor%3ERaj+GV%3BAuge+BK%3BAssimos+D%3BPreminger+GM%3C%2Fauthor%3E%3CAN%3E15072623%3C%2FAN%3E%3CDT%3EJournal+Article%3C%2FDT%3E>

140. **Effects of calcium supplements on the risk of renal stone formation in a population with low oxalate intake.**

**Item Type:** Journal Article

**Authors:** Stitchantrakul, Wasana;Sopassathit, Wichai;Prapaipanich, Surasing and Domrongkitchaiporn, Somnuek

**Publication Date:** De ,2004

**Journal:** Southeast Asian Journal of Tropical Medicine & Public Health 35(4), pp. 1028–1033

**Abstract:** It has been speculated that calcium supplement in subjects with low oxalate intake might increase the risk of calcium stone formation due to an increase in calcium absorption without a significant reduction in oxalate absorption. There have been no human studies addressing specifically the effects of taking calcium supplements in populations whose dietary oxalate is low. This study was conducted to determine the effects of calcium supplements on the risk of calcium stone formation in a population with low oxalate intake. Thirty-two healthy male navy privates, 22.7 +/- 1.9 (mean +/- SD) years old, who had oxalate intake of less than 1 mmol/day, a serum creatinine of less than 150 micromol/l, and no history of renal stones, participated in the study. Dietary oxalate was controlled to be under 1 mmol/day throughout the study. Twenty-four hour urine collections for the determination of urinary constituents were obtained at baseline and after taking calcium supplements. Detection of calcium oxalate was performed to assess the risk of calcium oxalate stone formation. The urinary excretion of calcium was significantly elevated above baseline values while taking the calcium supplements (3.48 +/- 2.13 vs 5.17 +/- 2.61 mmol/d, p < 0.05) and urinary oxalate was significantly decreased when the subjects took calcium supplements compared to the corresponding baseline value (0.13 +/- 0.05 vs 0.17 +/- 0.07 mmol/d, p = 0.01). Urinary citrate was significantly elevated when the subjects took calcium supplements compared to the baseline (0.83 +/- 0.57 vs 0.64 +/- 0.39 mmol/d, p = 0.03). There was no significant alteration in the activity products of calcium oxalate while taking the calcium supplements (0.54 +/- 0.25 vs 0.57 +/- 0.22, p = 0.54). The effect of calcium supplements with meals, for the reduction of the risk of calcium stone formation, was unchanged, even in a population whose oxalate intake is rather low. Taking calcium supplements resulted in a reduction in urinary oxalates and an elevation in urinary citrates. Both alterations in urinary constituents counterbalanced the elevation in urinary calcium which resulted from the calcium supplements.

**URL:** <https://ovidsp.ovid.com/ovidweb.cgi?T=JS&CSC=Y&NEWS=N&PAGE=fulltext&D=med5&AN=15916110> <https://libkey.io/libraries/2592/openurl?genre=article&sid=OVID:medline&id=pmid:15916110&id=doi:&issn=0125-1562&isbn=&volume=35&issue=4&spage=1028&pages=1028-33&date=2004&title=Southeast+Asian+Journal+of+Tropical+Medicine+%26+Public+Health&atitle=Effects+of+calcium+supplements+on+the+risk+of+renal+stone+formation+in+a+population+with+low+oxalate+intake.&aulast=Stitchantrakul&pid=%3Cauthor%3EStitchantrakul+W%3BSopassathit+W%3BPrapaipanich+S%3BDomrongkitchaiporn+S%3C%2Fauthor%3E%3CAN%3E15916110%3C%2FAN%3E%3CDT%3EClinical+Trial%3C%2FDT%3E>

141. **Prediction of upper urinary tract calculi using an artificial neural network.**

**Item Type:** Journal Article

**Authors:** Tanthanuch, Monthira and Tanthanuch, Sawit

**Publication Date:** Ma ,2004

**Journal:** Journal of the Medical Association of Thailand 87(5), pp. 515–518

**Abstract: OBJECTIVES**: To evaluate the possibility of using an artificial neural network (ANN) in upper urinary tract calculi prediction. **MATERIAL AND METHOD**: Data of 168 upper urinary tract calculi patients treated in the Division of Urology, Department of Surgery, Songklanagarind Hospital from January 1997 to December 2000 were reviewed and classified into 6 catagories and 20 characteristics. 100 items were used in training and 68 in testing for an ANN designed with 3 layers: 20 nodes for an input layer, 5 nodes for a hidden layer and a node for the output. **RESULTS**: Output data between 0-0.38 indicate free of calculi, 0.65-1 indicate prone to have calculi, 0.38-0.65 indicate probable calculi and further need investigation. **CONCLUSION**: An ANN with error back-propagation training can be used in diagnosing the presence of upper urinary tract calculi. The accuracy of prediction depends on a previous history of calculi, nephrocalcinosis, 24 hour urine assay for citrate and urine culture.

**URL:** <https://ovidsp.ovid.com/ovidweb.cgi?T=JS&CSC=Y&NEWS=N&PAGE=fulltext&D=med5&AN=15222521> <https://libkey.io/libraries/2592/openurl?genre=article&sid=OVID:medline&id=pmid:15222521&id=doi:&issn=0125-2208&isbn=&volume=87&issue=5&spage=515&pages=515-8&date=2004&title=Journal+of+the+Medical+Association+of+Thailand&atitle=Prediction+of+upper+urinary+tract+calculi+using+an+artificial+neural+network.&aulast=Tanthanuch&pid=%3Cauthor%3ETanthanuch+M%3BTanthanuch+S%3C%2Fauthor%3E%3CAN%3E15222521%3C%2FAN%3E%3CDT%3EEvaluation+Study%3C%2FDT%3E>

142. **Effects of calcium carbonate, magnesium oxide and sodium citrate bicarbonate health supplements on the urinary risk factors for kidney stone formation.**

**Item Type:** Journal Article

**Authors:** Allie, Shameez and Rodgers, Allen

**Publication Date:** Ja ,2003

**Journal:** Clinical Chemistry & Laboratory Medicine 41(1), pp. 39–45

**Abstract:** We describe a model to illustrate different chemical interactions that can occur in urine following ingestion of individual and combined health supplements. Two types of interactions are defined: synergism and addition. The model was applied to eight healthy males who participated in a study to investigate the chemical interactions between calcium carbonate, magnesium oxide and sodium citrate-bicarbonate health supplements on calcium oxalate urinary stone risk factors. Subjects ingested these components individually and in combination for 7 days. Twenty-four-hour urines were collected at baseline and during the final day of supplementation. These were analysed using standard laboratory techniques. Three different chemical interactions, all involving citrate, were identified: magnesium and citrate exerted a synergistic effect on lowering the relative superaturation (RS) of brushite; the same two components produced a synergistic effect on raising pH; finally, calcium and citrate exerted an additive effect on lowering the RS of uric acid. We propose that the novel approach described in this paper allows for the evaluation of individual, additive and synergistic interactions in the assessment of the efficacy of stone-risk reducing preparations.

**URL:** <https://ovidsp.ovid.com/ovidweb.cgi?T=JS&CSC=Y&NEWS=N&PAGE=fulltext&D=med5&DO=10.1515%2fCCLM.2003.008> <https://libkey.io/libraries/2592/openurl?genre=article&sid=OVID:medline&id=pmid:12636048&id=doi:10.1515%2FCCLM.2003.008&issn=1434-6621&isbn=&volume=41&issue=1&spage=39&pages=39-45&date=2003&title=Clinical+Chemistry+%26+Laboratory+Medicine&atitle=Effects+of+calcium+carbonate%2C+magnesium+oxide+and+sodium+citrate+bicarbonate+health+supplements+on+the+urinary+risk+factors+for+kidney+stone+formation.&aulast=Allie&pid=%3Cauthor%3EAllie+S%3BRodgers+A%3C%2Fauthor%3E%3CAN%3E12636048%3C%2FAN%3E%3CDT%3EComparative+Study%3C%2FDT%3E>

143. **Effect of vitamin C supplements on urinary oxalate and pH in calcium stone-forming patients.**

**Item Type:** Journal Article

**Authors:** Baxmann, Alessandra Calabria;De O G Mendonca, Claudia and Heilberg, Ita Pfeferman

**Publication Date:** Ma ,2003

**Journal:** Kidney International 63(3), pp. 1066–1071

**Abstract: BACKGROUND**: The contribution of ascorbate to urinary oxalate is controversial. The present study aimed to determine whether urinary oxalate and pH may be affected by vitamin C supplementation in calcium stone-forming patients. **METHODS**: Forty-seven adult calcium stone-forming patients received either 1 g (N=23) or 2 g (N=24) of vitamin C supplement for 3 days and 20 healthy subjects received 1 g. A 24-hour urine sample was obtained both before and after vitamin C for calcium, oxalate, magnesium, citrate, sodium, potassium, and creatinine determination. The Tiselius index was used as a calcium oxalate crystallization index. A spot fasting morning urine sample was also obtained to determine the urinary pH before and after vitamin C. **RESULTS**: Fasting urinary pH did not change after 1 g (5.8 +/- 0.6 vs. 5.8 +/- 0.7) or 2 g vitamin C (5.8 +/- 0.8 vs. 5.8 +/- 0.7). A significant increase in mean urinary oxalate was observed in calcium stone-forming patients receiving either 1 g (50 +/- 16 vs. 31 +/- 12 mg/24 hours) or 2 g (48 +/- 21 vs. 34 +/- 12 mg/24 hours) of vitamin C and in healthy subjects (25 +/- 12 vs. 39 +/- 13 mg/24 hours). A significant increase in mean Tiselius index was observed in calcium stone-forming patients after 1 g (1.43 +/- 0.70 vs. 0.92 +/- 0.65) or 2 g vitamin C (1.61 +/- 1.05 vs. 0.99 +/- 0.55) and in healthy subjects (1.50 +/- 0.69 vs. 0.91 +/- 0.46). Ancillary analyses of spot urine obtained after vitamin C were performed in 15 control subjects in vessels with or without ethylenediaminetetraacetic acid (EDTA) with no difference in urinary oxalate between them (28 +/- 23 vs. 26 +/- 21 mg/L), suggesting that the in vitro conversion of ascorbate to oxalate did not occur. **CONCLUSION**: These data suggest that vitamin C supplementation may increase urinary oxalate excretion and the risk of calcium oxalate crystallization in calcium stone-forming patients.

**URL:** <https://ovidsp.ovid.com/ovidweb.cgi?T=JS&CSC=Y&NEWS=N&PAGE=fulltext&D=med5&DO=10.1046%2fj.1523-1755.2003.00815.x> <https://libkey.io/libraries/2592/openurl?genre=article&sid=OVID:medline&id=pmid:12631089&id=doi:10.1046%2Fj.1523-1755.2003.00815.x&issn=0085-2538&isbn=&volume=63&issue=3&spage=1066&pages=1066-71&date=2003&title=Kidney+International&atitle=Effect+of+vitamin+C+supplements+on+urinary+oxalate+and+pH+in+calcium+stone-forming+patients.&aulast=Baxmann&pid=%3Cauthor%3EBaxmann+AC%3BDe+O+G+Mendonca+C%3BHeilberg+IP%3C%2Fauthor%3E%3CAN%3E12631089%3C%2FAN%3E%3CDT%3EClinical+Trial%3C%2FDT%3E>

144. **Lipid peroxidation and its correlations with urinary levels of oxalate, citric acid, and osteopontin in patients with renal calcium oxalate stones.**

**Item Type:** Journal Article

**Authors:** Huang, Ho-Shiang;Ma, Ming-Chieh;Chen, Chau-Fong and Chen, Jun

**Publication Date:** De ,2003

**Journal:** Urology 62(6), pp. 1123–1128

**Abstract: OBJECTIVES**: To determine whether lipid peroxidation plays a role in patients with calcium oxalate kidney stones and to determine the correlation of lipid peroxidation with tubular damage and the major urinary risk factors. We also used the isoenzymes of glutathione S-transferase (GST) to examine which parts of the renal tubules were injured in patients with renal stones. **METHODS**: This clinical study included two study groups. Group 1 included 32 normal volunteers, and group 2 included 32 patients with calcium oxalate kidney stones. A 24-hour urine sample was collected from each subject, and the levels of Ca, P, Mg, oxalate, citrate, N-acetyl-beta-glucosaminidase (NAG), beta-galactosidase (GAL), alphaGST, piGST, osteopontin (OPN), thiobarbituric acid-reactive substances (TBARS), and malondialdehyde (MDA) were examined. **RESULTS**: Hyperoxaluria, hypocitraturia, and low urinary OPN were the major abnormalities found in the patients with stones. Elevated urinary alphaGST, NAG, and GAL were also noted in the patients with stones; however, urinary piGST showed no statistically significant difference compared with the controls. Urinary TBARS and MDA had statistically significant correlations with alphaGST, GAL, NAG, Ca, and oxalate, but had no correlation with piGST, citrate, OPN, Mg, and P. Urinary citrate had a negative, linear, and statistically significant correlation with alphaGST, GAL, and NAG. **CONCLUSIONS**: Lipid peroxidation correlated with hyperoxaluria and renal tubular damage, indicating that hyperoxaluria can induce tubular cell injury and that this injury may be due to the production of free radicals in patients with calcium oxalate stones. Renal tubular damage in patients with stones may be limited to the proximal tubules.

**URL:** <https://ovidsp.ovid.com/ovidweb.cgi?T=JS&CSC=Y&NEWS=N&PAGE=fulltext&D=med5&DO=10.1016%2fs0090-4295%2803%2900764-7> <https://libkey.io/libraries/2592/openurl?genre=article&sid=OVID:medline&id=pmid:14665375&id=doi:10.1016%2Fs0090-4295%2803%2900764-7&issn=0090-4295&isbn=&volume=62&issue=6&spage=1123&pages=1123-8&date=2003&title=Urology&atitle=Lipid+peroxidation+and+its+correlations+with+urinary+levels+of+oxalate%2C+citric+acid%2C+and+osteopontin+in+patients+with+renal+calcium+oxalate+stones.&aulast=Huang&pid=%3Cauthor%3EHuang+HS%3BMa+MC%3BChen+CF%3BChen+J%3C%2Fauthor%3E%3CAN%3E14665375%3C%2FAN%3E%3CDT%3EJournal+Article%3C%2FDT%3E>

145. **Urine calcium and volume predict coverage of renal papilla by Randall's plaque.**

**Item Type:** Journal Article

**Authors:** Kuo, Ramsay L.;Lingeman, James E.;Evan, Andrew P.;Paterson, Ryan F.;Parks, Joan H.;Bledsoe, Sharon B.;Munch, Larry C. and Coe, Fredric L.

**Publication Date:** De ,2003

**Journal:** Kidney International 64(6), pp. 2150–2154

**Abstract: BACKGROUND**: Renal papillary plaques are common in calcium stone formers. We hypothesized that plaque should increase directly with urine calcium excretion, and inversely with urine volume. To test this, we measured papillary plaque areas in both idiopathic calcium oxalate stone formers and nonstone formers and examined 24-hour urine data to identify significant correlations. **METHODS**: Fourteen stone formers and four nonstone forming controls underwent papillary mapping with flexible nephroscopy. For each papillum, representative still images and moving pictures expert group (MPEG) movies were used to identify plaque extent and papillary borders. The mean fractional plaque coverage for each polar region (upper, inter, lower) and per papillum was calculated. The relationship of the plaque coverage data to urine measurements was assessed with general multivariate linear modeling. **RESULTS**: Mean polar fractional plaque coverage was higher in the calcium oxalate stone formers (7.4% vs. 0.5%, P= 0.012) as was mean fractional plaque per papillum (7.6% vs. 0.6%, P= 0.011). When correlating mean polar plaque coverage to urine data, urine volume and calcium excretion were the only measurements with independent relationships to plaque (P= 0.002, adjusted multiple R2= 0.521), with higher calcium and lower volume increasing coverage. The same relationships hold for mean plaque per papillum, except that urine pH also becomes an independent factor (P= 0.001, adjusted multiple R2= 0.606). **CONCLUSION**: Utilizing advanced digital video and endoscopic equipment, we have achieved the most accurate estimation of papillary plaque coverage to date. Our findings support the idea that urine volume and calcium are the main correlates of plaque coverage.

**URL:** <https://ovidsp.ovid.com/ovidweb.cgi?T=JS&CSC=Y&NEWS=N&PAGE=fulltext&D=med5&DO=10.1046%2fj.1523-1755.2003.00316.x> <https://libkey.io/libraries/2592/openurl?genre=article&sid=OVID:medline&id=pmid:14633137&id=doi:10.1046%2Fj.1523-1755.2003.00316.x&issn=0085-2538&isbn=&volume=64&issue=6&spage=2150&pages=2150-4&date=2003&title=Kidney+International&atitle=Urine+calcium+and+volume+predict+coverage+of+renal+papilla+by+Randall%27s+plaque.&aulast=Kuo&pid=%3Cauthor%3EKuo+RL%3BLingeman+JE%3BEvan+AP%3BPaterson+RF%3BParks+JH%3BBledsoe+SB%3BMunch+LC%3BCoe+FL%3C%2Fauthor%3E%3CAN%3E14633137%3C%2FAN%3E%3CDT%3EJournal+Article%3C%2FDT%3E>

146. **Urinary oxalate levels and the enteric bacterium Oxalobacter formigenes in patients with calcium oxalate urolithiasis.**

**Item Type:** Journal Article

**Authors:** Kwak, Cheol;Kim, Hee Kyung;Kim, Eui Chong;Choi, Myung Sik and Kim, Hyeon Hoe

**Publication Date:** Oc ,2003

**Journal:** European Urology 44(4), pp. 475–481

**Abstract: OBJECTIVES**: We performed a prospective study to evaluate the intestinal colonization of Oxalobacter formigenes and its relationship with urinary oxalate levels in patients with calcium oxalate stone disease. **METHODS**: One hundred and three patients with calcium oxalate urolithiasis, ranging in age from 21 to 73 years (mean age, 47 years) who were followed from August 2000 to September 2001 participated in this study. Fresh stool and 24-hour urine samples were collected. Genus specific oligonucleotide sequences corresponding to the homologous regions residing in the oxc gene were designed. In order to quantify O. formigenes in clinical specimens, a quantitative-PCR-based assay system utilizing a competitive DNA template as an internal standard was developed. Urine volume, pH, creatinine, oxalate, calcium, magnesium, phosphate, citrate and uric acid were measured. **RESULTS**: Intestinal Oxalobacteria were detected in 45.6% (n=47) of calcium oxalate stone patients by PCR. In stone formers who tested negative for Oxalobacteria, the average urinary oxalate level was 0.36 mmol/day, and this compared to 0.29 mmol/day for those patients that tested positive for Oxalobacteria (p: Intestinal Oxalobacteria were detected in 45.6% (n=47) of calcium oxalate stone patients by PCR. In stone formers who tested negative for Oxalobacteria, the average urinary oxalate level was 0.36 mmol/day, and this compared to 0.29 mmol/day for those patients that tested positive for Oxalobacteria (p**CONCLUSION**: Our results support the concept that O. formigenes is important in maintaining oxalate homeostasis and that its absence from the gut may be the risk of calcium oxalate urolithiasis.

**URL:** <https://ovidsp.ovid.com/ovidweb.cgi?T=JS&CSC=Y&NEWS=N&PAGE=fulltext&D=med5&DO=10.1016%2fs0302-2838%2803%2900318-x> <https://libkey.io/libraries/2592/openurl?genre=article&sid=OVID:medline&id=pmid:14499684&id=doi:10.1016%2Fs0302-2838%2803%2900318-x&issn=0302-2838&isbn=&volume=44&issue=4&spage=475&pages=475-81&date=2003&title=European+Urology&atitle=Urinary+oxalate+levels+and+the+enteric+bacterium+Oxalobacter+formigenes+in+patients+with+calcium+oxalate+urolithiasis.&aulast=Kwak&pid=%3Cauthor%3EKwak+C%3BKim+HK%3BKim+EC%3BChoi+MS%3BKim+HH%3C%2Fauthor%3E%3CAN%3E14499684%3C%2FAN%3E%3CDT%3EJournal+Article%3C%2FDT%3E>

147. **Influence of urinary stones on the composition of a 24-hour urine sample.**

**Item Type:** Journal Article

**Authors:** Laube, Norbert;Pullmann, Michael;Hergarten, Stefan and Hesse, Albrecht

**Publication Date:** Fe ,2003

**Journal:** Clinical Chemistry 49(2), pp. 281–285

**Abstract: BACKGROUND**: It can be assumed that stones in the urinary tract continuously increase in size by incorporating material from urine. Consequently, urine will exhibit depleted concentrations of lithogenic constituents when urinary stones are present in the patient's urinary tract. **METHODS**: To calculate the influence of the depletion effect, we considered two different models of stone growth. In the first model, the increase in stone size depends only on the urinary concentration of a lithogenic substance; the second model also considers the surface area of the growing stone. The case of only one kidney being affected by stone formation is considered separately. We discuss example calculations involving the formation of calcium oxalate. **RESULTS**: The calculated depletion effects are of a nonnegligible order of magnitude. Assuming both a measured oxalate concentration of, e.g., 0.37 mmol/L and a reasonable in vivo stone growing rate of 10 mm(3)/day, a relative underestimation of the real "in situ" oxalate concentration between approximately 21% (model 1) and approximately 42% (model 2) occurs. The depletion effect increases markedly with increasing stone growth rate. **CONCLUSIONS**: Metabolic status can be evaluated correctly only in patients who have been declared "stone-free", e.g., after stone removal. Because the expected stone-related depletion effect in most cases is of high clinical relevance, we recommend estimating the effect of the order of magnitude of the depletion on actual urinary composition.

**URL:** <https://ovidsp.ovid.com/ovidweb.cgi?T=JS&CSC=Y&NEWS=N&PAGE=fulltext&D=med5&DO=10.1373%2f49.2.281> <https://libkey.io/libraries/2592/openurl?genre=article&sid=OVID:medline&id=pmid:12560352&id=doi:10.1373%2F49.2.281&issn=0009-9147&isbn=&volume=49&issue=2&spage=281&pages=281-5&date=2003&title=Clinical+Chemistry&atitle=Influence+of+urinary+stones+on+the+composition+of+a+24-hour+urine+sample.&aulast=Laube&pid=%3Cauthor%3ELaube+N%3BPullmann+M%3BHergarten+S%3BHesse+A%3C%2Fauthor%3E%3CAN%3E12560352%3C%2FAN%3E%3CDT%3EJournal+Article%3C%2FDT%3E>

148. **Early changes of oxalate and calcium urine excretion in those with calcium oxalate stone formation after extracorporeal shock wave lithotripsy.**

**Item Type:** Journal Article

**Authors:** Oehlschlager, S.;Albrecht, S.;Hakenberg, O. W.;Schrodter, S.;Froehner, M.;Manseck, A. and Wirth, M. P.

**Publication Date:** Ju ,2003

**Journal:** Urology 62(1), pp. 17–21

**Abstract: OBJECTIVES**: To determine the extent of transient changes of tubular function in idiopathic calcium-oxalate (CaOx) stone-bearing patients after extracorporeal shock wave lithotripsy (ESWL), calcium and oxalate excretion were measured before and after ESWL. **METHODS**: In 22 patients with renal CaOx stones, the plasma values and urine excretion of creatinine, calcium, oxalate, magnesium, and citrate were measured before and on days 1 and 2 after ESWL under conditions of a standardized diet. Overnight urine collection for an 8-hour period was used to measure the urine excretion, and the values were extrapolated to a 24-hour period. For calculation of the urine ion activity, the AP(CaOx) index EQ(s) and the CaOx risk index were used. **RESULTS**: After ESWL, hyperoxaluria was noted in 10 patients compared with 2 before ESWL. Hypercalciuria was seen in 11 patients after ESWL compared with 3 before. Combined hyperoxaluria and hypercalciuria was found in 7 patients after ESWL compared with 1 before. Both the oxalate/creatinine and calcium/creatinine ratios were significantly increased after ESWL. The AP(CaOx) index EQ(s) and the CaOx risk index were significantly increased after ESWL in patients with increased post-ESWL calcium and/or oxalate excretion. **CONCLUSIONS**: Increased calcium and/or oxalate excretion can be seen in patients with CaOx stones early after ESWL. This increased excretion of lithogenic substances represents an increased risk of fragment apposition after ESWL for the group with a significantly increased AP(CaOx) index EQ(s) and CaOx risk index. Therefore, prophylactic measures in patients at risk early after ESWL might be warranted to prevent possible recurrent stone formation.

**URL:** <https://ovidsp.ovid.com/ovidweb.cgi?T=JS&CSC=Y&NEWS=N&PAGE=fulltext&D=med5&DO=10.1016%2fs0090-4295%2803%2900122-5> <https://libkey.io/libraries/2592/openurl?genre=article&sid=OVID:medline&id=pmid:12837412&id=doi:10.1016%2Fs0090-4295%2803%2900122-5&issn=0090-4295&isbn=&volume=62&issue=1&spage=17&pages=17-21&date=2003&title=Urology&atitle=Early+changes+of+oxalate+and+calcium+urine+excretion+in+those+with+calcium+oxalate+stone+formation+after+extracorporeal+shock+wave+lithotripsy.&aulast=Oehlschlager&pid=%3Cauthor%3EOehlschlager+S%3BAlbrecht+S%3BHakenberg+OW%3BSchrodter+S%3BFroehner+M%3BManseck+A%3BWirth+MP%3C%2Fauthor%3E%3CAN%3E12837412%3C%2FAN%3E%3CDT%3EComparative+Study%3C%2FDT%3E>

149. **Urinary saturation and risk factors for calcium oxalate stone disease based on spot and 24-hour urine specimens.**

**Item Type:** Journal Article

**Authors:** Ogawa, Yoshihide;Yonou, Hiroyuki;Hokama, Sanehiro;Oda, Masami;Morozumi, Makoto and Sugaya, Kimio

**Publication Date:** Sep 01 ,2003

**Journal:** Frontiers in Bioscience 8, pp. 167

**Abstract:** In 222 random spot urine specimens, the calcium concentration and calcium oxalate saturation [DG(CaOx)] were significantly higher among stone formers than among non-stone formers, while the citrate and creatinine-corrected citrate concentrations were lower. In 188 24-hour urine specimens, magnesium excretion was lower among stone formers than non-stone formers, while the creatinine-corrected calcium concentration and DG(CaOx) were higher. Among stone formers, there was no gender difference in the urinary concentrations of calcium, oxalate, citrate, magnesium, and DG(CaOx), but the creatinine-corrected calcium, citrate, and magnesium concentrations were higher in women, as well as 24-hour citrate excretion. The levels of calcium and oxalate have a major influence on DG(CaOx), while citrate and magnesium levels have a minor influence. DG(CaOx) was correlated with calcium and oxalate excretion, as well as with the creatinine-corrected calcium and oxalate concentrations. Approximately 5% of 24-hour urine specimens showed critical supersaturation, 80% showed metastable supersaturation, and 15% were unsaturated. Hypercalciuria or hyperoxaluria was fairly common (30% and 40%) in critically supersaturated urine, while it was less common (22.4% and 8.6%) in metastably supersaturated urine and was not detected in unsaturated urine. Hypocitraturia and/or hypomagnesiuria was more common (63.8-80%) at any saturation. The urinary calcium, oxalate, and citrate concentrations, as well as the creatinine-corrected calcium, oxalate, citrate, and magnesium concentrations and DG(CaOx), showed a significant correlation between 57 paired early morning spot urine and 24-hour urine specimens. The creatinine-corrected calcium and citrate concentrations of the early morning urine specimens were significantly correlated with the levels of calcium and citrate excretion in the paired 24-hour urine specimens. In conclusion, no parameter other than urinary saturation gives more than a vague indication of the risk of lithogenesis, so DG(CaOx) in either early morning urine or 24-hour urine specimens appears to be the best predictor of stone risk. Finally, the creatinine-corrected calcium and citrate concentrations in early morning urine can be used as a substitute for measuring 24-hour excretion.

**URL:** <https://ovidsp.ovid.com/ovidweb.cgi?T=JS&CSC=Y&NEWS=N&PAGE=fulltext&D=med5&DO=10.2741%2f1139> <https://libkey.io/libraries/2592/openurl?genre=article&sid=OVID:medline&id=pmid:12957883&id=doi:10.2741%2F1139&issn=1093-9946&isbn=&volume=8&issue=1&spage=a167&pages=a167-76&date=2003&title=Frontiers+in+Bioscience&atitle=Urinary+saturation+and+risk+factors+for+calcium+oxalate+stone+disease+based+on+spot+and+24-hour+urine+specimens.&aulast=Ogawa&pid=%3Cauthor%3EOgawa+Y%3BYonou+H%3BHokama+S%3BOda+M%3BMorozumi+M%3BSugaya+K%3C%2Fauthor%3E%3CAN%3E12957883%3C%2FAN%3E%3CDT%3EJournal+Article%3C%2FDT%3E>

150. **Thiazide does not affect urine oxalate excretion.**

**Item Type:** Journal Article

**Authors:** Parks, Joan H. and Coe, Frederic L.

**Publication Date:** Au ,2003

**Journal:** Journal of Urology 170(2 Pt 1), pp. 393–396

**Abstract: PURPOSE**: We assessed the effects of thiazide treatment on urine oxalate excretion in patients with kidney stones. **MATERIALS AND METHODS**: Of 537 patients (231 women), 209 (81 women) received thiazide for stone prevention. Urine oxalate was measured in 3, 24-hour urines before treatment, and in 1, 24-hour urine after 6 to 12 weeks of treatment. This was an observational cohort and treatment was not randomized. **RESULTS**: Urine oxalate increased in general. There was no difference in increase with or without thiazide, whether one considered simple t test comparisons or used ANOVA with pretreatment oxalate excretion as a covariate. **CONCLUSIONS**: Thiazide administration exerts no measurable effect on urine oxalate excretion that can be detected in clinical practice.

**URL:** <https://ovidsp.ovid.com/ovidweb.cgi?T=JS&CSC=Y&NEWS=N&PAGE=fulltext&D=med5&DO=10.1097%2f01.ju.0000076371.92938.9b> <https://libkey.io/libraries/2592/openurl?genre=article&sid=OVID:medline&id=pmid:12853783&id=doi:10.1097%2F01.ju.0000076371.92938.9b&issn=0022-5347&isbn=&volume=170&issue=2&spage=393&pages=393-6&date=2003&title=Journal+of+Urology&atitle=Thiazide+does+not+affect+urine+oxalate+excretion.&aulast=Parks&pid=%3Cauthor%3EParks+JH%3BCoe+FL%3C%2Fauthor%3E%3CAN%3E12853783%3C%2FAN%3E%3CDT%3EClinical+Trial%3C%2FDT%3E>

151. **Durability of the medical management of cystinuria.**

**Item Type:** Journal Article

**Authors:** Pietrow, Paul K.;Auge, Brian K.;Weizer, Alon Z.;Delvecchio, Fernando C.;Silverstein, Ari D.;Mathias, Barbara;Albala, David M. and Preminger, Glenn M.

**Publication Date:** Ja ,2003

**Journal:** Journal of Urology 169(1), pp. 68–70

**Abstract: PURPOSE**: Cystinuria is an autosomal recessive disorder of dibasic amino acid transport in the kidney that leads to an abundance of cystine in the urine. This molecule is poorly soluble in urine and it is prone to crystallization and stone formation at concentrations above 300 mg./l. Medical treatment in these patients has incorporated increasing urine volumes, alkalinization and thiol medications that decrease the availability of free cystine in urine. Despite a reasonable prognosis for reduced stone formation we and others have noted difficulties in patients complying with medical management recommendations. Therefore, we evaluated the durability of treatment success in our patients with cystinuria. **MATERIALS AND METHODS**: A retrospective chart review was performed in all patients with cystinuria referred to the comprehensive kidney stone center at our institution for an 8-year period. Medical therapy, stone recurrence rates, compliance with medications and scheduled followup, and the results of metabolic evaluations via 24-hour urine collections were reviewed. The average concentrations of urinary cystine in initial and followup 24-hour samples were compared in patients compliant and noncompliant with medical treatment. In addition, each patient was mailed a 1-page questionnaire to assess the self-perception of medical compliance. **RESULTS**: We identified 26 patients with a mean age of 32 years at referral (range 13 to 67) who were followed an average of 38.2 months (range 6 to 83). Females represented 58% of those with cystinuria. Overall compliance with medical recommendations was poor with a short duration of success. Of the 26 patients followed at our stone center only 4 (15%) achieved and maintained therapeutic success, as defined by urine cystine less than 300 mg./l. An additional 11 patients (42%) achieved therapeutic success but subsequently had failure at an average of 16 months (range 6 to 27). Of these patients 7 (64%) regained therapeutic success at an average of 9.4 months (range 4 to 20). Five patients (19%) never achieved therapeutic success, while an additional 6 (23%) failed to present to followup appointments or provide subsequent 24-hour urine studies despite referral to a tertiary care center. Patient self-assessment of medical compliance was uniformly high regardless of physician perceptions or treatment results. CONCLUSIONS The durability of medically treating patients with cystinuria is limited with only a small percent able to achieve and maintain the goal of decreasing cystine below the saturation concentration. Greater physician vigilance in these complicated stone formers is required to achieve successful prophylactic management. Furthermore, these patients require better insight into the own disease to improve compliance.

**URL:** <https://ovidsp.ovid.com/ovidweb.cgi?T=JS&CSC=Y&NEWS=N&PAGE=fulltext&D=med5&DO=10.1016%2fS0022-5347%2805%2964037-2> <https://libkey.io/libraries/2592/openurl?genre=article&sid=OVID:medline&id=pmid:12478105&id=doi:10.1016%2FS0022-5347%2805%2964037-2&issn=0022-5347&isbn=&volume=169&issue=1&spage=68&pages=68-70&date=2003&title=Journal+of+Urology&atitle=Durability+of+the+medical+management+of+cystinuria.&aulast=Pietrow&pid=%3Cauthor%3EPietrow+P%3BAuge+BK%3BWeizer+AZ%3BDelvecchio+FC%3BSilverstein+AD%3BMathias+B%3BAlbala+DM%3BPreminger+GM%3C%2Fauthor%3E%3CAN%3E12478105%3C%2FAN%3E%3CDT%3EJournal+Article%3C%2FDT%3E>

152. **Dietary risk factors for hyperoxaluria in calcium oxalate stone formers.**

**Item Type:** Journal Article

**Authors:** Siener, Roswitha;Ebert, Dagmar;Nicolay, Claudia and Hesse, Albrecht

**Publication Date:** Ma ,2003

**Journal:** Kidney International 63(3), pp. 1037–1043

**Abstract: BACKGROUND**: Hyperoxaluria is a major predisposing factor in calcium oxalate urolithiasis. The aim of the present study was to clarify the role of dietary oxalate in urinary oxalate excretion and to assess dietary risk factors for hyperoxaluria in calcium oxalate stone patients. **METHODS**: Dietary intakes of 186 calcium oxalate stone formers, 93 with hyperoxaluria (>or=0.5 mmol/day) and 93 with normal oxalate excretion (: Dietary intakes of 186 calcium oxalate stone formers, 93 with hyperoxaluria (>or=0.5 mmol/day) and 93 with normal oxalate excretion (**RESULTS**: The mean daily intakes of energy, total protein, fat and carbohydrates were similar in both groups. The diets of the patients with hyperoxaluria were estimated to contain 130 mg/day oxalate and 812 mg/day calcium as compared to 101 mg/day oxalate and 845 mg/day calcium among patients without hyperoxaluria. These differences were not significant. The mean daily intakes of water (in food and beverages), magnesium, potassium, dietary fiber and ascorbic acid were greater in patients with hyperoxaluria than in stone formers with normal oxalate excretion. Multiple logistic regression analysis revealed that urinary oxalate excretion was significantly associated with dietary ascorbate and fluid intake, and inversely related to calcium intake. Differences of estimated diet composition of both groups corresponded to differences in urinary parameters. **CONCLUSIONS**: These findings suggest that hyperoxaluria predominantly results from increased endogenous production and from intestinal hyperabsorption of oxalate, partly caused by an insufficient supply or low availability of calcium for complexation with oxalate in the intestinal lumen.

**URL:** <https://ovidsp.ovid.com/ovidweb.cgi?T=JS&CSC=Y&NEWS=N&PAGE=fulltext&D=med5&DO=10.1046%2fj.1523-1755.2003.00807.x> <https://libkey.io/libraries/2592/openurl?genre=article&sid=OVID:medline&id=pmid:12631085&id=doi:10.1046%2Fj.1523-1755.2003.00807.x&issn=0085-2538&isbn=&volume=63&issue=3&spage=1037&pages=1037-43&date=2003&title=Kidney+International&atitle=Dietary+risk+factors+for+hyperoxaluria+in+calcium+oxalate+stone+formers.&aulast=Siener&pid=%3Cauthor%3ESiener+R%3BEbert+D%3BNicolay+C%3BHesse+A%3C%2Fauthor%3E%3CAN%3E12631085%3C%2FAN%3E%3CDT%3EJournal+Article%3C%2FDT%3E>

153. **Metabolic risk factors in pediatric and adult calcium oxalate urinary stone formers: is there any difference?.**

**Item Type:** Journal Article

**Authors:** Tefekli, Ahmet;Esen, Tarik;Ziylan, Orhan;Erol, Bulent;Armagan, Abdullah;Ander, Haluk and Akinci, Mustafa

**Publication Date:** 2003

**Journal:** Urologia Internationalis 70(4), pp. 273–277

**Abstract: OBJECTIVES**: Urolithiasis in children is recognized with an increasing frequency, while exact etiological factors remain to be determined. The aim of this study is to compare the metabolic risk factors and saturation of urine in pediatric and adult calcium oxalate (Ca-Ox) stone formers. **METHODS**: A total of 33 pediatric (mean age: 6.8 +/- 3.1 years) and 120 adult patients (mean age: 39.7 +/- 5.7 years), with documented Ca-Ox urinary stone disease, underwent a comprehensive metabolic evaluation at our institution. Beside a broad serum analysis, concentrations of calcium, oxalate, magnesium, uric acid and citrate were measured in 24-hour collected urine. Saturation of urine was calculated by Marshall-Robertson's nomograms. **RESULTS**: Hypocitraturia, observed in 60.6%, and hypomagnesuria, detected in 39.4%, but not hypercalciuria, were the most common metabolic risk factors in the pediatric group. In adults, hypercalciuria still represented one of the major metabolic risk factors, detected in 44.1%, although hypocitraturia, observed in 45.8%, was the most prevalent metabolic risk factor, as it was in the pediatric group. Pediatric cases had significantly (p : Hypocitraturia, observed in 60.6%, and hypomagnesuria, detected in 39.4%, but not hypercalciuria, were the most common metabolic risk factors in the pediatric group. In adults, hypercalciuria still represented one of the major metabolic risk factors, detected in 44.1%, although hypocitraturia, observed in 45.8%, was the most prevalent metabolic risk factor, as it was in the pediatric group. Pediatric cases had significantly (p **CONCLUSIONS**: Metabolic risk factors significantly differ in pediatric and adult Ca-Ox stone formers. Hypocitraturia and hypomagnesuria seem to play a major role in stone formation, and metabolic abnormalities can be detected in a significant percentage of both primary and recurrent pediatric stone formers. Thus, a comprehensive metabolic evaluation is of utmost importance for all children with Ca-Ox stones. Copyright 2003 S. Karger AG, Basel

**URL:** <https://ovidsp.ovid.com/ovidweb.cgi?T=JS&CSC=Y&NEWS=N&PAGE=fulltext&D=med5&DO=10.1159%2f000070134> <https://libkey.io/libraries/2592/openurl?genre=article&sid=OVID:medline&id=pmid:12740490&id=doi:10.1159%2F000070134&issn=0042-1138&isbn=&volume=70&issue=4&spage=273&pages=273-7&date=2003&title=Urologia+Internationalis&atitle=Metabolic+risk+factors+in+pediatric+and+adult+calcium+oxalate+urinary+stone+formers%3A+is+there+any+difference%3F.&aulast=Tefekli&pid=%3Cauthor%3ETefekli+A%3BEsen+T%3BZiylan+O%3BErol+B%3BArmagan+A%3BAnder+H%3BAkinci+M%3C%2Fauthor%3E%3CAN%3E12740490%3C%2FAN%3E%3CDT%3EJournal+Article%3C%2FDT%3E>

154. **Effect of ascorbic acid consumption on urinary stone risk factors.**

**Item Type:** Journal Article

**Authors:** Traxer, Olivier;Huet, Beverley;Poindexter, John;Pak, Charles Y. C. and Pearle, Margaret S.

**Publication Date:** Au ,2003

**Journal:** Journal of Urology 170(2 Pt 1), pp. 397–401

**Abstract: PURPOSE**: Ascorbic acid (AA) has been implicated as a risk factor for calcium oxalate stones due to its conversion to oxalate and potential acidifying properties. We evaluated the effect of AA consumption on urinary saturation of calcium oxalate (CaOx) and urinary pH. **MATERIALS AND METHODS**: A total of 12 normal subjects (NS) and 12 CaOx stone formers (SF) underwent 2, 6-day phases of study while maintained on a controlled metabolic diet. In each phase subjects ingested 1 gm AA or an identical appearing placebo twice daily. On the last 2 days of each phase 2, 24-hour urine collections were analyzed for pH and stone risk factors, and blood specimens were submitted for serum chemistry studies. **RESULTS**: No difference in urinary pH was found between placebo and AA phases in NS (6.02 versus 6.02) and SF (6.0 versus 6.0). However, urinary oxalate was statistically significantly higher in the AA versus placebo phase for NS (34.7 versus 28.5 mg, p = 0.008) and SF (41.0 versus 30.5 mg, p : No difference in urinary pH was found between placebo and AA phases in NS (6.02 versus 6.02) and SF (6.0 versus 6.0). However, urinary oxalate was statistically significantly higher in the AA versus placebo phase for NS (34.7 versus 28.5 mg, p = 0.008) and SF (41.0 versus 30.5 mg, p **CONCLUSIONS**: Ingestion of 2 gm AA daily results in no change in urinary pH but a moderate though statistically significant increase in urinary oxalate in NS (20%) and SF (33%). Stone formers respond no differently to AA than normal subjects. We recommend limiting AA use to less than 2 gm daily in CaOx stone formers.

**URL:** <https://ovidsp.ovid.com/ovidweb.cgi?T=JS&CSC=Y&NEWS=N&PAGE=fulltext&D=med5&DO=10.1097%2f01.ju.0000076001.21606.53> <https://libkey.io/libraries/2592/openurl?genre=article&sid=OVID:medline&id=pmid:12853784&id=doi:10.1097%2F01.ju.0000076001.21606.53&issn=0022-5347&isbn=&volume=170&issue=2&spage=397&pages=397-401&date=2003&title=Journal+of+Urology&atitle=Effect+of+ascorbic+acid+consumption+on+urinary+stone+risk+factors.&aulast=Traxer&pid=%3Cauthor%3ETraxer+O%3BHuet+B%3BPoindexter+J%3BPak+CY%3BPearle+MS%3C%2Fauthor%3E%3CAN%3E12853784%3C%2FAN%3E%3CDT%3EClinical+Trial%3C%2FDT%3E>

155. **Intestinal Oxalobacter formigenes colonization in calcium oxalate stone formers and its relation to urinary oxalate.**

**Item Type:** Journal Article

**Authors:** Troxel, Scott A.;Sidhu, Harmeet;Kaul, Poonam and Low, Roger K.

**Publication Date:** Ap ,2003

**Journal:** Journal of Endourology 17(3), pp. 173–176

**Abstract: BACKGROUND AND PURPOSE**: Oxalobacter formigenes is an anaerobic commensal colonic bacterium capable of degrading oxalate through the enzyme oxalyl-CoA decarboxylase. It has been theorized that individuals who lack this bacterium have higher intestinal oxalate absorption, leading to a higher urinary oxalate concentration and an increased risk of calcium oxalate urolithiasis. We performed a prospective, controlled study to evaluate O. formigenes colonization in calcium oxalate stone formers and to correlate colonization with urinary oxalate and other standard urinary stone risk factors. **PATIENTS AND METHODS**: Thirty-five first-time calcium oxalate stone formers were compared with 10 control subjects having no history of urolithiasis and a normal renal ultrasound scan. All subjects underwent standard metabolic testing by submitting serum and 24-hour urine specimens. In addition, all subjects submitted stool samples for culture and detection of O. formigenes by Xentr(ix) O. formigenes Monitor. **RESULTS**: Intestinal Oxalobacter was detected in only 26% of the stone formers compared with 60% of the controls (p 44 mg/day) tested negative for O. formigenes (p 44 mg/day) tested negative for O. formigenes (p **CONCLUSIONS**: Calcium oxalate stone formers have a low rate of colonization with O. formigenes. Among stone formers, absence of intestinal Oxalobacter correlates with higher urinary oxalate concentration and an increased risk of hyperoxaluria. Introduction of the Oxalobacter bacterium or an analog of its enzyme oxalyl-CoA decarboxylase into the intestinal tract may be a treatment for calcium oxalate stone disease.

**URL:** <https://ovidsp.ovid.com/ovidweb.cgi?T=JS&CSC=Y&NEWS=N&PAGE=fulltext&D=med5&DO=10.1089%2f089277903321618743> <https://libkey.io/libraries/2592/openurl?genre=article&sid=OVID:medline&id=pmid:12803990&id=doi:10.1089%2F089277903321618743&issn=0892-7790&isbn=&volume=17&issue=3&spage=173&pages=173-6&date=2003&title=Journal+of+Endourology&atitle=Intestinal+Oxalobacter+formigenes+colonization+in+calcium+oxalate+stone+formers+and+its+relation+to+urinary+oxalate.&aulast=Troxel&pid=%3Cauthor%3ETroxel+SA%3BSidhu+H%3BKaul+P%3BLow+RK%3C%2Fauthor%3E%3CAN%3E12803990%3C%2FAN%3E%3CDT%3EJournal+Article%3C%2FDT%3E>

156. **Metabolic evaluation of children with urolithiasis: are adult references for supersaturation appropriate?.**

**Item Type:** Journal Article

**Authors:** Battino, Benjamin S.;DeFOOR, William;Coe, Frederic;Tackett, Leslie;Erhard, Michael;Wacksman, Jeffrey;Sheldon, Curtis A. and Minevich, Eugene

**Publication Date:** De ,2002

**Journal:** Journal of Urology 168(6), pp. 2568–2571

**Abstract: PURPOSE**: We determined the incidence of urinary stone risk factors in pediatric patients with urolithiasis. **MATERIALS AND METHODS**: Between 1998 and 2001, 71 children with urolithiasis at 2 pediatric institutions underwent metabolic evaluation. The 24-hour urine samples were analyzed outside central laboratory using adult and known pediatric references. Supersaturation and traditional metabolic parameters were determined and compared. **RESULTS**: All patients had metabolic abnormalities. Calcium related abnormalities were present in 92% of children, calcium oxalate supersaturation was abnormal in 69%, calcium phosphate supersaturation was elevated in 70% and traditional calcium parameters were abnormal in 80%. While 11% of the patients had abnormal calcium phosphate or oxalate supersaturation with normal traditional calcium parameters, 10% had normal calcium oxalate or phosphate supersaturation with abnormal traditional calcium parameters. Low urinary volume was identified in 75% of the children. **CONCLUSIONS**: Metabolic abnormalities are extremely common in pediatric patients with urolithiasis. Calcium related abnormalities are the most common abnormality. Urinary supersaturation values are complementary to traditional metabolic parameters and may be more sensitive predictors of recurrent stone risk. It is important to establish pediatric reference ranges to interpret these data more accurately.

**URL:** <https://ovidsp.ovid.com/ovidweb.cgi?T=JS&CSC=Y&NEWS=N&PAGE=fulltext&D=med4&DO=10.1016%2fS0022-5347%2805%2964217-6> <https://libkey.io/libraries/2592/openurl?genre=article&sid=OVID:medline&id=pmid:12441985&id=doi:10.1016%2FS0022-5347%2805%2964217-6&issn=0022-5347&isbn=&volume=168&issue=6&spage=2568&pages=2568-71&date=2002&title=Journal+of+Urology&atitle=Metabolic+evaluation+of+children+with+urolithiasis%3A+are+adult+references+for+supersaturation+appropriate%3F.&aulast=Battino&pid=%3Cauthor%3EBattino+BS%3BDeFOOR+W%3BCoe+F%3BTackett+L%3BErhard+M%3BWacksman+J%3BSheldon+CA%3BMinevich+E%3C%2Fauthor%3E%3CAN%3E12441985%3C%2FAN%3E%3CDT%3EJournal+Article%3C%2FDT%3E>

157. **Estrogen replacement increased the citrate and calcium excretion rates in postmenopausal women with recurrent urolithiasis.**

**Item Type:** Journal Article

**Authors:** Dey, Jayant;Creighton, Amery;Lindberg, Jill S.;Fuselier, Harold A.;Kok, Dirk J.;Cole, Francis E. and Hamm, Lee L.

**Publication Date:** Ja ,2002

**Journal:** Journal of Urology 167(1), pp. 169–171

**Abstract: PURPOSE**: Epidemiological data indicate a sharp increase in urinary calcium stone formation after menopause. We investigated the role of menopausal estrogen replacement therapy on the urinary constituents and characteristics that may influence recurrent calcium oxalate stone disease. **MATERIALS AND METHODS**: Urinary constituents in 28 postmenopausal women on estrogen replacement therapy for more than 6 months were compared with those in 41 women who had never been exposed to estrogen after menopause. These 2 groups had a history of recurrent calcium oxalate urolithiasis. A group of age matched, nonstone forming volunteers who were and were not on estrogen served as controls. **RESULTS**: The 24-hour urine collection revealed significantly higher mean calcium plus or minus standard deviation (188.8 +/- 101.5 versus 129.2 +/- 80.9 mg./24 hours, p : The 24-hour urine collection revealed significantly higher mean calcium plus or minus standard deviation (188.8 +/- 101.5 versus 129.2 +/- 80.9 mg./24 hours, p : The 24-hour urine collection revealed significantly higher mean calcium plus or minus standard deviation (188.8 +/- 101.5 versus 129.2 +/- 80.9 mg./24 hours, p : The 24-hour urine collection revealed significantly higher mean calcium plus or minus standard deviation (188.8 +/- 101.5 versus 129.2 +/- 80.9 mg./24 hours, p **CONCLUSIONS**: Higher urinary citrate and higher agglomeration inhibition in women exposed to estrogen may decrease the risk of subsequent calcium stone formation.

**URL:** <https://ovidsp.ovid.com/ovidweb.cgi?T=JS&CSC=Y&NEWS=N&PAGE=fulltext&D=med4&AN=11743298> <https://libkey.io/libraries/2592/openurl?genre=article&sid=OVID:medline&id=pmid:11743298&id=doi:&issn=0022-5347&isbn=&volume=167&issue=1&spage=169&pages=169-71&date=2002&title=Journal+of+Urology&atitle=Estrogen+replacement+increased+the+citrate+and+calcium+excretion+rates+in+postmenopausal+women+with+recurrent+urolithiasis.&aulast=Dey&pid=%3Cauthor%3EDey+J%3BCreighton+A%3BLindberg+JS%3BFuselier+HA%3BKok+DJ%3BCole+FE%3BHamm+L%3C%2Fauthor%3E%3CAN%3E11743298%3C%2FAN%3E%3CDT%3EComparative+Study%3C%2FDT%3E>

158. **The diagnostic value of the phosphate levels in serum and 24-hour urine samples in patients with recurrent renal stone disease.**

**Item Type:** Journal Article

**Authors:** Muezzinoglu, Talha;Gumus, Bilal;Sener, Ercument;Ari, Zeki and Buyuksu, Coskun

**Publication Date:** 2002

**Journal:** Scandinavian Journal of Urology & Nephrology 36(5), pp. 368–372

**Abstract: OBJECTIVE**: The aim of this study is to investigate the value of phosphate levels in serum and urine in patients with recurrent renal stone disease. **MATERIALS AND METHODS**: The patients (n:60) were divided into two groups as first-time stone disease (group 1) and recurrent renal stone disease (group 2). The demographical datas and their historical information were recorded and physical examination was done. The sera and urine for 24 hr were obtained from patients to measure electrolyte levels especially phosphorus. In addition, based on presenting serum phosphate levels, patients were divided into the hypophosphatemia group, less than 2.5 mg/dl; normophosphatemia group, between 2.5-5.2 mg/dl; and hyperphosphatemia group greater than 5.2 mg/dl. **RESULTS**: The mean age of study group was 45 (21-70) years. Thirty-six patients (60%) were in group 1 and 24 patients (40%) were in group 2. No statistically correlation was found between stone recurrence and phosphate levels both in serum and urine. There was only a statistical association of K levels in 24-hour urine samples between group 1 and 2. **CONCLUSION**: There was no significant association between stone recurrence and initial phosphate levels in the serum or in urine. We do not propose to determine phosphate levels routinely in management of patients with stone disease.

**URL:** <https://ovidsp.ovid.com/ovidweb.cgi?T=JS&CSC=Y&NEWS=N&PAGE=fulltext&D=med4&DO=10.1080%2f003655902320783881> <https://libkey.io/libraries/2592/openurl?genre=article&sid=OVID:medline&id=pmid:12487742&id=doi:10.1080%2F003655902320783881&issn=0036-5599&isbn=&volume=36&issue=5&spage=368&pages=368-72&date=2002&title=Scandinavian+Journal+of+Urology+%26+Nephrology&atitle=The+diagnostic+value+of+the+phosphate+levels+in+serum+and+24-hour+urine+samples+in+patients+with+recurrent+renal+stone+disease.&aulast=Muezzinoglu&pid=%3Cauthor%3EMuezzinoglu+T%3BGumus+B%3BSener+E%3BAri+Z%3BBuyuksu+C%3C%2Fauthor%3E%3CAN%3E12487742%3C%2FAN%3E%3CDT%3EComparative+Study%3C%2FDT%3E>

159. **Biochemical distinction between hyperuricosuric calcium urolithiasis and gouty diathesis.**

**Item Type:** Journal Article

**Authors:** Pak, Charles Y. C.;Poindexter, John R.;Peterson, Roy D.;Koska, Janice and Sakhaee, Khashayar

**Publication Date:** No ,2002

**Journal:** Urology 60(5), pp. 789–794

**Abstract: OBJECTIVES**: To determine whether the biochemical presentation and urinary physicochemical environment of patients with hyperuricosuria presenting with calcium stones (hyperuricosuric calcium urolithiasis [HUCU]) differs from those of patients with gouty diathesis (GD) or idiopathic uric acid urolithiasis. **METHODS**: A total of 122 patients with HUCU and 68 patients with GD were identified from our "stone registry" of patients who underwent a full ambulatory evaluation. All patients with HUCU had urinary uric acid greater than 800 mg/day in men and greater than 750 mg/day in women and presented with calcium stones. Those with GD had pure uric acid stones or mixed uric acid-calcium stones and did not have secondary causes of uric acid urolithiasis. Data derived from the fasting serum and 24-hour urine samples collected on a random diet and on a diet restricted in calcium, sodium, and oxalate were compared between the two groups. **RESULTS**: Compared with patients with HUCU, those with GD had significantly higher serum uric acid and lower urinary uric acid and pH levels (mean value 5.38 and 5.35 on random and restricted diets versus 6.09 and 6.14, respectively). The fractional excretion of urate and the discriminant score of the relationship between urinary pH and the fractional excretion of urate were significantly lower in those with GD than in those with HUCU. Patients with HUCU displayed a greater urinary saturation of sodium urate and calcium oxalate compared with those with GD, and those with GD had a higher urinary content of undissociated uric acid and lower urinary saturation of brushite (calcium phosphate). **CONCLUSIONS**: Patients with HUCU presented with normal urinary pH and hyperuricosuria, accompanied sometimes by hypercalciuria, which produced increased urinary saturation of sodium urate and calcium oxalate. In contrast, those with GD had a low fractional excretion of urate (that contributed to hyperuricemia) and low urinary pH (that led to increased amount of undissociated uric acid). The varying biochemical and physicochemical presentations of the two conditions can be ascribed to overindulgence with purine-rich foods in those with HUCU and underlying primary gout in those with GD.

**URL:** <https://ovidsp.ovid.com/ovidweb.cgi?T=JS&CSC=Y&NEWS=N&PAGE=fulltext&D=med4&DO=10.1016%2fs0090-4295%2802%2901908-8> <https://libkey.io/libraries/2592/openurl?genre=article&sid=OVID:medline&id=pmid:12429297&id=doi:10.1016%2Fs0090-4295%2802%2901908-8&issn=0090-4295&isbn=&volume=60&issue=5&spage=789&pages=789-94&date=2002&title=Urology&atitle=Biochemical+distinction+between+hyperuricosuric+calcium+urolithiasis+and+gouty+diathesis.&aulast=Pak&pid=%3Cauthor%3EPak+CY%3BPoindexter+JR%3BPeterson+RD%3BKoska+J%3BSakhaee+K%3C%2Fauthor%3E%3CAN%3E12429297%3C%2FAN%3E%3CDT%3EJournal+Article%3C%2FDT%3E>

160. **Urinary citrate excretion in idiopathic nephrolithiasis.**

**Item Type:** Journal Article

**Authors:** Ratan, Simmi K.;Bhatnagar, V.;Mitra, D. K.;Basu, N. and Malhotra, L. K.

**Publication Date:** Se ,2002

**Journal:** Indian Pediatrics 39(9), pp. 819–825

**Abstract: OBJECTIVE**: To determine urinary citrate excretion in children with nephrolithiasis and normal controls. **DESIGN**: Prospective. **SETTING**: Tertiary care center in New Delhi. **METHODS**: This study was done on 50 children, below the age of 12 years, with idiopathic urinary calculi and 150 age and weight matched controls. The children were divided into 3 groups: Group 1 (1-4 years), Group 2 (5-8 years) and Group 3 (9-12 years). Urinary citrate was estimated in a 24-hour urine sample using colorimetric method. The stones removed from these children were also analysed. **RESULTS**: There was a preponderance of urinary stones in males; the highest incidence being in Group 1. Excretion of citrate in 24-hour urine sample was significantly lower in patients compared to controls, for males in all age groups and for females in Group 3. However, there was no statistically significant difference in the urinary citrate value between males and females in a given age group for either controls or patients. The urinary citrate excretion increased with age in patients and controls, but the levels in patients were lower. Depending upon the constituents, four types of stones were identified, calcium phosphate, calcium oxalate, uric acid and magnesium ammonium phosphate. Nine stones had at least more than one major constituent. Hypocitraturia was detected in 43 percent cases. The incidence was 76 percent for calcium phosphate, 87 percent for calcium oxalate, 40 percent for uric acid stones and 50 percent for magnesium ammonium phosphate. **CONCLUSION**: This study shows that low urinary citrate is associated with urinary stones in children, especially in endemic areas, in the absence of obvious etiological factors. Urinary citrate excretion should be determined in all children with nephrolithiasis.

**URL:** <https://ovidsp.ovid.com/ovidweb.cgi?T=JS&CSC=Y&NEWS=N&PAGE=fulltext&D=med4&AN=12368525> <https://libkey.io/libraries/2592/openurl?genre=article&sid=OVID:medline&id=pmid:12368525&id=doi:&issn=0019-6061&isbn=&volume=39&issue=9&spage=819&pages=819-25&date=2002&title=Indian+Pediatrics&atitle=Urinary+citrate+excretion+in+idiopathic+nephrolithiasis.&aulast=Ratan&pid=%3Cauthor%3ERatan+SK%3BBhatnagar+V%3BMitra+DK%3BBasu+N%3BMalhotra+LK%3C%2Fauthor%3E%3CAN%3E12368525%3C%2FAN%3E%3CDT%3EJournal+Article%3C%2FDT%3E>

161. **Effects of 5 different diets on urinary risk factors for calcium oxalate kidney stone formation: evidence of different renal handling mechanisms in different race groups.**

**Item Type:** Journal Article

**Authors:** Rodgers, Allen L. and Lewandowski, Sonja

**Publication Date:** Se ,2002

**Journal:** Journal of Urology 168(3), pp. 931–936

**Abstract: PURPOSE**: Since the incidence of renal calculi in the South African black population is extremely rare while in white subjects it occurs at the same rate as elsewhere in the western world, we investigated the possibility that different renal handling mechanisms in response to different dietary challenges might occur in the 2 race groups. **MATERIALS AND METHODS**: We administered 5 different dietary protocols, including low calcium, high oxalate, vitamin C, high salt and lacto-vegetarian, to 10 healthy male subjects from each race group. We collected 24-hour urine at baseline and after 4 days on the prescribed diet which were analyzed for biochemical and physicochemical risk factors. Dietary intake was controlled throughout the experimental period. A 24-hour dietary recall questionnaire was recorded at baseline and analyzed using food composition tables. Statistical analysis of variance was performed on all the data. **RESULTS**: The low calcium diet caused statistically significant changes only in black subjects, which consisted of urinary oxalate increase (0.17 to 0.23 mmol./24 hours, p = 0.01), relative supersaturation of calcium oxalate decrease (1.88 to 0.97, p = 0.03) and relative supersaturation of brushite increase (0.85 to 1.69, p = 0.03). The high oxalate diet caused statistically significant changes in both race groups but these changes were different in the 2 groups. In white subjects urinary pH increased (6.24 to 6.62, p = 0.01), potassium excretion increased (40.01 to 73.49, p = 0.01) and relative supersaturation of brushite increased (1.34 to 2.12, p = 0.05). In black subjects urinary citrate increased (1.94 to 2.99 mmol./24 hours, p = 0.01). Clinically unimportant changes occurred in both race groups after the other 3 diets. **CONCLUSIONS**: Renal handling of dietary calcium and oxalate in South African black and white subjects is different and may explain the different stone incidence in the 2 race groups.

**URL:** <https://ovidsp.ovid.com/ovidweb.cgi?T=JS&CSC=Y&NEWS=N&PAGE=fulltext&D=med4&DO=10.1016%2fS0022-5347%2805%2964545-4> <https://libkey.io/libraries/2592/openurl?genre=article&sid=OVID:medline&id=pmid:12187193&id=doi:10.1016%2FS0022-5347%2805%2964545-4&issn=0022-5347&isbn=&volume=168&issue=3&spage=931&pages=931-6&date=2002&title=Journal+of+Urology&atitle=Effects+of+5+different+diets+on+urinary+risk+factors+for+calcium+oxalate+kidney+stone+formation%3A+evidence+of+different+renal+handling+mechanisms+in+different+race+groups.&aulast=Rodgers&pid=%3Cauthor%3ERodgers+AL%3BLewandowski+S%3C%2Fauthor%3E%3CAN%3E12187193%3C%2FAN%3E%3CDT%3EComparative+Study%3C%2FDT%3E>

162. **Proportional reduction of urine supersaturation during nephrolithiasis treatment.**

**Item Type:** Journal Article

**Authors:** Coe, F. L.;Wise, H.;Parks, J. H. and Asplin, J. R.

**Publication Date:** Oc ,2001

**Journal:** Journal of Urology 166(4), pp. 1247–1251

**Abstract: PURPOSE**: During metabolic stone therapy, urine supersaturation decreases in proportion to pretreatment levels. We gauge the quantitative contribution of regression to the mean for reducing urine supersaturation from high pretreatment to lower values during therapy. **MATERIALS AND METHODS**: The 24-hour urine supersaturations for calcium oxalate, calcium phosphate and uric acid were measured on 2 pretreatment and at least 1 treatment 24-hour collection for each of the 2,667 patients in 2 networks and at a university based specialty clinic. Changes in supersaturation between the first and second pretreatment collections were an estimate of random change and compared to therapeutic changes. **RESULTS**: Supersaturations decreased between the first and second pretreatment collections, proportional to the supersaturation in the first collection. However, the magnitude of this effect was minor compared to therapeutic changes. Also, mean change between pretreatment collections was 0, whereas mean change with therapy was greater than 0 for all 3 supersaturations. **CONCLUSIONS**: Although regression to the mean can be detected, it cannot be responsible for the decrease in urine supersaturation with therapy or the fact that the decrease is proportional to pretreatment mean supersaturation. The mechanisms responsible for proportional reduction remain to be clarified.

**URL:** <https://ovidsp.ovid.com/ovidweb.cgi?T=JS&CSC=Y&NEWS=N&PAGE=fulltext&D=med4&AN=11547051> <https://libkey.io/libraries/2592/openurl?genre=article&sid=OVID:medline&id=pmid:11547051&id=doi:&issn=0022-5347&isbn=&volume=166&issue=4&spage=1247&pages=1247-51&date=2001&title=Journal+of+Urology&atitle=Proportional+reduction+of+urine+supersaturation+during+nephrolithiasis+treatment.&aulast=Coe&pid=%3Cauthor%3ECoe+FL%3BWise+H%3BParks+JH%3BAsplin+JR%3C%2Fauthor%3E%3CAN%3E11547051%3C%2FAN%3E%3CDT%3EJournal+Article%3C%2FDT%3E>

163. **Contribution of dietary oxalate to urinary oxalate excretion.**

**Item Type:** Journal Article

**Authors:** Holmes, R. P.;Goodman, H. O. and Assimos, D. G.

**Publication Date:** Ja ,2001

**Journal:** Kidney International 59(1), pp. 270–276

**Abstract: BACKGROUND**: The amount of oxalate excreted in urine has a significant impact on calcium oxalate supersaturation and stone formation. Dietary oxalate is believed to make only a minor (10 to 20%) contribution to the amount of oxalate excreted in urine, but the validity of the experimental observations that support this conclusion can be questioned. An understanding of the actual contribution of dietary oxalate to urinary oxalate excretion is important, as it is potentially modifiable. **METHODS**: We varied the amount of dietary oxalate consumed by a group of adult individuals using formula diets and controlled, solid-food diets with a known oxalate content, determined by a recently developed analytical procedure. Controlled solid-food diets were consumed containing 10, 50, and 250 mg of oxalate/2500 kcal, as well as formula diets containing 0 and 180 mg oxalate/2500 kcal. Changes in the content of oxalate and other ions were assessed in 24-hour urine collections. **RESULTS**: Urinary oxalate excretion increased as dietary oxalate intake increased. With oxalate-containing diets, the mean contribution of dietary oxalate to urinary oxalate excretion ranged from 24.4 +/- 15.5% on the 10 mg/2500 kcal/day diet to 41.5 +/- 9.1% on the 250 mg/2500 kcal/day diet, much higher than previously estimated. When the calcium content of a diet containing 250 mg of oxalate was reduced from 1002 mg to 391 mg, urinary oxalate excretion increased by a mean of 28.2 +/- 4.8%, and the mean dietary contribution increased to 52.6 +/- 8.6%. **CONCLUSIONS**: These results suggest that dietary oxalate makes a much greater contribution to urinary oxalate excretion than previously recognized, that dietary calcium influences the bioavailability of ingested oxalate, and that the absorption of dietary oxalate may be an important factor in calcium oxalate stone formation.

**URL:** <https://ovidsp.ovid.com/ovidweb.cgi?T=JS&CSC=Y&NEWS=N&PAGE=fulltext&D=med4&DO=10.1046%2fj.1523-1755.2001.00488.x> <https://libkey.io/libraries/2592/openurl?genre=article&sid=OVID:medline&id=pmid:11135080&id=doi:10.1046%2Fj.1523-1755.2001.00488.x&issn=0085-2538&isbn=&volume=59&issue=1&spage=270&pages=270-6&date=2001&title=Kidney+International&atitle=Contribution+of+dietary+oxalate+to+urinary+oxalate+excretion.&aulast=Holmes&pid=%3Cauthor%3EHolmes+RP%3BGoodman+HO%3BAssimos+DG%3C%2Fauthor%3E%3CAN%3E11135080%3C%2FAN%3E%3CDT%3EJournal+Article%3C%2FDT%3E>

164. **Strong inhibition of crystal-cell attachment by pediatric urinary macromolecules: a close relationship with high urinary citrate secretion.**

**Item Type:** Journal Article

**Authors:** Miyake, O.;Kakimoto, K.;Tsujihata, M.;Yoshimura, K.;Takahara, S. and Okuyama, A.

**Publication Date:** Se ,2001

**Journal:** Urology 58(3), pp. 493–497

**Abstract: OBJECTIVES**: To investigate other reasons for the low incidence of pediatric urolithiasis, we evaluated the difference in the crystal-cell adhesion inhibitory activity of urinary macromolecules (UMMs) between children and adults. We also evaluated whether citrates influence the above inhibitory activity, because citrates are important in pediatric urine. **METHODS**: Urine samples were collected from children and healthy male adults during a 24-hour period, and urinary components with a molecular weight of 3 kDa or greater were extracted as UMMs to compare their inhibitory activity during the adhesion of calcium oxalate monohydrate crystals to cultured Madin-Darby canine kidney cells between children and adults. Subsequently, various concentrations of citrates were added to adult UMMs to evaluate the changes in the crystal-cell adhesion inhibitory activity of UMMs. **RESULTS**: Pediatric UMMs more strongly inhibited the adhesion of calcium oxalate monohydrate crystals to cultured Madin-Darby canine kidney cells at a concentration of 0.1 mg/mL compared with adult UMMs. In addition, pediatric UMMs contained higher proportions of fibronectin and glycosaminoglycans, both of which exhibit crystal-cell adhesion inhibitory activity. When citrates were added to adult UMMs, the crystal-cell adhesion inhibitory activity of UMMs was increased in a dose-dependent manner. However, citrates alone did not result in any differences in the inhibitory activity at any of the three different concentrations. **CONCLUSIONS**: We speculate that the incidence of pediatric urolithiasis is low because pediatric UMMs more potently inhibit the adhesion of calcium oxalate crystals to renal tubular cells or because the higher proportion of citrates in pediatric urine enhances the crystal-cell adhesion inhibitory activity of UMMs in a dose-dependent manner.

**URL:** <https://ovidsp.ovid.com/ovidweb.cgi?T=JS&CSC=Y&NEWS=N&PAGE=fulltext&D=med4&DO=10.1016%2fs0090-4295%2801%2901257-2> <https://libkey.io/libraries/2592/openurl?genre=article&sid=OVID:medline&id=pmid:11549512&id=doi:10.1016%2Fs0090-4295%2801%2901257-2&issn=0090-4295&isbn=&volume=58&issue=3&spage=493&pages=493-7&date=2001&title=Urology&atitle=Strong+inhibition+of+crystal-cell+attachment+by+pediatric+urinary+macromolecules%3A+a+close+relationship+with+high+urinary+citrate+secretion.&aulast=Miyake&pid=%3Cauthor%3EMiyake+O%3BKakimoto+K%3BTsujihata+M%3BYoshimura+K%3BTakahara+S%3BOkuyama+A%3C%2Fauthor%3E%3CAN%3E11549512%3C%2FAN%3E%3CDT%3EComparative+Study%3C%2FDT%3E>

165. **Adequacy of a single stone risk analysis in the medical evaluation of urolithiasis.**

**Item Type:** Journal Article

**Authors:** Pak, C. Y.;Peterson, R. and Poindexter, J. R.

**Publication Date:** Fe ,2001

**Journal:** Journal of Urology 165(2), pp. 378–381

**Abstract: PURPOSE**: We tested the hypothesis that a single 24-hour urine sample for stone risk analysis would be sufficient for the simplified medical evaluation of urolithiasis. **MATERIALS AND METHODS**: We retrospectively analyzed stone risk profile data on 24-hour urine samples obtained during random and restricted diets in 225 patients with recurrent urolithiasis. **RESULTS**: In 2 random samples we noted no significant difference in urinary calcium, oxalate, uric acid, citrate, pH, total volume, sodium, potassium, sulphate or phosphorus. For these risk factors there was a highly significant positive correlation in the 2 random samples (r > or = 0.68, p : In 2 random samples we noted no significant difference in urinary calcium, oxalate, uric acid, citrate, pH, total volume, sodium, potassium, sulphate or phosphorus. For these risk factors there was a highly significant positive correlation in the 2 random samples (r > or = 0.68, p **CONCLUSIONS**: The reproducibility of urinary stone risk factors is satisfactory in repeat urine samples. A single stone risk analysis is sufficient for the simplified medical evaluation of urolithiasis.

**URL:** <https://ovidsp.ovid.com/ovidweb.cgi?T=JS&CSC=Y&NEWS=N&PAGE=fulltext&D=med4&DO=10.1097%2f00005392-200102000-00006> <https://libkey.io/libraries/2592/openurl?genre=article&sid=OVID:medline&id=pmid:11176377&id=doi:10.1097%2F00005392-200102000-00006&issn=0022-5347&isbn=&volume=165&issue=2&spage=378&pages=378-81&date=2001&title=Journal+of+Urology&atitle=Adequacy+of+a+single+stone+risk+analysis+in+the+medical+evaluation+of+urolithiasis.&aulast=Pak&pid=%3Cauthor%3EPak+CY%3BPeterson+R%3BPoindexter+JR%3C%2Fauthor%3E%3CAN%3E11176377%3C%2FAN%3E%3CDT%3EJournal+Article%3C%2FDT%3E>

166. **Dietary supplementation with cranberry concentrate tablets may increase the risk of nephrolithiasis.**

**Item Type:** Journal Article

**Authors:** Terris, M. K.;Issa, M. M. and Tacker, J. R.

**Publication Date:** Ja ,2001

**Journal:** Urology 57(1), pp. 26–29

**Abstract: OBJECTIVES**: Cranberry juice has been recommended for patients with recurrent urinary tract infections. However, cranberry juice has a moderately high concentration of oxalate, a common component of kidney stones, and should be limited in patients with a history of nephrolithiasis. Cranberry concentrate tablets are currently available at nutrition stores and are sold as promoters of urinary tract health. After one of our patients with a distant history of calcium oxalate nephrolithiasis developed recurrent stones following self-administration of cranberry concentrate tablets, we sought to investigate the potential lithogenic properties of cranberry supplements. **METHODS**: Five healthy volunteers on a normal diet provided 24-hour urine collection for pH, volume, creatinine, oxalate, calcium, phosphate, uric acid, sodium, citrate, magnesium, and potassium. Cranberry tablets were administered to these volunteers at the manufacturer's recommended dosage for 7 days. On the seventh day, a second 24-hour urine collection was obtained. **RESULTS**: The urinary oxalate levels in the volunteers significantly increased (P = 0.01) by an average of 43.4% while receiving cranberry tablets. The excretion of potential lithogenic ions calcium, phosphate, and sodium also increased. However, inhibitors of stone formation, magnesium and potassium, rose as well. **CONCLUSIONS**: Cranberry concentrate tablets are marketed for urinary tract ailments. Physicians and manufacturers of cranberry products should make an effort to educate patients at risk for nephrolithiasis against ingestion of these dietary supplements.

**URL:** <https://ovidsp.ovid.com/ovidweb.cgi?T=JS&CSC=Y&NEWS=N&PAGE=fulltext&D=med4&DO=10.1016%2fs0090-4295%2800%2900884-0> <https://libkey.io/libraries/2592/openurl?genre=article&sid=OVID:medline&id=pmid:11164137&id=doi:10.1016%2Fs0090-4295%2800%2900884-0&issn=0090-4295&isbn=&volume=57&issue=1&spage=26&pages=26-9&date=2001&title=Urology&atitle=Dietary+supplementation+with+cranberry+concentrate+tablets+may+increase+the+risk+of+nephrolithiasis.&aulast=Terris&pid=%3Cauthor%3ETerris+MK%3BIssa+MM%3BTacker+JR%3C%2Fauthor%3E%3CAN%3E11164137%3C%2FAN%3E%3CDT%3EJournal+Article%3C%2FDT%3E>

167. **Effect of potential renal acid load of foods on calcium metabolism of renal calcium stone formers.**

**Item Type:** Journal Article

**Authors:** Trinchieri, A.;Zanetti, G.;Curro, A. and Lizzano, R.

**Publication Date:** 2001

**Journal:** European Urology 39(Suppl 2), pp. 33–36

**Abstract: OBJECTIVES**: Diet has been proposed as a causative factor of hypercalciuria in patients with calcium stones. The aim of this study was to investigate the influence of diet on calcium metabolism of renal stone formers. **METHODS**: Thirty-five renal calcium stone formers were entered in this study. A 2-day recall of dietary intake was obtained from each subject. The food records were coded and computer analyzed for total energy, protein, fat, carbohydrate, sodium, potassium, calcium, magnesium, phosphate, oxalate, vitamin C and fiber. Daily potential renal acid load (PRAL) of the diet was calculated considering the mineral and protein composition of foods, the mean intestinal absorption rate for each nutrient and the metabolism of sulfur-containing amino acids. A fasting blood sample was drawn and a 24-hour urine collection were obtained for analyses of calcium, phosphate and creatinine. Serum osteocalcin was also analyzed. A fasting 2-hour urine sample was collected in the morning for hydroxyproline, pyridinium cross-links and creatinine. **RESULTS**: The mean daily dietary PRAL of renal stone formers was 22.4 +/- 15.7 (range 4.2-65.8) mEq/day. Regression analysis demonstrated that urinary calcium excretion is dependent on daily protein intake and dietary PRAL, whereas the urinary pyridinium cross-links/creatinine ratio is inversely dependent on daily calcium intake. The urinary pyridinium cross-links/creatinine ratio was significantly lower in patients on a low calcium diet (: The mean daily dietary PRAL of renal stone formers was 22.4 +/- 15.7 (range 4.2-65.8) mEq/day. Regression analysis demonstrated that urinary calcium excretion is dependent on daily protein intake and dietary PRAL, whereas the urinary pyridinium cross-links/creatinine ratio is inversely dependent on daily calcium intake. The urinary pyridinium cross-links/creatinine ratio was significantly lower in patients on a low calcium diet (**CONCLUSIONS**: The urinary calcium excretion of renal stone formers seems to be dependent on dietary acid load rather than dietary calcium intake. In patients consuming an acidifying diet a restriction of calcium intake could increase bone resorption leading to a progressive bone loss.

**URL:** <https://ovidsp.ovid.com/ovidweb.cgi?T=JS&CSC=Y&NEWS=N&PAGE=fulltext&D=med4&DO=10.1159%2f000052556> <https://libkey.io/libraries/2592/openurl?genre=article&sid=OVID:medline&id=pmid:11223695&id=doi:10.1159%2F000052556&issn=0302-2838&isbn=&volume=39&issue=2&spage=33&pages=33-6%3B+discussion+36-7&date=2001&title=European+Urology&atitle=Effect+of+potential+renal+acid+load+of+foods+on+calcium+metabolism+of+renal+calcium+stone+formers.&aulast=Trinchieri&pid=%3Cauthor%3ETrinchieri+A%3BZanetti+G%3BCurro+A%3BLizzano+R%3C%2Fauthor%3E%3CAN%3E11223695%3C%2FAN%3E%3CDT%3EJournal+Article%3C%2FDT%3E>

168. **The risk of renal stone formation during and after long duration space flight.**

**Item Type:** Journal Article

**Authors:** Whitson, P. A.;Pietrzyk, R. A.;Morukov, B. V. and Sams, C. F.

**Publication Date:** No ,2001

**Journal:** Nephron 89(3), pp. 264–270

**Abstract: BACKGROUND**: The formation of a renal stone during space flight may have serious negative effects on the health of the crewmember and the success of the mission. Urinary biochemical factors and the influence of dietary factors associated with renal stone development were assessed during long duration Mir Space Station missions. **METHODS**: Twenty-four-hour urine samples were collected prior to, during and following long duration space flight. The relative urinary supersaturation of calcium oxalate, calcium phosphate (brushite), sodium urate, struvite and uric acid were determined. **RESULTS**: Changes in the urinary biochemistry of crewmembers during long duration spaceflight demonstrated increases in the supersaturation of the stone-forming salts. In-flight hypercalciuria was evident in a number of individual crewmembers and 24-hour dietary fluid intake and urine volume were significantly lower. During flight, there was a significant increase in brushite supersaturation. **CONCLUSIONS**: These data suggest acute effects of space flight and postflight changes in the urinary biochemistry favoring increased crystallization in the urine. The effects of dietary intake, especially fluid intake, may have a significant impact on the potential for renal stone formation. Efforts are now underway to assess the efficacy of a countermeasure to mitigate the increased risk. Copyright 2001 S. Karger AG, Basel

**URL:** <https://ovidsp.ovid.com/ovidweb.cgi?T=JS&CSC=Y&NEWS=N&PAGE=fulltext&D=med4&DO=10.1159%2f000046083> <https://libkey.io/libraries/2592/openurl?genre=article&sid=OVID:medline&id=pmid:11598387&id=doi:10.1159%2F000046083&issn=1660-8151&isbn=&volume=89&issue=3&spage=264&pages=264-70&date=2001&title=Nephron&atitle=The+risk+of+renal+stone+formation+during+and+after+long+duration+space+flight.&aulast=Whitson&pid=%3Cauthor%3EWhitson+PA%3BPietrzyk+RA%3BMorukov+BV%3BSams+CF%3C%2Fauthor%3E%3CAN%3E11598387%3C%2FAN%3E%3CDT%3EJournal+Article%3C%2FDT%3E>

169. **Why oral calcium supplements may reduce renal stone disease: report of a clinical pilot study.**

**Item Type:** Journal Article

**Authors:** Williams, C. P.;Child, D. F.;Hudson, P. R.;Davies, G. K.;Davies, M. G.;John, R.;Anandaram, P. S. and De Bolla, A. R.

**Publication Date:** Ja ,2001

**Journal:** Journal of Clinical Pathology 54(1), pp. 54–62

**Abstract: AIMS**: To investigate whether increasing the daily baseline of gut calcium can cause a gradual downregulation of the active intestinal transport of calcium via reduced parathyroid hormone (PTH) mediated activation of vitamin D, and to discuss why such a mechanism might prevent calcium oxalate rich stones. To demonstrate the importance of seasonal effects upon the evaluation of such data. **METHODS**: Within an intensive 24 hour urine collection regimen, daily calcium supplementation (500 mg) was given to five stone formers for a 10 week period during a six month crossover study. In a further population of patients on follow up for previous renal stone disease, observations were made on 1066 24 hour urine samples collected over five years in respect of seasonal effects relevant to the interpretation of the study. **RESULTS**: In the group of patients on calcium supplements the following results were found. During calcium supplementation, the proportion of urine calcium to oxalate was higher (increased calcium to oxalate molar ratio), the 24 hour urine product of calcium and oxalate did not rise, and urine oxalate was lower during the first six weeks of supplementation. Twenty four hour urine calcium was 10.2% higher than baseline in the final four weeks of the 10 weeks of supplementation. Twenty four hour urine phosphate was 11.4% lower during the first six weeks of supplementation, but then rose while the patients were still on supplementation; renal tubular reabsorption of phosphate (TmP/GFR) mirrored the urine phosphate changes inversely. PTH was higher after stopping supplementation, but 1,25-(OH)2-cholecalciferol changes were not detected. In the 1066 urine samples collected over five years the following results were found. Calcium and oxalate excretion correlated positively and not inversely. Urine calcium and phosphate excretion were 5.5% and 2.5% higher, respectively, in "light" months of the year compared with "dark" months. A post summer decline in both urine calcium and urine phosphate was relevant to the interpretation of the study. **CONCLUSIONS**: Regular calcium supplementation does not raise the product of calcium and oxalate in urine and the proportion of oxalate to calcium is reduced. The underlying mechanisms of the changes seen in phosphate, calcium, and PTH and the observations on 1,25-(OH)2-cholecalciferol are not clear. Observed changes in phosphate could possibly be part of a calcium regulating feedback loop operating over a period of weeks. In evaluating these mechanisms background seasonal effects are important. It is possible that "programming" of the gut mucosa in terms of calcium transport is a major determinant of the relation between calcium and oxalate concentrations in urine and their relative abundance. Increased oral calcium, in association with a reduction of the relative proportion absorbed, may be pertinent to the prevention of calcium oxalate rich stones.

**URL:** <https://ovidsp.ovid.com/ovidweb.cgi?T=JS&CSC=Y&NEWS=N&PAGE=fulltext&D=med4&DO=10.1136%2fjcp.54.1.54> <https://libkey.io/libraries/2592/openurl?genre=article&sid=OVID:medline&id=pmid:11271790&id=doi:10.1136%2Fjcp.54.1.54&issn=0021-9746&isbn=&volume=54&issue=1&spage=54&pages=54-62&date=2001&title=Journal+of+Clinical+Pathology&atitle=Why+oral+calcium+supplements+may+reduce+renal+stone+disease%3A+report+of+a+clinical+pilot+study.&aulast=Williams&pid=%3Cauthor%3EWilliams+CP%3BChild+DF%3BHudson+PR%3BDavies+GK%3BDavies+MG%3BJohn+R%3BAnandaram+PS%3BDe+Bolla+AR%3C%2Fauthor%3E%3CAN%3E11271790%3C%2FAN%3E%3CDT%3EClinical+Trial%3C%2FDT%3E>

170. **Higher urinary potassium is associated with decreased stone growth after shock wave lithotripsy.**

**Item Type:** Journal Article

**Authors:** Pierratos, A.;Dharamsi, N.;Carr, L. K.;Ibanez, D.;Jewett, M. A. and Honey, R. J.

**Publication Date:** No ,2000

**Journal:** Journal of Urology 164(5), pp. 1486–1489

**Abstract: PURPOSE**: We correlated serum and urinary biochemical parameters with radiological evidence of stone growth after shock wave lithotripsy. **MATERIALS AND METHODS**: Biochemical parameters in serum and 24-hour urine collections of 359 patients were correlated with stone growth for 2 years after shock wave lithotripsy. Each patient underwent a minimum of 2 radiological studies at 3 and 12 months and plain abdominal x-ray at 24 months. The presence and size of stones were documented by a radiologist in blinded fashion. Stone growth was defined as measurable growth of a preexisting stone or new stone formation. **RESULTS**: A total of 209 patients remained stone-free or had no existing stone growth, while stone size decreased in 30. Of the remaining 120 patients with stone growth 72 had new growth and 48 had growth of preexisting stones. Urinary excretion of potassium was significantly higher in those without than with stone growth (mean 24-hour urine collection plus or minus standard deviation 62 +/- 27 versus 54 +/- 23 mmol., p = 0.009). The only parameter significantly associated with stone growth was urinary potassium. Linear regression revealed that for each 10 unit increase in urinary potassium there was a corresponding 2 mm. decrease in stone growth (p = 0.013). **CONCLUSIONS**: Our results indicate that increased urinary potassium excretion correlates with a decreased risk of stone growth up to 2 years after shock wave lithotripsy, implying that a high potassium diet may be beneficial for preventing stone growth. The effect of potassium supplementation on stone formation and growth must be investigated further.

**URL:** <https://ovidsp.ovid.com/ovidweb.cgi?T=JS&CSC=Y&NEWS=N&PAGE=fulltext&D=med4&AN=11025688> <https://libkey.io/libraries/2592/openurl?genre=article&sid=OVID:medline&id=pmid:11025688&id=doi:&issn=0022-5347&isbn=&volume=164&issue=5&spage=1486&pages=1486-9&date=2000&title=Journal+of+Urology&atitle=Higher+urinary+potassium+is+associated+with+decreased+stone+growth+after+shock+wave+lithotripsy.&aulast=Pierratos&pid=%3Cauthor%3EPierratos+A%3BDharamsi+N%3BCarr+LK%3BIbanez+D%3BJewett+MA%3BHoney+RJ%3C%2Fauthor%3E%3CAN%3E11025688%3C%2FAN%3E%3CDT%3EJournal+Article%3C%2FDT%3E>

171. **Effects of low animal protein or high-fiber diets on urine composition in calcium nephrolithiasis.**

**Item Type:** Journal Article

**Authors:** Rotily, M.;Leonetti, F.;Iovanna, C.;Berthezene, P.;Dupuy, P.;Vazi, A. and Berland, Y.

**Publication Date:** Ma ,2000

**Journal:** Kidney International 57(3), pp. 1115–1123

**Abstract: BACKGROUND**: The purpose of this article is to evaluate the impact of low protein and high fiber intakes on risk factors of stone recurrence in idiopathic calcium stone formers (ICSFs). **METHODS**: Ninety-six ICSFs were randomly assigned a low animal protein diet ( 25 g/day), or a usual diet (control group); all patients were recommended to increase their fluid intake. Their daily urine compositions were analyzed at baseline and at four months. Compliance with dietary recommendations was checked by validated food frequency questionnaires. Compliance with total and animal protein intakes was assessed by 24-hour urea and sulphate outputs, respectively. The nutritional intervention (oral instructions, written leaflet, phoning) and food assessment were carried out by a research dietitian. **RESULTS**: At baseline, diets and the daily urine composition did not differ between the three groups. At four months, while diets differed significantly, the 24-hour output of calcium and oxalate did not differ significantly within and between groups after adjustment for potential confounders (age, sex, and personal and family history of calcium stones) and baseline values. However, as many as 12 out of 31 ICSFs (95% CI, 22 to 58%) assigned to a low animal protein diet achieved a reduction in the urine urea excretion rate of more than 50 mmol/day and also exhibited a significant decrease in urinary calcium excretion that averaged 1.8 mmol/day. A significant correlation between urea and calcium outputs was observed only among patients with hypercalciuria. **CONCLUSIONS**: These results show that only ICSFs who markedly decrease their animal protein intake, especially those with hypercalciuria, can expect to benefit from dietary recommendations.

**URL:** <https://ovidsp.ovid.com/ovidweb.cgi?T=JS&CSC=Y&NEWS=N&PAGE=fulltext&D=med4&DO=10.1046%2fj.1523-1755.2000.00939.x> <https://libkey.io/libraries/2592/openurl?genre=article&sid=OVID:medline&id=pmid:10720964&id=doi:10.1046%2Fj.1523-1755.2000.00939.x&issn=0085-2538&isbn=&volume=57&issue=3&spage=1115&pages=1115-23&date=2000&title=Kidney+International&atitle=Effects+of+low+animal+protein+or+high-fiber+diets+on+urine+composition+in+calcium+nephrolithiasis.&aulast=Rotily&pid=%3Cauthor%3ERotily+M%3BLeonetti+F%3BIovanna+C%3BBerthezene+P%3BDupuy+P%3BVazi+A%3BBerland+Y%3C%2Fauthor%3E%3CAN%3E10720964%3C%2FAN%3E%3CDT%3EClinical+Trial%3C%2FDT%3E>

172. **Tamm-Horsfall protein excretion and its relation to citrate in urine of stone-forming patients.**

**Item Type:** Journal Article

**Authors:** Ganter, K.;Bongartz, D. and Hesse, A.

**Publication Date:** Ma ,1999

**Journal:** Urology 53(3), pp. 492–495

**Abstract: OBJECTIVES**: To evaluate the relation of Tamm-Horsfall protein (THP) and citrate, both potent actors in the urinary stone forming process. **METHODS**: Quantitative determination of THP in calcium oxalate (CaOx) stone-forming patients and healthy subjects was carried out according to the enzyme-linked immunosorbent assay method. **RESULTS**: THP excretion in 24-hour urine samples of CaOx stone-forming patients was significantly reduced compared with healthy subjects. A significant correlation exists between the concentration of THP and citrate in the stone-forming group, as well as in the group of healthy subjects, and for the 24-hour excretion, this correlation persists in the group of CaOx stone-forming patients. **CONCLUSIONS**: Decreased THP and citrate excretions were found in CaOx stone-forming patients. They indicate a tubular dysfunction of the distal section.

**URL:** <https://ovidsp.ovid.com/ovidweb.cgi?T=JS&CSC=Y&NEWS=N&PAGE=fulltext&D=med4&DO=10.1016%2fs0090-4295%2898%2900581-0> <https://libkey.io/libraries/2592/openurl?genre=article&sid=OVID:medline&id=pmid:10096372&id=doi:10.1016%2Fs0090-4295%2898%2900581-0&issn=0090-4295&isbn=&volume=53&issue=3&spage=492&pages=492-5&date=1999&title=Urology&atitle=Tamm-Horsfall+protein+excretion+and+its+relation+to+citrate+in+urine+of+stone-forming+patients.&aulast=Ganter&pid=%3Cauthor%3EGanter+K%3BBongartz+D%3BHesse+A%3C%2Fauthor%3E%3CAN%3E10096372%3C%2FAN%3E%3CDT%3EJournal+Article%3C%2FDT%3E>

173. **Effects of a 'common sense diet' on urinary composition and supersaturation in patients with idiopathic calcium urolithiasis.**

**Item Type:** Journal Article

**Authors:** Hess, B.;Mauron, H.;Ackermann, D. and Jaeger, P.

**Publication Date:** Au ,1999

**Journal:** European Urology 36(2), pp. 136–143

**Abstract: OBJECTIVES**: To test whether dietary advice as the only treatment is able to reduce urinary supersaturation in patients with idiopathic calcium urolithiasis. **METHODS**: By means of EQUIL 2, we calculated relative supersaturations of calcium oxalate (RS(CaOx)), brushite (RS(Brushite)), apatite (RS(Apatite)) and uric acid (RS(UA)) of 24-hour urine samples of 68 healthy male controls as well as of 47 male idiopathic calcium stone formers (ICSF) before and after individualized dietary advice (DA). Main goals of DA were (1) to keep urine volume > 2,000 ml/day, and (2) to reduce meat protein intake to 1 g/kg body weight per day. Compliance was judged from changes in urine volume and U(Urea)xV. **RESULTS**: In the whole group of ICSF, DA did not alter supersaturations. Only in those 9 ICSF (19%) with good compliance (increase in volume and decrease in U(Urea)xV), RS(CaOx) and RS(UA) fell by 26 and 49%, respectively. Besides poor compliance, these findings can be explained by positive correlations between changes in volume and U(Urea)xV in ICSF (r = 0. 319, p = 0.037) or U(Na)xV (r = 0.342, p = 0.019). For instance, ICSF with volumes >/= 2,000 ml/day had evidence of elevated protein and salt intake; thus, DA mainly focused on protein and salt intake, but not on volume. This resulted in decreases in U(Urea)xV and U(Na)xV, but also in volume; thus, RS(CaOx) remained unaltered. **CONCLUSIONS**: DA is able to significantly lower RS(CaOx); however, because intakes of fluid and protein are directly and positively linked to each other, this only can be achieved if high fluid and lower meat protein intake are equally stressed in all ICSF.

**URL:** <https://ovidsp.ovid.com/ovidweb.cgi?T=JS&CSC=Y&NEWS=N&PAGE=fulltext&D=med4&DO=10.1159%2f000067985> <https://libkey.io/libraries/2592/openurl?genre=article&sid=OVID:medline&id=pmid:10420035&id=doi:10.1159%2F000067985&issn=0302-2838&isbn=&volume=36&issue=2&spage=136&pages=136-43&date=1999&title=European+Urology&atitle=Effects+of+a+%27common+sense+diet%27+on+urinary+composition+and+supersaturation+in+patients+with+idiopathic+calcium+urolithiasis.&aulast=Hess&pid=%3Cauthor%3EHess+B%3BMauron+H%3BAckermann+D%3BJaeger+P%3C%2Fauthor%3E%3CAN%3E10420035%3C%2FAN%3E%3CDT%3EJournal+Article%3C%2FDT%3E>

174. **Metabolic urinary correlates of calcium oxalate dihydrate in renal stones.**

**Item Type:** Journal Article

**Authors:** Asplin, J. R.;Lingeman, J.;Kahnoski, R.;Mardis, H.;Parks, J. H. and Coe, F. L.

**Publication Date:** Ma ,1998

**Journal:** Journal of Urology 159(3), pp. 664–668

**Abstract: PURPOSE**: We determined what metabolic features of the 24-hour urine predict calcium oxalate dihydrate in kidney stones. Prior studies have suggested that low urine magnesium, high urine calcium, high calcium-to-oxalate ratio and high urine supersaturation with respect to calcium oxalate monohydrate predict calcium oxalate dihydrate. **MATERIALS AND METHODS**: Stone analyses and results from 2, 24-hour pretreatment urine collections from 96 patients with nephrolithiasis were drawn from 3 kidney stone prevention centers. Standard stone risk measurements were made on the urine, including supersaturation for calcium oxalate monohydrate, brushite and uric acid. **RESULTS**: The main differences in metabolic urine findings were between patients with no calcium oxalate dihydrate and those with any calcium oxalate dihydrate in stones. Percent calcium oxalate dihydrate itself did not correlate with urine findings. Patients with no calcium oxalate dihydrate in stones showed a biphasic pattern of urine calcium oxalate monohydrate supersaturation, about half had values below almost any found among patients with calcium oxalate dihydrate in stones (less than 7) and the rest overlapped with the calcium oxalate dihydrate group. Except for higher calcium oxalate monohydrate supersaturation, patients with calcium oxalate dihydrate in stones had higher urine calcium excretion and lower urine citrate concentrations, even after calcium oxalate monohydrate supersaturation was considered. **CONCLUSIONS**: Patients with low calcium oxalate monohydrate supersaturation (less than 7) are unlikely to have calcium oxalate dihydrate in renal stones. However, many patients with no calcium oxalate dihydrate have higher calcium oxalate monohydrate supersaturation values, and so prediction of calcium oxalate dihydrate or its absence from urine findings is imperfect. Urine magnesium and the calcium-to-oxalate ratio are unrelated to calcium oxalate dihydrate.

**URL:** <https://ovidsp.ovid.com/ovidweb.cgi?T=JS&CSC=Y&NEWS=N&PAGE=fulltext&D=med4&AN=9474121> <https://libkey.io/libraries/2592/openurl?genre=article&sid=OVID:medline&id=pmid:9474121&id=doi:&issn=0022-5347&isbn=&volume=159&issue=3&spage=664&pages=664-8&date=1998&title=Journal+of+Urology&atitle=Metabolic+urinary+correlates+of+calcium+oxalate+dihydrate+in+renal+stones.&aulast=Asplin&pid=%3Cauthor%3EAsplin+JR%3BLingeman+J%3BKahnoski+R%3BMardis+H%3BParks+JH%3BCoe+FL%3C%2Fauthor%3E%3CAN%3E9474121%3C%2FAN%3E%3CDT%3EJournal+Article%3C%2FDT%3E>

175. **Repeated urine analysis in patients with calcium stone disease.**

**Item Type:** Journal Article

**Authors:** Bek-Jensen, H. and Tiselius, H. G.

**Publication Date:** 1998

**Journal:** European Urology 33(3), pp. 323–332

**Abstract: OBJECTIVE**: To study the variability in urine composition with respect to factors of importance for the calcium salt crystallization process and to test the reliability of using one or several urine samples in the clinical evaluation. **PATIENTS AND METHODS**: Twelve patients collected 16-hour daytime and 8-hour night urine samples during 4 days of the same week. The urine was analysed for calcium, oxalate, phosphate, magnesium, citrate and pH, and the ion activity products of CaOx [AP(CaOx) index] and CaP were calculated. The risk of CaOx crystallization, as well as the inhibition of CaOx crystal growth and aggregation, were assessed. **RESULTS**: There was a good correlation between estimates of the AP(CaOx) index in the different samples, as well as between the AP(CaOx) index and the direct assessment of the risk of CaOx crystallization in the night and daytime urine samples. There was, however, a pronounced intra-individual variation of all variables and parameters. With the assumption that an abnormality would appear in at least one of the four samples, we found that in more than 80% of the cases, two 24-hour (16 + 8 h) urine samples were sufficient to establish whether the patient had a normal or an abnormal urine composition. **CONCLUSION**: Urine samples collected during two 24-, 16- or 8-hour periods appear to be useful for detecting biochemical abnormalities considered of importance for CaOx stone formation.

**URL:** <https://ovidsp.ovid.com/ovidweb.cgi?T=JS&CSC=Y&NEWS=N&PAGE=fulltext&D=med4&DO=10.1159%2f000019568> <https://libkey.io/libraries/2592/openurl?genre=article&sid=OVID:medline&id=pmid:9555561&id=doi:10.1159%2F000019568&issn=0302-2838&isbn=&volume=33&issue=3&spage=323&pages=323-32&date=1998&title=European+Urology&atitle=Repeated+urine+analysis+in+patients+with+calcium+stone+disease.&aulast=Bek-Jensen&pid=%3Cauthor%3EBek-Jensen+H%3BTiselius+HG%3C%2Fauthor%3E%3CAN%3E9555561%3C%2FAN%3E%3CDT%3EJournal+Article%3C%2FDT%3E>

176. **Agglomeration inhibition reflected stone-forming activity during long-term potassium citrate therapy in calcium stone formers.**

**Item Type:** Journal Article

**Authors:** Fuselier, H. A.;Moore, K.;Lindberg, J.;Husserl, F. E.;Cole, F. E.;Kok, D. J.;Whitehead, D.;Galliano, D. J. and Erwin, D. T.

**Publication Date:** De ,1998

**Journal:** Urology 52(6), pp. 988–994

**Abstract: OBJECTIVES**: The agglomeration of preformed crystals of calcium oxalate has been hypothesized to be the rate-limiting step in renal stone-forming activity (SFA). The effect of urine on the in vitro inhibition of agglomeration of seed crystals of calcium oxalate monohydrate, designated [tm], has been used to monitor SFA in calcium oxalate stone formers (CaOxSF). The objective of the present study was to determine whether [tm] could be used to help monitor the long-term effectiveness of oral potassium citrate therapy (K-Cit-Rx) in CaOxSF. **METHODS**: Clinic and radiographic (or ultrasound) reports were evaluated for 80 patients, aged 20 to 72 years, 55 men and 25 women, who were treated with oral K-Cit for recurrent calcium oxalate urolithiasis at the Ochsner Stone Clinic between January 1992 and July 1996. Seventy-five of these patients had at least one 24-hour citrate excretion rate of less than 3.0 mm/day before or after K-Cit-Rx. SFA graded on a scale of -2 to +2 by radiographic criteria was combined with information on stone passage to evaluate clinical stone status, and 24-hour urine collections were evaluated for volume, pH, calcium, citrate, uric acid, oxalate, creatinine, and [tm] on free diet before and after 6 to 53 months of K-Cit-Rx. Historical information on procedures performed for urolithiasis before and on K-Cit-Rx was also reviewed. **RESULTS**: K-Cit-Rx resulted in increased urine pH (P : K-Cit-Rx resulted in increased urine pH (P : K-Cit-Rx resulted in increased urine pH (P : K-Cit-Rx resulted in increased urine pH (P : K-Cit-Rx resulted in increased urine pH (P **CONCLUSIONS**: The data confirm that [tm] can be used not only to verify previously documented stone formation rate but also to help evaluate the long-term effectiveness of therapy. In this report, changes in [tm] after K-Cit-Rx reflected decreased stone formation rate and decreased remedial procedures.

**URL:** <https://ovidsp.ovid.com/ovidweb.cgi?T=JS&CSC=Y&NEWS=N&PAGE=fulltext&D=med4&DO=10.1016%2fs0090-4295%2898%2900361-6> <https://libkey.io/libraries/2592/openurl?genre=article&sid=OVID:medline&id=pmid:9836542&id=doi:10.1016%2Fs0090-4295%2898%2900361-6&issn=0090-4295&isbn=&volume=52&issue=6&spage=988&pages=988-94&date=1998&title=Urology&atitle=Agglomeration+inhibition+reflected+stone-forming+activity+during+long-term+potassium+citrate+therapy+in+calcium+stone+formers.&aulast=Fuselier&pid=%3Cauthor%3EFuselier+HA%3BMoore+K%3BLindberg+J%3BHusserl+FE%3BCole+FE%3BKok+DJ%3BWhitehead+D%3BGalliano+DJ%3BErwin+DT%3C%2Fauthor%3E%3CAN%3E9836542%3C%2FAN%3E%3CDT%3EJournal+Article%3C%2FDT%3E>

177. **CLCN5 mutation Ser244Leu is associated with X-linked renal failure without X-linked recessive hypophosphatemic rickets.**

**Item Type:** Journal Article

**Authors:** Kelleher, C. L.;Buckalew, V. M.;Frederickson, E. D.;Rhodes, D. J.;Conner, D. A.;Seidman, J. G. and Seidman, C. E.

**Publication Date:** Ja ,1998

**Journal:** Kidney International 53(1), pp. 31–37

**Abstract:** This study demonstrates that a missense mutation in the voltage gated chloride channel, CLCN5, can cause X-linked renal failure without X-linked recessive hypophosphatemic rickets. A large kindred (Family A), initially evaluated in 1974 with an inherited syndrome characterized by hypercalciuria, nephrocalcinosis, low molecular weight proteinuria, renal tubular acidosis, and renal failure, was clinically re-evaluated and genetically characterized. Medical histories, physical examinations, blood chemistries, and 24-hour urine collections were obtained from 48 family members. Both female and male family members exhibited hypercalciuria, nephrolithiasis, and low molecular weight proteinuria. However, only men developed renal insufficiency, consistent with an X-linked recessive gene defect. Genetic linkage located the disease locus on the proximal short arm of the X chromosome (Xp11) where a voltage gated chloride channel gene, CLCN5, had previously been mapped. DNA sequence of the CLCN5 gene demonstrated a missense mutation (Ser244Leu) in affected family members. The same missense mutation has previously been shown to cause X-linked recessive hypophosphatemic rickets. No affected member of Family A had evidence of chronic hypophosphatemia, clinically significant rickets, or osteomalacia. We hypothesize that genetic background, environment, diet, or an unidentified modifying gene may account for the differing phenotypes resulting from this shared gene defect.

**URL:** <https://ovidsp.ovid.com/ovidweb.cgi?T=JS&CSC=Y&NEWS=N&PAGE=fulltext&D=med4&DO=10.1046%2fj.1523-1755.1998.00752.x> <https://libkey.io/libraries/2592/openurl?genre=article&sid=OVID:medline&id=pmid:9452997&id=doi:10.1046%2Fj.1523-1755.1998.00752.x&issn=0085-2538&isbn=&volume=53&issue=1&spage=31&pages=31-7&date=1998&title=Kidney+International&atitle=CLCN5+mutation+Ser244Leu+is+associated+with+X-linked+renal+failure+without+X-linked+recessive+hypophosphatemic+rickets.&aulast=Kelleher&pid=%3Cauthor%3EKelleher+CL%3BBuckalew+VM%3BFrederickson+ED%3BRhodes+DJ%3BConner+DA%3BSeidman+JG%3BSeidman+CE%3C%2Fauthor%3E%3CAN%3E9452997%3C%2FAN%3E%3CDT%3EJournal+Article%3C%2FDT%3E>

178. **A thermodynamic equilibrium model for calcium salt urolithiasis: clinical application.**

**Item Type:** Journal Article

**Authors:** Ashby, R. and Gyory, A. Z.

**Publication Date:** 1997

**Journal:** Experimental Nephrology 5(3), pp. 246–252

**Abstract:** It is proposed that urine is better modelled as a true equilibrium rather than in a supersaturated/metastable state and that the free citrate3- ion plays a major role in maintaining dispersion of the solid particles (reduced agglomeration). Published urinary chemistries, in conjunction with the computer programme SEQUIL, have been used to formulate a novel risk index for calcium stone formation independent of the traditional clinical classification of the stone former. Applying the risk index to three consecutive 24-hour urine samples of 39 untreated Ca stone formers showed that 35 (90%) patients produced at least one abnormal urine. Traditional single urinary parameter assessment, Ca/Cr, oxalate/Cr or citrate/Cr ratios, showed that only 17, 14, and 18% of the patients, respectively, had an abnormality, while taking all three together 24 (70%) had some abnormality and thus 30% were 'idiopathic' stone formers. Treatment regimens have been devised using the computer programme to return an abnormal urine to the normal according to the proposed risk index. In most urines two or more factors had to be changed simultaneously. Clinically there has been only one recurrence in 36 months, whereas before they had 4.4/3 years.

**URL:** <https://ovidsp.ovid.com/ovidweb.cgi?T=JS&CSC=Y&NEWS=N&PAGE=fulltext&D=med4&AN=9208285> <https://libkey.io/libraries/2592/openurl?genre=article&sid=OVID:medline&id=pmid:9208285&id=doi:&issn=1018-7782&isbn=&volume=5&issue=3&spage=246&pages=246-52&date=1997&title=Experimental+Nephrology&atitle=A+thermodynamic+equilibrium+model+for+calcium+salt+urolithiasis%3A+clinical+application.&aulast=Ashby&pid=%3Cauthor%3EAshby+R%3BGyory+AZ%3C%2Fauthor%3E%3CAN%3E9208285%3C%2FAN%3E%3CDT%3EJournal+Article%3C%2FDT%3E>

179. **Chronopharmacological studies on potassium citrate treatment of oxalocalcic urolithiasis.**

**Item Type:** Journal Article

**Authors:** Grases, F.;Conte, A.;March, J. G.;Garcia-Ferragut, L. and Villalonga, N.

**Publication Date:** 1997

**Journal:** International Urology & Nephrology 29(3), pp. 263–273

**Abstract:** The effect of minimum doses of extradietary potassium citrate ingestion on urolithogenic parameters has been studied. Separate urine fractions were collected in 24-hour periods. Five calcium oxalate stone formers have participated in the study. pH, calcium, citric acid, and the crystallization inhibitory capacity levels in fractional urine samples were determined before and during treatment. The most beneficial effect (increase in citraturia and crystallization inhibitory capacity) was produced by potassium citrate tablets ingested after dinner.

**URL:** <https://ovidsp.ovid.com/ovidweb.cgi?T=JS&CSC=Y&NEWS=N&PAGE=fulltext&D=med4&DO=10.1007%2fBF02550921> <https://libkey.io/libraries/2592/openurl?genre=article&sid=OVID:medline&id=pmid:9285296&id=doi:10.1007%2FBF02550921&issn=0301-1623&isbn=&volume=29&issue=3&spage=263&pages=263-73&date=1997&title=International+Urology+%26+Nephrology&atitle=Chronopharmacological+studies+on+potassium+citrate+treatment+of+oxalocalcic+urolithiasis.&aulast=Grases&pid=%3Cauthor%3EGrases+F%3BConte+A%3BMarch+JG%3BGarcia-Ferragut+L%3BVillalonga+N%3C%2Fauthor%3E%3CAN%3E9285296%3C%2FAN%3E%3CDT%3EJournal+Article%3C%2FDT%3E>

180. **Stone formation after augmentation cystoplasty: the role of intestinal mucus.**

**Item Type:** Journal Article

**Authors:** Khoury, A. E.;Salomon, M.;Doche, R.;Soboh, F.;Ackerley, C.;Jayanthi, R.;McLorie, G. A. and Mittelman, M. W.

**Publication Date:** Se ,1997

**Journal:** Journal of Urology 158(3 Pt 2), pp. 1133–1137

**Abstract: PURPOSE**: We evaluated the role of mucus in urine after bladder augmentation and hypothesize that mucus acts as a possible etiological factor in stone formation. **MATERIALS AND METHODS**: Mucus was collected via centrifugation from the 24-hour urine specimens of 8 stone forming and 10 nonstone forming patients who were randomly selected from our augmentation population. The mucus and stones were lyophilized, and then analyzed via scanning electron microscopy and energy dispersive x-ray spectrometry for calcium, phosphate, magnesium and sodium. The 24-hour urine collections were also analyzed to determine any metabolic differences between the 2 groups. **RESULTS**: Scanning electron microscopy and energy dispersive x-ray spectrometry spectra showed increased calcium, phosphate, and magnesium, and significantly higher (p : Scanning electron microscopy and energy dispersive x-ray spectrometry spectra showed increased calcium, phosphate, and magnesium, and significantly higher (p **CONCLUSIONS**: Mucus appears to have an important role in the genesis of bladder stones after augmentation, possibly acting as a nidus. Metabolic changes following augmentation were similar in stone and nonstone forming populations. Our data suggest that mucous calcium-to-phosphate ratios may be predictive of future stone formation. Furthermore, there may be a benefit in instituting more aggressive measures aimed at clearing mucus from the bladder.

**URL:** <https://ovidsp.ovid.com/ovidweb.cgi?T=JS&CSC=Y&NEWS=N&PAGE=fulltext&D=med4&AN=9258156> <https://libkey.io/libraries/2592/openurl?genre=article&sid=OVID:medline&id=pmid:9258156&id=doi:&issn=0022-5347&isbn=&volume=158&issue=3&spage=1133&pages=1133-7&date=1997&title=Journal+of+Urology&atitle=Stone+formation+after+augmentation+cystoplasty%3A+the+role+of+intestinal+mucus.&aulast=Khoury&pid=%3Cauthor%3EKhoury+AE%3BSalomon+M%3BDoche+R%3BSoboh+F%3BAckerley+C%3BJayanthi+R%3BMcLorie+GA%3BMittelman+MW%3C%2Fauthor%3E%3CAN%3E9258156%3C%2FAN%3E%3CDT%3EJournal+Article%3C%2FDT%3E>

181. **Effect of mineral water containing calcium and magnesium on calcium oxalate urolithiasis risk factors.**

**Item Type:** Journal Article

**Authors:** Rodgers, A. L.

**Publication Date:** 1997

**Journal:** Urologia Internationalis 58(2), pp. 93–99

**Abstract:** Calcium oxalate kidney stone formers are invariably advised to increase their fluid intake. In addition, magnesium therapy is often administered. Recently, a prospective study showed that a high dietary intake of calcium reduces the risk of symptomatic kidney stones. The present study was performed to test whether simultaneous delivery of these factors--high fluid intake, magnesium ingestion and increased dietary calcium--could reduce the risk of calcium oxalate kidney stone formation. A French mineral water, containing calcium and magnesium (202 and 36 ppm, respectively) was selected as the dietary vehicle. Twenty calcium oxalate stone-forming patients of each sex as well as 20 healthy volunteers of each sex participated in the study. Each subject provided a 24-hour urine collection after ingestion of mineral water over a period of 3 days; after a suitable rest period the protocol was repeated using local tap water (Ca: 13 ppm, Mg: 1 ppm). In addition, 24-hour urines were collected by each subject on their free diets. The entire cycle was repeated at least twice by each subject. Several risk factors (excretion of oxalate; relative supersaturations of calcium oxalate, brushite and uric acid; calcium oxalate metastable limit; oxalate:magnesium ratio and oxalate:metastable limit ratio) were favourably altered by the mineral water and tap water regimens but the former was more effective. In addition, the mineral water protocol produced favourable but unique changes in the excretion of citrate and magnesium as well as in the relative supersaturation of brushite which were not achieved by the tap water regimen. To the contrary, tap water produced an unfavourable change in the magnesium excretion. The group which benefitted most were male stone formers in whom 9 risk factors were favourably altered by the mineral water protocol. It is concluded that mineral water containing calcium and magnesium, such as that used in this study, deserves to be considered as a possible therapeutic or prophylactic agent in calcium oxalate kidney stone disease.

**URL:** <https://ovidsp.ovid.com/ovidweb.cgi?T=JS&CSC=Y&NEWS=N&PAGE=fulltext&D=med4&DO=10.1159%2f000282958> <https://libkey.io/libraries/2592/openurl?genre=article&sid=OVID:medline&id=pmid:9096270&id=doi:10.1159%2F000282958&issn=0042-1138&isbn=&volume=58&issue=2&spage=93&pages=93-9&date=1997&title=Urologia+Internationalis&atitle=Effect+of+mineral+water+containing+calcium+and+magnesium+on+calcium+oxalate+urolithiasis+risk+factors.&aulast=Rodgers&pid=%3Cauthor%3ERodgers+AL%3C%2Fauthor%3E%3CAN%3E9096270%3C%2FAN%3E%3CDT%3EClinical+Trial%3C%2FDT%3E>

182. **Spot urine samples for the metabolic evaluation of urolithiasis patients.**

**Item Type:** Journal Article

**Authors:** Strohmaier, W. L.;Hoelz, K. J. and Bichler, K. H.

**Publication Date:** 1997

**Journal:** European Urology 32(3), pp. 294–300

**Abstract: OBJECTIVES**: This prospective study was initiated to assess the significance of spot urine specimens (SU) for the metabolic evaluation of stone formers. **METHODS**: 68 stone patients (51 males, 17 females) and 20 controls (9 males, 11 females) participated. On 3 consecutive days, urine was collected. Fasting (SU1) and postprandial (SU2) SU were obtained. From these, aliquots were taken, all the other urine was mixed to obtain 24-hour timed specimens (24hU). In all specimens, pH, specific gravity, creatinine, calcium, magnesium, phosphate, citrate, uric and oxalic acids were measured. The latter analytes were related to creatinine (mmol/g creatinine). Pearson correlation coefficients with their levels of significance and the day-to-day variation were calculated. Using the values in the control group, normal values (means +/- 2 SD) were established. **RESULTS**: There was a significant correlation (p : There was a significant correlation (p **CONCLUSIONS**: Despite a minor inaccuracy by relating parameters to creatinine, SU are sufficient in the routine metabolic evaluation of stone formers, since a third of all 24hU has to be rejected because of considerable collection errors. SU circumvent this problem. Because of the day-to-day variation, 3 SU should be obtained to overcome the doubtful significance of one single specimen.

**URL:** <https://ovidsp.ovid.com/ovidweb.cgi?T=JS&CSC=Y&NEWS=N&PAGE=fulltext&D=med4&AN=9358216> <https://libkey.io/libraries/2592/openurl?genre=article&sid=OVID:medline&id=pmid:9358216&id=doi:&issn=0302-2838&isbn=&volume=32&issue=3&spage=294&pages=294-300&date=1997&title=European+Urology&atitle=Spot+urine+samples+for+the+metabolic+evaluation+of+urolithiasis+patients.&aulast=Strohmaier&pid=%3Cauthor%3EStrohmaier+WL%3BHoelz+KJ%3BBichler+KH%3C%2Fauthor%3E%3CAN%3E9358216%3C%2FAN%3E%3CDT%3EClinical+Trial%3C%2FDT%3E>

183. **Calcium oxalate crystallization in urine of healthy men and women: a comparative study.**

**Item Type:** Journal Article

**Authors:** Buchholz, N. P.;Kim, D. S.;Grover, P. K. and Ryall, R. L.

**Publication Date:** 1996

**Journal:** Scanning Microscopy 10(2), pp. 435–442

**Abstract:** This study aimed to compare calcium oxalate (CaOx) crystallization in undiluted urine from healthy men and women with the object of clarifying the difference in stone incidence between the two sexes. Twenty-four hour urine specimens were collected from 37 men and 28 women. Urinary pH, and concentrations of Ca, oxalate and urate were measured, and indices of crystallization determined by Coulter Counter particle analysis following induction of CaOx crystallization by addition of oxalate. The amount of oxalate required to induce crystallization was significantly (p < 0.01) higher in females than in males, as was the overall particle volume deposited after 90 minutes incubation (p < 0.006). Scanning electron microscopy revealed larger individual crystals in female urine, and a greater degree of crystal aggregation in male urine, although the average overall size of the precipitated crystal particles did not differ between the two sexes. There were no significant differences between men and women with regard to median pH, or Ca and oxalate concentrations, but the median urate concentrations were slightly, but significantly, higher (p < 0.05) in the women's urines than in the men's. It was concluded that the greater risk of CaOx stones in men is related to an increased propensity to nucleate CaOx crystals per se, rather than to a tendency to form larger crystalline particles.

**URL:** <https://ovidsp.ovid.com/ovidweb.cgi?T=JS&CSC=Y&NEWS=N&PAGE=fulltext&D=med4&AN=9813622> <https://libkey.io/libraries/2592/openurl?genre=article&sid=OVID:medline&id=pmid:9813622&id=doi:&issn=0891-7035&isbn=&volume=10&issue=2&spage=435&pages=435-42%3B+discussion+442-4&date=1996&title=Scanning+Microscopy&atitle=Calcium+oxalate+crystallization+in+urine+of+healthy+men+and+women%3A+a+comparative+study.&aulast=Buchholz&pid=%3Cauthor%3EBuchholz+NP%3BKim+DS%3BGrover+PK%3BRyall+RL%3C%2Fauthor%3E%3CAN%3E9813622%3C%2FAN%3E%3CDT%3EComparative+Study%3C%2FDT%3E>

184. **Comparison of relative risks of urinary stone formation after surgery for ulcerative colitis: conventional ileostomy vs. J-pouch. A comparative study.**

**Item Type:** Journal Article

**Authors:** Christie, P. M.;Knight, G. S. and Hill, G. L.

**Publication Date:** Ja ,1996

**Journal:** Diseases of the Colon & Rectum 39(1), pp. 50–54

**Abstract: PURPOSE**: Urinary stone formation is a widely recognized complication o f inflammatory bowel disease and the surgical management of these conditions. Previously the fecal volume and chemistry after restorative proctocolectomy with J-pouch were found to be similar to conventional panproctocolectomy with permanent ileostomy. The purpose of this study was to investigate whether the relative risks of urinary stone formation were less following J-pouch than following conventional ileostomy. **METHODS**: The risk of urinary stone formation was determined from the chemical composition of two consecutive 24-hour urine samples in 13 patients with well-functioning ileostomies, 15 patients with well-functioning J-pouches, and 17 control volunteers. **RESULTS**: Compared with controls, ileostomy and J-pouch patients had significantly lowered urinary volumes and pH, higher concentrations of calcium and oxalate, and an increased risk of forming uric acid stones. In addition there was an increased risk of forming calcium stones in the conventional ileostomy group. This risk was found not to be present in the J-pouch group. **CONCLUSIONS**: The risks of forming uric acid stones are high for both ileostomy and J-pouch patients, but our results suggest that there will be a reduction in calcium stone formation after J-pouch.

**URL:** <https://ovidsp.ovid.com/ovidweb.cgi?T=JS&CSC=Y&NEWS=N&PAGE=fulltext&D=med4&DO=10.1007%2fBF02048269> <https://libkey.io/libraries/2592/openurl?genre=article&sid=OVID:medline&id=pmid:8601357&id=doi:10.1007%2FBF02048269&issn=0012-3706&isbn=&volume=39&issue=1&spage=50&pages=50-4&date=1996&title=Diseases+of+the+Colon+%26+Rectum&atitle=Comparison+of+relative+risks+of+urinary+stone+formation+after+surgery+for+ulcerative+colitis%3A+conventional+ileostomy+vs.+J-pouch.+A+comparative+study.&aulast=Christie&pid=%3Cauthor%3EChristie+PM%3BKnight+GS%3BHill+GL%3C%2Fauthor%3E%3CAN%3E8601357%3C%2FAN%3E%3CDT%3EComparative+Study%3C%2FDT%3E>

185. **Urinary oxalate excretion increases with body size and decreases with increasing dietary calcium intake among healthy adults.**

**Item Type:** Journal Article

**Authors:** Lemann, J. Jr;Pleuss, J. A.;Worcester, E. M.;Hornick, L.;Schrab, D. and Hoffmann, R. G.

**Publication Date:** Ja ,1996

**Journal:** Kidney International 49(1), pp. 200–208

**Abstract:** Increasing dietary calcium intake decreases urinary oxalate excretion by increasing intestinal precipitation of dietary oxalate as calcium oxalate. This mechanism was speculated to account for the decreased prospective incidence of kidney stones as estimated dietary calcium intake, adjusted for caloric intake, increased among men in a recent large epidemiological study. To further assess the relationship between estimated diet calcium and urinary oxalate, we studied 94 health adults, 50 women and 44 men, ages 20 to 70 years with weights ranging form 47 to 104 kg while they ate their customary diets. Each subject completed a semiquantitative food frequency questionnaire and collected three 24-hour urines preserved with HCl. The urines were collected accurately as judged by a mean intrasubject CV for creatinine excretion of 9.8% and direct relations between urinary creatinine excretion and body wt (r = 0.62; P < 0.0001), or predicted urine creatinine content for sex, age and weight using the Cockcroft and Gault formulas (r = 0.76; P < 0.0001). Estimated diet calcium intake ranged from 6.8 to 68 mmol/day (272 to 2720 mg/day) and averaged 29.5 mmol/day (1180 mg/day). Individual mean urinary oxalate excretion ranged from 0.079 go 0.332 mmol/day (7 to 29 mg/day) and averaged 0.198 mmol/day (17 mg/day). Among all subjects, daily oxalate excretion was directly related to creatinine excretion as an estimate of lean body mass (r = 0.61; P < 0.0001). Thus, oxalate excretion among men averaged 0.228 +/- 0.051 SD mmol/day, a value significantly higher than the average among women of 0.173 +/- 0.045 mmol/day (P < 0.001). Daily urine oxalate excretion/creatinine decreased curvilinearly as estimated dietary Ca intake increased (r = -0.30; P = 0.0035) and as the ratio of estimated dietary calcium to dietary oxalate increased (r = -0.39; P = 0.0001). We conclude that body size is the major determinant of urinary oxalate excretion among healthy adults, presumably reflecting variations in endogenous oxalate synthesis with lean body mass. Increasing estimated diet calcium intake, especially up to the range of 15 to 20 mmol/day (600 to 800 mg/day) has an additional effect to decrease during oxalate excretion, presumably by limiting intestinal absorption of dietary oxalate.

**URL:** <https://ovidsp.ovid.com/ovidweb.cgi?T=JS&CSC=Y&NEWS=N&PAGE=fulltext&D=med4&DO=10.1038%2fki.1996.27> <https://libkey.io/libraries/2592/openurl?genre=article&sid=OVID:medline&id=pmid:8770968&id=doi:10.1038%2Fki.1996.27&issn=0085-2538&isbn=&volume=49&issue=1&spage=200&pages=200-8&date=1996&title=Kidney+International&atitle=Urinary+oxalate+excretion+increases+with+body+size+and+decreases+with+increasing+dietary+calcium+intake+among+healthy+adults.&aulast=Lemann&pid=%3Cauthor%3ELemann+J+Jr%3BPleuss+JA%3BWorcester+EM%3BHornick+L%3BSchrab+D%3BHoffmann+RG%3C%2Fauthor%3E%3CAN%3E8770968%3C%2FAN%3E%3CDT%3EJournal+Article%3C%2FDT%3E>

186. **Weekend versus weekday urine collections in assessment of stone-formers.**

**Item Type:** Journal Article

**Authors:** Norman, R. W.

**Publication Date:** Oc ,1996

**Journal:** Journal of the Royal Society of Medicine 89(10), pp. 561–562

**Abstract:** Twenty-four-hour urine collections are an important part of the metabolic evaluation of stone-formers, but are difficult for patients at work. At weekends the results might be different. Forty-five stone-formers who worked at day jobs from Monday to Friday collected urine for 24 h on a normal working day and also on a Saturday or Sunday and the differences were evaluated. Average 24 h urine volume was higher on weekdays than at weekends. Calcium, oxalate, and uric acid excretion did not differ. These results imply an increased risk of crystalluria at the weekend. Therefore weekend collections are most likely to show abnormalities and should be acceptable to clinicians.

**URL:** <https://ovidsp.ovid.com/ovidweb.cgi?T=JS&CSC=Y&NEWS=N&PAGE=fulltext&D=med4&DO=10.1177%2f014107689608901006> <https://libkey.io/libraries/2592/openurl?genre=article&sid=OVID:medline&id=pmid:8976890&id=doi:10.1177%2F014107689608901006&issn=0141-0768&isbn=&volume=89&issue=10&spage=561&pages=561-2&date=1996&title=Journal+of+the+Royal+Society+of+Medicine&atitle=Weekend+versus+weekday+urine+collections+in+assessment+of+stone-formers.&aulast=Norman&pid=%3Cauthor%3ENorman+RW%3C%2Fauthor%3E%3CAN%3E8976890%3C%2FAN%3E%3CDT%3EJournal+Article%3C%2FDT%3E>

187. **Some aspects of citrate metabolism in renal-stone patients from northeastern Thailand.**

**Item Type:** Journal Article

**Authors:** Sriboonlue, P.;Prasongwatana, V.;Borvornpadoongkitti, S. and Suwantrai, S.

**Publication Date:** No ,1996

**Journal:** Journal of the Medical Association of Thailand 79(11), pp. 737–743

**Abstract:** In our previous nephrolithiasis studies in the northeast region of Thailand, hypokaliurea and hypocitraturia were the 2 most commonly encountered metabolic abnormalities. This led us to believe that people prone to forming renal-stones in this area were in a state of potassium depletion, a condition which probably caused the low urinary excretion of citrate. Further studies on some aspects of citrate metabolism in these subjects were carried out. Two groups of adult male subjects were included in the study protocol. Group 1 consisted of 20 urban dwellers who were used as normal controls, and group 2 was comprised of 36 renal-stone patients residing in rural villages outside the municipal area. Fasting clotted venous blood and one 24-hour urine specimens were collected and analyzed for creatinine, citrate, calcium, phosphate, magnesium, sodium, potassium, chloride, bicarbonate and uric acid. Values for: creatinine and citrate clearances, the filtered load of citrate and the tubular reabsorption of citrate were then calculated. The results showed that, for both groups, the concentrations of most of the above parameters were within the normal ranges both serum and urine. An exception to this was that the levels of serum potassium and of urinary excretions of sodium, potassium and citrate in people in group 2 were significantly less than those in group 1 (p < 0.005, p < 0.001, p < 0.001 and p < 0.0001, respectively). With respect to citrate metabolism, while the serum citrate levels and the filtered load of citrate were not different between the 2 groups, the average percentage of renal tubular reabsorption of group 2 (95 +/- 1.1%) increased significantly (P < 0.0001)in comparison to group 1 (85 +/- 1.6%). Moreover when results from both groups were combined, a significant negative correlation between the renal tubular reabsorption of citrate and the urinary excretion of potassium was clearly seen (r = 0.4001, p < 0.007). Our data suggests that potassium depletion may affect the renal tubular cells in some manner which, consequently, causes an increase in renal tubular reabsorption of citrate. The final outcome of these changes in these renal stone subjects was hypocitraturia.

**URL:** <https://ovidsp.ovid.com/ovidweb.cgi?T=JS&CSC=Y&NEWS=N&PAGE=fulltext&D=med4&AN=8997014> <https://libkey.io/libraries/2592/openurl?genre=article&sid=OVID:medline&id=pmid:8997014&id=doi:&issn=0125-2208&isbn=&volume=79&issue=11&spage=737&pages=737-43&date=1996&title=Journal+of+the+Medical+Association+of+Thailand&atitle=Some+aspects+of+citrate+metabolism+in+renal-stone+patients+from+northeastern+Thailand.&aulast=Sriboonlue&pid=%3Cauthor%3ESriboonlue+P%3BPrasongwatana+V%3BBorvornpadoongkitti+S%3BSuwantrai+S%3C%2Fauthor%3E%3CAN%3E8997014%3C%2FAN%3E%3CDT%3EComparative+Study%3C%2FDT%3E>

188. **Idiopathic calcium oxalate urinary lithiasis: usefulness of Parks' and Tiselius' indices in the evaluation of the risk of stone formation.**

**Item Type:** Journal Article

**Authors:** Robert, M.;Roux, J. O.;Boularan, A. M.;Bourelly, F.;Monnier, L. and Grasset, D.

**Publication Date:** 1995

**Journal:** Urologia Internationalis 55(2), pp. 88–92

**Abstract:** Different indices of the risk of urinary calcium oxalate crystallization were compared to determine their usefulness in detecting the stone-formers particularly prone to recurrence. Urine volume and calcium, oxalate, citrate, magnesium or creatinine were determined in 55 patients presenting with an idiopathic calcium oxalate urolithiasis, as well as in 50 control subjects. On 24-hour urine samples, these elements allowed for the calculation and comparison of different indices of lithogenous risk as proposed by Parks and Tiselius. Both Parks' indices and the urinary citrate-calcium ratio varied significantly between the two groups, but conversely Tiselius' indices were statistically comparable. The three Tiselius' indices taking the 24-hour urine volume into account were also strongly correlated. Parks' index and the urinary citrate-calcium ratio are highly discriminating and potentially relevant to select the stone-formers with a high risk of relapse. Tiselius' indices basically reflect urinary calcium oxalate saturation, and can only be used clinically to control the treatment interfering with this. In this respect, the formula based simply on urine volume, calcium and oxalate over 24 h (Ca0.71.Ox.V-1.2) appears to be sufficient.

**URL:** <https://ovidsp.ovid.com/ovidweb.cgi?T=JS&CSC=Y&NEWS=N&PAGE=fulltext&D=med3&DO=10.1159%2f000282758> <https://libkey.io/libraries/2592/openurl?genre=article&sid=OVID:medline&id=pmid:8533202&id=doi:10.1159%2F000282758&issn=0042-1138&isbn=&volume=55&issue=2&spage=88&pages=88-92&date=1995&title=Urologia+Internationalis&atitle=Idiopathic+calcium+oxalate+urinary+lithiasis%3A+usefulness+of+Parks%27+and+Tiselius%27+indices+in+the+evaluation+of+the+risk+of+stone+formation.&aulast=Robert&pid=%3Cauthor%3ERobert+M%3BRoux+JO%3BBoularan+AM%3BBourelly+F%3BMonnier+L%3BGrasset+D%3C%2Fauthor%3E%3CAN%3E8533202%3C%2FAN%3E%3CDT%3EComparative+Study%3C%2FDT%3E>

189. **Calcium oxalate stone agglomeration reflects stone-forming activity: citrate inhibition depends on macromolecules larger than 30 kilodalton.**

**Item Type:** Journal Article

**Authors:** Erwin, D. T.;Kok, D. J.;Alam, J.;Vaughn, J.;Coker, O.;Carriere, B. T.;Lindberg, J.;Husserl, F. E.;Fuselier, H. Jr and Cole, F. E.

**Publication Date:** De ,1994

**Journal:** American Journal of Kidney Diseases 24(6), pp. 893–900

**Abstract:** To evaluate the clinical utility of in vitro calcium oxalate monohydrate (COM) crystallization kinetics measurements and to determine the effect of quantitative removal of urinary Tamm-Horsfall glycoprotein on such measurements, we examined 24-hour, room temperature urine collections of patients from our Stone Clinic and of normal subjects from our research laboratories at Ochsner Medical Institutions in New Orleans, LA, and compared their COM kinetic parameters in vitro before and after urine ultrafiltration (30 kd). Data from 53 calcium oxalate stone-forming patients (26% women; mean age, 47 years) who demonstrated radiographic or other evidence of forming at least one stone were compared with data from 22 healthy volunteers (25% women; mean age, 40 years). Hypercalciuria (> 7.5 mm/24 hr), hyperoxaluria (> 0.5 mm/24 hr), and hypocitraturia (< 2.0 mm/24 hr) were present in 38%, 26%, and 26% of the patient population, respectively. Urinary creatinine, urate, calcium, citrate, phosphate, oxalate, pH, volume, total immunoreactive-disaggregated Tamm-Horsfall glycoprotein, and the urine's effects on COM solubility, percent crystal growth inhibition, and crystal agglomeration inhibition [tm] were determined. Calcium oxalate monohydrate agglomeration inhibition, [tm], was reduced in stone-forming patients. It decreased with increasing stone frequency, making [tm] a useful tool for measuring the risk of stone recurrence. Urinary Tamm-Horsfall glycoprotein and citrate concentrations were linearly related to COM agglomeration inhibition. Their effects were synergistic. Tamm-Horsfall glycoprotein removal from urine reduced COM agglomeration inhibition dramatically. Alkali therapy increased urinary citrate concentration and increased [tm].(ABSTRACT TRUNCATED AT 250 WORDS)

**URL:** <https://ovidsp.ovid.com/ovidweb.cgi?T=JS&CSC=Y&NEWS=N&PAGE=fulltext&D=med3&DO=10.1016%2fs0272-6386%2812%2981057-2> <https://libkey.io/libraries/2592/openurl?genre=article&sid=OVID:medline&id=pmid:7985666&id=doi:10.1016%2Fs0272-6386%2812%2981057-2&issn=0272-6386&isbn=&volume=24&issue=6&spage=893&pages=893-900&date=1994&title=American+Journal+of+Kidney+Diseases&atitle=Calcium+oxalate+stone+agglomeration+reflects+stone-forming+activity%3A+citrate+inhibition+depends+on+macromolecules+larger+than+30+kilodalton.&aulast=Erwin&pid=%3Cauthor%3EErwin+DT%3BKok+DJ%3BAlam+J%3BVaughn+J%3BCoker+O%3BCarriere+BT%3BLindberg+J%3BHusserl+FE%3BFuselier+H+Jr%3BCole+FE%3C%2Fauthor%3E%3CAN%3E7985666%3C%2FAN%3E%3CDT%3EComparative+Study%3C%2FDT%3E>

190. **Value of repeated analyses of 24-hour urine in recurrent calcium urolithiasis.**

**Item Type:** Journal Article

**Authors:** Hobarth, K.;Hofbauer, J. and Szabo, N.

**Publication Date:** 1994

**Journal:** Urology 44(1), pp. 20–24

**Abstract: OBJECTIVES**: The value of repeated analyses of 24-hour urine collections for daily excretion of calcium, uric acid, citrate, phosphorus, and creatinine and for volume and pH performed to detect and classify metabolic disorders in a selected group of calcium stone formers with striking recurrence rates was assessed in a retrospective study. **METHODS**: A total of 441 urinalyses made over a mean period of 80.4 months of samples obtained from 49 patients were reviewed. Fifty-nine percent of patients were initially found to have metabolic disorders (absorptive hypercalciuria types I and II, hyperuricuria, hypocitraturia) and therefore received specific drug therapy (allopurinol, thiazide, alkali citrate, orthophosphate) for a mean of 45.7 months (group I). The remaining patients were classified as metabolically inactive and were given general metaphylactic instructions (group II). **RESULTS**: In 73% of patients recurrent stones developed, with no statistically significant difference between the two groups (79% vs 65%). In 55% of group I patients, urinalyses continued to yield abnormal findings during follow-up; however, subsequent abnormal findings were also seen in 40% of the metabolically inactive group II patients. Overall, metabolic disorders were observed at some point in 75% of patients. Only 27% remained recurrence free, and 62% thereof also had pathologic urinary findings. **CONCLUSIONS**: It does not appear that drug treatment in recurrent calcium urolithiasis based on urinary findings is superior to simple general metaphylactic recommendations, nor that repeated analyses of 24-hour urine collections furnish additional information on the risk of recurrent stone formation or on the presence of risk factors leading to recurrence in the long-term course of disease.

**URL:** <https://ovidsp.ovid.com/ovidweb.cgi?T=JS&CSC=Y&NEWS=N&PAGE=fulltext&D=med3&DO=10.1007%2f978-1-4615-2556-1_272> <https://libkey.io/libraries/2592/openurl?genre=article&sid=OVID:medline&id=pmid:8042263&id=doi:10.1007%2F978-1-4615-2556-1_272&issn=0090-4295&isbn=&volume=44&issue=1&spage=20&pages=20-4%3B+discussion+24-5&date=1994&title=Urology&atitle=Value+of+repeated+analyses+of+24-hour+urine+in+recurrent+calcium+urolithiasis.&aulast=Hobarth&pid=%3Cauthor%3EHobarth+K%3BHofbauer+J%3BSzabo+N%3C%2Fauthor%3E%3CAN%3E8042263%3C%2FAN%3E%3CDT%3EJournal+Article%3C%2FDT%3E>

191. **Evaluation of a simple test to estimate urine saturation with calcium oxalate in stone-forming patients.**

**Item Type:** Journal Article

**Authors:** Sanchez, A.;Gonzalez, A.;Sarano, H. D. and Reinaldi, S.

**Publication Date:** De ,1994

**Journal:** International Journal of Urology 1(4), pp. 316–318

**Abstract:** In 43 patients with recent-onset calcium oxalate nephrolithiasis (25 male, 18 female; age range: 17-61 yr), urine saturation with calcium oxalate (CaOx) was calculated using a computer program (Equil-AT). The results in kJ/mol (normal < or = 2.0) were compared with those obtained with a simple test consisting of the addition of increasing amounts of calcium and oxalate to 3 tubes containing 10 ml urine. Development of turbidity in tube I denotes urine supersaturation with CaOx; turbidity in tubes II or III indicates lesser degrees of saturation, while a lack of turbidity (NT: no turbidity) shows undersaturation. Both saturation estimates were performed using aliquots of 24-hour urine collections. Twenty-one samples developed turbidity in tube I, 9 in tube II, and 6 in tube III, while 7 showed NT. There were no significant differences in Equil-AT values between II and III and between III and NT (p < 0.05 for I vs II and I vs III). High computer-calculated saturation values were found in 86% of group I samples and 22% of group II samples. None of the urines exhibiting turbidity in tube III or NT had high saturation values as estimated by Equil-AT. The turbidity test was also performed using freshly voided morning urine samples from the same patients. The results agreed with those obtained using the same test on aliquots of 24-hour collections, although in 10.8% of patients the morning test indicated higher levels of saturation. It is concluded that the turbidity test is a rapid, inexpensive and accurate way of estimating urine saturation with CaOx in the outpatient clinic.

**URL:** <https://ovidsp.ovid.com/ovidweb.cgi?T=JS&CSC=Y&NEWS=N&PAGE=fulltext&D=med3&DO=10.1111%2fj.1442-2042.1994.tb00056.x> <https://libkey.io/libraries/2592/openurl?genre=article&sid=OVID:medline&id=pmid:7614393&id=doi:10.1111%2Fj.1442-2042.1994.tb00056.x&issn=0919-8172&isbn=&volume=1&issue=4&spage=316&pages=316-8&date=1994&title=International+Journal+of+Urology&atitle=Evaluation+of+a+simple+test+to+estimate+urine+saturation+with+calcium+oxalate+in+stone-forming+patients.&aulast=Sanchez&pid=%3Cauthor%3ESanchez+A%3BGonzalez+A%3BSarano+HD%3BReinaldi+S%3C%2Fauthor%3E%3CAN%3E7614393%3C%2FAN%3E%3CDT%3EComparative+Study%3C%2FDT%3E>

192. **Crystalluria and its possible significance. A patient-control study.**

**Item Type:** Journal Article

**Authors:** Abdel-Halim, R. E.

**Publication Date:** 1993

**Journal:** Scandinavian Journal of Urology & Nephrology 27(2), pp. 145–149

**Abstract:** The significance of crystalluria in the diagnosis and prognosis of urolithiasis remains a controversial subject in the current urological literature. In this study, in addition to the standard urolithiasis clinical and biochemical work-up, routine urine microscopy was performed to study crystals in 1 fresh and 2 stored morning urine samples from 140 urinary stone patients and 42 controls. Crystalluria was more frequently detected in patients (9.3% of the fresh samples) than in controls (2%). Storing the samples for 6 hours did not increase the frequency percent of detected crystalluria either in patients or controls. However, in the samples stored for 24 hours, the frequency of crystalluria increased to 27.1% in patients and only to 12% in controls, though the pH did not change from that of the fresh sample. In addition, while calcium oxalate crystals in patients formed aggregates whether in fresh or 24 hour samples, those of controls did not. This denotes a characteristic change in the physico-chemical properties of the urine of stone formers from that of controls. Accordingly, the study of crystalluria in patients with urolithiasis seems to help in the proper evaluation and, maybe, treatment of the disease.

**URL:** <https://ovidsp.ovid.com/ovidweb.cgi?T=JS&CSC=Y&NEWS=N&PAGE=fulltext&D=med3&DO=10.3109%2f00365599309181240> <https://libkey.io/libraries/2592/openurl?genre=article&sid=OVID:medline&id=pmid:8351464&id=doi:10.3109%2F00365599309181240&issn=0036-5599&isbn=&volume=27&issue=2&spage=145&pages=145-9&date=1993&title=Scandinavian+Journal+of+Urology+%26+Nephrology&atitle=Crystalluria+and+its+possible+significance.+A+patient-control+study.&aulast=Abdel-Halim&pid=%3Cauthor%3EAbdel-Halim+RE%3C%2Fauthor%3E%3CAN%3E8351464%3C%2FAN%3E%3CDT%3EJournal+Article%3C%2FDT%3E>

193. **Blockage of indwelling urinary catheters: the roles of urinary composition, the catheter, medication and diet.**

**Item Type:** Journal Article

**Authors:** Burr, R. G. and Nuseibeh, I. M.

**Publication Date:** Ap ,1993

**Journal:** Paraplegia 31(4), pp. 234–241

**Abstract:** The aim of this work was to study factors related to the blockage of indwelling urinary catheters. There were 40 patients with indwelling catheters, 20 of whom had catheters that blocked frequently. The other 20 were trouble free at the time of our study. The type and gauge of catheter and frequency of events were recorded. Urine samples for biochemical analysis comprised 24-hour collections, morning specimens on up to 10 different days and 5-8 samples at different times during the same day. Chemical analysis of debris removed from blocked catheters showed it to consist of mixed phosphates of calcium and magnesium, thus being similar to urinary stones that may be seen in spinal cord injury patients. Patients with frequent catheter blockage had significantly elevated urinary pH and ammonium and calcium concentrations. Discriminant analysis gave 78-94% separation of catheter blocking patients from nonblockers depending on the type of sample. We conclude that bacterial urease activity and urinary calcium concentration are the most important factors in catheter blockage. Elevation of urinary pH following ingestion of effervescent preparations, drug- or diet-induced increases in urinary calcium or magnesium excretion and inadequate or erratic fluid intake may be avoidable contributing factors.

**URL:** <https://ovidsp.ovid.com/ovidweb.cgi?T=JS&CSC=Y&NEWS=N&PAGE=fulltext&D=med3&DO=10.1038%2fsc.1993.42> <https://libkey.io/libraries/2592/openurl?genre=article&sid=OVID:medline&id=pmid:8493038&id=doi:10.1038%2Fsc.1993.42&issn=0031-1758&isbn=&volume=31&issue=4&spage=234&pages=234-41&date=1993&title=Paraplegia&atitle=Blockage+of+indwelling+urinary+catheters%3A+the+roles+of+urinary+composition%2C+the+catheter%2C+medication+and+diet.&aulast=Burr&pid=%3Cauthor%3EBurr+RG%3BNuseibeh+IM%3C%2Fauthor%3E%3CAN%3E8493038%3C%2FAN%3E%3CDT%3EJournal+Article%3C%2FDT%3E>

194. **Calcium oxalate crystallization properties in urine with different specific electrical conductivities.**

**Item Type:** Journal Article

**Authors:** Tiselius, H. G.

**Publication Date:** Se ,1992

**Journal:** Journal of Urology 148(3 Pt 2), pp. 990–994

**Abstract:** The relationship between the degree of urine dilution and the risk of calcium oxalate crystallization was studied in 32 urine samples collected from stone formers and normal subjects during an 8-hour period between 10 p.m. and 6 a.m., and in 4-hour urine samples collected during 24-hour periods from 6 patients with calcium stone disease. The risk of calcium oxalate crystallization was analyzed in terms of the increase in oxalate concentration required for a standardized precipitation of crystals of calcium oxalate. The degree of urine dilution was determined with a new instrument (urimho) designed for measuring the concentration, in terms of specific electrical conductivity, in urine samples of droplet size. With this device urine concentration can be expressed in urimho values between 1 and 5. There was a good correlation between recordings of specific electrical conductivity performed with the new device and with a conventional conductivity meter. There was a statistically significant positive correlation between urimho values and calcium oxalate crystallization. Calcium oxalate crystallization greater than 1.3 was not observed in any sample with a urimho value of 2 but it was noted in 31% of the samples with urimho values greater than 2. A positive relationship was also recorded between the urine pH and the urimho level, which is noteworthy because there was an inverse relationship between urine pH and calcium oxalate crystallization. A pH greater than 6 was observed in 78% of the samples with a urimho value of 2 but it was noted in only 27% of the samples with urimho readings between 3 and 5. A considerable variation in the response in urinary flow to ingested volumes was recorded. Therefore, monitoring of urine dilution by means of a sample device like the urimho might be of great help for patients with calcium stone disease in an effort to prevent recurrent stone formation by urine dilution, provided a urimho value of less than 3 can be maintained.

**URL:** <https://ovidsp.ovid.com/ovidweb.cgi?T=JS&CSC=Y&NEWS=N&PAGE=fulltext&D=med3&DO=10.1016%2fs0022-5347%2817%2936797-6> <https://libkey.io/libraries/2592/openurl?genre=article&sid=OVID:medline&id=pmid:1507357&id=doi:10.1016%2Fs0022-5347%2817%2936797-6&issn=0022-5347&isbn=&volume=148&issue=3&spage=990&pages=990-4&date=1992&title=Journal+of+Urology&atitle=Calcium+oxalate+crystallization+properties+in+urine+with+different+specific+electrical+conductivities.&aulast=Tiselius&pid=%3Cauthor%3ETiselius+HG%3C%2Fauthor%3E%3CAN%3E1507357%3C%2FAN%3E%3CDT%3EJournal+Article%3C%2FDT%3E>

195. **Effect of hydrochlorothiazide on renal hypercalciuria.**

**Item Type:** Journal Article

**Authors:** Voskaki, I.;al Qadreh, A.;Mengreli, C. and Sbyrakis, S.

**Publication Date:** 1992

**Journal:** Child Nephrology & Urology 12(1), pp. 6–9

**Abstract:** The long-term effect of hydrochlorothiazide (HCT) was studied in 30 children (17 boys and 13 girls), aged 4-13 years (mean 7.6 +/- 3.4), with renal hypercalciuria (HCU) for a period of 2.5 +/- 0.95 years. The purpose of the study was to evaluate the effect of HCT mainly on renal calcium excretion, but also on serum K+, Ca, P, Mg, cPTH, nephrogenic cAMP (ncAMP) and bone mineral content (BMC). In addition to the above-mentioned parameters, oxalate, uric acid, citrate and cystine levels in 24-hour urine collections were also measured, and UCa/UCr, UP/UCr, UMg/UCr in 3-hour urine collections were assayed. The results of the study showed that 1 mg/kg/day of HCT caused a rapid and long-lasting correction of HCU, a decrease in serum cPTH and ncAMP, a significant increase in BMC and prevention of the formation of new urinary stones or of the increase in size of stones already present. No side effects were observed.

**URL:** <https://ovidsp.ovid.com/ovidweb.cgi?T=JS&CSC=Y&NEWS=N&PAGE=fulltext&D=med3&AN=1606587> <https://libkey.io/libraries/2592/openurl?genre=article&sid=OVID:medline&id=pmid:1606587&id=doi:&issn=1012-6694&isbn=&volume=12&issue=1&spage=6&pages=6-9&date=1992&title=Child+Nephrology+%26+Urology&atitle=Effect+of+hydrochlorothiazide+on+renal+hypercalciuria.&aulast=Voskaki&pid=%3Cauthor%3EVoskaki+I%3Bal+Qadreh+A%3BMengreli+C%3BSbyrakis+S%3C%2Fauthor%3E%3CAN%3E1606587%3C%2FAN%3E%3CDT%3EJournal+Article%3C%2FDT%3E>

196. **Modification by food of the calcium absorbability and physicochemical effects of calcium citrate.**

**Item Type:** Journal Article

**Authors:** Wabner, C. L. and Pak, C. Y.

**Publication Date:** Oc ,1992

**Journal:** Journal of the American College of Nutrition 11(5), pp. 548–552

**Abstract:** The food-calcium (Ca) interaction was examined in 12 healthy women (mean age 38 years) maintained on a constant metabolic diet. They underwent three phases of study, comprised of control (no Ca), Ca citrate (1 g Ca/day) during meals, and Ca citrate separately from meals. Each phase was 7 days in length and two 24-hour urine samples were collected on days 6 and 7. The rise from the control phase in urinary Ca was slightly more prominent when Ca citrate was given with meals than without (68 and 62%, respectively). The fall in urinary phosphorus was equivalent at about 25% between Ca citrate phases. The rise in urinary citrate and pH and the decline in urinary ammonium were more prominent when Ca citrate was given with meals; however, the changes were small or nonsignificant. The urinary saturation of Ca oxalate, brushite or monosodium urate did not differ between the two Ca citrate phases. There was a nonsignificant rise in serum iron during Ca citrate phases. The results suggest that: 1) dissolution and absorption of Ca citrate might be slightly greater when given with food than without; 2) that the ability of Ca citrate to attenuate crystallization of stone-forming Ca salts in urine is not modified by food; and 3) that Ca citrate may not impair iron absorption from food.

**URL:** <https://ovidsp.ovid.com/ovidweb.cgi?T=JS&CSC=Y&NEWS=N&PAGE=fulltext&D=med3&DO=10.1080%2f07315724.1992.10718260> <https://libkey.io/libraries/2592/openurl?genre=article&sid=OVID:medline&id=pmid:1452953&id=doi:10.1080%2F07315724.1992.10718260&issn=0731-5724&isbn=&volume=11&issue=5&spage=548&pages=548-52&date=1992&title=Journal+of+the+American+College+of+Nutrition&atitle=Modification+by+food+of+the+calcium+absorbability+and+physicochemical+effects+of+calcium+citrate.&aulast=Wabner&pid=%3Cauthor%3EWabner+CL%3BPak+CY%3C%2Fauthor%3E%3CAN%3E1452953%3C%2FAN%3E%3CDT%3EJournal+Article%3C%2FDT%3E>

197. **Diet, vitamin D and vertebral mineral density in hypercalciuric calcium stone formers.**

**Item Type:** Journal Article

**Authors:** Bataille, P.;Achard, J. M.;Fournier, A.;Boudailliez, B.;Westeel, P. F.;el Esper, N.;Bergot, C.;Jans, I.;Lalau, J. D. and Petit, J.

**Publication Date:** Ju ,1991

**Journal:** Kidney International 39(6), pp. 1193–1205

**Abstract:** To elucidate the pathophysiology of dietary calcium independent hypercalciuria, 42 calcium stone formers (Ca SF) were selected because they had on free diet a calciuria greater than 0.1 mmol/kg/day. For four days they were put on a diet restricted in calcium (Ca RD) by exclusion of the dairy products. They collected 24 hour urines on free diet and on day 4 of Ca RD as well as the two-hour fasting urines on the morning of the day 5 and the four-hour urines passed after an oral calcium load of 1 g, for measurement of creatinine, Ca, PO4, urea and total hydroxyprolinuria (THP). On day 5 fasting plasma concentrations of Ca, PO4, intact PTH, Gla protein, calcidiol and calcitriol were measured. The patients were firstly classified into dietary hypercalciuria (DH, 18 patients) and dietary calcium-independent hypercalciuria (IH, 24 patients) on the basis of the disappearance or not of hypercalciuria on Ca RD. Then the patients with IH were subclassified into absorptive hypercalciuria (AH) because of normal fasting calciuria (8 patients) and into fasting hypercalciuria (16 patients). Fasting hypercalciuric patients were subsequently divided according to the PTH levels into renal hypercalciuria (RH, 1 patient) with elevated fasting PTH becoming normal after the Ca load and undetermined hypercalciuria (UH, 15 patients) with normal PTH levels. Furthermore, their vertebral mineral density (VMD) was measured by quantitative computerized tomography which was normal in DH (91 +/- 6% of the normal mean for age and sex) but was decreased in IH to 69 +/- 4%. No difference in VMD was observed between AH and UH. Urinary excretions of urea, phosphate and THP was higher in IH than in DH and comparable in AH and UH. Sodium excretion Ca RD was the same in all groups and subgroups as well as the plasma parameters. Plasma calcitriol was increased in IH and DH comparatively to normal in spite of normal plasma calcidiol. Calciuria increase after oral calcium load, an index of Ca absorption, was higher in IH than in controls and comparable in IH and DH as well as in the three subgroups of IH. From these data and correlation studies in IH it is concluded: (1.) VMD is decreased in Ca stone formers with IH but not in those with DH, making the distinction of these two groups of hypercalciuria patients clinically relevant.(ABSTRACT TRUNCATED AT 400 WORDS)

**URL:** <https://ovidsp.ovid.com/ovidweb.cgi?T=JS&CSC=Y&NEWS=N&PAGE=fulltext&D=med3&DO=10.1038%2fki.1991.151> <https://libkey.io/libraries/2592/openurl?genre=article&sid=OVID:medline&id=pmid:1895673&id=doi:10.1038%2Fki.1991.151&issn=0085-2538&isbn=&volume=39&issue=6&spage=1193&pages=1193-205&date=1991&title=Kidney+International&atitle=Diet%2C+vitamin+D+and+vertebral+mineral+density+in+hypercalciuric+calcium+stone+formers.&aulast=Bataille&pid=%3Cauthor%3EBataille+P%3BAchard+JM%3BFournier+A%3BBoudailliez+B%3BWesteel+PF%3Bel+Esper+N%3BBergot+C%3BJans+I%3BLalau+JD%3BPetit+J%3Bet+al%3C%2Fauthor%3E%3CAN%3E1895673%3C%2FAN%3E%3CDT%3EJournal+Article%3C%2FDT%3E>

198. **Metabolic evaluation in stone patients in relation to extracorporeal shock wave lithotripsy treatment.**

**Item Type:** Journal Article

**Authors:** Lycklama a Nijeholt, G. A.;Tan, H. K. and Papapoulos, S. E.

**Publication Date:** De ,1991

**Journal:** Journal of Urology 146(6), pp. 1478–1481

**Abstract:** No information exists in the literature about the optimal time for metabolic evaluation of stone patients in relation to extracorporeal shock wave lithotripsy (ESWL) treatment. It is uncertain whether the presence of a stone, ESWL treatment itself or subsequent colic episodes influence the urinary risk factors. A prospective study was performed to determine the optimal period for metabolic evaluation. Two 24-hour urine samples were collected directly before, and 1 week, 1 month and 3 months after therapy in an outpatient setting and tested for total volume, calcium, uric acid, oxalate, citrate and creatinine levels. A total of 66 patients was available for evaluation. Comparison of the 4 subsequent collecting periods showed no statistically significant differences in the excretion values. Also, in subgroups of patients with colic (16%), on a calcium oxalate restricted diet (12%) and with repeated treatments within 3 months (33%) no differences were noted. This means that the presence of a stone, treatment itself or subsequent colic episodes have no adverse effect on the urinary risk factors. For practical reasons metabolic evaluation directly before ESWL treatment seems most attractive. In the pre-ESWL samples hypercalciuria (greater than 7.5 mmol./24 hours), hyperuricosuria (greater than 6 mmol./24 hours), hyperoxaluria (greater than 0.5 mmol./24 hours) and hypocitraturia (less than 2 mmol./24 hours) were found in 31%, 12%, 18% and 27%, respectively, of the patients. It is concluded that metabolic evaluation before ESWL is practical, applicable and reliable.

**URL:** <https://ovidsp.ovid.com/ovidweb.cgi?T=JS&CSC=Y&NEWS=N&PAGE=fulltext&D=med3&DO=10.1016%2fs0022-5347%2817%2938144-2> <https://libkey.io/libraries/2592/openurl?genre=article&sid=OVID:medline&id=pmid:1942322&id=doi:10.1016%2Fs0022-5347%2817%2938144-2&issn=0022-5347&isbn=&volume=146&issue=6&spage=1478&pages=1478-81&date=1991&title=Journal+of+Urology&atitle=Metabolic+evaluation+in+stone+patients+in+relation+to+extracorporeal+shock+wave+lithotripsy+treatment.&aulast=Lycklama+a+Nijeholt&pid=%3Cauthor%3ELycklama+a+Nijeholt+GA%3BTan+HK%3BPapapoulos+SE%3C%2Fauthor%3E%3CAN%3E1942322%3C%2FAN%3E%3CDT%3EJournal+Article%3C%2FDT%3E>

199. **The urine specific gravity dipstick: a useful tool to increase fluid intake in stone forming patients.**

**Item Type:** Journal Article

**Authors:** McCormack, M.;Dessureault, J. and Guitard, M.

**Publication Date:** De ,1991

**Journal:** Journal of Urology 146(6), pp. 1475–1477

**Abstract:** High fluid intake is the only preventive dietary measure that can be recommended to all patients with stones. However, the efficacy of dietary advice given to patients is unknown. We compared the impact of dietary advice to increase hydration (group 1, 57 patients) and of no dietary advice (group 2, 83 patients) on 24-hour urine volume. No significant difference was noted between groups 1 (1,624 ml.) and 2 (1,732 ml.). We then determined if urine specific gravity dipsticks could help patients increase the 24-hour urine volume. A correlation between 24-hour urine volume and mean urine specific gravity was performed on 263 randomly chosen patients. There was an inverse relationship between urine specific gravity and 24-hour urine volume with a correlation coefficient of 0.522 (y = 1.0207 - 0.00374x). Most patients (81.6%) with 24-hour urine volumes of less than 2.1 had a urine specific gravity of more than 1.010. The use of specific gravity dipsticks was evaluated as a tool to help 24 patients increase the 24-hour urine volume. The 24-hour urine volume increased significantly (p less than 0.05, paired Student's t test) in patients after feedback from specific gravity dipsticks when they were instructed to keep the urine specific gravity at or less than 1.010 (average 24-hour urine volume increased 192%). We conclude that dietary advice may be insufficient to modify fluid intake habits in stone patients. However, modifications of fluid intake habits may be improved by feedback from specific gravity dipsticks.

**URL:** <https://ovidsp.ovid.com/ovidweb.cgi?T=JS&CSC=Y&NEWS=N&PAGE=fulltext&D=med3&DO=10.1016%2fs0022-5347%2817%2938143-0> <https://libkey.io/libraries/2592/openurl?genre=article&sid=OVID:medline&id=pmid:1942321&id=doi:10.1016%2Fs0022-5347%2817%2938143-0&issn=0022-5347&isbn=&volume=146&issue=6&spage=1475&pages=1475-7&date=1991&title=Journal+of+Urology&atitle=The+urine+specific+gravity+dipstick%3A+a+useful+tool+to+increase+fluid+intake+in+stone+forming+patients.&aulast=McCormack&pid=%3Cauthor%3EMcCormack+M%3BDessureault+J%3BGuitard+M%3C%2Fauthor%3E%3CAN%3E1942321%3C%2FAN%3E%3CDT%3EClinical+Trial%3C%2FDT%3E>

200. **Seasonal variations in urinary risk factors among patients with nephrolithiasis.**

**Item Type:** Journal Article

**Authors:** Stuart RO 2nd;Hill, K.;Poindexter, J. and Pak, C. Y.

**Publication Date:** Ja ,1991

**Journal:** Journal of Lithotripsy & Stone Disease 3(1), pp. 18–27

**Abstract:** Twenty-four hour urine specimens from 5,677 stone-forming patients throughout the United States were analyzed for seasonal variations in urinary risk factors for nephrolithiasis. Determinations were performed for urine volume, pH, calcium, oxalate, phosphorus, sodium, magnesium, citrate, sulphate, uric acid, and the relative supersaturation (RS) of calcium oxalate, brushite, monosodium urate, and uric acid. Criteria for significant seasonal variation included a significant difference in monthly means of risk factors, seasonal grouping of the data by the Student-Newman-Keuls multiple range test, consistent year-to-year trends and a physiologically significant range. Minimum urine volume of 1.54 +/- 0.70 SD L/day occurred in October while a maximum urine volume of 1.76 +/- 0.78 SD L/day was observed during February. Minimum urine pH of 5.94 +/- 0.64 SD was observed during July and August while a maximum pH of 6.18 +/- 0.61 SD was observed during February. Daily urinary excretion of sodium was lowest during August, 158 +/- 74 SD mEq/day and highest during February 177 +/- 70 SD mEq/day. The RS of brushite and uric acid were found to display significant pH-dependent seasonal variation with a maximum RS of uric acid 2.26 +/- 1.98 SD in June and a low of 1.48 +/- 1.30 SD in February. Maximum RS of brushite 2.75 +/- 2.58 was observed during February. Minimum RS of brushite 1.93 +/- 1.70 SD was observed in June. Phosphorus excretion displayed seasonal variation about a spring-fall axis with a maximum value 1042 +/- 373 SD mg/day in April and a minimum value of 895 +/- 289 SD mg/day. Urine volume, sodium, and pH were significantly lower during the summer (June, July, August) than in the winter (December, January, February). The RS of uric acid was higher, but that of brushite and monosodium urate was lower in the summer than in the winter. The seasonal changes observed in urine volume, pH, sodium, and the RS of brushite and uric acid are consistent with summertime sweating and increased physical activity. Seasonal variations in phosphorus excretion are probably dietary in origin. The summertime was characterized by an increased propensity for the crystallization of uric acid but not of calcium oxalate or calcium phosphate.

**URL:** <https://ovidsp.ovid.com/ovidweb.cgi?T=JS&CSC=Y&NEWS=N&PAGE=fulltext&D=med3&AN=11536932> <https://libkey.io/libraries/2592/openurl?genre=article&sid=OVID:medline&id=pmid:11536932&id=doi:&issn=1040-2152&isbn=&volume=3&issue=1&spage=18&pages=18-27&date=1991&title=Journal+of+Lithotripsy+%26+Stone+Disease&atitle=Seasonal+variations+in+urinary+risk+factors+among+patients+with+nephrolithiasis.&aulast=Hill&pid=%3Cauthor%3EStuart+RO+2nd%3BHill+K%3BPoindexter+J%3BPak+CY%3C%2Fauthor%3E%3CAN%3E11536932%3C%2FAN%3E%3CDT%3EComparative+Study%3C%2FDT%3E>

201. **Aspects on estimation of the risk of calcium oxalate crystallization in urine.**

**Item Type:** Journal Article

**Authors:** Tiselius, H. G.

**Publication Date:** 1991

**Journal:** Urologia Internationalis 47(4), pp. 255–259

**Abstract:** A previously formulated simplified estimate of the ion activity product of calcium oxalate, AP(CaOx) index, was modified in order to better correspond numerically to the ion activity product obtained by computed iterative approximation with the EQUIL 2 program. The new index, AP(CaOx) index EQ, was given the following form for a 4-hour urine: 1.9 x Ca(0.84) x Ox x Mg-(0.12) x Cit-(0.22) x V-(1.03), where the excretion of calcium, oxalate (Ox), magnesium and citrate (Cit) was expressed in millimoles excreted during the period, and urine volume (V) in liters. There was a good correlation between AP(CaOx) index and AP(CaOx) index EQ (r = 0.98). A standardized index calculated for a 24-hour urine volume of 1.5 liters, AP(CaOx) index EQ(s), was significantly higher in stoneforming men (p less than 0.001) and women (p less than 0.001) than in normal subjects. The mean ion activity products of CaOx at the start of crystallization in salt solutions and dialyzed urine were 2.74 +/- 0.25 x 10(-8) and 3.50 +/- 0.33 x 10(-8) (mol/l)2, respectively. In the urine from normal subjects, AP(CaOx) index EQ at the point of crystallization was 4.3 +/- 1.1 and in the urine from stone formers 2.8 +/- 0.5 (p less than 0.001). From calculation of AP(CaOx) index EQ in hourly collected urine, it was assumed that the risk of exceeding the level where crystallization starts during periods with the highest supersaturation can be anticipated when the 24-hour AP(CaOx) index EQ value exceeds 2.0.

**URL:** <https://ovidsp.ovid.com/ovidweb.cgi?T=JS&CSC=Y&NEWS=N&PAGE=fulltext&D=med3&DO=10.1159%2f000282232> <https://libkey.io/libraries/2592/openurl?genre=article&sid=OVID:medline&id=pmid:1781112&id=doi:10.1159%2F000282232&issn=0042-1138&isbn=&volume=47&issue=4&spage=255&pages=255-9&date=1991&title=Urologia+Internationalis&atitle=Aspects+on+estimation+of+the+risk+of+calcium+oxalate+crystallization+in+urine.&aulast=Tiselius&pid=%3Cauthor%3ETiselius+HG%3C%2Fauthor%3E%3CAN%3E1781112%3C%2FAN%3E%3CDT%3EJournal+Article%3C%2FDT%3E>

202. **Similarity of urinary risk factors among stone-forming patients in five regions of the United States.**

**Item Type:** Journal Article

**Authors:** Harvey, J. A.;Hill, K. D. and Pak, C. Y.

**Publication Date:** Ap ,1990

**Journal:** Journal of Lithotripsy & Stone Disease 2(2), pp. 124–132

**Abstract: STUDY OBJECTIVE**: To compare urinary biochemical risk factors among stone-forming patients in the Southeast (SE) or "stone belt" versus four other regions of the United States. **DESIGN**: Prospective biochemical survey for regional comparisons. **SETTING**: Referral-based nephrolithiasis clinics, urologists, nephrologists, and family practitioners. **PATIENTS**: Consecutive sample of 3473 stone-forming patients who submitted 24-hour urine collections for biochemical analyses of stone-forming risk factors. **INTERVENTIONS**: None. Subjects taking medication known to interfere with stone-forming risk factors were deleted from the final data compilation. **MEASUREMENTS AND MAIN RESULTS**: Overall, the mean values for each urinary parameter spanned a narrow range without significant difference between the five regions. Among "metabolic" factors, 40% in the SE had hypercalciuria (> 6.25 mmol/d), compared to 35%-43% in other regions, and hyperuricosuria (> 4.2 mmol/d) was found in 16% in the SE versus 17%-19% elsewhere. Among "environmental" factors, low urine volume ( : Overall, the mean values for each urinary parameter spanned a narrow range without significant difference between the five regions. Among "metabolic" factors, 40% in the SE had hypercalciuria (> 6.25 mmol/d), compared to 35%-43% in other regions, and hyperuricosuria (> 4.2 mmol/d) was found in 16% in the SE versus 17%-19% elsewhere. Among "environmental" factors, low urine volume ( **CONCLUSIONS**: Despite expected regional differences in nutritional and environmental influences, the results of this study showed a striking similarity in urinary biochemical risk factor profiles of stone-formers in all five regions of the United States.

**URL:** <https://ovidsp.ovid.com/ovidweb.cgi?T=JS&CSC=Y&NEWS=N&PAGE=fulltext&D=med3&AN=11536931> <https://libkey.io/libraries/2592/openurl?genre=article&sid=OVID:medline&id=pmid:11536931&id=doi:&issn=1040-2152&isbn=&volume=2&issue=2&spage=124&pages=124-32&date=1990&title=Journal+of+Lithotripsy+%26+Stone+Disease&atitle=Similarity+of+urinary+risk+factors+among+stone-forming+patients+in+five+regions+of+the+United+States.&aulast=Harvey&pid=%3Cauthor%3EHarvey+JA%3BHill+KD%3BPak+CY%3C%2Fauthor%3E%3CAN%3E11536931%3C%2FAN%3E%3CDT%3EComparative+Study%3C%2FDT%3E>

203. **Urinary constituents in an endemic area of stones and renal tubular acidosis in northeastern Thailand.**

**Item Type:** Journal Article

**Authors:** Nilwarangkur, S.;Malasit, P.;Nimmannit, S.;Susaengrat, W.;Ong-Aj-Yooth, S.;Vasuvattakul, S. and Pidetcha, P.

**Publication Date:** Se ,1990

**Journal:** Southeast Asian Journal of Tropical Medicine & Public Health 21(3), pp. 437–441

**Abstract:** Nephrolithiasis and endemic renal distal tubular acidosis are common in northeastern Thailand. The etiology is still unknown. It is generally accepted that urine electrolytes influence the capacity of urine to inhibit or promote renal and also bladder stones. The purpose of this study was to analyse the composition of the urine in the indigenous population in the northeast area and compare their values with data obtained from a group of age matched adults, living in Bangkok. Twenty-four hour urine samples from 23 normal adult villagers from six villages within the province of Khon Kaen and 34 normal adults living in Bangkok were collected, and the daily excretion of creatinine, uric acid, calcium and inorganic phosphate, sodium, potassium, chloride, magnesium and oxalate were assayed. Daily urinary sodium, potassium, chloride and phosphate of the villagers were significantly lower than those of Bangkokians. No difference in the urinary excretion of calcium, magnesium, uric acid, oxalate and creatinine was found. The Na/Ca, and Ca/PO4 ratios of villagers were significantly lower than those of the Bangkok subjects. The villagers excreted significantly lower amounts of Na in the face of relatively higher urinary Ca. The above data, combined with our previous study showing the low values of urinary citrate in the villagers in the same areas, strongly indicate that the indigeneous population is at high risk in developing urolithiasis. The causes for these electrolyte abnormalities are still unknown. Low contents of the major electrolytes in their diets might play an important role. Low phosphate output indicates low protein diets.(ABSTRACT TRUNCATED AT 250 WORDS)

**URL:** <https://ovidsp.ovid.com/ovidweb.cgi?T=JS&CSC=Y&NEWS=N&PAGE=fulltext&D=med3&AN=2075484> <https://libkey.io/libraries/2592/openurl?genre=article&sid=OVID:medline&id=pmid:2075484&id=doi:&issn=0125-1562&isbn=&volume=21&issue=3&spage=437&pages=437-41&date=1990&title=Southeast+Asian+Journal+of+Tropical+Medicine+%26+Public+Health&atitle=Urinary+constituents+in+an+endemic+area+of+stones+and+renal+tubular+acidosis+in+northeastern+Thailand.&aulast=Nilwarangkur&pid=%3Cauthor%3ENilwarangkur+S%3BMalasit+P%3BNimmannit+S%3BSusaengrat+W%3BOng-Aj-Yooth+S%3BVasuvattakul+S%3BPidetcha+P%3C%2Fauthor%3E%3CAN%3E2075484%3C%2FAN%3E%3CDT%3EComparative+Study%3C%2FDT%3E>

204. **Urinary excretion of glycosaminoglycans in normal and stone forming subjects.**

**Item Type:** Journal Article

**Authors:** Michelacci, Y. M.;Glashan, R. Q. and Schor, N.

**Publication Date:** De ,1989

**Journal:** Kidney International 36(6), pp. 1022–1028

**Abstract:** There is evidence suggesting that glycosaminoglycans (GAG) are potent inhibitors of growth and aggregation of calcium oxalate crystals in vitro. This finding raises the possibility that the urinary GAG could play an inhibitory role in the urolithiasis. To investigate this hypothesis, a study on the urinary excretion of GAG in normal and stone forming adults and children was undertaken. Different methods were compared, and the best results were obtained when the GAG were measured by densitometry after agarose gel electrophoresis. Although the GAG concentration was increased in the morning urine compared to the 24-hour urine samples, and in males compared to females, the GAG/creatinine ratio was independent of period of urine collection and of sex. So, it was advantageous to express the amounts of urinary GAG as mg/g of creatinine. Children excreted more GAG than adults, with a higher proportion of chondroitin sulphate. We have shown that the stone forming subjects, both adults and children, excreted lower levels of urinary GAG as compared to normal subjects, independently of the metabolic disorder. The proportions between chondroitin sulphate and heparan sulphate and the structures of these GAG were unaltered in the stone formers. These results indicate that there is a definite difference in terms of levels of GAG between normal and stone forming urines, and suggest a correlation between the urinary GAG concentration and urolithiasis.

**URL:** <https://ovidsp.ovid.com/ovidweb.cgi?T=JS&CSC=Y&NEWS=N&PAGE=fulltext&D=med3&DO=10.1038%2fki.1989.296> <https://libkey.io/libraries/2592/openurl?genre=article&sid=OVID:medline&id=pmid:2601252&id=doi:10.1038%2Fki.1989.296&issn=0085-2538&isbn=&volume=36&issue=6&spage=1022&pages=1022-8&date=1989&title=Kidney+International&atitle=Urinary+excretion+of+glycosaminoglycans+in+normal+and+stone+forming+subjects.&aulast=Michelacci&pid=%3Cauthor%3EMichelacci+YM%3BGlashan+RQ%3BSchor+N%3C%2Fauthor%3E%3CAN%3E2601252%3C%2FAN%3E%3CDT%3EJournal+Article%3C%2FDT%3E>

205. **Specific gravity test strips used in monitoring urine concentrations of urolithiasis patients.**

**Item Type:** Journal Article

**Authors:** Cadoff, R. E.;Drach, G. W. and LeBouton, J.

**Publication Date:** Fe ,1988

**Journal:** Journal of Urology 139(2), pp. 323–325

**Abstract:** Current therapy for urolithiasis patients includes instructions to increase water intake and 24-hour urine output. Previous studies have measured changes in the 24-hour urine volume to evaluate the efficacy of fluid therapy in each patient. We used paper test strips to monitor urine pH and specific gravity in 22 of our stone clinic patients: 10 were instructed to increase water intake just before the study (group 1) and 12 were not so instructed (group 2). Mean specific gravities of 1.0222 (1.0238 corrected for pH) for group 1 and 1.0197 (1.0220 corrected for pH) for group 2 did not differ significantly. Urine specific gravities also were compared for 3 intervals: 1 to 9 a.m., 9 a.m. to 5 p.m. and 5 p.m. to 1 a.m. Of the 22 patients 10 (3 from group 1 and 7 from group 2) had significant diurnal variations in the urine specific gravities, corrected and uncorrected, among these 3 periods. In addition, both groups had a significantly higher mean specific gravity from 1 to 9 a.m. (1.0234 uncorrected and 1.0248 corrected) than from 9 a.m. to 5 p.m. (1.0194 uncorrected and 1.0218 corrected). The 5 p.m. to 1 a.m. (mean of 1.0220 uncorrected and 1.0239 corrected) specific gravity did not differ significantly in either group. If 1.015 is the highest acceptable specific gravity of urine in stone patients, the findings suggest inadequate dilution of urine in these patients, whether or not they were instructed to increase water intake. Also, the significant diurnal variation in urine specific gravity would allow a nighttime triggering event at these hours of higher urine concentration.

**URL:** <https://ovidsp.ovid.com/ovidweb.cgi?T=JS&CSC=Y&NEWS=N&PAGE=fulltext&D=med3&DO=10.1016%2fs0022-5347%2817%2942398-6> <https://libkey.io/libraries/2592/openurl?genre=article&sid=OVID:medline&id=pmid:3339731&id=doi:10.1016%2Fs0022-5347%2817%2942398-6&issn=0022-5347&isbn=&volume=139&issue=2&spage=323&pages=323-5&date=1988&title=Journal+of+Urology&atitle=Specific+gravity+test+strips+used+in+monitoring+urine+concentrations+of+urolithiasis+patients.&aulast=Cadoff&pid=%3Cauthor%3ECadoff+RE%3BDrach+GW%3BLeBouton+J%3C%2Fauthor%3E%3CAN%3E3339731%3C%2FAN%3E%3CDT%3EJournal+Article%3C%2FDT%3E>

206. **Determination of urinary citrate by ion chromatography.**

**Item Type:** Journal Article

**Authors:** Ogawa, Y.;Morozumi, M.;Tanaka, T. and Yamaguchi, K.

**Publication Date:** Ja ,1986

**Journal:** Journal of Urology 135(1), pp. 178–181

**Abstract:** Ion chromatography was used to determine urinary citrate concentration. The technique is simple, requiring a 100-fold dilution of urine and 35 minutes at most for each analysis. The technique does, however, have the disadvantage that it requires a rather expensive automated system. The minimum detectable limit for citrate was 0.5 microgram/ml. in a standard solution, and the regression line for the standard curve from 0.5 to 50 micrograms/ml. citrate had a significant correlation coefficient (lnY = 1.048 X lnX-2.755, r = 0.997). The intra-run coefficient of variation was 0.9 per cent. The overall intra-run and inter-run coefficients of variation, including the sampling and dilution of urine, were 4.8 and 8.3 per cent respectively. There is a possibility that another ion is close enough to the citrate peak to interfere; however, this problem can be solved by the treatment of the sample with citrate lyase. When the present method was compared with the conventional enzyme method, a significant correlation between the results was obtained in both human and rat urine. The 24-hour urinary citrate excretion in eight normal human males was 73.8 to 378.4 mg./day (mean, 174.7 mg./day), while that in male Wistar-strain rats (approximately 150 gm.) was 1.5 to 18.6 mg./day (mean, 9.5 mg./day), as measured in more than 50 consecutive rat urine samples.

**URL:** <https://ovidsp.ovid.com/ovidweb.cgi?T=JS&CSC=Y&NEWS=N&PAGE=fulltext&D=med2&DO=10.1016%2fs0022-5347%2817%2945557-1> <https://libkey.io/libraries/2592/openurl?genre=article&sid=OVID:medline&id=pmid:3941460&id=doi:10.1016%2Fs0022-5347%2817%2945557-1&issn=0022-5347&isbn=&volume=135&issue=1&spage=178&pages=178-81&date=1986&title=Journal+of+Urology&atitle=Determination+of+urinary+citrate+by+ion+chromatography.&aulast=Ogawa&pid=%3Cauthor%3EOgawa+Y%3BMorozumi+M%3BTanaka+T%3BYamaguchi+K%3C%2Fauthor%3E%3CAN%3E3941460%3C%2FAN%3E%3CDT%3EComparative+Study%3C%2FDT%3E>

207. **Pattern of calcium metabolism in normo- and hypercalciuric patients with calcium urolithiasis in Saudi Arabia.**

**Item Type:** Journal Article

**Authors:** Hanash, K. A.;Bissada, N. K. and Woodhouse, N. J.

**Publication Date:** Ju ,1985

**Journal:** Urology 26(1), pp. 27–32

**Abstract:** Urinary calcium excretion was measured in 100 consecutive normocalcemic patients with calcium urolithiasis and 12 Saudi Arabian controls while the patients and the controls were eating their usual diet of unknown calcium content. Only 16 patients were hypercalciuric using the definition of twenty-four-hour urinary calcium of more than 300 mg for males and more than 250 mg for females, and one of these patients was subsequently found to have hyperparathyroidism. The twenty-four-hour urine calcium was less than 200 mg in 60 per cent of male patients and in all the male controls. Since the dietary intake of calcium during the twenty-four-hour urinary collection was unknown, a 1,000-mg calcium loading test was performed in an attempt to differentiate various patterns of abnormal calcium excretion. Of the twenty-four-hour normocalciuric patients 10 (18%) had "absorptive hypercalciuria" and 16 patients (29%) demonstrated a "renal hypercalciuria" pattern. Thirty-nine patients (57%) and all 12 controls had normocalciuria before and after calcium loading.

**URL:** <https://ovidsp.ovid.com/ovidweb.cgi?T=JS&CSC=Y&NEWS=N&PAGE=fulltext&D=med2&DO=10.1016%2f0090-4295%2885%2990249-3> <https://libkey.io/libraries/2592/openurl?genre=article&sid=OVID:medline&id=pmid:2990080&id=doi:10.1016%2F0090-4295%2885%2990249-3&issn=0090-4295&isbn=&volume=26&issue=1&spage=27&pages=27-32&date=1985&title=Urology&atitle=Pattern+of+calcium+metabolism+in+normo-+and+hypercalciuric+patients+with+calcium+urolithiasis+in+Saudi+Arabia.&aulast=Hanash&pid=%3Cauthor%3EHanash+KA%3BBissada+NK%3BWoodhouse+NJ%3C%2Fauthor%3E%3CAN%3E2990080%3C%2FAN%3E%3CDT%3EComparative+Study%3C%2FDT%3E>

208. **Potential pitfalls of the 2-hour calcium-to-creatinine ratio and urinary cyclic adenosine monophosphate excretion in the differential diagnosis of idiopathic hypercalciuria.**

**Item Type:** Journal Article

**Authors:** Heller, J. E.;Konnak, J. W. and Lau, Y. K.

**Publication Date:** Ma ,1984

**Journal:** Journal of Urology 131(5), pp. 911–913

**Abstract:** We studied 44 patients with calcium urolithiasis on high (900 mg. daily) and low (400 mg. daily) calcium diets. With 24-hour urinary data, we categorized the patients as normocalciuric or hypercalciuric and subdivided the hypercalciuric patients into absorptive and renal types. Abbreviated tests, including the 2-hour fasting urinary calcium-to-creatinine ratio and 24-hour urinary (nephrogenous) cyclic adenosine monophosphate, did not predict accurately whether hypercalciuria was of the idiopathic, absorptive or renal type. However, 24-hour urinary calcium excretions on the low calcium diet had a sensitivity and specificity of more than 90 per cent for reproducing the categorized diagnoses.

**URL:** <https://ovidsp.ovid.com/ovidweb.cgi?T=JS&CSC=Y&NEWS=N&PAGE=fulltext&D=med2&DO=10.1016%2fs0022-5347%2817%2950707-7> <https://libkey.io/libraries/2592/openurl?genre=article&sid=OVID:medline&id=pmid:6323773&id=doi:10.1016%2Fs0022-5347%2817%2950707-7&issn=0022-5347&isbn=&volume=131&issue=5&spage=911&pages=911-3&date=1984&title=Journal+of+Urology&atitle=Potential+pitfalls+of+the+2-hour+calcium-to-creatinine+ratio+and+urinary+cyclic+adenosine+monophosphate+excretion+in+the+differential+diagnosis+of+idiopathic+hypercalciuria.&aulast=Heller&pid=%3Cauthor%3EHeller+JE%3BKonnak+JW%3BLau+YK%3C%2Fauthor%3E%3CAN%3E6323773%3C%2FAN%3E%3CDT%3EJournal+Article%3C%2FDT%3E>

209. **When should patients with symptomatic urinary stone disease be evaluated metabolically?.**

**Item Type:** Journal Article

**Authors:** Norman, R. W.;Bath, S. S.;Robertson, W. G. and Peacock, M.

**Publication Date:** De ,1984

**Journal:** Journal of Urology 132(6), pp. 1137–1139

**Abstract:** To determine the optimal time at which to assess the urinary risk factors for calcium stone formation, we followed 11 patients after an episode of acute renal colic with sequential 24-hour urine collections, first in the hospital and then at regular intervals after they were discharged from the hospital. The changes that occurred in the urinary risk factors were compared to those of a control group in which samples were collected during the same interval. The in-hospital 24-hour urine volumes were high but decreased gradually to approach the relatively constant volume of the control group by 3 months. The opposite trend occurred with respect to the 24-hour urinary excretion of calcium. There were no significant changes in the 24-hour urinary pH or excretions of oxalate, uric acid and alcian blue precipitable polyanions. The over-all effect was to cause a progressive increase in the probability of stones forming in the patients. The accurate assessment of the urinary risk factors of calcium stone disease requires at least a 3-month delay following acute renal colic. This delay usually will provide sufficient time for the stone to pass and for the patient to return to the normal dietary and fluid intake.

**URL:** <https://ovidsp.ovid.com/ovidweb.cgi?T=JS&CSC=Y&NEWS=N&PAGE=fulltext&D=med2&DO=10.1016%2fs0022-5347%2817%2950064-6> <https://libkey.io/libraries/2592/openurl?genre=article&sid=OVID:medline&id=pmid:6502804&id=doi:10.1016%2Fs0022-5347%2817%2950064-6&issn=0022-5347&isbn=&volume=132&issue=6&spage=1137&pages=1137-9&date=1984&title=Journal+of+Urology&atitle=When+should+patients+with+symptomatic+urinary+stone+disease+be+evaluated+metabolically%3F.&aulast=Norman&pid=%3Cauthor%3ENorman+RW%3BBath+SS%3BRobertson+WG%3BPeacock+M%3C%2Fauthor%3E%3CAN%3E6502804%3C%2FAN%3E%3CDT%3EJournal+Article%3C%2FDT%3E>

210. **Heterozygous cystinuria and calcium oxalate urolithiasis.**

**Item Type:** Journal Article

**Authors:** Carpentier, P. J.;Kurth, K. H.;Blom, W. and Huijmans, J. G.

**Publication Date:** Au ,1983

**Journal:** Journal of Urology 130(2), pp. 302–304

**Abstract:** To evaluate the incidence of heterozygous cystinuria in patients with primary and recurrent calcium oxalate stone disease 24-hour urine samples from 140 patients were examined. The frequency of heterozygous cystinuria in this study was only 1.4 per cent (2 patients). The frequency of other metabolic disorders was in accordance with other reports: hypercalciuria (32 per cent), hyperuricosuria (32 per cent) and hyperoxaluria (17 per cent).

**URL:** <https://ovidsp.ovid.com/ovidweb.cgi?T=JS&CSC=Y&NEWS=N&PAGE=fulltext&D=med2&DO=10.1016%2fs0022-5347%2817%2951119-2> <https://libkey.io/libraries/2592/openurl?genre=article&sid=OVID:medline&id=pmid:6876277&id=doi:10.1016%2Fs0022-5347%2817%2951119-2&issn=0022-5347&isbn=&volume=130&issue=2&spage=302&pages=302-4&date=1983&title=Journal+of+Urology&atitle=Heterozygous+cystinuria+and+calcium+oxalate+urolithiasis.&aulast=Carpentier&pid=%3Cauthor%3ECarpentier+PJ%3BKurth+KH%3BBlom+W%3BHuijmans+JG%3C%2Fauthor%3E%3CAN%3E6876277%3C%2FAN%3E%3CDT%3EJournal+Article%3C%2FDT%3E>

211. **Methodological aspects of examination of 24 hour urinary excretions in outpatients with recurrent urolithiasis.**

**Item Type:** Journal Article

**Authors:** Laerum, E. and Palmer, H.

**Publication Date:** 1983

**Journal:** Scandinavian Journal of Urology & Nephrology 17(3), pp. 321–324

**Abstract:** In 85 recurrent stone formers, 2 consecutive 24 h urinary excretions were subjected to chemical examination for calcium, uric acid, magnesium and creatinine. 23 patients had hypercalciuria (greater than 6 mmol/24 h), 22 hyperuricosuria (greater than 3.5 mmol/24 h) and 5 hypomagnesuria (less than 2.4 mmol/24 h). In 1/3 of the cases, the diagnosis would have been missed if based on only one of the two collections, rather than on the mean values from both. Calcium-restricted diet 3 days before and during the sampling is not recommended. Hydrochloric acid appeared to be useful as a preservative and to prevent calcium and magnesium precipitation. No significant seasonal variations were found. Heating of the urine sample before analysis of uric acid did not influence the result.

**URL:** <https://ovidsp.ovid.com/ovidweb.cgi?T=JS&CSC=Y&NEWS=N&PAGE=fulltext&D=med2&DO=10.3109%2f00365598309182139> <https://libkey.io/libraries/2592/openurl?genre=article&sid=OVID:medline&id=pmid:6648379&id=doi:10.3109%2F00365598309182139&issn=0036-5599&isbn=&volume=17&issue=3&spage=321&pages=321-4&date=1983&title=Scandinavian+Journal+of+Urology+%26+Nephrology&atitle=Methodological+aspects+of+examination+of+24+hour+urinary+excretions+in+outpatients+with+recurrent+urolithiasis.&aulast=Laerum&pid=%3Cauthor%3ELaerum+E%3BPalmer+H%3C%2Fauthor%3E%3CAN%3E6648379%3C%2FAN%3E%3CDT%3EJournal+Article%3C%2FDT%3E>

212. **Effect of large doses of ascorbic acid in man on some nitrogenous components of urine.**

**Item Type:** Journal Article

**Authors:** Sutton, J. L.;Basu, T. K. and Dickerson, J. W.

**Publication Date:** Ap ,1983

**Journal:** Human Nutrition - Applied Nutrition 37(2), pp. 136–140

**Abstract:** Base-line values for the excretion of urea N, amine N, creatinine, uric acid and cysteine were measured in three consecutive 24-h urine collections from 16 healthy volunteers. The subjects then took 1 g ascorbic acid three times a day after meals for 7 days. Twenty-four hour urine samples were collected on the 1st, 3rd, 5th and 7th days of the study. Where possible, a further 24-h sample was collected a minimum of 10 d after the end of the study. Administration of 3 g/d of ascorbic acid had no effect on urine volume, pH or the excretion of urea, amino N or creatinine. It caused a transient increase in uric acid excretion but an immediate and sustained increase in the excretion of cysteine. This study provides no evidence for an effect of high doses of ascorbic acid on urinary stone formation but does suggest competition for important co-factors in the metabolism of drugs.

**URL:** <https://ovidsp.ovid.com/ovidweb.cgi?T=JS&CSC=Y&NEWS=N&PAGE=fulltext&D=med2&AN=6863023> <https://libkey.io/libraries/2592/openurl?genre=article&sid=OVID:medline&id=pmid:6863023&id=doi:&issn=0263-8495&isbn=&volume=37&issue=2&spage=136&pages=136-40&date=1983&title=Human+Nutrition+-+Applied+Nutrition&atitle=Effect+of+large+doses+of+ascorbic+acid+in+man+on+some+nitrogenous+components+of+urine.&aulast=Sutton&pid=%3Cauthor%3ESutton+JL%3BBasu+TK%3BDickerson+JW%3C%2Fauthor%3E%3CAN%3E6863023%3C%2FAN%3E%3CDT%3EJournal+Article%3C%2FDT%3E>

213. **Changes in electrolytes, urea and creatinine excretion during dry and wet seasons.**

**Item Type:** Journal Article

**Authors:** Taylor, G. O.;Suvarnabai, P. C.;Fajimi, J. L. and Adenaike, F. A.

**Publication Date:** 1983

**Journal:** African Journal of Medicine & Medical Sciences 12(3-4), pp. 133–137

**Abstract:** Twenty-four-hour urine samples were collected from 179 male and female medical students in Ibadan, and 112 medical students in Zaria. The values for 24-h urine volume, urinary electrolytes, urea and creatinine exhibited a skewed distribution and were normalized by logarithmic transformation. The geometric means values for the 24-h urine volume for the dry season were significantly less than those for the wet season in Ibadan and Zaria. The mean urinary electrolytes, urea, and creatinine levels were significantly less during the dry season than during the wet season at Ibadan. In the Zaria subjects the changes in these urinary constituents during the dry and wet seasons were the reverse of those at Ibadan. These results could explain the absence of significant seasonal changes in urinary specific gravity and the low occurrence of urinary calculi in the southern part of Nigeria.

**URL:** <https://ovidsp.ovid.com/ovidweb.cgi?T=JS&CSC=Y&NEWS=N&PAGE=fulltext&D=med2&AN=6326546> <https://libkey.io/libraries/2592/openurl?genre=article&sid=OVID:medline&id=pmid:6326546&id=doi:&issn=0309-3913&isbn=&volume=12&issue=3-4&spage=133&pages=133-7&date=1983&title=African+Journal+of+Medicine+%26+Medical+Sciences&atitle=Changes+in+electrolytes%2C+urea+and+creatinine+excretion+during+dry+and+wet+seasons.&aulast=Taylor&pid=%3Cauthor%3ETaylor+GO%3BSuvarnabai+PC%3BFajimi+JL%3BAdenaike+FA%3C%2Fauthor%3E%3CAN%3E6326546%3C%2FAN%3E%3CDT%3EJournal+Article%3C%2FDT%3E>

214. **The diagnostic evaluation of risk factors for urinary tract stones: an analysis of care patterns in five hospitals.**

**Item Type:** Journal Article

**Authors:** Jekel, J. F.;Chauncey, K. J.;Moore, N. L.;Gowdy, D. P.;Broadus, A. E. and Black, H. R.

**Publication Date:** 1982

**Journal:** Yale Journal of Biology & Medicine 55(2), pp. 89–100

**Abstract:** Medical records of hospitalized primary urolithiasis patients in five Connecticut hospitals were studied to determine the aggressiveness of the diagnostic search for metabolic and other renal stone risk factors. A total of 924 patients over three years were analyzed. Routine serum, urine, and X-ray tests were generally performed according to accepted criteria. However, medical histories were generally inadequate, and 24-hour urine studies for calcium and uric acid were performed on less than half of the patients for whom they were indicated. Neither the complexity of the stone problem nor the stone event number appeared to influence the intensity of the diagnostic search in any important way. Other deficiencies included the lack of followup of abnormal test findings and the lack of treatment of problems discovered. The diagnostic approach in each hospital was stable over time.

**URL:** <https://ovidsp.ovid.com/ovidweb.cgi?T=JS&CSC=Y&NEWS=N&PAGE=fulltext&D=med2&AN=7136009> <https://libkey.io/libraries/2592/openurl?genre=article&sid=OVID:medline&id=pmid:7136009&id=doi:&issn=0044-0086&isbn=&volume=55&issue=2&spage=89&pages=89-100&date=1982&title=Yale+Journal+of+Biology+%26+Medicine&atitle=The+diagnostic+evaluation+of+risk+factors+for+urinary+tract+stones%3A+an+analysis+of+care+patterns+in+five+hospitals.&aulast=Jekel&pid=%3Cauthor%3EJekel+JF%3BChauncey+KJ%3BMoore+NL%3BGowdy+DP%3BBroadus+AE%3BBlack+HR%3C%2Fauthor%3E%3CAN%3E7136009%3C%2FAN%3E%3CDT%3EJournal+Article%3C%2FDT%3E>

215. **Urolithiasis risk factors in continent reservoir ileostomy patients.**

**Item Type:** Journal Article

**Authors:** Stern, H.;Cohen, Z.;Wilson, D. R. and Mickle, D. A.

**Publication Date:** 1980

**Journal:** Diseases of the Colon & Rectum 23(8), pp. 556–558

**Abstract:** Conventional ileostomy patients are at an increased risk to urinary stone formation compared to normal controls. This study was designed to evaluate any further risk factors to urinary stone formation in patients with Kock ileostomies. Nine Kock ileostomy patients were matched for age, sex, and body weight with nine conventional ileostomy patients and nine controls. Two 24-hour urine samples from each patient were analyzed for volume, pH, Na+/K+ ratio, oxalate, and uric acid concentration. Both ileostomy groups demonstrated reduced urinary volume and Na+/K+ ratio as compared to the control groups (P less than 0.05). The Kock ileostomy group had the lowest urinary volume. There was no significant reduction in urinary pH or elevation in urine uric acid concentration in the Kock ileostomy group. The results suggest that there is no significantly added risk to uric acid stone formation in Kock ileostomy patients.

**URL:** <https://ovidsp.ovid.com/ovidweb.cgi?T=JS&CSC=Y&NEWS=N&PAGE=fulltext&D=med2&DO=10.1007%2fBF02988995> <https://libkey.io/libraries/2592/openurl?genre=article&sid=OVID:medline&id=pmid:7460692&id=doi:10.1007%2FBF02988995&issn=0012-3706&isbn=&volume=23&issue=8&spage=556&pages=556-8&date=1980&title=Diseases+of+the+Colon+%26+Rectum&atitle=Urolithiasis+risk+factors+in+continent+reservoir+ileostomy+patients.&aulast=Stern&pid=%3Cauthor%3EStern+H%3BCohen+Z%3BWilson+DR%3BMickle+DA%3C%2Fauthor%3E%3CAN%3E7460692%3C%2FAN%3E%3CDT%3EJournal+Article%3C%2FDT%3E>

216. **Validity of biochemical findings in the evaluation of patients with urolithiasis.**

**Item Type:** Journal Article

**Authors:** Tiselius, H. G. and Larsson, L.

**Publication Date:** 1980

**Journal:** European Urology 6(2), pp. 90–94

**Abstract:** Urinary excretion of calcium, magnesium and oxalate was studied in 38 patients with urolithiasis on two different occasions, and there appeared to be a good correlation between the biochemical findings in the two samples. All values were expressed per mole of creatinine and it was furthermore demonstrated that the variation in creatinine excretion was considerably less than the variation in urine volume. The calcium/magnesium quotients were calculated in 113 2-hour fasting urine samples and 24-hour urine samples and a good correlation was obtained. Biochemical grouping of the patients was performed by means of the two sets of values and the result obtained was approximately the same in both cases.

**URL:** <https://ovidsp.ovid.com/ovidweb.cgi?T=JS&CSC=Y&NEWS=N&PAGE=fulltext&D=med2&DO=10.1159%2f000473298> <https://libkey.io/libraries/2592/openurl?genre=article&sid=OVID:medline&id=pmid:7358065&id=doi:10.1159%2F000473298&issn=0302-2838&isbn=&volume=6&issue=2&spage=90&pages=90-4&date=1980&title=European+Urology&atitle=Validity+of+biochemical+findings+in+the+evaluation+of+patients+with+urolithiasis.&aulast=Tiselius&pid=%3Cauthor%3ETiselius+HG%3BLarsson+L%3C%2Fauthor%3E%3CAN%3E7358065%3C%2FAN%3E%3CDT%3EJournal+Article%3C%2FDT%3E>

217. **Renal excretion of hydrogen ions in stone formers.**

**Item Type:** Journal Article

**Authors:** Marya, R. K.;Dadoo, R. C.;Khurana, S.;Keshwani, R. K. and Chhabra, H. L.

**Publication Date:** 1979

**Journal:** Urologia Internationalis 34(5), pp. 363–368

**Abstract:** Urinary hydrogen ion excretion has been studied in 68 adult stone formers and in 50 controls. Under basal state, the stone formers showed statistically significant higher urinary pH, and lower 24-hour urinary titrable acid, ammonium and total hydrogen. On the 3rd day of ammonium chloride loading test, the stone formers showed statistically significant higher urinary pH and lower 24-hour urinary titrable acid, ammonium, total hydrogen and plasma bicarbonate. 'Clearance' indices of hydrogen ion excretion were also depressed in a great majority of the stone formers. The results indicate the frequent co-existence of defective urinary acidification with nephrolithiasis.

**URL:** <https://ovidsp.ovid.com/ovidweb.cgi?T=JS&CSC=Y&NEWS=N&PAGE=fulltext&D=med1&DO=10.1159%2f000280282> <https://libkey.io/libraries/2592/openurl?genre=article&sid=OVID:medline&id=pmid:40335&id=doi:10.1159%2F000280282&issn=0042-1138&isbn=&volume=34&issue=5&spage=363&pages=363-8&date=1979&title=Urologia+Internationalis&atitle=Renal+excretion+of+hydrogen+ions+in+stone+formers.&aulast=Marya&pid=%3Cauthor%3EMarya+RK%3BDadoo+RC%3BKhurana+S%3BKeshwani+RK%3BChhabra+HL%3C%2Fauthor%3E%3CAN%3E40335%3C%2FAN%3E%3CDT%3EJournal+Article%3C%2FDT%3E>

218. **Heterozygous cystinuria and calcium oxalate urolithiasis.**

**Item Type:** Journal Article

**Authors:** Resnick, M. I.;Goodman, H. O. and Boyce, W. H.

**Publication Date:** Ju ,1979

**Journal:** Journal of Urology 122(1), pp. 52–54

**Abstract:** Many variables are known to be associated with the formation of calcium oxalate urolithiasis but none is essential for the initiation or growth of stones. It is likely that the predisposition to stone formation is related to multiple factors. We herein describe still another metabolic state that seems to predispose to calcium oxalate stone disease, namely heterozygosity for cystinuria. Cystine screening tests were done on 24-hour urine specimens obtained from 126 patients in whom recurrent calcium oxalate stones form and 84 controls and quantitative amino acid determinations were done on all positive specimens. Of those studied 17 of 126 stone patients and 1 of 84 controls were heterozygous cystinurics. A test of the differences between the relative frequencies of cystinuria heterozygotes in the 2 groups with Fisher's exact test revealed them to be highly significant (p less than 0.001). Our study indicates that carrier status for 1 of the cystinuria genes predisposes to calcium oxalate stone formation but, like other factors related to urolithiasis, it is not a necessary cause of stone disease.

**URL:** <https://ovidsp.ovid.com/ovidweb.cgi?T=JS&CSC=Y&NEWS=N&PAGE=fulltext&D=med1&DO=10.1016%2fs0022-5347%2817%2956248-5> <https://libkey.io/libraries/2592/openurl?genre=article&sid=OVID:medline&id=pmid:458988&id=doi:10.1016%2Fs0022-5347%2817%2956248-5&issn=0022-5347&isbn=&volume=122&issue=1&spage=52&pages=52-4&date=1979&title=Journal+of+Urology&atitle=Heterozygous+cystinuria+and+calcium+oxalate+urolithiasis.&aulast=Resnick&pid=%3Cauthor%3EResnick+MI%3BGoodman+HO%3BBoyce+WH%3C%2Fauthor%3E%3CAN%3E458988%3C%2FAN%3E%3CDT%3EJournal+Article%3C%2FDT%3E>

219. **The effect of seasonal changes and socio-economic status on urinary pH and specific gravity.**

**Item Type:** Journal Article

**Authors:** Taylor, G. O.;Oyediran, A. B. and Adesina, H. A.

**Publication Date:** Ma ,1979

**Journal:** Tropical & Geographical Medicine 31(1), pp. 105–110

**Abstract:** Specific gravity and pH of urine in the dry and the wet season in three socio-economic Nigerian groups have been studied. Samples were collected at 7 a.m., 11 a.m. and 3 p.m. High-income subjects had a significantly lower urinary pH than low-income. During the wet season pH values were lower than in the dry, but not significantly so. During the day S.G. values did not differ, neither did they in different seasons. But there were significant differences between high and low income groups. S.G. and pH samples of high and low income groups were similar in children and in their parents. In medical students S.G. and pH in 24-hour urines were similar to the morning value of high and middle income groups and were not affected by season, but the volume was. The relevance of these findings is discussed relative to the rarity of urinary calculi in Nigerians.

**URL:** <https://ovidsp.ovid.com/ovidweb.cgi?T=JS&CSC=Y&NEWS=N&PAGE=fulltext&D=med1&AN=39374> <https://libkey.io/libraries/2592/openurl?genre=article&sid=OVID:medline&id=pmid:39374&id=doi:&issn=0041-3232&isbn=&volume=31&issue=1&spage=105&pages=105-10&date=1979&title=Tropical+%26+Geographical+Medicine&atitle=The+effect+of+seasonal+changes+and+socio-economic+status+on+urinary+pH+and+specific+gravity.&aulast=Taylor&pid=%3Cauthor%3ETaylor+GO%3BOyediran+AB%3BAdesina+HA%3C%2Fauthor%3E%3CAN%3E39374%3C%2FAN%3E%3CDT%3EJournal+Article%3C%2FDT%3E>

220. **Oral calcium tolerance and urinary cyclic AMP in urolithiasis.**

**Item Type:** Journal Article

**Authors:** Nelson, J. H. 3rd;Rehm, R. A.;Riemenschneider, H. W.;Wise, H. A. 2nd;Pflug, B.;Kemple, K.;Whitehouse, W. K. and Winter, C. C.

**Publication Date:** No ,1978

**Journal:** Urology 12(5), pp. 519–524

**Abstract:** Oral calcium tolerance and urinary cyclic AMP testing was used in the evaluation of 61 unselected patients with stones. The oral calcium tolerance test was easy to perform and was useful in defining several distinct metabolic abnormalities contributing to calculous formation. Oral calcium tolerance testing is more precise than twenty-four-hour urinary calcium determination and should provide a means of determining proper medical treatment of urolithiasis. Urinary cyclic AMP was disappointing as a measure of parathormone activity.

**URL:** <https://ovidsp.ovid.com/ovidweb.cgi?T=JS&CSC=Y&NEWS=N&PAGE=fulltext&D=med1&DO=10.1016%2f0090-4295%2878%2990464-8> <https://libkey.io/libraries/2592/openurl?genre=article&sid=OVID:medline&id=pmid:214923&id=doi:10.1016%2F0090-4295%2878%2990464-8&issn=0090-4295&isbn=&volume=12&issue=5&spage=519&pages=519-24&date=1978&title=Urology&atitle=Oral+calcium+tolerance+and+urinary+cyclic+AMP+in+urolithiasis.&aulast=Nelson&pid=%3Cauthor%3ENelson+JH+3rd%3BRehm+RA%3BRiemenschneider+HW%3BWise+HA+2nd%3BPflug+B%3BKemple+K%3BWhitehouse+WK%3BWinter+CC%3C%2Fauthor%3E%3CAN%3E214923%3C%2FAN%3E%3CDT%3EJournal+Article%3C%2FDT%3E>

221. **Seasonal variations in urinary crystals.**

**Item Type:** Journal Article

**Authors:** Hallson, P. C. and Rose, G. A.

**Publication Date:** Au ,1977

**Journal:** British Journal of Urology 49(4), pp. 277–284

**Abstract:** Individual urine samples from normal subjects and stone-formers with idiopathic hypercalciuria have been examined for crystals both qualitatively and quantitatively at 37 degrees C. The group as a whole showed a rise in incidence of urinary crystals in the summer months of June to August inclusive. This rise was seen most clearly in overnight urines, collected on rising in the morning, and the patients appeared to be at risk overnight during the summer. In the untreated patients the summer rise in incidence of phosphate crystals was quite dramatic but was only small in the cellulose phosphate treated group, who showed a rather constant and raised incidence of oxalate crystals right through the year. Seasonal crystal incidence has been compared with seasonal changes in urinary composition. The rise in crystal incidence during the summer was associated with increased creatinine concentration in the same urine samples and with increased oxalate concentration in 24-hour urine collections.

**URL:** <https://ovidsp.ovid.com/ovidweb.cgi?T=JS&CSC=Y&NEWS=N&PAGE=fulltext&D=med1&DO=10.1111%2fj.1464-410x.1977.tb04137.x> <https://libkey.io/libraries/2592/openurl?genre=article&sid=OVID:medline&id=pmid:912253&id=doi:10.1111%2Fj.1464-410x.1977.tb04137.x&issn=0007-1331&isbn=&volume=49&issue=4&spage=277&pages=277-84&date=1977&title=British+Journal+of+Urology&atitle=Seasonal+variations+in+urinary+crystals.&aulast=Hallson&pid=%3Cauthor%3EHallson+PC%3BRose+GA%3C%2Fauthor%3E%3CAN%3E912253%3C%2FAN%3E%3CDT%3EJournal+Article%3C%2FDT%3E>

222. **Urinary citrate excretion in stone-formers and normal controls.**

**Item Type:** Journal Article

**Authors:** Welshman, S. G. and McGeown, M. G.

**Publication Date:** Fe ,1976

**Journal:** British Journal of Urology 48(1), pp. 7–11

**Abstract:** A specific method was used for the estimation of citrate in 24-hour urine collections from 108 young adult controls, 158 patient controls and 164 stone-formers. Stone-formers excreted significantly less citrate in 24 hours than either patient controls or young adult controls. Stone-formers had a lower concentration of citrate in their urine than either of the control groups. The young adult females exhibited a much greater excretion of citrate relative to calcium than the young males. Because of the ability of citrate to complex with calcium ions and keep them in solution, the relatively low incidence of calcium-containing stones in females under 50 years of age could well be the result of their high excretion of citrate and their increased excretion of this substance relative to calcium.

**URL:** <https://ovidsp.ovid.com/ovidweb.cgi?T=JS&CSC=Y&NEWS=N&PAGE=fulltext&D=med1&DO=10.1111%2fj.1464-410x.1976.tb02731.x> <https://libkey.io/libraries/2592/openurl?genre=article&sid=OVID:medline&id=pmid:1268467&id=doi:10.1111%2Fj.1464-410x.1976.tb02731.x&issn=0007-1331&isbn=&volume=48&issue=1&spage=7&pages=7-11&date=1976&title=British+Journal+of+Urology&atitle=Urinary+citrate+excretion+in+stone-formers+and+normal+controls.&aulast=Welshman&pid=%3Cauthor%3EWelshman+SG%3BMcGeown+MG%3C%2Fauthor%3E%3CAN%3E1268467%3C%2FAN%3E%3CDT%3EComparative+Study%3C%2FDT%3E>

223. **Urinary calcium excretion in relation to kidney function in the adult.**

**Item Type:** Journal Article

**Authors:** Goldschmied, A.;Modan, B.;Greenberg, R. A.;Zurkowski, S. and Modan, M.

**Publication Date:** Ap ,1975

**Journal:** Journal of the American Geriatrics Society 23(4), pp. 155–160

**Abstract:** A study based on 55 hospital patients (age groups 40-59, 60-69, 70-79) demonstrated a significant correlation between the glomerular filtration rate (as expressed by endogenous creatinine clearance) and the mean urinary calcium concentration. Excluded were patients with thyroidal or hypophyseal disorders, osteoporosis, renal stones immobilizing conditions, or those treated with corticosteroids. The study patients had a normal fluid intake and did not receive any diuretics. The urinary calcium concentration test is less laborious and less liable to laboratory errors than the creatinine clearance test, which requires a 24-hour urine collection. Because of the gradual decrease in the glomerular filtration rate in the elderly, this test should be of value in geriatric practice.

**URL:** <https://ovidsp.ovid.com/ovidweb.cgi?T=JS&CSC=Y&NEWS=N&PAGE=fulltext&D=med1&DO=10.1111%2fj.1532-5415.1975.tb00161.x> <https://libkey.io/libraries/2592/openurl?genre=article&sid=OVID:medline&id=pmid:1117116&id=doi:10.1111%2Fj.1532-5415.1975.tb00161.x&issn=0002-8614&isbn=&volume=23&issue=4&spage=155&pages=155-60&date=1975&title=Journal+of+the+American+Geriatrics+Society&atitle=Urinary+calcium+excretion+in+relation+to+kidney+function+in+the+adult.&aulast=Goldschmied&pid=%3Cauthor%3EGoldschmied+A%3BModan+B%3BGreenberg+RA%3BZurkowski+S%3BModan+M%3C%2Fauthor%3E%3CAN%3E1117116%3C%2FAN%3E%3CDT%3EJournal+Article%3C%2FDT%3E>

224. **The relationship of the urinary cations, Calcium, Magnesium, Sodium and Potassium, in patients with Renal Calculi.**

**Item Type:** Journal Article

**Authors:** Welshman, S. G. and McGeown, M. G.

**Publication Date:** Ju ,1975

**Journal:** British Journal of Urology 47(3), pp. 237–242

**Abstract:** Calcium, magnesium, sodium and potassium were estimated in the 24-hour urine collections of 101 idiopathic male stone-formers, 89 male patient controls and 59 young male adult controls. The results were calculated in terms of 24-hour volume and 1 g of creatinine. The concentrations of the 4 cations, relative to a gram of creatinine, were also determined in the early-morning urines of 41 male stone-formers and 13 young male adult controls. No difference was observed in the 24-hour excretion of calcium, magnesium and sodium between the stone-formers and controls. The mean daily potassium excretion, however, was significantly reduced in the urine of stone-formers. The linear regression equations were calculated for calcium on magnesium, calcium on sodium, and calcium on potassium, using the 24-hour excretion values of these cations. Only the calcium on potassium line of stone-formers was significantly different from that of the normal subjects. A significant increase by stone-formers in the urinary calcium concentration of their early-morning specimens was found. The high concentration of urinary calcium in overnight urines of stone-formers combined with a low magnesium concentration might possibly contribute to the development of renal stone disease.

**URL:** <https://ovidsp.ovid.com/ovidweb.cgi?T=JS&CSC=Y&NEWS=N&PAGE=fulltext&D=med1&DO=10.1111%2fj.1464-410x.1975.tb03956.x> <https://libkey.io/libraries/2592/openurl?genre=article&sid=OVID:medline&id=pmid:1139114&id=doi:10.1111%2Fj.1464-410x.1975.tb03956.x&issn=0007-1331&isbn=&volume=47&issue=3&spage=237&pages=237-42&date=1975&title=British+Journal+of+Urology&atitle=The+relationship+of+the+urinary+cations%2C+Calcium%2C+Magnesium%2C+Sodium+and+Potassium%2C+in+patients+with+Renal+Calculi.&aulast=Welshman&pid=%3Cauthor%3EWelshman+SG%3BMcGeown+MG%3C%2Fauthor%3E%3CAN%3E1139114%3C%2FAN%3E%3CDT%3EComparative+Study%3C%2FDT%3E>

*You will need your* [*NHS OpenAthens account*](https://openathens.nice.org.uk/) *to access the full text of licenced content.*
*This service is provided to the NHS in England by NHSE Workforce, Training & Education.*
