## Appendix 1 for "Investigating 24-Hour Urine Expected Ranges of Non-Kidney Stone Formers: an EAU Endourology Research Group Systematic Review and Meta-Analysis"

**Search Terms:**

**Database:**
Ovid MEDLINE(R) ALL <1946 to November 27, 2024>

| **#** | **Query** | **Results from 29 Nov 2024** |
| --- | --- | --- |
| 1 | 24 hour.mp. [mp=title, book title, abstract, original title, name of substance word, subject heading word, floating sub-heading word, keyword heading word, organism supplementary concept word, protocol supplementary concept word, rare disease supplementary concept word, unique identifier, synonyms, population supplementary concept word, anatomy supplementary concept word] | 51,022 |
| 2 | 24-hour.mp. [mp=title, book title, abstract, original title, name of substance word, subject heading word, floating sub-heading word, keyword heading word, organism supplementary concept word, protocol supplementary concept word, rare disease supplementary concept word, unique identifier, synonyms, population supplementary concept word, anatomy supplementary concept word] | 51,022 |
| 3 | 24 - hour.mp. [mp=title, book title, abstract, original title, name of substance word, subject heading word, floating sub-heading word, keyword heading word, organism supplementary concept word, protocol supplementary concept word, rare disease supplementary concept word, unique identifier, synonyms, population supplementary concept word, anatomy supplementary concept word] | 51,022 |
| 4 | Twenty four hour.mp. [mp=title, book title, abstract, original title, name of substance word, subject heading word, floating sub-heading word, keyword heading word, organism supplementary concept word, protocol supplementary concept word, rare disease supplementary concept word, unique identifier, synonyms, population supplementary concept word, anatomy supplementary concept word] | 5,660 |
| 5 | Twenty-four hour.mp. [mp=title, book title, abstract, original title, name of substance word, subject heading word, floating sub-heading word, keyword heading word, organism supplementary concept word, protocol supplementary concept word, rare disease supplementary concept word, unique identifier, synonyms, population supplementary concept word, anatomy supplementary concept word] | 5,660 |
| 6 | Twenty - four hour.mp. [mp=title, book title, abstract, original title, name of substance word, subject heading word, floating sub-heading word, keyword heading word, organism supplementary concept word, protocol supplementary concept word, rare disease supplementary concept word, unique identifier, synonyms, population supplementary concept word, anatomy supplementary concept word] | 5,660 |
| 7 | 1 or 2 or 3 or 4 or 5 or 6 | 55,509 |
| 8 | urine collection.mp. [mp=title, book title, abstract, original title, name of substance word, subject heading word, floating sub-heading word, keyword heading word, organism supplementary concept word, protocol supplementary concept word, rare disease supplementary concept word, unique identifier, synonyms, population supplementary concept word, anatomy supplementary concept word] | 4,121 |
| 9 | (urine and collect*).mp. [mp=title, book title, abstract, original title, name of substance word, subject heading word, floating sub-heading word, keyword heading word, organism supplementary concept word, protocol supplementary concept word, rare disease supplementary concept word, unique identifier, synonyms, population supplementary concept word, anatomy supplementary concept word] | 52,079 |
| 10 | (urine and test*).mp. [mp=title, book title, abstract, original title, name of substance word, subject heading word, floating sub-heading word, keyword heading word, organism supplementary concept word, protocol supplementary concept word, rare disease supplementary concept word, unique identifier, synonyms, population supplementary concept word, anatomy supplementary concept word] | 93,322 |
| 11 | (urine and sample*).mp. [mp=title, book title, abstract, original title, name of substance word, subject heading word, floating sub-heading word, keyword heading word, organism supplementary concept word, protocol supplementary concept word, rare disease supplementary concept word, unique identifier, synonyms, population supplementary concept word, anatomy supplementary concept word] | 83,921 |
| 12 | 8 or 9 or 10 or 11 | 167,782 |
| 13 | exp Urine Specimen Collection/mt [Methods] | 328 |
| 14 | exp Urinalysis/mt [Methods] | 3,392 |
| 15 | exp Urine/ch [Chemistry] | 4,592 |
| 16 | exp Urinalysis/sn [Statistics & Numerical Data] | 247 |
| 17 | exp Urine Specimen Collection/sn [Statistics & Numerical Data] | 24 |
| 18 | 13 or 14 or 15 or 16 or 17 | 8,222 |
| 19 | Kidney Calculi/ | 22,268 |
| 20 | (kidney and calculi).mp. [mp=title, book title, abstract, original title, name of substance word, subject heading word, floating sub-heading word, keyword heading word, organism supplementary concept word, protocol supplementary concept word, rare disease supplementary concept word, unique identifier, synonyms, population supplementary concept word, anatomy supplementary concept word] | 26,882 |
| 21 | kidney stone*.mp. | 8,357 |
| 22 | kidney stone*.mp. | 8,357 |
| 23 | 19 or 20 or 21 or 22 | 30,096 |
| 24 | 7 and 12 and 23 | 373 |
| 25 | 7 and 18 and 23 | 41 |
| 26 | 24 or 25 | 378 |
| 27 | limit 26 to (english language and humans) | 303 |

24 hour.mp. [mp=title, book title, abstract, original title, name of substance word, subject heading word, floating sub-heading word, keyword heading word, organism supplementary concept word, protocol supplementary concept word, rare disease supplementary concept word, unique identifier, synonyms, population supplementary concept word, anatomy supplementary concept word]
24-hour.mp. [mp=title, book title, abstract, original title, name of substance word, subject heading word, floating sub-heading word, keyword heading word, organism supplementary concept word, protocol supplementary concept word, rare disease supplementary concept word, unique identifier, synonyms, population supplementary concept word, anatomy supplementary concept word]
24 - hour.mp. [mp=title, book title, abstract, original title, name of substance word, subject heading word, floating sub-heading word, keyword heading word, organism supplementary concept word, protocol supplementary concept word, rare disease supplementary concept word, unique identifier, synonyms, population supplementary concept word, anatomy supplementary concept word]
Twenty four hour.mp. [mp=title, book title, abstract, original title, name of substance word, subject heading word, floating sub-heading word, keyword heading word, organism supplementary concept word, protocol supplementary concept word, rare disease supplementary concept word, unique identifier, synonyms, population supplementary concept word, anatomy supplementary concept word]
Twenty-four hour.mp. [mp=title, book title, abstract, original title, name of substance word, subject heading word, floating sub-heading word, keyword heading word, organism supplementary concept word, protocol supplementary concept word, rare disease supplementary concept word, unique identifier, synonyms, population supplementary concept word, anatomy supplementary concept word]
Twenty - four hour.mp. [mp=title, book title, abstract, original title, name of substance word, subject heading word, floating sub-heading word, keyword heading word, organism supplementary concept word, protocol supplementary concept word, rare disease supplementary concept word, unique identifier, synonyms, population supplementary concept word, anatomy supplementary concept word]
1 or 2 or 3 or 4 or 5 or 6
urine collection.mp. [mp=title, book title, abstract, original title, name of substance word, subject heading word, floating sub-heading word, keyword heading word, organism supplementary concept word, protocol supplementary concept word, rare disease supplementary concept word, unique identifier, synonyms, population supplementary concept word, anatomy supplementary concept word]
(urine and collect*).mp. [mp=title, book title, abstract, original title, name of substance word, subject heading word, floating sub-heading word, keyword heading word, organism supplementary concept word, protocol supplementary concept word, rare disease supplementary concept word, unique identifier, synonyms, population supplementary concept word, anatomy supplementary concept word]
(urine and test*).mp. [mp=title, book title, abstract, original title, name of substance word, subject heading word, floating sub-heading word, keyword heading word, organism supplementary concept word, protocol supplementary concept word, rare disease supplementary concept word, unique identifier, synonyms, population supplementary concept word, anatomy supplementary concept word]
(urine and sample*).mp. [mp=title, book title, abstract, original title, name of substance word, subject heading word, floating sub-heading word, keyword heading word, organism supplementary concept word, protocol supplementary concept word, rare disease supplementary concept word, unique identifier, synonyms, population supplementary concept word, anatomy supplementary concept word]
8 or 9 or 10 or 11
exp Urine Specimen Collection/mt [Methods]
exp Urinalysis/mt [Methods]
exp Urine/ch [Chemistry]
exp Urinalysis/sn [Statistics & Numerical Data]
exp Urine Specimen Collection/sn [Statistics & Numerical Data]
13 or 14 or 15 or 16 or 17
Kidney Calculi/
(kidney and calculi).mp. [mp=title, book title, abstract, original title, name of substance word, subject heading word, floating sub-heading word, keyword heading word, organism supplementary concept word, protocol supplementary concept word, rare disease supplementary concept word, unique identifier, synonyms, population supplementary concept word, anatomy supplementary concept word]
kidney stone*.mp.
kidney stone*.mp.
19 or 20 or 21 or 22
7 and 12 and 23
7 and 18 and 23
24 or 25
limit 26 to (english language and humans)
