## Appendix 2 for "Investigating 24-Hour Urine Expected Ranges of Non-Kidney Stone Formers: an EAU Endourology Research Group Systematic Review and Meta-Analysis"

Evidence Search Results from your Knowledge & Library Service

1. **Predictive Modeling of Urinary Stone Composition Using Machine Learning and Clinical Data: Implications for Treatment Strategies and Pathophysiological Insights.**

**Item Type:** Journal Article

**Authors:** Chmiel, John A.;Stuivenberg, Gerrit A.;Wong, Jennifer F. W.;Nott, Linda;Burton, Jeremy P.;Razvi, Hassan and Bjazevic, Jennifer

**Publication Date:** Au ,2024

**Journal:** Journal of Endourology 38(8), pp. 778–787

**Abstract: Purpose:** Preventative strategies and surgical treatments for urolithiasis depend on stone composition. However, stone composition is often unknown until the stone is passed or surgically managed. Given that stone composition likely reflects the physiological parameters during its formation, we used clinical data from stone formers to predict stone composition. **Materials and Methods:** Data on stone composition, 24-hour urine, serum biochemistry, patient demographics, and medical history were prospectively collected from 777 kidney stone patients. Data were used to train gradient boosted machine and logistic regression models to distinguish calcium vs noncalcium, calcium oxalate monohydrate vs dihydrate, and calcium oxalate vs calcium phosphate vs uric acid stone types. Model performance was evaluated using the kappa score, and the influence of each predictor variable was assessed. **Results:** The calcium vs noncalcium model differentiated stone types with a kappa of 0.5231. The most influential predictors were 24-hour urine calcium, blood urate, and phosphate. The calcium oxalate monohydrate vs dihydrate model is the first of its kind and could discriminate stone types with a kappa of 0.2042. The key predictors were 24-hour urine urea, calcium, and oxalate. The multiclass model had a kappa of 0.3023 and the top predictors were age and 24-hour urine calcium and creatinine. **Conclusions:** Clinical data can be leveraged with machine learning algorithms to predict stone composition, which may help urologists determine stone type and guide their management plan before stone treatment. Investigating the most influential predictors of each classifier may improve the understanding of key clinical features of urolithiasis and shed light on pathophysiology.

**URL:** <https://ovidsp.ovid.com/ovidweb.cgi?T=JS&CSC=Y&NEWS=N&PAGE=fulltext&D=medl&DO=10.1089%2fend.2023.0446> <https://libkey.io/libraries/2592/openurl?genre=article&sid=OVID:medline&id=pmid:37975292&id=doi:10.1089%2Fend.2023.0446&issn=0892-7790&isbn=&volume=38&issue=8&spage=778&pages=778-787&date=2024&title=Journal+of+Endourology&atitle=Predictive+Modeling+of+Urinary+Stone+Composition+Using+Machine+Learning+and+Clinical+Data%3A+Implications+for+Treatment+Strategies+and+Pathophysiological+Insights.&aulast=Chmiel&pid=%3Cauthor%3EChmiel+JA%3BStuivenberg+GA%3BWong+JFW%3BNott+L%3BBurton+JP%3BRazvi+H%3BBjazevic+J%3C%2Fauthor%3E%3CAN%3E37975292%3C%2FAN%3E%3CDT%3EJournal+Article%3C%2FDT%3E>

2. **24-Hour Urinary Chemistries and Kidney Stone Risk.**

**Item Type:** Journal Article

**Authors:** Ferraro, Pietro Manuel;Taylor, Eric N. and Curhan, Gary C.

**Publication Date:** 2024

**Journal:** American Journal of Kidney Diseases 84(2), pp. 164–169

**Abstract: RATIONALE & OBJECTIVE**: Most previous studies of the relationship between urinary factors and kidney stone risk have either assumed a linear effect of urinary parameters on kidney stone risk or implemented arbitrary thresholds suggesting biologically implausible "all-or-nothing" effects. In addition, little is known about the hierarchy of effects of urinary factors on kidney stone risk. This study evaluated the independent associations between urine chemistries and kidney stone formation and examined their magnitude and shape. **STUDY DESIGN**: Prospective cohort study. **SETTING & PARTICIPANTS**: We analyzed 9,045 24-hour urine collections from 6,217 participants of the Health Professionals Follow-Up Study and Nurses' Health Studies I and II. **EXPOSURE**: Urine volume and pH, and concentrations of calcium, citrate, oxalate, potassium, magnesium, uric acid, phosphorus, and sodium. **OUTCOME**: Incident symptomatic kidney stones. **ANALYTICAL APPROACH**: Multivariable logistic regression analysis incorporating restricted cubic splines to explore potentially nonlinear relationships between urinary factors and the risk of forming a kidney stone. Optimal inflection point analysis was implemented for each factor, and dominance analysis was performed to establish the relative importance of each urinary factor. **RESULTS**: Each urinary factor was significantly associated with stone formation except for urine pH. Higher urinary levels of calcium, oxalate, phosphorus, and sodium were associated with a higher risk of stone formation whereas higher urine volume, uric acid, citrate, potassium, and magnesium were associated with a lower risk. The relationships were substantially linear for urine calcium, uric acid, and sodium. By contrast, the magnitudes of the relationships were modestly attenuated at levels above the inflection points for urine oxalate, citrate, volume, phosphorus, potassium, and magnesium. Dominance analysis identified 3 categories of factors' relative importance: higher (calcium, volume, and citrate), intermediate (oxalate, potassium, and magnesium), and lower (uric acid, phosphorus, and sodium). **LIMITATIONS**: Predominantly White participants, lack of information on stone composition. **CONCLUSIONS**: Urine chemistries have complex relationships and differential relative associations with the risk of kidney stone formation. **PLAIN-LANGUAGE SUMMARY**: Kidney stones are common and likely to recur. Certain urinary factors play a role in the development of stones, but their independent roles, relative importance, and shapes of association with stone formation are not well-characterized. We analyzed 24-hour urine collections from individuals with and without kidney stones. Stones were less likely in those with higher urine volume, citrate, potassium, magnesium, and uric acid and were more likely in those with higher calcium, oxalate, phosphorus, and sodium. The acidity of the urine was not related to stones. The urinary parameters showed different degrees of relative importance, with calcium, volume, and citrate being greatest. All parameters exhibited a linear or close-to-linear shape of association with stone formation. Copyright © 2024 The Authors. Published by Elsevier Inc. All rights reserved.

**URL:** <https://ovidsp.ovid.com/ovidweb.cgi?T=JS&CSC=Y&NEWS=N&PAGE=fulltext&D=medl&DO=10.1053%2fj.ajkd.2024.02.010> <https://libkey.io/libraries/2592/openurl?genre=article&sid=OVID:medline&id=pmid:38583757&id=doi:10.1053%2Fj.ajkd.2024.02.010&issn=0272-6386&isbn=&volume=84&issue=2&spage=164&pages=164-169&date=2024&title=American+Journal+of+Kidney+Diseases&atitle=24-Hour+Urinary+Chemistries+and+Kidney+Stone+Risk.&aulast=Ferraro&pid=%3Cauthor%3EFerraro+PM%3BTaylor+EN%3BCurhan+GC%3C%2Fauthor%3E%3CAN%3E38583757%3C%2FAN%3E%3CDT%3EJournal+Article%3C%2FDT%3E>

3. **Real-World Effectiveness of Preventive Pharmacological Therapy in Patients With Urolithiasis: A Retrospective Cohort Study.**

**Item Type:** Journal Article

**Authors:** Hollingsworth, John M.;Oerline, Mary K.;Hsi, Ryan S.;Crivelli, Joseph J.;Krampe, Noah;Asplin, John R. and Shahinian, Vahakn B.

**Publication Date:** 2024

**Journal:** American Journal of Kidney Diseases 84(1), pp. 83–93.e1

**Abstract: RATIONALE & OBJECTIVE**: Data supporting the efficacy of preventive pharmacological therapy (PPT) to reduce urolithiasis recurrence are based on clinical trials with composite outcomes that incorporate imaging findings and have uncertain clinical significance. This study evaluated whether the use of PPT leads to fewer symptomatic stone events. **STUDY DESIGN**: Retrospective cohort study. **SETTING & PARTICIPANTS**: Medicare enrollees with urolithiasis who completed 24-hour urine collections that revealed hypercalciuria, hypocitraturia, low urine pH, or hyperuricosuria. **EXPOSURE**: PPT (thiazide diuretics for hypercalciuria, alkali for hypocitraturia or low urine pH, or uric acid lowering drugs for hyperuricosuria) categorized as (1) adherent to guideline-concordant PPT, (2) nonadherent to guideline-concordant PPT, or (3) untreated. **OUTCOME**: Symptomatic stone event occurrence (emergency department [ED] visit or hospitalization for urolithiasis or stone-directed surgery). **ANALYTICAL APPROACH**: Cox proportional hazards regression. **RESULTS**: Among 13,942 patients, 31.0% were prescribed PPT. Compared with no treatment, concordant/adherent PPT use was associated with a significantly lower hazard of symptomatic stone events for patients with hypercalciuria (HR, 0.736 [95% CI, 0.593-0.915]) and low urine pH (HR, 0.804 [95% CI, 0.650-0.996]) but not for patients with hypocitraturia or hyperuricosuria. These associations were largely driven by significantly lower rates of ED visits after initiating PPT among the concordant/adherent group versus untreated patients. Patients with hypercalciuria had adjusted 2-year predicted probabilities of a visit of 3.8% [95% CI, 2.5%-5.2%%] and 6.9% [95% CI, 6.0%-7.7%] for the concordant/adherent PPT and no-treatment groups, respectively. Among patients with low urine pH, these probabilities were 4.3% (95% CI, 2.9%-5.7%) and 7.3% (95% CI, 6.5%-8.0%) for the concordant/adherent PPT and no-treatment groups, respectively. **LIMITATIONS**: Potential bias from the possibility that patients prescribed PPT had more severe disease than untreated patients. **CONCLUSIONS**: Patients with urolithiasis and hypercalciuria who were adherent to treatment with thiazide diuretics as well as those with low urine pH adherent to prescribed alkali therapy had fewer symptomatic stone events than untreated patients. **PLAIN-LANGUAGE SUMMARY**: Despite multiple clinical trials demonstrating the efficacy of thiazide diuretics and alkali for secondary prevention of kidney stones, they are infrequently prescribed due in part to a lack of data about their effectiveness in real-world settings. We analyzed medical claims from older adults with kidney stones for whom urine chemistry data were available. We found that patients who took prescribed thiazide diuretics for elevated urine calcium levels or alkali for low urinary pH were less likely to experience symptomatic stone recurrences than untreated patients. This benefit was expressed as lower rates of emergency department visits after initiating therapy. Our findings should inform the prescription of and adherence to treatment with thiazide diuretics and alkali for the prevention of recurrent kidney stones. Copyright © 2024 National Kidney Foundation, Inc. Published by Elsevier Inc. All rights reserved.

**URL:** <https://ovidsp.ovid.com/ovidweb.cgi?T=JS&CSC=Y&NEWS=N&PAGE=fulltext&D=med25&DO=10.1053%2fj.ajkd.2023.12.015> <https://libkey.io/libraries/2592/openurl?genre=article&sid=OVID:medline&id=pmid:38432593&id=doi:10.1053%2Fj.ajkd.2023.12.015&issn=0272-6386&isbn=&volume=84&issue=1&spage=83&pages=83-93.e1&date=2024&title=American+Journal+of+Kidney+Diseases&atitle=Real-World+Effectiveness+of+Preventive+Pharmacological+Therapy+in+Patients+With+Urolithiasis%3A+A+Retrospective+Cohort+Study.&aulast=Hollingsworth&pid=%3Cauthor%3EHollingsworth+JM%3BOerline+MK%3BHsi+RS%3BCrivelli+JJ%3BKrampe+N%3BAsplin+JR%3BShahinian+VB%3C%2Fauthor%3E%3CAN%3E38432593%3C%2FAN%3E%3CDT%3EJournal+Article%3C%2FDT%3E>

4. **Management of urinary stones: state of the art and future perspectives by experts in stone disease**

**Item Type:** Journal Article

**Authors:** Papatsoris, Athanasios;Alba, Alberto Budia;Galan Llopis, Juan Antonio;Musafer, Murtadha Al;Alameedee, Mohammed;Ather, Hammad;Caballero-Romeu, Juan Pablo;Costa-Bauza, Antonia;Dellis, Athanasios;El Howairis, Mohamed;Gambaro, Giovanni;Geavlete, Bogdan;Halinski, Adam;Hess, Bernhard;Jaffry, Syed;Kok, Dirk;Kouicem, Hichem;Llanes, Luis;Lopez Martinez, Juan M.;Popov, Elenko, et al

**Publication Date:** Jun 27 ,2024

**Journal:** Archivio Italiano Di Urologia, Andrologia 96(2), pp. 12703

**Abstract: AIM**: To present state of the art on the management of urinary stones from a panel of globally recognized urolithiasis experts who met during the Experts in Stone Disease Congress in Valencia in January 2024. Options of treatment: The surgical treatment modalities of renal and ureteral stones are well defined by the guidelines of international societies, although for some index cases more alternative options are possible. For 1.5 cm renal stones, both m-PCNL and RIRS have proven to be valid treatment alternatives with comparable stone-free rates. The m-PCNL has proven to be more cost effective and requires a shorter operative time, while the RIRS has demonstrated lower morbidity in terms of blood loss and shorter recovery times. SWL has proven to be less effective at least for lower calyceal stones but has the highest safety profile. For a 6mm obstructing stone of the pelviureteric junction (PUJ) stone, SWL should be the first choice for a stone less than 1 cm, due to less invasiveness and lower risk of complications although it has a lower stone free-rate. RIRS has advantages in certain conditions such as anticoagulant treatment, obesity, or body deformity. Technical issues of the surgical procedures for stone removal: In patients receiving antithrombotic therapy, SWL, PCN and open surgery are at elevated risk of hemorrhage or perinephric hematoma. URS, is associated with less morbidity in these cases. An individualized combined evaluation of risks of bleeding and thromboembolism should determine the perioperative thromboprophylactic strategy. Pre-interventional urine culture and antibiotic therapy are mandatory although UTI treatment is becoming more challenging due to increasing resistance to routinely applied antibiotics. The use of an intrarenal urine culture and stone culture is recommended to adapt antibiotic therapy in case of postoperative infectious complications. Measurements of temperature and pressure during RIRS are vital for ensuring patient safety and optimizing surgical outcomes although techniques of measurements and methods for data analysis are still to be refined. Ureteral stents were improved by the development of new biomaterials, new coatings, and new stent designs. Topics of current research are the development of drug eluting and bioresorbable stents. Complications of endoscopic treatment: PCNL is considered the most invasive surgical option. Fever and sepsis were observed in 11 and 0.5% and need for transfusion and embolization for bleeding in 7 and 0.4%. Major complications, as colonic, splenic, liver, gall bladder and bowel injuries are quite rare but are associated with significant morbidity. Ureteroscopy causes less complications, although some of them can be severe. They depend on high pressure in the urinary tract (sepsis or renal bleeding) or application of excessive force to the urinary tract (ureteral avulsion or stricture). Diagnostic work up: Genetic testing consents the diagnosis of monogenetic conditions causing stones. It should be carried out in children and in selected adults. In adults, monogenetic diseases can be diagnosed by systematic genetic testing in no more than 4%, when cystinuria, APRT deficiency, and xanthinuria are excluded. A reliable stone analysis by infrared spectroscopy or X-ray diffraction is mandatory and should be associated to examination of the stone under a stereomicroscope. The analysis of digital images of stones by deep convolutional neural networks in dry laboratory or during endoscopic examination could allow the classification of stones based on their color and texture. Scanning electron microscopy (SEM) in association with energy dispersive spectrometry (EDS) is another fundamental research tool for the study of kidney stones. The combination of metagenomic analysis using Next Generation Sequencing (NGS) techniques and the enhanced quantitative urine culture (EQUC) protocol can be used to evaluate the urobiome of renal stone formers. Twenty-four hour urine analysis has a place during patient evaluation together with repeated measurements of urinary pH with a digital pH meter. Urinary supersaturation is the most comprehensive physicochemical risk factor employed in urolithiasis research. Urinary macromolecules can act as both promoters or inhibitors of stone formation depending on the chemical composition of urine in which they are operating. At the moment, there are no clinical applications of macromolecules in stone management or prophylaxis. Patients should be evaluated for the association with systemic pathologies. **PROPHYLAXIS**: Personalized medicine and public health interventions are complementary to prevent stone recurrence. Personalized medicine addresses a small part of stone patients with a high risk of recurrence and systemic complications requiring specific dietary and pharmacological treatment to prevent stone recurrence and complications of associated systemic diseases. The more numerous subjects who form one or a few stones during their entire lifespan should be treated by modifications of diet and lifestyle. Primary prevention by public health interventions is advisable to reduce prevalence of stones in the general population. Renal stone formers at "high-risk" for recurrence need early diagnosis to start specific treatment. Stone analysis allows the identification of most "high-risk" patients forming non-calcium stones: infection stones (struvite), uric acid and urates, cystine and other rare stones (dihydroxyadenine, xanthine). Patients at "high-risk" forming calcium stones require a more difficult diagnosis by clinical and laboratory evaluation. Particularly, patients with cystinuria and primary hyperoxaluria should be actively searched. **FUTURE RESEARCH**: Application of Artificial Intelligence are promising for automated identification of ureteral stones on CT imaging, prediction of stone composition and 24-hour urinary risk factors by demographics and clinical parameters, assessment of stone composition by evaluation of endoscopic images and prediction of outcomes of stone treatments. The synergy between urologists, nephrologists, and scientists in basic kidney stone research will enhance the depth and breadth of investigations, leading to a more comprehensive understanding of kidney stone formation.

**URL:** <https://ovidsp.ovid.com/ovidweb.cgi?T=JS&CSC=Y&NEWS=N&PAGE=fulltext&D=med25&DO=10.4081%2faiua.2024.12703> <https://libkey.io/libraries/2592/openurl?genre=article&sid=OVID:medline&id=pmid:38934520&id=doi:10.4081%2Faiua.2024.12703&issn=1124-3562&isbn=&volume=96&issue=2&spage=12703&pages=12703&date=2024&title=Archivio+Italiano+di+Urologia%2C+Andrologia&atitle=Management+of+urinary+stones%3A+state+of+the+art+and+future+perspectives+by+experts+in+stone+disease.&aulast=Papatsoris&pid=%3Cauthor%3EPapatsoris+A%3BAlba+AB%3BGalan+Llopis+JA%3BMusafer+MA%3BAlameedee+M%3BAther+H%3BCaballero-Romeu+JP%3BCosta-Bauza+A%3BDellis+A%3BEl+Howairis+M%3BGambaro+G%3BGeavlete+B%3BHalinski+A%3BHess+B%3BJaffry+S%3BKok+D%3BKouicem+H%3BLlanes+L%3BLopez+Martinez+JM%3BPopov+E%3BRodgers+A%3BSoria+F%3BStamatelou+K%3BTrinchieri+A%3BTuerk+C%3C%2Fauthor%3E%3CAN%3E38934520%3C%2FAN%3E%3CDT%3EJournal+Article%3C%2FDT%3E>

5. **Comparison of metabolic parameters between pure-uric acid and mixed-uric acid kidney stone formers.**

**Item Type:** Journal Article

**Authors:** Ricapito, Anna;Gupta, Kavita;Zipkin, Jacob;Yim, Samuel;Connors, Christopher;Khargi, Raymond;Yaghoubian, Alan J.;Khusid, Johnathan A.;Atallah, William M. and Gupta, Mantu

**Publication Date:** Mar 13 ,2024

**Journal:** World Journal of Urology 42(1), pp. 138

**Abstract: PURPOSE**: We seek to compare clinical and 24-h urine parameters between pure-uric acid (UA) and UA-CaOx stone formers in our practice and explore how any differences in metabolic profiles could suggest different prevention strategies between the two groups. **METHODS**: We retrospectively reviewed patients with either pure- or mixed-UA nephrolithiasis from 2020 to 2023 at a tertiary care center. We included patients with a 24-h urine collection and a stone analysis detecting any amount of UA. Patients were organized into two cohorts: (1) those with 100% UA stones and (2) : We retrospectively reviewed patients with either pure- or mixed-UA nephrolithiasis from 2020 to 2023 at a tertiary care center. We included patients with a 24-h urine collection and a stone analysis detecting any amount of UA. Patients were organized into two cohorts: (1) those with 100% UA stones and (2) **RESULTS**: We identified 33 pure-UA patients and 33 mixed-UA patients. Patient demographics were similar between the groups (Table 1). Pure- and mixed-UA patients had a similar incidence of metabolic syndrome, diabetes, history of stones, and stone burden. Table 1 Demographic and baseline characteristics among pure- and mixed-uric acid stone formers Pure-uric acid stones (n = 33) Mixed-uric acid stones (n = 33) p-value Median age [IQR] 63.00 [58.00-72.50] 63.00 [53.50-68.00] 0.339 Median BMI [IQR] 28.79 [25.81-33.07] 27.96 [25.81-29.55] 0.534 Gender, n (%) 1.000 Male 21 (63.6) 21 (63.6) Female 12 (36.4) 12 (36.4) Metabolic syndrome, n (%) 17 (51.5) 16 (48.5) 0.806 Diabetes, n (%) 13 (39.4) 12 (36.4) 0.800 History of stones, n (%) 23 (69.7) 22 (66.7) 0.792 Median total stone burden, mm [IQR] 12.00 [6.00-26.50] 13.00 [7.05-20.00] 0.995 Median serum uric acid, mg/dL [IQR] 6.20 [4.80-7.15] 5.90 [4.98-6.89] 0.582 IQR Interquartile range BMI Body Mass Index n number We found the pure-UA cohort to have 24-h lower urine volume (1.53 vs. 1.96 L/day, p = 0.045) and citrate levels (286 vs. 457 mg/day, p = 0.036). UA-CaOx stone formers had higher urinary calcium levels (144 vs. 68 mg/day, p = 0.003), higher urinary oxalate levels (38 vs. 30 mg/day, p = 0.017), and higher median urinary calcium oxalate super-saturation (3.97 vs. 3.06, p = 0.047). **CONCLUSIONS**: Pure-UA kidney stone formers have different urinary metabolic parameters when compared with UA-CaOx stone formers, thus requiring different and tailored medical management. Copyright © 2024. The Author(s), under exclusive licence to Springer-Verlag GmbH Germany, part of Springer Nature.

**URL:** <https://ovidsp.ovid.com/ovidweb.cgi?T=JS&CSC=Y&NEWS=N&PAGE=fulltext&D=med25&DO=10.1007%2fs00345-024-04829-1> <https://libkey.io/libraries/2592/openurl?genre=article&sid=OVID:medline&id=pmid:38478092&id=doi:10.1007%2Fs00345-024-04829-1&issn=0724-4983&isbn=&volume=42&issue=1&spage=138&pages=138&date=2024&title=World+Journal+of+Urology&atitle=Comparison+of+metabolic+parameters+between+pure-uric+acid+and+mixed-uric+acid+kidney+stone+formers.&aulast=Ricapito&pid=%3Cauthor%3ERicapito+A%3BGupta+K%3BZipkin+J%3BYim+S%3BConnors+C%3BKhargi+R%3BYaghoubian+AJ%3BKhusid+JA%3BAtallah+WM%3BGupta+M%3C%2Fauthor%3E%3CAN%3E38478092%3C%2FAN%3E%3CDT%3EJournal+Article%3C%2FDT%3E>

6. **A Novel Machine-Learning Algorithm to Predict Stone Recurrence with 24-Hour Urine Data.**

**Item Type:** Journal Article

**Authors:** Shee, Kevin;Liu, Andrew W.;Chan, Carter;Yang, Heiko;Sui, Wilson;Desai, Manoj;Ho, Sunita;Chi, Thomas and Stoller, Marshall L.

**Publication Date:** Au ,2024

**Journal:** Journal of Endourology 38(8), pp. 809–816

**Abstract: Objectives:** The absence of predictive markers for kidney stone recurrence poses a challenge for the clinical management of stone disease. The unpredictability of stone events is also a significant limitation for clinical trials, where many patients must be enrolled to obtain sufficient stone events for analysis. In this study, we sought to use machine learning methods to identify a novel algorithm to predict stone recurrence. **Subjects/Patients and Methods:** Patients enrolled in the Registry for Stones of the Kidney and Ureter (ReSKU), a registry of nephrolithiasis patients collected between 2015-2020, with at least one prospectively collected 24-hour urine test (Litholink 24-hour urine test; Labcorp) were included in the training set. A validation set was obtained from chart review of stone patients not enrolled in ReSKU with 24-hour urine data. Stone events were defined as either an office visit where a patient reports symptomatic passage of stones or a surgical procedure for stone removal. Seven prediction classification methods were evaluated. Predictive analyses and receiver operator characteristics (ROC) curve generation were performed in R. **Results:** A training set of 423 kidney stone patients with stone event data and 24-hour urine samples were trained using the prediction classification methods. The highest performing prediction model was a Logistic Regression with ElasticNet machine learning model (area under curve [AUC] = 0.65). Restricting analysis to high confidence predictions significantly improved model accuracy (AUC = 0.82). The prediction model was validated on a validation set of 172 stone patients with stone event data and 24-hour urine samples. Prediction accuracy in the validation set demonstrated moderate discriminative ability (AUC = 0.64). Repeat modeling was performed with four of the highest scoring features, and ROC analyses demonstrated minimal loss in accuracy (AUC = 0.63). **Conclusion:** Machine-learning models based on 24-hour urine data can predict stone recurrences with a moderate degree of accuracy.

**URL:** <https://ovidsp.ovid.com/ovidweb.cgi?T=JS&CSC=Y&NEWS=N&PAGE=fulltext&D=medl&DO=10.1089%2fend.2023.0457> <https://libkey.io/libraries/2592/openurl?genre=article&sid=OVID:medline&id=pmid:39121452&id=doi:10.1089%2Fend.2023.0457&issn=0892-7790&isbn=&volume=38&issue=8&spage=809&pages=809-816&date=2024&title=Journal+of+Endourology&atitle=A+Novel+Machine-Learning+Algorithm+to+Predict+Stone+Recurrence+with+24-Hour+Urine+Data.&aulast=Shee&pid=%3Cauthor%3EShee+K%3BLiu+AW%3BChan+C%3BYang+H%3BSui+W%3BDesai+M%3BHo+S%3BChi+T%3BStoller+ML%3C%2Fauthor%3E%3CAN%3E39121452%3C%2FAN%3E%3CDT%3EJournal+Article%3C%2FDT%3E>

7. **Comparison of 24-hour urine composition prior to and after stone removal in nephrolithiasis: a prospective observational study.**

**Item Type:** Journal Article

**Authors:** Zhu, Wei;Zhang, Xin;Lu, Suilin;Ozsoy, Mehmet;Zhou, Zhen;Huang, Zhicong;Ai, Guoyao and Zeng, Guohua

**Publication Date:** Ju ,2024

**Journal:** Minerva Urology and Nephrology 76(3), pp. 351–356

**Abstract: BACKGROUND**: To evaluate 24-hour urine composition prior to and after complete stone removal in nephrolithiasis patients to determine potential relationship between kidney stones and patient metabolic status. **METHODS**: A prospective observational study was performed with patient enrollment from March 2019 to August 2020. 24-hour urine samples were collected prior to stone removal and 4 weeks after double-J stent removal, and examined the following urinary parameters: volume, creatinine, sodium, calcium, uric acid, citrate, oxalate, potassium, phosphorous, magnesium, and pH value. For each parameter, pairwise t test was performed to compare samples prior to and after stone removal. The number of cases that changed from normal to abnormal or vice versa was also evaluated for each parameter. The study was registered at <http://clinicaltrials.gov/> (NCT03846011). **RESULTS**: A total of 109 patients completed 24-hour urine collections prior to and after stone removal. The urinary calcium and phosphate levels increased significantly after stone removal, showing a mean difference of 0.55 mmol (P=0.015) and 2.35 mmol (P=0.001) respectively. None of the other urinary parameters demonstrated a statistically significant difference when means were compared. The percentage differences for all urinary parameters ranged from 5.4% to 14.1%. The percentages of patients that presented clinically significant changes in urinary parameter values from normal to abnormal or vice versa ranged from 4.6% to 20.1%. **CONCLUSIONS**: Ideally, evaluation of 24-hour urine compositions should be undertaken after total stone removal, especially for patients with calcium stones. For patients who cannot achieve total stone removal, 24-hour urine samples should be thoroughly interpreted as urinary calcium and phosphate levels might be depleted in the presence of urinary stones.

**URL:** <https://ovidsp.ovid.com/ovidweb.cgi?T=JS&CSC=Y&NEWS=N&PAGE=fulltext&D=med25&DO=10.23736%2fS2724-6051.22.05164-3> <https://libkey.io/libraries/2592/openurl?genre=article&sid=OVID:medline&id=pmid:36779823&id=doi:10.23736%2FS2724-6051.22.05164-3&issn=2724-6051&isbn=&volume=76&issue=3&spage=351&pages=351-356&date=2024&title=Minerva+Urology+and+Nephrology&atitle=Comparison+of+24-hour+urine+composition+prior+to+and+after+stone+removal+in+nephrolithiasis%3A+a+prospective+observational+study.&aulast=Zhu&pid=%3Cauthor%3EZhu+W%3BZhang+X%3BLu+S%3BOzsoy+M%3BZhou+Z%3BHuang+Z%3BAi+G%3BZeng+G%3C%2Fauthor%3E%3CAN%3E36779823%3C%2FAN%3E%3CDT%3EJournal+Article%3C%2FDT%3E>

8. **Associations Between Provider Specialty and Use of Follow-up Testing Among Patients on Preventive Pharmacological Therapy for Urinary Stone Disease.**

**Item Type:** Journal Article

**Authors:** Crivelli, Joseph J.;Yan, Phyllis L.;Shahinian, Vahakn;Hsi, Ryan S. and Hollingsworth, John M.

**Publication Date:** 2023

**Journal:** Urology Practice 10(4), pp. 400–406

**Abstract: INTRODUCTION**: The AUA Medical Management of Kidney Stones guideline outlines recommendations on follow-up testing for patients prescribed preventive pharmacological therapy. We evaluated adherence to these recommendations by provider specialty. **METHODS**: Using claims data from working-age adults with urinary stone disease (2008-2019), we identified patients prescribed a preventive pharmacological therapy agent (a thiazide diuretic, alkali citrate therapy, allopurinol, or a combination thereof) and the specialty of the prescribing physician (urology, nephrology, and general practice). Next, we identified patients who completed a 24-hour urine collection prior to their prescription fill. We then measured adherence to 3 recommendations outlined in the AUA guideline. Finally, we fit multivariable logistic regression models evaluating associations between prescribing provider specialty and adherence to recommended follow-up testing. **RESULTS**: Among 2,600 patients meeting study criteria, 1,523 (59%) adhered to >=1 follow-up testing recommendation, with a significant increase over the study period. Nephrologists had higher odds of adherence to >=1 follow-up test compared to urologists (odds ratio, 1.52; 95% confidence interval, 1.19-1.94; P : Among 2,600 patients meeting study criteria, 1,523 (59%) adhered to >=1 follow-up testing recommendation, with a significant increase over the study period. Nephrologists had higher odds of adherence to >=1 follow-up test compared to urologists (odds ratio, 1.52; 95% confidence interval, 1.19-1.94; P **CONCLUSIONS**: Following initiation of preventive pharmacological therapy, adherence to guideline-recommended follow-up testing was low overall. There exist meaningful specialty-specific differences in the use of this testing.

**URL:** <https://ovidsp.ovid.com/ovidweb.cgi?T=JS&CSC=Y&NEWS=N&PAGE=fulltext&D=med24&DO=10.1097%2fUPJ.0000000000000414> <https://libkey.io/libraries/2592/openurl?genre=article&sid=OVID:medline&id=pmid:37341368&id=doi:10.1097%2FUPJ.0000000000000414&issn=2352-0779&isbn=&volume=10&issue=4&spage=400&pages=400-406&date=2023&title=Urology+Practice&atitle=Associations+Between+Provider+Specialty+and+Use+of+Follow-up+Testing+Among+Patients+on+Preventive+Pharmacological+Therapy+for+Urinary+Stone+Disease.&aulast=Crivelli&pid=%3Cauthor%3ECrivelli+JJ%3BYan+PL%3BShahinian+V%3BHsi+RS%3BHollingsworth+JM%3C%2Fauthor%3E%3CAN%3E37341368%3C%2FAN%3E%3CDT%3EJournal+Article%3C%2FDT%3E>

9. **Associations between Net Gastrointestinal Alkali Absorption, 24-Hour Urine Lithogenic Factors, and Kidney Stones.**

**Item Type:** Journal Article

**Authors:** Ferraro, Pietro Manuel;Taylor, Eric N.;Asplin, John R. and Curhan, Gary C.

**Publication Date:** 2023

**Journal:** Clinical Journal of the American Society of Nephrology: CJASN 18(8), pp. 1068–1074

**Abstract: BACKGROUND**: It is not clear whether kidney stone formers have an abnormal handling of alkali and acid precursors in the gut, which might affect urine composition and ultimately stone formation. In this study, we aimed to investigate the determinants of net gastrointestinal alkali absorption and its associations with key urinary parameters in a large group of stone formers and non-stone formers. **METHODS**: Data were collected from three independent cohorts with at least one 24-hour urine collection. We explored potential determinants of net gastrointestinal alkali absorption and the association between net gastrointestinal alkali absorption, urinary parameters, and stone former status. Finally, we estimated the proportion of the association between urine parameters and stone former status explained by differences in net gastrointestinal alkali absorption. **RESULTS**: The analysis included 6067 participants (1102 men and 4965 women; 698 and 1804 of whom were stone formers, respectively). Average net gastrointestinal alkali absorption values were consistently lower in stone formers across the three cohorts (from -15.0 to -4.9 mEq/d). Age was directly associated with net gastrointestinal alkali absorption, whereas body mass index and net endogenous acid production were inversely associated. Net gastrointestinal alkali absorption was inversely associated with supersaturation for calcium oxalate, uric acid, and renal net acid excretion and directly associated with supersaturation for calcium phosphate, urine pH, and citrate. The odds of being a stone former was 15% (13%-17%) lower per 10 mEq/24 hours higher net gastrointestinal alkali absorption. Differences in net gastrointestinal alkali absorption explained a modest amount of the differences between stone formers and non-stone formers for supersaturation for calcium oxalate (6.3%) and a sizable amount for supersaturation for uric acid (15.2%), urine pH (38.3%), citrate (26.2%), and renal net acid excretion (63.4%). **CONCLUSIONS**: Kidney stone formers have lower net gastrointestinal alkali absorption, and this explains differences in urine composition and the likelihood of stone formation. Copyright © 2023 by the American Society of Nephrology.

**URL:** <https://ovidsp.ovid.com/ovidweb.cgi?T=JS&CSC=Y&NEWS=N&PAGE=fulltext&D=med24&DO=10.2215%2fCJN.0000000000000195> <https://libkey.io/libraries/2592/openurl?genre=article&sid=OVID:medline&id=pmid:37256914&id=doi:10.2215%2FCJN.0000000000000195&issn=1555-9041&isbn=&volume=18&issue=8&spage=1068&pages=1068-1074&date=2023&title=Clinical+Journal+of+The+American+Society+of+Nephrology%3A+CJASN&atitle=Associations+between+Net+Gastrointestinal+Alkali+Absorption%2C+24-Hour+Urine+Lithogenic+Factors%2C+and+Kidney+Stones.&aulast=Ferraro&pid=%3Cauthor%3EFerraro+PM%3BTaylor+EN%3BAsplin+JR%3BCurhan+GC%3C%2Fauthor%3E%3CAN%3E37256914%3C%2FAN%3E%3CDT%3EJournal+Article%3C%2FDT%3E>

10. **Specialist Care, Metabolic Testing, and Testing Completeness Among U.S. Veterans with Urinary Stone Disease.**

**Item Type:** Journal Article

**Authors:** Hsi, Ryan S.;Valicevic, Autumn N.;Patel, Sanjeevkumar R.;Oerline, Mary K.;Pao, Alan C.;Leppert, John T.;Hollingsworth, John M. and Shahinian, Vahakn

**Publication Date:** 2023

**Journal:** Urology Practice 10(1), pp. 49–56

**Abstract: Purpose**: Recent observational studies reporting a lack of benefit from 24-hour urine testing for urinary stone disease (USD) prevention assumed testing included all components recommended from clinical guidelines. We sought to assess the completeness of 24-hour urine testing in the VA population. **Materials and methods**: From the VHA Corporate Data Warehouse (2012-2019), we identified patients with USD (n=198,621) and determined those who saw a urologist and/or nephrologist, and received 24-hour urine testing within 12 months of their index USD encounter. Through Logical Observation Identifiers Names and Codes, we evaluated each collection's completeness, defined as including all of urine volume, calcium, oxalate, citrate, uric acid, and creatinine. We then fit a multilevel logistic regression model with random effects for VHA facility to evaluate factors associated with specialist follow-up, testing, and testing completeness. **Results**: Specialist follow-up occurred in 54.3% and was stable over time. Testing occurred in 8.4%, declining from 9.3% in 2012 to 7.2% in 2019. Of tests performed, 54.6% were complete (43.7% increasing to 62.7% from 2012-2019). In adjusted analysis, there was high between-facility variation in specialist follow-up (median OR 2.0; 95% CI 1.7-2.0), testing (median OR 2.2, 95% CI 1.9-2.4), and testing completeness (median OR, 6.0, 95% CI 4.5-7.3). Individual facilities contributed 52% (intraclass correlation coefficient, 0.52; 95% CI, 0.44-0.57) towards the observed variation in testing completeness. **Conclusions**: Approximately 1 in 12 U.S. Veterans with USD receive metabolic testing and half of these tests are complete. Addressing facility level variation in testing completeness may improve USD care.

**URL:** <https://ovidsp.ovid.com/ovidweb.cgi?T=JS&CSC=Y&NEWS=N&PAGE=fulltext&D=med23&DO=10.1097%2fupj.0000000000000356> <https://libkey.io/libraries/2592/openurl?genre=article&sid=OVID:medline&id=pmid:36545539&id=doi:10.1097%2Fupj.0000000000000356&issn=2352-0779&isbn=&volume=10&issue=1&spage=49&pages=49-56&date=2023&title=Urology+Practice&atitle=Specialist+Care%2C+Metabolic+Testing%2C+and+Testing+Completeness+Among+U.S.+Veterans+with+Urinary+Stone+Disease.&aulast=Hsi&pid=%3Cauthor%3EHsi+RS%3BValicevic+AN%3BPatel+SR%3BOerline+MK%3BPao+AC%3BLeppert+JT%3BHollingsworth+JM%3BShahinian+V%3C%2Fauthor%3E%3CAN%3E36545539%3C%2FAN%3E%3CDT%3EJournal+Article%3C%2FDT%3E>

11. **Potential for Urolithiasis-related Research Using the Novel Medicare-Litholink Database.**

**Item Type:** Journal Article

**Authors:** Krampe, Noah A.;Oerline, Mary K.;Asplin, John R.;Hsi, Ryan S.;Crivelli, Joseph J.;Shahinian, Vahakn B. and Hollingsworth, John M.

**Publication Date:** 2023

**Journal:** Urology Practice 10(2), pp. 147–152

**Abstract: INTRODUCTION**: To overcome the data availability hurdle of observational studies on urolithiasis, we linked claims data with 24-hour urine results from a large cohort of adults with urolithiasis. This database contains the sample size, clinical granularity, and long-term follow-up needed to study urolithiasis on a broad level. **METHODS**: We identified adults enrolled in Medicare with urolithiasis who had a 24-hour urine collection processed by Litholink (2011 to 2016). We created a linkage of their collections results and paid Medicare claims. We characterized them across a variety of sociodemographic and clinical factors. We measured frequencies of prescription fills for medications used to prevent stone recurrence, as well as frequencies of symptomatic stone events, among these patients. **RESULTS**: In total, there were 11,460 patients who performed 18,922 urine collections in the Medicare-Litholink cohort. The majority were male (57%), White (93.2%), and lived in a metropolitan county (51.5%). Results from their initial urine collections revealed abnormal pH to be the most common abnormality (77.2%), followed by low volume (63.8%), hypocitraturia (45.6%), hyperoxaluria (31.1%), hypercalciuria (28.4%), and hyperuricosuria (11.8%). Seventeen percent had prescription fills for alkali monotherapy, and 7.6% had prescription fills for thiazide diuretic monotherapy. Symptomatic stone events occurred in 23.1% at 2 years of follow-up. **CONCLUSIONS**: We successfully linked Medicare claims with results from 24-hour urine collections performed by adults that were processed by Litholink. The resulting database is a unique resource for future studies on the clinical effectiveness of stone prevention strategies and urolithiasis more broadly.

**URL:** <https://ovidsp.ovid.com/ovidweb.cgi?T=JS&CSC=Y&NEWS=N&PAGE=fulltext&D=med23&DO=10.1097%2fUPJ.0000000000000378> <https://libkey.io/libraries/2592/openurl?genre=article&sid=OVID:medline&id=pmid:37103409&id=doi:10.1097%2FUPJ.0000000000000378&issn=2352-0779&isbn=&volume=10&issue=2&spage=147&pages=147-152&date=2023&title=Urology+Practice&atitle=Potential+for+Urolithiasis-related+Research+Using+the+Novel+Medicare-Litholink+Database.&aulast=Krampe&pid=%3Cauthor%3EKrampe+NA%3BOerline+MK%3BAsplin+JR%3BHsi+RS%3BCrivelli+JJ%3BShahinian+VB%3BHollingsworth+JM%3C%2Fauthor%3E%3CAN%3E37103409%3C%2FAN%3E%3CDT%3EJournal+Article%3C%2FDT%3E>

12. **Serum and 24-hour urinary tests cost-effectiveness in stone formers.**

**Item Type:** Journal Article

**Authors:** Mohammadi, Abdolreza;Farabi, Hiro;Baghdadabad, Leila Zareian;Narouie, Behzad;Reis, Leonardo Oliveira and Aghamir, Seyed Mohammad Kazem

**Publication Date:** Aug 27 ,2023

**Journal:** BMC Urology 23(1), pp. 141

**Abstract: OBJECTIVE**: To assess the routine serum and 24-hour urine tests proficiency in diagnosing the baseline metabolic abnormality of kidney stone formers. **METHODS**: This study analyzes the routine serum and 24-hour urine tests proficiency in diagnosing the baseline metabolic abnormality of kidney stone formers. The sensitivity and specificity, false positive, and negative results of the tests are extracted from diagnostic kits used in the laboratories of the target community. To accurately infer the results, a simulation based on 1000 people was used through 22 standard laboratory tests (Additional File 2), including calcium, oxalate, phosphate, uric acid, sulphate, potassium, sodium, citrate, and magnesium in 24-hour urine; and calcium, creatinine, Vit D, uric acid, and intact parathyroid hormone (PTH) in serum. The incremental cost-effectiveness ratio (ICER) was calculated and compared for each diagnostic test versus other diagnostic tests according to the incremental cost required for correct diagnoses of stone causes. **RESULTS**: Urinary uric acid, citrate, and serum potassium constitute the cost-effectiveness boundary curve in this study. This means that other diagnostic tests are not cost-effective compared to these three tests in terms of indexing at least one item of cost and effectiveness. The ICER index for each correct diagnosis with the urinary uric acid test was $ 1.25 per diagnosis, the most cost-effective test compared to serum potassium and urinary citrate. **CONCLUSION**: The simplified blood and 24-hour urine metabolic evaluation, including urinary uric acid, citrate, and serum potassium, constitute the cost-effectiveness boundary curve. The most cost-effective test was urinary uric acid measurement. Copyright © 2023. BioMed Central Ltd., part of Springer Nature.

**URL:** <https://ovidsp.ovid.com/ovidweb.cgi?T=JS&CSC=Y&NEWS=N&PAGE=fulltext&D=med24&DO=10.1186%2fs12894-023-01310-w> <https://libkey.io/libraries/2592/openurl?genre=article&sid=OVID:medline&id=pmid:37635222&id=doi:10.1186%2Fs12894-023-01310-w&issn=1471-2490&isbn=&volume=23&issue=1&spage=141&pages=141&date=2023&title=BMC+Urology&atitle=Serum+and+24-hour+urinary+tests+cost-effectiveness+in+stone+formers.&aulast=Mohammadi&pid=%3Cauthor%3EMohammadi+A%3BFarabi+H%3BBaghdadabad+LZ%3BNarouie+B%3BReis+LO%3BAghamir+SMK%3C%2Fauthor%3E%3CAN%3E37635222%3C%2FAN%3E%3CDT%3EJournal+Article%3C%2FDT%3E>

13. **SPP1 and UMOD gene variants are synergistically associated with risk of renal stone disease.**

**Item Type:** Journal Article

**Authors:** Patel, Yash P.;Patel, Samir G.;Patel, Sandip B.;Parikh, Aditya;Soni, Shailesh;Srivastava, Ratika;Raval, Chintal;Raval, Manan A.;Nand Pandey, Sachchida;Ganpule, Arvind P. and Desai, Mahesh R.

**Publication Date:** May 05 ,2023

**Journal:** Gene 863, pp. 147264

**Abstract: OBJECTIVE**: Calcium and oxalate are the most abundant metabolites present in the stone matrix. The SPP1 and UMOD gene has specific expression in kidneys and are involved in various stages of stone formation. Therefore, genetic variants in the SPP1 and UMOD genes may enhance the development of renal stone disease. This study has been designed to understand the association of genetic variants of SPP1 and UMOD genes with renal stone disease. **MATERIALS AND METHOD**: A prospective study has been carried out, including 150 renal stone disease patients and 150 healthy individuals. Biochemical parameters were performed, including serum calcium levels, creatinine, parathyroid hormone, and 24-Hour urine metabolites. The genotyping of SPP1 (rs1126616) and UMOD (rs4293393) gene variants were performed using a customized TaqMan probe. T-test was used for continuous biochemical data analysis. The Chi-square test has been applied to assess the risk of a particular genotype associated with renal stone disease. In addition, correlation analysis for biochemical parameters and genetic variants with the renal stone disease has been performed using Shapley additive explanations (SHAP) values calculated with the help of the pycaret library. **RESULT**: Renal stone patients had significantly higher levels of parathyroid hormone (93.37 +/- 52.78 pg/ml vs 64.67 +/- 31.50 pg/ml, P=: Renal stone patients had significantly higher levels of parathyroid hormone (93.37 +/- 52.78 pg/ml vs 64.67 +/- 31.50 pg/ml, P=: Renal stone patients had significantly higher levels of parathyroid hormone (93.37 +/- 52.78 pg/ml vs 64.67 +/- 31.50 pg/ml, P=: Renal stone patients had significantly higher levels of parathyroid hormone (93.37 +/- 52.78 pg/ml vs 64.67 +/- 31.50 pg/ml, P=: Renal stone patients had significantly higher levels of parathyroid hormone (93.37 +/- 52.78 pg/ml vs 64.67 +/- 31.50 pg/ml, P=: Renal stone patients had significantly higher levels of parathyroid hormone (93.37 +/- 52.78 pg/ml vs 64.67 +/- 31.50 pg/ml, P=: Renal stone patients had significantly higher levels of parathyroid hormone (93.37 +/- 52.78 pg/ml vs 64.67 +/- 31.50 pg/ml, P=: Renal stone patients had significantly higher levels of parathyroid hormone (93.37 +/- 52.78 pg/ml vs 64.67 +/- 31.50 pg/ml, P=**CONCLUSION**: Genetic variants of the SPP1 and UMOD genes were associated with renal stone disease. In the presence of risk genotype and hyperparathyroidism, hypercalciuria, and hyperoxaluria, the susceptibility to develop the renal stone disease risk gets modulated. Copyright © 2023 Elsevier B.V. All rights reserved.

**URL:** <https://ovidsp.ovid.com/ovidweb.cgi?T=JS&CSC=Y&NEWS=N&PAGE=fulltext&D=med23&DO=10.1016%2fj.gene.2023.147264> <https://libkey.io/libraries/2592/openurl?genre=article&sid=OVID:medline&id=pmid:36804854&id=doi:10.1016%2Fj.gene.2023.147264&issn=0378-1119&isbn=&volume=863&issue=&spage=147264&pages=147264&date=2023&title=Gene&atitle=SPP1+and+UMOD+gene+variants+are+synergistically+associated+with+risk+of+renal+stone+disease.&aulast=Patel&pid=%3Cauthor%3EPatel+YP%3BPatel+SG%3BPatel+SB%3BParikh+A%3BSoni+S%3BSrivastava+R%3BRaval+C%3BRaval+MA%3BNand+Pandey+S%3BGanpule+AP%3BDesai+MR%3C%2Fauthor%3E%3CAN%3E36804854%3C%2FAN%3E%3CDT%3EJournal+Article%3C%2FDT%3E>

14. **Machine Learning Prediction of Kidney Stone Composition Using Electronic Health Record-Derived Features.**

**Item Type:** Journal Article

**Authors:** Abraham, Abin;Kavoussi, Nicholas L.;Sui, Wilson;Bejan, Cosmin;Capra, John A. and Hsi, Ryan

**Publication Date:** 2022

**Journal:** Journal of Endourology 36(2), pp. 243–250

**Abstract: Objectives:** To assess the accuracy of machine learning models in predicting kidney stone composition using variables extracted from the electronic health record (EHR). **Materials and Methods:** We identified kidney stone patients (n = 1296) with both stone composition and 24-hour (24H) urine testing. We trained machine learning models (XGBoost [XG] and logistic regression [LR]) to predict stone composition using 24H urine data and EHR-derived demographic and comorbidity data. Models predicted either binary (calcium vs noncalcium stone) or multiclass (calcium oxalate, uric acid, hydroxyapatite, or other) stone types. We evaluated performance using area under the receiver operating curve (ROC-AUC) and accuracy and identified predictors for each task. **Results:** For discriminating binary stone composition, XG outperformed LR with higher accuracy (91% vs 71%) with ROC-AUC of 0.80 for both models. Top predictors used by these models were supersaturations of uric acid and calcium phosphate, and urinary ammonium. For multiclass classification, LR outperformed XG with higher accuracy (0.64 vs 0.56) and ROC-AUC (0.79 vs 0.59), and urine pH had the highest predictive utility. Overall, 24H urine analyte data contributed more to the models' predictions of stone composition than EHR-derived variables. **Conclusion:** Machine learning models can predict calcium stone composition. LR outperforms XG in multiclass stone classification. Demographic and comorbidity data are predictive of stone composition; however, including 24H urine data improves performance. Further optimization of performance could lead to earlier directed medical therapy for kidney stone patients.

**URL:** <https://ovidsp.ovid.com/ovidweb.cgi?T=JS&CSC=Y&NEWS=N&PAGE=fulltext&D=med21&DO=10.1089%2fend.2021.0211> <https://libkey.io/libraries/2592/openurl?genre=article&sid=OVID:medline&id=pmid:34314237&id=doi:10.1089%2Fend.2021.0211&issn=0892-7790&isbn=&volume=36&issue=2&spage=243&pages=243-250&date=2022&title=Journal+of+Endourology&atitle=Machine+Learning+Prediction+of+Kidney+Stone+Composition+Using+Electronic+Health+Record-Derived+Features.&aulast=Abraham&pid=%3Cauthor%3EAbraham+A%3BKavoussi+NL%3BSui+W%3BBejan+C%3BCapra+JA%3BHsi+R%3C%2Fauthor%3E%3CAN%3E34314237%3C%2FAN%3E%3CDT%3EJournal+Article%3C%2FDT%3E>

15. **Agreement of Food Records and 24-Hour Urine Studies in Clinical Practice.**

**Item Type:** Journal Article

**Authors:** Betz, Melanie V.;Coe, Fredric L. and Chapman, Arlene B.

**Publication Date:** 2022

**Journal:** Journal of Renal Nutrition 32(1), pp. 51–57

**Abstract: OBJECTIVE**: The objective of this study was to determine the level of agreement between 3-day food records obtained as part of clinical care with 24-hour urine collections specifically assessing sodium, potassium, phosphorus, calcium, protein, and fluid intake. **DESIGN AND METHODS**: Data were collected from patients at a nephrology clinic in a metropolitan, academic medical center. Patients who completed both a 3-day food record and a 24-hour urine collection were analyzed. Food record and urine collection measurements were compared using a simple ratio, Pearson's correlation, and general linear models. **RESULTS**: Patients (n = 85) were 47.9 +/- 15.2 years of age, 54% were female, with a mean serum creatinine of 1.3 +/- 0.7 mg/dL and estimated glomerular filtration rate of 64.2 +/- 25.6 mL/min. Patients had autosomal-dominant polycystic kidney disease (48.2%), nephrolithiasis (31.1%), chronic kidney disease (4.7%), or other genetic or cystic conditions impacting the kidney (12.9%). Nutrient intake was measured utilizing a 3-day food record. Food records and urine collections were compared using the values, correlations, and general linear models. Fluid intake demonstrated the highest agreement (ratio 1.01) and calcium demonstrated the least agreement (ratio 6.30). Significant correlations were demonstrated for phosphorus (r = 0.321, P = .003), magnesium (r = 0.256, P = .018), protein (r = 0.555, P : Patients (n = 85) were 47.9 +/- 15.2 years of age, 54% were female, with a mean serum creatinine of 1.3 +/- 0.7 mg/dL and estimated glomerular filtration rate of 64.2 +/- 25.6 mL/min. Patients had autosomal-dominant polycystic kidney disease (48.2%), nephrolithiasis (31.1%), chronic kidney disease (4.7%), or other genetic or cystic conditions impacting the kidney (12.9%). Nutrient intake was measured utilizing a 3-day food record. Food records and urine collections were compared using the values, correlations, and general linear models. Fluid intake demonstrated the highest agreement (ratio 1.01) and calcium demonstrated the least agreement (ratio 6.30). Significant correlations were demonstrated for phosphorus (r = 0.321, P = .003), magnesium (r = 0.256, P = .018), protein (r = 0.555, P **CONCLUSION**: 3-day food records are useful tools to determine patient dietary patterns, but should be used with caution when assessing specific nutrient intake in clinical settings. Copyright © 2021 National Kidney Foundation, Inc. Published by Elsevier Inc. All rights reserved.

**URL:** <https://ovidsp.ovid.com/ovidweb.cgi?T=JS&CSC=Y&NEWS=N&PAGE=fulltext&D=med21&DO=10.1053%2fj.jrn.2021.07.005> <https://libkey.io/libraries/2592/openurl?genre=article&sid=OVID:medline&id=pmid:34429244&id=doi:10.1053%2Fj.jrn.2021.07.005&issn=1051-2276&isbn=&volume=32&issue=1&spage=51&pages=51-57&date=2022&title=Journal+of+Renal+Nutrition&atitle=Agreement+of+Food+Records+and+24-Hour+Urine+Studies+in+Clinical+Practice.&aulast=Betz&pid=%3Cauthor%3EBetz+MV%3BCoe+FL%3BChapman+AB%3C%2Fauthor%3E%3CAN%3E34429244%3C%2FAN%3E%3CDT%3EJournal+Article%3C%2FDT%3E>

16. **Acidification of 24-hour urine in urolithiasis risk testing: An obsolete relic?.**

**Item Type:** Journal Article

**Authors:** Cadamuro, Janne;Decho, Cosima;Frans, Glynis;Auer, Simon;von Meyer, Alexander;Kniewallner, Kathrin M.;Drerup, Martin;Heinrich, Elmar;Keppel, Martin H.;Mrazek, Cornelia;Felder, Thomas K.;Oberkofler, Hannes;Haschke-Becher, Elisabeth;Kipman, Ulrike;Salek, Tomas and Vermeersch, Pieter

**Publication Date:** Jul 01 ,2022

**Journal:** Clinica Chimica Acta 532, pp. 1–9

**Abstract: BACKGROUND**: Recommendations on the optimal preservation of 24 h urine for the metabolic work-up in urolithiasis patients are very heterogeneous. In case two such tests with different storage condition recommendations are being analysed, multiple collections would be needed, challenging especially elderly and very young patients. We therefore aimed to evaluate the stability of urine constituents under different storage conditions. **MATERIAL AND METHODS**: We collected urine samples from ten healthy volunteers and prepared aliquots to be stored either at room temperature or 4 degreeC. Some aliquots were preserved using hydrochloric acid prior to storage, some thereafter, some using the BD Urine preservation tube and some were not preserved at all. Storage duration was 0, 24, 48 or 72 h. In all samples calcium, magnesium, phosphorus, creatinine, oxalate, citrate and uric acid were measured and compared to the according reference sample. **RESULTS**: We could not find any significant deviation for any of the analytes and preanalytical treatment conditions compared to the associated reference sample. **CONCLUSION**: Preservation of 24 h urine for the metabolic evaluation in stone formers might not be necessary for sample storage up to 72 h. Copyright © 2022 Elsevier B.V. All rights reserved.

**URL:** <https://ovidsp.ovid.com/ovidweb.cgi?T=JS&CSC=Y&NEWS=N&PAGE=fulltext&D=med22&DO=10.1016%2fj.cca.2022.05.010> <https://libkey.io/libraries/2592/openurl?genre=article&sid=OVID:medline&id=pmid:35597305&id=doi:10.1016%2Fj.cca.2022.05.010&issn=0009-8981&isbn=&volume=532&issue=&spage=1&pages=1-9&date=2022&title=Clinica+Chimica+Acta&atitle=Acidification+of+24-hour+urine+in+urolithiasis+risk+testing%3A+An+obsolete+relic%3F.&aulast=Cadamuro&pid=%3Cauthor%3ECadamuro+J%3BDecho+C%3BFrans+G%3BAuer+S%3Bvon+Meyer+A%3BKniewallner+KM%3BDrerup+M%3BHeinrich+E%3BKeppel+MH%3BMrazek+C%3BFelder+TK%3BOberkofler+H%3BHaschke-Becher+E%3BKipman+U%3BSalek+T%3BVermeersch+P%3C%2Fauthor%3E%3CAN%3E35597305%3C%2FAN%3E%3CDT%3EJournal+Article%3C%2FDT%3E>

17. **Associations of Obesity and Neighborhood Factors With Urinary Stone Parameters.**

**Item Type:** Journal Article

**Authors:** Crivelli, Joseph J.;Redden, David T.;Johnson, Robert D.;Juarez, Lucia D.;Maalouf, Naim M.;Hughes, Amy E.;Wood, Kyle D.;Assimos, G. and Oates, Gabriela R.

**Publication Date:** 2022

**Journal:** American Journal of Preventive Medicine 63(1 Suppl 1), pp. S93–S102

**Abstract: INTRODUCTION**: Obesity is associated with kidney stone disease, but it is unknown whether this association differs by SES. This study assessed the extent to which obesity and neighborhood characteristics jointly contribute to urinary risk factors for kidney stone disease. **METHODS**: This was a retrospective analysis of adult patients with kidney stone disease evaluated with 24-hour urine collection (2001-2020). Neighborhood-level socioeconomic data were obtained for a principal component analysis, which identified 3 linearly independent factors. Associations between these factors and 24-hour urine measurements were assessed using linear regression as well as groupings of 24-hour urine results using multivariable logistic regression. Finally, multiplicative interactions were assessed testing effect modification by obesity, and analyses stratified by obesity were performed. Analyses were performed in 2021. **RESULTS**: In total, 1,264 patients met the study criteria. Factors retained on principal component analysis represented SES, family structure, and housing characteristics. On linear regression, there was a significant inverse correlation between SES and 24-hour urine sodium (p=0.0002). On multivariable logistic regression, obesity was associated with increased odds of multiple stone risk factors (OR=1.61; 95% CI=1.15, 2.26) and multiple dietary factors (OR=1.33; 95% CI=1.06, 1.67). No significant and consistent multiplicative interactions were observed between obesity and quartiles of neighborhood SES, family structure, or housing characteristics. **CONCLUSIONS**: Obesity was associated with the presence of multiple stone risk factors and multiple dietary factors; however, the strength and magnitude of these associations did not vary significantly by neighborhood SES, family structure, and housing characteristics. Copyright © 2022 American Journal of Preventive Medicine. Published by Elsevier Inc. All rights reserved.

**URL:** <https://ovidsp.ovid.com/ovidweb.cgi?T=JS&CSC=Y&NEWS=N&PAGE=fulltext&D=med22&DO=10.1016%2fj.amepre.2022.01.033> <https://libkey.io/libraries/2592/openurl?genre=article&sid=OVID:medline&id=pmid:35725147&id=doi:10.1016%2Fj.amepre.2022.01.033&issn=0749-3797&isbn=&volume=63&issue=1&spage=S93&pages=S93-S102&date=2022&title=American+Journal+of+Preventive+Medicine&atitle=Associations+of+Obesity+and+Neighborhood+Factors+With+Urinary+Stone+Parameters.&aulast=Crivelli&pid=%3Cauthor%3ECrivelli+JJ%3BRedden+DT%3BJohnson+RD%3BJuarez+LD%3BMaalouf+NM%3BHughes+AE%3BWood+KD%3BAssimos+G%3BOates+GR%3C%2Fauthor%3E%3CAN%3E35725147%3C%2FAN%3E%3CDT%3EJournal+Article%3C%2FDT%3E>

18. **Enteral nutrition and the risk of nephrolithiasis in complex pediatric patients.**

**Item Type:** Journal Article

**Authors:** DeFoor, William;Nehus, Edward;Schulte, Marion;Huesman, Sydney;Libs, Ashleigh;Niehaus, Renee and Devarajan, Prasad

**Publication Date:** 2022

**Journal:** Journal of Pediatric Urology 18(6), pp. 743.e1–743.e6

**Abstract: PURPOSE**: Medically complex, non-ambulatory children can often suffer from nephrolithiasis. The purpose of this study is to determine risk factors which are predictive for recurrent stone formation in this patient population. **MATERIAL AND METHODS**: A retrospective cohort study was performed on non-ambulatory patients with cerebral palsy and/or severe developmental delay presenting to a high-volume Pediatric Stone Center from 2015 to 2019. Two 24-hour urine collections were performed as a baseline prior to pharmacotherapeutic and/or dietary intervention. Healthy stone-forming children served as a control group. **RESULTS**: 28 non-ambulatory subjects and 38 healthy controls were evaluated. The study group had a higher rate of bilateral nephrolithiasis but a similar history of previous surgical procedures. 89% of the non-ambulatory children were fed via a gastrostomy. The median calcium excretion was the same in both groups (3.0 mg/kg/day). The median 24-hour excretion of oxalate was significantly increased in the study group (54 vs 31 mg/1.73 m^2^/day, p = 0.0001). Urinary citrate and phosphorus excretions, and the supersaturations of calcium oxalate and calcium phosphate were similar between study subjects and controls. Calcium oxalate stones were noted in 57% of those with known stone composition in the study group. Enteral feeding formulas were primarily based on soy protein, a known high oxalate food. **CONCLUSIONS**: Urinary oxalate excretion is significantly increased in a cohort of medically complex, non-ambulatory stone-forming children. Urinary calcium excretion was not elevated between study subjects and healthy controls. Further analysis is needed to assess if dietary intervention to limit oxalate excretion results in decreased stone formation. Copyright © 2022 Journal of Pediatric Urology Company. Published by Elsevier Ltd. All rights reserved.

**URL:** <https://ovidsp.ovid.com/ovidweb.cgi?T=JS&CSC=Y&NEWS=N&PAGE=fulltext&D=med22&DO=10.1016%2fj.jpurol.2022.06.003> <https://libkey.io/libraries/2592/openurl?genre=article&sid=OVID:medline&id=pmid:35760671&id=doi:10.1016%2Fj.jpurol.2022.06.003&issn=1477-5131&isbn=&volume=18&issue=6&spage=743.e1&pages=743.e1-743.e6&date=2022&title=Journal+of+pediatric+urology&atitle=Enteral+nutrition+and+the+risk+of+nephrolithiasis+in+complex+pediatric+patients.&aulast=DeFoor&pid=%3Cauthor%3EDeFoor+W%3BNehus+E%3BSchulte+M%3BHuesman+S%3BLibs+A%3BNiehaus+R%3BDevarajan+P%3C%2Fauthor%3E%3CAN%3E35760671%3C%2FAN%3E%3CDT%3EJournal+Article%3C%2FDT%3E>

19. **Estimating 24-hour urinary excretion using spot urine measurements in kidney stone formers.**

**Item Type:** Journal Article

**Authors:** Ferraro, Pietro Manuel;Lopez, Fernando;Petrarulo, Michele;Barbarini, Silvia;Curhan, Gary C.;Marangella, Martino and Taylor, Eric N.

**Publication Date:** 2022

**Journal:** Nephrology Dialysis Transplantation 37(11), pp. 2171–2179

**Abstract: BACKGROUND**: One limitation of the use of 24-hour collection is impracticality. We analysed the performance of spot urine measurements to estimate 24-hour excretion in patients with kidney stones. **METHODS**: A total of 74 adult patients from two centres performed a 24-hour urine collection. A sample of the last micturition was sent for spot urine analysis. Twenty patients were asked to collect two additional spot urine samples, one before dinner and the other after dinner. Urinary concentrations of creatinine, calcium, oxalate, uric acid, citrate and magnesium were measured in the 24-hour and each of the spot urine samples. Four approaches were used to estimate 24-hour urinary excretion, multiplying the ratio of the spot urinary analyte to creatinine concentration by (i) measured 24-hour urinary creatinine excretion (Prediction 1), (ii) estimated 24-hour urinary creatinine excretion (Prediction 2), (iii) assumed 1-g 24-hour urinary creatinine excretion (Prediction 3) or (iv) assumed 1.5-g 24-hour urinary creatinine excretion (Prediction 4). For each parameter we computed Lin's concordance correlation coefficients (CCCs), Bland-Altman plots and 95% limits of agreement. **RESULTS**: The performance of estimates obtained with Prediction 1 and Prediction 2 was similar, except for citrate and uric acid, for which Prediction 2 performed worse. Both approaches performed moderately well: citrate CCC {0.82 [95% confidence interval (CI) 0.75-0.90]}, oxalate [0.66 (95% CI 0.55-0.78)], magnesium [0.66 (95% CI 0.54-0.77)], calcium [0.63 (95% CI 0.50-0.75)] and uric acid [0.52 (95% CI 0.36-0.68)]. The performance of Predictions 3 and 4 was worse. **CONCLUSIONS**: Although spot urine samples may hold promise for clinical and population-based research, at present they have limited utility in clinical practice. Measuring or estimating 24-hour creatinine, rather than assuming a given creatinine excretion, will be necessary in future studies of spot urine samples. Copyright © The Author(s) 2022. Published by Oxford University Press on behalf of the ERA.

**URL:** <https://ovidsp.ovid.com/ovidweb.cgi?T=JS&CSC=Y&NEWS=N&PAGE=fulltext&D=med21&DO=10.1093%2fndt%2fgfab306> <https://libkey.io/libraries/2592/openurl?genre=article&sid=OVID:medline&id=pmid:35146503&id=doi:10.1093%2Fndt%2Fgfab306&issn=0931-0509&isbn=&volume=37&issue=11&spage=2171&pages=2171-2179&date=2022&title=Nephrology+Dialysis+Transplantation&atitle=Estimating+24-hour+urinary+excretion+using+spot+urine+measurements+in+kidney+stone+formers.&aulast=Ferraro&pid=%3Cauthor%3EFerraro+PM%3BLopez+F%3BPetrarulo+M%3BBarbarini+S%3BCurhan+GC%3BMarangella+M%3BTaylor+EN%3C%2Fauthor%3E%3CAN%3E35146503%3C%2FAN%3E%3CDT%3EJournal+Article%3C%2FDT%3E>

20. **Association of Urine Findings with Metabolic Syndrome Traits in a Population of Patients with Nephrolithiasis.**

**Item Type:** Journal Article

**Authors:** Hood, Virginia L.;Sternberg, Kevan M.;de Waal, Desiree;Asplin, John R.;Mulligan, Carley and Callas, Peter W.

**Publication Date:** 2022

**Journal:** Kidney360 3(2), pp. 317; 02 24–324

**Abstract: Background**: The odds of nephrolithiasis increase with more metabolic syndrome (MetS) traits. We evaluated associations of metabolic and dietary factors from urine studies and stone composition with MetS traits in a large cohort of stone-forming patients. **Methods**: Patients >18 years old who were evaluated for stones with 24-hour urine collections between July 2009 and December 2018 had their records reviewed retrospectively. Patient factors, laboratory values, and diagnoses were identified within 6 months of urine collection and stone composition within 1 year. Four groups with none, one, two, and three or four MetS traits (hypertension, obesity, dyslipidemia, and diabetes) were evaluated. Trends across groups were tested using linear contrasts in analysis of variance and analysis of covariance. **Results**: A total of 1473 patients met the inclusion criteria (835 with stone composition). MetS groups were 684 with no traits, 425 with one trait, 211 with two traits, and 153 with three or four traits. There were no differences among groups for urine volume, calcium, or ammonium excretion. There was a significant trend (P: A total of 1473 patients met the inclusion criteria (835 with stone composition). MetS groups were 684 with no traits, 425 with one trait, 211 with two traits, and 153 with three or four traits. There were no differences among groups for urine volume, calcium, or ammonium excretion. There was a significant trend (P: A total of 1473 patients met the inclusion criteria (835 with stone composition). MetS groups were 684 with no traits, 425 with one trait, 211 with two traits, and 153 with three or four traits. There were no differences among groups for urine volume, calcium, or ammonium excretion. There was a significant trend (P**Conclusions**: Stone-forming patients with MetS have a defined pattern of metabolic and dietary risk factors that contribute to an increased risk of stone formation, including higher acid excretion, largely the result of greater protein intake, and lower urine pH. Copyright © 2022 by the American Society of Nephrology.

**URL:** <https://ovidsp.ovid.com/ovidweb.cgi?T=JS&CSC=Y&NEWS=N&PAGE=fulltext&D=med21&DO=10.34067%2fKID.0002292021> <https://libkey.io/libraries/2592/openurl?genre=article&sid=OVID:medline&id=pmid:35373120&id=doi:10.34067%2FKID.0002292021&issn=2641-7650&isbn=&volume=3&issue=2&spage=317&pages=317-324&date=2022&title=Kidney360&atitle=Association+of+Urine+Findings+with+Metabolic+Syndrome+Traits+in+a+Population+of+Patients+with+Nephrolithiasis.&aulast=Hood&pid=%3Cauthor%3EHood+VL%3BSternberg+KM%3Bde+Waal+D%3BAsplin+JR%3BMulligan+C%3BCallas+PW%3C%2Fauthor%3E%3CAN%3E35373120%3C%2FAN%3E%3CDT%3EJournal+Article%3C%2FDT%3E>

21. **Comparison of Selective vs Empiric Pharmacologic Preventive Therapy of Kidney Stone Recurrence With High-Risk Features.**

**Item Type:** Journal Article

**Authors:** Hsi, Ryan S.;Yan, Phyllis L.;Crivelli, Joseph J.;Goldfarb, David S.;Shahinian, Vahakn and Hollingsworth, John M.

**Publication Date:** 2022

**Journal:** Urology 164, pp. 74–79

**Abstract: OBJECTIVE**: To compare the frequency of stone-related events among subgroups of high-risk patients with and without 24-hour urine testing before preventive pharmacological therapy (PPT) prescription. While recent studies show, on average, no benefit to a selective approach to PPT for urinary stone disease (USD), there could be heterogeneity in treatment effect across patient subgroups. **MATERIALS AND METHODS**: Using medical claims data from working-age adults and their dependents with USD (2008-2019), we identified those with a prescription fill for a PPT agent (thiazide diuretic, alkali therapy, or allopurinol). We then stratified patients into subgroups based on the presence of a concomitant condition or other factors that raised their stone recurrence risk. Finally, we fit multivariable regression models to measure the association between stone-related events (emergency department visit, hospitalization, and surgery) and 24-hour urine testing before PPT prescription by high-risk subgroup. **RESULTS**: Overall, 8369 adults with USD had a concomitant condition that raised their recurrence risk. Thirty-three percent (n = 2722) of these patients were prescribed PPT after 24-hour urine testing (median follow-up, 590 days), and 67% (n = 5647) received PPT empirically (median follow-up, 533 days). Compared to patients treated empirically, those with a history of recurrent USD had a significantly lower hazard of a subsequent stone-related event if they received selective PPT (hazard ratio, 0.83; 95% confidence interval, 0.71-0.96). No significant associations were noted for selective PPT in the other high-risk subgroups. **CONCLUSION**: Patients with a history of recurrent USD benefit from PPT when guided by findings from 24-hour urine testing. Copyright © 2022 Elsevier Inc. All rights reserved.

**URL:** <https://ovidsp.ovid.com/ovidweb.cgi?T=JS&CSC=Y&NEWS=N&PAGE=fulltext&D=med21&DO=10.1016%2fj.urology.2021.12.037> <https://libkey.io/libraries/2592/openurl?genre=article&sid=OVID:medline&id=pmid:35182586&id=doi:10.1016%2Fj.urology.2021.12.037&issn=0090-4295&isbn=&volume=164&issue=&spage=74&pages=74-79&date=2022&title=Urology&atitle=Comparison+of+Selective+vs+Empiric+Pharmacologic+Preventive+Therapy+of+Kidney+Stone+Recurrence+With+High-Risk+Features.&aulast=Hsi&pid=%3Cauthor%3EHsi+RS%3BYan+PL%3BCrivelli+JJ%3BGoldfarb+DS%3BShahinian+V%3BHollingsworth+JM%3C%2Fauthor%3E%3CAN%3E35182586%3C%2FAN%3E%3CDT%3EJournal+Article%3C%2FDT%3E>

22. **Urinary Metabolic Disturbances During Topiramate Use and their Reversibility Following Drug Cessation.**

**Item Type:** Journal Article

**Authors:** Pelzman, Daniel L.;Kazi, Eman;Jackman, Stephen V. and Semins, Michelle J.

**Publication Date:** 2022

**Journal:** Urology 165, pp. 139–143

**Abstract: OBJECTIVE**: To understand the metabolic disturbances of stone formers currently taking topiramate and to examine the reversibility of these disturbances with cessation of the medication. **MATERIALS AND METHODS**: All progress notes written by 5 endourologists from a single academic center were retrospectively reviewed from January 2010 to July 2020 containing the words "topiramate" or "topamax." Inclusion criteria were age >18 and presence of either a 24-hour urine sample or stone analysis while on topiramate. In addition, a subgroup of 18 patients with 24-hour urine samples before and after stopping topiramate were identified. **RESULTS**: A total of 93 patients were identified and included for final analysis. Twenty-four hour urine samples were available in 67 patients and showed mean citrate excretion of 331 +/- 322 mg/d, mean pH of 6.6 +/- 0.5, and mean calcium phosphate supersaturation of 1.9 +/- 1.1. In the subgroup analysis urinary citrate excretion increased from 225 mg/d to 614 mg/d (P =50%) calcium phosphate by composition. **CONCLUSION**: Hypocitraturia and elevated pH is seen during topiramate use with resultant higher rate of calcium phosphate stone formation compared to the general population. Stopping topiramate leads to significant increase in citrate excretion and normalization of pH. These metabolic disturbances appear to be reversible with medication cessation. Copyright © 2022 Elsevier Inc. All rights reserved.

**URL:** <https://ovidsp.ovid.com/ovidweb.cgi?T=JS&CSC=Y&NEWS=N&PAGE=fulltext&D=med21&DO=10.1016%2fj.urology.2022.01.027> <https://libkey.io/libraries/2592/openurl?genre=article&sid=OVID:medline&id=pmid:35093398&id=doi:10.1016%2Fj.urology.2022.01.027&issn=0090-4295&isbn=&volume=165&issue=&spage=139&pages=139-143&date=2022&title=Urology&atitle=Urinary+Metabolic+Disturbances+During+Topiramate+Use+and+their+Reversibility+Following+Drug+Cessation.&aulast=Pelzman&pid=%3Cauthor%3EPelzman+DL%3BKazi+E%3BJackman+SV%3BSemins+MJ%3C%2Fauthor%3E%3CAN%3E35093398%3C%2FAN%3E%3CDT%3EJournal+Article%3C%2FDT%3E>

23. **Defining 24-hour urine parameters and kidney stone risk of student athletes.**

**Item Type:** Journal Article

**Authors:** Theisen, Katherine M.;Ayyash, Omar;Pere, Maria;Ferroni, Matthew;Riley, Julie;Averch, Timothy D. and Semins, Michelle J.

**Publication Date:** Fe ,2022

**Journal:** Clinical Nephrology 97(2), pp. 86–92

**Abstract: PURPOSE**: Dehydration is a risk factor for kidney stone formation. Studying populations that may experience dehydration without a known increased incidence of stone formation may help understand stone formation and prevention. High-caliber athletes represent such a population. We characterized the urinary environment of high-caliber athletes utilizing 24-hour urine collections with comparison to non-athlete controls. **MATERIALS AND METHODS**: After Institutional Review Board (IRB) and National Collegiate Athletic Association (NCAA) compliance officer approval, 74 college-student athletes and 20 non-athletes were enrolled. Demographics, medical history, and sport of participation were recorded. Participants were asked to provide 24-hour urine collections as well as diet and activity logs at the time of urine collection. Standard stone risk parameters were assessed and compared to litholink reference standards. **RESULTS**: 34 athletes and 10 non-athletes provided at least one 24-hour urine specimen for evaluation. Athletes had a high prevalence of urinary risks for stone formation including low volume (median 1.46 L), low citrate, high sodium, high calcium (females), and high uric acid (males). However, athletes also had a high prevalence of known stone-protective factors such as high urine magnesium. Athletes had a lower urine pH but high supersaturation of uric acid and calcium oxalate compared to non-athletes. **CONCLUSION**: Student athletes appear to have a high prevalence of urinary risk factors for stone formation such as dehydration, high calcium, high uric acid, high sodium, and low citrate. Overall stone risk in this population may be offset by increased levels of stone-protective factors such as magnesium. Further study of this population may help generate hypotheses for effective stone prevention strategies in the general population.

**URL:** <https://ovidsp.ovid.com/ovidweb.cgi?T=JS&CSC=Y&NEWS=N&PAGE=fulltext&D=med21&DO=10.5414%2fCN110355> <https://libkey.io/libraries/2592/openurl?genre=article&sid=OVID:medline&id=pmid:34784999&id=doi:10.5414%2FCN110355&issn=0301-0430&isbn=&volume=97&issue=2&spage=86&pages=86-92&date=2022&title=Clinical+Nephrology&atitle=Defining+24-hour+urine+parameters+and+kidney+stone+risk+of+student+athletes.&aulast=Theisen&pid=%3Cauthor%3ETheisen+KM%3BAyyash+O%3BPere+M%3BFerroni+M%3BRiley+J%3BAverch+TD%3BSemins+MJ%3C%2Fauthor%3E%3CAN%3E34784999%3C%2FAN%3E%3CDT%3EJournal+Article%3C%2FDT%3E>

24. **Nuclear Magnetic Resonance Metabolomic Profiling and Urine Chemistries in Incident Kidney Stone Formers Compared with Controls.**

**Item Type:** Journal Article

**Authors:** Thongprayoon, Charat;Vuckovic, Ivan;Vaughan, Lisa E.;Macura, Slobodan;Larson, Nicholas B.;D'Costa, Matthew R.;Lieske, John C.;Rule, Andrew D. and Denic, Aleksandar

**Publication Date:** No ,2022

**Journal:** Journal of the American Society of Nephrology 33(11), pp. 2071–2086

**Abstract: BACKGROUND**: The urine metabolites and chemistries that contribute to kidney stone formation are not fully understood. This study examined differences between the urine metabolic and chemistries profiles of first-time stone formers and controls. **METHODS**: High-resolution ^1^H-nuclear magnetic resonance (NMR) spectroscopy-based metabolomic analysis was performed in 24-hour urine samples from a prospective cohort of 418 first-time symptomatic kidney stone formers and 440 controls. In total, 48 NMR-quantified metabolites in addition to 12 standard urine chemistries were assayed. Analysis of covariance was used to determine the association of stone former status with urine metabolites or chemistries after adjusting for age and sex and correcting for the false discovery rate. Gradient-boosted machine methods with nested cross-validation were applied to predict stone former status. **RESULTS**: Among the standard urine chemistries, stone formers had lower urine oxalate and potassium and higher urine calcium, phosphate, and creatinine. Among NMR urine metabolites, stone formers had lower hippuric acid, trigonelline, 2-furoylglycine, imidazole, and citrate and higher creatine and alanine. A cross-validated model using urine chemistries, age, and sex yielded a mean AUC of 0.76 (95% CI, 0.73 to 0.79). A cross-validated model using urine chemistries, NMR-quantified metabolites, age, and sex did not meaningfully improve the discrimination (mean AUC, 0.78; 95% CI, 0.75 to 0.81). In this combined model, among the top ten discriminating features, four were urine chemistries and six NMR-quantified metabolites. **CONCLUSIONS**: Although NMR-quantified metabolites did not improve discrimination, several urine metabolic profiles were identified that may improve understanding of kidney stone pathogenesis. Copyright © 2022 by the American Society of Nephrology.

**URL:** <https://ovidsp.ovid.com/ovidweb.cgi?T=JS&CSC=Y&NEWS=N&PAGE=fulltext&D=med22&DO=10.1681%2fASN.2022040416> <https://libkey.io/libraries/2592/openurl?genre=article&sid=OVID:medline&id=pmid:36316097&id=doi:10.1681%2FASN.2022040416&issn=1046-6673&isbn=&volume=33&issue=11&spage=2071&pages=2071-2086&date=2022&title=Journal+of+the+American+Society+of+Nephrology&atitle=Nuclear+Magnetic+Resonance+Metabolomic+Profiling+and+Urine+Chemistries+in+Incident+Kidney+Stone+Formers+Compared+with+Controls.&aulast=Thongprayoon&pid=%3Cauthor%3EThongprayoon+C%3BVuckovic+I%3BVaughan+LE%3BMacura+S%3BLarson+NB%3BD%27Costa+MR%3BLieske+JC%3BRule+AD%3BDenic+A%3C%2Fauthor%3E%3CAN%3E36316097%3C%2FAN%3E%3CDT%3EJournal+Article%3C%2FDT%3E>

25. **Stone Morphology Distinguishes Two Pathways of Idiopathic Calcium Oxalate Stone Pathogenesis.**

**Item Type:** Journal Article

**Authors:** Williams, James C. Jr;Al-Awadi, Haider;Muthenini, Manognya;Bledsoe, Sharon B.;El-Achkar, Tarek;Evan, Andrew P.;Coe, Fredric;Lingeman, James E. and Worcester, Elaine M.

**Publication Date:** 2022

**Journal:** Journal of Endourology 36(5), pp. 694–702

**Abstract: Introduction:** About 1 in 11 Americans will experience a kidney stone, but underlying causes remain obscure. The objective of the present study was to separate idiopathic calcium oxalate stone formers by whether or not they showed positive evidence of forming a stone on Randall's plaque (RP). **Materials and Methods:** In patients undergoing either percutaneous or ureteroscopic procedures for kidney stone removal, all stone material was extracted and analyzed using micro-CT imaging to identify those attached to RP. Twenty-four-hour urine samples were collected weeks after the stone removal procedure and patients were off of medications that would affect urine composition. The endoscopic video was analyzed for papillary pathology (RP, pitting, plugging, dilated ducts, and loss of papillary shape) by an observer blinded to the data on stone type. The percent papillary area occupied by RP and ductal plugging was quantified using image analysis software. **Results:** Patients having even one stone on RP (N = 36) did not differ from non-RP patients (N = 37) in age, sex, BMI, or other clinical characteristics. Compared with the non-RP group, RP stone formers had more numerous, but smaller, stones, more abundant papillary RP formation, and fewer ductal plugs, both by quantitative measurement of surface area (on average, three times more plaque area, but only 41% as much plug area as in non-RP patients) and by semiquantitative visual grading. Serum and blood values did not differ between RP and non-RP stone formers by any measure. **Conclusions:** Growth of many small stones on plaque seems the pathogenetic scheme for the RP stone-forming phenotype, whereas the non-RP phenotype stone pathogenesis pathway is less obvious. Higher papillary plugging in non-RP patients suggests that plugs play a role in stone formation and that these patients have a greater degree of papillary damage. Underlying mechanisms that create these distinctive phenotypes are presently unknown.

**URL:** <https://ovidsp.ovid.com/ovidweb.cgi?T=JS&CSC=Y&NEWS=N&PAGE=fulltext&D=med21&DO=10.1089%2fend.2021.0685> <https://libkey.io/libraries/2592/openurl?genre=article&sid=OVID:medline&id=pmid:34915736&id=doi:10.1089%2Fend.2021.0685&issn=0892-7790&isbn=&volume=36&issue=5&spage=694&pages=694-702&date=2022&title=Journal+of+Endourology&atitle=Stone+Morphology+Distinguishes+Two+Pathways+of+Idiopathic+Calcium+Oxalate+Stone+Pathogenesis.&aulast=Williams&pid=%3Cauthor%3EWilliams+JC+Jr%3BAl-Awadi+H%3BMuthenini+M%3BBledsoe+SB%3BEl-Achkar+T%3BEvan+AP%3BCoe+F%3BLingeman+JE%3BWorcester+EM%3C%2Fauthor%3E%3CAN%3E34915736%3C%2FAN%3E%3CDT%3EJournal+Article%3C%2FDT%3E>

26. **24-Hour urine collection for first time pediatric stone formers: Is it worth it?.**

**Item Type:** Journal Article

**Authors:** Carnes, Kevin;Howe, Adam;Feustel, Paul J.;Listman, James A.;White, Mark and Kogan, Barry A.

**Publication Date:** 2021

**Journal:** Journal of Pediatric Urology 17(3), pp. 387.e1–387.e7

**Abstract: INTRODUCTION/OBJECTIVE**: 24-h urine collections are recommended for motivated first-time stone formers. Given that children have a lifetime potential for recurrences, metabolic work-up has been recommended. 24-hour urine collections can be problematic, especially in children. We sought to study the benefits of 24-h urine collections in children with stones. **STUDY DESIGN**: We performed a single center, retrospective chart review of the most recent pediatric nephrolithiasis patients under age 18 at our center who supplied their first 24-h urine collection. We assessed whether 24-h urine results led to a change in management and if those patients were adherent to the recommendations. **RESULTS**: Seventy pediatric nephrolithiasis patients who had 24-h urine collection were reviewed. Recommendations other than standard dietary and fluid intake changes were made in 8/70 (11%). A low citrate/calcium ratio (327 vs. 525, p : Seventy pediatric nephrolithiasis patients who had 24-h urine collection were reviewed. Recommendations other than standard dietary and fluid intake changes were made in 8/70 (11%). A low citrate/calcium ratio (327 vs. 525, p : Seventy pediatric nephrolithiasis patients who had 24-h urine collection were reviewed. Recommendations other than standard dietary and fluid intake changes were made in 8/70 (11%). A low citrate/calcium ratio (327 vs. 525, p **DISCUSSION**: Our study aimed to evaluate the impact and value of 24-h urine collection in first time pediatric stone formers. We found that 24-h urine collections altered management from standard dietary recommendations in only 11% of cases. These collections were fraught with challenges - 55% of our samples appeared to be incorrectly collected, there was at least one abnormality noted in 100% of collections, these tests are expensive, and patients were poorly compliant with recommendations based on test results. Additionally, changes made based on the 24-h urine results seemed to vary depending on who evaluated the test results. Among cases in which changes were made, nephrologists made alterations at a far greater rate than urologists did. We do acknowledge there are several limitations to our study. First, this is a retrospective chart review. Second, for the urology patients, we were only able to review patient records that were available due to a transition from one electronic medical record to another, resulting in a loss of some earlier patient records. We highly doubt that those records we could not review were significantly different than those we did review. Third, this is a single center design and includes the practice patterns of the providers here. We acknowledge that our local practice patterns may or may not be reflective of national practice patterns, however, most clinicians are likely faced with similar interpretation issues and poor rates of compliance and could benefit from guidelines. **CONCLUSION**: 24-h urine collection for first time pediatric stone formers is expensive, difficult to accomplish and infrequently leads to treatment changes. Our data suggest it adds little for most children with stones and may be better reserved for those children with recurrent stone disease. Copyright © 2020 Journal of Pediatric Urology Company. Published by Elsevier Ltd. All rights reserved.

**URL:** <https://ovidsp.ovid.com/ovidweb.cgi?T=JS&CSC=Y&NEWS=N&PAGE=fulltext&D=med19&DO=10.1016%2fj.jpurol.2020.12.001> <https://libkey.io/libraries/2592/openurl?genre=article&sid=OVID:medline&id=pmid:33762156&id=doi:10.1016%2Fj.jpurol.2020.12.001&issn=1477-5131&isbn=&volume=17&issue=3&spage=387.e1&pages=387.e1-387.e7&date=2021&title=Journal+of+pediatric+urology&atitle=24-Hour+urine+collection+for+first+time+pediatric+stone+formers%3A+Is+it+worth+it%3F.&aulast=Carnes&pid=%3Cauthor%3ECarnes+K%3BHowe+A%3BFeustel+PJ%3BListman+JA%3BWhite+M%3BKogan+BA%3C%2Fauthor%3E%3CAN%3E33762156%3C%2FAN%3E%3CDT%3EJournal+Article%3C%2FDT%3E>

27. **Metabolic assessment in pure struvite stones formers: is it necessary?.**

**Item Type:** Journal Article

**Authors:** Danilovic, Alexandre;Ferreira, Thiago Augusto Cunha;Gomes, Samirah Abreu;Wei, Isabela Akemi;Vicentini, Fabio Carvalho;Torricelli, Fabio Cesar Miranda;Marchini, Giovanni Scala;Mazzucchi, Eduardo;Srougi, Miguel and Nahas, William Carlos

**Publication Date:** 2021

**Journal:** Jornal Brasileiro De Nefrologia 43(2), pp. 200–206

**Abstract: BACKGROUND AND OBJECTIVE**: Magnesium ammonium phosphate stones (MAP), also known as struvite stones, are associated with urinary infection and impairment of renal unit. The aim of this study is to evaluate the urinary metabolic risk factors for recurrence of renal calculi in patients submitted to nephrectomy due to MAP stones. **METHODS**: We retrospectively reviewed the charts of patients > 18 years old submitted to total nephrectomy due to pure MAP stones and pure calcium oxalate (CaOx) stones from July 2006 to July 2016. Urinary metabolic parameters were assessed through 24-hour urine exams >= 3 months after nephrectomy. Urinary metabolic parameters and new event related to lithiasis were compared. **RESULTS**: Twenty-eight and 39 patients were included in MAP and CaOx group, respectively. Abnormalities in 24-hour urine samples were similar between groups. Hypercalciuria occurred in 7.1 and 10.3% of patients in MAP and CaOx group, respectively (p = 0.66), whereas hypocitraturia was present in 65.2 and 59.0% of patients with MAP and CaOx group, respectively (p = 0.41). No significant difference in new events was found between MAP and CaOx groups (17.9 vs. 23.1%, respectively; p = 0.60). **CONCLUSION**: A 24-hour urine evaluation should be offered to patients submitted to nephrectomy due to pure MAP stones in order to detect metabolic risk, improve treatment, and prevent stone recurrence.

**URL:** <https://ovidsp.ovid.com/ovidweb.cgi?T=JS&CSC=Y&NEWS=N&PAGE=fulltext&D=med19&DO=10.1590%2f2175-8239-JBN-2020-0106> <https://libkey.io/libraries/2592/openurl?genre=article&sid=OVID:medline&id=pmid:33577639&id=doi:10.1590%2F2175-8239-JBN-2020-0106&issn=0101-2800&isbn=&volume=43&issue=2&spage=200&pages=200-206&date=2021&title=Jornal+Brasileiro+de+Nefrologia&atitle=Metabolic+assessment+in+pure+struvite+stones+formers%3A+is+it+necessary%3F.&aulast=Danilovic&pid=%3Cauthor%3EDanilovic+A%3BFerreira+TAC%3BGomes+SA%3BWei+IA%3BVicentini+FC%3BTorricelli+FCM%3BMarchini+GS%3BMazzucchi+E%3BSrougi+M%3BNahas+WC%3C%2Fauthor%3E%3CAN%3E33577639%3C%2FAN%3E%3CDT%3EJournal+Article%3C%2FDT%3E>

28. **Lumasiran, an RNAi Therapeutic for Primary Hyperoxaluria Type 1.**

**Item Type:** Journal Article

**Authors:** Garrelfs, Sander F.;Frishberg, Yaacov;Hulton, Sally A.;Koren, Michael J.;O'Riordan, William D.;Cochat, Pierre;Deschenes, Georges;Shasha-Lavsky, Hadas;Saland, Jeffrey M.;Van't Hoff, William G.;Fuster, Daniel G.;Magen, Daniella;Moochhala, Shabbir H.;Schalk, Gesa;Simkova, Eva;Groothoff, Jaap W.;Sas, David J.;Meliambro, Kristin A.;Lu, Jiandong;Sweetser, Marianne T., et al

**Publication Date:** 2021

**Journal:** New England Journal of Medicine 384(13), pp. 1216–1226

**Abstract: BACKGROUND**: Primary hyperoxaluria type 1 (PH1) is a rare genetic disease caused by hepatic overproduction of oxalate that leads to kidney stones, nephrocalcinosis, kidney failure, and systemic oxalosis. Lumasiran, an investigational RNA interference (RNAi) therapeutic agent, reduces hepatic oxalate production by targeting glycolate oxidase. **METHODS**: In this double-blind, phase 3 trial, we randomly assigned (in a 2:1 ratio) patients with PH1 who were 6 years of age or older to receive subcutaneous lumasiran or placebo for 6 months (with doses given at baseline and at months 1, 2, 3, and 6). The primary end point was the percent change in 24-hour urinary oxalate excretion from baseline to month 6 (mean percent change across months 3 through 6). Secondary end points included the percent change in the plasma oxalate level from baseline to month 6 (mean percent change across months 3 through 6) and the percentage of patients with 24-hour urinary oxalate excretion no higher than 1.5 times the upper limit of the normal range at month 6. **RESULTS**: A total of 39 patients underwent randomization; 26 were assigned to the lumasiran group and 13 to the placebo group. The least-squares mean difference in the change in 24-hour urinary oxalate excretion (lumasiran minus placebo) was -53.5 percentage points (P: A total of 39 patients underwent randomization; 26 were assigned to the lumasiran group and 13 to the placebo group. The least-squares mean difference in the change in 24-hour urinary oxalate excretion (lumasiran minus placebo) was -53.5 percentage points (P: A total of 39 patients underwent randomization; 26 were assigned to the lumasiran group and 13 to the placebo group. The least-squares mean difference in the change in 24-hour urinary oxalate excretion (lumasiran minus placebo) was -53.5 percentage points (P: A total of 39 patients underwent randomization; 26 were assigned to the lumasiran group and 13 to the placebo group. The least-squares mean difference in the change in 24-hour urinary oxalate excretion (lumasiran minus placebo) was -53.5 percentage points (P**CONCLUSIONS**: Lumasiran reduced urinary oxalate excretion, the cause of progressive kidney failure in PH1. The majority of patients who received lumasiran had normal or near-normal levels after 6 months of treatment. (Funded by Alnylam Pharmaceuticals; ILLUMINATE-A ClinicalTrials.gov number, NCT03681184.). Copyright © 2021 Massachusetts Medical Society.

**URL:** <https://ovidsp.ovid.com/ovidweb.cgi?T=JS&CSC=Y&NEWS=N&PAGE=fulltext&D=med19&DO=10.1056%2fNEJMoa2021712> <https://libkey.io/libraries/2592/openurl?genre=article&sid=OVID:medline&id=pmid:33789010&id=doi:10.1056%2FNEJMoa2021712&issn=0028-4793&isbn=&volume=384&issue=13&spage=1216&pages=1216-1226&date=2021&title=New+England+Journal+of+Medicine&atitle=Lumasiran%2C+an+RNAi+Therapeutic+for+Primary+Hyperoxaluria+Type+1.&aulast=Garrelfs&pid=%3Cauthor%3EGarrelfs+SF%3BFrishberg+Y%3BHulton+SA%3BKoren+MJ%3BO%27Riordan+WD%3BCochat+P%3BDeschenes+G%3BShasha-Lavsky+H%3BSaland+JM%3BVan%27t+Hoff+WG%3BFuster+DG%3BMagen+D%3BMoochhala+SH%3BSchalk+G%3BSimkova+E%3BGroothoff+JW%3BSas+DJ%3BMeliambro+KA%3BLu+J%3BSweetser+MT%3BGarg+PP%3BVaishnaw+AK%3BGansner+JM%3BMcGregor+TL%3BLieske+JC%3C%2Fauthor%3E%3CAN%3E33789010%3C%2FAN%3E%3CDT%3EClinical+Trial%2C+Phase+III%3C%2FDT%3E>

29. **Investigating Fluid Intake in an Underserved Community: What Factors Are Associated with Low Urine Volume on 24-Hour Urine Collection?.**

**Item Type:** Journal Article

**Authors:** Ghiraldi, Eric M.;Nourian, Alex;Chen, Michelle and Friedlander, Justin I.

**Publication Date:** 2021

**Journal:** Journal of Endourology 35(11), pp. 1723–1728

**Abstract: Background:** Stone prevention is dependent on high fluid intake, with evidence that low urine volume (LUV) can promote nephrolithiasis in the absence of other metabolic abnormalities. Herein, we investigate patient-related factors associated with LUV on initial 24-hour urine collection in an underserved population. **Materials and Methods:** A retrospective chart review was performed of patients treated by a single surgeon for nephrolithiasis from August 2014 to January 2019. Patients who submitted 24-hour urine samples were divided into two groups based on whether their initial collection was >2 L or not. Factors associated with 24-hour urine volume were analyzed using bivariate and multivariate analyses. As a secondary outcome, we investigated factors associated with corrected 24-hour urine volume on repeat 24-hour urine collection. **Results:** Two hundred eight patients (45.1%) submitted 24-hour urine collections, 63.9% (133/208) of which demonstrated a urine volume of Two hundred eight patients (45.1%) submitted 24-hour urine collections, 63.9% (133/208) of which demonstrated a urine volume of **Conclusions:** Female patients and those with no insurance or Medicaid were more likely to have LUV on an initial 24-hour urine collection. Further research into barriers to fluid intake is important for these two groups, along with directed patient education on strategies for increasing fluid intake.

**URL:** <https://ovidsp.ovid.com/ovidweb.cgi?T=JS&CSC=Y&NEWS=N&PAGE=fulltext&D=med20&DO=10.1089%2fend.2020.0879> <https://libkey.io/libraries/2592/openurl?genre=article&sid=OVID:medline&id=pmid:33899496&id=doi:10.1089%2Fend.2020.0879&issn=0892-7790&isbn=&volume=35&issue=11&spage=1723&pages=1723-1728&date=2021&title=Journal+of+Endourology&atitle=Investigating+Fluid+Intake+in+an+Underserved+Community%3A+What+Factors+Are+Associated+with+Low+Urine+Volume+on+24-Hour+Urine+Collection%3F.&aulast=Ghiraldi&pid=%3Cauthor%3EGhiraldi+EM%3BNourian+A%3BChen+M%3BFriedlander+JI%3C%2Fauthor%3E%3CAN%3E33899496%3C%2FAN%3E%3CDT%3EJournal+Article%3C%2FDT%3E>

30. **Prospective Randomized Evaluation of Idiopathic Hyperoxaluria Treatments.**

**Item Type:** Journal Article

**Authors:** Gupta, Mantu;Gallante, Blair;Bamberger, Jacob N.;Khusid, Johnathan A.;Parkhomenko, Egor;Chandhoke, Ryan;Capodice, Jillian and Atallah, William

**Publication Date:** 2021

**Journal:** Journal of Endourology 35(12), pp. 1844–1851

**Abstract: Purpose:** Calcium oxalate (CaOx) stone formation is influenced by urinary oxalate excretion. Stone formers with elevated urinary oxalate are commonly prescribed a low-oxalate diet or oral supplementation with vitamin B6 and magnesium to reduce urinary oxalate excretion. This study aims to compare the effects of dietary modification vs supplementation vs a combination of both on urinary oxalate. **Materials and Methods:** We enrolled patients with a documented history of CaOx stones and newly diagnosed idiopathic hyperoxaluria. Patients were randomized into three treatment groups: low oxalate diet (D), supplementation with 25 mg vitamin B6 and 400 mg magnesium oxide (S), or both low oxalate diet and B6/magnesium supplementation (DS). Baseline and 3-month postintervention 24-hour urine tests were obtained. The primary endpoint was change in 24-hour urinary oxalate (Ox24) at 12 weeks. Secondary endpoints included changes in other 24-hour urine parameters, compliance rates, and adverse effect rates. **Results:** In total, 164 patients were recruited and 62, 47, and 55 were enrolled into the D, S, and DS groups, respectively. Of these, 99 patients completed the study (56.5% of the D, 72.3% of the S, and 54.6% of the DS groups, respectively). Significant differences were noted in median percent reduction in Ox24 values (-31.1% vs -16.0% vs -23.9%, p = 0.007) in the D, S, and DS groups, respectively. Furthermore, the percentages of patients within each treatment arm who realized a decrease in Ox24 were also found to be significantly different: D = 91.4% vs. S = 67.6% vs DS = 86.7%, p = 0.027. No significant adverse events were observed in any of the study arms. **Conclusion:** Low oxalate diet is more effective than B6/magnesium supplementation at lowering urinary oxalate in idiopathic hyperoxaluric stone formers. Combination therapy did not produce greater reductions in urinary oxalate than either of the monotherapy arms suggesting it is of little clinical utility. Further study with long-term longitudinal follow-up is required to determine if these treatment strategies reduce recurrent stone events in this population.

**URL:** <https://ovidsp.ovid.com/ovidweb.cgi?T=JS&CSC=Y&NEWS=N&PAGE=fulltext&D=med20&DO=10.1089%2fend.2021.0122> <https://libkey.io/libraries/2592/openurl?genre=article&sid=OVID:medline&id=pmid:34254834&id=doi:10.1089%2Fend.2021.0122&issn=0892-7790&isbn=&volume=35&issue=12&spage=1844&pages=1844-1851&date=2021&title=Journal+of+Endourology&atitle=Prospective+Randomized+Evaluation+of+Idiopathic+Hyperoxaluria+Treatments.&aulast=Gupta&pid=%3Cauthor%3EGupta+M%3BGallante+B%3BBamberger+JN%3BKhusid+JA%3BParkhomenko+E%3BChandhoke+R%3BCapodice+J%3BAtallah+W%3C%2Fauthor%3E%3CAN%3E34254834%3C%2FAN%3E%3CDT%3EJournal+Article%3C%2FDT%3E>

31. **Comparison of Selective Versus Empiric Pharmacologic Preventative Therapy With Kidney Stone Recurrence.**

**Item Type:** Journal Article

**Authors:** Hsi, Ryan S.;Yan, Phyllis L.;Goldfarb, David S.;Egbuji, Ada;Si, Yajuan;Shahinian, Vahakn and Hollingsworth, John M.

**Publication Date:** 2021

**Journal:** Urology 149, pp. 81–88

**Abstract: OBJECTIVE**: To assess the effectiveness of an empiric approach to metabolic stone prevention. **METHODS**: Using medical claims from a cohort of working age adults with kidney stone diagnoses (2008-2017), we identified the subset who were prescribed thiazides, alkali therapy, or allopurinol-collectively known as preventive pharmacologic therapy (PPT). We distinguished between those who had 24-hour urine testing prior to initiating PPT (selective therapy) from those without it (empiric therapy). We conducted a survival analysis for time to first recurrence for stone-related events, including ED visits, hospitalizations, and surgery, up to 2 years after initiating PPT. **RESULTS**: Of 10,125 patients identified, 2744 (27%) and 7381 (73%) received selective and empiric therapy, respectively. The overall frequency of any stone-related event was 11%, and this did not differ between the 2 groups on bivariate analysis (P=.29). After adjusting for sociodemographic factors, comorbidities, medication class, and adherence, there was no difference in the hazard of a stone-related event between the selective and empiric therapy groups (hazard ratio, 0.97; 95% confidence interval, 0.84-1.12). When considered individually, the frequency of ED visits, hospitalizations, and surgeries did not differ between groups. Greater adherence to PPT and older age were associated with a lower hazard of a stone-related event (both P : Of 10,125 patients identified, 2744 (27%) and 7381 (73%) received selective and empiric therapy, respectively. The overall frequency of any stone-related event was 11%, and this did not differ between the 2 groups on bivariate analysis (P=.29). After adjusting for sociodemographic factors, comorbidities, medication class, and adherence, there was no difference in the hazard of a stone-related event between the selective and empiric therapy groups (hazard ratio, 0.97; 95% confidence interval, 0.84-1.12). When considered individually, the frequency of ED visits, hospitalizations, and surgeries did not differ between groups. Greater adherence to PPT and older age were associated with a lower hazard of a stone-related event (both P **CONCLUSION**: Compared to empiric therapy, PPT guided by 24-hour urine testing, on average, is not associated with a lower hazard of a stone-related event. These results suggest a need to identify kidney stone patients who benefit from 24-hour urine testing. Copyright © 2020 Elsevier Inc. All rights reserved.

**URL:** <https://ovidsp.ovid.com/ovidweb.cgi?T=JS&CSC=Y&NEWS=N&PAGE=fulltext&D=med19&DO=10.1016%2fj.urology.2020.11.054> <https://libkey.io/libraries/2592/openurl?genre=article&sid=OVID:medline&id=pmid:33352163&id=doi:10.1016%2Fj.urology.2020.11.054&issn=0090-4295&isbn=&volume=149&issue=&spage=81&pages=81-88&date=2021&title=Urology&atitle=Comparison+of+Selective+Versus+Empiric+Pharmacologic+Preventative+Therapy+With+Kidney+Stone+Recurrence.&aulast=Hsi&pid=%3Cauthor%3EHsi+RS%3BYan+PL%3BGoldfarb+DS%3BEgbuji+A%3BSi+Y%3BShahinian+V%3BHollingsworth+JM%3C%2Fauthor%3E%3CAN%3E33352163%3C%2FAN%3E%3CDT%3EComparative+Study%3C%2FDT%3E>

32. **Living Donor Gifted Lithiasis: Long-Term Outcomes in Recipients.**

**Item Type:** Journal Article

**Authors:** Jan, Muhammad Yahya;Sharfuddin, Asif;Mujtaba, Muhammad;Adebiyi, Oluwafisayo;Sundaram, Chandru P.;Yaqub, Muhammad;Taber, Tim;Powelson, John;Goggins, William and Doshi, Simit

**Publication Date:** Ap ,2021

**Journal:** Transplantation Proceedings 53(3), pp. 1091–1094

**Abstract: OBJECTIVES**: Presence of nephrolithiasis in a living donor has been at least a relative contraindication to living donor nephrectomy. The concern for stone recurrence and outcomes has been one of the reasons for reluctance to consider these medically complex donors. We evaluate long-term outcomes in recipients of kidney grafts from donors with nephrolithiasis, or history of nephrolithiasis, and provide results from our experience at Indiana University. **MATERIALS AND METHODS**: We retrospectively reviewed 57 donor-recipient pairs, where the allograft was received from a living donor with symptomatic calculi, or with imaging evidence of kidney stones, between 2003 and 2018. This research study was done in compliance with the ethical standards set forth in the Helsinki Congress. **RESULTS**: The mean age of recipients was 46+/-19 years and 58% were male. Kidney recipients were followed for a median of 3.5 years and 59.6% of patients had follow-up imaging studies. None of the recipients had obstructing renal calculi or related infections. None of the recipients required any interventions for recurrent calculi and no stone episode lead to adverse event to the graft. Hyperoxaluria and hypercalciuria were the most common risk factors in 24-hour urine collections obtained from donors. **CONCLUSIONS**: Our findings from a single large center looking at kidney recipient outcomes over a long follow-up period found that gifted lithiasis is a safe procedure. Careful selection of "medically complex donors" with kidney stones based on appropriate guidelines is a key step. Further studies are needed to help develop consensus guidelines. Copyright © 2021 Elsevier Inc. All rights reserved.

**URL:** <https://ovidsp.ovid.com/ovidweb.cgi?T=JS&CSC=Y&NEWS=N&PAGE=fulltext&D=med19&DO=10.1016%2fj.transproceed.2021.01.015> <https://libkey.io/libraries/2592/openurl?genre=article&sid=OVID:medline&id=pmid:33608128&id=doi:10.1016%2Fj.transproceed.2021.01.015&issn=0041-1345&isbn=&volume=53&issue=3&spage=1091&pages=1091-1094&date=2021&title=Transplantation+Proceedings&atitle=Living+Donor+Gifted+Lithiasis%3A+Long-Term+Outcomes+in+Recipients.&aulast=Jan&pid=%3Cauthor%3EJan+MY%3BSharfuddin+A%3BMujtaba+M%3BAdebiyi+O%3BSundaram+CP%3BYaqub+M%3BTaber+T%3BPowelson+J%3BGoggins+W%3BDoshi+S%3C%2Fauthor%3E%3CAN%3E33608128%3C%2FAN%3E%3CDT%3EEvaluation+Study%3C%2FDT%3E>

33. **Misleading localization by ^18^F-fluorocholine PET/CT in familial hypocalciuric hypercalcemia type-3: a case report.**

**Item Type:** Journal Article

**Authors:** Mukhtar, Noha N.;Abouzied, Mohei El-Din M.;Alqahtani, Mohammed H. and Hammami, Muhammad M.

**Publication Date:** Jan 26 ,2021

**Journal:** BMC Endocrine Disorders 21(1), pp. 20

**Abstract: BACKGROUND**: Familial hypocalciuric hypercalcemia (FHH) is a heterogeneous autosomal-dominant disorder of calcium hemostasis that may be difficult to distinguish clinically from mild primary hyperparathyroidism. Loss-of-function mutations mainly involving Arg15 residue of the adaptor-related protein complex 2, sigma subunit 1 (AP2S1) cause a rarer, more recently recognized form of FHH, FFH type-3. Recently, ^18^F-fluorocholine positron emission tomography/computed tomography (FCH-PET/CT) showed superior sensitivity to conventional imaging in localizing parathyroid adenomas. We report a new FFH type-3 patient who underwent unnecessary parathyroidectomy in association with misleading FCH-PET/CT imaging. **CASE PRESENTATION**: A 29-year old woman was initially evaluated for parathyroid hormone (PTH)-dependent hypercalcemia in 2013. Medical history was positive only for chronic constipation and malaise with no personal or family history of hypercalcemia, kidney stones, or neck surgery. Over seven years, serum calcium level was 2.51-2.89 mmol/L with concomitant PTH level of 58.7-94.8 mmol/L. Serum phosphate levels were in the low/low normal range. Serum creatinine and magnesium levels were normal. 25-hydroxy vitamin D level was 13 nmol/L. 24-hour urine calcium level was 1.92 mmol/day but increased to 6.99 mmol/day after treatment with cholecalciferol 1000 IU daily. Bone mineral density and renal ultrasound were normal. Parathyroid ultrasound showed two hypoechoic nodules inferior to the left and right thyroid lobes; however, ^99m^technitium-sestamibi scans (2013, 2016, 2018) were negative. FCH-PET/CT (2019) showed focal uptake co-localizing with the nodule inferior to the left thyroid lobe. The patient underwent left inferior parathyroidectomy and pathology was consistent with parathyroid hyperplasia. However, postoperatively, serum calcium and PTH levels remained elevated and FCH-PET/CT and ultrasound showed persistence of the uptake/nodule. Whole exome sequencing showed Arg15Cys mutation in the AP2S1 gene characteristic of FHH type-3. **CONCLUSIONS**: In this new case of FHH type-3, FCH-PET/CT failed to localize to the hyperplastic parathyroid glands and localized instead to apparently a lymph node. This, together with increased urinary calcium after vitamin D supplementation, led to unnecessary parathyroidectomy. Given the increasingly lower cost of genetic testing and the cost of follow up and unnecessary surgery, it may prudent to include genetic testing for FHH early on in patients with mild PTH-dependent hypercalcemia.

**URL:** <https://ovidsp.ovid.com/ovidweb.cgi?T=JS&CSC=Y&NEWS=N&PAGE=fulltext&D=med19&DO=10.1186%2fs12902-021-00683-z> <https://libkey.io/libraries/2592/openurl?genre=article&sid=OVID:medline&id=pmid:33499837&id=doi:10.1186%2Fs12902-021-00683-z&issn=1472-6823&isbn=&volume=21&issue=1&spage=20&pages=20&date=2021&title=BMC+Endocrine+Disorders&atitle=Misleading+localization+by+18F-fluorocholine+PET%2FCT+in+familial+hypocalciuric+hypercalcemia+type-3%3A+a+case+report.&aulast=Mukhtar&pid=%3Cauthor%3EMukhtar+NN%3BAbouzied+MEM%3BAlqahtani+MH%3BHammami+MM%3C%2Fauthor%3E%3CAN%3E33499837%3C%2FAN%3E%3CDT%3EJournal+Article%3C%2FDT%3E>

34. **Prevention of Urinary Stones With Hydration (PUSH): Design and Rationale of a Clinical Trial.**

**Item Type:** Journal Article

**Authors:** Scales, Charles D. Jr;Desai, Alana C.;Harper, Jonathan D.;Lai, H. Henry;Maalouf, Naim M.;Reese, Peter P.;Tasian, Gregory E.;Al-Khalidi, Hussein R.;Kirkali, Ziya and Wessells, Hunter

**Publication Date:** 2021

**Journal:** American Journal of Kidney Diseases 77(6), pp. 898–906.e1

**Abstract: RATIONALE & OBJECTIVE**: Although maintaining high fluid intake is an effective low-risk intervention for the secondary prevention of urinary stone disease, many patients with stones do not increase their fluid intake. **STUDY DESIGN**: We describe the rationale and design of the Prevention of Urinary Stones With Hydration (PUSH) Study, a randomized trial of a multicomponent behavioral intervention program to increase and maintain high fluid intake. Participants are randomly assigned (1:1 ratio) to the intervention or control arm. The target sample size is 1,642 participants. **SETTING & PARTICIPANTS**: Adults and adolescents 12 years and older with a symptomatic stone history and low urine volume are eligible. Exclusion criteria include infectious or monogenic causes of urinary stone disease and comorbid conditions precluding increased fluid intake. **INTERVENTIONS**: All participants receive usual care and a smart water bottle with smartphone application. Participants in the intervention arm receive a fluid intake prescription and an adaptive program of behavioral interventions, including financial incentives, structured problem solving, and other automated adherence interventions. Control arm participants receive guideline-based fluid instructions. **OUTCOMES**: The primary end point is recurrence of a symptomatic stone during 24 months of follow-up. Secondary end points include changes in radiographic stone burden, 24-hour urine output, and urinary symptoms. **LIMITATIONS**: Periodic 24-hour urine volumes may not fully reflect daily behavior. **CONCLUSIONS**: With its highly novel features, the PUSH Study will address an important health care problem. **FUNDING**: National Institute of Diabetes and Digestive and Kidney Diseases. **TRIAL REGISTRATION**: Registered at ClinicalTrials.gov with study number NCT03244189. Copyright © 2020 National Kidney Foundation, Inc. All rights reserved.

**URL:** <https://ovidsp.ovid.com/ovidweb.cgi?T=JS&CSC=Y&NEWS=N&PAGE=fulltext&D=med19&DO=10.1053%2fj.ajkd.2020.09.016> <https://libkey.io/libraries/2592/openurl?genre=article&sid=OVID:medline&id=pmid:33212205&id=doi:10.1053%2Fj.ajkd.2020.09.016&issn=0272-6386&isbn=&volume=77&issue=6&spage=898&pages=898-906.e1&date=2021&title=American+Journal+of+Kidney+Diseases&atitle=Prevention+of+Urinary+Stones+With+Hydration+%28PUSH%29%3A+Design+and+Rationale+of+a+Clinical+Trial.&aulast=Scales&pid=%3Cauthor%3EScales+CD+Jr%3BDesai+AC%3BHarper+JD%3BLai+HH%3BMaalouf+NM%3BReese+PP%3BTasian+GE%3BAl-Khalidi+HR%3BKirkali+Z%3BWessells+H%3C%2Fauthor%3E%3CAN%3E33212205%3C%2FAN%3E%3CDT%3EClinical+Trial+Protocol%3C%2FDT%3E>

35. **Are individual analyses of multiple short urine collections throughout the 24 hours superior to a standard 24-hour urine collection in precipitation risk assessment of healthy subjects?.**

**Item Type:** Journal Article

**Authors:** Shafiee, Mohammad A.;Shaker, Pouyan;Hosseini, Sayyideh F.;Alavinia, Mohammad;Aarabi, Mehdi;Rezaee, Ali J. and Halperin, Mitchel L.

**Publication Date:** Ma ,2021

**Journal:** Nephrology 26(3), pp. 234–238

**Abstract: PURPOSE**: The commonly used 24-hour collection technique has been the mainstay of diagnosis for supersaturation but has some certain limitations. Hence, superiority of multiple short urine collections as a new alternative in precipitation risk assessment was assessed compared to the standard 24-hour urine collection among healthy subjects. **MATERIALS AND METHODS**: Individual urine samples of 26 healthy subjects were acquired every 2 to 3 hours throughout the 24 hours. Urine samples were obtained and the time and volume of each sample were recorded. Urinary constituents involved in precipitation including, sodium-potassium, chloride, calcium, phosphate, citrate, magnesium, urea, creatinine and pH were measured. A simulated 24-hour collection was recalculated by the totalling of all shorter urine collections volume and urinary constituents excretions throughout the day. **RESULTS**: Urine pH, urine creatinine and precipitation rate had a significantly lower values in 24-hours urine collection compared to one individual value of multiple urine collections by -0.769 (P : Urine pH, urine creatinine and precipitation rate had a significantly lower values in 24-hours urine collection compared to one individual value of multiple urine collections by -0.769 (P : Urine pH, urine creatinine and precipitation rate had a significantly lower values in 24-hours urine collection compared to one individual value of multiple urine collections by -0.769 (P : Urine pH, urine creatinine and precipitation rate had a significantly lower values in 24-hours urine collection compared to one individual value of multiple urine collections by -0.769 (P : Urine pH, urine creatinine and precipitation rate had a significantly lower values in 24-hours urine collection compared to one individual value of multiple urine collections by -0.769 (P : Urine pH, urine creatinine and precipitation rate had a significantly lower values in 24-hours urine collection compared to one individual value of multiple urine collections by -0.769 (P : Urine pH, urine creatinine and precipitation rate had a significantly lower values in 24-hours urine collection compared to one individual value of multiple urine collections by -0.769 (P : Urine pH, urine creatinine and precipitation rate had a significantly lower values in 24-hours urine collection compared to one individual value of multiple urine collections by -0.769 (P **CONCLUSION**: Based on the results, individual analysis of multiple shorter urine collections throughout the day improves the ability of identifying supersaturation points, precipitation risk zones and may potentially improve risk assessment compared to the 24-hour urine collection method. Copyright © 2020 Asian Pacific Society of Nephrology.

**URL:** <https://ovidsp.ovid.com/ovidweb.cgi?T=JS&CSC=Y&NEWS=N&PAGE=fulltext&D=med19&DO=10.1111%2fnep.13842> <https://libkey.io/libraries/2592/openurl?genre=article&sid=OVID:medline&id=pmid:33314554&id=doi:10.1111%2Fnep.13842&issn=1320-5358&isbn=&volume=26&issue=3&spage=234&pages=234-238&date=2021&title=Nephrology&atitle=Are+individual+analyses+of+multiple+short+urine+collections+throughout+the+24+hours+superior+to+a+standard+24-hour+urine+collection+in+precipitation+risk+assessment+of+healthy+subjects%3F.&aulast=Shafiee&pid=%3Cauthor%3EShafiee+MA%3BShaker+P%3BHosseini+SF%3BAlavinia+M%3BAarabi+M%3BRezaee+AJ%3BHalperin+ML%3C%2Fauthor%3E%3CAN%3E33314554%3C%2FAN%3E%3CDT%3EJournal+Article%3C%2FDT%3E>

36. **Effect of a Probiotic Supplement Containing Lactobacillus Acidophilus and Bifidobacterium Animalis Lactis on Urine Oxalate in Calcium Stone Formers with Hyperoxaluria: A Randomized, Placebo-controlled, Double-blind and In-vitro Trial.**

**Item Type:** Journal Article

**Authors:** Tavasoli, Sanaz;Jalali, Saba;Naji, Mohammad;Borumandnia, Nasrin;Shakiba Majd, Ghazaleh;Basiri, Abbas;Khosravi Darani, Kianaoush;Karamad, Dina;Tajabadi-Ebrahimi, Maryam and Taheri, Maryam

**Publication Date:** Jun 15 ,2021

**Journal:** Urology Journal 19(3), pp. 179–188

**Abstract: PURPOSE**: To determine the effect of a probiotic supplement containing native Lactobacillus acidophilus (L. acidophilus) and Bifidobacterium animalis lactis (B. lactis) on 24-hour urine oxalate in recurrent calcium stone formers with hyperoxaluria. Moreover, the in-vitro oxalate degradation capacity and the intestinal colonization of consumed probiotics were evaluated. **MATERIALS AND METHODS**: The oxalate degrading activity of L. acidophilus and B. lactis were evaluated in-vitro. The presence of oxalyl-CoA decarboxylase (oxc) gene in the probiotic species was assessed. One hundred patients were randomized to receive the probiotic supplement or placebo for four weeks. The 24-hour urine oxalate and the colonization of consumed probiotics were assessed after weeks four and eight. **RESULTS**: Although the oxc gene was present in both species, only L. acidophilus had a good oxalate degrading activity, in-vitro. Thirty-four patients from the probiotic and thirty patients from the placebo group finished the study. The urine oxalate changes were not significantly different between groups (57.21 +/- 11.71 to 49.44 +/- 18.14 mg/day for probiotic, and 56.43 +/- 9.89 to 50.47 +/- 18.04 mg/day for placebo) (P = .776). The probiotic consumption had no significant effect on urine oxalate, both in univariable (P = .771) and multivariable analyses (P = .490). The consumed probiotics were not detected in the stool samples of most participants. **CONCLUSION**: Our results showed that the consumption of a probiotic supplement containing L. acidophilus and B. lactis did not affect urine oxalate. The results may be due to a lack of bacterial colonization in the intestine.

**URL:** <https://ovidsp.ovid.com/ovidweb.cgi?T=JS&CSC=Y&NEWS=N&PAGE=fulltext&D=med20&DO=10.22037%2fuj.v18i.6789> <https://libkey.io/libraries/2592/openurl?genre=article&sid=OVID:medline&id=pmid:34129232&id=doi:10.22037%2Fuj.v18i.6789&issn=1735-1308&isbn=&volume=19&issue=3&spage=179&pages=179-188&date=2021&title=Urology+Journal&atitle=Effect+of+a+Probiotic+Supplement+Containing+Lactobacillus+Acidophilus+and+Bifidobacterium+Animalis+Lactis+on+Urine+Oxalate+in+Calcium+Stone+Formers+with+Hyperoxaluria%3A+A+Randomized%2C+Placebo-controlled%2C+Double-blind+and+In-vitro+Trial.&aulast=Tavasoli&pid=%3Cauthor%3ETavasoli+S%3BJalali+S%3BNaji+M%3BBorumandnia+N%3BShakiba+Majd+G%3BBasiri+A%3BKhosravi+Darani+K%3BKaramad+D%3BTajabadi-Ebrahimi+M%3BTaheri+M%3C%2Fauthor%3E%3CAN%3E34129232%3C%2FAN%3E%3CDT%3EJournal+Article%3C%2FDT%3E>

37. **Renal lithiasis in pediatric patients: correlation of methods that depend on 24-hour collections with simpler methods that do not require timed urine.**

**Item Type:** Journal Article

**Authors:** Ubetagoyena Arrieta, M.;Areses Trapote, R.;Mendia Ubetagoyena, J.;Perez Revuelta, M. S. and Letona Luqui, M.

**Publication Date:** Ma ,2021

**Journal:** Actas Urologicas Espanolas 45(2), pp. 146–153

**Abstract: INTRODUCTION**: Daily practice requires quick, simple and accessible methods to appropriately assess the urinary excretion of solutes in diagnostic or follow-up evaluations of children with renal lithiasis. **OBJECTIVES**: The objective of this study was to correlate urine elimination of substances related to renal lithiasis that depend on the volume of excreted urine in a unit of time with other parameters that are calculated by measuring the concentration of these substances in blood and urine, such as urinary ratios, fractional excretions and excretion rates. **MATERIALS AND METHODS**: The study included 401 healthy children aged 3-14 years (187 boys and 214 girls), mean age 8.78+/-3.40 years. The analysis was carried out by Pearson's correlation coefficient. **RESULTS**: There was significant correlation between the elimination of sodium, potassium and chlorine in 24-hour urine sample and the urinary ratios and fractional excretions of these ions. Urinary ratios and rates of excretion of calcium, uric acid, phosphate, magnesium, citrate and oxalate were highly correlated with the determinations of these substances in 24-hour collections. **CONCLUSIONS**: These equations provide relevant information for the study of the etiology of renal lithiasis in children, as well as about compliance to dietary treatment. They also assess the effectiveness of the various treatments used in these patients, without having to resort to 24-hour collections, which pose a considerable challenge in the pediatric age group. Copyright © 2020 AEU. Publicado por Elsevier Espana, S.L.U. All rights reserved.

**URL:** <https://ovidsp.ovid.com/ovidweb.cgi?T=JS&CSC=Y&NEWS=N&PAGE=fulltext&D=med19&DO=10.1016%2fj.acuro.2020.09.006> <https://libkey.io/libraries/2592/openurl?genre=article&sid=OVID:medline&id=pmid:33160758&id=doi:10.1016%2Fj.acuro.2020.09.006&issn=2173-5786&isbn=&volume=45&issue=2&spage=146&pages=146-153&date=2021&title=Actas+Urologicas+Espanolas&atitle=Litiasis+en+pediatria%3A+correlacion+de+metodos+que+dependen+de+la+recogida+de+orina+de+24+horas+con+otros+mas+sencillos+que+no+requieren+orina+minutada.&aulast=Ubetagoyena+Arrieta&pid=%3Cauthor%3EUbetagoyena+Arrieta+M%3BAreses+Trapote+R%3BMendia+Ubetagoyena+J%3BPerez+Revuelta+MS%3BLetona+Luqui+M%3C%2Fauthor%3E%3CAN%3E33160758%3C%2FAN%3E%3CDT%3EJournal+Article%3C%2FDT%3E>

38. **24-Hour Urine Calcium Oxalate Supersaturation Risk Correlates with Computerized Tomography Volumetric Calcium Oxalate Stone Growth.**

**Item Type:** Journal Article

**Authors:** Yuzhakov, Stanislav;Steadman, Shavano D.;Otto, Brandon J.;Bird, Vincent G. and Canales, Benjamin K.

**Publication Date:** 2021

**Journal:** Journal of Urology 206(6), pp. 1438–1444

**Abstract: PURPOSE**: In vitro experiments demonstrate calcium oxalate (CaOx) supersaturation (SS) drives CaOx nucleation and growth. We investigated the link between 24-hour urine SS CaOx and in vivo stone growth through a natural history, imaging study. **MATERIALS AND METHODS**: Using an institutional review board-approved database, we sought >80% CaOx stone formers who prior to stone intervention obtained 2 separate computerized tomography (CT) scans with at least one 24-hour urine collection between scans. Two blinded reviewers calculated bilateral 3-dimensional stone volume using the Visage 7 R region of interest pen tool. CT volume difference was divided by time between scans, and SS CaOx was grouped into low (10). Statistical significance between groups was assessed by Kruskal-Wallis test. **RESULTS**: We identified 80 individuals with stone growth measured by 3-dimensional CT (mean ~7 months between studies). Inter-reviewer reliability of CT volume measurement was well correlated (0.98, Gwet's AC2), and an arbitrator was only needed in 13/160 (8%) cases. Median stone volume growth/year was 15%, 71% and 177% for low, medium and high risk groups, respectively (p : We identified 80 individuals with stone growth measured by 3-dimensional CT (mean ~7 months between studies). Inter-reviewer reliability of CT volume measurement was well correlated (0.98, Gwet's AC2), and an arbitrator was only needed in 13/160 (8%) cases. Median stone volume growth/year was 15%, 71% and 177% for low, medium and high risk groups, respectively (p : We identified 80 individuals with stone growth measured by 3-dimensional CT (mean ~7 months between studies). Inter-reviewer reliability of CT volume measurement was well correlated (0.98, Gwet's AC2), and an arbitrator was only needed in 13/160 (8%) cases. Median stone volume growth/year was 15%, 71% and 177% for low, medium and high risk groups, respectively (p **CONCLUSIONS**: In a population of pure CaOx stone formers, increased 24-hour SS CaOx risk was associated with increased in vivo stone growth. Further investigations using CT volumetric stone growth may allow for the noninvasive study of stone growth modulators, improved stone risk prediction and development of a kidney stone simulator.

**URL:** <https://ovidsp.ovid.com/ovidweb.cgi?T=JS&CSC=Y&NEWS=N&PAGE=fulltext&D=med20&DO=10.1097%2fJU.0000000000001961> <https://libkey.io/libraries/2592/openurl?genre=article&sid=OVID:medline&id=pmid:34288713&id=doi:10.1097%2FJU.0000000000001961&issn=0022-5347&isbn=&volume=206&issue=6&spage=1438&pages=1438-1444&date=2021&title=Journal+of+Urology&atitle=24-Hour+Urine+Calcium+Oxalate+Supersaturation+Risk+Correlates+with+Computerized+Tomography+Volumetric+Calcium+Oxalate+Stone+Growth.&aulast=Yuzhakov&pid=%3Cauthor%3EYuzhakov+S%3BSteadman+SD%3BOtto+BJ%3BBird+VG%3BCanales+BK%3C%2Fauthor%3E%3CAN%3E34288713%3C%2FAN%3E%3CDT%3EJournal+Article%3C%2FDT%3E>

39. **Association Between Daily Water Intake and 24-hour Urine Volume Among Adolescents With Kidney Stones.**

**Item Type:** Journal Article

**Authors:** Bernard, Joshua;Song, Lihai;Henderson, Brittney and Tasian, Gregory E.

**Publication Date:** 2020

**Journal:** Urology 140, pp. 150–154

**Abstract: OBJECTIVE**: To determine the association between daily water intake and 24-hour urine volume among adolescents with nephrolithiasis in order to estimate a "fluid prescription," the additional water intake needed to increase urine volume to a target goal. **METHODS**: We conducted a secondary analysis of an ecological momentary assessment study that prospectively measured daily water intake of 25 adolescents with nephrolithiasis over 7 days. We identified 24-hour urine volumes obtained for clinical care within 12 months of water intake assessment. A linear regression model was fit to estimate the magnitude of the association between daily water intake and 24-hour urine volume, adjusting for age, sex, race, and daily temperature. **RESULTS**: Twenty-two participants completed fifty-seven 24-hour urine collections within 12 months of the study period. Median daily water intake was 1.4 L (IQR 0.67-1.94). Median 24-hour urine volume was 2.01 L (IQR 1.20-2.73). A 1 L increase in daily water intake was associated with a 710 mL increase in 24-hour urine output (95%CI 0.55-0.87). Using the model output, the equation was generated to estimate the additional fluid intake needed fluid prescription (FP) to produce the desired increase in urine output (dUOP): FP = dUOP/0.71. **CONCLUSION**: The FP equation (FP = dUOP)/0.71), which reflects the relationship between water intake and urine volume, could be used to help adolescents with nephrolithiasis achieve urine output goals to decrease stone recurrence. Copyright © 2020 Elsevier Inc. All rights reserved.

**URL:** <https://ovidsp.ovid.com/ovidweb.cgi?T=JS&CSC=Y&NEWS=N&PAGE=fulltext&D=med17&DO=10.1016%2fj.urology.2020.01.024> <https://libkey.io/libraries/2592/openurl?genre=article&sid=OVID:medline&id=pmid:32004558&id=doi:10.1016%2Fj.urology.2020.01.024&issn=0090-4295&isbn=&volume=140&issue=&spage=150&pages=150-154&date=2020&title=Urology&atitle=Association+Between+Daily+Water+Intake+and+24-hour+Urine+Volume+Among+Adolescents+With+Kidney+Stones.&aulast=Bernard&pid=%3Cauthor%3EBernard+J%3BSong+L%3BHenderson+B%3BTasian+GE%3C%2Fauthor%3E%3CAN%3E32004558%3C%2FAN%3E%3CDT%3EJournal+Article%3C%2FDT%3E>

40. **Exploring the Potential Relationship Between Phytate Ingestion, Urinary Phytate Excretion, and Renal Stone Risk in a Unique Human Model: No Hard Evidence in Support of Phytate as a Stone Inhibitor.**

**Item Type:** Journal Article

**Authors:** Fakier, Saajidah and Rodgers, Allen

**Publication Date:** 2020

**Journal:** Journal of Renal Nutrition 30(5), pp. 396–403

**Abstract: OBJECTIVE**: Dietary phytate (IP6) enjoys a reputation as an inhibitor of calcium renal stone formation, although there are very few human studies to support this notion. In South Africa, urolithiasis occurs in the white (W) but is rare in the black (B) population. We undertook this unique human model to further investigate the IP6 theory. **METHODS**: Healthy W and B males completed baseline food-frequency recall questionnaires. Dietary intake of IP6 was restricted for 18 days. An IP6 dietary supplement was ingested on days 15-18. Twenty-four-hour urinary phytate and other urinary components were determined. Relative supersaturations of calcium salts were calculated. The urinary metastable limit (MSL) of calcium oxalate (CaOx) and its crystallisation kinetics were determined experimentally. **RESULTS**: Habitual dietary intake of IP6 and its urinary excretion were significantly higher in B than in W (1650 +/- 202 vs. 640 +/- 134 mg/d, P = .0002 and 1.13 +/- 0.12 vs. 0.75 +/- 0.13 muM, P : Habitual dietary intake of IP6 and its urinary excretion were significantly higher in B than in W (1650 +/- 202 vs. 640 +/- 134 mg/d, P = .0002 and 1.13 +/- 0.12 vs. 0.75 +/- 0.13 muM, P **CONCLUSIONS**: Despite notable differences in the renal handling of ingested IP6, there were no changes in any of the well-established urinary risk factors for calcium renal stone formation in either of our uniquely different test groups. We conclude that, in the absence of hard evidence, claims that IP6 is a stone inhibitor remain unproven. Copyright © 2019 National Kidney Foundation, Inc. Published by Elsevier Inc. All rights reserved.

**URL:** <https://ovidsp.ovid.com/ovidweb.cgi?T=JS&CSC=Y&NEWS=N&PAGE=fulltext&D=med17&DO=10.1053%2fj.jrn.2019.10.006> <https://libkey.io/libraries/2592/openurl?genre=article&sid=OVID:medline&id=pmid:31928801&id=doi:10.1053%2Fj.jrn.2019.10.006&issn=1051-2276&isbn=&volume=30&issue=5&spage=396&pages=396-403&date=2020&title=Journal+of+Renal+Nutrition&atitle=Exploring+the+Potential+Relationship+Between+Phytate+Ingestion%2C+Urinary+Phytate+Excretion%2C+and+Renal+Stone+Risk+in+a+Unique+Human+Model%3A+No+Hard+Evidence+in+Support+of+Phytate+as+a+Stone+Inhibitor.&aulast=Fakier&pid=%3Cauthor%3EFakier+S%3BRodgers+A%3C%2Fauthor%3E%3CAN%3E31928801%3C%2FAN%3E%3CDT%3EJournal+Article%3C%2FDT%3E>

41. **Factors Associated With Compliance With 24-Hour Urine Collection.**

**Item Type:** Journal Article

**Authors:** Ghiraldi, Eric M.;Braitman, Leonard E. and Friedlander, Justin I.

**Publication Date:** 2020

**Journal:** Urology 142, pp. 65–69

**Abstract: OBJECTIVE**: To identify how demographic factors, stone-associated medical comorbidities, and treatment predict compliance with 24-hour urine collection. **MATERIALS AND METHODS**: A retrospective medical record review of patients treated for urolithiasis between August 2014 and March 2017 was performed. Patient demographics, medical characteristics, stone factors, type of treatment, and compliance data were included for patients requested to submit a collection. Variables that were statistically significant on bivariate analysis were then used to formulate a model predicting submission of a 24-hour urine sample. **RESULTS**: Of the 303 patients who met inclusion criteria, 183 (60.4%) submitted an initial 24-hour urine collection. On bivariate analysis, patients older than 50 were more likely to submit a 24-hour urine collection (71.4% vs 51.5%; P : Of the 303 patients who met inclusion criteria, 183 (60.4%) submitted an initial 24-hour urine collection. On bivariate analysis, patients older than 50 were more likely to submit a 24-hour urine collection (71.4% vs 51.5%; P : Of the 303 patients who met inclusion criteria, 183 (60.4%) submitted an initial 24-hour urine collection. On bivariate analysis, patients older than 50 were more likely to submit a 24-hour urine collection (71.4% vs 51.5%; P : Of the 303 patients who met inclusion criteria, 183 (60.4%) submitted an initial 24-hour urine collection. On bivariate analysis, patients older than 50 were more likely to submit a 24-hour urine collection (71.4% vs 51.5%; P **CONCLUSIONS**: We suspect that patients perceive surgery as a more definitive treatment for kidney stones than conservative management. Patient education on the natural history and role of metabolic management in the prevention of nephrolithiasis is essential in improving compliance with 24-hour urine collection. Copyright © 2020 Elsevier Inc. All rights reserved.

**URL:** <https://ovidsp.ovid.com/ovidweb.cgi?T=JS&CSC=Y&NEWS=N&PAGE=fulltext&D=med17&DO=10.1016%2fj.urology.2020.03.047> <https://libkey.io/libraries/2592/openurl?genre=article&sid=OVID:medline&id=pmid:32305538&id=doi:10.1016%2Fj.urology.2020.03.047&issn=0090-4295&isbn=&volume=142&issue=&spage=65&pages=65-69&date=2020&title=Urology&atitle=Factors+Associated+With+Compliance+With+24-Hour+Urine+Collection.&aulast=Ghiraldi&pid=%3Cauthor%3EGhiraldi+EM%3BBraitman+LE%3BFriedlander+JI%3C%2Fauthor%3E%3CAN%3E32305538%3C%2FAN%3E%3CDT%3EJournal+Article%3C%2FDT%3E>

42. **What Predicts Recurrent Kidney Stone after Parathyroidectomy in Patients with Primary Hyperparathyroidism?.**

**Item Type:** Journal Article

**Authors:** Islam, Ana K.;Holt, Shelby;Reisch, Joan;Nwariaku, Fiemu;Antonelli, Jodi and Maalouf, Naim M.

**Publication Date:** 2020

**Journal:** Journal of the American College of Surgeons 231(1), pp. 74–82

**Abstract: BACKGROUND**: Some, but not all, patients with primary hyperparathyroidism (PHPT) and kidney stone disease (KSD) are cured of their nephrolithiasis after parathyroidectomy. The goal of this study was to identify risk factors for recurrent KSD despite successful parathyroidectomy in known stone formers with PHPT. **STUDY DESIGN**: We conducted a single-center retrospective review of patients presenting to urology clinic with KSD between January 2008 and July 2018, who were diagnosed with concurrent PHPT, and underwent definitive parathyroidectomy. Laboratory testing for serum calcium, PTH (parathyroid hormone), phosphorus and 25-OH-vitamin D, and 24-hour urine studies for volume, pH, calcium, citrate, oxalate, uric acid, sodium, and creatinine was performed pre- and post-parathyroidectomy. Stone recurrence was determined on routine diagnostic imaging or by symptomatic KSD. **RESULTS**: Mean age at parathyroidectomy was 57 +/- 14 years. Pre-parathyroidectomy, mean serum calcium, 24-hour urine calcium, and PTH were 10.6 +/- 0.5 mg/dL, 378 +/- 209 mg/day, and 114 +/- 97 pg/mL, respectively. Twenty-six of 69 patients (38%) had multigland parathyroid disease. After parathyroidectomy, serum calcium and PTH levels normalized in 69 of 69 and 62 of 69 patients, respectively. However, 37 of 69 patients (54%) had persistent hypercalciuria postoperatively, and 16 of 69 (23%) had recurrent KSD, on average, 2.0 +/- 1.6 years after parathyroidectomy. Patients with recurrent KSD post-parathyroidectomy were significantly younger compared with patients without recurrent KSD (51 +/- 15 vs 60 +/- 13 years, p = 0.02). In a logistic regression model, younger age remains a strong predictive factor for recurrent KSD. **CONCLUSIONS**: Nearly one-quarter of PHPT patients with KSD who undergo successful parathyroidectomy present with recurrent KSD despite normalization of serum calcium, and more than half exhibit persistent calciuria. These patients were younger and may require closer monitoring for stone recurrence after successful parathyroidectomy. Further studies are needed to better identify the etiology of KSD post-parathyroidectomy. Copyright © 2020 American College of Surgeons. Published by Elsevier Inc. All rights reserved.

**URL:** <https://ovidsp.ovid.com/ovidweb.cgi?T=JS&CSC=Y&NEWS=N&PAGE=fulltext&D=med17&DO=10.1016%2fj.jamcollsurg.2020.04.015> <https://libkey.io/libraries/2592/openurl?genre=article&sid=OVID:medline&id=pmid:32330575&id=doi:10.1016%2Fj.jamcollsurg.2020.04.015&issn=1072-7515&isbn=&volume=231&issue=1&spage=74&pages=74-82&date=2020&title=Journal+of+the+American+College+of+Surgeons&atitle=What+Predicts+Recurrent+Kidney+Stone+after+Parathyroidectomy+in+Patients+with+Primary+Hyperparathyroidism%3F.&aulast=Islam&pid=%3Cauthor%3EIslam+AK%3BHolt+S%3BReisch+J%3BNwariaku+F%3BAntonelli+J%3BMaalouf+NM%3C%2Fauthor%3E%3CAN%3E32330575%3C%2FAN%3E%3CDT%3EJournal+Article%3C%2FDT%3E>

43. **Lower Socioeconomic Status is Associated With Adverse Urinary Markers and Surgical Complexity in Kidney Stone Patients.**

**Item Type:** Journal Article

**Authors:** Quarrier, Scott;Li, Shuang;Penniston, Kristina L.;Best, Sara L.;Hedican, Sean P.;Jhagroo, Roy A. and Nakada, Stephen Y.

**Publication Date:** 2020

**Journal:** Urology 146, pp. 67–71

**Abstract: OBJECTIVE**: To determine if socioeconomic status (SES) correlates with severity of kidney stone disease and 24-hour urine parameters. **MATERIALS AND METHODS**: An IRB approved prospectively maintained database for nephrolithiasis was retrospectively analyzed for both 24-hour urine results and surgical procedures performed from 2009 to 2019. Severely distressed communities (SDC) were categorized as those with a Distressed Communities Index (DCI), a composite measure of SES, score in the top quartile (lowest for SES). Univariate and multivariate analyses were performed to evaluate the strength of the association of DCI on: stone size at presentation, need for and type of surgical procedure, need for staged surgery and specific stone risk factors in 24-hour urine collections. **RESULTS**: Surgical procedures were performed on 3939 patients (1978 women) who were not from SDC and 200 (97 women) from SDC. Patients from SDC were older (57.1 years vs 54.2 years; P = .009). Patients from SDC were more likely to undergo proportionally more invasive procedures (17.5% vs 11.6%; P = .011) and require staged surgery at a higher rate (13.0% vs 8.5%; P = .028). Men from SDC had larger stones (12.5 mm vs 9.7 mm; P = .001). Among 24-hour urine results from 2454 patients (1187 women), DCI was not correlated with sodium, calcium, magnesium, volume, oxalate, phosphate, and pH levels. Higher DCI (lower SES) correlated with lower urine citrate (P = .001) and lower urine potassium (P = .002). **CONCLUSION**: SES correlates with larger stone burden at the time of urologic intervention, requires proportionally more invasive procedures and more staged procedures. Lower SES correlated with lower urine citrate and potassium. Copyright © 2020 Elsevier Inc. All rights reserved.

**URL:** <https://ovidsp.ovid.com/ovidweb.cgi?T=JS&CSC=Y&NEWS=N&PAGE=fulltext&D=med18&DO=10.1016%2fj.urology.2020.09.025> <https://libkey.io/libraries/2592/openurl?genre=article&sid=OVID:medline&id=pmid:32991913&id=doi:10.1016%2Fj.urology.2020.09.025&issn=0090-4295&isbn=&volume=146&issue=&spage=67&pages=67-71&date=2020&title=Urology&atitle=Lower+Socioeconomic+Status+is+Associated+With+Adverse+Urinary+Markers+and+Surgical+Complexity+in+Kidney+Stone+Patients.&aulast=Quarrier&pid=%3Cauthor%3EQuarrier+S%3BLi+S%3BPenniston+KL%3BBest+SL%3BHedican+SP%3BJhagroo+RA%3BNakada+SY%3C%2Fauthor%3E%3CAN%3E32991913%3C%2FAN%3E%3CDT%3EJournal+Article%3C%2FDT%3E>

44. **A Prospective, Double-Blind, Randomized, Placebo-Controlled, Crossover Study Using an Orally Administered Oxalate Decarboxylase (OxDC).**

**Item Type:** Journal Article

**Authors:** Quintero, Emily;Bird, Victoria Yvonne;Liu, Howard;Stevens, Gary;Ryan, Alan S.;Buzzerd, Sabrina and Klimberg, Ira W.

**Publication Date:** 2020

**Journal:** Kidney360 1(11), pp. 1284; 11 25–1290

**Abstract: Background**: Hyperoxaluria is typically associated with excessive oxalate intake in the diet, decreased dietary calcium, hyperabsorption of oxalate, or increased endogenous production of oxalate. The disorder spectrum extends from recurrent kidney stones to ESKD. This clinical trial sought to evaluate the effectiveness of an acid stable oxalate decarboxylase (OxDC) to reduce urinary oxalate in healthy subjects on a high-oxalate diet. **Methods**: In this prospective, double-blind, randomized, placebo-controlled, crossover clinical trial, 33 healthy volunteers were randomized into two crossover sequences separated by a 2-day washout period. A controlled high-oxalate diet (750-800 mg oxalate, 500-550 mg calcium daily) was utilized, and six 24-hour urine collections were measured. Subjects were given approximately 1000 U (micromoles per minute per milligram) of OxDC or placebo with meals three times daily during the 4 days of treatment. **Results**: Urinary oxalate significantly decreased with OxDC treatment. The baseline corrected within-subject mean reduction in 24-hour urinary excretion (after OxDC dosing versus high-oxalate baseline preceding treatment) was 12.5 mg or 29% (P5% reduction) in 31 of 33 subjects (94%). Compared with placebo, OxDC produced a 24% reduction (P5% reduction) in 31 of 33 subjects (94%). Compared with placebo, OxDC produced a 24% reduction (P**Conclusions**: An orally administered OxDC is capable of significantly reducing urinary oxalate levels in healthy volunteers on a high-oxalate diet without affecting creatinine clearance, urine creatinine, or other solutes related to supersaturation of calcium oxalate. **Clinical Trial registry name and registration number**: Evaluation of Nephure, and the Reduction of Dietary Oxalate, in Healthy Volunteers, NCT03661216. Copyright © 2020 by the American Society of Nephrology.

**URL:** <https://ovidsp.ovid.com/ovidweb.cgi?T=JS&CSC=Y&NEWS=N&PAGE=fulltext&D=med18&DO=10.34067%2fKID.0001522020> <https://libkey.io/libraries/2592/openurl?genre=article&sid=OVID:medline&id=pmid:35372879&id=doi:10.34067%2FKID.0001522020&issn=2641-7650&isbn=&volume=1&issue=11&spage=1284&pages=1284-1290&date=2020&title=Kidney360&atitle=A+Prospective%2C+Double-Blind%2C+Randomized%2C+Placebo-Controlled%2C+Crossover+Study+Using+an+Orally+Administered+Oxalate+Decarboxylase+%28OxDC%29.&aulast=Quintero&pid=%3Cauthor%3EQuintero+E%3BBird+VY%3BLiu+H%3BStevens+G%3BRyan+AS%3BBuzzerd+S%3BKlimberg+IW%3C%2Fauthor%3E%3CAN%3E35372879%3C%2FAN%3E%3CDT%3EJournal+Article%3C%2FDT%3E>

45. **Metabolic evaluation of patients with recurrent, multiple or bilateral renal stones in Jaffna.**

**Item Type:** Journal Article

**Authors:** Rajendra, Sittampalam R.

**Publication Date:** Sep 30 ,2020

**Journal:** Ceylon Medical Journal 65(3), pp. 56–61

**Abstract: INTRODUCTION**: A comprehensive evaluation to identify metabolic abnormalities will help design management strategies for prevention of renal stone recurrences. The objective of this study is to identify the metabolic risk factors in a series of patients with recurrent, multiple or bilateral renal stone disease from the Northern Province, Sri Lanka. **METHODS**: This is a hospital based observational study on patients with recurrent, multiple or bilateral renal stones. Metabolic evaluation workout included, fasting non-tourniquet venous blood sample for serum ionized calcium, creatinine, and uric acid, freshly voided early morning urine sample for pH and laboratory urinalysis, and collection of two sets of 24 hour urine samples on two separate days, for phosphorous, creatinine, oxalate, uric acid, magnesium, calcium and citrate. **RESULTS**: The mean values of each metabolic parameter measured on the two sets of 24 hour urine samples, were found to be within the normal reference range for the entire 30 patient study group taken as whole, although the mean values for 24 hour urine magnesium (79.79+/-51.36 mg) and urine calcium (125.02+/-73.32 mg) were found to be towards the lower limit of normal reference range and the 24 hour urine oxalate (0.38+/-0.22 mmol) was found to be towards the upper limit of normal reference range. However, 80% of patients individually showed abnormal metabolic results either as single derangement (13 patients, 43.3%) or in multiple combinations of derangements (11 patients, 36.7%). Six patients (20%) did not have any metabolic abnormalities. Hyperoxaluria, hyperuricosuria, hypomagnesuria and hypocitraturia were the metabolic abnormalities identified among the study group. **CONCLUSION**: A comprehensive metabolic evaluation of each patient with recurrent, multiple or bilateral renal stone disease can help identify metabolic abnormalities, either as a single abnormality or combinations of abnormalities, specific for them. Such patient profiling will help develop bespoke management strategy based on appropriate dietary advice and specific medical treatment towards prevention of further stone formation.

**URL:** <https://ovidsp.ovid.com/ovidweb.cgi?T=JS&CSC=Y&NEWS=N&PAGE=fulltext&D=med18&DO=10.4038%2fcmj.v65i3.9186> <https://libkey.io/libraries/2592/openurl?genre=article&sid=OVID:medline&id=pmid:34800932&id=doi:10.4038%2Fcmj.v65i3.9186&issn=0009-0875&isbn=&volume=65&issue=3&spage=56&pages=56-61&date=2020&title=Ceylon+Medical+Journal&atitle=Metabolic+evaluation+of+patients+with+recurrent%2C+multiple+or+bilateral+renal+stones+in+Jaffna.&aulast=Rajendra&pid=%3Cauthor%3ERajendra+S+R%3C%2Fauthor%3E%3CAN%3E34800932%3C%2FAN%3E%3CDT%3EJournal+Article%3C%2FDT%3E>

46. **Extreme diet without calcium may lead to hyperoxaluria and kidney stone recurrence-A case study**

**Item Type:** Report

**Authors:** Salek, T.

**Publication Date:** 2020

**Accessed:**
**Abstract: BACKGROUND**: The aim of the study was to present a case study of a 56-year-old woman with hyperoxaluria induced by calcium-free diet that resulted in kidney stone recurrence. **METHODS**: A 24-hour urine collection and serum tests for kidney stone risk factors identification were performed. The monitoring of urine risk factors was done by untimed urine samples and 24-hour urine collections. Polarized light microscopy was performed for kidney stone analysis. **RESULTS**: The results of urine collection showed hyperoxaluria of 0.551 mmoL per 24 hours. After adding calcium-containing products to the diet the oxaluria decreased to reference range value of 0.352 mmoL/24 hours and all untimed oxalate to creatinine ratios returned to reference ranges. Polarized light microscopy revealed 100% calcium oxalate kidney stone composition (It was 50% Weddellite and 50% Whewellite). **CONCLUSIONS**: The case study shows the importance of calcium intake in the prevention of calcium oxalate kidney stone recurrence. Particularly, unsuitable diet without calcium can induce kidney stone recurrence. Knowledge of diet habits is important for interpretation of kidney stone risk factors and their inhibitors excreted in urine. Copyright © 2020 The Authors. Journal of Clinical Laboratory Analysis published by Wiley Periodicals LLC.

**URL:** <https://ovidsp.ovid.com/ovidweb.cgi?T=JS&CSC=Y&NEWS=N&PAGE=fulltext&D=med18&DO=10.1002%2fjcla.23512> <https://libkey.io/libraries/2592/openurl?genre=article&sid=OVID:medline&id=pmid:32761639&id=doi:10.1002%2Fjcla.23512&issn=0887-8013&isbn=&volume=34&issue=12&spage=e23512&pages=e23512&date=2020&title=Journal+of+Clinical+Laboratory+Analysis&atitle=Extreme+diet+without+calcium+may+lead+to+hyperoxaluria+and+kidney+stone+recurrence-A+case+study.&aulast=Salek&pid=%3Cauthor%3ESalek+T%3C%2Fauthor%3E%3CAN%3E32761639%3C%2FAN%3E%3CDT%3ECase+Reports%3C%2FDT%3E>

47. **The Association Between 24-Hour Urine and Stone Recurrence Among High Risk Kidney Stone Formers: A Population Level Assessment.**

**Item Type:** Journal Article

**Authors:** Samson, Patrick C.;Holt, Sarah K.;Hsi, Ryan S.;Sorensen, Mathew D. and Harper, Jonathan D.

**Publication Date:** 2020

**Journal:** Urology 144, pp. 71–76

**Abstract: OBJECTIVE**: To determine if obtaining a 24-hour urine collection (24HU) in stone formers is associated with decreased recurrent stone episodes. **METHODS**: Using the MarketScan database, adults 17-62 years old with nephrolithiasis were identified between 2007 and 2017 with a minimum of 3-year follow up. High-risk stone formers, those undergoing stone surgery, and those with history of recurrent stones were identified. The exposure was a 24HU within 6 months of primary diagnosis. The outcome was recurrent stone episodes-defined by stone-related emergency room visits, hospitalizations, or stone surgery 90 days to 3 years after diagnosis. Logistic regression was used to estimate recurrence risk by 24HU exposure for the overall cohort and sub-cohorts limited to known recurrent stone formers, high-risk subjects, and those having stone surgery. **RESULTS**: Of 434,055 subjects analyzed, 30,153 (6.9%) had a 24HU. An annual decline in 24HU utilization was seen (7.5%-5.8%). Regional variation in usage rate was also observed. On multivariate analysis, completing a 24HU was not associated with risk of recurrence in any of the following cohorts: recurrent stone formers (OR 0.98, 95% CI 0.9-1.07), both high risk and recurrent stone formers (OR 0.95 [0.8-1.13]), those undergoing surgery (OR 1.02 [0.97-1.07]); a positive association with 24HU and recurrence was seen in those labeled high-risk (OR 1.08 [1.01-1.16]) and in all-comers (OR 1.15 [1.12-1.19]). **CONCLUSION**: The 24HU was not associated with decreased recurrence rates in the overall population nor in higher risk sub-cohorts. Copyright © 2020 Elsevier Inc. All rights reserved.

**URL:** <https://ovidsp.ovid.com/ovidweb.cgi?T=JS&CSC=Y&NEWS=N&PAGE=fulltext&D=med18&DO=10.1016%2fj.urology.2020.05.064> <https://libkey.io/libraries/2592/openurl?genre=article&sid=OVID:medline&id=pmid:32540303&id=doi:10.1016%2Fj.urology.2020.05.064&issn=0090-4295&isbn=&volume=144&issue=&spage=71&pages=71-76&date=2020&title=Urology&atitle=The+Association+Between+24-Hour+Urine+and+Stone+Recurrence+Among+High+Risk+Kidney+Stone+Formers%3A+A+Population+Level+Assessment.&aulast=Samson&pid=%3Cauthor%3ESamson+PC%3BHolt+SK%3BHsi+RS%3BSorensen+MD%3BHarper+JD%3C%2Fauthor%3E%3CAN%3E32540303%3C%2FAN%3E%3CDT%3EJournal+Article%3C%2FDT%3E>

48. **Hypomagnesuria is Associated With Nephrolithiasis in Patients With Asymptomatic Primary Hyperparathyroidism.**

**Item Type:** Journal Article

**Authors:** Saponaro, Federica;Marcocci, Claudio;Apicella, Matteo;Mazoni, Laura;Borsari, Simona;Pardi, Elena;Di Giulio, Marina;Carlucci, Francesco;Scalese, Marco;Bilezikian, John P. and Cetani, Filomena

**Publication Date:** 2020

**Journal:** Journal of Clinical Endocrinology & Metabolism 105(8), pp. 08 01

**Abstract: CONTEXT**: The pathogenesis of nephrolithiasis in primary hyperparathyroidism (PHPT) remains to be elucidated. The latest guidelines suggest parathyroidectomy in patients with asymptomatic PHPT with hypercalciuria (> 400 mg/d) and increased stone risk profile. **OBJECTIVE**: The objective of this work is to evaluate the association of urinary stone risk factors and nephrolithiasis in patients with asymptomatic sporadic PHPT and its clinical relevance. **DESIGN**: A total of 157 consecutive patients with sporadic asymptomatic PHPT were evaluated by measurement of serum and 24-hour urinary parameters and kidney ultrasound. **RESULTS**: Urinary parameters were tested in the univariate analysis as continuous and categorical variables. Only hypercalciuria and hypomagnesuria were significantly associated with nephrolithiasis in the univariate and multivariate analysis adjusted for age, sex, body mass index, estimated glomerular filtration rate, parathyroid hormone, 25-hydroxyvitamin D, serum calcium, and urine volume (odds ratio, OR 2.14 [1.10-4.56]; P = .04; OR 3.06 [1.26-7.43]; P = .013, respectively). Hypomagnesuria remained associated with nephrolithiasis in the multivariate analysis (OR 6.09 [1.57-23.5], P = .009) even when the analysis was limited to patients without concomitant hypercalciuria. The urinary calcium/magnesium (Ca/Mg) ratio was also associated with nephrolithiasis (univariate OR 1.62 [1.27-2.08]; P = .001 and multivariate analysis OR 1.74 [1.25-2.42], P = .001). Hypomagnesuria and urinary Ca/Mg ratio had a better, but rather low, positive predictive value compared with hypercalciuria. **CONCLUSIONS**: Hypomagnesuria and urinary Ca/Mg ratio are each associated with silent nephrolithiasis and have potential clinical utility as risk factors, besides hypercalciuria, for kidney stones in asymptomatic PHPT patients. The other urinary indices that have been commonly thought to be associated with kidney stones in PHPT are not supported by our results. Copyright © Endocrine Society 2020. All rights reserved. For permissions, please e-mail: [](https://refworks.proquest.com/).

**URL:** <https://ovidsp.ovid.com/ovidweb.cgi?T=JS&CSC=Y&NEWS=N&PAGE=fulltext&D=med18&DO=10.1210%2fclinem%2fdgaa233> <https://libkey.io/libraries/2592/openurl?genre=article&sid=OVID:medline&id=pmid:32369583&id=doi:10.1210%2Fclinem%2Fdgaa233&issn=0021-972X&isbn=&volume=105&issue=8&spage=e2789&pages=&date=2020&title=Journal+of+Clinical+Endocrinology+%26+Metabolism&atitle=Hypomagnesuria+is+Associated+With+Nephrolithiasis+in+Patients+With+Asymptomatic+Primary+Hyperparathyroidism.&aulast=Saponaro&pid=%3Cauthor%3ESaponaro+F%3BMarcocci+C%3BApicella+M%3BMazoni+L%3BBorsari+S%3BPardi+E%3BDi+Giulio+M%3BCarlucci+F%3BScalese+M%3BBilezikian+JP%3BCetani+F%3C%2Fauthor%3E%3CAN%3E32369583%3C%2FAN%3E%3CDT%3EJournal+Article%3C%2FDT%3E>

49. **Association of Chronic Kidney Disease Stage with 24-Hour Urine Values Among Patients with Nephrolithiasis.**

**Item Type:** Journal Article

**Authors:** Sui, Wilson;Calvert, Josh K.;Kavoussi, Nicholas L.;Gould, Edward R.;Miller, Nicole L.;Bejan, Cosmin A. and Hsi, Ryan S.

**Publication Date:** 2020

**Journal:** Journal of Endourology 34(12), pp. 1263–1271

**Abstract: Introduction:** Nephrolithiasis is a known risk factor for chronic kidney disease (CKD); however, it is unknown how CKD affects urinary parameters related to stone risk. The purpose of this study was to assess the relationship of diminishing glomerular filtration rate (GFR) and kidney stone-related 24-hour urine (24H urine) composition. **Materials and Methods:** A single-institution retrospective review of patients (n = 2057) who underwent 24H urine analysis was performed. The serum creatinine within 1 year of the first 24H urine was used to determine estimated GFR and stratify patients by CKD stage. We performed analysis of variance and multivariable linear regression to assess the relationship of GFR and urinary analytes. **Results:** Among all patients, there were 184 (8.9%), 1537 (74.7%), 245 (11.9%), 70 (3.4%), 17 (0.8%), and 4 (0.2%) in CKD stage I, II, IIIa, IIIb, IV, and V groups, respectively. On analysis of 24H urine composition, as CKD increased, changes in urinary parameters protective against crystallization included decreased calcium and uric acid (UA) (P Among all patients, there were 184 (8.9%), 1537 (74.7%), 245 (11.9%), 70 (3.4%), 17 (0.8%), and 4 (0.2%) in CKD stage I, II, IIIa, IIIb, IV, and V groups, respectively. On analysis of 24H urine composition, as CKD increased, changes in urinary parameters protective against crystallization included decreased calcium and uric acid (UA) (P Among all patients, there were 184 (8.9%), 1537 (74.7%), 245 (11.9%), 70 (3.4%), 17 (0.8%), and 4 (0.2%) in CKD stage I, II, IIIa, IIIb, IV, and V groups, respectively. On analysis of 24H urine composition, as CKD increased, changes in urinary parameters protective against crystallization included decreased calcium and uric acid (UA) (P Among all patients, there were 184 (8.9%), 1537 (74.7%), 245 (11.9%), 70 (3.4%), 17 (0.8%), and 4 (0.2%) in CKD stage I, II, IIIa, IIIb, IV, and V groups, respectively. On analysis of 24H urine composition, as CKD increased, changes in urinary parameters protective against crystallization included decreased calcium and uric acid (UA) (P **Conclusions:** Higher CKD stage was associated with changes in urinary analytes that both promoted and inhibited stone formation, with the net effect of decreasing calcium oxalate and phosphate supersaturation. These patients may benefit from medical therapy that targets improving urinary citrate instead of lowering calcium or UA.

**URL:** <https://ovidsp.ovid.com/ovidweb.cgi?T=JS&CSC=Y&NEWS=N&PAGE=fulltext&D=med18&DO=10.1089%2fend.2020.0403> <https://libkey.io/libraries/2592/openurl?genre=article&sid=OVID:medline&id=pmid:32578450&id=doi:10.1089%2Fend.2020.0403&issn=0892-7790&isbn=&volume=34&issue=12&spage=1263&pages=1263-1271&date=2020&title=Journal+of+Endourology&atitle=Association+of+Chronic+Kidney+Disease+Stage+with+24-Hour+Urine+Values+Among+Patients+with+Nephrolithiasis.&aulast=Sui&pid=%3Cauthor%3ESui+W%3BCalvert+JK%3BKavoussi+NL%3BGould+ER%3BMiller+NL%3BBejan+CA%3BHsi+RS%3C%2Fauthor%3E%3CAN%3E32578450%3C%2FAN%3E%3CDT%3EJournal+Article%3C%2FDT%3E>

50. **Nephrolithiasis and Elevated Urinary Ammonium: A Matched Comparative Study.**

**Item Type:** Journal Article

**Authors:** Sui, Wilson;Hancock, Joel;Asplin, John R.;Gould, Edward R. and Hsi, Ryan S.

**Publication Date:** 2020

**Journal:** Urology 144, pp. 77–82

**Abstract: OBJECTIVE**: To describe the associations between elevated urinary ammonium and clinical characteristics of kidney stone formers. A 24-hour urine test is recommended in high-risk patients to identify urinary abnormalities and select interventions to reduce the recurrence risk. While elevations in urine ammonium may be seen in acidosis, diarrhea, high protein diets or due to pathogenic bacteria, the clinical characteristics of these patients have not been previously described. **METHODS**: We retrospectively identified adult patients with kidney stone disease who completed a 24-hour urine at our institution between 2006 and 2017. Patients with elevated urinary ammonium were identified (n = 121) and matched 1:1 by age and sex to controls for an overall cohort of n = 242. Differences in medical and surgical history, 24-hour urine analytes and stone composition were compared. **RESULTS**: Among 3625 24-hour urine studies screened, 7.1% of patients showed high urinary ammonium. In our study cohort, patients with elevated urinary ammonium also showed higher urine volume, oxalate, calcium, uric acid, sodium, chloride, and sulphate. Clinically, these patients had higher body mass index, and more often had a history of recurrent urinary tract infections, diabetes, gout, bowel resection, and urinary reconstruction history. Struvite stones tended to be more common in the elevated ammonium group vs control (n = 7 vs 1, P = .07). **CONCLUSION**: Elevated urinary ammonium among kidney stone patients is relatively uncommon. However, these patients have higher rates of comorbid metabolic conditions, urinary tract infections, and bowel surgery. This finding should prompt further review of the patient's history and may help direct prevention strategies. Copyright © 2020 Elsevier Inc. All rights reserved.

**URL:** <https://ovidsp.ovid.com/ovidweb.cgi?T=JS&CSC=Y&NEWS=N&PAGE=fulltext&D=med18&DO=10.1016%2fj.urology.2020.05.063> <https://libkey.io/libraries/2592/openurl?genre=article&sid=OVID:medline&id=pmid:32544550&id=doi:10.1016%2Fj.urology.2020.05.063&issn=0090-4295&isbn=&volume=144&issue=&spage=77&pages=77-82&date=2020&title=Urology&atitle=Nephrolithiasis+and+Elevated+Urinary+Ammonium%3A+A+Matched+Comparative+Study.&aulast=Sui&pid=%3Cauthor%3ESui+W%3BHancock+J%3BAsplin+JR%3BGould+ER%3BHsi+RS%3C%2Fauthor%3E%3CAN%3E32544550%3C%2FAN%3E%3CDT%3EJournal+Article%3C%2FDT%3E>

51. **Development and Validation of a New Gas Chromatography-Tandem Mass Spectrometry Method for the Measurement of Enrichment of Glyoxylate Metabolism Analytes in Hyperoxaluria Patients Using a Stable Isotope Procedure.**

**Item Type:** Journal Article

**Authors:** van Harskamp, Dewi;Garrelfs, Sander F.;Oosterveld, Michiel J. S.;Groothoff, Jaap W.;van Goudoever, Johannes B. and Schierbeek, Henk

**Publication Date:** 2020

**Journal:** Analytical Chemistry 92(2), pp. 1826–1832

**Abstract:** Primary hyperoxalurias (PH) are inborn errors of glyoxylate metabolism characterized by an increase in endogenous oxalate production. Oxalate overproduction may cause calcium-oxalate crystal formation leading to kidney stones, nephrocalcinosis, and ultimately kidney failure. Twenty-four hour urine oxalate excretion is an inaccurate measure for endogenous oxalate production in PH patients and not applicable in those with kidney failure. Treatment efficacy cannot be assessed with this measure during clinical trials. We describe the development and validation of a gas chromatography-tandem mass spectrometry method to analyze the samples obtained following a stable isotope infusion protocol of ^13^C_2_-oxalate and 1-^13^C-glycolate in both healthy individuals and PH patients. Isotopic enrichments of plasma oxalate, glycolate, and glyoxylate were measured on a gas chromatography-triple quadrupole mass spectrometry system using ethylhydroxylamine and N-tert-butyldimethylsilyl-N-methyltrifluoroacetamide (MTBSTFA) for analyte derivatization. Method precision was good for oxalate and glycolate (coefficients of variation [CV] were <6.3% and <4.2% for inter- and intraday precision, respectively) and acceptable for glyoxylate (CV <18.3% and <6.7% for inter- and intraday precision, respectively). The enrichment curves were linear over the specified range. Sensitivity was sufficient to accurately analyze enrichments. This new method allowed calculation of kinetic features of these metabolites, thus enabling a detailed analysis of the various pathways involved in glyoxylate metabolism. The method will further enhance the investigation of the metabolic PH derangements, provides a tool to accurately assess the therapeutic efficacy of new promising therapeutic interventions for PH, and could serve as a clinical tool to improve personalized therapeutic strategies.

**URL:** <https://ovidsp.ovid.com/ovidweb.cgi?T=JS&CSC=Y&NEWS=N&PAGE=fulltext&D=med17&DO=10.1021%2facs.analchem.9b03670> <https://libkey.io/libraries/2592/openurl?genre=article&sid=OVID:medline&id=pmid:31867958&id=doi:10.1021%2Facs.analchem.9b03670&issn=0003-2700&isbn=&volume=92&issue=2&spage=1826&pages=1826-1832&date=2020&title=Analytical+Chemistry&atitle=Development+and+Validation+of+a+New+Gas+Chromatography-Tandem+Mass+Spectrometry+Method+for+the+Measurement+of+Enrichment+of+Glyoxylate+Metabolism+Analytes+in+Hyperoxaluria+Patients+Using+a+Stable+Isotope+Procedure.&aulast=van+Harskamp&pid=%3Cauthor%3Evan+Harskamp+D%3BGarrelfs+SF%3BOosterveld+MJS%3BGroothoff+JW%3Bvan+Goudoever+JB%3BSchierbeek+H%3C%2Fauthor%3E%3CAN%3E31867958%3C%2FAN%3E%3CDT%3EJournal+Article%3C%2FDT%3E>

52. **Rising occurrence of hypocitraturia and hyperoxaluria associated with increasing prevalence of stone disease in calcium kidney stone formers.**

**Item Type:** Journal Article

**Authors:** Youssef, Ramy F.;Martin, Jeremy W.;Sakhaee, Khashayar;Poindexter, John;Dianatnejad, Sharmin;Scales, Charles D.;Preminger, Glenn M. and Lipkin, Michael E.

**Publication Date:** Oc ,2020

**Journal:** Scandinavian Journal of Urology 54(5), pp. 426–430

**Abstract: OBJECTIVE**: To evaluate metabolic risk factors in calcium kidney stone formers from two different decades, comparing changes in metabolic profiles over time. **METHODS**: A retrospective analysis was performed of calcium kidney stone formers who underwent metabolic evaluation of urolithiasis with 24-hour urine collections at a single institution. There were 309 patients evaluated from 1988 to 1994 (Group A), and 229 patients from 2007 to 2010 (Group B). A comparison between both groups was performed to assess changes in demographics and in metabolic stone profiles. **RESULTS**: Comparing Group A to Group B, the percentage of females increased from 43 to 56%, obese patients (BMI >= 30) increased from 22 to 35%, and patients >= 50 years increased from 29 to 47% (all p : Comparing Group A to Group B, the percentage of females increased from 43 to 56%, obese patients (BMI >= 30) increased from 22 to 35%, and patients >= 50 years increased from 29 to 47% (all p **CONCLUSIONS**: Urolithiasis has increased in females, obese, and older patients, consistent with population-based studies. We report a rising incidence of hypocitraturia and hyperoxaluria in the contemporary cohort, particularly in obese patients and in males, respectively. Further studies are needed to better characterize the metabolic changes corresponding to the increase in stone disease.

**URL:** <https://ovidsp.ovid.com/ovidweb.cgi?T=JS&CSC=Y&NEWS=N&PAGE=fulltext&D=med18&DO=10.1080%2f21681805.2020.1794955> <https://libkey.io/libraries/2592/openurl?genre=article&sid=OVID:medline&id=pmid:32715836&id=doi:10.1080%2F21681805.2020.1794955&issn=2168-1805&isbn=&volume=54&issue=5&spage=426&pages=426-430&date=2020&title=Scandinavian+Journal+of+Urology&atitle=Rising+occurrence+of+hypocitraturia+and+hyperoxaluria+associated+with+increasing+prevalence+of+stone+disease+in+calcium+kidney+stone+formers.&aulast=Youssef&pid=%3Cauthor%3EYoussef+RF%3BMartin+JW%3BSakhaee+K%3BPoindexter+J%3BDianatnejad+S%3BScales+CD%3BPreminger+GM%3BLipkin+ME%3C%2Fauthor%3E%3CAN%3E32715836%3C%2FAN%3E%3CDT%3EJournal+Article%3C%2FDT%3E>

53. **Long-term Recurrence Rates in Uric Acid Stone Formers With or Without Medical Management.**

**Item Type:** Journal Article

**Authors:** Canales, Benjamin K.;Sharma, Nitin;Yuzhakov, Stanislav V.;Bozorgmehri, Shahab;Otto, Brandon J. and Bird, Vincent G.

**Publication Date:** Se ,2019

**Journal:** Urology 131, pp. 46–52

**Abstract: OBJECTIVE**: To determine if medical therapy affects long-term clinical outcomes in uric acid stone formers (UASF). **METHODS**: We identified 53 UASF who had complete stone clearance following stone procedure by computed tomography (CT) and had >=1 postoperative 24-hour urine collection and a clinical follow-up >=6 months with a surveillance CT scan. Patients were divided into "adherent to medical therapy" (compliance with potassium citrate +/- allopurinol verified by computerized pharmacy data) or nonadherent groups. Primary outcomes were CT stone recurrence rate and need for surgical stone intervention. **RESULTS**: We found 28 of 53 (53%) adherent and 25 of 53 (47%) nonadherent individuals (14 declined medication, 11 intolerant). With median follow-up of 24 months, no significant differences were noted between groups in regards to stone recurrence (32%; P=.99) or in 24-hour urine pH compared to baseline or follow-up (range 5.46-5.62; P=0.06). Adherent patients, however, had smaller CT stone recurrence sizes (6.3 +/- 3.8 vs 11.8 +/- 6.2 mm, P=.02), were 28% less likely to require stone surgery compared to those without therapy (P : We found 28 of 53 (53%) adherent and 25 of 53 (47%) nonadherent individuals (14 declined medication, 11 intolerant). With median follow-up of 24 months, no significant differences were noted between groups in regards to stone recurrence (32%; P=.99) or in 24-hour urine pH compared to baseline or follow-up (range 5.46-5.62; P=0.06). Adherent patients, however, had smaller CT stone recurrence sizes (6.3 +/- 3.8 vs 11.8 +/- 6.2 mm, P=.02), were 28% less likely to require stone surgery compared to those without therapy (P **CONCLUSION**: UASF adherent to medical therapy had smaller recurrence sizes and fewer surgical interventions vs nonadherent, highlighting the protective role of potassium citrate in UA stone disease. The comparable urine pH and stone recurrence rates between groups, however, underscore areas for improvement in future UA stone prevention strategies. Copyright © 2019 Elsevier Inc. All rights reserved.

**URL:** <https://ovidsp.ovid.com/ovidweb.cgi?T=JS&CSC=Y&NEWS=N&PAGE=fulltext&D=med16&DO=10.1016%2fj.urology.2019.05.023> <https://libkey.io/libraries/2592/openurl?genre=article&sid=OVID:medline&id=pmid:31158354&id=doi:10.1016%2Fj.urology.2019.05.023&issn=0090-4295&isbn=&volume=131&issue=&spage=46&pages=46-52&date=2019&title=Urology&atitle=Long-term+Recurrence+Rates+in+Uric+Acid+Stone+Formers+With+or+Without+Medical+Management.&aulast=Canales&pid=%3Cauthor%3ECanales+BK%3BSharma+N%3BYuzhakov+SV%3BBozorgmehri+S%3BOtto+BJ%3BBird+VG%3C%2Fauthor%3E%3CAN%3E31158354%3C%2FAN%3E%3CDT%3EJournal+Article%3C%2FDT%3E>

54. **Initial collection of an inadequate 24-hour urine sample in children does not predict subsequent inadequate collections.**

**Item Type:** Journal Article

**Authors:** Chan, K. H.;Moser, E. A.;Whittam, B. M.;Misseri, R.;Cain, M. P.;Krambeck, A. and Schwaderer, A.

**Publication Date:** Fe ,2019

**Journal:** Journal of Pediatric Urology 15(1), pp. 74.e1–74.e7

**Abstract: INTRODUCTION**: Approximately half of adult stone formers submit specimens that are either under or over collections as determined by 24-h creatinine/kg. Previously identified predictors of inadequate collection in adults include female sex, older age, higher body mass index (BMI), vitamin D supplementation, and weekday collection. **OBJECTIVE**: The objective of this study is to determine risk factors for inadequate 24-h urinary specimen collection in the pediatric population. **STUDY DESIGN**: A retrospective analysis of all children (: A retrospective analysis of all children (**RESULTS**: Of 367 patients, 80 had an adequate collection (21.9%): median age, 13 years (interquartile range, 8-16); 61.1% female; 93.5% white; 19.5% obese; and 13.0% overweight. No parental or child factors were associated with inadequate collection (Summary Table). Of inadequate collections, more than 80% were over collections. In the 175 patients with more than one 24-h urinary specimen collection, the effect of an initial inadequate collection on subsequent inadequate collections was not significant after controlling for the correlation of samples from the same patient (p = 0.8). **DISCUSSION**: Any parental or child factors associated with the collection of inadequate 24-h urine specimens in children were not found. An initial inadequate collection does not predict subsequent inadequate collections. It was surprising that >80% of the inadequate collections were over collections rather than under collections. Possible explanations are that children collected urine samples for longer than the 24-h period or that stone-forming children produce more creatinine per 24-h period than healthy children due to hyperfiltration. **CONCLUSION**: Inadequate collections are very common, and the risk factors for them are unclear. A repeat collection would be suggested if the first is inadequate. Further studies must be planned to explore barriers to accurate specimen collection using qualitative research methodology. Copyright © 2018 Journal of Pediatric Urology Company. Published by Elsevier Ltd. All rights reserved.

**URL:** <https://ovidsp.ovid.com/ovidweb.cgi?T=JS&CSC=Y&NEWS=N&PAGE=fulltext&D=med16&DO=10.1016%2fj.jpurol.2018.10.019> <https://libkey.io/libraries/2592/openurl?genre=article&sid=OVID:medline&id=pmid:30467015&id=doi:10.1016%2Fj.jpurol.2018.10.019&issn=1477-5131&isbn=&volume=15&issue=1&spage=74.e1&pages=74.e1-74.e7&date=2019&title=Journal+of+pediatric+urology&atitle=Initial+collection+of+an+inadequate+24-hour+urine+sample+in+children+does+not+predict+subsequent+inadequate+collections.&aulast=Chan&pid=%3Cauthor%3EChan+KH%3BMoser+EA%3BWhittam+BM%3BMisseri+R%3BCain+MP%3BKrambeck+A%3BSchwaderer+A%3C%2Fauthor%3E%3CAN%3E30467015%3C%2FAN%3E%3CDT%3EJournal+Article%3C%2FDT%3E>

55. **The Effect of Lemonade and Diet Lemonade Upon Urinary Parameters Affecting Calcium Urinary Stone Formation.**

**Item Type:** Journal Article

**Authors:** Cheng, Julie W.;Wagner, Hillary;Asplin, John R.;Hodgkin, Georgia;Schlaifer, Amy;Fargusson, Michael;Fargusson, Joseph and Baldwin, D. Duane

**Publication Date:** 2019

**Journal:** Journal of Endourology 33(2), pp. 160–166

**Abstract: PURPOSE**: To determine the effects of regular and diet lemonade upon urinary parameters affecting kidney stone formation. **METHODS**: In this prospective blinded crossover study, 12 healthy participants consumed either 2 L of regular or diet lemonade daily while on a standardized low sodium moderate calcium diet. Twenty four-hour urine collections were obtained at baseline on the controlled diet only and on days 4 and 5 of each treatment phase. There was a 1-week washout period between regular and diet lemonade treatments. Primary outcomes were urine citrate, pH, and volume determined by 24-hour urine collections. Secondary outcomes included the supersaturation of calcium oxalate, calcium phosphate, and uric acid. **RESULTS**: Urine volume was significantly higher with both regular and diet lemonade consumption compared with baseline values. Urinary citrate significantly increased from baseline with diet lemonade only. Urine pH was unchanged with both beverages. The supersaturation of calcium oxalate significantly decreased with diet lemonade only, whereas supersaturation of calcium phosphate decreased with both beverages. Daily consumption of 2 L of regular and diet lemonade resulted in an intake of 168.4 and 170.2 mEq of citrate but a total alkali intake of 12.2 and 16.0 mEq, respectively. Compared with diet lemonade, regular lemonade provided subjects with 805 additional calories and 225 g of sugar per day. **CONCLUSIONS**: Diet lemonade may provide a low-calorie sugar-free cost-effective option for decreasing the risk of recurrent calcium nephrolithiasis through a significant increase in urine volume, increase in urinary citrate, and reduction in supersaturation of calcium oxalate and calcium phosphate.

**URL:** <https://ovidsp.ovid.com/ovidweb.cgi?T=JS&CSC=Y&NEWS=N&PAGE=fulltext&D=med16&DO=10.1089%2fend.2018.0623> <https://libkey.io/libraries/2592/openurl?genre=article&sid=OVID:medline&id=pmid:30585747&id=doi:10.1089%2Fend.2018.0623&issn=0892-7790&isbn=&volume=33&issue=2&spage=160&pages=160-166&date=2019&title=Journal+of+Endourology&atitle=The+Effect+of+Lemonade+and+Diet+Lemonade+Upon+Urinary+Parameters+Affecting+Calcium+Urinary+Stone+Formation.&aulast=Cheng&pid=%3Cauthor%3ECheng+JW%3BWagner+H%3BAsplin+JR%3BHodgkin+G%3BSchlaifer+A%3BFargusson+M%3BFargusson+J%3BBaldwin+DD%3C%2Fauthor%3E%3CAN%3E30585747%3C%2FAN%3E%3CDT%3EJournal+Article%3C%2FDT%3E>

56. **Antibiotic Use and Risk of Incident Kidney Stones in Female Nurses.**

**Item Type:** Journal Article

**Authors:** Ferraro, Pietro Manuel;Curhan, Gary C.;Gambaro, Giovanni and Taylor, Eric N.

**Publication Date:** 2019

**Journal:** American Journal of Kidney Diseases 74(6), pp. 736–741

**Abstract: RATIONALE & OBJECTIVE**: The intestinal microbiome may affect urinary stone disease by modulating the amount of oxalate absorbed from the intestine and subsequently excreted in urine. This study sought to explore the association between antibiotics, which alter the intestinal microbiota, and risk for urinary stone disease. **STUDY DESIGN**: Prospective cohort study. **SETTING & PARTICIPANTS**: 5,010 women in the Nurses' Health Study (NHS) I and II who had collected 24-hour urine samples. **EXPOSURES**: Use of antibiotics during the age range of 40 to 49 (NHS II), 40 to 59 (NHS I), and 20 to 39 years (both cohorts). **OUTCOMES**: Incident symptomatic urinary stone disease; urine composition. **ANALYTICAL APPROACH**: Cause-specific hazards regression adjusted for age, body mass index, comorbid conditions, thiazide use, and dietary factors. Follow-up was censored at the time of asymptomatic kidney stones, cancer, or death. **RESULTS**: Cumulative use of antibiotics for a total of 2 or more months during the age range of 40 to 49 years (NHS II) and 40 to 59 years (NHS II) was associated with significantly higher risk for developing incident stones compared with no use (pooled HR, 1.48; 95% CI, 1.12-1.96). Similar results were found for the period of 20 to 39 years (pooled HR, 1.36; 95% CI, 1.00-1.84). Results were unchanged after excluding participants who reported urinary tract infection with their stone event or as the most common reason for antibiotic use. Urine composition was generally similar across antibiotic groups except for marginally lower urine pH and citrate values among those taking antibiotics for 2 or more months. **LIMITATIONS**: Observational design; lack of information for type of antibiotic used; relatively large span of time between antibiotic use and urine collection. **CONCLUSIONS**: Use of antibiotics for more than 2 months in early adulthood and middle age is associated with higher risk for urinary stone disease in later life. Copyright © 2019 National Kidney Foundation, Inc. Published by Elsevier Inc. All rights reserved.

**URL:** <https://ovidsp.ovid.com/ovidweb.cgi?T=JS&CSC=Y&NEWS=N&PAGE=fulltext&D=med16&DO=10.1053%2fj.ajkd.2019.06.005> <https://libkey.io/libraries/2592/openurl?genre=article&sid=OVID:medline&id=pmid:31543288&id=doi:10.1053%2Fj.ajkd.2019.06.005&issn=0272-6386&isbn=&volume=74&issue=6&spage=736&pages=736-741&date=2019&title=American+Journal+of+Kidney+Diseases&atitle=Antibiotic+Use+and+Risk+of+Incident+Kidney+Stones+in+Female+Nurses.&aulast=Ferraro&pid=%3Cauthor%3EFerraro+PM%3BCurhan+GC%3BGambaro+G%3BTaylor+EN%3C%2Fauthor%3E%3CAN%3E31543288%3C%2FAN%3E%3CDT%3EComparative+Study%3C%2FDT%3E>

57. **Prevalence of twenty-four hour urine testing in Veterans with urinary stone disease.**

**Item Type:** Journal Article

**Authors:** Ganesan, Calyani;Thomas, I-Chun;Song, Shen;Sun, Andrew J.;Sohlberg, Ericka M.;Kurella Tamura, Manjula;Chertow, Glenn M.;Liao, Joseph C.;Conti, Simon;Elliott, Christopher S.;Leppert, John T. and Pao, Alan C.

**Publication Date:** 2019

**Journal:** PLoS ONE [Electronic Resource] 14(8), pp. e0220768

**Abstract: OBJECTIVE**: The American Urological Association guidelines recommend 24-hour urine testing in patients with urinary stone disease to decrease the risk of stone recurrence; however, national practice patterns for 24-hour urine testing are not well characterized. Our objective is to determine the prevalence of 24-hour urine testing in patients with urinary stone disease in the Veterans Health Administration and examine patient-specific and facility-level factors associated with 24-hour urine testing. Identifying variations in clinical practice can inform future quality improvement efforts in the management of urinary stone disease in integrated healthcare systems. **MATERIALS AND METHODS**: We accessed national Veterans Health Administration data through the Corporate Data Warehouse (CDW), hosted by the Veterans Affairs Informatics and Computing Infrastructure (VINCI), to identify patients with urinary stone disease. We defined stone formers as Veterans with one inpatient ICD-9 code for kidney or ureteral stones, two or more outpatient ICD-9 codes for kidney or ureteral stones, or one or more CPT codes for kidney or ureteral stone procedures from 2007 through 2013. We defined a 24-hour urine test as a 24-hour collection for calcium, oxalate, citrate or sulphate. We used multivariable regression to assess demographic, geographic, and selected clinical factors associated with 24-hour urine testing. **RESULTS**: We identified 130,489 Veterans with urinary stone disease; 19,288 (14.8%) underwent 24-hour urine testing. Patients who completed 24-hour urine testing were younger, had fewer comorbidities, and were more likely to be White. Utilization of 24-hour urine testing varied widely by geography and facility, the latter ranging from 1 to 40%. **CONCLUSIONS**: Fewer than one in six patients with urinary stone disease complete 24-hour urine testing in the Veterans Health Administration. In addition, utilization of 24-hour urine testing varies widely by facility identifying a target area for improvement in the care of patients with urinary stone disease. Future efforts to increase utilization of 24-hour urine testing and improve clinician awareness of targeted approaches to stone prevention may be warranted to reduce the morbidity and cost of urinary stone disease.

**URL:** <https://ovidsp.ovid.com/ovidweb.cgi?T=JS&CSC=Y&NEWS=N&PAGE=fulltext&D=med16&DO=10.1371%2fjournal.pone.0220768> <https://libkey.io/libraries/2592/openurl?genre=article&sid=OVID:medline&id=pmid:31393935&id=doi:10.1371%2Fjournal.pone.0220768&issn=1932-6203&isbn=&volume=14&issue=8&spage=e0220768&pages=e0220768&date=2019&title=PLoS+ONE+%5BElectronic+Resource%5D&atitle=Prevalence+of+twenty-four+hour+urine+testing+in+Veterans+with+urinary+stone+disease.&aulast=Ganesan&pid=%3Cauthor%3EGanesan+C%3BThomas+IC%3BSong+S%3BSun+AJ%3BSohlberg+EM%3BKurella+Tamura+M%3BChertow+GM%3BLiao+JC%3BConti+S%3BElliott+CS%3BLeppert+JT%3BPao+AC%3C%2Fauthor%3E%3CAN%3E31393935%3C%2FAN%3E%3CDT%3EJournal+Article%3C%2FDT%3E>

58. **Feasibility of a Telemedicine-Administered, Pharmacist-Staffed, Protocol-Driven, Multicenter Program for Kidney Stone Prevention in a Large Integrated Health Care System: Results of a Pilot Program.**

**Item Type:** Journal Article

**Authors:** Gasparini, Mark E.;Chang, Toby W.;St Lezin, Mark;Skerry, John E.;Chan, Andy and Ramaswamy, Krishna A.

**Publication Date:** 2019

**Journal:** Permanente Journal 23
**Abstract: INTRODUCTION**: Despite guidelines for prevention of recurrent renal calculi, routine dietary modification and metabolic evaluation are often not performed. **OBJECTIVE**: To determine feasibility of a multicenter, pharmacist-staffed program to enroll patients at high risk of recurrent kidney stones and provide dietary instruction, metabolic evaluation, and medical therapy via telemedicine. **METHODS**: A total of 536 consecutive adult patients were referred from 3 Northern California Kaiser Permanente facilities. We determined the proportion of patients who enrolled, received dietary counseling, and completed metabolic evaluation at 12 months. The program was staffed by a clinical pharmacist and supervised by urologists following a protocol based on the American Urological Association guidelines. Patients were contacted entirely via telemedicine. Cystine or struvite kidney stones, renal tubular acidosis, and primary hyperoxaluria were exclusion criteria. **RESULTS**: Of the 536 patients, 500 agreed to enrollment. Among patients enrolled for 3 months, 99% self-reported compliance with at least 3 of 5 aspects of dietary advice. A complete metabolic evaluation including 24-hour urine collection was performed in 80% of patients by 12 months. A significant improvement in all urinary parameters occurred in 52 patients with calcium stones who repeated 24-hour urine testing. The 12-month dropout rate was 12.4%. **CONCLUSION**: A telemedicine-administered, pharmacist-staffed, protocol-driven program can provide dietary advice and obtain compliance with metabolic testing for patients at high risk of recurrent kidney stones. Rates of metabolic testing and dropout compare favorably with previously reported rates. This report represents, to our knowledge, the first telemedicine-administered, pharmacist-staffed, kidney stone prevention program published in the literature.

**URL:** <https://ovidsp.ovid.com/ovidweb.cgi?T=JS&CSC=Y&NEWS=N&PAGE=fulltext&D=med16&DO=10.7812%2fTPP%2f19.023> <https://libkey.io/libraries/2592/openurl?genre=article&sid=OVID:medline&id=pmid:31702984&id=doi:10.7812%2FTPP%2F19.023&issn=1552-5767&isbn=&volume=23&issue=&spage=&pages=&date=2019&title=Permanente+Journal&atitle=Feasibility+of+a+Telemedicine-Administered%2C+Pharmacist-Staffed%2C+Protocol-Driven%2C+Multicenter+Program+for+Kidney+Stone+Prevention+in+a+Large+Integrated+Health+Care+System%3A+Results+of+a+Pilot+Program.&aulast=Gasparini&pid=%3Cauthor%3EGasparini+ME%3BChang+TW%3BSt+Lezin+M%3BSkerry+JE%3BChan+A%3BRamaswamy+KA%3C%2Fauthor%3E%3CAN%3E31702984%3C%2FAN%3E%3CDT%3EJournal+Article%3C%2FDT%3E>

59. **Trends in renal calculus composition and 24-hour urine analyses in patients with neurologically derived musculoskeletal deficiencies.**

**Item Type:** Journal Article

**Authors:** Hugar, Lee A.;Kafka, Ilan;Fuller, Thomas W.;Taan, Hassan;Averch, Timothy D. and Semins, Michelle J.

**Publication Date:** 2019

**Journal:** International Braz J Urol 45(3), pp. 572–580

**Abstract: PURPOSE**: To better characterize metabolic stone risk in patients with neurologically derived musculoskeletal deficiencies (NDMD) by determining how patient characteristics relate to renal calculus composition and 24-hour urine parameters. **MATERIALS AND METHODS**: We performed a retrospective cohort study of adult patients with neurologically derived musculoskeletal deficiencies presenting to our multidisciplinary Kidney Stone Clinic. Patients with a diagnosis of NDMD, at least one 24-hour urine collection, and one chemical stone analysis were included in the analysis. Calculi were classified as primarily metabolic or elevated pH. We assessed in clinical factors, demographics, and urine metabolites for differences between patients who formed primarily metabolic or elevated pH stones. **RESULTS**: Over a 16-year period, 100 patients with NDMD and nephrolithiasis were identified and 41 met inclusion criteria. Thirty percent (12 / 41) of patients had purely metabolic calculi. Patients with metabolic calculi were significantly more likely to be obese (median body mass index 30.3kg / m2 versus 25.9kg / m2), void spontaneously (75% vs. 6.9%), and have low urine volumes (100% vs. 69%). Patients who formed elevated pH stones were more likely to have positive preoperative urine cultures with urease splitting organisms (58.6% vs. 16.7%) and be hyperoxaluric and hypocitraturic on 24-hour urine analysis (37mg / day and 265mg / day versus 29mg / day and 523mg / day). **CONCLUSIONS**: Among patients with NDMD, metabolic factors may play a more significant role in renal calculus formation than previously believed. There is still a high incidence of carbonate apatite calculi, which could be attributed to bacteriuria. However, obesity, low urine volumes, hypocitraturia, and hyperoxaluria suggest an underrecognized metabolic contribution to stone formation in this population. Copyright R by the International Brazilian Journal of Urology.

**URL:** <https://ovidsp.ovid.com/ovidweb.cgi?T=JS&CSC=Y&NEWS=N&PAGE=fulltext&D=med16&DO=10.1590%2fS1677-5538.IBJU.2018.0531> <https://libkey.io/libraries/2592/openurl?genre=article&sid=OVID:medline&id=pmid:30676304&id=doi:10.1590%2FS1677-5538.IBJU.2018.0531&issn=1677-5538&isbn=&volume=45&issue=3&spage=572&pages=572-580&date=2019&title=International+Braz+J+Urol&atitle=Trends+in+renal+calculus+composition+and+24-hour+urine+analyses+in+patients+with+neurologically+derived+musculoskeletal+deficiencies.&aulast=Hugar&pid=%3Cauthor%3EHugar+LA%3BKafka+I%3BFuller+TW%3BTaan+H%3BAverch+TD%3BSemins+MJ%3C%2Fauthor%3E%3CAN%3E30676304%3C%2FAN%3E%3CDT%3EJournal+Article%3C%2FDT%3E>

60. **Patient Characteristics Associated With Completion of 24-hour Urine Analyses Among Children and Adolescents With Nephrolithiasis.**

**Item Type:** Journal Article

**Authors:** Lee, Albert S.;McGarry, Laura;Bowen, Diana K. and Tasian, Gregory E.

**Publication Date:** 2019

**Journal:** Urology 127, pp. 102–106

**Abstract: OBJECTIVE**: To inform the development of strategies to improve adherence to guidelines, we sought to identify characteristics of pediatric patients with nephrolithiasis associated with completing 24-hour urine analyses. **MATERIALS AND METHODS**: We performed a retrospective cohort study of patients with nephrolithiasis aged 3-18years treated in a large pediatric healthcare system from May 2012 to May 2017. Multivariable Cox models were fit to estimate the association between patient characteristics and completion of a 24-hour urine analysis. **RESULTS**: Among 623 patients, 317 (50.9%) completed a 24-hour urine collection. Median age was 14.4years (interquartile range [IQR] 10.5, 16.3). In adjusted analyses, age at diagnosis (hazard ratio [HR] 1.03; 95% confidence interval [CI] 1.01-1.07), renal colic on presentation (HR 1.72; 95% CI 1.15-2.58), and family history of nephrolithiasis (HR 1.50; 95% CI 1.17-1.93) were associated with an increased likelihood of completion of a 24-hour urine. Public/government assistance insurance (HR 0.68; 95% CI 0.48-0.96) was associated with decreased likelihood of completing a 24-hour urine. **CONCLUSION**: Patients who had prior painful experiences with stones (renal colic), and potential better understanding of nephrolithiasis (family history, older age on presentation) were more likely to complete a 24-hour urine. Those patients with public insurance/government assistance were less likely to complete a 24-hour urine. These results can be used to develop strategies to improve pediatric patients' adherence to completing 24-hour urine collections. Copyright © 2019. Published by Elsevier Inc.

**URL:** <https://ovidsp.ovid.com/ovidweb.cgi?T=JS&CSC=Y&NEWS=N&PAGE=fulltext&D=med16&DO=10.1016%2fj.urology.2019.02.008> <https://libkey.io/libraries/2592/openurl?genre=article&sid=OVID:medline&id=pmid:30796989&id=doi:10.1016%2Fj.urology.2019.02.008&issn=0090-4295&isbn=&volume=127&issue=&spage=102&pages=102-106&date=2019&title=Urology&atitle=Patient+Characteristics+Associated+With+Completion+of+24-hour+Urine+Analyses+Among+Children+and+Adolescents+With+Nephrolithiasis.&aulast=Lee&pid=%3Cauthor%3ELee+AS%3BMcGarry+L%3BBowen+DK%3BTasian+GE%3C%2Fauthor%3E%3CAN%3E30796989%3C%2FAN%3E%3CDT%3EJournal+Article%3C%2FDT%3E>

61. **Increased production and reduced urinary buffering of acid in uric acid stone formers is ameliorated by pioglitazone.**

**Item Type:** Journal Article

**Authors:** Maalouf, Naim M.;Poindexter, John R.;Adams-Huet, Beverley;Moe, Orson W. and Sakhaee, Khashayar

**Publication Date:** 2019

**Journal:** Kidney International 95(5), pp. 1262–1268

**Abstract:** Idiopathic uric acid nephrolithiasis is characterized by an overly acidic urine pH caused by the combination of increased acid production and inadequate buffering of urinary protons by ammonia. A large proportion of uric acid stone formers exhibit features of the metabolic syndrome. We previously demonstrated that thiazolidinediones improved the urinary biochemical profile in an animal model of the metabolic syndrome. In this proof-of-concept study, we examined whether the thiazolidinedione pioglitazone can also ameliorate the overly acidic urine in uric acid stone formers. Thirty-six adults with idiopathic uric acid nephrolithiasis were randomized to pioglitazone 30 mg/day or matching placebo for 24 weeks. At baseline and study end, participants underwent collection of blood and 24-hour urine in an inpatient research unit while consuming a fixed metabolic diet, followed by assessment of the ammoniagenic response to an acute oral acid load. Twenty-eight participants completed the study. Pioglitazone treatment improved features of the metabolic syndrome. Pioglitazone also reduced net acid excretion and increased urine pH (5.37 to 5.59), the proportion of net acid excreted as ammonium, and ammonium excretion in response to an acute acid load, whereas these parameters were unchanged with placebo. Treatment of patients with idiopathic uric acid nephrolithiasis with pioglitazone for 24 weeks led to a reduction in the acid load presented to the kidney and a more robust ammoniagenesis and ammonium excretion, resulting in significantly higher urine pH. Future studies should consider the impact of this targeted therapy on uric acid stone formation. Copyright © 2019 International Society of Nephrology. Published by Elsevier Inc. All rights reserved.

**URL:** <https://ovidsp.ovid.com/ovidweb.cgi?T=JS&CSC=Y&NEWS=N&PAGE=fulltext&D=med16&DO=10.1016%2fj.kint.2018.11.024> <https://libkey.io/libraries/2592/openurl?genre=article&sid=OVID:medline&id=pmid:30795852&id=doi:10.1016%2Fj.kint.2018.11.024&issn=0085-2538&isbn=&volume=95&issue=5&spage=1262&pages=1262-1268&date=2019&title=Kidney+International&atitle=Increased+production+and+reduced+urinary+buffering+of+acid+in+uric+acid+stone+formers+is+ameliorated+by+pioglitazone.&aulast=Maalouf&pid=%3Cauthor%3EMaalouf+NM%3BPoindexter+JR%3BAdams-Huet+B%3BMoe+OW%3BSakhaee+K%3C%2Fauthor%3E%3CAN%3E30795852%3C%2FAN%3E%3CDT%3EJournal+Article%3C%2FDT%3E>

62. **Comparison of metabolic changes for stone risks in 24-hour urine between non- and postmenopausal women.**

**Item Type:** Journal Article

**Authors:** Mai, Zanlin;Li, Xiaoxia;Jiang, Chonghe;Liu, Yongda;Chen, Yiwen;Wu, Wenqi and Zeng, Guohua

**Publication Date:** 2019

**Journal:** PLoS ONE [Electronic Resource] 14(1), pp. e0208893

**Abstract: BACKGROUND**: To explore the differences of 24-hour urine compositions associated with urolithiasis between non- and postmenopausal females. **METHODS**: The 24-hour urine samples of female participants were collected from May 2013 to July 2014 along with national cross-sectional study of urolithiasis among adults aged >=18 years in China. The exclusion criteria for the participants were: serum creatinine > 133mumol/L, with urinary tract infection, gout, hyperthyroidism, malignancy, had a history of cancer, kidney stones, enterectomy, had taken thiazide diuretics, allopurinol, vitamin supplement, potassium citrate or calcium supplements during the past two weeks. The compositions associated with urinary stone in 24-hour urine were measured and compared between non-and postmenopausal women. **RESULTS**: A total of 603 24-hour urine samples of female participants were analyzed. 354 women with a mean age of 52.5+/- 14.03 (range 19-84) years met the criteria, including 160 non-menopausal women and 194 postmenopausal women. Compared to the non-menopausal women, postmenopausal women had a lower secretion of citrate (p = 0.043), magnesium (p = 0.001) and creatinine (p = 0.001) in 24h urine. Multivariate linear regression analysis showed that the menopause status was associated with the changes in magnesium (p = 0.003) and creatinine (p = 0.002) secretion, whereas not with the changes in citrate (p = 0.402) secretion. **CONCLUSIONS**: Postmenopausal women have a significant lower secretion of magnesium in their 24-hour urine than non-menopausal ones. We suppose that might be associated with increased risk of urinary stone formation among postmenopausal women.

**URL:** <https://ovidsp.ovid.com/ovidweb.cgi?T=JS&CSC=Y&NEWS=N&PAGE=fulltext&D=med16&DO=10.1371%2fjournal.pone.0208893> <https://libkey.io/libraries/2592/openurl?genre=article&sid=OVID:medline&id=pmid:30677034&id=doi:10.1371%2Fjournal.pone.0208893&issn=1932-6203&isbn=&volume=14&issue=1&spage=e0208893&pages=e0208893&date=2019&title=PLoS+ONE+%5BElectronic+Resource%5D&atitle=Comparison+of+metabolic+changes+for+stone+risks+in+24-hour+urine+between+non-+and+postmenopausal+women.&aulast=Mai&pid=%3Cauthor%3EMai+Z%3BLi+X%3BJiang+C%3BLiu+Y%3BChen+Y%3BWu+W%3BZeng+G%3C%2Fauthor%3E%3CAN%3E30677034%3C%2FAN%3E%3CDT%3EJournal+Article%3C%2FDT%3E>

63. **Twenty-Four Hour Urine Testing and Prescriptions for Urinary Stone Disease-Related Medications in Veterans.**

**Item Type:** Journal Article

**Authors:** Song, Shen;Thomas, I-Chun;Ganesan, Calyani;Sohlberg, Ericka M.;Chertow, Glenn M.;Liao, Joseph C.;Conti, Simon;Elliott, Christopher S.;Pao, Alan C. and Leppert, John T.

**Publication Date:** 2019

**Journal:** Clinical Journal of the American Society of Nephrology: CJASN 14(12), pp. 1773–1780

**Abstract: BACKGROUND AND OBJECTIVES**: Current guidelines recommend 24-hour urine testing in the evaluation and treatment of persons with high-risk urinary stone disease. However, how much clinicians use information from 24-hour urine testing to guide secondary prevention strategies is unknown. We sought to determine the degree to which clinicians initiate or continue stone disease-related medications in response to 24-hour urine testing. **DESIGN, SETTING, PARTICIPANTS, & MEASUREMENTS**: We examined a national cohort of 130,489 patients with incident urinary stone disease in the Veterans Health Administration between 2007 and 2013 to determine whether prescription patterns for thiazide diuretics, alkali therapy, and allopurinol changed in response to 24-hour urine testing. **RESULTS**: Stone formers who completed 24-hour urine testing (n=17,303; 13%) were significantly more likely to be prescribed thiazide diuretics, alkali therapy, and allopurinol compared with those who did not complete a 24-hour urine test (n=113,186; 87%). Prescription of thiazide diuretics increased in patients with hypercalciuria (9% absolute increase if urine calcium 201-400 mg/d; 21% absolute increase if urine calcium >400 mg/d, P: Stone formers who completed 24-hour urine testing (n=17,303; 13%) were significantly more likely to be prescribed thiazide diuretics, alkali therapy, and allopurinol compared with those who did not complete a 24-hour urine test (n=113,186; 87%). Prescription of thiazide diuretics increased in patients with hypercalciuria (9% absolute increase if urine calcium 201-400 mg/d; 21% absolute increase if urine calcium >400 mg/d, P: Stone formers who completed 24-hour urine testing (n=17,303; 13%) were significantly more likely to be prescribed thiazide diuretics, alkali therapy, and allopurinol compared with those who did not complete a 24-hour urine test (n=113,186; 87%). Prescription of thiazide diuretics increased in patients with hypercalciuria (9% absolute increase if urine calcium 201-400 mg/d; 21% absolute increase if urine calcium >400 mg/d, P800 mg/d, P800 mg/d, P**CONCLUSIONS**: Clinicians adjust their treatment regimens in response to 24-hour urine testing by increasing the prescription of medications thought to reduce risk for urinary stone disease. Most patients who might benefit from targeted medications remain untreated. Copyright © 2019 by the American Society of Nephrology.

**URL:** <https://ovidsp.ovid.com/ovidweb.cgi?T=JS&CSC=Y&NEWS=N&PAGE=fulltext&D=med16&DO=10.2215%2fCJN.03580319> <https://libkey.io/libraries/2592/openurl?genre=article&sid=OVID:medline&id=pmid:31712387&id=doi:10.2215%2FCJN.03580319&issn=1555-9041&isbn=&volume=14&issue=12&spage=1773&pages=1773-1780&date=2019&title=Clinical+Journal+of+The+American+Society+of+Nephrology%3A+CJASN&atitle=Twenty-Four+Hour+Urine+Testing+and+Prescriptions+for+Urinary+Stone+Disease-Related+Medications+in+Veterans.&aulast=Song&pid=%3Cauthor%3ESong+S%3BThomas+IC%3BGanesan+C%3BSohlberg+EM%3BChertow+GM%3BLiao+JC%3BConti+S%3BElliott+CS%3BPao+AC%3BLeppert+JT%3C%2Fauthor%3E%3CAN%3E31712387%3C%2FAN%3E%3CDT%3EJournal+Article%3C%2FDT%3E>

64. **Effect of vitamin D supplementation on 24-hour urine calcium in patients with calcium Urolithiasis and vitamin D deficiency.**

**Item Type:** Journal Article

**Authors:** Taheri, Maryam;Tavasoli, Sanaz;Shokrzadeh, Fatemeh;Amiri, Fahimeh Bagheri and Basiri, Abbas

**Publication Date:** 2019

**Journal:** International Braz J Urol 45(2), pp. 340–346

**Abstract: PURPOSE**: Hypercalciuria is one of the risk factors for calcium kidney stone formation (the most common type of urinary stones). Although vitamin D deficiency is prevalent among urolithiasis patients, the effect of vitamin D supplementation on urine calcium in these patients is still unclear. **MATERIALS AND METHODS**: In this retrospective study, medical and laboratory tests records of 26 patients with recurrent calcium kidney stones and vitamin D deficiency treated with 50000IU vitamin D per week for 8-12 weeks were analyzed. The changes in 24-hour urine calcium (24-h Ca), serum 25-hydroxyvitamin D (25 (OH) D), serum parathormone (PTH), other 24-hour urine metabolites and calculated relative supersaturations of calcium oxalate (CaOxSS), calcium phosphate (CaPSS) and uric acid (UASS) were assessed. Moreover, correlations between changes in 24-h Ca and other aforementioned variables were assessed. **RESULTS**: Serum 25 (OH) D and 24-h Ca increased after vitamin D supplementation, while serum PTH decreased (p : Serum 25 (OH) D and 24-h Ca increased after vitamin D supplementation, while serum PTH decreased (p **CONCLUSIONS**: The result of current study suggests that although urine Ca increased in vitamin D supplemented patients, this increase was not associated with the increase in serum vitamin D and may be due to other factors such as dietary factors. Further randomized clinical trials considering other factors associated with urine Ca are warranted. Copyright R by the International Brazilian Journal of Urology.

**URL:** <https://ovidsp.ovid.com/ovidweb.cgi?T=JS&CSC=Y&NEWS=N&PAGE=fulltext&D=med16&DO=10.1590%2fS1677-5538.IBJU.2018.0522> <https://libkey.io/libraries/2592/openurl?genre=article&sid=OVID:medline&id=pmid:30735332&id=doi:10.1590%2FS1677-5538.IBJU.2018.0522&issn=1677-5538&isbn=&volume=45&issue=2&spage=340&pages=340-346&date=2019&title=International+Braz+J+Urol&atitle=Effect+of+vitamin+D+supplementation+on+24-hour+urine+calcium+in+patients+with+calcium+Urolithiasis+and+vitamin+D+deficiency.&aulast=Taheri&pid=%3Cauthor%3ETaheri+M%3BTavasoli+S%3BShokrzadeh+F%3BAmiri+FB%3BBasiri+A%3C%2Fauthor%3E%3CAN%3E30735332%3C%2FAN%3E%3CDT%3EJournal+Article%3C%2FDT%3E>

65. **What Factors Drive Staghorn vs Nonstaghorn Pattern Growth in Patients with Metabolic Stones?.**

**Item Type:** Journal Article

**Authors:** Winoker, Jared S.;Bamberger, Jacob N.;Chandhoke, Ryan A.;Atallah, William and Gupta, Mantu

**Publication Date:** 2019

**Journal:** Journal of Endourology 33(11), pp. 954–959

**Abstract: Purpose:** There has been an increase in the number of staghorn calculi that form in the absence of infection (metabolic staghorns). It is unknown why some large metabolic calculi form as solitary or multiple separate, nonbranching caliceal stones, whereas others develop as staghorn stones forming a cast of the collecting system. We sought to compare these two groups of metabolic stone formers (SFs) in an attempt to shed light on these disparate stone-forming phenomena. **Materials and Methods:** From January 2017 to September 2018, 190 patients underwent percutaneous nephrolithotomy for stones >2 cm. We identified 86 (45%) patients with a metabolic stone, defined as >=80% calcium oxalate monohydrate or dihydrate and/or calcium phosphate. Exclusion criteria included stones composed of cystine, >20% uric acid, or any infectious element (struvite or carbonate apatite). Metabolic staghorn and nonstaghorn SFs were compared with respect to medical comorbidity, 24-hour urine parameters, stone and urine microbiology, stone compositions, and intraoperative findings. Statistical differences were assessed using chi-square analysis, Fisher's exact test, and Student's t-test. **Results:** In total, 25 (29%) staghorn and 61 (71%) nonstaghorn SFs were included for analysis. The groups were statistically similar in age, sex, body mass index, and medical comorbidity. Staghorn SFs had larger stone burdens (p In total, 25 (29%) staghorn and 61 (71%) nonstaghorn SFs were included for analysis. The groups were statistically similar in age, sex, body mass index, and medical comorbidity. Staghorn SFs had larger stone burdens (p **Conclusions:** Although potentially driven by urinary oxalate, whether a metabolic stone will form into a staghorn configuration or not does not appear significantly influenced by standard determinants of stone development, including metabolic profile, cultures, hydronephrosis, and pelvicaliceal anatomy, among others. Further work is needed to elucidate the physicochemical factors that govern the pathogenesis of this increasingly prevalent entity.

**URL:** <https://ovidsp.ovid.com/ovidweb.cgi?T=JS&CSC=Y&NEWS=N&PAGE=fulltext&D=med16&DO=10.1089%2fend.2019.0079> <https://libkey.io/libraries/2592/openurl?genre=article&sid=OVID:medline&id=pmid:31436119&id=doi:10.1089%2Fend.2019.0079&issn=0892-7790&isbn=&volume=33&issue=11&spage=954&pages=954-959&date=2019&title=Journal+of+Endourology&atitle=What+Factors+Drive+Staghorn+vs+Nonstaghorn+Pattern+Growth+in+Patients+with+Metabolic+Stones%3F.&aulast=Winoker&pid=%3Cauthor%3EWinoker+JS%3BBamberger+JN%3BChandhoke+RA%3BAtallah+W%3BGupta+M%3C%2Fauthor%3E%3CAN%3E31436119%3C%2FAN%3E%3CDT%3EJournal+Article%3C%2FDT%3E>

66. **Renal Disorders in Pulmonary Sarcoidosis Patients.**

**Item Type:** Journal Article

**Authors:** Yassari, Fatemeh;Kiani, Arda;Taghavi, Kimia;Abdi, Ebrahim;Emami, Habib;Seifi, Sharareh and Abedini, Atefeh

**Publication Date:** Ja ,2019

**Journal:** Iranian Journal of Kidney Diseases 13(1), pp. 21–26

**Abstract: INTRODUCTION**: The frequency of kidney disorders varies in pulmonary sarcoidosis patients. Since the prevalence of kidney disorders among Iranian sarcoidosis patients is uncertain, this study aimed to evaluate kidney disorders and associated manifestations in Iranian pulmonary sarcoidosis patients. **MATERIALS AND METHODS**: One hundred patients with confirmed granuloma as pulmonary sarcoidosis were studied for renal disorders. Size of urinary tract and the presence of renal stones were checked via clinical examination and urinary organ ultrasonography. Patients' 24-hour urine sample was examined for pH, calcium, protein (over 250 mg) and creatinine (over 1.4 mg). **RESULTS**: Thirty-three percent of the patients expressed renal disorders simultaneously.Uric acid in pulmonary sarcoidosis patients could be correlated with the probability of developing renal stone. In addition, 1,25-dihydroxyvitamin D levels above 30 ng/mL and uric acid levels above 7 mg/dL in urine were directly correlated with renal disorders in sarcoidosis patients. **CONCLUSIONS**: Urinalysis is an easy and reliable method for assessing renal disorders in sarcoidosis patients. The current study proposes inclusion of urinalysis in routine checkups of sarcoidosis individuals.

**URL:** <https://ovidsp.ovid.com/ovidweb.cgi?T=JS&CSC=Y&NEWS=N&PAGE=fulltext&D=med16&AN=30851715> <https://libkey.io/libraries/2592/openurl?genre=article&sid=OVID:medline&id=pmid:30851715&id=doi:&issn=1735-8582&isbn=&volume=13&issue=1&spage=21&pages=21-26&date=2019&title=Iranian+journal+of+Kidney+Diseases&atitle=Renal+Disorders+in+Pulmonary+Sarcoidosis+Patients.&aulast=Yassari&pid=%3Cauthor%3EYassari+F%3BKiani+A%3BTaghavi+K%3BAbdi+E%3BEmami+H%3BSeifi+S%3BAbedini+A%3C%2Fauthor%3E%3CAN%3E30851715%3C%2FAN%3E%3CDT%3EJournal+Article%3C%2FDT%3E>

67. **Claudin-14 Gene Polymorphisms and Urine Calcium Excretion.**

**Item Type:** Journal Article

**Authors:** Arcidiacono, Teresa;Simonini, Marco;Lanzani, Chiara;Citterio, Lorena;Salvi, Erika;Barlassina, Cristina;Spotti, Donatella;Cusi, Daniele;Manunta, Paolo and Vezzoli, Giuseppe

**Publication Date:** 2018

**Journal:** Clinical Journal of the American Society of Nephrology: CJASN 13(10), pp. 1542–1549

**Abstract: BACKGROUND AND OBJECTIVES**: Claudin-16 and -19 are proteins forming pores for the paracellular reabsorption of divalent cations in the ascending limb of Henle loop; conversely, claudin-14 decreases ion permeability of these pores. Single-nucleotide polymorphisms in gene coding for claudin-14 were associated with kidney stones and calcium excretion. This study aimed to explore the association of claudin-14, claudin-16, and claudin-19 single-nucleotide polymorphisms with calcium excretion. **DESIGN, SETTING, PARTICIPANTS, & MEASUREMENTS**: We performed a retrospective observational study of 393 patients with hypertension who were naive to antihypertensive drugs, in whom we measured 24-hour urine calcium excretion; history of kidney stones was ascertained by interview; 370 of these patients underwent an intravenous 0.9% sodium chloride infusion (2 L in 2 hours) to evaluate the response of calcium excretion in three different 2-hour urine samples collected before, during, and after saline infusion. Genotypes of claudin-14, claudin-16, and claudin-19 were obtained from data of a previous genome-wide association study in the same patients. **RESULTS**: Thirty-one single-nucleotide polymorphisms of the 3' region of the claudin-14 gene were significantly associated with 24-hour calcium excretion and calcium excretion after saline infusion. The most significant associated single-nucleotide polymorphism was rs219755 (24-hour calcium excretion in GG, 225+/-124 mg/24 hours; 24-hour calcium excretion in GA, 194+/-100 mg/24 hours; 24-hour calcium excretion in AA, 124+/-73 mg/24 hours; P: Thirty-one single-nucleotide polymorphisms of the 3' region of the claudin-14 gene were significantly associated with 24-hour calcium excretion and calcium excretion after saline infusion. The most significant associated single-nucleotide polymorphism was rs219755 (24-hour calcium excretion in GG, 225+/-124 mg/24 hours; 24-hour calcium excretion in GA, 194+/-100 mg/24 hours; 24-hour calcium excretion in AA, 124+/-73 mg/24 hours; P**CONCLUSIONS**: Claudin-14 genotype at the 3' region is associated with calcium excretion in 24-hour urine and after the calciuretic stimulus of saline infusion. Copyright © 2018 by the American Society of Nephrology.

**URL:** <https://ovidsp.ovid.com/ovidweb.cgi?T=JS&CSC=Y&NEWS=N&PAGE=fulltext&D=med15&DO=10.2215%2fCJN.01770218> <https://libkey.io/libraries/2592/openurl?genre=article&sid=OVID:medline&id=pmid:30232134&id=doi:10.2215%2FCJN.01770218&issn=1555-9041&isbn=&volume=13&issue=10&spage=1542&pages=1542-1549&date=2018&title=Clinical+Journal+of+The+American+Society+of+Nephrology%3A+CJASN&atitle=Claudin-14+Gene+Polymorphisms+and+Urine+Calcium+Excretion.&aulast=Arcidiacono&pid=%3Cauthor%3EArcidiacono+T%3BSimonini+M%3BLanzani+C%3BCitterio+L%3BSalvi+E%3BBarlassina+C%3BSpotti+D%3BCusi+D%3BManunta+P%3BVezzoli+G%3C%2Fauthor%3E%3CAN%3E30232134%3C%2FAN%3E%3CDT%3EJournal+Article%3C%2FDT%3E>

68. **Seasonal variations in urinary calcium, volume, and vitamin d in kidney stone formers.**

**Item Type:** Journal Article

**Authors:** Attalla, Kyrollis;De, Shubha;Sarkissian, Carl and Monga, Manoj

**Publication Date:** 2018

**Journal:** International Braz J Urol 44(5), pp. 947–951

**Abstract: OBJECTIVES**: To investigate the seasonal variations in urinary calcium, serum vitamin D, and urinary volume in patients with a history of nephrolithiasis. **MATERIALS AND METHODS**: Patients included were those who completed a 24-hour urine metabolic evaluation on two occasions; one in summer (June-Aug) and one in winter (Nov-Jan), and who had not started any medications or been instructed on dietary modifications in the interval between the two tests that may have impacted the results. Patients were excluded if they were on thiazide diuretics or were taking calcium and / or Vitamin D supplementation. Welch's t-test was used to compare the difference in average summer and winter values. Unpaired Student t-test was used to compare baseline parameters (age, BMI), and Paired Student t-test was used to compare average seasonal measurements in men vs. women. **RESULTS**: 136 patients were identified who were not taking calcium or vitamin D supplements or thiazide diuretics, and who were not instructed on dietary modifications in the interval between the two measured parameters. No significant differences were observed when comparing male to female baseline parameters of age or BMI (Table-1). Average 24-hour urine calcium was higher (226.60) in the winter than in summer (194.18) and was significant in males (p = 0.014) and females (p : 136 patients were identified who were not taking calcium or vitamin D supplements or thiazide diuretics, and who were not instructed on dietary modifications in the interval between the two measured parameters. No significant differences were observed when comparing male to female baseline parameters of age or BMI (Table-1). Average 24-hour urine calcium was higher (226.60) in the winter than in summer (194.18) and was significant in males (p = 0.014) and females (p **CONCLUSIONS**: Urinary calcium is higher in winter months compared to summer months. As such, tailoring medical preventative strategies to the time of year may be helpful. Copyright R by the International Brazilian Journal of Urology.

**URL:** <https://ovidsp.ovid.com/ovidweb.cgi?T=JS&CSC=Y&NEWS=N&PAGE=fulltext&D=med15&DO=10.1590%2fS1677-5538.IBJU.2018.0095> <https://libkey.io/libraries/2592/openurl?genre=article&sid=OVID:medline&id=pmid:29757578&id=doi:10.1590%2FS1677-5538.IBJU.2018.0095&issn=1677-5538&isbn=&volume=44&issue=5&spage=947&pages=947-951&date=2018&title=International+Braz+J+Urol&atitle=Seasonal+variations+in+urinary+calcium%2C+volume%2C+and+vitamin+d+in+kidney+stone+formers.&aulast=Attalla&pid=%3Cauthor%3EAttalla+K%3BDe+S%3BSarkissian+C%3BMonga+M%3C%2Fauthor%3E%3CAN%3E29757578%3C%2FAN%3E%3CDT%3EJournal+Article%3C%2FDT%3E>

69. **The tolerability of Potassium Citrate Tablet in patients with intolerance to Potassium Citrate Powder form.**

**Item Type:** Journal Article

**Authors:** Basiri, Abbas;Taheri, Fatemeh and Taheri, Maryam

**Publication Date:** 2018

**Journal:** Urology Journal 15(1), pp. 16–20

**Abstract: PURPOSE**: To assess the tolerability of Potassium Citrate (KCit) tablet in patients with kidney stones that were not able to use the powder form of this drug due to unfavorable salty taste and gastrointestinal complications. **MATERIALS AND METHODS**: Twenty-three stone formers, with intolerance to potassium citrate powder form, which had referred to Labbafinejad stone preventive clinic (2015), have been included in this study. All of the patients took two Potassium citrate tablets (10 meq), three times a day for two weeks. Spot urine sample and the 24-hour urine collections were performed before and after KCit therapy. In addition, a visual analog taste scale was completed to gauge the taste and palatability of the KCit tablet in comparing with the powder form. **RESULTS**: All of the patients claimed that they consumed the tablets as prescribed. The urine pH and the 24-hour citrate and potassium were significantly higher after the treatment. In addition, the mean visual analog scale score was significantly improved in KCit therapy with tablet form versus to powder type (good vs. terrible score). **CONCLUSION**: Oral tolerance of KCit therapy is improved with the use of Potassium Citrate tablet, with beneficial effects on 24-hour urine citrate, potassium, and pH.

**URL:** <https://ovidsp.ovid.com/ovidweb.cgi?T=JS&CSC=Y&NEWS=N&PAGE=fulltext&D=med15&DO=10.22037%2fuj.v15i1.4356> <https://libkey.io/libraries/2592/openurl?genre=article&sid=OVID:medline&id=pmid:29363092&id=doi:10.22037%2Fuj.v15i1.4356&issn=1735-1308&isbn=&volume=15&issue=1&spage=16&pages=16-20&date=2018&title=Urology+Journal&atitle=The+tolerability+of+Potassium+Citrate+Tablet+in+patients+with+intolerance+to+Potassium+Citrate+Powder+form.&aulast=Basiri&pid=%3Cauthor%3EBasiri+A%3BTaheri+F%3BTaheri+M%3C%2Fauthor%3E%3CAN%3E29363092%3C%2FAN%3E%3CDT%3EClinical+Trial%3C%2FDT%3E>

70. **A multiregional Italian cohort of 24-hour urine metabolic evaluation in renal stone formers.**

**Item Type:** Journal Article

**Authors:** Esperto, Francesco;Marangella, Martino;Trinchieri, Alberto;Petrarulo, Michele and Miano, Roberto

**Publication Date:** Fe ,2018

**Journal:** Minerva Urologica E Nefrologica 70(1), pp. 87–94

**Abstract: BACKGROUND**: Nephrolithiasis is a common condition with several studies documenting an increased prevalence over the past four decades. EAU and AUA guidelines recommend 24-hour urine metabolic evaluation in high-risk stone formers. Aim of this study is to retrospectively evaluate the first three years of experience with LithoTest R (Biohealth Italia Srl, Turin, Italy) through the analysis of demographic, clinical and biochemical data collected from a large cohort of patients with kidney stones. **METHODS**: We retrospectively analyzed data from the LithoCenter database, including data from outpatient consultations, between January 2007 and December 2009 from all over Italy. LithoTest R was performed through a 24-hour urine collection and included measurements of urine volume and pH, 24-hour excretion of creatinine as well as main cations and anions, including calcium, magnesium sodium potassium, ammonium, uric acid, oxalate, citrate, phosphate, inorganic sulphate and chloride. Urine state of saturation for calcium oxalate (betaCaOx), calcium hydrogen phosphate or brushite (betabsh) and uric acid (betaUA) were also calculated by means of the computer program LithoRisk. Brand's test for cystinuria was also carried out. Statistical analysis was performed using the S-PSS software v. 22.0. **RESULTS**: The number of patients with data available for analysis was 435, of whom 236 were male (54%) and 199 female (46%). Complete 24-hour urine measurements were available for all 435 patients. Compared to men, women had significantly lower values for creatinine, urate, oxalate, phosphate, sodium, potassium, magnesium and chloride excretion, whereas 24-hour pH and citrate excretion were higher. No significant differences were found for the other examined variables. betaCaOx and betaUA were significantly higher in men than women, whereas no significant difference was found for betabsh. There was a direct relationship between calcium and sodium urine excretion. Excessive sodium excretion was recorded in 191 patients (44%) and low urine volumes in 201 (46.2%). Hyperoxaluria was observed in 118 patients (27.3%), hypercalciuria in 115 (26.6%), hyperuricosuria in 153 (35.4%), hypomagnesuria in 96 (22.2%), and hypocitraturia in 134 patients (31%). Hyperexcretion of sodium, hypocitraturia and hyperoxaluria were most frequent in males. betaCaOx was significantly higher in the setting of hypercalciuria, hypocitraturia, hyperoxaluria and urine pH below 5.5. **CONCLUSIONS**: Our findings in a large cohort of high-risk stone-forming patients show significant differences in urinary metabolic profiles between men and women. Carrying on the collection and analysis of data by LithoTest R from 2009 to 2015 and matching urinary and dietary data could eventually improve our understanding on the metabolic profile of stone-formers in Italy.

**URL:** <https://ovidsp.ovid.com/ovidweb.cgi?T=JS&CSC=Y&NEWS=N&PAGE=fulltext&D=med15&DO=10.23736%2fS0393-2249.17.02961-7> <https://libkey.io/libraries/2592/openurl?genre=article&sid=OVID:medline&id=pmid:28882031&id=doi:10.23736%2FS0393-2249.17.02961-7&issn=0393-2249&isbn=&volume=70&issue=1&spage=87&pages=87-94&date=2018&title=Minerva+Urologica+e+Nefrologica&atitle=A+multiregional+Italian+cohort+of+24-hour+urine+metabolic+evaluation+in+renal+stone+formers.&aulast=Esperto&pid=%3Cauthor%3EEsperto+F%3BMarangella+M%3BTrinchieri+A%3BPetrarulo+M%3BMiano+R%3C%2Fauthor%3E%3CAN%3E28882031%3C%2FAN%3E%3CDT%3EJournal+Article%3C%2FDT%3E>

71. **Intake of Trace Metals and the Risk of Incident Kidney Stones.**

**Item Type:** Journal Article

**Authors:** Ferraro, Pietro Manuel;Gambaro, Giovanni;Curhan, Gary C. and Taylor, Eric N.

**Publication Date:** 2018

**Journal:** Journal of Urology 199(6), pp. 1534–1539

**Abstract: PURPOSE**: The association between the intake of trace metals and the risk of incident stones has not been longitudinally investigated. **MATERIALS AND METHODS**: We performed a prospective analysis of 193,551 participants in the Health Professionals Follow-up Study, and the Nurses' Health Study I and II. During a followup of 3,316,580 person-years there was a total of 6,576 incident stones. We used multivariate regression models to identify associations of the intake of zinc, iron, copper and manganese with the risk of stones. In a subset of participants with 24-hour urine collections we examined the association between the intake of trace metals and urine composition. **RESULTS**: After multivariate adjustment total and dietary intakes of zinc and iron were not significantly associated with incident stones. A higher intake of manganese was associated with a lower risk of stones. The pooled HR of the highest quintile of total manganese intake compared with the lowest intake was 0.82 (95% CI 0.68-0.98, p = 0.02). Total but not dietary copper intake was marginally associated with a higher risk of stones (pooled HR 1.14, 95% CI 1.02-1.28, p = 0.01). There were no statistically significant associations of the total intake of manganese and copper with urinary supersaturation. **CONCLUSIONS**: Zinc and iron intake was not associated with a risk of stones. Copper intake may be associated with a higher risk in some individuals. Higher total manganese intake was associated with a lower risk of stones but not with traditional 24-hour urinary composite markers of stone risk. Further research is needed to elucidate the mechanisms by which manganese may reduce kidney stone formation. Copyright © 2018 American Urological Association Education and Research, Inc. Published by Elsevier Inc. All rights reserved.

**URL:** <https://ovidsp.ovid.com/ovidweb.cgi?T=JS&CSC=Y&NEWS=N&PAGE=fulltext&D=med15&DO=10.1016%2fj.juro.2018.01.077> <https://libkey.io/libraries/2592/openurl?genre=article&sid=OVID:medline&id=pmid:29391176&id=doi:10.1016%2Fj.juro.2018.01.077&issn=0022-5347&isbn=&volume=199&issue=6&spage=1534&pages=1534-1539&date=2018&title=Journal+of+Urology&atitle=Intake+of+Trace+Metals+and+the+Risk+of+Incident+Kidney+Stones.&aulast=Ferraro&pid=%3Cauthor%3EFerraro+PM%3BGambaro+G%3BCurhan+GC%3BTaylor+EN%3C%2Fauthor%3E%3CAN%3E29391176%3C%2FAN%3E%3CDT%3EJournal+Article%3C%2FDT%3E>

72. **Short-Term Changes in Urinary Relative Supersaturation Predict Recurrence of Kidney Stones: A Tool to Guide Preventive Measures in Urolithiasis.**

**Item Type:** Journal Article

**Authors:** Ferraro, Pietro Manuel;Ticinesi, Andrea;Meschi, Tiziana;Rodgers, Allen;Di Maio, Federica;Fulignati, Pierluigi;Borghi, Loris and Gambaro, Giovanni

**Publication Date:** 2018

**Journal:** Journal of Urology 200(5), pp. 1082–1087

**Abstract: PURPOSE**: Kidney stone disease is characterized by a relatively high rate of recurrence. In our study we analyzed the association between relative supersaturation and the risk of stone recurrence. Additionally, we examined the association between the risk of recurrence and changes in relative supersaturation and urinary composition after 1 week of medical treatment. **MATERIALS AND METHODS**: We performed a post hoc analysis of data from a previously published randomized controlled trial comparing the effect of 2 diets in 120 men with recurrent calcium oxalate stones and hypercalciuria. Baseline and followup 24-hour urine parameters were used to calculate the relative supersaturation of calcium oxalate, calcium phosphate and uric acid using the EQUIL2, JESS and LithoRisk computer programs. Cox models were used to calculate the estimated association between each baseline relative supersaturation, and 1-week changes and the risk of recurrence during followup. **RESULTS**: During a 5-year followup 35 patients (34%) experienced recurrence. A reduction in calcium oxalate relative supersaturation at 1 week was significantly associated with a lower risk of recurrence using the EQUIL2 calculation (for every 10% reduction from baseline HR 0.92, 95% CI 0.86-1.00, p = 0.044). However, there was no association for relative supersaturation calculated by other methods or for the relative supersaturation of other salts. Changes in the 24-hour urine excretion of citrate, potassium and magnesium were significantly associated with a risk of recurrence. **CONCLUSIONS**: In recurrent stone formers with hypercalciuria baseline values and changes in the relative supersaturation of calcium oxalate may be associated with the risk of recurrence. Changes in urinary citrate, potassium and magnesium following dietary intervention may also be predictive. Copyright © 2018 American Urological Association Education and Research, Inc. Published by Elsevier Inc. All rights reserved.

**URL:** <https://ovidsp.ovid.com/ovidweb.cgi?T=JS&CSC=Y&NEWS=N&PAGE=fulltext&D=med15&DO=10.1016%2fj.juro.2018.06.029> <https://libkey.io/libraries/2592/openurl?genre=article&sid=OVID:medline&id=pmid:29940247&id=doi:10.1016%2Fj.juro.2018.06.029&issn=0022-5347&isbn=&volume=200&issue=5&spage=1082&pages=1082-1087&date=2018&title=Journal+of+Urology&atitle=Short-Term+Changes+in+Urinary+Relative+Supersaturation+Predict+Recurrence+of+Kidney+Stones%3A+A+Tool+to+Guide+Preventive+Measures+in+Urolithiasis.&aulast=Ferraro&pid=%3Cauthor%3EFerraro+PM%3BTicinesi+A%3BMeschi+T%3BRodgers+A%3BDi+Maio+F%3BFulignati+P%3BBorghi+L%3BGambaro+G%3C%2Fauthor%3E%3CAN%3E29940247%3C%2FAN%3E%3CDT%3EEvaluation+Study%3C%2FDT%3E>

73. **Do Urinary Cystine Parameters Predict Clinical Stone Activity?.**

**Item Type:** Journal Article

**Authors:** Friedlander, Justin I.;Antonelli, Jodi A.;Canvasser, Noah E.;Morgan, Monica S. C.;Mollengarden, Daniel;Best, Sara and Pearle, Margaret S.

**Publication Date:** 2018

**Journal:** Journal of Urology 199(2), pp. 495–499

**Abstract: PURPOSE**: An accurate urinary predictor of stone recurrence would be clinically advantageous for patients with cystinuria. A proprietary assay (Litholink, Chicago, Illinois) measures cystine capacity as a potentially more reliable estimate of stone forming propensity. The recommended capacity level to prevent stone formation, which is greater than 150 mg/l, has not been directly correlated with clinical stone activity. We investigated the relationship between urinary cystine parameters and clinical stone activity. **MATERIALS AND METHODS**: We prospectively followed 48 patients with cystinuria using 24-hour urine collections and serial imaging, and recorded stone activity. We compared cystine urinary parameters at times of stone activity with those obtained during periods of stone quiescence. We then performed correlation and ROC analysis to evaluate the performance of cystine parameters to predict stone activity. **RESULTS**: During a median followup of 70.6 months (range 2.2 to 274.6) 85 stone events occurred which could be linked to a recent urine collection. Cystine capacity was significantly greater for quiescent urine than for stone event urine (mean +/- SD 48 +/- 107 vs -38 +/- 163 mg/l, p : During a median followup of 70.6 months (range 2.2 to 274.6) 85 stone events occurred which could be linked to a recent urine collection. Cystine capacity was significantly greater for quiescent urine than for stone event urine (mean +/- SD 48 +/- 107 vs -38 +/- 163 mg/l, p : During a median followup of 70.6 months (range 2.2 to 274.6) 85 stone events occurred which could be linked to a recent urine collection. Cystine capacity was significantly greater for quiescent urine than for stone event urine (mean +/- SD 48 +/- 107 vs -38 +/- 163 mg/l, p : During a median followup of 70.6 months (range 2.2 to 274.6) 85 stone events occurred which could be linked to a recent urine collection. Cystine capacity was significantly greater for quiescent urine than for stone event urine (mean +/- SD 48 +/- 107 vs -38 +/- 163 mg/l, p : During a median followup of 70.6 months (range 2.2 to 274.6) 85 stone events occurred which could be linked to a recent urine collection. Cystine capacity was significantly greater for quiescent urine than for stone event urine (mean +/- SD 48 +/- 107 vs -38 +/- 163 mg/l, p **CONCLUSIONS**: Our results suggest that the target for capacity should be lower than previously advised. Copyright © 2018 American Urological Association Education and Research, Inc. Published by Elsevier Inc. All rights reserved.

**URL:** <https://ovidsp.ovid.com/ovidweb.cgi?T=JS&CSC=Y&NEWS=N&PAGE=fulltext&D=med15&DO=10.1016%2fj.juro.2017.09.034> <https://libkey.io/libraries/2592/openurl?genre=article&sid=OVID:medline&id=pmid:28916274&id=doi:10.1016%2Fj.juro.2017.09.034&issn=0022-5347&isbn=&volume=199&issue=2&spage=495&pages=495-499&date=2018&title=Journal+of+Urology&atitle=Do+Urinary+Cystine+Parameters+Predict+Clinical+Stone+Activity%3F.&aulast=Friedlander&pid=%3Cauthor%3EFriedlander+JI%3BAntonelli+JA%3BCanvasser+NE%3BMorgan+MSC%3BMollengarden+D%3BBest+S%3BPearle+MS%3C%2Fauthor%3E%3CAN%3E28916274%3C%2FAN%3E%3CDT%3EClinical+Trial%3C%2FDT%3E>

74. **Quantitative Mineralogical Composition of Calculi and Urine Abnormalities for Calcium Oxalate Stone Formers: A Single-Center Results.**

**Item Type:** Journal Article

**Authors:** Kustov, Andrey V. and Strelnikov, Alexander I.

**Publication Date:** 2018

**Journal:** Urology Journal 15(3), pp. 87–91

**Abstract: PURPOSE**: The paper focuses on the relationship of risk factors and metabolic disorders with mineralogical composition of calculi, age and gender of calcium oxalate stone formers. **MATERIALS AND METHODS**: Stone mineralogical composition, 24 hour biochemistry and pH-profile of urine were examined for sixty four stone formers using powder X-ray diffraction, spectrophotometric and potentiometric techniques. **RESULTS**: The analysis indicated that 44 % of calculi were composed of pure calcium oxalate monohydrate, whereas other 56 % contained both monohydrate and dihydrate or usually their mixtures with hydroxyl apatite. Hypocitraturia, hypercalciuria and hyperuricosuria were identified as the most frequent disorders. Patients with pure calcium oxalate stones and calcium oxalate mixed with apatite revealed different patterns including age, acid-base balance of urine, calcium, citrate excretion etc.

**URL:** <https://ovidsp.ovid.com/ovidweb.cgi?T=JS&CSC=Y&NEWS=N&PAGE=fulltext&D=med15&DO=10.22037%2fuj.v0i0.3910> <https://libkey.io/libraries/2592/openurl?genre=article&sid=OVID:medline&id=pmid:29277881&id=doi:10.22037%2Fuj.v0i0.3910&issn=1735-1308&isbn=&volume=15&issue=3&spage=87&pages=87-91&date=2018&title=Urology+Journal&atitle=Quantitative+Mineralogical+Composition+of+Calculi+and+Urine+Abnormalities+for+Calcium+Oxalate+Stone+Formers%3A+A+Single-Center+Results.&aulast=Kustov&pid=%3Cauthor%3EKustov+AV%3BStrelnikov+AI%3C%2Fauthor%3E%3CAN%3E29277881%3C%2FAN%3E%3CDT%3EJournal+Article%3C%2FDT%3E>

75. **Successful Diabetic Control as Measured by Hemoglobin A1c Is Associated with Lower Urine Risk Factors for Uric Acid Calculi.**

**Item Type:** Journal Article

**Authors:** Maciolek, Kimberly A.;Penniston, Kristina L.;Jhagroo, Roy Allan and Best, Sara L.

**Publication Date:** 2018

**Journal:** Journal of Endourology 32(8), pp. 771–776

**Abstract: INTRODUCTION AND OBJECTIVES**: To examine the association of glycemic control, including strict glycemic control, with 24-hour urine risk factors for uric acid and calcium calculi. **MATERIALS AND METHODS**: With institutional review board (IRB) approval, we identified 183 stone formers (SFs) with 459 twenty-four-hour urine collections. Hemoglobin A1c (HgbA1c) measures were obtained within 3 months of the urine collection. Collections were categorized into normoglycemic (NG, HgbA1c = 6.5) cohorts; 24-hour urine parameters were compared. The NG cohort was further divided into patients with and without a history of diabetes mellitus (DM) type 2. Variables were analyzed using chi-square, Welch's t-test and multivariate linear regression to adjust for clustering, body mass index (BMI), age, gender, thiazide use, and potassium citrate use. **RESULTS**: Patients in the HG group were older with higher BMI. Multivariate analysis of the total study population revealed that hyperglycemia correlated with lower pH, higher uric acid relative saturation (RS), lower brushite RS, and higher citrate. NG SFs with and without a history of DM had similar risk factors for uric acid stone formation. Among NG SFs, those with DM had higher urine calcium and calcium oxalate RS than those without DM. However, this difference may be related to other factors since neither parameter correlated with DM on multivariate regression (p > 0.05). **CONCLUSIONS**: Successful glycemic control may be associated with reduced urinary risk factors for uric acid stone formation. Patients with well-controlled DM had equivalent risk factors to those without DM. Glycemic control should be considered a target of the multidisciplinary medical management of stone disease.

**URL:** <https://ovidsp.ovid.com/ovidweb.cgi?T=JS&CSC=Y&NEWS=N&PAGE=fulltext&D=med15&DO=10.1089%2fend.2018.0376> <https://libkey.io/libraries/2592/openurl?genre=article&sid=OVID:medline&id=pmid:29896970&id=doi:10.1089%2Fend.2018.0376&issn=0892-7790&isbn=&volume=32&issue=8&spage=771&pages=771-776&date=2018&title=Journal+of+Endourology&atitle=Successful+Diabetic+Control+as+Measured+by+Hemoglobin+A1c+Is+Associated+with+Lower+Urine+Risk+Factors+for+Uric+Acid+Calculi.&aulast=Maciolek&pid=%3Cauthor%3EMaciolek+KA%3BPenniston+KL%3BJhagroo+RA%3BBest+SL%3C%2Fauthor%3E%3CAN%3E29896970%3C%2FAN%3E%3CDT%3EJournal+Article%3C%2FDT%3E>

76. **Flexible ureterorenoscopy is associated with less stone recurrence rates over Shockwave lithotripsy in the management of 10-20 millimeter lower pole renal stone: medium follow-up results.**

**Item Type:** Journal Article

**Authors:** Ozgor, Faruk;Sahan, Murat;Yanaral, Fatih;Savun, Metin and Sarilar, Omer

**Publication Date:** 2018

**Journal:** International Braz J Urol 44(2), pp. 314–322

**Abstract: PURPOSE**: To identify the role of shock wave lithotripsy (SWL) and flexible ureterorenoscopy (f-URS) on the stone recurrence, in the management of 10-20 millimeter lower pole stone (LPS) with medium follow-up outcomes. **MATERIALS AND METHODS**: The patients' charts which were treated with SWL or f-URS for LPS between January 2011 and September 2013 were analyzed, retrospectively. Patients who had a solitary 10-20mm LPS were enrolled into the study. In both procedures, patient was accepted as stone free, if complete stone clearance was achieved in the 3rd month abdominal computed tomography. Only patients with a stone free status were evaluated in follow-ups. **RESULTS**: The stone-free rate was 77.9% (88/113 patients) for the SWL group and 89% (114/128 patients) for the f-URS group (p=0.029). Stone recurrence was detected in 28 (35.4%) patients in SWL group and in 17 (17.2%) patients in f-URS group (p=0.009). Stone types and 24 hour urine sample results were similar between groups (p=0.123 vs p=0.197, respectively). Multivariate regression analysis revealed that f-URS procedure and absence of abnormality in 24 hour urine analysis significantly decreased stone recurrence in medium term follow-up (p=0.001 and p: The stone-free rate was 77.9% (88/113 patients) for the SWL group and 89% (114/128 patients) for the f-URS group (p=0.029). Stone recurrence was detected in 28 (35.4%) patients in SWL group and in 17 (17.2%) patients in f-URS group (p=0.009). Stone types and 24 hour urine sample results were similar between groups (p=0.123 vs p=0.197, respectively). Multivariate regression analysis revealed that f-URS procedure and absence of abnormality in 24 hour urine analysis significantly decreased stone recurrence in medium term follow-up (p=0.001 and p**CONCLUSIONS**: Our study showed for the first time, that patients which underwent f-URS for LPS, faced less stone recurrence, independent from diet regimen and metabolic evaluation in medium term follow-up. Additionally, presence of abnormality in 24 hour urine analysis increase the stone recurrence risk in follow-ups. Copyright R by the International Brazilian Journal of Urology.

**URL:** <https://ovidsp.ovid.com/ovidweb.cgi?T=JS&CSC=Y&NEWS=N&PAGE=fulltext&D=med15&DO=10.1590%2fS1677-5538.IBJU.2017.0483> <https://libkey.io/libraries/2592/openurl?genre=article&sid=OVID:medline&id=pmid:29412548&id=doi:10.1590%2FS1677-5538.IBJU.2017.0483&issn=1677-5538&isbn=&volume=44&issue=2&spage=314&pages=314-322&date=2018&title=International+Braz+J+Urol&atitle=Flexible+ureterorenoscopy+is+associated+with+less+stone+recurrence+rates+over+Shockwave+lithotripsy+in+the+management+of+10-20+millimeter+lower+pole+renal+stone%3A+medium+follow-up+results.&aulast=Ozgor&pid=%3Cauthor%3EOzgor+F%3BSahan+M%3BYanaral+F%3BSavun+M%3BSarilar+O%3C%2Fauthor%3E%3CAN%3E29412548%3C%2FAN%3E%3CDT%3EComparative+Study%3C%2FDT%3E>

77. **Menopause and Risk of Kidney Stones.**

**Item Type:** Journal Article

**Authors:** Prochaska, Megan;Taylor, Eric N. and Curhan, Gary

**Publication Date:** 2018

**Journal:** Journal of Urology 200(4), pp. 823–828

**Abstract: PURPOSE**: Metabolic changes due to menopause may alter urine composition and kidney stone risk but results of prior work on this association have been mixed. We examined menopause and the risk of incident kidney stones, and changes in 24-hour urine composition in the NHS (Nurses' Health Study) II. **MATERIALS AND METHODS**: Using multivariate adjusted Cox proportional hazards models we prospectively analyzed 108,639 NHS II participants who provided information on menopause and kidney stones. We also analyzed 24-hour urine collections from 658 participants who performed a collection while premenopausal and a repeat collection after menopause. **RESULTS**: During 22 years of followup there were 3,456 incident kidney stones. The multivariate adjusted relative risk of an incident kidney stone in postmenopausal participants compared with premenopause was 1.27 (95% CI 1.08-1.46). On stratified analysis compared with premenopause the multivariate adjusted relative risk of natural and surgically induced menopause was 1.27 (95% CI 1.09-1.48) and 1.43 (95% CI 1.19-1.73), respectively. Among the 74,505 postmenopausal participants there was a total of 1,041 incident stone events. Compared with no hormone therapy neither current nor past use was significantly associated with kidney stone risk. Compared with premenopause the postmenopausal urine collections had lower mean calcium, citrate, phosphorus and uric acid, and higher mean volume. **CONCLUSIONS**: Postmenopausal status is associated with a higher risk of incident kidney stones. Natural menopause and surgical menopause are independently associated with higher risk. There are small but significant differences in urine composition between premenopausal and postmenopausal urine collections. Copyright © 2018 American Urological Association Education and Research, Inc. Published by Elsevier Inc. All rights reserved.

**URL:** <https://ovidsp.ovid.com/ovidweb.cgi?T=JS&CSC=Y&NEWS=N&PAGE=fulltext&D=med15&DO=10.1016%2fj.juro.2018.04.080> <https://libkey.io/libraries/2592/openurl?genre=article&sid=OVID:medline&id=pmid:29730204&id=doi:10.1016%2Fj.juro.2018.04.080&issn=0022-5347&isbn=&volume=200&issue=4&spage=823&pages=823-828&date=2018&title=Journal+of+Urology&atitle=Menopause+and+Risk+of+Kidney+Stones.&aulast=Prochaska&pid=%3Cauthor%3EProchaska+M%3BTaylor+EN%3BCurhan+G%3C%2Fauthor%3E%3CAN%3E29730204%3C%2FAN%3E%3CDT%3EComparative+Study%3C%2FDT%3E>

78. **Relative Supersaturation of 24-Hour Urine and Likelihood of Kidney Stones.**

**Item Type:** Journal Article

**Authors:** Prochaska, Megan;Taylor, Eric;Ferraro, Pietro Manuel and Curhan, Gary

**Publication Date:** 2018

**Journal:** Journal of Urology 199(5), pp. 1262–1266

**Abstract: PURPOSE**: The relative supersaturation of calcium oxalate, calcium phosphate and uric acid is used clinically in kidney stone prevention. The magnitude of the association between relative supersaturation and stone risk requires further quantification. **MATERIALS AND METHODS**: We performed a cross-sectional study using 24-hour urine collections from the NHS (Nurses' Health Study) I and II, and HPFS (Health Professionals Follow-up Study) cohorts to quantify the association between the relative supersaturation of calcium oxalate, calcium phosphate and uric acid, and the likelihood of stone formation. **RESULTS**: The OR of being a stone former was 5.85 (95% CI 3.40-10.04) in NHS I, 6.38 (95% CI 3.72-11.0) in NHS II and 6.95 (95% CI 3.56-13.6) in HPFS for the highest category of calcium oxalate relative supersaturation compared with less than 1.0. The OR of being a stone former was 1.86 (95% CI 0.94-3.71) in NHS I, 4.37 (95% CI 2.68-7.10) in NHS II and 3.59 (95% CI 2.04-6.31) in HPFS for the highest category of calcium phosphate relative supersaturation compared with less than 1.0. For uric acid relative supersaturation the OR of being a stone former was 4.30 (95% CI 2.34-7.90) in NHS I and 2.74 (95% CI 1.71-4.40) in NHS II for the highest relative supersaturation category compared with less than 1.0. In HPFS the uric acid relative supersaturation was not significantly associated with the likelihood of stone formation. **CONCLUSIONS**: The likelihood of being a stone former increases with higher relative supersaturation of calcium oxalate and calcium phosphate in men and women, and with higher relative supersaturation of uric acid in women. Copyright © 2018 American Urological Association Education and Research, Inc. Published by Elsevier Inc. All rights reserved.

**URL:** <https://ovidsp.ovid.com/ovidweb.cgi?T=JS&CSC=Y&NEWS=N&PAGE=fulltext&D=med15&DO=10.1016%2fj.juro.2017.10.046> <https://libkey.io/libraries/2592/openurl?genre=article&sid=OVID:medline&id=pmid:29132983&id=doi:10.1016%2Fj.juro.2017.10.046&issn=0022-5347&isbn=&volume=199&issue=5&spage=1262&pages=1262-1266&date=2018&title=Journal+of+Urology&atitle=Relative+Supersaturation+of+24-Hour+Urine+and+Likelihood+of+Kidney+Stones.&aulast=Prochaska&pid=%3Cauthor%3EProchaska+M%3BTaylor+E%3BFerraro+PM%3BCurhan+G%3C%2Fauthor%3E%3CAN%3E29132983%3C%2FAN%3E%3CDT%3EJournal+Article%3C%2FDT%3E>

79. **Effect of phyllanthus niruri on metabolic parameters of patients with kidney stone: a perspective for disease prevention.**

**Item Type:** Journal Article

**Authors:** Pucci, Nidia D.;Marchini, Giovanni S.;Mazzucchi, Eduardo;Reis, Sabrina T.;Srougi, Miguel;Evazian, Denise and Nahas, William C.

**Publication Date:** 2018

**Journal:** International Braz J Urol 44(4), pp. 758–764

**Abstract:** Phyllanthus niruri (P.niruri) or stone breaker is a plant commonly used to reduce stone risk, however, clinical studies on this issue are lacking. **OBJECTIVE**: To prospectively evaluate the effect of P. niruri on the urinary metabolic parameters of patients with urinary lithiasis. **MATERIALS AND METHODS**: We studied 56 patients with kidney stones : We studied 56 patients with kidney stones **RESULTS**: Mean age was 44+/-9.2 and BMI was 27.2+/-4.4kg/m2. Thirty-six patients (64%) were women. There were no significant changes in all periods for anthropometric and several serum measurements, including total blood count, creatinine, uric acid, sodium, potassium, calcium, urine volume and pH; a significant increase in urinary potassium from 50.5+/-20.4 to 56.2+/-21.8 mg/24-hour (p=0.017); magnesium/creatinine ratio 58+/-22.5 to 69.1+/-28.6mg/ gCr24-hour (p=0.013) and potassium/creatinine ratio 39.3+/-15.1 to 51.3+/-34.7mg/gCr24- hour (p=0.008) from baseline to wash out. The kidney stones decreased from 3.2+/-2 to 2.0+/-2per patient (p: Mean age was 44+/-9.2 and BMI was 27.2+/-4.4kg/m2. Thirty-six patients (64%) were women. There were no significant changes in all periods for anthropometric and several serum measurements, including total blood count, creatinine, uric acid, sodium, potassium, calcium, urine volume and pH; a significant increase in urinary potassium from 50.5+/-20.4 to 56.2+/-21.8 mg/24-hour (p=0.017); magnesium/creatinine ratio 58+/-22.5 to 69.1+/-28.6mg/ gCr24-hour (p=0.013) and potassium/creatinine ratio 39.3+/-15.1 to 51.3+/-34.7mg/gCr24- hour (p=0.008) from baseline to wash out. The kidney stones decreased from 3.2+/-2 to 2.0+/-2per patient (p**CONCLUSIONS**: P.niruri intake is safe and does not cause significant adverse effects on serum metabolic parameters. It increases urinary excretion of magnesium and potassium caused a significant decrease in urinary oxalate and uric acid in patients with hyperoxaluria and hyperuricosuria. The consumption of P.niruri contributed to the elimination of urinary calculi. Copyright R by the International Brazilian Journal of Urology.

**URL:** <https://ovidsp.ovid.com/ovidweb.cgi?T=JS&CSC=Y&NEWS=N&PAGE=fulltext&D=med15&DO=10.1590%2fS1677-5538.IBJU.2017.0521> <https://libkey.io/libraries/2592/openurl?genre=article&sid=OVID:medline&id=pmid:29617079&id=doi:10.1590%2FS1677-5538.IBJU.2017.0521&issn=1677-5538&isbn=&volume=44&issue=4&spage=758&pages=758-764&date=2018&title=International+Braz+J+Urol&atitle=Effect+of+phyllanthus+niruri+on+metabolic+parameters+of+patients+with+kidney+stone%3A+a+perspective+for+disease+prevention.&aulast=Pucci&pid=%3Cauthor%3EPucci+ND%3BMarchini+GS%3BMazzucchi+E%3BReis+ST%3BSrougi+M%3BEvazian+D%3BNahas+WC%3C%2Fauthor%3E%3CAN%3E29617079%3C%2FAN%3E%3CDT%3EJournal+Article%3C%2FDT%3E>

80. **Optimization of urinary dipstick pH: Are multiple dipstick pH readings reliably comparable to commercial 24-hour urinary pH?.**

**Item Type:** Journal Article

**Authors:** Abbott, Joel E.;Miller, Daniel L.;Shi, William;Wenzler, David;Elkhoury, Fuad F.;Patel, Nishant D. and Sur, Roger L.

**Publication Date:** 2017

**Journal:** Investigative and Clinical Urology 58(5), pp. 378–382

**Abstract: PURPOSE**: Accurate measurement of pH is necessary to guide medical management of nephrolithiasis. Urinary dipsticks offer a convenient method to measure pH, but prior studies have only assessed the accuracy of a single, spot dipstick. Given the known diurnal variation in pH, a single dipstick pH is unlikely to reflect the average daily urinary pH. Our goal was to determine whether multiple dipstick pH readings would be reliably comparable to pH from a 24-hour urine analysis. **MATERIALS AND METHODS**: Kidney stone patients undergoing a 24-hour urine collection were enrolled and took images of dipsticks from their first 3 voids concurrently with the 24-hour collection. Images were sent to and read by a study investigator. The individual and mean pH from the dipsticks were compared to the 24-hour urine pH and considered to be accurate if the dipstick readings were within 0.5 of the 24-hour urine pH. The Bland-Altman test of agreement was used to further compare dipstick pH relative to 24-hour urine pH. **RESULTS**: Fifty-nine percent of patients had mean urinary pH values within 0.5 pH units of their 24-hour urine pH. Bland-Altman analysis showed a mean difference between dipstick pH and 24-hour urine pH of -0.22, with an upper limit of agreement of 1.02 (95% confidence interval [CI], 0.45-1.59) and a lower limit of agreement of -1.47 (95% CI, -2.04 to -0.90). **CONCLUSIONS**: We concluded that urinary dipstick based pH measurement lacks the precision required to guide medical management of nephrolithiasis and physicians should use 24-hour urine analysis to base their metabolic therapy.

**URL:** <https://ovidsp.ovid.com/ovidweb.cgi?T=JS&CSC=Y&NEWS=N&PAGE=fulltext&D=med14&DO=10.4111%2ficu.2017.58.5.378> <https://libkey.io/libraries/2592/openurl?genre=article&sid=OVID:medline&id=pmid:28868511&id=doi:10.4111%2Ficu.2017.58.5.378&issn=2466-0493&isbn=&volume=58&issue=5&spage=378&pages=378-382&date=2017&title=Investigative+And+Clinical+Urology&atitle=Optimization+of+urinary+dipstick+pH%3A+Are+multiple+dipstick+pH+readings+reliably+comparable+to+commercial+24-hour+urinary+pH%3F.&aulast=Abbott&pid=%3Cauthor%3EAbbott+JE%3BMiller+DL%3BShi+W%3BWenzler+D%3BElkhoury+FF%3BPatel+ND%3BSur+RL%3C%2Fauthor%3E%3CAN%3E28868511%3C%2FAN%3E%3CDT%3EComparative+Study%3C%2FDT%3E>

81. **Do Overweight and Obese Pediatric Stone Formers Have Differences in Metabolic Abnormalities Compared With Normal-weight Stone Formers?.**

**Item Type:** Journal Article

**Authors:** Cambareri, Gina M.;Giel, Dana W.;Bayne, Aaron P.;Corbett, Sean;Schurtz, Elleson;Kovacevic, Larisa;Sukhu, Troy;Yap, Michael and Chiang, George

**Publication Date:** Ma ,2017

**Journal:** Urology 101, pp. 26–30

**Abstract: OBJECTIVE**: To determine if 24-hour urinary parameters in children with nephrolithiasis across 4 institutions were influenced by body mass index (BMI). **MATERIALS AND METHODS**: The 24-hour urinary parameters obtained from children with nephrolithiasis between 2000 and 2013 were stratified by BMI percentile >=85th and : The 24-hour urinary parameters obtained from children with nephrolithiasis between 2000 and 2013 were stratified by BMI percentile >=85th and **RESULTS**: Overweight and obese patients consisted of 35.4% of the cohort (n = 73). Metabolic abnormalities were present in 130 children (63.1%). The most common abnormality present in the =85th percentile, hyperoxaluria (37.0%). Univariable and multivariable analyses revealed that overweight and obese children were more likely to have low urinary volume and elevated uric acid compared to normal-weight children. **CONCLUSION**: Although there is a link between stone formation and BMI in adults, no definitive conclusions have been proven in the pediatric literature. Our study indicates that stone-forming children who are overweight or obese have low urinary volume and elevated uric acid compared to normal-weight stone-forming children. Copyright © 2016 Elsevier Inc. All rights reserved.

**URL:** <https://ovidsp.ovid.com/ovidweb.cgi?T=JS&CSC=Y&NEWS=N&PAGE=fulltext&D=med14&DO=10.1016%2fj.urology.2016.09.011> <https://libkey.io/libraries/2592/openurl?genre=article&sid=OVID:medline&id=pmid:27641936&id=doi:10.1016%2Fj.urology.2016.09.011&issn=0090-4295&isbn=&volume=101&issue=&spage=26&pages=26-30&date=2017&title=Urology&atitle=Do+Overweight+and+Obese+Pediatric+Stone+Formers+Have+Differences+in+Metabolic+Abnormalities+Compared+With+Normal-weight+Stone+Formers%3F.&aulast=Cambareri&pid=%3Cauthor%3ECambareri+GM%3BGiel+DW%3BBayne+AP%3BCorbett+S%3BSchurtz+E%3BKovacevic+L%3BSukhu+T%3BYap+M%3BChiang+G%3C%2Fauthor%3E%3CAN%3E27641936%3C%2FAN%3E%3CDT%3EComparative+Study%3C%2FDT%3E>

82. **Vitamin D Repletion in Kidney Stone Formers: A Randomized Controlled Trial.**

**Item Type:** Journal Article

**Authors:** Ferroni, Matthew C.;Rycyna, Kevin J.;Averch, Timothy D. and Semins, Michelle J.

**Publication Date:** 2017

**Journal:** Journal of Urology 197(4), pp. 1079–1083

**Abstract: PURPOSE**: Vitamin D deficiency is often detected during metabolic evaluation in the nephrolithiasis population. Multiple vitamin D repletion protocols exist, although their differing impact on urinary stone formation risk factors is unclear. **MATERIALS AND METHODS**: Patients with a history of calcium stones and vitamin D deficiency (less than 30 ng/ml) were randomized to receive either 1,000 IU daily or 50,000 IU weekly of vitamin D supplementation for 6 weeks. Patients completed a pretreatment and posttreatment serum vitamin D level evaluation and 24-hour urine collections to assess the response and any changes in urine stone formation risk parameters. **RESULTS**: A total of 21 patients completed the study, including 8 who received 1,000 IU daily and 13 who received 50,000 IU weekly. The 50,000 IU weekly group showed a significant increase in median serum vitamin D levels of 23 ng/ml (135%, p : A total of 21 patients completed the study, including 8 who received 1,000 IU daily and 13 who received 50,000 IU weekly. The 50,000 IU weekly group showed a significant increase in median serum vitamin D levels of 23 ng/ml (135%, p **CONCLUSIONS**: High dose and low dose vitamin D repletion had no effect on urine calcium excretion or the supersaturation of calcium salts in known stone formers. The higher dosing regimen, which had superior repletion, may be the optimal protocol in patients with vitamin D deficiency. Copyright © 2017 American Urological Association Education and Research, Inc. Published by Elsevier Inc. All rights reserved.

**URL:** <https://ovidsp.ovid.com/ovidweb.cgi?T=JS&CSC=Y&NEWS=N&PAGE=fulltext&D=med14&DO=10.1016%2fj.juro.2016.10.057> <https://libkey.io/libraries/2592/openurl?genre=article&sid=OVID:medline&id=pmid:27765695&id=doi:10.1016%2Fj.juro.2016.10.057&issn=0022-5347&isbn=&volume=197&issue=4&spage=1079&pages=1079-1083&date=2017&title=Journal+of+Urology&atitle=Vitamin+D+Repletion+in+Kidney+Stone+Formers%3A+A+Randomized+Controlled+Trial.&aulast=Ferroni&pid=%3Cauthor%3EFerroni+MC%3BRycyna+KJ%3BAverch+TD%3BSemins+MJ%3C%2Fauthor%3E%3CAN%3E27765695%3C%2FAN%3E%3CDT%3EJournal+Article%3C%2FDT%3E>

83. **Factors Associated with Compliance in Submitting 24-Hour Urine Collections in an Underserved Community.**

**Item Type:** Journal Article

**Authors:** Ghiraldi, Eric M.;Reddy, Madhumitha;Li, Tianyu;Lawler, Andrew C. and Friedlander, Justin I.

**Publication Date:** 2017

**Journal:** Journal of Endourology 31(S1), pp. S64–S68

**Abstract: PURPOSE**: Patients living in underserved areas do regularly express an interest in stone prevention; however, factors limiting participation, aside from obvious cost considerations, are largely unknown. To better understand factors associated with compliance with submitting 24-hour urine collections, we reviewed our patient experience at the kidney stone clinic at a hospital that provides care for an underserved urban community. **MATERIALS AND METHODS**: A retrospective chart review of patients treated for kidney and/or ureteral stones between August 2014 and May 2016 was performed. Patient demographics, medical characteristics, stone factors, and compliance data were compiled into our data set. Patients were divided into two groups: those who did and did not submit the requested initial 24-hour urine collection. Analysis of factors related to compliance was performed using univariate analysis and multivariate logistic regression. **RESULTS**: A total of 193 patients met inclusion criteria for our study, 42.5% (82/193) of whom submitted 24-hour urine samples. Of the 82 collections submitted, 34.1% (28/82) were considered inadequate by creatinine level. A second urine collection within 6 months was obtained in 14.0% (27/193) of patients. Univariate analysis demonstrated that African American (AA) patients were less likely to submit an initial 24-hour urine collection than Caucasian patients (collected: 30.9% vs 51.8%; p : A total of 193 patients met inclusion criteria for our study, 42.5% (82/193) of whom submitted 24-hour urine samples. Of the 82 collections submitted, 34.1% (28/82) were considered inadequate by creatinine level. A second urine collection within 6 months was obtained in 14.0% (27/193) of patients. Univariate analysis demonstrated that African American (AA) patients were less likely to submit an initial 24-hour urine collection than Caucasian patients (collected: 30.9% vs 51.8%; p : A total of 193 patients met inclusion criteria for our study, 42.5% (82/193) of whom submitted 24-hour urine samples. Of the 82 collections submitted, 34.1% (28/82) were considered inadequate by creatinine level. A second urine collection within 6 months was obtained in 14.0% (27/193) of patients. Univariate analysis demonstrated that African American (AA) patients were less likely to submit an initial 24-hour urine collection than Caucasian patients (collected: 30.9% vs 51.8%; p **CONCLUSIONS**: In our underserved patient population, AA patients were half as likely to submit a 24-hour urine collection than Caucasian patients, whereas patients with a positive family history of stones were more than twice as likely to submit than patients with no family history.

**URL:** <https://ovidsp.ovid.com/ovidweb.cgi?T=JS&CSC=Y&NEWS=N&PAGE=fulltext&D=med14&DO=10.1089%2fend.2016.0594> <https://libkey.io/libraries/2592/openurl?genre=article&sid=OVID:medline&id=pmid:27931114&id=doi:10.1089%2Fend.2016.0594&issn=0892-7790&isbn=&volume=31&issue=1&spage=S64&pages=S64-S68&date=2017&title=Journal+of+Endourology&atitle=Factors+Associated+with+Compliance+in+Submitting+24-Hour+Urine+Collections+in+an+Underserved+Community.&aulast=Ghiraldi&pid=%3Cauthor%3EGhiraldi+EM%3BReddy+M%3BLi+T%3BLawler+AC%3BFriedlander+JI%3C%2Fauthor%3E%3CAN%3E27931114%3C%2FAN%3E%3CDT%3EJournal+Article%3C%2FDT%3E>

84. **The Role of the 24-Hour Urine Collection in the Prevention of Kidney Stone Recurrence**

**Item Type:** Journal Article

**Authors:** Hsi, Ryan S.;Sanford, Thomas;Goldfarb, David S. and Stoller, Marshall L.

**Publication Date:** 2017

**Journal:** Journal of Urology 197(4), pp. 1084–1089

**Abstract: PURPOSE**: Kidney stone prevention relies on the 24-hour urine collection to diagnose metabolic abnormalities and direct dietary and pharmacological therapy. While its use is guideline supported for high risk and interested patients, evidence that the test can accurately predict recurrence or treatment response is limited. We sought to critically reassess the role of the 24-hour urine collection in stone prevention. **MATERIALS AND METHODS**: In addition to a MEDLINE R search to identify controlled studies of dietary and pharmacological interventions, evidence supporting the AUA (American Urological Association) and EAU (European Association of Urology) guidelines for metabolic stone prevention were evaluated. Additionally, the placebo arms of these studies were examined to assess the stone clinic effect, that is the impact of regular office visits without specific treatment on stone recurrence. **RESULTS**: The 24-hour urine test has several limitations, including the complexity of interpretation, the need for repeat collections, the inability to predict stone recurrence with individual parameters and supersaturation values, the unclear rationale of laboratory cutoff values and the difficulty of determining collection adequacy. Only 1 prospective trial has compared selective dietary recommendations based on 24-hour urine collection results vs general dietary instructions. While the trial supported the intervention arm, significant limitations to the study were found. Placebo arms of intervention trials have noted a 0% to 61% decrease in stone recurrence rate and a remission rate during the study of 20% to 86%. **CONCLUSIONS**: Whether all recurrent stone formers benefit from 24-hour urine collection has not been established. Additional comparative effectiveness trials are needed to determine which stone former benefits from selective therapy, as guided by the 24-hour urine collection. Copyright © 2017 American Urological Association Education and Research, Inc. Published by Elsevier Inc. All rights reserved.

**URL:** <https://ovidsp.ovid.com/ovidweb.cgi?T=JS&CSC=Y&NEWS=N&PAGE=fulltext&D=med14&DO=10.1016%2fj.juro.2016.10.052> <https://libkey.io/libraries/2592/openurl?genre=article&sid=OVID:medline&id=pmid:27746283&id=doi:10.1016%2Fj.juro.2016.10.052&issn=0022-5347&isbn=&volume=197&issue=4&spage=1084&pages=1084-1089&date=2017&title=Journal+of+Urology&atitle=The+Role+of+the+24-Hour+Urine+Collection+in+the+Prevention+of+Kidney+Stone+Recurrence.&aulast=Hsi&pid=%3Cauthor%3EHsi+RS%3BSanford+T%3BGoldfarb+DS%3BStoller+ML%3C%2Fauthor%3E%3CAN%3E27746283%3C%2FAN%3E%3CDT%3EJournal+Article%3C%2FDT%3E>

85. **Co-Occurrence of Asthma and Nephrolithiasis in Children.**

**Item Type:** Journal Article

**Authors:** Kartha, Ganesh K.;Li, Ina;Comhair, Suzy;Erzurum, Serpil C. and Monga, Manoj

**Publication Date:** 2017

**Journal:** PLoS ONE [Electronic Resource] 12(1), pp. e0168813

**Abstract:** It has been proposed that epithelial dysfunction and inflammation may predispose patients to kidney stone formation. Asthma is another chronic condition related to epithelial dysfunction and inflammation. We hypothesized that pediatric patients with asthma would have an increased prevalence of nephrolithiasis. Furthermore, we investigated if asthma patients with nephrolithiasis have clinical characteristics and urine profiles that point to mechanisms of stone formation. We evaluated 865 pediatric patients who had a diagnosis of nephrolithiasis. Clinical/demographic data and 24 hour urine samples were compared between asthma + stone (n = 142) and stone only patients. Data from asthmatics without stone were also available for evaluation of medication differences among asthma + stone and asthma only patients. The prevalence of nephrolithiasis in the pediatric population at our institution was 0.08% vs. 0.31% in our pediatric asthmatic population. The prevalence of asthma in our pediatric population was 6.8% vs. 26.7% in our pediatric stone patients. Asthma + stone patients were more likely to be on a combination inhaled corticosteroid + long acting beta agonist inhaler as compared to age/gender/BMI matched asthma patients without stone (29.7% vs. 13.7%, p = 0.0012). 259 kidney stone patients had 24 hour urine samples for comparison. There was no difference in 24 hour urine profiles between asthma + stone and stone only patients. Children with asthma have a 4-fold greater prevalence of kidney stones than the general pediatric population. Similarly, children with kidney stones have a 4-fold greater prevalence of asthma. This correlation may suggest a mechanistic link between asthma and nephrolithiasis. Further investigation is needed to elucidate the pathophysiologic origin of this relationship.

**URL:** <https://ovidsp.ovid.com/ovidweb.cgi?T=JS&CSC=Y&NEWS=N&PAGE=fulltext&D=med14&DO=10.1371%2fjournal.pone.0168813> <https://libkey.io/libraries/2592/openurl?genre=article&sid=OVID:medline&id=pmid:28081237&id=doi:10.1371%2Fjournal.pone.0168813&issn=1932-6203&isbn=&volume=12&issue=1&spage=e0168813&pages=e0168813&date=2017&title=PLoS+ONE+%5BElectronic+Resource%5D&atitle=Co-Occurrence+of+Asthma+and+Nephrolithiasis+in+Children.&aulast=Kartha&pid=%3Cauthor%3EKartha+GK%3BLi+I%3BComhair+S%3BErzurum+SC%3BMonga+M%3C%2Fauthor%3E%3CAN%3E28081237%3C%2FAN%3E%3CDT%3EComparative+Study%3C%2FDT%3E>

86. **Medical and dietary interventions for preventing recurrent urinary stones in children**

**Item Type:** Journal Article

**Authors:** Kern, Adam;Grimsby, Gwen;Mayo, Helen and Baker, Linda A.

**Publication Date:** Nov 09 ,2017

**Journal:** Cochrane Database of Systematic Reviews 11, pp. CD011252

**Abstract: BACKGROUND**: Nephrolithiasis, or urinary stone disease, in children causes significant morbidity, and is increasing in prevalence in the North American population. Therefore, medical and dietary interventions (MDI) for recurrent urinary stones in children are poised to gain increasing importance in the clinical armamentarium. **OBJECTIVES**: To assess the effects of medical and dietary interventions (MDI) for the prevention of idiopathic urinary stones in children aged from one to 18 years. **SEARCH METHODS**: We searched multiple databases using search terms relevant to this review, including studies identified from the Cochrane Central Register of Controlled Trials (CENTRAL, 2017, Issue 1), MEDLINE OvidSP (1946 to 14 February 2017), Embase OvidSP (1980 to 14 February 2017), International Clinical Trials Register (ICTRP) Search Portal and ClinicalTrials.gov. Additionally, we handsearched renal-related journals and the proceedings of major renal conferences, and reviewed weekly current awareness alerts for selected renal journals. The date of the last search was 14 February 2017. There were no language restrictions. **SELECTION CRITERIA**: Randomized controlled trials of at least one year of MDI versus control for prevention of recurrent idiopathic (non-syndromic) nephrolithiasis in children. **DATA COLLECTION AND ANALYSIS**: We used standard methodologic procedures expected by Cochrane. Titles and abstracts were identified by search criteria and then screened for relevance, and then data extraction and risk of bias assessment were carried out. We assessed the quality of evidence using GRADE. **MAIN RESULTS**: The search identified one study of 125 children (72 boys and 53 girls) with calcium-containing idiopathic nephrolithiasis and normal renal morphology following initial treatment with shockwave lithotripsy (SWL). Patients were randomized to oral potassium citrate 1 mEq/kg per day for 12 months versus no specific medication or preventive measure with results reported for a total of 96 patients (48 per group). This included children who were stone-free (n = 52) or had residual stone fragments (n = 44) following SWL. Primary outcomes:Medical therapy may lower rates of stone recurrence with a risk ratio (RR) of 0.19 (95% confidence interval (CI) 0.06 to 0.60; low quality evidence). This corresponds to 270 fewer stone recurrences per 1000 (133 fewer to 313 fewer) children. We downgraded the quality of evidence by two levels for very serious study limitations related to unclear allocation concealment (selection bias) and a high risk of performance, detection and attrition bias. While the data for adverse events were incomplete, they reported that six of 48 (12.5%) children receiving potassium citrate left the trial because of adverse effects. This corresponds to a RR of 13.0 (95% CI 0.75 to 224.53; very low quality evidence); an absolute effect size estimate could not be generated. We downgraded the quality of evidence for study limitations and imprecision. We found no information on retreatment rates. **SECONDARY OUTCOMES**: We found no evidence on serum electrolytes, 24-hour urine collection parameters or time to new stone formation. We were unable to perform any preplanned secondary analyses. **AUTHORS' CONCLUSIONS**: Oral potassium citrate supplementation may reduce recurrent calcium urinary stone formation in children following SWL; however, our confidence in this finding is limited. A substantial number of children stopped the medication due to adverse events. There is no trial evidence on retreatment rates. There is a critical need for additional well-designed trials in children with nephrolithiasis.

**URL:** <https://ovidsp.ovid.com/ovidweb.cgi?T=JS&CSC=Y&NEWS=N&PAGE=fulltext&D=med14&DO=10.1002%2f14651858.CD011252.pub2> <https://libkey.io/libraries/2592/openurl?genre=article&sid=OVID:medline&id=pmid:29117629&id=doi:10.1002%2F14651858.CD011252.pub2&issn=1361-6137&isbn=&volume=11&issue=11&spage=CD011252&pages=CD011252&date=2017&title=Cochrane+Database+of+Systematic+Reviews&atitle=Medical+and+dietary+interventions+for+preventing+recurrent+urinary+stones+in+children.&aulast=Kern&pid=%3Cauthor%3EKern+A%3BGrimsby+G%3BMayo+H%3BBaker+LA%3C%2Fauthor%3E%3CAN%3E29117629%3C%2FAN%3E%3CDT%3EJournal+Article%3C%2FDT%3E>

87. **Factors Associated with Compliance to Increased Fluid Intake and Urine Volume Following Dietary Counseling in First-Time Kidney Stone Patients.**

**Item Type:** Journal Article

**Authors:** Khambati, Aziz;Matulewicz, Richard S.;Perry, Kent T. and Nadler, Robert B.

**Publication Date:** 2017

**Journal:** Journal of Endourology 31(6), pp. 605–610

**Abstract: OBJECTIVE**: Increasing fluid intake to achieve a urine volume (UV) of >2.5 L/day decreases stone events. We assessed compliance rates and demographic and clinical variables associated with increased fluid intake and UV in patients with urolithiasis following dietary counseling. **MATERIALS AND METHODS**: In a retrospective study, patients with a low baseline UV (3 L/day and titrating it to achieve UV >2.5 L/day. A follow-up 24-hour urine collection was performed at 6 months (FU1) and 18 months (FU2) to assess compliance (UV >2.5 L/day) and predictors associated with it. **RESULTS**: Our cohort contained 363 patients, 53.4% men and 46.6% women. The mean UV of baseline, FU1, and FU2 urine collections was 1.63, 2.52, and 2.48 L, respectively. The compliance rate of obtaining UV of >2.5 L was 50.1% at 6 months. On logistic regression, male sex (odds ratio [OR] 3.27, 95% confidence interval [CI] 1.94, 5.52, p 1 L (OR 3.00, 95% CI 1.02, 8.76, p = 0.04) were associated with greater odds of compliance. Age >58 years (OR 0.39, 95% CI 0.20, 0.75, p = 0.005) and presence of lower urinary tract symptoms (OR 0.50, 95% CI 0.26, 0.95, p = 0.03) were associated with lower odds of compliance. Ethnicity, body mass index, renal function, marital status, renal stone burden, stone type, and household income were not associated with compliance. **CONCLUSIONS**: In stone formers, compliance to fluid intake recommendations as measured by 24-hour UV is roughly 50%. Understanding risk factors for noncompliance, especially in at risk patients, can be used for quality improvement initiatives and reducing stone events.

**URL:** <https://ovidsp.ovid.com/ovidweb.cgi?T=JS&CSC=Y&NEWS=N&PAGE=fulltext&D=med14&DO=10.1089%2fend.2016.0836> <https://libkey.io/libraries/2592/openurl?genre=article&sid=OVID:medline&id=pmid:28318298&id=doi:10.1089%2Fend.2016.0836&issn=0892-7790&isbn=&volume=31&issue=6&spage=605&pages=605-610&date=2017&title=Journal+of+Endourology&atitle=Factors+Associated+with+Compliance+to+Increased+Fluid+Intake+and+Urine+Volume+Following+Dietary+Counseling+in+First-Time+Kidney+Stone+Patients.&aulast=Khambati&pid=%3Cauthor%3EKhambati+A%3BMatulewicz+RS%3BPerry+KT%3BNadler+RB%3C%2Fauthor%3E%3CAN%3E28318298%3C%2FAN%3E%3CDT%3EJournal+Article%3C%2FDT%3E>

88. **Marked increase in urinary excretion of apolipoproteins in children with nephrolithiasis associated with hypercalciuria.**

**Item Type:** Journal Article

**Authors:** Kovacevic, Larisa;Lu, Hong;Caruso, Joseph A.;Govil-Dalela, Tuhina;Thomas, Ronald and Lakshmanan, Yegappan

**Publication Date:** 2017

**Journal:** Pediatric Nephrology 32(6), pp. 1029–1033

**Abstract: BACKGROUND**: Using a proteomic approach, we aimed to identify and compare the urinary excretion of proteins involved in lipid transport and metabolism in children with kidney stones and hypercalciuria (CAL), hypocitraturia (CIT), and normal metabolic work-up (NM), and in healthy controls (HCs). Additionally, we aimed to confirm these results using ELISA, and to examine the relationship between the urinary excretion of selected proteins with demographic, dietary, blood, and urinary parameters. **METHODS**: Prospective, controlled, pilot study of pooled urine from CAL, CIT, and NM versus age- and gender-matched HCs, using liquid chromatography-mass spectrometry. Relative protein abundance was estimated using spectral counting. Results were confirmed by ELISA performed on individual samples. **RESULTS**: Of the 1,813 proteins identified, 230 met the above criteria. Of those, 5 proteins (apolipoprotein A-II [APOA2]; apolipoprotein A-IV [APOA4]; apolipoprotein C-III [APOA3]; fatty acid-binding protein, liver [FABPL]; fatty acid-binding protein, adipocyte [FABP4]) involved in lipid metabolism and transport were found in the CAL group, with significant differences compared with HCs. ELISA analysis indicated statistically significant differences in the urinary excretion of APOC3, APOA4, and FABPL in the CAL group compared with HCs. Twenty-four-hour urinary calcium excretion correlated significantly with concentrations of ApoC3 (r = 0.77, p : Of the 1,813 proteins identified, 230 met the above criteria. Of those, 5 proteins (apolipoprotein A-II [APOA2]; apolipoprotein A-IV [APOA4]; apolipoprotein C-III [APOA3]; fatty acid-binding protein, liver [FABPL]; fatty acid-binding protein, adipocyte [FABP4]) involved in lipid metabolism and transport were found in the CAL group, with significant differences compared with HCs. ELISA analysis indicated statistically significant differences in the urinary excretion of APOC3, APOA4, and FABPL in the CAL group compared with HCs. Twenty-four-hour urinary calcium excretion correlated significantly with concentrations of ApoC3 (r = 0.77, p **CONCLUSIONS**: We provide proteomic data showing increased urinary excretion of lipid metabolism/transport-related proteins in children with kidney stones and hypercalciuria. These findings suggest that abnormalities in lipid metabolism might play a role in kidney stone formation.

**URL:** <https://ovidsp.ovid.com/ovidweb.cgi?T=JS&CSC=Y&NEWS=N&PAGE=fulltext&D=med14&DO=10.1007%2fs00467-016-3576-1> <https://libkey.io/libraries/2592/openurl?genre=article&sid=OVID:medline&id=pmid:28188435&id=doi:10.1007%2Fs00467-016-3576-1&issn=0931-041X&isbn=&volume=32&issue=6&spage=1029&pages=1029-1033&date=2017&title=Pediatric+Nephrology&atitle=Marked+increase+in+urinary+excretion+of+apolipoproteins+in+children+with+nephrolithiasis+associated+with+hypercalciuria.&aulast=Kovacevic&pid=%3Cauthor%3EKovacevic+L%3BLu+H%3BCaruso+JA%3BGovil-Dalela+T%3BThomas+R%3BLakshmanan+Y%3C%2Fauthor%3E%3CAN%3E28188435%3C%2FAN%3E%3CDT%3EClinical+Trial%3C%2FDT%3E>

89. **Age, Body Mass Index, and Gender Predict 24-Hour Urine Parameters in Recurrent Idiopathic Calcium Oxalate Stone Formers.**

**Item Type:** Journal Article

**Authors:** Otto, Brandon J.;Bozorgmehri, Shahab;Kuo, Jennifer;Canales, Muna;Bird, Vincent G. and Canales, Benjamin

**Publication Date:** De ,2017

**Journal:** Journal of Endourology 31(12), pp. 1335–1341

**Abstract: INTRODUCTION AND OBJECTIVE**: Despite guidelines, routine 24-hour urine testing is completed in : Despite guidelines, routine 24-hour urine testing is completed in **METHODS**: We performed a retrospective study of 392 consecutive patients from 2007 to 2014 with >=2 lifetime stone episodes, >70% calcium oxalate by mineral analysis, and >=1 24-hour urine collection. We compared mean 24-hour urine values by age in decades. We used logistic regression and receiver operating characteristic (ROC) curve analysis to assess the predictive ability of age, gender, body mass index (BMI), and comorbidities to detect abnormal 24-hour urine parameters. **RESULTS**: The mean age of the cohort was 51 +/- 16 years. Older age was associated with greater urinary oxalate (p-trend : The mean age of the cohort was 51 +/- 16 years. Older age was associated with greater urinary oxalate (p-trend : The mean age of the cohort was 51 +/- 16 years. Older age was associated with greater urinary oxalate (p-trend **CONCLUSIONS**: In our recurrent calcium oxalate cohort, age significantly impacted urinary calcium, oxalate, citrate, and pH. Along with gender and BMI, age can be used to predict key 24-hour urine stone risk results. These data lay the foundation for a risk prediction tool, which could be a surrogate for 24-hour urine results in recurrent stone formers, who are unwilling or unable to complete metabolic testing. Further validation of these findings is needed in other stone populations.

**URL:** <https://ovidsp.ovid.com/ovidweb.cgi?T=JS&CSC=Y&NEWS=N&PAGE=fulltext&D=med14&DO=10.1089%2fend.2017.0352> <https://libkey.io/libraries/2592/openurl?genre=article&sid=OVID:medline&id=pmid:29084490&id=doi:10.1089%2Fend.2017.0352&issn=0892-7790&isbn=&volume=31&issue=12&spage=1335&pages=1335-1341&date=2017&title=Journal+of+Endourology&atitle=Age%2C+Body+Mass+Index%2C+and+Gender+Predict+24-Hour+Urine+Parameters+in+Recurrent+Idiopathic+Calcium+Oxalate+Stone+Formers.&aulast=Otto&pid=%3Cauthor%3EOtto+BJ%3BBozorgmehri+S%3BKuo+J%3BCanales+M%3BBird+VG%3BCanales+B%3C%2Fauthor%3E%3CAN%3E29084490%3C%2FAN%3E%3CDT%3EJournal+Article%3C%2FDT%3E>

90. **Vascular Disease and Kidney Stones: Abdominal Aortic Calcifications Are Associated with Low Urine pH and Hypocitraturia.**

**Item Type:** Journal Article

**Authors:** Patel, Nishant D.;Ward, Ryan D.;Calle, Juan;Remer, Erick M. and Monga, Manoj

**Publication Date:** 2017

**Journal:** Journal of Endourology 31(9), pp. 956–961

**Abstract: OBJECTIVE**: Vascular calcifications are associated with nephrolithiasis. Although studies have demonstrated correlations with vascular disease and calcium stones in kidney stone formers (KSF), an etiologic link has remained elusive. As a noncontrast CT (NCCT) scan is typically part of a stone evaluation, our objective was to evaluate the association of NCCT-based assessment of abdominal aortic calcifications (AACs) with 24-hour urine parameters and stone composition. **MATERIALS AND METHODS**: Ninety-seven KSF were included with CT imaging and 24-hour urine studies. For each patient, semi-automated CT software was utilized to provide an AAC Agatston score from the celiac axis to the aortic bifurcation. Univariate analysis was performed to compare patients with or without AAC. Multivariate logistic regression was performed to assess for variables associated with 24-hour urine parameters and stone composition. **RESULTS**: The presence of AAC was associated with hypertension, diabetes, peripheral vascular disease, and coronary artery disease. Patients with any AAC showed lower 24-hour urine citrate (399 vs 593 mg/day, p : The presence of AAC was associated with hypertension, diabetes, peripheral vascular disease, and coronary artery disease. Patients with any AAC showed lower 24-hour urine citrate (399 vs 593 mg/day, p : The presence of AAC was associated with hypertension, diabetes, peripheral vascular disease, and coronary artery disease. Patients with any AAC showed lower 24-hour urine citrate (399 vs 593 mg/day, p : The presence of AAC was associated with hypertension, diabetes, peripheral vascular disease, and coronary artery disease. Patients with any AAC showed lower 24-hour urine citrate (399 vs 593 mg/day, p **CONCLUSIONS**: NCCT-based diagnosis of AAC is associated with low urine pH, hypocitraturia, and uric acid stone formation. The presence of AAC could be considered an additional prognosticator for the utility of alkalinization therapy.

**URL:** <https://ovidsp.ovid.com/ovidweb.cgi?T=JS&CSC=Y&NEWS=N&PAGE=fulltext&D=med14&DO=10.1089%2fend.2017.0350> <https://libkey.io/libraries/2592/openurl?genre=article&sid=OVID:medline&id=pmid:28605936&id=doi:10.1089%2Fend.2017.0350&issn=0892-7790&isbn=&volume=31&issue=9&spage=956&pages=956-961&date=2017&title=Journal+of+Endourology&atitle=Vascular+Disease+and+Kidney+Stones%3A+Abdominal+Aortic+Calcifications+Are+Associated+with+Low+Urine+pH+and+Hypocitraturia.&aulast=Patel&pid=%3Cauthor%3EPatel+ND%3BWard+RD%3BCalle+J%3BRemer+EM%3BMonga+M%3C%2Fauthor%3E%3CAN%3E28605936%3C%2FAN%3E%3CDT%3EJournal+Article%3C%2FDT%3E>

91. **A comparison of micro-PERC and retrograde intrarenal surgery results in pediatric patients with renal stones.**

**Item Type:** Journal Article

**Authors:** Sen, Haluk;Seckiner, Ilker;Bayrak, Omer;Dogan, Kazim and Erturhan, Sakip

**Publication Date:** De ,2017

**Journal:** Journal of Pediatric Urology 13(6), pp. 619.e1–619.e5

**Abstract: INTRODUCTION**: With advancements in endoscopic surgery, open surgical techniques for urinary system stones have paved the way for the application of less invasive treatment modalities in patients with pediatric kidney stone disease. These treatment options include extracorporeal shock wave lithotripsy (ESWL), percutaneous nephrolithotomy (PCNL), and retrograde intrarenal surgery (RIRS). **OBJECTIVE**: We aimed to compare the efficacy and safety of RIRS and micro-PCNL techniques in the surgical treatment of kidney stones in pediatric patients. **STUDY DESIGN**: A total of 48 pediatric patients, who underwent RIRS or micro-PERC for pediatric kidney stone disease, were retrospectively analyzed. Urinalysis, urine culture, serum creatinine (Cr), blood urea nitrogen (BUN), calcium, phosphorus, parathyroid hormone, 24-hour urine sample, complete blood count (CBC), urinary system X-ray, kidney ureter bladder (KUB), and urinary system ultrasonography (USG) test results were evaluated prior to the procedure. Intravenous pyelography (IVP), non-contrast computed tomography (CT), and renal scintigraphy evaluations were also performed, if necessary. The patients were divided into two groups: micro-PERC group (n = 25) and RIRS group (n = 23). Data relating to the duration of the operation, duration of fluoroscopy, length of hospitalization, complication rates, and stone-free rates were recorded. **RESULTS**: The mean ages of the micro-PERC and RIRS groups were 4 +/- 2.3 and 10.9 +/- 3 years, respectively (p = 0.001). However, the mean stone sizes were 12.2 +/- 2.8 and 13.7 +/- 3.5 mm, respectively (p > 0.05). The mean duration of operation was 75.1 +/- 18.9 min in the micro-PERC group and 62.3 +/- 15.3 min in the RIRS group (p > 0.05). In addition, the mean duration of fluoroscopy was 115 +/- 35.4 s in the micro-PERC group and 39.9 +/- 15.3 s in the RIRS group. The stone-free rates in the micro-PERC and RIRS groups following the procedure were reported to be 84% (21/25) and 82.6% (19/23), respectively (p > 0.05). In terms of the degree of preoperative hydronephrosis between the groups, the rates of mild and moderate-severe disease were 76% (19 out of 25) and 24% (6 out of 25), and 69.5% (16 out of 23) and 30.4% (7 out of 23) in the micro-PERC and RIRS groups, respectively. **CONCLUSION**: In recent years, technological developments in minimally invasive procedures, such as micro-PERC and RIRS, have facilitated choices made by urologists in the effective and safe first-line treatment in pediatric patients. Copyright © 2017 Journal of Pediatric Urology Company. Published by Elsevier Ltd. All rights reserved.

**URL:** <https://ovidsp.ovid.com/ovidweb.cgi?T=JS&CSC=Y&NEWS=N&PAGE=fulltext&D=med14&DO=10.1016%2fj.jpurol.2017.04.022> <https://libkey.io/libraries/2592/openurl?genre=article&sid=OVID:medline&id=pmid:28679474&id=doi:10.1016%2Fj.jpurol.2017.04.022&issn=1477-5131&isbn=&volume=13&issue=6&spage=619.e1&pages=619.e1-619.e5&date=2017&title=Journal+of+pediatric+urology&atitle=A+comparison+of+micro-PERC+and+retrograde+intrarenal+surgery+results+in+pediatric+patients+with+renal+stones.&aulast=Sen&pid=%3Cauthor%3ESen+H%3BSeckiner+I%3BBayrak+O%3BDogan+K%3BErturhan+S%3C%2Fauthor%3E%3CAN%3E28679474%3C%2FAN%3E%3CDT%3EComparative+Study%3C%2FDT%3E>

92. **Kidney Stone Recurrence among Children and Adolescents.**

**Item Type:** Journal Article

**Authors:** Tasian, Gregory E.;Kabarriti, Abdo E.;Kalmus, Angela and Furth, Susan L.

**Publication Date:** 2017

**Journal:** Journal of Urology 197(1), pp. 246–252

**Abstract: PURPOSE**: Kidney stone disease has become increasingly common during childhood and adolescence. However, the rate of symptomatic kidney stone recurrence for pediatric patients is uncertain. We sought to determine the recurrence rate of symptomatic kidney stones in a cohort of children with incident symptomatic nephrolithiasis. **MATERIALS AND METHODS**: We performed a retrospective cohort study of patients 3 to 18 years old without anatomical abnormalities or genetic causes of nephrolithiasis who presented with a first symptomatic kidney stone between 2008 and 2014. We determined recurrence rates of symptomatic nephrolithiasis, defined as a new kidney stone on ultrasound and/or computerized tomogram associated with pain and/or vomiting. We also estimated associations between completing 24-hour urinalysis and symptomatic kidney stone recurrence using Kaplan-Meier curves and multivariable Cox regression models. **RESULTS**: A total of 285 children with a median age of 14.8 years (IQR 11.3-16.6) at nephrolithiasis diagnosis were followed for 492 person-years. A total of 86 symptomatic recurrent stones developed in 68 patients (24%) during the followup period. The probability of symptomatic stone recurrence was 50% at 3 years after the index kidney stone. Median time to stone recurrence was 3 years at the first recurrence and 5 years at the second. Adjusting for confounders including adherence to followup, completing a 24-hour urinalysis after a kidney stone episode was associated with a 60% decreased risk of recurrence (hazard ratio 0.40, 95% CI 0.18-0.91). **CONCLUSIONS**: The risk of kidney stone recurrence is high during childhood, with approximately 50% of children presenting with symptomatic recurrence within 3 years of the first stone. The role and usefulness of analyzing 24-hour urine chemistries in decreasing kidney stone recurrence should be explored in future prospective studies. Copyright A© 2017 American Urological Association Education and Research, Inc. Published by Elsevier Inc. All rights reserved.

**URL:** <https://ovidsp.ovid.com/ovidweb.cgi?T=JS&CSC=Y&NEWS=N&PAGE=fulltext&D=med14&DO=10.1016%2fj.juro.2016.07.090> <https://libkey.io/libraries/2592/openurl?genre=article&sid=OVID:medline&id=pmid:27521691&id=doi:10.1016%2Fj.juro.2016.07.090&issn=0022-5347&isbn=&volume=197&issue=1&spage=246&pages=246-252&date=2017&title=Journal+of+Urology&atitle=Kidney+Stone+Recurrence+among+Children+and+Adolescents.&aulast=Tasian&pid=%3Cauthor%3ETasian+GE%3BKabarriti+AE%3BKalmus+A%3BFurth+SL%3C%2Fauthor%3E%3CAN%3E27521691%3C%2FAN%3E%3CDT%3EJournal+Article%3C%2FDT%3E>

93. **Physicochemical factors of the urine of children with hypercalciuria**

**Item Type:** Journal Article

**Authors:** Ubetagoyena-Arrieta, Mercedes;Martinez Saenz de Jubera, Jorge;Corera-Casty, Maria Nieves;Gonzalez Perez-Yarza, Eduardo and Garcia-Albizua, Irati

**Publication Date:** Ap ,2017

**Journal:** Archivos Espanoles De Urologia 70(3), pp. 342–348

**Abstract: OBJECTIVE**: Hypercalciuria is a common lithogenic risk factor. The aim of this study was, first, to study the characteristics of urine biochemical factors of children with hypercalciuria (HC) and compare them with those of children without hypercalciuria. Second, to analyze the differences between children with HC and lithiasis (HCL) and children with HC and no lithiasis (HCNL). **METHODS**: The sample was composed by 111 cases with HC, divided into 2 groups: HCNL group, consisting of 93 cases with no personal history of kidney stones, and HCL group, 18 cases with personal history of kidney stones. As a cohort control group, 113 healthy children were used. Creatinine, urea, sodium, potassium, chlorine, uric acid, calcium, phosphorus, magnesium and osmolality: blood and 24-hour urine following parameters were determined. Oxalate and citrate were determined in urine. **RESULTS**: The mean values of natriuresis, uricosuria, phosphaturia, magnesuria, citraturia, calcium oxalate and calcium phosphate saturation were higher in HCNL than in control group. The HCL group had phosphaturia, calcium oxalate and calcium phosphate saturations more elevated compared with the control group. There were no significant differences in urinary excretion of various parameters when the groups HCL and HCNL were compared. **CONCLUSIONS**: Our results show that children with hypercalciuria without lithiasis associated show an increase in natriuresis, phosphaturia, uricosuria, magnesuria and citraturia. We found no differences between these urinary abnormalities when compared hypercalciuric children without lithiasis with those with hypercalciuria and urolithiasis.

**URL:** <https://ovidsp.ovid.com/ovidweb.cgi?T=JS&CSC=Y&NEWS=N&PAGE=fulltext&D=med14&AN=28422036> <https://libkey.io/libraries/2592/openurl?genre=article&sid=OVID:medline&id=pmid:28422036&id=doi:&issn=0004-0614&isbn=&volume=70&issue=3&spage=342&pages=342-348&date=2017&title=Archivos+Espanoles+de+Urologia&atitle=Caracteristicas+de+los+factores+fisicoquimicos+que+componen+la+orina+de+los+ninos+con+hipercalciuria.&aulast=Ubetagoyena-Arrieta&pid=%3Cauthor%3EUbetagoyena-Arrieta+M%3BMartinez+Saenz+de+Jubera+J%3BCorera-Casty+MN%3BGonzalez+Perez-Yarza+E%3BGarcia-Albizua+I%3C%2Fauthor%3E%3CAN%3E28422036%3C%2FAN%3E%3CDT%3EJournal+Article%3C%2FDT%3E>

94. **Metabolic disturbances in Chinese children with urolithiasis: a single center report.**

**Item Type:** Journal Article

**Authors:** Yang, Dong;Tiselius, Hans-Goran;Lan, Chuangxin;Chen, Dong;Chen, Kang;Ou, Lili;Liu, Yang;Xu, Shaohong;Zeng, Guohua;Lei, Ming and Wu, Wenqi

**Publication Date:** Ju ,2017

**Journal:** Urolithiasis 45(3), pp. 285–290

**Abstract:** Urinary stones and urine composition are the first steps in the process of recurrence prevention, but data concerning the association between the two compositions are scarce in Chinese children with urolithiasis. We retrospectively analyzed the records of children (age range 0-18 years) with urolithiasis in our center between March 2004 and December 2013. Stone analysis was carried out in 382 children and 24-hour urine analysis in 80 children. Analysis of both stone and 24-hour urine composition was completed in 56 children. Stone samples were analyzed by Fourier transform-infrared spectrometry. The major stone constituents were calcium oxalate (78.8 %). Of 80 children with 24 h urine analysis, only 2.5 % were without urinary metabolic abnormalities. Hypocitraturia was recorded in 97.5 %, high sodium excretion in 50.0 %, cystinuria in 48.7 %, hypercalciuria in 18.8 %, small urine volumes in 12.5 %, hyperoxaluria in 5.0 % and hyperuricosuria in 1.3 %. Interestingly, higher urine volumes were recorded in girls than in boys (73.2 +/- 58.5 vs 51.3 +/- 45.3 mL/kg, p = 0.036). Urine sodium (p = 0.002) and oxalate (p = 0.004) were significantly higher in children >9 year old. Moreover, compared with calcium oxalate stone formers, the urine volume (p = 0.040), citrate (p = 0.007) and cystine (p = 0.004) were higher in patients with cystine stones. Hypocitraturia was the common abnormality among Chinese children with urolithiasis. The surprisingly high incidence of cystinuria is of note.

**URL:** <https://ovidsp.ovid.com/ovidweb.cgi?T=JS&CSC=Y&NEWS=N&PAGE=fulltext&D=med14&DO=10.1007%2fs00240-016-0910-9> <https://libkey.io/libraries/2592/openurl?genre=article&sid=OVID:medline&id=pmid:27510800&id=doi:10.1007%2Fs00240-016-0910-9&issn=2194-7228&isbn=&volume=45&issue=3&spage=285&pages=285-290&date=2017&title=Urolithiasis&atitle=Metabolic+disturbances+in+Chinese+children+with+urolithiasis%3A+a+single+center+report.&aulast=Yang&pid=%3Cauthor%3EYang+D%3BTiselius+HG%3BLan+C%3BChen+D%3BChen+K%3BOu+L%3BLiu+Y%3BXu+S%3BZeng+G%3BLei+M%3BWu+W%3C%2Fauthor%3E%3CAN%3E27510800%3C%2FAN%3E%3CDT%3EJournal+Article%3C%2FDT%3E>

95. **How Much Information is Lost When You Only Collect One 24-Hour Urine Sample during the Initial Metabolic Evaluation?.**

**Item Type:** Journal Article

**Authors:** Alruwaily, Abdulrahman F.;Dauw, Casey A.;Bierlein, Maggie J.;Yan, Phyllis;Asplin, John R.;Ghani, Khurshid R.;Wolf, J. Stuart Jr and Hollingsworth, John M.

**Publication Date:** Oc ,2016

**Journal:** Journal of Urology 196(4), pp. 1143–1148

**Abstract: PURPOSE**: During the initial metabolic evaluation the need for 1 vs 2, 24-hour urine collections is debated. While data suggest that mean urine chemistry measures are similar on consecutive samples, it remains unclear how much, if any, information is lost when only 1 sample is collected. **MATERIALS AND METHODS**: Using analytical files from Litholink Corporation R (1995 to 2013) we identified adults with kidney stones who underwent initial metabolic testing. Next we determined the subset of patients who collected 2, 24-hour urine samples with urine creatinine varying by 10% or less during a 7-day time window. We then examined the degree of variability in urine chemistry profiles. Specifically we calculated the mean absolute value of the difference between samples as well as the percent difference for individual urine parameters. **RESULTS**: We identified 70,192 patients meeting our eligibility criteria. While the overall means for individual urine parameters did not vary between samples, the percent difference between the samples varied widely. For example, nearly 1 in 3 patients had a 30% or greater difference in urine calcium and volume between 2 consecutive samples. We noted that inconsistencies between samples often involved multiple parameters. For instance, 29% and 25% of patients had a 20% difference in 2 and 3 or more parameters, respectively. **CONCLUSIONS**: We observed substantial differences between consecutive 24-hour urine samples that could affect clinical decision making. In light of these findings clinicians must weigh the information lost from only 1 collection vs the burden to the patient of collecting 2. Copyright © 2016 American Urological Association Education and Research, Inc. Published by Elsevier Inc. All rights reserved.

**URL:** <https://ovidsp.ovid.com/ovidweb.cgi?T=JS&CSC=Y&NEWS=N&PAGE=fulltext&D=med13&DO=10.1016%2fj.juro.2016.04.074> <https://libkey.io/libraries/2592/openurl?genre=article&sid=OVID:medline&id=pmid:27140069&id=doi:10.1016%2Fj.juro.2016.04.074&issn=0022-5347&isbn=&volume=196&issue=4&spage=1143&pages=1143-8&date=2016&title=Journal+of+Urology&atitle=How+Much+Information+is+Lost+When+You+Only+Collect+One+24-Hour+Urine+Sample+during+the+Initial+Metabolic+Evaluation%3F.&aulast=Alruwaily&pid=%3Cauthor%3EAlruwaily+AF%3BDauw+CA%3BBierlein+MJ%3BYan+P%3BAsplin+JR%3BGhani+KR%3BWolf+JS+Jr%3BHollingsworth+JM%3C%2Fauthor%3E%3CAN%3E27140069%3C%2FAN%3E%3CDT%3EJournal+Article%3C%2FDT%3E>

96. **Short-Term Tolvaptan Increases Water Intake and Effectively Decreases Urinary Calcium Oxalate, Calcium Phosphate and Uric Acid Supersaturations.**

**Item Type:** Journal Article

**Authors:** Cheungpasitporn, Wisit;Erickson, Stephen B.;Rule, Andrew D.;Enders, Felicity and Lieske, John C.

**Publication Date:** Ma ,2016

**Journal:** Journal of Urology 195(5), pp. 1476–1481

**Abstract: PURPOSE**: Some patients cannot effectively increase water intake and urine volume to prevent urinary stones. Tolvaptan, a V2 receptor antagonist, blocks water reabsorption in the collecting duct and should decrease urinary supersaturation of stone forming solutes, although this action has never been proved. **MATERIALS AND METHODS**: We conducted a double-blind, randomized, placebo controlled, crossover study of 21 calcium urinary stone formers stratified into majority calcium oxalate (10 patients) and calcium phosphate (11) groups. Patients received 45 mg tolvaptan per day or placebo for 1 week, followed by a washout week and crossover to tolvaptan or placebo for week 3. A 24-hour urine sample was collected at the end of weeks 1 and 3. **RESULTS**: Tolvaptan vs placebo decreased urinary osmolality (mean +/- SD 204 +/- 96 vs 529 +/- 213 mOsm/kg, p : Tolvaptan vs placebo decreased urinary osmolality (mean +/- SD 204 +/- 96 vs 529 +/- 213 mOsm/kg, p : Tolvaptan vs placebo decreased urinary osmolality (mean +/- SD 204 +/- 96 vs 529 +/- 213 mOsm/kg, p : Tolvaptan vs placebo decreased urinary osmolality (mean +/- SD 204 +/- 96 vs 529 +/- 213 mOsm/kg, p 0.05 for all interactions). **CONCLUSIONS**: Tolvaptan increases urine volume and decreases urinary supersaturation in calcium stone formers. Further study is needed to determine if long-term use of V2 receptor antagonists results in fewer stone events. Copyright © 2016 American Urological Association Education and Research, Inc. Published by Elsevier Inc. All rights reserved.

**URL:** <https://ovidsp.ovid.com/ovidweb.cgi?T=JS&CSC=Y&NEWS=N&PAGE=fulltext&D=med13&DO=10.1016%2fj.juro.2015.11.027> <https://libkey.io/libraries/2592/openurl?genre=article&sid=OVID:medline&id=pmid:26598423&id=doi:10.1016%2Fj.juro.2015.11.027&issn=0022-5347&isbn=&volume=195&issue=5&spage=1476&pages=1476-1481&date=2016&title=Journal+of+Urology&atitle=Short-Term+Tolvaptan+Increases+Water+Intake+and+Effectively+Decreases+Urinary+Calcium+Oxalate%2C+Calcium+Phosphate+and+Uric+Acid+Supersaturations.&aulast=Cheungpasitporn&pid=%3Cauthor%3ECheungpasitporn+W%3BErickson+SB%3BRule+AD%3BEnders+F%3BLieske+JC%3C%2Fauthor%3E%3CAN%3E26598423%3C%2FAN%3E%3CDT%3EJournal+Article%3C%2FDT%3E>

97. **Prospective evaluation of urinary metabolic indices in severely obese adolescents after weight loss surgery.**

**Item Type:** Journal Article

**Authors:** DeFoor, W. Robert;Asplin, John R.;Kollar, Linda;Jackson, Elizabeth;Jenkins, Todd;Schulte, Marion and Inge, Thomas

**Publication Date:** Fe ,2016

**Journal:** Surgery for Obesity & Related Diseases 12(2), pp. 363–367

**Abstract: BACKGROUND**: Observational studies in obese adults have found abnormal urinary metabolic indices that predispose to nephrolithiasis. Few studies have been performed in severely obese adolescents. **OBJECTIVES**: To assess urinary stone risk factors in severely obese adolescents and in those undergoing 2 types of weight loss surgery. **SETTING**: Children's hospital, United States. **METHODS**: A prospective cross-sectional study was performed to assess urinary metabolic profiles in severely obese adolescents who either have not undergone any gastrointestinal surgery or who have undergone Roux-en-Y gastric bypass (RYGB) or vertical sleeve gastrectomy (SG). Twenty-four-hour urine collections were performed at home and evaluated at a central laboratory. Established normal reference ranges for adults were used in the analysis. A linear regression analysis was performed assessing the relationship of the study group with each of the outcomes. **RESULTS**: A total of 55 samples were analyzed from 14 severely obese adolescents and from 17 severely obese adolescents after bariatric surgery (RYGB, 10; SG, 7). Median body mass index was similar between the RYGB and SG groups. The median 24-hour excretion of oxalate was significantly elevated in the RYGB group. Calcium and uric acid excretion and the median supersaturation of calcium oxalate, calcium phosphate, and uric acid were similar among all groups. **CONCLUSIONS**: Elevated excretion of oxalate in the urine of severely obese adolescents and in those who have undergone RYGB may portend increased risk for kidney stone formation. Larger longitudinal studies are needed to verify these findings and to determine the clinical risk of developing stone disease in these patient populations. Copyright © 2016 American Society for Metabolic and Bariatric Surgery. Published by Elsevier Inc. All rights reserved.

**URL:** <https://ovidsp.ovid.com/ovidweb.cgi?T=JS&CSC=Y&NEWS=N&PAGE=fulltext&D=med13&DO=10.1016%2fj.soard.2015.03.013> <https://libkey.io/libraries/2592/openurl?genre=article&sid=OVID:medline&id=pmid:26077697&id=doi:10.1016%2Fj.soard.2015.03.013&issn=1550-7289&isbn=&volume=12&issue=2&spage=363&pages=363-7&date=2016&title=Surgery+for+Obesity+%26+Related+Diseases&atitle=Prospective+evaluation+of+urinary+metabolic+indices+in+severely+obese+adolescents+after+weight+loss+surgery.&aulast=DeFoor&pid=%3Cauthor%3EDeFoor+WR%3BAsplin+JR%3BKollar+L%3BJackson+E%3BJenkins+T%3BSchulte+M%3BInge+T%3C%2Fauthor%3E%3CAN%3E26077697%3C%2FAN%3E%3CDT%3EJournal+Article%3C%2FDT%3E>

98. **Assessment of Urinary Inhibitor or Promoter Activity in Uric Acid Nephrolithiasis.**

**Item Type:** Journal Article

**Authors:** Doizi, Steeve;Rodgers, Kathy;Poindexter, John;Sakhaee, Khashayar and Maalouf, Naim M.

**Publication Date:** 2016

**Journal:** Journal of Urology 195(6), pp. 1799–1804

**Abstract: PURPOSE**: We assessed decreased inhibitor activity or increased promoter activity in the urine of idiopathic uric acid stone formers compared to nonstone formers independent of urinary pH. **MATERIALS AND METHODS**: A total of 30 idiopathic uric acid stone formers, and 9 obese and 12 lean nonstone formers collected 24-hour urine while on a metabolic diet. Three urine aliquots per subject were used to assess spontaneous nucleation (de novo crystal formation), crystal growth using a 0.1 mg/ml anhydrous uric acid seed and steady-state uric acid solubility (the maximum amount of uric acid dissolvable in urine) using a 5 mg/ml uric acid seed. All experiments were performed for 6 hours at a constant pH of 5.0. Uric acid concentration was measured in filtered aliquots at 0, 3 and 6 hours. **RESULTS**: At baseline 24-hour urinary pH was significantly lower and uric acid saturation was significantly higher in idiopathic uric acid stone formers. No significant spontaneous nucleation developed and similar uric acid steady-state solubility was reached in the 3 groups. Idiopathic uric acid stone formers and lean nonstone formers showed a similar decrease in uric acid concentration during crystal growth. Obese nonstone formers started with a higher uric acid concentration and consequently demonstrated a greater decrease in the uric acid concentration for crystal growth. **CONCLUSIONS**: This study suggests that there is no significant difference between idiopathic uric acid stone formers and nonstone formers in promoter or inhibitor activity in whole urine against uric acid stone formation when urine pH is maintained constant. The findings suggest that uric acid stone formation is dictated by high urinary saturation with respect to uric acid, which is driven primarily by low urine pH. Copyright © 2016 American Urological Association Education and Research, Inc. Published by Elsevier Inc. All rights reserved.

**URL:** <https://ovidsp.ovid.com/ovidweb.cgi?T=JS&CSC=Y&NEWS=N&PAGE=fulltext&D=med13&DO=10.1016%2fj.juro.2015.12.077> <https://libkey.io/libraries/2592/openurl?genre=article&sid=OVID:medline&id=pmid:26723865&id=doi:10.1016%2Fj.juro.2015.12.077&issn=0022-5347&isbn=&volume=195&issue=6&spage=1799&pages=1799-804&date=2016&title=Journal+of+Urology&atitle=Assessment+of+Urinary+Inhibitor+or+Promoter+Activity+in+Uric+Acid+Nephrolithiasis.&aulast=Doizi&pid=%3Cauthor%3EDoizi+S%3BRodgers+K%3BPoindexter+J%3BSakhaee+K%3BMaalouf+NM%3C%2Fauthor%3E%3CAN%3E26723865%3C%2FAN%3E%3CDT%3EJournal+Article%3C%2FDT%3E>

99. **The Effect of Disease Severity on 24-Hour Urine Parameters in Kidney Stone Patients With Type 2 Diabetes.**

**Item Type:** Journal Article

**Authors:** Fram, Ethan B.;Moazami, Saman and Stern, Joshua M.

**Publication Date:** Ja ,2016

**Journal:** Urology 87, pp. 52–59

**Abstract: OBJECTIVE**: To characterize the changes in urine composition associated with increasing severity of diabetes, we analyzed urine composition relative to glycated hemoglobin (HbA1c) and treatment strategy in a largely minority population. **METHODS**: Patients treated for kidney stones between 2001 and 2013 at a single tertiary institution and had 24-hour urine collections were included in the study. Patients with type 1 diabetes or taking either thiazide diuretics or alkalinizing agents were excluded. Analysis was performed in IBM SPSS Statistics version 20 using multivariate regression, and Kruskal-Wallis testing was used. **RESULTS**: Nine hundred fifty-five patients were included in this study-268 (28%) with type 2 diabetes mellitus, of whom 53 (19.8%) used insulin. Patients with diabetes had lower urine pH, calcium, and phosphate when compared with the control group, but no significant differences were found between the diabetes groups. Multivariate analysis found that HbA1c had a positive correlation with citrate (P = .008), creatinine (P = .037), urine volume (P = .044), and a trend toward a positive association with urinary calcium calcium (P = .064). Insulin use did not have a significant relationship with urinary parameters but trended toward an inverse relationship with calcium (P = .051). pH was not a significant predictor of any urine constituent. **CONCLUSION**: In an ethnically diverse inner city patient population, patients with diabetes mellitus type 2 who use insulin have no significant differences in urine parameters when compared with those on oral hypoglycemics. Worsening glucose control as measured by HbA1c levels predicts increased urine citrate and volume. Copyright © 2015 Elsevier Inc. All rights reserved.

**URL:** <https://ovidsp.ovid.com/ovidweb.cgi?T=JS&CSC=Y&NEWS=N&PAGE=fulltext&D=med13&DO=10.1016%2fj.urology.2015.10.013> <https://libkey.io/libraries/2592/openurl?genre=article&sid=OVID:medline&id=pmid:26525960&id=doi:10.1016%2Fj.urology.2015.10.013&issn=0090-4295&isbn=&volume=87&issue=&spage=52&pages=52-9&date=2016&title=Urology&atitle=The+Effect+of+Disease+Severity+on+24-Hour+Urine+Parameters+in+Kidney+Stone+Patients+With+Type+2+Diabetes.&aulast=Fram&pid=%3Cauthor%3EFram+EB%3BMoazami+S%3BStern+JM%3C%2Fauthor%3E%3CAN%3E26525960%3C%2FAN%3E%3CDT%3EJournal+Article%3C%2FDT%3E>

100. **Effect of L-Methionine on the Risk of Phosphate Stone Formation.**

**Item Type:** Journal Article

**Authors:** Siener, Roswitha;Struwe, Florian and Hesse, Albrecht

**Publication Date:** De ,2016

**Journal:** Urology 98, pp. 39–43

**Abstract: OBJECTIVE**: To determine the physiological effects following administration of the sulfur-containing amino acid L-methionine on urinary risk factors for calcium oxalate and phosphate stone formation under controlled, standardized conditions. **SUBJECTS AND METHODS**: Twelve healthy subjects were studied while consuming a standardized diet for 6 days. Day 5 was considered as control day. On day 6 (test day), participants received 1500 mg L-methionine at 9 AM. On the control and test days, 24-hour fractional urine collections were obtained. **RESULTS**: After single-dose administration of L-methionine, 24-hour fractional urinary pH decreased to values between 5.98 and 6.32. Urinary sulphate excretion, a direct marker for the metabolism of L-methionine, and ammonium excretion, which reflects the increase in net acid production, increased significantly after L-methionine administration. Urinary calcium excretion did not differ between control and test day (5.55 vs 5.45 mmol/24 hours; P = .754). The relative supersaturation of brushite and struvite decreased significantly by 25% (P = .010) and 34% (P = .049), respectively, whereas the risk of calcium oxalate stone formation was not affected by application of L-methionine (P = .477). **CONCLUSION**: The present findings show that L-methionine effectively decreases urinary pH and the risk of struvite and calcium phosphate stone formation in healthy subjects. Hypercalciuria is not expected to occur at physiological doses of L-methionine. Copyright © 2016 Elsevier Inc. All rights reserved.

**URL:** <https://ovidsp.ovid.com/ovidweb.cgi?T=JS&CSC=Y&NEWS=N&PAGE=fulltext&D=med13&DO=10.1016%2fj.urology.2016.08.007> <https://libkey.io/libraries/2592/openurl?genre=article&sid=OVID:medline&id=pmid:27521063&id=doi:10.1016%2Fj.urology.2016.08.007&issn=0090-4295&isbn=&volume=98&issue=&spage=39&pages=39-43&date=2016&title=Urology&atitle=Effect+of+L-Methionine+on+the+Risk+of+Phosphate+Stone+Formation.&aulast=Siener&pid=%3Cauthor%3ESiener+R%3BStruwe+F%3BHesse+A%3C%2Fauthor%3E%3CAN%3E27521063%3C%2FAN%3E%3CDT%3EJournal+Article%3C%2FDT%3E>

101. **Calculated insulin resistance correlates with stone-forming urinary metabolic changes and greater stone burden in high-risk stone patients.**

**Item Type:** Journal Article

**Authors:** Tran, Timothy Y.;Flynn, Mary;O'Bell, John and Pareek, Gyan

**Publication Date:** Ju ,2016

**Journal:** Clinical Nephrology 85(6), pp. 316–320

**Abstract: AIMS**: Metabolic syndrome and diabetes are associated with nephrolithiasis. Proposed mechanisms of lithogenesis include insulin resistance causing low urine pH and hyperinsulinemia leading to hypercalciuria. Herein, we sought to determine whether insulin resistance was associated with differences in stone burden and lithogenic changes on 24-hour urine samples. **MATERIALS AND METHODS**: All patients that underwent comprehensive metabolic workup including 24-hour urine samples and fasting insulin levels were included. Insulin resistance was defined as a homeostasis model assessment of insulin resistance value > 5 (HOMA-IR = (glucosexinsulin)/405). Patients on active metabolic therapy were excluded or the 24-hour urine sample predating treatment was utilized for analysis. Stone burden was determined by totaling the maximal diameter of all stones noted on CT. **RESULTS**: 18 of 30 patients (60.0%) had HOMA-IR > 5. Among patients with calculated insulin resistance, stone burden was greater (17.6 mm vs. 6.3 mm, p = 0.002) and 24-hour urine samples revealed higher urine calcium (293 mg/d vs. 159 mg/d, p = 0.02) and lower urine pH and citrate (454 mg/d vs. 639 mg/d, p = 0.04 and 5.83 vs. 6.33, p = 0.04, respectively). **CONCLUSIONS**: Previous studies have demonstrated a correlation between metabolic syndrome, diabetes, and nephrolithiasis. This report demonstrates a quantitative increase in stone burden among patients with calculated insulin resistance. The pathway for this greater stone burden may be related to the urinary metabolic changes noted among patients with insulin resistance. In the future, targeting reduction of fasting insulin levels may represent a key element of stone disease prevention.

**URL:** <https://ovidsp.ovid.com/ovidweb.cgi?T=JS&CSC=Y&NEWS=N&PAGE=fulltext&D=med13&DO=10.5414%2fCN108832> <https://libkey.io/libraries/2592/openurl?genre=article&sid=OVID:medline&id=pmid:27125629&id=doi:10.5414%2FCN108832&issn=0301-0430&isbn=&volume=85&issue=6&spage=316&pages=316-20&date=2016&title=Clinical+Nephrology&atitle=Calculated+insulin+resistance+correlates+with+stone-forming+urinary+metabolic+changes+and+greater+stone+burden+in+high-risk+stone+patients.&aulast=Tran&pid=%3Cauthor%3ETran+TY%3BFlynn+M%3BO%27Bell+J%3BPareek+G%3C%2Fauthor%3E%3CAN%3E27125629%3C%2FAN%3E%3CDT%3EJournal+Article%3C%2FDT%3E>

102. **Difference in 24-Hour Urine Composition between Diabetic and Non-Diabetic Adults without Nephrolithiasis.**

**Item Type:** Journal Article

**Authors:** Zhu, Wei;Mai, Zanlin;Qin, Jing;Duan, Xiaolu;Liu, Yang;Zhao, Zhijian;Yuan, Jian;Wan, Shaw P. and Zeng, Guohua

**Publication Date:** 2016

**Journal:** PLoS ONE [Electronic Resource] 11(2), pp. e0150006

**Abstract: BACKGROUND**: Diabetic patients are more likely to develop kidney stones than the general population. The underlying mechanisms for this disparity remain to be elucidated. Little is known about the relationship between urine composition and diabetes mellitus in non-stone-forming individuals. We sought to examine the differences in the 24-hour (24-h) urine composition between diabetic and non-diabetic adults who were not stone formers. **METHODS**: A convenience sample of 538 individuals without a history of nephrolithiasis, gout, hyperparathyroidism, or gastroenteric diseases participated in this study. The 24-h urine profiles of 115 diabetic adults were compared with those of 423 non-diabetic adults. Diabetes was defined by self-reported physician diagnosis or medication use. All participants were non-stone formers confirmed by urinary tract ultrasonography. Participants provided a fasting blood sample and a single 24-h urine collection for stone risk analysis. Student's t-test was used to compare mean urinary values. Linear regression models were adjusted for age, gender, body mass index, hypertension, fasting serum glucose, serum total cholesterol, estimated creatinine clearance rate and urinary factors. **RESULTS**: Univariable analysis showed that the diabetic participants had significantly higher 24-h urine volumes and lower urine calcium and magnesium excretions than non-diabetic participants (all P 0.05). The main limitation of this study is that the convenience samples and self-reported data may have been sources of bias. **CONCLUSION**: Our data showed that there were no differences in 24-h urine composition between diabetic and non-diabetic adults who are not stone formers. The reason for it might be the improved glycemic control in diabetic individuals in our study. Therefore, a tighter glycemic control might reduce stone formation in diabetic adults.

**URL:** <https://ovidsp.ovid.com/ovidweb.cgi?T=JS&CSC=Y&NEWS=N&PAGE=fulltext&D=med13&DO=10.1371%2fjournal.pone.0150006> <https://libkey.io/libraries/2592/openurl?genre=article&sid=OVID:medline&id=pmid:26906900&id=doi:10.1371%2Fjournal.pone.0150006&issn=1932-6203&isbn=&volume=11&issue=2&spage=e0150006&pages=e0150006&date=2016&title=PLoS+ONE+%5BElectronic+Resource%5D&atitle=Difference+in+24-Hour+Urine+Composition+between+Diabetic+and+Non-Diabetic+Adults+without+Nephrolithiasis.&aulast=Zhu&pid=%3Cauthor%3EZhu+W%3BMai+Z%3BQin+J%3BDuan+X%3BLiu+Y%3BZhao+Z%3BYuan+J%3BWan+SP%3BZeng+G%3C%2Fauthor%3E%3CAN%3E26906900%3C%2FAN%3E%3CDT%3EComparative+Study%3C%2FDT%3E>

103. **Retrospective review of serum and urinary lithogenic risk factors in patients with osteoporosis and osteopenia.**

**Item Type:** Journal Article

**Authors:** Arrabal-Polo, Miguel Angel;Giron-Prieto, Maria Sierra;Cano-Garcia, Maria Del Carmen;Poyatos-Andujar, Antonio;Quesada-Charneco, Miguel;Abad-Menor, Felix;Arias-Santiago, Salvador;Zuluaga-Gomez, Armando and Arrabal-Martin, Miguel

**Publication Date:** Ap ,2015

**Journal:** Urology 85(4), pp. 782–785

**Abstract: OBJECTIVE**: To analyze differences in bone remodeling markers, lithogenic factors and bone densitometry among the 3 groups of patients (controls, patients with relapsing calcium renal lithiasis, and patients with loss of bone mineral density without lithiasis). **MATERIAL AND METHODS**: This is a cross-sectional study including 203 patients who were divided in 3 groups: group 1 (controls), group 2 (patients with relapsing calcium renal lithiasis), and group 3 (patients with osteopenia and/or osteoporosis in the lumbar spine or hip). Bone densitometry, calcium-phosphorous and bone metabolism analysis, and analysis of lithogenic risk factors in fasting urine samples and 24-hour urine samples were performed. Statistical analysis was performed with SPSS 17.0. A P : This is a cross-sectional study including 203 patients who were divided in 3 groups: group 1 (controls), group 2 (patients with relapsing calcium renal lithiasis), and group 3 (patients with osteopenia and/or osteoporosis in the lumbar spine or hip). Bone densitometry, calcium-phosphorous and bone metabolism analysis, and analysis of lithogenic risk factors in fasting urine samples and 24-hour urine samples were performed. Statistical analysis was performed with SPSS 17.0. A P **RESULTS**: Patients in group 2 presented greater calcium excretion and a lower citrate excretion in 24-hour urine samples as compared with the other 2 groups. The proportion of hypercalciuria and hypocitraturia was higher in group 2. In addition, patients in group 2 presented a lower loss of bone mineral density as well as altered bone remodeling markers as compared with those in group 1. Patients in group 3 also presented alterations in urine calcium and citrate excretion with respect to the control group, with elevated fasting calcium and citrate levels and calcium-to-citrateratio. **CONCLUSION**: Lithogenic risk factors are altered in patients with osteopenia and/or osteoporosis without renal lithiasis although to a lesser extent than patients with calcium renal lithiasis. Copyright © 2015 Elsevier Inc. All rights reserved.

**URL:** <https://ovidsp.ovid.com/ovidweb.cgi?T=JS&CSC=Y&NEWS=N&PAGE=fulltext&D=med12&DO=10.1016%2fj.urology.2015.01.019> <https://libkey.io/libraries/2592/openurl?genre=article&sid=OVID:medline&id=pmid:25817102&id=doi:10.1016%2Fj.urology.2015.01.019&issn=0090-4295&isbn=&volume=85&issue=4&spage=782&pages=782-5&date=2015&title=Urology&atitle=Retrospective+review+of+serum+and+urinary+lithogenic+risk+factors+in+patients+with+osteoporosis+and+osteopenia.&aulast=Arrabal-Polo&pid=%3Cauthor%3EArrabal-Polo+MA%3BGiron-Prieto+MS%3BCano-Garcia+Mdel+C%3BPoyatos-Andujar+A%3BQuesada-Charneco+M%3BAbad-Menor+F%3BArias-Santiago+S%3BZuluaga-Gomez+A%3BArrabal-Martin+M%3C%2Fauthor%3E%3CAN%3E25817102%3C%2FAN%3E%3CDT%3EJournal+Article%3C%2FDT%3E>

104. **Factors Associated With the Lower Prevalence of Nephrolithiasis in Children Compared With Adults.**

**Item Type:** Journal Article

**Authors:** Grases, Felix;Rodriguez, Adrian;Costa-Bauza, Antonia;Saez-Torres, Concepcion;Rodrigo, Dolores;Gomez, Cristina;Mir-Perello, Concepcion and Frontera, Guiem

**Publication Date:** Se ,2015

**Journal:** Urology 86(3), pp. 587–592

**Abstract: OBJECTIVE**: To determine the reasons behind the lower prevalence of kidney stones in children by assessing urinary lithogenic parameters in healthy children, healthy adults, and 3 groups of stone-former patients. **METHODS**: The study subjects included 75 healthy adults, 105 healthy children, 62 patients with previous calcium oxalate monohydrate papillary stones, 120 patients with previous calcium oxalate monohydrate unattached stones, and 248 patients with previous calcium oxalate dihydrate stones. Twenty-four-hour urine samples were collected, and the urinary lithogenic parameters were measured. **RESULTS**: Calcium, magnesium, and phosphorous concentration differed significantly between healthy children and adults. Except citrate, all solute/creatinine ratios differed between healthy children and adults. However, these differences were much more important in the cases of calcium and magnesium. The calcium/creatinine ratio was 2-fold lower, whereas the magnesium/creatinine ratio was 2-fold higher, in healthy children than that in healthy adults (P : Calcium, magnesium, and phosphorous concentration differed significantly between healthy children and adults. Except citrate, all solute/creatinine ratios differed between healthy children and adults. However, these differences were much more important in the cases of calcium and magnesium. The calcium/creatinine ratio was 2-fold lower, whereas the magnesium/creatinine ratio was 2-fold higher, in healthy children than that in healthy adults (P **CONCLUSION**: Ratios of calcium and magnesium to creatinine, as well as morphoanatomic factors and lifestyle habits, may explain the lower prevalence of nephrolithiasis in children than those in adults. Copyright © 2015 Elsevier Inc. All rights reserved.

**URL:** <https://ovidsp.ovid.com/ovidweb.cgi?T=JS&CSC=Y&NEWS=N&PAGE=fulltext&D=med12&DO=10.1016%2fj.urology.2015.06.028> <https://libkey.io/libraries/2592/openurl?genre=article&sid=OVID:medline&id=pmid:26144339&id=doi:10.1016%2Fj.urology.2015.06.028&issn=0090-4295&isbn=&volume=86&issue=3&spage=587&pages=587-92&date=2015&title=Urology&atitle=Factors+Associated+With+the+Lower+Prevalence+of+Nephrolithiasis+in+Children+Compared+With+Adults.&aulast=Grases&pid=%3Cauthor%3EGrases+F%3BRodriguez+A%3BCosta-Bauza+A%3BSaez-Torres+C%3BRodrigo+D%3BGomez+C%3BMir-Perello+C%3BFrontera+G%3C%2Fauthor%3E%3CAN%3E26144339%3C%2FAN%3E%3CDT%3EComparative+Study%3C%2FDT%3E>

105. **Predicting Patients with Inadequate 24- or 48-Hour Urine Collections at Time of Metabolic Stone Evaluation.**

**Item Type:** Journal Article

**Authors:** McGuire, Barry B.;Bhanji, Yasin;Sharma, Vidit;Frainey, Brendan T.;McClean, Megan;Dong, Caroline;Rimar, Kalen;Perry, Kent T. and Nadler, Robert B.

**Publication Date:** Ju ,2015

**Journal:** Journal of Endourology 29(6), pp. 730–735

**Abstract: PURPOSE**: We aimed to understand the characteristics of patients who are less likely to submit adequate urine collections at metabolic stone evaluation. **METHODS**: Inadequate urine collection was defined using two definitions: (1) Reference ranges for 24-hour creatinine/kilogram (Cr/24) and (2) discrepancy in total 24-hour urine Cr between 24-hour urine collections. There were 1502 patients with >=1 kidney stone between 1998 and 2014 who performed a 24- or 48-hour urine collection at Northwestern Memorial Hospital and who were identified retrospectively. Multivariate analysis was performed to analyze predictor variables for adequate urine collection. **RESULTS**: A total of 2852 urine collections were analyzed. Mean age for males was 54.4 years (range 17-86), and for females was 50.2 years (range 8-90). One patient in the study was younger than 17 years old. (1) Analysis based on the Cr 24/kg definition: There were 50.7% of patients who supplied an inadequate sample. Females were nearly 50% less likely to supply an adequate sample compared with men, P=40%, and this was defined as the threshold for an inadequate sample. Female sex (OR 0.73 [0.54-0.98], P=0.037) predicted supplying inadequate samples. Adequate collections were more likely to be received on a Sunday (OR 1.6 [1.03-2.58], P=0.038) and by sedentary workers (OR 2.3 [1.12-4.72], P=0.023). **CONCLUSION**: Urine collections from patients during metabolic evaluation for nephrolithiasis may be considered inadequate based on two commonly used clinical definitions. This may have therapeutic or economic ramifications and the propensity for females to supply inadequate samples should be investigated further.

**URL:** <https://ovidsp.ovid.com/ovidweb.cgi?T=JS&CSC=Y&NEWS=N&PAGE=fulltext&D=med12&DO=10.1089%2fend.2014.0544> <https://libkey.io/libraries/2592/openurl?genre=article&sid=OVID:medline&id=pmid:25423010&id=doi:10.1089%2Fend.2014.0544&issn=0892-7790&isbn=&volume=29&issue=6&spage=730&pages=730-5&date=2015&title=Journal+of+Endourology&atitle=Predicting+Patients+with+Inadequate+24-+or+48-Hour+Urine+Collections+at+Time+of+Metabolic+Stone+Evaluation.&aulast=McGuire&pid=%3Cauthor%3EMcGuire+BB%3BBhanji+Y%3BSharma+V%3BFrainey+BT%3BMcClean+M%3BDong+C%3BRimar+K%3BPerry+KT%3BNadler+RB%3C%2Fauthor%3E%3CAN%3E25423010%3C%2FAN%3E%3CDT%3EEvaluation+Study%3C%2FDT%3E>

106. **Effect of Demographics on Excretion of Key Urinary Factors Related to Kidney Stone Risk.**

**Item Type:** Journal Article

**Authors:** Perinpam, Majuran;Ware, Erin B.;Smith, Jennifer A.;Turner, Stephen T.;Kardia, Sharon L. R. and Lieske, John C.

**Publication Date:** Oc ,2015

**Journal:** Urology 86(4), pp. 690–696

**Abstract: OBJECTIVE**: To investigate the effect of demographics including age and sex on excretion of 4 key urinary factors (calcium [Ca], magnesium [Mg], oxalate and uric acid [UA]) related to kidney stone risk. **METHODS**: Twenty-four-hour urine samples were collected from non-Hispanic white sibships in Rochester, MN. Height, weight, blood pressure, serum creatinine, and cystatin C were measured. Diet was assessed using the Viocare food frequency questionnaire. Effects of demographics and dietary elements on urinary excretions were evaluated in univariate, multivariate, and interaction models that included age, sex, and body mass index (BMI). **RESULTS**: Samples were available from 709 individuals. In multivariate models, sex was a significant predictor of all 4 urinary factors, age was significant for all but UA excretion, and serum creatinine was significant only for Ca and Mg excretion (P : Samples were available from 709 individuals. In multivariate models, sex was a significant predictor of all 4 urinary factors, age was significant for all but UA excretion, and serum creatinine was significant only for Ca and Mg excretion (P : Samples were available from 709 individuals. In multivariate models, sex was a significant predictor of all 4 urinary factors, age was significant for all but UA excretion, and serum creatinine was significant only for Ca and Mg excretion (P : Samples were available from 709 individuals. In multivariate models, sex was a significant predictor of all 4 urinary factors, age was significant for all but UA excretion, and serum creatinine was significant only for Ca and Mg excretion (P **CONCLUSION**: Age and sex influence excretion of key urinary factors related to kidney stone risk and should be taken into account when evaluating kidney stone patients. Copyright © 2015 Elsevier Inc. All rights reserved.

**URL:** <https://ovidsp.ovid.com/ovidweb.cgi?T=JS&CSC=Y&NEWS=N&PAGE=fulltext&D=med12&DO=10.1016%2fj.urology.2015.07.012> <https://libkey.io/libraries/2592/openurl?genre=article&sid=OVID:medline&id=pmid:26206452&id=doi:10.1016%2Fj.urology.2015.07.012&issn=0090-4295&isbn=&volume=86&issue=4&spage=690&pages=690-6&date=2015&title=Urology&atitle=Effect+of+Demographics+on+Excretion+of+Key+Urinary+Factors+Related+to+Kidney+Stone+Risk.&aulast=Perinpam&pid=%3Cauthor%3EPerinpam+M%3BWare+EB%3BSmith+JA%3BTurner+ST%3BKardia+SL%3BLieske+JC%3C%2Fauthor%3E%3CAN%3E26206452%3C%2FAN%3E%3CDT%3EJournal+Article%3C%2FDT%3E>

107. **100% uric acid stone formers: what makes them different?.**

**Item Type:** Journal Article

**Authors:** Reichard, Chad;Gill, Bradley C.;Sarkissian, Carl;De, Shubha and Monga, Manoj

**Publication Date:** Fe ,2015

**Journal:** Urology 85(2), pp. 296–298

**Abstract: OBJECTIVE**: To identify what risk factors on 24-hour urinalysis, if any, predispose patients to have higher percentages of uric acid (UA) stone composition in their stones, with specific emphasis on patients with pure UA stones. **METHODS**: We retrospectively identified 308 patients from review of a kidney stone analysis database. Patients were grouped according to the percentage UA composition: 10%-20%, 30%-50%, 60%-90%, and 100% UA. Data were extracted from 24-hour urine collections and serum chemistries. Patients taking allopurinol, citrates, or thiazide diuretics were excluded. **RESULTS**: The percentage UA stone composition increased as patients became older (P = .05) or heavier (P : The percentage UA stone composition increased as patients became older (P = .05) or heavier (P : The percentage UA stone composition increased as patients became older (P = .05) or heavier (P **CONCLUSION**: Older and heavier patients with higher serum UA levels are more likely to have a pure UA stone. This information combined with traditional predictors (urine pH, radiopacity of stone, and Hounsfield units) may help identify those most likely to respond to dissolution therapy. Copyright © 2015 Elsevier Inc. All rights reserved.

**URL:** <https://ovidsp.ovid.com/ovidweb.cgi?T=JS&CSC=Y&NEWS=N&PAGE=fulltext&D=med12&DO=10.1016%2fj.urology.2014.10.029> <https://libkey.io/libraries/2592/openurl?genre=article&sid=OVID:medline&id=pmid:25623669&id=doi:10.1016%2Fj.urology.2014.10.029&issn=0090-4295&isbn=&volume=85&issue=2&spage=296&pages=296-8&date=2015&title=Urology&atitle=100%25+uric+acid+stone+formers%3A+what+makes+them+different%3F.&aulast=Reichard&pid=%3Cauthor%3EReichard+C%3BGill+BC%3BSarkissian+C%3BDe+S%3BMonga+M%3C%2Fauthor%3E%3CAN%3E25623669%3C%2FAN%3E%3CDT%3EJournal+Article%3C%2FDT%3E>

108. **Calcium oxalate supersaturation increases early after Roux-en-Y gastric bypass.**

**Item Type:** Journal Article

**Authors:** Agrawal, Varun;Liu, Xiao J.;Campfield, Thomas;Romanelli, John;Enrique Silva, J. and Braden, Gregory L.

**Publication Date:** 2014

**Journal:** Surgery for Obesity & Related Diseases 10(1), pp. 88–94

**Abstract: BACKGROUND**: Calcium oxalate (CaOx) nephrolithiasis is an adverse effect of Roux-en-Y gastric bypass surgery (RYGB). It is unknown when the increased risk for CaOx stone formation occurs after surgery. **METHODS**: We studied 13 morbidly obese adults undergoing RYGB with 24-hour urine collections at 4 weeks before and 1, 2, 4, and 6 months after surgery and computed CaOx relative saturation ratio (RSR) by EQUIL2. **RESULTS**: Eleven patients were female, mean +/- standard deviation age was 41.1 +/- 7.2 years, and none had diabetes or chronic kidney disease. Median (interquartile range) urinary oxalate excretion increased linearly from 12.6 (10.9-37.9) mg/24 hr at baseline to 28.4 (14.4-44.0) mg/24 hr at 6 months (slope = .188; P = .005). CaOx RSR increased significantly at 2 months after RYGB (1.4 [1.2-2.4] to 4.9 [1.7-10.0]; P = .017) and rose throughout the study to 5.7 (3.7-12.2) at 6 months (P = .001) with a positive linear slope (.255; P = .001). One patient had critical CaOx supersaturation (RSR = 34.7) and severe hyperoxaluria (101.7 mg/24 hr) at 6 months after RYGB. Significant decreases over time were seen in urine volume and sodium and potassium excretion, but no changes were noted in urinary pH, calcium, magnesium, or citrate. **CONCLUSIONS**: Our data suggest that CaOx RSR, and thus risk for nephrolithiasis, rises as early as 2 months after RYGB and increases gradually in the first 6 months, largely because of reduced urine volume and increased urinary oxalate excretion. Interventions to reduce CaOx RSR, such as adequate fluid intake and agents to bind enteric oxalate, need to be evaluated in patients at risk for nephrolithiasis after RYGB. Copyright © 2014 American Society for Metabolic and Bariatric Surgery. Published by Elsevier Inc. All rights reserved.

**URL:** <https://ovidsp.ovid.com/ovidweb.cgi?T=JS&CSC=Y&NEWS=N&PAGE=fulltext&D=med11&DO=10.1016%2fj.soard.2013.03.014> <https://libkey.io/libraries/2592/openurl?genre=article&sid=OVID:medline&id=pmid:23953730&id=doi:10.1016%2Fj.soard.2013.03.014&issn=1550-7289&isbn=&volume=10&issue=1&spage=88&pages=88-94&date=2014&title=Surgery+for+Obesity+%26+Related+Diseases&atitle=Calcium+oxalate+supersaturation+increases+early+after+Roux-en-Y+gastric+bypass.&aulast=Agrawal&pid=%3Cauthor%3EAgrawal+V%3BLiu+XJ%3BCampfield+T%3BRomanelli+J%3BEnrique+Silva+J%3BBraden+GL%3C%2Fauthor%3E%3CAN%3E23953730%3C%2FAN%3E%3CDT%3EJournal+Article%3C%2FDT%3E>

109. **Predisposing factors for infantile urinary calculus in south-west of Iran.**

**Item Type:** Journal Article

**Authors:** Alemzadeh-Ansari, Mohammad Hasan;Valavi, Ehsan and Ahmadzadeh, Ali

**Publication Date:** Ja ,2014

**Journal:** Iranian Journal of Kidney Diseases 8(1), pp. 53–57

**Abstract: INTRODUCTION**: Urinary calculi in infants are relatively infrequent, but their incidence has increased in the recent decades. The aim of this study was to investigate the clinical presentation, metabolic risk factors, and urinary tract abnormalities in infants suffering from kidney calculus. **MATERIALS AND METHODS**: A total of 152 infants were admitted between 2009 and 2012 with ultrasonography-proven urolithiasis. A Foley catheter was fixed and 24-hour urine samples were analyzed for calcium, citrate, oxalate, uric acid, and magnesium. For detecting cystinuria, qualitative measurement of urinary cystine was done by nitroprusside test. Urinary tract structural abnormalities were also evaluated. **RESULTS**: The mean age at the diagnosis of kidney calculus was 5.46 months (range, 15 days to 12 months). The most common clinical findings were restlessness and urinary tract infection. A family history of calculi was found in 67.1% of the patients and 68.4% were born to consanguineous marriages. Metabolic abnormalities and urinary tract abnormalities were found in 96.1% and 15.1% of children, respectively. Urinary tract abnormalities were more common in girls. The most common metabolic risk factors were hypercalciuria (79.6%) and hypocitraturia (40.9%). Hyperoxaluria and hypomagnesuria were found in about 28% of patients, both of which were associated with bilateral urolithiasis. **CONCLUSIONS**: These findings show that urinary metabolic abnormalities are very common in infants with urolithiasis. Appropriate evaluation of urinary metabolic parameters can lead us to proper diagnosis and treatment.

**URL:** <https://ovidsp.ovid.com/ovidweb.cgi?T=JS&CSC=Y&NEWS=N&PAGE=fulltext&D=med11&AN=24413722> <https://libkey.io/libraries/2592/openurl?genre=article&sid=OVID:medline&id=pmid:24413722&id=doi:&issn=1735-8582&isbn=&volume=8&issue=1&spage=53&pages=53-7&date=2014&title=Iranian+journal+of+Kidney+Diseases&atitle=Predisposing+factors+for+infantile+urinary+calculus+in+south-west+of+Iran.&aulast=Alemzadeh-Ansari&pid=%3Cauthor%3EAlemzadeh-Ansari+MH%3BValavi+E%3BAhmadzadeh+A%3C%2Fauthor%3E%3CAN%3E24413722%3C%2FAN%3E%3CDT%3EJournal+Article%3C%2FDT%3E>

110. **The clinical pattern of primary hyperoxaluria in pediatric patient at Queen Rania Abdulla Children Hospital.**

**Item Type:** Journal Article

**Authors:** Almardini, Reham I.;Alfarah, Mahdi G. and Salaita, Ghazi M.

**Publication Date:** Ma ,2014

**Journal:** Arab Journal of Nephrology and Transplantation 7(2), pp. 119–123

**Abstract: INTRODUCTION**: Hyperoxaluria is a metabolic disorder that can lead to end stage renal disease (ESRD). It can be either inherited or acquired. Primary hyperoxaluria (PHO) is more common and characterized by an excessive production of oxalate leading to recurrent urolithiasis and progressive nephrocalcinosis. Due to the high rate of consanguineous marriage in Jordan this disease is commonly diagnosed in pediatric nephrology clinics. We aimed to demonstrate the clinical pattern and progression to ESRD in pediatric patients with hyperoxaluria at Queen Rania Abdulla Children Hospital. **METHODS**: Medical records of all patients followed up in the pediatric nephrology clinic with the diagnosis of PHO during the period between September 2007 and March 2013 were reviewed. **RESULTS**: There were 70 patients with the diagnosis of PHO, 52.9% were males. The median age at presentation was 3 years +/- 3 months with the youngest child being two months old. Diagnosis was made in the first year of life in 15.7% of patients. The most common presenting symptom was hematuria, while 14% of patients were asymptomatic and detected by family screening after the diagnosis of an index case. At the time of initial presentation, 15.7% of patients had ESRD and 25% had impaired renal function. Kidney stones were found in 57% of cases and nephrocalcinosis was found in 37%. **CONCLUSION**: High index of suspicion is needed to diagnose PHO in children presenting with kidney stone or unexplained hematuria. Twenty-four hour urine collection for oxalate are required to make the proper diagnosis. Family screening, when appropriate, is indicated for early detection of PHO.

**URL:** <https://ovidsp.ovid.com/ovidweb.cgi?T=JS&CSC=Y&NEWS=N&PAGE=fulltext&D=med11&AN=25366508> <https://libkey.io/libraries/2592/openurl?genre=article&sid=OVID:medline&id=pmid:25366508&id=doi:&issn=1858-554X&isbn=&volume=7&issue=2&spage=119&pages=119-23&date=2014&title=Arab+Journal+of+Nephrology+and+Transplantation&atitle=The+clinical+pattern+of+primary+hyperoxaluria+in+pediatric+patient+at+Queen+Rania+Abdulla+Children+Hospital.&aulast=Almardini&pid=%3Cauthor%3EAlmardini+RI%3BAlfarah+MG%3BSalaita+GM%3C%2Fauthor%3E%3CAN%3E25366508%3C%2FAN%3E%3CDT%3EJournal+Article%3C%2FDT%3E>

111. **Incidence of hypercalciuria and hypercalcemia during vitamin D and calcium supplementation in older women.**

**Item Type:** Journal Article

**Authors:** Gallagher, John Christopher;Smith, Lynette M. and Yalamanchili, Vinod

**Publication Date:** No ,2014

**Journal:** Menopause 21(11), pp. 1173–1180

**Abstract: OBJECTIVE**: This study aims to prospectively assess the incidence of hypercalciuria and hypercalcemia with different doses of vitamin D and with a calcium intake of approximately 1,200 mg/day. **METHODS**: This was a 1-year randomized placebo-controlled study of vitamin D (400-4,800 IU/d) in 163 white women aged 57 to 90 years. Calcium citrate tablets (200 mg) were added to the diet to achieve a total calcium intake of approximately 1,200 mg/day in all groups. All women had vitamin D insufficiency at baseline, with serum 25-hydroxyvitaminD levels lower than 20 ng/mL (50 nmol/L). Serum and 24-hour urine calcium were collected every 3 months on supplementation, any test result above the upper reference range represented an episode of hypercalcemia or hypercalciuria. Mixed-effects models and multivariate logistic regression were used in the analysis. **RESULTS**: Hypercalcemia (>10.2 mg/dL [2.55 mmol/L]) occurred in 8.8% of white women. Hypercalciuria (>300 mg/d [7.5 mmol]) occurred in 30.6% of white women. Episodes of hypercalciuria were transient in half of the group and recurrent in the other half. No relationship between hypercalcemia or hypercalciuria and vitamin D dose was found, and hypercalciuria was equally common in the placebo group. **CONCLUSIONS**: Hypercalciuria and hypercalcemia commonly occur with vitamin D and calcium supplements. Whether hypercalciuria and hypercalcemia are caused by calcium, vitamin D, or both is unclear. These findings may have relevance to the reported increase in kidney stones in the Women's Health Initiative trial. Because calcium 1,200 mg and vitamin D 800 IU/day are widely recommended in postmenopausal women, systematic evaluation of the safety of supplements is warranted in clinical management and in future studies.

**URL:** <https://ovidsp.ovid.com/ovidweb.cgi?T=JS&CSC=Y&NEWS=N&PAGE=fulltext&D=med11&DO=10.1097%2fGME.0000000000000270> <https://libkey.io/libraries/2592/openurl?genre=article&sid=OVID:medline&id=pmid:24937025&id=doi:10.1097%2FGME.0000000000000270&issn=1072-3714&isbn=&volume=21&issue=11&spage=1173&pages=1173-80&date=2014&title=Menopause&atitle=Incidence+of+hypercalciuria+and+hypercalcemia+during+vitamin+D+and+calcium+supplementation+in+older+women.&aulast=Gallagher&pid=%3Cauthor%3EGallagher+JC%3BSmith+LM%3BYalamanchili+V%3C%2Fauthor%3E%3CAN%3E24937025%3C%2FAN%3E%3CDT%3EJournal+Article%3C%2FDT%3E>

112. **Heritability of urinary traits that contribute to nephrolithiasis.**

**Item Type:** Journal Article

**Authors:** Lieske, John C.;Turner, Stephen T.;Edeh, Samuel N.;Smith, Jennifer A. and Kardia, Sharon L. R.

**Publication Date:** Ma ,2014

**Journal:** Clinical Journal of the American Society of Nephrology: CJASN 9(5), pp. 943–950

**Abstract: BACKGROUND AND OBJECTIVES**: Kidney stones and their risk factors aggregate in families, yet few studies have systematically estimated heritabilities and genetic correlations of the numerous urinary traits associated with risk of kidney stones. **DESIGN, SETTING, PARTICIPANTS, & MEASUREMENTS**: Twenty-four-hour urine samples were collected from the Genetic Epidemiology Network of Arteriopathy cohort of families in Rochester, Minnesota, to measure urinary determinants of supersaturation. Diet was assessed using the Viocare food frequency questionnaire. Heritabilities and genetic correlations among the urinary traits were estimated using variance components methods. **RESULTS**: Samples were available from 811 individuals (344 men, 467 women; mean age 66 +/- 9 years). Age, sex, and weight were significantly correlated with the majority of urinary traits. Many urine excretions (calcium, magnesium, citrate excretion) had strong evidence for heritability (P: Samples were available from 811 individuals (344 men, 467 women; mean age 66 +/- 9 years). Age, sex, and weight were significantly correlated with the majority of urinary traits. Many urine excretions (calcium, magnesium, citrate excretion) had strong evidence for heritability (P: Samples were available from 811 individuals (344 men, 467 women; mean age 66 +/- 9 years). Age, sex, and weight were significantly correlated with the majority of urinary traits. Many urine excretions (calcium, magnesium, citrate excretion) had strong evidence for heritability (P**CONCLUSIONS**: Evidence from this cohort suggests a strong heritable component to many urinary nephrolithiasis risk factors. Further study of genetic influences on urinary traits relevant for kidney stone pathogenesis is warranted.

**URL:** <https://ovidsp.ovid.com/ovidweb.cgi?T=JS&CSC=Y&NEWS=N&PAGE=fulltext&D=med11&DO=10.2215%2fCJN.08210813> <https://libkey.io/libraries/2592/openurl?genre=article&sid=OVID:medline&id=pmid:24578335&id=doi:10.2215%2FCJN.08210813&issn=1555-9041&isbn=&volume=9&issue=5&spage=943&pages=943-50&date=2014&title=Clinical+Journal+of+The+American+Society+of+Nephrology%3A+CJASN&atitle=Heritability+of+urinary+traits+that+contribute+to+nephrolithiasis.&aulast=Lieske&pid=%3Cauthor%3ELieske+JC%3BTurner+ST%3BEdeh+SN%3BSmith+JA%3BKardia+SL%3C%2Fauthor%3E%3CAN%3E24578335%3C%2FAN%3E%3CDT%3EJournal+Article%3C%2FDT%3E>

113. **Erythrocyte oxidative stress in patients with calcium oxalate stones correlates with stone size and renal tubular damage.**

**Item Type:** Journal Article

**Authors:** Ma, Ming-Chieh;Chen, Yih-Sharng and Huang, Ho-Shiang

**Publication Date:** Fe ,2014

**Journal:** Urology 83(2), pp. 510.e9–510.17

**Abstract: OBJECTIVE**: To investigate how erythrocyte oxidative stress relates to renal tubular damage and calcium oxalate stone size in patients as oxidative stress has been demonstrated to be associated with stone formation in disease progression. **METHODS**: The study included 29 controls, 29 patients with kidney stones, and 28 patients with ureteral stones. Venous blood samples were collected to measure the expression and activity of antioxidant enzymes in the isolated erythrocytes. A 24-hour urine sample was collected to measure urinary chemistry. The cellular levels of oxalate and the oxidative stress marker malondialdehyde (MDA) were determined to examine their correlations with stone size and renal tubule damage. **RESULTS**: Calcium oxalate stone deposition and high free radical levels in venous blood associated with high levels of urinary oxalate, glutathione S-transferases tubular damage markers, and MDA and low urinary citrate levels. Compared with the erythrocytes of controls, the erythrocytes of stone groups had significantly lower levels and activities of antioxidant proteins, namely, reduced glutathione, catalase, and copper- or zinc-superoxide dismutase. The ureteral stone group also had significantly lower erythrocyte glutathione peroxidase levels and glutathione reductase activity than the controls. Erythrocyte oxalate levels correlated positively with erythrocyte MDA levels and negatively with erythrocyte antioxidant protein activities. Erythrocyte oxidative stress, as indicated by cellular MDA levels, also correlated well with urinary glutathione S-transferases and stone size. **CONCLUSION**: These results suggest that oxalate-mediated oxidative stress in erythrocytes might contribute to the tubular damage and stone accumulation in patients with hyperoxaluria. Copyright © 2014 Elsevier Inc. All rights reserved.

**URL:** <https://ovidsp.ovid.com/ovidweb.cgi?T=JS&CSC=Y&NEWS=N&PAGE=fulltext&D=med11&DO=10.1016%2fj.urology.2013.09.050> <https://libkey.io/libraries/2592/openurl?genre=article&sid=OVID:medline&id=pmid:24360074&id=doi:10.1016%2Fj.urology.2013.09.050&issn=0090-4295&isbn=&volume=83&issue=2&spage=510.e9&pages=510.e9-17&date=2014&title=Urology&atitle=Erythrocyte+oxidative+stress+in+patients+with+calcium+oxalate+stones+correlates+with+stone+size+and+renal+tubular+damage.&aulast=Ma&pid=%3Cauthor%3EMa+MC%3BChen+YS%3BHuang+HS%3C%2Fauthor%3E%3CAN%3E24360074%3C%2FAN%3E%3CDT%3EJournal+Article%3C%2FDT%3E>

114. **Prevalence of 24-hour urine collection in high risk stone formers.**

**Item Type:** Journal Article

**Authors:** Milose, Jaclyn C.;Kaufman, Samuel R.;Hollenbeck, Brent K.;Wolf, J. Stuart Jr and Hollingsworth, John M.

**Publication Date:** Fe ,2014

**Journal:** Journal of Urology 191(2), pp. 376–380

**Abstract: PURPOSE**: Secondary prevention has an important role in urinary stone disease. The core of secondary prevention is the identification of modifiable risk factors by a 24-hour urine collection, which then directs selective medical therapy. While this decreases the recurrence rate, little is known about the frequency with which 24-hour urine collections are obtained. **MATERIALS AND METHODS**: Using medical claims from 2002 to 2006 we identified adults with incident urinary stone episodes. With appropriate diagnosis codes we determined those at high risk for recurrence. Of these patients we determined the proportion in whom a 24-hour urine collection was done within 6 months of diagnosis. Finally, we fitted regression models to measure associations between patient and provider level factors, and obtaining a 24-hour urine collection. **RESULTS**: We identified 28,836 patients at high risk for recurrence. The prevalence of 24-hour urine testing increased from 7.0% in 2003 to 7.9% in 2006 (p = 0.011), although the overall prevalence was exceedingly low at 7.4%. Multivariable regression revealed that region of residence and level of comorbid illness were independently associated with 24-hour urine collection, as was the type of physician who performed the followup. For instance, the odds of metabolic evaluation were 2.9 times higher when a patient was seen by a nephrologist (OR 2.92, 95% CI 2.32-3.67), and more than threefold higher when seen by a urologist (OR 3.87, 95% CI 3.48-4.30). **CONCLUSIONS**: Obtaining 24-hour urine collections in stone formers at high risk is uncommon, raising a quality of care concern. Copyright © 2014 American Urological Association Education and Research, Inc. Published by Elsevier Inc. All rights reserved.

**URL:** <https://ovidsp.ovid.com/ovidweb.cgi?T=JS&CSC=Y&NEWS=N&PAGE=fulltext&D=med11&DO=10.1016%2fj.juro.2013.08.080> <https://libkey.io/libraries/2592/openurl?genre=article&sid=OVID:medline&id=pmid:24018242&id=doi:10.1016%2Fj.juro.2013.08.080&issn=0022-5347&isbn=&volume=191&issue=2&spage=376&pages=376-80&date=2014&title=Journal+of+Urology&atitle=Prevalence+of+24-hour+urine+collection+in+high+risk+stone+formers.&aulast=Milose&pid=%3Cauthor%3EMilose+JC%3BKaufman+SR%3BHollenbeck+BK%3BWolf+JS+Jr%3BHollingsworth+JM%3C%2Fauthor%3E%3CAN%3E24018242%3C%2FAN%3E%3CDT%3EJournal+Article%3C%2FDT%3E>

115. **Urinary lithogenic risk profile in recurrent stone formers with hyperoxaluria: a randomized controlled trial comparing DASH (Dietary Approaches to Stop Hypertension)-style and low-oxalate diets.**

**Item Type:** Journal Article

**Authors:** Noori, Nazanin;Honarkar, Elaheh;Goldfarb, David S.;Kalantar-Zadeh, Kamyar;Taheri, Maryam;Shakhssalim, Nasser;Parvin, Mahmoud and Basiri, Abbas

**Publication Date:** Ma ,2014

**Journal:** American Journal of Kidney Diseases 63(3), pp. 456–463

**Abstract: BACKGROUND**: Patients with nephrolithiasis and hyperoxaluria generally are advised to follow a low-oxalate diet. However, most people do not eat isolated nutrients, but meals consisting of a variety of foods with complex combinations of nutrients. A more rational approach to nephrolithiasis prevention would be to base dietary advice on the cumulative effects of foods and different dietary patterns rather than single nutrients. **STUDY DESIGN**: Randomized controlled trial. **SETTING & PARTICIPANTS**: Recurrent stone formers with hyperoxaluria (urine oxalate > 40 mg/d). **INTERVENTION**: The intervention group was asked to follow a calorie-controlled Dietary Approaches to Stop Hypertension (DASH)-style diet (a diet high in fruit, vegetables, whole grains, and low-fat dairy products and low in saturated fat, total fat, cholesterol, refined grains, sweets, and meat), whereas the control group was prescribed a low-oxalate diet. Study length was 8 weeks. **OUTCOMES**: Primary: change in urinary calcium oxalate supersaturation. **SECONDARY**: Changes in 24-hour urinary composition. **RESULTS**: 57 participants were randomly assigned (DASH group, 29; low-oxalate group, 28). 41 participants completed the trial (DASH group, 21; low-oxalate group, 20). As-treated analysis showed a trend for urinary oxalate excretion to increase in the DASH versus the low-oxalate group (point estimate of difference, 9.0mg/d; 95% CI, -1.1 to 19.1mg/d; P=0.08). However, there was a trend for calcium oxalate supersaturation to decrease in the DASH versus the low-oxalate group (point estimate of difference, -1.24; 95% CI, -2.80 to 0.32; P=0.08) in association with an increase in magnesium and citrate excretion and urine pH in the DASH versus low-oxalate group. **LIMITATIONS**: Limited sample size, as-treated analysis, nonsignificant results. **CONCLUSIONS**: The DASH diet might be an effective alternative to the low-oxalate diet in reducing calcium oxalate supersaturation and should be studied more. Copyright © 2014. Published by Elsevier Inc.

**URL:** <https://ovidsp.ovid.com/ovidweb.cgi?T=JS&CSC=Y&NEWS=N&PAGE=fulltext&D=med11&DO=10.1053%2fj.ajkd.2013.11.022> <https://libkey.io/libraries/2592/openurl?genre=article&sid=OVID:medline&id=pmid:24560157&id=doi:10.1053%2Fj.ajkd.2013.11.022&issn=0272-6386&isbn=&volume=63&issue=3&spage=456&pages=456-63&date=2014&title=American+Journal+of+Kidney+Diseases&atitle=Urinary+lithogenic+risk+profile+in+recurrent+stone+formers+with+hyperoxaluria%3A+a+randomized+controlled+trial+comparing+DASH+%28Dietary+Approaches+to+Stop+Hypertension%29-style+and+low-oxalate+diets.&aulast=Noori&pid=%3Cauthor%3ENoori+N%3BHonarkar+E%3BGoldfarb+DS%3BKalantar-Zadeh+K%3BTaheri+M%3BShakhssalim+N%3BParvin+M%3BBasiri+A%3C%2Fauthor%3E%3CAN%3E24560157%3C%2FAN%3E%3CDT%3EJournal+Article%3C%2FDT%3E>

116. **Adequate dietary intake and nutritional status in patients with nephrolithiasis: new targets and objectives.**

**Item Type:** Journal Article

**Authors:** Oliveira, Larissa Marques Tondin de;Hauschild, Daniela Barbieri;Leite, Christiane de Mesquita Barros Almeida;Baptista, Deise Regina and Carvalho, Mauricio

**Publication Date:** No ,2014

**Journal:** Journal of Renal Nutrition 24(6), pp. 417–422

**Abstract: OBJECTIVES**: Modifiable risk factors for kidney stones, such as diet and nutritional status, are targets in the treatment and prevention of nephrolithiasis. The aim of the present study was to assess the nutritional state and energy and nutrient adequacy of patients with nephrolithiasis via anthropometric evaluation, dietary inquiry, and urine biochemistry. **DESIGN**: The present observational cross-sectional study. **SETTING**: An outpatient, nephrolithiasis clinic of a tertiary care, university hospital. **SUBJECTS**: A total of 31 patients with nephrolithiasis and 25 controls were assessed. **INTERVENTIONS**: All patients were subjected to nutritional evaluation, including dietary inquiry and chemical tests of blood and 24-hour urine samples. **MAIN OUTCOME MEASURE**: Nutritional state and nutrient ingestion adequacy. **RESULTS**: The group of patients with nephrolithiasis exhibited high body mass index, abdominal circumference, and body fat percentage values. The protein, sodium, calcium, potassium, and oxalate intakes differed significantly from the recommended values in both the nephrolithiasis and control groups. The nephrolithiasis and control groups differed only in terms of oxalate intake (159 +/- 119.27 vs. 112 +/- 47.9, respectively, P = .042). Regarding urine biochemistry, 30% of the patients with nephrolithiasis exhibited hypercalciuria, versus 12% of the control group. **CONCLUSIONS**: The prevalence of individuals with overweight/obesity was high in both the nephrolithiasis and control groups. This finding indicates that in addition to orientation regarding nonlithogenic dietary habits, continuous education by a multidisciplinary staff must also address the prevention and treatment of obesity. Copyright © 2014 National Kidney Foundation, Inc. Published by Elsevier Inc. All rights reserved.

**URL:** <https://ovidsp.ovid.com/ovidweb.cgi?T=JS&CSC=Y&NEWS=N&PAGE=fulltext&D=med11&DO=10.1053%2fj.jrn.2014.06.003> <https://libkey.io/libraries/2592/openurl?genre=article&sid=OVID:medline&id=pmid:25091136&id=doi:10.1053%2Fj.jrn.2014.06.003&issn=1051-2276&isbn=&volume=24&issue=6&spage=417&pages=417-22&date=2014&title=Journal+of+Renal+Nutrition&atitle=Adequate+dietary+intake+and+nutritional+status+in+patients+with+nephrolithiasis%3A+new+targets+and+objectives.&aulast=Oliveira&pid=%3Cauthor%3EOliveira+LM%3BHauschild+DB%3BLeite+Cde+M%3BBaptista+DR%3BCarvalho+M%3C%2Fauthor%3E%3CAN%3E25091136%3C%2FAN%3E%3CDT%3EJournal+Article%3C%2FDT%3E>

117. **The growing prevalence of kidney stones and opportunities for prevention.**

**Item Type:** Journal Article

**Authors:** Richman, Katherine;O'Bell, John and Pareek, Gyan

**Publication Date:** Dec 02 ,2014

**Journal:** Rhode Island Medicine 97(12), pp. 31–34

**Abstract:** The prevalence of kidney stones is climbing in the United States. Several investigators have demonstrated an association between metabolic syndrome and kidney stones and some have proposed a causal link. Risk factors for nephrolithiasis can be identified with a 24-hour urine collection and preventive measures can be customized to meet the needs of individual patients. Dietary and pharmacologic interventions are available to address urinary risk factors such as inadequate urine volume, hypercalcuria, hyperoxaluria, hyperuricosuria and hypocitraturia. Given that morbidity and healthcare costs associated with nephrolithiasis are on the rise, deterring stone formation is increasingly important. Multidisciplinary clinics that foster collaboration between urologists, nephrologists and dieticians offer patients effective prevention and treatment strategies.

**URL:** <https://ovidsp.ovid.com/ovidweb.cgi?T=JS&CSC=Y&NEWS=N&PAGE=fulltext&D=med11&AN=25463625> <https://libkey.io/libraries/2592/openurl?genre=article&sid=OVID:medline&id=pmid:25463625&id=doi:&issn=0363-7913&isbn=&volume=97&issue=12&spage=31&pages=31-4&date=2014&title=Rhode+Island+Medicine&atitle=The+growing+prevalence+of+kidney+stones+and+opportunities+for+prevention.&aulast=Richman&pid=%3Cauthor%3ERichman+K%3BO%27Bell+J%3BPareek+G%3C%2Fauthor%3E%3CAN%3E25463625%3C%2FAN%3E%3CDT%3EJournal+Article%3C%2FDT%3E>

118. **Type-2 diabetes and kidney stones: impact of diabetes medications and glycemic control.**

**Item Type:** Journal Article

**Authors:** Torricelli, Fabio C. M.;De, Shubha;Gebreselassie, Surafel;Li, Ina;Sarkissian, Carl and Monga, Manoj

**Publication Date:** Se ,2014

**Journal:** Urology 84(3), pp. 544–548

**Abstract: OBJECTIVE**: To evaluate the impact of diabetic medications and glycemic control on the urine pH, 24-hour urine stone risk profile, and stone composition. **PATIENTS AND METHODS**: We retrospectively reviewed our database searching for type-2 diabetic patients with kidney stones from July 2002 to January 2013. Patients were divided in 2 groups according to their diabetic medications: insulin vs oral antihyperglycemics. Patients were compared based on their urine collections and stone composition. A linear regression was done to assess which variables could predict a low urine pH. In a subgroup analysis, patients on thiazolidinediones (ie, pioglitazone) were compared with patients on other oral antihyperglycemics. **RESULTS**: We analyzed 1831 type-2 diabetic patients with stone disease; 375 (20.5%) were included in the insulin group and 1456 (79.5%) in the antihyperglycemics group. Linear regression revealed male gender (P = .011) and insulin therapy (P : We analyzed 1831 type-2 diabetic patients with stone disease; 375 (20.5%) were included in the insulin group and 1456 (79.5%) in the antihyperglycemics group. Linear regression revealed male gender (P = .011) and insulin therapy (P : We analyzed 1831 type-2 diabetic patients with stone disease; 375 (20.5%) were included in the insulin group and 1456 (79.5%) in the antihyperglycemics group. Linear regression revealed male gender (P = .011) and insulin therapy (P : We analyzed 1831 type-2 diabetic patients with stone disease; 375 (20.5%) were included in the insulin group and 1456 (79.5%) in the antihyperglycemics group. Linear regression revealed male gender (P = .011) and insulin therapy (P **CONCLUSION**: Urine pH is inversely related to HbA1c level. Insulin therapy is associated with higher urine pH than oral antihyperglycemic agents despite higher HbA1c suggesting that insulin may modify urine pH independent of glycemic control. Copyright © 2014 Elsevier Inc. All rights reserved.

**URL:** <https://ovidsp.ovid.com/ovidweb.cgi?T=JS&CSC=Y&NEWS=N&PAGE=fulltext&D=med11&DO=10.1016%2fj.urology.2014.02.074> <https://libkey.io/libraries/2592/openurl?genre=article&sid=OVID:medline&id=pmid:25168531&id=doi:10.1016%2Fj.urology.2014.02.074&issn=0090-4295&isbn=&volume=84&issue=3&spage=544&pages=544-8&date=2014&title=Urology&atitle=Type-2+diabetes+and+kidney+stones%3A+impact+of+diabetes+medications+and+glycemic+control.&aulast=Torricelli&pid=%3Cauthor%3ETorricelli+FC%3BDe+S%3BGebreselassie+S%3BLi+I%3BSarkissian+C%3BMonga+M%3C%2Fauthor%3E%3CAN%3E25168531%3C%2FAN%3E%3CDT%3EJournal+Article%3C%2FDT%3E>

119. **Can 24-hour urine stone risk profiles predict urinary stone composition?.**

**Item Type:** Journal Article

**Authors:** Torricelli, Fabio C. M.;De, Shubha;Liu, Xiaobo;Calle, Juan;Gebreselassie, Surafel and Monga, Manoj

**Publication Date:** Ju ,2014

**Journal:** Journal of Endourology 28(6), pp. 735–738

**Abstract: BACKGROUND AND PURPOSE**: Distinguishing calcium oxalate from uric acid stones is critical to identify those patients who may benefit from dissolution therapy and can also help direct preventive measures for stone growth. We aim to study whether 24-hour urine analysis may predict the urinary stone composition. **PATIENTS AND METHODS**: We retrospectively identified patients with calcium oxalate and uric acid stone compositions who also had a 24-hour urine collection within 3 months of stone analysis. Patients with calcium phosphate, cystine, and other stone compositions were excluded. Subjects were divided based on their stone type (calcium oxalate vs uric acid stones) and were compared according to demographic data and 24-hour urine analysis. Logistic regression analysis was performed to assess the association between stone composition and covariates. A nomogram was then constructed to predict uric acid stones over calcium oxalate stones. **RESULTS**: Of the 1163 patients identified, 1054 (90.6%) had calcium oxalate stones and 109 (9.4%) had uric acid stones. On logistic regression, body mass index (BMI) (odds ratio [OR] 1.351, 95% confidence interval [CI] 1.133-1.609; P: Of the 1163 patients identified, 1054 (90.6%) had calcium oxalate stones and 109 (9.4%) had uric acid stones. On logistic regression, body mass index (BMI) (odds ratio [OR] 1.351, 95% confidence interval [CI] 1.133-1.609; P**CONCLUSION**: Distinguishing uric acid from calcium oxalate stones can be performed with relative accuracy using parameters from the 24-hour urine stone risk profile and the patient's BMI and age.

**URL:** <https://ovidsp.ovid.com/ovidweb.cgi?T=JS&CSC=Y&NEWS=N&PAGE=fulltext&D=med11&DO=10.1089%2fend.2013.0769> <https://libkey.io/libraries/2592/openurl?genre=article&sid=OVID:medline&id=pmid:24460026&id=doi:10.1089%2Fend.2013.0769&issn=0892-7790&isbn=&volume=28&issue=6&spage=735&pages=735-8&date=2014&title=Journal+of+Endourology&atitle=Can+24-hour+urine+stone+risk+profiles+predict+urinary+stone+composition%3F.&aulast=Torricelli&pid=%3Cauthor%3ETorricelli+FC%3BDe+S%3BLiu+X%3BCalle+J%3BGebreselassie+S%3BMonga+M%3C%2Fauthor%3E%3CAN%3E24460026%3C%2FAN%3E%3CDT%3EJournal+Article%3C%2FDT%3E>

120. **Can obese stone formers follow dietary recommendations?.**

**Item Type:** Journal Article

**Authors:** Torricelli, Fabio Cesar Miranda;De, Shubha;Li, Ina;Sarkissian, Carl and Monga, Manoj

**Publication Date:** Fe ,2014

**Journal:** Journal of Endourology 28(2), pp. 248–251

**Abstract: INTRODUCTION**: Obese patients have an increased risk of kidney stones, and several studies have identified specific urinary derangements on 24-hour collections. The objective of this study was to assess obese and super-obese stone formers, and their compliance with dietary modifications over time, based on 24-hour urine outcomes. **PATIENTS AND METHODS**: A retrospective review was performed searching for all stone formers who completed a 24-hour urine collection before and after dietary counseling for stone prevention. Patients were excluded if placed on medical therapy in addition to dietary therapy. Patients were divided in three main groups according to their body mass index (BMI): 30, 30-40, and >40 kg/m(2). Demographic data and 24-hour urine stone risk parameters (volume, sodium, uric acid, citrate, and oxalate) were assessed. Initial 24-hour urine results were compared to follow-up results after dietary counseling. Then, the outcomes from each group were compared to each other. **RESULTS**: Two hundred and fourteen stone formers (67% male) were identified with a mean age of 49.5+/-15.0 years. One hundred twenty-eight (59.8%) patients had BMI 40 kg/m(2). Among patients with BMI 40 kg/m(2). Among patients with BMI 40 kg/m(2). Among patients with BMI 40 kg/m(2). Among patients with BMI 40 kg/m(2). Among patients with BMI 40 kg/m(2). Among patients with BMI **CONCLUSION**: Dietary recommendations can be an equally effective strategy in decreasing the risk or stone recurrence in obese and super-obese kidney stone formers as it is in those who are not obese.

**URL:** <https://ovidsp.ovid.com/ovidweb.cgi?T=JS&CSC=Y&NEWS=N&PAGE=fulltext&D=med11&DO=10.1089%2fend.2013.0536> <https://libkey.io/libraries/2592/openurl?genre=article&sid=OVID:medline&id=pmid:24032438&id=doi:10.1089%2Fend.2013.0536&issn=0892-7790&isbn=&volume=28&issue=2&spage=248&pages=248-51&date=2014&title=Journal+of+Endourology&atitle=Can+obese+stone+formers+follow+dietary+recommendations%3F.&aulast=Torricelli&pid=%3Cauthor%3ETorricelli+FC%3BDe+S%3BLi+I%3BSarkissian+C%3BMonga+M%3C%2Fauthor%3E%3CAN%3E24032438%3C%2FAN%3E%3CDT%3EJournal+Article%3C%2FDT%3E>

121. **Animal protein and the risk of kidney stones: a comparative metabolic study of animal protein sources.**

**Item Type:** Journal Article

**Authors:** Tracy, Chad R.;Best, Sara;Bagrodia, Aditya;Poindexter, John R.;Adams-Huet, Beverly;Sakhaee, Khashayar;Maalouf, Naim;Pak, Charles Y. C. and Pearle, Margaret S.

**Publication Date:** Ju ,2014

**Journal:** Journal of Urology 192(1), pp. 137–141

**Abstract: PURPOSE**: We compared the effect of 3 animal protein sources on urinary stone risk. **MATERIALS AND METHODS**: A total of 15 healthy subjects completed a 3-phase randomized, crossover metabolic study. During each 1-week phase subjects consumed a standard metabolic diet containing beef, chicken or fish. Serum chemistry and 24-hour urine samples collected at the end of each phase were compared using mixed model repeated measures analysis. **RESULTS**: Serum and urinary uric acid were increased for each phase. Beef was associated with lower serum uric acid than chicken or fish (6.5 vs 7.0 and 7.3 mg/dl, respectively, each p : Serum and urinary uric acid were increased for each phase. Beef was associated with lower serum uric acid than chicken or fish (6.5 vs 7.0 and 7.3 mg/dl, respectively, each p **CONCLUSIONS**: Consuming animal protein is associated with increased serum and urine uric acid in healthy individuals. The higher purine content of fish compared to beef or chicken is reflected in higher 24-hour urinary uric acid. However, as reflected in the saturation index, the stone forming propensity is marginally higher for beef compared to fish or chicken. Stone formers should be advised to limit the intake of all animal proteins, including fish. Copyright © 2014 American Urological Association Education and Research, Inc. Published by Elsevier Inc. All rights reserved.

**URL:** <https://ovidsp.ovid.com/ovidweb.cgi?T=JS&CSC=Y&NEWS=N&PAGE=fulltext&D=med11&DO=10.1016%2fj.juro.2014.01.093> <https://libkey.io/libraries/2592/openurl?genre=article&sid=OVID:medline&id=pmid:24518789&id=doi:10.1016%2Fj.juro.2014.01.093&issn=0022-5347&isbn=&volume=192&issue=1&spage=137&pages=137-41&date=2014&title=Journal+of+Urology&atitle=Animal+protein+and+the+risk+of+kidney+stones%3A+a+comparative+metabolic+study+of+animal+protein+sources.&aulast=Tracy&pid=%3Cauthor%3ETracy+CR%3BBest+S%3BBagrodia+A%3BPoindexter+JR%3BAdams-Huet+B%3BSakhaee+K%3BMaalouf+N%3BPak+CY%3BPearle+MS%3C%2Fauthor%3E%3CAN%3E24518789%3C%2FAN%3E%3CDT%3EClinical+Trial%2C+Phase+III%3C%2FDT%3E>

122. **Distinguishing characteristics of idiopathic calcium oxalate kidney stone formers with low amounts of Randall's plaque.**

**Item Type:** Journal Article

**Authors:** Wang, Xiangling;Krambeck, Amy E.;Williams, James C. Jr;Tang, Xiaojing;Rule, Andrew D.;Zhao, Fang;Bergstralh, Eric;Haskic, Zejfa;Edeh, Samuel;Holmes, David R. 3rd;Herrera Hernandez, Loren P. and Lieske, John C.

**Publication Date:** Oct 07 ,2014

**Journal:** Clinical Journal of the American Society of Nephrology: CJASN 9(10), pp. 1757–1763

**Abstract: BACKGROUND**: Overgrowth of calcium oxalate on Randall's plaque is a mechanism of stone formation among idiopathic calcium oxalate stone-formers (ICSFs). It is less clear how stones form when there is little or no plaque. **DESIGN, SETTING, PARTICIPANTS, & MEASUREMENTS**: Participants were a consecutive cohort of ICSFs who underwent percutaneous nephroscopic papillary mapping in the kidney or kidneys containing symptomatic stones and a papillary tip biopsy from a representative calyx during a stone removal procedure between 2009 and 2013. The distribution of Randall's plaque coverage was analyzed and used to divide ICSFs into those with a high (>=5%; mean, 10.5%; n=10) versus low (: Participants were a consecutive cohort of ICSFs who underwent percutaneous nephroscopic papillary mapping in the kidney or kidneys containing symptomatic stones and a papillary tip biopsy from a representative calyx during a stone removal procedure between 2009 and 2013. The distribution of Randall's plaque coverage was analyzed and used to divide ICSFs into those with a high (>=5%; mean, 10.5%; n=10) versus low (**RESULTS**: Low-plaque stone formers tended to be obese (50% versus 10%; P=0.03) and have a history of urinary tract infection (34% versus 0%; P=0.04). They were less likely to have multiple prior stone events (22% versus 80%; P=0.002) and had a lower mean 24-hour urine calcium excretion (187+/-86 mg versus 291+/-99 mg; P: Low-plaque stone formers tended to be obese (50% versus 10%; P=0.03) and have a history of urinary tract infection (34% versus 0%; P=0.04). They were less likely to have multiple prior stone events (22% versus 80%; P=0.002) and had a lower mean 24-hour urine calcium excretion (187+/-86 mg versus 291+/-99 mg; P**CONCLUSIONS**: These findings suggest that other pathways independent of Randall's plaque may contribute to stone pathogenesis among a subgroup of ICSFs who harbor low amounts of plaque. Copyright © 2014 by the American Society of Nephrology.

**URL:** <https://ovidsp.ovid.com/ovidweb.cgi?T=JS&CSC=Y&NEWS=N&PAGE=fulltext&D=med11&DO=10.2215%2fCJN.01490214> <https://libkey.io/libraries/2592/openurl?genre=article&sid=OVID:medline&id=pmid:25092598&id=doi:10.2215%2FCJN.01490214&issn=1555-9041&isbn=&volume=9&issue=10&spage=1757&pages=1757-63&date=2014&title=Clinical+Journal+of+The+American+Society+of+Nephrology%3A+CJASN&atitle=Distinguishing+characteristics+of+idiopathic+calcium+oxalate+kidney+stone+formers+with+low+amounts+of+Randall%27s+plaque.&aulast=Wang&pid=%3Cauthor%3EWang+X%3BKrambeck+AE%3BWilliams+JC+Jr%3BTang+X%3BRule+AD%3BZhao+F%3BBergstralh+E%3BHaskic+Z%3BEdeh+S%3BHolmes+DR+3rd%3BHerrera+Hernandez+LP%3BLieske+JC%3C%2Fauthor%3E%3CAN%3E25092598%3C%2FAN%3E%3CDT%3EComparative+Study%3C%2FDT%3E>

123. **Defining variation in urinary oxalate in hyperoxaluric stone formers.**

**Item Type:** Journal Article

**Authors:** Antonelli, Jodi A.;Langman, Craig B.;Odom, Christopher;Poindexter, John;Huet, Beverley and Pearle, Margaret S.

**Publication Date:** De ,2013

**Journal:** Journal of Endourology 27(12), pp. 1530–1534

**Abstract: BACKGROUND AND PURPOSE**: The development of effective preventive therapy for renal calculi in patients with secondary hyperoxaluria (2degreeHO) relies on establishing the pattern of normal variation in urinary oxalate (uOx) and attempting to reduce it. Therefore, we evaluated uOx at baseline and at subsequent time points in stone formers with 2degreeHO. **METHODS**: We reviewed the charts of 201 recurrent stone formers with 2degreeHO (uOx >= 40 mg/day). The 24-hour urine collections at baseline and after initiation of clinician-directed therapies were analyzed. Mixed models were constructed to analyze uOx over time for individual patients and as a group. Subgroup analyses were performed for enteric and idiopathic 2degreeHO. Coefficients of variation were computed using the root mean square error from linear models. **RESULTS**: The etiology of 2degreeHO was enteric in 17.9% and idiopathic in 82.1% of patients. Among the 943 urine collections analyzed, 196 oxalate values were derived from the enteric group and 747 from the idiopathic group. The median number of uOx values measured per person was four. The median 24-hour uOx (mg/day) was significantly higher for the enteric group than for the idiopathic group at the time of diagnosis: 64.4 (interquartile range [IQR]=48-90) vs 46.0 (IQR=38-56), P: The etiology of 2degreeHO was enteric in 17.9% and idiopathic in 82.1% of patients. Among the 943 urine collections analyzed, 196 oxalate values were derived from the enteric group and 747 from the idiopathic group. The median number of uOx values measured per person was four. The median 24-hour uOx (mg/day) was significantly higher for the enteric group than for the idiopathic group at the time of diagnosis: 64.4 (interquartile range [IQR]=48-90) vs 46.0 (IQR=38-56), P: The etiology of 2degreeHO was enteric in 17.9% and idiopathic in 82.1% of patients. Among the 943 urine collections analyzed, 196 oxalate values were derived from the enteric group and 747 from the idiopathic group. The median number of uOx values measured per person was four. The median 24-hour uOx (mg/day) was significantly higher for the enteric group than for the idiopathic group at the time of diagnosis: 64.4 (interquartile range [IQR]=48-90) vs 46.0 (IQR=38-56), P**CONCLUSIONS**: Among patients with 2degreeHO, uOx demonstrates significant random variation over time even with the incorporation of standard treatments, with enteric HO demonstrating higher values and greater variance than idiopathic HO.

**URL:** <https://ovidsp.ovid.com/ovidweb.cgi?T=JS&CSC=Y&NEWS=N&PAGE=fulltext&D=med10&DO=10.1089%2fend.2013.0199> <https://libkey.io/libraries/2592/openurl?genre=article&sid=OVID:medline&id=pmid:24147733&id=doi:10.1089%2Fend.2013.0199&issn=0892-7790&isbn=&volume=27&issue=12&spage=1530&pages=1530-4&date=2013&title=Journal+of+Endourology&atitle=Defining+variation+in+urinary+oxalate+in+hyperoxaluric+stone+formers.&aulast=Antonelli&pid=%3Cauthor%3EAntonelli+JA%3BLangman+CB%3BOdom+C%3BPoindexter+J%3BHuet+B%3BPearle+MS%3C%2Fauthor%3E%3CAN%3E24147733%3C%2FAN%3E%3CDT%3EJournal+Article%3C%2FDT%3E>

124. **Metabolic evaluation of first-time and recurrent stone formers**

**Item Type:** Journal Article

**Authors:** Goldfarb, David S. and Arowojolu, Omotayo

**Publication Date:** Fe ,2013

**Journal:** Urologic Clinics of North America 40(1), pp. 13–20

**Abstract:** Evaluation of stone formers should include careful attention to medications, past medical history, social history, family history, dietary evaluation, occupation, and laboratory evaluation. Laboratory evaluation requires at least serum chemistries and urinalysis. Twenty-four-hour urine collections are most appropriate for patients with recurrent stones or complex medical histories. However, these collections may be appropriate for some first-time stone formers, including those with comorbidities or large stones. Although twin studies demonstrate that heritability accounts for at least 50% of the kidney stone phenotype, the responsible genes are not clearly identified, and so genetic testing is rarely indicated. Copyright Published by Elsevier Inc.

**URL:** <https://ovidsp.ovid.com/ovidweb.cgi?T=JS&CSC=Y&NEWS=N&PAGE=fulltext&D=med10&DO=10.1016%2fj.ucl.2012.09.007> <https://libkey.io/libraries/2592/openurl?genre=article&sid=OVID:medline&id=pmid:23177631&id=doi:10.1016%2Fj.ucl.2012.09.007&issn=0094-0143&isbn=&volume=40&issue=1&spage=13&pages=13-20&date=2013&title=Urologic+Clinics+of+North+America&atitle=Metabolic+evaluation+of+first-time+and+recurrent+stone+formers.&aulast=Goldfarb&pid=%3Cauthor%3EGoldfarb+DS%3BArowojolu+O%3C%2Fauthor%3E%3CAN%3E23177631%3C%2FAN%3E%3CDT%3EJournal+Article%3C%2FDT%3E>

125. **Calcium and vitamin D supplementation and risk of kidney stone formation in postmenopausal women.**

**Item Type:** Journal Article

**Authors:** Haghighi, Anousheh;Samimagham, Hamidreza and Gohardehi, Golnar

**Publication Date:** May 21 ,2013

**Journal:** Iranian Journal of Kidney Diseases 7(3), pp. 210–213

**Abstract: INTRODUCTION**: Calcium and vitamin D are essential structural components of the skeletal system, which prevent osteoporosis after menopause. However, there is a controversial debate on the association between the intake of calcium and vitamin D supplements and the increased risk of formation of kidney calculi in postmenopausal women. which yet have to be confirmed. This study aimed to compare the metabolic changes after supplementation of calcium and vitamin D and examine the risk of stone formation. **MATERIALS AND METHODS**: Fifty-three postmenopausal women referred to rheumatology clinic who had no history of kidney calculi, bone diseases (apart from osteoporosis), metabolic, and rheumatic disorders and had not been receiving calcium, diuretics and calcitonin were investigated. Renal ultrasonography and blood tests were performed and the urine calcium levels were measured for a period of 24 hours for all patients. The examinations were repeated after a 1- year period of treatment with supplemental calcium (100 mg/d) and vitamin D (400 IU/d) and compared with the data before the treatment. **RESULTS**: After 1 year, asymptomatic lithiasis was confirmed in 1 of 53 patients (1.9%) using ultrasonographic examination. No significant differences were found between the 24-hour urine and blood calcium levels before and after the treatment. **CONCLUSIONS**: Our findings showed that oral intake of calcium and vitamin D after 1 year has no effect on the urinary calcium excretion rate and the formation of kidney calculi in postmenopausal women.

**URL:** <https://ovidsp.ovid.com/ovidweb.cgi?T=JS&CSC=Y&NEWS=N&PAGE=fulltext&D=med10&AN=23689153> <https://libkey.io/libraries/2592/openurl?genre=article&sid=OVID:medline&id=pmid:23689153&id=doi:&issn=1735-8582&isbn=&volume=7&issue=3&spage=210&pages=210-3&date=2013&title=Iranian+journal+of+Kidney+Diseases&atitle=Calcium+and+vitamin+D+supplementation+and+risk+of+kidney+stone+formation+in+postmenopausal+women.&aulast=Haghighi&pid=%3Cauthor%3EHaghighi+A%3BSamimagham+H%3BGohardehi+G%3C%2Fauthor%3E%3CAN%3E23689153%3C%2FAN%3E%3CDT%3EClinical+Trial%3C%2FDT%3E>

126. **Internet program for facilitating dietary modifications limiting kidney stone risk.**

**Item Type:** Journal Article

**Authors:** Lange, Jessica N.;Easter, Linda;Amoroso, Robert;Benfield, Debra;Mufarri, Patrick W.;Knight, John;Holmes, Ross P. and Assimos, Dean G.

**Publication Date:** Oc ,2013

**Journal:** Canadian Journal of Urology 20(5), pp. 6922–6926

**Abstract: INTRODUCTION**: Certain dietary modifications limit the risk of stone recurrence. Compliance is an important component of dietary therapy for stone prevention, and self-efficacy is an important ingredient of compliance. We developed an internet program to facilitate dietary compliance for stone prevention and performed a pilot study to assess its effectiveness. **MATERIALS AND METHODS**: The internet program provides information regarding dietary modifications including increased fluid consumption, limited animal protein, sodium, and oxalate intake, and adequate calcium consumption. Participants record their daily food and fluid intake and receive immediate feedback as to whether they were compliant or not. Five adult calcium stone formers collected three 24 hour urine specimens on self-selected diets, three 24 hour urine specimens while on a stone preventive metabolic diet, and three 24 hour urine specimens after utilizing the internet program for 1 month. Urinary stone risk parameters were measured, and data were analyzed using repeated measures ANOVA and Student's t test. **RESULTS**: All participants recorded their meals and snacks for each day and found the program easy to navigate. The mean time in hours from food consumption to log in was 35.25 +/- 70.8 hours. There were no statistically significant differences in stone risk factors between the controlled and internet dietary phases. Oxalate excretion was significantly higher during the self-selected dietary intake (p = 0.03). **CONCLUSIONS**: This pilot study demonstrates that subjects appear to be compliant with utilization of an interactive internet program for stone prevention with dietary modifications. In addition, improvement in certain stone risk parameters occurred.

**URL:** <https://ovidsp.ovid.com/ovidweb.cgi?T=JS&CSC=Y&NEWS=N&PAGE=fulltext&D=med10&AN=24128830> <https://libkey.io/libraries/2592/openurl?genre=article&sid=OVID:medline&id=pmid:24128830&id=doi:&issn=1195-9479&isbn=&volume=20&issue=5&spage=6922&pages=6922-6&date=2013&title=Canadian+Journal+of+Urology&atitle=Internet+program+for+facilitating+dietary+modifications+limiting+kidney+stone+risk.&aulast=Lange&pid=%3Cauthor%3ELange+JN%3BEaster+L%3BAmoroso+R%3BBenfield+D%3BMufarri+PW%3BKnight+J%3BHolmes+RP%3BAssimos+DG%3C%2Fauthor%3E%3CAN%3E24128830%3C%2FAN%3E%3CDT%3EJournal+Article%3C%2FDT%3E>

127. **Wu-Ling-San formula prophylaxis against recurrent calcium oxalate nephrolithiasis - a prospective randomized controlled trial.**

**Item Type:** Journal Article

**Authors:** Lin, Eugene;Ho, Lin;Lin, Mao-Sheng;Huang, Min-Ho and Chen, Wen-Chi

**Publication Date:** 2013

**Journal:** African Journal of Traditional, Complementary, & Alternative Medicines 10(5), pp. 199–209

**Abstract:** Wu-Ling-San (WLS) formula has been proved to prevent calcium oxalate nephrolithiasis both in vitro and in vivo. This is the first prospective, randomized and placebo-controlled clinical trial of WLS in calcium oxalate nephrolithiasis prevention. All patients who enrolled were asked to drink enough fluid to urinate at least 2 L daily during the study period. A 24-hour urine collection was performed to establish the baseline levels of multiple urinary parameters before taking the medicine. The patients were randomized and divided into two groups. The medication group took 2 gm WLS formula three times daily for 1 month. The control group took 2 gm placebo three times daily for 1 month. A 24-hour urine collection was performed to evaluate multiple urinary and serum parameters from all patients during the study period. A total of 39 patients were enrolled and 28 patients completed the study. Fourteen patients were allocated to WLS group and 14 patients to placebo group. After treatment, the mean urine output level increased to 2796.4 +/- 525.7 ml/day (percentage of change, 13.9 %) in the WLS formula group. With placebo therapy, the mean decreased slightly to 2521.4 +/- 762.7ml/day (percentage of change, -5.7 %). The percentage of change was significantly different between the two groups (independent t-test, P=0.02). No patient complained of side effects, such as fatigue, dizziness, musculoskeletal symptoms, or gastrointestinal disturbance. WLS formula is a promising adjunct to surgical and medical management of kidney stones. Active therapy with WLS formula has a positive effect on diuresis without leading to electrolyte imbalance.

**URL:** <https://ovidsp.ovid.com/ovidweb.cgi?T=JS&CSC=Y&NEWS=N&PAGE=fulltext&D=med10&DO=10.4314%2fajtcam.v10i5.1> <https://libkey.io/libraries/2592/openurl?genre=article&sid=OVID:medline&id=pmid:24311828&id=doi:10.4314%2Fajtcam.v10i5.1&issn=2505-0044&isbn=&volume=10&issue=5&spage=199&pages=199-209&date=2013&title=African+journal+of+traditional%2C+complementary%2C+%26+alternative+medicines&atitle=Wu-Ling-San+formula+prophylaxis+against+recurrent+calcium+oxalate+nephrolithiasis+-+a+prospective+randomized+controlled+trial.&aulast=Lin&pid=%3Cauthor%3ELin+E%3BHo+L%3BLin+MS%3BHuang+MH%3BChen+WC%3C%2Fauthor%3E%3CAN%3E24311828%3C%2FAN%3E%3CDT%3EJournal+Article%3C%2FDT%3E>

128. **Changes in urine parameters after desert exposure: assessment of stone risk in United States Marines transiently exposed to a desert environment.**

**Item Type:** Journal Article

**Authors:** Masterson, James H.;Jourdain, Victor J.;Collard, Daniel A.;Choe, Chong H.;Christman, Matthew S.;L'Esperance, James O. and Auge, Brian K.

**Publication Date:** Ja ,2013

**Journal:** Journal of Urology 189(1), pp. 165–170

**Abstract: PURPOSE**: Living in a desert environment has been associated with a higher incidence of kidney stone formation, likely because of concentrated urine output, higher production of vitamin D and genetic predisposition. We determined the changes in urinary parameters after a group of United States Marines temporarily transitioned from a temperate environment to a desert environment. **MATERIALS AND METHODS**: A total of 50 Marines completed a questionnaire and performed 3, 24-hour urine collections before mobilization to the desert, after 30 days in the desert and 2 weeks after returning from the desert. **RESULTS**: Daily urine output decreased 68% to 0.52 L despite marked increased fluid intake (17 L per day). Total daily urinary excretion of calcium, uric acid, sodium, magnesium and potassium in the desert decreased by 70%, 41%, 53%, 22% and 36%, respectively. Urinary pH decreased from 6.1 to 5.6 while in the desert, and citrate and oxalate had minimal changes. After their return from the desert, apart from a decrease of 22% in oxalate, there were no statistically significant differences from baseline. While in the desert, relative supersaturation risks of uric acid and sodium urate were increased 153% and 56%, respectively. Brushite relative supersaturation decreased 24%. After their return there was no statistical difference from baseline. **CONCLUSIONS**: Our findings suggest that the kidneys preserved water and electrolytes while the Marines were subjected to the desert environment. Despite this conservation, relative saturations indicate increased risk of stones in healthy men exposed to a desert environment with rapid resolution upon return. Copyright © 2013 American Urological Association Education and Research, Inc. Published by Elsevier Inc. All rights reserved.

**URL:** <https://ovidsp.ovid.com/ovidweb.cgi?T=JS&CSC=Y&NEWS=N&PAGE=fulltext&D=med10&DO=10.1016%2fj.juro.2012.08.097> <https://libkey.io/libraries/2592/openurl?genre=article&sid=OVID:medline&id=pmid:23174258&id=doi:10.1016%2Fj.juro.2012.08.097&issn=0022-5347&isbn=&volume=189&issue=1&spage=165&pages=165-70&date=2013&title=Journal+of+Urology&atitle=Changes+in+urine+parameters+after+desert+exposure%3A+assessment+of+stone+risk+in+United+States+Marines+transiently+exposed+to+a+desert+environment.&aulast=Masterson&pid=%3Cauthor%3EMasterson+JH%3BJourdain+VJ%3BCollard+DA%3BChoe+CH%3BChristman+MS%3BL%27Esperance+JO%3BAuge+BK%3C%2Fauthor%3E%3CAN%3E23174258%3C%2FAN%3E%3CDT%3EJournal+Article%3C%2FDT%3E>

129. **Using 24-hour urinalysis to predict stone type.**

**Item Type:** Journal Article

**Authors:** Moreira, Daniel M.;Friedlander, Justin I.;Hartman, Christopher;Elsamra, Sammy E.;Smith, Arthur D. and Okeke, Zeph

**Publication Date:** De ,2013

**Journal:** Journal of Urology 190(6), pp. 2106–2111

**Abstract: PURPOSE**: We determined the accuracy of 24-hour urinalysis in predicting stone type and identify the associations between 24-hour urine elements with stone type. **MATERIALS AND METHODS**: We performed a retrospective review of 503 stone formers with stone composition analysis and 24-hour urinalysis available. Analysis of 24-hour urine elements across stone types was performed using Fisher's exact test and ANOVA. Multinomial logistic regression was used to predict stone type based on 24-hour urinalysis. **RESULTS**: A total of 280 (56%) patients had predominantly calcium oxalate, 103 (20%) had uric acid, 93 (19%) had calcium phosphate, 16 (3%) had mixed and 11 (2%) had other stone types. There were several significant patient characteristics and 24-hour urinalysis differences across stone type groups. The statistical model predicted 371 (74%) calcium oxalate, 78 (16%) uric acid, 52 (10%) calcium phosphate, zero mixed and 2 (less than 1%) other stone types. The model correctly predicted calcium oxalate stones in 85%, uric acid in 51%, calcium phosphate in 31%, mixed in 0% and other stone types in 18% of the cases. Of the predicted stone types, correct predictions were 61%, 69%, 56% and 71% for calcium oxalate, uric acid, calcium phosphate and other stones types, respectively. The overall accuracy was 64%. Plots were used to explore the associations between each 24-hour urine element with each predicted stone type adjusted for all the others urinary elements. **CONCLUSIONS**: A 24-hour urinalysis alone does not accurately predict stone type. However, it may be used in conjunction with other variables to predict stone composition. Copyright © 2013 American Urological Association Education and Research, Inc. Published by Elsevier Inc. All rights reserved.

**URL:** <https://ovidsp.ovid.com/ovidweb.cgi?T=JS&CSC=Y&NEWS=N&PAGE=fulltext&D=med10&DO=10.1016%2fj.juro.2013.05.115> <https://libkey.io/libraries/2592/openurl?genre=article&sid=OVID:medline&id=pmid:23764079&id=doi:10.1016%2Fj.juro.2013.05.115&issn=0022-5347&isbn=&volume=190&issue=6&spage=2106&pages=2106-11&date=2013&title=Journal+of+Urology&atitle=Using+24-hour+urinalysis+to+predict+stone+type.&aulast=Moreira&pid=%3Cauthor%3EMoreira+DM%3BFriedlander+JI%3BHartman+C%3BElsamra+SE%3BSmith+AD%3BOkeke+Z%3C%2Fauthor%3E%3CAN%3E23764079%3C%2FAN%3E%3CDT%3EJournal+Article%3C%2FDT%3E>

130. **The efficacy of medical prophylaxis in children with calcium oxalate urolithiasis after percutaneous nephrolithotomy.**

**Item Type:** Journal Article

**Authors:** Oguz, Ural and Unsal, Ali

**Publication Date:** Ja ,2013

**Journal:** Journal of Endourology 27(1), pp. 92–95

**Abstract: OBJECTIVE**: To evaluate whether medical prophylaxis decreases calcium oxalate stone recurrences in children after percutaneous nephrolithotomy (PNL) or not. To our knowledge this is the first study that evaluates this topic in children after PNL. **PATIENTS AND METHODS**: We researched analysis of 42 children with calcium oxalate stone disease. They were divided into two groups. Twenty-two children who had a follow-up with medical prophylaxis after PNL were included in group I; 20 children who did not have medical prophylaxis were included in group II. They were all stone free and they were evaluated with 24-hour urine analysis and blood samples, abdominal X-ray, and ultrasonography. The average follow-up was 25.9 (12-42) months for group I and 24.6 (14-40) months for group II. **RESULTS**: Age, gender, follow-up time, and 24-hour urine samples were similar between groups (p>0.05). New stone formation was detected in two (9.1%) and seven (35%) patients in group I and II, respectively. New stone formation seemed to be higher in group II, but because of the less number of patients, this difference was not statistically significant (p=0.062). The number of patients with recurrence was statistically higher in group II (p=0.032). Stone formation rate (SFR) was calculated as 0.034 and 0.2 per patient per year for group I and II, respectively. This difference of SFR between groups was statistically significant (p=0.028). **CONCLUSION**: Medical prophylaxis of children after PNL can reduce new stone formation and SFR. Although it was not statistically significant because of low patient numbers, higher recurrence rate was detected in patients who did not have medical prophylaxis.

**URL:** <https://ovidsp.ovid.com/ovidweb.cgi?T=JS&CSC=Y&NEWS=N&PAGE=fulltext&D=med10&DO=10.1089%2fend.2012.0243> <https://libkey.io/libraries/2592/openurl?genre=article&sid=OVID:medline&id=pmid:22891769&id=doi:10.1089%2Fend.2012.0243&issn=0892-7790&isbn=&volume=27&issue=1&spage=92&pages=92-5&date=2013&title=Journal+of+Endourology&atitle=The+efficacy+of+medical+prophylaxis+in+children+with+calcium+oxalate+urolithiasis+after+percutaneous+nephrolithotomy.&aulast=Oguz&pid=%3Cauthor%3EOguz+U%3BUnsal+A%3C%2Fauthor%3E%3CAN%3E22891769%3C%2FAN%3E%3CDT%3EComparative+Study%3C%2FDT%3E>

131. **A pilot study of the effect of sodium thiosulphate on urinary lithogenicity and associated metabolic acid load in non-stone formers and stone formers with hypercalciuria.**

**Item Type:** Journal Article

**Authors:** Okonkwo, Onyeka W.;Batwara, Ruchika;Granja, Ignacio;Asplin, John R. and Goldfarb, David S.

**Publication Date:** 2013

**Journal:** PLoS ONE [Electronic Resource] 8(4), pp. e60380

**Abstract: BACKGROUND AND OBJECTIVES**: Sodium thiosulphate (STS) reduced calcium stone formation in both humans and genetic hypercalciuric stone forming (GHS) rats. We sought to measure urine chemistry changes resulting from STS administration in people. DESIGN SETTING PARTICIPANTS MEASUREMENTS: STS was given to healthy and hypercalciuric stone forming adults. Five normal non-stone forming adults (mean age 33 years), and 5 people with idiopathic hypercalciuria and calcium kidney stones (mean age 66 years) participated. Two baseline 24-hour urine collections were performed on days 2 and 3 of 3 days of self-selected diets. Subjects then drank STS 10 mmol twice a day for 7 days and did urine collections while repeating the self-selected diet. Results were compared by non-parametric Wilcoxon signed rank test. The primary outcome was the resulting change in urine chemistry. **RESULTS**: STS administration did not cause a significant change in urinary calcium excretion in either group. In both groups, 24 hour urinary ammonium (P = 0.005) and sulphate excretion (P = 0.007) increased, and urinary pH fell (P = 0.005); citrate excretion fell (P: STS administration did not cause a significant change in urinary calcium excretion in either group. In both groups, 24 hour urinary ammonium (P = 0.005) and sulphate excretion (P = 0.007) increased, and urinary pH fell (P = 0.005); citrate excretion fell (P**CONCLUSIONS**: The basis for studies demonstrating that STS prevented stones in rats and people was not reflected by the changes in urine chemistry reported here. Although serum HCO3 did not change, urine tests suggested an acid load in both non-stone forming and hypercalciuric stone-forming participants. The long term safety of STS needs to be determined before the drug can be tested in humans for long-term prevention of stone recurrence.

**URL:** <https://ovidsp.ovid.com/ovidweb.cgi?T=JS&CSC=Y&NEWS=N&PAGE=fulltext&D=med10&DO=10.1371%2fjournal.pone.0060380> [https://libkey.io/libraries/2592/openurl?genre=article&sid=OVID:medline&id=pmid:23593205&id=doi:10.1371%2Fjournal.pone.0060380&issn=1932-6203&isbn=&volume=8&issue=4&spage=e60380&pages=e60380&date=2013&title=PLoS+ONE+%5BElectronic+Resource%5D&atitle=A+pilot+study+of+the+effect+of+sodium+thiosulphate+on+urinary+lithogenicity+and+associated+metabolic+acid+load+in+non-stone+formers+and+stone+formers+with+hypercalciuria.&aulast=Okonkwo&pid=%3Cauthor%3EOkonkwo+OW%3BBatwara+R%3BGranja+I%3BAsplin+JR%3BGoldfarb+DS%3C%2Fauthor%3E%3CAN%3E23593205%3C%2FAN%3E%3CDT%3EClinical+Trial%3C%2FDT%3E](https://libkey.io/libraries/2592/openurl?genre=article&sid=OVID:medline&id=pmid:23593205&id=doi:10.1371%2Fjournal.pone.0060380&issn=1932-6203&isbn=&volume=8&issue=4&spage=e60380&pages=e60380&date=2013&title=PLoS+ONE+%5BElectronic+Resource%5D&atitle=A+pilot+study+of+the+effect+of+sodium+thiosulfate+on+urinary+lithogenicity+and+associated+metabolic+acid+load+in+non-stone+formers+and+stone+formers+with+hypercalciuria.&aulast=Okonkwo&pid=%3Cauthor%3EOkonkwo+OW%3BBatwara+R%3BGranja+I%3BAsplin+JR%3BGoldfarb+DS%3C%2Fauthor%3E%3CAN%3E23593205%3C%2FAN%3E%3CDT%3EClinical+Trial%3C%2FDT%3E)

132. **The effect of sodium bicarbonate upon urinary citrate excretion in calcium stone formers.**

**Item Type:** Journal Article

**Authors:** Pinheiro, Vivian Barbosa;Baxmann, Alessandra Calabria;Tiselius, Hans-Goran and Heilberg, Ita Pfeferman

**Publication Date:** Ju ,2013

**Journal:** Urology 82(1), pp. 33–37

**Abstract: OBJECTIVE**: To evaluate the effects of oral sodium bicarbonate (NaBic) supplementation upon urinary citrate excretion in calcium stone formers (CSFs). **METHODS**: Sixteen adult calcium stone formers with hypocitraturia were enrolled in a randomized, double-blind, crossover protocol using 60 mEq/day of NaBic during 3 days compared to the same period and doses of potassium citrate (KCit) supplementation. Blood and 24-hour urine samples were collected at baseline and during the third day of each alkali salt. **RESULTS**: NaBic, similarly to KCit supplementation, led to an equivalent and significant increase in urinary citrate and pH. Compared to baseline, NaBic led to a significant increase in sodium excretion without concomitant increases in urinary calcium excretion, whereas KCit induced a significant increase in potassium excretion coupled with a significant reduction in urinary calcium. Although NaBic and KCit both reduced calcium oxalate supersaturation (CaOxSS) significantly vs baseline, KCit reduced calcium oxalate supersaturation significantly further vs NaBic. Both KCit and NaBic significantly reduced urinary phosphate and increased calcium phosphate supersaturation (CaPSS) compared to baseline. Finally, a significantly higher sodium urate supersaturation (NaUrSS) was observed after the use of the 2 drugs. **CONCLUSION**: This short-term study suggests that NaBic represents an effective alternative for the treatment of hypocitraturic calcium oxalate stone formers who cannot tolerate or afford the cost of KCit. In view of the increased sodium urate supersaturation, patients with pure uric acid stones and high urate excretion may be less suited for treatment with NaBic. Copyright © 2013 Elsevier Inc. All rights reserved.

**URL:** <https://ovidsp.ovid.com/ovidweb.cgi?T=JS&CSC=Y&NEWS=N&PAGE=fulltext&D=med10&DO=10.1016%2fj.urology.2013.03.002> <https://libkey.io/libraries/2592/openurl?genre=article&sid=OVID:medline&id=pmid:23602798&id=doi:10.1016%2Fj.urology.2013.03.002&issn=0090-4295&isbn=&volume=82&issue=1&spage=33&pages=33-7&date=2013&title=Urology&atitle=The+effect+of+sodium+bicarbonate+upon+urinary+citrate+excretion+in+calcium+stone+formers.&aulast=Pinheiro&pid=%3Cauthor%3EPinheiro+VB%3BBaxmann+AC%3BTiselius+HG%3BHeilberg+IP%3C%2Fauthor%3E%3CAN%3E23602798%3C%2FAN%3E%3CDT%3EJournal+Article%3C%2FDT%3E>

133. **Stone disease in living-related renal donors: long-term outcomes for transplant donors and recipients.**

**Item Type:** Journal Article

**Authors:** Rizkala, Emad;Coleman, Sarah;Tran, Christine;Isac, Wahib;Flechner, Stuart M.;Goldfarb, David and Monga, Manoj

**Publication Date:** De ,2013

**Journal:** Journal of Endourology 27(12), pp. 1520–1524

**Abstract: BACKGROUND AND PURPOSE**: Historically, patients wishing to donate their kidney to living related recipients were deemed ineligible if preoperative imaging demonstrated nephrolithiasis. We assess the outcomes of donors with nephrolithiasis and the outcomes of their recipients. **METHODS**: Donors undergoing nephrectomy between 2001 and 2011 who had nephrolithiasis on preoperative computed tomography (CT) imaging or a history of stone passage were identified. A retrospective chart review documented donor and recipient demographics, donor 24-hour urine collections, stone size and location, stone events after transplant, and graft function. A seven-question telephone survey regarding development and/or presence of symptomatic nephrolithiasis was conducted. **RESULTS**: Fifty-four donor-recipient pairs met the inclusion criteria. Twenty-eight (51.9%) patients had valid preoperative 24-hour urine collection, seven (25%) of whom had hypercalciuria. Seven (13%) patients had previous symptomatic nephrolithiasis, but no stones on imaging. Forty-one patients donated a kidney with at least one stone, with a mean stone size of 2.4 mm (range 1-6 mm). Median follow-up for donors and recipients was 22.5 months (interquartile range [IQR] 1-79.3) and 47.4 months (IQR 25.1-76.1), with 50% and 77.7% having a follow-up of more than 2 years, respectively. One donor with nephrolithiasis on preoperative imaging who donated the contralateral kidney passed a stone spontaneously after visiting the emergency department. Otherwise, no other donors or recipients experienced any stone episodes during the follow-up period. **CONCLUSION**: The risk of clinical stone recurrence in donors and recipients is low: As such, presence of small caliceal stones should not constitute an exclusion for living-related kidney donation.

**URL:** <https://ovidsp.ovid.com/ovidweb.cgi?T=JS&CSC=Y&NEWS=N&PAGE=fulltext&D=med10&DO=10.1089%2fend.2013.0203> <https://libkey.io/libraries/2592/openurl?genre=article&sid=OVID:medline&id=pmid:24261656&id=doi:10.1089%2Fend.2013.0203&issn=0892-7790&isbn=&volume=27&issue=12&spage=1520&pages=1520-4&date=2013&title=Journal+of+Endourology&atitle=Stone+disease+in+living-related+renal+donors%3A+long-term+outcomes+for+transplant+donors+and+recipients.&aulast=Rizkala&pid=%3Cauthor%3ERizkala+E%3BColeman+S%3BTran+C%3BIsac+W%3BFlechner+SM%3BGoldfarb+D%3BMonga+M%3C%2Fauthor%3E%3CAN%3E24261656%3C%2FAN%3E%3CDT%3EJournal+Article%3C%2FDT%3E>

134. **Adequate or not? A comparison of 24-hour urine studies for renal stone prevention by creatinine to weight ratio.**

**Item Type:** Journal Article

**Authors:** Sawyer, Mark D.;Dietrich, Mary S.;Pickens, Ryan B.;Herrell, S. Duke and Miller, Nicole L.

**Publication Date:** Ma ,2013

**Journal:** Journal of Endourology 27(3), pp. 366–369

**Abstract: PURPOSE**: The intent of this study was to compare urine parameters between kidney stone formers with low, expected, and high creatinine to weight ratio (Cr/kg) and to discuss implications for treatment. **PATIENTS AND METHODS**: In this retrospective study, 381 stone formers (205 females and 176 males) with urine collections for nephrolithiasis evaluation were included. Using the first collected sample, patients were grouped into low, expected, and high Cr/kg using sex appropriate reference ranges (15.0-20.0 mg/kg for females and 18.0-24.0 mg/kg for males) and evaluated. **RESULTS**: Of initial collections, 50.7% were outside the Cr/kg reference range. Median age increased as Cr/kg decreased with 13 years age difference between low (56) and high (43) groups. Body mass index and weight also increased significantly with decreased ratio. No significant difference between groups was found for urine volume, total oxalate, oxalate concentration, pH, and supersaturations of calcium oxalate and uric acid. Total calcium, sodium, citrate, calcium concentration, supersaturation of calcium phosphate, and other parameters significantly increased with increased Cr/kg for the cohort. **CONCLUSIONS**: To our knowledge, this is the first study to thoroughly evaluate the impact of Cr/kg on urine parameters. Clinicians should be cognizant of limitations of Cr/kg for evaluating collection adequacy. It is especially important to consider Cr/kg when there is potential for undertreatment or overtreatment of patients with abnormal Cr/kg. Studies demonstrating hypercalciuria and high Cr/kg may need to be repeated before starting thiazide diuretics. Further studies are needed to determine how to better interpret urine components that vary with Cr/kg ratio.

**URL:** <https://ovidsp.ovid.com/ovidweb.cgi?T=JS&CSC=Y&NEWS=N&PAGE=fulltext&D=med10&DO=10.1089%2fend.2012.0203> <https://libkey.io/libraries/2592/openurl?genre=article&sid=OVID:medline&id=pmid:22967263&id=doi:10.1089%2Fend.2012.0203&issn=0892-7790&isbn=&volume=27&issue=3&spage=366&pages=366-9&date=2013&title=Journal+of+Endourology&atitle=Adequate+or+not%3F+A+comparison+of+24-hour+urine+studies+for+renal+stone+prevention+by+creatinine+to+weight+ratio.&aulast=Sawyer&pid=%3Cauthor%3ESawyer+MD%3BDietrich+MS%3BPickens+RB%3BHerrell+SD%3BMiller+NL%3C%2Fauthor%3E%3CAN%3E22967263%3C%2FAN%3E%3CDT%3EComparative+Study%3C%2FDT%3E>

135. **Dietary management of idiopathic hyperoxaluria and the influence of patient characteristics and compliance.**

**Item Type:** Journal Article

**Authors:** Schwen, Zeyad R.;Riley, Julie M.;Shilo, Yaniv and Averch, Timothy D.

**Publication Date:** De ,2013

**Journal:** Urology 82(6), pp. 1220–1225

**Abstract: OBJECTIVE**: To assess the efficacy of dietary management for the treatment of idiopathic hyperoxaluria in a large tertiary care center and examine the influence of patient factors, compliance, and follow-up on oxalate reduction, which has not been previously investigated. **METHODS**: Retrospectively, 149 patients with kidney stones with idiopathic hyperoxaluria who received dietary management at our stone clinic were evaluated. Changes in urinary parameters on 24-hour urine collections were calculated for all patients and those with abnormal values in the overall short-term (30-240 days) and long-term (>240 days) time periods. Changes in urinary oxalate were evaluated with respect to patient characteristics and compliance measures. **RESULTS**: Urine oxalate and supersaturation of calcium oxalate were significantly (P : Urine oxalate and supersaturation of calcium oxalate were significantly (P **CONCLUSION**: This study confirms that meaningful reductions of urine oxalate and supersaturation of calcium oxalate can be achieved with dietary management of hyperoxaluria on a larger clinical scale. Furthermore, we identified that women and patients with low BMIs had greater urine oxalate reductions and urine volume may also be used by clinicians as a measure of dietary compliance. Copyright © 2013 Elsevier Inc. All rights reserved.

**URL:** <https://ovidsp.ovid.com/ovidweb.cgi?T=JS&CSC=Y&NEWS=N&PAGE=fulltext&D=med10&DO=10.1016%2fj.urology.2013.08.002> <https://libkey.io/libraries/2592/openurl?genre=article&sid=OVID:medline&id=pmid:24054440&id=doi:10.1016%2Fj.urology.2013.08.002&issn=0090-4295&isbn=&volume=82&issue=6&spage=1220&pages=1220-5&date=2013&title=Urology&atitle=Dietary+management+of+idiopathic+hyperoxaluria+and+the+influence+of+patient+characteristics+and+compliance.&aulast=Schwen&pid=%3Cauthor%3ESchwen+ZR%3BRiley+JM%3BShilo+Y%3BAverch+TD%3C%2Fauthor%3E%3CAN%3E24054440%3C%2FAN%3E%3CDT%3EJournal+Article%3C%2FDT%3E>

136. **Frequency of rare allelic variation in candidate genes among individuals with low and high urinary calcium excretion.**

**Item Type:** Journal Article

**Authors:** Toka, Hakan R.;Genovese, Giulio;Mount, David B.;Pollak, Martin R. and Curhan, Gary C.

**Publication Date:** 2013

**Journal:** PLoS ONE [Electronic Resource] 8(8), pp. e71885

**Abstract:** Our study investigated the association of rare allelic variants with extremes of 24-hour urinary calcium excretion because higher urinary calcium excretion is a dominant risk factor for calcium-based kidney stone formation. We resequenced 40 candidate genes potentially related to urinary calcium excretion in individuals from the Nurses' Health Studies I & II and the Health Professionals Follow-up Study. A total of 960 participants were selected based on availability of 24-hour urine collection data and level of urinary calcium excretion (low vs. high). We utilized DNA sample pooling, droplet-based target gene enrichment, multiplexing, and high-throughput sequencing. Approximately 64% of samples (n = 615) showed both successful target enrichment and sequencing data with >20-fold deep coverage. A total of 259 novel allelic variants were identified. None of the rare gene variants (allele frequencies = 2% suggested an association of the Claudin14 SNP rs113831133 with lower urinary calcium excretion (6/520 versus 29/710 haplotypes, P value = 0.003). Our data, together with previous human and animal studies, suggest a possible role for Claudin14 in urinary calcium excretion. Genetic validation studies in larger sample sets will be necessary to confirm our findings for rs113831133. In the tested set of candidate genes, rare allelic variants do not appear to contribute significantly to differences in urinary calcium excretion between individuals.

**URL:** <https://ovidsp.ovid.com/ovidweb.cgi?T=JS&CSC=Y&NEWS=N&PAGE=fulltext&D=med10&DO=10.1371%2fjournal.pone.0071885> <https://libkey.io/libraries/2592/openurl?genre=article&sid=OVID:medline&id=pmid:23991001&id=doi:10.1371%2Fjournal.pone.0071885&issn=1932-6203&isbn=&volume=8&issue=8&spage=e71885&pages=e71885&date=2013&title=PLoS+ONE+%5BElectronic+Resource%5D&atitle=Frequency+of+rare+allelic+variation+in+candidate+genes+among+individuals+with+low+and+high+urinary+calcium+excretion.&aulast=Toka&pid=%3Cauthor%3EToka+HR%3BGenovese+G%3BMount+DB%3BPollak+MR%3BCurhan+GC%3C%2Fauthor%3E%3CAN%3E23991001%3C%2FAN%3E%3CDT%3EJournal+Article%3C%2FDT%3E>

137. **Changing stone composition profile of children with nephrolithiasis.**

**Item Type:** Journal Article

**Authors:** Wood, Kyle D.;Stanasel, Irina S.;Koslov, David S.;Mufarrij, Patrick W.;McLorie, Gordon A. and Assimos, Dean G.

**Publication Date:** Ju ,2013

**Journal:** Urology 82(1), pp. 210–213

**Abstract: OBJECTIVE**: To determine if this trend toward calcium phosphate stone formation exists in children. **METHODS**: This is a retrospective study of medical records of 179 children managed at our medical center from 1992-2010 for whom stone analysis and other pertinent laboratory data were available. A comparison of patients managed from 1992-2000 (P1) and 2001-2010 (P2) was undertaken. Statistical analysis included nonparametric tests. **RESULTS**: There were no significant differences in the mean age of the 2 cohorts. During both periods, boys comprised a significantly higher proportion during the first decade of life, whereas girls comprised a significantly higher proportion during the second decade. A higher percentage of patients had calcium oxalate (CaOx) stones in P1 compared to P2 (60% vs 47%, P = .0019). There was a significant increase in the percentage of patients having calcium phosphate stones in P2 compared to P1 (27% vs 18.5%, P = .008). Twenty-seven patients had recurrent stones. A comparison of the compositions of the first and last stones of patients within this group demonstrated an increasing proportion of brushite stones (3.7% vs 11.1%, P = .04). Twenty-four hour urine testing results were similar for those with CaOx and calcium phosphate stones. **CONCLUSION**: An increasing proportion of children have calcium phosphate calculi. Brushite stones are more prevalent in children with recurrent stone events. The impetus of these shifts is not readily apparent. Copyright © 2013 Elsevier Inc. All rights reserved.

**URL:** <https://ovidsp.ovid.com/ovidweb.cgi?T=JS&CSC=Y&NEWS=N&PAGE=fulltext&D=med10&DO=10.1016%2fj.urology.2013.02.033> <https://libkey.io/libraries/2592/openurl?genre=article&sid=OVID:medline&id=pmid:23561713&id=doi:10.1016%2Fj.urology.2013.02.033&issn=0090-4295&isbn=&volume=82&issue=1&spage=210&pages=210-3&date=2013&title=Urology&atitle=Changing+stone+composition+profile+of+children+with+nephrolithiasis.&aulast=Wood&pid=%3Cauthor%3EWood+KD%3BStanasel+IS%3BKoslov+DS%3BMufarrij+PW%3BMcLorie+GA%3BAssimos+DG%3C%2Fauthor%3E%3CAN%3E23561713%3C%2FAN%3E%3CDT%3EJournal+Article%3C%2FDT%3E>

138. **Hyperinsulinemia and urinary calcium excretion in calcium stone formers with idiopathic hypercalciuria.**

**Item Type:** Journal Article

**Authors:** Yoon, Vivienne;Adams-Huet, Beverley;Sakhaee, Khashayar and Maalouf, Naim M.

**Publication Date:** Ju ,2013

**Journal:** Journal of Clinical Endocrinology & Metabolism 98(6), pp. 2589–2594

**Abstract: CONTEXT**: Calcium stone formers with idiopathic hypercalciuria (IH) are known to exhibit an exaggerated postprandial rise in urine calcium excretion compared with non-stone-forming individuals, and insulin has been proposed to mediate this difference. **OBJECTIVE**: Our objective was to investigate the impact of hyperinsulinemia on urine calcium excretion in IH compared with non-stone-forming controls. **PARTICIPANTS AND SETTING**: Ten IH patients and 22 control non-stone-forming subjects (8 lean and 14 overweight and obese) participated at the University of Texas Southwestern Clinical and Translational Research Center. **DESIGN**: After stabilization on a fixed metabolic diet, subjects underwent a hyperinsulinemic-euglycemic clamp. Fasting 2-hour urine specimens were collected before and during the clamp. **MAIN OUTCOME MEASURES**: Changes in fractional calcium excretion (F(E)Ca) during the clamp were compared between the 3 groups of subjects (IH, overweight/obese controls, and lean controls). Insulin sensitivity was measured by glucose disposal rate. **RESULTS**: IH had significantly higher 24-hour urine calcium excretion than controls, and exhibited similar age, body mass index, and insulin sensitivity as overweight/obese controls. The hyperinsulinemic-euglycemic clamp resulted in a significant increase in serum insulin with no significant changes in serum calcium and glucose. This was accompanied by a small increase in F(E)Ca, with no significant differences between the 3 groups. There was no correlation between insulin sensitivity and 24-hour urine calcium or the change in F(E)Ca during the hyperinsulinemic clamp. **CONCLUSIONS**: The rise in urine calcium associated with euglycemic hyperinsulinemia was small and not statistically different between IH and non-stone-forming controls. Insulin is therefore unlikely to play a significant pathogenetic role in IH.

**URL:** <https://ovidsp.ovid.com/ovidweb.cgi?T=JS&CSC=Y&NEWS=N&PAGE=fulltext&D=med10&DO=10.1210%2fjc.2013-1301> <https://libkey.io/libraries/2592/openurl?genre=article&sid=OVID:medline&id=pmid:23553859&id=doi:10.1210%2Fjc.2013-1301&issn=0021-972X&isbn=&volume=98&issue=6&spage=2589&pages=2589-94&date=2013&title=Journal+of+Clinical+Endocrinology+%26+Metabolism&atitle=Hyperinsulinemia+and+urinary+calcium+excretion+in+calcium+stone+formers+with+idiopathic+hypercalciuria.&aulast=Yoon&pid=%3Cauthor%3EYoon+V%3BAdams-Huet+B%3BSakhaee+K%3BMaalouf+NM%3C%2Fauthor%3E%3CAN%3E23553859%3C%2FAN%3E%3CDT%3EJournal+Article%3C%2FDT%3E>

139. **Biochemical determinants of severe lithogenic activity in patients with idiopathic calcium nephrolithiasis.**

**Item Type:** Journal Article

**Authors:** Arrabal-Polo, Miguel Angel;Arrabal-Martin, Miguel;de Haro-Munoz, Tomas;Poyatos-Andujar, Antonio;Palaeo-Yago, Francisco and Zuluaga-Gomez, Armando

**Publication Date:** Ja ,2012

**Journal:** Urology 79(1), pp. 48–54

**Abstract: OBJECTIVE**: To analyze the biochemical alterations in plasma and the urine determinants of severe lithogenic activity in patients with idiopathic calcium nephrolithiasis. **METHODS**: We performed a cross-sectional study of 120 patients divided into 2 groups: group 1, 60 patients without nephrolithiasis; and group 2, 60 patients with severe and/or recurrent calcium nephrolithiasis. In all patients, a study of renal function, calcium metabolism, and bone remodeling markers, and a study of the lithogenic factors were performed in urine after fasting and in 24-hour urine samples. **RESULTS**: We observed greater values for phosphorus in group 1 than in group 2 (P=.03). Also, we found greater values for intact parathyroid hormone (P=.01), osteocalcin (P=.000), and beta-crosslaps (P=.000) in group 2 than in group 1. In the 24-hour urine samples, significant differences were found between groups 1 and 2 in calciuria (11.7 vs 17.4 mg/dL; P=.000), citraturia (50.6 vs 33.5 mg/dL; P=.002), calcium/creatinine quotient (0.14 vs 0.20; P=.001), calcium/citrate quotient (0.05 vs 0.13; P=.04), and calcium/creatinine quotient after fasting (0.09 vs 0.16; P=.000). **CONCLUSION**: We consider the determinants of severe and/or recurrent calcium lithiasis to be hypercalciuria and hypocitraturia and a calcium/citrate quotient>0.06. As risk markers we can consider phosphatemia35, alkaline phosphatase>80 U/L, intact parathyroid hormone>60 pg/mL, osteocalcin>16 ng/mL, beta-crosslaps>0.400 ng/mL, and beta-crosslaps/osteocalcin quotient>0.028. Copyright © 2012 Elsevier Inc. All rights reserved.

**URL:** <https://ovidsp.ovid.com/ovidweb.cgi?T=JS&CSC=Y&NEWS=N&PAGE=fulltext&D=med9&DO=10.1016%2fj.urology.2011.07.1382> <https://libkey.io/libraries/2592/openurl?genre=article&sid=OVID:medline&id=pmid:21908029&id=doi:10.1016%2Fj.urology.2011.07.1382&issn=0090-4295&isbn=&volume=79&issue=1&spage=48&pages=48-54&date=2012&title=Urology&atitle=Biochemical+determinants+of+severe+lithogenic+activity+in+patients+with+idiopathic+calcium+nephrolithiasis.&aulast=Arrabal-Polo&pid=%3Cauthor%3EArrabal-Polo+MA%3BArrabal-Martin+M%3Bde+Haro-Munoz+T%3BPoyatos-Andujar+A%3BPalaeo-Yago+F%3BZuluaga-Gomez+A%3C%2Fauthor%3E%3CAN%3E21908029%3C%2FAN%3E%3CDT%3EComparative+Study%3C%2FDT%3E>

140. **Practice patterns in evaluation of living kidney donors in United Network for Organ Sharing-approved kidney transplant centers.**

**Item Type:** Journal Article

**Authors:** Brar, Amarpali;Jindal, Rahul M.;Abbott, Kevin C.;Hurst, Frank P. and Salifu, Moro O.

**Publication Date:** 2012

**Journal:** American Journal of Nephrology 35(5), pp. 466–473

**Abstract: INTRODUCTION**: The current pattern of evaluation for living kidney donors was investigated. **METHODS**: We designed a 37-question electronic survey to collect information about living kidney donor evaluation. Of the 181 United Network for Organ Sharing (UNOS)-approved centers, 72 responded. Survey responses were coded and downloaded into SPSS. Data was expressed as means and standard deviations or the percentage of centers with specific responses. **RESULTS**: 66% of the centers used a cut-off of 140/90, whereas 22.7 and 7.1% excluded patients with pre-hypertension with a cut-off BP of 130/85 and 120/80, respectively. 66% of the centers used 24-hour urine creatinine to assess for proteinuria. 20% of the centers accepted living kidney donors with microalbuminuria and 84% accepted patients with a history of nephrolithiasis. 24% of the centers reported use of formal cognitive testing of potential living donors. **DISCUSSION**: There were significant variations in exclusion criteria based on GFR, history of kidney stones, body mass index, BP and donors with urinary abnormalities. The definitions for hematuria and proteinuria were variable. There is a need for uniformity in selection and for a living donor registry. We also recommend raising the cut-off for estimated GFR to 90 ml/min to account for 10-15% overestimation when CrCl is used. Copyright © 2012 S. Karger AG, Basel.

**URL:** <https://ovidsp.ovid.com/ovidweb.cgi?T=JS&CSC=Y&NEWS=N&PAGE=fulltext&D=med9&DO=10.1159%2f000338450> <https://libkey.io/libraries/2592/openurl?genre=article&sid=OVID:medline&id=pmid:22555113&id=doi:10.1159%2F000338450&issn=0250-8095&isbn=&volume=35&issue=5&spage=466&pages=466-73&date=2012&title=American+Journal+of+Nephrology&atitle=Practice+patterns+in+evaluation+of+living+kidney+donors+in+United+Network+for+Organ+Sharing-approved+kidney+transplant+centers.&aulast=Brar&pid=%3Cauthor%3EBrar+A%3BJindal+RM%3BAbbott+KC%3BHurst+FP%3BSalifu+MO%3C%2Fauthor%3E%3CAN%3E22555113%3C%2FAN%3E%3CDT%3EJournal+Article%3C%2FDT%3E>

141. **Effect of vitamin D repletion on urinary calcium excretion among kidney stone formers.**

**Item Type:** Journal Article

**Authors:** Leaf, David E.;Korets, Ruslan;Taylor, Eric N.;Tang, Jie;Asplin, John R.;Goldfarb, David S.;Gupta, Mantu and Curhan, Gary C.

**Publication Date:** Ma ,2012

**Journal:** Clinical Journal of the American Society of Nephrology: CJASN 7(5), pp. 829–834

**Abstract: BACKGROUND AND OBJECTIVES**: Despite the important role of vitamin D in maintaining bone health, many clinicians are reluctant to treat vitamin D deficiency in kidney stone formers because of the theoretical risk of increasing urinary calcium excretion. This study examined the effect of vitamin D repletion on urinary calcium excretion among stone formers. **DESIGN, SETTING, PARTICIPANTS, & MEASUREMENTS**: Participants (n=29) were recruited from urology clinics affiliated with New York Presbyterian Hospital. Enrollment criteria included a history of nephrolithiasis, urinary calcium excretion between 150 and 400 mg/d, and a serum 25-hydroxyvitamin D level : Participants (n=29) were recruited from urology clinics affiliated with New York Presbyterian Hospital. Enrollment criteria included a history of nephrolithiasis, urinary calcium excretion between 150 and 400 mg/d, and a serum 25-hydroxyvitamin D level **RESULTS**: Levels of 25-hydroxyvitamin D increased significantly after vitamin D repletion (17+/-6 and 35+/-10 ng/ml, P=20 mg/d; these participants also had an increase in urine sodium excretion, likely reflecting dietary variability. No participant experienced adverse effects from vitamin D, including hypercalcemia. **CONCLUSIONS**: Among stone formers with vitamin D deficiency, a limited course of vitamin D repletion does not seem to increase mean urinary calcium excretion, although a subset of individuals may have an increase. These data suggest that vitamin D therapy, if indicated, should not be withheld solely on the basis of stone disease, but 24-hour urinary calcium excretion should be monitored after repletion.

**URL:** <https://ovidsp.ovid.com/ovidweb.cgi?T=JS&CSC=Y&NEWS=N&PAGE=fulltext&D=med9&DO=10.2215%2fCJN.11331111> <https://libkey.io/libraries/2592/openurl?genre=article&sid=OVID:medline&id=pmid:22422535&id=doi:10.2215%2FCJN.11331111&issn=1555-9041&isbn=&volume=7&issue=5&spage=829&pages=829-34&date=2012&title=Clinical+Journal+of+The+American+Society+of+Nephrology%3A+CJASN&atitle=Effect+of+vitamin+D+repletion+on+urinary+calcium+excretion+among+kidney+stone+formers.&aulast=Leaf&pid=%3Cauthor%3ELeaf+DE%3BKorets+R%3BTaylor+EN%3BTang+J%3BAsplin+JR%3BGoldfarb+DS%3BGupta+M%3BCurhan+GC%3C%2Fauthor%3E%3CAN%3E22422535%3C%2FAN%3E%3CDT%3EClinical+Trial%3C%2FDT%3E>

142. **Gas-containing renal stones: findings from five consecutive patients.**

**Item Type:** Journal Article

**Authors:** Manny, Ted B.;Mufarrij, Patrick W.;Lange, Jessica N.;Mirzazadeh, Majid;Hemal, Ashok K. and Assimos, Dean G.

**Publication Date:** De ,2012

**Journal:** Urology 80(6), pp. 1203–1208

**Abstract: OBJECTIVE**: To describe the clinical course, microbiology, and metabolic findings of 5 patients presenting to our institution with gas-containing renal stones. **MATERIALS AND METHODS**: During a 20-month period beginning in 2009, 5 patients were identified by computed tomography scanning to harbor gas-containing renal calculi. Despite similar imaging and referral practice patterns, no such cases had been seen at our institution in the preceding 20 years. The records of these patients were reviewed to better characterize this unique condition. **RESULTS**: All 5 subjects were premenopausal women. One patient presented with urosepsis and 4 presented with flank pain. All had urinary tract infections, and Escherichia coli was isolated from a voided urine specimen in 3. Stone culture was positive in 2 and was concordant with the voided specimen in 1. The stones were solitary in 4 and multiple in 1 patient. All the stones were composed of calcium phosphate. Of the 5 patients, 3 had pure calcium phosphate stones and 2 had stones with calcium oxalate monohydrate components. Also, 3 subjects had diabetes mellitus, 3 had hypertension, and 1 had a history of gout. Two subjects underwent 24-hour urine metabolic testing, and abnormalities were identified in both. All patients were rendered stone free: 4 with percutaneous nephrostolithotomy and 1 using robotic pyelolithotomy. **CONCLUSION**: Gas-containing renal stones are rare but might be increasing in prevalence. The pathophysiology is unknown but is most likely influenced by a combination of metabolic and infectious factors. Copyright © 2012 Elsevier Inc. All rights reserved.

**URL:** <https://ovidsp.ovid.com/ovidweb.cgi?T=JS&CSC=Y&NEWS=N&PAGE=fulltext&D=med9&DO=10.1016%2fj.urology.2012.08.035> <https://libkey.io/libraries/2592/openurl?genre=article&sid=OVID:medline&id=pmid:23102439&id=doi:10.1016%2Fj.urology.2012.08.035&issn=0090-4295&isbn=&volume=80&issue=6&spage=1203&pages=1203-8&date=2012&title=Urology&atitle=Gas-containing+renal+stones%3A+findings+from+five+consecutive+patients.&aulast=Manny&pid=%3Cauthor%3EManny+TB%3BMufarrij+PW%3BLange+JN%3BMirzazadeh+M%3BHemal+AK%3BAssimos+DG%3C%2Fauthor%3E%3CAN%3E23102439%3C%2FAN%3E%3CDT%3EJournal+Article%3C%2FDT%3E>

143. **Effect of Ramadan fasting on urinary risk factors for calculus formation.**

**Item Type:** Journal Article

**Authors:** Miladipour, Amir Hossein;Shakhssalim, Nasser;Parvin, Mahmoud and Azadvari, Mohaddeseh

**Publication Date:** Ja ,2012

**Journal:** Iranian Journal of Kidney Diseases 6(1), pp. 33–38

**Abstract: INTRODUCTION**: Even though dehydration could aggravate formation of urinary calculi, the effects of fluid and food restriction on calculus formation is not thoroughly defined. The purpose of this study is to evaluate the effects of fluid and food restriction in Ramadan fasting on urinary factors in kidney and urinary calculus formation. **MATERIALS AND METHODS**: Fifty-seven men aged 30 to 55 years old, including 37 recurrent calcium calculus formers and 20 with no history of kidney calculi were evaluated for blood tests, ultrasonography investigations, urinalysis, urine culture, and also 24-hour urine collection test. Metabolites including calcium, oxalate, citrate, uric acid, magnesium, phosphate, potassium, sodium, and creatinine were measured before and during Ramadan fasting. The values of calculus-precipitating solutes as well as inhibitory factors were documented thoroughly. **RESULTS**: Total excretion of calcium, phosphate, and magnesium in 24-hour urine and also urine volume during fasting were significantly lower than those in the nonfasting period. Urine concentration of calcium during fasting was significantly lower than nonfasting (P : Total excretion of calcium, phosphate, and magnesium in 24-hour urine and also urine volume during fasting were significantly lower than those in the nonfasting period. Urine concentration of calcium during fasting was significantly lower than nonfasting (P **CONCLUSIONS**: Fasting during Ramadan has different effects on total excretion and concentrations of urinary precipitate and inhibitory factors contributing to calculus formation. We did not find enough evidence in favor of increased risks of calculus formation during Ramadan fasting.

**URL:** <https://ovidsp.ovid.com/ovidweb.cgi?T=JS&CSC=Y&NEWS=N&PAGE=fulltext&D=med9&AN=22218117> <https://libkey.io/libraries/2592/openurl?genre=article&sid=OVID:medline&id=pmid:22218117&id=doi:&issn=1735-8582&isbn=&volume=6&issue=1&spage=33&pages=33-8&date=2012&title=Iranian+journal+of+Kidney+Diseases&atitle=Effect+of+Ramadan+fasting+on+urinary+risk+factors+for+calculus+formation.&aulast=Miladipour&pid=%3Cauthor%3EMiladipour+AH%3BShakhssalim+N%3BParvin+M%3BAzadvari+M%3C%2Fauthor%3E%3CAN%3E22218117%3C%2FAN%3E%3CDT%3EJournal+Article%3C%2FDT%3E>

144. **Controlled metabolic diet reduces calcium oxalate supersaturation but not oxalate excretion after bariatric surgery.**

**Item Type:** Journal Article

**Authors:** Pang, Ran;Linnes, Michael P.;O'Connor, Helen M.;Li, Xujian;Bergstralh, Eric and Lieske, John C.

**Publication Date:** Au ,2012

**Journal:** Urology 80(2), pp. 250–254

**Abstract: OBJECTIVE**: To identify the effect of a controlled metabolic diet on reducing urinary calcium oxalate (CaOx) supersaturation in subjects with hyperoxaluric nephrolithiasis after potentially malabsorptive forms of bariatric surgery. **METHODS**: Subjects with a history of CaOx kidney stones and mild hyperoxaluria after bariatric surgery (n = 9) collected baseline 24-hour urine samples while consuming a free choice diet. They were then instructed to consume a controlled diet low in oxalate (70-80 mg/d), normal in calcium (1000 mg/d), and moderate in protein before 2 final 24-hour urine collections. **RESULTS**: Overall, the urinary CaOx supersaturation decreased from 1.97 +/- 0.49 delta Gibbs (DG) with the free choice diet to 1.13 +/- 0.75 DG with the controlled diet (P .05), contributing to the significant CaOx supersaturation change. **CONCLUSION**: A controlled metabolic diet normal in calcium, moderate in protein, and reduced in oxalate can positively affect urinary CaOx supersaturation after bariatric surgery. However, this diet did not appear to decrease urinary oxalate excretion. Therefore, restriction of dietary oxalate alone might not be enough to reduce urinary oxalate excretion to normal levels in this group of patients with known enteric hyperoxaluria. Additional strategies could be necessary, such as the use of oral calcium supplements as oxalate binders and a lower fat diet. Copyright © 2012 Elsevier Inc. All rights reserved.

**URL:** <https://ovidsp.ovid.com/ovidweb.cgi?T=JS&CSC=Y&NEWS=N&PAGE=fulltext&D=med9&DO=10.1016%2fj.urology.2012.02.052> <https://libkey.io/libraries/2592/openurl?genre=article&sid=OVID:medline&id=pmid:22554593&id=doi:10.1016%2Fj.urology.2012.02.052&issn=0090-4295&isbn=&volume=80&issue=2&spage=250&pages=250-4&date=2012&title=Urology&atitle=Controlled+metabolic+diet+reduces+calcium+oxalate+supersaturation+but+not+oxalate+excretion+after+bariatric+surgery.&aulast=Pang&pid=%3Cauthor%3EPang+R%3BLinnes+MP%3BO%27Connor+HM%3BLi+X%3BBergstralh+E%3BLieske+JC%3C%2Fauthor%3E%3CAN%3E22554593%3C%2FAN%3E%3CDT%3EJournal+Article%3C%2FDT%3E>

145. **Weight, age and coefficients of variation in renal solute excretion.**

**Item Type:** Journal Article

**Authors:** Perry, Guy M. L.;Scheinman, Steven J. and Asplin, John R.

**Publication Date:** 2012

**Journal:** Nephron.Physiology 122(1-2), pp. 13–18

**Abstract: BACKGROUND**: Homoscedasticity (constant variance over axes or among statistical factors) is an integral assumption of most statistical analyses. However, a number of empirical studies in model organisms and humans demonstrate significant differences in residual variance (that component of phenotype unexplained by known factors) or intra-individual variation among genotypes. Our work suggests that renal traits may be particularly susceptible to randomization by genetic and non-genetic factors, including endogenous variables like age and weight. **METHODS**: We tested associations between age, weight and intra-individual variation in urinary calcium, citrate, chloride, creatinine, potassium, magnesium, sodium, ammonium, oxalate, phosphorus, sulphate, uric acid and urea nitrogen in 9,024 male and 6,758 female kidney stone patients. Coefficients of variation (CVs) were calculated for each individual for each solute from paired 24-hour urines. Analysis of CVs was corrected for inter-measurement collection variance in creatinine and urine volume. CVs for sodium and urea nitrogen were included to correct for dietary salt and protein. **RESULTS**: Age was positively associated with individual CVs for calcium and negatively associated with CVs for potassium, ammonium and phosphorus (p(FDR) : Age was positively associated with individual CVs for calcium and negatively associated with CVs for potassium, ammonium and phosphorus (p(FDR) : Age was positively associated with individual CVs for calcium and negatively associated with CVs for potassium, ammonium and phosphorus (p(FDR) **CONCLUSION**: Intra-individual variation changes over age and weight axes for numerous urinary solutes. Changing residual variance over age and weight could cause bias in the detection or estimation of genetic or environmental effects. New methodologies may need to account for such residual unpredictability, especially in diverse collections. Copyright © 2013 S. Karger AG, Basel.

**URL:** <https://ovidsp.ovid.com/ovidweb.cgi?T=JS&CSC=Y&NEWS=N&PAGE=fulltext&D=med9&DO=10.1159%2f000346148> <https://libkey.io/libraries/2592/openurl?genre=article&sid=OVID:medline&id=pmid:23595094&id=doi:10.1159%2F000346148&issn=1660-2137&isbn=&volume=122&issue=1-2&spage=13&pages=13-8&date=2012&title=Nephron.+Physiology&atitle=Weight%2C+age+and+coefficients+of+variation+in+renal+solute+excretion.&aulast=Perry&pid=%3Cauthor%3EPerry+GM%3BScheinman+SJ%3BAsplin+JR%3C%2Fauthor%3E%3CAN%3E23595094%3C%2FAN%3E%3CDT%3EJournal+Article%3C%2FDT%3E>

146. **Biochemical control of bone loss and stone-forming propensity by potassium-calcium citrate after bariatric surgery.**

**Item Type:** Journal Article

**Authors:** Sakhaee, Khashayar;Griffith, Carolyn and Pak, Charles Y. C.

**Publication Date:** 2012

**Journal:** Surgery for Obesity & Related Diseases 8(1), pp. 67–72

**Abstract: BACKGROUND**: Patients undergoing Roux-en-Y gastric bypass (RYGB) surgery are prone to developing bone loss and kidney stones. The goal of the present study was to test the hypothesis that an effervescent formulation of potassium calcium citrate (PCC) would avert metabolic complications by providing bioavailable calcium and alkali. **METHODS**: A total of 24 patients with RYGB underwent a 2-phase crossover randomized trial comparing PCC and placebo. During the last 2 days of each 2-week phase, the serum and 24-hour urine samples were analyzed for calcium and bone turnover markers, acid base status, and urinary stone risk factors. **RESULTS**: Compared with placebo, PCC marginally reduced the serum parathyroid hormone level and significantly decreased urinary deoxypyridinoline by 12% (P : Compared with placebo, PCC marginally reduced the serum parathyroid hormone level and significantly decreased urinary deoxypyridinoline by 12% (P : Compared with placebo, PCC marginally reduced the serum parathyroid hormone level and significantly decreased urinary deoxypyridinoline by 12% (P : Compared with placebo, PCC marginally reduced the serum parathyroid hormone level and significantly decreased urinary deoxypyridinoline by 12% (P : Compared with placebo, PCC marginally reduced the serum parathyroid hormone level and significantly decreased urinary deoxypyridinoline by 12% (P **CONCLUSION**: In RYGB patients, PCC supplementation inhibited bone resorption by providing bioavailable calcium, reduced the urinary saturation of uric acid, and increased the inhibitor activity against calcium oxalate agglomeration by providing alkali that increased urinary pH and citrate. Copyright © 2012 American Society for Metabolic and Bariatric Surgery. Published by Elsevier Inc. All rights reserved.

**URL:** <https://ovidsp.ovid.com/ovidweb.cgi?T=JS&CSC=Y&NEWS=N&PAGE=fulltext&D=med9&DO=10.1016%2fj.soard.2011.05.001> <https://libkey.io/libraries/2592/openurl?genre=article&sid=OVID:medline&id=pmid:21703942&id=doi:10.1016%2Fj.soard.2011.05.001&issn=1550-7289&isbn=&volume=8&issue=1&spage=67&pages=67-72&date=2012&title=Surgery+for+Obesity+%26+Related+Diseases&atitle=Biochemical+control+of+bone+loss+and+stone-forming+propensity+by+potassium-calcium+citrate+after+bariatric+surgery.&aulast=Sakhaee&pid=%3Cauthor%3ESakhaee+K%3BGriffith+C%3BPak+CY%3C%2Fauthor%3E%3CAN%3E21703942%3C%2FAN%3E%3CDT%3EJournal+Article%3C%2FDT%3E>

147. **Persistence of hypercalciuria after successful surgical treatment for primary hyperparathyroidism.**

**Item Type:** Journal Article

**Authors:** Spivacow, Francisco Rodolfo;Negri, Armando Luis;del Valle, Elisa Elena;Fradinger, Erich;Martinez, Carolina and Polonsky, Ana

**Publication Date:** Ju ,2012

**Journal:** International Urology & Nephrology 44(3), pp. 857–863

**Abstract: UNLABELLED**: Primary hyperparathyroidism (PHPT) causes hypercalciuria and stone disease in a subset of patients. Hypercalciuria typically normalizes after surgery, although the risk of stone formation may persist up to 10 years. There are few reports in the literature that show persistent hypercalciuria despite normalization of serum calcium after parathyroid surgery. We retrospectively analyzed 111 patients with PHPT from the osteoporosis, and stone clinics seen between 1999 and 2006. We selected only patients who had a complete metabolic profile that included 24-hour collections before and at least 3 months after parathyroidectomy. We excluded patients who had creatinine clearance : Primary hyperparathyroidism (PHPT) causes hypercalciuria and stone disease in a subset of patients. Hypercalciuria typically normalizes after surgery, although the risk of stone formation may persist up to 10 years. There are few reports in the literature that show persistent hypercalciuria despite normalization of serum calcium after parathyroid surgery. We retrospectively analyzed 111 patients with PHPT from the osteoporosis, and stone clinics seen between 1999 and 2006. We selected only patients who had a complete metabolic profile that included 24-hour collections before and at least 3 months after parathyroidectomy. We excluded patients who had creatinine clearance **CONCLUSIONS**: Persistently increased fractional excretion of calcium could explain the sustained increased risk of stone disease in patients with PHPT for many years after successful parathyroidectomy.

**URL:** <https://ovidsp.ovid.com/ovidweb.cgi?T=JS&CSC=Y&NEWS=N&PAGE=fulltext&D=med9&DO=10.1007%2fs11255-011-9953-6> <https://libkey.io/libraries/2592/openurl?genre=article&sid=OVID:medline&id=pmid:21505756&id=doi:10.1007%2Fs11255-011-9953-6&issn=0301-1623&isbn=&volume=44&issue=3&spage=857&pages=857-63&date=2012&title=International+Urology+%26+Nephrology&atitle=Persistence+of+hypercalciuria+after+successful+surgical+treatment+for+primary+hyperparathyroidism.&aulast=Spivacow&pid=%3Cauthor%3ESpivacow+FR%3BNegri+AL%3Bdel+Valle+EE%3BFradinger+E%3BMartinez+C%3BPolonsky+A%3C%2Fauthor%3E%3CAN%3E21505756%3C%2FAN%3E%3CDT%3EJournal+Article%3C%2FDT%3E>

148. **A child with atypical celiac disease and recurrent urolithiasis.**

**Item Type:** Journal Article

**Authors:** Yarnell, Eric L.

**Publication Date:** Ma ,2012

**Journal:** Iranian Journal of Kidney Diseases 6(2), pp. 146–148

**Abstract:** A young boy with prior constipation developed recurrent severe calcium phosphate kidney calculi, sometimes sufficient to cause acute kidney failure and hydronephrosis. After several major surgeries, food allergies were determined by serum immunoglobulin E testing, and when he finally went on a gluten-free diet, he stopped forming calculi and has had no surgeries related to kidney calculi since. Hyperoxaluria was not identified in this child by 24-hour urine analysis, unlike most other reports of kidney calculus formation in individuals with gluten intolerance.

**URL:** <https://ovidsp.ovid.com/ovidweb.cgi?T=JS&CSC=Y&NEWS=N&PAGE=fulltext&D=med9&AN=22388615> <https://libkey.io/libraries/2592/openurl?genre=article&sid=OVID:medline&id=pmid:22388615&id=doi:&issn=1735-8582&isbn=&volume=6&issue=2&spage=146&pages=146-8&date=2012&title=Iranian+journal+of+Kidney+Diseases&atitle=A+child+with+atypical+celiac+disease+and+recurrent+urolithiasis.&aulast=Yarnell&pid=%3Cauthor%3EYarnell+EL%3C%2Fauthor%3E%3CAN%3E22388615%3C%2FAN%3E%3CDT%3ECase+Reports%3C%2FDT%3E>

149. **Treatment and prevention of kidney stones: an update**

**Item Type:** Journal Article

**Authors:** Frassetto, Lynda and Kohlstadt, Ingrid

**Publication Date:** Dec 01 ,2011

**Journal:** American Family Physician 84(11), pp. 1234–1242

**Abstract:** The incidence of nephrolithiasis (kidney stones) is rising worldwide, especially in women and with increasing age. Kidney stones are associated with chronic kidney disease. Preventing recurrence is largely specific to the type of stone (e.g., calcium oxalate, calcium phosphate, cystine, struvite [magnesium ammonium phosphate]), and uric acid stones); however, even when the stone cannot be retrieved, urine pH and 24-hour urine assessment provide information about stone-forming factors that can guide prevention. Medications, such as protease inhibitors, antibiotics, and some diuretics, increase the risk of some types of kidney stones, and patients should be counseled about the risks of using these medications. Managing diet, medication use, and nutrient intake can help prevent the formation of kidney stones. Obesity increases the risk of kidney stones. However, weight loss could undermine prevention of kidney stones if associated with a high animal protein intake, laxative abuse, rapid loss of lean tissue, or poor hydration. For prevention of calcium oxalate, cystine, and uric acid stones, urine should be alkalinized by eating a diet high in fruits and vegetables, taking supplemental or prescription citrate, or drinking alkaline mineral waters. For prevention of calcium phosphate and struvite stones, urine should be acidified; cranberry juice or betaine can lower urine pH. Antispasmodic medications, ureteroscopy, and metabolic testing are increasingly being used to augment fluid and pain medications in the acute management of kidney stones.

**URL:** <https://ovidsp.ovid.com/ovidweb.cgi?T=JS&CSC=Y&NEWS=N&PAGE=fulltext&D=med8&AN=22150656> <https://libkey.io/libraries/2592/openurl?genre=article&sid=OVID:medline&id=pmid:22150656&id=doi:&issn=0002-838X&isbn=&volume=84&issue=11&spage=1234&pages=1234-42&date=2011&title=American+Family+Physician&atitle=Treatment+and+prevention+of+kidney+stones%3A+an+update.&aulast=Frassetto&pid=%3Cauthor%3EFrassetto+L%3BKohlstadt+I%3C%2Fauthor%3E%3CAN%3E22150656%3C%2FAN%3E%3CDT%3EJournal+Article%3C%2FDT%3E>

150. **Impact of dietary calcium and oxalate, and Oxalobacter formigenes colonization on urinary oxalate excretion.**

**Item Type:** Journal Article

**Authors:** Jiang, Juquan;Knight, John;Easter, Linda H.;Neiberg, Rebecca;Holmes, Ross P. and Assimos, Dean G.

**Publication Date:** Ju ,2011

**Journal:** Journal of Urology 186(1), pp. 135–139

**Abstract: PURPOSE**: Enteric colonization with Oxalobacter formigenes, a bacterium whose main energy source is oxalate, has been demonstrated to decrease the risk of recurrent calcium oxalate kidney stone formation. We assessed the impact of diets controlled in calcium and oxalate contents on urinary and fecal analytes in healthy subjects who were naturally colonized with O. formigenes or not colonized with O. formigenes. **MATERIALS AND METHODS**: A total of 11 O. formigenes colonized and 11 noncolonized subjects were administered diets controlled in calcium and oxalate contents. We assayed 24-hour urine collections and stool samples obtained on the last 4 days of each 1-week diet for stone risk parameters and O. formigenes levels. Mixed model analysis was used to determine the effects of colonization status on these variables. **RESULTS**: Urinary calcium and oxalate excretion were significantly altered by the dietary changes in O. formigenes colonized and noncolonized individuals. Mixed model analysis showed significant interaction between colonization status and oxalate excretion on a low calcium (400 mg daily)/moderate oxalate (250 mg daily) diet (p = 0.026). Urinary oxalate excretion was 19.5% lower in O. formigenes colonized subjects than in noncolonized subjects on the low calcium/moderate oxalate diet (mean +/- SE 34.9 +/- 2.6 vs 43.6 +/- 2.6 mg, p = 0.031). **CONCLUSIONS**: Results suggest that O. formigenes colonization decreases oxalate excretion during periods of low calcium and moderate oxalate intake. Copyright © 2011 American Urological Association Education and Research, Inc. Published by Elsevier Inc. All rights reserved.

**URL:** <https://ovidsp.ovid.com/ovidweb.cgi?T=JS&CSC=Y&NEWS=N&PAGE=fulltext&D=med8&DO=10.1016%2fj.juro.2011.03.006> <https://libkey.io/libraries/2592/openurl?genre=article&sid=OVID:medline&id=pmid:21575973&id=doi:10.1016%2Fj.juro.2011.03.006&issn=0022-5347&isbn=&volume=186&issue=1&spage=135&pages=135-9&date=2011&title=Journal+of+Urology&atitle=Impact+of+dietary+calcium+and+oxalate%2C+and+Oxalobacter+formigenes+colonization+on+urinary+oxalate+excretion.&aulast=Jiang&pid=%3Cauthor%3EJiang+J%3BKnight+J%3BEaster+LH%3BNeiberg+R%3BHolmes+RP%3BAssimos+DG%3C%2Fauthor%3E%3CAN%3E21575973%3C%2FAN%3E%3CDT%3EJournal+Article%3C%2FDT%3E>

151. **Effect of renal function on urinary mineral excretion and stone composition.**

**Item Type:** Journal Article

**Authors:** Kadlec, Adam O.;Greco, Kristin A.;Fridirici, Zachary C.;Gerber, Daniel and Turk, Thomas M. T.

**Publication Date:** Oc ,2011

**Journal:** Urology 78(4), pp. 744–747

**Abstract: OBJECTIVE**: To determine the effect of renal function on urinary mineral stone excretion and composition of kidney stones in patients undergoing urologic intervention for nephrolithiasis. **METHODS**: We performed a retrospective analysis of 158 patients with 193 kidney stones who underwent endourologic intervention for nephrolithiasis. The patients were grouped by the estimated glomerular filtration rate (eGFR). The kidney stone composition and results of the 24-hour urinalysis were reported for each group. Statistical analysis was performed using the chi-square test, Kruskal-Wallis test, and analysis of variance. **RESULTS**: The kidney stone composition profile differed significantly between the groups, with uric acid stones associated with a lower eGFR and calcium phosphate stones associated with a greater eGFR. A lower eGFR correlated with lower urine pH, lower calcium excretion, and greater oxalate excretion. Excretion of uric acid and the body mass index did not differ between groups. **CONCLUSION**: Different degrees of renal function correlated with certain types of kidney stones. The urine pH was lower in patients with impaired renal function. Renal function might be an underappreciated factor that affects kidney stone composition and urinary mineral excretion. Copyright © 2011 Elsevier Inc. All rights reserved.

**URL:** <https://ovidsp.ovid.com/ovidweb.cgi?T=JS&CSC=Y&NEWS=N&PAGE=fulltext&D=med8&DO=10.1016%2fj.urology.2011.04.007> <https://libkey.io/libraries/2592/openurl?genre=article&sid=OVID:medline&id=pmid:21664652&id=doi:10.1016%2Fj.urology.2011.04.007&issn=0090-4295&isbn=&volume=78&issue=4&spage=744&pages=744-7&date=2011&title=Urology&atitle=Effect+of+renal+function+on+urinary+mineral+excretion+and+stone+composition.&aulast=Kadlec&pid=%3Cauthor%3EKadlec+AO%3BGreco+KA%3BFridirici+ZC%3BGerber+D%3BTurk+TM%3C%2Fauthor%3E%3CAN%3E21664652%3C%2FAN%3E%3CDT%3EJournal+Article%3C%2FDT%3E>

152. **Determinants of osteopenia in male renal-stone-disease patients with idiopathic hypercalciuria.**

**Item Type:** Journal Article

**Authors:** Letavernier, Emmanuel;Traxer, Olivier;Daudon, Michel;Tligui, Mohammed;Hubert-Brierre, Jerome;Guerrot, Dominique;Sebag, Aline;Baud, Laurent and Haymann, Jean-Philippe

**Publication Date:** Ma ,2011

**Journal:** Clinical Journal of the American Society of Nephrology: CJASN 6(5), pp. 1149–1154

**Abstract: BACKGROUND AND OBJECTIVES**: Bone demineralization is frequent in renal-stone formers with hypercalciuria. Although this pathologic link has been recognized for decades, the underlying mechanisms and risk factors associated with osteopenia/osteoporosis in this population remain partially understood. **DESIGN, SETTING, PARTICIPANTS, & MEASUREMENTS**: This study retrospectively analyzed determinants of low bone mineral density (BMD) in 65 idiopathic hypercalciuric male renal-stone formers. Clinical and biologic evaluation included BMD measurement, bone-remodeling markers, analysis of calcium metabolism with oral calcium load test, and dietary inquiry. **RESULTS**: Patients with osteopenia (n=23, 35% of the population) presented significantly higher fasting calciuria as compared with normal bone density patients (n=42) (calcium/creatinine ratio was 0.32 versus 0.24 mmol/mmol; P=0.006). Analysis of the whole population revealed a negative association between fasting hypercalciuria and BMD (P = 0.003), independent of confounding variables including body-mass index and tobacco consumption. The fasting calcium/creatinine ratio above 0.25 mmol/mmol was associated with a 3.8-fold increase in the risk of low BMD. **CONCLUSION**: In our study, fasting hypercalciuria after a 2-day calcium-restricted diet appears as the only biologic factor associated with low BMD, suggesting a bone-calcium efflux. Our results support the view of a parathyroid-independent pathologic process that remains to be identified. Hypercalciuric patients with low BMD do not excrete more calcium in 24-hour urine samples than patients without low BMD. Copyright © 2011 by the American Society of Nephrology

**URL:** <https://ovidsp.ovid.com/ovidweb.cgi?T=JS&CSC=Y&NEWS=N&PAGE=fulltext&D=med8&DO=10.2215%2fCJN.10191110> <https://libkey.io/libraries/2592/openurl?genre=article&sid=OVID:medline&id=pmid:21441130&id=doi:10.2215%2FCJN.10191110&issn=1555-9041&isbn=&volume=6&issue=5&spage=1149&pages=1149-54&date=2011&title=Clinical+Journal+of+The+American+Society+of+Nephrology%3A+CJASN&atitle=Determinants+of+osteopenia+in+male+renal-stone-disease+patients+with+idiopathic+hypercalciuria.&aulast=Letavernier&pid=%3Cauthor%3ELetavernier+E%3BTraxer+O%3BDaudon+M%3BTligui+M%3BHubert-Brierre+J%3BGuerrot+D%3BSebag+A%3BBaud+L%3BHaymann+JP%3C%2Fauthor%3E%3CAN%3E21441130%3C%2FAN%3E%3CDT%3EJournal+Article%3C%2FDT%3E>

153. **Splenda R improves tolerance of oral potassium citrate supplementation for prevention of stone formation: results of a randomized double-blind trial.**

**Item Type:** Journal Article

**Authors:** Mechlin, Clay;Kalorin, Carmin;Asplin, John and White, Mark

**Publication Date:** Se ,2011

**Journal:** Journal of Endourology 25(9), pp. 1541–1545

**Abstract: BACKGROUND AND PURPOSE**: Oral citrate supplements have been shown to decrease kidney stone recurrence rates in both laboratory and clinical studies. The taste of the citrate supplements, however, is poor, and long-term compliance is low. Our objective was to determine if Splenda( R) added to potassium citrate (KCit) improves palatability without changing 24-hour urine parameters. **PATIENTS AND METHODS**: 12 subjects were randomly assigned to receive either KCit alone for 3 days or KCit + Splenda in a double-blind trial. The 24-hour urine collections were performed before and after 3 days of therapy. After 1 week, the two groups switched treatments. After each treatment, a visual analog taste scale was completed to gauge the taste and palatability. The 24-hour urine parameters of kidney stone risk factors were compared between groups. The primary end points were to determine whether Splenda improved palatability of citrate supplementation and whether it altered 24-hour urine parameters. **RESULTS**: Taste was judged to be 2.5 +/- 0.9 points better in the Splenda + KCit compared with KCit alone (P=0.02). The 24-hour Cit, K, and pH were significantly higher in the KCit and KCit + Splenda groups compared with baseline, but not significantly different from each other. **CONCLUSION**: Splenda significantly improves the palatability of KCit therapy and does not alter the beneficial effects of KCit on 24-hour urine Cit, K, or pH. The addition of Splenda altered the average taste score from one that might prohibit compliance to one that would not.

**URL:** <https://ovidsp.ovid.com/ovidweb.cgi?T=JS&CSC=Y&NEWS=N&PAGE=fulltext&D=med8&DO=10.1089%2fend.2010.0663> <https://libkey.io/libraries/2592/openurl?genre=article&sid=OVID:medline&id=pmid:21815827&id=doi:10.1089%2Fend.2010.0663&issn=0892-7790&isbn=&volume=25&issue=9&spage=1541&pages=1541-5&date=2011&title=Journal+of+Endourology&atitle=Splenda+R+improves+tolerance+of+oral+potassium+citrate+supplementation+for+prevention+of+stone+formation%3A+results+of+a+randomized+double-blind+trial.&aulast=Mechlin&pid=%3Cauthor%3EMechlin+C%3BKalorin+C%3BAsplin+J%3BWhite+M%3C%2Fauthor%3E%3CAN%3E21815827%3C%2FAN%3E%3CDT%3EJournal+Article%3C%2FDT%3E>

154. **Effect of n-3 fatty acid supplementation on urinary risk factors for calcium oxalate stone formation.**

**Item Type:** Journal Article

**Authors:** Siener, Roswitha;Jansen, Brigitte;Watzer, Bernhard and Hesse, Albrecht

**Publication Date:** Fe ,2011

**Journal:** Journal of Urology 185(2), pp. 719–724

**Abstract: PURPOSE**: Findings are inconsistent in a few studies of the effect of n-3 fatty acid supplementation on urinary calcium and oxalate excretion in stone formers. We evaluated the physiological effects of supplementation with eicosapentaenoic acid and docosahexaenoic acid on urinary risk factors for calcium oxalate stone formation under standardized conditions. **MATERIALS AND METHODS**: We studied 15 healthy subjects initially while consuming a standardized diet for 5 days (control phase). During consecutive intervention phases 1-5-day standardized diet, 2-20-day free diet and 3-5-day standardized diet participants received 900 mg eicosapentaenoic acid and 600 mg docosahexaenoic acid daily. While ingesting the standardized diets, daily 24-hour urine samples were collected. **RESULTS**: After short-term supplementation with eicosapentaenoic acid and docosahexaenoic acid in phase 1 we noted no changes in urinary parameters compared to the control phase. After 30-day supplementation with eicosapentaenoic acid and docosahexaenoic acid in phase 3 relative supersaturation with calcium oxalate decreased significantly by 23% from a mean +/- SD of 2.01 +/- 1.26 to 1.55 +/- 0.84 due to significantly decreased urinary oxalate excretion (p = 0.023). Other urinary variables were not affected by supplementation. **CONCLUSIONS**: Results show that 30-day n-3 fatty acid supplementation effectively decreases urinary oxalate excretion and the risk of calcium oxalate crystallization. The mechanism of the physiological effect may be decreased cellular oxalic acid exchange attributable to an altered fatty acid pattern of membrane phospholipids with concomitant changes in oxalate transporter activity. Calcium oxalate stone formers may benefit from long-term n-3 fatty acid supplementation. Copyright A© 2011 American Urological Association Education and Research, Inc. Published by Elsevier Inc. All rights reserved.

**URL:** <https://ovidsp.ovid.com/ovidweb.cgi?T=JS&CSC=Y&NEWS=N&PAGE=fulltext&D=med8&DO=10.1016%2fj.juro.2010.09.074> <https://libkey.io/libraries/2592/openurl?genre=article&sid=OVID:medline&id=pmid:21168878&id=doi:10.1016%2Fj.juro.2010.09.074&issn=0022-5347&isbn=&volume=185&issue=2&spage=719&pages=719-24&date=2011&title=Journal+of+Urology&atitle=Effect+of+n-3+fatty+acid+supplementation+on+urinary+risk+factors+for+calcium+oxalate+stone+formation.&aulast=Siener&pid=%3Cauthor%3ESiener+R%3BJansen+B%3BWatzer+B%3BHesse+A%3C%2Fauthor%3E%3CAN%3E21168878%3C%2FAN%3E%3CDT%3EComparative+Study%3C%2FDT%3E>

155. **Regressive course of oxalate deposition in primary hyperoxaluria after kidney transplantation.**

**Item Type:** Journal Article

**Authors:** Celik, G.;Sen, S.;Sipahi, S.;Akkin, C.;Tamsel, S.;Toz, H. and Hoscoskun, C.

**Publication Date:** 2010

**Journal:** Renal Failure 32(9), pp. 1131–1136

**Abstract:** Primary hyperoxaluria (PH) is a rare autosomal recessive disease caused by the functional defect of alanine-glyoxylate aminotransferase (AGT) enzyme in the liver and it is characterized by the deposition of diffuse calcium oxalate crystals. A 38-year-old male patient presented with history of recurrent nephrolithiasis and has received chronic hemodialysis treatment for 2 years. Cadaveric renal transplantation was applied to the case. The patient was reoperated on postoperative day 13 because of the collection surrounding the urethra. During this operation, kidney biopsy was made due to late decrease in creatinine levels. Deposition of diffuse oxalate crystal was detected in allograft kidney biopsy, whereas in the 0-hour biopsy there were no oxalate crystals. Oxalate level was found to be high in a 24-hour urine specimen (118 mg/L, normal level: 7-44 mg/L). The patient was identified with primary hyperoxaluria and followed up in terms of systemic oxalate deposition as well as allograft kidney. In the kidney biopsy taken after 18 months, we detected that oxalate crystals almost entirely disappeared. In our case, bilateral preretinal, intraretinal, and intravascular diffuse oxalate crystals were detected, and argon laser photocoagulation treatments were needed for choroidal and retinal neovascularization. Repeated ophthalmic examinations showed the regressive nature of oxalate depositions. In the 18th month, fundus examination and fluorescein angiography revealed that oxalate crystals were significantly regressed. To increase the quality of life and slow down the systemic effects of oxalosis, kidney-only transplantation is beneficial.

**URL:** <https://ovidsp.ovid.com/ovidweb.cgi?T=JS&CSC=Y&NEWS=N&PAGE=fulltext&D=med8&DO=10.3109%2f0886022X.2010.509900> <https://libkey.io/libraries/2592/openurl?genre=article&sid=OVID:medline&id=pmid:20863224&id=doi:10.3109%2F0886022X.2010.509900&issn=0886-022X&isbn=&volume=32&issue=9&spage=1131&pages=1131-6&date=2010&title=Renal+Failure&atitle=Regressive+course+of+oxalate+deposition+in+primary+hyperoxaluria+after+kidney+transplantation.&aulast=Celik&pid=%3Cauthor%3ECelik+G%3BSen+S%3BSipahi+S%3BAkkin+C%3BTamsel+S%3BToz+H%3BHoscoskun+C%3C%2Fauthor%3E%3CAN%3E20863224%3C%2FAN%3E%3CDT%3ECase+Reports%3C%2FDT%3E>

156. **Metabolic abnormalities in patients with nephrolithiasis: comparison of first-episode with recurrent cases in Southern Iran.**

**Item Type:** Journal Article

**Authors:** Hosseini, Mohammad Mehdi;Eshraghian, Ahad;Dehghanian, Ilnaz;Irani, Dariush and Amini, Mitra

**Publication Date:** Ma ,2010

**Journal:** International Urology & Nephrology 42(1), pp. 127–131

**Abstract: BACKGROUND**: Metabolic disorders are one of the etiologic factors in renal stone formation. The aim of present study was to evaluate prevalence of metabolic disorders in patients with renal stone. **METHODS**: From 572 patients referring to our urologic clinics, 376 patients participated in the study. Patients were divided to first time stone former (group A) and recurrent renal stone (group B). Twenty-four hour urine specimens were obtained for urinary calcium, oxalate, magnesium, citrate, uric acid, phosphor and 24-h urine volume. Venous blood samples were obtained evaluating serum phosphate, uric acid, and calcium. The prevalence of each metabolic disorder was detected, and two groups were compared regarding metabolic disorders. **RESULTS**: The prevalence of renal stone in male patients was 63.04% versus 36.96% in female patients (P 0.05). **CONCLUSION**: Low 24-h urine volume was the most common abnormalities in patients with nephrolithiasis in our region. Metabolic evaluation must be performed in all patients with renal stone even those with first time stone formation.

**URL:** <https://ovidsp.ovid.com/ovidweb.cgi?T=JS&CSC=Y&NEWS=N&PAGE=fulltext&D=med8&DO=10.1007%2fs11255-009-9599-9> <https://libkey.io/libraries/2592/openurl?genre=article&sid=OVID:medline&id=pmid:19548107&id=doi:10.1007%2Fs11255-009-9599-9&issn=0301-1623&isbn=&volume=42&issue=1&spage=127&pages=127-31&date=2010&title=International+Urology+%26+Nephrology&atitle=Metabolic+abnormalities+in+patients+with+nephrolithiasis%3A+comparison+of+first-episode+with+recurrent+cases+in+Southern+Iran.&aulast=Hosseini&pid=%3Cauthor%3EHosseini+MM%3BEshraghian+A%3BDehghanian+I%3BIrani+D%3BAmini+M%3C%2Fauthor%3E%3CAN%3E19548107%3C%2FAN%3E%3CDT%3EComparative+Study%3C%2FDT%3E>

157. **Profile of the brushite stone former.**

**Item Type:** Journal Article

**Authors:** Krambeck, Amy E.;Handa, Shelly E.;Evan, Andrew P. and Lingeman, James E.

**Publication Date:** Oc ,2010

**Journal:** Journal of Urology 184(4), pp. 1367–1371

**Abstract: PURPOSE**: The incidence of brushite stones has increased during the last 3 decades and we report our experience with brushite stone formers. **MATERIALS AND METHODS**: From 1996 to 2008 we identified 82 patients with brushite urinary calculi. After institutional review board approval a review of our prospectively collected database was performed. **RESULTS**: There were 54 (65.9%) male and 28 (34.1%) female stone formers. Mean age was 44 years (range 4 to 84). Prior stone events were reported by 69 (84.1%) patients with 54 (78.3%) having received shock wave lithotripsy. Bilateral calculi were present in 28 (34.1%) patients. Mean stone area was 29.2 mm(2) (range 2 to 130). Surgery was performed in 80 patients including 63 (76.8%) percutaneous nephrolithotomy, 8 (9.8%) ureteroscopy, 3 (3.7%) shock wave lithotripsy, and 6 (7.3%) ureteroscopy and percutaneous nephrolithotomy. After primary and secondary procedures 76 (92.7%) patients were rendered stone-free. Metabolic urine studies were available in 45 patients. All patients demonstrated 1 or more abnormalities, with hypercalciuria (greater than 250 mg daily for women and greater than 275 mg daily for men) in 38 (80.9%), urine pH greater than 6.2 in 29 (61.7%), urine volume less than 2 l in 27 (57.4%), hypocitraturia (less than 320 mg daily) in 22 (46.8%), hyperuricosuria (greater than 750 mg daily in women, greater than 800 mg daily in men) in 8 (17%) and hyperoxaluria (greater than 32 mg daily in women and greater than 43 mg daily in men) in 5 (10.6%). Recurrent stone events occurred in 31 (37.8%) patients at a mean of 33 (range 2 to 118) months from treatment. **CONCLUSIONS**: Brushite stone formers are a treatment challenge. Almost a third will present with bilateral stones and the stone burden is sizeable. Nearly 80% of patients report having prior shock wave lithotripsy and recurrent stone events occurred approximately 3 years after treatment. All patients with brushite stones in this cohort had an underlying metabolic abnormality and specifically brushite stones should be heralded as a marker for hypercalciuria. Based on these data we recommend all brushite stone formers undergo 24-hour urine studies and have close long-term followup. Copyright © 2010 American Urological Association Education and Research, Inc. Published by Elsevier Inc. All rights reserved.

**URL:** <https://ovidsp.ovid.com/ovidweb.cgi?T=JS&CSC=Y&NEWS=N&PAGE=fulltext&D=med8&DO=10.1016%2fj.juro.2010.05.094> <https://libkey.io/libraries/2592/openurl?genre=article&sid=OVID:medline&id=pmid:20719342&id=doi:10.1016%2Fj.juro.2010.05.094&issn=0022-5347&isbn=&volume=184&issue=4&spage=1367&pages=1367-71&date=2010&title=Journal+of+Urology&atitle=Profile+of+the+brushite+stone+former.&aulast=Krambeck&pid=%3Cauthor%3EKrambeck+AE%3BHanda+SE%3BEvan+AP%3BLingeman+JE%3C%2Fauthor%3E%3CAN%3E20719342%3C%2FAN%3E%3CDT%3EJournal+Article%3C%2FDT%3E>

158. **Analysis of 24-hour urine parameters as it relates to age of onset of cystine stone formation.**

**Item Type:** Journal Article

**Authors:** Lambert, Erica H.;Asplin, John R.;Herrell, Stanley Duke and Miller, Nicole L.

**Publication Date:** Ju ,2010

**Journal:** Journal of Endourology 24(7), pp. 1179–1182

**Abstract: INTRODUCTION**: Cystinuria is an inherited disorder characterized by the impaired reabsorption of the dibasic amino acid, cystine, in the proximal tubule of the nephron. Cystine stones most frequently occur in the first through third decades of life with a decreased incidence in old age. We hypothesize that patients presenting with first stone event at an older age would have more favorable 24-hour urine parameters compared with those patients who present early. **PATIENTS**: This is a retrospective review of 94 patients with cystinuria and cystine stones who underwent a 24-hour urine test. The patients were divided into four groups based on age at first clinical stone event: group 1, children aged 50 years (n = 19). All 24-hour urines were collected prior to any medical therapy. Statistical analysis was performed using analysis of variance and Tukey pairwise comparison test. **RESULTS**: There was a significant increase in cystine supersaturation (SS) for the young adults (group 2) as opposed to older adults (groups 3 and 4) though cystine excretion was not different among the groups. A higher urine volume appeared to account for the lower cystine SS in the older adults. The children were found to have cystine SS similar to the young adults. **DISCUSSION**: In this retrospective study, a correlation was found between the age of onset of cystine stones and 24-hour urine parameters. Cystinuric patients presenting with first stone event at an older age have lower cystine SS and increased urinary volume compared with patients presenting earlier, likely conferring a protective effect.

**URL:** <https://ovidsp.ovid.com/ovidweb.cgi?T=JS&CSC=Y&NEWS=N&PAGE=fulltext&D=med8&DO=10.1089%2fend.2010.0133> <https://libkey.io/libraries/2592/openurl?genre=article&sid=OVID:medline&id=pmid:20624086&id=doi:10.1089%2Fend.2010.0133&issn=0892-7790&isbn=&volume=24&issue=7&spage=1179&pages=1179-82&date=2010&title=Journal+of+Endourology&atitle=Analysis+of+24-hour+urine+parameters+as+it+relates+to+age+of+onset+of+cystine+stone+formation.&aulast=Lambert&pid=%3Cauthor%3ELambert+EH%3BAsplin+JR%3BHerrell+SD%3BMiller+NL%3C%2Fauthor%3E%3CAN%3E20624086%3C%2FAN%3E%3CDT%3EComparative+Study%3C%2FDT%3E>

159. **Effects of a low-salt diet on idiopathic hypercalciuria in calcium-oxalate stone formers: a 3-mo randomized controlled trial.**

**Item Type:** Journal Article

**Authors:** Nouvenne, Antonio;Meschi, Tiziana;Prati, Beatrice;Guerra, Angela;Allegri, Franca;Vezzoli, Giuseppe;Soldati, Laura;Gambaro, Giovanni;Maggiore, Umberto and Borghi, Loris

**Publication Date:** Ma ,2010

**Journal:** American Journal of Clinical Nutrition 91(3), pp. 565–570

**Abstract: BACKGROUND**: A direct relation exists between sodium and calcium excretion, but randomized studies evaluating the sustained effect of a low-salt diet on idiopathic hypercalciuria, one of the main risk factors for calcium-oxalate stone formation, are still lacking. **OBJECTIVE**: Our goal was to evaluate the effect of a low-salt diet on urinary calcium excretion in patients affected by idiopathic calcium nephrolithiasis. **DESIGN**: Patients affected by idiopathic calcium stone disease and hypercalciuria (>300 mg Ca/d in men and >250 mg Ca/d in women) were randomly assigned to receive either water therapy alone (control diet) or water therapy and a low-salt diet (low-sodium diet) for 3 mo. Twenty-four-hour urine samples were obtained twice from all patients: one sample at baseline on a free diet and one sample after 3 mo of treatment. **RESULTS**: A total of 210 patients were randomly assigned to receive a control diet (n = 102) or a low-sodium diet (n = 108); 13 patients (2 on the control diet, 11 on the low-sodium diet) withdrew from the trial. At the follow-up visit, patients on the low-sodium diet had lower urinary sodium (mean +/- SD: 68 +/- 43 mmol/d at 3 mo compared with 228 +/- 57 mmol/d at baseline; P : A total of 210 patients were randomly assigned to receive a control diet (n = 102) or a low-sodium diet (n = 108); 13 patients (2 on the control diet, 11 on the low-sodium diet) withdrew from the trial. At the follow-up visit, patients on the low-sodium diet had lower urinary sodium (mean +/- SD: 68 +/- 43 mmol/d at 3 mo compared with 228 +/- 57 mmol/d at baseline; P : A total of 210 patients were randomly assigned to receive a control diet (n = 102) or a low-sodium diet (n = 108); 13 patients (2 on the control diet, 11 on the low-sodium diet) withdrew from the trial. At the follow-up visit, patients on the low-sodium diet had lower urinary sodium (mean +/- SD: 68 +/- 43 mmol/d at 3 mo compared with 228 +/- 57 mmol/d at baseline; P : A total of 210 patients were randomly assigned to receive a control diet (n = 102) or a low-sodium diet (n = 108); 13 patients (2 on the control diet, 11 on the low-sodium diet) withdrew from the trial. At the follow-up visit, patients on the low-sodium diet had lower urinary sodium (mean +/- SD: 68 +/- 43 mmol/d at 3 mo compared with 228 +/- 57 mmol/d at baseline; P **CONCLUSION**: A low-salt diet can reduce calcium excretion in hypercalciuric stone formers. This trial was registered at clinicaltrials.gov as NCT01005082.

**URL:** <https://ovidsp.ovid.com/ovidweb.cgi?T=JS&CSC=Y&NEWS=N&PAGE=fulltext&D=med8&DO=10.3945%2fajcn.2009.28614> <https://libkey.io/libraries/2592/openurl?genre=article&sid=OVID:medline&id=pmid:20042524&id=doi:10.3945%2Fajcn.2009.28614&issn=0002-9165&isbn=&volume=91&issue=3&spage=565&pages=565-70&date=2010&title=American+Journal+of+Clinical+Nutrition&atitle=Effects+of+a+low-salt+diet+on+idiopathic+hypercalciuria+in+calcium-oxalate+stone+formers%3A+a+3-mo+randomized+controlled+trial.&aulast=Nouvenne&pid=%3Cauthor%3ENouvenne+A%3BMeschi+T%3BPrati+B%3BGuerra+A%3BAllegri+F%3BVezzoli+G%3BSoldati+L%3BGambaro+G%3BMaggiore+U%3BBorghi+L%3C%2Fauthor%3E%3CAN%3E20042524%3C%2FAN%3E%3CDT%3EJournal+Article%3C%2FDT%3E>

160. **The effect of restrictive bariatric surgery on urinary stone risk factors.**

**Item Type:** Journal Article

**Authors:** Semins, Michelle J.;Asplin, John R.;Steele, Kimberly;Assimos, Dean G.;Lingeman, James E.;Donahue, Susan;Magnuson, Thomas;Schweitzer, Michael and Matlaga, Brian R.

**Publication Date:** Oc ,2010

**Journal:** Urology 76(4), pp. 826–829

**Abstract: OBJECTIVES**: Malabsorptive bariatric procedures such as Roux-en-Y gastric bypass (RYGB) place patients at risk for developing kidney stones. Stone risk factors after purely restrictive procedures such as gastric banding and sleeve gastrectomy are not well characterized. Therefore, we performed a study to examine urinary risk factors of patients who underwent restrictive gastric surgery for bariatric indications. **METHODS**: A total of 18 patients were enrolled in the study; 14 underwent gastric banding and 4 underwent sleeve gastrectomy. All subjects collected 24-hour urine specimens; at least 6 months had elapsed between surgery and urine collection. Standard stone risk parameters were assessed, and comparisons were made with a group of normal adult nonstone-formers, routine stone-formers, and RYGB bariatric surgery subjects. **RESULTS**: Urinary oxalate excretion of the restrictive cohort was significantly less than the RYGB cohort (35.4 vs. 60.7 mg/d; P : Urinary oxalate excretion of the restrictive cohort was significantly less than the RYGB cohort (35.4 vs. 60.7 mg/d; P **CONCLUSIONS**: Restrictive bariatric surgery does not appear to be associated with an increased risk for kidney stone disease. In particular, urinary oxalate levels were significantly less than those of RYGB subjects and not significantly different from routine stone-formers and nonstone-forming controls. Copyright © 2010 Elsevier Inc. All rights reserved.

**URL:** <https://ovidsp.ovid.com/ovidweb.cgi?T=JS&CSC=Y&NEWS=N&PAGE=fulltext&D=med8&DO=10.1016%2fj.urology.2010.01.037> <https://libkey.io/libraries/2592/openurl?genre=article&sid=OVID:medline&id=pmid:20381135&id=doi:10.1016%2Fj.urology.2010.01.037&issn=0090-4295&isbn=&volume=76&issue=4&spage=826&pages=826-9&date=2010&title=Urology&atitle=The+effect+of+restrictive+bariatric+surgery+on+urinary+stone+risk+factors.&aulast=Semins&pid=%3Cauthor%3ESemins+MJ%3BAsplin+JR%3BSteele+K%3BAssimos+DG%3BLingeman+JE%3BDonahue+S%3BMagnuson+T%3BSchweitzer+M%3BMatlaga+BR%3C%2Fauthor%3E%3CAN%3E20381135%3C%2FAN%3E%3CDT%3EComparative+Study%3C%2FDT%3E>

161. **Metabolic investigation of patients with staghorn calculus: is it necessary?.**

**Item Type:** Journal Article

**Authors:** Amaro, Carmen R. P.;Goldberg, Jose;Agostinho, Aparecido D.;Damasio, Patricia;Kawano, Paulo R.;Fugita, Oscar E. H. and Amaro, Joao L.

**Publication Date:** 2009

**Journal:** International Braz J Urol 35(6), pp. 658–661

**Abstract: OBJECTIVE**: To evaluate the prevalence of metabolic disorders in patients with staghorn calculi treated at the Regional Center of Lithiasis Metabolic Studies in central region of Sao Paulo State, Brazil. **MATERIALS AND METHODS**: Between February 2000 and February 2008, 630 patients with urinary calculi were evaluated in the lithiasis outpatient clinic. Thirty-seven of them had staghorn calculi (35 women and 2 men). The inclusion criteria for the metabolic investigation included the absence of urological manipulation 30 days before the examination, negative urine culture and creatinine clearance > 60 mL/min. The protocol for metabolic investigation consisted of qualitative search for cystinuria. Two non-consecutive 24-hour urine samples collected to measure calcium, phosphorus, uric acid, sodium, potassium, magnesium, oxalate and citrate, and serum calcium levels, phosphorus, uric acid, sodium, potassium, magnesium, chloride, parathormone and urine pH. **RESULTS**: Among patients with lithiasis, 5.9% (37/630) had staghorn calculus and in 48.6% (18/37) were diagnosed with urinary infection. The females were predominant for 94.5% of cases. The calculi were unilateral in 31 of cases and bilateral in six. Metabolic abnormalities were found in 68.2% of patients with hypercalciuria (64.2%) and hypocitraturia (53.3%) being the most common disorders. **CONCLUSIONS**: The presence of metabolic disorders in nearly 70% of patients with staghorn calculus reinforces the necessity for evaluation of these patients. The diagnosis and treatment of identified metabolic abnormalities can contribute to the prevention of recurrent staghorn calculi.

**URL:** <https://ovidsp.ovid.com/ovidweb.cgi?T=JS&CSC=Y&NEWS=N&PAGE=fulltext&D=med7&DO=10.1590%2fs1677-55382009000600004> <https://libkey.io/libraries/2592/openurl?genre=article&sid=OVID:medline&id=pmid:20028571&id=doi:10.1590%2Fs1677-55382009000600004&issn=1677-5538&isbn=&volume=35&issue=6&spage=658&pages=658-61%3B+discussion+662-3&date=2009&title=International+Braz+J+Urol&atitle=Metabolic+investigation+of+patients+with+staghorn+calculus%3A+is+it+necessary%3F.&aulast=Amaro&pid=%3Cauthor%3EAmaro+CR%3BGoldberg+J%3BAgostinho+AD%3BDamasio+P%3BKawano+PR%3BFugita+OE%3BAmaro+JL%3C%2Fauthor%3E%3CAN%3E20028571%3C%2FAN%3E%3CDT%3EJournal+Article%3C%2FDT%3E>

162. **Patients with primary hyperparathyroidism--why do some form stones?.**

**Item Type:** Journal Article

**Authors:** Berger, Aaron D.;Wu, Wilfred;Eisner, Brian H.;Cooperberg, Matthew R.;Duh, Quan-Yang and Stoller, Marshall L.

**Publication Date:** Ma ,2009

**Journal:** Journal of Urology 181(5), pp. 2141–2145

**Abstract: PURPOSE**: Primary hyperparathyroidism is a common cause of hypercalcemia with resulting hypercalciuria and a subset of these patients with primary hyperparathyroidism present with urolithiasis. We evaluated the metabolic stone profile in patients presenting for parathyroid surgery and determined whether there were differences between those with and without a history of urinary stones. **MATERIALS AND METHODS**: A total of 60 patients with primary hyperparathyroidism presenting for parathyroid surgery were prospectively enrolled in the study, of whom 54 completed a preoperative 24-hour urine collection. Of these 54 patients 27 completed an additional postoperative 24-hour urine collection. We compared preoperative metabolic urinary profiles and serum calcium in patients with primary hyperparathyroidism with and without a history of urolithiasis. In addition, we compared changes in urine metabolic profiles that occurred after successful parathyroid surgery. **RESULTS**: There were no significant differences in preoperative 24-hour urine metabolic profiles or serum calcium between patients who had primary hyperparathyroidism with and without a history of urolithiasis. Compared to preoperative levels after successful parathyroid surgery there were significant decreases in serum calcium (10.8 to 9.3 mg/dl, p : There were no significant differences in preoperative 24-hour urine metabolic profiles or serum calcium between patients who had primary hyperparathyroidism with and without a history of urolithiasis. Compared to preoperative levels after successful parathyroid surgery there were significant decreases in serum calcium (10.8 to 9.3 mg/dl, p : There were no significant differences in preoperative 24-hour urine metabolic profiles or serum calcium between patients who had primary hyperparathyroidism with and without a history of urolithiasis. Compared to preoperative levels after successful parathyroid surgery there were significant decreases in serum calcium (10.8 to 9.3 mg/dl, p **CONCLUSIONS**: Other etiological factors must exist that cause some patients with primary hyperparathyroidism to form stones, while most never have stones despite profound hypercalcemia and hypercalciuria. Routine 24-hour urine evaluation cannot predict which patients with primary hyperparathyroidism will have kidney stones.

**URL:** <https://ovidsp.ovid.com/ovidweb.cgi?T=JS&CSC=Y&NEWS=N&PAGE=fulltext&D=med7&DO=10.1016%2fj.juro.2009.01.028> <https://libkey.io/libraries/2592/openurl?genre=article&sid=OVID:medline&id=pmid:19296981&id=doi:10.1016%2Fj.juro.2009.01.028&issn=0022-5347&isbn=&volume=181&issue=5&spage=2141&pages=2141-5&date=2009&title=Journal+of+Urology&atitle=Patients+with+primary+hyperparathyroidism--why+do+some+form+stones%3F.&aulast=Berger&pid=%3Cauthor%3EBerger+AD%3BWu+W%3BEisner+BH%3BCooperberg+MR%3BDuh+QY%3BStoller+ML%3C%2Fauthor%3E%3CAN%3E19296981%3C%2FAN%3E%3CDT%3EComparative+Study%3C%2FDT%3E>

163. **Effect of soda consumption on urinary stone risk parameters.**

**Item Type:** Journal Article

**Authors:** Passman, Corey M.;Holmes, Ross P.;Knight, John;Easter, Linda;Pais, Vernon and Assimos, Dean G.

**Publication Date:** Ma ,2009

**Journal:** Journal of Endourology 23(3), pp. 347–350

**Abstract: BACKGROUND AND PURPOSE**: Fluid consumption has been demonstrated to influence kidney stone formation. Studies have shown that consumption of cola may be a risk factor for stone disease, while fluids containing citric acid may attenuate stone activity. Diet was not always controlled in these investigations, however. We undertook a study to determine the impact of three different fluids on urinary stone risk factors. **SUBJECTS AND METHODS**: Six healthy nonstone-forming adults were placed on a standardized metabolic diet and consumed three different types of fluid during three 5-day periods. There was a 2-day washout between each sequence. The three fluids administered during these periods were Le Bleu water, caffeine-free Diet Coke, and Fresca (citrate containing). These two soda preparations were chosen to prevent the known increase in calcium excretion promoted by carbohydrates and caffeine. Twenty-four hour urine specimens were collected on days 4 and 5 of each sequence. The following urinary parameters were measured: Volume, calcium, oxalate, creatinine, uric acid, citrate, sodium, magnesium, phosphorus, sulphate, urea nitrogen, pH, and supersaturation indices. A paired t test was used for statistical analysis. **RESULTS**: Urinary volumes were significantly higher and supersaturation of calcium oxalate significantly lower compared with a self-selected dietary regimen. A decrease in uric acid was also seen in the Fresca cohort. There were no statistically significant differences for any of the urinary parameters. **CONCLUSION**: There is no increased risk or benefit to consuming Fresca or caffeine-free Diet Coke compared with Le Bleu bottled water with respect to stone formation.

**URL:** <https://ovidsp.ovid.com/ovidweb.cgi?T=JS&CSC=Y&NEWS=N&PAGE=fulltext&D=med7&DO=10.1089%2fend.2008.0225> <https://libkey.io/libraries/2592/openurl?genre=article&sid=OVID:medline&id=pmid:19275488&id=doi:10.1089%2Fend.2008.0225&issn=0892-7790&isbn=&volume=23&issue=3&spage=347&pages=347-50&date=2009&title=Journal+of+Endourology&atitle=Effect+of+soda+consumption+on+urinary+stone+risk+parameters.&aulast=Passman&pid=%3Cauthor%3EPassman+CM%3BHolmes+RP%3BKnight+J%3BEaster+L%3BPais+V%3BAssimos+DG%3C%2Fauthor%3E%3CAN%3E19275488%3C%2FAN%3E%3CDT%3EClinical+Trial%3C%2FDT%3E>

164. **Prevalence of hyperoxaluria after bariatric surgery.**

**Item Type:** Journal Article

**Authors:** Patel, Bhavin N.;Passman, Corey M.;Fernandez, Adolfo;Asplin, John R.;Coe, Fredric L.;Kim, Sam C.;Lingeman, James E. and Assimos, Dean G.

**Publication Date:** Ja ,2009

**Journal:** Journal of Urology 181(1), pp. 161–166

**Abstract: PURPOSE**: Recent investigations have shown increased oxalate excretion in patients in whom kidney stones formed after contemporary bariatric surgery. We determined whether there is an increased prevalence of hyperoxaluria after such procedures performed in nonstone formers. **MATERIALS AND METHODS**: A total of 58 nonstone forming adults who underwent laparoscopic Roux-en-Y (52) or a biliopancreatic diversion-duodenal switch procedure (6) collected 24-hour urine specimens 6 months or greater after bariatric surgery. Standard stone risk parameters were assessed. Comparisons were made with a group of healthy nonstone forming adults and stone formers in a commercial database. **RESULTS**: The bariatric group had a significantly higher mean urinary oxalate excretion compared to that in controls and stone formers (67.2 vs 34.1 and 37.0 mg per day, respectively, p : The bariatric group had a significantly higher mean urinary oxalate excretion compared to that in controls and stone formers (67.2 vs 34.1 and 37.0 mg per day, respectively, p : The bariatric group had a significantly higher mean urinary oxalate excretion compared to that in controls and stone formers (67.2 vs 34.1 and 37.0 mg per day, respectively, p **CONCLUSIONS**: There is a high prevalence of hyperoxaluria in patients without a history of kidney stones who undergo bariatric surgery. A significant proportion of these patients have profound hyperoxaluria, which is not uniformly expressed.

**URL:** <https://ovidsp.ovid.com/ovidweb.cgi?T=JS&CSC=Y&NEWS=N&PAGE=fulltext&D=med7&DO=10.1016%2fj.juro.2008.09.028> <https://libkey.io/libraries/2592/openurl?genre=article&sid=OVID:medline&id=pmid:19013593&id=doi:10.1016%2Fj.juro.2008.09.028&issn=0022-5347&isbn=&volume=181&issue=1&spage=161&pages=161-6&date=2009&title=Journal+of+Urology&atitle=Prevalence+of+hyperoxaluria+after+bariatric+surgery.&aulast=Patel&pid=%3Cauthor%3EPatel+BN%3BPassman+CM%3BFernandez+A%3BAsplin+JR%3BCoe+FL%3BKim+SC%3BLingeman+JE%3BAssimos+DG%3C%2Fauthor%3E%3CAN%3E19013593%3C%2FAN%3E%3CDT%3EJournal+Article%3C%2FDT%3E>

165. **Risk factors for renal calcium stone formation in South African and European young adults.**

**Item Type:** Journal Article

**Authors:** Rodgers, Allen;Allie-Hamdulay, Shameez;Pinnock, Diane;Baretta, Gretchen and Trinchieri, Alberto

**Publication Date:** Se ,2009

**Journal:** Archivio Italiano Di Urologia, Andrologia 81(3), pp. 171–174

**Abstract: OBJECTIVES**: The different susceptibility to renal stone disease of white and black people has been previously explained in terms of intrinsic (genetics) and extrinsic (diet, lifestyle) factors. However, in South Africa, the absence of stone disease in the black population has not yet been fully explained by either of these. The aim of the present study was to identify potential differences between black and white subjects in South Africa and white subjects in Europe with respect to their relative dietary and urinary risk factors for renal stone formation. **MATERIALS AND METHODS**: A total of 72 healthy subjects (45 males and 27 females, age range 21-30 years) with no previous history of renal stone disease or specific diseases predisposing to renal stone formation were recruited in South Africa (SA) and in Italy (IT). They were divided in three groups: South African blacks (SA-B), South African whites (SA-W) and Italian whites (IT-W). Each participant provided a 24-hour dietary record and 24-hour urine sample taken over the same period. Nutrients and calories were calculated by means of food composition tables using a computerised procedure. Urinary concentrations of potassium, sodium, calcium, phosphate, oxalate, urate, citrate, magnesium, and creatinine, together with the pH and urinary volumes, were measured. **RESULTS**: The mean carbohydrate intake was significantly higher in SA-B (293+90 g/day) than in both SA-W (194+74, p = 0.002) and IT-W (212 +/- 81; p = 0.000). Daily magnesium intake was higher in SA-B (290+124 mg/day) than in IT-W (176+73 mg/day, p = 0.002). The mean daily urinary excretion of calcium was significantly (p = 0.029) lower in SA-B (3.07 +/- 1.68 mmol/day) with respect to SA-W (4.65 +/- 2.44 mmol/day) and IT-W (4.51 +/- 1.89 mmol/day) whereas mean daily urinary excretion of citrate was significantly (P = 0.012) higher in SA-B (3.36 +/- 1.4 mmol/day) than in SA-W (3.09 +/- 1.45 mmol/day) and IT-W (2.36 +/- 0.98 mmol/day). **CONCLUSION**: Although the carbohydrate intake and the percent of energy from carbohydrate of black subjects in this study were higher with respect to white controls, we were not able to show any other relevant difference of the known dietary stone risk patterns between black and white subjects. On the other hand the urinary patterns of black controls seem to be more favourable in term of risk for stone formation than those of white controls showing a lower calcium excretion and a higher citrate excretion in the urine. Our result of higher carbohydrate intake in black subjects is counter-intuitive as it suggests a higher risk of stone formation in this group. This puzzling result may have arisen because our subjects were recruited from the urban population rather than from rural areas, suggesting that western diets and lifestyles may ultimately change the stone incidence profile in the black population.

**URL:** <https://ovidsp.ovid.com/ovidweb.cgi?T=JS&CSC=Y&NEWS=N&PAGE=fulltext&D=med7&AN=19911680> <https://libkey.io/libraries/2592/openurl?genre=article&sid=OVID:medline&id=pmid:19911680&id=doi:&issn=1124-3562&isbn=&volume=81&issue=3&spage=171&pages=171-4&date=2009&title=Archivio+Italiano+di+Urologia%2C+Andrologia&atitle=Risk+factors+for+renal+calcium+stone+formation+in+South+African+and+European+young+adults.&aulast=Rodgers&pid=%3Cauthor%3ERodgers+A%3BAllie-Hamdulay+S%3BPinnock+D%3BBaretta+G%3BTrinchieri+A%3C%2Fauthor%3E%3CAN%3E19911680%3C%2FAN%3E%3CDT%3EComparative+Study%3C%2FDT%3E>

166. **Changes in urinary stone risk factors in hypocitraturic calcium oxalate stone formers treated with dietary sodium supplementation.**

**Item Type:** Journal Article

**Authors:** Stoller, Marshall L.;Chi, Thomas;Eisner, Brian H.;Shami, Gina and Gentle, Donald L.

**Publication Date:** Ma ,2009

**Journal:** Journal of Urology 181(3), pp. 1140–1144

**Abstract: PURPOSE**: We investigated the effects of supplemental dietary sodium on risk factors for urinary stone disease in stone forming patients with hypocitraturia. **MATERIALS AND METHODS**: Ten patients diagnosed with recurrent isolated hypocitraturic calcium urolithiasis were identified. Baseline 24-hour urinalysis was performed with patients on their regular diet, including citrate replacement with 20 mEq potassium citrate 3 times per day. Strict daily dietary logs were kept for a 7-day period, during which patients had normal oral intake and potassium citrate replacement. Patients then received supplemental sodium chloride for 1 week (1 gm orally 3 times per day), in addition to their regular diets and potassium citrate supplementation. Dietary logs were continued and 24-hour urinalysis was performed at the end of 1 week of supplemental sodium. Risk factors for urinary stone disease were compared using the Student t test and ANOVA. **RESULTS**: Two patients were unable to comply with sodium supplementation based on 24-hour urinalysis and, therefore, they were excluded from study. The remaining 8 patients were analyzed. Patients on supplemental dietary sodium demonstrated significantly increased mean urinary voided volume (933 ml per day above baseline, p : Two patients were unable to comply with sodium supplementation based on 24-hour urinalysis and, therefore, they were excluded from study. The remaining 8 patients were analyzed. Patients on supplemental dietary sodium demonstrated significantly increased mean urinary voided volume (933 ml per day above baseline, p : Two patients were unable to comply with sodium supplementation based on 24-hour urinalysis and, therefore, they were excluded from study. The remaining 8 patients were analyzed. Patients on supplemental dietary sodium demonstrated significantly increased mean urinary voided volume (933 ml per day above baseline, p : Two patients were unable to comply with sodium supplementation based on 24-hour urinalysis and, therefore, they were excluded from study. The remaining 8 patients were analyzed. Patients on supplemental dietary sodium demonstrated significantly increased mean urinary voided volume (933 ml per day above baseline, p **CONCLUSIONS**: Dietary sodium supplementation resulted in an increased voided urine volume and decreased the relative risk supersaturation ratio for calcium oxalate stones in patients with a history of hypocitraturic calcium oxalate nephrolithiasis. Urinary calcium excretion as well as other urine parameters that are risk factors for nephrolithiasis was not changed. Sodium restriction may be inappropriate in patients with hypocitraturia and recurrent urinary stones. Sodium supplementation may be beneficial in these patients because it results in voluntary increased fluid intake.

**URL:** <https://ovidsp.ovid.com/ovidweb.cgi?T=JS&CSC=Y&NEWS=N&PAGE=fulltext&D=med7&DO=10.1016%2fj.juro.2008.11.020> <https://libkey.io/libraries/2592/openurl?genre=article&sid=OVID:medline&id=pmid:19152919&id=doi:10.1016%2Fj.juro.2008.11.020&issn=0022-5347&isbn=&volume=181&issue=3&spage=1140&pages=1140-4&date=2009&title=Journal+of+Urology&atitle=Changes+in+urinary+stone+risk+factors+in+hypocitraturic+calcium+oxalate+stone+formers+treated+with+dietary+sodium+supplementation.&aulast=Stoller&pid=%3Cauthor%3EStoller+ML%3BChi+T%3BEisner+BH%3BShami+G%3BGentle+DL%3C%2Fauthor%3E%3CAN%3E19152919%3C%2FAN%3E%3CDT%3EJournal+Article%3C%2FDT%3E>

167. **Dissolution of radiolucent renal stones by oral alkalinization with potassium citrate/potassium bicarbonate.**

**Item Type:** Journal Article

**Authors:** Trinchieri, Alberto;Esposito, Nicola and Castelnuovo, Chiara

**Publication Date:** Se ,2009

**Journal:** Archivio Italiano Di Urologia, Andrologia 81(3), pp. 188–191

**Abstract: AIM**: Uric acid stone disease is dependent on three pathogenetic factors: acid urine pH, low urine volume, and hyperuricosuria. However, the most important factor for uric acid stone formation is persistently acidic urine that represents a prerequisite for uric acid stone formation and growth. Urinary alkalization with alkali administration has been advocated for dissolution of stones on the basis of estabilished clinical experience. The aim of this study was to evaluate the clinical efficacy of therapy with potassium citrate/potassium bicarbonate for dissolution of radiolucent stones. **PATIENTS AND METHODS**: A total of 8 patients with radiolucent stones (: A total of 8 patients with radiolucent stones (**RESULTS**: During the first period of treatment stone burden remained unchanged in all patients. On the contrary after 6 weeks of potassium citrate/bicarbonate treatment, complete stone dissolution was found in three of the patients. In the other five cases a partial dissolution was observed and in two of them complete dissolution of the stone was achieved after prolongation of the treatment for 4 and 6 month respectively. Mean urinary volumes were unchanged during all the two study periods. Mean urinary pH was significantly higher during the potassium citrate/bicarbonate treatment period in comparison to the first study period (morning 6.60 +/- 1.06 vs 5.53 +/- 0.51, p = 0.030; afternoon 6.53 +/- 0.70 vs 5.63 +/- 0.41, p = 0.007; night 6.57 +/- 0.51 vs 5.98 +/- 0.80, p = 0.092). Tolerance of the drug was good, and no serious effects were observed sufficient to interrupt treatment. None of the patients required subsequent interventions for stone treatment. **CONCLUSION**: Urinary alkalization with potassium citrate/bicarbonate is a well tolerated and highly effective treatment, resulting in dissolution of nonobstructing uric acid stones.

**URL:** <https://ovidsp.ovid.com/ovidweb.cgi?T=JS&CSC=Y&NEWS=N&PAGE=fulltext&D=med7&AN=19911683> <https://libkey.io/libraries/2592/openurl?genre=article&sid=OVID:medline&id=pmid:19911683&id=doi:&issn=1124-3562&isbn=&volume=81&issue=3&spage=188&pages=188-91&date=2009&title=Archivio+Italiano+di+Urologia%2C+Andrologia&atitle=Dissolution+of+radiolucent+renal+stones+by+oral+alkalinization+with+potassium+citrate%2Fpotassium+bicarbonate.&aulast=Trinchieri&pid=%3Cauthor%3ETrinchieri+A%3BEsposito+N%3BCastelnuovo+C%3C%2Fauthor%3E%3CAN%3E19911683%3C%2FAN%3E%3CDT%3EJournal+Article%3C%2FDT%3E>

168. **Effect of potassium citrate therapy on the risk of renal stone formation during spaceflight.**

**Item Type:** Journal Article

**Authors:** Whitson, Peggy A.;Pietrzyk, Robert A.;Jones, Jeffrey A.;Nelman-Gonzalez, Mayra;Hudson, Edgar K. and Sams, Clarence F.

**Publication Date:** No ,2009

**Journal:** Journal of Urology 182(5), pp. 2490–2496

**Abstract: PURPOSE**: Exposure to microgravity affects human physiology and results in changes in urinary chemical composition during and after spaceflight, favoring an increased risk of renal stones. We assessed the efficacy of potassium citrate to decrease the stone risk during and after spaceflight. **MATERIALS AND METHODS**: The study was done in 30 long duration spaceflight crew members to the space stations Mir and International Space Station. Before, during and after spaceflight 24-hour urine samples were collected to assess the renal stone risk. Potassium citrate (20 mEq) was ingested daily by International Space Station crew members in a double-blind, placebo controlled study. Mir crew members performed the identical protocol but did not ingest medication. **RESULTS**: Potassium citrate treated crew members had decreased urinary calcium excretion and maintained the calcium oxalate supersaturation risk at preflight levels compared to that in controls. Increased urinary pH in the treatment group decreased the risk of uric acid stones. **CONCLUSIONS**: Results from this investigation suggest that supplementation with potassium citrate may decrease the risk of renal stone formation during and immediately after spaceflight.

**URL:** <https://ovidsp.ovid.com/ovidweb.cgi?T=JS&CSC=Y&NEWS=N&PAGE=fulltext&D=med7&DO=10.1016%2fj.juro.2009.07.010> <https://libkey.io/libraries/2592/openurl?genre=article&sid=OVID:medline&id=pmid:19765769&id=doi:10.1016%2Fj.juro.2009.07.010&issn=0022-5347&isbn=&volume=182&issue=5&spage=2490&pages=2490-6&date=2009&title=Journal+of+Urology&atitle=Effect+of+potassium+citrate+therapy+on+the+risk+of+renal+stone+formation+during+spaceflight.&aulast=Whitson&pid=%3Cauthor%3EWhitson+PA%3BPietrzyk+RA%3BJones+JA%3BNelman-Gonzalez+M%3BHudson+EK%3BSams+CF%3C%2Fauthor%3E%3CAN%3E19765769%3C%2FAN%3E%3CDT%3EJournal+Article%3C%2FDT%3E>

169. **New methods of assessing crystal growth and saturation of brushite in whole urine: effect of pH, calcium and citrate.**

**Item Type:** Journal Article

**Authors:** Pak, Charles Y. C.;Rodgers, Kathy;Poindexter, John R. and Sakhaee, Khashayar

**Publication Date:** Oc ,2008

**Journal:** Journal of Urology 180(4), pp. 1532–1537

**Abstract: PURPOSE**: Brushite crystallization might be important in stone formation and prevention. To explore this question new methods for the saturation and crystal growth of brushite were devised that are applicable to whole urine without any computer program. **MATERIALS AND METHODS**: The saturation value (concentration-to-product ratio) was determined by dividing the molar concentration product of Ca ([Ca]) and phosphate ([P]), that is [Ca] x [P], of original urine by the steady state solubility obtained after incubating with an excess of brushite (10 mg/ml) for 5 hours. Crystal growth was measured from the depletion of filtrate ([Ca] x [P]) 3 hours after seeding with brushite (0.25 mg/ml). To test the effect of pH, Ca and citrate the saturation value and crystal growth were determined in 24-hour urine samples from 4 normal volunteers and 2 stone formers, and modified artificially to produce 4 ranges of pH, Ca and citrate by adding acid, base, Ca or citrate. **RESULTS**: The saturation value and crystal growth of brushite increased with an increase in pH or the Ca concentration but they decreased when the citrate concentration increased. The saturation value correlated strongly with crystal growth. **CONCLUSIONS**: The new methods of brushite saturation value and crystal growth should help discern how abnormalities in urinary pH, Ca and citrate interact to influence the formation of Ca stones in cases of distal renal tubular acidosis and alkali therapy.

**URL:** <https://ovidsp.ovid.com/ovidweb.cgi?T=JS&CSC=Y&NEWS=N&PAGE=fulltext&D=med7&DO=10.1016%2fj.juro.2008.06.011> <https://libkey.io/libraries/2592/openurl?genre=article&sid=OVID:medline&id=pmid:18710766&id=doi:10.1016%2Fj.juro.2008.06.011&issn=0022-5347&isbn=&volume=180&issue=4&spage=1532&pages=1532-7&date=2008&title=Journal+of+Urology&atitle=New+methods+of+assessing+crystal+growth+and+saturation+of+brushite+in+whole+urine%3A+effect+of+pH%2C+calcium+and+citrate.&aulast=Pak&pid=%3Cauthor%3EPak+CY%3BRodgers+K%3BPoindexter+JR%3BSakhaee+K%3C%2Fauthor%3E%3CAN%3E18710766%3C%2FAN%3E%3CDT%3EComparative+Study%3C%2FDT%3E>

170. **Comparison between lemonade and potassium citrate and impact on urine pH and 24-hour urine parameters in patients with kidney stone formation.**

**Item Type:** Journal Article

**Authors:** Koff, Stacey G.;Paquette, Edmond L.;Cullen, Jenny;Gancarczyk, Kevin K.;Tucciarone, Paulette R. and Schenkman, Noah S.

**Publication Date:** Ju ,2007

**Journal:** Urology 69(6), pp. 1013–1016

**Abstract: OBJECTIVES**: The past few decades have been notable for advances in minimally invasive techniques to treat kidney stones. Despite successful surgical management of calculi, the impact of diet on stone disease prevention remains an important factor. Dietary modifications can be applied to prevent stone recurrence in an economical manner that is acceptable to patients. Lemonade has been studied in the past and found to increase urinary citrate levels in small groups of patients. The present study examined the effect of lemonade intake on a change in urinary pH and improvement of urinary stone risk factors compared with potassium citrate treatment. **METHODS**: A total of 21 stone-forming patients were treated with lemonade therapy and potassium citrate supplementation in a prospective cross-over trial. we performed 24-hour urine collections at baseline and with each treatment arm. the results between the two groups were examined to determine whether differences existed in the urinary stone risk factors. **RESULTS**: Potassium citrate, but not lemonade, improved citrate levels and urinary ph to a significant degree. patients taking potassium citrate had a significantly decreased urine volume compared with their urine volume drinking lemonade. neither lemonade nor potassium citrate affected the uric acid levels to a significant degree. **CONCLUSIONS**: The results of our study have shown that lemonade did not provide improvements in urinary citrate or pH levels but did assist patients in maintaining urine output compared with potassium citrate therapy.

**URL:** <https://ovidsp.ovid.com/ovidweb.cgi?T=JS&CSC=Y&NEWS=N&PAGE=fulltext&D=med6&DO=10.1016%2fj.urology.2007.02.008> <https://libkey.io/libraries/2592/openurl?genre=article&sid=OVID:medline&id=pmid:17572176&id=doi:10.1016%2Fj.urology.2007.02.008&issn=0090-4295&isbn=&volume=69&issue=6&spage=1013&pages=1013-6&date=2007&title=Urology&atitle=Comparison+between+lemonade+and+potassium+citrate+and+impact+on+urine+pH+and+24-hour+urine+parameters+in+patients+with+kidney+stone+formation.&aulast=Koff&pid=%3Cauthor%3EKoff+SG%3BPaquette+EL%3BCullen+J%3BGancarczyk+KK%3BTucciarone+PR%3BSchenkman+NS%3C%2Fauthor%3E%3CAN%3E17572176%3C%2FAN%3E%3CDT%3EClinical+Trial%3C%2FDT%3E>

171. **Update on nephrolithiasis management**

**Item Type:** Journal Article

**Authors:** Long, L. O. and Park, S.

**Publication Date:** Se ,2007

**Journal:** Minerva Urologica E Nefrologica 59(3), pp. 317–325

**Abstract:** Urolithiasis affects 10%-15% of the population in their lifetime. After spontaneous passage or surgical treatment, a subset of these patients will have recurrent calculi. These recurrent stone events are significantly morbid and can potentially lead to serious chronic renal disease, thus prevention is a very important treatment goal. Fortunately, a reversible metabolic abnormality can be identified in over 90% of recurrent stone formers. Thus, a detailed metabolic evaluation using 24 hour urine collections and serum tests is indicated in patients at high risk for stone recurrence. Once the patient's underlying urinary physicochemical and physiologic derangements are defined, targeted medical therapy can be initiated in order to prevent growth of pre-existing stones and recurrent stone formation. In this paper, we provide a review of the currently available selective and nonselective pharmacologic treatments for urolithiasis. Furthermore, we discuss a number of investigational agents for kidney stone prevention. Although many of these agents are effective, there remain numerous clinical scenarios where currently available therapies are inadequate. [References: 59]

**URL:** <https://ovidsp.ovid.com/ovidweb.cgi?T=JS&CSC=Y&NEWS=N&PAGE=fulltext&D=med6&AN=17912227> <https://libkey.io/libraries/2592/openurl?genre=article&sid=OVID:medline&id=pmid:17912227&id=doi:&issn=0393-2249&isbn=&volume=59&issue=3&spage=317&pages=317-25&date=2007&title=Minerva+Urologica+e+Nefrologica&atitle=Update+on+nephrolithiasis+management.&aulast=Long&pid=%3Cauthor%3ELong+LO%3BPark+S%3C%2Fauthor%3E%3CAN%3E17912227%3C%2FAN%3E%3CDT%3EJournal+Article%3C%2FDT%3E>

172. **Urolithiasis in pediatric patients: a single center study of incidence, clinical presentation and outcome.**

**Item Type:** Journal Article

**Authors:** VanDervoort, Kristy;Wiesen, Jonathan;Frank, Rachel;Vento, Suzanne;Crosby, Virginia;Chandra, Manju and Trachtman, Howard

**Publication Date:** Ju ,2007

**Journal:** Journal of Urology 177(6), pp. 2300–2305

**Abstract: PURPOSE**: The incidence of kidney stones in adults has increased in the last 30 years. This retrospective, single site review was done to test the hypotheses that the incidence of urolithiasis in pediatric patients increased from 1994 to 2005, and that metabolic abnormalities were more common in patients with renal stones in the final 3 years of the study period. **MATERIALS AND METHODS**: Charts from 2 time periods were reviewed, 1994 to 1996 (period 1) and 2003 to 2005 (period 2). Clinical and laboratory data, including demographics, presenting complaints, laboratory assessment, treatment and outcome, were tabulated in patients with confirmed urolithiasis. **RESULTS**: The number of patients with urolithiasis increased from 7 in period 1 to 61 in period 2. When expressed as cases per 100 new patients the incidence increased 4.6 times (p = 0.014). Focusing on period 2, 28% of patients were younger than 10 years. While blood tests were generally normal, 76% of patients had at least 1 abnormality in the 24-hour urine collection. Hypocitraturia, which was the most common metabolic abnormality, was noted in 52% of patients. The small number of patients in period 1 precluded determination as to whether metabolic abnormalities were more common in period 2. Surgery and/or lithotripsy was required in 12 children. Stone disease recurred in 39% of the patients. **CONCLUSIONS**: The incidence of urolithiasis in the pediatric population increased nearly 5-fold at our institution during the last decade. We recommend that the primary diagnostic test be a 24-hour urine collection. The most common metabolic abnormality was hypocitraturia, followed by hypercalciuria. Recurrence of stones is common (approximately 40% rate) and followup is advised.

**URL:** <https://ovidsp.ovid.com/ovidweb.cgi?T=JS&CSC=Y&NEWS=N&PAGE=fulltext&D=med6&DO=10.1016%2fj.juro.2007.02.002> <https://libkey.io/libraries/2592/openurl?genre=article&sid=OVID:medline&id=pmid:17509344&id=doi:10.1016%2Fj.juro.2007.02.002&issn=0022-5347&isbn=&volume=177&issue=6&spage=2300&pages=2300-5&date=2007&title=Journal+of+Urology&atitle=Urolithiasis+in+pediatric+patients%3A+a+single+center+study+of+incidence%2C+clinical+presentation+and+outcome.&aulast=VanDervoort&pid=%3Cauthor%3EVanDervoort+K%3BWiesen+J%3BFrank+R%3BVento+S%3BCrosby+V%3BChandra+M%3BTrachtman+H%3C%2Fauthor%3E%3CAN%3E17509344%3C%2FAN%3E%3CDT%3EJournal+Article%3C%2FDT%3E>

173. **Reduction of renal stone risk by potassium-magnesium citrate during 5 weeks of bed rest.**

**Item Type:** Journal Article

**Authors:** Zerwekh, Joseph E.;Odvina, Clarita V.;Wuermser, Lisa-Ann and Pak, Charles Y. C.

**Publication Date:** Ju ,2007

**Journal:** Journal of Urology 177(6), pp. 2179–2184

**Abstract: PURPOSE**: Exposure to the microgravity environment of space increases the risk of kidney stone formation, particularly for calcium oxalate and uric acid stones. This study was performed to evaluate the efficacy of potassium alkali as potassium-magnesium citrate in reducing renal stone risk and bone turnover. **MATERIALS AND METHODS**: This study was performed as a double-blind, placebo controlled trial. We studied 20 normocalciuric subjects randomized to either placebo or potassium-magnesium citrate (42 mEq potassium, 21 mEq magnesium, 63 mEq citrate per day) before and during 5 weeks of strict bed rest. The study was performed in the General Clinical Research Center and under a controlled dietary regimen composed of 100 mEq of sodium, 800 mg of calcium, 0.8 gm/kg animal protein and 2,200 kcal per day. Two 24-hour urine collections were obtained under oil each week for assessment of stone risk parameters and relative saturation of calcium oxalate, brushite and undissociated uric acid. Blood was also collected for determination of serum immunoreactive parathyroid hormone and vitamin D metabolites. **RESULTS**: Bed rest promoted a rapid increase in urinary calcium excretion of approximately 50 mg per day in both groups. Despite this increase subjects treated with potassium-magnesium citrate demonstrated significant decreases in the relative saturation of calcium oxalate and in the concentration of undissociated uric acid compared to placebo. Immunoreactive parathyroid hormone, serum 1,25-dihydroxyvitamin D and intestinal calcium absorption all decreased in both groups with no difference in response between the 2 treatment arms. **CONCLUSIONS**: Provision of alkali as potassium-magnesium citrate is an effective countermeasure for the increased risk of renal stone disease associated with immobilization. Despite an increase in urine calcium concentration, the relative saturation of calcium oxalate decreased due to citrate chelation of calcium and the concentration of undissociated uric acid decreased due to the significant increase in urine pH.

**URL:** <https://ovidsp.ovid.com/ovidweb.cgi?T=JS&CSC=Y&NEWS=N&PAGE=fulltext&D=med6&DO=10.1016%2fj.juro.2007.01.156> <https://libkey.io/libraries/2592/openurl?genre=article&sid=OVID:medline&id=pmid:17509313&id=doi:10.1016%2Fj.juro.2007.01.156&issn=0022-5347&isbn=&volume=177&issue=6&spage=2179&pages=2179-84&date=2007&title=Journal+of+Urology&atitle=Reduction+of+renal+stone+risk+by+potassium-magnesium+citrate+during+5+weeks+of+bed+rest.&aulast=Zerwekh&pid=%3Cauthor%3EZerwekh+JE%3BOdvina+CV%3BWuermser+LA%3BPak+CY%3C%2Fauthor%3E%3CAN%3E17509313%3C%2FAN%3E%3CDT%3EJournal+Article%3C%2FDT%3E>

174. **Urine composition in type 2 diabetes: predisposition to uric acid nephrolithiasis.**

**Item Type:** Journal Article

**Authors:** Cameron, Mary Ann;Maalouf, Naim M.;Adams-Huet, Beverley;Moe, Orson W. and Sakhaee, Khashayar

**Publication Date:** Ma ,2006

**Journal:** Journal of the American Society of Nephrology 17(5), pp. 1422–1428

**Abstract:** Type 2 diabetes is a risk factor for nephrolithiasis in general and has been associated with uric acid stones in particular. The purpose of this study was to identify the metabolic features that place patients with type 2 diabetes at increased risk for uric acid nephrolithiasis. Three groups of individuals were recruited for this outpatient study: patients who have type 2 diabetes and are not stone formers (n = 24), patients who do not have diabetes and are uric acid stone formers (UASF; n = 8), and normal volunteers (NV; n = 59). Participants provided a fasting blood sample and a single 24-h urine collection for stone risk analysis. Twenty-four-hour urine volume and total uric acid did not differ among the three groups. Patients with type 2 diabetes and UASF had lower 24-h urine pH than NV. Urine pH inversely correlated with both body weight and 24-h urine sulphate in all groups. Urine pH remained significantly lower in patients with type 2 diabetes and UASF than NV after adjustment for weight and urine sulphate (P < 0.01). For a given urine sulphate, urine net acid excretion tended to be higher in patients with type 2 diabetes versus NV. With increasing urine sulphate, NV and patients with type 2 diabetes had a similar rise in urine ammonium, whereas in UASF, ammonium excretion remained unchanged. The main risk factor for uric acid nephrolithiasis in patients with type 2 diabetes is a low urine pH. Higher body mass and increased acid intake can contribute to but cannot entirely account for the lower urine pH in patients with type 2 diabetes.

**URL:** <https://ovidsp.ovid.com/ovidweb.cgi?T=JS&CSC=Y&NEWS=N&PAGE=fulltext&D=med6&DO=10.1681%2fASN.2005121246> <https://libkey.io/libraries/2592/openurl?genre=article&sid=OVID:medline&id=pmid:16597681&id=doi:10.1681%2FASN.2005121246&issn=1046-6673&isbn=&volume=17&issue=5&spage=1422&pages=1422-8&date=2006&title=Journal+of+the+American+Society+of+Nephrology&atitle=Urine+composition+in+type+2+diabetes%3A+predisposition+to+uric+acid+nephrolithiasis.&aulast=Cameron&pid=%3Cauthor%3ECameron+MA%3BMaalouf+NM%3BAdams-Huet+B%3BMoe+OW%3BSakhaee+K%3C%2Fauthor%3E%3CAN%3E16597681%3C%2FAN%3E%3CDT%3EJournal+Article%3C%2FDT%3E>

175. **Causes of hypocitraturia in recurrent calcium stone formers: focusing on urinary potassium excretion.**

**Item Type:** Journal Article

**Authors:** Domrongkitchaiporn, Somnuek;Stitchantrakul, Wasana and Kochakarn, Wachira

**Publication Date:** Oc ,2006

**Journal:** American Journal of Kidney Diseases 48(4), pp. 546–554

**Abstract: BACKGROUND**: Multiple factors associated with hypocitraturia have been identified. However, limited studies addressing the causal relationship to hypocitraturia are available. We therefore conducted this study to determine factors associated with hypocitraturia and show their causal relationship in recurrent calcium stone formers. **METHODS**: Dietary review and 24-hour urine samples were obtained from all recurrent calcium stone formers referred for metabolic workup in the stone clinic. One month of oral potassium chloride supplementation was prescribed to stone formers to determine the causal relationship between urinary potassium and citrate levels. **RESULTS**: Eighty-three subjects, 44 men and 39 women, were recruited to participate in this study. Hypocitraturia (citrate : Eighty-three subjects, 44 men and 39 women, were recruited to participate in this study. Hypocitraturia (citrate : Eighty-three subjects, 44 men and 39 women, were recruited to participate in this study. Hypocitraturia (citrate 0.05). **CONCLUSION**: Hypocitraturia was found to be a common risk factor associated with recurrent calcium stone formation and low urinary potassium level, low alkaline absorption, low urinary calcium level, and high titratable acid excretion. Hypocitraturia is predominantly of dietary origin. Estimation of fruit intake should be included in the metabolic evaluation for recurrent calcium stone formation.

**URL:** <https://ovidsp.ovid.com/ovidweb.cgi?T=JS&CSC=Y&NEWS=N&PAGE=fulltext&D=med6&DO=10.1053%2fj.ajkd.2006.06.008> <https://libkey.io/libraries/2592/openurl?genre=article&sid=OVID:medline&id=pmid:16997050&id=doi:10.1053%2Fj.ajkd.2006.06.008&issn=0272-6386&isbn=&volume=48&issue=4&spage=546&pages=546-54&date=2006&title=American+Journal+of+Kidney+Diseases&atitle=Causes+of+hypocitraturia+in+recurrent+calcium+stone+formers%3A+focusing+on+urinary+potassium+excretion.&aulast=Domrongkitchaiporn&pid=%3Cauthor%3EDomrongkitchaiporn+S%3BStitchantrakul+W%3BKochakarn+W%3C%2Fauthor%3E%3CAN%3E16997050%3C%2FAN%3E%3CDT%3EEvaluation+Study%3C%2FDT%3E>

176. **Changes in urinary compositions among children after consuming prepared oral doses of aloe (Aloe vera Linn).**

**Item Type:** Journal Article

**Authors:** Kirdpon, Sukachart;Kirdpon, Wichit;Airarat, Wanchai;Thepsuthammarat, Kaewjai and Nanakorn, Somsong

**Publication Date:** Au ,2006

**Journal:** Journal of the Medical Association of Thailand 89(8), pp. 1199–1205

**Abstract: OBJECTIVES**: 1) To investigate the amount of citrate and tartrate in aloe gel, and in the urine of healthy normal children, before and after consuming fresh aloe gel. 2) To evaluate the changes in the chemical composition of urine among subjects after taking aloe gel. 3) To determine the value of consuming aloe gel for prevention of renal stone formation. **DESIGN**: Experimental study. **MATERIAL AND METHOD**: Thirteen healthy boys between 9 and 13 years of age were enrolled (with informed permission) in the clinical trial. Subjects ingested 100 g of fresh prepared aloe gel twice a day for seven consecutive days. The 24-hour urine was collected one day prior to taking the gel (Day 0), Days 2 and 5 of consumption, and Day 8 (one day after completion). The authors determined the urine volume, osmolality, potassium, sodium, phosphate, calcium, magnesium, uric acid, citrate, tartrate, oxalate, Permissible Increment in Calcium (PI Ca), Permissible Increment in Oxalate (PI Ox), Concentration Product Ratio of Calcium Phosphate (CPR CaPO4) and the citrate per creatinine ratio. **RESULTS**: The citrate and tartrate concentration in 100 g of fresh aloe gel was 96.3 and 158.9 mg, respectively. The 24-hr urine volume and urinary citrate excretion were significantly increased (p : The citrate and tartrate concentration in 100 g of fresh aloe gel was 96.3 and 158.9 mg, respectively. The 24-hr urine volume and urinary citrate excretion were significantly increased (p : The citrate and tartrate concentration in 100 g of fresh aloe gel was 96.3 and 158.9 mg, respectively. The 24-hr urine volume and urinary citrate excretion were significantly increased (p **CONCLUSION**: One hundred grams of fresh Aloe vera gel contains 96.3 milligrams of citrate and 158.9 milligrams of tartrate and were in the mid-range among Thai fruits. Changes in chemical compositions of urine after aloe gel consumption shows potential for preventing kidney stone formation among children.

**URL:** <https://ovidsp.ovid.com/ovidweb.cgi?T=JS&CSC=Y&NEWS=N&PAGE=fulltext&D=med6&AN=17048430> <https://libkey.io/libraries/2592/openurl?genre=article&sid=OVID:medline&id=pmid:17048430&id=doi:&issn=0125-2208&isbn=&volume=89&issue=8&spage=1199&pages=1199-205&date=2006&title=Journal+of+the+Medical+Association+of+Thailand&atitle=Changes+in+urinary+compositions+among+children+after+consuming+prepared+oral+doses+of+aloe+%28Aloe+vera+Linn%29.&aulast=Kirdpon&pid=%3Cauthor%3EKirdpon+S%3BKirdpon+W%3BAirarat+W%3BThepsuthammarat+K%3BNanakorn+S%3C%2Fauthor%3E%3CAN%3E17048430%3C%2FAN%3E%3CDT%3EClinical+Trial%3C%2FDT%3E>

177. **Renal tubular impairment in children with idiopathic hypercalciuria.**

**Item Type:** Journal Article

**Authors:** Skalova, Sylva and Kutilek, Stepan

**Publication Date:** 2006

**Journal:** Acta Medica (Hradec Kralove) 49(2), pp. 109–111

**Abstract:** Idiopathic hypercalciuria (IH) is defined as hypercalciuria that persists after correction of dietary inbalances and has no detectable cause. The excretion of urinary N-acetyl-beta-D-glucosaminidase (U-NAG), a marker of proximal tubular damage, has been previously reported as either increased or normal in children with IH. We evaluated U-NAG in 20 children (13 boys and 7 girls, mean age 10.3 years +/- 5.7 SD) with IH (urinary calcium excretion above 0.1 mmol/kg/24 hours, with no detectable cause) and with otherwise normal renal function tests. Ultrasound examination revealed urolithiasis (n=4) and nephrocalcinosis (n=1). The U-NAG values were evaluated in the spot urine collected from the second morning void and calculated as the urinary NAG/creatinine ratio (U-NAG/Cr) and expressed in nkat/mmol. The 24-hour urinary calcium excretion (U-Ca/24h) was assessed in a urinary sample from 24-hour collected urine and calculated in mmol/kg. The obtained results of U-Ca/24h and U-NAG/Cr were expressed as Z-scores. When compared to the reference data, the U-Ca/24h and U-NAG/Cr were significantly higher (p = 0.0004 and p = 0.006, respectively). There was no correlation between the U-NAG/Cr and U-Ca/24h (r = 0.18, p = 0.20). The U-NAG/Cr values were significantly higher in the 5 patients with urolithiasis/nephrocalcinosis, whether compared to the rest of the group (p = 0.02), or to the reference data (p = 0.01). The U-NAG/Cr activity was higher in 15 children without urolithiasis/nephrocalcinosis when compared to reference data (p < 0.01). There was no difference in U-Ca/24h between the children with and without urolithiasis/nephrocalcinosis (p = 0.58). These findings suggest that tubular impairment, as reflected by U-NAG/Cr, might occur in children with IH, especially in patients with urolithiasis/nephrocalcinosis. There doesn't seem to be a direct relationship between the U-NAG/Cr activity and the degree of calcium leakage.

**URL:** <https://ovidsp.ovid.com/ovidweb.cgi?T=JS&CSC=Y&NEWS=N&PAGE=fulltext&D=med6&AN=16956119> <https://libkey.io/libraries/2592/openurl?genre=article&sid=OVID:medline&id=pmid:16956119&id=doi:&issn=1211-4286&isbn=&volume=49&issue=2&spage=109&pages=109-11&date=2006&title=Acta+Medica+%28Hradec+Kralove%29&atitle=Renal+tubular+impairment+in+children+with+idiopathic+hypercalciuria.&aulast=Skalova&pid=%3Cauthor%3ESkalova+S%3BKutilek+S%3C%2Fauthor%3E%3CAN%3E16956119%3C%2FAN%3E%3CDT%3EJournal+Article%3C%2FDT%3E>

178. **Body size and 24-hour urine composition.**

**Item Type:** Journal Article

**Authors:** Taylor, Eric N. and Curhan, Gary C.

**Publication Date:** De ,2006

**Journal:** American Journal of Kidney Diseases 48(6), pp. 905–915

**Abstract: BACKGROUND**: Greater body mass index (BMI) is a risk factor for kidney stones. However, the relation between BMI and the urinary excretion of many lithogenic factors remains unclear. **METHODS**: We studied urine pH, urine volume, and 24-hour urinary excretion of calcium, oxalate, citrate, uric acid, sodium, magnesium, potassium, phosphate, and creatinine in stone-forming and non-stone-forming participants in the Health Professionals Follow-Up Study (599 stone-forming and 404 non-stone-forming men), Nurses' Health Study (888 stone-forming and 398 non-stone-forming older women), and Nurses' Health Study II (689 stone-forming and 295 non-stone-forming younger women). Each cohort was divided into quintiles of BMI. Tests of linear trend were conducted by 1-way analysis of variance. Linear regression models were adjusted for age, history of stone disease, dietary intake, and urinary factors. **RESULTS**: Participants with greater BMIs excreted more urinary oxalate (P for trend : Participants with greater BMIs excreted more urinary oxalate (P for trend : Participants with greater BMIs excreted more urinary oxalate (P for trend : Participants with greater BMIs excreted more urinary oxalate (P for trend : Participants with greater BMIs excreted more urinary oxalate (P for trend : Participants with greater BMIs excreted more urinary oxalate (P for trend : Participants with greater BMIs excreted more urinary oxalate (P for trend : Participants with greater BMIs excreted more urinary oxalate (P for trend **CONCLUSION**: Positive associations between BMI and urinary calcium excretion likely are due to differences in animal protein and sodium intake. The greater incidence of kidney stones in the obese may be due to an increase in uric acid nephrolithiasis.

**URL:** <https://ovidsp.ovid.com/ovidweb.cgi?T=JS&CSC=Y&NEWS=N&PAGE=fulltext&D=med6&DO=10.1053%2fj.ajkd.2006.09.004> <https://libkey.io/libraries/2592/openurl?genre=article&sid=OVID:medline&id=pmid:17162145&id=doi:10.1053%2Fj.ajkd.2006.09.004&issn=0272-6386&isbn=&volume=48&issue=6&spage=905&pages=905-15&date=2006&title=American+Journal+of+Kidney+Diseases&atitle=Body+size+and+24-hour+urine+composition.&aulast=Taylor&pid=%3Cauthor%3ETaylor+EN%3BCurhan+GC%3C%2Fauthor%3E%3CAN%3E17162145%3C%2FAN%3E%3CDT%3EJournal+Article%3C%2FDT%3E>

179. **Association of prevalent hypertension with 24-hour urinary excretion of calcium, citrate, and other factors.**

**Item Type:** Journal Article

**Authors:** Taylor, Eric N.;Mount, David B.;Forman, John P. and Curhan, Gary C.

**Publication Date:** Ma ,2006

**Journal:** American Journal of Kidney Diseases 47(5), pp. 780–789

**Abstract: BACKGROUND**: The relation between hypertension and the urinary excretion of calcium, citrate, and other factors is unclear. It has been proposed that increased urinary calcium excretion is a central feature of essential hypertension. Metabolic acidosis also may be associated with hypertension and decreases urinary citrate levels. **METHODS**: To compare the urine composition of individuals with and without hypertension, we studied 24-hour urinary excretion of calcium, citrate, oxalate, uric acid, sodium, magnesium, potassium, phosphorus, and creatinine and pH in a subset of participants with and without nephrolithiasis in the Nurses' Health Study I (older women; N = 1,284), Nurses' Health Study II (younger women; N = 952), and the Health Professionals Follow-up Study (men; N = 788). Logistic regression models adjusted for age, weight, dietary intake, and urinary factors. **RESULTS**: In participants with and without nephrolithiasis, citrate was the only urinary factor consistently related to hypertension. Compared with those in the lowest quartile of urinary citrate excretion, multivariate odds ratios of prevalent hypertension in the highest quartile were 0.37 (95% confidence interval [CI], 0.24 to 0.55; P trend : In participants with and without nephrolithiasis, citrate was the only urinary factor consistently related to hypertension. Compared with those in the lowest quartile of urinary citrate excretion, multivariate odds ratios of prevalent hypertension in the highest quartile were 0.37 (95% confidence interval [CI], 0.24 to 0.55; P trend : In participants with and without nephrolithiasis, citrate was the only urinary factor consistently related to hypertension. Compared with those in the lowest quartile of urinary citrate excretion, multivariate odds ratios of prevalent hypertension in the highest quartile were 0.37 (95% confidence interval [CI], 0.24 to 0.55; P trend **CONCLUSION**: Lower urinary citrate excretion is associated independently with prevalent hypertension. Factors that regulate urinary citrate excretion may play a role in hypertension.

**URL:** <https://ovidsp.ovid.com/ovidweb.cgi?T=JS&CSC=Y&NEWS=N&PAGE=fulltext&D=med6&DO=10.1053%2fj.ajkd.2006.01.024> <https://libkey.io/libraries/2592/openurl?genre=article&sid=OVID:medline&id=pmid:16632016&id=doi:10.1053%2Fj.ajkd.2006.01.024&issn=0272-6386&isbn=&volume=47&issue=5&spage=780&pages=780-9&date=2006&title=American+Journal+of+Kidney+Diseases&atitle=Association+of+prevalent+hypertension+with+24-hour+urinary+excretion+of+calcium%2C+citrate%2C+and+other+factors.&aulast=Taylor&pid=%3Cauthor%3ETaylor+EN%3BMount+DB%3BForman+JP%3BCurhan+GC%3C%2Fauthor%3E%3CAN%3E16632016%3C%2FAN%3E%3CDT%3EJournal+Article%3C%2FDT%3E>

180. **Biochemical and stone-risk profiles with topiramate treatment.**

**Item Type:** Journal Article

**Authors:** Welch, Brian J.;Graybeal, Dion;Moe, Orson W.;Maalouf, Naim M. and Sakhaee, Khashayar

**Publication Date:** Oc ,2006

**Journal:** American Journal of Kidney Diseases 48(4), pp. 555–563

**Abstract: BACKGROUND**: Topiramate is a novel neuromodulatory agent commonly prescribed for the treatment of seizure disorders and for migraine headache prophylaxis. Calcium phosphate kidney stones have been observed with topiramate treatment, but a comprehensive elucidation of stone-risk profile was not reported previously. This study explores the relationship between topiramate treatment and propensity for kidney stone formation. **METHODS**: Thirty-two topiramate-treated subjects and 50 healthy volunteers participated in a cross-sectional study in which serum chemistry test and 24-hour urine collection results were evaluated for stone risk. Furthermore, a short-term longitudinal study was conducted in 7 patients to assess stone risk before and 3 months after topiramate treatment. **RESULTS**: Serum bicarbonate levels were lower with topiramate treatment. Urinary pH, urinary bicarbonate excretion, and fractional excretion of bicarbonate increased, whereas urinary citrate excretion was significantly lower (737 +/- 329 versus 278 +/- 226 mg/d; P : Serum bicarbonate levels were lower with topiramate treatment. Urinary pH, urinary bicarbonate excretion, and fractional excretion of bicarbonate increased, whereas urinary citrate excretion was significantly lower (737 +/- 329 versus 278 +/- 226 mg/d; P : Serum bicarbonate levels were lower with topiramate treatment. Urinary pH, urinary bicarbonate excretion, and fractional excretion of bicarbonate increased, whereas urinary citrate excretion was significantly lower (737 +/- 329 versus 278 +/- 226 mg/d; P : Serum bicarbonate levels were lower with topiramate treatment. Urinary pH, urinary bicarbonate excretion, and fractional excretion of bicarbonate increased, whereas urinary citrate excretion was significantly lower (737 +/- 329 versus 278 +/- 226 mg/d; P **CONCLUSION**: Treatment with topiramate causes systemic metabolic acidosis, markedly lower urinary citrate excretion, and increased urinary pH. These changes increase the propensity to form calcium phosphate stones.

**URL:** <https://ovidsp.ovid.com/ovidweb.cgi?T=JS&CSC=Y&NEWS=N&PAGE=fulltext&D=med6&DO=10.1053%2fj.ajkd.2006.07.003> <https://libkey.io/libraries/2592/openurl?genre=article&sid=OVID:medline&id=pmid:16997051&id=doi:10.1053%2Fj.ajkd.2006.07.003&issn=0272-6386&isbn=&volume=48&issue=4&spage=555&pages=555-63&date=2006&title=American+Journal+of+Kidney+Diseases&atitle=Biochemical+and+stone-risk+profiles+with+topiramate+treatment.&aulast=Welch&pid=%3Cauthor%3EWelch+BJ%3BGraybeal+D%3BMoe+OW%3BMaalouf+NM%3BSakhaee+K%3C%2Fauthor%3E%3CAN%3E16997051%3C%2FAN%3E%3CDT%3EClinical+Trial%3C%2FDT%3E>

181. **Renal function in patients with nephrolithiasis.**

**Item Type:** Journal Article

**Authors:** Worcester, Elaine M.;Parks, Joan H.;Evan, Andrew P. and Coe, Fredric L.

**Publication Date:** 2006

**Journal:** Journal of Urology 176(2), pp. 600–603

**Abstract: PURPOSE**: We describe kidney function, as measured by creatinine clearance in stone formers, and classified by type of stone formed and systemic etiologies of stone formation. **MATERIALS AND METHODS**: The mean of 3 pretreatment 24-hour creatinine clearance measurements in each of 1,856 stone formers and creatinine clearance in 153 normal individuals were used. Clearance was adjusted for patient sex, age and body weight using general linear modeling. **RESULTS**: As a group, all stone formers had decreased clearance adjusted for age, sex and body weight compared to that in normal individuals. Although clearance was particularly low in cystine and struvite stone formers, they were below normal in even common CaOx stone formers. **CONCLUSIONS**: As a rule, patients with kidney stones do not have normal kidney function. In clinical management all efforts must be made to minimize renal injury, balancing the risks of obstruction from stones against those of urological procedures.

**URL:** <https://ovidsp.ovid.com/ovidweb.cgi?T=JS&CSC=Y&NEWS=N&PAGE=fulltext&D=med6&DO=10.1016%2fj.juro.2006.03.095> <https://libkey.io/libraries/2592/openurl?genre=article&sid=OVID:medline&id=pmid:16813897&id=doi:10.1016%2Fj.juro.2006.03.095&issn=0022-5347&isbn=&volume=176&issue=2&spage=600&pages=600-3%3B+discussion+603&date=2006&title=Journal+of+Urology&atitle=Renal+function+in+patients+with+nephrolithiasis.&aulast=Worcester&pid=%3Cauthor%3EWorcester+EM%3BParks+JH%3BEvan+AP%3BCoe+FL%3C%2Fauthor%3E%3CAN%3E16813897%3C%2FAN%3E%3CDT%3EJournal+Article%3C%2FDT%3E>

182. **High kidney stone risk in men working in steel industry at hot temperatures.**

**Item Type:** Journal Article

**Authors:** Atan, Luiz;Andreoni, Cassio;Ortiz, Valdemar;Silva, Edina Koga;Pitta, Ricardo;Atan, Fernanda and Srougi, Miguel

**Publication Date:** Ma ,2005

**Journal:** Urology 65(5), pp. 858–861

**Abstract: OBJECTIVES**: To study the incidence of urinary lithiasis and metabolic alterations among male employees from a steel industry who were exposed to high temperatures in the work environment. **METHODS**: A retrospective cross-sectional study was performed and consisted of two stages. First, the incidence of urolithiasis among the industry's 10,326 employees was assessed. These employees were divided into two group's: group 1 (n = 1289) consisted of the hot-area workers (temperature greater than 45 degrees C) and group 2 (n = 9037) consisted of those working in areas at room temperature. In the second stage, 59 workers without urolithiasis who underwent a metabolic evaluation were divided into two group's: group 3 (n = 34) consisted of hot-area workers and group 4 (n = 25) consisted of those working in areas at room temperature. Evaluations were made of calcium, creatinine, and uric acid in serum; in the 24-hour urine samples, we assessed the volume, calcium, uric acid, citrate, and oxalate. **RESULTS**: Of the 10,326 workers, 181 (1.75%) had presented with at least one episode of urinary stones. Of these, 103 were among the hot-area workers (8.0%) and 78 among the room-temperature workers (0.9%; P : Of the 10,326 workers, 181 (1.75%) had presented with at least one episode of urinary stones. Of these, 103 were among the hot-area workers (8.0%) and 78 among the room-temperature workers (0.9%; P **CONCLUSIONS**: Workers exposed to high temperatures presented with a ninefold risk of lithiasis. Hypocitraturia and low urine volumes were the metabolic alterations observed.

**URL:** <https://ovidsp.ovid.com/ovidweb.cgi?T=JS&CSC=Y&NEWS=N&PAGE=fulltext&D=med6&DO=10.1016%2fj.urology.2004.11.048> <https://libkey.io/libraries/2592/openurl?genre=article&sid=OVID:medline&id=pmid:15882711&id=doi:10.1016%2Fj.urology.2004.11.048&issn=0090-4295&isbn=&volume=65&issue=5&spage=858&pages=858-61&date=2005&title=Urology&atitle=High+kidney+stone+risk+in+men+working+in+steel+industry+at+hot+temperatures.&aulast=Atan&pid=%3Cauthor%3EAtan+L%3BAndreoni+C%3BOrtiz+V%3BSilva+EK%3BPitta+R%3BAtan+F%3BSrougi+M%3C%2Fauthor%3E%3CAN%3E15882711%3C%2FAN%3E%3CDT%3EJournal+Article%3C%2FDT%3E>

183. **Serial crystalluria determination and the risk of recurrence in calcium stone formers.**

**Item Type:** Journal Article

**Authors:** Daudon, Michel;Hennequin, Carole;Boujelben, Ghazi;Lacour, Bernard and Jungers, Paul

**Publication Date:** Ma ,2005

**Journal:** Kidney International 67(5), pp. 1934–1943

**Abstract: BACKGROUND**: Urinary crystal precipitation is the necessary initial step in kidney stone formation. However, clinical relevance of crystalluria in the evaluation of stone formers is disputed. **METHODS**: We serially determined crystalluria in first-voided morning urine samples, together with full 24-hour urine biochemistry, in 181 patients with idiopathic calcium nephrolithiasis who had formed at least one calcium-oxalate stone and were followed for at least 3 years under our care. All stone events which occurred prior to referral, then after entry in the study were recorded. **RESULTS**: As compared with 109 patients who had no evidence of stone recurrence during follow-up, the 72 patients who experienced >/= one recurrent stone event had a lower daily urine volume (1.74 +/- 0.06 vs. 2.26 +/- 0.05 L/day (mean +/- SEM) (P : As compared with 109 patients who had no evidence of stone recurrence during follow-up, the 72 patients who experienced >/= one recurrent stone event had a lower daily urine volume (1.74 +/- 0.06 vs. 2.26 +/- 0.05 L/day (mean +/- SEM) (P : As compared with 109 patients who had no evidence of stone recurrence during follow-up, the 72 patients who experienced >/= one recurrent stone event had a lower daily urine volume (1.74 +/- 0.06 vs. 2.26 +/- 0.05 L/day (mean +/- SEM) (P **CONCLUSION**: These data provide evidence that crystalluria, when repeatedly found in early morning urine samples, is highly predictive of the risk of stone recurrence in calcium stone formers. Serial search for crystalluria, a simple and cheap method, may be proposed as a useful tool for the monitoring of calcium stone formers, in addition to urine biochemistry.

**URL:** <https://ovidsp.ovid.com/ovidweb.cgi?T=JS&CSC=Y&NEWS=N&PAGE=fulltext&D=med6&DO=10.1111%2fj.1523-1755.2005.00292.x> <https://libkey.io/libraries/2592/openurl?genre=article&sid=OVID:medline&id=pmid:15840041&id=doi:10.1111%2Fj.1523-1755.2005.00292.x&issn=0085-2538&isbn=&volume=67&issue=5&spage=1934&pages=1934-43&date=2005&title=Kidney+International&atitle=Serial+crystalluria+determination+and+the+risk+of+recurrence+in+calcium+stone+formers.&aulast=Daudon&pid=%3Cauthor%3EDaudon+M%3BHennequin+C%3BBoujelben+G%3BLacour+B%3BJungers+P%3C%2Fauthor%3E%3CAN%3E15840041%3C%2FAN%3E%3CDT%3EJournal+Article%3C%2FDT%3E>

184. **Jejunoileal bypass reversal: effect on renal function, metabolic parameters and stone formation.**

**Item Type:** Journal Article

**Authors:** Dhar, Nivedita Bhatta;Grundfest, Sharon;Jones, Jace S. and Streem, Stevan B.

**Publication Date:** 2005

**Journal:** Journal of Urology 174(5), pp. 1844–1846

**Abstract: PURPOSE**: While the effect of jejunoileal bypass (JIB) reversal has been well studied regarding hepatic function, there is little information regarding the effect of reversal on renal function and even less data regarding the metabolic urinary stone environment. We evaluated the results of JIB reversal on renal function, the urinary stone milieu and the clinical development of recurrent calculi in affected patients. **MATERIALS AND METHODS**: From 1995 to 2003, 4 female patients with a mean age of 48.2 years underwent JIB reversal primarily for refractory stone disease. The clinical and metabolic courses prior to and following bypass reversal were reviewed specifically to evaluate renal function, serum and urinary metabolic stone profiles, and clinical stone formation. **RESULTS**: At initial presentation following JIB all 4 patients had significantly increased 24-hour urinary oxalate (range 80 to 160 mg, mean 112.5, normal less than 50) and significantly low 24-hour urinary citrate (range 5 to 62 mg, mean 21.5, normal greater than 320). Following reversal 24-hour urinary oxalate normalized to between 31 and 36 mg (mean 33.75). However, 24-hour urinary citrate continued to be low (range 215 to 248 mg, mean 226.5). After JIB reversal all 4 patients continued to have new stones until the commencement of urinary alkalization, following which only 1 had 1 calculus, which occurred 47 months after reversal. After JIB mean serum creatinine was 1.48 mg/dl (range 0.8 to 1.9) and mean urinary creatinine excretion was 0.91 mg per hour (range 0.69 to 1.15). After JIB reversal mean serum creatinine was 1.28 mg/dl (range 0.6 to 2.0) and mean urinary creatinine excretion was 1.0 mg per hour (range 0.85 to 1.10). **CONCLUSIONS**: JIB reversal normalizes 24-hour urinary oxalate. While urinary citrate improves, it continues to be low and such patients are at high risk for recurrent stone formation. However, in this setting appropriate replacement therapy has a significant and positive impact on that propensity.

**URL:** <https://ovidsp.ovid.com/ovidweb.cgi?T=JS&CSC=Y&NEWS=N&PAGE=fulltext&D=med6&DO=10.1097%2f01.ju.0000177079.56949.1a> <https://libkey.io/libraries/2592/openurl?genre=article&sid=OVID:medline&id=pmid:16217311&id=doi:10.1097%2F01.ju.0000177079.56949.1a&issn=0022-5347&isbn=&volume=174&issue=5&spage=1844&pages=1844-6%3B+discussion+1846&date=2005&title=Journal+of+Urology&atitle=Jejunoileal+bypass+reversal%3A+effect+on+renal+function%2C+metabolic+parameters+and+stone+formation.&aulast=Dhar&pid=%3Cauthor%3EDhar+NB%3BGrundfest+S%3BJones+JS%3BStreem+SB%3C%2Fauthor%3E%3CAN%3E16217311%3C%2FAN%3E%3CDT%3EJournal+Article%3C%2FDT%3E>

185. **Increased frequency of ultrasonographic findings suggestive of renal stones in patients with ankylosing spondylitis.**

**Item Type:** Journal Article

**Authors:** Korkmaz, C.;Ozcan, A. and Akcar, N.

**Publication Date:** 2005

**Journal:** Clinical & Experimental Rheumatology 23(3), pp. 389–392

**Abstract: OBJECTIVE**: To determine the frequency of hypercalciuria and renal stones in ankylosing spondylitis (AS) sufferers. **METHODS**: This study involved 83 consecutive AS patients (21 female, 62 male; mean age 36.7 yr), 72 consecutive Behcet's disease (BD) patients (29 female, 43 male; mean age 37.7 yr) as disease control and 92 healthy control (HC) (26 female, 66 male; mean age 32.9 yr.) Twenty-four hour urine analyses for urinary calcium and uric acid levels were performed in each patient. Likewise, blood samples for erythrocyte sedimentation rate (ESR), C-reactive protein (CRP), parathyroid hormone (PTH), calcium and uric acid evaluation were taken. Renal ultrasonography to evaluate the presence of renal stone was performed in patients with AS, as well as in the BD patients and HC individuals after a fasting period of 12 hours. **RESULTS**: 20 of the 80 (25%) patients with AS were diagnosed with renal stones. Only 4/72 (5.5%) BD patients, and 3/98 (3.3%) HC individuals had renal stones by ultrasonography. AS patients had a significantly higher frequency of renal stones compared with BD (p : 20 of the 80 (25%) patients with AS were diagnosed with renal stones. Only 4/72 (5.5%) BD patients, and 3/98 (3.3%) HC individuals had renal stones by ultrasonography. AS patients had a significantly higher frequency of renal stones compared with BD (p : 20 of the 80 (25%) patients with AS were diagnosed with renal stones. Only 4/72 (5.5%) BD patients, and 3/98 (3.3%) HC individuals had renal stones by ultrasonography. AS patients had a significantly higher frequency of renal stones compared with BD (p : 20 of the 80 (25%) patients with AS were diagnosed with renal stones. Only 4/72 (5.5%) BD patients, and 3/98 (3.3%) HC individuals had renal stones by ultrasonography. AS patients had a significantly higher frequency of renal stones compared with BD (p **CONCLUSION**: Renal stone prevalence was found to increase in AS patients. The likelihood of renal stone formation was also found to increase with the extension of the disease duration of AS sufferers.

**URL:** <https://ovidsp.ovid.com/ovidweb.cgi?T=JS&CSC=Y&NEWS=N&PAGE=fulltext&D=med6&AN=15971429> <https://libkey.io/libraries/2592/openurl?genre=article&sid=OVID:medline&id=pmid:15971429&id=doi:&issn=0392-856X&isbn=&volume=23&issue=3&spage=389&pages=389-92&date=2005&title=Clinical+%26+Experimental+Rheumatology&atitle=Increased+frequency+of+ultrasonographic+findings+suggestive+of+renal+stones+in+patients+with+ankylosing+spondylitis.&aulast=Korkmaz&pid=%3Cauthor%3EKorkmaz+C%3BOzcan+A%3BAkcar+N%3C%2Fauthor%3E%3CAN%3E15971429%3C%2FAN%3E%3CDT%3EJournal+Article%3C%2FDT%3E>

186. **Use of a probiotic to decrease enteric hyperoxaluria.**

**Item Type:** Journal Article

**Authors:** Lieske, John C.;Goldfarb, David S.;De Simone, Claudio and Regnier, Cynthia

**Publication Date:** Se ,2005

**Journal:** Kidney International 68(3), pp. 1244–1249

**Abstract: BACKGROUND**: Patients with inflammatory bowel disease have a 10- to 100-fold increased risk of nephrolithiasis, with enteric hyperoxaluria being the major risk factor for these and other patients with fat malabsorptive states. Endogenous components of the intestinal microflora can potentially limit dietary oxalate absorption. **METHODS**: Ten patients were studied with chronic fat malabsorption, calcium oxalate stones, and hyperoxaluria thought to be caused by jejunoileal bypass (1) and Roux-en-Y gastric bypass surgery for obesity (4), dumping syndrome secondary to gastrectomy (2), celiac sprue (1), chronic pancreatitis (1), and ulcerative colitis in remission (1). For 3 months, patients received increasing doses of a lactic acid bacteria mixture (Oxadrop), VSL Pharmaceuticals), followed by a washout month. Twenty-four-hour urine collections were performed at baseline and after each month. **RESULTS**: Mean urinary oxalate excretion fell by 19% after 1 month (1 dose per day, P : Mean urinary oxalate excretion fell by 19% after 1 month (1 dose per day, P : Mean urinary oxalate excretion fell by 19% after 1 month (1 dose per day, P **CONCLUSION**: Manipulation of gastrointestinal (GI) flora can influence urinary oxalate excretion to reduce urinary supersaturation levels. These changes could have a salutary effect on stone formation rates. Further studies will be needed to establish the optimal dosing regimen.

**URL:** <https://ovidsp.ovid.com/ovidweb.cgi?T=JS&CSC=Y&NEWS=N&PAGE=fulltext&D=med6&DO=10.1111%2fj.1523-1755.2005.00520.x> <https://libkey.io/libraries/2592/openurl?genre=article&sid=OVID:medline&id=pmid:16105057&id=doi:10.1111%2Fj.1523-1755.2005.00520.x&issn=0085-2538&isbn=&volume=68&issue=3&spage=1244&pages=1244-9&date=2005&title=Kidney+International&atitle=Use+of+a+probiotic+to+decrease+enteric+hyperoxaluria.&aulast=Lieske&pid=%3Cauthor%3ELieske+JC%3BGoldfarb+DS%3BDe+Simone+C%3BRegnier+C%3C%2Fauthor%3E%3CAN%3E16105057%3C%2FAN%3E%3CDT%3EClinical+Trial%3C%2FDT%3E>

187. **Segregation of urine calcium excretion in families ascertained for nephrolithiasis: evidence for a major gene.**

**Item Type:** Journal Article

**Authors:** Loredo-Osti, J. C.;Roslin, Nicole M.;Tessier, Josee;Fujiwara, T. Mary;Morgan, Kenneth and Bonnardeaux, Alain

**Publication Date:** Se ,2005

**Journal:** Kidney International 68(3), pp. 966–971

**Abstract: BACKGROUND**: The quantitative genetics of urine calcium excretion has not been established. It is a trait of interest because hypercalciuria is commonly found in subjects with nephrolithiasis. The aim of this study was to model the segregation of this trait in a sample of French-Canadian families ascertained through a stone former. **METHODS**: Major gene, polygenic, and mixed models were fit to 24-hour urine calcium excretion from 567 individuals in 221 nuclear families, while simultaneously taking into account gender, age at examination, body mass index (BMI), and the use of thiazide drugs. The nuclear families were extracted from 154 pedigrees, some of which were four generations, with at least two siblings with a history of calcium stones. **RESULTS**: All the proposed genetic models fit the data significantly better than the null model. The most parsimonious model was the mixed codominant/polygenic model but it was statistically indistinguishable from the single-gene codominant model. In both of these models the heritability attributable to the major gene was estimated to be 0.58. **CONCLUSION**: Our results suggest that a major gene with a relatively large effect on variation in urine calcium excretion is segregating in French-Canadian families with stone formers. This implies that the power of quantitative trait segregation analysis of urine calcium excretion may be increased in these families, and results indicate that it should be feasible to genetically map the quantitative trait locus.

**URL:** <https://ovidsp.ovid.com/ovidweb.cgi?T=JS&CSC=Y&NEWS=N&PAGE=fulltext&D=med6&DO=10.1111%2fj.1523-1755.2005.00490.x> <https://libkey.io/libraries/2592/openurl?genre=article&sid=OVID:medline&id=pmid:16105027&id=doi:10.1111%2Fj.1523-1755.2005.00490.x&issn=0085-2538&isbn=&volume=68&issue=3&spage=966&pages=966-71&date=2005&title=Kidney+International&atitle=Segregation+of+urine+calcium+excretion+in+families+ascertained+for+nephrolithiasis%3A+evidence+for+a+major+gene.&aulast=Loredo-Osti&pid=%3Cauthor%3ELoredo-Osti+JC%3BRoslin+NM%3BTessier+J%3BFujiwara+TM%3BMorgan+K%3BBonnardeaux+A%3C%2Fauthor%3E%3CAN%3E16105027%3C%2FAN%3E%3CDT%3EJournal+Article%3C%2FDT%3E>

188. **Quantification of hypercalciuria with the urine calcium osmolality ratio in children.**

**Item Type:** Journal Article

**Authors:** Mir, Sevgi and Serdaroglu, Erkin

**Publication Date:** No ,2005

**Journal:** Pediatric Nephrology 20(11), pp. 1562–1565

**Abstract:** Hypercalciuria is a major risk factor for kidney stones and hematuria in children. Calcium measurements in 24-h collected urine (mg/kg/d) is still essential for the hypercalciuria diagnose but collection of 24 hour urine is difficult especially in young children. A prospective study was conducted to compare calcium/osmolality (Uca/Uosm) ratio to calcium/creatinine (Uea/Ucr) and to correlate both ratios to 24-hour urine calcium excretion (mg/kg/d) and the value of Uca/Uosm ratio for diagnosis of hypercalciuria in children. Two hundred and fifteen patients aged between 3 to 15 years (median 8 years) were included in the study. Early morning second urine samples and 24-hour collected urine samples were analyzed. The 24-hour urinary calcium excretions were significantly correlated with Uca/Ucr and Uca/Uosm ratio (r =0.67, P<0.001 and r =0.61, P<0.001 respectively). The accepted hypercalciuria level of 24-hour urine calcium of 4 mg/kg/day coincides with Uca/Ucr ratio of 0.18 mg/mg (sensitivity 90.0%, specificity 84.4%) and with Uca/Uosm ratio of 0.14 mg/l/mOsm/kg (sensitivity 90%, specificity 81.0%) in the ROC curve analysis. Positive and negative predictive values were found 98.3% and 39.1% for Uca/Uosm ratio and respectively 43.9% and 98.4% for Uca/Ucr ratio. In conclusion Uca/Uosm ratio is a good marker of hypercalciuria as well as Uca/Ucr ratio and may be used for screening of hypercalciuria.

**URL:** <https://ovidsp.ovid.com/ovidweb.cgi?T=JS&CSC=Y&NEWS=N&PAGE=fulltext&D=med6&DO=10.1007%2fs00467-005-1992-8> <https://libkey.io/libraries/2592/openurl?genre=article&sid=OVID:medline&id=pmid:16133062&id=doi:10.1007%2Fs00467-005-1992-8&issn=0931-041X&isbn=&volume=20&issue=11&spage=1562&pages=1562-5&date=2005&title=Pediatric+Nephrology&atitle=Quantification+of+hypercalciuria+with+the+urine+calcium+osmolality+ratio+in+children.&aulast=Mir&pid=%3Cauthor%3EMir+S%3BSerdaroglu+E%3C%2Fauthor%3E%3CAN%3E16133062%3C%2FAN%3E%3CDT%3EJournal+Article%3C%2FDT%3E>

189. **Enteric hyperoxaluria, nephrolithiasis, and oxalate nephropathy: potentially serious and unappreciated complications of Roux-en-Y gastric bypass.**

**Item Type:** Journal Article

**Authors:** Nelson, Wayne K.;Houghton, Scott G.;Milliner, Dawn S.;Lieske, John C. and Sarr, Michael G.

**Publication Date:** 2005

**Journal:** Surgery for Obesity & Related Diseases 1(5), pp. 481–485

**Abstract: BACKGROUND**: Neither the presence nor prevalence of enteric hyperoxaluria has been recognized after Roux-en-Y gastric bypass (RYGBP). We have noted a high rate of oxalate nephrolithiasis and even 2 patients with oxalate nephropathy in this patient population postoperatively. Our aim was to determine the frequency of the occurrence and effects of enteric hyperoxaluria after RYGBP. **METHODS**: Retrospective review of all patients at our institution diagnosed with calcium oxalate nephrolithiasis or oxalate nephropathy after standard (n = 14) or distal (n = 9) RYGBP. The mean postoperative follow-up was 55 months. **RESULTS**: A total of 23 patients (14 men and 9 women; mean age 45 years; mean preoperative body mass index 55 kg/m(2)) developed enteric hyperoxaluria after RYGBP, defined by the presence of oxalate nephropathy (n = 2) or calcium oxalate nephrolithiasis (n = 21) and increased 24-hour excretion of urinary oxalate and/or calcium oxalate supersaturation. Enteric hyperoxaluria was recognized after a mean weight loss of 46 kg at 29 months (range 2-85) after RYGBP. Two patients developed renal failure and required chronic hemodialysis. Of the 21 patients with nephrolithiasis, 14 had no history of nephrolithiasis preoperatively, and 19 of 21 required lithotripsy or other intervention. Of the 23 patients, 20 tested had increased oxalate excretion, and 14 of 15 tested had high urine calcium oxalate supersaturation. **CONCLUSION**: Enteric hyperoxaluria, nephrolithiasis, and oxalate nephropathy must be considered with the other risks of RYGBP. Efforts should be made to identify factors that predispose patients to developing hyperoxaluria.

**URL:** <https://ovidsp.ovid.com/ovidweb.cgi?T=JS&CSC=Y&NEWS=N&PAGE=fulltext&D=med6&DO=10.1016%2fj.soard.2005.07.002> <https://libkey.io/libraries/2592/openurl?genre=article&sid=OVID:medline&id=pmid:16925274&id=doi:10.1016%2Fj.soard.2005.07.002&issn=1550-7289&isbn=&volume=1&issue=5&spage=481&pages=481-5&date=2005&title=Surgery+for+Obesity+%26+Related+Diseases&atitle=Enteric+hyperoxaluria%2C+nephrolithiasis%2C+and+oxalate+nephropathy%3A+potentially+serious+and+unappreciated+complications+of+Roux-en-Y+gastric+bypass.&aulast=Nelson&pid=%3Cauthor%3ENelson+WK%3BHoughton+SG%3BMilliner+DS%3BLieske+JC%3BSarr+MG%3C%2Fauthor%3E%3CAN%3E16925274%3C%2FAN%3E%3CDT%3EJournal+Article%3C%2FDT%3E>

190. **Genetic and dietary factors in urinary citrate excretion.**

**Item Type:** Journal Article

**Authors:** Shah, Ojas;Assimos, Dean G. and Holmes, Ross P.

**Publication Date:** Ma ,2005

**Journal:** Journal of Endourology 19(2), pp. 177–182

**Abstract: BACKGROUND AND PURPOSE**: Hypocitraturia, an important risk factor for calcium oxalate nephrolithiasis, is the result of numerous factors. We studied citrate excretion by patients with and without stones consuming normal and controlled formula diets. **SUBJECTS AND METHODS**: Subjects with and without a history of calcium oxalate stones (N = 101 per group) provided two or three 24-hour urine specimens during consumption of self-selected diets. Data also were collected on subsets of subjects consuming formula (Ensure) diets. Citrate was determined using the citrate lyase method of Petrarulo and associates, and values for multiple specimens were averaged. The data were adjusted for creatinine excretion and examined on a per-day basis. **RESULTS**: The mean citrate excretion of the non-stone formers was slightly but not significantly higher than that of the stone formers (442 +/- 217 versus 378 +/- 153 mg/g of creatinine). All statistical analyses revealed highly significant differences between, but not within, individuals, a result compatible with a genetic influence. In the normal population, 5% of subjects had a citrate excretion : The mean citrate excretion of the non-stone formers was slightly but not significantly higher than that of the stone formers (442 +/- 217 versus 378 +/- 153 mg/g of creatinine). All statistical analyses revealed highly significant differences between, but not within, individuals, a result compatible with a genetic influence. In the normal population, 5% of subjects had a citrate excretion : The mean citrate excretion of the non-stone formers was slightly but not significantly higher than that of the stone formers (442 +/- 217 versus 378 +/- 153 mg/g of creatinine). All statistical analyses revealed highly significant differences between, but not within, individuals, a result compatible with a genetic influence. In the normal population, 5% of subjects had a citrate excretion **CONCLUSION**: These findings imply a genetic influence on citrate excretion, as has already been demonstrated for calcium excretion. Further studies of genetic influences on calcium oxalate stone formation are warranted.

**URL:** <https://ovidsp.ovid.com/ovidweb.cgi?T=JS&CSC=Y&NEWS=N&PAGE=fulltext&D=med6&DO=10.1089%2fend.2005.19.177> <https://libkey.io/libraries/2592/openurl?genre=article&sid=OVID:medline&id=pmid:15798414&id=doi:10.1089%2Fend.2005.19.177&issn=0892-7790&isbn=&volume=19&issue=2&spage=177&pages=177-82&date=2005&title=Journal+of+Endourology&atitle=Genetic+and+dietary+factors+in+urinary+citrate+excretion.&aulast=Shah&pid=%3Cauthor%3EShah+O%3BAssimos+DG%3BHolmes+RP%3C%2Fauthor%3E%3CAN%3E15798414%3C%2FAN%3E%3CDT%3EJournal+Article%3C%2FDT%3E>

191. **The metabolic syndrome and uric acid nephrolithiasis: novel features of renal manifestation of insulin resistance.**

**Item Type:** Journal Article

**Authors:** Abate, Nicola;Chandalia, Manisha;Cabo-Chan, Alberto V. Jr;Moe, Orson W. and Sakhaee, Khashayar

**Publication Date:** Fe ,2004

**Journal:** Kidney International 65(2), pp. 386–392

**Abstract: BACKGROUND**: Uric acid nephrolithiasis primarily results from low urinary pH, which increases the concentration of the insoluble undissociated uric acid, causing formation of both uric acid and mixed uric acid/calcium oxalate stones. These patients have recently been described as exhibiting features of insulin resistance. This study was designed to evaluate if insulin resistance is associated with excessively low urinary pH in overtly healthy volunteers (non-stone formers) and if insulin resistance may explain the excessively low urinary pH in patients with uric acid nephrolithiasis. **METHODS**: Fifty-five healthy volunteers (non stone-formers) with a large range of body mass index and 13 patients with recurrent uric acid nephrolithiasis underwent hyperinsulinemic euglycemic clamp, 24-hour urinary studies, and anthropometric measurements of adiposity. A subgroup of 35 non-stone formers had 2-hour timed urinary collection before and during the hyperinsulinemic phase of the clamp studies. **RESULTS**: For the non-stone former population, low insulin sensitivity measured as glucose disposal rate significantly correlated with low 24-hour urinary pH (r= 0. 35; P= 0.01). In addition to the previously described acidic urine pH and hypouricosuria, patients with recurrent uric acid nephrolithiasis were found to be severely insulin resistant (glucose disposal rate: uric acid stone-formers vs. normals; 4.1 +/- 1.3 vs. 6.9 +/- 2.1 mg/min/kg of lean body mass, P= 0.008). Acute hyperinsulinemia was associated with higher urinary pH (6.1 +/- 0.7 at baseline to 6.8 +/- 0.7 during hyperinsulinemia; P : For the non-stone former population, low insulin sensitivity measured as glucose disposal rate significantly correlated with low 24-hour urinary pH (r= 0. 35; P= 0.01). In addition to the previously described acidic urine pH and hypouricosuria, patients with recurrent uric acid nephrolithiasis were found to be severely insulin resistant (glucose disposal rate: uric acid stone-formers vs. normals; 4.1 +/- 1.3 vs. 6.9 +/- 2.1 mg/min/kg of lean body mass, P= 0.008). Acute hyperinsulinemia was associated with higher urinary pH (6.1 +/- 0.7 at baseline to 6.8 +/- 0.7 during hyperinsulinemia; P : For the non-stone former population, low insulin sensitivity measured as glucose disposal rate significantly correlated with low 24-hour urinary pH (r= 0. 35; P= 0.01). In addition to the previously described acidic urine pH and hypouricosuria, patients with recurrent uric acid nephrolithiasis were found to be severely insulin resistant (glucose disposal rate: uric acid stone-formers vs. normals; 4.1 +/- 1.3 vs. 6.9 +/- 2.1 mg/min/kg of lean body mass, P= 0.008). Acute hyperinsulinemia was associated with higher urinary pH (6.1 +/- 0.7 at baseline to 6.8 +/- 0.7 during hyperinsulinemia; P **CONCLUSION**: We conclude that one renal manifestation of insulin resistance may be low urinary ammonium and pH. This defect can result in increased risk of uric acid precipitation despite normouricosuria.

**URL:** <https://ovidsp.ovid.com/ovidweb.cgi?T=JS&CSC=Y&NEWS=N&PAGE=fulltext&D=med5&DO=10.1111%2fj.1523-1755.2004.00386.x> <https://libkey.io/libraries/2592/openurl?genre=article&sid=OVID:medline&id=pmid:14717908&id=doi:10.1111%2Fj.1523-1755.2004.00386.x&issn=0085-2538&isbn=&volume=65&issue=2&spage=386&pages=386-92&date=2004&title=Kidney+International&atitle=The+metabolic+syndrome+and+uric+acid+nephrolithiasis%3A+novel+features+of+renal+manifestation+of+insulin+resistance.&aulast=Abate&pid=%3Cauthor%3EAbate+N%3BChandalia+M%3BCabo-Chan+AV+Jr%3BMoe+OW%3BSakhaee+K%3C%2Fauthor%3E%3CAN%3E14717908%3C%2FAN%3E%3CDT%3EJournal+Article%3C%2FDT%3E>

192. **Assessment of oxalate absorption from almonds and black beans with and without the use of an extrinsic label.**

**Item Type:** Journal Article

**Authors:** Chai, Weiwen and Liebman, Michael

**Publication Date:** Se ,2004

**Journal:** Journal of Urology 172(3), pp. 953–957

**Abstract: PURPOSE**: Oxalate bioavailability is an important determinant of whether the consumption of a particular food is a high risk in individuals predisposed to kidney stones. We estimated and compared oxalate absorption from a high oxalate containing legume (black beans) and a high oxalate containing nut (almonds). We also compared an isotope method using extrinsically labeled oxalate and an oxalate load method to assess oxalate absorption. **MATERIALS AND METHODS**: Six male and 5 female subjects participated in the 4 oxalate load tests, namely almonds, almonds with 20 mg C2-oxalic acid, black beans and black beans with 20 mg C2-oxalic acid. Each treatment provided a total of 120 mg oxalate, after which timed urine samples were collected for the analysis of oxalate, calcium and creatinine. **RESULTS**: Average oxalate absorption from the 2 almond treatments (5.9%) using the oxalate load method was significantly higher than that from the 2 black bean treatments (1.8%) during the 24-hour post-oxalate load collection period. In contrast, C2-oxalic acid absorption from the almond (7.9%) and black bean (8.6%) treatments did not significantly differ. **CONCLUSIONS**: The higher oxalate absorption from almonds than from black beans suggests that the relative amount of soluble and insoluble oxalate in food has an important role in the determination of oxalate absorption. Since extrinsically provided C2-oxalate and oxalate naturally occurring in the high oxalate test foods appeared to be differentially absorbed, the data do not support the use of extrinsically labeled oxalate to assess food oxalate absorption.

**URL:** <https://ovidsp.ovid.com/ovidweb.cgi?T=JS&CSC=Y&NEWS=N&PAGE=fulltext&D=med5&DO=10.1097%2f01.ju.0000135918.00761.8a> <https://libkey.io/libraries/2592/openurl?genre=article&sid=OVID:medline&id=pmid:15311007&id=doi:10.1097%2F01.ju.0000135918.00761.8a&issn=0022-5347&isbn=&volume=172&issue=3&spage=953&pages=953-7&date=2004&title=Journal+of+Urology&atitle=Assessment+of+oxalate+absorption+from+almonds+and+black+beans+with+and+without+the+use+of+an+extrinsic+label.&aulast=Chai&pid=%3Cauthor%3EChai+W%3BLiebman+M%3C%2Fauthor%3E%3CAN%3E15311007%3C%2FAN%3E%3CDT%3EJournal+Article%3C%2FDT%3E>

193. **Schedule of taking calcium supplement and the risk of nephrolithiasis.**

**Item Type:** Journal Article

**Authors:** Domrongkitchaiporn, Somnuek;Sopassathit, Wichai;Stitchantrakul, Wasana;Prapaipanich, Surasing;Ingsathit, Atiporn and Rajatanavin, Rajata

**Publication Date:** Ma ,2004

**Journal:** Kidney International 65(5), pp. 1835–1841

**Abstract: BACKGROUND**: Variation in the timing of calcium supplement may affect gastrointestinal absorption of both calcium and oxalate differently and may associate with variable risk of calcium oxalate nephrolithiasis. There are few human studies addressing specifically the appropriate time for taking calcium supplement. Therefore, this study was performed to compare calcium bioavailability and the risk of calcium oxalate stone formation for calcium supplement taken with meal vs. taken at bedtime. **METHODS**: Thirty-two healthy male navy privates, 22.7 +/- 1.9 years old (mean +/- SD), who had normal renal function (serum creatinine less than 150 umol/L) and no history of renal stone, participated in the study. The subjects were randomly allocated into two groups of 16 each. Group A took 1 g of calcium carbonate with meal, 3 times/day; and group B took 3 g/day of calcium carbonate at bedtime. After taking the regimens for 1 week, followed by 1 month of washout period, crossover between both groups was done. The diet was controlled throughout the study. Twenty-four-hour urine collections for the determination of urinary constituents were obtained at baseline and after taking both regimens of calcium supplement. Activity product for calcium oxalate was determined to assess the risk of calcium oxalate stone formation. **RESULTS**: Urinary excretions of calcium were significantly elevated above the baseline values when taking calcium supplement both with meal (3.48 +/- 2.13 mmol/day vs. 5.17 +/- 2.61 mmol/day, P : Urinary excretions of calcium were significantly elevated above the baseline values when taking calcium supplement both with meal (3.48 +/- 2.13 mmol/day vs. 5.17 +/- 2.61 mmol/day, P : Urinary excretions of calcium were significantly elevated above the baseline values when taking calcium supplement both with meal (3.48 +/- 2.13 mmol/day vs. 5.17 +/- 2.61 mmol/day, P : Urinary excretions of calcium were significantly elevated above the baseline values when taking calcium supplement both with meal (3.48 +/- 2.13 mmol/day vs. 5.17 +/- 2.61 mmol/day, P **CONCLUSION**: Calcium supplement should be taken with meal in order to avoid increasing the risk of calcium oxalate nephrolithiasis.

**URL:** <https://ovidsp.ovid.com/ovidweb.cgi?T=JS&CSC=Y&NEWS=N&PAGE=fulltext&D=med5&DO=10.1111%2fj.1523-1755.2004.00587.x> <https://libkey.io/libraries/2592/openurl?genre=article&sid=OVID:medline&id=pmid:15086924&id=doi:10.1111%2Fj.1523-1755.2004.00587.x&issn=0085-2538&isbn=&volume=65&issue=5&spage=1835&pages=1835-41&date=2004&title=Kidney+International&atitle=Schedule+of+taking+calcium+supplement+and+the+risk+of+nephrolithiasis.&aulast=Domrongkitchaiporn&pid=%3Cauthor%3EDomrongkitchaiporn+S%3BSopassathit+W%3BStitchantrakul+W%3BPrapaipanich+S%3BIngsathit+A%3BRajatanavin+R%3C%2Fauthor%3E%3CAN%3E15086924%3C%2FAN%3E%3CDT%3EClinical+Trial%3C%2FDT%3E>

194. **Changes in urinary parameters after oral administration of potassium-sodium citrate and magnesium oxide to prevent urolithiasis.**

**Item Type:** Journal Article

**Authors:** Kato, Yuji;Yamaguchi, Satoshi;Yachiku, Sunao;Nakazono, Shusaku;Hori, Jun-ichi;Wada, Naoki and Hou, Kyokushin

**Publication Date:** 2004

**Journal:** Urology 63(1), pp. 7–11

**Abstract: OBJECTIVES**: To examine urinary parameters among normal individuals and patients with calcium oxalate (CaOx) stones after oral administration of potassium-sodium citrate (KNa-Cit) and magnesium oxide (MgO). Urinary citrate and magnesium have been known as the inhibitors of CaOx stone formation. Supplementation with potassium-magnesium citrate prevents the recurrence of CaOx stones. **METHODS**: Twenty-five male volunteers aged 21 to 42 years without a history of urinary stones were given either KNa-Cit or MgO, or both. Fourteen patients with recurrent CaOx stones were also given both supplements, and 24-hour urine samples were collected to determine the urinary parameters. **RESULTS**: The administration of both KNa-Cit and MgO to the normal individuals increased the excretion of citrate, magnesium, and potassium by 70.0%, 44.2%, and 50.0%, respectively. These parameters increased less when KNa-Cit or MgO was administered individually. After administration of both supplements to the patients with stones, the citrate, magnesium, and potassium levels increased by 62.1%, 63.3%, and 25.3%, respectively, and oxalate decreased by 66.5%. In both normal individuals and patients, the ion activity product index of CaOx decreased significantly more after administration of the combination than with either compound alone or before administration. **CONCLUSION**: The combination of KNa-Cit and MgO is more effective than either supplement alone in inhibiting the crystallization of CaOx stones. The combination may improve the urinary parameters of patients with stones accompanied by hypocitraturia and/or hypomagnesuria.

**URL:** <https://ovidsp.ovid.com/ovidweb.cgi?T=JS&CSC=Y&NEWS=N&PAGE=fulltext&D=med5&DO=10.1016%2fj.urology.2003.09.057> <https://libkey.io/libraries/2592/openurl?genre=article&sid=OVID:medline&id=pmid:14751336&id=doi:10.1016%2Fj.urology.2003.09.057&issn=0090-4295&isbn=&volume=63&issue=1&spage=7&pages=7-11%3B+discussion+11-2&date=2004&title=Urology&atitle=Changes+in+urinary+parameters+after+oral+administration+of+potassium-sodium+citrate+and+magnesium+oxide+to+prevent+urolithiasis.&aulast=Kato&pid=%3Cauthor%3EKato+Y%3BYamaguchi+S%3BYachiku+S%3BNakazono+S%3BHori+J%3BWada+N%3BHou+K%3C%2Fauthor%3E%3CAN%3E14751336%3C%2FAN%3E%3CDT%3EClinical+Trial%3C%2FDT%3E>

195. **Association of urinary pH with body weight in nephrolithiasis.**

**Item Type:** Journal Article

**Authors:** Maalouf, Naim M.;Sakhaee, Khashayar;Parks, Joan H.;Coe, Frederic L.;Adams-Huet, Beverley and Pak, Charles Y. C.

**Publication Date:** Ap ,2004

**Journal:** Kidney International 65(4), pp. 1422–1425

**Abstract: BACKGROUND**: The prevalence of kidney stone disease in the United States is progressively increasing, paralleling the growing rate of obesity. Uric acid nephrolithiasis, a condition associated with a low urinary pH, has been linked to obesity and insulin resistance. Based on these observations, we hypothesized that urinary pH may be inversely associated to body weight in nephrolithiasis. **METHODS**: Data were retrieved from 4883 patients with nephrolithiasis who underwent ambulatory evaluation at two established stone clinics in Dallas and Chicago. The patients collected 24-hour urine samples on an outpatient basis, while avoiding any drug that could alter urinary pH. Patients were divided in increasing sextiles of body weight, and urinary pH was adjusted for urinary creatinine and for age. **RESULTS**: Urinary pH had a strong, graded inverse association with body weight. Urinary creatinine and age were both found to be significant covariates of urinary pH, while gender was not a significant independent variable after adjustment for urinary creatinine. Mean 24-hour urinary pH, adjusted for age and urinary creatinine, were 6.09, 6.04, 6.01, 5.99, 5.97, and 5.91 for sextiles of body weight in increasing order from Dallas (P for linear trend : Urinary pH had a strong, graded inverse association with body weight. Urinary creatinine and age were both found to be significant covariates of urinary pH, while gender was not a significant independent variable after adjustment for urinary creatinine. Mean 24-hour urinary pH, adjusted for age and urinary creatinine, were 6.09, 6.04, 6.01, 5.99, 5.97, and 5.91 for sextiles of body weight in increasing order from Dallas (P for linear trend : Urinary pH had a strong, graded inverse association with body weight. Urinary creatinine and age were both found to be significant covariates of urinary pH, while gender was not a significant independent variable after adjustment for urinary creatinine. Mean 24-hour urinary pH, adjusted for age and urinary creatinine, were 6.09, 6.04, 6.01, 5.99, 5.97, and 5.91 for sextiles of body weight in increasing order from Dallas (P for linear trend **CONCLUSION**: We conclude that urinary pH is inversely related to body weight among patients with stones. The results confirm the previously proposed scheme that obesity may sometimes cause uric acid nephrolithiasis by producing excessively acid urine due to insulin resistance.

**URL:** <https://ovidsp.ovid.com/ovidweb.cgi?T=JS&CSC=Y&NEWS=N&PAGE=fulltext&D=med5&DO=10.1111%2fj.1523-1755.2004.00522.x> <https://libkey.io/libraries/2592/openurl?genre=article&sid=OVID:medline&id=pmid:15086484&id=doi:10.1111%2Fj.1523-1755.2004.00522.x&issn=0085-2538&isbn=&volume=65&issue=4&spage=1422&pages=1422-5&date=2004&title=Kidney+International&atitle=Association+of+urinary+pH+with+body+weight+in+nephrolithiasis.&aulast=Maalouf&pid=%3Cauthor%3EMaalouf+NM%3BSakhaee+K%3BParks+JH%3BCoe+FL%3BAdams-Huet+B%3BPak+CY%3C%2Fauthor%3E%3CAN%3E15086484%3C%2FAN%3E%3CDT%3EJournal+Article%3C%2FDT%3E>

196. **Metabolic abnormalities associated with renal calculi in patients with horseshoe kidneys.**

**Item Type:** Journal Article

**Authors:** Raj, Ganesh V.;Auge, Brian K.;Assimos, Dean and Preminger, Glenn M.

**Publication Date:** Ma ,2004

**Journal:** Journal of Endourology 18(2), pp. 157–161

**Abstract: BACKGROUND AND PURPOSE**: Horseshoe kidneys are a complex anatomic variant of fused kidneys, with a 20% reported incidence of associated calculi. Anatomic causes such as high insertion of the ureter on the renal pelvis and obstruction of the ureteropelvic junction are thought to contribute to stone formation via impaired drainage, with urinary stasis, and an increased incidence of infection. In this multi-institutional study, we evaluated whether metabolic factors contributed to stone development in patients with horseshoe kidneys. **PATIENTS AND METHODS**: A retrospective review of 37 patients with horseshoe kidneys was performed to determine if these patients had metabolic derangements that might have contributed to calculus formation. Stone compositions as well as 24-hour urine collections were examined. Specific data points of interest were total urine volume; urine pH; urine concentrations of calcium, sodium, uric acid, oxalate, and citrate; and number of abnormalities per patient per 24-hour urine collection. These data were compared with those of a group of 13 patients with stones in caliceal diverticula as well as 24 age-, race-, and sex-matched controls with stones in anatomically normal kidneys. **RESULTS**: Eleven (9 men and 2 women) of the 37 patients (30%) with renal calculi in horseshoe kidneys had complete metabolic evaluations available for review. All patients were noted to have at least one abnormality, with an average of 2.68 abnormalities per 24-hour urine collection (range 1-4). One patient had primary hyperparathyroidism and underwent a parathyroidectomy. Low urine volumes were noted in eight patients on at least one of the two specimens (range 350-1640 mL/day). Hypercalciuria, hyperoxaluria, hyperuricosuria, and hypocitraturia were noted in seven, three, six, and six patients, respectively. No patients were found to have gouty diathesis or developed cystine stones. Comparative metabolic analyses of patients with renal calculi in caliceal diverticula or normal kidneys revealed a distinct profile in patients with horseshoe kidneys, with a higher incidence of hypocitraturia. **CONCLUSIONS**: All patients with renal calculi in horseshoe kidneys were noted to have metabolic abnormalities predisposing to stone formation. In this initial series of 11 patients, hypovolemia, hypercalcuria and hypocitraturia were most common metabolic defects. These findings suggest that metabolic derangements play a role in stone formation in patients with a horseshoe kidney. Patients with calculi in anatomically abnormal kidneys should be considered for a metabolic evaluation to identify their stone-forming risk factors in order to initiate preventative selective medical therapy and reduce the risk of recurrent calculus formation.

**URL:** <https://ovidsp.ovid.com/ovidweb.cgi?T=JS&CSC=Y&NEWS=N&PAGE=fulltext&D=med5&DO=10.1089%2f089277904322959798> <https://libkey.io/libraries/2592/openurl?genre=article&sid=OVID:medline&id=pmid:15072623&id=doi:10.1089%2F089277904322959798&issn=0892-7790&isbn=&volume=18&issue=2&spage=157&pages=157-61&date=2004&title=Journal+of+Endourology&atitle=Metabolic+abnormalities+associated+with+renal+calculi+in+patients+with+horseshoe+kidneys.&aulast=Raj&pid=%3Cauthor%3ERaj+GV%3BAuge+BK%3BAssimos+D%3BPreminger+GM%3C%2Fauthor%3E%3CAN%3E15072623%3C%2FAN%3E%3CDT%3EJournal+Article%3C%2FDT%3E>

197. **Effects of calcium supplements on the risk of renal stone formation in a population with low oxalate intake.**

**Item Type:** Journal Article

**Authors:** Stitchantrakul, Wasana;Sopassathit, Wichai;Prapaipanich, Surasing and Domrongkitchaiporn, Somnuek

**Publication Date:** De ,2004

**Journal:** Southeast Asian Journal of Tropical Medicine & Public Health 35(4), pp. 1028–1033

**Abstract:** It has been speculated that calcium supplement in subjects with low oxalate intake might increase the risk of calcium stone formation due to an increase in calcium absorption without a significant reduction in oxalate absorption. There have been no human studies addressing specifically the effects of taking calcium supplements in populations whose dietary oxalate is low. This study was conducted to determine the effects of calcium supplements on the risk of calcium stone formation in a population with low oxalate intake. Thirty-two healthy male navy privates, 22.7 +/- 1.9 (mean +/- SD) years old, who had oxalate intake of less than 1 mmol/day, a serum creatinine of less than 150 micromol/l, and no history of renal stones, participated in the study. Dietary oxalate was controlled to be under 1 mmol/day throughout the study. Twenty-four hour urine collections for the determination of urinary constituents were obtained at baseline and after taking calcium supplements. Detection of calcium oxalate was performed to assess the risk of calcium oxalate stone formation. The urinary excretion of calcium was significantly elevated above baseline values while taking the calcium supplements (3.48 +/- 2.13 vs 5.17 +/- 2.61 mmol/d, p < 0.05) and urinary oxalate was significantly decreased when the subjects took calcium supplements compared to the corresponding baseline value (0.13 +/- 0.05 vs 0.17 +/- 0.07 mmol/d, p = 0.01). Urinary citrate was significantly elevated when the subjects took calcium supplements compared to the baseline (0.83 +/- 0.57 vs 0.64 +/- 0.39 mmol/d, p = 0.03). There was no significant alteration in the activity products of calcium oxalate while taking the calcium supplements (0.54 +/- 0.25 vs 0.57 +/- 0.22, p = 0.54). The effect of calcium supplements with meals, for the reduction of the risk of calcium stone formation, was unchanged, even in a population whose oxalate intake is rather low. Taking calcium supplements resulted in a reduction in urinary oxalates and an elevation in urinary citrates. Both alterations in urinary constituents counterbalanced the elevation in urinary calcium which resulted from the calcium supplements.

**URL:** <https://ovidsp.ovid.com/ovidweb.cgi?T=JS&CSC=Y&NEWS=N&PAGE=fulltext&D=med5&AN=15916110> <https://libkey.io/libraries/2592/openurl?genre=article&sid=OVID:medline&id=pmid:15916110&id=doi:&issn=0125-1562&isbn=&volume=35&issue=4&spage=1028&pages=1028-33&date=2004&title=Southeast+Asian+Journal+of+Tropical+Medicine+%26+Public+Health&atitle=Effects+of+calcium+supplements+on+the+risk+of+renal+stone+formation+in+a+population+with+low+oxalate+intake.&aulast=Stitchantrakul&pid=%3Cauthor%3EStitchantrakul+W%3BSopassathit+W%3BPrapaipanich+S%3BDomrongkitchaiporn+S%3C%2Fauthor%3E%3CAN%3E15916110%3C%2FAN%3E%3CDT%3EClinical+Trial%3C%2FDT%3E>

198. **Effects of calcium carbonate, magnesium oxide and sodium citrate bicarbonate health supplements on the urinary risk factors for kidney stone formation.**

**Item Type:** Journal Article

**Authors:** Allie, Shameez and Rodgers, Allen

**Publication Date:** Ja ,2003

**Journal:** Clinical Chemistry & Laboratory Medicine 41(1), pp. 39–45

**Abstract:** We describe a model to illustrate different chemical interactions that can occur in urine following ingestion of individual and combined health supplements. Two types of interactions are defined: synergism and addition. The model was applied to eight healthy males who participated in a study to investigate the chemical interactions between calcium carbonate, magnesium oxide and sodium citrate-bicarbonate health supplements on calcium oxalate urinary stone risk factors. Subjects ingested these components individually and in combination for 7 days. Twenty-four-hour urines were collected at baseline and during the final day of supplementation. These were analysed using standard laboratory techniques. Three different chemical interactions, all involving citrate, were identified: magnesium and citrate exerted a synergistic effect on lowering the relative superaturation (RS) of brushite; the same two components produced a synergistic effect on raising pH; finally, calcium and citrate exerted an additive effect on lowering the RS of uric acid. We propose that the novel approach described in this paper allows for the evaluation of individual, additive and synergistic interactions in the assessment of the efficacy of stone-risk reducing preparations.

**URL:** <https://ovidsp.ovid.com/ovidweb.cgi?T=JS&CSC=Y&NEWS=N&PAGE=fulltext&D=med5&DO=10.1515%2fCCLM.2003.008> <https://libkey.io/libraries/2592/openurl?genre=article&sid=OVID:medline&id=pmid:12636048&id=doi:10.1515%2FCCLM.2003.008&issn=1434-6621&isbn=&volume=41&issue=1&spage=39&pages=39-45&date=2003&title=Clinical+Chemistry+%26+Laboratory+Medicine&atitle=Effects+of+calcium+carbonate%2C+magnesium+oxide+and+sodium+citrate+bicarbonate+health+supplements+on+the+urinary+risk+factors+for+kidney+stone+formation.&aulast=Allie&pid=%3Cauthor%3EAllie+S%3BRodgers+A%3C%2Fauthor%3E%3CAN%3E12636048%3C%2FAN%3E%3CDT%3EComparative+Study%3C%2FDT%3E>

199. **Effect of vitamin C supplements on urinary oxalate and pH in calcium stone-forming patients.**

**Item Type:** Journal Article

**Authors:** Baxmann, Alessandra Calabria;De O G Mendonca, Claudia and Heilberg, Ita Pfeferman

**Publication Date:** Ma ,2003

**Journal:** Kidney International 63(3), pp. 1066–1071

**Abstract: BACKGROUND**: The contribution of ascorbate to urinary oxalate is controversial. The present study aimed to determine whether urinary oxalate and pH may be affected by vitamin C supplementation in calcium stone-forming patients. **METHODS**: Forty-seven adult calcium stone-forming patients received either 1 g (N=23) or 2 g (N=24) of vitamin C supplement for 3 days and 20 healthy subjects received 1 g. A 24-hour urine sample was obtained both before and after vitamin C for calcium, oxalate, magnesium, citrate, sodium, potassium, and creatinine determination. The Tiselius index was used as a calcium oxalate crystallization index. A spot fasting morning urine sample was also obtained to determine the urinary pH before and after vitamin C. **RESULTS**: Fasting urinary pH did not change after 1 g (5.8 +/- 0.6 vs. 5.8 +/- 0.7) or 2 g vitamin C (5.8 +/- 0.8 vs. 5.8 +/- 0.7). A significant increase in mean urinary oxalate was observed in calcium stone-forming patients receiving either 1 g (50 +/- 16 vs. 31 +/- 12 mg/24 hours) or 2 g (48 +/- 21 vs. 34 +/- 12 mg/24 hours) of vitamin C and in healthy subjects (25 +/- 12 vs. 39 +/- 13 mg/24 hours). A significant increase in mean Tiselius index was observed in calcium stone-forming patients after 1 g (1.43 +/- 0.70 vs. 0.92 +/- 0.65) or 2 g vitamin C (1.61 +/- 1.05 vs. 0.99 +/- 0.55) and in healthy subjects (1.50 +/- 0.69 vs. 0.91 +/- 0.46). Ancillary analyses of spot urine obtained after vitamin C were performed in 15 control subjects in vessels with or without ethylenediaminetetraacetic acid (EDTA) with no difference in urinary oxalate between them (28 +/- 23 vs. 26 +/- 21 mg/L), suggesting that the in vitro conversion of ascorbate to oxalate did not occur. **CONCLUSION**: These data suggest that vitamin C supplementation may increase urinary oxalate excretion and the risk of calcium oxalate crystallization in calcium stone-forming patients.

**URL:** <https://ovidsp.ovid.com/ovidweb.cgi?T=JS&CSC=Y&NEWS=N&PAGE=fulltext&D=med5&DO=10.1046%2fj.1523-1755.2003.00815.x> <https://libkey.io/libraries/2592/openurl?genre=article&sid=OVID:medline&id=pmid:12631089&id=doi:10.1046%2Fj.1523-1755.2003.00815.x&issn=0085-2538&isbn=&volume=63&issue=3&spage=1066&pages=1066-71&date=2003&title=Kidney+International&atitle=Effect+of+vitamin+C+supplements+on+urinary+oxalate+and+pH+in+calcium+stone-forming+patients.&aulast=Baxmann&pid=%3Cauthor%3EBaxmann+AC%3BDe+O+G+Mendonca+C%3BHeilberg+IP%3C%2Fauthor%3E%3CAN%3E12631089%3C%2FAN%3E%3CDT%3EClinical+Trial%3C%2FDT%3E>

200. **Effects of an oxalate load on urinary oxalate excretion in calcium stone formers.**

**Item Type:** Journal Article

**Authors:** de O G Mendonca, Claudia;Martini, Ligia Araujo;Baxmann, Alessandra Calabria;Nishiura, Jose Luiz;Cuppari, Lilian;Sigulem, Dirce Maria and Heilberg, Ita Pfeferman

**Publication Date:** Ja ,2003

**Journal:** Journal of Renal Nutrition 13(1), pp. 39–46

**Abstract: OBJECTIVE**: To investigate the oxalate intake and the effect of an oxalate load on urinary oxalate excretion in calcium stone-forming (CSF) patients. **DESIGN**: Prospective study. **SETTING**: University-affiliated outpatient Renal Lithiasis Unit. **PATIENTS AND CONTROLS**: Seventy (70) CSF and 41 healthy subjects (HS) collected a 24-hour urine sample and were submitted to a 3-day dietary record to determine mean oxalate (Ox), calcium (Ca) and vitamin C intake. Fifty-eight (58) CSF patients were randomly selected to receive milk (N = 28) or dark (N = 30) chocolate as an oxalate load. **INTERVENTION**: Administration of either milk (94 mg Ox + 430 mg Ca) or dark chocolate (94 mg Ox + 26 mg Ca) for 3 days. A 24-hour urine sample was obtained before and after the load to determine calcium, oxalate, sodium, potassium, urea, and creatinine. **MAIN OUTCOME MEASURE**: Oxalate intake and excretion. **RESULTS**: CSF patients presented mean Ox intake of 98 +/- 137 mg/d, similar to that of HS (108 +/- 139 mg/d). Mean Ox and vitamin C intake was directly correlated with Ox excretion only in CSF. The consumption of dark chocolate induced a significant increase in mean urinary Ox (36 +/- 14 versus 30 +/- 10 mg/24 hr) not observed in the milk chocolate group. Thus, a 2-fold increase in Ox intake in this population of CSF patients produced a significant 20% increase in oxaluria, not observed when Ca was consumed simultaneously. **CONCLUSION**: The present study suggests that even small increases in Ox intake affect oxalate excretion and the mitigation of urinary oxalate increase by Ca consumption reinforces that Ca and Ox intakes for CSF patients should be in balance. Further studies are necessary to assess whether or not a 20% increase in oxaluria will lead to a higher risk of stone formation. Copyright 2003 by the National Kidney Foundation, Inc.

**URL:** <https://ovidsp.ovid.com/ovidweb.cgi?T=JS&CSC=Y&NEWS=N&PAGE=fulltext&D=med5&DO=10.1053%2fjren.2003.50002> <https://libkey.io/libraries/2592/openurl?genre=article&sid=OVID:medline&id=pmid:12563622&id=doi:10.1053%2Fjren.2003.50002&issn=1051-2276&isbn=&volume=13&issue=1&spage=39&pages=39-46&date=2003&title=Journal+of+Renal+Nutrition&atitle=Effects+of+an+oxalate+load+on+urinary+oxalate+excretion+in+calcium+stone+formers.&aulast=de+O+G+Mendonca&pid=%3Cauthor%3Ede+O+G+Mendonca+C%3BMartini+LA%3BBaxmann+AC%3BNishiura+JL%3BCuppari+L%3BSigulem+DM%3BHeilberg+IP%3C%2Fauthor%3E%3CAN%3E12563622%3C%2FAN%3E%3CDT%3EClinical+Trial%3C%2FDT%3E>

201. **Influence of urine pH and citrate concentration on the upper limit of metastability for calcium phosphate.**

**Item Type:** Journal Article

**Authors:** Greischar, Amy;Nakagawa, Y. and Coe, Frederic L.

**Publication Date:** Ma ,2003

**Journal:** Journal of Urology 169(3), pp. 867–870

**Abstract: PURPOSE**: We determined the effects on the urine upper limit of metastability for calcium phosphate of citrate concentration and pH, and achievement of the upper limit of metastability by adding calcium or phosphate. **MATERIALS AND METHODS**: The citrate concentration in aliquots of 24-hour urine samples from normal males without a history of kidney stones was increased. The upper limit of metastability was determined by the point of visible crystal formation, as confirmed by increased optical density at 620 nm. when calcium or pH was increased. In additional experiments the upper limit of metastability was determined by adding calcium or phosphate at pH 5.9 and 6.4. **RESULTS**: Regardless of how the upper limit of metastability was achieved increasing the citrate concentration increased the former value by about 0.4 units per mM. citrate per l. The upper limit of metastability achieved in a given urine sample by adding phosphate or calcium did not differ. Increasing urine pH increased the upper limit of metastability. **CONCLUSIONS**: Treatment with alkaline citrate salts may decrease stone formation via an increase in calcium phosphate upper limit of metastability by increasing urine citrate and by directly affecting increased pH.

**URL:** <https://ovidsp.ovid.com/ovidweb.cgi?T=JS&CSC=Y&NEWS=N&PAGE=fulltext&D=med5&DO=10.1097%2f01.ju.0000049801.32639.ff> <https://libkey.io/libraries/2592/openurl?genre=article&sid=OVID:medline&id=pmid:12576801&id=doi:10.1097%2F01.ju.0000049801.32639.ff&issn=0022-5347&isbn=&volume=169&issue=3&spage=867&pages=867-70&date=2003&title=Journal+of+Urology&atitle=Influence+of+urine+pH+and+citrate+concentration+on+the+upper+limit+of+metastability+for+calcium+phosphate.&aulast=Greischar&pid=%3Cauthor%3EGreischar+A%3BNakagawa+Y%3BCoe+FL%3C%2Fauthor%3E%3CAN%3E12576801%3C%2FAN%3E%3CDT%3EJournal+Article%3C%2FDT%3E>

202. **Lipid peroxidation and its correlations with urinary levels of oxalate, citric acid, and osteopontin in patients with renal calcium oxalate stones.**

**Item Type:** Journal Article

**Authors:** Huang, Ho-Shiang;Ma, Ming-Chieh;Chen, Chau-Fong and Chen, Jun

**Publication Date:** De ,2003

**Journal:** Urology 62(6), pp. 1123–1128

**Abstract: OBJECTIVES**: To determine whether lipid peroxidation plays a role in patients with calcium oxalate kidney stones and to determine the correlation of lipid peroxidation with tubular damage and the major urinary risk factors. We also used the isoenzymes of glutathione S-transferase (GST) to examine which parts of the renal tubules were injured in patients with renal stones. **METHODS**: This clinical study included two study groups. Group 1 included 32 normal volunteers, and group 2 included 32 patients with calcium oxalate kidney stones. A 24-hour urine sample was collected from each subject, and the levels of Ca, P, Mg, oxalate, citrate, N-acetyl-beta-glucosaminidase (NAG), beta-galactosidase (GAL), alphaGST, piGST, osteopontin (OPN), thiobarbituric acid-reactive substances (TBARS), and malondialdehyde (MDA) were examined. **RESULTS**: Hyperoxaluria, hypocitraturia, and low urinary OPN were the major abnormalities found in the patients with stones. Elevated urinary alphaGST, NAG, and GAL were also noted in the patients with stones; however, urinary piGST showed no statistically significant difference compared with the controls. Urinary TBARS and MDA had statistically significant correlations with alphaGST, GAL, NAG, Ca, and oxalate, but had no correlation with piGST, citrate, OPN, Mg, and P. Urinary citrate had a negative, linear, and statistically significant correlation with alphaGST, GAL, and NAG. **CONCLUSIONS**: Lipid peroxidation correlated with hyperoxaluria and renal tubular damage, indicating that hyperoxaluria can induce tubular cell injury and that this injury may be due to the production of free radicals in patients with calcium oxalate stones. Renal tubular damage in patients with stones may be limited to the proximal tubules.

**URL:** <https://ovidsp.ovid.com/ovidweb.cgi?T=JS&CSC=Y&NEWS=N&PAGE=fulltext&D=med5&DO=10.1016%2fs0090-4295%2803%2900764-7> <https://libkey.io/libraries/2592/openurl?genre=article&sid=OVID:medline&id=pmid:14665375&id=doi:10.1016%2Fs0090-4295%2803%2900764-7&issn=0090-4295&isbn=&volume=62&issue=6&spage=1123&pages=1123-8&date=2003&title=Urology&atitle=Lipid+peroxidation+and+its+correlations+with+urinary+levels+of+oxalate%2C+citric+acid%2C+and+osteopontin+in+patients+with+renal+calcium+oxalate+stones.&aulast=Huang&pid=%3Cauthor%3EHuang+HS%3BMa+MC%3BChen+CF%3BChen+J%3C%2Fauthor%3E%3CAN%3E14665375%3C%2FAN%3E%3CDT%3EJournal+Article%3C%2FDT%3E>

203. **Urine calcium and volume predict coverage of renal papilla by Randall's plaque.**

**Item Type:** Journal Article

**Authors:** Kuo, Ramsay L.;Lingeman, James E.;Evan, Andrew P.;Paterson, Ryan F.;Parks, Joan H.;Bledsoe, Sharon B.;Munch, Larry C. and Coe, Fredric L.

**Publication Date:** De ,2003

**Journal:** Kidney International 64(6), pp. 2150–2154

**Abstract: BACKGROUND**: Renal papillary plaques are common in calcium stone formers. We hypothesized that plaque should increase directly with urine calcium excretion, and inversely with urine volume. To test this, we measured papillary plaque areas in both idiopathic calcium oxalate stone formers and nonstone formers and examined 24-hour urine data to identify significant correlations. **METHODS**: Fourteen stone formers and four nonstone forming controls underwent papillary mapping with flexible nephroscopy. For each papillum, representative still images and moving pictures expert group (MPEG) movies were used to identify plaque extent and papillary borders. The mean fractional plaque coverage for each polar region (upper, inter, lower) and per papillum was calculated. The relationship of the plaque coverage data to urine measurements was assessed with general multivariate linear modeling. **RESULTS**: Mean polar fractional plaque coverage was higher in the calcium oxalate stone formers (7.4% vs. 0.5%, P= 0.012) as was mean fractional plaque per papillum (7.6% vs. 0.6%, P= 0.011). When correlating mean polar plaque coverage to urine data, urine volume and calcium excretion were the only measurements with independent relationships to plaque (P= 0.002, adjusted multiple R2= 0.521), with higher calcium and lower volume increasing coverage. The same relationships hold for mean plaque per papillum, except that urine pH also becomes an independent factor (P= 0.001, adjusted multiple R2= 0.606). **CONCLUSION**: Utilizing advanced digital video and endoscopic equipment, we have achieved the most accurate estimation of papillary plaque coverage to date. Our findings support the idea that urine volume and calcium are the main correlates of plaque coverage.

**URL:** <https://ovidsp.ovid.com/ovidweb.cgi?T=JS&CSC=Y&NEWS=N&PAGE=fulltext&D=med5&DO=10.1046%2fj.1523-1755.2003.00316.x> <https://libkey.io/libraries/2592/openurl?genre=article&sid=OVID:medline&id=pmid:14633137&id=doi:10.1046%2Fj.1523-1755.2003.00316.x&issn=0085-2538&isbn=&volume=64&issue=6&spage=2150&pages=2150-4&date=2003&title=Kidney+International&atitle=Urine+calcium+and+volume+predict+coverage+of+renal+papilla+by+Randall%27s+plaque.&aulast=Kuo&pid=%3Cauthor%3EKuo+RL%3BLingeman+JE%3BEvan+AP%3BPaterson+RF%3BParks+JH%3BBledsoe+SB%3BMunch+LC%3BCoe+FL%3C%2Fauthor%3E%3CAN%3E14633137%3C%2FAN%3E%3CDT%3EJournal+Article%3C%2FDT%3E>

204. **Influence of urinary stones on the composition of a 24-hour urine sample.**

**Item Type:** Journal Article

**Authors:** Laube, Norbert;Pullmann, Michael;Hergarten, Stefan and Hesse, Albrecht

**Publication Date:** Fe ,2003

**Journal:** Clinical Chemistry 49(2), pp. 281–285

**Abstract: BACKGROUND**: It can be assumed that stones in the urinary tract continuously increase in size by incorporating material from urine. Consequently, urine will exhibit depleted concentrations of lithogenic constituents when urinary stones are present in the patient's urinary tract. **METHODS**: To calculate the influence of the depletion effect, we considered two different models of stone growth. In the first model, the increase in stone size depends only on the urinary concentration of a lithogenic substance; the second model also considers the surface area of the growing stone. The case of only one kidney being affected by stone formation is considered separately. We discuss example calculations involving the formation of calcium oxalate. **RESULTS**: The calculated depletion effects are of a nonnegligible order of magnitude. Assuming both a measured oxalate concentration of, e.g., 0.37 mmol/L and a reasonable in vivo stone growing rate of 10 mm(3)/day, a relative underestimation of the real "in situ" oxalate concentration between approximately 21% (model 1) and approximately 42% (model 2) occurs. The depletion effect increases markedly with increasing stone growth rate. **CONCLUSIONS**: Metabolic status can be evaluated correctly only in patients who have been declared "stone-free", e.g., after stone removal. Because the expected stone-related depletion effect in most cases is of high clinical relevance, we recommend estimating the effect of the order of magnitude of the depletion on actual urinary composition.

**URL:** <https://ovidsp.ovid.com/ovidweb.cgi?T=JS&CSC=Y&NEWS=N&PAGE=fulltext&D=med5&DO=10.1373%2f49.2.281> <https://libkey.io/libraries/2592/openurl?genre=article&sid=OVID:medline&id=pmid:12560352&id=doi:10.1373%2F49.2.281&issn=0009-9147&isbn=&volume=49&issue=2&spage=281&pages=281-5&date=2003&title=Clinical+Chemistry&atitle=Influence+of+urinary+stones+on+the+composition+of+a+24-hour+urine+sample.&aulast=Laube&pid=%3Cauthor%3ELaube+N%3BPullmann+M%3BHergarten+S%3BHesse+A%3C%2Fauthor%3E%3CAN%3E12560352%3C%2FAN%3E%3CDT%3EJournal+Article%3C%2FDT%3E>

205. **The etiology of urolithiasis in HIV infected patients.**

**Item Type:** Journal Article

**Authors:** Nadler, Robert B.;Rubenstein, Jonathan N.;Eggener, Scott E.;Loor, Michelle M. and Smith, Norm D.

**Publication Date:** Fe ,2003

**Journal:** Journal of Urology 169(2), pp. 475–477

**Abstract: PURPOSE**: It is commonly thought that urinary lithiasis in HIV infected patients on protease inhibitor therapy is composed primarily of the protease inhibitor itself. Since many HIV infected patients on protease inhibitors presenting to our institution had nonprotease inhibitor stones, we investigated potential underlying metabolic abnormalities that may account for the lithogenesis. **MATERIALS AND METHODS**: We retrospectively reviewed all HIV infected patients on protease inhibitors with renal colic and evidence of nephrolithiasis who presented to our institution between June 1996 and January 2001. Patients were evaluated for stone composition and metabolic abnormalities of blood and urine when possible. **RESULTS**: A total of 24 patients were identified, and all were or had been on protease inhibitors (indinavir 14, ritonavir 3, nelfnavir 2, unspecified 5). Of the 14 patients on indinavir only 4 (28.6%) had indinavir containing stones. The remaining stones in this group and in those not on indinavir contained various amounts of calcium oxalate monohydrate and dihydrate, ammonium acid urate and uric acid. Of 10 patients who underwent 24-hour urine collection for metabolic evaluation 8 (80%) had abnormalities, including hypocitraturia in 5, hyperoxaluria in 4, hypomagnesuria in 4, hypercalciuria in 3, increased supersaturation of calcium oxalate in 3 and hyperuricosuria in 2. Abnormalities in the levels of urinary phosphate and sodium were also observed. **CONCLUSIONS**: HIV infected patients form many types of stones, which probably are attributable to underlying metabolic abnormalities rather than the use of protease inhibitors. A complete metabolic evaluation is warranted in these patients, as a means of guiding treatment to prevent future stone episodes, while avoiding the need to alter antiretroviral regimens.

**URL:** <https://ovidsp.ovid.com/ovidweb.cgi?T=JS&CSC=Y&NEWS=N&PAGE=fulltext&D=med5&DO=10.1097%2f01.ju.0000046021.43205.d6> <https://libkey.io/libraries/2592/openurl?genre=article&sid=OVID:medline&id=pmid:12544290&id=doi:10.1097%2F01.ju.0000046021.43205.d6&issn=0022-5347&isbn=&volume=169&issue=2&spage=475&pages=475-7&date=2003&title=Journal+of+Urology&atitle=The+etiology+of+urolithiasis+in+HIV+infected+patients.&aulast=Nadler&pid=%3Cauthor%3ENadler+RB%3BRubenstein+JN%3BEggener+SE%3BLoor+MM%3BSmith+ND%3C%2Fauthor%3E%3CAN%3E12544290%3C%2FAN%3E%3CDT%3EJournal+Article%3C%2FDT%3E>

206. **Early changes of oxalate and calcium urine excretion in those with calcium oxalate stone formation after extracorporeal shock wave lithotripsy.**

**Item Type:** Journal Article

**Authors:** Oehlschlager, S.;Albrecht, S.;Hakenberg, O. W.;Schrodter, S.;Froehner, M.;Manseck, A. and Wirth, M. P.

**Publication Date:** Ju ,2003

**Journal:** Urology 62(1), pp. 17–21

**Abstract: OBJECTIVES**: To determine the extent of transient changes of tubular function in idiopathic calcium-oxalate (CaOx) stone-bearing patients after extracorporeal shock wave lithotripsy (ESWL), calcium and oxalate excretion were measured before and after ESWL. **METHODS**: In 22 patients with renal CaOx stones, the plasma values and urine excretion of creatinine, calcium, oxalate, magnesium, and citrate were measured before and on days 1 and 2 after ESWL under conditions of a standardized diet. Overnight urine collection for an 8-hour period was used to measure the urine excretion, and the values were extrapolated to a 24-hour period. For calculation of the urine ion activity, the AP(CaOx) index EQ(s) and the CaOx risk index were used. **RESULTS**: After ESWL, hyperoxaluria was noted in 10 patients compared with 2 before ESWL. Hypercalciuria was seen in 11 patients after ESWL compared with 3 before. Combined hyperoxaluria and hypercalciuria was found in 7 patients after ESWL compared with 1 before. Both the oxalate/creatinine and calcium/creatinine ratios were significantly increased after ESWL. The AP(CaOx) index EQ(s) and the CaOx risk index were significantly increased after ESWL in patients with increased post-ESWL calcium and/or oxalate excretion. **CONCLUSIONS**: Increased calcium and/or oxalate excretion can be seen in patients with CaOx stones early after ESWL. This increased excretion of lithogenic substances represents an increased risk of fragment apposition after ESWL for the group with a significantly increased AP(CaOx) index EQ(s) and CaOx risk index. Therefore, prophylactic measures in patients at risk early after ESWL might be warranted to prevent possible recurrent stone formation.

**URL:** <https://ovidsp.ovid.com/ovidweb.cgi?T=JS&CSC=Y&NEWS=N&PAGE=fulltext&D=med5&DO=10.1016%2fs0090-4295%2803%2900122-5> <https://libkey.io/libraries/2592/openurl?genre=article&sid=OVID:medline&id=pmid:12837412&id=doi:10.1016%2Fs0090-4295%2803%2900122-5&issn=0090-4295&isbn=&volume=62&issue=1&spage=17&pages=17-21&date=2003&title=Urology&atitle=Early+changes+of+oxalate+and+calcium+urine+excretion+in+those+with+calcium+oxalate+stone+formation+after+extracorporeal+shock+wave+lithotripsy.&aulast=Oehlschlager&pid=%3Cauthor%3EOehlschlager+S%3BAlbrecht+S%3BHakenberg+OW%3BSchrodter+S%3BFroehner+M%3BManseck+A%3BWirth+MP%3C%2Fauthor%3E%3CAN%3E12837412%3C%2FAN%3E%3CDT%3EComparative+Study%3C%2FDT%3E>

207. **Thiazide does not affect urine oxalate excretion.**

**Item Type:** Journal Article

**Authors:** Parks, Joan H. and Coe, Frederic L.

**Publication Date:** Au ,2003

**Journal:** Journal of Urology 170(2 Pt 1), pp. 393–396

**Abstract: PURPOSE**: We assessed the effects of thiazide treatment on urine oxalate excretion in patients with kidney stones. **MATERIALS AND METHODS**: Of 537 patients (231 women), 209 (81 women) received thiazide for stone prevention. Urine oxalate was measured in 3, 24-hour urines before treatment, and in 1, 24-hour urine after 6 to 12 weeks of treatment. This was an observational cohort and treatment was not randomized. **RESULTS**: Urine oxalate increased in general. There was no difference in increase with or without thiazide, whether one considered simple t test comparisons or used ANOVA with pretreatment oxalate excretion as a covariate. **CONCLUSIONS**: Thiazide administration exerts no measurable effect on urine oxalate excretion that can be detected in clinical practice.

**URL:** <https://ovidsp.ovid.com/ovidweb.cgi?T=JS&CSC=Y&NEWS=N&PAGE=fulltext&D=med5&DO=10.1097%2f01.ju.0000076371.92938.9b> <https://libkey.io/libraries/2592/openurl?genre=article&sid=OVID:medline&id=pmid:12853783&id=doi:10.1097%2F01.ju.0000076371.92938.9b&issn=0022-5347&isbn=&volume=170&issue=2&spage=393&pages=393-6&date=2003&title=Journal+of+Urology&atitle=Thiazide+does+not+affect+urine+oxalate+excretion.&aulast=Parks&pid=%3Cauthor%3EParks+JH%3BCoe+FL%3C%2Fauthor%3E%3CAN%3E12853783%3C%2FAN%3E%3CDT%3EClinical+Trial%3C%2FDT%3E>

208. **Durability of the medical management of cystinuria.**

**Item Type:** Journal Article

**Authors:** Pietrow, Paul K.;Auge, Brian K.;Weizer, Alon Z.;Delvecchio, Fernando C.;Silverstein, Ari D.;Mathias, Barbara;Albala, David M. and Preminger, Glenn M.

**Publication Date:** Ja ,2003

**Journal:** Journal of Urology 169(1), pp. 68–70

**Abstract: PURPOSE**: Cystinuria is an autosomal recessive disorder of dibasic amino acid transport in the kidney that leads to an abundance of cystine in the urine. This molecule is poorly soluble in urine and it is prone to crystallization and stone formation at concentrations above 300 mg./l. Medical treatment in these patients has incorporated increasing urine volumes, alkalinization and thiol medications that decrease the availability of free cystine in urine. Despite a reasonable prognosis for reduced stone formation we and others have noted difficulties in patients complying with medical management recommendations. Therefore, we evaluated the durability of treatment success in our patients with cystinuria. **MATERIALS AND METHODS**: A retrospective chart review was performed in all patients with cystinuria referred to the comprehensive kidney stone center at our institution for an 8-year period. Medical therapy, stone recurrence rates, compliance with medications and scheduled followup, and the results of metabolic evaluations via 24-hour urine collections were reviewed. The average concentrations of urinary cystine in initial and followup 24-hour samples were compared in patients compliant and noncompliant with medical treatment. In addition, each patient was mailed a 1-page questionnaire to assess the self-perception of medical compliance. **RESULTS**: We identified 26 patients with a mean age of 32 years at referral (range 13 to 67) who were followed an average of 38.2 months (range 6 to 83). Females represented 58% of those with cystinuria. Overall compliance with medical recommendations was poor with a short duration of success. Of the 26 patients followed at our stone center only 4 (15%) achieved and maintained therapeutic success, as defined by urine cystine less than 300 mg./l. An additional 11 patients (42%) achieved therapeutic success but subsequently had failure at an average of 16 months (range 6 to 27). Of these patients 7 (64%) regained therapeutic success at an average of 9.4 months (range 4 to 20). Five patients (19%) never achieved therapeutic success, while an additional 6 (23%) failed to present to followup appointments or provide subsequent 24-hour urine studies despite referral to a tertiary care center. Patient self-assessment of medical compliance was uniformly high regardless of physician perceptions or treatment results. CONCLUSIONS The durability of medically treating patients with cystinuria is limited with only a small percent able to achieve and maintain the goal of decreasing cystine below the saturation concentration. Greater physician vigilance in these complicated stone formers is required to achieve successful prophylactic management. Furthermore, these patients require better insight into the own disease to improve compliance.

**URL:** <https://ovidsp.ovid.com/ovidweb.cgi?T=JS&CSC=Y&NEWS=N&PAGE=fulltext&D=med5&DO=10.1016%2fS0022-5347%2805%2964037-2> <https://libkey.io/libraries/2592/openurl?genre=article&sid=OVID:medline&id=pmid:12478105&id=doi:10.1016%2FS0022-5347%2805%2964037-2&issn=0022-5347&isbn=&volume=169&issue=1&spage=68&pages=68-70&date=2003&title=Journal+of+Urology&atitle=Durability+of+the+medical+management+of+cystinuria.&aulast=Pietrow&pid=%3Cauthor%3EPietrow+P%3BAuge+BK%3BWeizer+AZ%3BDelvecchio+FC%3BSilverstein+AD%3BMathias+B%3BAlbala+DM%3BPreminger+GM%3C%2Fauthor%3E%3CAN%3E12478105%3C%2FAN%3E%3CDT%3EJournal+Article%3C%2FDT%3E>

209. **Influence of gender and age on calcium oxalate crystal growth inhibition by urine from relatives of stone forming patients.**

**Item Type:** Journal Article

**Authors:** Bergsland, Kristin J.;Kinder, Jennifer M.;Asplin, John R.;Coe, Brian J. and Coe, Fredric L.

**Publication Date:** Ju ,2002

**Journal:** Journal of Urology 167(6), pp. 2372–2376

**Abstract: PURPOSE**: We define the relationships between urine inhibition of calcium oxalate crystal growth and age, gender, urine chemistries and stone formation among relatives of calcium stone forming patients. **MATERIALS AND METHODS**: We collected 24-hour urine samples from 366 first degree relatives of calcium stone formers. Calcium oxalate crystal growth inhibition was studied using a constant amount of dialyzed urine protein in a seeded crystallization system. Standard stone risk measurements were also performed on the urine, including supersaturation for calcium oxalate, calcium phosphate and uric acid. **RESULTS**: By multivariate analysis crystal growth inhibition is strongly inversely related to the amount of protein excreted per day, and the age of the subject. When corrected for protein excretion and age, urine proteins from nonstone forming male subjects inhibited crystal growth more strongly than those from corresponding female subjects. Among stone formers the sex difference was not present. **CONCLUSIONS**: Inhibition of calcium oxalate crystal growth is influenced by a complex combination of gender, age, stone formation and assay conditions. The effect of daily protein excretion is most likely a consequence of using a fixed amount of urine protein per assay. The influence of age is significant and unexplained, with the urine of young people (less than 20 years) demonstrating a vigorous ability to inhibit crystallization. In addition, the urine of nonstone forming male relatives appears to have a greater ability to inhibit crystallization than that of nonstone forming female relatives. Further use of this assay in clinical investigations must take age and gender into proper account.

**URL:** <https://ovidsp.ovid.com/ovidweb.cgi?T=JS&CSC=Y&NEWS=N&PAGE=fulltext&D=med4&AN=11992040> <https://libkey.io/libraries/2592/openurl?genre=article&sid=OVID:medline&id=pmid:11992040&id=doi:&issn=0022-5347&isbn=&volume=167&issue=6&spage=2372&pages=2372-6&date=2002&title=Journal+of+Urology&atitle=Influence+of+gender+and+age+on+calcium+oxalate+crystal+growth+inhibition+by+urine+from+relatives+of+stone+forming+patients.&aulast=Bergsland&pid=%3Cauthor%3EBergsland+KJ%3BKinder+JM%3BAsplin+JR%3BCoe+BJ%3BCoe+FL%3C%2Fauthor%3E%3CAN%3E11992040%3C%2FAN%3E%3CDT%3EJournal+Article%3C%2FDT%3E>

210. **Vitamin D receptor gene polymorphism and bone mineral density in hypercalciuric calcium-stone-forming patients.**

**Item Type:** Journal Article

**Authors:** Heilberg, Ita Pfeferman;Teixeira, Sergio Henrique;Martini, Ligia Araujo and Boim, Mirian Aparecia

**Publication Date:** Ja ,2002

**Journal:** Nephron 90(1), pp. 51–57

**Abstract:** Reduced bone mineral density (BMD) and an increased risk of vertebral fracture have been reported in calcium-stone-forming (CSF) patients presenting with idiopathic hypercalciuria. We investigated the association between BsmI vitamin D receptor (VDR) polymorphism and BMD in 68 hypercalciuric CSF patients (35 males and 33 premenopausal females, mean age +/- SD = 39 +/- 10 years). BMD was measured at lumbar spine (L2-L4) and femur neck sites using dual energy X-ray absorptiometry. A 72-hour dietary record and a 24-hour urine sample were obtained from each patient to determine calcium intake and excretion. The allelic frequency found for the sample as a whole was 16% BB, 44% Bb and 40% bb. Mean BMD values did not significantly differ among BB, Bb and bb patients at L2-L4 (1.162 +/- 0.10, 1.133 +/- 0.11 and 1.194 +/- 0.19 g/cm2, mean +/- SD, respectively) or at neck sites (0.920 +/- 0.11, 0.931 +/- 0.15 and 0.982 +/- 0.15 g/cm2, respectively). Calcium intake and excretion were also not significantly different among the three genotypes. Patients were then divided into two groups, normal BMD, T-score > or =-1 (n = 34) and low BMD, T-score <-1 (n = 34), to further evaluate the allele influence on previous bone loss. Despite a trend for a higher mean BMD at spine or neck sites for patients with one or two b alleles when compared to BB patients, the difference did not reach statistical significance. The distribution of BB, Bb and bb genotypes in the low-bone-mass group (15, 47 and 38%, respectively) was similar to that in the normal-bone-mass group (18, 41 and 14%, respectively). These data suggest that BsmI VDR polymorphism does not play an important role in the bone loss seen in hypercalciuric CSF patients. Copyright 2002 S. Karger AG, Basel

**URL:** <https://ovidsp.ovid.com/ovidweb.cgi?T=JS&CSC=Y&NEWS=N&PAGE=fulltext&D=med4&DO=10.1159%2f000046314> <https://libkey.io/libraries/2592/openurl?genre=article&sid=OVID:medline&id=pmid:11744805&id=doi:10.1159%2F000046314&issn=1660-8151&isbn=&volume=90&issue=1&spage=51&pages=51-7&date=2002&title=Nephron&atitle=Vitamin+D+receptor+gene+polymorphism+and+bone+mineral+density+in+hypercalciuric+calcium-stone-forming+patients.&aulast=Heilberg&pid=%3Cauthor%3EHeilberg+IP%3BTeixeira+SH%3BMartini+LA%3BBoim+MA%3C%2Fauthor%3E%3CAN%3E11744805%3C%2FAN%3E%3CDT%3EJournal+Article%3C%2FDT%3E>

211. **Etiological role of estrogen status in renal stone formation.**

**Item Type:** Journal Article

**Authors:** Heller, Howard J.;Sakhaee, Khashayar;Moe, Orson W. and Pak, Charles Y. C.

**Publication Date:** No ,2002

**Journal:** Journal of Urology 168(5), pp. 1923–1927

**Abstract: PURPOSE**: Estrogen may protect against kidney stone formation since nephrolithiasis is more common in men than in women. Moreover, the incidence of stones rises after menopause in women. We examined the contribution of estrogen to kidney stone risk by comparing outpatient evaluations in the 2 genders, and in estrogen treated and untreated postmenopausal women. **MATERIALS AND METHODS**: We reviewed the results of the initial evaluation of 1,454 adult calcium oxalate stone formers, including 1,050 men and 404 women. Of the postmenopausal women 39 and 50 were estrogen treated and untreated, respectively. Samples of urine and blood were collected 1 week after the imposition of a diet restricted moderately in sodium and calcium, and modestly in oxalate and animal protein. **RESULTS**: Compared with men the daily excretion of urinary calcium, oxalate and uric acid was lower in women. Women had lower saturations of calcium oxalate and brushite as well as lower excretion of undissociated uric acid. Compared with men urinary calcium was lower in women until age 50 years, when it equaled that of men. Citrate was equal in the genders until the age 60 years, when it tended to decrease in women. Compared with nontreated postmenopausal women those treated with estrogen had lower mean 24-hour calcium plus or minus SD (155 +/- 62 versus 193 +/- 90 mg. per day, p : Compared with men the daily excretion of urinary calcium, oxalate and uric acid was lower in women. Women had lower saturations of calcium oxalate and brushite as well as lower excretion of undissociated uric acid. Compared with men urinary calcium was lower in women until age 50 years, when it equaled that of men. Citrate was equal in the genders until the age 60 years, when it tended to decrease in women. Compared with nontreated postmenopausal women those treated with estrogen had lower mean 24-hour calcium plus or minus SD (155 +/- 62 versus 193 +/- 90 mg. per day, p : Compared with men the daily excretion of urinary calcium, oxalate and uric acid was lower in women. Women had lower saturations of calcium oxalate and brushite as well as lower excretion of undissociated uric acid. Compared with men urinary calcium was lower in women until age 50 years, when it equaled that of men. Citrate was equal in the genders until the age 60 years, when it tended to decrease in women. Compared with nontreated postmenopausal women those treated with estrogen had lower mean 24-hour calcium plus or minus SD (155 +/- 62 versus 193 +/- 90 mg. per day, p : Compared with men the daily excretion of urinary calcium, oxalate and uric acid was lower in women. Women had lower saturations of calcium oxalate and brushite as well as lower excretion of undissociated uric acid. Compared with men urinary calcium was lower in women until age 50 years, when it equaled that of men. Citrate was equal in the genders until the age 60 years, when it tended to decrease in women. Compared with nontreated postmenopausal women those treated with estrogen had lower mean 24-hour calcium plus or minus SD (155 +/- 62 versus 193 +/- 90 mg. per day, p **CONCLUSIONS**: The lower risk of stone formation in women may be due to the lower urinary saturation of stone forming salts. Estrogen treatment may decrease the risk of stone recurrence in postmenopausal women by lowering urinary calcium and calcium oxalate saturation.

**URL:** <https://ovidsp.ovid.com/ovidweb.cgi?T=JS&CSC=Y&NEWS=N&PAGE=fulltext&D=med4&DO=10.1016%2fS0022-5347%2805%2964264-4> <https://libkey.io/libraries/2592/openurl?genre=article&sid=OVID:medline&id=pmid:12394677&id=doi:10.1016%2FS0022-5347%2805%2964264-4&issn=0022-5347&isbn=&volume=168&issue=5&spage=1923&pages=1923-7&date=2002&title=Journal+of+Urology&atitle=Etiological+role+of+estrogen+status+in+renal+stone+formation.&aulast=Heller&pid=%3Cauthor%3EHeller+HJ%3BSakhaee+K%3BMoe+OW%3BPak+CY%3C%2Fauthor%3E%3CAN%3E12394677%3C%2FAN%3E%3CDT%3EJournal+Article%3C%2FDT%3E>

212. **Urinary stone risk factors in the siblings of patients with calcium renal stones.**

**Item Type:** Journal Article

**Authors:** Kinder, Jennifer M.;Clark, Cyndi D.;Coe, Brian J.;Asplin, John R.;Parks, Joan H. and Coe, Fredric L.

**Publication Date:** Ma ,2002

**Journal:** Journal of Urology 167(5), pp. 1965–1967

**Abstract: PURPOSE**: We determined which, if any, urinary stone risk factors accurately discriminate stone forming and nonstone forming siblings of patients with calcium renal stones. **MATERIALS AND METHODS**: A total of 252 siblings of stone formers provided 2, 24-hour urine samples, which were sent overnight and analyzed at a central laboratory. Standard stone risk factors were measured and the supersaturation of calcium oxalate, calcium phosphate and uric acid was calculated. **RESULTS**: Discriminant functions were derived for each gender by multivariate analysis. In stone forming sisters higher urinary calcium and pH discriminated with a success rate of 70%. In stone forming brothers higher urinary calcium, lower urinary potassium and older age discriminated with a success rate of 79%. **CONCLUSIONS**: Select urinary measurements as well as age classify siblings into those with and without stones with fair accuracy. Calcium excretion and urinary pH in females, and calcium excretion, urinary potassium and age in males are feasible identifiers of stone forming siblings. To determine whether these measurements can be used to predict new stone onset may require years of observation of our current cohort.

**URL:** <https://ovidsp.ovid.com/ovidweb.cgi?T=JS&CSC=Y&NEWS=N&PAGE=fulltext&D=med4&AN=11956418> <https://libkey.io/libraries/2592/openurl?genre=article&sid=OVID:medline&id=pmid:11956418&id=doi:&issn=0022-5347&isbn=&volume=167&issue=5&spage=1965&pages=1965-7&date=2002&title=Journal+of+Urology&atitle=Urinary+stone+risk+factors+in+the+siblings+of+patients+with+calcium+renal+stones.&aulast=Kinder&pid=%3Cauthor%3EKinder+JM%3BClark+CD%3BCoe+BJ%3BAsplin+JR%3BParks+JH%3BCoe+FL%3C%2Fauthor%3E%3CAN%3E11956418%3C%2FAN%3E%3CDT%3EJournal+Article%3C%2FDT%3E>

213. **The diagnostic value of the phosphate levels in serum and 24-hour urine samples in patients with recurrent renal stone disease.**

**Item Type:** Journal Article

**Authors:** Muezzinoglu, Talha;Gumus, Bilal;Sener, Ercument;Ari, Zeki and Buyuksu, Coskun

**Publication Date:** 2002

**Journal:** Scandinavian Journal of Urology & Nephrology 36(5), pp. 368–372

**Abstract: OBJECTIVE**: The aim of this study is to investigate the value of phosphate levels in serum and urine in patients with recurrent renal stone disease. **MATERIALS AND METHODS**: The patients (n:60) were divided into two groups as first-time stone disease (group 1) and recurrent renal stone disease (group 2). The demographical datas and their historical information were recorded and physical examination was done. The sera and urine for 24 hr were obtained from patients to measure electrolyte levels especially phosphorus. In addition, based on presenting serum phosphate levels, patients were divided into the hypophosphatemia group, less than 2.5 mg/dl; normophosphatemia group, between 2.5-5.2 mg/dl; and hyperphosphatemia group greater than 5.2 mg/dl. **RESULTS**: The mean age of study group was 45 (21-70) years. Thirty-six patients (60%) were in group 1 and 24 patients (40%) were in group 2. No statistically correlation was found between stone recurrence and phosphate levels both in serum and urine. There was only a statistical association of K levels in 24-hour urine samples between group 1 and 2. **CONCLUSION**: There was no significant association between stone recurrence and initial phosphate levels in the serum or in urine. We do not propose to determine phosphate levels routinely in management of patients with stone disease.

**URL:** <https://ovidsp.ovid.com/ovidweb.cgi?T=JS&CSC=Y&NEWS=N&PAGE=fulltext&D=med4&DO=10.1080%2f003655902320783881> <https://libkey.io/libraries/2592/openurl?genre=article&sid=OVID:medline&id=pmid:12487742&id=doi:10.1080%2F003655902320783881&issn=0036-5599&isbn=&volume=36&issue=5&spage=368&pages=368-72&date=2002&title=Scandinavian+Journal+of+Urology+%26+Nephrology&atitle=The+diagnostic+value+of+the+phosphate+levels+in+serum+and+24-hour+urine+samples+in+patients+with+recurrent+renal+stone+disease.&aulast=Muezzinoglu&pid=%3Cauthor%3EMuezzinoglu+T%3BGumus+B%3BSener+E%3BAri+Z%3BBuyuksu+C%3C%2Fauthor%3E%3CAN%3E12487742%3C%2FAN%3E%3CDT%3EComparative+Study%3C%2FDT%3E>

214. **Impact of renal cryoablation on urine composition.**

**Item Type:** Journal Article

**Authors:** Ng, Christopher S. and Gill, Inderbir S.

**Publication Date:** Ju ,2002

**Journal:** Urology 59(6), pp. 831–834

**Abstract: OBJECTIVES**: To examine prospectively serial urine biochemical parameters in 14 patients (9 men, 5 women) undergoing laparoscopic cryoablation of a small, exophytic solid renal mass. Prior studies have shown that various types of renal injury may predispose to the formation of urinary calculi. The metabolic effects of cryoenergy on the surrounding normal renal parenchyma are unknown. **METHODS**: Timed 24-hour urine collections were obtained preoperatively and postoperatively on days 1, 30, and 60 to evaluate the following parameters: light microscopic findings, volume, pH, creatinine, protein, beta(2)-microglobulin, calcium, citrate, oxalate, phosphate, uric acid, sodium, and potassium. **RESULTS**: Urinary beta(2)-microglobulin excretion increased from a preoperative baseline value of 114.8 to 1931.2 microg/L on postoperative day 1, an increase of more than 15-fold (P = 0.05), thus confirming major renal injury. These values sharply decreased at 30 days and returned to near-baseline levels at 60 days postoperatively (P = 0.76). Nevertheless, all lithogenic parameters remained within the normal range throughout the follow-up period, with no significant change in any value. **CONCLUSIONS**: Our findings suggest that renal cryoablation does not adversely alter urine composition with respect to lithogenic parameters for up to 2 months after surgery. Elevated beta(2)-microglobulin levels indicating significant renal injury immediately postoperatively spontaneously revert to baseline levels within 2 months.

**URL:** <https://ovidsp.ovid.com/ovidweb.cgi?T=JS&CSC=Y&NEWS=N&PAGE=fulltext&D=med4&DO=10.1016%2fs0090-4295%2802%2901541-8> <https://libkey.io/libraries/2592/openurl?genre=article&sid=OVID:medline&id=pmid:12031362&id=doi:10.1016%2Fs0090-4295%2802%2901541-8&issn=0090-4295&isbn=&volume=59&issue=6&spage=831&pages=831-4&date=2002&title=Urology&atitle=Impact+of+renal+cryoablation+on+urine+composition.&aulast=Ng&pid=%3Cauthor%3ENg+CS%3BGill+IS%3C%2Fauthor%3E%3CAN%3E12031362%3C%2FAN%3E%3CDT%3EJournal+Article%3C%2FDT%3E>

215. **A single 24-hour urine collection is inadequate for the medical evaluation of nephrolithiasis.**

**Item Type:** Journal Article

**Authors:** Parks, Joan H.;Goldfisher, Evan;Asplin, John R. and Coe, Fredric L.

**Publication Date:** Ap ,2002

**Journal:** Journal of Urology 167(4), pp. 1607–1612

**Abstract: PURPOSE**: We determined the adequacy of a single 24-hour urine sample for evaluating patients for medical renal stone prevention. **MATERIALS AND METHODS**: A total of 459 patients from a private urology practice specializing in the treatment of urolithiasis and 683 from a university stone research clinic provided 2 and 3, 24-hour urine samples, respectively. We used samples 1 and 2 from private practice patients, and 1 and 3 from university clinic patients for analysis, and compared each to the others by correlation coefficients and calculation of the mean difference plus or minus standard deviation (SD) of the difference. Urine risk factors were measured by standard methods. **RESULTS**: Although the correlation of urine values 1 and 2 was excellent for all stone risk factors, SD values for the differences were large enough that within 1 SD on either side of 0, which included 68.8% of cases, by chance urine 1 would depart from urine 2 by clinically important amounts. These departures would be more than sufficient to misdiagnose common metabolic disorders. **CONCLUSIONS**: A single 24-hour sample is not sufficient for evaluating patients before metabolic treatment for stone prevention because misdiagnosis is common, leading to inappropriate treatment.

**URL:** <https://ovidsp.ovid.com/ovidweb.cgi?T=JS&CSC=Y&NEWS=N&PAGE=fulltext&D=med4&AN=11912373> <https://libkey.io/libraries/2592/openurl?genre=article&sid=OVID:medline&id=pmid:11912373&id=doi:&issn=0022-5347&isbn=&volume=167&issue=4&spage=1607&pages=1607-12&date=2002&title=Journal+of+Urology&atitle=A+single+24-hour+urine+collection+is+inadequate+for+the+medical+evaluation+of+nephrolithiasis.&aulast=Parks&pid=%3Cauthor%3EParks+JH%3BGoldfisher+E%3BAsplin+JR%3BCoe+FL%3C%2Fauthor%3E%3CAN%3E11912373%3C%2FAN%3E%3CDT%3EJournal+Article%3C%2FDT%3E>

216. **Metabolic risk factors for stone formation in patients with cystic fibrosis.**

**Item Type:** Journal Article

**Authors:** Perez-Brayfield, Marcos R.;Caplan, Daniel;Gatti, John M.;Smith, Edwin A. and Kirsch, Andrew J.

**Publication Date:** Fe ,2002

**Journal:** Journal of Urology 167(2 Pt 1), pp. 480–484

**Abstract: PURPOSE**: Cystic fibrosis is characterized by chronic pulmonary disease, insufficient pancreatic and digestive function, and abnormal sweat concentrations. Patients with cystic fibrosis also have an increased incidence of nephrolithiasis. We compared the results of metabolic evaluation in patients with cystic fibrosis with and without nephrolithiasis. **MATERIALS AND METHODS**: A total of 496 patients were evaluated at our center, including 98 with a mean age of 25 years who had cystic fibrosis and complete metabolic evaluation available between 1996 and 2000. Of these patients 13 (13%) had a history of nephrolithiasis. The records were reviewed for clinical characteristics and all patients underwent metabolic evaluation, including serum electrolyte measurement and 24-hour urine collection. Statistical analysis was done to compare the stone versus nonstone groups. **RESULTS**: The incidence of nephrolithiasis in our study was 3%. We identified 13 patients 16 to 41 years old (mean age 27) with nephrolithiasis, of whom 62% had had multiple episodes. Flank pain was the presenting symptom in 9 of the 13 cases (69%). Renal ultrasound and computerized tomography were the most common imaging modalities. In 9 cases stones were passed without intervention, extracorporeal shock wave lithotripsy was required in 2 and ureteroscopy with stone extraction was done in 2. Calcium oxalate was the dominant stone composition in the 9 patients in whom stone analysis was performed. Metabolic evaluation of the stone versus no stone groups showed elevated urinary oxalate (45.5 versus 42.5 mg./24 hours), relative calcium oxalate supersaturation (5.3 versus 7.2) and decreased urinary citrate in the 2 groups. There was a statistical difference in citrate excretion with lower levels in stone formers (102 versus 218 mg./24 hours, p = 0.0007). **CONCLUSIONS**: Patients with cystic fibrosis have an increased incidence of nephrolithiasis and are at high risk for recurrence. Metabolic evaluation is indicated in all patients with cystic fibrosis and urolithiasis since most have hyperoxaluria, urinary calcium oxalate supersaturation and decreased urinary citrate. Correcting detected stone risk factors in these cases may decrease stone recurrence.

**URL:** <https://ovidsp.ovid.com/ovidweb.cgi?T=JS&CSC=Y&NEWS=N&PAGE=fulltext&D=med4&DO=10.1016%2fS0022-5347%2801%2969068-2> <https://libkey.io/libraries/2592/openurl?genre=article&sid=OVID:medline&id=pmid:11792901&id=doi:10.1016%2FS0022-5347%2801%2969068-2&issn=0022-5347&isbn=&volume=167&issue=2&spage=480&pages=480-4&date=2002&title=Journal+of+Urology&atitle=Metabolic+risk+factors+for+stone+formation+in+patients+with+cystic+fibrosis.&aulast=Perez-Brayfield&pid=%3Cauthor%3EPerez-Brayfield+MR%3BCaplan+D%3BGatti+JM%3BSmith+EA%3BKirsch+AJ%3C%2Fauthor%3E%3CAN%3E11792901%3C%2FAN%3E%3CDT%3EJournal+Article%3C%2FDT%3E>

217. **Urinary citrate excretion in idiopathic nephrolithiasis.**

**Item Type:** Journal Article

**Authors:** Ratan, Simmi K.;Bhatnagar, V.;Mitra, D. K.;Basu, N. and Malhotra, L. K.

**Publication Date:** Se ,2002

**Journal:** Indian Pediatrics 39(9), pp. 819–825

**Abstract: OBJECTIVE**: To determine urinary citrate excretion in children with nephrolithiasis and normal controls. **DESIGN**: Prospective. **SETTING**: Tertiary care center in New Delhi. **METHODS**: This study was done on 50 children, below the age of 12 years, with idiopathic urinary calculi and 150 age and weight matched controls. The children were divided into 3 groups: Group 1 (1-4 years), Group 2 (5-8 years) and Group 3 (9-12 years). Urinary citrate was estimated in a 24-hour urine sample using colorimetric method. The stones removed from these children were also analysed. **RESULTS**: There was a preponderance of urinary stones in males; the highest incidence being in Group 1. Excretion of citrate in 24-hour urine sample was significantly lower in patients compared to controls, for males in all age groups and for females in Group 3. However, there was no statistically significant difference in the urinary citrate value between males and females in a given age group for either controls or patients. The urinary citrate excretion increased with age in patients and controls, but the levels in patients were lower. Depending upon the constituents, four types of stones were identified, calcium phosphate, calcium oxalate, uric acid and magnesium ammonium phosphate. Nine stones had at least more than one major constituent. Hypocitraturia was detected in 43 percent cases. The incidence was 76 percent for calcium phosphate, 87 percent for calcium oxalate, 40 percent for uric acid stones and 50 percent for magnesium ammonium phosphate. **CONCLUSION**: This study shows that low urinary citrate is associated with urinary stones in children, especially in endemic areas, in the absence of obvious etiological factors. Urinary citrate excretion should be determined in all children with nephrolithiasis.

**URL:** <https://ovidsp.ovid.com/ovidweb.cgi?T=JS&CSC=Y&NEWS=N&PAGE=fulltext&D=med4&AN=12368525> <https://libkey.io/libraries/2592/openurl?genre=article&sid=OVID:medline&id=pmid:12368525&id=doi:&issn=0019-6061&isbn=&volume=39&issue=9&spage=819&pages=819-25&date=2002&title=Indian+Pediatrics&atitle=Urinary+citrate+excretion+in+idiopathic+nephrolithiasis.&aulast=Ratan&pid=%3Cauthor%3ERatan+SK%3BBhatnagar+V%3BMitra+DK%3BBasu+N%3BMalhotra+LK%3C%2Fauthor%3E%3CAN%3E12368525%3C%2FAN%3E%3CDT%3EJournal+Article%3C%2FDT%3E>

218. **Effects of 5 different diets on urinary risk factors for calcium oxalate kidney stone formation: evidence of different renal handling mechanisms in different race groups.**

**Item Type:** Journal Article

**Authors:** Rodgers, Allen L. and Lewandowski, Sonja

**Publication Date:** Se ,2002

**Journal:** Journal of Urology 168(3), pp. 931–936

**Abstract: PURPOSE**: Since the incidence of renal calculi in the South African black population is extremely rare while in white subjects it occurs at the same rate as elsewhere in the western world, we investigated the possibility that different renal handling mechanisms in response to different dietary challenges might occur in the 2 race groups. **MATERIALS AND METHODS**: We administered 5 different dietary protocols, including low calcium, high oxalate, vitamin C, high salt and lacto-vegetarian, to 10 healthy male subjects from each race group. We collected 24-hour urine at baseline and after 4 days on the prescribed diet which were analyzed for biochemical and physicochemical risk factors. Dietary intake was controlled throughout the experimental period. A 24-hour dietary recall questionnaire was recorded at baseline and analyzed using food composition tables. Statistical analysis of variance was performed on all the data. **RESULTS**: The low calcium diet caused statistically significant changes only in black subjects, which consisted of urinary oxalate increase (0.17 to 0.23 mmol./24 hours, p = 0.01), relative supersaturation of calcium oxalate decrease (1.88 to 0.97, p = 0.03) and relative supersaturation of brushite increase (0.85 to 1.69, p = 0.03). The high oxalate diet caused statistically significant changes in both race groups but these changes were different in the 2 groups. In white subjects urinary pH increased (6.24 to 6.62, p = 0.01), potassium excretion increased (40.01 to 73.49, p = 0.01) and relative supersaturation of brushite increased (1.34 to 2.12, p = 0.05). In black subjects urinary citrate increased (1.94 to 2.99 mmol./24 hours, p = 0.01). Clinically unimportant changes occurred in both race groups after the other 3 diets. **CONCLUSIONS**: Renal handling of dietary calcium and oxalate in South African black and white subjects is different and may explain the different stone incidence in the 2 race groups.

**URL:** <https://ovidsp.ovid.com/ovidweb.cgi?T=JS&CSC=Y&NEWS=N&PAGE=fulltext&D=med4&DO=10.1016%2fS0022-5347%2805%2964545-4> <https://libkey.io/libraries/2592/openurl?genre=article&sid=OVID:medline&id=pmid:12187193&id=doi:10.1016%2FS0022-5347%2805%2964545-4&issn=0022-5347&isbn=&volume=168&issue=3&spage=931&pages=931-6&date=2002&title=Journal+of+Urology&atitle=Effects+of+5+different+diets+on+urinary+risk+factors+for+calcium+oxalate+kidney+stone+formation%3A+evidence+of+different+renal+handling+mechanisms+in+different+race+groups.&aulast=Rodgers&pid=%3Cauthor%3ERodgers+AL%3BLewandowski+S%3C%2Fauthor%3E%3CAN%3E12187193%3C%2FAN%3E%3CDT%3EComparative+Study%3C%2FDT%3E>

219. **Urinary patterns of patients with renal stones associated with chronic inflammatory bowel disease.**

**Item Type:** Journal Article

**Authors:** Trinchieri, Alberto;Lizzano, Renata;Castelnuovo, Chiara;Zanetti, Giampaolo and Pisani, Enrico

**Publication Date:** Ju ,2002

**Journal:** Archivio Italiano Di Urologia, Andrologia 74(2), pp. 61–64

**Abstract: OBJECTIVE**: The aim of this study was to analyze the frequency of renal stone patients with chronic inflammatory bowel disease and their urinary patterns. **METHODS**: During a 20-year period, 1941 consecutive patients with renal stone disease underwent routine laboratory procedures including a fasting blood sample for chemistry profile and a 24-hour urine collection for analyses of electrolytes. Thorough histories including chronic inflammatory disease or ileal resection were obtained. Patients with inflammatory bowel disease together with a control group comprising 47 idiopathic renal calcium stone formers were submitted to a xylose absorption test for evaluation of intestinal absorption. **RESULTS**: We observed 10 patients with Crohn's disease, 12 with ulcerative colitis and one patient with ileal bypass for obesity. Six patients underwent ileal resection and 10 patients total colectomy. Urinary oxalate excretion was significantly higher and urinary citrate lower in stone patients with ileal disease (Ox 60 +/- 23, Cit 113 + 7-118 mg/day) than in idiopathic stone formers (Ox 28.2 +/- 11.5, Cit 381 +/- 205) and stone patients with ulcerative colitis (Ox 20.3 +/- 14.8, Cit 369 +/- 247). Urinary volume was significantly lower in patients with ulcerative colitis. A significant inverse correlation (-0.38, p : We observed 10 patients with Crohn's disease, 12 with ulcerative colitis and one patient with ileal bypass for obesity. Six patients underwent ileal resection and 10 patients total colectomy. Urinary oxalate excretion was significantly higher and urinary citrate lower in stone patients with ileal disease (Ox 60 +/- 23, Cit 113 + 7-118 mg/day) than in idiopathic stone formers (Ox 28.2 +/- 11.5, Cit 381 +/- 205) and stone patients with ulcerative colitis (Ox 20.3 +/- 14.8, Cit 369 +/- 247). Urinary volume was significantly lower in patients with ulcerative colitis. A significant inverse correlation (-0.38, p **CONCLUSIONS**: Crohn's disease and ulcerative colitis are characterized by recurrent inflammatory involvement of different intestinal segments involving distinctive urinary patterns. Malabosorption associated with ileal disease causes increased oxalate absorption by increasing oxalate solubility in the intestinal lumen and permeability of the colonic mucosa; a reduced citrate excretion is associated in relation to mild acidosis due to the loss of bicarbonate in the liquid stool. In ulcerative colitis, especially if an ileostomy is present, urine are scanty and concentrated, and urine pH falls, leading to uric acid or mixed stones. Mild hyperoxaluria of idiopathic renal stone formers is not related to subtle intestinal malabsorption.

**URL:** <https://ovidsp.ovid.com/ovidweb.cgi?T=JS&CSC=Y&NEWS=N&PAGE=fulltext&D=med4&AN=12161938> <https://libkey.io/libraries/2592/openurl?genre=article&sid=OVID:medline&id=pmid:12161938&id=doi:&issn=1124-3562&isbn=&volume=74&issue=2&spage=61&pages=61-4&date=2002&title=Archivio+Italiano+di+Urologia%2C+Andrologia&atitle=Urinary+patterns+of+patients+with+renal+stones+associated+with+chronic+inflammatory+bowel+disease.&aulast=Trinchieri&pid=%3Cauthor%3ETrinchieri+A%3BLizzano+R%3BCastelnuovo+C%3BZanetti+G%3BPisani+E%3C%2Fauthor%3E%3CAN%3E12161938%3C%2FAN%3E%3CDT%3EComparative+Study%3C%2FDT%3E>

220. **Reduction of oxaluria after an oral course of lactic acid bacteria at high concentration.**

**Item Type:** Journal Article

**Authors:** Campieri, C.;Campieri, M.;Bertuzzi, V.;Swennen, E.;Matteuzzi, D.;Stefoni, S.;Pirovano, F.;Centi, C.;Ulisse, S.;Famularo, G. and De Simone, C.

**Publication Date:** Se ,2001

**Journal:** Kidney International 60(3), pp. 1097–1105

**Abstract: BACKGROUND**: Hyperoxaluria is a major risk factor for renal stones, and in most cases, it appears to be sustained by increased dietary load or increased intestinal absorption. Previous studies have shown that components of the endogenous digestive microflora, in particular Oxalobacter formigenes, utilize oxalate in the gut, thus limiting its absorption. We tested the hypothesis of whether oxaluria can be reduced by means of reducing intestinal absorption through feeding a mixture of freeze-dried lactic acid bacteria. **METHODS**: Six patients with idiopathic calcium-oxalate urolithiasis and mild hyperoxaluria (>40 mg/24 h) received daily a mixture containing 8 x 10(11) freeze-dried lactic acid bacteria (L. acidophilus, L. plantarum, L. brevis, S. thermophilus, B. infantis) for four weeks. The 24-hour urinary excretion of oxalate was determined at the end of the study period and then one month after ending the treatment. The ability of bacteria to degrade oxalate and grow in oxalate-containing media, and the gene expression of Ox1T, an enzyme that catalyzes the transmembrane exchange of oxalate, also were investigated. **RESULTS**: The treatment resulted in a great reduction of the 24-hour excretion of oxalate in all six patients enrolled. Mean levels +/- SD were 33.5 +/- 15.9 mg/24 h at the end of the study period and 28.3 +/- 14.6 mg/24 h one month after treatment was interrupted compared with baseline values of 55.5 +/- 19.6 mg/24 h (P : The treatment resulted in a great reduction of the 24-hour excretion of oxalate in all six patients enrolled. Mean levels +/- SD were 33.5 +/- 15.9 mg/24 h at the end of the study period and 28.3 +/- 14.6 mg/24 h one month after treatment was interrupted compared with baseline values of 55.5 +/- 19.6 mg/24 h (P **CONCLUSIONS**: The urinary excretion of oxalate, a major risk factor for renal stone formation and growth in patients with idiopathic calcium-oxalate urolithiasis, can be greatly reduced with treatment using a high concentration of freeze-dried lactic acid bacteria. We postulate that the biological manipulation of the endogenous digestive microflora can be a novel approach for the prevention of urinary stone formation.

**URL:** <https://ovidsp.ovid.com/ovidweb.cgi?T=JS&CSC=Y&NEWS=N&PAGE=fulltext&D=med4&DO=10.1046%2fj.1523-1755.2001.0600031097.x> <https://libkey.io/libraries/2592/openurl?genre=article&sid=OVID:medline&id=pmid:11532105&id=doi:10.1046%2Fj.1523-1755.2001.0600031097.x&issn=0085-2538&isbn=&volume=60&issue=3&spage=1097&pages=1097-105&date=2001&title=Kidney+International&atitle=Reduction+of+oxaluria+after+an+oral+course+of+lactic+acid+bacteria+at+high+concentration.&aulast=Campieri&pid=%3Cauthor%3ECampieri+C%3BCampieri+M%3BBertuzzi+V%3BSwennen+E%3BMatteuzzi+D%3BStefoni+S%3BPirovano+F%3BCenti+C%3BUlisse+S%3BFamularo+G%3BDe+Simone+C%3C%2Fauthor%3E%3CAN%3E11532105%3C%2FAN%3E%3CDT%3EClinical+Trial%3C%2FDT%3E>

221. **Differences in urine volume and supersaturation in 2 physician networks.**

**Item Type:** Journal Article

**Authors:** Coe, F. L.;Wise, H.;Parks, J. H.;Fleury, P. J. and Asplin, J. R.

**Publication Date:** Au ,2001

**Journal:** Journal of Urology 166(2), pp. 423–428

**Abstract: PURPOSE**: We determined whether a network of practices devoted to a broad range of urological care would achieve a decrease in metabolic stone risk comparable to that achieved by a network of similar practices that emphasized kidney stone management as a distinct specialized interest, provided that each was given equivalent access to high level urine testing and software support. **MATERIALS AND METHODS**: Pretreatment and treatment 24-hour urine samples were obtained from patients in a large network of practices related by the shared use of lithotripsy facilities and instruments (group 2) and a contrasting network of practices that emphasize stone treatment over other concerns (group 1). All known urine risk factors, including supersaturation, were measured and calculated. **RESULTS**: Treatment supersaturation values in group 2 exceeded those in group 1. The reason was unpredicted and unexplained but highly consistent lower urine volume in group 2 patients that was present before and persisted during treatment. Group 2 physicians mostly achieved changes in urine volume and stone risk factors equivalent to those of group 1 physicians but began with higher supersaturation due to lower urine volume. **CONCLUSIONS**: A network of physicians not specialized for stone care may achieve a decreased risk equivalent to that of more specialized physicians. Initial patient characteristics may vary significantly in the groups for reasons that are unknown to date, greatly affecting treatment outcome.

**URL:** <https://ovidsp.ovid.com/ovidweb.cgi?T=JS&CSC=Y&NEWS=N&PAGE=fulltext&D=med4&AN=11458041> <https://libkey.io/libraries/2592/openurl?genre=article&sid=OVID:medline&id=pmid:11458041&id=doi:&issn=0022-5347&isbn=&volume=166&issue=2&spage=423&pages=423-8&date=2001&title=Journal+of+Urology&atitle=Differences+in+urine+volume+and+supersaturation+in+2+physician+networks.&aulast=Coe&pid=%3Cauthor%3ECoe+FL%3BWise+H%3BParks+JH%3BFleury+PJ%3BAsplin+JR%3C%2Fauthor%3E%3CAN%3E11458041%3C%2FAN%3E%3CDT%3EComparative+Study%3C%2FDT%3E>

222. **Twenty-four-hour urine chemistries and the risk of kidney stones among women and men.**

**Item Type:** Journal Article

**Authors:** Curhan, G. C.;Willett, W. C.;Speizer, F. E. and Stampfer, M. J.

**Publication Date:** Ju ,2001

**Journal:** Kidney International 59(6), pp. 2290–2298

**Abstract: BACKGROUND**: Results of a 24-hour urine collection are integral to the selection of the most appropriate intervention to prevent kidney stone recurrence. However, the currently accepted definitions of normal urine values are not firmly supported by the literature. In addition, little information is available about the relationship between risk of stone formation and the levels of urinary factors. Unfortunately, the majority of previous studies of 24-hour urine chemistries were limited by the inclusion of recurrent stone formers and poorly defined controls. **METHODS**: We obtained 24-hour urine collections from 807 men and women with a history of kidney stone disease and 239 without a history who were participants in three large ongoing cohort studies: the Nurses' Health Study I (NHS I; mean age of 61 years), the Nurses' Health Study II (NHS II; mean age of 42 years), and the Health Professionals Follow-up Study (HPFS; mean age of 59 years). **RESULTS**: Mean 24-hour urine calcium excretion was higher and urine volume was lower in cases than controls in NHS I (P : Mean 24-hour urine calcium excretion was higher and urine volume was lower in cases than controls in NHS I (P : Mean 24-hour urine calcium excretion was higher and urine volume was lower in cases than controls in NHS I (P : Mean 24-hour urine calcium excretion was higher and urine volume was lower in cases than controls in NHS I (P or =200 mg/L for NHS I was 4.34 (95% CI, 1.59 to 11.88), for NHS II was 51.09 (4.27 to 611.1), and for HPFS was 4.30 (1.71 to 10.84). There was substantial variation in the relative risks for stone formation for the concentration of other urine factors within the different cohorts. **CONCLUSIONS**: The traditional definitions of normal 24-hour urine values need to be reassessed, as a substantial proportion of controls would be defined as abnormal, and the association with risk of stone formation may be continuous rather than dichotomous. The 24-hour urine chemistries are important for predicting risk of stone formation, but the significance and the magnitudes of the associations appear to differ by age and gender.

**URL:** <https://ovidsp.ovid.com/ovidweb.cgi?T=JS&CSC=Y&NEWS=N&PAGE=fulltext&D=med4&DO=10.1046%2fj.1523-1755.2001.00746.x> <https://libkey.io/libraries/2592/openurl?genre=article&sid=OVID:medline&id=pmid:11380833&id=doi:10.1046%2Fj.1523-1755.2001.00746.x&issn=0085-2538&isbn=&volume=59&issue=6&spage=2290&pages=2290-8&date=2001&title=Kidney+International&atitle=Twenty-four-hour+urine+chemistries+and+the+risk+of+kidney+stones+among+women+and+men.&aulast=Curhan&pid=%3Cauthor%3ECurhan+GC%3BWillett+WC%3BSpeizer+FE%3BStampfer+MJ%3C%2Fauthor%3E%3CAN%3E11380833%3C%2FAN%3E%3CDT%3EJournal+Article%3C%2FDT%3E>

223. **Diets with either beef or plant proteins reduce risk of calcium oxalate precipitation in patients with a history of calcium kidney stones.**

**Item Type:** Journal Article

**Authors:** Massey, L. K. and Kynast-Gales, S. A.

**Publication Date:** Ma ,2001

**Journal:** Journal of the American Dietetic Association 101(3), pp. 326–331

**Abstract: OBJECTIVE**: To determine the effect of substituting equal amounts of dietary protein as animal protein (beef) for plant protein (legumes, seeds, nuts, and grains) on urinary components associated with calcium oxalate precipitability risk. **DESIGN**: Randomized crossover trial. **SUBJECTS**: Twenty-three normocalciuric patients with a history of calcium kidney stones (8 women and 15 men, mean age 50.7+/-14.6 years) with 24-hour urinary calcium : Twenty-three normocalciuric patients with a history of calcium kidney stones (8 women and 15 men, mean age 50.7+/-14.6 years) with 24-hour urinary calcium : Twenty-three normocalciuric patients with a history of calcium kidney stones (8 women and 15 men, mean age 50.7+/-14.6 years) with 24-hour urinary calcium **SETTING**: Four-day, free-living adaptation period, followed by 2-day metabolic unit study. **INTERVENTION**: The study compared consumption of 2 servings of beef (43 g protein for women and 50 g for men) daily with an equal amount of protein from plant foods including legumes, nuts, and grains. **MAIN OUTCOME MEASURES**: Tiselius risk index (TRI) for calcium oxalate precipitability calculated from urinary calcium, oxalate, magnesium, citrate, and volume. **STATISTICAL ANALYSES**: Paired t tests. **RESULTS**: Urinary calcium, oxalate, magnesium, citrate, phosphorus, volume, and TRI did not differ between diets. Urinary sodium and potassium were higher for patients on the plant protein diet. After correcting for variations in urinary sodium and potassium between diets, the difference in urinary calcium remained insignificant. TRI was lower on both beef- and plant-protein diets compared with self-selected prestudy diets for all participants. CONCLUSION /APPLICATIONS: Balanced diets containing moderate amounts of either beef or plant protein are equally effective in reducing calcium oxalate kidney stone risk based on changes in urinary composition.

**URL:** <https://ovidsp.ovid.com/ovidweb.cgi?T=JS&CSC=Y&NEWS=N&PAGE=fulltext&D=med4&DO=10.1016%2fS0002-8223%2801%2900085-2> <https://libkey.io/libraries/2592/openurl?genre=article&sid=OVID:medline&id=pmid:11269613&id=doi:10.1016%2FS0002-8223%2801%2900085-2&issn=0002-8223&isbn=&volume=101&issue=3&spage=326&pages=326-31&date=2001&title=Journal+of+the+American+Dietetic+Association&atitle=Diets+with+either+beef+or+plant+proteins+reduce+risk+of+calcium+oxalate+precipitation+in+patients+with+a+history+of+calcium+kidney+stones.&aulast=Massey&pid=%3Cauthor%3EMassey+LK%3BKynast-Gales+SA%3C%2Fauthor%3E%3CAN%3E11269613%3C%2FAN%3E%3CDT%3EClinical+Trial%3C%2FDT%3E>

224. **Strong inhibition of crystal-cell attachment by pediatric urinary macromolecules: a close relationship with high urinary citrate secretion.**

**Item Type:** Journal Article

**Authors:** Miyake, O.;Kakimoto, K.;Tsujihata, M.;Yoshimura, K.;Takahara, S. and Okuyama, A.

**Publication Date:** Se ,2001

**Journal:** Urology 58(3), pp. 493–497

**Abstract: OBJECTIVES**: To investigate other reasons for the low incidence of pediatric urolithiasis, we evaluated the difference in the crystal-cell adhesion inhibitory activity of urinary macromolecules (UMMs) between children and adults. We also evaluated whether citrates influence the above inhibitory activity, because citrates are important in pediatric urine. **METHODS**: Urine samples were collected from children and healthy male adults during a 24-hour period, and urinary components with a molecular weight of 3 kDa or greater were extracted as UMMs to compare their inhibitory activity during the adhesion of calcium oxalate monohydrate crystals to cultured Madin-Darby canine kidney cells between children and adults. Subsequently, various concentrations of citrates were added to adult UMMs to evaluate the changes in the crystal-cell adhesion inhibitory activity of UMMs. **RESULTS**: Pediatric UMMs more strongly inhibited the adhesion of calcium oxalate monohydrate crystals to cultured Madin-Darby canine kidney cells at a concentration of 0.1 mg/mL compared with adult UMMs. In addition, pediatric UMMs contained higher proportions of fibronectin and glycosaminoglycans, both of which exhibit crystal-cell adhesion inhibitory activity. When citrates were added to adult UMMs, the crystal-cell adhesion inhibitory activity of UMMs was increased in a dose-dependent manner. However, citrates alone did not result in any differences in the inhibitory activity at any of the three different concentrations. **CONCLUSIONS**: We speculate that the incidence of pediatric urolithiasis is low because pediatric UMMs more potently inhibit the adhesion of calcium oxalate crystals to renal tubular cells or because the higher proportion of citrates in pediatric urine enhances the crystal-cell adhesion inhibitory activity of UMMs in a dose-dependent manner.

**URL:** <https://ovidsp.ovid.com/ovidweb.cgi?T=JS&CSC=Y&NEWS=N&PAGE=fulltext&D=med4&DO=10.1016%2fs0090-4295%2801%2901257-2> <https://libkey.io/libraries/2592/openurl?genre=article&sid=OVID:medline&id=pmid:11549512&id=doi:10.1016%2Fs0090-4295%2801%2901257-2&issn=0090-4295&isbn=&volume=58&issue=3&spage=493&pages=493-7&date=2001&title=Urology&atitle=Strong+inhibition+of+crystal-cell+attachment+by+pediatric+urinary+macromolecules%3A+a+close+relationship+with+high+urinary+citrate+secretion.&aulast=Miyake&pid=%3Cauthor%3EMiyake+O%3BKakimoto+K%3BTsujihata+M%3BYoshimura+K%3BTakahara+S%3BOkuyama+A%3C%2Fauthor%3E%3CAN%3E11549512%3C%2FAN%3E%3CDT%3EComparative+Study%3C%2FDT%3E>

225. **Sensitivity to meat protein intake and hyperoxaluria in idiopathic calcium stone formers.**

**Item Type:** Journal Article

**Authors:** Nguyen, Q. V.;Kalin, A.;Drouve, U.;Casez, J. P. and Jaeger, P.

**Publication Date:** Ju ,2001

**Journal:** Kidney International 59(6), pp. 2273–2281

**Abstract: BACKGROUND**: High protein intake is an accepted risk factor for renal stone disease. Whether meat protein intake affects oxaluria, however, remains controversial in healthy subjects and in stone formers. This study was designed (1) to test the oxaluric response to a meat protein load in male recurrent idiopathic calcium stone formers (ICSFs) with and without mild metabolic hyperoxaluria (MMH and non-MMH, respectively), as well as in healthy controls, and (2) to seek for possible disturbed vitamin B(6) metabolism in MMH, in analogy with primary hyperoxaluria. **METHODS**: Twelve MMH, 8 non-MMH, and 13 healthy males were studied after five days on a high meat protein diet (HPD; 700 g meat/fish daily) following a run-in phase of five days on a moderate protein diet (MPD; 160 g meat/fish daily). In both diets, oxalate-rich nutrients were avoided, as well as sweeteners and vitamin C-containing medicines. Twenty-four-hour urinary excretion of oxalate was measured on the last day of each period, along with 4-pyridoxic acid (U(4PA)) and markers of protein intake, that is, urea, phosphate, uric acid, and sulphate. Serum pyridoxal 5' phosphate (S(P5P)) was measured after protein loading. **RESULTS**: Switching from MPD (0.97 +/- 0.18 g protein/kg/day) to HPD (2.26 +/- 0.38 g protein/kg/day) led to the expected rise in the urinary excretion rates of all markers of protein intake in all subjects. Concurrently, the mean urinary excretion of oxalate increased in ICSFs taken as a whole (+73 +/- 134 micromol/24 h, P = 0.024) as well as in the MMH subgroup (+100 +/- 144 micromol/24 h, P = 0.034) but not in controls (-17 +/- 63 micromol/24 h). In seven ICSFs (4 MMH and 3 non-MMH) but in none of the healthy controls (P = 0.016, chi square), an increment in oxaluria was observed and considered as significant based on the intra-assay coefficient of variation at our laboratory (8.5%). There was no difference in S(P5P)nd U(4PA)etween the groups after protein loading. **CONCLUSION**: Approximately one third of ICSFs with or without so-called MMH are sensitive to meat protein in terms of oxalate excretion, as opposed to healthy subjects. Mechanisms underlying this sensitivity to meat protein remain to be elucidated and do not seem to involve vitamin B(6) deficiency.

**URL:** <https://ovidsp.ovid.com/ovidweb.cgi?T=JS&CSC=Y&NEWS=N&PAGE=fulltext&D=med4&DO=10.1046%2fj.1523-1755.2001.00744.x> <https://libkey.io/libraries/2592/openurl?genre=article&sid=OVID:medline&id=pmid:11380831&id=doi:10.1046%2Fj.1523-1755.2001.00744.x&issn=0085-2538&isbn=&volume=59&issue=6&spage=2273&pages=2273-81&date=2001&title=Kidney+International&atitle=Sensitivity+to+meat+protein+intake+and+hyperoxaluria+in+idiopathic+calcium+stone+formers.&aulast=Nguyen&pid=%3Cauthor%3ENguyen+QV%3BKalin+A%3BDrouve+U%3BCasez+JP%3BJaeger+P%3C%2Fauthor%3E%3CAN%3E11380831%3C%2FAN%3E%3CDT%3EClinical+Trial%3C%2FDT%3E>

226. **Adequacy of a single stone risk analysis in the medical evaluation of urolithiasis.**

**Item Type:** Journal Article

**Authors:** Pak, C. Y.;Peterson, R. and Poindexter, J. R.

**Publication Date:** Fe ,2001

**Journal:** Journal of Urology 165(2), pp. 378–381

**Abstract: PURPOSE**: We tested the hypothesis that a single 24-hour urine sample for stone risk analysis would be sufficient for the simplified medical evaluation of urolithiasis. **MATERIALS AND METHODS**: We retrospectively analyzed stone risk profile data on 24-hour urine samples obtained during random and restricted diets in 225 patients with recurrent urolithiasis. **RESULTS**: In 2 random samples we noted no significant difference in urinary calcium, oxalate, uric acid, citrate, pH, total volume, sodium, potassium, sulphate or phosphorus. For these risk factors there was a highly significant positive correlation in the 2 random samples (r > or = 0.68, p : In 2 random samples we noted no significant difference in urinary calcium, oxalate, uric acid, citrate, pH, total volume, sodium, potassium, sulphate or phosphorus. For these risk factors there was a highly significant positive correlation in the 2 random samples (r > or = 0.68, p **CONCLUSIONS**: The reproducibility of urinary stone risk factors is satisfactory in repeat urine samples. A single stone risk analysis is sufficient for the simplified medical evaluation of urolithiasis.

**URL:** <https://ovidsp.ovid.com/ovidweb.cgi?T=JS&CSC=Y&NEWS=N&PAGE=fulltext&D=med4&DO=10.1097%2f00005392-200102000-00006> <https://libkey.io/libraries/2592/openurl?genre=article&sid=OVID:medline&id=pmid:11176377&id=doi:10.1097%2F00005392-200102000-00006&issn=0022-5347&isbn=&volume=165&issue=2&spage=378&pages=378-81&date=2001&title=Journal+of+Urology&atitle=Adequacy+of+a+single+stone+risk+analysis+in+the+medical+evaluation+of+urolithiasis.&aulast=Pak&pid=%3Cauthor%3EPak+CY%3BPeterson+R%3BPoindexter+JR%3C%2Fauthor%3E%3CAN%3E11176377%3C%2FAN%3E%3CDT%3EJournal+Article%3C%2FDT%3E>

227. **Patient adherence to long-term medical treatment of kidney stones.**

**Item Type:** Journal Article

**Authors:** Parks, J. H.;Asplin, J. R. and Coe, F. L.

**Publication Date:** De ,2001

**Journal:** Journal of Urology 166(6), pp. 2057–2060

**Abstract: PURPOSE**: We determine patient adherence to and quality of outcome of medical kidney stone treatment during a 30-year duration at a single university based referral clinic. We also analyze time trends in adherence and timing of followup measurements, and supersaturation reduction during treatment. **MATERIALS AND METHODS**: Data on all patients who entered the University of Chicago Kidney Stone Prevention Program from 1970 to 2000 were analyzed. Fractions of new patients who had any followup and those remaining in followup at increasing intervals were analyzed. Timing of followup was measured. Changes in adherence during the 3 decades were also analyzed, as was reduction in supersaturation in regard to calcium oxalate, calcium phosphate and uric acid. **RESULTS**: A total of 70% to 80% of patients were retained at each successive followup cycle with 2 physicians, and a clinical protocol that always required 6-week followup with 24-hour urine collection and a yearly one thereafter for stone risk factors. Retention decreased during the last 5 years of the 1990s. Supersaturation reduction was present by the first followup and remained constant or improved with time. Timing of followup measurements was in accord with our protocol. **CONCLUSIONS**: At best, one can retain only 70% to 80% of patients in a followup program at each interval, and achieve supersaturation reductions that are constant and significant during the long term. Timing of followup measurements can be close to that of the protocol in use.

**URL:** <https://ovidsp.ovid.com/ovidweb.cgi?T=JS&CSC=Y&NEWS=N&PAGE=fulltext&D=med4&AN=11696706> <https://libkey.io/libraries/2592/openurl?genre=article&sid=OVID:medline&id=pmid:11696706&id=doi:&issn=0022-5347&isbn=&volume=166&issue=6&spage=2057&pages=2057-60&date=2001&title=Journal+of+Urology&atitle=Patient+adherence+to+long-term+medical+treatment+of+kidney+stones.&aulast=Parks&pid=%3Cauthor%3EParks+JH%3BAsplin+JR%3BCoe+FL%3C%2Fauthor%3E%3CAN%3E11696706%3C%2FAN%3E%3CDT%3EJournal+Article%3C%2FDT%3E>

228. **Dietary supplementation with cranberry concentrate tablets may increase the risk of nephrolithiasis.**

**Item Type:** Journal Article

**Authors:** Terris, M. K.;Issa, M. M. and Tacker, J. R.

**Publication Date:** Ja ,2001

**Journal:** Urology 57(1), pp. 26–29

**Abstract: OBJECTIVES**: Cranberry juice has been recommended for patients with recurrent urinary tract infections. However, cranberry juice has a moderately high concentration of oxalate, a common component of kidney stones, and should be limited in patients with a history of nephrolithiasis. Cranberry concentrate tablets are currently available at nutrition stores and are sold as promoters of urinary tract health. After one of our patients with a distant history of calcium oxalate nephrolithiasis developed recurrent stones following self-administration of cranberry concentrate tablets, we sought to investigate the potential lithogenic properties of cranberry supplements. **METHODS**: Five healthy volunteers on a normal diet provided 24-hour urine collection for pH, volume, creatinine, oxalate, calcium, phosphate, uric acid, sodium, citrate, magnesium, and potassium. Cranberry tablets were administered to these volunteers at the manufacturer's recommended dosage for 7 days. On the seventh day, a second 24-hour urine collection was obtained. **RESULTS**: The urinary oxalate levels in the volunteers significantly increased (P = 0.01) by an average of 43.4% while receiving cranberry tablets. The excretion of potential lithogenic ions calcium, phosphate, and sodium also increased. However, inhibitors of stone formation, magnesium and potassium, rose as well. **CONCLUSIONS**: Cranberry concentrate tablets are marketed for urinary tract ailments. Physicians and manufacturers of cranberry products should make an effort to educate patients at risk for nephrolithiasis against ingestion of these dietary supplements.

**URL:** <https://ovidsp.ovid.com/ovidweb.cgi?T=JS&CSC=Y&NEWS=N&PAGE=fulltext&D=med4&DO=10.1016%2fs0090-4295%2800%2900884-0> <https://libkey.io/libraries/2592/openurl?genre=article&sid=OVID:medline&id=pmid:11164137&id=doi:10.1016%2Fs0090-4295%2800%2900884-0&issn=0090-4295&isbn=&volume=57&issue=1&spage=26&pages=26-9&date=2001&title=Urology&atitle=Dietary+supplementation+with+cranberry+concentrate+tablets+may+increase+the+risk+of+nephrolithiasis.&aulast=Terris&pid=%3Cauthor%3ETerris+MK%3BIssa+MM%3BTacker+JR%3C%2Fauthor%3E%3CAN%3E11164137%3C%2FAN%3E%3CDT%3EJournal+Article%3C%2FDT%3E>

229. **A family-based study of metabolic phenotypes in calcium urolithiasis.**

**Item Type:** Journal Article

**Authors:** Tessier, J.;Petrucci, M.;Trouve, M. L.;Valiquette, L.;Guay, G.;Ouimet, D. and Bonnardeaux, A.

**Publication Date:** Se ,2001

**Journal:** Kidney International 60(3), pp. 1141–1147

**Abstract: BACKGROUND**: A family history increases the risk of kidney stone passage independent of dietary risk factors. However, the metabolic basis for familial aggregation of urolithiasis is unknown. **METHODS**: We evaluated metabolic risk factors in families with at least two sibs with a history of calcium stones. Sibs underwent outpatient evaluations simultaneously, including 24-hour urine collection and oral calcium loading. Phenotypes were compared between affected and unaffected sibs from the same sibship. **RESULTS**: Eighty-three sibships comprising 388 sibs (212 affected sibs, 114 males and 98 females, and 176 unaffected sibs, 68 males and 108 females) from 71 families were analyzed. Daily urine calcium excretion was higher in affected compared with unaffected sibs (0.64 +/- 0.33 vs. 0.50 +/- 0.22 mmol Ca(2+)/mmol creatinine, respectively, P : Eighty-three sibships comprising 388 sibs (212 affected sibs, 114 males and 98 females, and 176 unaffected sibs, 68 males and 108 females) from 71 families were analyzed. Daily urine calcium excretion was higher in affected compared with unaffected sibs (0.64 +/- 0.33 vs. 0.50 +/- 0.22 mmol Ca(2+)/mmol creatinine, respectively, P : Eighty-three sibships comprising 388 sibs (212 affected sibs, 114 males and 98 females, and 176 unaffected sibs, 68 males and 108 females) from 71 families were analyzed. Daily urine calcium excretion was higher in affected compared with unaffected sibs (0.64 +/- 0.33 vs. 0.50 +/- 0.22 mmol Ca(2+)/mmol creatinine, respectively, P **CONCLUSION**: Increased urine calcium excretion is the only phenotype associated with a kidney stone formation in these French-Canadian families.

**URL:** <https://ovidsp.ovid.com/ovidweb.cgi?T=JS&CSC=Y&NEWS=N&PAGE=fulltext&D=med4&DO=10.1046%2fj.1523-1755.2001.0600031141.x> <https://libkey.io/libraries/2592/openurl?genre=article&sid=OVID:medline&id=pmid:11532110&id=doi:10.1046%2Fj.1523-1755.2001.0600031141.x&issn=0085-2538&isbn=&volume=60&issue=3&spage=1141&pages=1141-7&date=2001&title=Kidney+International&atitle=A+family-based+study+of+metabolic+phenotypes+in+calcium+urolithiasis.&aulast=Tessier&pid=%3Cauthor%3ETessier+J%3BPetrucci+M%3BTrouve+ML%3BValiquette+L%3BGuay+G%3BOuimet+D%3BBonnardeaux+A%3C%2Fauthor%3E%3CAN%3E11532110%3C%2FAN%3E%3CDT%3EJournal+Article%3C%2FDT%3E>

230. **Effect of potential renal acid load of foods on calcium metabolism of renal calcium stone formers.**

**Item Type:** Journal Article

**Authors:** Trinchieri, A.;Zanetti, G.;Curro, A. and Lizzano, R.

**Publication Date:** 2001

**Journal:** European Urology 39(Suppl 2), pp. 33–36

**Abstract: OBJECTIVES**: Diet has been proposed as a causative factor of hypercalciuria in patients with calcium stones. The aim of this study was to investigate the influence of diet on calcium metabolism of renal stone formers. **METHODS**: Thirty-five renal calcium stone formers were entered in this study. A 2-day recall of dietary intake was obtained from each subject. The food records were coded and computer analyzed for total energy, protein, fat, carbohydrate, sodium, potassium, calcium, magnesium, phosphate, oxalate, vitamin C and fiber. Daily potential renal acid load (PRAL) of the diet was calculated considering the mineral and protein composition of foods, the mean intestinal absorption rate for each nutrient and the metabolism of sulfur-containing amino acids. A fasting blood sample was drawn and a 24-hour urine collection were obtained for analyses of calcium, phosphate and creatinine. Serum osteocalcin was also analyzed. A fasting 2-hour urine sample was collected in the morning for hydroxyproline, pyridinium cross-links and creatinine. **RESULTS**: The mean daily dietary PRAL of renal stone formers was 22.4 +/- 15.7 (range 4.2-65.8) mEq/day. Regression analysis demonstrated that urinary calcium excretion is dependent on daily protein intake and dietary PRAL, whereas the urinary pyridinium cross-links/creatinine ratio is inversely dependent on daily calcium intake. The urinary pyridinium cross-links/creatinine ratio was significantly lower in patients on a low calcium diet (: The mean daily dietary PRAL of renal stone formers was 22.4 +/- 15.7 (range 4.2-65.8) mEq/day. Regression analysis demonstrated that urinary calcium excretion is dependent on daily protein intake and dietary PRAL, whereas the urinary pyridinium cross-links/creatinine ratio is inversely dependent on daily calcium intake. The urinary pyridinium cross-links/creatinine ratio was significantly lower in patients on a low calcium diet (**CONCLUSIONS**: The urinary calcium excretion of renal stone formers seems to be dependent on dietary acid load rather than dietary calcium intake. In patients consuming an acidifying diet a restriction of calcium intake could increase bone resorption leading to a progressive bone loss.

**URL:** <https://ovidsp.ovid.com/ovidweb.cgi?T=JS&CSC=Y&NEWS=N&PAGE=fulltext&D=med4&DO=10.1159%2f000052556> <https://libkey.io/libraries/2592/openurl?genre=article&sid=OVID:medline&id=pmid:11223695&id=doi:10.1159%2F000052556&issn=0302-2838&isbn=&volume=39&issue=2&spage=33&pages=33-6%3B+discussion+36-7&date=2001&title=European+Urology&atitle=Effect+of+potential+renal+acid+load+of+foods+on+calcium+metabolism+of+renal+calcium+stone+formers.&aulast=Trinchieri&pid=%3Cauthor%3ETrinchieri+A%3BZanetti+G%3BCurro+A%3BLizzano+R%3C%2Fauthor%3E%3CAN%3E11223695%3C%2FAN%3E%3CDT%3EJournal+Article%3C%2FDT%3E>

231. **Decreased urinary citrate excretion in type 1a glycogen storage disease.**

**Item Type:** Journal Article

**Authors:** Weinstein, D. A.;Somers, M. J. and Wolfsdorf, J. I.

**Publication Date:** Ma ,2001

**Journal:** Journal of Pediatrics 138(3), pp. 378–382

**Abstract: OBJECTIVES**: To quantify urinary citrate and calcium excretion and systemic acid-base status in patients with type 1a glycogen storage disease (GSD1a) and to investigate their relationship to renal complications. **STUDY DESIGN**: Fifteen patients (7 male and 8 female; age range, 3--28 years) were studied during annual evaluations of metabolic control. All were treated with intermittent doses of uncooked cornstarch. Hourly blood sampling and a 24-hour urine collection were obtained while subjects followed their usual home dietary regimen. **RESULTS**: All but the youngest subject had low levels of citrate excretion (mean 2.4 +/- 1.8 mg/kg/d; 129 +/- 21 mg citrate/g creatinine). Normally, urinary citrate excretion increases with age; however, in patients with GSD1a, a strong inverse exponential relationship was found between age and citrate excretion (r = -0.84, P : All but the youngest subject had low levels of citrate excretion (mean 2.4 +/- 1.8 mg/kg/d; 129 +/- 21 mg citrate/g creatinine). Normally, urinary citrate excretion increases with age; however, in patients with GSD1a, a strong inverse exponential relationship was found between age and citrate excretion (r = -0.84, P **CONCLUSIONS**: Hypocitraturia that worsens with age occurs in metabolically compensated patients with GSD1a. The combination of low citrate excretion and hypercalciuria appears to be important in the pathogenesis of nephrocalcinosis and nephrolithiasis. Citrate supplementation may be beneficial in preventing or ameliorating nephrocalcinosis and the development of urinary calculi in GSD1a.

**URL:** <https://ovidsp.ovid.com/ovidweb.cgi?T=JS&CSC=Y&NEWS=N&PAGE=fulltext&D=med4&DO=10.1067%2fmpd.2001.111322> <https://libkey.io/libraries/2592/openurl?genre=article&sid=OVID:medline&id=pmid:11241046&id=doi:10.1067%2Fmpd.2001.111322&issn=0022-3476&isbn=&volume=138&issue=3&spage=378&pages=378-82&date=2001&title=Journal+of+Pediatrics&atitle=Decreased+urinary+citrate+excretion+in+type+1a+glycogen+storage+disease.&aulast=Weinstein&pid=%3Cauthor%3EWeinstein+DA%3BSomers+MJ%3BWolfsdorf+JI%3C%2Fauthor%3E%3CAN%3E11241046%3C%2FAN%3E%3CDT%3EJournal+Article%3C%2FDT%3E>

232. **The risk of renal stone formation during and after long duration space flight.**

**Item Type:** Journal Article

**Authors:** Whitson, P. A.;Pietrzyk, R. A.;Morukov, B. V. and Sams, C. F.

**Publication Date:** No ,2001

**Journal:** Nephron 89(3), pp. 264–270

**Abstract: BACKGROUND**: The formation of a renal stone during space flight may have serious negative effects on the health of the crewmember and the success of the mission. Urinary biochemical factors and the influence of dietary factors associated with renal stone development were assessed during long duration Mir Space Station missions. **METHODS**: Twenty-four-hour urine samples were collected prior to, during and following long duration space flight. The relative urinary supersaturation of calcium oxalate, calcium phosphate (brushite), sodium urate, struvite and uric acid were determined. **RESULTS**: Changes in the urinary biochemistry of crewmembers during long duration spaceflight demonstrated increases in the supersaturation of the stone-forming salts. In-flight hypercalciuria was evident in a number of individual crewmembers and 24-hour dietary fluid intake and urine volume were significantly lower. During flight, there was a significant increase in brushite supersaturation. **CONCLUSIONS**: These data suggest acute effects of space flight and postflight changes in the urinary biochemistry favoring increased crystallization in the urine. The effects of dietary intake, especially fluid intake, may have a significant impact on the potential for renal stone formation. Efforts are now underway to assess the efficacy of a countermeasure to mitigate the increased risk. Copyright 2001 S. Karger AG, Basel

**URL:** <https://ovidsp.ovid.com/ovidweb.cgi?T=JS&CSC=Y&NEWS=N&PAGE=fulltext&D=med4&DO=10.1159%2f000046083> <https://libkey.io/libraries/2592/openurl?genre=article&sid=OVID:medline&id=pmid:11598387&id=doi:10.1159%2F000046083&issn=1660-8151&isbn=&volume=89&issue=3&spage=264&pages=264-70&date=2001&title=Nephron&atitle=The+risk+of+renal+stone+formation+during+and+after+long+duration+space+flight.&aulast=Whitson&pid=%3Cauthor%3EWhitson+PA%3BPietrzyk+RA%3BMorukov+BV%3BSams+CF%3C%2Fauthor%3E%3CAN%3E11598387%3C%2FAN%3E%3CDT%3EJournal+Article%3C%2FDT%3E>

233. **Why oral calcium supplements may reduce renal stone disease: report of a clinical pilot study.**

**Item Type:** Journal Article

**Authors:** Williams, C. P.;Child, D. F.;Hudson, P. R.;Davies, G. K.;Davies, M. G.;John, R.;Anandaram, P. S. and De Bolla, A. R.

**Publication Date:** Ja ,2001

**Journal:** Journal of Clinical Pathology 54(1), pp. 54–62

**Abstract: AIMS**: To investigate whether increasing the daily baseline of gut calcium can cause a gradual downregulation of the active intestinal transport of calcium via reduced parathyroid hormone (PTH) mediated activation of vitamin D, and to discuss why such a mechanism might prevent calcium oxalate rich stones. To demonstrate the importance of seasonal effects upon the evaluation of such data. **METHODS**: Within an intensive 24 hour urine collection regimen, daily calcium supplementation (500 mg) was given to five stone formers for a 10 week period during a six month crossover study. In a further population of patients on follow up for previous renal stone disease, observations were made on 1066 24 hour urine samples collected over five years in respect of seasonal effects relevant to the interpretation of the study. **RESULTS**: In the group of patients on calcium supplements the following results were found. During calcium supplementation, the proportion of urine calcium to oxalate was higher (increased calcium to oxalate molar ratio), the 24 hour urine product of calcium and oxalate did not rise, and urine oxalate was lower during the first six weeks of supplementation. Twenty four hour urine calcium was 10.2% higher than baseline in the final four weeks of the 10 weeks of supplementation. Twenty four hour urine phosphate was 11.4% lower during the first six weeks of supplementation, but then rose while the patients were still on supplementation; renal tubular reabsorption of phosphate (TmP/GFR) mirrored the urine phosphate changes inversely. PTH was higher after stopping supplementation, but 1,25-(OH)2-cholecalciferol changes were not detected. In the 1066 urine samples collected over five years the following results were found. Calcium and oxalate excretion correlated positively and not inversely. Urine calcium and phosphate excretion were 5.5% and 2.5% higher, respectively, in "light" months of the year compared with "dark" months. A post summer decline in both urine calcium and urine phosphate was relevant to the interpretation of the study. **CONCLUSIONS**: Regular calcium supplementation does not raise the product of calcium and oxalate in urine and the proportion of oxalate to calcium is reduced. The underlying mechanisms of the changes seen in phosphate, calcium, and PTH and the observations on 1,25-(OH)2-cholecalciferol are not clear. Observed changes in phosphate could possibly be part of a calcium regulating feedback loop operating over a period of weeks. In evaluating these mechanisms background seasonal effects are important. It is possible that "programming" of the gut mucosa in terms of calcium transport is a major determinant of the relation between calcium and oxalate concentrations in urine and their relative abundance. Increased oral calcium, in association with a reduction of the relative proportion absorbed, may be pertinent to the prevention of calcium oxalate rich stones.

**URL:** <https://ovidsp.ovid.com/ovidweb.cgi?T=JS&CSC=Y&NEWS=N&PAGE=fulltext&D=med4&DO=10.1136%2fjcp.54.1.54> <https://libkey.io/libraries/2592/openurl?genre=article&sid=OVID:medline&id=pmid:11271790&id=doi:10.1136%2Fjcp.54.1.54&issn=0021-9746&isbn=&volume=54&issue=1&spage=54&pages=54-62&date=2001&title=Journal+of+Clinical+Pathology&atitle=Why+oral+calcium+supplements+may+reduce+renal+stone+disease%3A+report+of+a+clinical+pilot+study.&aulast=Williams&pid=%3Cauthor%3EWilliams+CP%3BChild+DF%3BHudson+PR%3BDavies+GK%3BDavies+MG%3BJohn+R%3BAnandaram+PS%3BDe+Bolla+AR%3C%2Fauthor%3E%3CAN%3E11271790%3C%2FAN%3E%3CDT%3EClinical+Trial%3C%2FDT%3E>

234. **Contributory metabolic factors in the development of nephrolithiasis in patients with medullary sponge kidney.**

**Item Type:** Journal Article

**Authors:** Yagisawa, T.;Kobayashi, C.;Hayashi, T.;Yoshida, A. and Toma, H.

**Publication Date:** Ju ,2001

**Journal:** American Journal of Kidney Diseases 37(6), pp. 1140–1143

**Abstract:** Whether specific metabolic abnormalities are related to nephrolithiasis in patients with medullary sponge kidney (MSK) remains a debated issue. The purpose of this study is to determine metabolic disorders in patients with MSK and nephrolithiasis compared with idiopathic calcium-stone-forming patients. One hundred eighty-four patients with recurrent calcium-stone formations were investigated with regard to metabolic abnormalities. Of these, 22 patients (11.9%; 13 men, 9 women) showed MSK by radiological examination. MSK was defined as a kidney that presented at least three linear or round papillary opacities in the affected papilla on urography. Multiple stones (more than five) existed in both kidneys in all patients with MSK. The remaining 162 patients (109 men, 53 women) were idiopathic calcium-stone formers. Frequencies of low urine volume (urine 40 mg/24 h) were similar between the groups. Hypercalciuria (men, calcium > 300 mg/24 h; women, calcium of 250 mg/24 h) was found less frequently in the MSK group. The frequency of hypocitraturia (citrate < 300 mg/24 h) was significantly greater in the MSK group than the idiopathic group (77.3% versus 33.9%, respectively). Mean 24-hour urinary excretions of calcium, citrate, uric acid, and magnesium were significantly less in the MSK group. No differences were found in serum calcium, phosphate, and parathyroid hormone levels between the groups. Low urinary excretions of citrate and magnesium are the most typical metabolic disorders that distinguish MSK stone patients from idiopathic calcium-stone-forming patients. In addition to such anatomic abnormalities as ectatic collecting ducts, low levels of urinary inhibitors of stones seem to contribute to the pathogenesis of nephrolithiasis in patients with MSK.

**URL:** <https://ovidsp.ovid.com/ovidweb.cgi?T=JS&CSC=Y&NEWS=N&PAGE=fulltext&D=med4&DO=10.1053%2fajkd.2001.24515> <https://libkey.io/libraries/2592/openurl?genre=article&sid=OVID:medline&id=pmid:11382681&id=doi:10.1053%2Fajkd.2001.24515&issn=0272-6386&isbn=&volume=37&issue=6&spage=1140&pages=1140-3&date=2001&title=American+Journal+of+Kidney+Diseases&atitle=Contributory+metabolic+factors+in+the+development+of+nephrolithiasis+in+patients+with+medullary+sponge+kidney.&aulast=Yagisawa&pid=%3Cauthor%3EYagisawa+T%3BKobayashi+C%3BHayashi+T%3BYoshida+A%3BToma+H%3C%2Fauthor%3E%3CAN%3E11382681%3C%2FAN%3E%3CDT%3EJournal+Article%3C%2FDT%3E>

235. **Comparative effects of potassium chloride and bicarbonate on thiazide-induced reduction in urinary calcium excretion.**

**Item Type:** Journal Article

**Authors:** Frassetto, L. A.;Nash, E.;Morris, R. C. Jr and Sebastian, A.

**Publication Date:** Au ,2000

**Journal:** Kidney International 58(2), pp. 748–752

**Abstract: BACKGROUND**: The chronic low-grade metabolic acidosis that occurs in various renal disorders and in normal people, and that is related both to dietary net acid load and age-related renal functional decline, may contribute to osteoporosis by increasing urine calcium excretion. Administration of potassium (K) alkali salts neutralizes acid and lowers urine calcium excretion. Urine calcium excretion also can be reduced by the administration of thiazide diuretics, which are often given with supplemental K to avoid hypokalemia. We determined whether the K alkali salt potassium bicarbonate (KHCO3) and the thiazide diuretic hydrochlorothiazide (HCTZ) combined is more effective in reducing urinary calcium than KHCO3 alone or HCTZ combined with the conventionally coadministered nonalkalinizing K salt potassium chloride (KCl). **METHODS**: Thirty-one healthy men and women aged 50 or greater were recruited for a four-week, double-blind, randomized study. After a baseline period of 10 days with three 24-hour urine and arterialized blood collections, subjects were randomized to receive either HCTZ (50 mg) plus potassium (60 mmol daily) as either the chloride or bicarbonate salt. Another 19 women received potassium bicarbonate (60 mmol) alone. After two weeks, triplicate collections of 24-hour urines and arterialized bloods were repeated. **RESULTS**: Urinary calcium excretion decreased significantly in all groups. KHCO3 alone and HCTZ + KCl induced similar decreases (-0.70 +/- 0.60 vs. -0.80 +/- 1. 0 mmol/day, respectively). Compared with those treatments, the combination of HCTZ + KHCO3 induced more than a twofold greater decrease in urinary calcium excretion (-1.8 +/- 1.2 mmol/day, P : Urinary calcium excretion decreased significantly in all groups. KHCO3 alone and HCTZ + KCl induced similar decreases (-0.70 +/- 0.60 vs. -0.80 +/- 1. 0 mmol/day, respectively). Compared with those treatments, the combination of HCTZ + KHCO3 induced more than a twofold greater decrease in urinary calcium excretion (-1.8 +/- 1.2 mmol/day, P : Urinary calcium excretion decreased significantly in all groups. KHCO3 alone and HCTZ + KCl induced similar decreases (-0.70 +/- 0.60 vs. -0.80 +/- 1. 0 mmol/day, respectively). Compared with those treatments, the combination of HCTZ + KHCO3 induced more than a twofold greater decrease in urinary calcium excretion (-1.8 +/- 1.2 mmol/day, P **CONCLUSIONS**: KHCO3 was superior to KCl as an adjunct to HCTZ, inducing a twofold greater reduction in urine calcium excretion, and completely neutralizing endogenous acid production so as to correct the pre-existing mild metabolic acidosis that an acid-producing diet usually induces in older people. Accordingly, for reducing urine calcium excretion in stone disease and osteoporosis, the combination of HCTZ + KHCO3 may be preferable to that of HCTZ + KCl.

**URL:** <https://ovidsp.ovid.com/ovidweb.cgi?T=JS&CSC=Y&NEWS=N&PAGE=fulltext&D=med4&DO=10.1046%2fj.1523-1755.2000.00221.x> <https://libkey.io/libraries/2592/openurl?genre=article&sid=OVID:medline&id=pmid:10916098&id=doi:10.1046%2Fj.1523-1755.2000.00221.x&issn=0085-2538&isbn=&volume=58&issue=2&spage=748&pages=748-52&date=2000&title=Kidney+International&atitle=Comparative+effects+of+potassium+chloride+and+bicarbonate+on+thiazide-induced+reduction+in+urinary+calcium+excretion.&aulast=Frassetto&pid=%3Cauthor%3EFrassetto+LA%3BNash+E%3BMorris+RC+Jr%3BSebastian+A%3C%2Fauthor%3E%3CAN%3E10916098%3C%2FAN%3E%3CDT%3EClinical+Trial%3C%2FDT%3E>

236. **Effects of calcium and magnesium on urinary oxalate excretion after oxalate loads.**

**Item Type:** Journal Article

**Authors:** Liebman, M. and Costa, G.

**Publication Date:** Ma ,2000

**Journal:** Journal of Urology 163(5), pp. 1565–1569

**Abstract: PURPOSE**: Urinary oxalate is a primary determinant of the level of calcium oxalate saturation and the formation of calcium oxalate crystals, a key event in kidney stone formation. The primary objective of this study was to compare the effects of calcium carbonate and magnesium oxide on oxalate absorption. **MATERIALS AND METHODS**: An experimental model was used that allowed differentiation between endogenously and oxalate load-derived urinary oxalate. Twenty-four healthy subjects (10 males, 14 females) participated in three oxalate load (OL) tests: control (OL alone), calcium carbonate (OL with concomitant calcium carbonate ingestion), and magnesium oxide (OL with concomitant magnesium oxide ingestion). Oxalate loads consisted of 180 mg. unlabeled and 18 mg. 1,2[13C2] oxalic acid. Timed urine samples were collected after the OL for analysis of oxalate, calcium, magnesium, and creatinine. **RESULTS**: Both the calcium carbonate and magnesium oxide treatments were associated with significantly lower load-derived oxalate levels at all time points within the initial 24-hour post-oxalate ingestion period compared with levels observed for the control treatment. There were no treatment effects on endogenous oxalate levels. The efficiency of oxalate absorption for the calcium carbonate (5.1%) and magnesium oxide (7.6%) treatments was significantly lower than that for the control treatment (13.5%). **CONCLUSIONS**: The results suggested that magnesium was nearly as effective as calcium in reducing oxalate absorption and urinary excretion. Higher levels of urinary oxalate, calcium, and magnesium in males appeared to be largely a function of body size since gender differences either disappeared or were reversed when a correction was made for urinary creatinine excretion.

**URL:** <https://ovidsp.ovid.com/ovidweb.cgi?T=JS&CSC=Y&NEWS=N&PAGE=fulltext&D=med4&AN=10751889> <https://libkey.io/libraries/2592/openurl?genre=article&sid=OVID:medline&id=pmid:10751889&id=doi:&issn=0022-5347&isbn=&volume=163&issue=5&spage=1565&pages=1565-9&date=2000&title=Journal+of+Urology&atitle=Effects+of+calcium+and+magnesium+on+urinary+oxalate+excretion+after+oxalate+loads.&aulast=Liebman&pid=%3Cauthor%3ELiebman+M%3BCosta+G%3C%2Fauthor%3E%3CAN%3E10751889%3C%2FAN%3E%3CDT%3EComparative+Study%3C%2FDT%3E>

237. **High sodium chloride intake is associated with low bone density in calcium stone-forming patients.**

**Item Type:** Journal Article

**Authors:** Martini, L. A.;Cuppari, L.;Colugnati, F. A.;Sigulem, D. M.;Szejnfeld, V. L.;Schor, N. and Heilberg, I. P.

**Publication Date:** Au ,2000

**Journal:** Clinical Nephrology 54(2), pp. 85–93

**Abstract: BACKGROUND**: Although renal stone disease has been associated with reduced bone mass, the impact of nutrient intake on bone loss is unknown. **SUBJECTS AND METHODS**: The present study was undertaken to investigate the influence of nutrient intake on bone density of 85 calcium stone-forming (CSF) patients (47 male and 38 premenopausal females) aged 41+/-11 years (X+/-SD). Bone mineral density (BMD) was measured using dual energy X-ray absorptiometry at the lumbar spine (L2-L4) and femoral neck sites, and low BMD was defined as a T score : The present study was undertaken to investigate the influence of nutrient intake on bone density of 85 calcium stone-forming (CSF) patients (47 male and 38 premenopausal females) aged 41+/-11 years (X+/-SD). Bone mineral density (BMD) was measured using dual energy X-ray absorptiometry at the lumbar spine (L2-L4) and femoral neck sites, and low BMD was defined as a T score **RESULTS**: Forty-eight patients (56%) presented normal BMD and 37 (44%) low BMD. There were no statistical differences regarding age, weight, height, body mass index, protein, calcium and phosphorus intakes between both groups. The mean U(Ca), phosphorus and nitrogen appearance also did not differ between groups. However, there was a higher percentage of hypercalciuria among low vs normal BMD patients (62 vs 33%, p : Forty-eight patients (56%) presented normal BMD and 37 (44%) low BMD. There were no statistical differences regarding age, weight, height, body mass index, protein, calcium and phosphorus intakes between both groups. The mean U(Ca), phosphorus and nitrogen appearance also did not differ between groups. However, there was a higher percentage of hypercalciuria among low vs normal BMD patients (62 vs 33%, p or = 16 g/day was also higher among low vs normal BMD patients (35 vs 12%, p or = 16 g/day) was the single variable that was predictive of risk of low bone density in CSF patients (odds ratio = 3.8). **CONCLUSION**: These data suggest that reducing salt intake should be recommended for CSF patients presenting hypercalciuria and osteopenia.

**URL:** <https://ovidsp.ovid.com/ovidweb.cgi?T=JS&CSC=Y&NEWS=N&PAGE=fulltext&D=med4&AN=10968683> <https://libkey.io/libraries/2592/openurl?genre=article&sid=OVID:medline&id=pmid:10968683&id=doi:&issn=0301-0430&isbn=&volume=54&issue=2&spage=85&pages=85-93&date=2000&title=Clinical+Nephrology&atitle=High+sodium+chloride+intake+is+associated+with+low+bone+density+in+calcium+stone-forming+patients.&aulast=Martini&pid=%3Cauthor%3EMartini+LA%3BCuppari+L%3BColugnati+FA%3BSigulem+DM%3BSzejnfeld+VL%3BSchor+N%3BHeilberg+IP%3C%2Fauthor%3E%3CAN%3E10968683%3C%2FAN%3E%3CDT%3EJournal+Article%3C%2FDT%3E>

238. **Metabolic risk factors in patients with ureteropelvic junction obstruction and renal calculi.**

**Item Type:** Journal Article

**Authors:** Matin, S. F. and Streem, S. B.

**Publication Date:** Ju ,2000

**Journal:** Journal of Urology 163(6), pp. 1676–1678

**Abstract: PURPOSE**: We performed a prospective study to determine the incidence and spectrum of metabolic abnormalities predisposing to stone formation in patients with ureteropelvic junction obstruction and renal calculi. **MATERIALS AND METHODS**: A total of 47 consecutive patients with congenital ureteropelvic junction obstruction underwent metabolic evaluation of stone risk factors. Of these patients 21 had associated stones (study group), while 26 did not (control group). Logistical regression, Wilcoxon rank sum and Fisher's exact tests were performed to determine whether there was a significant difference between these groups in regard to the presence of metabolic risk factors. **RESULTS**: Demographically and symptomatically the 2 groups were equivalent except that the study patients were older. The 24-hour urinary excretion of calcium was significantly higher in study than in the control patients (p = 0.007). While the incidence of hypercalciuria and hyperuricosuria was also higher in the study population, these differences were not significant (p = 0.08 and 0.07, respectively). **CONCLUSIONS**: Metabolic abnormalities predisposing to stone formation are present more frequently in patients with ureteropelvic junction obstruction who have associated stones compared to those who do not. As such, urinary stasis alone does not explain stone formation in these cases. Rather, the local physiological environment of urine likely has a predisposing role. In addition to restoring unobstructed urinary flow, consideration should be given to metabolic evaluation and prophylactic treatment for affected patients.

**URL:** <https://ovidsp.ovid.com/ovidweb.cgi?T=JS&CSC=Y&NEWS=N&PAGE=fulltext&D=med4&AN=10799158> <https://libkey.io/libraries/2592/openurl?genre=article&sid=OVID:medline&id=pmid:10799158&id=doi:&issn=0022-5347&isbn=&volume=163&issue=6&spage=1676&pages=1676-8&date=2000&title=Journal+of+Urology&atitle=Metabolic+risk+factors+in+patients+with+ureteropelvic+junction+obstruction+and+renal+calculi.&aulast=Matin&pid=%3Cauthor%3EMatin+SF%3BStreem+SB%3C%2Fauthor%3E%3CAN%3E10799158%3C%2FAN%3E%3CDT%3EJournal+Article%3C%2FDT%3E>

239. **Clinical use of cystine supersaturation measurements.**

**Item Type:** Journal Article

**Authors:** Nakagawa, Y.;Asplin, J. R.;Goldfarb, D. S.;Parks, J. H. and Coe, F. L.

**Publication Date:** No ,2000

**Journal:** Journal of Urology 164(5), pp. 1481–1485

**Abstract: PURPOSE**: We measured the concentration and solubility of cystine in urine from patients with cystinuria or calcium stones and from normal subjects to determine whether urine cystine supersaturation can be calculated from a standard nomogram of solubility versus pH or needs to be measured directly. We also evaluated whether increasing pH of the 24-hour collection recovered enough crystallized cystine to increase cystine supersaturation. **MATERIALS AND METHODS**: Cystine concentration, pH and usual stone risk factors were measured on 50 ml. aliquots of 24-hour collections from 24 patients with cystinuria, 22 calcium stone formers and 15 normal subjects. After 48 hours of incubation with sodium bicarbonate, a second aliquot was taken from the 24-hour collection for cystine concentration. The original urine at its ambient pH was incubated with an excess of cystine crystals for 24, 48, 72 or 96 hours at 37C to determine solubility and kinetics of equilibration. **RESULTS**: Cystine solubility varied so widely at any pH range that no predictive nomogram could be relied on for calculating supersaturation. Addition of sodium bicarbonate to the 24-hour urine significantly increased cystine concentration. Urine from stone formers had higher cystine solubility than urine from normal subjects. **CONCLUSIONS**: Clinical management of cystinuria can be improved by direct measurement of cystine solubility because it varies widely at any given pH. Increasing 24-hour collection pH with sodium bicarbonate additionally improves accuracy of supersaturation measurement by recovering crystallized cystine.

**URL:** <https://ovidsp.ovid.com/ovidweb.cgi?T=JS&CSC=Y&NEWS=N&PAGE=fulltext&D=med4&AN=11025687> <https://libkey.io/libraries/2592/openurl?genre=article&sid=OVID:medline&id=pmid:11025687&id=doi:&issn=0022-5347&isbn=&volume=164&issue=5&spage=1481&pages=1481-5&date=2000&title=Journal+of+Urology&atitle=Clinical+use+of+cystine+supersaturation+measurements.&aulast=Nakagawa&pid=%3Cauthor%3ENakagawa+Y%3BAsplin+JR%3BGoldfarb+DS%3BParks+JH%3BCoe+FL%3C%2Fauthor%3E%3CAN%3E11025687%3C%2FAN%3E%3CDT%3EJournal+Article%3C%2FDT%3E>

240. **Higher urinary potassium is associated with decreased stone growth after shock wave lithotripsy.**

**Item Type:** Journal Article

**Authors:** Pierratos, A.;Dharamsi, N.;Carr, L. K.;Ibanez, D.;Jewett, M. A. and Honey, R. J.

**Publication Date:** No ,2000

**Journal:** Journal of Urology 164(5), pp. 1486–1489

**Abstract: PURPOSE**: We correlated serum and urinary biochemical parameters with radiological evidence of stone growth after shock wave lithotripsy. **MATERIALS AND METHODS**: Biochemical parameters in serum and 24-hour urine collections of 359 patients were correlated with stone growth for 2 years after shock wave lithotripsy. Each patient underwent a minimum of 2 radiological studies at 3 and 12 months and plain abdominal x-ray at 24 months. The presence and size of stones were documented by a radiologist in blinded fashion. Stone growth was defined as measurable growth of a preexisting stone or new stone formation. **RESULTS**: A total of 209 patients remained stone-free or had no existing stone growth, while stone size decreased in 30. Of the remaining 120 patients with stone growth 72 had new growth and 48 had growth of preexisting stones. Urinary excretion of potassium was significantly higher in those without than with stone growth (mean 24-hour urine collection plus or minus standard deviation 62 +/- 27 versus 54 +/- 23 mmol., p = 0.009). The only parameter significantly associated with stone growth was urinary potassium. Linear regression revealed that for each 10 unit increase in urinary potassium there was a corresponding 2 mm. decrease in stone growth (p = 0.013). **CONCLUSIONS**: Our results indicate that increased urinary potassium excretion correlates with a decreased risk of stone growth up to 2 years after shock wave lithotripsy, implying that a high potassium diet may be beneficial for preventing stone growth. The effect of potassium supplementation on stone formation and growth must be investigated further.

**URL:** <https://ovidsp.ovid.com/ovidweb.cgi?T=JS&CSC=Y&NEWS=N&PAGE=fulltext&D=med4&AN=11025688> <https://libkey.io/libraries/2592/openurl?genre=article&sid=OVID:medline&id=pmid:11025688&id=doi:&issn=0022-5347&isbn=&volume=164&issue=5&spage=1486&pages=1486-9&date=2000&title=Journal+of+Urology&atitle=Higher+urinary+potassium+is+associated+with+decreased+stone+growth+after+shock+wave+lithotripsy.&aulast=Pierratos&pid=%3Cauthor%3EPierratos+A%3BDharamsi+N%3BCarr+LK%3BIbanez+D%3BJewett+MA%3BHoney+RJ%3C%2Fauthor%3E%3CAN%3E11025688%3C%2FAN%3E%3CDT%3EJournal+Article%3C%2FDT%3E>

241. **When and how to evaluate a patient with nephrolithiasis.**

**Item Type:** Journal Article

**Authors:** Rivers, K.;Shetty, S. and Menon, M.

**Publication Date:** Ma ,2000

**Journal:** Urologic Clinics of North America 27(2), pp. 203–213

**Abstract:** Nephrolithiasis is a common disorder that afflicts up to 12% of the population and continues to be a significant cause of patient injury. Evaluation of these patients should include the assessment of their comorbidities and underlying medical conditions. Patients who form stones can undergo a streamlined evaluation. A cause can be found in more than 90% of these patients. With medical treatment, stone-recurrence rates can be decreased by 85% for calcium oxalate stone formation, which affects a large proportion of patients. Introduction of nonspecific medical therapy in uncomplicated calcium stone disease may improve the quality of life for these patients and allow utilization a simple diagnostic protocol. This is in contrast to the previous recommendations of disease-specific therapy. Because patients without complications represent the majority of those who form stones, a simplified approach greatly reduces the cost of evaluation and treatment. Underlying medical conditions, however, require disease-specific therapy. The protocol represented here should aid physicians and patients in the approach to management of stone disease. The fasting calcium-load test is not required, and the entire evaluation can be performed in an ambulatory setting in two visits. Two 24-hour urine samples should be obtained on a random and restricted diet. Patients who form calcium stones can be subdivided into those who form hypercalciuric and normocalciuric stones. Patients who form normocalciuric stones are treated with conservative measures (increased fluid intake) and potassium and magnesium citrate. Patients who form hypercalciuric stones are treated with a combination of thiazides and potassium and magnesium citrate.

**URL:** <https://ovidsp.ovid.com/ovidweb.cgi?T=JS&CSC=Y&NEWS=N&PAGE=fulltext&D=med4&DO=10.1016%2fs0094-0143%2805%2970251-2> <https://libkey.io/libraries/2592/openurl?genre=article&sid=OVID:medline&id=pmid:10778464&id=doi:10.1016%2Fs0094-0143%2805%2970251-2&issn=0094-0143&isbn=&volume=27&issue=2&spage=203&pages=203-13&date=2000&title=Urologic+Clinics+of+North+America&atitle=When+and+how+to+evaluate+a+patient+with+nephrolithiasis.&aulast=Rivers&pid=%3Cauthor%3ERivers+K%3BShetty+S%3BMenon+M%3C%2Fauthor%3E%3CAN%3E10778464%3C%2FAN%3E%3CDT%3EJournal+Article%3C%2FDT%3E>

242. **Effects of low animal protein or high-fiber diets on urine composition in calcium nephrolithiasis.**

**Item Type:** Journal Article

**Authors:** Rotily, M.;Leonetti, F.;Iovanna, C.;Berthezene, P.;Dupuy, P.;Vazi, A. and Berland, Y.

**Publication Date:** Ma ,2000

**Journal:** Kidney International 57(3), pp. 1115–1123

**Abstract: BACKGROUND**: The purpose of this article is to evaluate the impact of low protein and high fiber intakes on risk factors of stone recurrence in idiopathic calcium stone formers (ICSFs). **METHODS**: Ninety-six ICSFs were randomly assigned a low animal protein diet ( 25 g/day), or a usual diet (control group); all patients were recommended to increase their fluid intake. Their daily urine compositions were analyzed at baseline and at four months. Compliance with dietary recommendations was checked by validated food frequency questionnaires. Compliance with total and animal protein intakes was assessed by 24-hour urea and sulphate outputs, respectively. The nutritional intervention (oral instructions, written leaflet, phoning) and food assessment were carried out by a research dietitian. **RESULTS**: At baseline, diets and the daily urine composition did not differ between the three groups. At four months, while diets differed significantly, the 24-hour output of calcium and oxalate did not differ significantly within and between groups after adjustment for potential confounders (age, sex, and personal and family history of calcium stones) and baseline values. However, as many as 12 out of 31 ICSFs (95% CI, 22 to 58%) assigned to a low animal protein diet achieved a reduction in the urine urea excretion rate of more than 50 mmol/day and also exhibited a significant decrease in urinary calcium excretion that averaged 1.8 mmol/day. A significant correlation between urea and calcium outputs was observed only among patients with hypercalciuria. **CONCLUSIONS**: These results show that only ICSFs who markedly decrease their animal protein intake, especially those with hypercalciuria, can expect to benefit from dietary recommendations.

**URL:** <https://ovidsp.ovid.com/ovidweb.cgi?T=JS&CSC=Y&NEWS=N&PAGE=fulltext&D=med4&DO=10.1046%2fj.1523-1755.2000.00939.x> <https://libkey.io/libraries/2592/openurl?genre=article&sid=OVID:medline&id=pmid:10720964&id=doi:10.1046%2Fj.1523-1755.2000.00939.x&issn=0085-2538&isbn=&volume=57&issue=3&spage=1115&pages=1115-23&date=2000&title=Kidney+International&atitle=Effects+of+low+animal+protein+or+high-fiber+diets+on+urine+composition+in+calcium+nephrolithiasis.&aulast=Rotily&pid=%3Cauthor%3ERotily+M%3BLeonetti+F%3BIovanna+C%3BBerthezene+P%3BDupuy+P%3BVazi+A%3BBerland+Y%3C%2Fauthor%3E%3CAN%3E10720964%3C%2FAN%3E%3CDT%3EClinical+Trial%3C%2FDT%3E>

243. **Prevention of recurrent nephrolithiasis**

**Item Type:** Journal Article

**Authors:** Goldfarb, D. S. and Coe, F. L.

**Publication Date:** Nov 15 ,1999

**Journal:** American Family Physician 60(8), pp. 2269–2276

**Abstract:** The first episode of nephrolithiasis provides an opportunity to advise patients about measures for preventing future stones. Low fluid intake and excessive intake of protein, salt and oxalate are important modifiable risk factors for kidney stones. Calcium restriction is not useful and may potentiate osteoporosis. Diseases such as hyperparathyroidism, sarcoidosis and renal tubular acidosis should be considered in patients with nephrolithiasis. A 24-hour urine collection with measurement of the important analytes is usually reserved for use in patients with recurrent stone formation. In these patients, the major urinary risk factors include hypercalciuria, hyperoxaluria, hypocitraturia and hyperuricosuria. Effective preventive and treatment measures include thiazide therapy to lower the urinary calcium level, citrate supplementation to increase the urinary citrate level and, sometimes, allopurinol therapy to lower uric acid excretion. Uric acid stones are most often treated with citrate supplementation. Data now support the cost-effectiveness of evaluation and treatment of patients with recurrent stones. [References: 34]

**URL:** <https://ovidsp.ovid.com/ovidweb.cgi?T=JS&CSC=Y&NEWS=N&PAGE=fulltext&D=med4&AN=10593318> <https://libkey.io/libraries/2592/openurl?genre=article&sid=OVID:medline&id=pmid:10593318&id=doi:&issn=0002-838X&isbn=&volume=60&issue=8&spage=2269&pages=2269-76&date=1999&title=American+Family+Physician&atitle=Prevention+of+recurrent+nephrolithiasis.&aulast=Goldfarb&pid=%3Cauthor%3EGoldfarb+DS%3BCoe+FL%3C%2Fauthor%3E%3CAN%3E10593318%3C%2FAN%3E%3CDT%3EJournal+Article%3C%2FDT%3E>

244. **Metabolic evaluation of stone disease patients: a practical approach**

**Item Type:** Journal Article

**Authors:** Lifshitz, D. A.;Shalhav, A. L.;Lingeman, J. E. and Evan, A. P.

**Publication Date:** No ,1999

**Journal:** Journal of Endourology 13(9), pp. 669–678

**Abstract:** In the last three decades, minimally invasive techniques have progressed significantly, replacing traditional open surgery as the mainstay of stone disease surgical treatment. The challenge for the next millennium remains medical prevention of calcium urolithiasis, a field where less dramatic progress has been achieved during the same period of time. The purpose of this article is to provide the practicing urologist with current practical guidelines for the assessment and management of calcium urolithiasis patients. The recommendations are based on the latest available information regarding the pathogenesis, medical treatment options, and decision-making rationale when managing these challenging patients. Every urolithiasis patient should undergo a basic evaluation, which is considered the minimal essential diagnostic work-up, in order to rule out obvious, treatable systemic causes of urinary stone disease. All patients should be advised about conservative nonspecific preventive measures. High-risk stone patients should have a more extensive metabolic evaluation based on two 24-hour urine samples. Treatment protocols for each patient are tailored individually according to the metabolic evaluation findings. [References: 79]

**URL:** <https://ovidsp.ovid.com/ovidweb.cgi?T=JS&CSC=Y&NEWS=N&PAGE=fulltext&D=med4&DO=10.1089%2fend.1999.13.669> <https://libkey.io/libraries/2592/openurl?genre=article&sid=OVID:medline&id=pmid:10608520&id=doi:10.1089%2Fend.1999.13.669&issn=0892-7790&isbn=&volume=13&issue=9&spage=669&pages=669-78&date=1999&title=Journal+of+Endourology&atitle=Metabolic+evaluation+of+stone+disease+patients%3A+a+practical+approach.&aulast=Lifshitz&pid=%3Cauthor%3ELifshitz+DA%3BShalhav+AL%3BLingeman+JE%3BEvan+AP%3C%2Fauthor%3E%3CAN%3E10608520%3C%2FAN%3E%3CDT%3EJournal+Article%3C%2FDT%3E>

245. **A prospective study of recurrence rate and risk factors for recurrence after a first renal stone.**

**Item Type:** Journal Article

**Authors:** Trinchieri, A.;Ostini, F.;Nespoli, R.;Rovera, F.;Montanari, E. and Zanetti, G.

**Publication Date:** Ju ,1999

**Journal:** Journal of Urology 162(1), pp. 27–30

**Abstract: PURPOSE**: We investigate further the recurrence rate and risk factors for recurrence in 300 consecutive patients who presented to our stone clinic after a first stone episode 7 to 17 years ago. **MATERIALS AND METHODS**: The medical records of the patients who presented consecutively with a first stone episode from 1980 to 1990 were studied and supplemented by a followup mail questionnaire and telephone interviews. At first visit serum samples were taken from all patients and 24-hour urine samples were collected for metabolic testing. **RESULTS**: A total of 195 patients were followed successfully, of whom 52 (27%) experienced symptomatic stone recurrence after a mean plus or minus standard deviation of 7.5+/-5.9 years. However, ultrasound examination of 36 symptom-free patients showed recurrent stones in 28%. Comparison of patients with or without recurrence confirmed that recurrence was not influenced by sex, family history of stones and urinary risk factors. However, age at onset of the disease was lower for patients who had 2 or more stones during followup than those who had only 1 stone or no recurrence. **CONCLUSIONS**: Stones can recur as long as 10 years after the first episode, although the rate is lower than previously reported. The metabolic evaluation after a first stone episode needs to be reappraised in terms of its cost-effectiveness, since recurrences do not seem to be predictable from standard laboratory tests.

**URL:** <https://ovidsp.ovid.com/ovidweb.cgi?T=JS&CSC=Y&NEWS=N&PAGE=fulltext&D=med4&DO=10.1097%2f00005392-199907000-00007> <https://libkey.io/libraries/2592/openurl?genre=article&sid=OVID:medline&id=pmid:10379732&id=doi:10.1097%2F00005392-199907000-00007&issn=0022-5347&isbn=&volume=162&issue=1&spage=27&pages=27-30&date=1999&title=Journal+of+Urology&atitle=A+prospective+study+of+recurrence+rate+and+risk+factors+for+recurrence+after+a+first+renal+stone.&aulast=Trinchieri&pid=%3Cauthor%3ETrinchieri+A%3BOstini+F%3BNespoli+R%3BRovera+F%3BMontanari+E%3BZanetti+G%3C%2Fauthor%3E%3CAN%3E10379732%3C%2FAN%3E%3CDT%3EJournal+Article%3C%2FDT%3E>

246. **Metabolic urinary correlates of calcium oxalate dihydrate in renal stones.**

**Item Type:** Journal Article

**Authors:** Asplin, J. R.;Lingeman, J.;Kahnoski, R.;Mardis, H.;Parks, J. H. and Coe, F. L.

**Publication Date:** Ma ,1998

**Journal:** Journal of Urology 159(3), pp. 664–668

**Abstract: PURPOSE**: We determined what metabolic features of the 24-hour urine predict calcium oxalate dihydrate in kidney stones. Prior studies have suggested that low urine magnesium, high urine calcium, high calcium-to-oxalate ratio and high urine supersaturation with respect to calcium oxalate monohydrate predict calcium oxalate dihydrate. **MATERIALS AND METHODS**: Stone analyses and results from 2, 24-hour pretreatment urine collections from 96 patients with nephrolithiasis were drawn from 3 kidney stone prevention centers. Standard stone risk measurements were made on the urine, including supersaturation for calcium oxalate monohydrate, brushite and uric acid. **RESULTS**: The main differences in metabolic urine findings were between patients with no calcium oxalate dihydrate and those with any calcium oxalate dihydrate in stones. Percent calcium oxalate dihydrate itself did not correlate with urine findings. Patients with no calcium oxalate dihydrate in stones showed a biphasic pattern of urine calcium oxalate monohydrate supersaturation, about half had values below almost any found among patients with calcium oxalate dihydrate in stones (less than 7) and the rest overlapped with the calcium oxalate dihydrate group. Except for higher calcium oxalate monohydrate supersaturation, patients with calcium oxalate dihydrate in stones had higher urine calcium excretion and lower urine citrate concentrations, even after calcium oxalate monohydrate supersaturation was considered. **CONCLUSIONS**: Patients with low calcium oxalate monohydrate supersaturation (less than 7) are unlikely to have calcium oxalate dihydrate in renal stones. However, many patients with no calcium oxalate dihydrate have higher calcium oxalate monohydrate supersaturation values, and so prediction of calcium oxalate dihydrate or its absence from urine findings is imperfect. Urine magnesium and the calcium-to-oxalate ratio are unrelated to calcium oxalate dihydrate.

**URL:** <https://ovidsp.ovid.com/ovidweb.cgi?T=JS&CSC=Y&NEWS=N&PAGE=fulltext&D=med4&AN=9474121> <https://libkey.io/libraries/2592/openurl?genre=article&sid=OVID:medline&id=pmid:9474121&id=doi:&issn=0022-5347&isbn=&volume=159&issue=3&spage=664&pages=664-8&date=1998&title=Journal+of+Urology&atitle=Metabolic+urinary+correlates+of+calcium+oxalate+dihydrate+in+renal+stones.&aulast=Asplin&pid=%3Cauthor%3EAsplin+JR%3BLingeman+J%3BKahnoski+R%3BMardis+H%3BParks+JH%3BCoe+FL%3C%2Fauthor%3E%3CAN%3E9474121%3C%2FAN%3E%3CDT%3EJournal+Article%3C%2FDT%3E>

247. **Supersaturation and stone composition in a network of dispersed treatment sites.**

**Item Type:** Journal Article

**Authors:** Asplin, J.;Parks, J.;Lingeman, J.;Kahnoski, R.;Mardis, H.;Lacey, S.;Goldfarb, D.;Grasso, M. and Coe, F.

**Publication Date:** Ju ,1998

**Journal:** Journal of Urology 159(6), pp. 1821–1825

**Abstract: PURPOSE**: We determined the validity of urine supersaturation assessed from 2, 24-hour urine collections from outpatients eating uncontrolled diets and receiving care at a network of treatment sites that uses a central laboratory. We compared supersaturation to stone composition to determine whether supersaturation values correlate with composition. **MATERIALS AND METHODS**: Two 24-hour urine samples collected from 183 patients at 6 treatment sites were shipped to a single central laboratory. Complexations and crystallizations in vitro from aging during the transport step were interrupted by pH change in acid and alkaline directions. Relevant analytes were measured, and supersaturation was calculated for calcium oxalate, calcium phosphate as brushite and uric acid. Stone analysis was done at various laboratories. **RESULTS**: Urine supersaturation values correlated well with stone composition. Higher calcium phosphate and uric acid supersaturation was noted when stones contained higher amounts of calcium phosphate and any uric acid, respectively. In a validation study values of relevant urine materials were unchanged after 48 hours of aging. **CONCLUSIONS**: Despite the need for sample transport, resulting in the inevitable aging of samples, and variations in diet and details of sample collection, supersaturation values measured in only 2, 24-hour urine collections accurately reflected stone composition. This finding indicates that supersaturation values are reasonably stable in most patients during the months to years required for stones to form. In addition, samples collected in standard practice settings and sent to a central laboratory may accurately reflect these supersaturation values.

**URL:** <https://ovidsp.ovid.com/ovidweb.cgi?T=JS&CSC=Y&NEWS=N&PAGE=fulltext&D=med4&DO=10.1016%2fS0022-5347%2801%2963164-1> <https://libkey.io/libraries/2592/openurl?genre=article&sid=OVID:medline&id=pmid:9598467&id=doi:10.1016%2FS0022-5347%2801%2963164-1&issn=0022-5347&isbn=&volume=159&issue=6&spage=1821&pages=1821-5&date=1998&title=Journal+of+Urology&atitle=Supersaturation+and+stone+composition+in+a+network+of+dispersed+treatment+sites.&aulast=Asplin&pid=%3Cauthor%3EAsplin+J%3BParks+J%3BLingeman+J%3BKahnoski+R%3BMardis+H%3BLacey+S%3BGoldfarb+D%3BGrasso+M%3BCoe+F%3C%2Fauthor%3E%3CAN%3E9598467%3C%2FAN%3E%3CDT%3EJournal+Article%3C%2FDT%3E>

248. **Relative hyperoxaluria, crystalluria and haematuria after megadose ingestion of vitamin C.**

**Item Type:** Journal Article

**Authors:** Auer, B. L.;Auer, D. and Rodgers, A. L.

**Publication Date:** Se ,1998

**Journal:** European Journal of Clinical Investigation 28(9), pp. 695–700

**Abstract: BACKGROUND**: Long-term or high-dosage consumption of vitamin C may play a role in calcium oxalate kidney stone formation. The present study was undertaken to determine the biochemical and physicochemical risk factors in a male subject who developed haematuria and calcium oxalate crystalluria after ingestion of large doses of ascorbic acid for 8 consecutive days. **METHODS**: Twenty-four-hour urine samples were collected before and during the ascorbic acid ingestion period as well as after the detection of haematuria. A special procedure was implemented for urine collections to allow for oxalate, ascorbate and other urinalysis. Oxalate was determined in the presence of EDTA to prevent in vitro conversion to ascorbic acid, whereas ascorbate itself was determined by manual titration in a redox method using the dye dichlorophenolindophenol. Urinalysis data were used to compute calcium oxalate relative supersaturations and Tiselius risk indices, whereas scanning electron microscopy was used to examine urinary deposits. **RESULTS**: Oxalate excretion increased by about 350% during ascorbate ingestion before haematuria. Ascorbate concentrations also increased dramatically but appeared to reach a plateau maximum. Increasing calcium excretion was accompanied by decreasing potassium and phosphate values. The calcium oxalate relative supersaturation and Tiselius risk index increased during vitamin C ingestion and large aggregates of calcium oxalate dihydrate crystals were observed by scanning electron microscopy immediately after the detection of haematuria. **CONCLUSION**: High percentage metabolic conversion of ascorbate to oxalate in this subject caused relative hyperoxaluria and crystalluria, the latter manifesting itself as haematuria. Clinicians need to be alerted to the potential dangers of large dose ingestion of vitamin C in some individuals.

**URL:** <https://ovidsp.ovid.com/ovidweb.cgi?T=JS&CSC=Y&NEWS=N&PAGE=fulltext&D=med4&DO=10.1046%2fj.1365-2362.1998.00349.x> <https://libkey.io/libraries/2592/openurl?genre=article&sid=OVID:medline&id=pmid:9767367&id=doi:10.1046%2Fj.1365-2362.1998.00349.x&issn=0014-2972&isbn=&volume=28&issue=9&spage=695&pages=695-700&date=1998&title=European+Journal+of+Clinical+Investigation&atitle=Relative+hyperoxaluria%2C+crystalluria+and+haematuria+after+megadose+ingestion+of+vitamin+C.&aulast=Auer&pid=%3Cauthor%3EAuer+BL%3BAuer+D%3BRodgers+AL%3C%2Fauthor%3E%3CAN%3E9767367%3C%2FAN%3E%3CDT%3ECase+Reports%3C%2FDT%3E>

249. **CLCN5 mutation Ser244Leu is associated with X-linked renal failure without X-linked recessive hypophosphatemic rickets.**

**Item Type:** Journal Article

**Authors:** Kelleher, C. L.;Buckalew, V. M.;Frederickson, E. D.;Rhodes, D. J.;Conner, D. A.;Seidman, J. G. and Seidman, C. E.

**Publication Date:** Ja ,1998

**Journal:** Kidney International 53(1), pp. 31–37

**Abstract:** This study demonstrates that a missense mutation in the voltage gated chloride channel, CLCN5, can cause X-linked renal failure without X-linked recessive hypophosphatemic rickets. A large kindred (Family A), initially evaluated in 1974 with an inherited syndrome characterized by hypercalciuria, nephrocalcinosis, low molecular weight proteinuria, renal tubular acidosis, and renal failure, was clinically re-evaluated and genetically characterized. Medical histories, physical examinations, blood chemistries, and 24-hour urine collections were obtained from 48 family members. Both female and male family members exhibited hypercalciuria, nephrolithiasis, and low molecular weight proteinuria. However, only men developed renal insufficiency, consistent with an X-linked recessive gene defect. Genetic linkage located the disease locus on the proximal short arm of the X chromosome (Xp11) where a voltage gated chloride channel gene, CLCN5, had previously been mapped. DNA sequence of the CLCN5 gene demonstrated a missense mutation (Ser244Leu) in affected family members. The same missense mutation has previously been shown to cause X-linked recessive hypophosphatemic rickets. No affected member of Family A had evidence of chronic hypophosphatemia, clinically significant rickets, or osteomalacia. We hypothesize that genetic background, environment, diet, or an unidentified modifying gene may account for the differing phenotypes resulting from this shared gene defect.

**URL:** <https://ovidsp.ovid.com/ovidweb.cgi?T=JS&CSC=Y&NEWS=N&PAGE=fulltext&D=med4&DO=10.1046%2fj.1523-1755.1998.00752.x> <https://libkey.io/libraries/2592/openurl?genre=article&sid=OVID:medline&id=pmid:9452997&id=doi:10.1046%2Fj.1523-1755.1998.00752.x&issn=0085-2538&isbn=&volume=53&issue=1&spage=31&pages=31-7&date=1998&title=Kidney+International&atitle=CLCN5+mutation+Ser244Leu+is+associated+with+X-linked+renal+failure+without+X-linked+recessive+hypophosphatemic+rickets.&aulast=Kelleher&pid=%3Cauthor%3EKelleher+CL%3BBuckalew+VM%3BFrederickson+ED%3BRhodes+DJ%3BConner+DA%3BSeidman+JG%3BSeidman+CE%3C%2Fauthor%3E%3CAN%3E9452997%3C%2FAN%3E%3CDT%3EJournal+Article%3C%2FDT%3E>

250. **Medical reduction of stone risk in a network of treatment centers compared to a research clinic.**

**Item Type:** Journal Article

**Authors:** Lingeman, J.;Mardis, H.;Kahnoski, R.;Goldfarb, D. S.;Lacy, S.;Grasso, M.;Scheinman, S. J.;Parks, J. H.;Asplin, J. R. and Coe, F. L.

**Publication Date:** No ,1998

**Journal:** Journal of Urology 160(5), pp. 1629–1634

**Abstract: PURPOSE**: We determined whether a network of 7 comprehensive kidney stone treatment centers supported by specialized stone management software and laboratory resources could achieve reductions in urine supersaturation comparable to those in a single research clinic devoted to metabolic stone prevention. **MATERIALS AND METHODS**: Supersaturation values for calcium oxalate, calcium phosphate and uric acid in 24-hour urine samples were calculated from a set of kidney stone risk factor measurements made at a central laboratory site for the network and research laboratory for the clinic. Individual results and group outcomes were presented to each center in time sequential table graphics. The decrease in supersaturation with treatment was compared in the network and clinic using analysis of variance. **RESULTS**: Supersaturation was effectively reduced in the network and clinic, and the reduction was proportional to the initial supersaturation value and increase in urine volume. The clinic achieved a greater supersaturation reduction, higher fraction of patient followup and greater increase in urine volume but the treatment effects in the network were, nevertheless, substantial and significant. **CONCLUSIONS**: Given proper software and laboratory support, a network of treatment centers can rival but not quite match results in a dedicated metabolic stone research and prevention clinic. Therefore, large scale stone prevention in a network system appears feasible and effective.

**URL:** <https://ovidsp.ovid.com/ovidweb.cgi?T=JS&CSC=Y&NEWS=N&PAGE=fulltext&D=med4&AN=9783920> <https://libkey.io/libraries/2592/openurl?genre=article&sid=OVID:medline&id=pmid:9783920&id=doi:&issn=0022-5347&isbn=&volume=160&issue=5&spage=1629&pages=1629-34&date=1998&title=Journal+of+Urology&atitle=Medical+reduction+of+stone+risk+in+a+network+of+treatment+centers+compared+to+a+research+clinic.&aulast=Lingeman&pid=%3Cauthor%3ELingeman+J%3BMardis+H%3BKahnoski+R%3BGoldfarb+DS%3BLacy+S%3BGrasso+M%3BScheinman+SJ%3BParks+JH%3BAsplin+JR%3BCoe+FL%3C%2Fauthor%3E%3CAN%3E9783920%3C%2FAN%3E%3CDT%3EComparative+Study%3C%2FDT%3E>

251. **Potassium and sodium intake and excretion in calcium stone forming patients.**

**Item Type:** Journal Article

**Authors:** Martini, L. A.;Cuppari, L.;Cunha, M. A.;Schor, N. and Heilberg, I. P.

**Publication Date:** Ju ,1998

**Journal:** Journal of Renal Nutrition 8(3), pp. 127–131

**Abstract: OBJECTIVE**: To determine mean potassium (K) intake and its correlation with urinary calcium (uCa) and citrate excretion, as well as uCa, sodium (Na), and K levels of calcium stone forming patients. We determined the K-rich foods most commonly consumed by these patients. **DESIGN**: Case-control. **SETTING**: University-affiliated outpatient renal Lithiasis Unit. **PATIENTS AND CONTROLS**: One hundred hypercalciuric calcium stone forming patients (CSF, 54 men/46 women), 37 with associated hypocitraturia, were sequentially enrolled in the study that was performed before the initiation of any care for their renal stones. The control group consisted of 100 age-matched healthy subjects (HS, 47 men/53 women) who were laboratory employees with no history of renal stones. **INTERVENTION**: The analyses consisted of a 3-day dietary record to determine the mean K and calcium (Ca) intakes, and a 24-hour urine sample with measurements of K, Ca, Na, and citrate. **MAIN OUTCOME MEASURE**: K and Na intake determined by dietary record. **RESULTS**: uCa and Na levels and the Na/K ratio were significantly higher for CSF versus HS (238 +/- 118 v 148 +/- 74 mg/24 hours, 238 +/- 100 v 181 +/- 68 mEq/24 hours, 6.6 +/- 3.5 v 5.1 +/- 2.3, respectively, P : uCa and Na levels and the Na/K ratio were significantly higher for CSF versus HS (238 +/- 118 v 148 +/- 74 mg/24 hours, 238 +/- 100 v 181 +/- 68 mEq/24 hours, 6.6 +/- 3.5 v 5.1 +/- 2.3, respectively, P : uCa and Na levels and the Na/K ratio were significantly higher for CSF versus HS (238 +/- 118 v 148 +/- 74 mg/24 hours, 238 +/- 100 v 181 +/- 68 mEq/24 hours, 6.6 +/- 3.5 v 5.1 +/- 2.3, respectively, P : uCa and Na levels and the Na/K ratio were significantly higher for CSF versus HS (238 +/- 118 v 148 +/- 74 mg/24 hours, 238 +/- 100 v 181 +/- 68 mEq/24 hours, 6.6 +/- 3.5 v 5.1 +/- 2.3, respectively, P : uCa and Na levels and the Na/K ratio were significantly higher for CSF versus HS (238 +/- 118 v 148 +/- 74 mg/24 hours, 238 +/- 100 v 181 +/- 68 mEq/24 hours, 6.6 +/- 3.5 v 5.1 +/- 2.3, respectively, P **CONCLUSION**: Despite the consumption of K-rich foods at least once a day, the mean K intake by CSF patients was 58 mEq/day. This intake can still be considered to be low, although it meets recommended daily dietary allowance requirements. Therefore, we describe herein a population of CSF with high-Na intake and normal- to low-K intake, which may contribute to stone formation.

**URL:** <https://ovidsp.ovid.com/ovidweb.cgi?T=JS&CSC=Y&NEWS=N&PAGE=fulltext&D=med4&DO=10.1016%2fs1051-2276%2898%2990003-6> <https://libkey.io/libraries/2592/openurl?genre=article&sid=OVID:medline&id=pmid:9724501&id=doi:10.1016%2Fs1051-2276%2898%2990003-6&issn=1051-2276&isbn=&volume=8&issue=3&spage=127&pages=127-31&date=1998&title=Journal+of+Renal+Nutrition&atitle=Potassium+and+sodium+intake+and+excretion+in+calcium+stone+forming+patients.&aulast=Martini&pid=%3Cauthor%3EMartini+LA%3BCuppari+L%3BCunha+MA%3BSchor+N%3BHeilberg+IP%3C%2Fauthor%3E%3CAN%3E9724501%3C%2FAN%3E%3CDT%3EJournal+Article%3C%2FDT%3E>

252. **Substituting milk for apple juice does not increase kidney stone risk in most normocalciuric adults who form calcium oxalate stones.**

**Item Type:** Journal Article

**Authors:** Massey, L. K. and Kynast-Gales, S. A.

**Publication Date:** Ma ,1998

**Journal:** Journal of the American Dietetic Association 98(3), pp. 303–308

**Abstract: OBJECTIVE**: Increasing intake of dietary calcium from less than 400 mg to 800 mg daily may decrease the absorption of dietary oxalate, which in turn would decrease urinary oxalate excretion. The effect of substituting milk for apple juice on urine composition and risk of calcium oxalate precipitability was studied. **SUBJECTS**: Twenty-one normocalciuric adults with a history of at least 1 calcium oxalate stone and urinary oxalate excretion exceeding 275 micromol/day on their self-selected diet. **DESIGN**: Randomized crossover trial. **INTERVENTION**: Each participant consumed two moderate-oxalate (2,011 micromol/day) study diets, which were identical except that one contained 360 mL milk and the other contained 540 mL apple juice as the beverage with meals. **SETTING**: Four days free-living then 2 days in the metabolic unit of a university nutrition department. **MAIN OUTCOME MEASURE**: Tiselius risk index for calcium oxalate precipitability calculated from urine composition. **STATISTICAL ANALYSES**: Paired t tests. **RESULTS**: Twenty-four hour urinary oxalate excretion was 18% lower (P: Twenty-four hour urinary oxalate excretion was 18% lower (P: Twenty-four hour urinary oxalate excretion was 18% lower (P**APPLICATIONS**: Substituting 360 mL milk daily for apple juice with meals in a diet containing moderate amounts of dietary oxalate from whole grains, legumes, fruits, and vegetables does not increase the risk index of calcium oxalate precipitability in most normocalciuric adults who form stones.

**URL:** <https://ovidsp.ovid.com/ovidweb.cgi?T=JS&CSC=Y&NEWS=N&PAGE=fulltext&D=med4&DO=10.1016%2fS0002-8223%2898%2900071-6> <https://libkey.io/libraries/2592/openurl?genre=article&sid=OVID:medline&id=pmid:9508013&id=doi:10.1016%2FS0002-8223%2898%2900071-6&issn=0002-8223&isbn=&volume=98&issue=3&spage=303&pages=303-8&date=1998&title=Journal+of+the+American+Dietetic+Association&atitle=Substituting+milk+for+apple+juice+does+not+increase+kidney+stone+risk+in+most+normocalciuric+adults+who+form+calcium+oxalate+stones.&aulast=Massey&pid=%3Cauthor%3EMassey+LK%3BKynast-Gales+SA%3C%2Fauthor%3E%3CAN%3E9508013%3C%2FAN%3E%3CDT%3EClinical+Trial%3C%2FDT%3E>

253. **Nutritional potassium status of healthy adult males residing in the rural northeast Thailand.**

**Item Type:** Journal Article

**Authors:** Sriboonlue, P.;Prasongwatana, V.;Suwantrai, S.;Bovornpadungkitti, S.;Tungsanga, K. and Tosukhowong, P.

**Publication Date:** Ma ,1998

**Journal:** Journal of the Medical Association of Thailand 81(3), pp. 223–232

**Abstract:** Sixteen villages from rural areas of 8 provinces in the northeastern region of Thailand were randomly selected as study sites. Data on potassium (K) contents in 24-hour urine and serum samples of 93 healthy adult volunteer males aged 20-50 years old were completely collected and covered all 3 seasons of the year. The method of direct weighing of food was used to assess K intake in 13 subjects. K loss through sweat during working (9 hours) in the field was measured in 14 subjects by soaking their worn-clothes in distilled water after which K contents were measured by the flame photometry method. The results showed that the means urinary K excretion of 93 subjects were less than that of the cut-off value for normal (> or = 30 mmol/day) in all seasons of which 76.71%, 90.71% and 81.02% of the urine were categorized as hypokaliuria in the hot, rainy and cold seasons, respectively. In the case of serum K of these subjects, though the mean values were within a normal limit (> or = 3.5 mmol/l), 36.56%, 34.41% and 29.03% of the serum were classified as hypokalemia in the hot, rainy and cold seasons, respectively. In the assessment of K intake, it was clearly demonstrated that the values in all 3 seasons were much lower than that of the estimated safe and adequate daily dietary intake (ESADI) of K for the westerners (1975-5625 mg/day), i.e., the means of intake in the hot, rainy and cold seasons were only 807 +/- 172, 877 +/- 257 and 902 +/- 227 mg/day, respectively. Furthermore, K loss through sweat in the cold and the hot seasons were as high as a third (7.4 +/- 2.4 mmol/day) and a half (11.5 +/- 1.6 mmol/day), respectively, of the urinary excretion. Though the total body K contents were not evaluated in this study, our results suggest rural people in the northeast region of Thailand may be in a state of K deficiency. The severity is probably worsened in the hot season as seen from the tendency of decrease in serum K levels among 650 renal stone formers and 260 blood donors in this season.

**URL:** <https://ovidsp.ovid.com/ovidweb.cgi?T=JS&CSC=Y&NEWS=N&PAGE=fulltext&D=med4&AN=9623015> <https://libkey.io/libraries/2592/openurl?genre=article&sid=OVID:medline&id=pmid:9623015&id=doi:&issn=0125-2208&isbn=&volume=81&issue=3&spage=223&pages=223-32&date=1998&title=Journal+of+the+Medical+Association+of+Thailand&atitle=Nutritional+potassium+status+of+healthy+adult+males+residing+in+the+rural+northeast+Thailand.&aulast=Sriboonlue&pid=%3Cauthor%3ESriboonlue+P%3BPrasongwatana+V%3BSuwantrai+S%3BBovornpadungkitti+S%3BTungsanga+K%3BTosukhowong+P%3C%2Fauthor%3E%3CAN%3E9623015%3C%2FAN%3E%3CDT%3EJournal+Article%3C%2FDT%3E>

254. **A study of dietary calcium and other nutrients in idiopathic renal calcium stone formers with low bone mineral content.**

**Item Type:** Journal Article

**Authors:** Trinchieri, A.;Nespoli, R.;Ostini, F.;Rovera, F.;Zanetti, G. and Pisani, E.

**Publication Date:** Ma ,1998

**Journal:** Journal of Urology 159(3), pp. 654–657

**Abstract: PURPOSE**: Patients with calcium renal stone are reported to have lower bone mineral density. The state of bone density in patients with renal stones have different explanations but the role of nutritional factors seems to be crucial. A group of 48 consecutive male calcium renal stone formers was studied to investigate the relationship between bone density and dietary intake. **MATERIALS AND METHODS**: Patients completed a dietary diary for a 3-day period during normal diet. Nutrients and calories were calculated by food composition tables using a computerized procedure. Bone densitometry was assessed at the lumbar spine and femoral neck, and expressed as Z score. A blood sample was collected and was analyzed for serum biochemistry including alkaline phosphatase, parathyroid hormone and 1,25 vitamin D. A 24-hour urine sample was analyzed for calcium, phosphate, oxalate, citrate and other electrolytes. **RESULTS**: Dietary calcium intake was significantly lower (p : Dietary calcium intake was significantly lower (p **CONCLUSIONS**: A primary abnormality of bone metabolism could be a reasonable explanation of reduced bone density observed in renal stone formers on a low calcium diet since serum parathyroid hormone levels are in the normal range. From a therapeutic point of view these data confirm that restriction of dairy products in renal stone formers should be avoided.

**URL:** <https://ovidsp.ovid.com/ovidweb.cgi?T=JS&CSC=Y&NEWS=N&PAGE=fulltext&D=med4&AN=9474119> <https://libkey.io/libraries/2592/openurl?genre=article&sid=OVID:medline&id=pmid:9474119&id=doi:&issn=0022-5347&isbn=&volume=159&issue=3&spage=654&pages=654-7&date=1998&title=Journal+of+Urology&atitle=A+study+of+dietary+calcium+and+other+nutrients+in+idiopathic+renal+calcium+stone+formers+with+low+bone+mineral+content.&aulast=Trinchieri&pid=%3Cauthor%3ETrinchieri+A%3BNespoli+R%3BOstini+F%3BRovera+F%3BZanetti+G%3BPisani+E%3C%2Fauthor%3E%3CAN%3E9474119%3C%2FAN%3E%3CDT%3EJournal+Article%3C%2FDT%3E>

255. **Hyperoxaluria in patients with idiopathic calcium nephrolithiasis.**

**Item Type:** Journal Article

**Authors:** Trinchieri, A.;Ostini, F.;Nespoli, R.;Rovera, F.;Zanetti, G. and Pisani, E.

**Publication Date:** 1998

**Journal:** Journal of Nephrology 11(Suppl 1), pp. 70–72

**Abstract:** We studied 476 patients with idiopathic renal calcium stone disease (286 M, 190 F) while they ate their customary diets. Each subject collected a 24-hour urine sample and completed a dietary diary for a 3-day period. Daily urinary oxalate excretion (M 0.24 +/- 0.15 mg/dl, F 0.23 +/- 0.15 mg/dl) and nutrient intake values were calculated and multiple regression analyses were performed. Daily urinary oxalate excretion was significantly (p < 0.001) related to urinary volume (R = 0.24), vitamin C intake (R = 0.33) and body mass index (R = 0.37) and inversely related to calcium intake (R = -0.35). We conclude that urinary oxalate reflects endogenous oxalate production, presumably related to body size, but also intestinal absorption of oxalate, related to dietary intake and to the effect of dietary calcium intake which reduces intestinal oxalate absorption.

**URL:** <https://ovidsp.ovid.com/ovidweb.cgi?T=JS&CSC=Y&NEWS=N&PAGE=fulltext&D=med4&AN=9604817> <https://libkey.io/libraries/2592/openurl?genre=article&sid=OVID:medline&id=pmid:9604817&id=doi:&issn=1121-8428&isbn=&volume=11&issue=1&spage=70&pages=70-2&date=1998&title=Journal+of+Nephrology&atitle=Hyperoxaluria+in+patients+with+idiopathic+calcium+nephrolithiasis.&aulast=Trinchieri&pid=%3Cauthor%3ETrinchieri+A%3BOstini+F%3BNespoli+R%3BRovera+F%3BZanetti+G%3BPisani+E%3C%2Fauthor%3E%3CAN%3E9604817%3C%2FAN%3E%3CDT%3EJournal+Article%3C%2FDT%3E>

256. **Correspondence between stone composition and urine supersaturation in nephrolithiasis.**

**Item Type:** Journal Article

**Authors:** Parks, J. H.;Coward, M. and Coe, F. L.

**Publication Date:** Ma ,1997

**Journal:** Kidney International 51(3), pp. 894–900

**Abstract:** Supersaturation (SS) with respect to calcium oxalate monohydrate (COM), brushite (Br) and uric acid (UA), obtained in three 24-hour pretreatment urine samples from patients with stone disease were compared to the mineral composition of stones passed by the same patients to determine whether sparse urine SS measurements accurately reflect the long-term average SS values in the kidney and final urine. Among males and females elevation of SS above same sex normals corresponded to composition. As well, treatments that reduced stone rates also reduced these SS values. The degree of calcium phosphate (CaP) admixture was accurately matched by shifting magnitudes of COM and Br SS. As well, increasing CaP content was associated with falling urine citrate and rising urine pH, suggesting renal tubular acidosis. We conclude that sparse urine SS measurements accurately track stone admixtures, and are a reliable index of average renal and urine SS.

**URL:** <https://ovidsp.ovid.com/ovidweb.cgi?T=JS&CSC=Y&NEWS=N&PAGE=fulltext&D=med4&DO=10.1038%2fki.1997.126> <https://libkey.io/libraries/2592/openurl?genre=article&sid=OVID:medline&id=pmid:9067927&id=doi:10.1038%2Fki.1997.126&issn=0085-2538&isbn=&volume=51&issue=3&spage=894&pages=894-900&date=1997&title=Kidney+International&atitle=Correspondence+between+stone+composition+and+urine+supersaturation+in+nephrolithiasis.&aulast=Parks&pid=%3Cauthor%3EParks+JH%3BCoward+M%3BCoe+FL%3C%2Fauthor%3E%3CAN%3E9067927%3C%2FAN%3E%3CDT%3EJournal+Article%3C%2FDT%3E>

257. **Effect of mineral water containing calcium and magnesium on calcium oxalate urolithiasis risk factors.**

**Item Type:** Journal Article

**Authors:** Rodgers, A. L.

**Publication Date:** 1997

**Journal:** Urologia Internationalis 58(2), pp. 93–99

**Abstract:** Calcium oxalate kidney stone formers are invariably advised to increase their fluid intake. In addition, magnesium therapy is often administered. Recently, a prospective study showed that a high dietary intake of calcium reduces the risk of symptomatic kidney stones. The present study was performed to test whether simultaneous delivery of these factors--high fluid intake, magnesium ingestion and increased dietary calcium--could reduce the risk of calcium oxalate kidney stone formation. A French mineral water, containing calcium and magnesium (202 and 36 ppm, respectively) was selected as the dietary vehicle. Twenty calcium oxalate stone-forming patients of each sex as well as 20 healthy volunteers of each sex participated in the study. Each subject provided a 24-hour urine collection after ingestion of mineral water over a period of 3 days; after a suitable rest period the protocol was repeated using local tap water (Ca: 13 ppm, Mg: 1 ppm). In addition, 24-hour urines were collected by each subject on their free diets. The entire cycle was repeated at least twice by each subject. Several risk factors (excretion of oxalate; relative supersaturations of calcium oxalate, brushite and uric acid; calcium oxalate metastable limit; oxalate:magnesium ratio and oxalate:metastable limit ratio) were favourably altered by the mineral water and tap water regimens but the former was more effective. In addition, the mineral water protocol produced favourable but unique changes in the excretion of citrate and magnesium as well as in the relative supersaturation of brushite which were not achieved by the tap water regimen. To the contrary, tap water produced an unfavourable change in the magnesium excretion. The group which benefitted most were male stone formers in whom 9 risk factors were favourably altered by the mineral water protocol. It is concluded that mineral water containing calcium and magnesium, such as that used in this study, deserves to be considered as a possible therapeutic or prophylactic agent in calcium oxalate kidney stone disease.

**URL:** <https://ovidsp.ovid.com/ovidweb.cgi?T=JS&CSC=Y&NEWS=N&PAGE=fulltext&D=med4&DO=10.1159%2f000282958> <https://libkey.io/libraries/2592/openurl?genre=article&sid=OVID:medline&id=pmid:9096270&id=doi:10.1159%2F000282958&issn=0042-1138&isbn=&volume=58&issue=2&spage=93&pages=93-9&date=1997&title=Urologia+Internationalis&atitle=Effect+of+mineral+water+containing+calcium+and+magnesium+on+calcium+oxalate+urolithiasis+risk+factors.&aulast=Rodgers&pid=%3Cauthor%3ERodgers+AL%3C%2Fauthor%3E%3CAN%3E9096270%3C%2FAN%3E%3CDT%3EClinical+Trial%3C%2FDT%3E>

258. **Sensitivity to calcium intake in calcium stone forming patients.**

**Item Type:** Journal Article

**Authors:** Heilberg, I. P.;Martini, L. A.;Draibe, S. A.;Ajzen, H.;Ramos, O. L. and Schor, N.

**Publication Date:** 1996

**Journal:** Nephron 73(2), pp. 145–153

**Abstract:** The absorptive or renal origin of hypercalciuria can be discriminated using an acute oral calcium load test (ACLT). Of 86 patients with calcium oxalate kidney stones, 28 (23%) were found to be hypercalciuric (HCa) and 58 (67%) normocalciuric (NCa) on their customary free diet, containing 542 +/- 29 mg/day (mean +/- SE) of calcium. Since the apparently normal 24-hour calcium excretion of many calcium stone formers (CSF) may be due to a combination of high calcium absorption with moderately low calcium intake, all patients were investigated by ACLT. Of 28 HCa patients, 13 (46%) were classified as absorptive (AH) and 15 (54%) as renal hypercalciuria (RH). Of the 58 NCa patients, 38 (65%) presented features of intestinal hyperabsorption and were therefore designated as AH-like, and 20 (35%) as RH-like. To further elucidate the role of dietary calcium in these CSF, a chronic calcium load test (CCLT), consisting of 1 g/day of oral Ca for 7 days, was designed. A positive response to the CCLT was considered to occur when urinary calcium (uCa) was > or = 4 mg/ kg/24 h on the 7th day. Among NCa patients, 29% of AH-like subjects responded to the CCLT and 71% did not; 50% of RH-like subjects also responded and 50% did not. In HCa patients, 85% of AH and 67% of RH subjects maintained uCa > or = 4 mg/kg/24 h after the CCLT and 15% of AH and 23% of RH subjects did not. However, a significant additional increase in mean uCa was not observed among HCa patients. All patients were submitted to a second evaluation of fasting calciuria (Ca/Cr). A modification of this parameter was noticed in 89% of RH-like and 78% of RH patients. In conclusion, these data suggest the presence of subpopulations of patients sensitive or not to calcium intake, regardless of whether the acute response to a calcium overload test suggested AH or RH. The CCLT disclosed dietary hypercalciuria in 21/58 (36%) of previously NCa patients. In these NCa patients, the ACLT may be replaced by the CCLT. The distinction between AH and RH initially evidenced by the ACLT was not further confirmed. These data suggest that either fasting Ca/Cr is not adequate for subclassification of HCa or that AH and RH represent a different spectrum of the same disease, and that a primary resorptive component should also be considered.

**URL:** <https://ovidsp.ovid.com/ovidweb.cgi?T=JS&CSC=Y&NEWS=N&PAGE=fulltext&D=med4&DO=10.1159%2f000189031> <https://libkey.io/libraries/2592/openurl?genre=article&sid=OVID:medline&id=pmid:8773335&id=doi:10.1159%2F000189031&issn=1660-8151&isbn=&volume=73&issue=2&spage=145&pages=145-53&date=1996&title=Nephron&atitle=Sensitivity+to+calcium+intake+in+calcium+stone+forming+patients.&aulast=Heilberg&pid=%3Cauthor%3EHeilberg+IP%3BMartini+LA%3BDraibe+SA%3BAjzen+H%3BRamos+OL%3BSchor+N%3C%2Fauthor%3E%3CAN%3E8773335%3C%2FAN%3E%3CDT%3EClinical+Trial%3C%2FDT%3E>

259. **Struvite stones: long term follow up under metaphylaxis.**

**Item Type:** Journal Article

**Authors:** Jarrar, K.;Boedeker, R. H. and Weidner, W.

**Publication Date:** 1996

**Journal:** Annales D Urologie 30(3), pp. 112–117

**Abstract:** This study presents the results of a 10-year metaphylaxis of 19 former struvite stone formers, each having had 2-3 stone operations. In these patients, urine was acidified with L-methionine (Acimethin) using a dose of three to six tablets 500 mg/day. Every three months, 11 laboratory parameters were checked in 24-hour urine. 6 parameters were determined in serum. In addition, urine samples were tested for infection. Statistical analysis of analytical data, which was supported by computer graphs, provided the results for the urinary parameters. They were described by geometric means and 95% confidence intervals using ANOVA (analysis of variance). During therapy, the mean pH values decreased significantly from 7.5 to 5.5. Signific-ant increases were found in the excretion of citrate, magnesium, potassium, and uric acid. An increase was also found for calcium, which, however, could not be confirmed to be statistically significant (p = 0.08). In serum, changes of parameters could only be registered for calcium and phosphate. However, at all times, total serum concentrations stayed within their normal limits. Three patients suffered occasional infections (16%), only two of them formed recurrent stones (10%). In assessing the efficacy of L-methionine therapy, the drop in urinary pH to acidic values was the most relevant factor for metaphylaxis.

**URL:** <https://ovidsp.ovid.com/ovidweb.cgi?T=JS&CSC=Y&NEWS=N&PAGE=fulltext&D=med4&AN=8766146> <https://libkey.io/libraries/2592/openurl?genre=article&sid=OVID:medline&id=pmid:8766146&id=doi:&issn=0003-4401&isbn=&volume=30&issue=3&spage=112&pages=112-7&date=1996&title=Annales+d+Urologie&atitle=Struvite+stones%3A+long+term+follow+up+under+metaphylaxis.&aulast=Jarrar&pid=%3Cauthor%3EJarrar+K%3BBoedeker+RH%3BWeidner+W%3C%2Fauthor%3E%3CAN%3E8766146%3C%2FAN%3E%3CDT%3EJournal+Article%3C%2FDT%3E>

260. **Urinary oxalate excretion increases with body size and decreases with increasing dietary calcium intake among healthy adults.**

**Item Type:** Journal Article

**Authors:** Lemann, J. Jr;Pleuss, J. A.;Worcester, E. M.;Hornick, L.;Schrab, D. and Hoffmann, R. G.

**Publication Date:** Ja ,1996

**Journal:** Kidney International 49(1), pp. 200–208

**Abstract:** Increasing dietary calcium intake decreases urinary oxalate excretion by increasing intestinal precipitation of dietary oxalate as calcium oxalate. This mechanism was speculated to account for the decreased prospective incidence of kidney stones as estimated dietary calcium intake, adjusted for caloric intake, increased among men in a recent large epidemiological study. To further assess the relationship between estimated diet calcium and urinary oxalate, we studied 94 health adults, 50 women and 44 men, ages 20 to 70 years with weights ranging form 47 to 104 kg while they ate their customary diets. Each subject completed a semiquantitative food frequency questionnaire and collected three 24-hour urines preserved with HCl. The urines were collected accurately as judged by a mean intrasubject CV for creatinine excretion of 9.8% and direct relations between urinary creatinine excretion and body wt (r = 0.62; P < 0.0001), or predicted urine creatinine content for sex, age and weight using the Cockcroft and Gault formulas (r = 0.76; P < 0.0001). Estimated diet calcium intake ranged from 6.8 to 68 mmol/day (272 to 2720 mg/day) and averaged 29.5 mmol/day (1180 mg/day). Individual mean urinary oxalate excretion ranged from 0.079 go 0.332 mmol/day (7 to 29 mg/day) and averaged 0.198 mmol/day (17 mg/day). Among all subjects, daily oxalate excretion was directly related to creatinine excretion as an estimate of lean body mass (r = 0.61; P < 0.0001). Thus, oxalate excretion among men averaged 0.228 +/- 0.051 SD mmol/day, a value significantly higher than the average among women of 0.173 +/- 0.045 mmol/day (P < 0.001). Daily urine oxalate excretion/creatinine decreased curvilinearly as estimated dietary Ca intake increased (r = -0.30; P = 0.0035) and as the ratio of estimated dietary calcium to dietary oxalate increased (r = -0.39; P = 0.0001). We conclude that body size is the major determinant of urinary oxalate excretion among healthy adults, presumably reflecting variations in endogenous oxalate synthesis with lean body mass. Increasing estimated diet calcium intake, especially up to the range of 15 to 20 mmol/day (600 to 800 mg/day) has an additional effect to decrease during oxalate excretion, presumably by limiting intestinal absorption of dietary oxalate.

**URL:** <https://ovidsp.ovid.com/ovidweb.cgi?T=JS&CSC=Y&NEWS=N&PAGE=fulltext&D=med4&DO=10.1038%2fki.1996.27> <https://libkey.io/libraries/2592/openurl?genre=article&sid=OVID:medline&id=pmid:8770968&id=doi:10.1038%2Fki.1996.27&issn=0085-2538&isbn=&volume=49&issue=1&spage=200&pages=200-8&date=1996&title=Kidney+International&atitle=Urinary+oxalate+excretion+increases+with+body+size+and+decreases+with+increasing+dietary+calcium+intake+among+healthy+adults.&aulast=Lemann&pid=%3Cauthor%3ELemann+J+Jr%3BPleuss+JA%3BWorcester+EM%3BHornick+L%3BSchrab+D%3BHoffmann+RG%3C%2Fauthor%3E%3CAN%3E8770968%3C%2FAN%3E%3CDT%3EJournal+Article%3C%2FDT%3E>

261. **Evaluation of the risk of stone formation: study on crystalluria in patients with recurrent calcium oxalate urolithiasis.**

**Item Type:** Journal Article

**Authors:** Robert, M.;Boularan, A. M.;Delbos, O.;Monnier, L. and Grasset, D.

**Publication Date:** 1996

**Journal:** European Urology 29(4), pp. 456–461

**Abstract: OBJECTIVE**: The aim of this study was to determine the usefulness of the morning calcium oxalate crystalluria in detecting stone formers particularly prone to recurrence. **METHODS**: Over a 24-hour period of urine collection, the morning calcium oxalate crystalluria was evaluated as well as the risk of stone formation, established with Tiselius and Parks indices, for 25 recurrent stone formers (group 1) and 25 normal controls (group 2). **RESULTS**: Morning crystalluria (type, size, number/ml and state of aggregate) and the Tiselius index were comparable in the two groups. Conversely, calciuria as well as the citrate/ calcium ratio and the Parks index varied significantly for stone formers and normal controls. No particular correlation appeared between crystalluria and indices of Tiselius and Parks, calciuria, calcium-oxalate product or calcium/ oxalate and citrate/calcium ratios. **CONCLUSIONS**: Morning calcium oxalate crystalluria does not enable an efficient characterization of recurrent stone formers. Its discordance with others potential indicators of the risk of stone formation poses the problem of their respective validity and evokes the prevalence of still unknown inhibiting agents in the phenomenon of crystallization.

**URL:** <https://ovidsp.ovid.com/ovidweb.cgi?T=JS&CSC=Y&NEWS=N&PAGE=fulltext&D=med4&DO=10.1159%2f000473796> <https://libkey.io/libraries/2592/openurl?genre=article&sid=OVID:medline&id=pmid:8791054&id=doi:10.1159%2F000473796&issn=0302-2838&isbn=&volume=29&issue=4&spage=456&pages=456-61&date=1996&title=European+Urology&atitle=Evaluation+of+the+risk+of+stone+formation%3A+study+on+crystalluria+in+patients+with+recurrent+calcium+oxalate+urolithiasis.&aulast=Robert&pid=%3Cauthor%3ERobert+M%3BBoularan+AM%3BDelbos+O%3BMonnier+L%3BGrasset+D%3C%2Fauthor%3E%3CAN%3E8791054%3C%2FAN%3E%3CDT%3EJournal+Article%3C%2FDT%3E>

262. **Effect of extracorporeal shock wave lithotripsy on glomerular and tubular functions.**

**Item Type:** Journal Article

**Authors:** Sen, S.;Erdem, Y.;Oymak, O.;Yalcin, A. U.;Turgan, C.;Ersoy, H.;Bingol, N. and Tamer, S.

**Publication Date:** 1996

**Journal:** International Urology & Nephrology 28(3), pp. 309–313

**Abstract:** To evaluate the early and late effects of extracorporeal shock wave lithotripsy on renal function, we prospectively designed a controlled study using a Direx lithotriptor. Twenty-five patients with renal stones and 16 healthy volunteers as the control group were included in the study. Blood and urine samples were collected before and after 24 hours, seven days and 8 months in the patient group. White blood cell count, serum levels of haemoglobin, urea, creatinine, SGOT, SGPT, AP, and LDH were determined. 24-hour urine specimens were collected to be tested for volume, excretion of creatinine, albumin, N-acetyl-beta-glucosaminidase, gamma-glutamyltransferase and beta-2-microglobulin. There were statistically significant increments in the secretion of urinary enzymes and albumin in the early period after ESWL, no longer lasting 8 months after the procedure. At 8 months one patient was hypertensive as judged by the diastolic pressure above 95 mm Hg. The results of this study showed that, although there was a transient glomerular and tubular damage after extracorporeal shock wave lithotripsy, the procedure seems safe and causes no permanent deterioration in renal function.

**URL:** <https://ovidsp.ovid.com/ovidweb.cgi?T=JS&CSC=Y&NEWS=N&PAGE=fulltext&D=med4&DO=10.1007%2fBF02550491> <https://libkey.io/libraries/2592/openurl?genre=article&sid=OVID:medline&id=pmid:8899469&id=doi:10.1007%2FBF02550491&issn=0301-1623&isbn=&volume=28&issue=3&spage=309&pages=309-13&date=1996&title=International+Urology+%26+Nephrology&atitle=Effect+of+extracorporeal+shock+wave+lithotripsy+on+glomerular+and+tubular+functions.&aulast=Sen&pid=%3Cauthor%3ESen+S%3BErdem+Y%3BOymak+O%3BYalcin+AU%3BTurgan+C%3BErsoy+H%3BBingol+N%3BTamer+S%3C%2Fauthor%3E%3CAN%3E8899469%3C%2FAN%3E%3CDT%3EClinical+Trial%3C%2FDT%3E>

263. **Characteristics of patients surgically treated for primary hyperparathyroidism with and without renal stones.**

**Item Type:** Journal Article

**Authors:** Soreide, J. A.;van Heerden, J. A.;Grant, C. S.;Lo, C. Y. and Ilstrup, D. M.

**Publication Date:** 1996

**Journal:** Surgery 120(6), pp. 1033–1037

**Abstract: BACKGROUND**: Between 1980 and 1984, 312 (30%) of 1038 patients undergoing initial cervical exploration for primary hyperparathyroidism (pHPT) at our institution had proven renal stones. **METHODS**: In this retrospective study we focused on clinical characteristics, biochemical tests, perioperative and pathologic findings, and immediate outcomes of operation, comparing findings in patients with and without renal stones. **RESULTS**: Patients with renal stones were more often younger male patients, had serum phosphorus levels significantly lower (p : Patients with renal stones were more often younger male patients, had serum phosphorus levels significantly lower (p : Patients with renal stones were more often younger male patients, had serum phosphorus levels significantly lower (p : Patients with renal stones were more often younger male patients, had serum phosphorus levels significantly lower (p : Patients with renal stones were more often younger male patients, had serum phosphorus levels significantly lower (p : Patients with renal stones were more often younger male patients, had serum phosphorus levels significantly lower (p **CONCLUSIONS**: In patients with renal stones the diagnosis of pHPT should be considered. If the diagnosis is confirmed, a liberal approach to cervical exploration should be taken.

**URL:** <https://ovidsp.ovid.com/ovidweb.cgi?T=JS&CSC=Y&NEWS=N&PAGE=fulltext&D=med4&DO=10.1016%2fs0039-6060%2896%2980051-1> <https://libkey.io/libraries/2592/openurl?genre=article&sid=OVID:medline&id=pmid:8957491&id=doi:10.1016%2Fs0039-6060%2896%2980051-1&issn=0039-6060&isbn=&volume=120&issue=6&spage=1033&pages=1033-7%3B+discussion+1037-8&date=1996&title=Surgery&atitle=Characteristics+of+patients+surgically+treated+for+primary+hyperparathyroidism+with+and+without+renal+stones.&aulast=Soreide&pid=%3Cauthor%3ESoreide+JA%3Bvan+Heerden+JA%3BGrant+CS%3BLo+CY%3BIlstrup+DM%3C%2Fauthor%3E%3CAN%3E8957491%3C%2FAN%3E%3CDT%3EJournal+Article%3C%2FDT%3E>

264. **Some aspects of citrate metabolism in renal-stone patients from northeastern Thailand.**

**Item Type:** Journal Article

**Authors:** Sriboonlue, P.;Prasongwatana, V.;Borvornpadoongkitti, S. and Suwantrai, S.

**Publication Date:** No ,1996

**Journal:** Journal of the Medical Association of Thailand 79(11), pp. 737–743

**Abstract:** In our previous nephrolithiasis studies in the northeast region of Thailand, hypokaliurea and hypocitraturia were the 2 most commonly encountered metabolic abnormalities. This led us to believe that people prone to forming renal-stones in this area were in a state of potassium depletion, a condition which probably caused the low urinary excretion of citrate. Further studies on some aspects of citrate metabolism in these subjects were carried out. Two groups of adult male subjects were included in the study protocol. Group 1 consisted of 20 urban dwellers who were used as normal controls, and group 2 was comprised of 36 renal-stone patients residing in rural villages outside the municipal area. Fasting clotted venous blood and one 24-hour urine specimens were collected and analyzed for creatinine, citrate, calcium, phosphate, magnesium, sodium, potassium, chloride, bicarbonate and uric acid. Values for: creatinine and citrate clearances, the filtered load of citrate and the tubular reabsorption of citrate were then calculated. The results showed that, for both groups, the concentrations of most of the above parameters were within the normal ranges both serum and urine. An exception to this was that the levels of serum potassium and of urinary excretions of sodium, potassium and citrate in people in group 2 were significantly less than those in group 1 (p < 0.005, p < 0.001, p < 0.001 and p < 0.0001, respectively). With respect to citrate metabolism, while the serum citrate levels and the filtered load of citrate were not different between the 2 groups, the average percentage of renal tubular reabsorption of group 2 (95 +/- 1.1%) increased significantly (P < 0.0001)in comparison to group 1 (85 +/- 1.6%). Moreover when results from both groups were combined, a significant negative correlation between the renal tubular reabsorption of citrate and the urinary excretion of potassium was clearly seen (r = 0.4001, p < 0.007). Our data suggests that potassium depletion may affect the renal tubular cells in some manner which, consequently, causes an increase in renal tubular reabsorption of citrate. The final outcome of these changes in these renal stone subjects was hypocitraturia.

**URL:** <https://ovidsp.ovid.com/ovidweb.cgi?T=JS&CSC=Y&NEWS=N&PAGE=fulltext&D=med4&AN=8997014> <https://libkey.io/libraries/2592/openurl?genre=article&sid=OVID:medline&id=pmid:8997014&id=doi:&issn=0125-2208&isbn=&volume=79&issue=11&spage=737&pages=737-43&date=1996&title=Journal+of+the+Medical+Association+of+Thailand&atitle=Some+aspects+of+citrate+metabolism+in+renal-stone+patients+from+northeastern+Thailand.&aulast=Sriboonlue&pid=%3Cauthor%3ESriboonlue+P%3BPrasongwatana+V%3BBorvornpadoongkitti+S%3BSuwantrai+S%3C%2Fauthor%3E%3CAN%3E8997014%3C%2FAN%3E%3CDT%3EComparative+Study%3C%2FDT%3E>

265. **Inappropriate phosphate excretion in idiopathic hypercalciuria: the key to a common cause and future treatment?.**

**Item Type:** Journal Article

**Authors:** Williams, C. P.;Child, D. F.;Hudson, P. R.;Soysa, L. D.;Davies, G. K.;Davies, M. G. and De Bolla, A. R.

**Publication Date:** No ,1996

**Journal:** Journal of Clinical Pathology 49(11), pp. 881–888

**Abstract: AIMS**: To present experimental evidence in support of a proposed common cause for absorptive hypercalciuria, renal hypercalciuria, renal phosphate leak and enhancement of 1,25-(OH)2-vitamin D concentrations in patients presenting with renal stone disease; and to suggest further investigation with a view to new management. **METHODS**: An oral calcium loading test was administered to 15 patients with renal stones and 10 normal controls in the fasting state: urine and blood were collected hourly. After the second urine sample, 400 mg calcium dissolved in water was administered orally. Serum calcium, albumin, parathyroid hormone (PTH), and phosphate were measured together with urine calcium clearance and urinary phosphate from which the TmPO4/glomerular filtration rate (GFR) ratio was calculated. Serum 1,25-(OH)2-vitamin D was measured in the first serum sample. In addition, 24 hour urine calcium results were collected retrospectively from the patients' case notes over the previous 18 months. **RESULTS**: In the basal state, renal stone patients had an overall greater phosphaturia (lower TmPO4/GFR: median 1.72 compared with 2.10 in controls) and increased calcium clearance. Serum corrected calcium and PTH concentrations did not differ between the groups. After calcium loading, serum calcium and urine calcium clearance rose in both groups, with patients with renal stones experiencing a greater percentage fall in phosphaturia. In both groups TmPO4/GFR fell (greater phosphaturia) with increased serum corrected calcium, with the patients showing notably greater phosphaturia for any given calcium concentration. Patients also had notably greater phosphaturia compared with the serum calcium concentration for any given PTH value. Serum 1,25-(OH)2-vitamin D was higher in patients than controls and for any 1,25-(OH)2-vitamin D concentration phosphaturia measured against serum calcium was greater in patients than controls. 1,25-(OH)2-vitamin D did not correlate with phosphaturia relative to serum calcium concentrations within the patient and control groups. **CONCLUSIONS**: It is proposed that patients with idiopathic hypercalciuria have an "inappropriately' high phosphate excretion for any given serum calcium concentration. Loss of phosphate may induce increased activation of 1,25-(OH)2-vitamin D. Some of the commonly described causes of stone formation may be manifestations of a single mechanism.

**URL:** <https://ovidsp.ovid.com/ovidweb.cgi?T=JS&CSC=Y&NEWS=N&PAGE=fulltext&D=med4&DO=10.1136%2fjcp.49.11.881> <https://libkey.io/libraries/2592/openurl?genre=article&sid=OVID:medline&id=pmid:8944605&id=doi:10.1136%2Fjcp.49.11.881&issn=0021-9746&isbn=&volume=49&issue=11&spage=881&pages=881-8&date=1996&title=Journal+of+Clinical+Pathology&atitle=Inappropriate+phosphate+excretion+in+idiopathic+hypercalciuria%3A+the+key+to+a+common+cause+and+future+treatment%3F.&aulast=Williams&pid=%3Cauthor%3EWilliams+CP%3BChild+DF%3BHudson+PR%3BSoysa+LD%3BDavies+GK%3BDavies+MG%3BDe+Bolla+AR%3C%2Fauthor%3E%3CAN%3E8944605%3C%2FAN%3E%3CDT%3EJournal+Article%3C%2FDT%3E>

266. **Urinary Tamm-Horsfall protein increased after potassium citrate therapy in calcium stone formers.**

**Item Type:** Journal Article

**Authors:** Fuselier, H. A.;Ward, D. M.;Lindberg, J. S.;Allen, J. M.;Husserl, F. E.;Marcucci, P. A.;Cole, F. E.;Turnipseed, J.;Alam, J. and Kok, D. J.

**Publication Date:** Ju ,1995

**Journal:** Urology 45(6), pp. 942–946

**Abstract: OBJECTIVES**: To evaluate the effect of oral potassium citrate therapy on urinary excretion rates of citrate. Tamm-Horsfall protein (THP), and on calcium oxalate monohydrate crystal agglomeration inhibition [tm], in patients with recurrent calcium stone formation. **METHODS**: To evaluate the effect of oral therapy with potassium citrate on urinary citrate, THP, and [tm], 24-hour urine samples were collected before and at least 2 months after initiation of oral potassium citrate therapy in 33 calcium stone-forming patients who had no dietary restrictions. The citrate concentration was measured by an adaptation of a citrate lyase method. Urinary disaggregated THP concentration was determined with a quantitative enzyme-linked immunosorbent assay. The [tm] was determined by observing the effects of patients' urine, before and after oral potassium citrate therapy, on the uptake of 45Ca2+ onto the surfaces of added preformed calcium oxalate crystals in a supersaturated solution of calcium oxalate, using the in vitro kinetic method described by other investigators. **RESULTS**: We observed an increased urinary excretion rate of citrate from a mean of 1.9 mmol/24 h prealkali to 2.6 mmol/24 h postalkali (P : We observed an increased urinary excretion rate of citrate from a mean of 1.9 mmol/24 h prealkali to 2.6 mmol/24 h postalkali (P : We observed an increased urinary excretion rate of citrate from a mean of 1.9 mmol/24 h prealkali to 2.6 mmol/24 h postalkali (P : We observed an increased urinary excretion rate of citrate from a mean of 1.9 mmol/24 h prealkali to 2.6 mmol/24 h postalkali (P **CONCLUSIONS**: To our knowledge this is the first report correlating increased urinary citrate with THP excretion rate following oral alkalinization with potassium citrate in calcium stone formers. Of clinical importance is the corresponding increase in [tm], which was previously shown to be inversely related to stone-forming activity. Moreover, urinary citrate and THP are known to have a synergistic effect on [tm]. Our data suggest that the effectiveness of potassium citrate therapy in calcium stone-forming patients may, at least in part, be due to increased levels of THP.

**URL:** <https://ovidsp.ovid.com/ovidweb.cgi?T=JS&CSC=Y&NEWS=N&PAGE=fulltext&D=med3&DO=10.1016%2fs0090-4295%2899%2980112-5> <https://libkey.io/libraries/2592/openurl?genre=article&sid=OVID:medline&id=pmid:7771027&id=doi:10.1016%2Fs0090-4295%2899%2980112-5&issn=0090-4295&isbn=&volume=45&issue=6&spage=942&pages=942-6&date=1995&title=Urology&atitle=Urinary+Tamm-Horsfall+protein+increased+after+potassium+citrate+therapy+in+calcium+stone+formers.&aulast=Fuselier&pid=%3Cauthor%3EFuselier+HA%3BWard+DM%3BLindberg+JS%3BAllen+JM%3BHusserl+FE%3BMarcucci+PA%3BCole+FE%3BTurnipseed+J%3BAlam+J%3BKok+DJ%3Bet+al%3C%2Fauthor%3E%3CAN%3E7771027%3C%2FAN%3E%3CDT%3EJournal+Article%3C%2FDT%3E>

267. **Diagnosis and treatment of calcium kidney stones**

**Item Type:** Journal Article

**Authors:** Klugman, V. and Favus, M. J.

**Publication Date:** 1995

**Journal:** Advances in Endocrinology & Metabolism 6, pp. 117–142

**Abstract:** Calcium oxalate nephrolithiasis is a common syndrome that recurs and may be complicated by infection, obstruction, bleeding, and rarely, impairment in renal function. The formation of Ca oxalate stones depends on the state of urinary supersaturation with respect to Ca and oxalate and the action of urinary inhibitors of crystal nucleation, aggregation, and growth. Idiopathic hypercalciuria is the most common cause of Ca oxalate stones and is characterized by hypercalciuria, normocalcemia, and intestinal Ca hyperabsorption with or without elevated serum 1,25(OH)2D3 levels in the absence of other known causes of hypercalciuria. Current diagnostic evaluation of recurrent Ca oxalate nephrolithiasis should be conducted while the patients follow their usual diets and includes the following: 1. Analysis of stone composition by polarization microscopy. 2. Measurement of serum Ca, phosphate, uric acid, 1,25(OH)2D3, and creatinine. 3. Twenty-four-hour urine collection for an analysis of volume, pH, and excretion of Ca, phosphorus, magnesium, uric acid, citrate, sodium, oxalate, and creatinine. Therapy to prevent stone recurrence is designed to reduce urinary supersaturation of Ca oxalate by increasing urine volume, reducing urine Ca to below 200 mg/24 hr with thiazide, maintaining dietary Ca intake at 600 to 800 mg/day, and adding potassium citrate if urine citrate levels are reduced. If elevated, urine oxalate excretion can be reduced by dietary oxalate restriction. Stones less than 2 cm in diameter located in the renal parenchyma or upper urinary tract can be fragmented with ESWL, whereas larger stones or those in the lower urinary tract should be removed by either percutaneous nephrolithotomy or ureteroscopic procedures. [References: 73]

**URL:** <https://ovidsp.ovid.com/ovidweb.cgi?T=JS&CSC=Y&NEWS=N&PAGE=fulltext&D=med3&AN=7671093> <https://libkey.io/libraries/2592/openurl?genre=article&sid=OVID:medline&id=pmid:7671093&id=doi:&issn=1049-6734&isbn=&volume=6&issue=&spage=117&pages=117-42&date=1995&title=Advances+in+Endocrinology+%26+Metabolism&atitle=Diagnosis+and+treatment+of+calcium+kidney+stones.&aulast=Klugman&pid=%3Cauthor%3EKlugman+V%3BFavus+MJ%3C%2Fauthor%3E%3CAN%3E7671093%3C%2FAN%3E%3CDT%3EJournal+Article%3C%2FDT%3E>

268. **Clinical and biochemical differences in patients with pure calcium oxalate monohydrate and calcium oxalate dihydrate kidney stones.**

**Item Type:** Journal Article

**Authors:** Pierratos, A. E.;Khalaff, H.;Cheng, P. T.;Psihramis, K. and Jewett, M. A.

**Publication Date:** Ma ,1994

**Journal:** Journal of Urology 151(3), pp. 571–574

**Abstract:** To examine the factors and patient characteristics predisposing to formation of calcium oxalate monohydrate or calcium oxalate dihydrate kidney stones, we compared blood and 24-hour urine tests, gender distribution and patient age in 2 groups of patients with pure calcium oxalate monohydrate (422) and calcium oxalate dihydrate (68) stones treated at the lithotripsy unit of the Wellesley Hospital and University of Toronto during 4 years. The calcium oxalate monohydrate group included relatively more women (31% versus 16% in the calcium oxalate dihydrate group, chi-square 7.89, p = 0.005). Patients were older in the calcium oxalate monohydrate group (59 +/- 13 versus 51 +/- 13 years, p = 0.001). The calcium oxalate monohydrate group had lower urinary calcium (4.19 +/- 2.34 versus 7.19 +/- 3.38 mmol. per day, p < 0.0001), calcium oxalate relative saturation rate (6.9 +/- 3.9 versus 8.9 +/- 3.3, p = 0.001), brushite relative saturation rate (0.7 +/- 0.8 versus 1.2 +/- 0.9, p = 0.0001) and urinary pH (5.72 +/- 0.75 versus 5.93 +/- 0.72). When corrected for patient age and gender, the calcium oxalate dihydrate group still had higher urine calcium levels. Higher urine pH in the calcium oxalate dihydrate group was age-related. In summary, we present evidence that calcium oxalate dihydrate stones are relatively more common among younger male patients with higher urine calcium levels and higher urine pH.

**URL:** <https://ovidsp.ovid.com/ovidweb.cgi?T=JS&CSC=Y&NEWS=N&PAGE=fulltext&D=med3&DO=10.1016%2fs0022-5347%2817%2935017-6> <https://libkey.io/libraries/2592/openurl?genre=article&sid=OVID:medline&id=pmid:8308959&id=doi:10.1016%2Fs0022-5347%2817%2935017-6&issn=0022-5347&isbn=&volume=151&issue=3&spage=571&pages=571-4&date=1994&title=Journal+of+Urology&atitle=Clinical+and+biochemical+differences+in+patients+with+pure+calcium+oxalate+monohydrate+and+calcium+oxalate+dihydrate+kidney+stones.&aulast=Pierratos&pid=%3Cauthor%3EPierratos+AE%3BKhalaff+H%3BCheng+PT%3BPsihramis+K%3BJewett+MA%3C%2Fauthor%3E%3CAN%3E8308959%3C%2FAN%3E%3CDT%3EComparative+Study%3C%2FDT%3E>

269. **Evaluation of a simple test to estimate urine saturation with calcium oxalate in stone-forming patients.**

**Item Type:** Journal Article

**Authors:** Sanchez, A.;Gonzalez, A.;Sarano, H. D. and Reinaldi, S.

**Publication Date:** De ,1994

**Journal:** International Journal of Urology 1(4), pp. 316–318

**Abstract:** In 43 patients with recent-onset calcium oxalate nephrolithiasis (25 male, 18 female; age range: 17-61 yr), urine saturation with calcium oxalate (CaOx) was calculated using a computer program (Equil-AT). The results in kJ/mol (normal < or = 2.0) were compared with those obtained with a simple test consisting of the addition of increasing amounts of calcium and oxalate to 3 tubes containing 10 ml urine. Development of turbidity in tube I denotes urine supersaturation with CaOx; turbidity in tubes II or III indicates lesser degrees of saturation, while a lack of turbidity (NT: no turbidity) shows undersaturation. Both saturation estimates were performed using aliquots of 24-hour urine collections. Twenty-one samples developed turbidity in tube I, 9 in tube II, and 6 in tube III, while 7 showed NT. There were no significant differences in Equil-AT values between II and III and between III and NT (p < 0.05 for I vs II and I vs III). High computer-calculated saturation values were found in 86% of group I samples and 22% of group II samples. None of the urines exhibiting turbidity in tube III or NT had high saturation values as estimated by Equil-AT. The turbidity test was also performed using freshly voided morning urine samples from the same patients. The results agreed with those obtained using the same test on aliquots of 24-hour collections, although in 10.8% of patients the morning test indicated higher levels of saturation. It is concluded that the turbidity test is a rapid, inexpensive and accurate way of estimating urine saturation with CaOx in the outpatient clinic.

**URL:** <https://ovidsp.ovid.com/ovidweb.cgi?T=JS&CSC=Y&NEWS=N&PAGE=fulltext&D=med3&DO=10.1111%2fj.1442-2042.1994.tb00056.x> <https://libkey.io/libraries/2592/openurl?genre=article&sid=OVID:medline&id=pmid:7614393&id=doi:10.1111%2Fj.1442-2042.1994.tb00056.x&issn=0919-8172&isbn=&volume=1&issue=4&spage=316&pages=316-8&date=1994&title=International+Journal+of+Urology&atitle=Evaluation+of+a+simple+test+to+estimate+urine+saturation+with+calcium+oxalate+in+stone-forming+patients.&aulast=Sanchez&pid=%3Cauthor%3ESanchez+A%3BGonzalez+A%3BSarano+HD%3BReinaldi+S%3C%2Fauthor%3E%3CAN%3E7614393%3C%2FAN%3E%3CDT%3EComparative+Study%3C%2FDT%3E>

270. **Investigation of single and recurrent stone formers**

**Item Type:** Journal Article

**Authors:** Tiselius, H. G.

**Publication Date:** 1994

**Journal:** Mineral & Electrolyte Metabolism 20(6), pp. 321–327

**Abstract:** A routine programme for investigation of patients with renal stone disease is presented. The different steps in this process are determined by the stone composition and the patient's previous history of stone formation. Assessment of the supersaturation and the crystallization properties are carried out in urine collected during a 24-hour period in one 16-hour and one 8-hour sample. The programme described in this paper has been formulated with a therapeutic goal in mind, whereby expensive and unnecessary procedures are avoided. [References: 24]

**URL:** <https://ovidsp.ovid.com/ovidweb.cgi?T=JS&CSC=Y&NEWS=N&PAGE=fulltext&D=med3&AN=7783694> <https://libkey.io/libraries/2592/openurl?genre=article&sid=OVID:medline&id=pmid:7783694&id=doi:&issn=0378-0392&isbn=&volume=20&issue=6&spage=321&pages=321-7&date=1994&title=Mineral+%26+Electrolyte+Metabolism&atitle=Investigation+of+single+and+recurrent+stone+formers.&aulast=Tiselius&pid=%3Cauthor%3ETiselius+HG%3C%2Fauthor%3E%3CAN%3E7783694%3C%2FAN%3E%3CDT%3EComparative+Study%3C%2FDT%3E>

271. **Seasonal changes in serum and erythrocyte potassium among renal stone formers from northeastern Thailand.**

**Item Type:** Journal Article

**Authors:** Sriboonlue, P.;Tungsanga, K.;Tosukhowong, P. and Sitprija, V.

**Publication Date:** Ju ,1993

**Journal:** Southeast Asian Journal of Tropical Medicine & Public Health 24(2), pp. 287–292

**Abstract:** Hypocitraturia, hypokalemia and low urinary excretion of potassium are common findings in nephrolithiasis in Northeastern Thailand. However, intracellular potassium has not been studied. We measured serum potassium, erythrocyte potassium, 24-hour urinary excretion of sodium, potassium, citrate, ammonium, titratable acid and pH in 17 nephrolithiasis patients from Northeastern Thailand during 2 seasons: the cool and hot months. There were no significant differences in urinary pH, ammonium, titratable acid, citrate and potassium during these 2 seasons. However, hypocitraturia and hypokaliuria were observed in the majority of cases in both periods. Seasonal variation in serum potassium, erythrocyte potassium, urine volume and urinary excretion of sodium was observed. Values were significantly lower in the hot months. In the cool months, the urinary excretion of citrate correlated linearly with that of potassium (r = 0.696, p < 0.002). Such a correlation was not found in the hot months. We concluded that extracellular and intracellular potassium depletion might be present in these patients and is greater during the hot than during the cool months. The pathogenesis may be multi-factorial.

**URL:** <https://ovidsp.ovid.com/ovidweb.cgi?T=JS&CSC=Y&NEWS=N&PAGE=fulltext&D=med3&AN=8266231> <https://libkey.io/libraries/2592/openurl?genre=article&sid=OVID:medline&id=pmid:8266231&id=doi:&issn=0125-1562&isbn=&volume=24&issue=2&spage=287&pages=287-92&date=1993&title=Southeast+Asian+Journal+of+Tropical+Medicine+%26+Public+Health&atitle=Seasonal+changes+in+serum+and+erythrocyte+potassium+among+renal+stone+formers+from+northeastern+Thailand.&aulast=Sriboonlue&pid=%3Cauthor%3ESriboonlue+P%3BTungsanga+K%3BTosukhowong+P%3BSitprija+V%3C%2Fauthor%3E%3CAN%3E8266231%3C%2FAN%3E%3CDT%3EJournal+Article%3C%2FDT%3E>

272. **Alterations in renal stone risk factors after space flight.**

**Item Type:** Journal Article

**Authors:** Whitson, P. A.;Pietrzyk, R. A.;Pak, C. Y. and Cintron, N. M.

**Publication Date:** Se ,1993

**Journal:** Journal of Urology 150(3), pp. 803–807

**Abstract:** Exposure to the microgravity environment of space produces a number of physiological changes of metabolic and environmental origin that could increase the potential for renal stone formation. Metabolic, environmental and physicochemical factors that influence renal stone risk potential were examined in 24-hour urine samples from astronauts 10 days before launch and on landing day to provide an immediate postflight assessment of these factors. In addition, comparisons were made between male and female crewmembers, and between crewmembers on missions of less than 6 days and those on 6 to 10-day missions. Results suggest that immediately after space flight the risk of calcium oxalate and uric acid stone formation is increased as a result of metabolic (hypercalciuria, hypocitraturia, pH) and environmental (lower urine volume) derangements, some of which could reflect residual effects of having been exposed to microgravity.

**URL:** <https://ovidsp.ovid.com/ovidweb.cgi?T=JS&CSC=Y&NEWS=N&PAGE=fulltext&D=med3&DO=10.1016%2fs0022-5347%2817%2935618-5> <https://libkey.io/libraries/2592/openurl?genre=article&sid=OVID:medline&id=pmid:8345588&id=doi:10.1016%2Fs0022-5347%2817%2935618-5&issn=0022-5347&isbn=&volume=150&issue=3&spage=803&pages=803-7&date=1993&title=Journal+of+Urology&atitle=Alterations+in+renal+stone+risk+factors+after+space+flight.&aulast=Whitson&pid=%3Cauthor%3EWhitson+PA%3BPietrzyk+RA%3BPak+CY%3BCintron+NM%3C%2Fauthor%3E%3CAN%3E8345588%3C%2FAN%3E%3CDT%3EJournal+Article%3C%2FDT%3E>

273. **Effect of hydrochlorothiazide on renal hypercalciuria.**

**Item Type:** Journal Article

**Authors:** Voskaki, I.;al Qadreh, A.;Mengreli, C. and Sbyrakis, S.

**Publication Date:** 1992

**Journal:** Child Nephrology & Urology 12(1), pp. 6–9

**Abstract:** The long-term effect of hydrochlorothiazide (HCT) was studied in 30 children (17 boys and 13 girls), aged 4-13 years (mean 7.6 +/- 3.4), with renal hypercalciuria (HCU) for a period of 2.5 +/- 0.95 years. The purpose of the study was to evaluate the effect of HCT mainly on renal calcium excretion, but also on serum K+, Ca, P, Mg, cPTH, nephrogenic cAMP (ncAMP) and bone mineral content (BMC). In addition to the above-mentioned parameters, oxalate, uric acid, citrate and cystine levels in 24-hour urine collections were also measured, and UCa/UCr, UP/UCr, UMg/UCr in 3-hour urine collections were assayed. The results of the study showed that 1 mg/kg/day of HCT caused a rapid and long-lasting correction of HCU, a decrease in serum cPTH and ncAMP, a significant increase in BMC and prevention of the formation of new urinary stones or of the increase in size of stones already present. No side effects were observed.

**URL:** <https://ovidsp.ovid.com/ovidweb.cgi?T=JS&CSC=Y&NEWS=N&PAGE=fulltext&D=med3&AN=1606587> <https://libkey.io/libraries/2592/openurl?genre=article&sid=OVID:medline&id=pmid:1606587&id=doi:&issn=1012-6694&isbn=&volume=12&issue=1&spage=6&pages=6-9&date=1992&title=Child+Nephrology+%26+Urology&atitle=Effect+of+hydrochlorothiazide+on+renal+hypercalciuria.&aulast=Voskaki&pid=%3Cauthor%3EVoskaki+I%3Bal+Qadreh+A%3BMengreli+C%3BSbyrakis+S%3C%2Fauthor%3E%3CAN%3E1606587%3C%2FAN%3E%3CDT%3EJournal+Article%3C%2FDT%3E>

274. **Metabolic studies in kidney stone disease.**

**Item Type:** Journal Article

**Authors:** Wong, S. Y.;Slater, S. R.;Evans, R. A.;Mason, R.;Lancaster, E. K.;Acland, S. M.;Eade, Y.;Hills, E. and Dunstan, C. R.

**Publication Date:** Ma ,1992

**Journal:** Quarterly Journal of Medicine 82(299), pp. 247–258

**Abstract:** Patients with kidney stones (n = 59) and healthy controls (n = 31) collected a 24-hour urine sample and later underwent a 6-hour 'fast and load' test in which an oral calcium load was taken after 2 hours. In the 24-hour urine sample, mean calcium excretion was higher in patients than controls, while mean urate, oxalate and citrate levels were similar. The patients had higher levels of fasting plasma calcium, serum calcitriol and fasting urinary calcium, and lower levels of plasma phosphate than did the controls. Following the calcium load, plasma and urinary calcium increased similarly in both groups. Serum parathyroid hormone (PTH) levels were similar in both groups and decreased similarly following the calcium load. Multiple linear regression, relating the presence or absence of stone formation to all variables, found the only variables significantly related to stone formation to be plasma levels of calcium (p less than 0.001) and phosphate (p = 0.001) and fasting urinary urea (p less than 0.001), and 24-hour urinary calcium excretion (p less than 0.05). Urinary oxalate and citrate were not related to stone formation. The data do not support the hypothesis that primary stimulation by calcitriol produces a normal fasting plasma calcium level, with an exaggerated increase after an oral calcium load. The findings instead suggest an abnormality of parathyroid cell 'set point', such that PTH secretion continues until the plasma calcium level is a little higher and the phosphate a little lower than in controls.

**URL:** <https://ovidsp.ovid.com/ovidweb.cgi?T=JS&CSC=Y&NEWS=N&PAGE=fulltext&D=med3&AN=1631259> <https://libkey.io/libraries/2592/openurl?genre=article&sid=OVID:medline&id=pmid:1631259&id=doi:&issn=0033-5622&isbn=&volume=82&issue=299&spage=247&pages=247-58&date=1992&title=Quarterly+Journal+of+Medicine&atitle=Metabolic+studies+in+kidney+stone+disease.&aulast=Wong&pid=%3Cauthor%3EWong+SY%3BSlater+SR%3BEvans+RA%3BMason+R%3BLancaster+EK%3BAcland+SM%3BEade+Y%3BHills+E%3BDunstan+CR%3C%2Fauthor%3E%3CAN%3E1631259%3C%2FAN%3E%3CDT%3EJournal+Article%3C%2FDT%3E>

275. **Diet, vitamin D and vertebral mineral density in hypercalciuric calcium stone formers.**

**Item Type:** Journal Article

**Authors:** Bataille, P.;Achard, J. M.;Fournier, A.;Boudailliez, B.;Westeel, P. F.;el Esper, N.;Bergot, C.;Jans, I.;Lalau, J. D. and Petit, J.

**Publication Date:** Ju ,1991

**Journal:** Kidney International 39(6), pp. 1193–1205

**Abstract:** To elucidate the pathophysiology of dietary calcium independent hypercalciuria, 42 calcium stone formers (Ca SF) were selected because they had on free diet a calciuria greater than 0.1 mmol/kg/day. For four days they were put on a diet restricted in calcium (Ca RD) by exclusion of the dairy products. They collected 24 hour urines on free diet and on day 4 of Ca RD as well as the two-hour fasting urines on the morning of the day 5 and the four-hour urines passed after an oral calcium load of 1 g, for measurement of creatinine, Ca, PO4, urea and total hydroxyprolinuria (THP). On day 5 fasting plasma concentrations of Ca, PO4, intact PTH, Gla protein, calcidiol and calcitriol were measured. The patients were firstly classified into dietary hypercalciuria (DH, 18 patients) and dietary calcium-independent hypercalciuria (IH, 24 patients) on the basis of the disappearance or not of hypercalciuria on Ca RD. Then the patients with IH were subclassified into absorptive hypercalciuria (AH) because of normal fasting calciuria (8 patients) and into fasting hypercalciuria (16 patients). Fasting hypercalciuric patients were subsequently divided according to the PTH levels into renal hypercalciuria (RH, 1 patient) with elevated fasting PTH becoming normal after the Ca load and undetermined hypercalciuria (UH, 15 patients) with normal PTH levels. Furthermore, their vertebral mineral density (VMD) was measured by quantitative computerized tomography which was normal in DH (91 +/- 6% of the normal mean for age and sex) but was decreased in IH to 69 +/- 4%. No difference in VMD was observed between AH and UH. Urinary excretions of urea, phosphate and THP was higher in IH than in DH and comparable in AH and UH. Sodium excretion Ca RD was the same in all groups and subgroups as well as the plasma parameters. Plasma calcitriol was increased in IH and DH comparatively to normal in spite of normal plasma calcidiol. Calciuria increase after oral calcium load, an index of Ca absorption, was higher in IH than in controls and comparable in IH and DH as well as in the three subgroups of IH. From these data and correlation studies in IH it is concluded: (1.) VMD is decreased in Ca stone formers with IH but not in those with DH, making the distinction of these two groups of hypercalciuria patients clinically relevant.(ABSTRACT TRUNCATED AT 400 WORDS)

**URL:** <https://ovidsp.ovid.com/ovidweb.cgi?T=JS&CSC=Y&NEWS=N&PAGE=fulltext&D=med3&DO=10.1038%2fki.1991.151> <https://libkey.io/libraries/2592/openurl?genre=article&sid=OVID:medline&id=pmid:1895673&id=doi:10.1038%2Fki.1991.151&issn=0085-2538&isbn=&volume=39&issue=6&spage=1193&pages=1193-205&date=1991&title=Kidney+International&atitle=Diet%2C+vitamin+D+and+vertebral+mineral+density+in+hypercalciuric+calcium+stone+formers.&aulast=Bataille&pid=%3Cauthor%3EBataille+P%3BAchard+JM%3BFournier+A%3BBoudailliez+B%3BWesteel+PF%3Bel+Esper+N%3BBergot+C%3BJans+I%3BLalau+JD%3BPetit+J%3Bet+al%3C%2Fauthor%3E%3CAN%3E1895673%3C%2FAN%3E%3CDT%3EJournal+Article%3C%2FDT%3E>

276. **Comparison of patients with idiopathic calcium phosphate and calcium oxalate stones.**

**Item Type:** Journal Article

**Authors:** Gault, M. H.;Chafe, L. L.;Morgan, J. M.;Parfrey, P. S.;Harnett, J. D.;Walsh, E. A.;Prabhakaran, V. M.;Dow, D. and Colpitts, A.

**Publication Date:** No ,1991

**Journal:** Medicine 70(6), pp. 345–359

**Abstract:** Our primary objective was to test the hypothesis that a defect in acidification is more common in patients who have idiopathic calcium phosphate kidney stones than in those whose stones are formed mainly of calcium oxalate. Additionally, other risk factors might differ for these 2 stone types. Urine pH was measured serially over 24 hours, and along with ammonium and titratable acid, it was measured before and serially after ingestion of ammonium chloride in 3 groups of subjects: 24 patients with predominantly calcium phosphate stones, 30 patients with calcium oxalate stones, and 15 health non-stone-formers. Twenty-six parameters potentially related to stone formation and acidification were assayed on urines collected over 24 hours, and 15 parameters on blood. The data base was a computerized list of 5900 analyses of stones from patients living in Newfoundland. Patients not known by their physician to have had urinary tract infection, anatomical abnormality, hyperparathyroidism, or renal tubular acidosis were asked to participate in the study. Differences between means were considered significant if p values were less than 0.05 for F by analysis of variance and also less than 0.01 by t-test. In all patients with calcium oxalate stones and all non-stone-formers, urine acidified to pH less than 5.25, but in 8 of the 23 phosphate stone formers who completed the ammonium chloride study urine failed to acidify to pH less than 5.25. As all 8 had normal values for venous pH, total CO2, and chloride, they were considered to have incomplete renal tubular acidosis (IRTA). The 8 phosphate stone formers with IRTA had greater mean values for urine pH on all 9 specimens collected serially over 24 hours (all means greater than 6.2), and after administration of ammonium chloride (p less than 0.01), as well as lower mean values for urine titratable acid excretion (p less than 0.01), both after administration of ammonium chloride and in 24-hour urine samples, compared with the remaining phosphate stone formers whose urine acidified and the oxalate and non-stone-forming control groups. Nearly all the phosphate stone formers had 1 or more risk factors for stone formation, but with frequencies not significantly higher than those found in the oxalate group. Hypercalciuria and hypocitruria were the commonest, but increased oxalate or urate also occurred. Thus, idiopathic calcium phosphate stone formation can be associated with 1 or more of several risk factors, and, with the possible exception of those with IRTA, treatment should be similar to that given to patients with calcium oxalate stones.

**URL:** <https://ovidsp.ovid.com/ovidweb.cgi?T=JS&CSC=Y&NEWS=N&PAGE=fulltext&D=med3&DO=10.1097%2f00005792-199111000-00001> <https://libkey.io/libraries/2592/openurl?genre=article&sid=OVID:medline&id=pmid:1956278&id=doi:10.1097%2F00005792-199111000-00001&issn=0025-7974&isbn=&volume=70&issue=6&spage=345&pages=345-59&date=1991&title=Medicine&atitle=Comparison+of+patients+with+idiopathic+calcium+phosphate+and+calcium+oxalate+stones.&aulast=Gault&pid=%3Cauthor%3EGault+MH%3BChafe+LL%3BMorgan+JM%3BParfrey+PS%3BHarnett+JD%3BWalsh+EA%3BPrabhakaran+VM%3BDow+D%3BColpitts+A%3C%2Fauthor%3E%3CAN%3E1956278%3C%2FAN%3E%3CDT%3EComparative+Study%3C%2FDT%3E>

277. **A retrospective analysis of the metabolic status of stone formers in the New York City metropolitan areas.**

**Item Type:** Journal Article

**Authors:** Hatch, M.;Schepers, A.;Grunberger, I. and Godec, C. J.

**Publication Date:** Ma ,1991

**Journal:** New York State Journal of Medicine 91(5), pp. 196–200

**Abstract:** Eighty stone-forming patients were evaluated as outpatients for risk factors involved in stone formation. Analyses of the major stone risk parameters in the 24-hour collections indicated low volume (less than 2L) in about 79% of these patients. Forty percent of all patients were found to be hypocitraturic, 30% were hyperoxaluric, 18% were hypercalciuric, and 13% were hyperuricosuric. Sixty-one percent of the patients had a normal calcium load study; of these, four patients were uric acid stone formers. The remaining 39% of patients were classified as either absorptive hypercalciuric (16 patients), or fasting hypercalciuric with normal (12 patients) or elevated (3 patients) serum parathyroid hormone (PTH) levels. Of these 31 patients, 61% were not hypercalciuric on the basis of the 24-hour urine collections. The protocol provided a diagnosis in over 98% of patients. Low urinary volume was the only defect found in 5% of patients, while more than one risk factor was identified in 93%.

**URL:** <https://ovidsp.ovid.com/ovidweb.cgi?T=JS&CSC=Y&NEWS=N&PAGE=fulltext&D=med3&AN=1857571> <https://libkey.io/libraries/2592/openurl?genre=article&sid=OVID:medline&id=pmid:1857571&id=doi:&issn=0028-7628&isbn=&volume=91&issue=5&spage=196&pages=196-200&date=1991&title=New+York+State+Journal+of+Medicine&atitle=A+retrospective+analysis+of+the+metabolic+status+of+stone+formers+in+the+New+York+City+metropolitan+areas.&aulast=Hatch&pid=%3Cauthor%3EHatch+M%3BSchepers+A%3BGrunberger+I%3BGodec+CJ%3C%2Fauthor%3E%3CAN%3E1857571%3C%2FAN%3E%3CDT%3EJournal+Article%3C%2FDT%3E>

278. **Urinary excretion of oxalate by patients with renal hypercalciuric stone disease. Effect of chronic treatment with hydrochlorothiazide.**

**Item Type:** Journal Article

**Authors:** Urivetzky, M.;Braverman, S.;Motola, J. A. and Smith, A. D.

**Publication Date:** Ap ,1991

**Journal:** Urology 37(4), pp. 327–330

**Abstract:** Hydrochlorothiazide is employed to reduce calcium excretion in patients with urinary stone disease secondary to renal leak hypercalciuria. Because the drug also has been reported to be a competitive inhibitor of oxalate excretion by the renal tubules, we sought to determine whether chronic use indeed affected the amount of oxalate excreted. Patients taking hydrochlorothiazide 50 mg daily did not have a statistically significant reduction in twenty-four-hour urinary oxalate on their customary diets (pretreatment 37 +/- 3 mg/day [mean +/- S.E.M.; N = 22]; at one year 36 +/- 3 mg/day [N = 22]; at two years 37 +/- 3 mg/day [N = 16]). In 12 patients who voluntarily collected twelve-hour urine specimens after dinner on the third day of a low-oxalate diet and again the next day after a 1 g oxalate load, hydrochlorothiazide had no significant effect on oxalate excretion (19 +/- 2.3 mmol oxalate/mol creatinine on hydrochlorothiazide versus 20.6 +/- 2.6 mmol off the drug after low oxalate meal; 50 +/- 7.8 mmol/mol creatinine on hydrochlorothiazide versus 56.2 +/- 7.5 mmol off the drug after an oxalate load). As expected, there was a significant reduction in urinary calcium excretion and thus of calcium oxalate urinary saturation during hydrochlorothiazide administration. Hydrochlorothiazide by itself is not sufficient to reduce oxalate excretion in patients with renal leak hypercalciuria.

**URL:** <https://ovidsp.ovid.com/ovidweb.cgi?T=JS&CSC=Y&NEWS=N&PAGE=fulltext&D=med3&DO=10.1016%2f0090-4295%2891%2980258-9> <https://libkey.io/libraries/2592/openurl?genre=article&sid=OVID:medline&id=pmid:2014597&id=doi:10.1016%2F0090-4295%2891%2980258-9&issn=0090-4295&isbn=&volume=37&issue=4&spage=327&pages=327-30&date=1991&title=Urology&atitle=Urinary+excretion+of+oxalate+by+patients+with+renal+hypercalciuric+stone+disease.+Effect+of+chronic+treatment+with+hydrochlorothiazide.&aulast=Urivetzky&pid=%3Cauthor%3EUrivetzky+M%3BBraverman+S%3BMotola+JA%3BSmith+AD%3C%2Fauthor%3E%3CAN%3E2014597%3C%2FAN%3E%3CDT%3EJournal+Article%3C%2FDT%3E>

279. **Cystinuria**

**Item Type:** Journal Article

**Authors:** Milliner, D. S.

**Publication Date:** De ,1990

**Journal:** Endocrinology & Metabolism Clinics of North America 19(4), pp. 889–907

**Abstract:** Cystinuria is an hereditary disorder of renal and intestinal transport characterized by the excessive urinary excretion of cystine, arginine, lysine, and ornithine. It is inherited as a common recessive gene with allelic mutations. Complementary studies of the plasma response to oral cystine loading, intestinal mucosal transport patterns, and urine cystine excretion allow separation of homozygous cystinuric subjects into three groups. In type I, the most common form, there is no active transport of cystine or dibasic amino acids across the mucosal gradient, and heterozygous subjects show normal urine cystine values. Type II is characterized by markedly reduced or absent intestinal transport of cystine. Heterozygotes for type II show significantly elevated urine cystine but less than is seen in homozygotes. In type III there is diminished, although demonstrable, intestinal absorption of cystine and dibasic amino acids. Urine cystine in heterozygotes is intermediate between types I and II. Urolithiasis with its attendant complications is the sole clinical manifestation of cystinuria and is due to the relative insolubility of cystine in the urine. The urolithiasis may become clinically manifest at any time from infancy through the ninth decade, although the mean age is the second to third decade. Clinical presentation is similar to that of other types of urolithiasis. Although cystinuria accounts for only 1% to 2% of all urolithiasis and 6% to 8% of urolithiasis in pediatric populations, repeated stone formation in affected patients often causes considerable morbidity. Cystine crystals in the urine are diagnostic but show up in only 19% to 26% of homozygous cystinuric patients. Sodium cyanide nitroprusside is a suitable screening test that should identify homozygous stone formers but will not detect all heterozygotes. A positive screening test should be followed by quantitation of urinary amino acids. A homozygous patient can be functionally defined as one who excretes 250 mg or more of cystine/g of creatinine in a 24-hour urine collection. Other causes of excess urinary cystine must be excluded. Medical therapy will be directed toward dissolution of existing calculi and prevention of new stone formation. Increasing urine volume by generous oral fluid intake is beneficial. Dietary sodium restriction has a favorable effect on urinary cystine excretion. Cystine solubility can be improved by urinary alkalinization and where necessary by the administration of thiol chelators, particularly D-penicillamine or mercaptopropionylglycine. Because these chelators have significant adverse effects, they should be reserved for patients who do not respond to a more conservative program. Patients with infected, symptomatic, or obstructing stones require surgical intervention.(ABSTRACT TRUNCATED AT 400 WORDS) [References: 112]

**URL:** <https://ovidsp.ovid.com/ovidweb.cgi?T=JS&CSC=Y&NEWS=N&PAGE=fulltext&D=med3&AN=2081517> <https://libkey.io/libraries/2592/openurl?genre=article&sid=OVID:medline&id=pmid:2081517&id=doi:&issn=0889-8529&isbn=&volume=19&issue=4&spage=889&pages=889-907&date=1990&title=Endocrinology+%26+Metabolism+Clinics+of+North+America&atitle=Cystinuria.&aulast=Milliner&pid=%3Cauthor%3EMilliner+DS%3C%2Fauthor%3E%3CAN%3E2081517%3C%2FAN%3E%3CDT%3EJournal+Article%3C%2FDT%3E>

280. **Urinary constituents in an endemic area of stones and renal tubular acidosis in northeastern Thailand.**

**Item Type:** Journal Article

**Authors:** Nilwarangkur, S.;Malasit, P.;Nimmannit, S.;Susaengrat, W.;Ong-Aj-Yooth, S.;Vasuvattakul, S. and Pidetcha, P.

**Publication Date:** Se ,1990

**Journal:** Southeast Asian Journal of Tropical Medicine & Public Health 21(3), pp. 437–441

**Abstract:** Nephrolithiasis and endemic renal distal tubular acidosis are common in northeastern Thailand. The etiology is still unknown. It is generally accepted that urine electrolytes influence the capacity of urine to inhibit or promote renal and also bladder stones. The purpose of this study was to analyse the composition of the urine in the indigenous population in the northeast area and compare their values with data obtained from a group of age matched adults, living in Bangkok. Twenty-four hour urine samples from 23 normal adult villagers from six villages within the province of Khon Kaen and 34 normal adults living in Bangkok were collected, and the daily excretion of creatinine, uric acid, calcium and inorganic phosphate, sodium, potassium, chloride, magnesium and oxalate were assayed. Daily urinary sodium, potassium, chloride and phosphate of the villagers were significantly lower than those of Bangkokians. No difference in the urinary excretion of calcium, magnesium, uric acid, oxalate and creatinine was found. The Na/Ca, and Ca/PO4 ratios of villagers were significantly lower than those of the Bangkok subjects. The villagers excreted significantly lower amounts of Na in the face of relatively higher urinary Ca. The above data, combined with our previous study showing the low values of urinary citrate in the villagers in the same areas, strongly indicate that the indigeneous population is at high risk in developing urolithiasis. The causes for these electrolyte abnormalities are still unknown. Low contents of the major electrolytes in their diets might play an important role. Low phosphate output indicates low protein diets.(ABSTRACT TRUNCATED AT 250 WORDS)

**URL:** <https://ovidsp.ovid.com/ovidweb.cgi?T=JS&CSC=Y&NEWS=N&PAGE=fulltext&D=med3&AN=2075484> <https://libkey.io/libraries/2592/openurl?genre=article&sid=OVID:medline&id=pmid:2075484&id=doi:&issn=0125-1562&isbn=&volume=21&issue=3&spage=437&pages=437-41&date=1990&title=Southeast+Asian+Journal+of+Tropical+Medicine+%26+Public+Health&atitle=Urinary+constituents+in+an+endemic+area+of+stones+and+renal+tubular+acidosis+in+northeastern+Thailand.&aulast=Nilwarangkur&pid=%3Cauthor%3ENilwarangkur+S%3BMalasit+P%3BNimmannit+S%3BSusaengrat+W%3BOng-Aj-Yooth+S%3BVasuvattakul+S%3BPidetcha+P%3C%2Fauthor%3E%3CAN%3E2075484%3C%2FAN%3E%3CDT%3EComparative+Study%3C%2FDT%3E>

281. **Nephrolithiasis and urine ion changes in ulcerative colitis patients undergoing colectomy and endorectal ileal pullthrough.**

**Item Type:** Journal Article

**Authors:** Stelzner, M.;Phillips, J. D.;Saleh, S. and Fonkalsrud, E. W.

**Publication Date:** Ju ,1990

**Journal:** Journal of Surgical Research 48(6), pp. 552–556

**Abstract:** Nephrolithiasis occurs in 5 to 13% of patients with ulcerative colitis (UC) who undergo colectomy and abdominal ileostomy, presumably from chronic dehydration and urinary concentration. Whether endorectal ileal pullthrough with ileal reservoir (PTR) changes the incidence of stones (primarily calcium oxalate) after colectomy is not known. Urinary excretion of Na2+, K+, Ca2+, Mg2+, phosphate, urate, oxalate, and citrate was measured in a prospective study of 12 UC patients undergoing PTR with temporary end ileostomy. Twenty-four-hour urine samples were obtained before colectomy (t1), after colectomy but before ileostomy closure (t2), and 5 months after ileostomy closure (t3). Urine volumes decreased from 831 +/- 101 cc (mean +/- SE) at t1 to 715 +/- 101 cc at t2 and then increased to 1278 +/- 421 cc at t3 (significant, with P less than 0.01 by t test). Urinary excretions of Mg2+, oxalate, and citrate were low in UC patients compared to those in controls (15 healthy adult volunteers). Excretion of Ca2+ increased significantly following temporary ileostomy while excretion of Mg2+ fell. Excretion of Ca2+ fell and excretion of Mg2+ and citrate increased following PTR. We conclude that PTR patients have increased urine volumes and urinary ion changes known to decrease the risk of developing renal stones.

**URL:** <https://ovidsp.ovid.com/ovidweb.cgi?T=JS&CSC=Y&NEWS=N&PAGE=fulltext&D=med3&DO=10.1016%2f0022-4804%2890%2990229-u> <https://libkey.io/libraries/2592/openurl?genre=article&sid=OVID:medline&id=pmid:2362415&id=doi:10.1016%2F0022-4804%2890%2990229-u&issn=0022-4804&isbn=&volume=48&issue=6&spage=552&pages=552-6&date=1990&title=Journal+of+Surgical+Research&atitle=Nephrolithiasis+and+urine+ion+changes+in+ulcerative+colitis+patients+undergoing+colectomy+and+endorectal+ileal+pullthrough.&aulast=Stelzner&pid=%3Cauthor%3EStelzner+M%3BPhillips+JD%3BSaleh+S%3BFonkalsrud+EW%3C%2Fauthor%3E%3CAN%3E2362415%3C%2FAN%3E%3CDT%3EJournal+Article%3C%2FDT%3E>

282. **Studies on citrate metabolism in normal subjects and kidney stone patients.**

**Item Type:** Journal Article

**Authors:** Minisola, S.;Rossi, W.;Pacitti, M. T.;Scarnecchia, L.;Bigi, F.;Carnevale, V. and Mazzuoli, G.

**Publication Date:** 1989

**Journal:** Mineral & Electrolyte Metabolism 15(5), pp. 303–308

**Abstract:** The study was carried out in order to investigate citrate metabolism (i.e. serum levels, filtered load, tubular reabsorption, total and fasting urinary excretion) in a large group of normal subjects (n = 39) and kidney stone patients (n = 45). We found that mean serum citrate levels and filtered loads were similar in the two groups studied. On the other hand, both the 24-hour urinary citrate and the fasting citrate/creatinine ratio were significantly reduced, whereas the mean tubular reabsorption of citrate appeared to be significantly increased in kidney stone patients in respect to control subjects. A good correlation was found between the fasting and the 24-hour citrate/creatinine ratio in both normal subjects (r = 0.84, p less than 0.001) and kidney stone patients (r = 0.89, p less than 0.001). Our data indicate that the kidney appears to be involved in the pathogenesis of hypocitraturia in kidney stone patients and that a substantial proportion of these patients show a reduced urinary excretion of citrate. Finally, the evaluation of the fasting citrate/creatinine ratio may replace the measurement of the substance on the basis of a 24-hour urine collection.

**URL:** <https://ovidsp.ovid.com/ovidweb.cgi?T=JS&CSC=Y&NEWS=N&PAGE=fulltext&D=med3&AN=2811789> <https://libkey.io/libraries/2592/openurl?genre=article&sid=OVID:medline&id=pmid:2811789&id=doi:&issn=0378-0392&isbn=&volume=15&issue=5&spage=303&pages=303-8&date=1989&title=Mineral+%26+Electrolyte+Metabolism&atitle=Studies+on+citrate+metabolism+in+normal+subjects+and+kidney+stone+patients.&aulast=Minisola&pid=%3Cauthor%3EMinisola+S%3BRossi+W%3BPacitti+MT%3BScarnecchia+L%3BBigi+F%3BCarnevale+V%3BMazzuoli+G%3C%2Fauthor%3E%3CAN%3E2811789%3C%2FAN%3E%3CDT%3EComparative+Study%3C%2FDT%3E>

283. **Comparative study of the circadian rhythmicity in the urinary concentration of glycosaminoglycans in patients of calcium oxalate nephrolithiasis and in healthy adults.**

**Item Type:** Journal Article

**Authors:** Sidhu, H.;Thind, S. K.;Nath, R.;Vaidyanathan, S.;Hemal, A. K.;Mandal, A. K. and Krishan, K.

**Publication Date:** 1989

**Journal:** Urologia Internationalis 44(4), pp. 218–221

**Abstract:** Glycosaminoglycans (GAGs) are potent inhibitors of calcium oxalate crystallisation and/or crystal aggregation. Urinary concentration of GAGs has been shown to vary during 24 h; therefore, circadian rhythmicity in urinary concentration of GAGs was investigated in 33 healthy male adults in the age group of 20-40 years and in 27 male patients of a similar age group with calcium nephrolithiasis. Three-hourly urine samples were collected for 24 h beginning from 00.00 h for estimation of urinary concentration of GAGs. The data of each patient was analysed by single cosinor rhythmometry and population mean-cosinor rhythmometry was then applied to each group. Twenty-four-hour urinary excretion of GAGs was significantly less in renal calculus patients (16.867 +/- 5.89 mumol) than in healthy subjects (22.588 +/- 5.32 mumol; p less than 0.001). A statistically validated circadian rhythm in urinary concentration of GAGs was demonstrated in both the groups. However, the amplitude-acrophase test revealed a significant difference between the two groups (F2,57 = 8.305; p less than 0.001); the amplitude was 2.354 mumol/l in patients with nephrolithiasis, whereas the amplitude was 7.028 mumol/l in healthy adults. The mesor test also revealed a significant difference in the 3-hour urinary concentration of GAGs between the two groups (18.536 mumol/l in healthy adults vs. 9.728 mumol/l in patients with nephrolithiasis). Thus in patients with nephrolithiasis, not only is the 24-hour urinary excretion of GAGs significantly low but the 3-hourly urinary concentration of GAGs is also significantly decreased as compared to healthy subjects.(ABSTRACT TRUNCATED AT 250 WORDS)

**URL:** <https://ovidsp.ovid.com/ovidweb.cgi?T=JS&CSC=Y&NEWS=N&PAGE=fulltext&D=med3&DO=10.1159%2f000281507> <https://libkey.io/libraries/2592/openurl?genre=article&sid=OVID:medline&id=pmid:2800054&id=doi:10.1159%2F000281507&issn=0042-1138&isbn=&volume=44&issue=4&spage=218&pages=218-21&date=1989&title=Urologia+Internationalis&atitle=Comparative+study+of+the+circadian+rhythmicity+in+the+urinary+concentration+of+glycosaminoglycans+in+patients+of+calcium+oxalate+nephrolithiasis+and+in+healthy+adults.&aulast=Sidhu&pid=%3Cauthor%3ESidhu+H%3BThind+SK%3BNath+R%3BVaidyanathan+S%3BHemal+AK%3BMandal+AK%3BKrishan+K%3C%2Fauthor%3E%3CAN%3E2800054%3C%2FAN%3E%3CDT%3EComparative+Study%3C%2FDT%3E>

284. **Standardized estimate of the ion activity product of calcium oxalate in urine from renal stone formers.**

**Item Type:** Journal Article

**Authors:** Tiselius, H. G.

**Publication Date:** 1989

**Journal:** European Urology 16(1), pp. 48–50

**Abstract:** An estimate of the ion activity product of calcium oxalate [AP(CaOx) index(s)] was calculated for a standardized 24-hour urine volume of 1,500 ml. AP(CaOx) index(s) was higher in stone formers than in normal subjects and higher in men than women. There was a good correlation between AP(CaOx) index(s) and our previously described CaOx risk index. AP(CaOx) index(s) might be particularly useful for comparison of different groups of patients as well as for comparison of men and women. It can be assumed that such an index will be very useful for evaluation of patients with short-term urine collections.

**URL:** <https://ovidsp.ovid.com/ovidweb.cgi?T=JS&CSC=Y&NEWS=N&PAGE=fulltext&D=med3&DO=10.1159%2f000471529> <https://libkey.io/libraries/2592/openurl?genre=article&sid=OVID:medline&id=pmid:2714318&id=doi:10.1159%2F000471529&issn=0302-2838&isbn=&volume=16&issue=1&spage=48&pages=48-50&date=1989&title=European+Urology&atitle=Standardized+estimate+of+the+ion+activity+product+of+calcium+oxalate+in+urine+from+renal+stone+formers.&aulast=Tiselius&pid=%3Cauthor%3ETiselius+HG%3C%2Fauthor%3E%3CAN%3E2714318%3C%2FAN%3E%3CDT%3EComparative+Study%3C%2FDT%3E>

285. **Osteocalcin, parathormone and hypercalciuria.**

**Item Type:** Journal Article

**Authors:** Rico, H.;Paramo, P.;Perez del Molino, J.;Nacarino, L. and Yague, M.

**Publication Date:** 1988

**Journal:** European Urology 15(3-4), pp. 239–242

**Abstract:** Osteocalcin synthesis is dependent on the influence of the renal vitamin D metabolite, 1,25(OH)2D3. This metabolite is an etiological factor in some hypercalciurias, and osteocalcin may thus be a parameter for discovering them. In turn, parathormone, which stimulates 1,25(OH)2D3 synthesis, is also implicated in the hypercalciurias. Mean molecular parathormone, osteocalcin, 24-hour calciuria and the calcium/creatinine and hydroxyproline/creatinine ratios were determined in urine samples obtained after a 12-hour fast from 18 patients with absorptive hypercalciuria and 11 patients with renal hypercalciuria out of a total of 62 patients with renal lithiasis. No changes were observed in osteocalcin or parathormone, indicating that neither is valid for the diagnosis of hypercalciuria. Significant differences were only found in the Ca/Cr ratio (p less than 0.001), which was higher (0.31 +/- 0.07 vs. 0.13 +/- 0.04 mg/mg) in renal hypercalciuria than in absorptive hypercalciuria. No changes in osteocalcin have been reported in the hypercalciurias, but variations in parathormone have been reported, therefore requiring further study and thought to understand the processes involved.

**URL:** <https://ovidsp.ovid.com/ovidweb.cgi?T=JS&CSC=Y&NEWS=N&PAGE=fulltext&D=med3&DO=10.1159%2f000473443> <https://libkey.io/libraries/2592/openurl?genre=article&sid=OVID:medline&id=pmid:3265105&id=doi:10.1159%2F000473443&issn=0302-2838&isbn=&volume=15&issue=3-4&spage=239&pages=239-42&date=1988&title=European+Urology&atitle=Osteocalcin%2C+parathormone+and+hypercalciuria.&aulast=Rico&pid=%3Cauthor%3ERico+H%3BParamo+P%3BPerez+del+Molino+J%3BNacarino+L%3BYague+M%3C%2Fauthor%3E%3CAN%3E3265105%3C%2FAN%3E%3CDT%3EJournal+Article%3C%2FDT%3E>

286. **Evaluation of factors involved in calcium stone formation.**

**Item Type:** Journal Article

**Authors:** Abraham, P. A. and Smith, C. L.

**Publication Date:** 1987

**Journal:** Mineral & Electrolyte Metabolism 13(3), pp. 201–208

**Abstract:** The role of urinary calcium oxalate supersaturation in the initiation of calcium stone formation is unclear since 24-hour urine collections from both stone-formers and nonstone-formers are supersaturated for calcium oxalate. Urinary supersaturation for calcium phosphate, uric acid, and monosodium urate or decreased excretion of inhibitors of stone formation, citrate and magnesium, may also contribute to calcium stone formation. To assess these possibilities we studied variations within 24 h of determinants for calcium stone formation. Six consecutive 4-hour urine specimens were collected from stone-formers (n = 15) and nonstone-formers (n = 11) during their usual diet and activities. Compared to nonstone-formers, urine from stone-formers had increased calcium concentrations, while pH and oxalate, phosphorus, uric acid, magnesium and citrate concentrations were not different. Urine from both stone-formers and nonstone-formers was always supersaturated for calcium oxalate, uric acid and monosodium urate with no difference between the groups. Urine from stone-formers was always supersaturated for calcium phosphate as brushite, while urine from nonstone-formers was mostly undersaturated for brushite. In addition to these physicochemical studies, we reviewed the composition of 235 stones containing calcium oxalate and analyzed it by crystallography. The center of 67% of these stones contained calcium phosphate. These physicochemical and stone analysis data suggest that precipitation of a calcium phosphate crystal nidus may contribute to the initiation of many calcium stones.

**URL:** <https://ovidsp.ovid.com/ovidweb.cgi?T=JS&CSC=Y&NEWS=N&PAGE=fulltext&D=med2&AN=3627051> <https://libkey.io/libraries/2592/openurl?genre=article&sid=OVID:medline&id=pmid:3627051&id=doi:&issn=0378-0392&isbn=&volume=13&issue=3&spage=201&pages=201-8&date=1987&title=Mineral+%26+Electrolyte+Metabolism&atitle=Evaluation+of+factors+involved+in+calcium+stone+formation.&aulast=Abraham&pid=%3Cauthor%3EAbraham+PA%3BSmith+CL%3C%2Fauthor%3E%3CAN%3E3627051%3C%2FAN%3E%3CDT%3EJournal+Article%3C%2FDT%3E>

287. **Type I primary hyperoxaluria associated with type I renal tubular acidosis.**

**Item Type:** Journal Article

**Authors:** Cheng, J. H.;Fu, Y. K.;Chen, W. P.;Hwang, B. T.;Chen, S. M. and Lin, C. Y.

**Publication Date:** 1987

**Journal:** International Journal of Pediatric Nephrology 8(4), pp. 235–238

**Abstract:** An 8-year-old boy who had suffered from recurrent stone formation since the age of 4 years, was admitted as an emergency due to anuria for a half day on November 20, 1986. Kidney-ureter-bladder film showed that the urethra was obstructed by a stone, and emergent cystoscopy was performed to remove it. He is the product of consanguinous marriage, his parents being first cousins. There was no family history of renal stone. Laboratory investigations showed hypokalemic, hyperchloremic metabolic acidosis. The ammonium chloride loading test revealed inability to acidify the urine and a markedly decreased excretion of titrable hydrogen ion and ammonium ion in the urine. These results indicate that this is a case of Type I renal tubular acidosis. His 24-hour urinary excretion of oxalate and glyoxylate were also markedly increased. There were no underlying causes leading to the development of secondary hyperoxaluria. These results also establish the diagnosis of Type I primary hyperoxaluria. The patient then received regimens of Polycitra 1ml/kg/day and Vitamin B6 50mg/day for 4 months. However, urinary stone developed again in this patient 4 months later. To our knowledge, Type I primary hyperoxaluria in association with Type I renal tubular acidosis has not been previously reported.

**URL:** <https://ovidsp.ovid.com/ovidweb.cgi?T=JS&CSC=Y&NEWS=N&PAGE=fulltext&D=med2&AN=3449474> <https://libkey.io/libraries/2592/openurl?genre=article&sid=OVID:medline&id=pmid:3449474&id=doi:&issn=0391-6510&isbn=&volume=8&issue=4&spage=235&pages=235-8&date=1987&title=International+Journal+of+Pediatric+Nephrology&atitle=Type+I+primary+hyperoxaluria+associated+with+type+I+renal+tubular+acidosis.&aulast=Cheng&pid=%3Cauthor%3ECheng+JH%3BFu+YK%3BChen+WP%3BHwang+BT%3BChen+SM%3BLin+CY%3C%2Fauthor%3E%3CAN%3E3449474%3C%2FAN%3E%3CDT%3ECase+Reports%3C%2FDT%3E>

288. **Significance of the calcium to creatinine concentration ratio of a single-voided urine specimen in patients with hypercalciuric urolithiasis.**

**Item Type:** Journal Article

**Authors:** Matsushita, K. and Tanikawa, K.

**Publication Date:** Se ,1987

**Journal:** Tokai Journal of Experimental & Clinical Medicine 12(3), pp. 167–171

**Abstract:** Thirty-six patients with recurrent calcium oxalate nephrolithiasis were selected from the stone clinic. Fourteen were normocalcemic and had normal daily urinary calcium excretion. Among 22 patients with idiopathic hypercalciuria, 10 received thiazide diuretics for the prevention of new stone formation. Single-voided urine samples were collected at the outpatient clinic and 24-hour urine at the patients' homes. In hypercalciuric patients, irrespective of thiazide diuretic therapy, the mean value of the calcium/creatinine concentration ratios of postprandial single-voided urine specimens had a meaningful correlation with the man value of 24-hour urinary calcium excretion rates. Also in hypercalciuric patients with thiazide diuretics, a negative correlation was observed between the calcium/creatinine concentration ratio and the index for urinary saturation with calcium oxalate of a postprandial single-voided urine sample. Thus, in the hypercalciuric stone formers, 24-hour urinary calcium excretion rates and the degree of urinary saturation with calcium oxalate can be estimated from the calcium/creatinine concentration ratios of single-voided urinary samples.

**URL:** <https://ovidsp.ovid.com/ovidweb.cgi?T=JS&CSC=Y&NEWS=N&PAGE=fulltext&D=med2&AN=3454077> <https://libkey.io/libraries/2592/openurl?genre=article&sid=OVID:medline&id=pmid:3454077&id=doi:&issn=0385-0005&isbn=&volume=12&issue=3&spage=167&pages=167-71&date=1987&title=Tokai+Journal+of+Experimental+%26+Clinical+Medicine&atitle=Significance+of+the+calcium+to+creatinine+concentration+ratio+of+a+single-voided+urine+specimen+in+patients+with+hypercalciuric+urolithiasis.&aulast=Matsushita&pid=%3Cauthor%3EMatsushita+K%3BTanikawa+K%3C%2Fauthor%3E%3CAN%3E3454077%3C%2FAN%3E%3CDT%3EJournal+Article%3C%2FDT%3E>

289. **Determination of urinary citrate by ion chromatography.**

**Item Type:** Journal Article

**Authors:** Ogawa, Y.;Morozumi, M.;Tanaka, T. and Yamaguchi, K.

**Publication Date:** Ja ,1986

**Journal:** Journal of Urology 135(1), pp. 178–181

**Abstract:** Ion chromatography was used to determine urinary citrate concentration. The technique is simple, requiring a 100-fold dilution of urine and 35 minutes at most for each analysis. The technique does, however, have the disadvantage that it requires a rather expensive automated system. The minimum detectable limit for citrate was 0.5 microgram/ml. in a standard solution, and the regression line for the standard curve from 0.5 to 50 micrograms/ml. citrate had a significant correlation coefficient (lnY = 1.048 X lnX-2.755, r = 0.997). The intra-run coefficient of variation was 0.9 per cent. The overall intra-run and inter-run coefficients of variation, including the sampling and dilution of urine, were 4.8 and 8.3 per cent respectively. There is a possibility that another ion is close enough to the citrate peak to interfere; however, this problem can be solved by the treatment of the sample with citrate lyase. When the present method was compared with the conventional enzyme method, a significant correlation between the results was obtained in both human and rat urine. The 24-hour urinary citrate excretion in eight normal human males was 73.8 to 378.4 mg./day (mean, 174.7 mg./day), while that in male Wistar-strain rats (approximately 150 gm.) was 1.5 to 18.6 mg./day (mean, 9.5 mg./day), as measured in more than 50 consecutive rat urine samples.

**URL:** <https://ovidsp.ovid.com/ovidweb.cgi?T=JS&CSC=Y&NEWS=N&PAGE=fulltext&D=med2&DO=10.1016%2fs0022-5347%2817%2945557-1> <https://libkey.io/libraries/2592/openurl?genre=article&sid=OVID:medline&id=pmid:3941460&id=doi:10.1016%2Fs0022-5347%2817%2945557-1&issn=0022-5347&isbn=&volume=135&issue=1&spage=178&pages=178-81&date=1986&title=Journal+of+Urology&atitle=Determination+of+urinary+citrate+by+ion+chromatography.&aulast=Ogawa&pid=%3Cauthor%3EOgawa+Y%3BMorozumi+M%3BTanaka+T%3BYamaguchi+K%3C%2Fauthor%3E%3CAN%3E3941460%3C%2FAN%3E%3CDT%3EComparative+Study%3C%2FDT%3E>

290. **Investigation and treatment of renal calculi.**

**Item Type:** Journal Article

**Authors:** Evans, R. A.;Maher, P. O.;Agostino, M.;Wong, S. Y.;Marel, G. M. and Hills, E.

**Publication Date:** Sep 30 ,1985

**Journal:** Medical Journal of Australia 143(7), pp. 278–281

**Abstract:** We have investigated and treated 176 patients who were suffering from renal calculi. The stones contained calcium in 87% of patients, predominantly urate in 11%, and rarely contained magnesium ammonium phosphate or cystine. Of the patients with calcium stones, hypercalciuria was present in 75% and was identified in 57% by the measurement of the 24-hour urinary calcium excretion, and in a further 18% by a standardization calcium "fast-and-load" test. Nine patients were found to have primary hyperparathyroidism and were treated surgically. A further 21% were suspected to have normocalcaemic hyperparathyroidism, and metabolic studies are being developed to clarify this. The treatment of hypercalciuria included a low-calcium diet, and various combinations of a thiazide diuretic, phosphate supplements and sodium cellulose phosphate. Hypercalciuria was controlled in all compliant patients, and only two developed further stones. Hyperuricosuria was rarely the sole metabolic abnormality in patients with calcium stones, though this might reflect the referral pattern of the Unit. Uric acid stones were frequently, but not invariably, associated with hyperuricosuria and acid urine, and even large uric acid calculi dissolved with a combined therapy of high fluid intake, allopurinol and an alkalinizing agent. Surgical treatment was rarely required in these patients. A stone in the renal pelvis of one patient was removed percutaneously and did not require ultrasonic fragmentation. Modern methods of investigation and treatment have greatly improved the outlook for patients with recurrent renal calculi.

**URL:** <https://ovidsp.ovid.com/ovidweb.cgi?T=JS&CSC=Y&NEWS=N&PAGE=fulltext&D=med2&DO=10.5694%2fj.1326-5377.1985.tb123006.x> <https://libkey.io/libraries/2592/openurl?genre=article&sid=OVID:medline&id=pmid:4046915&id=doi:10.5694%2Fj.1326-5377.1985.tb123006.x&issn=0025-729X&isbn=&volume=143&issue=7&spage=278&pages=278-81&date=1985&title=Medical+Journal+of+Australia&atitle=Investigation+and+treatment+of+renal+calculi.&aulast=Evans&pid=%3Cauthor%3EEvans+RA%3BMaher+PO%3BAgostino+M%3BWong+SY%3BMarel+GM%3BHills+E%3C%2Fauthor%3E%3CAN%3E4046915%3C%2FAN%3E%3CDT%3EJournal+Article%3C%2FDT%3E>

291. **Kidney stones. Medical management and newer options for stone 'removal'.**

**Item Type:** Journal Article

**Authors:** Kanig, S. P. and Conn, R. L.

**Publication Date:** 1985

**Journal:** Postgraduate Medicine 78(6), pp. 38–44

**Abstract:** Patients with recurrent kidney stone disease or stone formers at increased risk of recurrence deserve a thorough metabolic workup. This should be based on a careful history and include urinalysis, serum chemistry studies, and analysis of 24-hour urine collections. Measures to prevent recurrent stone formation are aimed at correcting the metabolic imbalances detected in the workup. A variety of drugs are available that target one or more of the metabolic abnormalities that may be involved. For "surgically active" renal and ureteral stone disease, newer techniques make surgery unnecessary in most cases. Extracorporeal shock wave lithotripsy is becoming the preferred technique for disintegration of upper urinary tract stones. Percutaneous ultrasonic lithotripsy and electrohydraulic disintegration also are widely used. For lower urinary tract stones, the ureteroscope permits either extraction under visualization or ultrasonic disintegration.

**URL:** <https://ovidsp.ovid.com/ovidweb.cgi?T=JS&CSC=Y&NEWS=N&PAGE=fulltext&D=med2&DO=10.1080%2f00325481.1985.11699183> <https://libkey.io/libraries/2592/openurl?genre=article&sid=OVID:medline&id=pmid:4059131&id=doi:10.1080%2F00325481.1985.11699183&issn=0032-5481&isbn=&volume=78&issue=6&spage=38&pages=38-44%2C+47-51&date=1985&title=Postgraduate+Medicine&atitle=Kidney+stones.+Medical+management+and+newer+options+for+stone+%27removal%27.&aulast=Kanig&pid=%3Cauthor%3EKanig+SP%3BConn+RL%3C%2Fauthor%3E%3CAN%3E4059131%3C%2FAN%3E%3CDT%3EJournal+Article%3C%2FDT%3E>

292. **Influence of dietary calcium on serum 1,25-dihydroxyvitamin D concentrations in renal stone formers.**

**Item Type:** Journal Article

**Authors:** D'Amour, P.;Gascon-Barre, M.;Dufresne, L. and Perreault, J. P.

**Publication Date:** No ,1984

**Journal:** Clinical Endocrinology 21(5), pp. 549–562

**Abstract:** The role of 1,25-dihydroxyvitamin D (1,25(OH)2D) in the pathogenesis of idiopathic hypercalciuria was studied in 37 renal stone formers who, during two 10-day periods, followed first a normal and then a low calcium diet. The following samples were taken during each diet; 24 h urine; fasting blood and urine; blood and urine following a 1 g oral calcium load. Patients were divided according to serum calcium level, 24 h urinary calcium excretion on the first diet and fasting calcium excretion on the second diet. Eight patients were found to be normocalciuric (NSF), 16 had absorptive hypercalciuria (AH), five renal hypercalciuria (RH) and eight primary hyperparathyroidism. In NSF and AH, a positive correlation was found between the fasting and the 24 hour urinary calcium (r = 0.787, P less than 0.001), while negative correlations were found between the fasting urinary calcium and the serum parathyroid hormone (r = -0.703, P less than 0.001) or the fasting urinary cyclic AMP (r = -0.434, P less than 0.01). Patients with RH had higher serum PTH and urinary cAMP levels for a given degree of fasting calciuria mainly on the low calcium diet. Mean serum 1,25(OH)2D was similar in NSF (43.6 +/- 4.5 pg/ml), AH (43.6 +/- 2.3 pg/ml) and RH (40.4 +/- 4.8 pg/ml) on the first diet; increases were similar in all groups after 10 d of calcium restriction. A positive correlation was found between the serum 1,25(OH)2D concentrations and the 24 h urinary calcium excretion on the first diet in NSF (r = 0.889, P less than 0.001) but not in AH or RH. There was no evidence of such correlation with the low calcium diet. No correlation between the calciuric response to calcium loading and the serum concentrations of 1,25(OH)2D was found. The results suggest that serum concentrations of 1,25(OH)2D may be related to urinary calcium excretion in NSF more than in AH or RH. The factors responsible for the hyperabsorption of calcium in the latter patients remain to be elucidated.

**URL:** <https://ovidsp.ovid.com/ovidweb.cgi?T=JS&CSC=Y&NEWS=N&PAGE=fulltext&D=med2&DO=10.1111%2fj.1365-2265.1984.tb01394.x> <https://libkey.io/libraries/2592/openurl?genre=article&sid=OVID:medline&id=pmid:6094046&id=doi:10.1111%2Fj.1365-2265.1984.tb01394.x&issn=0300-0664&isbn=&volume=21&issue=5&spage=549&pages=549-62&date=1984&title=Clinical+Endocrinology&atitle=Influence+of+dietary+calcium+on+serum+1%2C25-dihydroxyvitamin+D+concentrations+in+renal+stone+formers.&aulast=D%27Amour&pid=%3Cauthor%3ED%27Amour+P%3BGascon-Barre+M%3BDufresne+L%3BPerreault+JP%3C%2Fauthor%3E%3CAN%3E6094046%3C%2FAN%3E%3CDT%3EJournal+Article%3C%2FDT%3E>

293. **Potential pitfalls of the 2-hour calcium-to-creatinine ratio and urinary cyclic adenosine monophosphate excretion in the differential diagnosis of idiopathic hypercalciuria.**

**Item Type:** Journal Article

**Authors:** Heller, J. E.;Konnak, J. W. and Lau, Y. K.

**Publication Date:** Ma ,1984

**Journal:** Journal of Urology 131(5), pp. 911–913

**Abstract:** We studied 44 patients with calcium urolithiasis on high (900 mg. daily) and low (400 mg. daily) calcium diets. With 24-hour urinary data, we categorized the patients as normocalciuric or hypercalciuric and subdivided the hypercalciuric patients into absorptive and renal types. Abbreviated tests, including the 2-hour fasting urinary calcium-to-creatinine ratio and 24-hour urinary (nephrogenous) cyclic adenosine monophosphate, did not predict accurately whether hypercalciuria was of the idiopathic, absorptive or renal type. However, 24-hour urinary calcium excretions on the low calcium diet had a sensitivity and specificity of more than 90 per cent for reproducing the categorized diagnoses.

**URL:** <https://ovidsp.ovid.com/ovidweb.cgi?T=JS&CSC=Y&NEWS=N&PAGE=fulltext&D=med2&DO=10.1016%2fs0022-5347%2817%2950707-7> <https://libkey.io/libraries/2592/openurl?genre=article&sid=OVID:medline&id=pmid:6323773&id=doi:10.1016%2Fs0022-5347%2817%2950707-7&issn=0022-5347&isbn=&volume=131&issue=5&spage=911&pages=911-3&date=1984&title=Journal+of+Urology&atitle=Potential+pitfalls+of+the+2-hour+calcium-to-creatinine+ratio+and+urinary+cyclic+adenosine+monophosphate+excretion+in+the+differential+diagnosis+of+idiopathic+hypercalciuria.&aulast=Heller&pid=%3Cauthor%3EHeller+JE%3BKonnak+JW%3BLau+YK%3C%2Fauthor%3E%3CAN%3E6323773%3C%2FAN%3E%3CDT%3EJournal+Article%3C%2FDT%3E>

294. **Collection and handling of 24-hour urine specimens for measurement of analytes related to renal calculi.**

**Item Type:** Journal Article

**Authors:** Ng, R. H.;Menon, M. and Ladenson, J. H.

**Publication Date:** Ma ,1984

**Journal:** Clinical Chemistry 30(3), pp. 467–471

**Abstract:** Analysis for calcium, magnesium, phosphorus, oxalate, uric acid, and creatinine in 24-h urine collections is often needed for the differential diagnosis of patients with renal calculi. Considerable attention has been given to improving the methods of analysis, but improper sample collection and processing can cause significant errors for calcium and oxalate in urine samples not treated with acid and for uric acid in urine samples not treated with base. The errors are related to the concentration of the analyte, the interval the sample is stored before analysis, and the original pH of the urine sample. We describe here a system of sequential acidification (to pH 1.5) and alkalinization (to pH 9) of 24-h urine samples, followed by heating at 56 degrees C for 10 min. This procedure allows accurate analysis for all the above analytes in the same 24-h collection of urine. We validated the sample-treatment protocol for 80 24-h urine collections.

**URL:** <https://ovidsp.ovid.com/ovidweb.cgi?T=JS&CSC=Y&NEWS=N&PAGE=fulltext&D=med2&AN=6697501> <https://libkey.io/libraries/2592/openurl?genre=article&sid=OVID:medline&id=pmid:6697501&id=doi:&issn=0009-9147&isbn=&volume=30&issue=3&spage=467&pages=467-71&date=1984&title=Clinical+Chemistry&atitle=Collection+and+handling+of+24-hour+urine+specimens+for+measurement+of+analytes+related+to+renal+calculi.&aulast=Ng&pid=%3Cauthor%3ENg+RH%3BMenon+M%3BLadenson+JH%3C%2Fauthor%3E%3CAN%3E6697501%3C%2FAN%3E%3CDT%3EComparative+Study%3C%2FDT%3E>

295. **When should patients with symptomatic urinary stone disease be evaluated metabolically?.**

**Item Type:** Journal Article

**Authors:** Norman, R. W.;Bath, S. S.;Robertson, W. G. and Peacock, M.

**Publication Date:** De ,1984

**Journal:** Journal of Urology 132(6), pp. 1137–1139

**Abstract:** To determine the optimal time at which to assess the urinary risk factors for calcium stone formation, we followed 11 patients after an episode of acute renal colic with sequential 24-hour urine collections, first in the hospital and then at regular intervals after they were discharged from the hospital. The changes that occurred in the urinary risk factors were compared to those of a control group in which samples were collected during the same interval. The in-hospital 24-hour urine volumes were high but decreased gradually to approach the relatively constant volume of the control group by 3 months. The opposite trend occurred with respect to the 24-hour urinary excretion of calcium. There were no significant changes in the 24-hour urinary pH or excretions of oxalate, uric acid and alcian blue precipitable polyanions. The over-all effect was to cause a progressive increase in the probability of stones forming in the patients. The accurate assessment of the urinary risk factors of calcium stone disease requires at least a 3-month delay following acute renal colic. This delay usually will provide sufficient time for the stone to pass and for the patient to return to the normal dietary and fluid intake.

**URL:** <https://ovidsp.ovid.com/ovidweb.cgi?T=JS&CSC=Y&NEWS=N&PAGE=fulltext&D=med2&DO=10.1016%2fs0022-5347%2817%2950064-6> <https://libkey.io/libraries/2592/openurl?genre=article&sid=OVID:medline&id=pmid:6502804&id=doi:10.1016%2Fs0022-5347%2817%2950064-6&issn=0022-5347&isbn=&volume=132&issue=6&spage=1137&pages=1137-9&date=1984&title=Journal+of+Urology&atitle=When+should+patients+with+symptomatic+urinary+stone+disease+be+evaluated+metabolically%3F.&aulast=Norman&pid=%3Cauthor%3ENorman+RW%3BBath+SS%3BRobertson+WG%3BPeacock+M%3C%2Fauthor%3E%3CAN%3E6502804%3C%2FAN%3E%3CDT%3EJournal+Article%3C%2FDT%3E>

296. **Postprandial hyperoxaluria and intestinal oxalate absorption in idiopathic renal stone disease.**

**Item Type:** Journal Article

**Authors:** Schwille, P. O.;Hanisch, E. and Scholz, D.

**Publication Date:** Oc ,1984

**Journal:** Journal of Urology 132(4), pp. 650–655

**Abstract:** Calcium and oxalate were studied in daily, fasting and postprandial urine specimens from healthy subjects and patients with idiopathic renal calcium stones in response to a test meal free of oxalate, and supplemented with calcium and 14carbon-oxalic acid. The data showed that the amount of oxalate in fasting urine of patients with stones did not differ from that in controls. Generally, patients with stones had considerable postprandial hyperoxaluria in terms of excretion and concentration, associated with a significantly higher degree of supersaturation with regard to calcium oxalate compared to controls. These findings were paralleled by decreased intestinal absorption of 14carbon-oxalate and by unchanged 24-hour urinary oxalate. Although the source of increased postprandial oxalate in patients with stones is not clear the possibility of enhanced de novo synthesis from oxalate precursors is discussed. In patients with different types of calciuria the 2 main risk factors (hyperoxaluria and hypercalciuria) for the process of stone formation are recognizable more readily in the postprandial urine specimens than in fasting or daily urine specimens.

**URL:** <https://ovidsp.ovid.com/ovidweb.cgi?T=JS&CSC=Y&NEWS=N&PAGE=fulltext&D=med2&DO=10.1016%2fs0022-5347%2817%2949808-9> <https://libkey.io/libraries/2592/openurl?genre=article&sid=OVID:medline&id=pmid:6471206&id=doi:10.1016%2Fs0022-5347%2817%2949808-9&issn=0022-5347&isbn=&volume=132&issue=4&spage=650&pages=650-5&date=1984&title=Journal+of+Urology&atitle=Postprandial+hyperoxaluria+and+intestinal+oxalate+absorption+in+idiopathic+renal+stone+disease.&aulast=Schwille&pid=%3Cauthor%3ESchwille+PO%3BHanisch+E%3BScholz+D%3C%2Fauthor%3E%3CAN%3E6471206%3C%2FAN%3E%3CDT%3EJournal+Article%3C%2FDT%3E>

297. **The regional educational impact of a Renal Stone Center.**

**Item Type:** Journal Article

**Authors:** Jekel, J. F.;Chauncey, K. J.;Moore, N. L.;Broadus, A. E. and Gowdy, D. P.

**Publication Date:** 1983

**Journal:** Yale Journal of Biology & Medicine 56(2), pp. 97–108

**Abstract:** This study evaluates the impact of the Renal Stone Center (RSC) at Yale University School of Medicine on the extent of the diagnostic search for urolithiasis risk factors at the Yale-New Haven Medical Center and four affiliated community hospitals. Using a multiple time series design and a Physician Performance Index (PPI) based on multiple normative practice criteria, the hospital and outpatient charts of 1,924 patients with a primary diagnosis of urolithiasis at the five hospitals were examined to determine whether there was a change over time that could be related to the RSC. There was no statistically significant improvement in the PPI at four of the five hospitals. At the fifth, there was a statistically significant improvement that possibly was related to factors other than the RSC. Routine blood, urine, and X-ray tests usually were performed well. However, the history of urolithiasis risk factors and the examination of 24-hour urines were generally done poorly or not at all, and this generalization held true (although modified somewhat) even when physician office records were examined as well.

**URL:** <https://ovidsp.ovid.com/ovidweb.cgi?T=JS&CSC=Y&NEWS=N&PAGE=fulltext&D=med2&AN=6636839> <https://libkey.io/libraries/2592/openurl?genre=article&sid=OVID:medline&id=pmid:6636839&id=doi:&issn=0044-0086&isbn=&volume=56&issue=2&spage=97&pages=97-108&date=1983&title=Yale+Journal+of+Biology+%26+Medicine&atitle=The+regional+educational+impact+of+a+Renal+Stone+Center.&aulast=Jekel&pid=%3Cauthor%3EJekel+JF%3BChauncey+KJ%3BMoore+NL%3BBroadus+AE%3BGowdy+DP%3C%2Fauthor%3E%3CAN%3E6636839%3C%2FAN%3E%3CDT%3EJournal+Article%3C%2FDT%3E>

298. **Renal handling of urate in patients with calcium stone disease.**

**Item Type:** Journal Article

**Authors:** Fellstrom, B.;Backman, U.;Danielson, B. G.;Johansson, G.;Ljunghall, S. and Wikstrom, B.

**Publication Date:** 1982

**Journal:** Nephron 31(1), pp. 31–36

**Abstract:** The renal handling of urate was investigated in 28 recurrent calcium stone formers and 12 healthy controls. 12 patients were hyperuricosuric and 16 patients had incomplete proximal or distal acidification defects. Measurements of the glomerular filtration and the tubular reabsorption of filtered urate were made through the pyrazinamide (PZA)-suppression test of urate secretion. Hyperuricosuria could not be explained by defects in the renal handling of urate. Patients with proximal acidification defects had a higher tubular reabsorption of filtered urate than the other subjects. Tubular reabsorption of filtered urate was inversely correlated, and filtered urate escaping reabsorption positively correlated to the 24-hour excretion of urate. In contrast, the PZA-suppressible fraction of urate excretion, which is the net effect of tubular secretion and post-secretory reabsorption and thought to be the main regulator of urate excretion, was not correlated to the 24-hour excretion of urate. It is concluded that the renal handling of urate is basically normal in calcium stone disease, whereas minor deviations may co-exist with renal tubular acidification dysfunction.

**URL:** <https://ovidsp.ovid.com/ovidweb.cgi?T=JS&CSC=Y&NEWS=N&PAGE=fulltext&D=med2&DO=10.1159%2f000182610> <https://libkey.io/libraries/2592/openurl?genre=article&sid=OVID:medline&id=pmid:7110473&id=doi:10.1159%2F000182610&issn=1660-8151&isbn=&volume=31&issue=1&spage=31&pages=31-6&date=1982&title=Nephron&atitle=Renal+handling+of+urate+in+patients+with+calcium+stone+disease.&aulast=Fellstrom&pid=%3Cauthor%3EFellstrom+B%3BBackman+U%3BDanielson+BG%3BJohansson+G%3BLjunghall+S%3BWikstrom+B%3C%2Fauthor%3E%3CAN%3E7110473%3C%2FAN%3E%3CDT%3EJournal+Article%3C%2FDT%3E>

299. **Renal calculi associated with incomplete distal renal tubular acidosis.**

**Item Type:** Journal Article

**Authors:** Konnak, J. W.;Kogan, B. A. and Lau, K.

**Publication Date:** No ,1982

**Journal:** Journal of Urology 128(5), pp. 900–902

**Abstract:** We report 5 cases of renal calculi associated with incomplete type I renal tubular acidosis. The patients presented with a history of recurrent calculi and demonstrated bilateral calculi with corticomedullary junction calcification on x-ray. Stone analysis revealed primarily calcium phosphate stones and metabolic studies demonstrated hypercalciuria, mild depression of serum bicarbonate and inability to lower the pH of a fasting morning urine to less than 5.8. The 24-hour urinary citrate excretion was depressed markedly. The diagnosis was confirmed by an ammonium chloride loading test. Treatment consisted of alkalization of the patients, which corrects the hypercalciuria and increases the urinary citrate level and should decrease the tendency to form stones. The diagnosis of incomplete type I renal tubular acidosis should be considered in patients with recurrent renal calculi.

**URL:** <https://ovidsp.ovid.com/ovidweb.cgi?T=JS&CSC=Y&NEWS=N&PAGE=fulltext&D=med2&DO=10.1016%2fs0022-5347%2817%2953268-1> <https://libkey.io/libraries/2592/openurl?genre=article&sid=OVID:medline&id=pmid:7176046&id=doi:10.1016%2Fs0022-5347%2817%2953268-1&issn=0022-5347&isbn=&volume=128&issue=5&spage=900&pages=900-2&date=1982&title=Journal+of+Urology&atitle=Renal+calculi+associated+with+incomplete+distal+renal+tubular+acidosis.&aulast=Konnak&pid=%3Cauthor%3EKonnak+JW%3BKogan+BA%3BLau+K%3C%2Fauthor%3E%3CAN%3E7176046%3C%2FAN%3E%3CDT%3EJournal+Article%3C%2FDT%3E>

300. **Citrate in urine and serum and associated variables in subgroups of urolithiasis. Results from an outpatient stone clinic.**

**Item Type:** Journal Article

**Authors:** Schwille, P. O.;Scholz, D.;Schwille, K.;Leutschaft, R.;Goldberg, I. and Sigel, A.

**Publication Date:** 1982

**Journal:** Nephron 31(3), pp. 194–202

**Abstract:** Outpatient renal stone formers belonging to the established urolithiasis subgroups and controls were examined with respect to urinary and serum citrate (Cit) and several associated variables. Only in the normocalciuric majority of calcium and in uric acid stone formers was Cit in 24-hour urine decreased, but was normal in 2-hour fasting morning, and in 3-hour postprandial urine following a Cit-free test meal. Serum Cit was elevated in normocalciuria, renal and resorptive hypercalciuria. This Cit constellation was associated with either normal (absorptive, renal hypercalciuria) or low (normocalciuria, uric acid stone formers) parathyroid gland function as assessed by serum parathyroid hormone and nephrogenous urinary cyclic AMP, except in patients with primary hyperparathyroidism. In 2-hour morning urine the magnesium/creatinine ratio (normocalciuria) and ammonia excretion (uric acid stone formers) were decreased, while ammonia in 24-hour urine was low in all stone formers. It is suggested that Cit metabolism is altered in renal stone disease in general, and that in normocalciuria, stone inhibitors (Cit; magnesium) may be deficient.

**URL:** <https://ovidsp.ovid.com/ovidweb.cgi?T=JS&CSC=Y&NEWS=N&PAGE=fulltext&D=med2&DO=10.1159%2f000182646> <https://libkey.io/libraries/2592/openurl?genre=article&sid=OVID:medline&id=pmid:7121665&id=doi:10.1159%2F000182646&issn=1660-8151&isbn=&volume=31&issue=3&spage=194&pages=194-202&date=1982&title=Nephron&atitle=Citrate+in+urine+and+serum+and+associated+variables+in+subgroups+of+urolithiasis.+Results+from+an+outpatient+stone+clinic.&aulast=Schwille&pid=%3Cauthor%3ESchwille+PO%3BScholz+D%3BSchwille+K%3BLeutschaft+R%3BGoldberg+I%3BSigel+A%3C%2Fauthor%3E%3CAN%3E7121665%3C%2FAN%3E%3CDT%3EJournal+Article%3C%2FDT%3E>

301. **The stone-forming kidney: a study of functional differences between individual kidneys in idiopathic renal lithiasis.**

**Item Type:** Journal Article

**Authors:** Malek, R. S.;Wilkiemeyer, R. M. and Boyce, W. H.

**Publication Date:** Ju ,1976

**Journal:** Journal of Urology 116(1), pp. 11–14

**Abstract:** The 24-hour urinary excretion of various electrolytes, as well as the glomerular filtration rate, renal plasma flow and maximal rate of tubular excretion of para-aminohippuric acid was measured in each kidney of 10 patients with idiopathic renal lithiasis. Results in hypercalciuric patients indicate that, in contrast to earlier views, actively stone-forming, normally functioning kidneys are normocalciuric but hypophosphaturic in comparison to their mates with inactive or no stone disease, which are hypercalciuric but normophosphaturic. Therefore, hypophosphaturia rather than hypercalciuria may be an important calculogenic factor in some patients.

**URL:** <https://ovidsp.ovid.com/ovidweb.cgi?T=JS&CSC=Y&NEWS=N&PAGE=fulltext&D=med1&DO=10.1016%2fs0022-5347%2817%2958652-8> <https://libkey.io/libraries/2592/openurl?genre=article&sid=OVID:medline&id=pmid:933268&id=doi:10.1016%2Fs0022-5347%2817%2958652-8&issn=0022-5347&isbn=&volume=116&issue=1&spage=11&pages=11-4&date=1976&title=Journal+of+Urology&atitle=The+stone-forming+kidney%3A+a+study+of+functional+differences+between+individual+kidneys+in+idiopathic+renal+lithiasis.&aulast=Malek&pid=%3Cauthor%3EMalek+RS%3BWilkiemeyer+RM%3BBoyce+WH%3C%2Fauthor%3E%3CAN%3E933268%3C%2FAN%3E%3CDT%3EJournal+Article%3C%2FDT%3E>

302. **Excretion of inhibitors of calcification in urine. Part I. Findings in control subjects and patients with renal stones.**

**Item Type:** Journal Article

**Authors:** Oreopoulos, D. G.;Walker, D.;Akriotis, D. J.;Roncari, D. A.;Husdan, H.;Symvoulidis, A.;Deveber, G. A.;Rapoport, A. and Reid, D. B.

**Publication Date:** Apr 05 ,1975

**Journal:** Canadian Medical Association Journal 112(7), pp. 827–831

**Abstract:** The total excretion of inhibitors of in vitro calcification was measured (in inhibiting units per day) in 24-hour urine samples of 11 control subjects and 20 patients with renal calculi. A semiquantitative method incorporating the rachitic rat cartilage technique was used. In both groups there was a significant positive correlation between the number of inhibiting units per day and the daily urine volume. The mean number of inhibiting units per day was significantly (P smaller than 0.05) higher in the stone patients than in the controls. However, the stone-formers had significantly larger (P smaller than 0.01) 24-hour urine volumes. When corrections were made for urine volume there was no significant difference between the two groups. These data suggest that the underlying abnormality responsible for renal stone formation is not a persistent decrease in the total concentration of urinary inhibitors of calcification.

**URL:** <https://ovidsp.ovid.com/ovidweb.cgi?T=JS&CSC=Y&NEWS=N&PAGE=fulltext&D=med1&AN=1122457> <https://libkey.io/libraries/2592/openurl?genre=article&sid=OVID:medline&id=pmid:1122457&id=doi:&issn=0008-4409&isbn=&volume=112&issue=7&spage=827&pages=827-31&date=1975&title=Canadian+Medical+Association+Journal&atitle=Excretion+of+inhibitors+of+calcification+in+urine.+Part+I.+Findings+in+control+subjects+and+patients+with+renal+stones.&aulast=Oreopoulos&pid=%3Cauthor%3EOreopoulos+DG%3BWalker+D%3BAkriotis+DJ%3BRoncari+DA%3BHusdan+H%3BSymvoulidis+A%3BDeveber+GA%3BRapoport+A%3BReid+DB%3C%2Fauthor%3E%3CAN%3E1122457%3C%2FAN%3E%3CDT%3EJournal+Article%3C%2FDT%3E>

303. **The relationship of the urinary cations, Calcium, Magnesium, Sodium and Potassium, in patients with Renal Calculi.**

**Item Type:** Journal Article

**Authors:** Welshman, S. G. and McGeown, M. G.

**Publication Date:** Ju ,1975

**Journal:** British Journal of Urology 47(3), pp. 237–242

**Abstract:** Calcium, magnesium, sodium and potassium were estimated in the 24-hour urine collections of 101 idiopathic male stone-formers, 89 male patient controls and 59 young male adult controls. The results were calculated in terms of 24-hour volume and 1 g of creatinine. The concentrations of the 4 cations, relative to a gram of creatinine, were also determined in the early-morning urines of 41 male stone-formers and 13 young male adult controls. No difference was observed in the 24-hour excretion of calcium, magnesium and sodium between the stone-formers and controls. The mean daily potassium excretion, however, was significantly reduced in the urine of stone-formers. The linear regression equations were calculated for calcium on magnesium, calcium on sodium, and calcium on potassium, using the 24-hour excretion values of these cations. Only the calcium on potassium line of stone-formers was significantly different from that of the normal subjects. A significant increase by stone-formers in the urinary calcium concentration of their early-morning specimens was found. The high concentration of urinary calcium in overnight urines of stone-formers combined with a low magnesium concentration might possibly contribute to the development of renal stone disease.

**URL:** <https://ovidsp.ovid.com/ovidweb.cgi?T=JS&CSC=Y&NEWS=N&PAGE=fulltext&D=med1&DO=10.1111%2fj.1464-410x.1975.tb03956.x> <https://libkey.io/libraries/2592/openurl?genre=article&sid=OVID:medline&id=pmid:1139114&id=doi:10.1111%2Fj.1464-410x.1975.tb03956.x&issn=0007-1331&isbn=&volume=47&issue=3&spage=237&pages=237-42&date=1975&title=British+Journal+of+Urology&atitle=The+relationship+of+the+urinary+cations%2C+Calcium%2C+Magnesium%2C+Sodium+and+Potassium%2C+in+patients+with+Renal+Calculi.&aulast=Welshman&pid=%3Cauthor%3EWelshman+SG%3BMcGeown+MG%3C%2Fauthor%3E%3CAN%3E1139114%3C%2FAN%3E%3CDT%3EComparative+Study%3C%2FDT%3E>
