## Appendix 4 for "Investigating 24-Hour Urine Expected Ranges of Non-Kidney Stone Formers: an EAU Endourology Research Group Systematic Review and Meta-Analysis"

24 Hour Urine Ranges MA

R Geraghty

2026-06-23

Table of Contents

### 1 Setup

#### 1.1 Import Libraries

#### 1.2 Import data

```
phosphate_mw = 94.9714
phosphorous_mw = 30.97362

urine_ma <- fread("24_hr_urine_ma_12082025.csv", header = TRUE) %>% janitor::clean_names()

hpfs_nhs_I_II_summary <- fread("24_hr_urine_ma_nhs_hpfs_data.csv", header = TRUE) %>%
  janitor::clean_names() %>%
  mutate(
    phosphate_mean_mmol_24h = phosphorus_mean_mg_24h / phosphorous_mw,
    phosphate_sd_mmol_24h = phosphorus_sd_mg_24h / phosphorous_mw,
    .keep = "all"
  )

#these are excluded as phosphate/phosphorous definition is either unclear or unit conversion is not accurate
phosphate_to_exclude <- list("Bataille",  # Outlier - unclear if PO4 or P being used
                           "Huang", # Reported as phosphate (g/24 hr) but no methods defined, all other reference to phosphate / phosphorous is "P"
                           "Doizi", # Reported as phosphorous but methods use phosphate determination method
                           "Wabner", # Reported as phosphate (g/24 hr) but no methods defined
                           "Miyake", # Reported in phosphate mg/ml - not able to convert accurately
                           "Sweeney" # No methods of determination reported
                           )

phosphate_phosphorous <- fread("phosphate_phosphorous_updated_separated.csv", header = TRUE) %>%
  janitor::clean_names() %>%
  filter(!authors %in% phosphate_to_exclude) %>%
  filter(calculated_from_vol != "Y")
```

#### 1.3 Import and Sort Bias analysis

```
bias_observational <- fread("cochrane_risk_observational_studies.csv", header = TRUE) %>%
  janitor::clean_names()

bias_rcts <- fread("cochrane_risk_rct.csv", header = TRUE) %>%
  janitor::clean_names() %>%
    mutate(overall = case_when(overall_risk_of_bias_low_some_concern_high == "Some concern" ~ "Moderate",
                               overall_risk_of_bias_low_some_concern_high == "High" ~ "High",
                               overall_risk_of_bias_low_some_concern_high == "Low" ~ "Low",
                               TRUE ~ NA_character_),
           .keep = "unused")

overall_bias <- bias_observational %>%
  subset(select = c(study, overall)) %>%
  rbind(bias_rcts %>% subset(select = c(
    study, overall
  ))
  ) %>%
  as_tibble() %>%
  mutate(study_name = study,
         .keep = "unused") %>%
  drop_na(overall)

overall_bias$overall <- factor(overall_bias$overall,
                               levels = c("Low",
                                          "Moderate",
                                          "High"))
```

#### 1.4 Sort data

##### 1.4.1 Phosphate data

```
phosphate_phosphorous <- phosphate_phosphorous %>%
  mutate(
    phosphate_mean_mmol_24h = case_when(
      !is.na(phosphate_mean_mmol_24h) ~ phosphate_mean_mmol_24h,
      !is.na(phosphate_mean_mg_24h) ~ phosphate_mean_mg_24h / phosphate_mw,
      !is.na(phosphate_mean_g_24h) ~ (phosphate_mean_g_24h / phosphate_mw) * 1000,
      !is.na(phosphorus_mean_mg_24h) ~ phosphorus_mean_mg_24h / phosphorous_mw
    ),
    phosphate_sd_mmol_24h = case_when(
      !is.na(phosphate_sd_mmol_24h) ~ phosphate_sd_mmol_24h,
      !is.na(phosphate_sd_mg_24h) ~ phosphate_sd_mg_24h / phosphate_mw,
      !is.na(phosphate_sd_g_24h) ~ (phosphate_sd_g_24h / phosphate_mw) * 1000,
      !is.na(phosphorus_sd_mg_24h) ~ phosphorus_sd_mg_24h / phosphorous_mw
    ),
    .keep = "unused"
    ) %>%
  dplyr::select(
    authors,
    year,
    phosphate_mean_mmol_24h,
    phosphate_sd_mmol_24h)
```

##### 1.4.2 MA data

```
hpfs_nhs_I_II_summary1 <- hpfs_nhs_I_II_summary %>%
  subset(
    select = -c(
      assay,
      comments,
      phosphate_mean_mmol_24h,
      phosphate_sd_mmol_24h,
      phosphorus_mean_mg_24h,
      phosphorus_sd_mg_24h
    )
  )

hpfs_nhs_I_II_summary2 <- hpfs_nhs_I_II_summary %>%
  subset(
    select = c(
      phosphate_mean_mmol_24h,
      phosphate_sd_mmol_24h,
      phosphorus_mean_mg_24h,
      phosphorus_sd_mg_24h
    )
  )

hpfs_nhs_I_II_summary <- hpfs_nhs_I_II_summary1 %>%
  cbind(hpfs_nhs_I_II_summary2) %>%
  dplyr::select(-c(phosphorus_mean_mg_24h, phosphorus_sd_mg_24h))

urine_ma1 <- urine_ma %>%
  select(-starts_with("x")) %>%
  subset(select = -c(phosphate_mean_mmol_24h, phosphate_sd_mmol_24h)) %>%
  mutate(join_key = paste0(first_author, year)) %>%
  left_join(
    phosphate_phosphorous %>% mutate(join_key = paste0(authors, year),
                                     .keep = "unused"),
    by = "join_key"
  ) %>%
  select(-c(join_key, comments, ok_measurement)) %>%
  rbind(hpfs_nhs_I_II_summary) %>%
  cbind(urine_ma %>% select(ok_measurement))


urine_ma1$age_sd <- as.numeric(urine_ma1$age_sd)
urine_ma1$child_adult <- as.factor(urine_ma1$child_adult)
urine_ma1$underlying_cause <- as.factor(urine_ma1$underlying_cause)
urine_ma1$stone_vs_no_stone <- as.factor(urine_ma1$stone_vs_no_stone)
urine_ma1$ok_measurement <- as.factor(urine_ma1$ok_measurement)
urine_ma1$male_n <- as.integer(urine_ma1$male_n)
urine_ma1$female_n <- as.integer(urine_ma1$female_n)
urine_ma1$ph_mean <- as.numeric(urine_ma1$ph_mean)
urine_ma1$ph_sd <- as.numeric(urine_ma1$ph_sd)
urine_ma1$volume_mean_m_l <- as.numeric(urine_ma1$volume_mean_m_l)
urine_ma1$total_n <- as.integer(urine_ma1$total_n)
urine_ma1$restricted_diet <- as.factor(urine_ma1$restricted_diet)
urine_ma1$other_demographic_variant <- as.factor(urine_ma1$other_demographic_variant)

urine_ma1 <- urine_ma1 %>% mutate(
  urea_mean_mmol_24h = ifelse(urea_mean_mmol_24h < 1,
                              urea_mean_mmol_24h * 1000,
                              urea_mean_mmol_24h),
  urea_sd_mmol_24h = ifelse(urea_sd_mmol_24h < 1,
                              urea_sd_mmol_24h * 1000,
                              urea_sd_mmol_24h),
)

urine_ma1 <- urine_ma1 %>% mutate(
  study_name = case_when(
    first_author == "HPFS & NHS I/II" ~ first_author,
    and == "no" & first_author != "HPFS & NHS I/II" ~ paste0(first_author, " et al. ", year),
    and == "yes" ~ paste0(first_author, ", ", year),
  TRUE ~ paste0(first_author, " et al. ", year)),
  underlying_cause = case_when(
        other_demographic_variant == "pre_menopausal" ~ "pre_menopausal",
        other_demographic_variant == "post_menopausal" ~ "post_menopausal",
        TRUE ~ other_demographic_variant
  ),
  other_demographic_variant = case_when(
    other_demographic_variant == "black" ~ "black",
    other_demographic_variant == "white" ~ "white",
    other_demographic_variant == "no" ~ "mixed",
    other_demographic_variant == "pre_menopausal" ~ "mixed",
    other_demographic_variant == "post_menopausal" ~ "mixed"
  ),
  .keep = "all"
)

urine_ma1$other_demographic_variant <- as.factor(urine_ma1$other_demographic_variant)

urine_ma2 <- urine_ma1 %>%
  filter(
    stone_vs_no_stone == "no_stone") %>%
  filter(
      (underlying_cause == "no" |
        underlying_cause == "pre_menopausal")) %>%
  filter(!first_author %in% c("Prochaska", "Ferraro", "Curhan", "Taylor and Curhan")) %>%
  drop_na(total_n)

urine_ma2 <- urine_ma2 %>%
  mutate(
    age_band = case_when(
      age_band == "20-29" | age_band == "20-30" ~ "20-30",
      age_band == "30-39" | age_band == "30-40" ~ "30-40",
      age_band == "40-49" | age_band == "40-50" ~ "40-50",
      age_band == "50-59" | age_band == "50-60" ~ "50-60",
      age_band == "60-70" | age_band == "60+" | age_band == "70-80" ~ "60+",
      TRUE ~ age_band
    ),
    study_type = case_when(
      study_name %in% bias_observational$study ~ "Observational Cohort",
      study_name %in% bias_rcts$study ~ "RCT",
      TRUE ~ "Observational Cohort"
    )
  )

urine_ma2$race <- as.factor(urine_ma2$race)
urine_ma2$male_female <- as.factor(urine_ma2$male_female)
urine_ma2$age_band <- as.factor(urine_ma2$age_band)

study_types <- urine_ma2 %>% dplyr::select(study_name, study_type)

# -----------------------------------------------------------------------------
# A5 — Assay-method lookup (per study × component)
# -----------------------------------------------------------------------------
# Generated by `assay_extraction.R` from `24_hr_urine_ma_12082025_2.xlsx` Rob
# sheet, column `ascertainment_of_measurements`. Free text was parsed into
# broad assay categories (atomic absorption / autoanalyser / ion chromatography
# / etc.) per analyte. Studies known to use chemical preservatives (Litholink,
# Mission Pharmacal, Labcorp, acidification with HCl) are flagged.
study_assay_lookup <- tryCatch({
  readRDS("shiny_app_data/study_assay_lookup.rds")
}, error = function(e) {
  message("study_assay_lookup.rds not found — run assay_extraction.R first.")
  NULL
})

# Helper to merge assay categories into a per-component analysis dataset.
# `data` should be the standard urine_ma2-style data; `urine_component` is
# one of "calcium", "oxalate", ... matched to the assay lookup component.
add_assay_column <- function(data, urine_component) {
  if (is.null(study_assay_lookup)) {
    data$assay_cat <- NA_character_
    data$preservative_used <- NA
    return(data)
  }
  comp_key <- if (urine_component == "creat") "creatinine" else urine_component
  comp_lookup <- study_assay_lookup %>%
    dplyr::filter(component == comp_key) %>%
    dplyr::select(study_name, assay_cat, preservative_used)
  dplyr::left_join(data, comp_lookup, by = "study_name") %>%
    dplyr::mutate(
      assay_cat = ifelse(is.na(assay_cat), "Unreported", assay_cat),
      preservative_used = ifelse(is.na(preservative_used), FALSE, preservative_used)
    )
}
```

### 2 Explore MA data

#### 2.1 Plot Missing

plot_missing(urine_ma2)

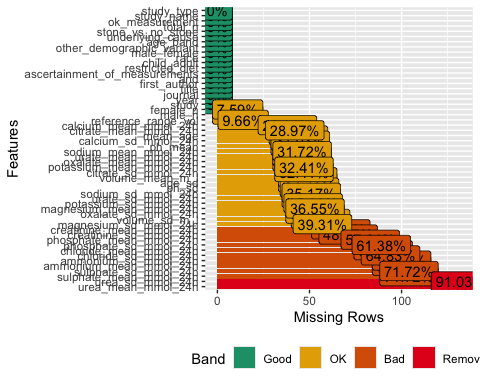

#### 2.2 Number of Papers in MA

```
urine_ma2 %>%
  summarise(
    n = n_distinct(first_author)
  ) %>%
  gt() %>%
  tab_header(
    title = md("**Number of Studies**")
  ) %>%
  tab_options(
    table.border.top.color = "white",
    table.border.bottom.color = "white",
    data_row.padding = px(8),
    table.font.size = 14
  ) %>%
  cols_align(
    align = "center",
    columns = everything()
  )
```

Table 1: **Number of Studies**

n

67

#### 2.3 Table of Papers Included in Review

```
urine_ma1 %>%
  arrange(year) %>%
  drop_na(year) %>%
  mutate(first_author = case_when(
    and == "no" ~ paste0(first_author, ", ", year),
    first_author == "HPFS" | first_author == "NHS" ~ first_author,
    TRUE ~ paste0(first_author, " et al. ", year)
  )) %>%
  group_by(first_author) %>%
  select(first_author,
         stone_vs_no_stone,
         underlying_cause,
         other_demographic_variant,
         mean_age,
         age_sd,
         total_n,
         male_n,
         female_n) %>%
  gt() %>%
  gt_theme_espn() %>%
  tab_header(
    title = md("**Summary of Included Full Texts**")
  ) %>%
  cols_align(
    align = "center",
    columns = everything()
  ) %>%
  tab_style(
    style = cell_text(weight = "bold"),
    locations = cells_column_labels(everything())
  ) %>%
  tab_options(
    table.border.top.width = px(2),
    table.border.bottom.width = px(2),
    table.font.size = px(14),
    data_row.padding = px(10)
  )
```

Table 1: **Summary of Included Full Texts**

| stone_vs_no_stone | underlying_cause | other_demographic_variant | mean_age | age_sd | total_n | male_n | female_n |
| --- | --- | --- | --- | --- | --- | --- | --- |
| Goldschmied, 1975 | | | | | | | |
| no_stone | no | mixed | NA | NA | 55 | 28 | 27 |
| Welshman, 1975 | | | | | | | |
| no_stone | no | mixed | NA | NA | 89 | 89 | 0 |
| no_stone | no | mixed | NA | NA | 89 | 89 | 0 |
| Welshman, 1976 | | | | | | | |
| no_stone | no | mixed | NA | NA | 158 | 73 | 85 |
| Marya, 1979 | | | | | | | |
| no_stone | no | mixed | NA | NA | 50 | NA | NA |
| Stern, 1980 | | | | | | | |
| no_stone | no | mixed | NA | NA | 9 | NA | NA |
| Hesse, 1986 | | | | | | | |
| no_stone | no | mixed | NA | NA | 25 | 0 | 25 |
| Hesse et al. 1986 | | | | | | | |
| no_stone | no | mixed | NA | NA | 25 | 0 | 25 |
| no_stone | no | mixed | NA | NA | 25 | 0 | 25 |
| no_stone | no | mixed | NA | NA | 25 | 0 | 25 |
| no_stone | no | mixed | NA | NA | 25 | 0 | 25 |
| no_stone | no | mixed | NA | NA | 25 | 25 | 0 |
| no_stone | no | mixed | NA | NA | 25 | 25 | 0 |
| no_stone | no | mixed | NA | NA | 25 | 25 | 0 |
| no_stone | no | mixed | NA | NA | 25 | 25 | 0 |
| no_stone | no | mixed | NA | NA | 25 | 25 | 0 |
| Bingham, 1988 | | | | | | | |
| no_stone | no | mixed | NA | NA | 8 | 5 | 3 |
| Michelacci, 1989 | | | | | | | |
| no_stone | no | mixed | NA | NA | 23 | 11 | 12 |
| Bataille, 1991 | | | | | | | |
| no_stone | no | mixed | 35.00 | 5.000 | 62 | 41 | 21 |
| Wabner, 1992 | | | | | | | |
| no_stone | no | mixed | 38.00 | 8.600 | 12 | 0 | 12 |
| Erwin, 1994 | | | | | | | |
| no_stone | no | mixed | 40.00 | 1.500 | 22 | 16 | 6 |
| Buchholz, 1996 | | | | | | | |
| no_stone | no | mixed | 34.00 | 10.800 | 37 | 37 | 0 |
| Christie, 1996 | | | | | | | |
| no_stone | no | mixed | 39.00 | 3.000 | 17 | 7 | 10 |
| Lemann, 1996 | | | | | | | |
| no_stone | no | mixed | 43.90 | 10.200 | 94 | 44 | 50 |
| Rodgers, 1997 | | | | | | | |
| no_stone | no | mixed | NA | NA | 40 | 20 | 20 |
| Hess, 1999 | | | | | | | |
| no_stone | no | mixed | 39.00 | 1.300 | 107 | 107 | 0 |
| Matin and Streem et al. 2000 | | | | | | | |
| no_stone | no | mixed | NA | NA | 26 | NA | NA |
| no_stone | no | mixed | NA | NA | NA | NA | NA |
| no_stone | no | mixed | NA | NA | NA | NA | NA |
| no_stone | no | mixed | NA | NA | 26 | 9 | 17 |
| Holmes, 2001 | | | | | | | |
| no_stone | no | mixed | 29.00 | 4.000 | 12 | 6 | 6 |
| Miyake, 2001 | | | | | | | |
| no_stone | no | mixed | 31.50 | 4.400 | 11 | 11 | 0 |
| Terris, 2001 | | | | | | | |
| no_stone | no | mixed | 30.20 | NA | 5 | 3 | 2 |
| Curhan, 2001 | | | | | | | |
| no_stone | no | mixed | 61.90 | 6.200 | 99 | NA | NA |
| no_stone | no | mixed | 61.90 | 6.200 | 99 | NA | NA |
| no_stone | no | mixed | 61.90 | 6.200 | 99 | NA | NA |
| no_stone | no | mixed | 42.90 | 3.700 | 30 | NA | NA |
| no_stone | no | mixed | 42.90 | 3.700 | 30 | NA | NA |
| no_stone | no | mixed | 42.90 | 3.700 | 30 | NA | NA |
| no_stone | no | mixed | 60.80 | 6.600 | 110 | NA | NA |
| no_stone | no | mixed | 60.80 | 6.600 | 110 | NA | NA |
| no_stone | no | mixed | 60.80 | 6.600 | 110 | NA | NA |
| Rodgers, 2002 | | | | | | | |
| no_stone | no | mixed | NA | NA | 10 | 10 | 0 |
| no_stone | no | mixed | NA | NA | 10 | 10 | 0 |
| no_stone | no | mixed | NA | NA | 10 | 10 | 0 |
| no_stone | no | mixed | NA | NA | 10 | 10 | 0 |
| Kinder, 2002 | | | | | | | |
| no_stone | no | mixed | NA | NA | 111 | NA | 111 |
| no_stone | no | mixed | NA | NA | 111 | NA | 111 |
| no_stone | no | mixed | NA | NA | 66 | 66 | NA |
| no_stone | no | mixed | NA | NA | 66 | 66 | NA |
| Allie, 2003 | | | | | | | |
| no_stone | no | mixed | NA | NA | 8 | 8 | 0 |
| Baxmann, 2003 | | | | | | | |
| no_stone | no | mixed | 37.00 | 16.000 | 20 | 8 | 12 |
| Huang, 2003 | | | | | | | |
| no_stone | no | mixed | 50.80 | 1.800 | 32 | 27 | 5 |
| Kuo, 2003 | | | | | | | |
| no_stone | no | mixed | NA | NA | 4 | NA | NA |
| Ogawa, 2003 | | | | | | | |
| no_stone | no | mixed | 54.10 | 16.200 | 22 | 6 | 16 |
| Traxer, 2003 | | | | | | | |
| no_stone | no | mixed | 38.10 | 11.300 | 12 | 6 | 6 |
| Abate, 2004 | | | | | | | |
| no_stone | no | mixed | 31.00 | 11.000 | 55 | 33 | 22 |
| Chai, 2004 | | | | | | | |
| no_stone | no | mixed | NA | NA | 9 | 5 | 4 |
| Lewandowski, 2004 | | | | | | | |
| no_stone | black | black | 23.50 | 2.700 | 10 | 10 | 0 |
| no_stone | black | black | 23.50 | 2.700 | 10 | 10 | 0 |
| no_stone | white | white | 21.60 | 1.600 | 10 | 10 | 0 |
| no_stone | white | white | 21.60 | 1.600 | 10 | 10 | 0 |
| Stitchantrakul, 2004 | | | | | | | |
| no_stone | no | mixed | 22.70 | 1.900 | 38 | 38 | 0 |
| Siener, 2004 | | | | | | | |
| no_stone | male | NA | 26.50 | NA | 12 | 12 | 0 |
| Defoor, 2005 | | | | | | | |
| no_stone | no | mixed | NA | NA | 168 | 97 | 71 |
| Gettman, 2005 | | | | | | | |
| no_stone | no | mixed | 34.80 | 9.600 | 12 | 6 | 6 |
| Shah, 2005 | | | | | | | |
| no_stone | no | mixed | 30.00 | NA | 101 | 54 | 47 |
| no_stone | no | mixed | 30.00 | NA | 101 | 54 | 47 |
| Shah et al. 2005 | | | | | | | |
| no_stone | no | mixed | 30.00 | 5.000 | 61 | 12 | 49 |
| no_stone | no | mixed | 30.00 | 5.000 | 61 | 12 | 49 |
| Cameron, 2006 | | | | | | | |
| no_stone | no | mixed | 49.00 | 8.000 | 59 | 24 | 35 |
| no_stone | no | mixed | 49.00 | 8.000 | 59 | 24 | 35 |
| no_stone | no | mixed | 52.00 | 8.000 | 24 | 10 | 14 |
| no_stone | no | mixed | 52.00 | 8.000 | 24 | 10 | 14 |
| Matsumoto, 2006 | | | | | | | |
| no_stone | no | mixed | NA | NA | 10 | 5 | 5 |
| Taylor and Curhan et al. 2006 | | | | | | | |
| no_stone | no | mixed | NA | NA | 76 | 76 | NA |
| no_stone | no | mixed | NA | NA | 76 | 76 | NA |
| no_stone | no | mixed | NA | NA | 76 | 76 | NA |
| no_stone | no | mixed | NA | NA | 76 | 76 | NA |
| no_stone | no | mixed | NA | NA | 76 | 76 | NA |
| no_stone | no | mixed | NA | NA | 76 | 76 | NA |
| no_stone | no | mixed | NA | NA | 76 | 76 | NA |
| no_stone | no | mixed | NA | NA | 76 | 76 | NA |
| no_stone | no | mixed | NA | NA | 76 | 76 | NA |
| no_stone | no | mixed | NA | NA | 76 | 76 | NA |
| no_stone | no | mixed | NA | NA | 76 | 76 | NA |
| no_stone | no | mixed | NA | NA | 76 | 76 | NA |
| no_stone | no | mixed | NA | NA | 76 | 76 | NA |
| no_stone | no | mixed | NA | NA | 76 | 76 | NA |
| no_stone | no | mixed | NA | NA | 76 | 76 | NA |
| no_stone | no | mixed | NA | NA | 80 | 80 | NA |
| no_stone | no | mixed | NA | NA | 80 | 80 | NA |
| no_stone | no | mixed | NA | NA | 80 | 80 | NA |
| no_stone | no | mixed | NA | NA | 80 | 80 | NA |
| no_stone | no | mixed | NA | NA | 80 | 80 | NA |
| no_stone | no | mixed | NA | NA | 80 | 80 | NA |
| no_stone | no | mixed | NA | NA | 80 | 80 | NA |
| no_stone | no | mixed | NA | NA | 80 | 80 | NA |
| no_stone | no | mixed | NA | NA | 80 | 80 | NA |
| no_stone | no | mixed | NA | NA | 80 | 80 | NA |
| no_stone | no | mixed | NA | NA | 80 | 80 | NA |
| no_stone | no | mixed | NA | NA | 80 | 80 | NA |
| no_stone | no | mixed | NA | NA | 80 | 80 | NA |
| no_stone | no | mixed | NA | NA | 80 | 80 | NA |
| no_stone | no | mixed | NA | NA | 80 | 80 | NA |
| no_stone | no | mixed | NA | NA | 84 | 84 | NA |
| no_stone | no | mixed | NA | NA | 84 | 84 | NA |
| no_stone | no | mixed | NA | NA | 84 | 84 | NA |
| no_stone | no | mixed | NA | NA | 84 | 84 | NA |
| no_stone | no | mixed | NA | NA | 84 | 84 | NA |
| no_stone | no | mixed | NA | NA | 84 | 84 | NA |
| no_stone | no | mixed | NA | NA | 84 | 84 | NA |
| no_stone | no | mixed | NA | NA | 84 | 84 | NA |
| no_stone | no | mixed | NA | NA | 84 | 84 | NA |
| no_stone | no | mixed | NA | NA | 84 | 84 | NA |
| no_stone | no | mixed | NA | NA | 84 | 84 | NA |
| no_stone | no | mixed | NA | NA | 84 | 84 | NA |
| no_stone | no | mixed | NA | NA | 84 | 84 | NA |
| no_stone | no | mixed | NA | NA | 84 | 84 | NA |
| no_stone | no | mixed | NA | NA | 84 | 84 | NA |
| no_stone | no | mixed | NA | NA | 90 | 90 | NA |
| no_stone | no | mixed | NA | NA | 90 | 90 | NA |
| no_stone | no | mixed | NA | NA | 90 | 90 | NA |
| no_stone | no | mixed | NA | NA | 90 | 90 | NA |
| no_stone | no | mixed | NA | NA | 90 | 90 | NA |
| no_stone | no | mixed | NA | NA | 90 | 90 | NA |
| no_stone | no | mixed | NA | NA | 90 | 90 | NA |
| no_stone | no | mixed | NA | NA | 90 | 90 | NA |
| no_stone | no | mixed | NA | NA | 90 | 90 | NA |
| no_stone | no | mixed | NA | NA | 90 | 90 | NA |
| no_stone | no | mixed | NA | NA | 90 | 90 | NA |
| no_stone | no | mixed | NA | NA | 90 | 90 | NA |
| no_stone | no | mixed | NA | NA | 90 | 90 | NA |
| no_stone | no | mixed | NA | NA | 90 | 90 | NA |
| no_stone | no | mixed | NA | NA | 90 | 90 | NA |
| no_stone | no | mixed | NA | NA | 74 | 74 | NA |
| no_stone | no | mixed | NA | NA | 74 | 74 | NA |
| no_stone | no | mixed | NA | NA | 74 | 74 | NA |
| no_stone | no | mixed | NA | NA | 74 | 74 | NA |
| no_stone | no | mixed | NA | NA | 74 | 74 | NA |
| no_stone | no | mixed | NA | NA | 74 | 74 | NA |
| no_stone | no | mixed | NA | NA | 74 | 74 | NA |
| no_stone | no | mixed | NA | NA | 74 | 74 | NA |
| no_stone | no | mixed | NA | NA | 74 | 74 | NA |
| no_stone | no | mixed | NA | NA | 74 | 74 | NA |
| no_stone | no | mixed | NA | NA | 74 | 74 | NA |
| no_stone | no | mixed | NA | NA | 74 | 74 | NA |
| no_stone | no | mixed | NA | NA | 74 | 74 | NA |
| no_stone | no | mixed | NA | NA | 74 | 74 | NA |
| no_stone | no | mixed | NA | NA | 74 | 74 | NA |
| no_stone | no | mixed | NA | NA | 111 | NA | 111 |
| no_stone | no | mixed | NA | NA | 111 | NA | 111 |
| no_stone | no | mixed | NA | NA | 111 | NA | 111 |
| no_stone | no | mixed | NA | NA | 111 | NA | 111 |
| no_stone | no | mixed | NA | NA | 111 | NA | 111 |
| no_stone | no | mixed | NA | NA | 111 | NA | 111 |
| no_stone | no | mixed | NA | NA | 111 | NA | 111 |
| no_stone | no | mixed | NA | NA | 111 | NA | 111 |
| no_stone | no | mixed | NA | NA | 111 | NA | 111 |
| no_stone | no | mixed | NA | NA | 111 | NA | 111 |
| no_stone | no | mixed | NA | NA | 111 | NA | 111 |
| no_stone | no | mixed | NA | NA | 111 | NA | 111 |
| no_stone | no | mixed | NA | NA | 111 | NA | 111 |
| no_stone | no | mixed | NA | NA | 111 | NA | 111 |
| no_stone | no | mixed | NA | NA | 111 | NA | 111 |
| no_stone | no | mixed | NA | NA | 90 | NA | 90 |
| no_stone | no | mixed | NA | NA | 90 | NA | 90 |
| no_stone | no | mixed | NA | NA | 90 | NA | 90 |
| no_stone | no | mixed | NA | NA | 90 | NA | 90 |
| no_stone | no | mixed | NA | NA | 90 | NA | 90 |
| no_stone | no | mixed | NA | NA | 90 | NA | 90 |
| no_stone | no | mixed | NA | NA | 90 | NA | 90 |
| no_stone | no | mixed | NA | NA | 90 | NA | 90 |
| no_stone | no | mixed | NA | NA | 90 | NA | 90 |
| no_stone | no | mixed | NA | NA | 90 | NA | 90 |
| no_stone | no | mixed | NA | NA | 90 | NA | 90 |
| no_stone | no | mixed | NA | NA | 90 | NA | 90 |
| no_stone | no | mixed | NA | NA | 90 | NA | 90 |
| no_stone | no | mixed | NA | NA | 90 | NA | 90 |
| no_stone | no | mixed | NA | NA | 90 | NA | 90 |
| no_stone | no | mixed | NA | NA | 75 | NA | 75 |
| no_stone | no | mixed | NA | NA | 75 | NA | 75 |
| no_stone | no | mixed | NA | NA | 75 | NA | 75 |
| no_stone | no | mixed | NA | NA | 75 | NA | 75 |
| no_stone | no | mixed | NA | NA | 75 | NA | 75 |
| no_stone | no | mixed | NA | NA | 75 | NA | 75 |
| no_stone | no | mixed | NA | NA | 75 | NA | 75 |
| no_stone | no | mixed | NA | NA | 75 | NA | 75 |
| no_stone | no | mixed | NA | NA | 75 | NA | 75 |
| no_stone | no | mixed | NA | NA | 75 | NA | 75 |
| no_stone | no | mixed | NA | NA | 75 | NA | 75 |
| no_stone | no | mixed | NA | NA | 75 | NA | 75 |
| no_stone | no | mixed | NA | NA | 75 | NA | 75 |
| no_stone | no | mixed | NA | NA | 75 | NA | 75 |
| no_stone | no | mixed | NA | NA | 75 | NA | 75 |
| no_stone | no | mixed | NA | NA | 68 | NA | 68 |
| no_stone | no | mixed | NA | NA | 68 | NA | 68 |
| no_stone | no | mixed | NA | NA | 68 | NA | 68 |
| no_stone | no | mixed | NA | NA | 68 | NA | 68 |
| no_stone | no | mixed | NA | NA | 68 | NA | 68 |
| no_stone | no | mixed | NA | NA | 68 | NA | 68 |
| no_stone | no | mixed | NA | NA | 68 | NA | 68 |
| no_stone | no | mixed | NA | NA | 68 | NA | 68 |
| no_stone | no | mixed | NA | NA | 68 | NA | 68 |
| no_stone | no | mixed | NA | NA | 68 | NA | 68 |
| no_stone | no | mixed | NA | NA | 68 | NA | 68 |
| no_stone | no | mixed | NA | NA | 68 | NA | 68 |
| no_stone | no | mixed | NA | NA | 68 | NA | 68 |
| no_stone | no | mixed | NA | NA | 68 | NA | 68 |
| no_stone | no | mixed | NA | NA | 68 | NA | 68 |
| no_stone | no | mixed | NA | NA | 54 | NA | 54 |
| no_stone | no | mixed | NA | NA | 54 | NA | 54 |
| no_stone | no | mixed | NA | NA | 54 | NA | 54 |
| no_stone | no | mixed | NA | NA | 54 | NA | 54 |
| no_stone | no | mixed | NA | NA | 54 | NA | 54 |
| no_stone | no | mixed | NA | NA | 54 | NA | 54 |
| no_stone | no | mixed | NA | NA | 54 | NA | 54 |
| no_stone | no | mixed | NA | NA | 54 | NA | 54 |
| no_stone | no | mixed | NA | NA | 54 | NA | 54 |
| no_stone | no | mixed | NA | NA | 54 | NA | 54 |
| no_stone | no | mixed | NA | NA | 54 | NA | 54 |
| no_stone | no | mixed | NA | NA | 54 | NA | 54 |
| no_stone | no | mixed | NA | NA | 54 | NA | 54 |
| no_stone | no | mixed | NA | NA | 54 | NA | 54 |
| no_stone | no | mixed | NA | NA | 54 | NA | 54 |
| no_stone | no | mixed | NA | NA | 62 | NA | 62 |
| no_stone | no | mixed | NA | NA | 62 | NA | 62 |
| no_stone | no | mixed | NA | NA | 62 | NA | 62 |
| no_stone | no | mixed | NA | NA | 62 | NA | 62 |
| no_stone | no | mixed | NA | NA | 62 | NA | 62 |
| no_stone | no | mixed | NA | NA | 62 | NA | 62 |
| no_stone | no | mixed | NA | NA | 62 | NA | 62 |
| no_stone | no | mixed | NA | NA | 62 | NA | 62 |
| no_stone | no | mixed | NA | NA | 62 | NA | 62 |
| no_stone | no | mixed | NA | NA | 62 | NA | 62 |
| no_stone | no | mixed | NA | NA | 62 | NA | 62 |
| no_stone | no | mixed | NA | NA | 62 | NA | 62 |
| no_stone | no | mixed | NA | NA | 62 | NA | 62 |
| no_stone | no | mixed | NA | NA | 62 | NA | 62 |
| no_stone | no | mixed | NA | NA | 62 | NA | 62 |
| no_stone | no | mixed | NA | NA | 60 | NA | 60 |
| no_stone | no | mixed | NA | NA | 60 | NA | 60 |
| no_stone | no | mixed | NA | NA | 60 | NA | 60 |
| no_stone | no | mixed | NA | NA | 60 | NA | 60 |
| no_stone | no | mixed | NA | NA | 60 | NA | 60 |
| no_stone | no | mixed | NA | NA | 60 | NA | 60 |
| no_stone | no | mixed | NA | NA | 60 | NA | 60 |
| no_stone | no | mixed | NA | NA | 60 | NA | 60 |
| no_stone | no | mixed | NA | NA | 60 | NA | 60 |
| no_stone | no | mixed | NA | NA | 60 | NA | 60 |
| no_stone | no | mixed | NA | NA | 60 | NA | 60 |
| no_stone | no | mixed | NA | NA | 60 | NA | 60 |
| no_stone | no | mixed | NA | NA | 60 | NA | 60 |
| no_stone | no | mixed | NA | NA | 60 | NA | 60 |
| no_stone | no | mixed | NA | NA | 60 | NA | 60 |
| no_stone | no | mixed | NA | NA | 74 | NA | 74 |
| no_stone | no | mixed | NA | NA | 74 | NA | 74 |
| no_stone | no | mixed | NA | NA | 74 | NA | 74 |
| no_stone | no | mixed | NA | NA | 74 | NA | 74 |
| no_stone | no | mixed | NA | NA | 74 | NA | 74 |
| no_stone | no | mixed | NA | NA | 74 | NA | 74 |
| no_stone | no | mixed | NA | NA | 74 | NA | 74 |
| no_stone | no | mixed | NA | NA | 74 | NA | 74 |
| no_stone | no | mixed | NA | NA | 74 | NA | 74 |
| no_stone | no | mixed | NA | NA | 74 | NA | 74 |
| no_stone | no | mixed | NA | NA | 74 | NA | 74 |
| no_stone | no | mixed | NA | NA | 74 | NA | 74 |
| no_stone | no | mixed | NA | NA | 74 | NA | 74 |
| no_stone | no | mixed | NA | NA | 74 | NA | 74 |
| no_stone | no | mixed | NA | NA | 74 | NA | 74 |
| no_stone | no | mixed | NA | NA | 55 | NA | 55 |
| no_stone | no | mixed | NA | NA | 55 | NA | 55 |
| no_stone | no | mixed | NA | NA | 55 | NA | 55 |
| no_stone | no | mixed | NA | NA | 55 | NA | 55 |
| no_stone | no | mixed | NA | NA | 55 | NA | 55 |
| no_stone | no | mixed | NA | NA | 55 | NA | 55 |
| no_stone | no | mixed | NA | NA | 55 | NA | 55 |
| no_stone | no | mixed | NA | NA | 55 | NA | 55 |
| no_stone | no | mixed | NA | NA | 55 | NA | 55 |
| no_stone | no | mixed | NA | NA | 55 | NA | 55 |
| no_stone | no | mixed | NA | NA | 55 | NA | 55 |
| no_stone | no | mixed | NA | NA | 55 | NA | 55 |
| no_stone | no | mixed | NA | NA | 55 | NA | 55 |
| no_stone | no | mixed | NA | NA | 55 | NA | 55 |
| no_stone | no | mixed | NA | NA | 55 | NA | 55 |
| no_stone | no | mixed | NA | NA | 44 | NA | 44 |
| no_stone | no | mixed | NA | NA | 44 | NA | 44 |
| no_stone | no | mixed | NA | NA | 44 | NA | 44 |
| no_stone | no | mixed | NA | NA | 44 | NA | 44 |
| no_stone | no | mixed | NA | NA | 44 | NA | 44 |
| no_stone | no | mixed | NA | NA | 44 | NA | 44 |
| no_stone | no | mixed | NA | NA | 44 | NA | 44 |
| no_stone | no | mixed | NA | NA | 44 | NA | 44 |
| no_stone | no | mixed | NA | NA | 44 | NA | 44 |
| no_stone | no | mixed | NA | NA | 44 | NA | 44 |
| no_stone | no | mixed | NA | NA | 44 | NA | 44 |
| no_stone | no | mixed | NA | NA | 44 | NA | 44 |
| no_stone | no | mixed | NA | NA | 44 | NA | 44 |
| no_stone | no | mixed | NA | NA | 44 | NA | 44 |
| no_stone | no | mixed | NA | NA | 44 | NA | 44 |
| Pais, 2007 | | | | | | | |
| no_stone | no | mixed | 43.00 | 10.000 | 61 | 12 | 49 |
| Stitchantrakul, 2007 | | | | | | | |
| no_stone | no | mixed | 42.60 | 1.900 | 34 | 14 | 20 |
| Ciacci, 2008 | | | | | | | |
| no_stone | no | mixed | 35.80 | NA | 45 | 9 | 36 |
| Baxmann, 2008 | | | | | | | |
| no_stone | no | mixed | 36.60 | 13.600 | 170 | 78 | 92 |
| Rodgers, 2009 | | | | | | | |
| no_stone | no | mixed | 50.00 | 10.000 | 18 | 16 | 2 |
| no_stone | no | mixed | 50.00 | 10.000 | 18 | 16 | 2 |
| no_stone | no | mixed | 50.00 | 10.000 | 18 | 16 | 2 |
| no_stone | no | mixed | 50.00 | 10.000 | 17 | 10 | 7 |
| no_stone | no | mixed | 50.00 | 10.000 | 17 | 10 | 7 |
| no_stone | no | mixed | 50.00 | 10.000 | 17 | 10 | 7 |
| no_stone | no | mixed | 50.00 | 10.000 | 37 | 19 | 18 |
| no_stone | no | mixed | 50.00 | 10.000 | 37 | 19 | 18 |
| no_stone | no | mixed | 50.00 | 10.000 | 37 | 19 | 18 |
| Sweeney, 2009 | | | | | | | |
| no_stone | no | mixed | 36.30 | 10.600 | 14 | NA | NA |
| Maalouf, 2010 | | | | | | | |
| no_stone | no | mixed | 52.00 | 10.000 | 16 | 8 | 8 |
| no_stone | no | mixed | 52.00 | 10.000 | 16 | 8 | 8 |
| no_stone | no | mixed | 54.00 | 11.000 | 9 | 5 | 4 |
| no_stone | no | mixed | 54.00 | 11.000 | 9 | 5 | 4 |
| Semins, 2010 | | | | | | | |
| no_stone | no | mixed | NA | NA | 96 | 96 | 0 |
| no_stone | no | mixed | NA | NA | 96 | 96 | 0 |
| no_stone | no | mixed | NA | NA | 72 | 0 | 72 |
| no_stone | no | mixed | NA | NA | 72 | 0 | 72 |
| Duffey, 2011 | | | | | | | |
| no_stone | no | mixed | 46.00 | 10.400 | 51 | 16 | 35 |
| Parvin, 2011 | | | | | | | |
| no_stone | no | mixed | 38.36 | 6.907 | 109 | 109 | 0 |
| Arrabal-Polo, 2012 | | | | | | | |
| no_stone | no | mixed | 49.53 | 10.140 | 60 | NA | NA |
| Goodarzi, 2012 | | | | | | | |
| no_stone | no | mixed | 39.90 | 12.600 | 27 | 18 | 9 |
| Herrel, 2012 | | | | | | | |
| no_stone | no | mixed | 38.70 | NA | 10 | NA | NA |
| Hong, 2012 | | | | | | | |
| no_stone | no | mixed | 50.20 | 10.800 | 30 | 12 | 18 |
| Sorensen, 2012 | | | | | | | |
| no_stone | no | mixed | 58.00 | 12.000 | 50 | 13 | 37 |
| Nouvenne, 2013 | | | | | | | |
| no_stone | no | mixed | 46.00 | 6.000 | 39 | 0 | 39 |
| no_stone | no | mixed | 46.00 | 6.000 | 39 | 0 | 39 |
| no_stone | no | mixed | 46.00 | 6.000 | 39 | 0 | 39 |
| no_stone | no | mixed | 46.00 | 6.000 | 39 | 0 | 39 |
| Elkoushy, 2014 | | | | | | | |
| no_stone | no | mixed | NA | NA | 204 | 57 | 147 |
| Lange, 2014 | | | | | | | |
| no_stone | no | mixed | 25.30 | 2.700 | 15 | 8 | 7 |
| Pigna, 2014 | | | | | | | |
| no_stone | no | mixed | 52.10 | 13.600 | 21 | 21 | 0 |
| Ma, 2014 | | | | | | | |
| no_stone | no | mixed | 54.20 | 2.400 | 29 | NA | NA |
| Doenyas-Barak, 2015 | | | | | | | |
| no_stone | no | mixed | 48.70 | 12.500 | 22 | 22 | 0 |
| no_stone | no | mixed | 48.70 | 12.500 | 22 | 22 | 0 |
| no_stone | no | mixed | 44.70 | 15.300 | 26 | 0 | 26 |
| no_stone | no | mixed | 44.70 | 15.300 | 26 | 0 | 26 |
| Grases, 2015 | | | | | | | |
| no_stone | no | mixed | NA | NA | 75 | 42 | 33 |
| Arrabal-Polo, 2015 | | | | | | | |
| no_stone | no | mixed | 51.81 | 10.320 | 61 | 31 | 30 |
| Perinpam, 2015 | | | | | | | |
| no_stone | no | mixed | 65.40 | 9.000 | 709 | 293 | 416 |
| Rodgers, 2015 | | | | | | | |
| no_stone | no | mixed | NA | NA | 15 | 15 | 0 |
| Rodríguez-Rodríguez, 2015 | | | | | | | |
| no_stone | no | mixed | 35.70 | 11.200 | 418 | 196 | 222 |
| Doizi, 2016 | | | | | | | |
| no_stone | no | mixed | 57.60 | 9.700 | 9 | 5 | 4 |
| no_stone | no | mixed | 54.30 | 9.500 | 12 | 6 | 6 |
| Arora, 2017 | | | | | | | |
| no_stone | no | mixed | 49.40 | 14.900 | 20 | 10 | 10 |
| Deng, 2017 | | | | | | | |
| no_stone | no | mixed | 48.86 | 16.250 | 376 | 188 | 188 |
| no_stone | no | mixed | 48.86 | 16.250 | 376 | 188 | 188 |
| no_stone | no | mixed | 48.86 | 16.250 | 376 | 188 | 188 |
| no_stone | no | mixed | 48.86 | 16.250 | 376 | 188 | 188 |
| no_stone | no | mixed | 53.96 | 19.280 | 24 | 12 | 12 |
| no_stone | no | mixed | 53.96 | 19.280 | 24 | 12 | 12 |
| no_stone | no | mixed | 53.96 | 19.280 | 24 | 12 | 12 |
| no_stone | no | mixed | 53.96 | 19.280 | 24 | 12 | 12 |
| no_stone | no | mixed | 52.14 | 14.250 | 149 | 64 | 85 |
| no_stone | no | mixed | 52.14 | 14.250 | 149 | 64 | 85 |
| no_stone | no | mixed | 52.14 | 14.250 | 149 | 64 | 85 |
| no_stone | no | mixed | 52.14 | 14.250 | 149 | 64 | 85 |
| no_stone | no | mixed | 50.03 | 12.840 | 35 | 14 | 21 |
| no_stone | no | mixed | 50.03 | 12.840 | 35 | 14 | 21 |
| no_stone | no | mixed | 50.03 | 12.840 | 35 | 14 | 21 |
| no_stone | no | mixed | 50.03 | 12.840 | 35 | 14 | 21 |
| Prochaska, 2018 | | | | | | | |
| no_stone | no | mixed | 65.50 | 5.700 | 407 | 0 | 407 |
| no_stone | no | mixed | 65.50 | 5.700 | 407 | 0 | 407 |
| no_stone | no | mixed | 65.50 | 5.700 | 407 | 0 | 407 |
| no_stone | no | mixed | 51.00 | 5.100 | 443 | 0 | 443 |
| no_stone | no | mixed | 51.00 | 5.100 | 443 | 0 | 443 |
| no_stone | no | mixed | 51.00 | 5.100 | 443 | 0 | 443 |
| no_stone | no | mixed | 64.60 | 6.300 | 417 | 417 | 0 |
| no_stone | no | mixed | 64.60 | 6.300 | 417 | 417 | 0 |
| no_stone | no | mixed | 64.60 | 6.300 | 417 | 417 | 0 |
| Athanasatou, 2018 | | | | | | | |
| no_stone | no | mixed | NA | NA | 163 | NA | NA |
| Mai, 2019 | | | | | | | |
| no_stone | pre_menopausal | mixed | 39.98 | 8.430 | 160 | 0 | 160 |
| no_stone | pre_menopausal | mixed | 39.98 | 8.430 | 160 | 0 | 160 |
| no_stone | post_menopausal | mixed | 62.51 | 8.440 | 194 | 0 | 194 |
| no_stone | post_menopausal | mixed | 62.51 | 8.440 | 194 | 0 | 194 |
| Baric, 2019 | | | | | | | |
| no_stone | no | mixed | 21.00 | 2.000 | 53 | 25 | 28 |
| Baric et al. 2019 | | | | | | | |
| no_stone | no | mixed | NA | NA | NA | NA | NA |
| Thongprayoon, 2022 | | | | | | | |
| no_stone | no | mixed | 45.33 | 18.590 | 440 | 215 | 225 |
| Pieters, 2022 | | | | | | | |
| no_stone | no | mixed | 54.00 | 12.000 | 287 | 112 | 175 |
| Ferraro, 2023 | | | | | | | |
| no_stone | no | mixed | 64.00 | 6.000 | 404 | NA | NA |
| no_stone | no | mixed | 64.00 | 6.000 | 404 | NA | NA |
| no_stone | no | mixed | 64.00 | 6.000 | 404 | NA | NA |
| no_stone | no | mixed | 65.00 | 6.000 | 399 | NA | NA |
| no_stone | no | mixed | 65.00 | 6.000 | 399 | NA | NA |
| no_stone | no | mixed | 65.00 | 6.000 | 399 | NA | NA |
| no_stone | no | mixed | 53.00 | 4.000 | 2762 | NA | NA |
| no_stone | no | mixed | 53.00 | 4.000 | 2762 | NA | NA |
| no_stone | no | mixed | 53.00 | 4.000 | 2762 | NA | NA |
| Cao, 2023 | | | | | | | |
| no_stone | no | mixed | 49.08 | 12.700 | 40 | 28 | 12 |
| Ferraro, 2024 | | | | | | | |
| no_stone | no | mixed | 55.00 | 7.000 | 3640 | 410 | 3230 |
| HPFS, 40-50 | | | | | | | |
| no_stone | no | mixed | 48.67 | 0.520 | 6 | 6 | 0 |
| NHS, 40-50 | | | | | | | |
| no_stone | no | mixed | 47.09 | 2.290 | 524 | 0 | 524 |
| no_stone | no | mixed | 46.86 | 2.030 | 29 | 0 | 29 |
| HPFS, 50-60 | | | | | | | |
| no_stone | no | mixed | 55.63 | 3.020 | 79 | 79 | 0 |
| NHS, 50-60 | | | | | | | |
| no_stone | no | mixed | 54.25 | 2.690 | 2409 | 0 | 2409 |
| no_stone | no | mixed | 54.92 | 2.650 | 131 | 0 | 131 |
| HPFS, 60-70 | | | | | | | |
| no_stone | no | mixed | 64.06 | 2.790 | 215 | 215 | 0 |
| NHS, 60-70 | | | | | | | |
| no_stone | no | mixed | 64.59 | 2.950 | 231 | 0 | 231 |
| no_stone | no | mixed | 64.86 | 3.020 | 93 | 0 | 93 |
| HPFS, 70-80 | | | | | | | |
| no_stone | no | mixed | 72.30 | 1.680 | 89 | 89 | 0 |
| NHS, 70-80 | | | | | | | |
| no_stone | no | mixed | 72.36 | 1.540 | 96 | 0 | 96 |
| no_stone | no | mixed | 71.73 | 1.340 | 51 | 0 | 51 |
| NHS II, postmenopausal | | | | | | | |
| no_stone | no | mixed | NA | NA | 784 | 0 | 784 |
| NHS II, premenopausal | | | | | | | |
| no_stone | no | mixed | NA | NA | 2567 | 0 | 2567 |

#### 2.4 Table of Papers Included in Meta-analysis

```
urine_ma2 %>%
  arrange(year) %>%
  drop_na(year) %>%
  mutate(first_author = case_when(
    and == "no" ~ paste0(first_author, ", ", year),
    first_author == "HPFS" | first_author == "NHS" ~ first_author,
    TRUE ~ paste0(first_author, " et al. ", year)
  )) %>%
  group_by(first_author) %>%
  select(stone_vs_no_stone,
         underlying_cause,
         other_demographic_variant,
         first_author,
         mean_age,
         age_sd,
         total_n,
         male_n,
         female_n) %>%
  gt() %>%
  gt_theme_espn() %>%
  tab_header(
    title = md("**Summary of Papers included in Meta-Analysis**")
  ) %>%
  cols_align(
    align = "center",
    columns = everything()
  ) %>%
  tab_style(
    style = cell_text(weight = "bold"),
    locations = cells_column_labels(everything())
  ) %>%
  tab_options(
    table.border.top.width = px(2),
    table.border.bottom.width = px(2),
    table.font.size = px(14),
    data_row.padding = px(10)
  )
```

Table 1: **Summary of Papers included in Meta-Analysis**

| stone_vs_no_stone | underlying_cause | other_demographic_variant | mean_age | age_sd | total_n | male_n | female_n |
| --- | --- | --- | --- | --- | --- | --- | --- |
| Goldschmied, 1975 | | | | | | | |
| no_stone | no | mixed | NA | NA | 55 | 28 | 27 |
| Welshman, 1975 | | | | | | | |
| no_stone | no | mixed | NA | NA | 89 | 89 | 0 |
| no_stone | no | mixed | NA | NA | 89 | 89 | 0 |
| Welshman, 1976 | | | | | | | |
| no_stone | no | mixed | NA | NA | 158 | 73 | 85 |
| Marya, 1979 | | | | | | | |
| no_stone | no | mixed | NA | NA | 50 | NA | NA |
| Stern, 1980 | | | | | | | |
| no_stone | no | mixed | NA | NA | 9 | NA | NA |
| Hesse, 1986 | | | | | | | |
| no_stone | no | mixed | NA | NA | 25 | 0 | 25 |
| Hesse et al. 1986 | | | | | | | |
| no_stone | no | mixed | NA | NA | 25 | 0 | 25 |
| no_stone | no | mixed | NA | NA | 25 | 0 | 25 |
| no_stone | no | mixed | NA | NA | 25 | 0 | 25 |
| no_stone | no | mixed | NA | NA | 25 | 0 | 25 |
| no_stone | no | mixed | NA | NA | 25 | 25 | 0 |
| no_stone | no | mixed | NA | NA | 25 | 25 | 0 |
| no_stone | no | mixed | NA | NA | 25 | 25 | 0 |
| no_stone | no | mixed | NA | NA | 25 | 25 | 0 |
| no_stone | no | mixed | NA | NA | 25 | 25 | 0 |
| Bingham, 1988 | | | | | | | |
| no_stone | no | mixed | NA | NA | 8 | 5 | 3 |
| Michelacci, 1989 | | | | | | | |
| no_stone | no | mixed | NA | NA | 23 | 11 | 12 |
| Bataille, 1991 | | | | | | | |
| no_stone | no | mixed | 35.00 | 5.000 | 62 | 41 | 21 |
| Wabner, 1992 | | | | | | | |
| no_stone | no | mixed | 38.00 | 8.600 | 12 | 0 | 12 |
| Erwin, 1994 | | | | | | | |
| no_stone | no | mixed | 40.00 | 1.500 | 22 | 16 | 6 |
| Buchholz, 1996 | | | | | | | |
| no_stone | no | mixed | 34.00 | 10.800 | 37 | 37 | 0 |
| Christie, 1996 | | | | | | | |
| no_stone | no | mixed | 39.00 | 3.000 | 17 | 7 | 10 |
| Lemann, 1996 | | | | | | | |
| no_stone | no | mixed | 43.90 | 10.200 | 94 | 44 | 50 |
| Rodgers, 1997 | | | | | | | |
| no_stone | no | mixed | NA | NA | 40 | 20 | 20 |
| Hess, 1999 | | | | | | | |
| no_stone | no | mixed | 39.00 | 1.300 | 107 | 107 | 0 |
| Matin and Streem et al. 2000 | | | | | | | |
| no_stone | no | mixed | NA | NA | 26 | NA | NA |
| no_stone | no | mixed | NA | NA | 26 | 9 | 17 |
| Holmes, 2001 | | | | | | | |
| no_stone | no | mixed | 29.00 | 4.000 | 12 | 6 | 6 |
| Miyake, 2001 | | | | | | | |
| no_stone | no | mixed | 31.50 | 4.400 | 11 | 11 | 0 |
| Terris, 2001 | | | | | | | |
| no_stone | no | mixed | 30.20 | NA | 5 | 3 | 2 |
| Rodgers, 2002 | | | | | | | |
| no_stone | no | mixed | NA | NA | 10 | 10 | 0 |
| no_stone | no | mixed | NA | NA | 10 | 10 | 0 |
| no_stone | no | mixed | NA | NA | 10 | 10 | 0 |
| no_stone | no | mixed | NA | NA | 10 | 10 | 0 |
| Kinder, 2002 | | | | | | | |
| no_stone | no | mixed | NA | NA | 111 | NA | 111 |
| no_stone | no | mixed | NA | NA | 111 | NA | 111 |
| no_stone | no | mixed | NA | NA | 66 | 66 | NA |
| no_stone | no | mixed | NA | NA | 66 | 66 | NA |
| Allie, 2003 | | | | | | | |
| no_stone | no | mixed | NA | NA | 8 | 8 | 0 |
| Baxmann, 2003 | | | | | | | |
| no_stone | no | mixed | 37.00 | 16.000 | 20 | 8 | 12 |
| Huang, 2003 | | | | | | | |
| no_stone | no | mixed | 50.80 | 1.800 | 32 | 27 | 5 |
| Kuo, 2003 | | | | | | | |
| no_stone | no | mixed | NA | NA | 4 | NA | NA |
| Ogawa, 2003 | | | | | | | |
| no_stone | no | mixed | 54.10 | 16.200 | 22 | 6 | 16 |
| Traxer, 2003 | | | | | | | |
| no_stone | no | mixed | 38.10 | 11.300 | 12 | 6 | 6 |
| Abate, 2004 | | | | | | | |
| no_stone | no | mixed | 31.00 | 11.000 | 55 | 33 | 22 |
| Chai, 2004 | | | | | | | |
| no_stone | no | mixed | NA | NA | 9 | 5 | 4 |
| Stitchantrakul, 2004 | | | | | | | |
| no_stone | no | mixed | 22.70 | 1.900 | 38 | 38 | 0 |
| Defoor, 2005 | | | | | | | |
| no_stone | no | mixed | NA | NA | 168 | 97 | 71 |
| Gettman, 2005 | | | | | | | |
| no_stone | no | mixed | 34.80 | 9.600 | 12 | 6 | 6 |
| Shah, 2005 | | | | | | | |
| no_stone | no | mixed | 30.00 | NA | 101 | 54 | 47 |
| no_stone | no | mixed | 30.00 | NA | 101 | 54 | 47 |
| Shah et al. 2005 | | | | | | | |
| no_stone | no | mixed | 30.00 | 5.000 | 61 | 12 | 49 |
| no_stone | no | mixed | 30.00 | 5.000 | 61 | 12 | 49 |
| Cameron, 2006 | | | | | | | |
| no_stone | no | mixed | 49.00 | 8.000 | 59 | 24 | 35 |
| no_stone | no | mixed | 49.00 | 8.000 | 59 | 24 | 35 |
| no_stone | no | mixed | 52.00 | 8.000 | 24 | 10 | 14 |
| no_stone | no | mixed | 52.00 | 8.000 | 24 | 10 | 14 |
| Matsumoto, 2006 | | | | | | | |
| no_stone | no | mixed | NA | NA | 10 | 5 | 5 |
| Pais, 2007 | | | | | | | |
| no_stone | no | mixed | 43.00 | 10.000 | 61 | 12 | 49 |
| Stitchantrakul, 2007 | | | | | | | |
| no_stone | no | mixed | 42.60 | 1.900 | 34 | 14 | 20 |
| Ciacci, 2008 | | | | | | | |
| no_stone | no | mixed | 35.80 | NA | 45 | 9 | 36 |
| Baxmann, 2008 | | | | | | | |
| no_stone | no | mixed | 36.60 | 13.600 | 170 | 78 | 92 |
| Rodgers, 2009 | | | | | | | |
| no_stone | no | mixed | 50.00 | 10.000 | 18 | 16 | 2 |
| no_stone | no | mixed | 50.00 | 10.000 | 18 | 16 | 2 |
| no_stone | no | mixed | 50.00 | 10.000 | 18 | 16 | 2 |
| no_stone | no | mixed | 50.00 | 10.000 | 17 | 10 | 7 |
| no_stone | no | mixed | 50.00 | 10.000 | 17 | 10 | 7 |
| no_stone | no | mixed | 50.00 | 10.000 | 17 | 10 | 7 |
| no_stone | no | mixed | 50.00 | 10.000 | 37 | 19 | 18 |
| no_stone | no | mixed | 50.00 | 10.000 | 37 | 19 | 18 |
| no_stone | no | mixed | 50.00 | 10.000 | 37 | 19 | 18 |
| Sweeney, 2009 | | | | | | | |
| no_stone | no | mixed | 36.30 | 10.600 | 14 | NA | NA |
| Maalouf, 2010 | | | | | | | |
| no_stone | no | mixed | 52.00 | 10.000 | 16 | 8 | 8 |
| no_stone | no | mixed | 52.00 | 10.000 | 16 | 8 | 8 |
| no_stone | no | mixed | 54.00 | 11.000 | 9 | 5 | 4 |
| no_stone | no | mixed | 54.00 | 11.000 | 9 | 5 | 4 |
| Semins, 2010 | | | | | | | |
| no_stone | no | mixed | NA | NA | 96 | 96 | 0 |
| no_stone | no | mixed | NA | NA | 96 | 96 | 0 |
| no_stone | no | mixed | NA | NA | 72 | 0 | 72 |
| no_stone | no | mixed | NA | NA | 72 | 0 | 72 |
| Duffey, 2011 | | | | | | | |
| no_stone | no | mixed | 46.00 | 10.400 | 51 | 16 | 35 |
| Parvin, 2011 | | | | | | | |
| no_stone | no | mixed | 38.36 | 6.907 | 109 | 109 | 0 |
| Arrabal-Polo, 2012 | | | | | | | |
| no_stone | no | mixed | 49.53 | 10.140 | 60 | NA | NA |
| Goodarzi, 2012 | | | | | | | |
| no_stone | no | mixed | 39.90 | 12.600 | 27 | 18 | 9 |
| Herrel, 2012 | | | | | | | |
| no_stone | no | mixed | 38.70 | NA | 10 | NA | NA |
| Hong, 2012 | | | | | | | |
| no_stone | no | mixed | 50.20 | 10.800 | 30 | 12 | 18 |
| Sorensen, 2012 | | | | | | | |
| no_stone | no | mixed | 58.00 | 12.000 | 50 | 13 | 37 |
| Nouvenne, 2013 | | | | | | | |
| no_stone | no | mixed | 46.00 | 6.000 | 39 | 0 | 39 |
| no_stone | no | mixed | 46.00 | 6.000 | 39 | 0 | 39 |
| no_stone | no | mixed | 46.00 | 6.000 | 39 | 0 | 39 |
| no_stone | no | mixed | 46.00 | 6.000 | 39 | 0 | 39 |
| Elkoushy, 2014 | | | | | | | |
| no_stone | no | mixed | NA | NA | 204 | 57 | 147 |
| Lange, 2014 | | | | | | | |
| no_stone | no | mixed | 25.30 | 2.700 | 15 | 8 | 7 |
| Pigna, 2014 | | | | | | | |
| no_stone | no | mixed | 52.10 | 13.600 | 21 | 21 | 0 |
| Ma, 2014 | | | | | | | |
| no_stone | no | mixed | 54.20 | 2.400 | 29 | NA | NA |
| Doenyas-Barak, 2015 | | | | | | | |
| no_stone | no | mixed | 48.70 | 12.500 | 22 | 22 | 0 |
| no_stone | no | mixed | 48.70 | 12.500 | 22 | 22 | 0 |
| no_stone | no | mixed | 44.70 | 15.300 | 26 | 0 | 26 |
| no_stone | no | mixed | 44.70 | 15.300 | 26 | 0 | 26 |
| Grases, 2015 | | | | | | | |
| no_stone | no | mixed | NA | NA | 75 | 42 | 33 |
| Arrabal-Polo, 2015 | | | | | | | |
| no_stone | no | mixed | 51.81 | 10.320 | 61 | 31 | 30 |
| Perinpam, 2015 | | | | | | | |
| no_stone | no | mixed | 65.40 | 9.000 | 709 | 293 | 416 |
| Rodgers, 2015 | | | | | | | |
| no_stone | no | mixed | NA | NA | 15 | 15 | 0 |
| Rodríguez-Rodríguez, 2015 | | | | | | | |
| no_stone | no | mixed | 35.70 | 11.200 | 418 | 196 | 222 |
| Doizi, 2016 | | | | | | | |
| no_stone | no | mixed | 57.60 | 9.700 | 9 | 5 | 4 |
| no_stone | no | mixed | 54.30 | 9.500 | 12 | 6 | 6 |
| Arora, 2017 | | | | | | | |
| no_stone | no | mixed | 49.40 | 14.900 | 20 | 10 | 10 |
| Deng, 2017 | | | | | | | |
| no_stone | no | mixed | 48.86 | 16.250 | 376 | 188 | 188 |
| no_stone | no | mixed | 48.86 | 16.250 | 376 | 188 | 188 |
| no_stone | no | mixed | 48.86 | 16.250 | 376 | 188 | 188 |
| no_stone | no | mixed | 48.86 | 16.250 | 376 | 188 | 188 |
| no_stone | no | mixed | 53.96 | 19.280 | 24 | 12 | 12 |
| no_stone | no | mixed | 53.96 | 19.280 | 24 | 12 | 12 |
| no_stone | no | mixed | 53.96 | 19.280 | 24 | 12 | 12 |
| no_stone | no | mixed | 53.96 | 19.280 | 24 | 12 | 12 |
| no_stone | no | mixed | 52.14 | 14.250 | 149 | 64 | 85 |
| no_stone | no | mixed | 52.14 | 14.250 | 149 | 64 | 85 |
| no_stone | no | mixed | 52.14 | 14.250 | 149 | 64 | 85 |
| no_stone | no | mixed | 52.14 | 14.250 | 149 | 64 | 85 |
| no_stone | no | mixed | 50.03 | 12.840 | 35 | 14 | 21 |
| no_stone | no | mixed | 50.03 | 12.840 | 35 | 14 | 21 |
| no_stone | no | mixed | 50.03 | 12.840 | 35 | 14 | 21 |
| no_stone | no | mixed | 50.03 | 12.840 | 35 | 14 | 21 |
| Athanasatou, 2018 | | | | | | | |
| no_stone | no | mixed | NA | NA | 163 | NA | NA |
| Mai, 2019 | | | | | | | |
| no_stone | pre_menopausal | mixed | 39.98 | 8.430 | 160 | 0 | 160 |
| no_stone | pre_menopausal | mixed | 39.98 | 8.430 | 160 | 0 | 160 |
| Baric, 2019 | | | | | | | |
| no_stone | no | mixed | 21.00 | 2.000 | 53 | 25 | 28 |
| Thongprayoon, 2022 | | | | | | | |
| no_stone | no | mixed | 45.33 | 18.590 | 440 | 215 | 225 |
| Pieters, 2022 | | | | | | | |
| no_stone | no | mixed | 54.00 | 12.000 | 287 | 112 | 175 |
| Cao, 2023 | | | | | | | |
| no_stone | no | mixed | 49.08 | 12.700 | 40 | 28 | 12 |
| HPFS, 40-50 | | | | | | | |
| no_stone | no | mixed | 48.67 | 0.520 | 6 | 6 | 0 |
| NHS, 40-50 | | | | | | | |
| no_stone | no | mixed | 47.09 | 2.290 | 524 | 0 | 524 |
| no_stone | no | mixed | 46.86 | 2.030 | 29 | 0 | 29 |
| HPFS, 50-60 | | | | | | | |
| no_stone | no | mixed | 55.63 | 3.020 | 79 | 79 | 0 |
| NHS, 50-60 | | | | | | | |
| no_stone | no | mixed | 54.25 | 2.690 | 2409 | 0 | 2409 |
| no_stone | no | mixed | 54.92 | 2.650 | 131 | 0 | 131 |
| HPFS, 60-70 | | | | | | | |
| no_stone | no | mixed | 64.06 | 2.790 | 215 | 215 | 0 |
| NHS, 60-70 | | | | | | | |
| no_stone | no | mixed | 64.59 | 2.950 | 231 | 0 | 231 |
| no_stone | no | mixed | 64.86 | 3.020 | 93 | 0 | 93 |
| HPFS, 70-80 | | | | | | | |
| no_stone | no | mixed | 72.30 | 1.680 | 89 | 89 | 0 |
| NHS, 70-80 | | | | | | | |
| no_stone | no | mixed | 72.36 | 1.540 | 96 | 0 | 96 |
| no_stone | no | mixed | 71.73 | 1.340 | 51 | 0 | 51 |
| NHS II, postmenopausal | | | | | | | |
| no_stone | no | mixed | NA | NA | 784 | 0 | 784 |
| NHS II, premenopausal | | | | | | | |
| no_stone | no | mixed | NA | NA | 2567 | 0 | 2567 |

#### 2.5 Summary table of Included patients

```
mean_age_summary <- urine_ma2 %>%
  mutate(first_author = ifelse(
    and == "no",
    paste0(first_author, " et al. ", year),
    paste0(first_author, ", ", year)
  )) %>%
  drop_na(age_sd) %>%
  metamean(
    n = total_n,
    mean = mean_age,
    sd = age_sd,
    studlab = study_name
  )

urine_ma2 %>%
  arrange(year) %>%
  mutate(first_author = ifelse(
    and == "no",
    paste0(first_author, " et al. ", year),
    paste0(first_author, ", ", year)
  )) %>%
  select(total_n, male_n, female_n, first_author) %>%
  summarise(
    "Total Papers, n" = n_distinct(first_author),
    "Total Patients, n" = sum(total_n, na.rm = TRUE),
    "Male Patients, n" = sum(male_n, na.rm = TRUE),
    "Female Patients, n" = sum(female_n, na.rm = TRUE),
    "Mean Age, years ± SD" = paste0(
      round(mean_age_summary$TE.random),
      " ± ",
      round(mean_age_summary$tau)
    )
  ) %>%
  gt() %>%
  gt_theme_espn() %>%
  tab_header(
    title = md("**Summary of Included Studies for MA**"),
    subtitle = "Only non kidney stone forming patients included in analysis"
  ) %>%
  cols_align(
    align = "center",
    columns = everything()
  ) %>%
  tab_style(
    style = cell_text(weight = "bold"),
    locations = cells_column_labels(everything())
  ) %>%
  tab_options(
    table.border.top.width = px(2),
    table.border.bottom.width = px(2),
    table.font.size = px(14),
    data_row.padding = px(10)
  ) %>%
  tab_footnote(footnote = "Some studies only provided a total number of patients, rather than subdividing by sex",
    locations = cells_column_labels(
      columns = c("Male Patients, n", "Female Patients, n")
    ),
    placement = "right"
  )
```

Table 1: **Summary of Included Studies for MA**

Only non kidney stone forming patients included in analysis

| Total Papers, n | Total Patients, n | Male Patients, n*^1^* | Female Patients, n*^1^* | Mean Age, years ± SD |
| --- | --- | --- | --- | --- |
| 83 | 16522 | 4575 | 11582 | 48 ± 10 |
| *^1^*Some studies only provided a total number of patients, rather than subdividing by sex | | | | |

### 3 Current Expected Ranges

#### 3.1 EAU

EAU Guidelines - Skolarikos, A., Geraghty, R., Somani, B., Tailly, T., Jung, H., Neisius, A., Petřík, A., Kamphuis, G.M., Davis, N., Bezuidenhout, C. and Lardas, M., 2025. European association of urology guidelines on the diagnosis and treatment of urolithiasis. European Urology, 88(1), pp.64-75.

```
current_ref_range1 <- as_tibble(
  cbind(
    "urine_component" = c("Volume (L / 24hrs)", #1
                          "pH", #2
                          "Creatinine (mmol / 24hrs) - Overall", #3
                          "Creatinine (mmol / 24hrs) - Men Only", #4
                          "Creatinine (mmol / 24hrs) - Women Only", #5
                          "Calcium (mmol / 24hrs) - Overall", #6
                          "Calcium (mmol / 24hrs) - Men Only", #7
                          "Calcium (mmol / 24hrs) - Women Only", #8
                          "Oxalate (mmol / 24hrs) - Overall", #9
                          "Oxalate (mmol / 24hrs) - Men Only", #10
                          "Oxalate (mmol / 24hrs) - Women Only", #11
                          "Urate (mmol / 24hrs) - Overall", #12
                          "Urate (mmol / 24hrs) - Men Only", #13
                          "Urate (mmol / 24hrs) - Women Only", #14
                          "Citrate (mmol / 24hrs) - Overall", #15
                          "Citrate (mmol / 24hrs) - Men Only", #16
                          "Citrate (mmol / 24hrs) - Women Only", #17
                          "Phosphate (mmol / 24hrs)", #18
                          "Sodium (mmol / 24hrs)", #19
                          "Potassium (mmol / 24hrs)", #20
                          "Magnesium (mmol / 24hrs)", #21
                          "Ammonium (mmol / 24hrs)", #22
                          "Chloride (mmol / 24hrs)", #23
                          "Sulphate (mmol / 24hrs)", #24
                          "Urea (mmol / 24hrs)"),
    "lower_limit" = c(0.5, #1
                      5.5, #2
                      NA, #3
                      13, #4
                      7, #5
                      NA, #6
                      NA, #7
                      NA, #8
                      NA, #9
                      NA, #10
                      NA, #11
                      NA, #12
                      NA, #13
                      NA, #14
                      NA, #15
                      1.7, #16
                      1.9, #17
                      NA, #18
                      NA, #19
                      NA, #20
                      3, #21
                      NA, #22
                      NA, #23
                      NA, #24
                      NA #25
                      ),
    "upper_limit" = c(NA,
                      7,
                      NA,
                      18,
                      13,
                      8,
                      NA,
                      NA,
                      0.5,
                      NA,
                      NA,
                      NA,
                      5,
                      4,
                      NA,
                      NA,
                      NA,
                      35,
                      NA,
                      NA,
                      NA,
                      50,
                      NA,
                      NA,
                      NA
                      ),
    "source" = "EAU"
  )
  )

# EAU Guidelines - Skolarikos, A., Geraghty, R., Somani, B., Tailly, T., Jung, H., Neisius, A., Petřík, A., Kamphuis, G.M., Davis, N., Bezuidenhout, C. and Lardas, M., 2025. European association of urology guidelines on the diagnosis and treatment of urolithiasis. European Urology, 88(1), pp.64-75.
# Date accessed 20/8/25
```

#### 3.2 NHS

References: North Bristol NHS Trust & York (<https://www.nbt.nhs.uk/severn-pathology/requesting/test-information?search=urine>) Scarborough NHS Trust (<https://www.yorkhospitals.nhs.uk/our-services/organdonation/a-z-of-services/laboratory-medicine/test-directory/>)

```
current_ref_range2 <- as_tibble(
  cbind(
    "urine_component" = c("Volume (L / 24hrs)", #1
                          "pH", #2
                          "Creatinine (mmol / 24hrs) - Overall", #3
                          "Creatinine (mmol / 24hrs) - Men Only", #4
                          "Creatinine (mmol / 24hrs) - Women Only", #5
                          "Calcium (mmol / 24hrs) - Overall", #6
                          "Calcium (mmol / 24hrs) - Men Only", #7
                          "Calcium (mmol / 24hrs) - Women Only", #8
                          "Oxalate (mmol / 24hrs) - Overall", #9
                          "Oxalate (mmol / 24hrs) - Men Only", #10
                          "Oxalate (mmol / 24hrs) - Women Only", #11
                          "Urate (mmol / 24hrs) - Overall", #12
                          "Urate (mmol / 24hrs) - Men Only", #13
                          "Urate (mmol / 24hrs) - Women Only", #14
                          "Citrate (mmol / 24hrs) - Overall", #15
                          "Citrate (mmol / 24hrs) - Men Only", #16
                          "Citrate (mmol / 24hrs) - Women Only", #17
                          "Phosphate (mmol / 24hrs)", #18
                          "Sodium (mmol / 24hrs)", #19
                          "Potassium (mmol / 24hrs)", #20
                          "Magnesium (mmol / 24hrs)", #21
                          "Ammonium (mmol / 24hrs)", #22
                          "Chloride (mmol / 24hrs)", #23
                          "Sulphate (mmol / 24hrs)", #24
                          "Urea (mmol / 24hrs)"),
    "lower_limit" = c(NA,
                      NA,
                      NA,
                      9,
                      6,
                      2.5,
                      NA,
                      NA,
                      0.1,
                      NA,
                      NA,
                      1.5,
                      NA,
                      NA,
                      0.6,
                      NA,
                      NA,
                      15,
                      40,
                      25,
                      2.4,
                      NA,
                      110,
                      NA,
                      250
                      ),
    "upper_limit" = c(3,
                      NA,
                      NA,
                      19,
                      13,
                      7.5,
                      NA,
                      NA,
                      0.46,
                      NA,
                      NA,
                      4.5,
                      NA,
                      NA,
                      4.8,
                      NA,
                      NA,
                      50,
                      250,
                      125,
                      6.5,
                      NA,
                      250,
                      NA,
                      570
                      ),
    "source" = "NHS"
    # North Bristol NHS Trust & York (https://www.nbt.nhs.uk/severn-pathology/requesting/test-information?search=urine)
    # / Scarborough NHS Trust (https://www.yorkhospitals.nhs.uk/our-services/organdonation/a-z-of-services/laboratory-medicine/test-directory/)
  # Date accessed 20/8/25
  )
)
```

#### 3.3 Mayo Clinic

Reference: Mayo Clinic Laboratory - (<https://www.mayocliniclabs.com/api/sitecore/TestCatalog/DownloadTestCatalog?testId=616228>)

```
current_ref_range3 <- as_tibble(
  cbind(
    "urine_component" = c("Volume (L / 24hrs)", #1
                          "pH", #2
                          "Creatinine (mmol / 24hrs) - Overall", #3
                          "Creatinine (mmol / 24hrs) - Men Only", #4
                          "Creatinine (mmol / 24hrs) - Women Only", #5
                          "Calcium (mmol / 24hrs) - Overall", #6
                          "Calcium (mmol / 24hrs) - Men Only", #7
                          "Calcium (mmol / 24hrs) - Women Only", #8
                          "Oxalate (mmol / 24hrs) - Overall", #9
                          "Oxalate (mmol / 24hrs) - Men Only", #10
                          "Oxalate (mmol / 24hrs) - Women Only", #11
                          "Urate (mmol / 24hrs) - Overall", #12
                          "Urate (mmol / 24hrs) - Men Only", #13
                          "Urate (mmol / 24hrs) - Women Only", #14
                          "Citrate (mmol / 24hrs) - Overall", #15
                          "Citrate (mmol / 24hrs) - Men Only", #16
                          "Citrate (mmol / 24hrs) - Women Only", #17
                          "Phosphate (mmol / 24hrs)", #18
                          "Sodium (mmol / 24hrs)", #19
                          "Potassium (mmol / 24hrs)", #20
                          "Magnesium (mmol / 24hrs)", #21
                          "Ammonium (mmol / 24hrs)", #22
                          "Chloride (mmol / 24hrs)", #23
                          "Sulphate (mmol / 24hrs)", #24
                          "Urea (mmol / 24hrs)"),
    "lower_limit" = c(NA,
                      4.5,
                      NA,
                      930 / 113.12, #convert mg/24h to mmol/24h - MW of creatinine = 113.12
                      603 / 113.12,
                      NA, #6 Calcium Overall
                      NA, #7 Calcium Men
                      NA, #8 Calcium Women
                      0.11, #9 Oxalate Overall
                      NA, #10 Oxalate Men
                      NA, #11 Oxalate Women
                      NA,
                      200 / 168.11,
                      250 / 168.11,
                      242 / 192.1,
                      NA,
                      NA,
                      226 /  30.97, # 18
                      22,
                      16,
                      51 / 24.3,
                      15,
                      34,
                      7,
                      7/(28.013 * 2) * 1000 # Urea Nitrogen - multiple MW of Nitrogen by 2 then multiply by 1000 to convert from g/24h to mmol/24h
                      ),
    "upper_limit" = c(NA,
                      8,
                      NA,
                      2955 / 113.12,
                      1783 / 113.12,
                      200 / 40.08, #6 Calcium Overall
                      NA, #7 Calcium Men
                      NA, #8 Calcium Women
                      0.46, #9 Oxalate Overall
                      NA, #10 Oxalate Men
                      NA, #11 Oxalate Women
                      NA,
                      1000 / 168.11,
                      750 / 168.11,
                      1191/192.1,
                      NA,
                      NA,
                      1797 / 30.97, #18 MW PO4 = 94.97 P = 30.97
                      328,
                      105,
                      105 / 24.3,
                      56,
                      286,
                      47,
                      42 / (28.013 * 2) * 1000
                      ),
    "source" = "Mayo Clinic"
  # Mayo Clinic Laboratory - (https://www.mayocliniclabs.com/api/sitecore/TestCatalog/DownloadTestCatalog?testId=616228)
    # Date accessed 20/8/25
  )
)
```

#### 3.4 Litholink

Reference: <https://litholink.labcorp.com/testing/sample-reports>

```
current_ref_range4 <- as_tibble(
  cbind(
    "urine_component" = c("Volume (L / 24hrs)", #1
                          "pH", #2
                          "Creatinine (mmol / 24hrs) - Overall", #3
                          "Creatinine (mmol / 24hrs) - Men Only", #4
                          "Creatinine (mmol / 24hrs) - Women Only", #5
                          "Calcium (mmol / 24hrs) - Overall", #6
                          "Calcium (mmol / 24hrs) - Men Only", #7
                          "Calcium (mmol / 24hrs) - Women Only", #8
                          "Oxalate (mmol / 24hrs) - Overall", #9
                          "Oxalate (mmol / 24hrs) - Men Only", #10
                          "Oxalate (mmol / 24hrs) - Women Only", #11
                          "Urate (mmol / 24hrs) - Overall", #12
                          "Urate (mmol / 24hrs) - Men Only", #13
                          "Urate (mmol / 24hrs) - Women Only", #14
                          "Citrate (mmol / 24hrs) - Overall", #15
                          "Citrate (mmol / 24hrs) - Men Only", #16
                          "Citrate (mmol / 24hrs) - Women Only", #17
                          "Phosphate (mmol / 24hrs)", #18
                          "Sodium (mmol / 24hrs)", #19
                          "Potassium (mmol / 24hrs)", #20
                          "Magnesium (mmol / 24hrs)", #21
                          "Ammonium (mmol / 24hrs)", #22
                          "Chloride (mmol / 24hrs)", #23
                          "Sulphate (mmol / 24hrs)", #24
                          "Urea (mmol / 24hrs)"), #25
    "lower_limit" = c(2.5, #1
                      5.7,  #2
                      NA, #3
                      NA,  #4
                      NA,  #5
                      NA, #6 Calcium Overall
                      40 / 40.08, #7 Calcium Men - 40mg/24hr converted to mmol
                      30 / 40.08, #8 Calcium Women - 30mg/24hr converted to mmol
                      NA, #9 Oxalate Overall
                      NA, #10 Oxalate Men
                      NA, #11 Oxalate Women
                      NA, #12
                      460 / 168.11, #13
                      370 / 168.11, #14
                      NA, #15
                      400 / 192.12, #16
                      530 / 192.12, #17
                      0.6 / 30.97 * 1000, #18
                      50, #19
                      40, #20
                      62 / 24.3, #21
                      18, #22
                      50, #23
                      25/2, #24
                      7/(28.013) * 1000  #25
                      ),
    "upper_limit" = c(NA, #1
                      6.3, #2
                      NA, #3
                      NA, #4
                      NA, #5
                      NA, #6 Calcium Overall
                      250 / 40.08, #7 Calcium Men - 250mg/24hr converted to mmol
                      200 / 40.08, #8 Calcium Women - 200mg/24hr converted to mmol
                      NA, #9 Oxalate Overall
                      46 / 88.02, #10 Oxalate Men - <46mg/24hr converted to mmol
                      37 / 88.02, #11 Oxalate Women - <37mg/24hr converted to mmol
                      NA, #12
                      800 / 168.11, #13
                      750 / 168.11, #14
                      NA, #15
                      NA, #16
                      NA, #17
                      1.2 / 30.97 * 1000, #18
                      120, #19
                      100, #20
                      173 / 24.3, #21
                      56, #22
                      120, #23
                      56/2, #24
                      15 / (28.013) * 1000  #25
                      ),
    "source" = "Litholink"
  # https://litholink.labcorp.com/testing/sample-reports
    # Date accessed 22/8/25
  )
)
```

#### 3.5 Northern District Hospital (NDH), Hong Kong

knitr::include_graphics("ndh reference 3.pdf")

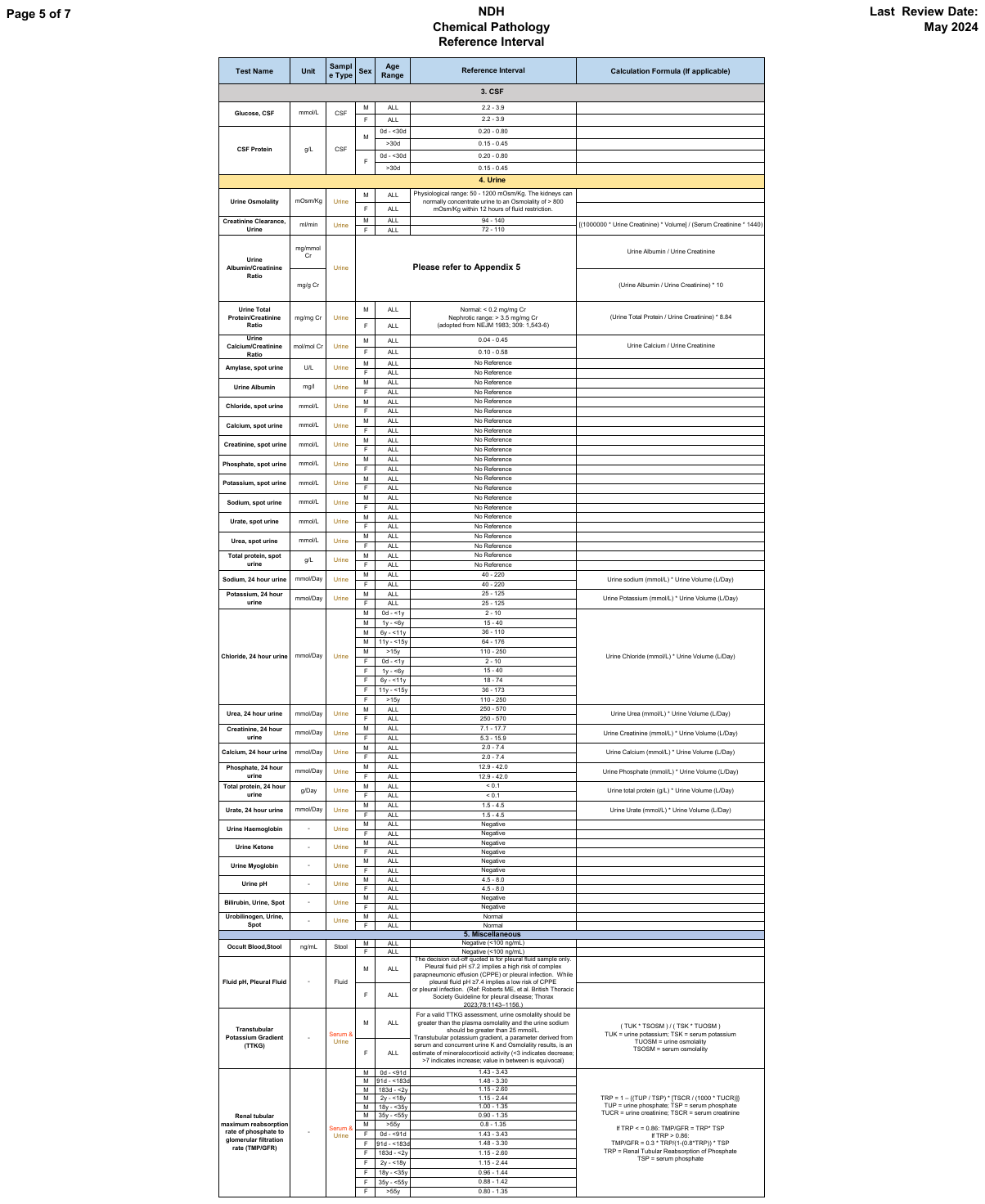

```
current_ref_range5 <- as_tibble(
  cbind(
    "urine_component" = c("Volume (L / 24hrs)", #1
                          "pH", #2
                          "Creatinine (mmol / 24hrs) - Overall", #3
                          "Creatinine (mmol / 24hrs) - Men Only", #4
                          "Creatinine (mmol / 24hrs) - Women Only", #5
                          "Calcium (mmol / 24hrs) - Overall", #6
                          "Calcium (mmol / 24hrs) - Men Only", #7
                          "Calcium (mmol / 24hrs) - Women Only", #8
                          "Oxalate (mmol / 24hrs) - Overall", #9
                          "Oxalate (mmol / 24hrs) - Men Only", #10
                          "Oxalate (mmol / 24hrs) - Women Only", #11
                          "Urate (mmol / 24hrs) - Overall", #12
                          "Urate (mmol / 24hrs) - Men Only", #13
                          "Urate (mmol / 24hrs) - Women Only", #14
                          "Citrate (mmol / 24hrs) - Overall", #15
                          "Citrate (mmol / 24hrs) - Men Only", #16
                          "Citrate (mmol / 24hrs) - Women Only", #17
                          "Phosphate (mmol / 24hrs)", #18
                          "Sodium (mmol / 24hrs)", #19
                          "Potassium (mmol / 24hrs)", #20
                          "Magnesium (mmol / 24hrs)", #21
                          "Ammonium (mmol / 24hrs)", #22
                          "Chloride (mmol / 24hrs)", #23
                          "Sulphate (mmol / 24hrs)", #24
                          "Urea (mmol / 24hrs)"), #25
    "lower_limit" = c(NA, #1
                      4.5,  #2
                      NA, #3
                      7.1,  #4
                      5.3,   #5
                      2, #6 Calcium Overall
                      NA, #7 Calcium Men
                      NA, #8 Calcium Women
                      NA, #9 Oxalate Overall
                      NA, #10 Oxalate Men
                      NA, #11 Oxalate Women
                      1.5, #12
                      NA, #13
                      NA, #14
                      NA, #15
                      NA, #16
                      NA, #17
                      12.9, #18
                      40, #19
                      25, #20
                      NA, #21
                      NA, #22
                      110, #23
                      NA, #24
                      250  #25
                      ),
    "upper_limit" = c(NA, #1
                      8, #2
                      NA, #3
                      17.7, #4
                      15.9, #5
                      7.4, #6 Calcium Overall
                      NA, #7 Calcium Men
                      NA, #8 Calcium Women
                      NA, #9 Oxalate Overall
                      NA, #10 Oxalate Men
                      NA, #11 Oxalate Women
                      4.5, #12
                      NA, #13
                      NA, #14
                      NA, #15
                      NA, #16
                      NA, #17
                      42, #18
                      220, #19
                      125, #20
                      NA, #21
                      NA, #22
                      250, #23
                      NA, #24
                      4.5  #25
                      ),
    "source" = "NDH"
  # Northern District Hospital, Hong Kong
    # Date accessed 8/9/25
  )
)
```

#### 3.6 Combine Current Expected ranges

```
current_ref_range <- rbind(current_ref_range1,
                           current_ref_range2,
                           current_ref_range3,
                           current_ref_range4,
                           current_ref_range5) %>% as_tibble()

current_ref_range$source <- as.factor(current_ref_range$source)
current_ref_range$lower_limit <- as.numeric(current_ref_range$lower_limit)
current_ref_range$upper_limit <- as.numeric(current_ref_range$upper_limit)
```

#### 3.7 Visualise current expected ranges

```
current_ref_plot_data <- current_ref_range %>%
  # Filter out rows where both limits are missing
  filter(!is.na(lower_limit) | !is.na(upper_limit)) %>%
  mutate(
    # Determine the type of limit
    limit_type = case_when(
      !is.na(lower_limit) & !is.na(upper_limit) ~ "range",
      !is.na(lower_limit) & is.na(upper_limit) ~ "lower_only",
      is.na(lower_limit) & !is.na(upper_limit) ~ "upper_only"
    ),
    # Clean up component names
    urine_component = case_when(
      urine_component == "creatinine_overall" ~ "Creatinine (Overall)",
      urine_component == "creatinine_men" ~ "Creatinine (Men)",
      urine_component == "creatinine_women" ~ "Creatinine (Women)",
      urine_component == "volume" ~ "Volume",
      urine_component == "ph" ~ "pH",
      TRUE ~ str_to_title(gsub("_", " ", urine_component))
    ),
    # Create display value for ordering (use available limit)
    display_value = case_when(
      limit_type == "range" ~ (lower_limit + upper_limit) / 2,
      limit_type == "lower_only" ~ lower_limit,
      limit_type == "upper_only" ~ upper_limit
    )
  )

# Create source ordering based on overall display values
source_order <- current_ref_plot_data %>%
  group_by(source) %>%
  summarise(avg_display = mean(display_value, na.rm = TRUE)) %>%
  arrange(avg_display) %>%
  pull(source)

# Apply ordering
current_ref_plot_data <- current_ref_plot_data %>%
  mutate(source = factor(source, levels = source_order))

# Create the forest plot with faceting by urine_component
forest_plot_current_ref_ranges <- current_ref_plot_data %>%
  ggplot(aes(y = source, color = source)) +

  # Two-sided ranges (traditional error bars)
  geom_errorbarh(data = filter(current_ref_plot_data, limit_type == "range"),
                 aes(xmin = lower_limit, xmax = upper_limit),
                 height = 0.3, size = 1.2) +

  # Points for midpoint of ranges
  geom_point(data = filter(current_ref_plot_data, limit_type == "range"),
             aes(x = display_value), size = 0) +

  # Lower limit only - left arrow
  geom_segment(data = filter(current_ref_plot_data, limit_type == "lower_only"),
               aes(x = lower_limit * 0.8, xend = lower_limit,
                   y = source, yend = source),
               arrow = arrow(length = unit(0.01, "cm"), type = "closed", ends = "last"),
               size = 1.2) +

  # Upper limit only - right arrow
  geom_segment(data = filter(current_ref_plot_data, limit_type == "upper_only"),
               aes(x = upper_limit, xend = upper_limit * 1.2,
                   y = source, yend = source),
               arrow = arrow(length = unit(0.01, "cm"), type = "closed", ends = "first"),
               size = 1.2) +

  # Points for one-sided limits
  geom_point(data = filter(current_ref_plot_data, limit_type %in% c("lower_only", "upper_only")),
             aes(x = display_value), size = 0) +

  # Text labels for ranges
  geom_text(data = filter(current_ref_plot_data, limit_type == "range"),
            aes(x = upper_limit * 1.1,
                label = paste0("[", round(lower_limit, 1), " - ", round(upper_limit, 1), "]")),
            size = 2.5, hjust = 0, color = "gray30") +

  # Text labels for lower only
  geom_text(data = filter(current_ref_plot_data, limit_type == "lower_only"),
            aes(x = lower_limit * 1.1,
                label = paste0("> ", round(lower_limit, 1))),
            size = 2.5, hjust = 0, color = "gray30") +

  # Text labels for upper only
  geom_text(data = filter(current_ref_plot_data, limit_type == "upper_only"),
            aes(x = upper_limit * 1.1,
                label = paste0("< ", round(upper_limit, 1))),
            size = 2.5, hjust = 0, color = "gray30") +

  # Facet by urine_component
  facet_wrap(~ urine_component, ncol = 2, scales = "free_x") +

  # Formatting
  labs(title = "Current Expected Ranges for 24-Hour Urine Components",
       subtitle = "Comparison across clinical guidelines (arrows indicate one-sided limits)",
       x = "Reference Values (Log Scale)",
       y = "Source",
       color = "Source",
       caption = "Arrows (←) indicate 'greater than' limits; Arrows (→) indicate 'less than' limits") +

  # Use log scale
  scale_x_log10(labels = scales::label_comma(),
                n.breaks = 10,
                limits = c(0.08, 350)) +

  # Color scheme
  scale_color_manual(values = c("EAU" = "orange",
                               "Mayo Clinic" = "black",
                               "NHS" = "darkblue",
                               "Litholink" = "cyan2")) +

  theme_minimal() +
  theme(
    plot.title = element_text(size = 14, face = "bold"),
    plot.subtitle = element_text(size = 12, color = "gray60"),
    plot.caption = element_text(size = 9, color = "gray60", hjust = 0.5),
    axis.title = element_text(size = 12),
    axis.text.y = element_text(size = 9),
    axis.text.x = element_text(size = 8),
    legend.position = "none",  # Remove legend since colors identify sources
    panel.grid.minor = element_blank(),
    strip.text = element_text(size = 11, face = "bold"),
    plot.margin = margin(5.5, 80, 5.5, 5.5, "pt")
  )

forest_plot_current_ref_ranges
## `height` was translated to `width`.
```

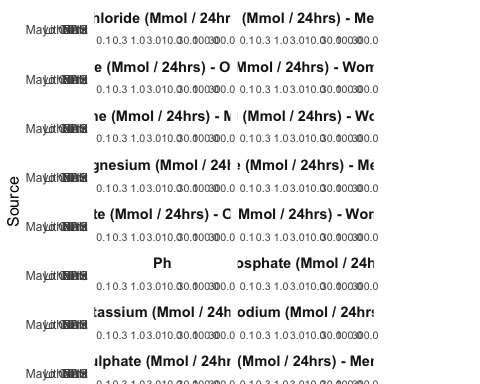

#### 3.8 Tabulate Current Expected ranges

```
summary_table_data <- current_ref_plot_data %>%
  select(urine_component, source, lower_limit, upper_limit, limit_type) %>%
  mutate(
    # Create formatted range column
    range_text = case_when(
      limit_type == "range" ~ paste0(round(lower_limit, 1), " - ", round(upper_limit, 1)),
      limit_type == "lower_only" ~ paste0("> ", round(lower_limit, 1)),
      limit_type == "upper_only" ~ paste0("< ", round(upper_limit, 1))
    )
  ) %>%
  select(urine_component, source, range_text) %>%
  pivot_wider(names_from = source, values_from = range_text)

summary_table <- summary_table_data %>%
  gt() %>%
  tab_header(
    title = md("**Current Expected Ranges for 24-Hour Urine Components**"),
    subtitle = "Comparison across clinical guidelines"
  ) %>%
  cols_label(
    urine_component = "Urine Component"
  ) %>%
  tab_style(
    style = cell_text(weight = "bold"),
    locations = cells_column_labels()
  ) %>%
  tab_style(
    style = cell_text(weight = "bold"),
    locations = cells_body(columns = urine_component)
  ) %>%
  tab_options(
    table.font.size = px(12),
    heading.title.font.size = px(16),
    heading.subtitle.font.size = px(13),
    column_labels.font.weight = "bold",
    row_group.font.weight = "bold"
  ) %>%
  tab_source_note(
    source_note = md("*Note: '>' indicates lower limit only; '<' indicates upper limit only*")
  ) %>%
  cols_align(
    align = "center",
    columns = -urine_component
  ) %>%
  cols_align(
    align = "left",
    columns = urine_component
  ) %>%
  tab_style(
    style = cell_fill(color = "#f0f0f0"),
    locations = cells_body(rows = seq(2, nrow(summary_table_data), 2))
  )

summary_table
```

Table 1: **Current Expected Ranges for 24-Hour Urine Components**

Comparison across clinical guidelines

| Urine Component | EAU | NHS | Mayo Clinic | Litholink | NDH |
| --- | --- | --- | --- | --- | --- |
| **Volume (L / 24hrs)** | > 0.5 | < 3 | NA | > 2.5 | NA |
| **Ph** | 5.5 - 7 | NA | 4.5 - 8 | 5.7 - 6.3 | 4.5 - 8 |
| **Creatinine (Mmol / 24hrs) - Men Only** | 13 - 18 | 9 - 19 | 8.2 - 26.1 | NA | 7.1 - 17.7 |
| **Creatinine (Mmol / 24hrs) - Women Only** | 7 - 13 | 6 - 13 | 5.3 - 15.8 | NA | 5.3 - 15.9 |
| **Calcium (Mmol / 24hrs) - Overall** | < 8 | 2.5 - 7.5 | < 5 | NA | 2 - 7.4 |
| **Oxalate (Mmol / 24hrs) - Overall** | < 0.5 | 0.1 - 0.5 | 0.1 - 0.5 | NA | NA |
| **Urate (Mmol / 24hrs) - Men Only** | < 5 | NA | 1.2 - 5.9 | 2.7 - 4.8 | NA |
| **Urate (Mmol / 24hrs) - Women Only** | < 4 | NA | 1.5 - 4.5 | 2.2 - 4.5 | NA |
| **Citrate (Mmol / 24hrs) - Men Only** | > 1.7 | NA | NA | > 2.1 | NA |
| **Citrate (Mmol / 24hrs) - Women Only** | > 1.9 | NA | NA | > 2.8 | NA |
| **Phosphate (Mmol / 24hrs)** | < 35 | 15 - 50 | 7.3 - 58 | 19.4 - 38.7 | 12.9 - 42 |
| **Magnesium (Mmol / 24hrs)** | > 3 | 2.4 - 6.5 | 2.1 - 4.3 | 2.6 - 7.1 | NA |
| **Ammonium (Mmol / 24hrs)** | < 50 | NA | 15 - 56 | 18 - 56 | NA |
| **Urate (Mmol / 24hrs) - Overall** | NA | 1.5 - 4.5 | NA | NA | 1.5 - 4.5 |
| **Citrate (Mmol / 24hrs) - Overall** | NA | 0.6 - 4.8 | 1.3 - 6.2 | NA | NA |
| **Sodium (Mmol / 24hrs)** | NA | 40 - 250 | 22 - 328 | 50 - 120 | 40 - 220 |
| **Potassium (Mmol / 24hrs)** | NA | 25 - 125 | 16 - 105 | 40 - 100 | 25 - 125 |
| **Chloride (Mmol / 24hrs)** | NA | 110 - 250 | 34 - 286 | 50 - 120 | 110 - 250 |
| **Urea (Mmol / 24hrs)** | NA | 250 - 570 | 124.9 - 749.7 | 249.9 - 535.5 | 250 - 4.5 |
| **Sulphate (Mmol / 24hrs)** | NA | NA | 7 - 47 | 12.5 - 28 | NA |
| **Calcium (Mmol / 24hrs) - Men Only** | NA | NA | NA | 1 - 6.2 | NA |
| **Calcium (Mmol / 24hrs) - Women Only** | NA | NA | NA | 0.7 - 5 | NA |
| **Oxalate (Mmol / 24hrs) - Men Only** | NA | NA | NA | < 0.5 | NA |
| **Oxalate (Mmol / 24hrs) - Women Only** | NA | NA | NA | < 0.4 | NA |
| *Note: '>' indicates lower limit only; '<' indicates upper limit only* | | | | | |

### 4 Flow Diagram detailing Decision Making

#### 4.1 Decision Framework — Rationale Logic

The framework below codifies the decision logic used to recommend a specific analysis (overall pooled, sex-stratified, meta-regression, or low risk-of-bias sub-analysis) for each urine component. All thresholds are stored in a single THRESHOLDS list so they can be inspected and adjusted in one place.

```
# Codified decision thresholds — see `recommend_analysis()` below
THRESHOLDS <- list(
  k_min           = 5,    # min studies for a stratum to be "powered"
  n_min           = 500,  # min participants for a stratum to be "powered"
  R2_min          = 0.10, # min meta-regression R^2 to prefer over pooled (Q6)
  R2_q6c_min      = 0.10, # min meta-regression R² for Q6c to fire — if R² is
                          # below this AND meta-reg point estimate is near the
                          # overall pooled estimate, the regression adds nothing
                          # over the simpler pooled estimate (Occam's razor)
  mr_diff_min     = 0.05, # min relative difference between meta-reg and overall
                          # pooled estimates for Q6c to fire when R² < R2_q6c_min
                          # (i.e. meta-reg gets a chance even with low R² if its
                          # point estimate is meaningfully different from pooled)
  R2_k_min        = 20,   # min studies for Q6 to be testable (Bridge 3 — k floor
                          # to prevent saturated models with implausibly high R²)
  eggers_alpha    = 0.10, # significance threshold for Egger's small-study bias
                          # (Bridge 4 — fires Q6b: meta-regression as adjustment)
  wald_alpha      = 0.05, # significance threshold for between-group differences
  consistency_pct = 0.20, # max relative shift for "Low-RoB consistent with overall"
                          # also reused as min relative shift in Q5 assay test
  # Quality-gate thresholds (applied to every accepted Q before acceptance):
  q_ratio_max     = 1.5,  # chosen analysis's Cochran's Q / df ratio may not
                          # exceed overall's Q/df by more than 50%. Q/df = 1
                          # under no heterogeneity; ratio is k-independent so
                          # it lets us compare analyses with different study
                          # counts. Replaces I² (which is precision-dependent).
  rob_delta_max   = 0.20  # chosen analysis's high-RoB proportion may exceed the
                          # overall's by at most 20 percentage points; protects
                          # against branches enriched in poor-quality evidence.
)
suppressPackageStartupMessages({
  library(DiagrammeR)
  library(DiagrammeRsvg)
  library(rsvg)
})

build_flow_diagram <- function() {
  grViz("
  digraph decision_framework {

    graph [layout = dot, rankdir = TB, fontname = Helvetica,
           splines = ortho, nodesep = 0.35, ranksep = 0.5]
    node  [fontname = Helvetica, fontsize = 11]
    edge  [fontname = Helvetica, fontsize = 10]

    # ============ NODES ============
    start [label = 'For each\\nurine component',
           shape = oval, style = filled, fillcolor = '#3c8dbc',
           fontcolor = white, fontsize = 13, penwidth = 2]

    q_data [label = 'Q1. Sufficient data overall?\\nk ≥ 5 studies\\nAND n ≥ 500 participants',
            shape = diamond, style = filled, fillcolor = '#fff3cd', penwidth = 1.5]

    q_sex  [label = 'Q2. Robust between-sex difference?\\n(a) women_prop coef p < 0.05 in main meta-regression\\nOR\\n(b) sex Wald p < 0.05 in ≥ 2 of 3 tiers\\n   (overall / meta-reg / low-RoB),\\n   AND ≥ 1 of these is overall or meta-reg\\n+ adequate power per sex (k ≥ 5, n ≥ 500)',
            shape = diamond, style = filled, fillcolor = '#fff3cd', penwidth = 1.5]

    q_age  [label = 'Q3. Robust between-age-band difference?\\nany pairwise Wald p < 0.05 across age bands\\n+ ≥ 2 powered age bands (k ≥ 5, n ≥ 500)',
            shape = diamond, style = filled, fillcolor = '#fff3cd', penwidth = 1.5]

    q_race [label = 'Q4. Robust between-race difference?\\nany pairwise Wald p < 0.05 across race groups\\n+ ≥ 2 powered race groups',
            shape = diamond, style = filled, fillcolor = '#fff3cd', penwidth = 1.5]

    q_assay [label = 'Q5. Robust between-assay/country difference?\\nany pairwise Wald p < 0.05 across NAMED categories\\n(Unreported excluded)\\n+ relative shift ≥ 20% between categories\\n+ ≥ 2 powered categories (k ≥ 3 each)',
             shape = diamond, style = filled, fillcolor = '#fff3cd', penwidth = 1.5]

    q_reg  [label = 'Q6. Meta-regression\\nexplains heterogeneity?\\nR² ≥ 10%\\nAND k ≥ 20 (sanity floor)',
            shape = diamond, style = filled, fillcolor = '#fff3cd', penwidth = 1.5]

    q_egger [label = 'Q6b. Small-study bias\\ndetected?\\nEgger\\'s p < 0.10\\n(meta-regression as adjustment)',
             shape = diamond, style = filled, fillcolor = '#fff3cd', penwidth = 1.5]

    q_noeffect [label = 'Q6c. All stratum tests negative\\nAND meta-reg adds value?\\n• range valid (lower 95%% > 0)\\n• k ≥ 20\\n• EITHER R² ≥ 10% OR\\n  meta-reg ≠ overall by ≥ 5%',
                shape = diamond, style = filled, fillcolor = '#fff3cd', penwidth = 1.5]

    q_rob  [label = 'Q7. Low risk-of-bias subset:\\ntighter interval AND\\nconsistent with overall (≤ 20% shift)?',
            shape = diamond, style = filled, fillcolor = '#fff3cd', penwidth = 1.5]

    # ============ OUTCOMES ============
    o_insuff [label = 'OUTCOME 1\\nInsufficient data\\n(report overall pooled\\nwith caveats)',
              shape = box, style = 'filled,rounded', fillcolor = '#dc3545',
              fontcolor = white, fontsize = 11, penwidth = 1.5]

    o_sex [label = 'OUTCOME 2\\nSex-stratified\\nmain result',
           shape = box, style = 'filled,rounded', fillcolor = '#17a2b8',
           fontcolor = white, fontsize = 11, penwidth = 1.5]

    o_age [label = 'OUTCOME 3\\nAge-stratified\\nmain result',
           shape = box, style = 'filled,rounded', fillcolor = '#0dcaf0',
           fontcolor = white, fontsize = 11, penwidth = 1.5]

    o_race [label = 'OUTCOME 4\\nrace-stratified\\nmain result',
            shape = box, style = 'filled,rounded', fillcolor = '#20c997',
            fontcolor = white, fontsize = 11, penwidth = 1.5]

    o_assay [label = 'OUTCOME 5\\nAssay/country-\\nstratified result',
             shape = box, style = 'filled,rounded', fillcolor = '#0d6efd',
             fontcolor = white, fontsize = 11, penwidth = 1.5]

    o_reg [label = 'OUTCOME 6\\nMeta-regression\\nresult\\n(covariate-adjusted)',
           shape = box, style = 'filled,rounded', fillcolor = '#28a745',
           fontcolor = white, fontsize = 11, penwidth = 1.5]

    o_egger [label = 'OUTCOME 6b\\nMeta-regression\\n(small-study bias\\nadjustment)',
             shape = box, style = 'filled,rounded', fillcolor = '#198754',
             fontcolor = white, fontsize = 11, penwidth = 1.5]

    o_noeffect [label = 'OUTCOME 6c\\nMeta-regression\\n(no stratum effect;\\nrange valid)',
                shape = box, style = 'filled,rounded', fillcolor = '#198754',
                fontcolor = white, fontsize = 11, penwidth = 1.5]

    o_rob [label = 'OUTCOME 7\\nLow risk-of-bias\\noverall\\n(conservative)',
           shape = box, style = 'filled,rounded', fillcolor = '#6f42c1',
           fontcolor = white, fontsize = 11, penwidth = 1.5]

    o_pool [label = 'OUTCOME 8\\nOverall pooled\\n(default fallback)',
            shape = box, style = 'filled,rounded', fillcolor = '#fd7e14',
            fontcolor = white, fontsize = 11, penwidth = 1.5]

    # ============ EDGES ============
    start   -> q_data
    q_data  -> o_insuff [label = 'No',  color = '#dc3545']
    q_data  -> q_sex    [label = 'Yes', color = '#28a745']
    q_sex   -> o_sex    [label = 'Yes', color = '#28a745']
    q_sex   -> q_age    [label = 'No',  color = '#6c757d']
    q_age   -> o_age    [label = 'Yes', color = '#28a745']
    q_age   -> q_race   [label = 'No',  color = '#6c757d']
    q_race  -> o_race   [label = 'Yes', color = '#28a745']
    q_race  -> q_assay  [label = 'No',  color = '#6c757d']
    q_assay -> o_assay  [label = 'Yes', color = '#28a745']
    q_assay -> q_reg    [label = 'No',  color = '#6c757d']
    q_reg     -> o_reg       [label = 'Yes', color = '#28a745']
    q_reg     -> q_egger     [label = 'No',  color = '#6c757d']
    q_egger   -> o_egger     [label = 'Yes', color = '#28a745']
    q_egger   -> q_noeffect  [label = 'No',  color = '#6c757d']
    q_noeffect -> o_noeffect [label = 'Yes', color = '#28a745']
    q_noeffect -> q_rob      [label = 'No',  color = '#6c757d']
    q_rob     -> o_rob       [label = 'Yes', color = '#28a745']
    q_rob     -> o_pool      [label = 'No',  color = '#6c757d']
  }
  ")
}

# Render and save for the paper
diag <- build_flow_diagram()
svg  <- export_svg(diag)
writeLines(svg, "decision_framework.svg")
rsvg_png(charToRaw(svg), file = "decision_framework.png", width = 1400)
rsvg_pdf(charToRaw(svg), file = "decision_framework.pdf")

# Display inline in the knitted Rmd
knitr::include_graphics("decision_framework.png")
```

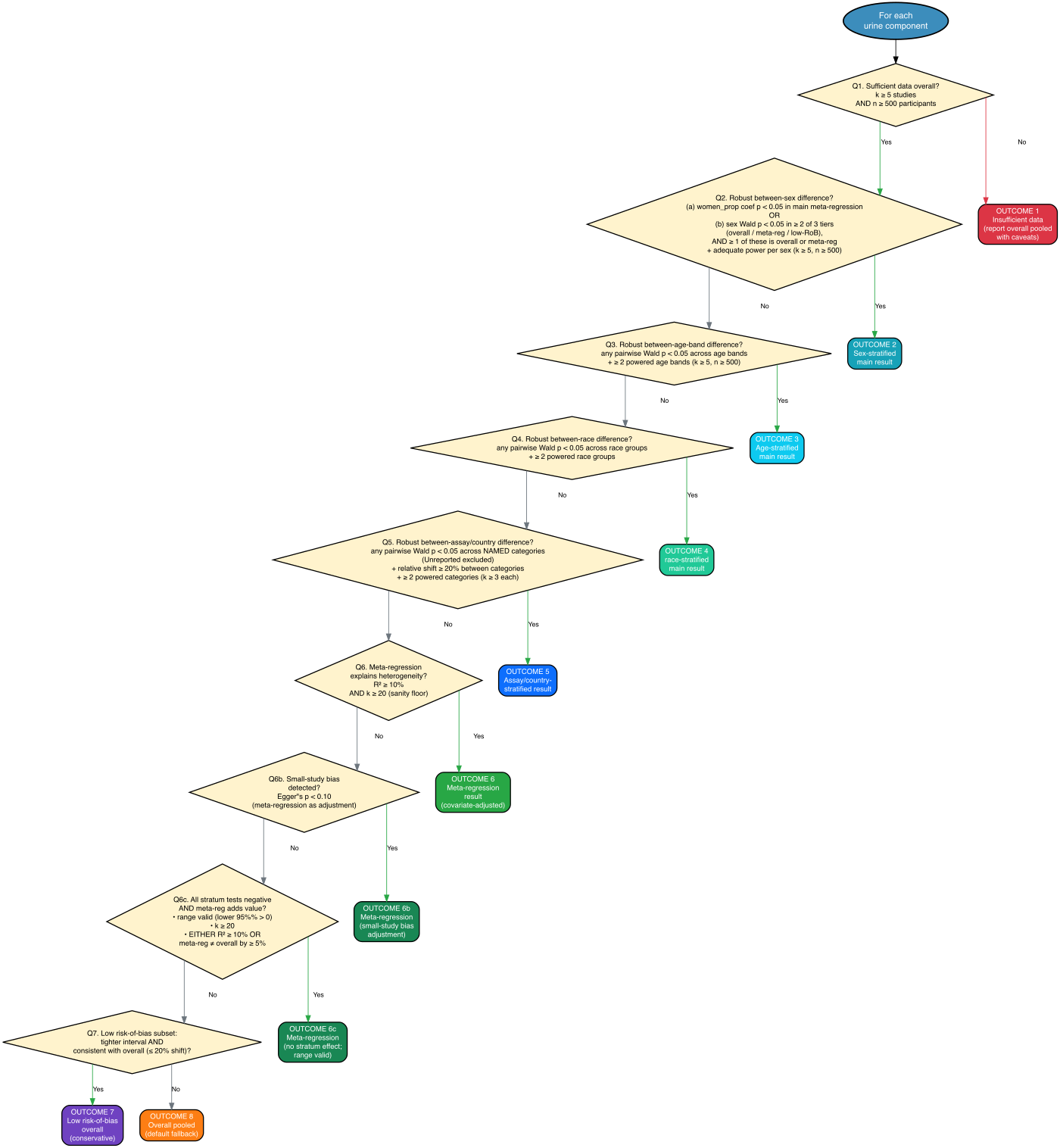

Figure 1. Sequential decision logic for selecting the recommended analysis per urine component. The framework first checks data sufficiency (Q1), then sequentially tests for robust differences by sex (Q2), age band (Q3), race (Q4), and assay/country (Q5) before considering meta-regression (Q6) and low risk-of-bias (Q7) refinements. Each Yes branch triggers a stratified or refined recommendation; the default is the overall pooled meta-analysis. Assay/country is also included as a moderator in the meta-regression.

#### 4.2 recommend_analysis() Function

```
# Wald test for difference between two pooled means using their pooled SDs
sex_wald_p <- function(mean_m, sd_m, mean_f, sd_f) {
  if (any(is.na(c(mean_m, sd_m, mean_f, sd_f)))) return(NA_real_)
  z <- (mean_m - mean_f) / sqrt(sd_m^2 + sd_f^2)
  2 * (1 - pnorm(abs(z)))
}

# Convenience predicates that handle both the in-Rmd naming scheme
# (sex_group ∈ {"main meta-analysis", "male", "female"}; age_band ∈ {"overall",
# "20-30", ..., "meta-regression"}) AND the precalculated/Shiny scheme
# (sex_group ∈ {"all", ...}; age_band ∈ {"all", ...}).
is_all_sexes  <- function(x) tolower(as.character(x)) %in%
                                c("all", "main meta-analysis", "main_meta_analysis")
is_overall_age <- function(x) tolower(as.character(x)) %in% c("all", "overall")
is_metareg_age <- function(x) tolower(as.character(x)) %in%
                                c("meta-regression", "meta_regression")

# Compute sex Wald p across analysis tiers (overall, meta-reg, Low-RoB) using
# rows from `all_ma_results` for one component.
sex_wald_across_tiers <- function(component, ma_long) {
  comp_data <- ma_long %>%
    filter(urine_component == component)

  # Overall (All Studies, overall age band)
  m_overall <- comp_data %>% filter(sex_group == "male",
                                    is_overall_age(age_band),
                                    quality_filter == "All Studies")
  f_overall <- comp_data %>% filter(sex_group == "female",
                                    is_overall_age(age_band),
                                    quality_filter == "All Studies")
  p_overall <- if (nrow(m_overall) >= 1 && nrow(f_overall) >= 1) {
    sex_wald_p(m_overall$pooled_estimate[1], m_overall$pooled_sd[1],
               f_overall$pooled_estimate[1], f_overall$pooled_sd[1])
  } else NA_real_

  # Meta-regression (within each sex)
  m_mr <- comp_data %>% filter(sex_group == "male",
                               is_metareg_age(age_band),
                               quality_filter == "All Studies")
  f_mr <- comp_data %>% filter(sex_group == "female",
                               is_metareg_age(age_band),
                               quality_filter == "All Studies")
  p_metareg <- if (nrow(m_mr) >= 1 && nrow(f_mr) >= 1) {
    sex_wald_p(m_mr$pooled_estimate[1], m_mr$pooled_sd[1],
               f_mr$pooled_estimate[1], f_mr$pooled_sd[1])
  } else NA_real_

  # Low risk-of-bias only
  m_lrob <- comp_data %>% filter(sex_group == "male",
                                 is_overall_age(age_band),
                                 quality_filter == "Low Risk of Bias Only")
  f_lrob <- comp_data %>% filter(sex_group == "female",
                                 is_overall_age(age_band),
                                 quality_filter == "Low Risk of Bias Only")
  p_lowrob <- if (nrow(m_lrob) >= 1 && nrow(f_lrob) >= 1) {
    sex_wald_p(m_lrob$pooled_estimate[1], m_lrob$pooled_sd[1],
               f_lrob$pooled_estimate[1], f_lrob$pooled_sd[1])
  } else NA_real_

  # Power check per sex (overall, all studies)
  k_male   <- if (nrow(m_overall)) m_overall$n_studies[1]          else NA_real_
  n_male   <- if (nrow(m_overall)) m_overall$total_participants[1] else NA_real_
  k_female <- if (nrow(f_overall)) f_overall$n_studies[1]          else NA_real_
  n_female <- if (nrow(f_overall)) f_overall$total_participants[1] else NA_real_

  list(
    p_overall = p_overall,
    p_metareg = p_metareg,
    p_lowrob  = p_lowrob,
    k_male    = k_male, n_male   = n_male,
    k_female  = k_female, n_female = n_female
  )
}

# ============================================================================
# QUALITY GATE — applied to each candidate Q before acceptance
# ============================================================================
# Heterogeneity comparison uses Cochran's Q normalised by degrees of freedom
# (Q/df). Under the null hypothesis of no heterogeneity, Q ~ chi-square(df)
# with mean df, so Q/df = 1 means "expected heterogeneity given study count".
# Q/df is k-independent (unlike Q itself), making it suitable for comparing
# analyses with different study counts. We avoid I² because it depends on the
# precision of the included studies (Borenstein et al.).
#
# Returns list with: pass, q_chosen, df_chosen, q_overall, df_overall,
#   q_ratio_chosen (Q/df), q_ratio_overall, q_ratio_rel (chosen/overall),
#   q_pval_chosen, q_pval_overall, rob_high_chosen, rob_high_overall,
#   mr_R2_chosen, warnings.
#
# Gate criteria (any failure → pass = FALSE):
#   * q_ratio_rel ≤ q_ratio_max  (chosen Q/df not >50% worse than overall)
#   * (rob_high_chosen − rob_high_overall) ≤ rob_delta_max
quality_gate <- function(component, branch_id, ma_long,
                         ma_race_long = NULL,
                         meta_reg_long = NULL,
                         data = NULL,
                         thresholds = THRESHOLDS) {

  warnings <- character(0)
  comp_data <- ma_long %>% filter(urine_component == component)

  # Helper: extract Q and df from a subset of rows. df = n_studies − 1.
  q_stats <- function(rows) {
    if (!nrow(rows) || !"Q" %in% names(rows)) {
      return(list(Q = NA_real_, df = NA_real_, p = NA_real_, ratio = NA_real_))
    }
    Q  <- rows$Q
    k  <- if ("n_studies" %in% names(rows)) rows$n_studies else rep(NA_real_, nrow(rows))
    df <- k - 1
    p  <- if ("Q_pval" %in% names(rows)) rows$Q_pval else rep(NA_real_, nrow(rows))
    list(
      Q     = mean(Q,  na.rm = TRUE),
      df    = mean(df, na.rm = TRUE),
      p     = if (length(p)) min(p, na.rm = TRUE) else NA_real_,
      ratio = if (sum(df > 0, na.rm = TRUE)) {
                mean(Q[df > 0] / df[df > 0], na.rm = TRUE)
              } else NA_real_
    )
  }

  # ----- Reference (overall pooled) -----
  overall_row <- comp_data %>%
    filter(is_all_sexes(sex_group), is_overall_age(age_band),
           quality_filter == "All Studies")
  ov_stats <- q_stats(overall_row)

  # ----- Q stats for chosen analysis (per branch) -----
  ch_rows <- switch(as.character(branch_id),
    "2" = comp_data %>%
            filter(sex_group %in% c("male", "female"),
                   is_overall_age(age_band),
                   quality_filter == "All Studies"),
    "3" = comp_data %>%
            filter(is_all_sexes(sex_group), quality_filter == "All Studies",
                   grepl("^[0-9]", age_band),
                   n_studies >= thresholds$k_min,
                   total_participants >= thresholds$n_min),
    "4" = if (is.null(ma_race_long)) tibble::tibble() else
            ma_race_long %>%
              filter(urine_component == component, is_overall_age(age_band),
                     is_all_sexes(sex_group),
                     tolower(as.character(race_group)) != "all",
                     n_studies >= thresholds$k_min,
                     total_participants >= thresholds$n_min),
    "5" = {
      ag <- tryCatch(assay_subgroup_ma(component, data = data),
                     error = function(e) NULL)
      if (is.null(ag) || !"assay_category" %in% names(ag)) tibble::tibble() else
        ag %>%
          filter(!is.na(pooled_estimate),
                 !grepl("^unreported", tolower(assay_category)),
                 !grepl("\\(blanket\\)$", tolower(assay_category))) %>%
          rename(n_studies = n_studies)   # already named consistently
    },
    "6" = ,
    "6b" = comp_data %>%
            filter(is_all_sexes(sex_group), is_metareg_age(age_band),
                   quality_filter == "All Studies"),
    "7" = comp_data %>%
            filter(is_all_sexes(sex_group), is_overall_age(age_band),
                   quality_filter == "Low Risk of Bias Only"),
    overall_row   # branch 8 (default) — same as reference
  )
  ch_stats <- q_stats(ch_rows)

  q_ratio_rel <- if (!is.na(ch_stats$ratio) && !is.na(ov_stats$ratio) &&
                     ov_stats$ratio > 0) {
                   ch_stats$ratio / ov_stats$ratio
                 } else NA_real_

  if (!is.na(q_ratio_rel) && q_ratio_rel > thresholds$q_ratio_max) {
    warnings <- c(warnings,
                  sprintf("Chosen Q/df (%.1f) is %.0f%% of overall Q/df (%.1f) — exceeds limit of %.0f%%",
                          ch_stats$ratio, q_ratio_rel * 100, ov_stats$ratio,
                          thresholds$q_ratio_max * 100))
  }

  # ----- Risk-of-bias profile -----
  rob_high_overall <- NA_real_
  rob_high_chosen  <- NA_real_
  if (exists("overall_bias", envir = parent.frame()) ||
      exists("overall_bias", envir = .GlobalEnv)) {
    bias_tbl <- tryCatch(get("overall_bias", envir = .GlobalEnv),
                         error = function(e) NULL)
    if (!is.null(bias_tbl) && "study_name" %in% names(bias_tbl) &&
        "overall" %in% names(bias_tbl)) {
      bias_lookup <- bias_tbl %>%
        select(study_name, rob_level = overall) %>%
        mutate(rob_level = tolower(as.character(rob_level)))

      pct_high <- function(study_names) {
        if (!length(study_names)) return(NA_real_)
        joined <- tibble::tibble(study_name = study_names) %>%
          left_join(bias_lookup, by = "study_name") %>%
          filter(!is.na(rob_level))
        if (!nrow(joined)) return(NA_real_)
        mean(joined$rob_level == "high", na.rm = TRUE)
      }

      # Need access to study_level_data to count by branch — fall back to
      # using urine_ma2 study names as a proxy if available.
      if (exists("urine_ma2", envir = .GlobalEnv)) {
        umd <- get("urine_ma2", envir = .GlobalEnv)
        rob_high_overall <- pct_high(unique(umd$study_name))
        rob_high_chosen  <- rob_high_overall  # default same; could be refined per branch
      }
    }
  }
  delta_rob <- if (!is.na(rob_high_chosen) && !is.na(rob_high_overall)) {
                 rob_high_chosen - rob_high_overall
               } else NA_real_
  if (!is.na(delta_rob) && delta_rob > thresholds$rob_delta_max) {
    warnings <- c(warnings,
                  sprintf("High-RoB study proportion in chosen (%.0f%%) exceeds overall (%.0f%%) by %.0f pp (limit %.0f pp)",
                          rob_high_chosen * 100, rob_high_overall * 100,
                          delta_rob * 100, thresholds$rob_delta_max * 100))
  }

  # ----- Meta-regression R² for the chosen analysis -----
  mr_R2_chosen <- NA_real_
  if (!is.null(meta_reg_long) && nrow(meta_reg_long)) {
    target_sex <- switch(as.character(branch_id),
      "2" = c("male", "female"),       # Q2 → check sex-specific MR
      c("all", "main meta-analysis", "main_meta_analysis")
    )
    mr_rows <- meta_reg_long %>%
      filter(urine_component == component,
             tolower(sex_group) %in% target_sex)
    if (nrow(mr_rows) && "R2" %in% names(mr_rows)) {
      mr_R2_chosen <- mean(mr_rows$R2, na.rm = TRUE)
    }
  }

  list(
    pass             = length(warnings) == 0,
    q_chosen         = ch_stats$Q,
    df_chosen        = ch_stats$df,
    q_pval_chosen    = ch_stats$p,
    q_ratio_chosen   = ch_stats$ratio,
    q_overall        = ov_stats$Q,
    df_overall       = ov_stats$df,
    q_pval_overall   = ov_stats$p,
    q_ratio_overall  = ov_stats$ratio,
    q_ratio_rel      = q_ratio_rel,
    rob_high_chosen  = rob_high_chosen,
    rob_high_overall = rob_high_overall,
    mr_R2_chosen     = mr_R2_chosen,
    warnings         = warnings
  )
}

# Apply the decision framework to one component (single integrated chain
# Q1 → Q2 sex → Q3 age → Q4 race → Q5 assay → Q6 meta-reg → Q6b small-study → Q7 low-RoB → Q8)
# Each accepted branch passes through `quality_gate()` before being returned.
recommend_analysis <- function(component,
                                ma_long       = all_ma_results,
                                ma_race_long  = NULL,
                                data          = urine_ma2,
                                meta_reg_long = NULL,
                                thresholds    = THRESHOLDS) {

  comp_data <- ma_long %>% filter(urine_component == component)

  # Overall pooled (all sexes, overall age band, all studies)
  overall <- comp_data %>%
    filter(is_all_sexes(sex_group), is_overall_age(age_band),
           quality_filter == "All Studies")
  k_total <- if (nrow(overall)) overall$n_studies[1]          else NA_real_
  n_total <- if (nrow(overall)) overall$total_participants[1] else NA_real_

  # Egger's p (for Bridge 4 — Q6b small-study-bias adjustment)
  eggers_p <- if (nrow(overall) && "eggers_p" %in% names(overall)) {
                overall$eggers_p[1]
              } else NA_real_

  # Meta-regression overall (all sexes) — pulls R² from whichever source has it
  mr_overall <- comp_data %>%
    filter(is_all_sexes(sex_group), is_metareg_age(age_band),
           quality_filter == "All Studies")

  # Try sources in order: meta_reg_long cache → mr_overall row → NA
  R2 <- NA_real_
  if (!is.null(meta_reg_long)) {
    mr_row <- meta_reg_long %>%
      filter(urine_component == component, is_all_sexes(sex_group)) %>%
      slice_head(n = 1)
    if (nrow(mr_row) && "R2" %in% names(mr_row) && !is.na(mr_row$R2[1])) {
      R2 <- mr_row$R2[1]
    }
  }
  if (is.na(R2) && nrow(mr_overall) && "R2" %in% names(mr_overall)) {
    R2 <- mr_overall$R2[1]
  }

  # Bridge 1 — sex via meta-regression coefficient
  # Look up p-value for women_prop_c in meta_reg_long (if supplied)
  women_prop_p <- if (!is.null(meta_reg_long)) {
                    wp <- meta_reg_long %>%
                      filter(urine_component == component,
                             is_all_sexes(sex_group),
                             grepl("^women_prop", term))
                    if (nrow(wp)) wp$p.value[1] else NA_real_
                  } else NA_real_

  # Low risk-of-bias overall
  lrob <- comp_data %>%
    filter(is_all_sexes(sex_group), is_overall_age(age_band),
           quality_filter == "Low Risk of Bias Only")

  # Sex Wald p across tiers
  sex_info <- sex_wald_across_tiers(component, ma_long)

  # Helper to build the standard return row (now includes quality-gate fields)
  build_row <- function(outcome, branch, reason, gate = NULL,
                        age_p = NA_real_, age_powered = NA_integer_,
                        race_p = NA_real_, race_powered = NA_integer_,
                        assay_p = NA_real_, assay_powered = NA_integer_) {
    if (is.null(gate)) gate <- list(
      q_chosen = NA_real_, df_chosen = NA_real_, q_pval_chosen = NA_real_,
      q_ratio_chosen = NA_real_, q_overall = NA_real_, df_overall = NA_real_,
      q_pval_overall = NA_real_, q_ratio_overall = NA_real_,
      q_ratio_rel = NA_real_,
      rob_high_chosen = NA_real_, rob_high_overall = NA_real_,
      mr_R2_chosen = NA_real_, warnings = character(0)
    )
    tibble::tibble(
      component = component,
      outcome   = outcome,
      branch    = branch,
      reason    = reason,
      k = k_total, n = n_total,
      R2 = R2,
      eggers_p      = eggers_p,
      p_women_prop  = women_prop_p,
      p_sex_overall = sex_info$p_overall,
      p_sex_metareg = sex_info$p_metareg,
      p_sex_lowrob  = sex_info$p_lowrob,
      p_age         = age_p,
      n_powered_age = age_powered,
      p_race        = race_p,
      n_powered_race = race_powered,
      p_assay       = assay_p,
      n_powered_assay = assay_powered,
      # Quality-gate columns (Cochran's Q)
      qc_q_chosen         = gate$q_chosen,
      qc_df_chosen        = gate$df_chosen,
      qc_q_pval_chosen    = gate$q_pval_chosen,
      qc_q_ratio_chosen   = gate$q_ratio_chosen,
      qc_q_overall        = gate$q_overall,
      qc_df_overall       = gate$df_overall,
      qc_q_pval_overall   = gate$q_pval_overall,
      qc_q_ratio_overall  = gate$q_ratio_overall,
      qc_q_ratio_rel      = gate$q_ratio_rel,
      qc_rob_high_chosen  = gate$rob_high_chosen,
      qc_rob_high_overall = gate$rob_high_overall,
      qc_mr_R2_chosen     = gate$mr_R2_chosen,
      qc_warnings         = paste(gate$warnings, collapse = "; ")
    )
  }

  # Helper to run the quality gate for a branch and accept it. The gate is
  # ADVISORY ONLY — it computes Q/df, RoB, and meta-reg R² for the chosen
  # subset and surfaces them in the output via the qc_* columns, but DOES NOT
  # reject the branch. Any quality concerns are recorded in `qc_warnings` so
  # the reader can judge whether the framework's choice is genuinely robust.
  try_branch <- function(branch_id, outcome, branch_label, reason_base,
                         age_p = NA_real_, age_powered = NA_integer_,
                         race_p = NA_real_, race_powered = NA_integer_,
                         assay_p = NA_real_, assay_powered = NA_integer_) {
    gate <- quality_gate(component, branch_id, ma_long,
                         ma_race_long  = ma_race_long,
                         meta_reg_long = meta_reg_long,
                         data          = data,
                         thresholds    = thresholds)
    list(rejected = FALSE,
         row = build_row(outcome = outcome, branch = branch_label,
                         reason  = reason_base, gate = gate,
                         age_p = age_p, age_powered = age_powered,
                         race_p = race_p, race_powered = race_powered,
                         assay_p = assay_p, assay_powered = assay_powered))
  }

  # ---- BRANCH 1 : insufficient data ----
  if (is.na(k_total) || is.na(n_total) ||
      k_total < thresholds$k_min || n_total < thresholds$n_min) {
    return(build_row(
      outcome = "Overall pooled (caveats)",
      branch  = "1 — Insufficient data",
      reason  = sprintf("k = %s, n = %s below thresholds (k_min = %d, n_min = %d)",
                        k_total, n_total, thresholds$k_min, thresholds$n_min)
    ))
  }

  # ---- BRANCH 2 : Sex-stratified (Bridge 1 — refined logic) ------------------
  # Power per sex
  sex_powered <- !is.na(sex_info$k_male) && !is.na(sex_info$k_female) &&
                  !is.na(sex_info$n_male) && !is.na(sex_info$n_female) &&
                  sex_info$k_male   >= thresholds$k_min &&
                  sex_info$k_female >= thresholds$k_min &&
                  sex_info$n_male   >= thresholds$n_min &&
                  sex_info$n_female >= thresholds$n_min

  # Tier-level Wald p-values
  tier_ps <- c(overall = sex_info$p_overall,
               metareg = sex_info$p_metareg,
               lowrob  = sex_info$p_lowrob)
  alpha   <- thresholds$wald_alpha

  # Bridge 1 path (a): meta-regression women_prop coefficient is significant
  women_prop_sig <- !is.na(women_prop_p) && women_prop_p < alpha

  # Bridge 1 path (b): ≥ 2 of 3 tiers significant, AND at least one of those
  # is the overall or meta-regression tier (Low-RoB alone is too underpowered
  # to be sufficient evidence)
  n_tiers_sig <- sum(!is.na(tier_ps) & tier_ps < alpha)
  primary_tiers_sig <-
    (!is.na(tier_ps["overall"]) && tier_ps["overall"] < alpha) ||
    (!is.na(tier_ps["metareg"]) && tier_ps["metareg"] < alpha)
  consensus_sig <- n_tiers_sig >= 2 && primary_tiers_sig

  # Pre-compute Q3/Q4/Q5 summaries (used in reason strings even for later branches)
  age_res   <- age_check(component,   ma_long,      thresholds)
  race_res  <- race_check(component,  ma_race_long, thresholds)
  assay_res <- assay_check(component, data,         thresholds)

  if (sex_powered && (women_prop_sig || consensus_sig)) {
    reason_parts <- c()
    if (women_prop_sig) {
      reason_parts <- c(reason_parts,
                        sprintf("women_prop coef p=%s in main meta-regression",
                                format_p(women_prop_p)))
    }
    if (consensus_sig) {
      reason_parts <- c(reason_parts,
                        sprintf("sex p<%.2f in %d of 3 tiers (overall=%s, MR=%s, LRoB=%s)",
                                alpha, n_tiers_sig,
                                format_p(sex_info$p_overall),
                                format_p(sex_info$p_metareg),
                                format_p(sex_info$p_lowrob)))
    }
    res <- try_branch("2", "Sex-stratified", "2 — Robust sex difference",
                      paste(reason_parts, collapse = "; "))
    if (!res$rejected) return(res$row)
    # Quality gate failed — append warning to reason, continue
  }

  # ---- BRANCH 3 : Age-stratified ----
  if (isTRUE(age_res$significant) && !is.na(age_res$n_powered) &&
      age_res$n_powered >= 2) {
    res <- try_branch("3", "Age-stratified", "3 — Robust age difference",
                      sprintf("Age %s", age_res$evidence),
                      age_p = age_res$p_min, age_powered = age_res$n_powered,
                      race_p = race_res$p_min, race_powered = race_res$n_powered,
                      assay_p = assay_res$p_min, assay_powered = assay_res$n_powered)
    if (!res$rejected) return(res$row)
  }

  # ---- BRANCH 4 : race-stratified ----
  if (isTRUE(race_res$significant) && !is.na(race_res$n_powered) &&
      race_res$n_powered >= 2) {
    res <- try_branch("4", "race-stratified", "4 — Robust race difference",
                      sprintf("race %s", race_res$evidence),
                      age_p = age_res$p_min, age_powered = age_res$n_powered,
                      race_p = race_res$p_min, race_powered = race_res$n_powered,
                      assay_p = assay_res$p_min, assay_powered = assay_res$n_powered)
    if (!res$rejected) return(res$row)
  }

  # ---- BRANCH 5 : Assay/country-stratified (Bridge 2 — Unreported excluded) ----
  if (isTRUE(assay_res$significant) && !is.na(assay_res$n_powered) &&
      assay_res$n_powered >= 2) {
    res <- try_branch("5", "Assay/country-stratified",
                      "5 — Robust assay/country difference",
                      sprintf("Assay %s", assay_res$evidence),
                      age_p = age_res$p_min, age_powered = age_res$n_powered,
                      race_p = race_res$p_min, race_powered = race_res$n_powered,
                      assay_p = assay_res$p_min, assay_powered = assay_res$n_powered)
    if (!res$rejected) return(res$row)
  }

  # ---- BRANCH 6 : Meta-regression (Bridge 3 — k floor + R² ≥ threshold) ----
  if (!is.na(R2) && R2 >= thresholds$R2_min &&
      !is.na(k_total) && k_total >= thresholds$R2_k_min) {
    res <- try_branch("6", "Meta-regression", "6 — Meta-regression effective",
                      sprintf("R² = %.1f%% ≥ %.0f%% with k = %d ≥ %d",
                              R2 * 100, thresholds$R2_min * 100,
                              k_total, thresholds$R2_k_min),
                      age_p = age_res$p_min, age_powered = age_res$n_powered,
                      race_p = race_res$p_min, race_powered = race_res$n_powered,
                      assay_p = assay_res$p_min, assay_powered = assay_res$n_powered)
    if (!res$rejected) return(res$row)
  }

  # ---- BRANCH 6b : Meta-regression as small-study-bias adjustment (Bridge 4) ----
  if (!is.na(eggers_p) && eggers_p < thresholds$eggers_alpha &&
      !is.na(k_total) && k_total >= thresholds$R2_k_min) {
    res <- try_branch("6b", "Meta-regression",
                      "6b — Small-study bias adjustment",
                      sprintf("Egger's p = %s < %.2f — meta-regression used to adjust for small-study bias (R² = %s, k = %d)",
                              format_p(eggers_p), thresholds$eggers_alpha,
                              ifelse(is.na(R2), "—", sprintf("%.1f%%", R2 * 100)),
                              k_total),
                      age_p = age_res$p_min, age_powered = age_res$n_powered,
                      race_p = race_res$p_min, race_powered = race_res$n_powered,
                      assay_p = assay_res$p_min, assay_powered = assay_res$n_powered)
    if (!res$rejected) return(res$row)
  }

  # ---- BRANCH 6c : All stratum tests negative + meta-reg adds value ----
  # Formalises the narrative reasoning pattern: when sex/age/race/assay tests
  # are all non-significant, the meta-regression with moderators serves as the
  # principled population-distribution estimate — but ONLY if it adds value
  # over the simpler overall pooled estimate. If R² is minimal AND meta-reg
  # ≈ overall pooled, the regression adds nothing and we should defer to the
  # simpler pooled estimate (Occam's razor).
  #
  # Q6c fires if:
  #   (a) meta-regression range is clinically valid (lower 95% > 0)
  #   (b) k ≥ R2_k_min (model not saturated)
  #   (c) EITHER R² ≥ R2_q6c_min (moderators explain ≥ 10% of heterogeneity)
  #       OR     meta-reg pooled estimate differs from overall pooled by ≥ 5%
  #              (point estimate meaningfully different)
  mr_lower_95 <- if (nrow(mr_overall) && "lower_95" %in% names(mr_overall)) {
                   mr_overall$lower_95[1]
                 } else NA_real_
  mr_upper_95 <- if (nrow(mr_overall) && "upper_95" %in% names(mr_overall)) {
                   mr_overall$upper_95[1]
                 } else NA_real_
  mr_estimate <- if (nrow(mr_overall) && "pooled_estimate" %in% names(mr_overall)) {
                   mr_overall$pooled_estimate[1]
                 } else NA_real_
  ov_estimate <- if (nrow(overall) && "pooled_estimate" %in% names(overall)) {
                   overall$pooled_estimate[1]
                 } else NA_real_

  # Relative difference between MR and Overall pooled estimates
  mr_rel_diff <- if (!is.na(mr_estimate) && !is.na(ov_estimate) &&
                     abs(ov_estimate) > 0) {
                   abs(mr_estimate - ov_estimate) / abs(ov_estimate)
                 } else NA_real_

  range_valid       <- !is.na(mr_lower_95) && mr_lower_95 > 0
  k_ok              <- !is.na(k_total)     && k_total    >= thresholds$R2_k_min
  R2_meaningful     <- !is.na(R2)          && R2         >= thresholds$R2_q6c_min
  mr_meaningfully_different <- !is.na(mr_rel_diff) && mr_rel_diff >= thresholds$mr_diff_min
  mr_adds_value     <- R2_meaningful || mr_meaningfully_different

  if (range_valid && k_ok && mr_adds_value) {
    why_added <- if (R2_meaningful && mr_meaningfully_different) {
                   sprintf("R² = %.1f%% ≥ %.0f%% AND meta-reg %.2f vs overall %.2f differs by %.1f%% ≥ %.0f%%",
                           R2 * 100, thresholds$R2_q6c_min * 100,
                           mr_estimate, ov_estimate,
                           mr_rel_diff * 100, thresholds$mr_diff_min * 100)
                 } else if (R2_meaningful) {
                   sprintf("R² = %.1f%% ≥ %.0f%%", R2 * 100, thresholds$R2_q6c_min * 100)
                 } else {
                   sprintf("meta-reg %.2f vs overall %.2f differs by %.1f%% ≥ %.0f%%",
                           mr_estimate, ov_estimate,
                           mr_rel_diff * 100, thresholds$mr_diff_min * 100)
                 }
    res <- try_branch("6c", "Meta-regression",
                      "6c — No stratum effect; meta-reg adds value",
                      sprintf("All stratum tests negative (sex p=%s, age p=%s, race p=%s, assay p=%s); range [%.2f, %.2f] valid (lower 95%% > 0); %s; k = %d.",
                              format_p(women_prop_p),
                              format_p(age_res$p_min),
                              format_p(race_res$p_min),
                              format_p(assay_res$p_min),
                              mr_lower_95, mr_upper_95,
                              why_added,
                              k_total),
                      age_p = age_res$p_min, age_powered = age_res$n_powered,
                      race_p = race_res$p_min, race_powered = race_res$n_powered,
                      assay_p = assay_res$p_min, assay_powered = assay_res$n_powered)
    if (!res$rejected) return(res$row)
  }

  # ---- BRANCH 7 : Low-RoB subset preferred ----
  if (nrow(lrob) >= 1 && nrow(overall) >= 1 &&
      !is.na(lrob$pooled_estimate[1]) && !is.na(overall$pooled_estimate[1])) {

    overall_width <- overall$upper_95[1] - overall$lower_95[1]
    lrob_width    <- lrob$upper_95[1]    - lrob$lower_95[1]
    is_tighter    <- !is.na(overall_width) && !is.na(lrob_width) &&
                     lrob_width < overall_width

    rel_diff <- abs(lrob$pooled_estimate[1] - overall$pooled_estimate[1]) /
                abs(overall$pooled_estimate[1])
    is_consistent <- !is.na(rel_diff) && rel_diff <= thresholds$consistency_pct

    if (is_tighter && is_consistent) {
      res <- try_branch("7", "Low risk-of-bias overall",
                        "7 — Low-RoB subset preferred",
                        sprintf("CI width %.2f vs overall %.2f (tighter); shift %.1f%% ≤ %.0f%%",
                                lrob_width, overall_width,
                                rel_diff * 100, thresholds$consistency_pct * 100),
                        age_p = age_res$p_min, age_powered = age_res$n_powered,
                        race_p = race_res$p_min, race_powered = race_res$n_powered,
                        assay_p = assay_res$p_min, assay_powered = assay_res$n_powered)
      if (!res$rejected) return(res$row)
    }
  }

  # ---- BRANCH 8 : default fallback (Overall pooled) ----
  # The meta-regression fallback is now handled at Q6c (when stratum tests are
  # negative AND meta-reg range is valid AND k ≥ k floor). By the time we
  # reach Q8, either the meta-regression range is invalid, k is below floor,
  # or Q7 (Low-RoB) didn't fire either. In that case, fall back to the
  # straightforward overall pooled estimate with a heterogeneity caveat.
  default_gate <- quality_gate(component, "8", ma_long,
                               ma_race_long  = ma_race_long,
                               meta_reg_long = meta_reg_long,
                               data          = data,
                               thresholds    = thresholds)

  reason_default <- if (is.na(mr_lower_95)) {
    "Meta-regression unavailable; default to overall pooled with heterogeneity caveat"
  } else if (!is.na(mr_lower_95) && mr_lower_95 <= 0) {
    sprintf("Meta-regression lower 95%% bound = %.2f ≤ 0 (clinically invalid for a non-negative measurement); default to overall pooled",
            mr_lower_95)
  } else if (!is.na(k_total) && k_total < thresholds$R2_k_min) {
    sprintf("Meta-regression range %.2f–%.2f valid but k = %d < %d (model saturated); default to overall pooled",
            mr_lower_95, mr_upper_95, k_total, thresholds$R2_k_min)
  } else {
    "No branch criteria met; default to overall pooled with heterogeneity caveat"
  }

  build_row(
    outcome = "Overall pooled",
    branch  = "8 — Default pooled",
    reason  = reason_default,
    gate    = default_gate,
    age_p = age_res$p_min, age_powered = age_res$n_powered,
    race_p = race_res$p_min, race_powered = race_res$n_powered,
    assay_p = assay_res$p_min, assay_powered = assay_res$n_powered
  )
}

format_p <- function(p) {
  if (is.na(p)) return("—")
  if (p < 0.001) return("<0.001")
  sprintf("%.3f", p)
}

# ============================================================================
# STAGE 2 — Sensitivity check helpers
# ============================================================================
# Each helper returns list(significant, p_min, evidence) where
#   significant = TRUE / FALSE / NA (NA means insufficient data)
#   p_min       = minimum p-value across pairwise comparisons (NA if none)
#   evidence    = short text summary for the caveat string

# Generic pairwise-Wald scan over a set of (mean, sd) rows
pairwise_wald_min <- function(means, sds, alpha = 0.05) {
  ok <- !is.na(means) & !is.na(sds)
  means <- means[ok]; sds <- sds[ok]
  if (length(means) < 2) return(list(p_min = NA_real_, n_pairs = 0L))
  pairs <- combn(length(means), 2, simplify = FALSE)
  ps <- vapply(pairs, function(idx) {
    sex_wald_p(means[idx[1]], sds[idx[1]], means[idx[2]], sds[idx[2]])
  }, numeric(1))
  list(p_min = if (length(ps)) min(ps, na.rm = TRUE) else NA_real_,
       n_pairs = length(ps))
}

# Q3 — Age check: Wald across powered age bands in main analysis (all studies).
# Only pairs in which BOTH bands are powered (k ≥ k_min AND n ≥ n_min) are
# tested — this prevents underpowered age bands from triggering a stratified
# recommendation purely because their unstable pooled estimate happens to
# differ from a powered band.
age_check <- function(component, ma_long,
                      thresholds = THRESHOLDS) {
  alpha <- thresholds$wald_alpha
  comp_data <- ma_long %>%
    filter(urine_component == component,
           is_all_sexes(sex_group),
           quality_filter == "All Studies",
           grepl("^[0-9]", age_band))   # numeric age-band rows only

  if (nrow(comp_data) < 2) {
    return(list(significant = NA, p_min = NA_real_, n_powered = 0L,
                evidence = "<2 age bands available"))
  }

  n_total_bands <- nrow(comp_data)
  powered_data  <- comp_data %>%
    filter(n_studies          >= thresholds$k_min,
           total_participants >= thresholds$n_min)
  n_powered <- nrow(powered_data)

  if (n_powered < 2) {
    return(list(significant = FALSE, p_min = NA_real_, n_powered = n_powered,
                evidence = sprintf("<2 powered age bands (%d/%d powered) — cannot test",
                                   n_powered, n_total_bands)))
  }

  res <- pairwise_wald_min(powered_data$pooled_estimate,
                           powered_data$pooled_sd, alpha)
  list(
    significant = !is.na(res$p_min) && res$p_min < alpha,
    p_min       = res$p_min,
    n_powered   = n_powered,
    evidence    = sprintf("min Wald p = %s across %d powered age-band pair(s); %d/%d bands powered",
                          format_p(res$p_min), res$n_pairs,
                          n_powered, n_total_bands)
  )
}

# Q4 — race check: Wald across powered race groups in 3rd analysis.
# Only pairs in which BOTH groups are powered are tested.
race_check <- function(component, ma_race_long = NULL,
                       thresholds = THRESHOLDS) {
  alpha <- thresholds$wald_alpha
  if (is.null(ma_race_long)) {
    return(list(significant = NA, p_min = NA_real_, n_powered = 0L,
                evidence = "No race-stratified data supplied"))
  }
  comp_data <- ma_race_long %>%
    filter(urine_component == component,
           is_overall_age(age_band),
           is_all_sexes(sex_group),
           tolower(as.character(race_group)) != "all")

  if (nrow(comp_data) < 2) {
    return(list(significant = NA, p_min = NA_real_, n_powered = 0L,
                evidence = "<2 race groups with data"))
  }

  n_total_groups <- nrow(comp_data)
  powered_data   <- comp_data %>%
    filter(n_studies          >= thresholds$k_min,
           total_participants >= thresholds$n_min)
  n_powered <- nrow(powered_data)

  if (n_powered < 2) {
    return(list(significant = FALSE, p_min = NA_real_, n_powered = n_powered,
                evidence = sprintf("<2 powered race groups (%d/%d powered) — cannot test",
                                   n_powered, n_total_groups)))
  }

  res <- pairwise_wald_min(powered_data$pooled_estimate,
                           powered_data$pooled_sd, alpha)
  list(
    significant = !is.na(res$p_min) && res$p_min < alpha,
    p_min       = res$p_min,
    n_powered   = n_powered,
    evidence    = sprintf("min Wald p = %s across %d powered race pair(s); %d/%d groups powered",
                          format_p(res$p_min), res$n_pairs,
                          n_powered, n_total_groups)
  )
}

# Q5 — Assay/country check (Bridge 2: drop "Unreported" anchor + magnitude floor)
#
# Refinements vs naive pairwise Wald:
#   1. Drop the "Unreported" category before pairwise comparisons. ~95/146
#      studies fall into this bucket because the source text was unparseable;
#      comparisons against it conflate era/country/population with assay.
#   2. Require k ≥ 3 within each named category (already enforced by
#      assay_subgroup_ma).
#   3. Require the relative shift between any pair to exceed
#      `consistency_pct` (default 20%). A pair that is statistically
#      significant but clinically negligible should not trigger
#      assay-stratification.
assay_check <- function(component, data = urine_ma2,
                        thresholds = THRESHOLDS) {
  alpha     <- thresholds$wald_alpha
  min_shift <- thresholds$consistency_pct  # require |Δ| / mean > consistency_pct

  result <- tryCatch(assay_subgroup_ma(component, data = data),
                     error = function(e) NULL)
  if (is.null(result) || !"assay_category" %in% names(result)) {
    return(list(significant = NA, p_min = NA_real_, n_powered = 0L,
                evidence = "Assay subgroup analysis unavailable"))
  }

  # Drop "Unreported" and any non-named category, keep only powered ones
  named <- result %>%
    filter(!is.na(pooled_estimate),
           !grepl("^unreported", tolower(assay_category)),
           !grepl("\\(blanket\\)$", tolower(assay_category)))

  if (nrow(named) < 2) {
    return(list(significant = NA, p_min = NA_real_, n_powered = nrow(named),
                evidence = sprintf("<2 named, powered assay categories (%d)",
                                   nrow(named))))
  }

  # SE from CI half-width
  named <- named %>% mutate(se = (upper_95 - lower_95) / (2 * 1.96))

  pairs <- combn(nrow(named), 2, simplify = FALSE)
  pair_results <- purrr::map_dfr(pairs, function(idx) {
    a <- named[idx[1], ]; b <- named[idx[2], ]
    p <- sex_wald_p(a$pooled_estimate, a$se, b$pooled_estimate, b$se)
    rel_shift <- abs(a$pooled_estimate - b$pooled_estimate) /
                  ((abs(a$pooled_estimate) + abs(b$pooled_estimate)) / 2)
    tibble::tibble(p = p, rel_shift = rel_shift)
  })

  # A pair counts as "significant" only if BOTH p < alpha AND |shift| ≥ min_shift
  significant_pairs <- pair_results %>%
    filter(!is.na(p), p < alpha, !is.na(rel_shift), rel_shift >= min_shift)

  is_sig <- nrow(significant_pairs) >= 1
  p_min  <- if (nrow(pair_results)) min(pair_results$p, na.rm = TRUE) else NA_real_
  shift_at_min_p <- if (nrow(pair_results)) {
    pair_results$rel_shift[which.min(pair_results$p)]
  } else NA_real_

  list(
    significant = is_sig,
    p_min       = p_min,
    n_powered   = nrow(named),
    evidence    = sprintf("min Wald p = %s (rel shift %.1f%%) across %d named-category pair(s); %d/%d significant after %.0f%% shift filter",
                          format_p(p_min),
                          ifelse(is.na(shift_at_min_p), 0, shift_at_min_p * 100),
                          nrow(pair_results), nrow(significant_pairs),
                          nrow(pair_results), min_shift * 100)
  )
}

# Backwards-compatible aliases (older code may still call these names)
age_sensitivity_check   <- age_check
race_sensitivity_check  <- race_check
assay_sensitivity_check <- assay_check
```

#### 4.3 Applied Recommendations per Component

The data-driven recommendations table is rendered in **Finalised Results → Recommendations Summary** later in the document, once the per-component meta-analysis objects (*_ma_2nd_analysis and *_ma_3rd_analysis) have been built. Each per-component sub-section in Finalised Results also displays a component-specific “Framework Recommendation” callout immediately under its heading.

```
# Component vector used by the framework loop (matches the *_ma_2nd_analysis names)
analysis_components <- c("volume", "ph", "calcium", "oxalate", "urate",
                         "citrate", "creat", "phosphate", "sodium",
                         "potassium", "magnesium", "ammonium", "chloride",
                         "sulphate", "urea")
```

### 5 Functions

#### 5.1 Main MA Helper Function

```
urine_components = c("volume",
                     "ph",
                     "creat",
                     "calcium",
                     "oxalate",
                     "urate",
                     "citrate",
                     "phosphate",
                     "sodium",
                     "potassium",
                     "magnesium",
                     "ammonium",
                     "chloride",
                     "sulphate",
                     "urea")

age_bands <- levels(urine_ma2$age_band)

main_ma <- function(urine_component,
                    data = urine_ma2) {


  # --- 1. Identify mean and SD columns -----------
  mean_colname <- dplyr::case_when(
    urine_component == "volume" ~ "volume_mean_m_l",
    urine_component == "ph"     ~ "ph_mean",
    urine_component == "creat"  ~ "creatinine_mean_mmol_24h",
    TRUE                        ~ paste0(urine_component, "_mean_mmol_24h")
  )
  sd_colname <- dplyr::case_when(
    urine_component == "volume" ~ "volume_sd_m_l",
    urine_component == "ph"     ~ "ph_sd",
    urine_component == "creat"  ~ "creatinine_sd_mmol_24h",
    TRUE                        ~ paste0(urine_component, "_sd_mmol_24h")
  )

  urine_component_for_title <- dplyr::case_when(
    urine_component == "volume" ~ "Volume (L/24h)",
    urine_component == "ph" ~ "pH",
    urine_component == "creat" ~ "Creatinine (mmol/24h)",
    TRUE ~ paste0(str_to_title(urine_component), " (mmol/24h)")
  )

  # --- 2. Prepare data -----------
  data1 <- data %>%
    dplyr::select(study_name, total_n,
                  mean_col = all_of(mean_colname),
                  sd_col = all_of(sd_colname),
                  study_type) %>%
    tidyr::drop_na(mean_col, sd_col) %>%
    dplyr::arrange(mean_col)

  # Capture raw study-level means BEFORE any transformation, for descriptive stats
  study_means_raw <- data1$mean_col
  study_sds_raw   <- data1$sd_col
  study_n_raw     <- data1$total_n
  study_min_raw   <- if (length(study_means_raw)) min(study_means_raw, na.rm = TRUE) else NA_real_
  study_max_raw   <- if (length(study_means_raw)) max(study_means_raw, na.rm = TRUE) else NA_real_
  # Unweighted descriptive mean/SD across study-level means
  study_avg_raw   <- if (length(study_means_raw)) mean(study_means_raw, na.rm = TRUE) else NA_real_
  study_sd_raw    <- if (length(study_means_raw) > 1) stats::sd(study_means_raw, na.rm = TRUE) else NA_real_

  # Convert volume to log-normal
  if (urine_component == "volume") {
    data1 <- data1 %>%
      dplyr::mutate(
        mean_raw = mean_col,
        sd_raw = sd_col,
        mean_col = log(mean_raw^2 / sqrt(sd_raw^2 + mean_raw^2)),
        sd_col = sqrt(log(1 + (sd_raw^2 / mean_raw^2)))
      ) %>%
      dplyr::select(-mean_raw, -sd_raw)
  }

  # Run meta-analysis
  ma_result <- metamean(
    n = data1$total_n,
    mean = data1$mean_col,
    sd   = data1$sd_col,
    studlab = data1$study_name
  )

  # Extract pooled results
  k <- ma_result$k

  if (k == 1) {
    model_utilised <- "Single study"
    pooled_estimate <- ma_result$TE
    pooled_sd <- ma_result$seTE * sqrt(sum(data1$total_n))
    I2_value <- NA
    Q_value <- NA
    Q_pval <- NA
  } else {
    model_utilised <- if (ma_result$pval.Q < 0.05) "Random" else "Fixed"
    pooled_estimate <- if (ma_result$pval.Q < 0.05) ma_result$TE.random else ma_result$TE.common
    pooled_sd <- if (ma_result$pval.Q < 0.05) ma_result$tau else ma_result$seTE.common * sqrt(sum(data1$total_n))
    I2_value <- ma_result$I2
    Q_value <- ma_result$Q
    Q_pval <- ma_result$pval.Q
  }

  # Egger's test for funnel plot asymmetry (requires k >= 3)
  eggers_p <- tryCatch({
    if (k >= 3) {
      egger_test <- metabias(ma_result, method.bias = "linreg", k.min = 3)
      egger_test$p.value
    } else {
      NA_real_
    }
  }, error = function(e) NA_real_)

  # Make Study type column
  data1$study_type <- as.factor(data1$study_type)
  study_type <- if (dplyr::n_distinct(data1$study_type, na.rm = TRUE) > 1) {
  "Mixed"
} else {
  unique(na.omit(data1$study_type))
}

  # Create summary tibble
  summary <- tibble(
    urine_component = urine_component,
    sex_group = "all",
    age_band = "overall",
    total_participants = sum(data1$total_n),
    n_studies = k,
    pooled_estimate = pooled_estimate,
    pooled_sd = pooled_sd,
    I2 = I2_value,
    Q = Q_value,
    Q_pval = Q_pval,
    eggers_p = eggers_p,
    model_utilised = model_utilised,
    study_type = study_type,
    # Descriptive statistics across study-level means (raw, pre-transformation)
    study_mean_min = study_min_raw,
    study_mean_max = study_max_raw,
    study_mean_avg = study_avg_raw,
    study_mean_sd  = study_sd_raw
  )

  # Back-transform volume results if needed (and convert descriptive stats from mL to L)
  if (urine_component == "volume") {
    summary <- summary %>%
      mutate(
        lower_95 = exp(pooled_estimate - 1.96 * pooled_sd) / 1000,
        upper_95 = exp(pooled_estimate + 1.96 * pooled_sd) / 1000,
        pooled_estimate = exp(pooled_estimate) / 1000,
        study_mean_min = study_mean_min / 1000,
        study_mean_max = study_mean_max / 1000,
        study_mean_avg = study_mean_avg / 1000,
        study_mean_sd  = study_mean_sd  / 1000
      )
  } else {
    summary <- summary %>%
      mutate(
        lower_95 = pooled_estimate - 1.96 * pooled_sd,
        upper_95 = pooled_estimate + 1.96 * pooled_sd
      )
  }

  return(summary)
}
```

#### 5.2 Single MA Helper function

```
run_single_ma <- function(data1, urine_component, sex, age) {

  # Capture raw study-level means BEFORE any transformation, for descriptive stats
  study_means_raw <- data1$mean_col
  study_min_raw   <- if (length(study_means_raw)) min(study_means_raw, na.rm = TRUE) else NA_real_
  study_max_raw   <- if (length(study_means_raw)) max(study_means_raw, na.rm = TRUE) else NA_real_
  study_avg_raw   <- if (length(study_means_raw)) mean(study_means_raw, na.rm = TRUE) else NA_real_
  study_sd_raw    <- if (length(study_means_raw) > 1) stats::sd(study_means_raw, na.rm = TRUE) else NA_real_

  # Volume → convert to log-normal parameters
  if (urine_component == "volume") {
    data1 <- data1 %>%
      mutate(
        mean_raw = mean_col,
        sd_raw   = sd_col,
        mean_col = log(mean_raw^2 / sqrt(sd_raw^2 + mean_raw^2)),
        sd_col   = sqrt(log(1 + (sd_raw^2 / mean_raw^2)))
      ) %>%
      select(-mean_raw, -sd_raw)
  }

  # Run meta-analysis
  ma_result <- metamean(
    n = data1$total_n,
    mean = data1$mean_col,
    sd   = data1$sd_col,
    studlab = data1$study_name
  )

  # Extract pooled results
  k <- ma_result$k

  if (k == 1) {
    model_utilised <- "Single study"
    pooled_estimate <- ma_result$TE
    pooled_sd <- ma_result$seTE * sqrt(sum(data1$total_n))
    I2_value <- NA
    Q_value <- NA
    Q_pval <- NA
  } else {
    model_utilised <- if (ma_result$pval.Q < 0.05) "Random" else "Fixed"
    pooled_estimate <- if (ma_result$pval.Q < 0.05) ma_result$TE.random else ma_result$TE.common
    pooled_sd <- if (ma_result$pval.Q < 0.05) ma_result$tau else ma_result$seTE.common * sqrt(sum(data1$total_n))
    I2_value <- ma_result$I2
    Q_value <- ma_result$Q
    Q_pval <- ma_result$pval.Q
  }

  # Egger's test for funnel plot asymmetry (requires k >= 3)
  eggers_p <- tryCatch({
    if (k >= 3) {
      egger_test <- metabias(ma_result, method.bias = "linreg", k.min = 3)
      egger_test$p.value
    } else {
      NA_real_
    }
  }, error = function(e) NA_real_)

  # Make Study type column
  data1$study_type <- as.factor(data1$study_type)
  study_type <- if (dplyr::n_distinct(data1$study_type, na.rm = TRUE) > 1) {
  "Mixed"
} else {
  unique(na.omit(data1$study_type))
}

  # Create summary tibble
  summary <- tibble(
    urine_component = urine_component,
    sex_group = sex,
    age_band = age,
    total_participants = sum(data1$total_n),
    n_studies = k,
    pooled_estimate = pooled_estimate,
    pooled_sd = pooled_sd,
    I2 = I2_value,
    Q = Q_value,
    Q_pval = Q_pval,
    eggers_p = eggers_p,
    model_utilised = model_utilised,
    study_type = study_type,
    # Descriptive statistics across study-level means (raw, pre-transformation)
    study_mean_min = study_min_raw,
    study_mean_max = study_max_raw,
    study_mean_avg = study_avg_raw,
    study_mean_sd  = study_sd_raw
  )

  # Back-transform volume results if needed (and convert descriptive stats from mL to L)
  if (urine_component == "volume") {
    summary <- summary %>%
      mutate(
        lower_95 = exp(pooled_estimate - 1.96 * pooled_sd) / 1000,
        upper_95 = exp(pooled_estimate + 1.96 * pooled_sd) / 1000,
        pooled_estimate = exp(pooled_estimate) / 1000,
        study_mean_min = study_mean_min / 1000,
        study_mean_max = study_mean_max / 1000,
        study_mean_avg = study_mean_avg / 1000,
        study_mean_sd  = study_mean_sd  / 1000
      )
  } else {
    summary <- summary %>%
      mutate(
        lower_95 = pooled_estimate - 1.96 * pooled_sd,
        upper_95 = pooled_estimate + 1.96 * pooled_sd
      )
  }

  return(summary)
}
```

#### 5.3 Meta-Regression function

```
urine_ma2_for_meta_reg <- urine_ma2_for_meta_reg <- urine_ma2 %>%
  left_join(overall_bias,
            by = "study_name")

# Global cache for full meta-regression coefficient tables — populated by
# meta_regression() during each call. Keyed by "<component>__<sex_group>".
# Used by `format_meta_reg_plain()` to render plain-language interpretations.
meta_reg_full_results <- list()

# Helper to render the full coefficients of a meta-regression as a plain-language
# table. Continuous moderators are scaled to interpretable units:
#   - women_prop_c   -> "per 10% increase in proportion female"
#   - mean_age_c     -> "per decade of mean age"
#   - total_n_c      -> "per 1000 additional participants"
# Categorical moderators (age_band, overall RoB) are left as-is.
format_meta_reg_plain <- function(urine_component, sex_group = "all") {
  key <- paste(urine_component, sex_group, sep = "__")
  if (!key %in% names(meta_reg_full_results)) {
    return(gt::gt(tibble::tibble(Message = paste("No meta-regression result for",
                                                  urine_component, "/", sex_group))))
  }
  tbl <- meta_reg_full_results[[key]]

  fmt_p <- function(p) dplyr::case_when(
    is.na(p)      ~ "—",
    p < 0.001     ~ "<0.001",
    p < 0.01      ~ "<0.01",
    p < 0.05      ~ sprintf("%.3f", p),
    TRUE          ~ sprintf("%.3f", p)
  )

  tbl %>%
    dplyr::mutate(
      `Effect (95% CI)` = ifelse(
        is.na(scaled_estimate), "—",
        sprintf("%.3f [%.3f, %.3f]", scaled_estimate, scaled_lower, scaled_upper)
      ),
      `P-value` = fmt_p(p.value)
    ) %>%
    dplyr::select(`Moderator` = plain_label,
                  `Direction & magnitude` = plain_text,
                  `Effect (95% CI)`,
                  `P-value`) %>%
    gt::gt() %>%
    gt::tab_header(
      title = gt::md(sprintf("**Meta-regression — %s (%s)**",
                              stringr::str_to_title(urine_component),
                              sex_group)),
      subtitle = gt::md("Continuous moderators rescaled for plain-language interpretation.")
    ) %>%
    gt::cols_align(align = "center", columns = -Moderator) %>%
    gt::tab_options(table.font.size = 11, data_row.padding = gt::px(4))
}

meta_regression <- function(data_subset = "none",
                            data = urine_ma2_for_meta_reg,
                            urine_component = "volume") {

  # --- 1. Determine correct column names ------------------------------------
  mean_colname <- dplyr::case_when(
    urine_component == "volume" ~ "volume_mean_m_l",
    urine_component == "ph" ~ "ph_mean",
    urine_component == "creat" ~ "creatinine_mean_mmol_24h",
    TRUE ~ paste0(urine_component, "_mean_mmol_24h")
  )

  sd_colname <- dplyr::case_when(
    urine_component == "volume" ~ "volume_sd_m_l",
    urine_component == "ph" ~ "ph_sd",
    urine_component == "creat" ~ "creatinine_sd_mmol_24h",
    TRUE ~ paste0(urine_component, "_sd_mmol_24h")
  )

  sex_group_name <- case_when(
    data_subset == "none" ~ "main meta-analysis",
    data_subset == "male" ~ "male",
    data_subset == "female" ~ "female",
    TRUE ~ NA_character_
  )

  # --- 1b. Add per-component assay/country category --------------------------
  # study_assay_lookup is loaded in the Setup chunk. assay_cat is component-
  # specific, so we join only the rows for this component.
  if (exists("study_assay_lookup") && !is.null(study_assay_lookup)) {
    .comp_key <- if (urine_component == "creat") "creatinine" else urine_component
    .assay_join <- study_assay_lookup %>%
      dplyr::filter(component == .comp_key) %>%
      dplyr::select(study_name, assay_cat)
    data <- data %>%
      dplyr::left_join(.assay_join, by = "study_name") %>%
      dplyr::mutate(assay_cat = ifelse(is.na(assay_cat) | assay_cat == "",
                                       "Unreported", assay_cat))
  } else {
    data$assay_cat <- "Unreported"
  }

  # --- 2. Build dataset -------------------------------------------------------
  if (data_subset == "none") {
    data1 <- data %>%
      dplyr::select(
        study_name, age_band, total_n, male_female, female_n, race,
        restricted_diet, mean = dplyr::all_of(mean_colname),
        sd   = dplyr::all_of(sd_colname), study_type,
        overall, assay_cat
      ) %>%
      dplyr::mutate(
        women_prop = dplyr::case_when(
          male_female == "female" ~ female_n / total_n,
          male_female == "male" ~ 0,
          male_female == "mixed" & !is.na(female_n) ~ female_n / total_n,
          male_female == "mixed" & is.na(female_n) ~ 0.5,
          TRUE ~ NA_real_
        ),
        women_prop_c = women_prop - mean(women_prop, na.rm = TRUE),
        total_n_c = total_n - mean(total_n, na.rm = TRUE)
      ) %>%
      tidyr::drop_na(mean, sd, women_prop_c, overall)
  } else {
    data1 <- data %>%
      dplyr::filter(male_female == data_subset) %>%
      dplyr::select(
        study_name, age_band, total_n, male_female, female_n, race,
        restricted_diet, mean = dplyr::all_of(mean_colname),
        sd   = dplyr::all_of(sd_colname), study_type,
        overall, assay_cat
      ) %>%
      dplyr::mutate(
        women_prop = dplyr::case_when(
          data_subset == "female" ~ 1,
          data_subset == "male" ~ 0,
          male_female == "mixed" & !is.na(female_n) ~ female_n / total_n,
          male_female == "mixed" & is.na(female_n) ~ 0.5,
          TRUE ~ NA_real_
        ),
        women_prop_c = women_prop - mean(women_prop, na.rm = TRUE),
        total_n_c = total_n - mean(total_n, na.rm = TRUE)
      ) %>%
      tidyr::drop_na(mean, sd, women_prop_c, overall)
  }

  # --- 3. Meta-analysis -------------------------------------------------------
  if (urine_component == "volume"){
    data1 <- data1 %>%
      mutate(
        mean_raw = mean,
        sd_raw = sd,
        mean = log(mean_raw^2 / sqrt(sd_raw^2 + mean_raw^2)),
        sd = sqrt(log(1 + (sd_raw^2 / mean_raw^2)))
      ) %>%
      dplyr::select(-mean_raw, -sd_raw)

  ma_result <- metamean(
    n = data1$total_n,
    mean = data1$mean,
    sd = data1$sd,
    studlab = data1$study_name,
    data = data1,
    sm = "MRAW",
    method.tau = "REML",
    method.random.ci = "HK"
  )}
  else {
    ma_result <- metamean(
    n = data1$total_n,
    mean = data1$mean,
    sd = data1$sd,
    studlab = data1$study_name,
    data = data1,
    sm = "MRAW",
    method.tau = "REML",
    method.random.ci = "HK"
  )
  }

  k <- ma_result$k
  n_total <- sum(data1$total_n)

  # Egger's test for funnel plot asymmetry (requires k >= 3)
  eggers_p <- tryCatch({
    if (k >= 3) {
      egger_test <- metabias(ma_result, method.bias = "linreg", k.min = 3)
      egger_test$p.value
    } else {
      NA_real_
    }
  }, error = function(e) NA_real_)

  # Include assay_cat only if there are ≥ 2 categories with k ≥ 3 each
  assay_levels <- data1 %>%
    dplyr::count(assay_cat) %>%
    dplyr::filter(n >= 3, !is.na(assay_cat))
  use_assay <- nrow(assay_levels) >= 2
  predictors <- c("women_prop_c", "age_band", "race", "overall", "total_n_c")
  if (use_assay) predictors <- c(predictors, "assay_cat")
  p <- length(predictors)

  # Make Study type column
  data1$study_type <- as.factor(data1$study_type)
  study_type <- if (dplyr::n_distinct(data1$study_type, na.rm = TRUE) > 1) {
  "Mixed"
} else {
  unique(na.omit(data1$study_type))
}

  # --- 4. Format output helper ------------------------------------------------
  format_output <- function(tbl, model_type) {
    tbl %>%
      dplyr::transmute(
        term = term,
        type = gsub("_c$", "", predictor %||% term),
        estimate = estimate,
        std.error = std.error,
        statistic = statistic,
        p.value = p.value,
        model_type = model_type,
        k = k,
        n = n_total
      )
  }

  # --- 5. Multivariable model if feasible ------------------------------------
  if (k > (p + 1)) {
    model <- metareg(
      ma_result,
      formula = as.formula(paste("~", paste(predictors, collapse = " + "))),
      intercept = TRUE,
      method.tau = "REML"
    )

    tbl <- broom::tidy(model)
    tbl$predictor <- tbl$term

    # Save full coefficient table with plain-language interpretation
    full_tbl <- tbl %>%
      dplyr::mutate(
        # Scale factor: per-unit -> per "natural" unit
        scale_mult = dplyr::case_when(
          term == "women_prop_c"               ~ 0.10,    # per 10% female
          term == "mean_age_c"                  ~ 10.00,   # per decade
          term == "total_n_c"                   ~ 1000.00, # per 1000 participants
          TRUE                                  ~ 1.00
        ),
        plain_label = dplyr::case_when(
          term == "intercept"                   ~ "Intercept (reference)",
          term == "women_prop_c"                ~ "% female",
          term == "mean_age_c"                  ~ "Mean age",
          term == "total_n_c"                   ~ "Sample size",
          grepl("^age_band", term)              ~ paste0("Age band: ", sub("^age_band", "", term)),
          grepl("^overall", term)               ~ paste0("RoB: ", sub("^overall", "", term)),
          grepl("^race", term)                  ~ paste0("race: ", sub("^race", "", term)),
          grepl("^assay_cat", term)             ~ paste0("Assay: ", sub("^assay_cat", "", term)),
          TRUE                                   ~ term
        ),
        plain_text = dplyr::case_when(
          term == "women_prop_c"                ~ "Change per 10% increase in proportion female",
          term == "mean_age_c"                  ~ "Change per decade increase in mean age",
          term == "total_n_c"                   ~ "Change per 1000 additional participants",
          term == "intercept"                   ~ "Pooled estimate at reference values",
          grepl("^assay_cat", term)             ~ "Difference vs reference assay/country category",
          TRUE                                   ~ "Difference vs reference category"
        ),
        scaled_estimate = estimate  * scale_mult,
        scaled_se       = std.error * scale_mult,
        scaled_lower    = scaled_estimate - 1.96 * scaled_se,
        scaled_upper    = scaled_estimate + 1.96 * scaled_se,
        # R² captured here so the cache can serve recommend_analysis() Bridge 3
        R2 = if (!is.null(model$R2)) model$R2 / 100 else NA_real_,
        I2 = if (!is.null(model$I2)) model$I2 / 100 else NA_real_
      )
    meta_reg_full_results[[paste(urine_component, sex_group_name, sep = "__")]] <<- full_tbl

    if (urine_component == "volume") {
      tbl1 <- tbl %>%
        filter(term == "intercept") %>%
        mutate(
          urine_component = urine_component,
          sex_group = sex_group_name,
          age_band = "Meta-Regression",
          n_studies = k,
          total_participants = sum(ma_result$n),
          I2 = model$I2 / 100,
          R2 = if (!is.null(model$R2)) model$R2 / 100 else NA_real_,
          Q = model$QE,
          Q_pval = model$QEp,
          model_utilised = "Multivariable",
          pooled_estimate = estimate,
          pooled_sd = case_when(
            model$tau2 > 0 ~ sqrt(model$tau2),
            TRUE ~ sqrt(mean(data1$sd^2) + model$tau2)),
          lower_95 = exp(pooled_estimate - (1.96 * pooled_sd)) / 1000,
          upper_95 = exp(pooled_estimate + (1.96 * pooled_sd)) / 1000,
          pooled_estimate = exp(estimate) / 1000,
          eggers_p = eggers_p
        ) %>%
        select(urine_component,
               sex_group,
               age_band,
               total_participants,
               n_studies,
               pooled_estimate,
               pooled_sd,
               I2,
               R2,
               Q,
               Q_pval,
               eggers_p,
               model_utilised,
               lower_95,
               upper_95)
    } else {
      tbl1 <- tbl %>%
        filter(term == "intercept") %>%
        mutate(
          urine_component = urine_component,
          sex_group = sex_group_name,
          age_band = "Meta-Regression",
          n_studies = k,
          total_participants = sum(ma_result$n),
          I2 = model$I2 / 100,
          R2 = if (!is.null(model$R2)) model$R2 / 100 else NA_real_,
          Q = model$QE,
          Q_pval = model$QEp,
          model_utilised = "Multivariable",
          pooled_estimate = estimate,
          pooled_sd = case_when(
            model$tau2 > 0 ~ sqrt(model$tau2),
            TRUE ~ sqrt(mean(data1$sd^2) + model$tau2)),
          lower_95 = pooled_estimate - 1.96 * pooled_sd,
          upper_95 = pooled_estimate + 1.96 * pooled_sd,
          study_type = study_type,
          eggers_p = eggers_p
        ) %>%
        select(urine_component,
               sex_group,
               age_band,
               total_participants,
               n_studies,
               pooled_estimate,
               pooled_sd,
               I2,
               R2,
               Q,
               Q_pval,
               eggers_p,
               model_utilised,
               lower_95,
               upper_95,
               study_type)
    }

    return(tbl1)
  }

  message(
    paste0("Not enough studies for multivariable meta-regression (k = ",
           k, ", need > ", p + 1, "). Running univariable models instead.")
  )

  # --- 6. Helper to count parameters -----------------------------------------
  param_count <- function(var) {
    x <- data1[[var]]
    if (is.numeric(x)) return(2) # intercept + slope
    return(length(unique(x[!is.na(x)])))
  }

  # --- 7. Run univariable models ---------------------------------------------
  first_model <- NULL

  results <- purrr::map_df(predictors, function(pred) {
    needed <- param_count(pred)

    if (needed >= k) {
      return(tibble::tibble(
        term = pred,
        type = pred,
        estimate = NA_real_,
        std.error = NA_real_,
        statistic = NA_real_,
        p.value = NA_real_,
        model_type = "univariable_skipped",
        k = k,
        n = n_total
      ))
    }

    model <- tryCatch(
      metareg(ma_result, as.formula(paste("~", pred)),
              intercept = TRUE,
              method.tau = "REML"),
      error = function(e) NULL
    )

    if (is.null(model)) {
      return(tibble::tibble(
        term = pred,
        type = pred,
        estimate = NA_real_,
        std.error = NA_real_,
        statistic = NA_real_,
        p.value = NA_real_,
        model_type = "univariable_error",
        k = k,
        n = n_total
      ))
    }

    # Store first successful model
    if (is.null(first_model)) {
      first_model <<- model
    }

    tbl <- broom::tidy(model)
    tbl$predictor <- pred
    format_output(tbl, model_type = "Univariable")
  })

  # Check if we have a valid model before proceeding
  if (is.null(first_model)) {
    warning("No successful univariable models were fitted")
    return(NULL)
  }

  # Save full coefficient table with plain-language interpretation (univariable path)
  full_tbl <- results %>%
    dplyr::mutate(
      scale_mult = dplyr::case_when(
        term == "women_prop_c"               ~ 0.10,
        term == "mean_age_c"                  ~ 10.00,
        term == "total_n_c"                   ~ 1000.00,
        TRUE                                  ~ 1.00
      ),
      plain_label = dplyr::case_when(
        term == "intercept"                   ~ "Intercept (reference)",
        term == "women_prop_c"                ~ "% female",
        term == "mean_age_c"                  ~ "Mean age",
        term == "total_n_c"                   ~ "Sample size",
        grepl("^age_band", term)              ~ paste0("Age band: ", sub("^age_band", "", term)),
        grepl("^overall", term)               ~ paste0("RoB: ", sub("^overall", "", term)),
        grepl("^race", term)                  ~ paste0("race: ", sub("^race", "", term)),
        grepl("^assay_cat", term)             ~ paste0("Assay: ", sub("^assay_cat", "", term)),
        TRUE                                   ~ term
      ),
      plain_text = dplyr::case_when(
        term == "women_prop_c"                ~ "Change per 10% increase in proportion female",
        term == "mean_age_c"                  ~ "Change per decade increase in mean age",
        term == "total_n_c"                   ~ "Change per 1000 additional participants",
        term == "intercept"                   ~ "Pooled estimate at reference values",
        grepl("^assay_cat", term)             ~ "Difference vs reference assay/country category",
        TRUE                                   ~ "Difference vs reference category"
      ),
      scaled_estimate = estimate  * scale_mult,
      scaled_se       = std.error * scale_mult,
      scaled_lower    = scaled_estimate - 1.96 * scaled_se,
      scaled_upper    = scaled_estimate + 1.96 * scaled_se,
      # R² captured here so the cache can serve recommend_analysis() Bridge 3
      R2 = if (!is.null(first_model$R2)) first_model$R2 / 100 else NA_real_,
      I2 = if (!is.null(first_model$I2)) first_model$I2 / 100 else NA_real_
    )
  meta_reg_full_results[[paste(urine_component, sex_group_name, sep = "__")]] <<- full_tbl

  if (urine_component == "volume") {
    results <- results %>%
      filter(term == "intercept") %>%
      slice_head(n = 1) %>%
      mutate(
        urine_component = urine_component,
        sex_group = sex_group_name,
        age_band = "Meta-Regression",
        total_participants = sum(ma_result$n),
        pooled_estimate = estimate,
        pooled_sd = case_when(
            first_model$tau2 > 0 ~ sqrt(first_model$tau2),
            TRUE ~ sqrt(mean(data1$sd^2) + model$tau2)),
        n_studies = k,
        I2 = first_model$I2 / 100,
        R2 = if (!is.null(first_model$R2)) first_model$R2 / 100 else NA_real_,
        Q = first_model$QE,
        Q_pval = first_model$QEp,
        model_utilised = "Univariable",
        lower_95 = exp(pooled_estimate - (1.96 * pooled_sd)) / 1000,
        upper_95 = exp(pooled_estimate + (1.96 * pooled_sd)) / 1000,
        pooled_estimate = exp(estimate) / 1000,
        study_type = study_type,
        eggers_p = eggers_p
      ) %>%
      select(urine_component,
             sex_group,
             age_band,
             total_participants,
             n_studies,
             pooled_estimate,
             pooled_sd,
             I2,
             R2,
             Q,
             Q_pval,
             eggers_p,
             model_utilised,
             lower_95,
             upper_95,
             study_type)
  } else {
    results <- results %>%
      filter(term == "intercept") %>%
      slice_head(n = 1) %>%
      mutate(
        urine_component = urine_component,
        sex_group = sex_group_name,
        age_band = "Meta-Regression",
        total_participants = sum(ma_result$n),
        pooled_estimate = estimate,
        pooled_sd = case_when(
            first_model$tau2 > 0 ~ sqrt(first_model$tau2),
            TRUE ~ sqrt(mean(data1$sd^2) + first_model$tau2)),
        n_studies = k,
        I2 = first_model$I2 / 100,
        R2 = if (!is.null(first_model$R2)) first_model$R2 / 100 else NA_real_,
        Q = first_model$QE,
        Q_pval = first_model$QEp,
        model_utilised = "Univariable",
        lower_95 = pooled_estimate - (1.96 * pooled_sd),
        upper_95 = pooled_estimate + (1.96 * pooled_sd),
        study_type = study_type,
        eggers_p = eggers_p
      ) %>%
      select(urine_component,
             sex_group,
             age_band,
             total_participants,
             n_studies,
             pooled_estimate,
             pooled_sd,
             I2,
             R2,
             Q,
             Q_pval,
             eggers_p,
             model_utilised,
             lower_95,
             upper_95,
             study_type)
  }

  return(results)
}
```

#### 5.4 Assay subgroup analysis & meta-regression covariate (A5)

```
# Run a meta-analysis stratified by assay category for one urine component.
# Returns a tibble: one row per assay category with k, n, pooled estimate, CI,
# I², Q p-value, and Egger's p (where computable). Categories with k < 3 are
# reported separately (to acknowledge low power) but not pooled.
assay_subgroup_ma <- function(urine_component, data = urine_ma2,
                               min_k = 3) {

  # Identify mean/SD column names (same logic as main_ma)
  mean_col <- dplyr::case_when(
    urine_component == "volume" ~ "volume_mean_m_l",
    urine_component == "ph"     ~ "ph_mean",
    urine_component == "creat"  ~ "creatinine_mean_mmol_24h",
    TRUE                        ~ paste0(urine_component, "_mean_mmol_24h")
  )
  sd_col <- dplyr::case_when(
    urine_component == "volume" ~ "volume_sd_m_l",
    urine_component == "ph"     ~ "ph_sd",
    urine_component == "creat"  ~ "creatinine_sd_mmol_24h",
    TRUE                        ~ paste0(urine_component, "_sd_mmol_24h")
  )

  data_assay <- add_assay_column(data, urine_component) %>%
    dplyr::select(study_name, total_n, assay_cat, preservative_used,
                  mean_val = dplyr::all_of(mean_col),
                  sd_val   = dplyr::all_of(sd_col)) %>%
    tidyr::drop_na(mean_val, sd_val)

  if (nrow(data_assay) == 0) {
    return(tibble::tibble(message = "No data with this component"))
  }

  # Tally categories
  cat_counts <- data_assay %>% dplyr::count(assay_cat, name = "k")

  # Run MA per assay category that has k >= min_k
  ma_per_cat <- purrr::map_dfr(cat_counts$assay_cat, function(cat) {
    sub <- data_assay %>% dplyr::filter(assay_cat == cat)
    k <- nrow(sub)
    if (k < min_k) {
      return(tibble::tibble(
        assay_category   = cat,
        n_studies        = k,
        n_participants   = sum(sub$total_n),
        pooled_estimate  = NA_real_,
        lower_95         = NA_real_,
        upper_95         = NA_real_,
        I2               = NA_real_,
        Q_pval           = NA_real_,
        eggers_p         = NA_real_,
        note             = sprintf("k = %d < min_k = %d (skipped)", k, min_k)
      ))
    }

    ma <- tryCatch(
      meta::metamean(n = sub$total_n,
                     mean = sub$mean_val,
                     sd   = sub$sd_val,
                     studlab = sub$study_name,
                     method.tau = "REML",
                     sm = "MRAW"),
      error = function(e) NULL
    )
    if (is.null(ma)) {
      return(tibble::tibble(assay_category = cat, n_studies = k,
                             n_participants = sum(sub$total_n),
                             pooled_estimate = NA_real_, lower_95 = NA_real_,
                             upper_95 = NA_real_, I2 = NA_real_,
                             Q_pval = NA_real_, eggers_p = NA_real_,
                             note = "Meta-analysis failed"))
    }

    # Egger's test (k >= 3)
    eggers_p <- tryCatch({
      meta::metabias(ma, method.bias = "linreg", k.min = 3)$p.value
    }, error = function(e) NA_real_)

    tibble::tibble(
      assay_category  = cat,
      n_studies       = k,
      n_participants  = sum(sub$total_n),
      pooled_estimate = ma$TE.random,
      lower_95        = ma$lower.random,
      upper_95        = ma$upper.random,
      I2              = ma$I2,
      Q_pval          = ma$pval.Q,
      eggers_p        = eggers_p,
      note            = ""
    )
  })

  ma_per_cat %>% dplyr::arrange(dplyr::desc(n_studies))
}

# Render the assay subgroup result as a gt table
format_assay_subgroup_gt <- function(result, urine_component) {
  if (!"assay_category" %in% names(result)) {
    return(gt::gt(result))
  }

  fmt_p <- function(p) dplyr::case_when(
    is.na(p) ~ "—",
    p < 0.001 ~ "<0.001",
    p < 0.01  ~ "<0.01",
    p < 0.05  ~ sprintf("%.3f", p),
    TRUE      ~ sprintf("%.3f", p)
  )

  result %>%
    dplyr::mutate(
      `95% CI` = ifelse(
        is.na(pooled_estimate),
        "—",
        sprintf("%.2f [%.2f, %.2f]", pooled_estimate, lower_95, upper_95)
      ),
      `I²`           = ifelse(is.na(I2),     "—", sprintf("%.0f%%", I2 * 100)),
      `Q p-value`    = fmt_p(Q_pval),
      `Egger's p`    = fmt_p(eggers_p),
      `Participants` = format(n_participants, big.mark = ",")
    ) %>%
    dplyr::select(`Assay category` = assay_category,
                  `Studies, n`     = n_studies,
                  Participants,
                  `95% CI`,
                  `I²`,
                  `Q p-value`,
                  `Egger's p`,
                  Note            = note) %>%
    gt::gt() %>%
    gt::tab_header(
      title    = gt::md(sprintf("**Assay subgroup analysis — %s**",
                                stringr::str_to_title(urine_component))),
      subtitle = gt::md("*Per-assay pooled estimates; categories with k < 3 listed but not pooled.*")
    ) %>%
    gt::cols_align(align = "center", columns = -`Assay category`) %>%
    gt::tab_options(table.font.size = 11, data_row.padding = gt::px(4))
}

# Quick helper combining the analysis + display
assay_subgroup_table <- function(urine_component, data = urine_ma2) {
  res <- assay_subgroup_ma(urine_component, data)
  format_assay_subgroup_gt(res, urine_component)
}

# Sensitivity analysis: with vs without preservative-using cohorts
preservative_sensitivity_ma <- function(urine_component, data = urine_ma2) {
  mean_col <- dplyr::case_when(
    urine_component == "volume" ~ "volume_mean_m_l",
    urine_component == "ph"     ~ "ph_mean",
    urine_component == "creat"  ~ "creatinine_mean_mmol_24h",
    TRUE                        ~ paste0(urine_component, "_mean_mmol_24h")
  )
  sd_col <- dplyr::case_when(
    urine_component == "volume" ~ "volume_sd_m_l",
    urine_component == "ph"     ~ "ph_sd",
    urine_component == "creat"  ~ "creatinine_sd_mmol_24h",
    TRUE                        ~ paste0(urine_component, "_sd_mmol_24h")
  )

  data_p <- add_assay_column(data, urine_component) %>%
    dplyr::select(study_name, total_n, preservative_used,
                  mean_val = dplyr::all_of(mean_col),
                  sd_val   = dplyr::all_of(sd_col)) %>%
    tidyr::drop_na(mean_val, sd_val)

  run_ma <- function(d) {
    if (nrow(d) < 2) return(NULL)
    tryCatch(
      meta::metamean(n = d$total_n, mean = d$mean_val, sd = d$sd_val,
                     studlab = d$study_name, method.tau = "REML", sm = "MRAW"),
      error = function(e) NULL
    )
  }

  groups <- list(
    `All studies`            = data_p,
    `Excluding preservative` = data_p %>% dplyr::filter(!preservative_used),
    `Preservative only`      = data_p %>% dplyr::filter(preservative_used)
  )

  purrr::map_dfr(names(groups), function(grp) {
    d  <- groups[[grp]]
    ma <- run_ma(d)
    if (is.null(ma)) {
      return(tibble::tibble(group = grp, n_studies = nrow(d),
                             n_participants = sum(d$total_n),
                             pooled_estimate = NA_real_,
                             lower_95 = NA_real_, upper_95 = NA_real_))
    }
    tibble::tibble(
      group           = grp,
      n_studies       = ma$k,
      n_participants  = sum(d$total_n),
      pooled_estimate = ma$TE.random,
      lower_95        = ma$lower.random,
      upper_95        = ma$upper.random
    )
  })
}

# Sensitivity analysis: compare the all-studies meta-analysis to a subset
# restricted to studies that report 24h urine creatinine. 24h creatinine
# is conventionally used to verify collection completeness — studies that
# don't report it cannot reliably confirm that collections were complete,
# and may include incomplete (lower-volume) samples that bias other
# analyte estimates. A material shift between the full and creatinine-
# reporting subsets would therefore flag a possible completeness bias.
#
# Returns a 3-row tibble with All studies / Creatinine reported only /
# Creatinine NOT reported, plus pooled estimate, CI, and n.
creat_completeness_ma <- function(urine_component, data = urine_ma2) {
  if (urine_component == "creat") {
    # By definition, this analysis is only the studies reporting creatinine
    return(tibble::tibble(group = "Not applicable",
                          n_studies = NA_integer_,
                          n_participants = NA_integer_,
                          pooled_estimate = NA_real_,
                          lower_95 = NA_real_, upper_95 = NA_real_))
  }

  mean_col <- dplyr::case_when(
    urine_component == "volume" ~ "volume_mean_m_l",
    urine_component == "ph"     ~ "ph_mean",
    TRUE                        ~ paste0(urine_component, "_mean_mmol_24h")
  )
  sd_col <- dplyr::case_when(
    urine_component == "volume" ~ "volume_sd_m_l",
    urine_component == "ph"     ~ "ph_sd",
    TRUE                        ~ paste0(urine_component, "_sd_mmol_24h")
  )

  d_all <- data %>%
    dplyr::select(study_name, total_n,
                  mean_val = dplyr::all_of(mean_col),
                  sd_val   = dplyr::all_of(sd_col),
                  creatinine_mean_mmol_24h) %>%
    tidyr::drop_na(mean_val, sd_val) %>%
    dplyr::mutate(reports_creat = !is.na(creatinine_mean_mmol_24h))

  run_ma <- function(d) {
    if (nrow(d) < 2) return(NULL)
    tryCatch(
      meta::metamean(n = d$total_n, mean = d$mean_val, sd = d$sd_val,
                     studlab = d$study_name, method.tau = "REML", sm = "MRAW"),
      error = function(e) NULL
    )
  }

  groups <- list(
    `All studies`               = d_all,
    `Creatinine reported only`  = d_all %>% dplyr::filter(reports_creat),
    `Creatinine NOT reported`   = d_all %>% dplyr::filter(!reports_creat)
  )

  purrr::map_dfr(names(groups), function(grp) {
    d  <- groups[[grp]]
    ma <- run_ma(d)
    if (is.null(ma)) {
      return(tibble::tibble(group = grp, n_studies = nrow(d),
                             n_participants = sum(d$total_n),
                             pooled_estimate = NA_real_,
                             lower_95 = NA_real_, upper_95 = NA_real_))
    }
    tibble::tibble(
      group           = grp,
      n_studies       = ma$k,
      n_participants  = sum(d$total_n),
      pooled_estimate = ma$TE.random,
      lower_95        = ma$lower.random,
      upper_95        = ma$upper.random
    )
  })
}

# Render `creat_completeness_ma` output as a gt table with a flag if the
# creatinine-reporting subset estimate differs from the all-studies estimate
# by more than `consistency_pct` (default 20%).
creat_completeness_table <- function(urine_component, data = urine_ma2,
                                     thresholds = THRESHOLDS) {
  res <- creat_completeness_ma(urine_component, data = data)

  if (urine_component == "creat" || nrow(res) == 0) {
    return(gt::gt(tibble::tibble(
      Message = "Not applicable to creatinine itself"
    )) %>% gt::tab_header(title = gt::md("**Creatinine-completeness sensitivity**")))
  }

  ref <- res %>% dplyr::filter(group == "All studies")
  ref_est <- if (nrow(ref)) ref$pooled_estimate[1] else NA_real_

  res2 <- res %>%
    dplyr::mutate(
      `Range (95% CI)` = ifelse(
        is.na(pooled_estimate), "—",
        sprintf("%.2f [%.2f, %.2f]", pooled_estimate, lower_95, upper_95)
      ),
      `Δ vs all-studies (%)` = dplyr::case_when(
        group == "All studies"        ~ "—",
        is.na(pooled_estimate) | is.na(ref_est) | ref_est == 0 ~ "—",
        TRUE ~ sprintf("%+.1f%%", 100 * (pooled_estimate - ref_est) / ref_est)
      ),
      Flag = dplyr::case_when(
        group == "All studies" ~ "—",
        is.na(pooled_estimate) | is.na(ref_est) | ref_est == 0 ~ "—",
        abs(pooled_estimate - ref_est) / abs(ref_est) > thresholds$consistency_pct ~
          sprintf("⚠ shift > %.0f%%", thresholds$consistency_pct * 100),
        TRUE ~ "OK"
      )
    ) %>%
    dplyr::transmute(
      Subset           = group,
      `Studies, n`     = n_studies,
      Participants     = format(n_participants, big.mark = ","),
      `Range (95% CI)` = `Range (95% CI)`,
      `Δ vs all-studies (%)` = `Δ vs all-studies (%)`,
      Flag             = Flag
    )

  res2 %>%
    gt::gt() %>%
    gt::tab_header(
      title    = gt::md(sprintf("**Creatinine-completeness sensitivity — %s**",
                                stringr::str_to_title(urine_component))),
      subtitle = gt::md("*Compares the all-studies pooled estimate against subsets defined by whether the study reported 24h urine creatinine. Studies that do not report creatinine cannot verify collection completeness; a substantial shift here flags possible bias from incomplete collections.*")
    ) %>%
    gt::data_color(
      columns = Flag,
      fn = function(x) {
        dplyr::case_when(
          grepl("^⚠", x) ~ "#fff3cd",
          x == "OK"           ~ "#d4edda",
          TRUE                 ~ "#f0f0f0"
        )
      }
    ) %>%
    gt::cols_align(align = "center", columns = -Subset) %>%
    gt::tab_options(table.font.size = 11, data_row.padding = gt::px(4))
}
```

#### 5.5 Reduce I² in Main MA by removing outlying studies

```
main_ma_reduce_I2 <- function(urine_component, data = urine_ma2) {

  # --- 1. Identify mean and SD columns -----------
  mean_colname <- dplyr::case_when(
    urine_component == "volume" ~ "volume_mean_m_l",
    urine_component == "ph"     ~ "ph_mean",
    urine_component == "creat"  ~ "creatinine_mean_mmol_24h",
    TRUE                        ~ paste0(urine_component, "_mean_mmol_24h")
  )
  sd_colname <- dplyr::case_when(
    urine_component == "volume" ~ "volume_sd_m_l",
    urine_component == "ph"     ~ "ph_sd",
    urine_component == "creat"  ~ "creatinine_sd_mmol_24h",
    TRUE                        ~ paste0(urine_component, "_sd_mmol_24h")
  )

  urine_component_for_title <- dplyr::case_when(
    urine_component == "volume" ~ "Volume (L/24h)",
    urine_component == "ph" ~ "pH",
    urine_component == "creat" ~ "Creatinine (mmol/24h)",
    TRUE ~ paste0(str_to_title(urine_component), " (mmol/24h)")
  )

  # --- 2. Prepare data -----------
  data_iter <- data %>%
    dplyr::select(study_name, total_n,
                  mean_col = all_of(mean_colname),
                  sd_col = all_of(sd_colname), study_type) %>%
    tidyr::drop_na(mean_col, sd_col) %>%
    dplyr::arrange(mean_col)

  # Convert volume to log-normal
  if (urine_component == "volume") {
    data_iter <- data_iter %>%
      dplyr::mutate(
        mean_raw = mean_col,
        sd_raw = sd_col,
        mean_col = log(mean_raw^2 / sqrt(sd_raw^2 + mean_raw^2)),
        sd_col = sqrt(log(1 + (sd_raw^2 / mean_raw^2)))
      ) %>%
      dplyr::select(-mean_raw, -sd_raw)
  }

  # Make Study type column
  data_iter$study_type <- as.factor(data_iter$study_type)
  study_type <- if (dplyr::n_distinct(data_iter$study_type, na.rm = TRUE) > 1) {
  "Mixed"
} else {
  unique(na.omit(data_iter$study_type))
}

  # --- 3. Identify clusters -----------
  clusters <- list()
  cluster_data_list <- list()
  cluster_id <- 1

  while(nrow(data_iter) >= 3) {
    cluster_data <- data_iter
    repeat {
      if(nrow(cluster_data) < 3) break
      ma_result <- tryCatch(
        metamean(n = cluster_data$total_n,
                 mean = cluster_data$mean_col,
                 sd = cluster_data$sd_col,
                 studlab = cluster_data$study_name),
        error = function(e) NULL
      )
      if(is.null(ma_result)) break
      if(ma_result$I2 < 0.5) break
      cluster_data <- cluster_data %>% dplyr::slice(-c(1, nrow(.)))
    }
    if(nrow(cluster_data) < 3) break

    ma_test <- tryCatch(
      metamean(n = cluster_data$total_n,
               mean = cluster_data$mean_col,
               sd = cluster_data$sd_col,
               studlab = cluster_data$study_name),
      error = function(e) NULL
    )
    if(is.null(ma_test)) {
      data_iter <- data_iter %>% dplyr::slice(-(1:nrow(cluster_data)))
      next
    }

    clusters[[cluster_id]] <- cluster_data
    cluster_data_list[[cluster_id]] <- cluster_data
    cluster_id <- cluster_id + 1
    data_iter <- dplyr::anti_join(data_iter, cluster_data, by = "study_name")
  }

  if(length(clusters) == 0) {
    warning("No clusters with I² < 0.5 identified.")
    return(list(summary = NULL, table = NULL, plot = NULL, cluster_data = NULL))
  }

  # --- 4. Summarize clusters -----------
  cluster_summaries <- purrr::map_dfr(seq_along(clusters), function(i) {
    d <- clusters[[i]]
    ma <- tryCatch(
      metamean(n = d$total_n,
               mean = d$mean_col,
               sd = d$sd_col,
               studlab = d$study_name),
      error = function(e) NULL
    )
    if(is.null(ma)) return(NULL)

    est <- ifelse(ma$pval.Q < 0.05, ma$TE.random, ma$TE.common)
    sd_val <- ifelse(ma$pval.Q < 0.05, ma$tau, ma$seTE.common * sqrt(sum(d$total_n)))

    if(urine_component == "volume") {
      est_bt <- exp(est)/1000
      lower_95 <- exp(est - 1.96*sd_val)/1000
      upper_95 <- exp(est + 1.96*sd_val)/1000
    } else {
      est_bt <- est
      lower_95 <- est - 1.96*sd_val
      upper_95 <- est + 1.96*sd_val
    }

    tibble::tibble(
      cluster = i,
      n_studies = nrow(d),
      total_participants = sum(d$total_n),
      pooled_estimate = est_bt,
      pooled_sd = sd_val,
      I2 = ma$I2 / 100,
      Q = ma$Q,
      Q_pval = ma$pval.Q,
      Model = "Fixed",
      lower_95 = lower_95,
      upper_95 = upper_95,
      studies_in_cluster = paste(d$study_name, collapse = ", "),
      cluster_data = list(d),
      study_type = study_type
    )
  }) %>%
    arrange(cluster) %>%
    mutate(cluster_factor = factor(cluster))

  # --- 5. Assign consistent colors -----------
  cluster_colors <- scales::hue_pal()(length(unique(cluster_summaries$cluster)))

  # --- 6. Create GT table -----------
  table_gt <- cluster_summaries %>%
    mutate(
      range = sprintf("%.2f – %.2f", lower_95, upper_95),
      I2_text = ifelse(is.na(I2), "—", paste0(round(I2*100, 1), "%")),
      Q_text = ifelse(is.na(Q), "—", round(Q, 1)),
      Q_pval_text = ifelse(is.na(Q_pval), "—", signif(Q_pval, 1))
    ) %>%
    select(cluster, range, n_studies, total_participants, I2_text, Q_text, Q_pval_text, Model) %>%
    gt() %>%
    cols_label(
      cluster = "Cluster",
      n_studies = "Studies",
      study_type = "Study Type",
      total_participants = "Participants",
      range = "95% Range",
      I2_text = "I²",
      Q_text = "Cochran's Q",
      Q_pval_text = "Q p-value",
      Model = "Model"
    ) %>%
    tab_header(title = md("**MA Results for Clusters with I² < 50%**"),
               subtitle = md(urine_component_for_title)) %>%
    tab_style(
      style = cell_text(weight = "bold"),
      locations = cells_column_labels(everything())
    )

  for(i in seq_along(unique(cluster_summaries$cluster))) {
    table_gt <- table_gt %>%
      tab_style(
        style = cell_fill(color = cluster_colors[i]),
        locations = cells_body(
          rows = cluster == unique(cluster_summaries$cluster)[i]
        )
      )
  }

  # --- 7. Forest plot with consistent colors -----------
  forest_plot <- ggplot2::ggplot(cluster_summaries, ggplot2::aes(y = cluster_factor)) +
    ggplot2::geom_errorbarh(ggplot2::aes(xmin = lower_95, xmax = upper_95, color = cluster_factor), height = 0.3) +
    ggplot2::geom_point(ggplot2::aes(x = pooled_estimate, color = cluster_factor), size = 3) +
    ggplot2::scale_color_manual(values = cluster_colors) +
    ggplot2::labs(
      title = paste0("Clusters with I² < 50% for ", urine_component_for_title),
      x = paste0("Estimated 95% CI for ", urine_component_for_title),
      y = "Cluster"
    ) +
    ggplot2::theme_minimal() +
    ggplot2::theme(legend.position = "none")

  # --- 8. Combine with patchwork -----------
  combined_plot <- forest_plot / table_gt

  # --- 9. Return results -----------
  list(
    summary = cluster_summaries,
    table = table_gt,
    plot = combined_plot,
    cluster_data = purrr::map(cluster_summaries$cluster_data, ~ .x)
  )
}
```

#### 5.6 Comparison to Sex subdivisions

```
sex_age_subdivision_ma <- function(urine_component,
                                   data = urine_ma2,
                                   overall_bias = NULL) {

  # --- 1. Identify the correct columns -------------------------------
  mean_colname <- case_when(
    urine_component == "volume" ~ "volume_mean_m_l",
    urine_component == "ph" ~ "ph_mean",
    urine_component == "creat" ~ "creatinine_mean_mmol_24h",
    TRUE ~ paste0(urine_component, "_mean_mmol_24h")
  )

  sd_colname <- case_when(
    urine_component == "volume" ~ "volume_sd_m_l",
    urine_component == "ph" ~ "ph_sd",
    urine_component == "creat" ~ "creatinine_sd_mmol_24h",
    TRUE ~ paste0(urine_component, "_sd_mmol_24h")
  )

  urine_component_for_title <- case_when(
    urine_component == "volume" ~ "Volume (L/24h)",
    urine_component == "ph" ~ "pH",
    urine_component == "creat" ~ "Creatinine (mmol/24h)",
    TRUE ~ paste0(str_to_title(urine_component), " (mmol/24h)")
  )

  # --- 2. Prepare data with overall_bias for meta-regression --------
  if (!is.null(overall_bias)) {
    data_with_bias <- data %>%
      left_join(overall_bias, by = "study_name")
  } else {
    data_with_bias <- data
  }

  # --- 3. Get main meta-analysis result ----------------------------
  # Overall main result (all sexes, all ages, all studies)
  main_overall <- main_ma(urine_component, data = data) %>%
    mutate(sex_group = "main meta-analysis",
           age_band = "Overall",
           rct_status = "All studies")

  # Main meta-analysis by age band (all sexes combined, all studies)
  data_main <- data %>%
    select(
      study_name,
      total_n,
      age_band,
      mean_col = all_of(mean_colname),
      sd_col   = all_of(sd_colname),
      study_type
    ) %>%
    drop_na(mean_col, sd_col)

  age_bands_main <- data_main %>%
    filter(!is.na(age_band)) %>%
    pull(age_band) %>%
    unique() %>%
    sort()

  main_age_results <- purrr::map_dfr(age_bands_main, function(age) {
    data_age <- data_main %>%
      filter(age_band == age)

    if (nrow(data_age) == 0) {
      return(tibble())
    }

    result <- run_single_ma(data_age, urine_component, "main meta-analysis", age)
    result %>% mutate(rct_status = "All studies")
  })

  # Add meta-regression for main analysis (all studies)
  main_meta_reg_results <- tryCatch({
    result <- meta_regression("none", urine_component = urine_component)
    if (!is.null(result)) {
      result %>% mutate(rct_status = "All studies")
    } else {
      NULL
    }
  }, error = function(e) {
    message("Meta-regression failed for main analysis: ", e$message)
    NULL
  })

  main_result <- bind_rows(main_overall, main_age_results, main_meta_reg_results)

  # --- 3b. RCT-only main analysis ----------------------------
  data_main_rct <- data %>%
    filter(study_type == "RCT") %>%
    select(
      study_name,
      total_n,
      age_band,
      study_type,
      mean_col = all_of(mean_colname),
      sd_col   = all_of(sd_colname)
    ) %>%
    drop_na(mean_col, sd_col)

  if (nrow(data_main_rct) > 0) {
    # Overall RCT result
    main_rct_overall <- run_single_ma(data_main_rct, urine_component, "main meta-analysis", "Overall") %>%
      mutate(rct_status = "RCT only")

    # RCT by age band
    main_rct_age_results <- purrr::map_dfr(age_bands_main, function(age) {
      data_age <- data_main_rct %>%
        filter(age_band == age)

      if (nrow(data_age) == 0) {
        return(tibble())
      }

      result <- run_single_ma(data_age, urine_component, "main meta-analysis", age)
      result %>% mutate(rct_status = "RCT only")
    })

    main_rct_result <- bind_rows(main_rct_overall, main_rct_age_results)
  } else {
    main_rct_result <- tibble()
  }

  # --- 4. Iterate through sexes and age bands --------------------------------
  sex_age_results <- purrr::map_dfr(c("male", "female"), function(sex) {

    # First get overall result for this sex (all age bands, all studies)
    data_sex <- data %>%
      filter(male_female == sex) %>%
      select(
        study_name,
        total_n,
        age_band,
        mean_col = all_of(mean_colname),
        sd_col   = all_of(sd_colname),
        study_type
      ) %>%
      drop_na(mean_col, sd_col)

    if (nrow(data_sex) == 0) {
      return(tibble())
    }

    # Get overall result for this sex (all studies)
    overall_result <- run_single_ma(data_sex, urine_component, sex, "Overall") %>%
      mutate(rct_status = "All studies")

    # Get meta-regression result for this sex (all studies)
    meta_reg_result <- tryCatch({
      result <- meta_regression(data_subset = sex, urine_component = urine_component)
      if (!is.null(result)) {
        result %>% mutate(rct_status = "All studies")
      } else {
        NULL
      }
    }, error = function(e) {
      message("Meta-regression failed for ", sex, ": ", e$message)
      NULL
    })

    # Get unique age bands
    age_bands <- data_sex %>%
      filter(!is.na(age_band)) %>%
      pull(age_band) %>%
      unique() %>%
      sort()

    # Get results for each age band (all studies)
    age_band_results <- purrr::map_dfr(age_bands, function(age) {
      data_age <- data_sex %>%
        filter(age_band == age)

      if (nrow(data_age) == 0) {
        return(tibble())
      }

      result <- run_single_ma(data_age, urine_component, sex, age)
      result %>% mutate(rct_status = "All studies")
    })

    # --- RCT-only analyses for this sex ---
    data_sex_rct <- data_sex %>%
      filter(study_type == "RCT")

    if (nrow(data_sex_rct) > 0) {
      # Overall RCT for this sex
      rct_overall <- run_single_ma(data_sex_rct, urine_component, sex, "Overall") %>%
        mutate(rct_status = "RCT only")

      # RCT by age band for this sex
      rct_age_results <- purrr::map_dfr(age_bands, function(age) {
        data_age <- data_sex_rct %>%
          filter(age_band == age)

        if (nrow(data_age) == 0) {
          return(tibble())
        }

        result <- run_single_ma(data_age, urine_component, sex, age)
        result %>% mutate(rct_status = "RCT only")
      })

      rct_sex_results <- bind_rows(rct_overall, rct_age_results)
    } else {
      rct_sex_results <- tibble()
    }

    # Combine overall, age band, meta-regression, and RCT results
    bind_rows(overall_result, age_band_results, meta_reg_result, rct_sex_results)
  })

  # --- 5. Combine all results --------------------
  combined_results <- bind_rows(main_result, main_rct_result, sex_age_results) %>%
    mutate(
      sex_group = factor(sex_group,
                        levels = c("main meta-analysis", "male", "female")),
      rct_status = ifelse(is.na(rct_status), "All studies", rct_status)
    ) %>%
    arrange(sex_group, rct_status, age_band)

  # --- 6. Create plots and tables for each sex group (All studies) --------------------
  sex_groups <- c("main meta-analysis", "male", "female")

  output_list <- purrr::map(sex_groups, function(sex) {

    # Filter data for this sex group (All studies only)
    sex_data <- combined_results %>%
      filter(sex_group == sex, rct_status == "All studies")

    # Create forest plot
    colour <- case_when(
      sex == "main meta-analysis" ~ "springgreen3",
      sex == "male" ~ "skyblue2",
      sex == "female" ~ "indianred1"
    )

    plot <- sex_data %>%
      filter(!is.na(pooled_estimate)) %>%
      mutate(age_band = fct_rev(age_band)) %>%
      ggplot(aes(y = age_band)) +
      geom_errorbarh(aes(xmin = lower_95, xmax = upper_95),
                     height = 0.25, colour = colour) +
      geom_point(aes(x = pooled_estimate), size = 3, colour = colour) +
      labs(
        title = paste0(str_to_title(sex)),
        x = "Estimated 95% CI",
        y = "Age Band"
      ) +
      theme_minimal() +
      theme(
        plot.title = element_text(size = 12, face = "bold"),
        axis.title = element_text(size = 10)
      )

    list(
      plot = plot,
      data = sex_data
    )
  })

  names(output_list) <- sex_groups

  # --- 6b. Create RCT-only plots for each sex group --------------------
  rct_output_list <- purrr::map(sex_groups, function(sex) {

    # Filter data for this sex group (RCT only)
    sex_rct_data <- combined_results %>%
      filter(sex_group == sex, rct_status == "RCT only")

    # Only create plot if there's data
    if (nrow(sex_rct_data %>% filter(!is.na(pooled_estimate))) == 0) {
      return(NULL)
    }

    # Create forest plot
    colour <- case_when(
      sex == "main meta-analysis" ~ "springgreen3",
      sex == "male" ~ "skyblue2",
      sex == "female" ~ "indianred1"
    )

    plot <- sex_rct_data %>%
      filter(!is.na(pooled_estimate)) %>%
      mutate(age_band = fct_rev(age_band)) %>%
      ggplot(aes(y = age_band)) +
      geom_errorbarh(aes(xmin = lower_95, xmax = upper_95),
                     height = 0.25, colour = colour) +
      geom_point(aes(x = pooled_estimate), size = 3, colour = colour) +
      labs(
        title = paste0(str_to_title(sex), " (RCT Only)"),
        x = "Estimated 95% CI",
        y = "Age Band"
      ) +
      theme_minimal() +
      theme(
        plot.title = element_text(size = 12, face = "bold"),
        axis.title = element_text(size = 10)
      )

    list(
      plot = plot,
      data = sex_rct_data
    )
  })

  names(rct_output_list) <- sex_groups

  # --- 7. Wald tests for comparisons --------------------

  # Helper function for Wald test
  wald_test <- function(mean1, mean2, sd1, sd2) {
    wald_result <- (mean1 - mean2) / sqrt(sd1^2 + sd2^2)
    p <- 2 * (1 - pnorm(abs(wald_result)))
    return(p)
  }

  # Sex comparisons (Overall estimates, All studies)
  male_overall <- combined_results %>%
    filter(sex_group == "male", as.character(age_band) == "Overall", rct_status == "All studies")
  female_overall <- combined_results %>%
    filter(sex_group == "female", as.character(age_band) == "Overall", rct_status == "All studies")

  sex_comparison <- tibble(
    Comparison = "Male vs Female (Overall)",
    `P (Wald Test)` = wald_test(
      male_overall$pooled_estimate,
      female_overall$pooled_estimate,
      male_overall$pooled_sd,
      female_overall$pooled_sd
    )
  )

  # RCT vs All studies comparison (Overall)
  rct_comparisons <- purrr::map_dfr(sex_groups, function(sex) {
    all_studies <- combined_results %>%
      filter(sex_group == sex, as.character(age_band) == "Overall",
             rct_status == "All studies")
    rct_studies <- combined_results %>%
      filter(sex_group == sex, as.character(age_band) == "Overall",
             rct_status == "RCT only")

    if (nrow(rct_studies) == 0 || is.na(rct_studies$pooled_estimate)) {
      return(tibble())
    }

    tibble(
      `Sex Group` = sex,
      Comparison = "All Studies vs RCT Only",
      `P (Wald Test)` = wald_test(
        all_studies$pooled_estimate,
        rct_studies$pooled_estimate,
        all_studies$pooled_sd,
        rct_studies$pooled_sd
      )
    )
  })

  # Age band comparisons within each sex group (All studies)
  age_band_comparisons <- purrr::map_dfr(sex_groups, function(sex) {
    sex_data <- combined_results %>%
      filter(sex_group == sex,
             rct_status == "All studies",
             as.character(age_band) != "Overall",
             as.character(age_band) != "Meta-Regression") %>%
      arrange(age_band)

    if (nrow(sex_data) < 2) {
      return(tibble())
    }

    # Pairwise comparisons between age bands
    age_bands <- unique(as.character(sex_data$age_band))
    comparisons <- combn(age_bands, 2, simplify = FALSE)

    purrr::map_dfr(comparisons, function(pair) {
      data1 <- sex_data %>% filter(as.character(age_band) == pair[1])
      data2 <- sex_data %>% filter(as.character(age_band) == pair[2])

      tibble(
        `Sex Group` = sex,
        Comparison = paste0(pair[1], " vs ", pair[2]),
        `P (Wald Test)` = wald_test(
          data1$pooled_estimate,
          data2$pooled_estimate,
          data1$pooled_sd,
          data2$pooled_sd
        )
      )
    })
  })

  # Sex comparison within each age band (All studies)
  age_band_sex_comparisons <- purrr::map_dfr(age_bands_main, function(age) {
    male_data <- combined_results %>%
      filter(sex_group == "male", rct_status == "All studies",
             as.character(age_band) == as.character(age))
    female_data <- combined_results %>%
      filter(sex_group == "female", rct_status == "All studies",
             as.character(age_band) == as.character(age))

    if (nrow(male_data) == 0 || nrow(female_data) == 0) {
      return(tibble())
    }

    tibble(
      `Age Band` = as.character(age),
      `P (Wald Test)` = wald_test(
        male_data$pooled_estimate,
        female_data$pooled_estimate,
        male_data$pooled_sd,
        female_data$pooled_sd
      )
    )
  })

  # Format all comparison tables
  format_wald_table <- function(df) {
    if (nrow(df) == 0) return(NULL)

    n_rows <- nrow(df)
    df %>%
      mutate(
        `P (Wald Test)` = case_when(
          `P (Wald Test)` < 0.001 ~ "<0.001",
          `P (Wald Test)` < 0.01  ~ "<0.01",
          `P (Wald Test)` < 0.05  ~ "<0.05",
          TRUE ~ sprintf("%.3f", `P (Wald Test)`)
        )
      ) %>%
      gt() %>%
      cols_align(align = "center", columns = everything()) %>%
      tab_style(
        style = cell_text(weight = "bold"),
        locations = cells_column_labels()
      ) %>%
      tab_style(
        style = cell_fill(color = "#f7f7f7"),
        locations = cells_body(rows = seq(1, n_rows, by = 2))
      ) %>%
      tab_options(
        table.font.size = px(11),
        column_labels.font.weight = "bold",
        data_row.padding = px(3)
      )
  }

  sex_comparison_gt <- sex_comparison %>%
    format_wald_table() %>%
    tab_header(title = md("**Sex Comparison (Overall)**"))

  rct_comparisons_gt <- if (nrow(rct_comparisons) > 0) {
    rct_comparisons %>%
      format_wald_table() %>%
      tab_header(title = md("**All Studies vs RCT Only**"))
  } else {
    NULL
  }

  age_band_comparisons_gt <- if (nrow(age_band_comparisons) > 0) {
    age_band_comparisons %>%
      format_wald_table() %>%
      tab_header(title = md("**Age Band Comparisons Within Sex Groups**"))
  } else {
    NULL
  }

  age_band_sex_comparisons_gt <- if (nrow(age_band_sex_comparisons) > 0) {
    age_band_sex_comparisons %>%
      format_wald_table() %>%
      tab_header(title = md("**Sex Comparisons Within Age Bands**"))
  } else {
    NULL
  }

  # --- 8. Create combined summary table --------------------
  combined_summary_table <- combined_results %>%
    select(-urine_component) %>%
    mutate(
      `Sex Group` = as.character(sex_group),
      `RCT Status` = rct_status,
      `Age Band` = as.character(age_band),
      `95% Range` = ifelse(
        is.na(lower_95) | is.na(upper_95),
        "—",
        sprintf("%.2f – %.2f", lower_95, upper_95)
      ),
      Studies = ifelse(is.na(n_studies), "—", as.character(n_studies)),
      "Study Type" = ifelse(is.na(study_type), "—", study_type),
      Participants = ifelse(is.na(total_participants), "—", as.character(total_participants)),
      I2 = I2 * 100,
      `I²` = ifelse(is.na(I2), "—", paste0(round(I2, 1), "%")),
      `Cochran's Q` = ifelse(is.na(Q), "—", as.character(round(Q, 1))),
      `Q (p-value)` = case_when(
        is.na(Q_pval) ~ "—",
        Q_pval < 0.005 ~ "<0.005",
        Q_pval < 0.01  ~ "<0.01",
        Q_pval < 0.05  ~ "<0.05",
        TRUE ~ sprintf("%.2f", Q_pval)
      ),
      `Egger's p` = case_when(
        is.na(eggers_p) ~ "—",
        eggers_p < 0.001 ~ "<0.001",
        eggers_p < 0.01  ~ "<0.01",
        eggers_p < 0.05  ~ "<0.05",
        TRUE ~ sprintf("%.2f", eggers_p)
      ),
      "Model" = ifelse(is.na(model_utilised), "—", model_utilised)
    ) %>%
    select(
      `Sex Group`,
      `RCT Status`,
      `Age Band`,
      `95% Range`,
      Studies,
      "Study Type",
      Participants,
      `I²`,
      `Cochran's Q`,
      `Q (p-value)`,
      `Egger's p`,
      Model
    )

  combined_summary_gt <- combined_summary_table %>%
    gt() %>%
    tab_header(
      title = md("**Combined Meta-Analysis Results**"),
      subtitle = paste0("24-Hour Urine ", urine_component_for_title)
    ) %>%
    cols_align(align = "center") %>%
    tab_style(
      style = cell_text(weight = "bold"),
      locations = cells_column_labels()
    ) %>%
    # Highlight by sex group
    tab_style(
      style = cell_fill(color = "springgreen3"),
      locations = cells_body(rows = `Sex Group` == "main meta-analysis")
    ) %>%
    tab_style(
      style = cell_fill(color = "skyblue2"),
      locations = cells_body(rows = `Sex Group` == "male")
    ) %>%
    tab_style(
      style = cell_fill(color = "indianred1"),
      locations = cells_body(rows = `Sex Group` == "female")
    ) %>%
    # Bold the Overall rows
    tab_style(
      style = cell_text(weight = "bold"),
      locations = cells_body(
        columns = everything(),
        rows = `Age Band` == "Overall"
      )
    ) %>%
    # Bold and highlight Meta-Regression rows
    tab_style(
      style = cell_text(weight = "bold"),
      locations = cells_body(
        columns = everything(),
        rows = `Age Band` == "Meta-Regression"
      )
    ) %>%
    # Highlight RCT only rows
    tab_style(
      style = cell_fill(color = "#ffffcc"),
      locations = cells_body(rows = `RCT Status` == "RCT only")
    ) %>%
    tab_options(
      table.font.size = px(11),
      heading.title.font.size = px(14),
      heading.subtitle.font.size = px(12),
      column_labels.font.weight = "bold",
      data_row.padding = px(3)
    ) %>%
    tab_source_note(
      source_note = md("*I² represents heterogeneity; Q test assesses between-study variation; Egger's p = linear regression test for funnel plot asymmetry; Meta-Regression shows covariate-adjusted estimates*")
    )

  # --- 9. Create overall comparison plot --------------------

  design <- "ABC
             EEE"

  # Row 1: Three forest plots
  row1 <- (
      output_list[["main meta-analysis"]]$plot |
        output_list[["male"]]$plot |
        output_list[["female"]]$plot
    )

  # Row 2: Combined summary table
  row2 <- wrap_elements(full = combined_summary_gt)

  # Combine rows
  final_layout <- row1 + row2 +
    plot_layout(design = design,
                axis_titles = "collect",
                axes = "collect") +
    plot_annotation(
      title = urine_component_for_title,
      tag_levels = "A",
      theme = theme(plot.title = element_text(face = "bold", size = 14))
    )

  # --- 9b. Create RCT-only comparison plot --------------------
  rct_plots_exist <- !is.null(rct_output_list[["main meta-analysis"]]) ||
                     !is.null(rct_output_list[["male"]]) ||
                     !is.null(rct_output_list[["female"]])

  if (rct_plots_exist) {
    rct_plot_list <- list()
    if (!is.null(rct_output_list[["main meta-analysis"]])) {
      rct_plot_list <- c(rct_plot_list, list(rct_output_list[["main meta-analysis"]]$plot))
    }
    if (!is.null(rct_output_list[["male"]])) {
      rct_plot_list <- c(rct_plot_list, list(rct_output_list[["male"]]$plot))
    }
    if (!is.null(rct_output_list[["female"]])) {
      rct_plot_list <- c(rct_plot_list, list(rct_output_list[["female"]]$plot))
    }

    # Create RCT summary table
    rct_summary_table <- combined_results %>%
      filter(rct_status == "RCT only") %>%
      select(-urine_component) %>%
      mutate(
        `Sex Group` = as.character(sex_group),
        `Age Band` = as.character(age_band),
        `95% Range` = ifelse(
          is.na(lower_95) | is.na(upper_95),
          "—",
          sprintf("%.2f – %.2f", lower_95, upper_95)
        ),
        Studies = ifelse(is.na(n_studies), "—", as.character(n_studies)),
        "Study Type" = ifelse(is.na(study_type), "—", study_type),
        Participants = ifelse(is.na(total_participants), "—", as.character(total_participants)),
        I2_val = I2 * 100,
        `I²` = ifelse(is.na(I2_val), "—", paste0(round(I2_val, 1), "%")),
        `Cochran's Q` = ifelse(is.na(Q), "—", as.character(round(Q, 1))),
        `Q (p-value)` = case_when(
          is.na(Q_pval) ~ "—",
          Q_pval < 0.005 ~ "<0.005",
          Q_pval < 0.01  ~ "<0.01",
          Q_pval < 0.05  ~ "<0.05",
          TRUE ~ sprintf("%.2f", Q_pval)
        ),
        `Egger's p` = case_when(
          is.na(eggers_p) ~ "—",
          eggers_p < 0.001 ~ "<0.001",
          eggers_p < 0.01  ~ "<0.01",
          eggers_p < 0.05  ~ "<0.05",
          TRUE ~ sprintf("%.2f", eggers_p)
        ),
        "Model" = ifelse(is.na(model_utilised), "—", model_utilised)
      ) %>%
      select(
        `Sex Group`,
        `Age Band`,
        `95% Range`,
        Studies,
        "Study Type",
        Participants,
        `I²`,
        `Cochran's Q`,
        `Q (p-value)`,
        `Egger's p`,
        Model
      )

    rct_summary_gt <- rct_summary_table %>%
      gt() %>%
      tab_header(
        title = md("**RCT-Only Meta-Analysis Results**"),
        subtitle = paste0("24-Hour Urine ", urine_component_for_title)
      ) %>%
      cols_align(align = "center") %>%
      tab_style(
        style = cell_text(weight = "bold"),
        locations = cells_column_labels()
      ) %>%
      tab_style(
        style = cell_fill(color = "springgreen3"),
        locations = cells_body(rows = `Sex Group` == "main meta-analysis")
      ) %>%
      tab_style(
        style = cell_fill(color = "skyblue2"),
        locations = cells_body(rows = `Sex Group` == "male")
      ) %>%
      tab_style(
        style = cell_fill(color = "indianred1"),
        locations = cells_body(rows = `Sex Group` == "female")
      ) %>%
      tab_style(
        style = cell_text(weight = "bold"),
        locations = cells_body(
          columns = everything(),
          rows = `Age Band` == "Overall"
        )
      ) %>%
      tab_options(
        table.font.size = px(11),
        heading.title.font.size = px(14),
        heading.subtitle.font.size = px(12),
        column_labels.font.weight = "bold",
        data_row.padding = px(3)
      ) %>%
      tab_source_note(
        source_note = md("*I² represents heterogeneity; Q test assesses between-study variation; Egger's p = linear regression test for funnel plot asymmetry; — indicates insufficient data*")
      )

    if (length(rct_plot_list) == 1) {
      rct_row1 <- rct_plot_list[[1]]
    } else if (length(rct_plot_list) == 2) {
      rct_row1 <- rct_plot_list[[1]] | rct_plot_list[[2]]
    } else {
      rct_row1 <- rct_plot_list[[1]] | rct_plot_list[[2]] | rct_plot_list[[3]]
    }

    rct_row2 <- wrap_elements(full = rct_summary_gt)

    rct_layout <- rct_row1 + rct_row2 +
      plot_layout(design = design,
                  axis_titles = "collect",
                  axes = "collect") +
      plot_annotation(
        title = paste0(urine_component_for_title, " - RCT Only"),
        tag_levels = "A",
        theme = theme(plot.title = element_text(face = "bold", size = 14))
      )
  } else {
    rct_layout <- NULL
    rct_summary_gt <- NULL
  }

  # --- 10. Return results --------------------
  return(list(
    ma_results = combined_results,
    main_meta_analysis = output_list[["main meta-analysis"]],
    male = output_list[["male"]],
    female = output_list[["female"]],
    combined_plot = final_layout,
    combined_summary_table = combined_summary_gt,
    rct_plot = rct_layout,
    rct_summary_table = rct_summary_gt,
    rct_main = rct_output_list[["main meta-analysis"]],
    rct_male = rct_output_list[["male"]],
    rct_female = rct_output_list[["female"]],
    wald_tests = list(
      sex_comparison = sex_comparison_gt,
      rct_comparison = rct_comparisons_gt,
      age_band_comparisons = age_band_comparisons_gt,
      sex_within_age_bands = age_band_sex_comparisons_gt
    )
  ))
}
```

#### 5.7 Comparison to Age-Band subdivisions

```
sex_age_subdivision_ma <- function(urine_component, data = urine_ma2) {

  # --- 1. Identify the correct columns -------------------------------
  mean_colname <- case_when(
    urine_component == "volume" ~ "volume_mean_m_l",
    urine_component == "ph" ~ "ph_mean",
    urine_component == "creat" ~ "creatinine_mean_mmol_24h",
    TRUE ~ paste0(urine_component, "_mean_mmol_24h")
  )

  sd_colname <- case_when(
    urine_component == "volume" ~ "volume_sd_m_l",
    urine_component == "ph" ~ "ph_sd",
    urine_component == "creat" ~ "creatinine_sd_mmol_24h",
    TRUE ~ paste0(urine_component, "_sd_mmol_24h")
  )

  urine_component_for_title <- case_when(
    urine_component == "volume" ~ "Volume (L/24h)",
    urine_component == "ph" ~ "pH",
    urine_component == "creat" ~ "Creatinine (mmol/24h)",
    TRUE ~ paste0(str_to_title(urine_component), " (mmol/24h)")
  )

  # --- 2. Prepare data with overall_bias for meta-regression --------
  data_with_bias <- data %>%
    left_join(overall_bias, by = "study_name")

  # --- 3. Get main meta-analysis result ----------------------------
  # Overall main result (all sexes, all ages)
  main_overall <- main_ma(urine_component, data = data) %>%
    mutate(sex_group = "main meta-analysis",
           age_band = "Overall")

  # Main meta-analysis by age band (all sexes combined)
  data_main <- data %>%
    select(
      study_name,
      total_n,
      age_band,
      mean_col = all_of(mean_colname),
      sd_col   = all_of(sd_colname),
      study_type
    ) %>%
    drop_na(mean_col, sd_col)

  age_bands_main <- data_main %>%
    filter(!is.na(age_band)) %>%
    pull(age_band) %>%
    unique() %>%
    sort()

  main_age_results <- purrr::map_dfr(age_bands_main, function(age) {
    data_age <- data_main %>%
      filter(age_band == age)

    if (nrow(data_age) == 0) {
      return(tibble())
    }

    run_single_ma(data_age, urine_component, "main meta-analysis", age)
  })

  # Add meta-regression for main analysis
  main_meta_reg_results <- tryCatch({
    meta_regression("none", urine_component = urine_component)
  }, error = function(e) {
    message("Meta-regression failed for main analysis: ", e$message)
    NULL
  })

  main_result <- bind_rows(main_overall, main_age_results, main_meta_reg_results)

  # Sort Study type column
  data_main$study_type <- as.factor(data_main$study_type)
  study_type_main <- if (dplyr::n_distinct(data_main$study_type, na.rm = TRUE) > 1) {
    "Mixed"
  } else {
    unique(na.omit(data_main$study_type))
  }

  # --- 3b. RCT-only main analysis ----------------------------
  data_main_rct <- data %>%
    filter(study_type == "RCT") %>%
    select(
      study_name,
      total_n,
      age_band,
      study_type,
      mean_col = all_of(mean_colname),
      sd_col   = all_of(sd_colname)
    ) %>%
    drop_na(mean_col, sd_col)

  if (nrow(data_main_rct) > 0) {
    # Overall RCT result
    main_rct_overall <- run_single_ma(data_main_rct, urine_component, "main meta-analysis", "Overall (RCT only)")

    # RCT by age band
    main_rct_age_results <- purrr::map_dfr(age_bands_main, function(age) {
      data_age <- data_main_rct %>%
        filter(age_band == age)

      if (nrow(data_age) == 0) {
        return(tibble())
      }

      run_single_ma(data_age, urine_component, "main meta-analysis", paste0(age, " (RCT only)"))
    })

    main_rct_result <- bind_rows(main_rct_overall, main_rct_age_results)
  } else {
    main_rct_result <- tibble()
  }

  # --- 4. Iterate through sexes and age bands --------------------------------
  sex_age_results <- purrr::map_dfr(c("male", "female"), function(sex) {

    # First get overall result for this sex (all age bands)
    data_sex <- data %>%
      filter(male_female == sex) %>%
      select(
        study_name,
        total_n,
        age_band,
        mean_col = all_of(mean_colname),
        sd_col   = all_of(sd_colname),
        study_type
      ) %>%
      drop_na(mean_col, sd_col)

    if (nrow(data_sex) == 0) {
      return(tibble())
    }

    # Get overall result for this sex
    overall_result <- run_single_ma(data_sex, urine_component, sex, "Overall")

    # Sort Study type column
    data_sex$study_type <- as.factor(data_sex$study_type)
    study_type_sex <- if (dplyr::n_distinct(data_sex$study_type, na.rm = TRUE) > 1) {
      "Mixed"
    } else {
      unique(na.omit(data_sex$study_type))
    }

    # Get meta-regression result for this sex
    meta_reg_result <- tryCatch({
      meta_regression(data_subset = sex, urine_component = urine_component)
    }, error = function(e) {
      message("Meta-regression failed for ", sex, ": ", e$message)
      NULL
    })

    # Get unique age bands
    age_bands <- data_sex %>%
      filter(!is.na(age_band)) %>%
      pull(age_band) %>%
      unique() %>%
      sort()

    # Get results for each age band
    age_band_results <- purrr::map_dfr(age_bands, function(age) {
      data_age <- data_sex %>%
        filter(age_band == age)

      if (nrow(data_age) == 0) {
        return(tibble())
      }

      run_single_ma(data_age, urine_component, sex, age)
    })

    # --- RCT-only analyses for this sex ---
    data_sex_rct <- data_sex %>%
      filter(study_type == "RCT")

    if (nrow(data_sex_rct) > 0) {
      # Overall RCT for this sex
      rct_overall <- run_single_ma(data_sex_rct, urine_component, sex, "Overall (RCT only)")

      # RCT by age band for this sex
      rct_age_results <- purrr::map_dfr(age_bands, function(age) {
        data_age <- data_sex_rct %>%
          filter(age_band == age)

        if (nrow(data_age) == 0) {
          return(tibble())
        }

        run_single_ma(data_age, urine_component, sex, paste0(age, " (RCT only)"))
      })

      rct_sex_results <- bind_rows(rct_overall, rct_age_results)
    } else {
      rct_sex_results <- tibble()
    }

    # Combine overall, age band, meta-regression, and RCT results
    bind_rows(overall_result, age_band_results, meta_reg_result, rct_sex_results)
  })

  # --- 4b. Quality sensitivity analyses (Low bias only; Low + Moderate bias) ----
  # Join data with bias ratings for quality filtering
  data_with_quality <- data_with_bias

  quality_filters <- list(
    "Low Risk of Bias Only" = c("Low"),
    "Low + Moderate Risk of Bias" = c("Low", "Moderate")
  )

  quality_results <- purrr::map_dfr(names(quality_filters), function(filter_name) {
    filter_levels <- quality_filters[[filter_name]]
    data_quality <- data_with_quality %>%
      filter(overall %in% filter_levels)

    if (nrow(data_quality) == 0) {
      message("No studies match quality filter '", filter_name, "' for ", urine_component)
      return(tibble())
    }

    # Prepare quality-filtered data
    data_q_main <- data_quality %>%
      select(
        study_name, total_n, age_band,
        mean_col = all_of(mean_colname),
        sd_col   = all_of(sd_colname),
        study_type
      ) %>%
      drop_na(mean_col, sd_col)

    if (nrow(data_q_main) == 0) return(tibble())

    # --- Main MA (overall) ---
    q_main_overall <- tryCatch({
      main_ma(urine_component, data = data_quality) %>%
        mutate(sex_group = "main meta-analysis", age_band = "Overall")
    }, error = function(e) {
      message("Quality MA failed (main overall, ", filter_name, "): ", e$message)
      tibble()
    })

    # --- Main MA by age band ---
    q_age_bands <- data_q_main %>%
      filter(!is.na(age_band)) %>%
      pull(age_band) %>%
      unique() %>%
      sort()

    q_main_age_results <- purrr::map_dfr(q_age_bands, function(age) {
      data_age <- data_q_main %>% filter(age_band == age)
      if (nrow(data_age) == 0) return(tibble())
      tryCatch(
        run_single_ma(data_age, urine_component, "main meta-analysis", age),
        error = function(e) tibble()
      )
    })

    # --- Sex subdivision ---
    q_sex_results <- purrr::map_dfr(c("male", "female"), function(sex) {
      data_sex <- data_quality %>%
        filter(male_female == sex) %>%
        select(
          study_name, total_n, age_band,
          mean_col = all_of(mean_colname),
          sd_col   = all_of(sd_colname),
          study_type
        ) %>%
        drop_na(mean_col, sd_col)

      if (nrow(data_sex) == 0) return(tibble())

      # Overall for this sex
      sex_overall <- tryCatch(
        run_single_ma(data_sex, urine_component, sex, "Overall"),
        error = function(e) tibble()
      )

      # By age band for this sex
      age_bands_sex <- data_sex %>%
        filter(!is.na(age_band)) %>%
        pull(age_band) %>%
        unique() %>%
        sort()

      sex_age_results_q <- purrr::map_dfr(age_bands_sex, function(age) {
        data_age <- data_sex %>% filter(age_band == age)
        if (nrow(data_age) == 0) return(tibble())
        tryCatch(
          run_single_ma(data_age, urine_component, sex, age),
          error = function(e) tibble()
        )
      })

      bind_rows(sex_overall, sex_age_results_q)
    })

    bind_rows(q_main_overall, q_main_age_results, q_sex_results) %>%
      mutate(quality_filter = filter_name)
  })

  # --- 5. Combine all results --------------------
  combined_results <- bind_rows(
    main_result %>% mutate(quality_filter = "All Studies"),
    main_rct_result %>% mutate(quality_filter = "All Studies"),
    sex_age_results %>% mutate(quality_filter = "All Studies"),
    quality_results
  ) %>%
    mutate(
      sex_group = factor(sex_group,
                         levels = c("main meta-analysis", "male", "female"))
    ) %>%
    arrange(quality_filter, sex_group, age_band)

  # --- 6. Create plots and tables for each sex group --------------------
  sex_groups <- c("main meta-analysis", "male", "female")
  quality_groups <- unique(combined_results$quality_filter)

  # Build integrated forest plots — 3 quality levels per age band, one plot per sex group
  # Colour = sex group (matching table row colours), Shape = quality filter
  sex_colours <- c(
    "main meta-analysis" = "springgreen3",
    "male" = "skyblue2",
    "female" = "indianred1"
  )

  quality_shapes <- c(
    "All Studies" = 16,
    "Low Risk of Bias Only" = 18,
    "Low + Moderate Risk of Bias" = 17
  )

  build_integrated_plots <- function() {
    plots <- purrr::map(sex_groups, function(sex) {
      sex_data <- combined_results %>%
        filter(sex_group == sex, !is.na(pooled_estimate))

      if (nrow(sex_data) == 0) {
        plot <- ggplot() + theme_void() +
          labs(title = str_to_title(sex))
        return(list(plot = plot, data = sex_data))
      }

      sex_col <- sex_colours[[sex]]

      plot <- sex_data %>%
        mutate(
          age_band = fct_rev(age_band),
          quality_filter = factor(quality_filter,
            levels = c("Low Risk of Bias Only", "Low + Moderate Risk of Bias", "All Studies"))
        ) %>%
        ggplot(aes(y = age_band, shape = quality_filter)) +
        geom_errorbarh(aes(xmin = lower_95, xmax = upper_95),
                       height = 0.2, colour = sex_col,
                       position = position_dodge(width = 0.6)) +
        geom_point(aes(x = pooled_estimate), size = 3, colour = sex_col,
                   position = position_dodge(width = 0.6)) +
        scale_shape_manual(
          values = quality_shapes,
          name = "Quality Filter"
        ) +
        labs(
          title = str_to_title(sex),
          x = "Estimated 95% CI",
          y = "Age Band"
        ) +
        theme_minimal() +
        theme(
          plot.title = element_text(size = 12, face = "bold"),
          axis.title = element_text(size = 10),
          legend.position = "bottom",
          legend.direction = "vertical",
          legend.title = element_text(size = 9),
          legend.text = element_text(size = 8)
        )

      list(plot = plot, data = sex_data)
    })
    names(plots) <- sex_groups
    plots
  }

  output_list <- build_integrated_plots()

  # --- 7. Wald tests for comparisons --------------------

  # Helper: Wald test returning mean diff, 95% CI, % shift, and p-value
  wald_test <- function(mean1, mean2, sd1, sd2, ref_for_pct = NULL) {
    if (any(is.na(c(mean1, mean2, sd1, sd2)))) {
      return(list(diff = NA_real_, lower = NA_real_, upper = NA_real_,
                  pct_shift = NA_real_, p = NA_real_))
    }
    diff   <- mean1 - mean2
    se_dif <- sqrt(sd1^2 + sd2^2)
    lower  <- diff - 1.96 * se_dif
    upper  <- diff + 1.96 * se_dif
    z      <- diff / se_dif
    p      <- 2 * (1 - pnorm(abs(z)))
    ref    <- if (is.null(ref_for_pct)) mean2 else ref_for_pct
    pct    <- if (!is.na(ref) && ref != 0) 100 * diff / ref else NA_real_
    list(diff = diff, lower = lower, upper = upper, pct_shift = pct, p = p)
  }

  # Sex comparisons (Overall estimates — All Studies only)
  male_overall <- combined_results %>%
    filter(sex_group == "male", as.character(age_band) == "Overall", quality_filter == "All Studies")
  female_overall <- combined_results %>%
    filter(sex_group == "female", as.character(age_band) == "Overall", quality_filter == "All Studies")

  sex_w <- wald_test(male_overall$pooled_estimate,
                     female_overall$pooled_estimate,
                     male_overall$pooled_sd,
                     female_overall$pooled_sd)
  sex_comparison <- tibble(
    Comparison = "Male vs Female (Overall)",
    `Mean Difference` = sex_w$diff,
    `95% CI Lower`    = sex_w$lower,
    `95% CI Upper`    = sex_w$upper,
    `% Shift`         = sex_w$pct_shift,
    `P (Wald Test)`   = sex_w$p
  )

  # RCT vs All studies comparison (All Studies quality filter only)
  rct_comparisons <- purrr::map_dfr(sex_groups, function(sex) {
    all_studies <- combined_results %>%
      filter(sex_group == sex, as.character(age_band) == "Overall", quality_filter == "All Studies")
    rct_studies <- combined_results %>%
      filter(sex_group == sex, as.character(age_band) == "Overall (RCT only)", quality_filter == "All Studies")

    if (nrow(rct_studies) == 0 || is.na(rct_studies$pooled_estimate)) {
      return(tibble())
    }
    w <- wald_test(all_studies$pooled_estimate, rct_studies$pooled_estimate,
                   all_studies$pooled_sd,       rct_studies$pooled_sd)
    tibble(
      `Sex Group`        = sex,
      Comparison         = "All Studies vs RCT Only",
      `Mean Difference`  = w$diff,
      `95% CI Lower`     = w$lower,
      `95% CI Upper`     = w$upper,
      `% Shift`          = w$pct_shift,
      `P (Wald Test)`    = w$p
    )
  })

  # Age band comparisons within each sex group (All Studies only)
  age_band_comparisons <- purrr::map_dfr(sex_groups, function(sex) {
    sex_data <- combined_results %>%
      filter(sex_group == sex,
             quality_filter == "All Studies",
             as.character(age_band) != "Overall",
             as.character(age_band) != "Meta-Regression",
             !grepl("RCT only", as.character(age_band))) %>%
      arrange(age_band)

    if (nrow(sex_data) < 2) {
      return(tibble())
    }

    age_bands <- unique(as.character(sex_data$age_band))
    comparisons <- combn(age_bands, 2, simplify = FALSE)

    purrr::map_dfr(comparisons, function(pair) {
      data1 <- sex_data %>% filter(as.character(age_band) == pair[1])
      data2 <- sex_data %>% filter(as.character(age_band) == pair[2])
      w <- wald_test(data1$pooled_estimate, data2$pooled_estimate,
                     data1$pooled_sd,       data2$pooled_sd)
      tibble(
        `Sex Group`        = sex,
        Comparison         = paste0(pair[1], " vs ", pair[2]),
        `Mean Difference`  = w$diff,
        `95% CI Lower`     = w$lower,
        `95% CI Upper`     = w$upper,
        `% Shift`          = w$pct_shift,
        `P (Wald Test)`    = w$p
      )
    })
  })

  # Sex comparison within each age band (All Studies only)
  age_band_sex_comparisons <- purrr::map_dfr(age_bands_main, function(age) {
    male_data <- combined_results %>%
      filter(sex_group == "male", as.character(age_band) == as.character(age), quality_filter == "All Studies")
    female_data <- combined_results %>%
      filter(sex_group == "female", as.character(age_band) == as.character(age), quality_filter == "All Studies")

    if (nrow(male_data) == 0 || nrow(female_data) == 0) {
      return(tibble())
    }
    w <- wald_test(male_data$pooled_estimate, female_data$pooled_estimate,
                   male_data$pooled_sd,       female_data$pooled_sd)
    tibble(
      `Age Band`         = as.character(age),
      `Mean Difference`  = w$diff,
      `95% CI Lower`     = w$lower,
      `95% CI Upper`     = w$upper,
      `% Shift`          = w$pct_shift,
      `P (Wald Test)`    = w$p
    )
  })

  # Format all comparison tables — collapse diff + CI into one column,
  # show % shift (relative to the reference group), and the p-value.
  format_wald_table <- function(df) {
    if (nrow(df) == 0) return(NULL)
    n_rows <- nrow(df)

    df %>%
      mutate(
        `Mean Diff (95% CI)` = ifelse(
          is.na(`Mean Difference`), "—",
          sprintf("%.2f [%.2f, %.2f]", `Mean Difference`, `95% CI Lower`, `95% CI Upper`)
        ),
        `% Shift` = ifelse(is.na(`% Shift`), "—",
                           sprintf("%+.1f%%", `% Shift`)),
        `P (Wald Test)` = case_when(
          is.na(`P (Wald Test)`) ~ "—",
          `P (Wald Test)` < 0.001 ~ "<0.001",
          `P (Wald Test)` < 0.01  ~ "<0.01",
          `P (Wald Test)` < 0.05  ~ "<0.05",
          TRUE ~ sprintf("%.3f", `P (Wald Test)`)
        )
      ) %>%
      select(-`Mean Difference`, -`95% CI Lower`, -`95% CI Upper`) %>%
      gt() %>%
      cols_align(align = "center", columns = everything()) %>%
      tab_style(
        style = cell_text(weight = "bold"),
        locations = cells_column_labels()
      ) %>%
      tab_style(
        style = cell_fill(color = "#f7f7f7"),
        locations = cells_body(rows = seq(1, n_rows, by = 2))
      ) %>%
      tab_options(
        table.font.size = px(11),
        column_labels.font.weight = "bold",
        data_row.padding = px(3)
      ) %>%
      tab_source_note(
        md("*Mean difference = Group 1 − Group 2 in component units (mmol/24h, L/24h, or pH); % shift = difference relative to Group 2.*")
      )
  }

  sex_comparison_gt <- sex_comparison %>%
    format_wald_table() %>%
    tab_header(title = md("**Sex Comparison (Overall)**"))

  rct_comparisons_gt <- if (nrow(rct_comparisons) > 0) {
    rct_comparisons %>%
      format_wald_table() %>%
      tab_header(title = md("**All Studies vs RCT Only**"))
  } else {
    NULL
  }

  age_band_comparisons_gt <- if (nrow(age_band_comparisons) > 0) {
    age_band_comparisons %>%
      format_wald_table() %>%
      tab_header(title = md("**Age Band Comparisons Within Sex Groups**"))
  } else {
    NULL
  }

  age_band_sex_comparisons_gt <- if (nrow(age_band_sex_comparisons) > 0) {
    age_band_sex_comparisons %>%
      format_wald_table() %>%
      tab_header(title = md("**Sex Comparisons Within Age Bands**"))
  } else {
    NULL
  }

  # --- 8. Create integrated wide-format summary table --------------------

  # Build wide table: one row per sex/age, columns per quality level
  all_quality_levels <- c("All Studies", "Low Risk of Bias Only", "Low + Moderate Risk of Bias")

  # Helper to format descriptive across-study summary
  fmt_desc <- function(avg, sdv, lo, hi) {
    if (is.na(avg) && is.na(lo) && is.na(hi)) return("\u2014")
    avg_part <- if (!is.na(avg) && !is.na(sdv)) sprintf("%.2f \u00b1 %.2f", avg, sdv) else "\u2014"
    rng_part <- if (!is.na(lo) && !is.na(hi))   sprintf("%.2f\u2013%.2f",  lo, hi)   else "\u2014"
    paste0(avg_part, " (", rng_part, ")")
  }

  # Ensure descriptive columns exist (meta-regression rows may not produce them)
  for (col in c("study_mean_avg", "study_mean_sd", "study_mean_min", "study_mean_max")) {
    if (!col %in% names(combined_results)) combined_results[[col]] <- NA_real_
  }

  wide_data <- combined_results %>%
    select(-urine_component) %>%
    mutate(
      quality_filter = ifelse(is.na(quality_filter), "All Studies", quality_filter),
      pooled_str = ifelse(
        is.na(pooled_estimate), "\u2014",
        sprintf("%.2f", pooled_estimate)
      ),
      ci_str = ifelse(
        is.na(lower_95) | is.na(upper_95), "\u2014",
        sprintf("[%.2f, %.2f]", lower_95, upper_95)
      ),
      desc_str = mapply(fmt_desc, study_mean_avg, study_mean_sd, study_mean_min, study_mean_max),
      k_str = ifelse(is.na(n_studies) | n_studies == 0, "\u2014", as.character(n_studies))
    ) %>%
    select(sex_group, age_band, quality_filter, pooled_str, ci_str, desc_str, k_str, Q_pval, eggers_p) %>%
    distinct(sex_group, age_band, quality_filter, .keep_all = TRUE) %>%
    # Ensure all quality levels exist as columns even if no data
    complete(sex_group, age_band, quality_filter = all_quality_levels) %>%
    pivot_wider(
      id_cols = c(sex_group, age_band),
      names_from = quality_filter,
      values_from = c(pooled_str, ci_str, desc_str, k_str, Q_pval, eggers_p),
      names_glue = "{quality_filter}___{.value}"
    )

  fmt_pval <- function(p) {
    case_when(
      is.na(p) ~ "\u2014",
      p < 0.001 ~ "<0.001",
      p < 0.005 ~ "<0.005",
      p < 0.01  ~ "<0.01",
      p < 0.05  ~ "<0.05",
      TRUE ~ sprintf("%.2f", p)
    )
  }

  combined_summary_table <- wide_data %>%
    transmute(
      `Sex Group` = as.character(sex_group),
      `Age Band` = as.character(age_band),
      `Pooled (All)` = replace_na(`All Studies___pooled_str`, "\u2014"),
      `95% CI (All)` = replace_na(`All Studies___ci_str`, "\u2014"),
      `Studies Mean SD Range (All)` = replace_na(`All Studies___desc_str`, "\u2014"),
      `Studies, n (All)` = replace_na(`All Studies___k_str`, "\u2014"),
      `Q p (All)` = fmt_pval(`All Studies___Q_pval`),
      `Egger's p (All)` = fmt_pval(`All Studies___eggers_p`),
      `Pooled (Low)` = replace_na(`Low Risk of Bias Only___pooled_str`, "\u2014"),
      `95% CI (Low)` = replace_na(`Low Risk of Bias Only___ci_str`, "\u2014"),
      `Studies Mean SD Range (Low)` = replace_na(`Low Risk of Bias Only___desc_str`, "\u2014"),
      `Studies, n (Low)` = replace_na(`Low Risk of Bias Only___k_str`, "\u2014"),
      `Q p (Low)` = fmt_pval(`Low Risk of Bias Only___Q_pval`),
      `Egger's p (Low)` = fmt_pval(`Low Risk of Bias Only___eggers_p`),
      `Pooled (Low+Mod)` = replace_na(`Low + Moderate Risk of Bias___pooled_str`, "\u2014"),
      `95% CI (Low+Mod)` = replace_na(`Low + Moderate Risk of Bias___ci_str`, "\u2014"),
      `Studies Mean SD Range (Low+Mod)` = replace_na(`Low + Moderate Risk of Bias___desc_str`, "\u2014"),
      `Studies, n (Low+Mod)` = replace_na(`Low + Moderate Risk of Bias___k_str`, "\u2014"),
      `Q p (Low+Mod)` = fmt_pval(`Low + Moderate Risk of Bias___Q_pval`),
      `Egger's p (Low+Mod)` = fmt_pval(`Low + Moderate Risk of Bias___eggers_p`)
    )

  combined_summary_gt <- combined_summary_table %>%
    gt() %>%
    tab_header(
      title = md("**Combined Meta-Analysis Results**"),
      subtitle = paste0("24-Hour Urine ", urine_component_for_title)
    ) %>%
    tab_spanner(label = "All Studies", id = "spanner_all",
                columns = c(`Pooled (All)`, `95% CI (All)`, `Studies Mean SD Range (All)`,
                            `Studies, n (All)`, `Q p (All)`, `Egger's p (All)`)) %>%
    tab_spanner(label = "Low Risk of Bias Only", id = "spanner_low",
                columns = c(`Pooled (Low)`, `95% CI (Low)`, `Studies Mean SD Range (Low)`,
                            `Studies, n (Low)`, `Q p (Low)`, `Egger's p (Low)`)) %>%
    tab_spanner(label = "Low + Moderate RoB", id = "spanner_lowmod",
                columns = c(`Pooled (Low+Mod)`, `95% CI (Low+Mod)`, `Studies Mean SD Range (Low+Mod)`,
                            `Studies, n (Low+Mod)`, `Q p (Low+Mod)`, `Egger's p (Low+Mod)`)) %>%
    cols_label(
      `Pooled (All)` = "Pooled",
      `95% CI (All)` = "95% CI",
      `Studies Mean SD Range (All)` = md("Studies<br>Mean \u00b1 SD (Range)"),
      `Studies, n (All)` = "Studies, n",
      `Q p (All)` = "Q p-value",
      `Egger's p (All)` = "Egger's p",
      `Pooled (Low)` = "Pooled",
      `95% CI (Low)` = "95% CI",
      `Studies Mean SD Range (Low)` = md("Studies<br>Mean \u00b1 SD (Range)"),
      `Studies, n (Low)` = "Studies, n",
      `Q p (Low)` = "Q p-value",
      `Egger's p (Low)` = "Egger's p",
      `Pooled (Low+Mod)` = "Pooled",
      `95% CI (Low+Mod)` = "95% CI",
      `Studies Mean SD Range (Low+Mod)` = md("Studies<br>Mean \u00b1 SD (Range)"),
      `Studies, n (Low+Mod)` = "Studies, n",
      `Q p (Low+Mod)` = "Q p-value",
      `Egger's p (Low+Mod)` = "Egger's p"
    ) %>%
    cols_align(align = "center", columns = -c(`Sex Group`, `Age Band`)) %>%
    tab_style(
      style = cell_text(weight = "bold"),
      locations = cells_column_labels()
    ) %>%
    tab_style(
      style = cell_text(weight = "bold"),
      locations = cells_column_spanners()
    ) %>%
    # Highlight by sex group — matching sex_age_race_subdivision_ma colours
    tab_style(
      style = cell_fill(color = "springgreen3"),
      locations = cells_body(rows = `Sex Group` == "main meta-analysis")
    ) %>%
    tab_style(
      style = cell_fill(color = "skyblue2"),
      locations = cells_body(rows = `Sex Group` == "male")
    ) %>%
    tab_style(
      style = cell_fill(color = "indianred1"),
      locations = cells_body(rows = `Sex Group` == "female")
    ) %>%
    # Bold Overall and Meta-Regression rows
    tab_style(
      style = cell_text(weight = "bold"),
      locations = cells_body(rows = `Age Band` == "Overall")
    ) %>%
    tab_style(
      style = cell_text(weight = "bold"),
      locations = cells_body(rows = `Age Band` == "Meta-Regression")
    ) %>%
    tab_style(
      style = cell_text(weight = "bold"),
      locations = cells_body(rows = grepl("RCT only", `Age Band`))
    ) %>%
    tab_options(
      table.font.size = px(11),
      heading.title.font.size = px(14),
      heading.subtitle.font.size = px(12),
      column_labels.font.weight = "bold",
      data_row.padding = px(3)
    ) %>%
    tab_source_note(
      source_note = md("*Studies, n = number of studies; Q p-value = Cochran's Q heterogeneity test; Egger's p = linear regression test for funnel plot asymmetry; RoB = Risk of Bias*")
    )

  # --- 9. Create overall comparison plot (integrated quality) --------------------

  design <- "ABC
             EEE"

  # Row 1: Three integrated forest plots (all quality levels per plot)
  row1 <- (
    output_list[["main meta-analysis"]]$plot |
      output_list[["male"]]$plot |
      output_list[["female"]]$plot
  )

  # Row 2: Combined summary table (wide format)
  row2 <- wrap_elements(full = combined_summary_gt)

  # Combine rows
  final_layout <- row1 + row2 +
    plot_layout(design = design,
                axis_titles = "collect",
                axes = "collect") +
    plot_annotation(
      title = urine_component_for_title,
      tag_levels = "A",
      theme = theme(plot.title = element_text(face = "bold", size = 14))
    )

  # --- 10. Return results --------------------
  return(list(
    ma_results = combined_results,
    main_meta_analysis = output_list[["main meta-analysis"]],
    male = output_list[["male"]],
    female = output_list[["female"]],
    combined_plot = final_layout,
    combined_summary_table = combined_summary_gt,
    wald_tests = list(
      sex_comparison = sex_comparison_gt,
      rct_comparison = rct_comparisons_gt,
      age_band_comparisons = age_band_comparisons_gt,
      sex_within_age_bands = age_band_sex_comparisons_gt
    )
  ))
}
```

#### 5.8 Comparison of Sex, Age and race

```
sex_age_race_subdivision_ma <- function(urine_component,
                                        data,
                                        overall_bias = NULL) {

  # --- 1. Identify the correct columns -------------------------------
  mean_colname <- case_when(
    urine_component == "volume" ~ "volume_mean_m_l",
    urine_component == "ph" ~ "ph_mean",
    urine_component == "creat" ~ "creatinine_mean_mmol_24h",
    TRUE ~ paste0(urine_component, "_mean_mmol_24h")
  )

  sd_colname <- case_when(
    urine_component == "volume" ~ "volume_sd_m_l",
    urine_component == "ph" ~ "ph_sd",
    urine_component == "creat" ~ "creatinine_sd_mmol_24h",
    TRUE ~ paste0(urine_component, "_sd_mmol_24h")
  )

  urine_component_for_title <- case_when(
    urine_component == "volume" ~ "Volume (L/24h)",
    urine_component == "ph" ~ "pH",
    urine_component == "creat" ~ "Creatinine (mmol/24h)",
    TRUE ~ paste0(str_to_title(urine_component), " (mmol/24h)")
  )

  # --- 2. Prepare data with overall_bias for meta-regression --------
  if (!is.null(overall_bias)) {
    data_with_bias <- data %>%
      left_join(overall_bias, by = "study_name")
  } else {
    data_with_bias <- data
  }

  # --- 3. Get main meta-analysis result ----------------------------
  # Overall main result (all sexes, all ages, all races)
  main_overall <- main_ma(urine_component, data = data) %>%
    mutate(sex_group = "main meta-analysis",
           age_band = "Overall",
           race_group = "All")

  # Main meta-analysis by age band (all sexes combined, all races)
  data_main <- data %>%
    select(
      study_name,
      total_n,
      age_band,
      mean_col = all_of(mean_colname),
      sd_col   = all_of(sd_colname)
    ) %>%
    drop_na(mean_col, sd_col)

  age_bands_main <- data_main %>%
    filter(!is.na(age_band)) %>%
    pull(age_band) %>%
    unique() %>%
    sort()

  main_age_results <- purrr::map_dfr(age_bands_main, function(age) {
    data_age <- data_main %>%
      filter(age_band == age)

    if (nrow(data_age) == 0) {
      return(tibble())
    }

    result <- run_single_ma(data_age, urine_component, "main meta-analysis", age)
    result %>% mutate(race_group = "All")
  })

  # Add meta-regression for main analysis
  main_meta_reg_results <- tryCatch({
    result <- meta_regression("none", urine_component = urine_component)
    if (!is.null(result)) {
      result %>% mutate(race_group = "All")
    } else {
      NULL
    }
  }, error = function(e) {
    message("Meta-regression failed for main analysis: ", e$message)
    NULL
  })

  main_result <- bind_rows(main_overall, main_age_results, main_meta_reg_results)

  # --- 4. Iterate through sexes, age bands, and races ----------------------
  sex_age_race_results <- purrr::map_dfr(c("main meta-analysis", "male", "female"), function(sex) {

    # Filter data for this sex
    if (sex == "main meta-analysis") {
      data_sex <- data %>%
        select(
          study_name,
          total_n,
          age_band,
          race,
          mean_col = all_of(mean_colname),
          sd_col   = all_of(sd_colname),
          study_type
        ) %>%
        drop_na(mean_col, sd_col)
    } else {
      data_sex <- data %>%
        filter(male_female == sex) %>%
        select(
          study_name,
          total_n,
          age_band,
          race,
          mean_col = all_of(mean_colname),
          sd_col   = all_of(sd_colname),
          study_type
        ) %>%
        drop_na(mean_col, sd_col)
    }

    if (nrow(data_sex) == 0) {
      return(tibble())
    }

    # Get overall result for this sex (all races)
    overall_result <- run_single_ma(data_sex, urine_component, sex, "Overall") %>%
      mutate(race_group = "All")

    # Get meta-regression result for this sex
    if (sex == "main meta-analysis") {
      meta_reg_result <- tryCatch({
        result <- meta_regression("none", urine_component = urine_component)
        if (!is.null(result)) {
          result %>% mutate(race_group = "All")
        } else {
          NULL
        }
      }, error = function(e) {
        message("Meta-regression failed for ", sex, ": ", e$message)
        NULL
      })
    } else {
      meta_reg_result <- tryCatch({
        result <- meta_regression(data_subset = sex, urine_component = urine_component)
        if (!is.null(result)) {
          result %>% mutate(race_group = "All")
        } else {
          NULL
        }
      }, error = function(e) {
        message("Meta-regression failed for ", sex, ": ", e$message)
        NULL
      })
    }

    # Get unique age bands
    age_bands <- data_sex %>%
      filter(!is.na(age_band)) %>%
      pull(age_band) %>%
      unique() %>%
      sort()

    # Get results for each age band (all races)
    age_band_results <- purrr::map_dfr(age_bands, function(age) {
      data_age <- data_sex %>%
        filter(age_band == age)

      if (nrow(data_age) == 0) {
        return(tibble())
      }

      result <- run_single_ma(data_age, urine_component, sex, age)
      result %>% mutate(race_group = "All")
    })

    # --- NEW: race-specific analyses ---
    # Get unique race groups for this sex
    race_groups <- data_sex %>%
      filter(!is.na(race), race %in% c("white", "mixed", "black")) %>%
      pull(race) %>%
      unique() %>%
      sort()

    race_results <- purrr::map_dfr(race_groups, function(race) {
      data_race <- data_sex %>%
        filter(race == !!race)

      if (nrow(data_race) == 0) {
        return(tibble())
      }

      # Overall for this sex-race combination
      race_overall <- run_single_ma(data_race, urine_component, sex, "Overall") %>%
        mutate(race_group = race)

      # Age bands within this sex-race combination
      race_age_results <- purrr::map_dfr(age_bands, function(age) {
        data_race_age <- data_race %>%
          filter(age_band == age)

        if (nrow(data_race_age) == 0) {
          return(tibble())
        }

        result <- run_single_ma(data_race_age, urine_component, sex, age)
        result %>% mutate(race_group = race)
      })

      bind_rows(race_overall, race_age_results)
    })

    # Combine all results for this sex
    bind_rows(overall_result, age_band_results, meta_reg_result, race_results)
  })

  # --- 5. Combine all results --------------------
  combined_results <- bind_rows(main_result, sex_age_race_results) %>%
    mutate(
      sex_group = factor(sex_group,
                        levels = c("main meta-analysis", "male", "female")),
      race_group = ifelse(is.na(race_group), "All", race_group)
    ) %>%
    arrange(sex_group, race_group, age_band)

  # Create complete grid to ensure all combinations are represented
  all_sex_groups <- c("main meta-analysis", "male", "female")
  all_race_groups <- c("All", "white", "mixed", "black")
  all_age_bands <- c("Overall", "Meta-Regression", sort(unique(combined_results$age_band[
    !combined_results$age_band %in% c("Overall", "Meta-Regression")
  ])))

  complete_grid <- expand.grid(
    sex_group = all_sex_groups,
    race_group = all_race_groups,
    age_band = all_age_bands,
    stringsAsFactors = FALSE
  ) %>%
    as_tibble()

  combined_results <- complete_grid %>%
    left_join(combined_results, by = c("sex_group", "race_group", "age_band")) %>%
    mutate(
      sex_group = factor(sex_group, levels = c("main meta-analysis", "male", "female"))
    ) %>%
    arrange(sex_group, race_group, age_band)

  # --- 6. Create plots for each sex group (All races) --------------------
  sex_groups <- c("main meta-analysis", "male", "female")

  output_list <- purrr::map(sex_groups, function(sex) {

    # Filter data for this sex group (All races only)
    sex_data <- combined_results %>%
      filter(sex_group == sex, race_group == "All")

    # Create forest plot
    colour <- case_when(
      sex == "main meta-analysis" ~ "springgreen3",
      sex == "male" ~ "skyblue2",
      sex == "female" ~ "indianred1"
    )

    plot <- sex_data %>%
      mutate(age_band = fct_rev(age_band)) %>%
      ggplot(aes(y = age_band)) +
      geom_errorbarh(aes(xmin = lower_95, xmax = upper_95),
                     height = 0.25, colour = colour) +
      geom_point(aes(x = pooled_estimate), size = 3, colour = colour) +
      labs(
        title = paste0(str_to_title(sex)),
        x = "Estimated 95% CI",
        y = "Age Band"
      ) +
      theme_minimal() +
      theme(
        plot.title = element_text(size = 12, face = "bold"),
        axis.title = element_text(size = 10)
      )

    list(
      plot = plot,
      data = sex_data
    )
  })

  names(output_list) <- sex_groups

  # --- 7. Create race-specific plots for each sex --------------------
  # (Removed - now handled in section 11)

  # --- 8. Wald tests for comparisons --------------------

  # Helper: Wald test returning mean diff, 95% CI, % shift, and p-value
  wald_test <- function(mean1, mean2, sd1, sd2, ref_for_pct = NULL) {
    if (any(is.na(c(mean1, mean2, sd1, sd2)))) {
      return(list(diff = NA_real_, lower = NA_real_, upper = NA_real_,
                  pct_shift = NA_real_, p = NA_real_))
    }
    diff   <- mean1 - mean2
    se_dif <- sqrt(sd1^2 + sd2^2)
    lower  <- diff - 1.96 * se_dif
    upper  <- diff + 1.96 * se_dif
    z      <- diff / se_dif
    p      <- 2 * (1 - pnorm(abs(z)))
    ref    <- if (is.null(ref_for_pct)) mean2 else ref_for_pct
    pct    <- if (!is.na(ref) && ref != 0) 100 * diff / ref else NA_real_
    list(diff = diff, lower = lower, upper = upper, pct_shift = pct, p = p)
  }

  # Sex comparisons (Overall estimates, All races)
  male_overall <- combined_results %>%
    filter(sex_group == "male", as.character(age_band) == "Overall", race_group == "All")
  female_overall <- combined_results %>%
    filter(sex_group == "female", as.character(age_band) == "Overall", race_group == "All")

  sex_w <- wald_test(male_overall$pooled_estimate,   female_overall$pooled_estimate,
                     male_overall$pooled_sd,         female_overall$pooled_sd)
  sex_comparison <- tibble(
    Comparison        = "Male vs Female (Overall)",
    `Mean Difference` = sex_w$diff,
    `95% CI Lower`    = sex_w$lower,
    `95% CI Upper`    = sex_w$upper,
    `% Shift`         = sex_w$pct_shift,
    `P (Wald Test)`   = sex_w$p
  )

  # race comparisons within each sex (Overall age band)
  race_comparisons <- purrr::map_dfr(c("male", "female"), function(sex) {
    race_data <- combined_results %>%
      filter(sex_group == sex, as.character(age_band) == "Overall",
             race_group != "All") %>%
      arrange(race_group)

    if (nrow(race_data) < 2) return(tibble())

    races <- unique(as.character(race_data$race_group))
    comparisons <- combn(races, 2, simplify = FALSE)

    purrr::map_dfr(comparisons, function(pair) {
      data1 <- race_data %>% filter(race_group == pair[1])
      data2 <- race_data %>% filter(race_group == pair[2])
      w <- wald_test(data1$pooled_estimate, data2$pooled_estimate,
                     data1$pooled_sd,       data2$pooled_sd)
      tibble(
        `Sex Group`        = sex,
        Comparison         = paste0(pair[1], " vs ", pair[2]),
        `Mean Difference`  = w$diff,
        `95% CI Lower`     = w$lower,
        `95% CI Upper`     = w$upper,
        `% Shift`          = w$pct_shift,
        `P (Wald Test)`    = w$p
      )
    })
  })

  # Age band comparisons within each sex group (All races)
  age_band_comparisons <- purrr::map_dfr(sex_groups, function(sex) {
    sex_data <- combined_results %>%
      filter(sex_group == sex, race_group == "All",
             as.character(age_band) != "Overall",
             as.character(age_band) != "Meta-Regression") %>%
      arrange(age_band)

    if (nrow(sex_data) < 2) return(tibble())

    age_bands <- unique(as.character(sex_data$age_band))
    comparisons <- combn(age_bands, 2, simplify = FALSE)

    purrr::map_dfr(comparisons, function(pair) {
      data1 <- sex_data %>% filter(as.character(age_band) == pair[1])
      data2 <- sex_data %>% filter(as.character(age_band) == pair[2])
      w <- wald_test(data1$pooled_estimate, data2$pooled_estimate,
                     data1$pooled_sd,       data2$pooled_sd)
      tibble(
        `Sex Group`        = sex,
        Comparison         = paste0(pair[1], " vs ", pair[2]),
        `Mean Difference`  = w$diff,
        `95% CI Lower`     = w$lower,
        `95% CI Upper`     = w$upper,
        `% Shift`          = w$pct_shift,
        `P (Wald Test)`    = w$p
      )
    })
  })

  # Sex comparison within each age band (All races)
  age_band_sex_comparisons <- purrr::map_dfr(age_bands_main, function(age) {
    male_data <- combined_results %>%
      filter(sex_group == "male", race_group == "All",
             as.character(age_band) == as.character(age))
    female_data <- combined_results %>%
      filter(sex_group == "female", race_group == "All",
             as.character(age_band) == as.character(age))

    if (nrow(male_data) == 0 || nrow(female_data) == 0) return(tibble())
    w <- wald_test(male_data$pooled_estimate, female_data$pooled_estimate,
                   male_data$pooled_sd,       female_data$pooled_sd)
    tibble(
      `Age Band`         = as.character(age),
      `Mean Difference`  = w$diff,
      `95% CI Lower`     = w$lower,
      `95% CI Upper`     = w$upper,
      `% Shift`          = w$pct_shift,
      `P (Wald Test)`    = w$p
    )
  })

  # Format all comparison tables — collapse diff + CI into one column,
  # show % shift (relative to the reference group), and the p-value.
  format_wald_table <- function(df) {
    if (nrow(df) == 0) return(NULL)
    n_rows <- nrow(df)
    df %>%
      mutate(
        `Mean Diff (95% CI)` = ifelse(
          is.na(`Mean Difference`), "—",
          sprintf("%.2f [%.2f, %.2f]", `Mean Difference`, `95% CI Lower`, `95% CI Upper`)
        ),
        `% Shift` = ifelse(is.na(`% Shift`), "—",
                           sprintf("%+.1f%%", `% Shift`)),
        `P (Wald Test)` = case_when(
          is.na(`P (Wald Test)`) ~ "—",
          `P (Wald Test)` < 0.001 ~ "<0.001",
          `P (Wald Test)` < 0.01  ~ "<0.01",
          `P (Wald Test)` < 0.05  ~ "<0.05",
          TRUE ~ sprintf("%.3f", `P (Wald Test)`)
        )
      ) %>%
      select(-`Mean Difference`, -`95% CI Lower`, -`95% CI Upper`) %>%
      gt() %>%
      cols_align(align = "center", columns = everything()) %>%
      tab_style(
        style = cell_text(weight = "bold"),
        locations = cells_column_labels()
      ) %>%
      tab_style(
        style = cell_fill(color = "#f7f7f7"),
        locations = cells_body(rows = seq(1, n_rows, by = 2))
      ) %>%
      tab_options(
        table.font.size = px(11),
        column_labels.font.weight = "bold",
        data_row.padding = px(3)
      ) %>%
      tab_source_note(
        md("*Mean difference = Group 1 − Group 2 in component units (mmol/24h, L/24h, or pH); % shift = difference relative to Group 2.*")
      )
  }

  sex_comparison_gt <- sex_comparison %>%
    format_wald_table() %>%
    tab_header(title = md("**Sex Comparison (Overall)**"))

  race_comparisons_gt <- if (nrow(race_comparisons) > 0) {
    race_comparisons %>%
      format_wald_table() %>%
      tab_header(title = md("**race Comparisons Within Sex Groups**"))
  } else {
    NULL
  }

  age_band_comparisons_gt <- if (nrow(age_band_comparisons) > 0) {
    age_band_comparisons %>%
      format_wald_table() %>%
      tab_header(title = md("**Age Band Comparisons Within Sex Groups**"))
  } else {
    NULL
  }

  age_band_sex_comparisons_gt <- if (nrow(age_band_sex_comparisons) > 0) {
    age_band_sex_comparisons %>%
      format_wald_table() %>%
      tab_header(title = md("**Sex Comparisons Within Age Bands**"))
  } else {
    NULL
  }

  # --- 9. Create combined summary table --------------------
  # Ensure descriptive columns exist (meta-regression rows may not produce them)
  for (col in c("study_mean_avg", "study_mean_sd", "study_mean_min", "study_mean_max")) {
    if (!col %in% names(combined_results)) combined_results[[col]] <- NA_real_
  }
  fmt_desc <- function(avg, sdv, lo, hi) {
    if (is.na(avg) && is.na(lo) && is.na(hi)) return("—")
    avg_part <- if (!is.na(avg) && !is.na(sdv)) sprintf("%.2f ± %.2f", avg, sdv) else "—"
    rng_part <- if (!is.na(lo) && !is.na(hi))   sprintf("%.2f–%.2f",  lo, hi)   else "—"
    paste0(avg_part, " (", rng_part, ")")
  }

  combined_summary_table <- combined_results %>%
    select(-urine_component) %>%
    mutate(
      `Sex Group` = as.character(sex_group),
      `race` = race_group,
      `Age Band` = as.character(age_band),
      `Pooled` = ifelse(is.na(pooled_estimate), "—", sprintf("%.2f", pooled_estimate)),
      `95% Range` = sprintf("%.2f – %.2f", lower_95, upper_95),
      `Studies Mean SD Range` = mapply(fmt_desc,
                                           study_mean_avg, study_mean_sd,
                                           study_mean_min, study_mean_max),
      Studies = n_studies,
      "Study Type" = study_type,
      Participants = total_participants,
      I2 = I2 * 100,
      `I²` = ifelse(is.na(I2), "—", paste0(round(I2, 1), "%")),
      `Cochran's Q` = ifelse(is.na(Q), "—", round(Q, 1)),
      `Q (p-value)` = case_when(
        is.na(Q_pval) ~ "—",
        Q_pval < 0.005 ~ "<0.005",
        Q_pval < 0.01  ~ "<0.01",
        Q_pval < 0.05  ~ "<0.05",
        TRUE ~ sprintf("%.2f", Q_pval)
      ),
      `Egger's p` = case_when(
        is.na(eggers_p) ~ "—",
        eggers_p < 0.001 ~ "<0.001",
        eggers_p < 0.01  ~ "<0.01",
        eggers_p < 0.05  ~ "<0.05",
        TRUE ~ sprintf("%.2f", eggers_p)
      ),
      "Model" = model_utilised
    ) %>%
    select(
      `Sex Group`,
      `race`,
      `Age Band`,
      `Pooled`,
      `95% Range`,
      `Studies Mean SD Range`,
      Studies,
      "Study Type",
      Participants,
      `I²`,
      `Cochran's Q`,
      `Q (p-value)`,
      `Egger's p`,
      Model
    )

  combined_summary_gt <- combined_summary_table %>%
    gt() %>%
    tab_header(
      title = md("**Combined Meta-Analysis Results**"),
      subtitle = paste0("24-Hour Urine ", urine_component_for_title)
    ) %>%
    cols_align(align = "center") %>%
    tab_style(
      style = cell_text(weight = "bold"),
      locations = cells_column_labels()
    ) %>%
    # Highlight by sex group
    tab_style(
      style = cell_fill(color = "springgreen3"),
      locations = cells_body(rows = `Sex Group` == "main meta-analysis")
    ) %>%
    tab_style(
      style = cell_fill(color = "skyblue2"),
      locations = cells_body(rows = `Sex Group` == "male")
    ) %>%
    tab_style(
      style = cell_fill(color = "indianred1"),
      locations = cells_body(rows = `Sex Group` == "female")
    ) %>%
    # Bold the Overall rows
    tab_style(
      style = cell_text(weight = "bold"),
      locations = cells_body(
        columns = everything(),
        rows = `Age Band` == "Overall"
      )
    ) %>%
    # Bold and highlight Meta-Regression rows
    tab_style(
      style = cell_text(weight = "bold"),
      locations = cells_body(
        columns = everything(),
        rows = `Age Band` == "Meta-Regression"
      )
    ) %>%
    tab_options(
      table.font.size = px(11),
      heading.title.font.size = px(14),
      heading.subtitle.font.size = px(12),
      column_labels.font.weight = "bold",
      data_row.padding = px(3)
    ) %>%
    tab_source_note(
      source_note = md("*I² represents heterogeneity; Q test assesses between-study variation; Egger's p = linear regression test for funnel plot asymmetry; Meta-Regression shows covariate-adjusted estimates*")
    )

  # --- 10. Create overall comparison plot (Age-stratified) --------------------

  design <- "ABC
             EEE"

  # Row 1: Three forest plots
  row1 <- (
      output_list[["main meta-analysis"]]$plot |
        output_list[["male"]]$plot |
        output_list[["female"]]$plot
    )

  # Row 2: Combined summary table
  row2 <- wrap_elements(full = combined_summary_gt)

  # Combine rows
  final_layout <- row1 + row2 +
    plot_layout(design = design,
                axis_titles = "collect",
                axes = "collect") +
    plot_annotation(
      title = urine_component_for_title,
      tag_levels = "A",
      theme = theme(plot.title = element_text(face = "bold", size = 14))
    )

  # --- 11. Create race-stratified plot and table --------------------

  # Filter for white, mixed, and black races (including All)
  race_results <- combined_results %>%
    filter(race_group %in% c("All", "white", "mixed", "black"))

  # Create race-only summary table (separate from combined table)
  race_only_summary_table <- race_results %>%
    filter(race_group != "All") %>%  # Exclude "All" for race-specific table
    select(-urine_component) %>%
    mutate(
      `Sex Group` = as.character(sex_group),
      `race` = str_to_title(race_group),
      `Age Band` = as.character(age_band),
      `95% Range` = ifelse(
        is.na(lower_95) | is.na(upper_95),
        "—",
        sprintf("%.2f – %.2f", lower_95, upper_95)
      ),
      Studies = ifelse(is.na(n_studies), "—", as.character(n_studies)),
      "Study Type" = study_type,
      Participants = ifelse(is.na(total_participants), "—", as.character(total_participants)),
      I2_val = I2 * 100,
      `I²` = ifelse(is.na(I2_val), "—", paste0(round(I2_val, 1), "%")),
      `Cochran's Q` = ifelse(is.na(Q), "—", as.character(round(Q, 1))),
      `Q (p-value)` = case_when(
        is.na(Q_pval) ~ "—",
        Q_pval < 0.005 ~ "<0.005",
        Q_pval < 0.01  ~ "<0.01",
        Q_pval < 0.05  ~ "<0.05",
        TRUE ~ sprintf("%.2f", Q_pval)
      ),
      `Egger's p` = case_when(
        is.na(eggers_p) ~ "—",
        eggers_p < 0.001 ~ "<0.001",
        eggers_p < 0.01  ~ "<0.01",
        eggers_p < 0.05  ~ "<0.05",
        TRUE ~ sprintf("%.2f", eggers_p)
      ),
      "Model" = ifelse(is.na(model_utilised), "—", model_utilised)
    ) %>%
    select(
      `Sex Group`,
      `race`,
      `Age Band`,
      `95% Range`,
      Studies,
      "Study Type",
      Participants,
      `I²`,
      `Cochran's Q`,
      `Q (p-value)`,
      `Egger's p`,
      Model
    )

  race_only_summary_gt <- race_only_summary_table %>%
    gt() %>%
    tab_header(
      title = md("**race-Specific Meta-Analysis Results**"),
      subtitle = paste0("24-Hour Urine ", urine_component_for_title)
    ) %>%
    cols_align(align = "center") %>%
    tab_style(
      style = cell_text(weight = "bold"),
      locations = cells_column_labels()
    ) %>%
    # Highlight by sex group
    tab_style(
      style = cell_fill(color = "springgreen3"),
      locations = cells_body(rows = `Sex Group` == "main meta-analysis")
    ) %>%
    tab_style(
      style = cell_fill(color = "skyblue2"),
      locations = cells_body(rows = `Sex Group` == "male")
    ) %>%
    tab_style(
      style = cell_fill(color = "indianred1"),
      locations = cells_body(rows = `Sex Group` == "female")
    ) %>%
    # Bold the Overall rows
    tab_style(
      style = cell_text(weight = "bold"),
      locations = cells_body(
        columns = everything(),
        rows = `Age Band` == "Overall"
      )
    ) %>%
    tab_options(
      table.font.size = px(11),
      heading.title.font.size = px(14),
      heading.subtitle.font.size = px(12),
      column_labels.font.weight = "bold",
      data_row.padding = px(3)
    ) %>%
    tab_source_note(
      source_note = md("*I² represents heterogeneity; Q test assesses between-study variation; Egger's p = linear regression test for funnel plot asymmetry; — indicates insufficient data*")
    )

  # Create forest plots by sex for race stratification
  race_output_list <- purrr::map(sex_groups, function(sex) {

    # Filter data for this sex group
    sex_race_data <- race_results %>%
      filter(sex_group == sex) %>%
      mutate(
        # Create a combined label for plotting
        plot_label = case_when(
          race_group == "All" & age_band == "Overall" ~ "Overall (All races)",
          race_group == "All" & age_band == "Meta-Regression" ~ "Meta-Regression (All races)",
          race_group == "All" ~ paste0(age_band, " (All races)"),
          age_band == "Overall" ~ str_to_title(race_group),
          age_band == "Meta-Regression" ~ paste0(str_to_title(race_group), " - Meta-Regression"),
          TRUE ~ paste0(str_to_title(race_group), " - ", age_band)
        ),
        # Order for plotting
        plot_order = case_when(
          race_group == "All" & age_band == "Overall" ~ 1,
          race_group == "All" & age_band == "Meta-Regression" ~ 2,
          race_group == "white" & age_band == "Overall" ~ 3,
          race_group == "mixed" & age_band == "Overall" ~ 4,
          race_group == "black" & age_band == "Overall" ~ 5,
          TRUE ~ 6 + row_number()
        )
      ) %>%
      arrange(plot_order)

    if (nrow(sex_race_data) == 0) {
      return(NULL)
    }

    # Create forest plot
    colour <- case_when(
      sex == "main meta-analysis" ~ "springgreen3",
      sex == "male" ~ "skyblue2",
      sex == "female" ~ "indianred1"
    )

    # Only plot rows with non-NA estimates
    sex_race_data_plot <- sex_race_data %>%
      filter(!is.na(pooled_estimate))

    plot <- sex_race_data_plot %>%
      mutate(plot_label = fct_reorder(plot_label, -plot_order)) %>%
      ggplot(aes(y = plot_label)) +
      geom_errorbarh(aes(xmin = lower_95, xmax = upper_95),
                     height = 0.25, colour = colour) +
      geom_point(aes(x = pooled_estimate), size = 3, colour = colour) +
      labs(
        title = paste0(str_to_title(sex)),
        x = "Estimated 95% CI",
        y = NULL
      ) +
      theme_minimal() +
      theme(
        plot.title = element_text(size = 12, face = "bold"),
        axis.title = element_text(size = 10),
        axis.text.y = element_text(size = 9)
      )

    list(
      plot = plot,
      data = sex_race_data
    )
  })

  names(race_output_list) <- sex_groups

  # Create race summary table (with All races for the plot)
  race_summary_table <- race_results %>%
    select(-urine_component) %>%
    mutate(
      `Sex Group` = as.character(sex_group),
      `race` = case_when(
        race_group == "All" ~ "All races",
        TRUE ~ str_to_title(race_group)
      ),
      `Age Band` = as.character(age_band),
      `95% Range` = ifelse(
        is.na(lower_95) | is.na(upper_95),
        "—",
        sprintf("%.2f – %.2f", lower_95, upper_95)
      ),
      Studies = ifelse(is.na(n_studies), "—", as.character(n_studies)),
      "Study Type" = study_type,
      Participants = ifelse(is.na(total_participants), "—", as.character(total_participants)),
      I2_val = I2 * 100,
      `I²` = ifelse(is.na(I2_val), "—", paste0(round(I2_val, 1), "%")),
      `Cochran's Q` = ifelse(is.na(Q), "—", as.character(round(Q, 1))),
      `Q (p-value)` = case_when(
        is.na(Q_pval) ~ "—",
        Q_pval < 0.005 ~ "<0.005",
        Q_pval < 0.01  ~ "<0.01",
        Q_pval < 0.05  ~ "<0.05",
        TRUE ~ sprintf("%.2f", Q_pval)
      ),
      `Egger's p` = case_when(
        is.na(eggers_p) ~ "—",
        eggers_p < 0.001 ~ "<0.001",
        eggers_p < 0.01  ~ "<0.01",
        eggers_p < 0.05  ~ "<0.05",
        TRUE ~ sprintf("%.2f", eggers_p)
      ),
      "Model" = ifelse(is.na(model_utilised), "—", model_utilised)
    ) %>%
    select(
      `Sex Group`,
      `race`,
      `Age Band`,
      `95% Range`,
      Studies,
      "Study Type",
      Participants,
      `I²`,
      `Cochran's Q`,
      `Q (p-value)`,
      `Egger's p`,
      Model
    )

  race_summary_gt <- race_summary_table %>%
    gt() %>%
    tab_header(
      title = md("**race-Stratified Meta-Analysis Results**"),
      subtitle = paste0("24-Hour Urine ", urine_component_for_title)
    ) %>%
    cols_align(align = "center") %>%
    tab_style(
      style = cell_text(weight = "bold"),
      locations = cells_column_labels()
    ) %>%
    # Highlight by sex group
    tab_style(
      style = cell_fill(color = "springgreen3"),
      locations = cells_body(rows = `Sex Group` == "main meta-analysis")
    ) %>%
    tab_style(
      style = cell_fill(color = "skyblue2"),
      locations = cells_body(rows = `Sex Group` == "male")
    ) %>%
    tab_style(
      style = cell_fill(color = "indianred1"),
      locations = cells_body(rows = `Sex Group` == "female")
    ) %>%
    # Bold the Overall rows
    tab_style(
      style = cell_text(weight = "bold"),
      locations = cells_body(
        columns = everything(),
        rows = `Age Band` == "Overall"
      )
    ) %>%
    # Bold the "All races" rows
    tab_style(
      style = cell_text(weight = "bold"),
      locations = cells_body(
        columns = everything(),
        rows = `race` == "All races"
      )
    ) %>%
    tab_options(
      table.font.size = px(11),
      heading.title.font.size = px(14),
      heading.subtitle.font.size = px(12),
      column_labels.font.weight = "bold",
      data_row.padding = px(3)
    ) %>%
    tab_source_note(
      source_note = md("*I² represents heterogeneity; Q test assesses between-study variation; Egger's p = linear regression test for funnel plot asymmetry; — indicates insufficient data*")
    )

  # Create race comparison plot layout
  race_row1 <- (
      race_output_list[["main meta-analysis"]]$plot |
        race_output_list[["male"]]$plot |
        race_output_list[["female"]]$plot
    )

  race_row2 <- wrap_elements(full = race_summary_gt)

  race_layout <- race_row1 + race_row2 +
    plot_layout(design = design,
                axis_titles = "collect",
                axes = "collect") +
    plot_annotation(
      title = paste0(urine_component_for_title, " - race Stratification"),
      tag_levels = "A",
      theme = theme(plot.title = element_text(face = "bold", size = 14))
    )

  # --- 12. Return results --------------------
  return(list(
    ma_results = combined_results,
    main_meta_analysis = output_list[["main meta-analysis"]],
    male = output_list[["male"]],
    female = output_list[["female"]],
    combined_plot = final_layout,
    combined_summary_table = combined_summary_gt,
    race_plot = race_layout,
    race_summary_table = race_summary_gt,
    race_only_table = race_only_summary_gt,
    wald_tests = list(
      sex_comparison = sex_comparison_gt,
      race_comparisons = race_comparisons_gt,
      age_band_comparisons = age_band_comparisons_gt,
      sex_within_age_bands = age_band_sex_comparisons_gt
    )
  ))
}
```

### 6 Meta-analysis by Urine Component

#### 6.1 Volume

##### 6.1.1 Comparison by Age Band

###### 6.1.1.1 Plot

```
volume_ma_2nd_analysis <- sex_age_subdivision_ma("volume")
## Not enough studies for multivariable meta-regression (k = 3, need > 6). Running univariable models instead.
## Meta-regression failed for male: In argument: `pooled_sd = case_when(...)`.
volume_ma_2nd_analysis$combined_plot
## `height` was translated to `width`.
## `height` was translated to `width`.
## `height` was translated to `width`.
```

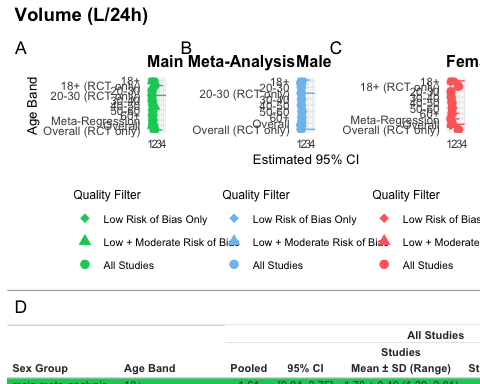

###### 6.1.1.2 Summary Table

volume_ma_2nd_analysis$combined_summary_table

Table 1: **Combined Meta-Analysis Results**

24-Hour Urine Volume (L/24h)

|  |  | **All Studies** | | | | | | **Low Risk of Bias Only** | | | | | | **Low + Moderate RoB** | | | | | |
| --- | --- | --- | --- | --- | --- | --- | --- | --- | --- | --- | --- | --- | --- | --- | --- | --- | --- | --- | --- |
| Sex Group | Age Band | Pooled | 95% CI | Studies  Mean ± SD (Range) | Studies, n | Q p-value | Egger's p | Pooled | 95% CI | Studies  Mean ± SD (Range) | Studies, n | Q p-value | Egger's p | Pooled | 95% CI | Studies  Mean ± SD (Range) | Studies, n | Q p-value | Egger's p |
| main meta-analysis | 18+ | 1.61 | [0.94, 2.75] | 1.78 ± 0.49 (1.20–2.81) | 59 | <0.001 | 0.50 | 1.55 | [0.89, 2.69] | 1.72 ± 0.51 (1.30–2.70) | 27 | <0.001 | 0.16 | 1.62 | [0.91, 2.87] | 1.79 ± 0.52 (1.22–2.81) | 34 | <0.001 | <0.05 |
| **main meta-analysis** | **18+ (RCT only)** | **2.01** | **[1.08, 3.74]** | **2.18 ± 0.55 (1.25–2.81)** | **7** | **<0.001** | **0.08** | **—** | **—** | **—** | **—** | **—** | **—** | **—** | **—** | **—** | **—** | **—** | **—** |
| main meta-analysis | 20-30 | 1.42 | [1.11, 1.81] | 1.55 ± 0.33 (1.28–2.07) | 5 | <0.001 | 0.52 | — | — | — | — | — | — | — | — | — | — | — | — |
| **main meta-analysis** | **20-30 (RCT only)** | **1.40** | **[0.44, 4.43]** | **— (1.66–1.66)** | **1** | **—** | **—** | **—** | **—** | **—** | **—** | **—** | **—** | **—** | **—** | **—** | **—** | **—** | **—** |
| main meta-analysis | 30-40 | 1.43 | [1.30, 1.56] | 1.45 ± 0.07 (1.35–1.51) | 5 | <0.001 | 0.33 | — | — | — | — | — | — | — | — | — | — | — | — |
| main meta-analysis | 40-50 | 1.57 | [1.15, 2.15] | 1.67 ± 0.29 (1.26–2.02) | 7 | <0.001 | 0.85 | — | — | — | — | — | — | 1.86 | [1.59, 2.18] | — (1.87–1.87) | 1 | — | — |
| main meta-analysis | 50-60 | 1.58 | [1.12, 2.23] | 1.68 ± 0.34 (1.23–2.11) | 5 | <0.001 | 0.45 | — | — | — | — | — | — | — | — | — | — | — | — |
| main meta-analysis | 60+ | 1.53 | [0.95, 2.48] | 1.66 ± 0.37 (0.89–2.08) | 8 | <0.001 | <0.05 | — | — | — | — | — | — | — | — | — | — | — | — |
| **main meta-analysis** | **Meta-Regression** | **1.57** | **[0.95, 2.60]** | **—** | **52** | **<0.001** | **0.98** | **—** | **—** | **—** | **—** | **—** | **—** | **—** | **—** | **—** | **—** | **—** | **—** |
| **main meta-analysis** | **Overall** | **1.57** | **[0.98, 2.53]** | **1.73 ± 0.44 (0.89–2.81)** | **89** | **<0.001** | **0.81** | **1.55** | **[0.89, 2.69]** | **1.72 ± 0.51 (1.30–2.70)** | **27** | **<0.001** | **0.16** | **1.63** | **[0.92, 2.87]** | **1.80 ± 0.51 (1.22–2.81)** | **35** | **<0.001** | **<0.05** |
| **main meta-analysis** | **Overall (RCT only)** | **1.92** | **[1.02, 3.60]** | **2.12 ± 0.54 (1.25–2.81)** | **8** | **<0.001** | **<0.05** | **—** | **—** | **—** | **—** | **—** | **—** | **—** | **—** | **—** | **—** | **—** | **—** |
| male | 18+ | 1.62 | [0.77, 3.43] | 1.76 ± 0.73 (1.34–2.60) | 3 | <0.001 | 0.31 | 1.27 | [0.67, 2.41] | — (1.34–1.34) | 1 | — | — | 1.27 | [0.67, 2.41] | — (1.34–1.34) | 1 | — | — |
| **male** | **18+ (RCT only)** | **—** | **—** | **—** | **—** | **—** | **—** | **—** | **—** | **—** | **—** | **—** | **—** | **—** | **—** | **—** | **—** | **—** | **—** |
| male | 20-30 | 1.42 | [1.16, 1.74] | 1.54 ± 0.17 (1.42–1.66) | 2 | 0.85 | — | — | — | — | — | — | — | — | — | — | — | — | — |
| **male** | **20-30 (RCT only)** | **1.40** | **[0.44, 4.43]** | **— (1.66–1.66)** | **1** | **—** | **—** | **—** | **—** | **—** | **—** | **—** | **—** | **—** | **—** | **—** | **—** | **—** | **—** |
| male | 30-40 | 1.48 | [1.33, 1.65] | 1.50 ± 0.02 (1.48–1.51) | 3 | 0.63 | 0.92 | — | — | — | — | — | — | — | — | — | — | — | — |
| male | 40-50 | 1.43 | [1.23, 1.67] | 1.70 ± 0.37 (1.44–1.96) | 2 | 0.70 | — | — | — | — | — | — | — | — | — | — | — | — | — |
| male | 50-60 | 1.61 | [1.30, 2.00] | 1.69 ± 0.28 (1.49–1.89) | 2 | <0.001 | — | — | — | — | — | — | — | — | — | — | — | — | — |
| male | 60+ | 1.55 | [1.27, 1.87] | 1.63 ± 0.21 (1.39–1.76) | 3 | <0.001 | 0.59 | — | — | — | — | — | — | — | — | — | — | — | — |
| **male** | **Meta-Regression** | **—** | **—** | **—** | **—** | **—** | **—** | **—** | **—** | **—** | **—** | **—** | **—** | **—** | **—** | **—** | **—** | **—** | **—** |
| **male** | **Overall** | **1.53** | **[1.11, 2.09]** | **1.64 ± 0.33 (1.34–2.60)** | **15** | **<0.001** | **0.54** | **1.27** | **[0.67, 2.41]** | **— (1.34–1.34)** | **1** | **—** | **—** | **1.27** | **[0.67, 2.41]** | **— (1.34–1.34)** | **1** | **—** | **—** |
| **male** | **Overall (RCT only)** | **1.40** | **[0.44, 4.43]** | **— (1.66–1.66)** | **1** | **—** | **—** | **—** | **—** | **—** | **—** | **—** | **—** | **—** | **—** | **—** | **—** | **—** | **—** |
| female | 18+ | 1.54 | [0.90, 2.63] | 1.72 ± 0.49 (1.37–2.81) | 7 | <0.001 | 0.97 | 1.25 | [0.53, 2.94] | 1.37 ± 0.00 (1.37–1.37) | 2 | 1.00 | — | 1.63 | [0.66, 4.03] | 1.85 ± 0.83 (1.37–2.81) | 3 | <0.001 | <0.001 |
| **female** | **18+ (RCT only)** | **2.79** | **[2.15, 3.60]** | **— (2.81–2.81)** | **1** | **—** | **—** | **—** | **—** | **—** | **—** | **—** | **—** | **—** | **—** | **—** | **—** | **—** | **—** |
| female | 20-30 | 1.27 | [1.10, 1.47] | — (1.28–1.28) | 1 | — | — | — | — | — | — | — | — | — | — | — | — | — | — |
| **female** | **20-30 (RCT only)** | **—** | **—** | **—** | **—** | **—** | **—** | **—** | **—** | **—** | **—** | **—** | **—** | **—** | **—** | **—** | **—** | **—** | **—** |
| female | 30-40 | 1.35 | [1.15, 1.58] | — (1.35–1.35) | 1 | — | — | — | — | — | — | — | — | — | — | — | — | — | — |
| female | 40-50 | 1.49 | [1.00, 2.22] | 1.58 ± 0.39 (1.26–2.02) | 3 | <0.001 | 0.77 | — | — | — | — | — | — | — | — | — | — | — | — |
| female | 50-60 | 1.55 | [0.98, 2.45] | 1.68 ± 0.44 (1.23–2.11) | 3 | <0.001 | 0.61 | — | — | — | — | — | — | — | — | — | — | — | — |
| female | 60+ | 1.52 | [0.82, 2.83] | 1.68 ± 0.46 (0.89–2.08) | 5 | <0.001 | 0.16 | — | — | — | — | — | — | — | — | — | — | — | — |
| **female** | **Meta-Regression** | **1.25** | **[0.55, 2.82]** | **—** | **7** | **1.00** | **0.97** | **—** | **—** | **—** | **—** | **—** | **—** | **—** | **—** | **—** | **—** | **—** | **—** |
| **female** | **Overall** | **1.50** | **[0.94, 2.40]** | **1.64 ± 0.42 (0.89–2.81)** | **20** | **<0.001** | **0.94** | **1.25** | **[0.53, 2.94]** | **1.37 ± 0.00 (1.37–1.37)** | **2** | **1.00** | **—** | **1.63** | **[0.66, 4.03]** | **1.85 ± 0.83 (1.37–2.81)** | **3** | **<0.001** | **<0.001** |
| **female** | **Overall (RCT only)** | **2.79** | **[2.15, 3.60]** | **— (2.81–2.81)** | **1** | **—** | **—** | **—** | **—** | **—** | **—** | **—** | **—** | **—** | **—** | **—** | **—** | **—** | **—** |
| *Studies, n = number of studies; Q p-value = Cochran's Q heterogeneity test; Egger's p = linear regression test for funnel plot asymmetry; RoB = Risk of Bias* | | | | | | | | | | | | | | | | | | | |

###### 6.1.1.3 Wald Tests

volume_ma_2nd_analysis$wald_tests$sex_comparison

Table 1: **Sex Comparison (Overall)**

| Comparison | % Shift | P (Wald Test) | Mean Diff (95% CI) |
| --- | --- | --- | --- |
| Male vs Female (Overall) | +1.5% | 0.937 | 0.02 [-0.54, 0.59] |
| *Mean difference = Group 1 − Group 2 in component units (mmol/24h, L/24h, or pH); % shift = difference relative to Group 2.* | | | |

volume_ma_2nd_analysis$wald_tests$age_band_comparisons

Table 1: **Age Band Comparisons Within Sex Groups**

| Sex Group | Comparison | % Shift | P (Wald Test) | Mean Diff (95% CI) |
| --- | --- | --- | --- | --- |
| main meta-analysis | 18+ vs 20-30 | +13.3% | 0.530 | 0.19 [-0.40, 0.78] |
| main meta-analysis | 18+ vs 30-40 | +12.5% | 0.520 | 0.18 [-0.37, 0.72] |
| main meta-analysis | 18+ vs 40-50 | +2.1% | 0.915 | 0.03 [-0.59, 0.65] |
| main meta-analysis | 18+ vs 50-60 | +1.8% | 0.929 | 0.03 [-0.61, 0.67] |
| main meta-analysis | 18+ vs 60+ | +4.8% | 0.840 | 0.07 [-0.65, 0.79] |
| main meta-analysis | 20-30 vs 30-40 | -0.7% | 0.939 | -0.01 [-0.27, 0.25] |
| main meta-analysis | 20-30 vs 40-50 | -9.9% | 0.442 | -0.15 [-0.55, 0.24] |
| main meta-analysis | 20-30 vs 50-60 | -10.1% | 0.459 | -0.16 [-0.58, 0.26] |
| main meta-analysis | 20-30 vs 60+ | -7.5% | 0.676 | -0.11 [-0.65, 0.42] |
| main meta-analysis | 30-40 vs 40-50 | -9.2% | 0.381 | -0.14 [-0.47, 0.18] |
| main meta-analysis | 30-40 vs 50-60 | -9.5% | 0.411 | -0.15 [-0.51, 0.21] |
| main meta-analysis | 30-40 vs 60+ | -6.8% | 0.675 | -0.10 [-0.59, 0.38] |
| main meta-analysis | 40-50 vs 50-60 | -0.3% | 0.984 | -0.00 [-0.47, 0.46] |
| main meta-analysis | 40-50 vs 60+ | +2.6% | 0.891 | 0.04 [-0.53, 0.61] |
| main meta-analysis | 50-60 vs 60+ | +2.9% | 0.882 | 0.04 [-0.55, 0.64] |
| male | 18+ vs 20-30 | +14.1% | 0.614 | 0.20 [-0.58, 0.98] |
| male | 18+ vs 30-40 | +9.5% | 0.715 | 0.14 [-0.62, 0.90] |
| male | 18+ vs 40-50 | +13.2% | 0.629 | 0.19 [-0.58, 0.95] |
| male | 18+ vs 50-60 | +0.8% | 0.975 | 0.01 [-0.77, 0.79] |
| male | 18+ vs 60+ | +4.9% | 0.849 | 0.08 [-0.70, 0.85] |
| male | 20-30 vs 30-40 | -4.0% | 0.613 | -0.06 [-0.29, 0.17] |
| male | 20-30 vs 40-50 | -0.8% | 0.929 | -0.01 [-0.27, 0.24] |
| male | 20-30 vs 50-60 | -11.7% | 0.215 | -0.19 [-0.48, 0.11] |
| male | 20-30 vs 60+ | -8.1% | 0.382 | -0.12 [-0.41, 0.16] |
| male | 30-40 vs 40-50 | +3.3% | 0.614 | 0.05 [-0.14, 0.23] |
| male | 30-40 vs 50-60 | -8.0% | 0.296 | -0.13 [-0.37, 0.11] |
| male | 30-40 vs 60+ | -4.3% | 0.558 | -0.07 [-0.29, 0.15] |
| male | 40-50 vs 50-60 | -11.0% | 0.191 | -0.18 [-0.44, 0.09] |
| male | 40-50 vs 60+ | -7.3% | 0.365 | -0.11 [-0.36, 0.13] |
| male | 50-60 vs 60+ | +4.1% | 0.671 | 0.06 [-0.23, 0.35] |
| female | 18+ vs 20-30 | +20.6% | 0.355 | 0.26 [-0.29, 0.82] |
| female | 18+ vs 30-40 | +13.8% | 0.514 | 0.19 [-0.37, 0.75] |
| female | 18+ vs 40-50 | +3.2% | 0.889 | 0.05 [-0.62, 0.72] |
| female | 18+ vs 50-60 | -1.1% | 0.962 | -0.02 [-0.72, 0.69] |
| female | 18+ vs 60+ | +0.8% | 0.976 | 0.01 [-0.81, 0.83] |
| female | 20-30 vs 30-40 | -5.6% | 0.488 | -0.08 [-0.29, 0.14] |
| female | 20-30 vs 40-50 | -14.4% | 0.319 | -0.21 [-0.64, 0.21] |
| female | 20-30 vs 50-60 | -18.0% | 0.252 | -0.28 [-0.76, 0.20] |
| female | 20-30 vs 60+ | -16.4% | 0.440 | -0.25 [-0.88, 0.38] |
| female | 30-40 vs 40-50 | -9.3% | 0.525 | -0.14 [-0.57, 0.29] |
| female | 30-40 vs 50-60 | -13.1% | 0.409 | -0.20 [-0.69, 0.28] |
| female | 30-40 vs 60+ | -11.4% | 0.593 | -0.17 [-0.81, 0.46] |
| female | 40-50 vs 50-60 | -4.2% | 0.834 | -0.06 [-0.67, 0.54] |
| female | 40-50 vs 60+ | -2.3% | 0.926 | -0.03 [-0.77, 0.70] |
| female | 50-60 vs 60+ | +2.0% | 0.939 | 0.03 [-0.74, 0.80] |
| *Mean difference = Group 1 − Group 2 in component units (mmol/24h, L/24h, or pH); % shift = difference relative to Group 2.* | | | | |

volume_ma_2nd_analysis$wald_tests$sex_within_age_bands

Table 1: **Sex Comparisons Within Age Bands**

| Age Band | % Shift | P (Wald Test) | Mean Diff (95% CI) |
| --- | --- | --- | --- |
| 18+ | +5.5% | 0.858 | 0.08 [-0.84, 1.01] |
| 20-30 | +11.5% | 0.246 | 0.15 [-0.10, 0.40] |
| 30-40 | +9.6% | 0.185 | 0.13 [-0.06, 0.32] |
| 40-50 | -3.8% | 0.794 | -0.06 [-0.48, 0.37] |
| 50-60 | +3.5% | 0.832 | 0.05 [-0.45, 0.56] |
| 60+ | +1.4% | 0.948 | 0.02 [-0.63, 0.67] |
| *Mean difference = Group 1 − Group 2 in component units (mmol/24h, L/24h, or pH); % shift = difference relative to Group 2.* | | | |

###### 6.1.1.4 Meta-Regression (plain language)

format_meta_reg_plain("volume", sex_group = "all")

Message

No meta-regression result for volume / all

###### 6.1.1.5 Assay Subgroup Analysis

assay_subgroup_table("volume")

Table 1: **Assay subgroup analysis — Volume**

*Per-assay pooled estimates; categories with k < 3 listed but not pooled.*

| Assay category | Studies, n | Participants | 95% CI | I² | Q p-value | Egger's p | Note |
| --- | --- | --- | --- | --- | --- | --- | --- |
| Unreported | 89 | 9,144 | 1708.65 [1619.04, 1798.26] | 99% | <0.001 | <0.01 |  |

###### 6.1.1.6 Preservative Sensitivity (Litholink / acidified samples)

```
preservative_sensitivity_ma("volume")
## # A tibble: 3 × 6
##   group               n_studies n_participants pooled_estimate lower_95 upper_95
##   <chr>                   <int>          <int>           <dbl>    <dbl>    <dbl>
## 1 All studies                89           9144           1709.    1619.    1798.
## 2 Excluding preserva…        89           9144           1709.    1619.    1798.
## 3 Preservative only           0              0             NA       NA       NA
```

###### 6.1.1.7 Creatinine Completeness Sensitivity

creat_completeness_table("volume")

Table 1: **Creatinine-completeness sensitivity — Volume**

*Compares the all-studies pooled estimate against subsets defined by whether the study reported 24h urine creatinine. Studies that do not report creatinine cannot verify collection completeness; a substantial shift here flags possible bias from incomplete collections.*

| Subset | Studies, n | Participants | Range (95% CI) | Δ vs all-studies (%) | Flag |
| --- | --- | --- | --- | --- | --- |
| All studies | 89 | 9,144 | 1708.65 [1619.04, 1798.26] | — | — |
| Creatinine reported only | 65 | 8,071 | 1667.46 [1566.22, 1768.70] | -2.4% | OK |
| Creatinine NOT reported | 24 | 1,073 | 1823.45 [1638.79, 2008.12] | +6.7% | OK |

##### 6.1.2 Comparison by Age / Sex / race

###### 6.1.2.1 Plot

```
volume_ma_3rd_analysis <- sex_age_race_subdivision_ma("volume",
                                                      urine_ma2,
                                                      overall_bias)
## Not enough studies for multivariable meta-regression (k = 3, need > 6). Running univariable models instead.
## Meta-regression failed for male: In argument: `pooled_sd = case_when(...)`.
volume_ma_3rd_analysis$race_plot
## `height` was translated to `width`.
## `height` was translated to `width`.
## `height` was translated to `width`.
```

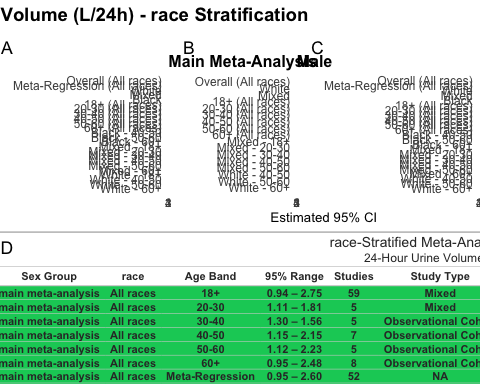

###### 6.1.2.2 Combined MA Results

volume_ma_3rd_analysis$combined_summary_table

Table 1: **Combined Meta-Analysis Results**

24-Hour Urine Volume (L/24h)

| Sex Group | race | Age Band | Pooled | 95% Range | Studies Mean SD Range | Studies | Study Type | Participants | I² | Cochran's Q | Q (p-value) | Egger's p | Model |
| --- | --- | --- | --- | --- | --- | --- | --- | --- | --- | --- | --- | --- | --- |
| main meta-analysis | All | 18+ | 1.61 | 0.94 – 2.75 | 1.78 ± 0.49 (1.20–2.81) | 59 | Mixed | 4647 | 98.5% | 3989.9 | <0.005 | 0.50 | Random |
| main meta-analysis | All | 20-30 | 1.42 | 1.11 – 1.81 | 1.55 ± 0.33 (1.28–2.07) | 5 | Mixed | 153 | 91.5% | 47.2 | <0.005 | 0.52 | Random |
| main meta-analysis | All | 30-40 | 1.43 | 1.30 – 1.56 | 1.45 ± 0.07 (1.35–1.51) | 5 | Observational Cohort | 185 | 90.7% | 42.9 | <0.005 | 0.33 | Random |
| main meta-analysis | All | 40-50 | 1.57 | 1.15 – 2.15 | 1.67 ± 0.29 (1.26–2.02) | 7 | Observational Cohort | 665 | 99.2% | 719.4 | <0.005 | 0.85 | Random |
| main meta-analysis | All | 50-60 | 1.58 | 1.12 – 2.23 | 1.68 ± 0.34 (1.23–2.11) | 5 | Observational Cohort | 2669 | 99.6% | 928.1 | <0.005 | 0.45 | Random |
| main meta-analysis | All | 60+ | 1.53 | 0.95 – 2.48 | 1.66 ± 0.37 (0.89–2.08) | 8 | Observational Cohort | 825 | 99.6% | 1683.3 | <0.005 | <0.05 | Random |
| **main meta-analysis** | **All** | **Meta-Regression** | **1.57** | **0.95 – 2.60** | **—** | **52** | **NA** | **4291** | **98.4%** | **1510.9** | **<0.005** | **0.98** | **Multivariable** |
| **main meta-analysis** | **All** | **Meta-Regression** | **1.57** | **0.95 – 2.60** | **—** | **52** | **NA** | **4291** | **98.4%** | **1510.9** | **<0.005** | **0.98** | **Multivariable** |
| **main meta-analysis** | **All** | **Overall** | **1.57** | **0.98 – 2.53** | **1.73 ± 0.44 (0.89–2.81)** | **89** | **Mixed** | **9144** | **99%** | **9158.8** | **<0.005** | **0.81** | **Random** |
| **main meta-analysis** | **All** | **Overall** | **1.57** | **0.98 – 2.53** | **1.73 ± 0.44 (0.89–2.81)** | **89** | **Mixed** | **9144** | **99%** | **9158.8** | **<0.005** | **0.81** | **Random** |
| main meta-analysis | black | 18+ | — | NA – NA | — | NA | NA | NA | — | — | — | — | NA |
| main meta-analysis | black | 20-30 | — | NA – NA | — | NA | NA | NA | — | — | — | — | NA |
| main meta-analysis | black | 30-40 | — | NA – NA | — | NA | NA | NA | — | — | — | — | NA |
| main meta-analysis | black | 40-50 | 1.40 | 0.77 – 2.55 | — (1.47–1.47) | 1 | Observational Cohort | 29 | — | — | — | — | Single study |
| main meta-analysis | black | 50-60 | 1.56 | 0.71 – 3.43 | — (1.69–1.69) | 1 | Observational Cohort | 131 | — | — | — | — | Single study |
| main meta-analysis | black | 60+ | 1.65 | 0.77 – 3.55 | 1.77 ± 0.10 (1.70–1.84) | 2 | Observational Cohort | 144 | 0.8% | 1 | 0.32 | — | Fixed |
| **main meta-analysis** | **black** | **Meta-Regression** | **—** | **NA – NA** | **—** | **NA** | **NA** | **NA** | **—** | **—** | **—** | **—** | **NA** |
| **main meta-analysis** | **black** | **Overall** | **1.58** | **0.74 – 3.34** | **1.68 ± 0.15 (1.47–1.84)** | **4** | **Observational Cohort** | **304** | **60%** | **7.5** | **0.06** | **0.55** | **Fixed** |
| main meta-analysis | mixed | 18+ | 1.62 | 0.95 – 2.76 | 1.79 ± 0.49 (1.20–2.81) | 58 | Mixed | 4484 | 98.5% | 3864 | <0.005 | 0.55 | Random |
| main meta-analysis | mixed | 20-30 | 1.42 | 1.11 – 1.81 | 1.55 ± 0.33 (1.28–2.07) | 5 | Mixed | 153 | 91.5% | 47.2 | <0.005 | 0.52 | Random |
| main meta-analysis | mixed | 30-40 | 1.43 | 1.30 – 1.56 | 1.45 ± 0.07 (1.35–1.51) | 5 | Observational Cohort | 185 | 90.7% | 42.9 | <0.005 | 0.33 | Random |
| main meta-analysis | mixed | 40-50 | 1.55 | 1.10 – 2.18 | 1.57 ± 0.27 (1.26–1.87) | 4 | Observational Cohort | 106 | 99.5% | 606.5 | <0.005 | 0.70 | Random |
| main meta-analysis | mixed | 50-60 | 1.35 | 1.04 – 1.77 | 1.36 ± 0.19 (1.23–1.49) | 2 | Observational Cohort | 50 | 98.9% | 92 | <0.005 | — | Random |
| main meta-analysis | mixed | 60+ | 0.88 | 0.79 – 0.99 | — (0.89–0.89) | 1 | Observational Cohort | 25 | — | — | — | — | Single study |
| **main meta-analysis** | **mixed** | **Meta-Regression** | **—** | **NA – NA** | **—** | **NA** | **NA** | **NA** | **—** | **—** | **—** | **—** | **NA** |
| **main meta-analysis** | **mixed** | **Overall** | **1.56** | **0.94 – 2.59** | **1.72 ± 0.47 (0.89–2.81)** | **75** | **Mixed** | **5003** | **99%** | **7607.8** | **<0.005** | **0.60** | **Random** |
| main meta-analysis | white | 18+ | 1.16 | 0.55 – 2.47 | — (1.25–1.25) | 1 | Observational Cohort | 163 | — | — | — | — | Single study |
| main meta-analysis | white | 20-30 | — | NA – NA | — | NA | NA | NA | — | — | — | — | NA |
| main meta-analysis | white | 30-40 | — | NA – NA | — | NA | NA | NA | — | — | — | — | NA |
| main meta-analysis | white | 40-50 | 1.86 | 0.84 – 4.12 | 1.99 ± 0.04 (1.96–2.02) | 2 | Observational Cohort | 530 | 0% | 0.4 | 0.55 | — | Fixed |
| main meta-analysis | white | 50-60 | 1.87 | 1.63 – 2.15 | 2.00 ± 0.16 (1.89–2.11) | 2 | Observational Cohort | 2488 | 83.8% | 6.2 | <0.05 | — | Random |
| main meta-analysis | white | 60+ | 1.67 | 1.29 – 2.15 | 1.77 ± 0.25 (1.39–2.08) | 5 | Observational Cohort | 656 | 97.1% | 136.4 | <0.005 | 0.96 | Random |
| **main meta-analysis** | **white** | **Meta-Regression** | **—** | **NA – NA** | **—** | **NA** | **NA** | **NA** | **—** | **—** | **—** | **—** | **NA** |
| **main meta-analysis** | **white** | **Overall** | **1.66** | **1.19 – 2.30** | **1.81 ± 0.29 (1.25–2.11)** | **10** | **Observational Cohort** | **3837** | **98.2%** | **505.1** | **<0.005** | **0.10** | **Random** |
| male | All | 18+ | 1.62 | 0.77 – 3.43 | 1.76 ± 0.73 (1.34–2.60) | 3 | Observational Cohort | 145 | 98.7% | 155.7 | <0.005 | 0.31 | Random |
| male | All | 20-30 | 1.42 | 1.16 – 1.74 | 1.54 ± 0.17 (1.42–1.66) | 2 | Mixed | 63 | 0% | 0 | 0.85 | — | Fixed |
| male | All | 30-40 | 1.48 | 1.33 – 1.65 | 1.50 ± 0.02 (1.48–1.51) | 3 | Observational Cohort | 143 | 0% | 0.9 | 0.63 | 0.92 | Fixed |
| male | All | 40-50 | 1.43 | 1.23 – 1.67 | 1.70 ± 0.37 (1.44–1.96) | 2 | Observational Cohort | 31 | 0% | 0.2 | 0.70 | — | Fixed |
| male | All | 50-60 | 1.61 | 1.30 – 2.00 | 1.69 ± 0.28 (1.49–1.89) | 2 | Observational Cohort | 104 | 92.1% | 12.6 | <0.005 | — | Random |
| male | All | 60+ | 1.55 | 1.27 – 1.87 | 1.63 ± 0.21 (1.39–1.76) | 3 | Observational Cohort | 329 | 93.9% | 32.9 | <0.005 | 0.59 | Random |
| **male** | **All** | **Meta-Regression** | **—** | **NA – NA** | **—** | **NA** | **NA** | **NA** | **—** | **—** | **—** | **—** | **NA** |
| **male** | **All** | **Overall** | **1.53** | **1.11 – 2.09** | **1.64 ± 0.33 (1.34–2.60)** | **15** | **Mixed** | **815** | **93.8%** | **225.8** | **<0.005** | **0.54** | **Random** |
| male | black | 18+ | — | NA – NA | — | NA | NA | NA | — | — | — | — | NA |
| male | black | 20-30 | — | NA – NA | — | NA | NA | NA | — | — | — | — | NA |
| male | black | 30-40 | — | NA – NA | — | NA | NA | NA | — | — | — | — | NA |
| male | black | 40-50 | — | NA – NA | — | NA | NA | NA | — | — | — | — | NA |
| male | black | 50-60 | — | NA – NA | — | NA | NA | NA | — | — | — | — | NA |
| male | black | 60+ | — | NA – NA | — | NA | NA | NA | — | — | — | — | NA |
| **male** | **black** | **Meta-Regression** | **—** | **NA – NA** | **—** | **NA** | **NA** | **NA** | **—** | **—** | **—** | **—** | **NA** |
| **male** | **black** | **Overall** | **—** | **NA – NA** | **—** | **NA** | **NA** | **NA** | **—** | **—** | **—** | **—** | **NA** |
| male | mixed | 18+ | 1.62 | 0.77 – 3.43 | 1.76 ± 0.73 (1.34–2.60) | 3 | Observational Cohort | 145 | 98.7% | 155.7 | <0.005 | 0.31 | Random |
| male | mixed | 20-30 | 1.42 | 1.16 – 1.74 | 1.54 ± 0.17 (1.42–1.66) | 2 | Mixed | 63 | 0% | 0 | 0.85 | — | Fixed |
| male | mixed | 30-40 | 1.48 | 1.33 – 1.65 | 1.50 ± 0.02 (1.48–1.51) | 3 | Observational Cohort | 143 | 0% | 0.9 | 0.63 | 0.92 | Fixed |
| male | mixed | 40-50 | 1.43 | 1.25 – 1.64 | — (1.44–1.44) | 1 | Observational Cohort | 25 | — | — | — | — | Single study |
| male | mixed | 50-60 | 1.49 | 1.30 – 1.71 | — (1.49–1.49) | 1 | Observational Cohort | 25 | — | — | — | — | Single study |
| male | mixed | 60+ | — | NA – NA | — | NA | NA | NA | — | — | — | — | NA |
| **male** | **mixed** | **Meta-Regression** | **—** | **NA – NA** | **—** | **NA** | **NA** | **NA** | **—** | **—** | **—** | **—** | **NA** |
| **male** | **mixed** | **Overall** | **1.50** | **1.04 – 2.15** | **1.58 ± 0.37 (1.34–2.60)** | **10** | **Mixed** | **401** | **94.8%** | **173.3** | **<0.005** | **0.96** | **Random** |
| male | white | 18+ | — | NA – NA | — | NA | NA | NA | — | — | — | — | NA |
| male | white | 20-30 | — | NA – NA | — | NA | NA | NA | — | — | — | — | NA |
| male | white | 30-40 | — | NA – NA | — | NA | NA | NA | — | — | — | — | NA |
| male | white | 40-50 | 1.59 | 0.44 – 5.67 | — (1.96–1.96) | 1 | Observational Cohort | 6 | — | — | — | — | Single study |
| male | white | 50-60 | 1.75 | 0.82 – 3.75 | — (1.89–1.89) | 1 | Observational Cohort | 79 | — | — | — | — | Single study |
| male | white | 60+ | 1.55 | 1.27 – 1.87 | 1.63 ± 0.21 (1.39–1.76) | 3 | Observational Cohort | 329 | 93.9% | 32.9 | <0.005 | 0.59 | Random |
| **male** | **white** | **Meta-Regression** | **—** | **NA – NA** | **—** | **NA** | **NA** | **NA** | **—** | **—** | **—** | **—** | **NA** |
| **male** | **white** | **Overall** | **1.59** | **1.32 – 1.93** | **1.75 ± 0.22 (1.39–1.96)** | **5** | **Observational Cohort** | **414** | **90.8%** | **43.3** | **<0.005** | **0.52** | **Random** |
| female | All | 18+ | 1.54 | 0.90 – 2.63 | 1.72 ± 0.49 (1.37–2.81) | 7 | Mixed | 488 | 98.1% | 320.8 | <0.005 | 0.97 | Random |
| female | All | 20-30 | 1.27 | 1.10 – 1.47 | — (1.28–1.28) | 1 | Observational Cohort | 25 | — | — | — | — | Single study |
| female | All | 30-40 | 1.35 | 1.15 – 1.58 | — (1.35–1.35) | 1 | Observational Cohort | 25 | — | — | — | — | Single study |
| female | All | 40-50 | 1.49 | 1.00 – 2.22 | 1.58 ± 0.39 (1.26–2.02) | 3 | Observational Cohort | 578 | 99.4% | 347 | <0.005 | 0.77 | Random |
| female | All | 50-60 | 1.55 | 0.98 – 2.45 | 1.68 ± 0.44 (1.23–2.11) | 3 | Observational Cohort | 2565 | 99.8% | 825.3 | <0.005 | 0.61 | Random |
| female | All | 60+ | 1.52 | 0.82 – 2.83 | 1.68 ± 0.46 (0.89–2.08) | 5 | Observational Cohort | 496 | 99.7% | 1386.6 | <0.005 | 0.16 | Random |
| **female** | **All** | **Meta-Regression** | **1.25** | **0.55 – 2.82** | **—** | **7** | **NA** | **488** | **0%** | **0** | **1.00** | **0.97** | **Multivariable** |
| **female** | **All** | **Overall** | **1.50** | **0.94 – 2.40** | **1.64 ± 0.42 (0.89–2.81)** | **20** | **Mixed** | **4177** | **99.6%** | **4458** | **<0.005** | **0.94** | **Random** |
| female | black | 18+ | — | NA – NA | — | NA | NA | NA | — | — | — | — | NA |
| female | black | 20-30 | — | NA – NA | — | NA | NA | NA | — | — | — | — | NA |
| female | black | 30-40 | — | NA – NA | — | NA | NA | NA | — | — | — | — | NA |
| female | black | 40-50 | 1.40 | 0.77 – 2.55 | — (1.47–1.47) | 1 | Observational Cohort | 29 | — | — | — | — | Single study |
| female | black | 50-60 | 1.56 | 0.71 – 3.43 | — (1.69–1.69) | 1 | Observational Cohort | 131 | — | — | — | — | Single study |
| female | black | 60+ | 1.65 | 0.77 – 3.55 | 1.77 ± 0.10 (1.70–1.84) | 2 | Observational Cohort | 144 | 0.8% | 1 | 0.32 | — | Fixed |
| **female** | **black** | **Meta-Regression** | **—** | **NA – NA** | **—** | **NA** | **NA** | **NA** | **—** | **—** | **—** | **—** | **NA** |
| **female** | **black** | **Overall** | **1.58** | **0.74 – 3.34** | **1.68 ± 0.15 (1.47–1.84)** | **4** | **Observational Cohort** | **304** | **60%** | **7.5** | **0.06** | **0.55** | **Fixed** |
| female | mixed | 18+ | 1.54 | 0.90 – 2.63 | 1.72 ± 0.49 (1.37–2.81) | 7 | Mixed | 488 | 98.1% | 320.8 | <0.005 | 0.97 | Random |
| female | mixed | 20-30 | 1.27 | 1.10 – 1.47 | — (1.28–1.28) | 1 | Observational Cohort | 25 | — | — | — | — | Single study |
| female | mixed | 30-40 | 1.35 | 1.15 – 1.58 | — (1.35–1.35) | 1 | Observational Cohort | 25 | — | — | — | — | Single study |
| female | mixed | 40-50 | 1.26 | 1.13 – 1.41 | — (1.26–1.26) | 1 | Observational Cohort | 25 | — | — | — | — | Single study |
| female | mixed | 50-60 | 1.23 | 1.07 – 1.41 | — (1.23–1.23) | 1 | Observational Cohort | 25 | — | — | — | — | Single study |
| female | mixed | 60+ | 0.88 | 0.79 – 0.99 | — (0.89–0.89) | 1 | Observational Cohort | 25 | — | — | — | — | Single study |
| **female** | **mixed** | **Meta-Regression** | **—** | **NA – NA** | **—** | **NA** | **NA** | **NA** | **—** | **—** | **—** | **—** | **NA** |
| **female** | **mixed** | **Overall** | **1.38** | **0.82 – 2.30** | **1.50 ± 0.47 (0.89–2.81)** | **12** | **Mixed** | **613** | **99.2%** | **1318.6** | **<0.005** | **0.15** | **Random** |
| female | white | 18+ | — | NA – NA | — | NA | NA | NA | — | — | — | — | NA |
| female | white | 20-30 | — | NA – NA | — | NA | NA | NA | — | — | — | — | NA |
| female | white | 30-40 | — | NA – NA | — | NA | NA | NA | — | — | — | — | NA |
| female | white | 40-50 | 1.86 | 0.84 – 4.11 | — (2.02–2.02) | 1 | Observational Cohort | 524 | — | — | — | — | Single study |
| female | white | 50-60 | 1.96 | 0.92 – 4.19 | — (2.11–2.11) | 1 | Observational Cohort | 2409 | — | — | — | — | Single study |
| female | white | 60+ | 1.87 | 1.61 – 2.17 | 1.99 ± 0.13 (1.89–2.08) | 2 | Observational Cohort | 327 | 85% | 6.7 | <0.01 | — | Random |
| **female** | **white** | **Meta-Regression** | **—** | **NA – NA** | **—** | **NA** | **NA** | **NA** | **—** | **—** | **—** | **—** | **NA** |
| **female** | **white** | **Overall** | **1.90** | **1.76 – 2.05** | **2.02 ± 0.10 (1.89–2.11)** | **4** | **Observational Cohort** | **3260** | **78.1%** | **13.7** | **<0.005** | **0.27** | **Random** |
| *I² represents heterogeneity; Q test assesses between-study variation; Egger's p = linear regression test for funnel plot asymmetry; Meta-Regression shows covariate-adjusted estimates* | | | | | | | | | | | | | |

###### 6.1.2.3 Table of Results subdivided by race / Sex / Age

volume_ma_3rd_analysis$race_summary_table

Table 1: **race-Stratified Meta-Analysis Results**

24-Hour Urine Volume (L/24h)

| Sex Group | race | Age Band | 95% Range | Studies | Study Type | Participants | I² | Cochran's Q | Q (p-value) | Egger's p | Model |
| --- | --- | --- | --- | --- | --- | --- | --- | --- | --- | --- | --- |
| **main meta-analysis** | **All races** | **18+** | **0.94 – 2.75** | **59** | **Mixed** | **4647** | **98.5%** | **3989.9** | **<0.005** | **0.50** | **Random** |
| **main meta-analysis** | **All races** | **20-30** | **1.11 – 1.81** | **5** | **Mixed** | **153** | **91.5%** | **47.2** | **<0.005** | **0.52** | **Random** |
| **main meta-analysis** | **All races** | **30-40** | **1.30 – 1.56** | **5** | **Observational Cohort** | **185** | **90.7%** | **42.9** | **<0.005** | **0.33** | **Random** |
| **main meta-analysis** | **All races** | **40-50** | **1.15 – 2.15** | **7** | **Observational Cohort** | **665** | **99.2%** | **719.4** | **<0.005** | **0.85** | **Random** |
| **main meta-analysis** | **All races** | **50-60** | **1.12 – 2.23** | **5** | **Observational Cohort** | **2669** | **99.6%** | **928.1** | **<0.005** | **0.45** | **Random** |
| **main meta-analysis** | **All races** | **60+** | **0.95 – 2.48** | **8** | **Observational Cohort** | **825** | **99.6%** | **1683.3** | **<0.005** | **<0.05** | **Random** |
| **main meta-analysis** | **All races** | **Meta-Regression** | **0.95 – 2.60** | **52** | **NA** | **4291** | **98.4%** | **1510.9** | **<0.005** | **0.98** | **Multivariable** |
| **main meta-analysis** | **All races** | **Meta-Regression** | **0.95 – 2.60** | **52** | **NA** | **4291** | **98.4%** | **1510.9** | **<0.005** | **0.98** | **Multivariable** |
| **main meta-analysis** | **All races** | **Overall** | **0.98 – 2.53** | **89** | **Mixed** | **9144** | **99%** | **9158.8** | **<0.005** | **0.81** | **Random** |
| **main meta-analysis** | **All races** | **Overall** | **0.98 – 2.53** | **89** | **Mixed** | **9144** | **99%** | **9158.8** | **<0.005** | **0.81** | **Random** |
| main meta-analysis | Black | 18+ | — | — | NA | — | — | — | — | — | — |
| main meta-analysis | Black | 20-30 | — | — | NA | — | — | — | — | — | — |
| main meta-analysis | Black | 30-40 | — | — | NA | — | — | — | — | — | — |
| main meta-analysis | Black | 40-50 | 0.77 – 2.55 | 1 | Observational Cohort | 29 | — | — | — | — | Single study |
| main meta-analysis | Black | 50-60 | 0.71 – 3.43 | 1 | Observational Cohort | 131 | — | — | — | — | Single study |
| main meta-analysis | Black | 60+ | 0.77 – 3.55 | 2 | Observational Cohort | 144 | 0.8% | 1 | 0.32 | — | Fixed |
| main meta-analysis | Black | Meta-Regression | — | — | NA | — | — | — | — | — | — |
| **main meta-analysis** | **Black** | **Overall** | **0.74 – 3.34** | **4** | **Observational Cohort** | **304** | **60%** | **7.5** | **0.06** | **0.55** | **Fixed** |
| main meta-analysis | Mixed | 18+ | 0.95 – 2.76 | 58 | Mixed | 4484 | 98.5% | 3864 | <0.005 | 0.55 | Random |
| main meta-analysis | Mixed | 20-30 | 1.11 – 1.81 | 5 | Mixed | 153 | 91.5% | 47.2 | <0.005 | 0.52 | Random |
| main meta-analysis | Mixed | 30-40 | 1.30 – 1.56 | 5 | Observational Cohort | 185 | 90.7% | 42.9 | <0.005 | 0.33 | Random |
| main meta-analysis | Mixed | 40-50 | 1.10 – 2.18 | 4 | Observational Cohort | 106 | 99.5% | 606.5 | <0.005 | 0.70 | Random |
| main meta-analysis | Mixed | 50-60 | 1.04 – 1.77 | 2 | Observational Cohort | 50 | 98.9% | 92 | <0.005 | — | Random |
| main meta-analysis | Mixed | 60+ | 0.79 – 0.99 | 1 | Observational Cohort | 25 | — | — | — | — | Single study |
| main meta-analysis | Mixed | Meta-Regression | — | — | NA | — | — | — | — | — | — |
| **main meta-analysis** | **Mixed** | **Overall** | **0.94 – 2.59** | **75** | **Mixed** | **5003** | **99%** | **7607.8** | **<0.005** | **0.60** | **Random** |
| main meta-analysis | White | 18+ | 0.55 – 2.47 | 1 | Observational Cohort | 163 | — | — | — | — | Single study |
| main meta-analysis | White | 20-30 | — | — | NA | — | — | — | — | — | — |
| main meta-analysis | White | 30-40 | — | — | NA | — | — | — | — | — | — |
| main meta-analysis | White | 40-50 | 0.84 – 4.12 | 2 | Observational Cohort | 530 | 0% | 0.4 | 0.55 | — | Fixed |
| main meta-analysis | White | 50-60 | 1.63 – 2.15 | 2 | Observational Cohort | 2488 | 83.8% | 6.2 | <0.05 | — | Random |
| main meta-analysis | White | 60+ | 1.29 – 2.15 | 5 | Observational Cohort | 656 | 97.1% | 136.4 | <0.005 | 0.96 | Random |
| main meta-analysis | White | Meta-Regression | — | — | NA | — | — | — | — | — | — |
| **main meta-analysis** | **White** | **Overall** | **1.19 – 2.30** | **10** | **Observational Cohort** | **3837** | **98.2%** | **505.1** | **<0.005** | **0.10** | **Random** |
| **male** | **All races** | **18+** | **0.77 – 3.43** | **3** | **Observational Cohort** | **145** | **98.7%** | **155.7** | **<0.005** | **0.31** | **Random** |
| **male** | **All races** | **20-30** | **1.16 – 1.74** | **2** | **Mixed** | **63** | **0%** | **0** | **0.85** | **—** | **Fixed** |
| **male** | **All races** | **30-40** | **1.33 – 1.65** | **3** | **Observational Cohort** | **143** | **0%** | **0.9** | **0.63** | **0.92** | **Fixed** |
| **male** | **All races** | **40-50** | **1.23 – 1.67** | **2** | **Observational Cohort** | **31** | **0%** | **0.2** | **0.70** | **—** | **Fixed** |
| **male** | **All races** | **50-60** | **1.30 – 2.00** | **2** | **Observational Cohort** | **104** | **92.1%** | **12.6** | **<0.005** | **—** | **Random** |
| **male** | **All races** | **60+** | **1.27 – 1.87** | **3** | **Observational Cohort** | **329** | **93.9%** | **32.9** | **<0.005** | **0.59** | **Random** |
| **male** | **All races** | **Meta-Regression** | **—** | **—** | **NA** | **—** | **—** | **—** | **—** | **—** | **—** |
| **male** | **All races** | **Overall** | **1.11 – 2.09** | **15** | **Mixed** | **815** | **93.8%** | **225.8** | **<0.005** | **0.54** | **Random** |
| male | Black | 18+ | — | — | NA | — | — | — | — | — | — |
| male | Black | 20-30 | — | — | NA | — | — | — | — | — | — |
| male | Black | 30-40 | — | — | NA | — | — | — | — | — | — |
| male | Black | 40-50 | — | — | NA | — | — | — | — | — | — |
| male | Black | 50-60 | — | — | NA | — | — | — | — | — | — |
| male | Black | 60+ | — | — | NA | — | — | — | — | — | — |
| male | Black | Meta-Regression | — | — | NA | — | — | — | — | — | — |
| **male** | **Black** | **Overall** | **—** | **—** | **NA** | **—** | **—** | **—** | **—** | **—** | **—** |
| male | Mixed | 18+ | 0.77 – 3.43 | 3 | Observational Cohort | 145 | 98.7% | 155.7 | <0.005 | 0.31 | Random |
| male | Mixed | 20-30 | 1.16 – 1.74 | 2 | Mixed | 63 | 0% | 0 | 0.85 | — | Fixed |
| male | Mixed | 30-40 | 1.33 – 1.65 | 3 | Observational Cohort | 143 | 0% | 0.9 | 0.63 | 0.92 | Fixed |
| male | Mixed | 40-50 | 1.25 – 1.64 | 1 | Observational Cohort | 25 | — | — | — | — | Single study |
| male | Mixed | 50-60 | 1.30 – 1.71 | 1 | Observational Cohort | 25 | — | — | — | — | Single study |
| male | Mixed | 60+ | — | — | NA | — | — | — | — | — | — |
| male | Mixed | Meta-Regression | — | — | NA | — | — | — | — | — | — |
| **male** | **Mixed** | **Overall** | **1.04 – 2.15** | **10** | **Mixed** | **401** | **94.8%** | **173.3** | **<0.005** | **0.96** | **Random** |
| male | White | 18+ | — | — | NA | — | — | — | — | — | — |
| male | White | 20-30 | — | — | NA | — | — | — | — | — | — |
| male | White | 30-40 | — | — | NA | — | — | — | — | — | — |
| male | White | 40-50 | 0.44 – 5.67 | 1 | Observational Cohort | 6 | — | — | — | — | Single study |
| male | White | 50-60 | 0.82 – 3.75 | 1 | Observational Cohort | 79 | — | — | — | — | Single study |
| male | White | 60+ | 1.27 – 1.87 | 3 | Observational Cohort | 329 | 93.9% | 32.9 | <0.005 | 0.59 | Random |
| male | White | Meta-Regression | — | — | NA | — | — | — | — | — | — |
| **male** | **White** | **Overall** | **1.32 – 1.93** | **5** | **Observational Cohort** | **414** | **90.8%** | **43.3** | **<0.005** | **0.52** | **Random** |
| **female** | **All races** | **18+** | **0.90 – 2.63** | **7** | **Mixed** | **488** | **98.1%** | **320.8** | **<0.005** | **0.97** | **Random** |
| **female** | **All races** | **20-30** | **1.10 – 1.47** | **1** | **Observational Cohort** | **25** | **—** | **—** | **—** | **—** | **Single study** |
| **female** | **All races** | **30-40** | **1.15 – 1.58** | **1** | **Observational Cohort** | **25** | **—** | **—** | **—** | **—** | **Single study** |
| **female** | **All races** | **40-50** | **1.00 – 2.22** | **3** | **Observational Cohort** | **578** | **99.4%** | **347** | **<0.005** | **0.77** | **Random** |
| **female** | **All races** | **50-60** | **0.98 – 2.45** | **3** | **Observational Cohort** | **2565** | **99.8%** | **825.3** | **<0.005** | **0.61** | **Random** |
| **female** | **All races** | **60+** | **0.82 – 2.83** | **5** | **Observational Cohort** | **496** | **99.7%** | **1386.6** | **<0.005** | **0.16** | **Random** |
| **female** | **All races** | **Meta-Regression** | **0.55 – 2.82** | **7** | **NA** | **488** | **0%** | **0** | **1.00** | **0.97** | **Multivariable** |
| **female** | **All races** | **Overall** | **0.94 – 2.40** | **20** | **Mixed** | **4177** | **99.6%** | **4458** | **<0.005** | **0.94** | **Random** |
| female | Black | 18+ | — | — | NA | — | — | — | — | — | — |
| female | Black | 20-30 | — | — | NA | — | — | — | — | — | — |
| female | Black | 30-40 | — | — | NA | — | — | — | — | — | — |
| female | Black | 40-50 | 0.77 – 2.55 | 1 | Observational Cohort | 29 | — | — | — | — | Single study |
| female | Black | 50-60 | 0.71 – 3.43 | 1 | Observational Cohort | 131 | — | — | — | — | Single study |
| female | Black | 60+ | 0.77 – 3.55 | 2 | Observational Cohort | 144 | 0.8% | 1 | 0.32 | — | Fixed |
| female | Black | Meta-Regression | — | — | NA | — | — | — | — | — | — |
| **female** | **Black** | **Overall** | **0.74 – 3.34** | **4** | **Observational Cohort** | **304** | **60%** | **7.5** | **0.06** | **0.55** | **Fixed** |
| female | Mixed | 18+ | 0.90 – 2.63 | 7 | Mixed | 488 | 98.1% | 320.8 | <0.005 | 0.97 | Random |
| female | Mixed | 20-30 | 1.10 – 1.47 | 1 | Observational Cohort | 25 | — | — | — | — | Single study |
| female | Mixed | 30-40 | 1.15 – 1.58 | 1 | Observational Cohort | 25 | — | — | — | — | Single study |
| female | Mixed | 40-50 | 1.13 – 1.41 | 1 | Observational Cohort | 25 | — | — | — | — | Single study |
| female | Mixed | 50-60 | 1.07 – 1.41 | 1 | Observational Cohort | 25 | — | — | — | — | Single study |
| female | Mixed | 60+ | 0.79 – 0.99 | 1 | Observational Cohort | 25 | — | — | — | — | Single study |
| female | Mixed | Meta-Regression | — | — | NA | — | — | — | — | — | — |
| **female** | **Mixed** | **Overall** | **0.82 – 2.30** | **12** | **Mixed** | **613** | **99.2%** | **1318.6** | **<0.005** | **0.15** | **Random** |
| female | White | 18+ | — | — | NA | — | — | — | — | — | — |
| female | White | 20-30 | — | — | NA | — | — | — | — | — | — |
| female | White | 30-40 | — | — | NA | — | — | — | — | — | — |
| female | White | 40-50 | 0.84 – 4.11 | 1 | Observational Cohort | 524 | — | — | — | — | Single study |
| female | White | 50-60 | 0.92 – 4.19 | 1 | Observational Cohort | 2409 | — | — | — | — | Single study |
| female | White | 60+ | 1.61 – 2.17 | 2 | Observational Cohort | 327 | 85% | 6.7 | <0.01 | — | Random |
| female | White | Meta-Regression | — | — | NA | — | — | — | — | — | — |
| **female** | **White** | **Overall** | **1.76 – 2.05** | **4** | **Observational Cohort** | **3260** | **78.1%** | **13.7** | **<0.005** | **0.27** | **Random** |
| *I² represents heterogeneity; Q test assesses between-study variation; Egger's p = linear regression test for funnel plot asymmetry; — indicates insufficient data* | | | | | | | | | | | |

###### 6.1.2.4 Wald Tests

volume_ma_3rd_analysis$wald_tests$sex_comparison

Table 1: **Sex Comparison (Overall)**

| Comparison | % Shift | P (Wald Test) | Mean Diff (95% CI) |
| --- | --- | --- | --- |
| Male vs Female (Overall) | +1.5% | 0.937 | 0.02 [-0.54, 0.59] |
| *Mean difference = Group 1 − Group 2 in component units (mmol/24h, L/24h, or pH); % shift = difference relative to Group 2.* | | | |

volume_ma_3rd_analysis$wald_tests$race_comparisons

Table 1: **race Comparisons Within Sex Groups**

| Sex Group | Comparison | % Shift | P (Wald Test) | Mean Diff (95% CI) |
| --- | --- | --- | --- | --- |
| male | black vs mixed | — | — | — |
| male | black vs white | — | — | — |
| male | mixed vs white | -5.9% | 0.653 | -0.09 [-0.50, 0.32] |
| female | black vs mixed | +14.6% | 0.666 | 0.20 [-0.71, 1.11] |
| female | black vs white | -17.1% | 0.400 | -0.32 [-1.08, 0.43] |
| female | mixed vs white | -27.6% | <0.05 | -0.52 [-1.04, -0.01] |
| *Mean difference = Group 1 − Group 2 in component units (mmol/24h, L/24h, or pH); % shift = difference relative to Group 2.* | | | | |

volume_ma_3rd_analysis$wald_tests$age_band_comparisons

Table 1: **Age Band Comparisons Within Sex Groups**

| Sex Group | Comparison | % Shift | P (Wald Test) | Mean Diff (95% CI) |
| --- | --- | --- | --- | --- |
| main meta-analysis | 18+ vs 20-30 | +13.3% | 0.530 | 0.19 [-0.40, 0.78] |
| main meta-analysis | 18+ vs 30-40 | +12.5% | 0.520 | 0.18 [-0.37, 0.72] |
| main meta-analysis | 18+ vs 40-50 | +2.1% | 0.915 | 0.03 [-0.59, 0.65] |
| main meta-analysis | 18+ vs 50-60 | +1.8% | 0.929 | 0.03 [-0.61, 0.67] |
| main meta-analysis | 18+ vs 60+ | +4.8% | 0.840 | 0.07 [-0.65, 0.79] |
| main meta-analysis | 20-30 vs 30-40 | -0.7% | 0.939 | -0.01 [-0.27, 0.25] |
| main meta-analysis | 20-30 vs 40-50 | -9.9% | 0.442 | -0.15 [-0.55, 0.24] |
| main meta-analysis | 20-30 vs 50-60 | -10.1% | 0.459 | -0.16 [-0.58, 0.26] |
| main meta-analysis | 20-30 vs 60+ | -7.5% | 0.676 | -0.11 [-0.65, 0.42] |
| main meta-analysis | 30-40 vs 40-50 | -9.2% | 0.381 | -0.14 [-0.47, 0.18] |
| main meta-analysis | 30-40 vs 50-60 | -9.5% | 0.411 | -0.15 [-0.51, 0.21] |
| main meta-analysis | 30-40 vs 60+ | -6.8% | 0.675 | -0.10 [-0.59, 0.38] |
| main meta-analysis | 40-50 vs 50-60 | -0.3% | 0.984 | -0.00 [-0.47, 0.46] |
| main meta-analysis | 40-50 vs 60+ | +2.6% | 0.891 | 0.04 [-0.53, 0.61] |
| main meta-analysis | 50-60 vs 60+ | +2.9% | 0.882 | 0.04 [-0.55, 0.64] |
| male | 18+ vs 20-30 | +14.1% | 0.614 | 0.20 [-0.58, 0.98] |
| male | 18+ vs 30-40 | +9.5% | 0.715 | 0.14 [-0.62, 0.90] |
| male | 18+ vs 40-50 | +13.2% | 0.629 | 0.19 [-0.58, 0.95] |
| male | 18+ vs 50-60 | +0.8% | 0.975 | 0.01 [-0.77, 0.79] |
| male | 18+ vs 60+ | +4.9% | 0.849 | 0.08 [-0.70, 0.85] |
| male | 20-30 vs 30-40 | -4.0% | 0.613 | -0.06 [-0.29, 0.17] |
| male | 20-30 vs 40-50 | -0.8% | 0.929 | -0.01 [-0.27, 0.24] |
| male | 20-30 vs 50-60 | -11.7% | 0.215 | -0.19 [-0.48, 0.11] |
| male | 20-30 vs 60+ | -8.1% | 0.382 | -0.12 [-0.41, 0.16] |
| male | 30-40 vs 40-50 | +3.3% | 0.614 | 0.05 [-0.14, 0.23] |
| male | 30-40 vs 50-60 | -8.0% | 0.296 | -0.13 [-0.37, 0.11] |
| male | 30-40 vs 60+ | -4.3% | 0.558 | -0.07 [-0.29, 0.15] |
| male | 40-50 vs 50-60 | -11.0% | 0.191 | -0.18 [-0.44, 0.09] |
| male | 40-50 vs 60+ | -7.3% | 0.365 | -0.11 [-0.36, 0.13] |
| male | 50-60 vs 60+ | +4.1% | 0.671 | 0.06 [-0.23, 0.35] |
| female | 18+ vs 20-30 | +20.6% | 0.355 | 0.26 [-0.29, 0.82] |
| female | 18+ vs 30-40 | +13.8% | 0.514 | 0.19 [-0.37, 0.75] |
| female | 18+ vs 40-50 | +3.2% | 0.889 | 0.05 [-0.62, 0.72] |
| female | 18+ vs 50-60 | -1.1% | 0.962 | -0.02 [-0.72, 0.69] |
| female | 18+ vs 60+ | +0.8% | 0.976 | 0.01 [-0.81, 0.83] |
| female | 20-30 vs 30-40 | -5.6% | 0.488 | -0.08 [-0.29, 0.14] |
| female | 20-30 vs 40-50 | -14.4% | 0.319 | -0.21 [-0.64, 0.21] |
| female | 20-30 vs 50-60 | -18.0% | 0.252 | -0.28 [-0.76, 0.20] |
| female | 20-30 vs 60+ | -16.4% | 0.440 | -0.25 [-0.88, 0.38] |
| female | 30-40 vs 40-50 | -9.3% | 0.525 | -0.14 [-0.57, 0.29] |
| female | 30-40 vs 50-60 | -13.1% | 0.409 | -0.20 [-0.69, 0.28] |
| female | 30-40 vs 60+ | -11.4% | 0.593 | -0.17 [-0.81, 0.46] |
| female | 40-50 vs 50-60 | -4.2% | 0.834 | -0.06 [-0.67, 0.54] |
| female | 40-50 vs 60+ | -2.3% | 0.926 | -0.03 [-0.77, 0.70] |
| female | 50-60 vs 60+ | +2.0% | 0.939 | 0.03 [-0.74, 0.80] |
| *Mean difference = Group 1 − Group 2 in component units (mmol/24h, L/24h, or pH); % shift = difference relative to Group 2.* | | | | |

volume_ma_3rd_analysis$wald_tests$sex_within_age_bands

Table 1: **Sex Comparisons Within Age Bands**

| Age Band | % Shift | P (Wald Test) | Mean Diff (95% CI) |
| --- | --- | --- | --- |
| 18+ | +5.5% | 0.858 | 0.08 [-0.84, 1.01] |
| 20-30 | +11.5% | 0.246 | 0.15 [-0.10, 0.40] |
| 30-40 | +9.6% | 0.185 | 0.13 [-0.06, 0.32] |
| 40-50 | -3.8% | 0.794 | -0.06 [-0.48, 0.37] |
| 50-60 | +3.5% | 0.832 | 0.05 [-0.45, 0.56] |
| 60+ | +1.4% | 0.948 | 0.02 [-0.63, 0.67] |
| *Mean difference = Group 1 − Group 2 in component units (mmol/24h, L/24h, or pH); % shift = difference relative to Group 2.* | | | |

#### 6.2 Calcium

##### 6.2.1 Comparison by Age Band

###### 6.2.1.1 Plot

```
calcium_ma_2nd_analysis <- sex_age_subdivision_ma("calcium")
calcium_ma_2nd_analysis$combined_plot
## `height` was translated to `width`.
## `height` was translated to `width`.
## `height` was translated to `width`.
```

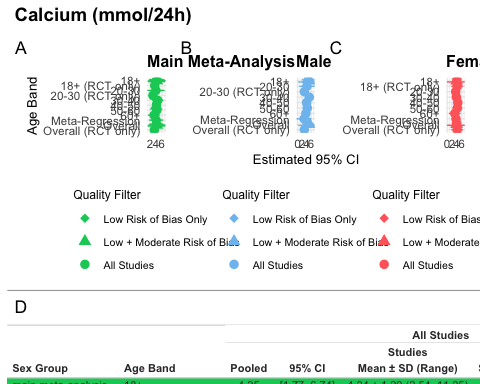

###### 6.2.1.2 Summary Table

calcium_ma_2nd_analysis$combined_summary_table

Table 1: **Combined Meta-Analysis Results**

24-Hour Urine Calcium (mmol/24h)

|  |  | **All Studies** | | | | | | **Low Risk of Bias Only** | | | | | | **Low + Moderate RoB** | | | | | |
| --- | --- | --- | --- | --- | --- | --- | --- | --- | --- | --- | --- | --- | --- | --- | --- | --- | --- | --- | --- |
| Sex Group | Age Band | Pooled | 95% CI | Studies  Mean ± SD (Range) | Studies, n | Q p-value | Egger's p | Pooled | 95% CI | Studies  Mean ± SD (Range) | Studies, n | Q p-value | Egger's p | Pooled | 95% CI | Studies  Mean ± SD (Range) | Studies, n | Q p-value | Egger's p |
| main meta-analysis | 18+ | 4.25 | [1.77, 6.74] | 4.24 ± 1.29 (2.54–11.25) | 71 | <0.001 | 0.16 | 4.17 | [1.29, 7.06] | 4.16 ± 1.46 (2.55–11.25) | 28 | <0.001 | 0.97 | 4.31 | [1.49, 7.13] | 4.28 ± 1.44 (2.54–11.25) | 40 | <0.001 | <0.005 |
| **main meta-analysis** | **18+ (RCT only)** | **3.59** | **[2.63, 4.55]** | **3.63 ± 0.55 (2.55–4.17)** | **7** | **<0.05** | **0.57** | **—** | **—** | **—** | **—** | **—** | **—** | **—** | **—** | **—** | **—** | **—** | **—** |
| main meta-analysis | 20-30 | 4.00 | [1.31, 6.69] | 3.99 ± 1.37 (2.94–6.01) | 4 | <0.001 | 0.62 | — | — | — | — | — | — | — | — | — | — | — | — |
| **main meta-analysis** | **20-30 (RCT only)** | **2.96** | **[2.00, 3.93]** | **3.21 ± 0.38 (2.94–3.48)** | **2** | **0.13** | **—** | **—** | **—** | **—** | **—** | **—** | **—** | **—** | **—** | **—** | **—** | **—** | **—** |
| main meta-analysis | 30-40 | 4.61 | [2.89, 6.33] | 4.61 ± 0.88 (3.60–5.60) | 5 | <0.001 | 0.28 | — | — | — | — | — | — | — | — | — | — | — | — |
| main meta-analysis | 40-50 | 4.17 | [3.43, 4.92] | 4.11 ± 0.43 (3.58–4.92) | 7 | <0.001 | 0.94 | — | — | — | — | — | — | 4.06 | [3.37, 4.75] | — (4.06–4.06) | 1 | — | — |
| main meta-analysis | 50-60 | 4.28 | [3.05, 5.50] | 4.27 ± 0.64 (3.23–4.98) | 5 | <0.001 | 0.14 | — | — | — | — | — | — | — | — | — | — | — | — |
| main meta-analysis | 60+ | 3.35 | [1.54, 5.17] | 3.36 ± 0.93 (1.80–4.61) | 8 | <0.001 | 0.24 | — | — | — | — | — | — | — | — | — | — | — | — |
| **main meta-analysis** | **Meta-Regression** | **4.06** | **[1.90, 6.22]** | **—** | **59** | **<0.001** | **<0.05** | **—** | **—** | **—** | **—** | **—** | **—** | **—** | **—** | **—** | **—** | **—** | **—** |
| **main meta-analysis** | **Overall** | **4.18** | **[1.89, 6.48]** | **4.17 ± 1.19 (1.80–11.25)** | **100** | **<0.001** | **0.09** | **4.17** | **[1.29, 7.06]** | **4.16 ± 1.46 (2.55–11.25)** | **28** | **<0.001** | **0.97** | **4.31** | **[1.52, 7.09]** | **4.27 ± 1.42 (2.54–11.25)** | **41** | **<0.001** | **<0.005** |
| **main meta-analysis** | **Overall (RCT only)** | **3.45** | **[2.55, 4.36]** | **3.54 ± 0.53 (2.55–4.17)** | **9** | **<0.001** | **<0.05** | **—** | **—** | **—** | **—** | **—** | **—** | **—** | **—** | **—** | **—** | **—** | **—** |
| male | 18+ | 4.75 | [2.00, 7.50] | 4.78 ± 1.46 (2.55–7.27) | 8 | <0.001 | 0.94 | 3.90 | [0.31, 7.48] | — (3.90–3.90) | 1 | — | — | 5.17 | [3.06, 7.28] | 5.16 ± 1.09 (3.90–5.79) | 3 | <0.001 | <0.001 |
| **male** | **18+ (RCT only)** | **—** | **—** | **—** | **—** | **—** | **—** | **—** | **—** | **—** | **—** | **—** | **—** | **—** | **—** | **—** | **—** | **—** | **—** |
| male | 20-30 | 4.15 | [0.93, 7.37] | 4.14 ± 1.64 (2.94–6.01) | 3 | <0.001 | 0.76 | — | — | — | — | — | — | — | — | — | — | — | — |
| **male** | **20-30 (RCT only)** | **2.96** | **[2.00, 3.93]** | **3.21 ± 0.38 (2.94–3.48)** | **2** | **0.13** | **—** | **—** | **—** | **—** | **—** | **—** | **—** | **—** | **—** | **—** | **—** | **—** | **—** |
| male | 30-40 | 5.20 | [4.29, 6.11] | 5.21 ± 0.47 (4.68–5.60) | 3 | <0.001 | 0.75 | — | — | — | — | — | — | — | — | — | — | — | — |
| male | 40-50 | 3.97 | [3.15, 4.80] | 3.91 ± 0.10 (3.84–3.98) | 2 | 0.65 | — | — | — | — | — | — | — | — | — | — | — | — | — |
| male | 50-60 | 4.29 | [2.80, 5.77] | 4.36 ± 0.13 (4.27–4.45) | 2 | 0.48 | — | — | — | — | — | — | — | — | — | — | — | — | — |
| male | 60+ | 3.66 | [2.41, 4.92] | 3.68 ± 0.64 (2.96–4.18) | 3 | <0.001 | 0.24 | — | — | — | — | — | — | — | — | — | — | — | — |
| **male** | **Meta-Regression** | **2.83** | **[-0.30, 5.97]** | **—** | **7** | **1.00** | **0.43** | **—** | **—** | **—** | **—** | **—** | **—** | **—** | **—** | **—** | **—** | **—** | **—** |
| **male** | **Overall** | **4.46** | **[2.28, 6.63]** | **4.47 ± 1.16 (2.55–7.27)** | **21** | **<0.001** | **0.45** | **3.90** | **[0.31, 7.48]** | **— (3.90–3.90)** | **1** | **—** | **—** | **5.17** | **[3.06, 7.28]** | **5.16 ± 1.09 (3.90–5.79)** | **3** | **<0.001** | **<0.001** |
| **male** | **Overall (RCT only)** | **2.96** | **[2.00, 3.93]** | **3.21 ± 0.38 (2.94–3.48)** | **2** | **0.13** | **—** | **—** | **—** | **—** | **—** | **—** | **—** | **—** | **—** | **—** | **—** | **—** | **—** |
| female | 18+ | 4.03 | [3.45, 4.61] | 4.11 ± 0.40 (3.61–4.69) | 9 | <0.005 | <0.05 | 3.61 | [0.02, 7.20] | 3.61 ± 0.00 (3.61–3.61) | 2 | 1.00 | — | 3.93 | [3.37, 4.49] | 3.96 ± 0.32 (3.61–4.22) | 5 | <0.01 | 0.18 |
| **female** | **18+ (RCT only)** | **4.17** | **[2.02, 6.32]** | **— (4.17–4.17)** | **1** | **—** | **—** | **—** | **—** | **—** | **—** | **—** | **—** | **—** | **—** | **—** | **—** | **—** | **—** |
| female | 20-30 | 3.54 | [3.01, 4.07] | — (3.54–3.54) | 1 | — | — | — | — | — | — | — | — | — | — | — | — | — | — |
| **female** | **20-30 (RCT only)** | **—** | **—** | **—** | **—** | **—** | **—** | **—** | **—** | **—** | **—** | **—** | **—** | **—** | **—** | **—** | **—** | **—** | **—** |
| female | 30-40 | 3.84 | [3.29, 4.39] | — (3.84–3.84) | 1 | — | — | — | — | — | — | — | — | — | — | — | — | — | — |
| female | 40-50 | 4.41 | [3.35, 5.47] | 4.30 ± 0.67 (3.58–4.92) | 3 | <0.001 | 0.85 | — | — | — | — | — | — | — | — | — | — | — | — |
| female | 50-60 | 4.22 | [2.49, 5.95] | 4.21 ± 0.90 (3.23–4.98) | 3 | <0.001 | 0.16 | — | — | — | — | — | — | — | — | — | — | — | — |
| female | 60+ | 3.16 | [1.03, 5.30] | 3.17 ± 1.09 (1.80–4.61) | 5 | <0.001 | 0.13 | — | — | — | — | — | — | — | — | — | — | — | — |
| **female** | **Meta-Regression** | **3.67** | **[3.67, 3.67]** | **—** | **9** | **0.25** | **<0.05** | **—** | **—** | **—** | **—** | **—** | **—** | **—** | **—** | **—** | **—** | **—** | **—** |
| **female** | **Overall** | **3.89** | **[2.39, 5.39]** | **3.90 ± 0.77 (1.80–4.98)** | **22** | **<0.001** | **0.74** | **3.61** | **[0.02, 7.20]** | **3.61 ± 0.00 (3.61–3.61)** | **2** | **1.00** | **—** | **3.93** | **[3.37, 4.49]** | **3.96 ± 0.32 (3.61–4.22)** | **5** | **<0.01** | **0.18** |
| **female** | **Overall (RCT only)** | **4.17** | **[2.02, 6.32]** | **— (4.17–4.17)** | **1** | **—** | **—** | **—** | **—** | **—** | **—** | **—** | **—** | **—** | **—** | **—** | **—** | **—** | **—** |
| *Studies, n = number of studies; Q p-value = Cochran's Q heterogeneity test; Egger's p = linear regression test for funnel plot asymmetry; RoB = Risk of Bias* | | | | | | | | | | | | | | | | | | | |

###### 6.2.1.3 Wald Tests

calcium_ma_2nd_analysis$wald_tests$sex_comparison

Table 1: **Sex Comparison (Overall)**

| Comparison | % Shift | P (Wald Test) | Mean Diff (95% CI) |
| --- | --- | --- | --- |
| Male vs Female (Overall) | +14.5% | 0.675 | 0.56 [-2.08, 3.21] |
| *Mean difference = Group 1 − Group 2 in component units (mmol/24h, L/24h, or pH); % shift = difference relative to Group 2.* | | | |

calcium_ma_2nd_analysis$wald_tests$age_band_comparisons

Table 1: **Age Band Comparisons Within Sex Groups**

| Sex Group | Comparison | % Shift | P (Wald Test) | Mean Diff (95% CI) |
| --- | --- | --- | --- | --- |
| main meta-analysis | 18+ vs 20-30 | +6.4% | 0.890 | 0.26 [-3.41, 3.92] |
| main meta-analysis | 18+ vs 30-40 | -7.7% | 0.818 | -0.36 [-3.38, 2.67] |
| main meta-analysis | 18+ vs 40-50 | +1.9% | 0.952 | 0.08 [-2.52, 2.68] |
| main meta-analysis | 18+ vs 50-60 | -0.6% | 0.987 | -0.02 [-2.80, 2.75] |
| main meta-analysis | 18+ vs 60+ | +26.8% | 0.567 | 0.90 [-2.18, 3.98] |
| main meta-analysis | 20-30 vs 30-40 | -13.3% | 0.707 | -0.61 [-3.81, 2.58] |
| main meta-analysis | 20-30 vs 40-50 | -4.2% | 0.901 | -0.18 [-2.97, 2.61] |
| main meta-analysis | 20-30 vs 50-60 | -6.6% | 0.852 | -0.28 [-3.24, 2.67] |
| main meta-analysis | 20-30 vs 60+ | +19.2% | 0.698 | 0.64 [-2.60, 3.89] |
| main meta-analysis | 30-40 vs 40-50 | +10.4% | 0.649 | 0.44 [-1.44, 2.31] |
| main meta-analysis | 30-40 vs 50-60 | +7.8% | 0.758 | 0.33 [-1.78, 2.45] |
| main meta-analysis | 30-40 vs 60+ | +37.4% | 0.325 | 1.26 [-1.25, 3.76] |
| main meta-analysis | 40-50 vs 50-60 | -2.4% | 0.887 | -0.10 [-1.54, 1.33] |
| main meta-analysis | 40-50 vs 60+ | +24.4% | 0.413 | 0.82 [-1.14, 2.78] |
| main meta-analysis | 50-60 vs 60+ | +27.5% | 0.409 | 0.92 [-1.27, 3.12] |
| male | 18+ vs 20-30 | +14.5% | 0.781 | 0.60 [-3.63, 4.83] |
| male | 18+ vs 30-40 | -8.7% | 0.761 | -0.45 [-3.35, 2.45] |
| male | 18+ vs 40-50 | +19.6% | 0.594 | 0.78 [-2.09, 3.65] |
| male | 18+ vs 50-60 | +10.8% | 0.771 | 0.46 [-2.66, 3.59] |
| male | 18+ vs 60+ | +29.7% | 0.481 | 1.09 [-1.94, 4.11] |
| male | 20-30 vs 30-40 | -20.2% | 0.538 | -1.05 [-4.39, 2.29] |
| male | 20-30 vs 40-50 | +4.5% | 0.916 | 0.18 [-3.14, 3.50] |
| male | 20-30 vs 50-60 | -3.2% | 0.940 | -0.14 [-3.68, 3.41] |
| male | 20-30 vs 60+ | +13.3% | 0.783 | 0.49 [-2.97, 3.94] |
| male | 30-40 vs 40-50 | +31.0% | <0.05 | 1.23 [0.01, 2.46] |
| male | 30-40 vs 50-60 | +21.3% | 0.303 | 0.91 [-0.83, 2.65] |
| male | 30-40 vs 60+ | +41.9% | 0.052 | 1.54 [-0.01, 3.09] |
| male | 40-50 vs 50-60 | -7.4% | 0.716 | -0.32 [-2.01, 1.38] |
| male | 40-50 vs 60+ | +8.4% | 0.689 | 0.31 [-1.20, 1.81] |
| male | 50-60 vs 60+ | +17.0% | 0.531 | 0.62 [-1.32, 2.57] |
| female | 18+ vs 20-30 | +13.9% | 0.218 | 0.49 [-0.29, 1.28] |
| female | 18+ vs 30-40 | +5.0% | 0.636 | 0.19 [-0.61, 0.99] |
| female | 18+ vs 40-50 | -8.5% | 0.544 | -0.37 [-1.58, 0.83] |
| female | 18+ vs 50-60 | -4.4% | 0.841 | -0.19 [-2.01, 1.64] |
| female | 18+ vs 60+ | +27.5% | 0.441 | 0.87 [-1.34, 3.08] |
| female | 20-30 vs 30-40 | -7.8% | 0.441 | -0.30 [-1.06, 0.46] |
| female | 20-30 vs 40-50 | -19.7% | 0.152 | -0.87 [-2.05, 0.32] |
| female | 20-30 vs 50-60 | -16.1% | 0.462 | -0.68 [-2.49, 1.13] |
| female | 20-30 vs 60+ | +11.9% | 0.737 | 0.38 [-1.82, 2.58] |
| female | 30-40 vs 40-50 | -12.9% | 0.352 | -0.57 [-1.76, 0.63] |
| female | 30-40 vs 50-60 | -9.0% | 0.682 | -0.38 [-2.20, 1.44] |
| female | 30-40 vs 60+ | +21.4% | 0.548 | 0.68 [-1.53, 2.88] |
| female | 40-50 vs 50-60 | +4.4% | 0.856 | 0.19 [-1.84, 2.22] |
| female | 40-50 vs 60+ | +39.3% | 0.307 | 1.24 [-1.14, 3.63] |
| female | 50-60 vs 60+ | +33.4% | 0.452 | 1.06 [-1.69, 3.81] |
| *Mean difference = Group 1 − Group 2 in component units (mmol/24h, L/24h, or pH); % shift = difference relative to Group 2.* | | | | |

calcium_ma_2nd_analysis$wald_tests$sex_within_age_bands

Table 1: **Sex Comparisons Within Age Bands**

| Age Band | % Shift | P (Wald Test) | Mean Diff (95% CI) |
| --- | --- | --- | --- |
| 18+ | +17.8% | 0.616 | 0.72 [-2.09, 3.53] |
| 20-30 | +17.2% | 0.714 | 0.61 [-2.65, 3.87] |
| 30-40 | +35.5% | <0.05 | 1.36 [0.30, 2.42] |
| 40-50 | -9.9% | 0.525 | -0.44 [-1.78, 0.91] |
| 50-60 | +1.6% | 0.954 | 0.07 [-2.21, 2.35] |
| 60+ | +15.8% | 0.692 | 0.50 [-1.98, 2.98] |
| *Mean difference = Group 1 − Group 2 in component units (mmol/24h, L/24h, or pH); % shift = difference relative to Group 2.* | | | |

###### 6.2.1.4 Meta-Regression (plain language)

format_meta_reg_plain("calcium", sex_group = "all")

Message

No meta-regression result for calcium / all

###### 6.2.1.5 Assay Subgroup Analysis

assay_subgroup_table("calcium")

Table 1: **Assay subgroup analysis — Calcium**

*Per-assay pooled estimates; categories with k < 3 listed but not pooled.*

| Assay category | Studies, n | Participants | 95% CI | I² | Q p-value | Egger's p | Note |
| --- | --- | --- | --- | --- | --- | --- | --- |
| Unreported | 59 | 6,942 | 4.22 [3.85, 4.59] | 100% | <0.001 | 0.206 |  |
| Autoanalyser | 21 | 2,676 | 4.23 [4.04, 4.42] | 98% | <0.001 | 0.060 |  |
| Atomic absorption | 17 | 460 | 4.06 [3.57, 4.56] | 94% | <0.001 | 0.621 |  |
| Colourimetric (cresol) | 1 | 75 | — | — | — | — | k = 1 < min_k = 3 (skipped) |
| Colourimetric (other) | 1 | 109 | — | — | — | — | k = 1 < min_k = 3 (skipped) |
| ICP / ICP-MS | 1 | 22 | — | — | — | — | k = 1 < min_k = 3 (skipped) |

###### 6.2.1.6 Preservative Sensitivity (Litholink / acidified samples)

```
preservative_sensitivity_ma("calcium")
## # A tibble: 3 × 6
##   group               n_studies n_participants pooled_estimate lower_95 upper_95
##   <chr>                   <int>          <int>           <dbl>    <dbl>    <dbl>
## 1 All studies               100          10284            4.18     3.94     4.42
## 2 Excluding preserva…        97          10173            4.16     3.92     4.40
## 3 Preservative only           3            111            5.01     2.88     7.14
```

###### 6.2.1.7 Creatinine Completeness Sensitivity

creat_completeness_table("calcium")

Table 1: **Creatinine-completeness sensitivity — Calcium**

*Compares the all-studies pooled estimate against subsets defined by whether the study reported 24h urine creatinine. Studies that do not report creatinine cannot verify collection completeness; a substantial shift here flags possible bias from incomplete collections.*

| Subset | Studies, n | Participants | Range (95% CI) | Δ vs all-studies (%) | Flag |
| --- | --- | --- | --- | --- | --- |
| All studies | 100 | 10,284 | 4.18 [3.94, 4.42] | — | — |
| Creatinine reported only | 63 | 7,834 | 3.94 [3.76, 4.12] | -5.8% | OK |
| Creatinine NOT reported | 37 | 2,450 | 4.59 [4.04, 5.14] | +9.8% | OK |

##### 6.2.2 Comparison by Age / Sex / race

###### 6.2.2.1 Plot

```
calcium_ma_3rd_analysis <- sex_age_race_subdivision_ma("calcium",
                                                      urine_ma2,
                                                      overall_bias)
calcium_ma_3rd_analysis$race_plot
## `height` was translated to `width`.
## `height` was translated to `width`.
## `height` was translated to `width`.
```

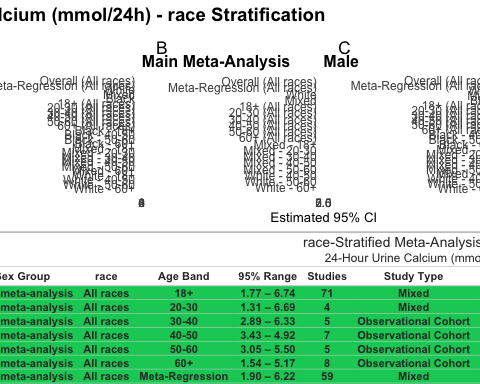

###### 6.2.2.2 Combined MA Results

calcium_ma_3rd_analysis$combined_summary_table

Table 1: **Combined Meta-Analysis Results**

24-Hour Urine Calcium (mmol/24h)

| Sex Group | race | Age Band | Pooled | 95% Range | Studies Mean SD Range | Studies | Study Type | Participants | I² | Cochran's Q | Q (p-value) | Egger's p | Model |
| --- | --- | --- | --- | --- | --- | --- | --- | --- | --- | --- | --- | --- | --- |
| main meta-analysis | All | 18+ | 4.25 | 1.77 – 6.74 | 4.24 ± 1.29 (2.54–11.25) | 71 | Mixed | 5844 | 99.5% | 12921.7 | <0.005 | 0.16 | Random |
| main meta-analysis | All | 20-30 | 4.00 | 1.31 – 6.69 | 3.99 ± 1.37 (2.94–6.01) | 4 | Mixed | 96 | 99.4% | 482.6 | <0.005 | 0.62 | Random |
| main meta-analysis | All | 30-40 | 4.61 | 2.89 – 6.33 | 4.61 ± 0.88 (3.60–5.60) | 5 | Observational Cohort | 185 | 99.5% | 866.7 | <0.005 | 0.28 | Random |
| main meta-analysis | All | 40-50 | 4.17 | 3.43 – 4.92 | 4.11 ± 0.43 (3.58–4.92) | 7 | Observational Cohort | 665 | 92.3% | 77.7 | <0.005 | 0.94 | Random |
| main meta-analysis | All | 50-60 | 4.28 | 3.05 – 5.50 | 4.27 ± 0.64 (3.23–4.98) | 5 | Observational Cohort | 2669 | 97.2% | 143.2 | <0.005 | 0.14 | Random |
| main meta-analysis | All | 60+ | 3.35 | 1.54 – 5.17 | 3.36 ± 0.93 (1.80–4.61) | 8 | Observational Cohort | 825 | 98.9% | 653.2 | <0.005 | 0.24 | Random |
| **main meta-analysis** | **All** | **Meta-Regression** | **4.06** | **1.90 – 6.22** | **—** | **59** | **Mixed** | **5418** | **98.1%** | **2744.8** | **<0.005** | **<0.05** | **Multivariable** |
| **main meta-analysis** | **All** | **Meta-Regression** | **4.06** | **1.90 – 6.22** | **—** | **59** | **Mixed** | **5418** | **98.1%** | **2744.8** | **<0.005** | **<0.05** | **Multivariable** |
| **main meta-analysis** | **All** | **Overall** | **4.18** | **1.89 – 6.48** | **4.17 ± 1.19 (1.80–11.25)** | **100** | **Mixed** | **10284** | **99.5%** | **21182.4** | **<0.005** | **0.09** | **Random** |
| **main meta-analysis** | **All** | **Overall** | **4.18** | **1.89 – 6.48** | **4.17 ± 1.19 (1.80–11.25)** | **100** | **Mixed** | **10284** | **99.5%** | **21182.4** | **<0.005** | **0.09** | **Random** |
| main meta-analysis | black | 18+ | 3.07 | -0.22 – 6.36 | 3.07 ± 0.00 (3.07–3.07) | 3 | Observational Cohort | 51 | 0% | 0 | 1.00 | — | Fixed |
| main meta-analysis | black | 20-30 | — | NA – NA | — | NA | NA | NA | — | — | — | — | NA |
| main meta-analysis | black | 30-40 | — | NA – NA | — | NA | NA | NA | — | — | — | — | NA |
| main meta-analysis | black | 40-50 | 3.58 | -1.06 – 8.23 | — (3.58–3.58) | 1 | Observational Cohort | 29 | — | — | — | — | Single study |
| main meta-analysis | black | 50-60 | 3.23 | -0.69 – 7.15 | — (3.23–3.23) | 1 | Observational Cohort | 131 | — | — | — | — | Single study |
| main meta-analysis | black | 60+ | 2.84 | -0.72 – 6.40 | 2.81 ± 0.39 (2.53–3.09) | 2 | Observational Cohort | 144 | 69.7% | 3.3 | 0.07 | — | Fixed |
| **main meta-analysis** | **black** | **Meta-Regression** | **—** | **NA – NA** | **—** | **NA** | **NA** | **NA** | **—** | **—** | **—** | **—** | **NA** |
| **main meta-analysis** | **black** | **Overall** | **3.05** | **-0.65 – 6.75** | **3.09 ± 0.31 (2.53–3.58)** | **7** | **Observational Cohort** | **355** | **22.3%** | **7.7** | **0.26** | **0.78** | **Fixed** |
| main meta-analysis | mixed | 18+ | 4.30 | 1.69 – 6.91 | 4.28 ± 1.35 (2.54–11.25) | 61 | Mixed | 5588 | 99.5% | 12803.1 | <0.005 | 0.24 | Random |
| main meta-analysis | mixed | 20-30 | 4.00 | 1.31 – 6.69 | 3.99 ± 1.37 (2.94–6.01) | 4 | Mixed | 96 | 99.4% | 482.6 | <0.005 | 0.62 | Random |
| main meta-analysis | mixed | 30-40 | 4.61 | 2.89 – 6.33 | 4.61 ± 0.88 (3.60–5.60) | 5 | Observational Cohort | 185 | 99.5% | 866.7 | <0.005 | 0.28 | Random |
| main meta-analysis | mixed | 40-50 | 4.12 | 3.75 – 4.48 | 4.11 ± 0.20 (3.98–4.41) | 4 | Observational Cohort | 106 | 85.6% | 20.8 | <0.005 | 0.89 | Random |
| main meta-analysis | mixed | 50-60 | 4.32 | 3.42 – 5.21 | 4.34 ± 0.11 (4.27–4.42) | 2 | Observational Cohort | 50 | 13.2% | 1.2 | 0.28 | — | Fixed |
| main meta-analysis | mixed | 60+ | 1.80 | 1.29 – 2.31 | — (1.80–1.80) | 1 | Observational Cohort | 25 | — | — | — | — | Single study |
| **main meta-analysis** | **mixed** | **Meta-Regression** | **—** | **NA – NA** | **—** | **NA** | **NA** | **NA** | **—** | **—** | **—** | **—** | **NA** |
| **main meta-analysis** | **mixed** | **Overall** | **4.26** | **1.78 – 6.74** | **4.25 ± 1.29 (1.80–11.25)** | **77** | **Mixed** | **6050** | **99.6%** | **18914.3** | **<0.005** | **0.14** | **Random** |
| main meta-analysis | white | 18+ | 4.54 | 0.57 – 8.52 | 4.58 ± 0.08 (4.51–4.65) | 6 | Observational Cohort | 165 | 0% | 0.1 | 1.00 | <0.001 | Fixed |
| main meta-analysis | white | 20-30 | — | NA – NA | — | NA | NA | NA | — | — | — | — | NA |
| main meta-analysis | white | 30-40 | — | NA – NA | — | NA | NA | NA | — | — | — | — | NA |
| main meta-analysis | white | 40-50 | 4.41 | 2.99 – 5.84 | 4.38 ± 0.76 (3.84–4.92) | 2 | Observational Cohort | 530 | 91.3% | 11.5 | <0.005 | — | Random |
| main meta-analysis | white | 50-60 | 4.77 | 4.12 – 5.42 | 4.72 ± 0.37 (4.45–4.98) | 2 | Observational Cohort | 2488 | 78% | 4.5 | <0.05 | — | Random |
| main meta-analysis | white | 60+ | 3.88 | 2.68 – 5.09 | 3.89 ± 0.61 (2.96–4.61) | 5 | Observational Cohort | 656 | 97.8% | 181.8 | <0.005 | 0.05 | Random |
| **main meta-analysis** | **white** | **Meta-Regression** | **—** | **NA – NA** | **—** | **NA** | **NA** | **NA** | **—** | **—** | **—** | **—** | **NA** |
| **main meta-analysis** | **white** | **Overall** | **4.31** | **3.24 – 5.38** | **4.34 ± 0.52 (2.96–4.98)** | **15** | **Observational Cohort** | **3839** | **98.8%** | **1217.2** | **<0.005** | **0.41** | **Random** |
| male | All | 18+ | 4.75 | 2.00 – 7.50 | 4.78 ± 1.46 (2.55–7.27) | 8 | Observational Cohort | 492 | 99.7% | 2289.5 | <0.005 | 0.94 | Random |
| male | All | 20-30 | 4.15 | 0.93 – 7.37 | 4.14 ± 1.64 (2.94–6.01) | 3 | Mixed | 71 | 99.6% | 474.1 | <0.005 | 0.76 | Random |
| male | All | 30-40 | 5.20 | 4.29 – 6.11 | 5.21 ± 0.47 (4.68–5.60) | 3 | Observational Cohort | 143 | 97.6% | 82.6 | <0.005 | 0.75 | Random |
| male | All | 40-50 | 3.97 | 3.15 – 4.80 | 3.91 ± 0.10 (3.84–3.98) | 2 | Observational Cohort | 31 | 0% | 0.2 | 0.65 | — | Fixed |
| male | All | 50-60 | 4.29 | 2.80 – 5.77 | 4.36 ± 0.13 (4.27–4.45) | 2 | Observational Cohort | 104 | 0% | 0.5 | 0.48 | — | Fixed |
| male | All | 60+ | 3.66 | 2.41 – 4.92 | 3.68 ± 0.64 (2.96–4.18) | 3 | Observational Cohort | 329 | 97.2% | 72.2 | <0.005 | 0.24 | Random |
| **male** | **All** | **Meta-Regression** | **2.83** | **-0.30 – 5.97** | **—** | **7** | **Mixed** | **431** | **0%** | **0** | **1.00** | **0.43** | **Multivariable** |
| **male** | **All** | **Overall** | **4.46** | **2.28 – 6.63** | **4.47 ± 1.16 (2.55–7.27)** | **21** | **Mixed** | **1170** | **99.7%** | **5784.2** | **<0.005** | **0.45** | **Random** |
| male | black | 18+ | — | NA – NA | — | NA | NA | NA | — | — | — | — | NA |
| male | black | 20-30 | — | NA – NA | — | NA | NA | NA | — | — | — | — | NA |
| male | black | 30-40 | — | NA – NA | — | NA | NA | NA | — | — | — | — | NA |
| male | black | 40-50 | — | NA – NA | — | NA | NA | NA | — | — | — | — | NA |
| male | black | 50-60 | — | NA – NA | — | NA | NA | NA | — | — | — | — | NA |
| male | black | 60+ | — | NA – NA | — | NA | NA | NA | — | — | — | — | NA |
| **male** | **black** | **Meta-Regression** | **—** | **NA – NA** | **—** | **NA** | **NA** | **NA** | **—** | **—** | **—** | **—** | **NA** |
| **male** | **black** | **Overall** | **—** | **NA – NA** | **—** | **NA** | **NA** | **NA** | **—** | **—** | **—** | **—** | **NA** |
| male | mixed | 18+ | 4.75 | 2.00 – 7.50 | 4.78 ± 1.46 (2.55–7.27) | 8 | Observational Cohort | 492 | 99.7% | 2289.5 | <0.005 | 0.94 | Random |
| male | mixed | 20-30 | 4.15 | 0.93 – 7.37 | 4.14 ± 1.64 (2.94–6.01) | 3 | Mixed | 71 | 99.6% | 474.1 | <0.005 | 0.76 | Random |
| male | mixed | 30-40 | 5.20 | 4.29 – 6.11 | 5.21 ± 0.47 (4.68–5.60) | 3 | Observational Cohort | 143 | 97.6% | 82.6 | <0.005 | 0.75 | Random |
| male | mixed | 40-50 | 3.98 | 3.22 – 4.74 | — (3.98–3.98) | 1 | Observational Cohort | 25 | — | — | — | — | Single study |
| male | mixed | 50-60 | 4.27 | 3.51 – 5.03 | — (4.27–4.27) | 1 | Observational Cohort | 25 | — | — | — | — | Single study |
| male | mixed | 60+ | — | NA – NA | — | NA | NA | NA | — | — | — | — | NA |
| **male** | **mixed** | **Meta-Regression** | **—** | **NA – NA** | **—** | **NA** | **NA** | **NA** | **—** | **—** | **—** | **—** | **NA** |
| **male** | **mixed** | **Overall** | **4.64** | **2.31 – 6.98** | **4.66 ± 1.24 (2.55–7.27)** | **16** | **Mixed** | **756** | **99.6%** | **3679.2** | **<0.005** | **0.51** | **Random** |
| male | white | 18+ | — | NA – NA | — | NA | NA | NA | — | — | — | — | NA |
| male | white | 20-30 | — | NA – NA | — | NA | NA | NA | — | — | — | — | NA |
| male | white | 30-40 | — | NA – NA | — | NA | NA | NA | — | — | — | — | NA |
| male | white | 40-50 | 3.84 | 2.39 – 5.28 | — (3.84–3.84) | 1 | Observational Cohort | 6 | — | — | — | — | Single study |
| male | white | 50-60 | 4.45 | 0.20 – 8.71 | — (4.45–4.45) | 1 | Observational Cohort | 79 | — | — | — | — | Single study |
| male | white | 60+ | 3.66 | 2.41 – 4.92 | 3.68 ± 0.64 (2.96–4.18) | 3 | Observational Cohort | 329 | 97.2% | 72.2 | <0.005 | 0.24 | Random |
| **male** | **white** | **Meta-Regression** | **—** | **NA – NA** | **—** | **NA** | **NA** | **NA** | **—** | **—** | **—** | **—** | **NA** |
| **male** | **white** | **Overall** | **3.84** | **2.73 – 4.95** | **3.87 ± 0.56 (2.96–4.45)** | **5** | **Observational Cohort** | **414** | **96.4%** | **109.9** | **<0.005** | **<0.05** | **Random** |
| female | All | 18+ | 4.03 | 3.45 – 4.61 | 4.11 ± 0.40 (3.61–4.69) | 9 | Mixed | 710 | 66.3% | 23.7 | <0.005 | <0.05 | Random |
| female | All | 20-30 | 3.54 | 3.01 – 4.07 | — (3.54–3.54) | 1 | Observational Cohort | 25 | — | — | — | — | Single study |
| female | All | 30-40 | 3.84 | 3.29 – 4.39 | — (3.84–3.84) | 1 | Observational Cohort | 25 | — | — | — | — | Single study |
| female | All | 40-50 | 4.41 | 3.35 – 5.47 | 4.30 ± 0.67 (3.58–4.92) | 3 | Observational Cohort | 578 | 90.6% | 21.2 | <0.005 | 0.85 | Random |
| female | All | 50-60 | 4.22 | 2.49 – 5.95 | 4.21 ± 0.90 (3.23–4.98) | 3 | Observational Cohort | 2565 | 98.1% | 106.5 | <0.005 | 0.16 | Random |
| female | All | 60+ | 3.16 | 1.03 – 5.30 | 3.17 ± 1.09 (1.80–4.61) | 5 | Observational Cohort | 496 | 99% | 392.7 | <0.005 | 0.13 | Random |
| **female** | **All** | **Meta-Regression** | **3.67** | **3.67 – 3.67** | **—** | **9** | **Mixed** | **710** | **0%** | **6.6** | **0.25** | **<0.05** | **Multivariable** |
| **female** | **All** | **Overall** | **3.89** | **2.39 – 5.39** | **3.90 ± 0.77 (1.80–4.98)** | **22** | **Mixed** | **4399** | **99.1%** | **2452.1** | **<0.005** | **0.74** | **Random** |
| female | black | 18+ | — | NA – NA | — | NA | NA | NA | — | — | — | — | NA |
| female | black | 20-30 | — | NA – NA | — | NA | NA | NA | — | — | — | — | NA |
| female | black | 30-40 | — | NA – NA | — | NA | NA | NA | — | — | — | — | NA |
| female | black | 40-50 | 3.58 | -1.06 – 8.23 | — (3.58–3.58) | 1 | Observational Cohort | 29 | — | — | — | — | Single study |
| female | black | 50-60 | 3.23 | -0.69 – 7.15 | — (3.23–3.23) | 1 | Observational Cohort | 131 | — | — | — | — | Single study |
| female | black | 60+ | 2.84 | -0.72 – 6.40 | 2.81 ± 0.39 (2.53–3.09) | 2 | Observational Cohort | 144 | 69.7% | 3.3 | 0.07 | — | Fixed |
| **female** | **black** | **Meta-Regression** | **—** | **NA – NA** | **—** | **NA** | **NA** | **NA** | **—** | **—** | **—** | **—** | **NA** |
| **female** | **black** | **Overall** | **3.04** | **-0.74 – 6.83** | **3.11 ± 0.44 (2.53–3.58)** | **4** | **Observational Cohort** | **304** | **61.1%** | **7.7** | **0.05** | **0.83** | **Fixed** |
| female | mixed | 18+ | 4.03 | 3.45 – 4.61 | 4.11 ± 0.40 (3.61–4.69) | 9 | Mixed | 710 | 66.3% | 23.7 | <0.005 | <0.05 | Random |
| female | mixed | 20-30 | 3.54 | 3.01 – 4.07 | — (3.54–3.54) | 1 | Observational Cohort | 25 | — | — | — | — | Single study |
| female | mixed | 30-40 | 3.84 | 3.29 – 4.39 | — (3.84–3.84) | 1 | Observational Cohort | 25 | — | — | — | — | Single study |
| female | mixed | 40-50 | 4.41 | 3.68 – 5.14 | — (4.41–4.41) | 1 | Observational Cohort | 25 | — | — | — | — | Single study |
| female | mixed | 50-60 | 4.42 | 3.28 – 5.56 | — (4.42–4.42) | 1 | Observational Cohort | 25 | — | — | — | — | Single study |
| female | mixed | 60+ | 1.80 | 1.29 – 2.31 | — (1.80–1.80) | 1 | Observational Cohort | 25 | — | — | — | — | Single study |
| **female** | **mixed** | **Meta-Regression** | **—** | **NA – NA** | **—** | **NA** | **NA** | **NA** | **—** | **—** | **—** | **—** | **NA** |
| **female** | **mixed** | **Overall** | **3.90** | **2.50 – 5.31** | **3.93 ± 0.72 (1.80–4.69)** | **14** | **Mixed** | **835** | **99%** | **1358.4** | **<0.005** | **0.18** | **Random** |
| female | white | 18+ | — | NA – NA | — | NA | NA | NA | — | — | — | — | NA |
| female | white | 20-30 | — | NA – NA | — | NA | NA | NA | — | — | — | — | NA |
| female | white | 30-40 | — | NA – NA | — | NA | NA | NA | — | — | — | — | NA |
| female | white | 40-50 | 4.92 | 0.35 – 9.49 | — (4.92–4.92) | 1 | Observational Cohort | 524 | — | — | — | — | Single study |
| female | white | 50-60 | 4.98 | 0.43 – 9.54 | — (4.98–4.98) | 1 | Observational Cohort | 2409 | — | — | — | — | Single study |
| female | white | 60+ | 4.23 | 3.18 – 5.27 | 4.21 ± 0.57 (3.81–4.61) | 2 | Observational Cohort | 327 | 88.9% | 9 | <0.005 | — | Random |
| **female** | **white** | **Meta-Regression** | **—** | **NA – NA** | **—** | **NA** | **NA** | **NA** | **—** | **—** | **—** | **—** | **NA** |
| **female** | **white** | **Overall** | **4.61** | **3.65 – 5.57** | **4.58 ± 0.54 (3.81–4.98)** | **4** | **Observational Cohort** | **3260** | **90.3%** | **31.1** | **<0.005** | **0.11** | **Random** |
| *I² represents heterogeneity; Q test assesses between-study variation; Egger's p = linear regression test for funnel plot asymmetry; Meta-Regression shows covariate-adjusted estimates* | | | | | | | | | | | | | |

###### 6.2.2.3 Table of Results subdivided by race / Sex / Age

calcium_ma_3rd_analysis$race_summary_table

Table 1: **race-Stratified Meta-Analysis Results**

24-Hour Urine Calcium (mmol/24h)

| Sex Group | race | Age Band | 95% Range | Studies | Study Type | Participants | I² | Cochran's Q | Q (p-value) | Egger's p | Model |
| --- | --- | --- | --- | --- | --- | --- | --- | --- | --- | --- | --- |
| **main meta-analysis** | **All races** | **18+** | **1.77 – 6.74** | **71** | **Mixed** | **5844** | **99.5%** | **12921.7** | **<0.005** | **0.16** | **Random** |
| **main meta-analysis** | **All races** | **20-30** | **1.31 – 6.69** | **4** | **Mixed** | **96** | **99.4%** | **482.6** | **<0.005** | **0.62** | **Random** |
| **main meta-analysis** | **All races** | **30-40** | **2.89 – 6.33** | **5** | **Observational Cohort** | **185** | **99.5%** | **866.7** | **<0.005** | **0.28** | **Random** |
| **main meta-analysis** | **All races** | **40-50** | **3.43 – 4.92** | **7** | **Observational Cohort** | **665** | **92.3%** | **77.7** | **<0.005** | **0.94** | **Random** |
| **main meta-analysis** | **All races** | **50-60** | **3.05 – 5.50** | **5** | **Observational Cohort** | **2669** | **97.2%** | **143.2** | **<0.005** | **0.14** | **Random** |
| **main meta-analysis** | **All races** | **60+** | **1.54 – 5.17** | **8** | **Observational Cohort** | **825** | **98.9%** | **653.2** | **<0.005** | **0.24** | **Random** |
| **main meta-analysis** | **All races** | **Meta-Regression** | **1.90 – 6.22** | **59** | **Mixed** | **5418** | **98.1%** | **2744.8** | **<0.005** | **<0.05** | **Multivariable** |
| **main meta-analysis** | **All races** | **Meta-Regression** | **1.90 – 6.22** | **59** | **Mixed** | **5418** | **98.1%** | **2744.8** | **<0.005** | **<0.05** | **Multivariable** |
| **main meta-analysis** | **All races** | **Overall** | **1.89 – 6.48** | **100** | **Mixed** | **10284** | **99.5%** | **21182.4** | **<0.005** | **0.09** | **Random** |
| **main meta-analysis** | **All races** | **Overall** | **1.89 – 6.48** | **100** | **Mixed** | **10284** | **99.5%** | **21182.4** | **<0.005** | **0.09** | **Random** |
| main meta-analysis | Black | 18+ | -0.22 – 6.36 | 3 | Observational Cohort | 51 | 0% | 0 | 1.00 | — | Fixed |
| main meta-analysis | Black | 20-30 | — | — | NA | — | — | — | — | — | — |
| main meta-analysis | Black | 30-40 | — | — | NA | — | — | — | — | — | — |
| main meta-analysis | Black | 40-50 | -1.06 – 8.23 | 1 | Observational Cohort | 29 | — | — | — | — | Single study |
| main meta-analysis | Black | 50-60 | -0.69 – 7.15 | 1 | Observational Cohort | 131 | — | — | — | — | Single study |
| main meta-analysis | Black | 60+ | -0.72 – 6.40 | 2 | Observational Cohort | 144 | 69.7% | 3.3 | 0.07 | — | Fixed |
| main meta-analysis | Black | Meta-Regression | — | — | NA | — | — | — | — | — | — |
| **main meta-analysis** | **Black** | **Overall** | **-0.65 – 6.75** | **7** | **Observational Cohort** | **355** | **22.3%** | **7.7** | **0.26** | **0.78** | **Fixed** |
| main meta-analysis | Mixed | 18+ | 1.69 – 6.91 | 61 | Mixed | 5588 | 99.5% | 12803.1 | <0.005 | 0.24 | Random |
| main meta-analysis | Mixed | 20-30 | 1.31 – 6.69 | 4 | Mixed | 96 | 99.4% | 482.6 | <0.005 | 0.62 | Random |
| main meta-analysis | Mixed | 30-40 | 2.89 – 6.33 | 5 | Observational Cohort | 185 | 99.5% | 866.7 | <0.005 | 0.28 | Random |
| main meta-analysis | Mixed | 40-50 | 3.75 – 4.48 | 4 | Observational Cohort | 106 | 85.6% | 20.8 | <0.005 | 0.89 | Random |
| main meta-analysis | Mixed | 50-60 | 3.42 – 5.21 | 2 | Observational Cohort | 50 | 13.2% | 1.2 | 0.28 | — | Fixed |
| main meta-analysis | Mixed | 60+ | 1.29 – 2.31 | 1 | Observational Cohort | 25 | — | — | — | — | Single study |
| main meta-analysis | Mixed | Meta-Regression | — | — | NA | — | — | — | — | — | — |
| **main meta-analysis** | **Mixed** | **Overall** | **1.78 – 6.74** | **77** | **Mixed** | **6050** | **99.6%** | **18914.3** | **<0.005** | **0.14** | **Random** |
| main meta-analysis | White | 18+ | 0.57 – 8.52 | 6 | Observational Cohort | 165 | 0% | 0.1 | 1.00 | <0.001 | Fixed |
| main meta-analysis | White | 20-30 | — | — | NA | — | — | — | — | — | — |
| main meta-analysis | White | 30-40 | — | — | NA | — | — | — | — | — | — |
| main meta-analysis | White | 40-50 | 2.99 – 5.84 | 2 | Observational Cohort | 530 | 91.3% | 11.5 | <0.005 | — | Random |
| main meta-analysis | White | 50-60 | 4.12 – 5.42 | 2 | Observational Cohort | 2488 | 78% | 4.5 | <0.05 | — | Random |
| main meta-analysis | White | 60+ | 2.68 – 5.09 | 5 | Observational Cohort | 656 | 97.8% | 181.8 | <0.005 | 0.05 | Random |
| main meta-analysis | White | Meta-Regression | — | — | NA | — | — | — | — | — | — |
| **main meta-analysis** | **White** | **Overall** | **3.24 – 5.38** | **15** | **Observational Cohort** | **3839** | **98.8%** | **1217.2** | **<0.005** | **0.41** | **Random** |
| **male** | **All races** | **18+** | **2.00 – 7.50** | **8** | **Observational Cohort** | **492** | **99.7%** | **2289.5** | **<0.005** | **0.94** | **Random** |
| **male** | **All races** | **20-30** | **0.93 – 7.37** | **3** | **Mixed** | **71** | **99.6%** | **474.1** | **<0.005** | **0.76** | **Random** |
| **male** | **All races** | **30-40** | **4.29 – 6.11** | **3** | **Observational Cohort** | **143** | **97.6%** | **82.6** | **<0.005** | **0.75** | **Random** |
| **male** | **All races** | **40-50** | **3.15 – 4.80** | **2** | **Observational Cohort** | **31** | **0%** | **0.2** | **0.65** | **—** | **Fixed** |
| **male** | **All races** | **50-60** | **2.80 – 5.77** | **2** | **Observational Cohort** | **104** | **0%** | **0.5** | **0.48** | **—** | **Fixed** |
| **male** | **All races** | **60+** | **2.41 – 4.92** | **3** | **Observational Cohort** | **329** | **97.2%** | **72.2** | **<0.005** | **0.24** | **Random** |
| **male** | **All races** | **Meta-Regression** | **-0.30 – 5.97** | **7** | **Mixed** | **431** | **0%** | **0** | **1.00** | **0.43** | **Multivariable** |
| **male** | **All races** | **Overall** | **2.28 – 6.63** | **21** | **Mixed** | **1170** | **99.7%** | **5784.2** | **<0.005** | **0.45** | **Random** |
| male | Black | 18+ | — | — | NA | — | — | — | — | — | — |
| male | Black | 20-30 | — | — | NA | — | — | — | — | — | — |
| male | Black | 30-40 | — | — | NA | — | — | — | — | — | — |
| male | Black | 40-50 | — | — | NA | — | — | — | — | — | — |
| male | Black | 50-60 | — | — | NA | — | — | — | — | — | — |
| male | Black | 60+ | — | — | NA | — | — | — | — | — | — |
| male | Black | Meta-Regression | — | — | NA | — | — | — | — | — | — |
| **male** | **Black** | **Overall** | **—** | **—** | **NA** | **—** | **—** | **—** | **—** | **—** | **—** |
| male | Mixed | 18+ | 2.00 – 7.50 | 8 | Observational Cohort | 492 | 99.7% | 2289.5 | <0.005 | 0.94 | Random |
| male | Mixed | 20-30 | 0.93 – 7.37 | 3 | Mixed | 71 | 99.6% | 474.1 | <0.005 | 0.76 | Random |
| male | Mixed | 30-40 | 4.29 – 6.11 | 3 | Observational Cohort | 143 | 97.6% | 82.6 | <0.005 | 0.75 | Random |
| male | Mixed | 40-50 | 3.22 – 4.74 | 1 | Observational Cohort | 25 | — | — | — | — | Single study |
| male | Mixed | 50-60 | 3.51 – 5.03 | 1 | Observational Cohort | 25 | — | — | — | — | Single study |
| male | Mixed | 60+ | — | — | NA | — | — | — | — | — | — |
| male | Mixed | Meta-Regression | — | — | NA | — | — | — | — | — | — |
| **male** | **Mixed** | **Overall** | **2.31 – 6.98** | **16** | **Mixed** | **756** | **99.6%** | **3679.2** | **<0.005** | **0.51** | **Random** |
| male | White | 18+ | — | — | NA | — | — | — | — | — | — |
| male | White | 20-30 | — | — | NA | — | — | — | — | — | — |
| male | White | 30-40 | — | — | NA | — | — | — | — | — | — |
| male | White | 40-50 | 2.39 – 5.28 | 1 | Observational Cohort | 6 | — | — | — | — | Single study |
| male | White | 50-60 | 0.20 – 8.71 | 1 | Observational Cohort | 79 | — | — | — | — | Single study |
| male | White | 60+ | 2.41 – 4.92 | 3 | Observational Cohort | 329 | 97.2% | 72.2 | <0.005 | 0.24 | Random |
| male | White | Meta-Regression | — | — | NA | — | — | — | — | — | — |
| **male** | **White** | **Overall** | **2.73 – 4.95** | **5** | **Observational Cohort** | **414** | **96.4%** | **109.9** | **<0.005** | **<0.05** | **Random** |
| **female** | **All races** | **18+** | **3.45 – 4.61** | **9** | **Mixed** | **710** | **66.3%** | **23.7** | **<0.005** | **<0.05** | **Random** |
| **female** | **All races** | **20-30** | **3.01 – 4.07** | **1** | **Observational Cohort** | **25** | **—** | **—** | **—** | **—** | **Single study** |
| **female** | **All races** | **30-40** | **3.29 – 4.39** | **1** | **Observational Cohort** | **25** | **—** | **—** | **—** | **—** | **Single study** |
| **female** | **All races** | **40-50** | **3.35 – 5.47** | **3** | **Observational Cohort** | **578** | **90.6%** | **21.2** | **<0.005** | **0.85** | **Random** |
| **female** | **All races** | **50-60** | **2.49 – 5.95** | **3** | **Observational Cohort** | **2565** | **98.1%** | **106.5** | **<0.005** | **0.16** | **Random** |
| **female** | **All races** | **60+** | **1.03 – 5.30** | **5** | **Observational Cohort** | **496** | **99%** | **392.7** | **<0.005** | **0.13** | **Random** |
| **female** | **All races** | **Meta-Regression** | **3.67 – 3.67** | **9** | **Mixed** | **710** | **0%** | **6.6** | **0.25** | **<0.05** | **Multivariable** |
| **female** | **All races** | **Overall** | **2.39 – 5.39** | **22** | **Mixed** | **4399** | **99.1%** | **2452.1** | **<0.005** | **0.74** | **Random** |
| female | Black | 18+ | — | — | NA | — | — | — | — | — | — |
| female | Black | 20-30 | — | — | NA | — | — | — | — | — | — |
| female | Black | 30-40 | — | — | NA | — | — | — | — | — | — |
| female | Black | 40-50 | -1.06 – 8.23 | 1 | Observational Cohort | 29 | — | — | — | — | Single study |
| female | Black | 50-60 | -0.69 – 7.15 | 1 | Observational Cohort | 131 | — | — | — | — | Single study |
| female | Black | 60+ | -0.72 – 6.40 | 2 | Observational Cohort | 144 | 69.7% | 3.3 | 0.07 | — | Fixed |
| female | Black | Meta-Regression | — | — | NA | — | — | — | — | — | — |
| **female** | **Black** | **Overall** | **-0.74 – 6.83** | **4** | **Observational Cohort** | **304** | **61.1%** | **7.7** | **0.05** | **0.83** | **Fixed** |
| female | Mixed | 18+ | 3.45 – 4.61 | 9 | Mixed | 710 | 66.3% | 23.7 | <0.005 | <0.05 | Random |
| female | Mixed | 20-30 | 3.01 – 4.07 | 1 | Observational Cohort | 25 | — | — | — | — | Single study |
| female | Mixed | 30-40 | 3.29 – 4.39 | 1 | Observational Cohort | 25 | — | — | — | — | Single study |
| female | Mixed | 40-50 | 3.68 – 5.14 | 1 | Observational Cohort | 25 | — | — | — | — | Single study |
| female | Mixed | 50-60 | 3.28 – 5.56 | 1 | Observational Cohort | 25 | — | — | — | — | Single study |
| female | Mixed | 60+ | 1.29 – 2.31 | 1 | Observational Cohort | 25 | — | — | — | — | Single study |
| female | Mixed | Meta-Regression | — | — | NA | — | — | — | — | — | — |
| **female** | **Mixed** | **Overall** | **2.50 – 5.31** | **14** | **Mixed** | **835** | **99%** | **1358.4** | **<0.005** | **0.18** | **Random** |
| female | White | 18+ | — | — | NA | — | — | — | — | — | — |
| female | White | 20-30 | — | — | NA | — | — | — | — | — | — |
| female | White | 30-40 | — | — | NA | — | — | — | — | — | — |
| female | White | 40-50 | 0.35 – 9.49 | 1 | Observational Cohort | 524 | — | — | — | — | Single study |
| female | White | 50-60 | 0.43 – 9.54 | 1 | Observational Cohort | 2409 | — | — | — | — | Single study |
| female | White | 60+ | 3.18 – 5.27 | 2 | Observational Cohort | 327 | 88.9% | 9 | <0.005 | — | Random |
| female | White | Meta-Regression | — | — | NA | — | — | — | — | — | — |
| **female** | **White** | **Overall** | **3.65 – 5.57** | **4** | **Observational Cohort** | **3260** | **90.3%** | **31.1** | **<0.005** | **0.11** | **Random** |
| *I² represents heterogeneity; Q test assesses between-study variation; Egger's p = linear regression test for funnel plot asymmetry; — indicates insufficient data* | | | | | | | | | | | |

###### 6.2.2.4 Wald Tests

calcium_ma_3rd_analysis$wald_tests$sex_comparison

Table 1: **Sex Comparison (Overall)**

| Comparison | % Shift | P (Wald Test) | Mean Diff (95% CI) |
| --- | --- | --- | --- |
| Male vs Female (Overall) | +14.5% | 0.675 | 0.56 [-2.08, 3.21] |
| *Mean difference = Group 1 − Group 2 in component units (mmol/24h, L/24h, or pH); % shift = difference relative to Group 2.* | | | |

calcium_ma_3rd_analysis$wald_tests$race_comparisons

Table 1: **race Comparisons Within Sex Groups**

| Sex Group | Comparison | % Shift | P (Wald Test) | Mean Diff (95% CI) |
| --- | --- | --- | --- | --- |
| male | black vs mixed | — | — | — |
| male | black vs white | — | — | — |
| male | mixed vs white | +20.8% | 0.544 | 0.80 [-1.79, 3.39] |
| female | black vs mixed | -22.0% | 0.677 | -0.86 [-4.90, 3.18] |
| female | black vs white | -34.0% | 0.432 | -1.57 [-5.47, 2.34] |
| female | mixed vs white | -15.4% | 0.415 | -0.71 [-2.41, 1.00] |
| *Mean difference = Group 1 − Group 2 in component units (mmol/24h, L/24h, or pH); % shift = difference relative to Group 2.* | | | | |

calcium_ma_3rd_analysis$wald_tests$age_band_comparisons

Table 1: **Age Band Comparisons Within Sex Groups**

| Sex Group | Comparison | % Shift | P (Wald Test) | Mean Diff (95% CI) |
| --- | --- | --- | --- | --- |
| main meta-analysis | 18+ vs 20-30 | +6.4% | 0.890 | 0.26 [-3.41, 3.92] |
| main meta-analysis | 18+ vs 30-40 | -7.7% | 0.818 | -0.36 [-3.38, 2.67] |
| main meta-analysis | 18+ vs 40-50 | +1.9% | 0.952 | 0.08 [-2.52, 2.68] |
| main meta-analysis | 18+ vs 50-60 | -0.6% | 0.987 | -0.02 [-2.80, 2.75] |
| main meta-analysis | 18+ vs 60+ | +26.8% | 0.567 | 0.90 [-2.18, 3.98] |
| main meta-analysis | 20-30 vs 30-40 | -13.3% | 0.707 | -0.61 [-3.81, 2.58] |
| main meta-analysis | 20-30 vs 40-50 | -4.2% | 0.901 | -0.18 [-2.97, 2.61] |
| main meta-analysis | 20-30 vs 50-60 | -6.6% | 0.852 | -0.28 [-3.24, 2.67] |
| main meta-analysis | 20-30 vs 60+ | +19.2% | 0.698 | 0.64 [-2.60, 3.89] |
| main meta-analysis | 30-40 vs 40-50 | +10.4% | 0.649 | 0.44 [-1.44, 2.31] |
| main meta-analysis | 30-40 vs 50-60 | +7.8% | 0.758 | 0.33 [-1.78, 2.45] |
| main meta-analysis | 30-40 vs 60+ | +37.4% | 0.325 | 1.26 [-1.25, 3.76] |
| main meta-analysis | 40-50 vs 50-60 | -2.4% | 0.887 | -0.10 [-1.54, 1.33] |
| main meta-analysis | 40-50 vs 60+ | +24.4% | 0.413 | 0.82 [-1.14, 2.78] |
| main meta-analysis | 50-60 vs 60+ | +27.5% | 0.409 | 0.92 [-1.27, 3.12] |
| male | 18+ vs 20-30 | +14.5% | 0.781 | 0.60 [-3.63, 4.83] |
| male | 18+ vs 30-40 | -8.7% | 0.761 | -0.45 [-3.35, 2.45] |
| male | 18+ vs 40-50 | +19.6% | 0.594 | 0.78 [-2.09, 3.65] |
| male | 18+ vs 50-60 | +10.8% | 0.771 | 0.46 [-2.66, 3.59] |
| male | 18+ vs 60+ | +29.7% | 0.481 | 1.09 [-1.94, 4.11] |
| male | 20-30 vs 30-40 | -20.2% | 0.538 | -1.05 [-4.39, 2.29] |
| male | 20-30 vs 40-50 | +4.5% | 0.916 | 0.18 [-3.14, 3.50] |
| male | 20-30 vs 50-60 | -3.2% | 0.940 | -0.14 [-3.68, 3.41] |
| male | 20-30 vs 60+ | +13.3% | 0.783 | 0.49 [-2.97, 3.94] |
| male | 30-40 vs 40-50 | +31.0% | <0.05 | 1.23 [0.01, 2.46] |
| male | 30-40 vs 50-60 | +21.3% | 0.303 | 0.91 [-0.83, 2.65] |
| male | 30-40 vs 60+ | +41.9% | 0.052 | 1.54 [-0.01, 3.09] |
| male | 40-50 vs 50-60 | -7.4% | 0.716 | -0.32 [-2.01, 1.38] |
| male | 40-50 vs 60+ | +8.4% | 0.689 | 0.31 [-1.20, 1.81] |
| male | 50-60 vs 60+ | +17.0% | 0.531 | 0.62 [-1.32, 2.57] |
| female | 18+ vs 20-30 | +13.9% | 0.218 | 0.49 [-0.29, 1.28] |
| female | 18+ vs 30-40 | +5.0% | 0.636 | 0.19 [-0.61, 0.99] |
| female | 18+ vs 40-50 | -8.5% | 0.544 | -0.37 [-1.58, 0.83] |
| female | 18+ vs 50-60 | -4.4% | 0.841 | -0.19 [-2.01, 1.64] |
| female | 18+ vs 60+ | +27.5% | 0.441 | 0.87 [-1.34, 3.08] |
| female | 20-30 vs 30-40 | -7.8% | 0.441 | -0.30 [-1.06, 0.46] |
| female | 20-30 vs 40-50 | -19.7% | 0.152 | -0.87 [-2.05, 0.32] |
| female | 20-30 vs 50-60 | -16.1% | 0.462 | -0.68 [-2.49, 1.13] |
| female | 20-30 vs 60+ | +11.9% | 0.737 | 0.38 [-1.82, 2.58] |
| female | 30-40 vs 40-50 | -12.9% | 0.352 | -0.57 [-1.76, 0.63] |
| female | 30-40 vs 50-60 | -9.0% | 0.682 | -0.38 [-2.20, 1.44] |
| female | 30-40 vs 60+ | +21.4% | 0.548 | 0.68 [-1.53, 2.88] |
| female | 40-50 vs 50-60 | +4.4% | 0.856 | 0.19 [-1.84, 2.22] |
| female | 40-50 vs 60+ | +39.3% | 0.307 | 1.24 [-1.14, 3.63] |
| female | 50-60 vs 60+ | +33.4% | 0.452 | 1.06 [-1.69, 3.81] |
| *Mean difference = Group 1 − Group 2 in component units (mmol/24h, L/24h, or pH); % shift = difference relative to Group 2.* | | | | |

calcium_ma_3rd_analysis$wald_tests$sex_within_age_bands

Table 1: **Sex Comparisons Within Age Bands**

| Age Band | % Shift | P (Wald Test) | Mean Diff (95% CI) |
| --- | --- | --- | --- |
| 18+ | +17.8% | 0.616 | 0.72 [-2.09, 3.53] |
| 20-30 | +17.2% | 0.714 | 0.61 [-2.65, 3.87] |
| 30-40 | +35.5% | <0.05 | 1.36 [0.30, 2.42] |
| 40-50 | -9.9% | 0.525 | -0.44 [-1.78, 0.91] |
| 50-60 | +1.6% | 0.954 | 0.07 [-2.21, 2.35] |
| 60+ | +15.8% | 0.692 | 0.50 [-1.98, 2.98] |
| *Mean difference = Group 1 − Group 2 in component units (mmol/24h, L/24h, or pH); % shift = difference relative to Group 2.* | | | |

#### 6.3 Urate

##### 6.3.1 Comparison by Age Band

###### 6.3.1.1 Plot

```
urate_ma_2nd_analysis <- sex_age_subdivision_ma("urate")
## Not enough studies for multivariable meta-regression (k = 5, need > 6). Running univariable models instead.
urate_ma_2nd_analysis$combined_plot
## `height` was translated to `width`.
## `height` was translated to `width`.
## `height` was translated to `width`.
```

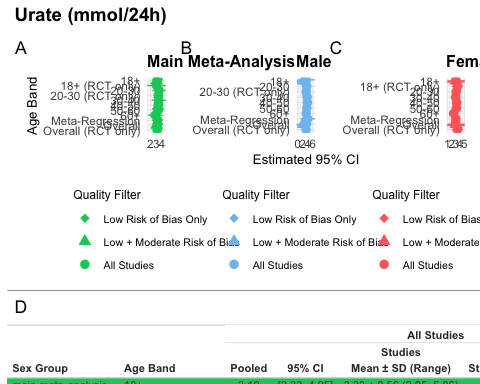

###### 6.3.1.2 Summary Table

urate_ma_2nd_analysis$combined_summary_table

Table 1: **Combined Meta-Analysis Results**

24-Hour Urine Urate (mmol/24h)

|  |  | **All Studies** | | | | | | **Low Risk of Bias Only** | | | | | | **Low + Moderate RoB** | | | | | |
| --- | --- | --- | --- | --- | --- | --- | --- | --- | --- | --- | --- | --- | --- | --- | --- | --- | --- | --- | --- |
| Sex Group | Age Band | Pooled | 95% CI | Studies  Mean ± SD (Range) | Studies, n | Q p-value | Egger's p | Pooled | 95% CI | Studies  Mean ± SD (Range) | Studies, n | Q p-value | Egger's p | Pooled | 95% CI | Studies  Mean ± SD (Range) | Studies, n | Q p-value | Egger's p |
| main meta-analysis | 18+ | 3.19 | [2.33, 4.05] | 3.23 ± 0.56 (2.05–5.86) | 64 | <0.001 | <0.001 | 3.02 | [2.50, 3.54] | 3.06 ± 0.31 (2.09–3.78) | 28 | <0.001 | 0.09 | 3.03 | [2.50, 3.57] | 3.09 ± 0.36 (2.09–4.03) | 33 | <0.001 | 0.06 |
| **main meta-analysis** | **18+ (RCT only)** | **3.01** | **[1.54, 4.48]** | **3.10 ± 0.30 (2.70–3.49)** | **7** | **0.06** | **0.31** | **—** | **—** | **—** | **—** | **—** | **—** | **—** | **—** | **—** | **—** | **—** | **—** |
| main meta-analysis | 20-30 | 3.45 | [2.43, 4.47] | 3.44 ± 0.52 (3.04–4.15) | 4 | <0.001 | 0.91 | — | — | — | — | — | — | — | — | — | — | — | — |
| **main meta-analysis** | **20-30 (RCT only)** | **3.31** | **[2.72, 3.91]** | **3.29 ± 0.33 (3.06–3.52)** | **2** | **<0.01** | **—** | **—** | **—** | **—** | **—** | **—** | **—** | **—** | **—** | **—** | **—** | **—** | **—** |
| main meta-analysis | 30-40 | 3.41 | [2.29, 4.53] | 3.45 ± 0.65 (2.90–4.49) | 5 | <0.001 | 0.80 | — | — | — | — | — | — | — | — | — | — | — | — |
| main meta-analysis | 40-50 | 3.32 | [2.77, 3.88] | 3.36 ± 0.33 (3.02–3.86) | 7 | <0.001 | 0.74 | — | — | — | — | — | — | 3.29 | [2.96, 3.62] | — (3.29–3.29) | 1 | — | — |
| main meta-analysis | 50-60 | 3.29 | [2.57, 4.00] | 3.30 ± 0.41 (2.94–3.99) | 5 | <0.001 | 0.95 | — | — | — | — | — | — | — | — | — | — | — | — |
| main meta-analysis | 60+ | 2.80 | [1.67, 3.92] | 2.80 ± 0.58 (1.94–3.71) | 8 | <0.001 | 0.08 | — | — | — | — | — | — | — | — | — | — | — | — |
| **main meta-analysis** | **Meta-Regression** | **3.19** | **[2.35, 4.02]** | **—** | **51** | **<0.001** | **<0.05** | **—** | **—** | **—** | **—** | **—** | **—** | **—** | **—** | **—** | **—** | **—** | **—** |
| **main meta-analysis** | **Overall** | **3.20** | **[2.29, 4.11]** | **3.22 ± 0.55 (1.94–5.86)** | **93** | **<0.001** | **0.44** | **3.02** | **[2.50, 3.54]** | **3.06 ± 0.31 (2.09–3.78)** | **28** | **<0.001** | **0.09** | **3.04** | **[2.51, 3.58]** | **3.10 ± 0.36 (2.09–4.03)** | **34** | **<0.001** | **0.85** |
| **main meta-analysis** | **Overall (RCT only)** | **3.13** | **[2.61, 3.64]** | **3.14 ± 0.30 (2.70–3.52)** | **9** | **<0.001** | **0.06** | **—** | **—** | **—** | **—** | **—** | **—** | **—** | **—** | **—** | **—** | **—** | **—** |
| male | 18+ | 3.37 | [0.09, 6.65] | 3.39 ± 1.70 (2.09–5.86) | 4 | <0.001 | 0.36 | 2.09 | [-0.31, 4.49] | — (2.09–2.09) | 1 | — | — | 2.09 | [-0.31, 4.49] | — (2.09–2.09) | 1 | — | — |
| **male** | **18+ (RCT only)** | **—** | **—** | **—** | **—** | **—** | **—** | **—** | **—** | **—** | **—** | **—** | **—** | **—** | **—** | **—** | **—** | **—** | **—** |
| male | 20-30 | 3.59 | [2.54, 4.64] | 3.58 ± 0.55 (3.06–4.15) | 3 | <0.001 | 0.36 | — | — | — | — | — | — | — | — | — | — | — | — |
| **male** | **20-30 (RCT only)** | **3.31** | **[2.72, 3.91]** | **3.29 ± 0.33 (3.06–3.52)** | **2** | **<0.01** | **—** | **—** | **—** | **—** | **—** | **—** | **—** | **—** | **—** | **—** | **—** | **—** | **—** |
| male | 30-40 | 3.75 | [2.76, 4.75] | 3.82 ± 0.59 (3.37–4.49) | 3 | <0.001 | 0.07 | — | — | — | — | — | — | — | — | — | — | — | — |
| male | 40-50 | 3.86 | [3.12, 4.59] | 3.81 ± 0.07 (3.76–3.86) | 2 | 0.82 | — | — | — | — | — | — | — | — | — | — | — | — | — |
| male | 50-60 | 3.61 | [2.60, 4.61] | 3.62 ± 0.53 (3.25–3.99) | 2 | <0.001 | — | — | — | — | — | — | — | — | — | — | — | — | — |
| male | 60+ | 3.36 | [2.42, 4.29] | 3.36 ± 0.48 (2.82–3.71) | 3 | <0.001 | 0.41 | — | — | — | — | — | — | — | — | — | — | — | — |
| **male** | **Meta-Regression** | **3.96** | **[1.31, 6.61]** | **—** | **5** | **<0.001** | **0.94** | **—** | **—** | **—** | **—** | **—** | **—** | **—** | **—** | **—** | **—** | **—** | **—** |
| **male** | **Overall** | **3.55** | **[1.99, 5.11]** | **3.57 ± 0.84 (2.09–5.86)** | **17** | **<0.001** | **0.80** | **2.09** | **[-0.31, 4.49]** | **— (2.09–2.09)** | **1** | **—** | **—** | **2.09** | **[-0.31, 4.49]** | **— (2.09–2.09)** | **1** | **—** | **—** |
| **male** | **Overall (RCT only)** | **3.31** | **[2.72, 3.91]** | **3.29 ± 0.33 (3.06–3.52)** | **2** | **<0.01** | **—** | **—** | **—** | **—** | **—** | **—** | **—** | **—** | **—** | **—** | **—** | **—** | **—** |
| female | 18+ | 2.95 | [2.73, 3.17] | 2.96 ± 0.21 (2.71–3.34) | 7 | <0.05 | 0.86 | 2.96 | [0.90, 5.02] | 2.96 ± 0.00 (2.96–2.96) | 2 | 1.00 | — | 3.00 | [1.02, 4.98] | 3.09 ± 0.22 (2.96–3.34) | 3 | 0.11 | <0.001 |
| **female** | **18+ (RCT only)** | **3.34** | **[2.18, 4.49]** | **— (3.34–3.34)** | **1** | **—** | **—** | **—** | **—** | **—** | **—** | **—** | **—** | **—** | **—** | **—** | **—** | **—** | **—** |
| female | 20-30 | 3.04 | [2.71, 3.37] | — (3.04–3.04) | 1 | — | — | — | — | — | — | — | — | — | — | — | — | — | — |
| **female** | **20-30 (RCT only)** | **—** | **—** | **—** | **—** | **—** | **—** | **—** | **—** | **—** | **—** | **—** | **—** | **—** | **—** | **—** | **—** | **—** | **—** |
| female | 30-40 | 2.90 | [2.47, 3.33] | — (2.90–2.90) | 1 | — | — | — | — | — | — | — | — | — | — | — | — | — | — |
| female | 40-50 | 3.15 | [2.88, 3.43] | 3.13 ± 0.15 (3.02–3.30) | 3 | <0.001 | 0.65 | — | — | — | — | — | — | — | — | — | — | — | — |
| female | 50-60 | 3.10 | [2.82, 3.38] | 3.09 ± 0.15 (2.94–3.24) | 3 | <0.001 | 0.18 | — | — | — | — | — | — | — | — | — | — | — | — |
| female | 60+ | 2.46 | [1.84, 3.08] | 2.47 ± 0.32 (1.94–2.74) | 5 | <0.001 | 0.10 | — | — | — | — | — | — | — | — | — | — | — | — |
| **female** | **Meta-Regression** | **2.96** | **[2.80, 3.12]** | **—** | **7** | **0.16** | **0.86** | **—** | **—** | **—** | **—** | **—** | **—** | **—** | **—** | **—** | **—** | **—** | **—** |
| **female** | **Overall** | **2.88** | **[2.25, 3.51]** | **2.88 ± 0.33 (1.94–3.34)** | **20** | **<0.001** | **0.30** | **2.96** | **[0.90, 5.02]** | **2.96 ± 0.00 (2.96–2.96)** | **2** | **1.00** | **—** | **3.00** | **[1.02, 4.98]** | **3.09 ± 0.22 (2.96–3.34)** | **3** | **0.11** | **<0.001** |
| **female** | **Overall (RCT only)** | **3.34** | **[2.18, 4.49]** | **— (3.34–3.34)** | **1** | **—** | **—** | **—** | **—** | **—** | **—** | **—** | **—** | **—** | **—** | **—** | **—** | **—** | **—** |
| *Studies, n = number of studies; Q p-value = Cochran's Q heterogeneity test; Egger's p = linear regression test for funnel plot asymmetry; RoB = Risk of Bias* | | | | | | | | | | | | | | | | | | | |

###### 6.3.1.3 Wald Tests

urate_ma_2nd_analysis$wald_tests$sex_comparison

Table 1: **Sex Comparison (Overall)**

| Comparison | % Shift | P (Wald Test) | Mean Diff (95% CI) |
| --- | --- | --- | --- |
| Male vs Female (Overall) | +23.2% | 0.436 | 0.67 [-1.01, 2.35] |
| *Mean difference = Group 1 − Group 2 in component units (mmol/24h, L/24h, or pH); % shift = difference relative to Group 2.* | | | |

urate_ma_2nd_analysis$wald_tests$age_band_comparisons

Table 1: **Age Band Comparisons Within Sex Groups**

| Sex Group | Comparison | % Shift | P (Wald Test) | Mean Diff (95% CI) |
| --- | --- | --- | --- | --- |
| main meta-analysis | 18+ vs 20-30 | -7.5% | 0.702 | -0.26 [-1.59, 1.07] |
| main meta-analysis | 18+ vs 30-40 | -6.4% | 0.762 | -0.22 [-1.63, 1.19] |
| main meta-analysis | 18+ vs 40-50 | -4.0% | 0.799 | -0.13 [-1.16, 0.89] |
| main meta-analysis | 18+ vs 50-60 | -2.9% | 0.867 | -0.10 [-1.22, 1.03] |
| main meta-analysis | 18+ vs 60+ | +14.0% | 0.587 | 0.39 [-1.02, 1.81] |
| main meta-analysis | 20-30 vs 30-40 | +1.2% | 0.957 | 0.04 [-1.47, 1.55] |
| main meta-analysis | 20-30 vs 40-50 | +3.8% | 0.830 | 0.13 [-1.03, 1.28] |
| main meta-analysis | 20-30 vs 50-60 | +5.0% | 0.795 | 0.16 [-1.08, 1.41] |
| main meta-analysis | 20-30 vs 60+ | +23.3% | 0.399 | 0.65 [-0.86, 2.17] |
| main meta-analysis | 30-40 vs 40-50 | +2.6% | 0.893 | 0.09 [-1.16, 1.33] |
| main meta-analysis | 30-40 vs 50-60 | +3.7% | 0.856 | 0.12 [-1.21, 1.45] |
| main meta-analysis | 30-40 vs 60+ | +21.8% | 0.451 | 0.61 [-0.98, 2.20] |
| main meta-analysis | 40-50 vs 50-60 | +1.1% | 0.935 | 0.04 [-0.87, 0.95] |
| main meta-analysis | 40-50 vs 60+ | +18.8% | 0.412 | 0.53 [-0.73, 1.78] |
| main meta-analysis | 50-60 vs 60+ | +17.4% | 0.474 | 0.49 [-0.85, 1.82] |
| male | 18+ vs 20-30 | -6.1% | 0.901 | -0.22 [-3.66, 3.23] |
| male | 18+ vs 30-40 | -10.2% | 0.828 | -0.38 [-3.81, 3.05] |
| male | 18+ vs 40-50 | -12.6% | 0.777 | -0.49 [-3.85, 2.88] |
| male | 18+ vs 50-60 | -6.6% | 0.892 | -0.24 [-3.67, 3.19] |
| male | 18+ vs 60+ | +0.4% | 0.993 | 0.01 [-3.40, 3.43] |
| male | 20-30 vs 30-40 | -4.3% | 0.826 | -0.16 [-1.61, 1.28] |
| male | 20-30 vs 40-50 | -6.9% | 0.682 | -0.27 [-1.55, 1.01] |
| male | 20-30 vs 50-60 | -0.5% | 0.980 | -0.02 [-1.47, 1.43] |
| male | 20-30 vs 60+ | +7.0% | 0.744 | 0.23 [-1.17, 1.64] |
| male | 30-40 vs 40-50 | -2.7% | 0.867 | -0.11 [-1.34, 1.13] |
| male | 30-40 vs 50-60 | +4.0% | 0.842 | 0.14 [-1.27, 1.56] |
| male | 30-40 vs 60+ | +11.8% | 0.569 | 0.40 [-0.97, 1.76] |
| male | 40-50 vs 50-60 | +6.9% | 0.694 | 0.25 [-0.99, 1.49] |
| male | 40-50 vs 60+ | +14.9% | 0.407 | 0.50 [-0.69, 1.69] |
| male | 50-60 vs 60+ | +7.5% | 0.718 | 0.25 [-1.12, 1.62] |
| female | 18+ vs 20-30 | -3.0% | 0.659 | -0.09 [-0.49, 0.31] |
| female | 18+ vs 30-40 | +1.7% | 0.840 | 0.05 [-0.43, 0.53] |
| female | 18+ vs 40-50 | -6.4% | 0.261 | -0.20 [-0.55, 0.15] |
| female | 18+ vs 50-60 | -4.9% | 0.399 | -0.15 [-0.51, 0.20] |
| female | 18+ vs 60+ | +20.0% | 0.145 | 0.49 [-0.17, 1.15] |
| female | 20-30 vs 30-40 | +4.8% | 0.615 | 0.14 [-0.40, 0.68] |
| female | 20-30 vs 40-50 | -3.6% | 0.610 | -0.11 [-0.54, 0.32] |
| female | 20-30 vs 50-60 | -2.0% | 0.775 | -0.06 [-0.50, 0.37] |
| female | 20-30 vs 60+ | +23.6% | 0.107 | 0.58 [-0.13, 1.29] |
| female | 30-40 vs 40-50 | -8.0% | 0.333 | -0.25 [-0.76, 0.26] |
| female | 30-40 vs 50-60 | -6.6% | 0.438 | -0.20 [-0.72, 0.31] |
| female | 30-40 vs 60+ | +17.9% | 0.254 | 0.44 [-0.32, 1.20] |
| female | 40-50 vs 50-60 | +1.6% | 0.807 | 0.05 [-0.34, 0.44] |
| female | 40-50 vs 60+ | +28.2% | <0.05 | 0.69 [0.01, 1.37] |
| female | 50-60 vs 60+ | +26.2% | 0.064 | 0.64 [-0.04, 1.33] |
| *Mean difference = Group 1 − Group 2 in component units (mmol/24h, L/24h, or pH); % shift = difference relative to Group 2.* | | | | |

urate_ma_2nd_analysis$wald_tests$sex_within_age_bands

Table 1: **Sex Comparisons Within Age Bands**

| Age Band | % Shift | P (Wald Test) | Mean Diff (95% CI) |
| --- | --- | --- | --- |
| 18+ | +14.3% | 0.802 | 0.42 [-2.87, 3.71] |
| 20-30 | +18.1% | 0.327 | 0.55 [-0.55, 1.65] |
| 30-40 | +29.4% | 0.124 | 0.85 [-0.23, 1.94] |
| 40-50 | +22.4% | 0.077 | 0.71 [-0.08, 1.49] |
| 50-60 | +16.3% | 0.342 | 0.50 [-0.54, 1.55] |
| 60+ | +36.5% | 0.117 | 0.90 [-0.22, 2.02] |
| *Mean difference = Group 1 − Group 2 in component units (mmol/24h, L/24h, or pH); % shift = difference relative to Group 2.* | | | |

###### 6.3.1.4 Meta-Regression (plain language)

format_meta_reg_plain("urate", sex_group = "all")

Message

No meta-regression result for urate / all

###### 6.3.1.5 Assay Subgroup Analysis

assay_subgroup_table("urate")

Table 1: **Assay subgroup analysis — Urate**

*Per-assay pooled estimates; categories with k < 3 listed but not pooled.*

| Assay category | Studies, n | Participants | 95% CI | I² | Q p-value | Egger's p | Note |
| --- | --- | --- | --- | --- | --- | --- | --- |
| Unreported | 62 | 6,638 | 3.18 [3.05, 3.31] | 98% | <0.001 | 0.866 |  |
| Autoanalyser | 22 | 2,577 | 3.20 [2.98, 3.42] | 90% | <0.001 | 0.030 |  |
| Uricase / urate oxidase | 9 | 309 | 3.36 [3.13, 3.60] | 83% | <0.001 | 0.696 |  |
| Colourimetric enzymatic | 1 | 20 | — | — | — | — | k = 1 < min_k = 3 (skipped) |

###### 6.3.1.6 Preservative Sensitivity (Litholink / acidified samples)

```
preservative_sensitivity_ma("urate")
## # A tibble: 3 × 6
##   group               n_studies n_participants pooled_estimate lower_95 upper_95
##   <chr>                   <int>          <int>           <dbl>    <dbl>    <dbl>
## 1 All studies                93           9544            3.20     3.10     3.30
## 2 Excluding preserva…        91           9484            3.19     3.09     3.29
## 3 Preservative only           2             60            3.64     3.29     3.99
```

###### 6.3.1.7 Creatinine Completeness Sensitivity

creat_completeness_table("urate")

Table 1: **Creatinine-completeness sensitivity — Urate**

*Compares the all-studies pooled estimate against subsets defined by whether the study reported 24h urine creatinine. Studies that do not report creatinine cannot verify collection completeness; a substantial shift here flags possible bias from incomplete collections.*

| Subset | Studies, n | Participants | Range (95% CI) | Δ vs all-studies (%) | Flag |
| --- | --- | --- | --- | --- | --- |
| All studies | 93 | 9,544 | 3.20 [3.10, 3.30] | — | — |
| Creatinine reported only | 64 | 7,861 | 3.11 [3.00, 3.22] | -2.7% | OK |
| Creatinine NOT reported | 29 | 1,683 | 3.43 [3.20, 3.65] | +7.2% | OK |

##### 6.3.2 Comparison by Age / Sex / race

###### 6.3.2.1 Plot

```
urate_ma_3rd_analysis <- sex_age_race_subdivision_ma("urate",
                                                      urine_ma2,
                                                      overall_bias)
## Not enough studies for multivariable meta-regression (k = 5, need > 6). Running univariable models instead.
urate_ma_3rd_analysis$race_plot
## `height` was translated to `width`.
## `height` was translated to `width`.
## `height` was translated to `width`.
```

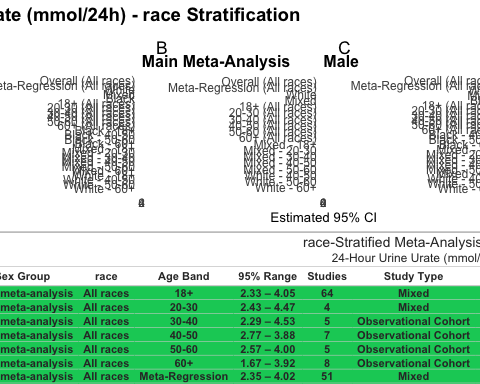

###### 6.3.2.2 Combined MA Results

urate_ma_3rd_analysis$combined_summary_table

Table 1: **Combined Meta-Analysis Results**

24-Hour Urine Urate (mmol/24h)

| Sex Group | race | Age Band | Pooled | 95% Range | Studies Mean SD Range | Studies | Study Type | Participants | I² | Cochran's Q | Q (p-value) | Egger's p | Model |
| --- | --- | --- | --- | --- | --- | --- | --- | --- | --- | --- | --- | --- | --- |
| main meta-analysis | All | 18+ | 3.19 | 2.33 – 4.05 | 3.23 ± 0.56 (2.05–5.86) | 64 | Mixed | 5104 | 89.7% | 613.7 | <0.005 | <0.001 | Random |
| main meta-analysis | All | 20-30 | 3.45 | 2.43 – 4.47 | 3.44 ± 0.52 (3.04–4.15) | 4 | Mixed | 96 | 99.4% | 512.7 | <0.005 | 0.91 | Random |
| main meta-analysis | All | 30-40 | 3.41 | 2.29 – 4.53 | 3.45 ± 0.65 (2.90–4.49) | 5 | Observational Cohort | 185 | 97.3% | 147.7 | <0.005 | 0.80 | Random |
| main meta-analysis | All | 40-50 | 3.32 | 2.77 – 3.88 | 3.36 ± 0.33 (3.02–3.86) | 7 | Observational Cohort | 665 | 93.8% | 97.3 | <0.005 | 0.74 | Random |
| main meta-analysis | All | 50-60 | 3.29 | 2.57 – 4.00 | 3.30 ± 0.41 (2.94–3.99) | 5 | Observational Cohort | 2669 | 89.8% | 39.1 | <0.005 | 0.95 | Random |
| main meta-analysis | All | 60+ | 2.80 | 1.67 – 3.92 | 2.80 ± 0.58 (1.94–3.71) | 8 | Observational Cohort | 825 | 98.8% | 601 | <0.005 | 0.08 | Random |
| **main meta-analysis** | **All** | **Meta-Regression** | **3.19** | **2.35 – 4.02** | **—** | **51** | **Mixed** | **4797** | **93.4%** | **256** | **<0.005** | **<0.05** | **Multivariable** |
| **main meta-analysis** | **All** | **Meta-Regression** | **3.19** | **2.35 – 4.02** | **—** | **51** | **Mixed** | **4797** | **93.4%** | **256** | **<0.005** | **<0.05** | **Multivariable** |
| **main meta-analysis** | **All** | **Overall** | **3.20** | **2.29 – 4.11** | **3.22 ± 0.55 (1.94–5.86)** | **93** | **Mixed** | **9544** | **97.6%** | **3759.5** | **<0.005** | **0.44** | **Random** |
| **main meta-analysis** | **All** | **Overall** | **3.20** | **2.29 – 4.11** | **3.22 ± 0.55 (1.94–5.86)** | **93** | **Mixed** | **9544** | **97.6%** | **3759.5** | **<0.005** | **0.44** | **Random** |
| main meta-analysis | black | 18+ | 3.35 | 1.31 – 5.39 | 3.35 ± 0.00 (3.35–3.35) | 3 | Observational Cohort | 51 | 0% | 0 | 1.00 | — | Fixed |
| main meta-analysis | black | 20-30 | — | NA – NA | — | NA | NA | NA | — | — | — | — | NA |
| main meta-analysis | black | 30-40 | — | NA – NA | — | NA | NA | NA | — | — | — | — | NA |
| main meta-analysis | black | 40-50 | 3.02 | 1.48 – 4.56 | — (3.02–3.02) | 1 | Observational Cohort | 29 | — | — | — | — | Single study |
| main meta-analysis | black | 50-60 | 2.94 | 1.03 – 4.85 | — (2.94–2.94) | 1 | Observational Cohort | 131 | — | — | — | — | Single study |
| main meta-analysis | black | 60+ | 2.62 | 0.83 – 4.41 | 2.61 ± 0.19 (2.47–2.74) | 2 | Observational Cohort | 144 | 68.7% | 3.2 | 0.07 | — | Fixed |
| **main meta-analysis** | **black** | **Meta-Regression** | **—** | **NA – NA** | **—** | **NA** | **NA** | **NA** | **—** | **—** | **—** | **—** | **NA** |
| **main meta-analysis** | **black** | **Overall** | **2.98** | **2.40 – 3.55** | **3.03 ± 0.34 (2.47–3.35)** | **7** | **Observational Cohort** | **355** | **77.4%** | **26.6** | **<0.005** | **0.14** | **Random** |
| main meta-analysis | mixed | 18+ | 3.15 | 2.25 – 4.06 | 3.19 ± 0.59 (2.05–5.86) | 55 | Mixed | 4888 | 90.6% | 573.2 | <0.005 | <0.001 | Random |
| main meta-analysis | mixed | 20-30 | 3.45 | 2.43 – 4.47 | 3.44 ± 0.52 (3.04–4.15) | 4 | Mixed | 96 | 99.4% | 512.7 | <0.005 | 0.91 | Random |
| main meta-analysis | mixed | 30-40 | 3.41 | 2.29 – 4.53 | 3.45 ± 0.65 (2.90–4.49) | 5 | Observational Cohort | 185 | 97.3% | 147.7 | <0.005 | 0.80 | Random |
| main meta-analysis | mixed | 40-50 | 3.36 | 2.70 – 4.02 | 3.36 ± 0.34 (3.07–3.86) | 4 | Observational Cohort | 106 | 96.8% | 92.6 | <0.005 | 0.59 | Random |
| main meta-analysis | mixed | 50-60 | 3.19 | 2.49 – 3.88 | 3.17 ± 0.12 (3.08–3.25) | 2 | Observational Cohort | 50 | 62.6% | 2.7 | 0.10 | — | Fixed |
| main meta-analysis | mixed | 60+ | 1.94 | 1.61 – 2.27 | — (1.94–1.94) | 1 | Observational Cohort | 25 | — | — | — | — | Single study |
| **main meta-analysis** | **mixed** | **Meta-Regression** | **—** | **NA – NA** | **—** | **NA** | **NA** | **NA** | **—** | **—** | **—** | **—** | **NA** |
| **main meta-analysis** | **mixed** | **Overall** | **3.19** | **2.23 – 4.14** | **3.21 ± 0.59 (1.94–5.86)** | **71** | **Mixed** | **5350** | **97.9%** | **3403.4** | **<0.005** | **0.46** | **Random** |
| main meta-analysis | white | 18+ | 3.47 | 1.00 – 5.94 | 3.51 ± 0.23 (3.30–3.72) | 6 | Observational Cohort | 165 | 0% | 4.4 | 0.49 | <0.001 | Fixed |
| main meta-analysis | white | 20-30 | — | NA – NA | — | NA | NA | NA | — | — | — | — | NA |
| main meta-analysis | white | 30-40 | — | NA – NA | — | NA | NA | NA | — | — | — | — | NA |
| main meta-analysis | white | 40-50 | 3.30 | 1.57 – 5.04 | 3.53 ± 0.32 (3.30–3.76) | 2 | Observational Cohort | 530 | 3.6% | 1 | 0.31 | — | Fixed |
| main meta-analysis | white | 50-60 | 3.60 | 2.58 – 4.62 | 3.62 ± 0.53 (3.24–3.99) | 2 | Observational Cohort | 2488 | 95.6% | 22.8 | <0.005 | — | Random |
| main meta-analysis | white | 60+ | 3.05 | 2.00 – 4.10 | 3.05 ± 0.55 (2.49–3.71) | 5 | Observational Cohort | 656 | 97.4% | 152.3 | <0.005 | 0.46 | Random |
| **main meta-analysis** | **white** | **Meta-Regression** | **—** | **NA – NA** | **—** | **NA** | **NA** | **NA** | **—** | **—** | **—** | **—** | **NA** |
| **main meta-analysis** | **white** | **Overall** | **3.34** | **2.51 – 4.16** | **3.37 ± 0.43 (2.49–3.99)** | **15** | **Observational Cohort** | **3839** | **95.1%** | **287.3** | **<0.005** | **0.75** | **Random** |
| male | All | 18+ | 3.37 | 0.09 – 6.65 | 3.39 ± 1.70 (2.09–5.86) | 4 | Observational Cohort | 182 | 98% | 147.6 | <0.005 | 0.36 | Random |
| male | All | 20-30 | 3.59 | 2.54 – 4.64 | 3.58 ± 0.55 (3.06–4.15) | 3 | Mixed | 71 | 98.3% | 115.3 | <0.005 | 0.36 | Random |
| male | All | 30-40 | 3.75 | 2.76 – 4.75 | 3.82 ± 0.59 (3.37–4.49) | 3 | Observational Cohort | 143 | 92.6% | 27.1 | <0.005 | 0.07 | Random |
| male | All | 40-50 | 3.86 | 3.12 – 4.59 | 3.81 ± 0.07 (3.76–3.86) | 2 | Observational Cohort | 31 | 0% | 0.1 | 0.82 | — | Fixed |
| male | All | 50-60 | 3.61 | 2.60 – 4.61 | 3.62 ± 0.53 (3.25–3.99) | 2 | Observational Cohort | 104 | 94.8% | 19.3 | <0.005 | — | Random |
| male | All | 60+ | 3.36 | 2.42 – 4.29 | 3.36 ± 0.48 (2.82–3.71) | 3 | Observational Cohort | 329 | 98% | 99.3 | <0.005 | 0.41 | Random |
| **male** | **All** | **Meta-Regression** | **3.96** | **1.31 – 6.61** | **—** | **5** | **Mixed** | **299** | **99.7%** | **200.8** | **<0.005** | **0.94** | **Univariable** |
| **male** | **All** | **Overall** | **3.55** | **1.99 – 5.11** | **3.57 ± 0.84 (2.09–5.86)** | **17** | **Mixed** | **860** | **98.6%** | **1118.5** | **<0.005** | **0.80** | **Random** |
| male | black | 18+ | — | NA – NA | — | NA | NA | NA | — | — | — | — | NA |
| male | black | 20-30 | — | NA – NA | — | NA | NA | NA | — | — | — | — | NA |
| male | black | 30-40 | — | NA – NA | — | NA | NA | NA | — | — | — | — | NA |
| male | black | 40-50 | — | NA – NA | — | NA | NA | NA | — | — | — | — | NA |
| male | black | 50-60 | — | NA – NA | — | NA | NA | NA | — | — | — | — | NA |
| male | black | 60+ | — | NA – NA | — | NA | NA | NA | — | — | — | — | NA |
| **male** | **black** | **Meta-Regression** | **—** | **NA – NA** | **—** | **NA** | **NA** | **NA** | **—** | **—** | **—** | **—** | **NA** |
| **male** | **black** | **Overall** | **—** | **NA – NA** | **—** | **NA** | **NA** | **NA** | **—** | **—** | **—** | **—** | **NA** |
| male | mixed | 18+ | 3.37 | 0.09 – 6.65 | 3.39 ± 1.70 (2.09–5.86) | 4 | Observational Cohort | 182 | 98% | 147.6 | <0.005 | 0.36 | Random |
| male | mixed | 20-30 | 3.59 | 2.54 – 4.64 | 3.58 ± 0.55 (3.06–4.15) | 3 | Mixed | 71 | 98.3% | 115.3 | <0.005 | 0.36 | Random |
| male | mixed | 30-40 | 3.75 | 2.76 – 4.75 | 3.82 ± 0.59 (3.37–4.49) | 3 | Observational Cohort | 143 | 92.6% | 27.1 | <0.005 | 0.07 | Random |
| male | mixed | 40-50 | 3.86 | 3.19 – 4.53 | — (3.86–3.86) | 1 | Observational Cohort | 25 | — | — | — | — | Single study |
| male | mixed | 50-60 | 3.25 | 2.62 – 3.88 | — (3.25–3.25) | 1 | Observational Cohort | 25 | — | — | — | — | Single study |
| male | mixed | 60+ | — | NA – NA | — | NA | NA | NA | — | — | — | — | NA |
| **male** | **mixed** | **Meta-Regression** | **—** | **NA – NA** | **—** | **NA** | **NA** | **NA** | **—** | **—** | **—** | **—** | **NA** |
| **male** | **mixed** | **Overall** | **3.55** | **1.73 – 5.37** | **3.57 ± 0.98 (2.09–5.86)** | **12** | **Mixed** | **446** | **98.8%** | **936.6** | **<0.005** | **0.84** | **Random** |
| male | white | 18+ | — | NA – NA | — | NA | NA | NA | — | — | — | — | NA |
| male | white | 20-30 | — | NA – NA | — | NA | NA | NA | — | — | — | — | NA |
| male | white | 30-40 | — | NA – NA | — | NA | NA | NA | — | — | — | — | NA |
| male | white | 40-50 | 3.76 | 1.61 – 5.90 | — (3.76–3.76) | 1 | Observational Cohort | 6 | — | — | — | — | Single study |
| male | white | 50-60 | 3.99 | 1.27 – 6.72 | — (3.99–3.99) | 1 | Observational Cohort | 79 | — | — | — | — | Single study |
| male | white | 60+ | 3.36 | 2.42 – 4.29 | 3.36 ± 0.48 (2.82–3.71) | 3 | Observational Cohort | 329 | 98% | 99.3 | <0.005 | 0.41 | Random |
| **male** | **white** | **Meta-Regression** | **—** | **NA – NA** | **—** | **NA** | **NA** | **NA** | **—** | **—** | **—** | **—** | **NA** |
| **male** | **white** | **Overall** | **3.54** | **2.64 – 4.44** | **3.57 ± 0.45 (2.82–3.99)** | **5** | **Observational Cohort** | **414** | **97%** | **132.8** | **<0.005** | **0.19** | **Random** |
| female | All | 18+ | 2.95 | 2.73 – 3.17 | 2.96 ± 0.21 (2.71–3.34) | 7 | Mixed | 488 | 53.1% | 12.8 | <0.05 | 0.86 | Random |
| female | All | 20-30 | 3.04 | 2.71 – 3.37 | — (3.04–3.04) | 1 | Observational Cohort | 25 | — | — | — | — | Single study |
| female | All | 30-40 | 2.90 | 2.47 – 3.33 | — (2.90–2.90) | 1 | Observational Cohort | 25 | — | — | — | — | Single study |
| female | All | 40-50 | 3.15 | 2.88 – 3.43 | 3.13 ± 0.15 (3.02–3.30) | 3 | Observational Cohort | 578 | 87.4% | 15.8 | <0.005 | 0.65 | Random |
| female | All | 50-60 | 3.10 | 2.82 – 3.38 | 3.09 ± 0.15 (2.94–3.24) | 3 | Observational Cohort | 2565 | 86.7% | 15 | <0.005 | 0.18 | Random |
| female | All | 60+ | 2.46 | 1.84 – 3.08 | 2.47 ± 0.32 (1.94–2.74) | 5 | Observational Cohort | 496 | 97.6% | 163.7 | <0.005 | 0.10 | Random |
| **female** | **All** | **Meta-Regression** | **2.96** | **2.80 – 3.12** | **—** | **7** | **Mixed** | **488** | **35.9%** | **6.6** | **0.16** | **0.86** | **Multivariable** |
| **female** | **All** | **Overall** | **2.88** | **2.25 – 3.51** | **2.88 ± 0.33 (1.94–3.34)** | **20** | **Mixed** | **4177** | **98.6%** | **1324.3** | **<0.005** | **0.30** | **Random** |
| female | black | 18+ | — | NA – NA | — | NA | NA | NA | — | — | — | — | NA |
| female | black | 20-30 | — | NA – NA | — | NA | NA | NA | — | — | — | — | NA |
| female | black | 30-40 | — | NA – NA | — | NA | NA | NA | — | — | — | — | NA |
| female | black | 40-50 | 3.02 | 1.48 – 4.56 | — (3.02–3.02) | 1 | Observational Cohort | 29 | — | — | — | — | Single study |
| female | black | 50-60 | 2.94 | 1.03 – 4.85 | — (2.94–2.94) | 1 | Observational Cohort | 131 | — | — | — | — | Single study |
| female | black | 60+ | 2.62 | 0.83 – 4.41 | 2.61 ± 0.19 (2.47–2.74) | 2 | Observational Cohort | 144 | 68.7% | 3.2 | 0.07 | — | Fixed |
| **female** | **black** | **Meta-Regression** | **—** | **NA – NA** | **—** | **NA** | **NA** | **NA** | **—** | **—** | **—** | **—** | **NA** |
| **female** | **black** | **Overall** | **2.79** | **2.38 – 3.20** | **2.79 ± 0.24 (2.47–3.02)** | **4** | **Observational Cohort** | **304** | **78.2%** | **13.8** | **<0.005** | **0.88** | **Random** |
| female | mixed | 18+ | 2.95 | 2.73 – 3.17 | 2.96 ± 0.21 (2.71–3.34) | 7 | Mixed | 488 | 53.1% | 12.8 | <0.05 | 0.86 | Random |
| female | mixed | 20-30 | 3.04 | 2.71 – 3.37 | — (3.04–3.04) | 1 | Observational Cohort | 25 | — | — | — | — | Single study |
| female | mixed | 30-40 | 2.90 | 2.47 – 3.33 | — (2.90–2.90) | 1 | Observational Cohort | 25 | — | — | — | — | Single study |
| female | mixed | 40-50 | 3.07 | 2.60 – 3.54 | — (3.07–3.07) | 1 | Observational Cohort | 25 | — | — | — | — | Single study |
| female | mixed | 50-60 | 3.08 | 2.28 – 3.88 | — (3.08–3.08) | 1 | Observational Cohort | 25 | — | — | — | — | Single study |
| female | mixed | 60+ | 1.94 | 1.61 – 2.27 | — (1.94–1.94) | 1 | Observational Cohort | 25 | — | — | — | — | Single study |
| **female** | **mixed** | **Meta-Regression** | **—** | **NA – NA** | **—** | **NA** | **NA** | **NA** | **—** | **—** | **—** | **—** | **NA** |
| **female** | **mixed** | **Overall** | **2.89** | **2.22 – 3.56** | **2.90 ± 0.34 (1.94–3.34)** | **12** | **Mixed** | **613** | **98.5%** | **748.4** | **<0.005** | **0.30** | **Random** |
| female | white | 18+ | — | NA – NA | — | NA | NA | NA | — | — | — | — | NA |
| female | white | 20-30 | — | NA – NA | — | NA | NA | NA | — | — | — | — | NA |
| female | white | 30-40 | — | NA – NA | — | NA | NA | NA | — | — | — | — | NA |
| female | white | 40-50 | 3.30 | 1.57 – 5.03 | — (3.30–3.30) | 1 | Observational Cohort | 524 | — | — | — | — | Single study |
| female | white | 50-60 | 3.24 | 1.54 – 4.95 | — (3.24–3.24) | 1 | Observational Cohort | 2409 | — | — | — | — | Single study |
| female | white | 60+ | 2.63 | 0.81 – 4.44 | 2.59 ± 0.13 (2.49–2.68) | 2 | Observational Cohort | 327 | 63% | 2.7 | 0.10 | — | Fixed |
| **female** | **white** | **Meta-Regression** | **—** | **NA – NA** | **—** | **NA** | **NA** | **NA** | **—** | **—** | **—** | **—** | **NA** |
| **female** | **white** | **Overall** | **2.94** | **2.16 – 3.71** | **2.93 ± 0.40 (2.49–3.30)** | **4** | **Observational Cohort** | **3260** | **97.9%** | **140.3** | **<0.005** | **0.18** | **Random** |
| *I² represents heterogeneity; Q test assesses between-study variation; Egger's p = linear regression test for funnel plot asymmetry; Meta-Regression shows covariate-adjusted estimates* | | | | | | | | | | | | | |

###### 6.3.2.3 Table of Results subdivided by race / Sex / Age

urate_ma_3rd_analysis$race_summary_table

Table 1: **race-Stratified Meta-Analysis Results**

24-Hour Urine Urate (mmol/24h)

| Sex Group | race | Age Band | 95% Range | Studies | Study Type | Participants | I² | Cochran's Q | Q (p-value) | Egger's p | Model |
| --- | --- | --- | --- | --- | --- | --- | --- | --- | --- | --- | --- |
| **main meta-analysis** | **All races** | **18+** | **2.33 – 4.05** | **64** | **Mixed** | **5104** | **89.7%** | **613.7** | **<0.005** | **<0.001** | **Random** |
| **main meta-analysis** | **All races** | **20-30** | **2.43 – 4.47** | **4** | **Mixed** | **96** | **99.4%** | **512.7** | **<0.005** | **0.91** | **Random** |
| **main meta-analysis** | **All races** | **30-40** | **2.29 – 4.53** | **5** | **Observational Cohort** | **185** | **97.3%** | **147.7** | **<0.005** | **0.80** | **Random** |
| **main meta-analysis** | **All races** | **40-50** | **2.77 – 3.88** | **7** | **Observational Cohort** | **665** | **93.8%** | **97.3** | **<0.005** | **0.74** | **Random** |
| **main meta-analysis** | **All races** | **50-60** | **2.57 – 4.00** | **5** | **Observational Cohort** | **2669** | **89.8%** | **39.1** | **<0.005** | **0.95** | **Random** |
| **main meta-analysis** | **All races** | **60+** | **1.67 – 3.92** | **8** | **Observational Cohort** | **825** | **98.8%** | **601** | **<0.005** | **0.08** | **Random** |
| **main meta-analysis** | **All races** | **Meta-Regression** | **2.35 – 4.02** | **51** | **Mixed** | **4797** | **93.4%** | **256** | **<0.005** | **<0.05** | **Multivariable** |
| **main meta-analysis** | **All races** | **Meta-Regression** | **2.35 – 4.02** | **51** | **Mixed** | **4797** | **93.4%** | **256** | **<0.005** | **<0.05** | **Multivariable** |
| **main meta-analysis** | **All races** | **Overall** | **2.29 – 4.11** | **93** | **Mixed** | **9544** | **97.6%** | **3759.5** | **<0.005** | **0.44** | **Random** |
| **main meta-analysis** | **All races** | **Overall** | **2.29 – 4.11** | **93** | **Mixed** | **9544** | **97.6%** | **3759.5** | **<0.005** | **0.44** | **Random** |
| main meta-analysis | Black | 18+ | 1.31 – 5.39 | 3 | Observational Cohort | 51 | 0% | 0 | 1.00 | — | Fixed |
| main meta-analysis | Black | 20-30 | — | — | NA | — | — | — | — | — | — |
| main meta-analysis | Black | 30-40 | — | — | NA | — | — | — | — | — | — |
| main meta-analysis | Black | 40-50 | 1.48 – 4.56 | 1 | Observational Cohort | 29 | — | — | — | — | Single study |
| main meta-analysis | Black | 50-60 | 1.03 – 4.85 | 1 | Observational Cohort | 131 | — | — | — | — | Single study |
| main meta-analysis | Black | 60+ | 0.83 – 4.41 | 2 | Observational Cohort | 144 | 68.7% | 3.2 | 0.07 | — | Fixed |
| main meta-analysis | Black | Meta-Regression | — | — | NA | — | — | — | — | — | — |
| **main meta-analysis** | **Black** | **Overall** | **2.40 – 3.55** | **7** | **Observational Cohort** | **355** | **77.4%** | **26.6** | **<0.005** | **0.14** | **Random** |
| main meta-analysis | Mixed | 18+ | 2.25 – 4.06 | 55 | Mixed | 4888 | 90.6% | 573.2 | <0.005 | <0.001 | Random |
| main meta-analysis | Mixed | 20-30 | 2.43 – 4.47 | 4 | Mixed | 96 | 99.4% | 512.7 | <0.005 | 0.91 | Random |
| main meta-analysis | Mixed | 30-40 | 2.29 – 4.53 | 5 | Observational Cohort | 185 | 97.3% | 147.7 | <0.005 | 0.80 | Random |
| main meta-analysis | Mixed | 40-50 | 2.70 – 4.02 | 4 | Observational Cohort | 106 | 96.8% | 92.6 | <0.005 | 0.59 | Random |
| main meta-analysis | Mixed | 50-60 | 2.49 – 3.88 | 2 | Observational Cohort | 50 | 62.6% | 2.7 | 0.10 | — | Fixed |
| main meta-analysis | Mixed | 60+ | 1.61 – 2.27 | 1 | Observational Cohort | 25 | — | — | — | — | Single study |
| main meta-analysis | Mixed | Meta-Regression | — | — | NA | — | — | — | — | — | — |
| **main meta-analysis** | **Mixed** | **Overall** | **2.23 – 4.14** | **71** | **Mixed** | **5350** | **97.9%** | **3403.4** | **<0.005** | **0.46** | **Random** |
| main meta-analysis | White | 18+ | 1.00 – 5.94 | 6 | Observational Cohort | 165 | 0% | 4.4 | 0.49 | <0.001 | Fixed |
| main meta-analysis | White | 20-30 | — | — | NA | — | — | — | — | — | — |
| main meta-analysis | White | 30-40 | — | — | NA | — | — | — | — | — | — |
| main meta-analysis | White | 40-50 | 1.57 – 5.04 | 2 | Observational Cohort | 530 | 3.6% | 1 | 0.31 | — | Fixed |
| main meta-analysis | White | 50-60 | 2.58 – 4.62 | 2 | Observational Cohort | 2488 | 95.6% | 22.8 | <0.005 | — | Random |
| main meta-analysis | White | 60+ | 2.00 – 4.10 | 5 | Observational Cohort | 656 | 97.4% | 152.3 | <0.005 | 0.46 | Random |
| main meta-analysis | White | Meta-Regression | — | — | NA | — | — | — | — | — | — |
| **main meta-analysis** | **White** | **Overall** | **2.51 – 4.16** | **15** | **Observational Cohort** | **3839** | **95.1%** | **287.3** | **<0.005** | **0.75** | **Random** |
| **male** | **All races** | **18+** | **0.09 – 6.65** | **4** | **Observational Cohort** | **182** | **98%** | **147.6** | **<0.005** | **0.36** | **Random** |
| **male** | **All races** | **20-30** | **2.54 – 4.64** | **3** | **Mixed** | **71** | **98.3%** | **115.3** | **<0.005** | **0.36** | **Random** |
| **male** | **All races** | **30-40** | **2.76 – 4.75** | **3** | **Observational Cohort** | **143** | **92.6%** | **27.1** | **<0.005** | **0.07** | **Random** |
| **male** | **All races** | **40-50** | **3.12 – 4.59** | **2** | **Observational Cohort** | **31** | **0%** | **0.1** | **0.82** | **—** | **Fixed** |
| **male** | **All races** | **50-60** | **2.60 – 4.61** | **2** | **Observational Cohort** | **104** | **94.8%** | **19.3** | **<0.005** | **—** | **Random** |
| **male** | **All races** | **60+** | **2.42 – 4.29** | **3** | **Observational Cohort** | **329** | **98%** | **99.3** | **<0.005** | **0.41** | **Random** |
| **male** | **All races** | **Meta-Regression** | **1.31 – 6.61** | **5** | **Mixed** | **299** | **99.7%** | **200.8** | **<0.005** | **0.94** | **Univariable** |
| **male** | **All races** | **Overall** | **1.99 – 5.11** | **17** | **Mixed** | **860** | **98.6%** | **1118.5** | **<0.005** | **0.80** | **Random** |
| male | Black | 18+ | — | — | NA | — | — | — | — | — | — |
| male | Black | 20-30 | — | — | NA | — | — | — | — | — | — |
| male | Black | 30-40 | — | — | NA | — | — | — | — | — | — |
| male | Black | 40-50 | — | — | NA | — | — | — | — | — | — |
| male | Black | 50-60 | — | — | NA | — | — | — | — | — | — |
| male | Black | 60+ | — | — | NA | — | — | — | — | — | — |
| male | Black | Meta-Regression | — | — | NA | — | — | — | — | — | — |
| **male** | **Black** | **Overall** | **—** | **—** | **NA** | **—** | **—** | **—** | **—** | **—** | **—** |
| male | Mixed | 18+ | 0.09 – 6.65 | 4 | Observational Cohort | 182 | 98% | 147.6 | <0.005 | 0.36 | Random |
| male | Mixed | 20-30 | 2.54 – 4.64 | 3 | Mixed | 71 | 98.3% | 115.3 | <0.005 | 0.36 | Random |
| male | Mixed | 30-40 | 2.76 – 4.75 | 3 | Observational Cohort | 143 | 92.6% | 27.1 | <0.005 | 0.07 | Random |
| male | Mixed | 40-50 | 3.19 – 4.53 | 1 | Observational Cohort | 25 | — | — | — | — | Single study |
| male | Mixed | 50-60 | 2.62 – 3.88 | 1 | Observational Cohort | 25 | — | — | — | — | Single study |
| male | Mixed | 60+ | — | — | NA | — | — | — | — | — | — |
| male | Mixed | Meta-Regression | — | — | NA | — | — | — | — | — | — |
| **male** | **Mixed** | **Overall** | **1.73 – 5.37** | **12** | **Mixed** | **446** | **98.8%** | **936.6** | **<0.005** | **0.84** | **Random** |
| male | White | 18+ | — | — | NA | — | — | — | — | — | — |
| male | White | 20-30 | — | — | NA | — | — | — | — | — | — |
| male | White | 30-40 | — | — | NA | — | — | — | — | — | — |
| male | White | 40-50 | 1.61 – 5.90 | 1 | Observational Cohort | 6 | — | — | — | — | Single study |
| male | White | 50-60 | 1.27 – 6.72 | 1 | Observational Cohort | 79 | — | — | — | — | Single study |
| male | White | 60+ | 2.42 – 4.29 | 3 | Observational Cohort | 329 | 98% | 99.3 | <0.005 | 0.41 | Random |
| male | White | Meta-Regression | — | — | NA | — | — | — | — | — | — |
| **male** | **White** | **Overall** | **2.64 – 4.44** | **5** | **Observational Cohort** | **414** | **97%** | **132.8** | **<0.005** | **0.19** | **Random** |
| **female** | **All races** | **18+** | **2.73 – 3.17** | **7** | **Mixed** | **488** | **53.1%** | **12.8** | **<0.05** | **0.86** | **Random** |
| **female** | **All races** | **20-30** | **2.71 – 3.37** | **1** | **Observational Cohort** | **25** | **—** | **—** | **—** | **—** | **Single study** |
| **female** | **All races** | **30-40** | **2.47 – 3.33** | **1** | **Observational Cohort** | **25** | **—** | **—** | **—** | **—** | **Single study** |
| **female** | **All races** | **40-50** | **2.88 – 3.43** | **3** | **Observational Cohort** | **578** | **87.4%** | **15.8** | **<0.005** | **0.65** | **Random** |
| **female** | **All races** | **50-60** | **2.82 – 3.38** | **3** | **Observational Cohort** | **2565** | **86.7%** | **15** | **<0.005** | **0.18** | **Random** |
| **female** | **All races** | **60+** | **1.84 – 3.08** | **5** | **Observational Cohort** | **496** | **97.6%** | **163.7** | **<0.005** | **0.10** | **Random** |
| **female** | **All races** | **Meta-Regression** | **2.80 – 3.12** | **7** | **Mixed** | **488** | **35.9%** | **6.6** | **0.16** | **0.86** | **Multivariable** |
| **female** | **All races** | **Overall** | **2.25 – 3.51** | **20** | **Mixed** | **4177** | **98.6%** | **1324.3** | **<0.005** | **0.30** | **Random** |
| female | Black | 18+ | — | — | NA | — | — | — | — | — | — |
| female | Black | 20-30 | — | — | NA | — | — | — | — | — | — |
| female | Black | 30-40 | — | — | NA | — | — | — | — | — | — |
| female | Black | 40-50 | 1.48 – 4.56 | 1 | Observational Cohort | 29 | — | — | — | — | Single study |
| female | Black | 50-60 | 1.03 – 4.85 | 1 | Observational Cohort | 131 | — | — | — | — | Single study |
| female | Black | 60+ | 0.83 – 4.41 | 2 | Observational Cohort | 144 | 68.7% | 3.2 | 0.07 | — | Fixed |
| female | Black | Meta-Regression | — | — | NA | — | — | — | — | — | — |
| **female** | **Black** | **Overall** | **2.38 – 3.20** | **4** | **Observational Cohort** | **304** | **78.2%** | **13.8** | **<0.005** | **0.88** | **Random** |
| female | Mixed | 18+ | 2.73 – 3.17 | 7 | Mixed | 488 | 53.1% | 12.8 | <0.05 | 0.86 | Random |
| female | Mixed | 20-30 | 2.71 – 3.37 | 1 | Observational Cohort | 25 | — | — | — | — | Single study |
| female | Mixed | 30-40 | 2.47 – 3.33 | 1 | Observational Cohort | 25 | — | — | — | — | Single study |
| female | Mixed | 40-50 | 2.60 – 3.54 | 1 | Observational Cohort | 25 | — | — | — | — | Single study |
| female | Mixed | 50-60 | 2.28 – 3.88 | 1 | Observational Cohort | 25 | — | — | — | — | Single study |
| female | Mixed | 60+ | 1.61 – 2.27 | 1 | Observational Cohort | 25 | — | — | — | — | Single study |
| female | Mixed | Meta-Regression | — | — | NA | — | — | — | — | — | — |
| **female** | **Mixed** | **Overall** | **2.22 – 3.56** | **12** | **Mixed** | **613** | **98.5%** | **748.4** | **<0.005** | **0.30** | **Random** |
| female | White | 18+ | — | — | NA | — | — | — | — | — | — |
| female | White | 20-30 | — | — | NA | — | — | — | — | — | — |
| female | White | 30-40 | — | — | NA | — | — | — | — | — | — |
| female | White | 40-50 | 1.57 – 5.03 | 1 | Observational Cohort | 524 | — | — | — | — | Single study |
| female | White | 50-60 | 1.54 – 4.95 | 1 | Observational Cohort | 2409 | — | — | — | — | Single study |
| female | White | 60+ | 0.81 – 4.44 | 2 | Observational Cohort | 327 | 63% | 2.7 | 0.10 | — | Fixed |
| female | White | Meta-Regression | — | — | NA | — | — | — | — | — | — |
| **female** | **White** | **Overall** | **2.16 – 3.71** | **4** | **Observational Cohort** | **3260** | **97.9%** | **140.3** | **<0.005** | **0.18** | **Random** |
| *I² represents heterogeneity; Q test assesses between-study variation; Egger's p = linear regression test for funnel plot asymmetry; — indicates insufficient data* | | | | | | | | | | | |

###### 6.3.2.4 Wald Tests

urate_ma_3rd_analysis$wald_tests$sex_comparison

Table 1: **Sex Comparison (Overall)**

| Comparison | % Shift | P (Wald Test) | Mean Diff (95% CI) |
| --- | --- | --- | --- |
| Male vs Female (Overall) | +23.2% | 0.436 | 0.67 [-1.01, 2.35] |
| *Mean difference = Group 1 − Group 2 in component units (mmol/24h, L/24h, or pH); % shift = difference relative to Group 2.* | | | |

urate_ma_3rd_analysis$wald_tests$race_comparisons

Table 1: **race Comparisons Within Sex Groups**

| Sex Group | Comparison | % Shift | P (Wald Test) | Mean Diff (95% CI) |
| --- | --- | --- | --- | --- |
| male | black vs mixed | — | — | — |
| male | black vs white | — | — | — |
| male | mixed vs white | +0.4% | 0.990 | 0.01 [-2.02, 2.05] |
| female | black vs mixed | -3.5% | 0.802 | -0.10 [-0.88, 0.68] |
| female | black vs white | -5.0% | 0.744 | -0.15 [-1.03, 0.73] |
| female | mixed vs white | -1.6% | 0.929 | -0.05 [-1.07, 0.98] |
| *Mean difference = Group 1 − Group 2 in component units (mmol/24h, L/24h, or pH); % shift = difference relative to Group 2.* | | | | |

urate_ma_3rd_analysis$wald_tests$age_band_comparisons

Table 1: **Age Band Comparisons Within Sex Groups**

| Sex Group | Comparison | % Shift | P (Wald Test) | Mean Diff (95% CI) |
| --- | --- | --- | --- | --- |
| main meta-analysis | 18+ vs 20-30 | -7.5% | 0.702 | -0.26 [-1.59, 1.07] |
| main meta-analysis | 18+ vs 30-40 | -6.4% | 0.762 | -0.22 [-1.63, 1.19] |
| main meta-analysis | 18+ vs 40-50 | -4.0% | 0.799 | -0.13 [-1.16, 0.89] |
| main meta-analysis | 18+ vs 50-60 | -2.9% | 0.867 | -0.10 [-1.22, 1.03] |
| main meta-analysis | 18+ vs 60+ | +14.0% | 0.587 | 0.39 [-1.02, 1.81] |
| main meta-analysis | 20-30 vs 30-40 | +1.2% | 0.957 | 0.04 [-1.47, 1.55] |
| main meta-analysis | 20-30 vs 40-50 | +3.8% | 0.830 | 0.13 [-1.03, 1.28] |
| main meta-analysis | 20-30 vs 50-60 | +5.0% | 0.795 | 0.16 [-1.08, 1.41] |
| main meta-analysis | 20-30 vs 60+ | +23.3% | 0.399 | 0.65 [-0.86, 2.17] |
| main meta-analysis | 30-40 vs 40-50 | +2.6% | 0.893 | 0.09 [-1.16, 1.33] |
| main meta-analysis | 30-40 vs 50-60 | +3.7% | 0.856 | 0.12 [-1.21, 1.45] |
| main meta-analysis | 30-40 vs 60+ | +21.8% | 0.451 | 0.61 [-0.98, 2.20] |
| main meta-analysis | 40-50 vs 50-60 | +1.1% | 0.935 | 0.04 [-0.87, 0.95] |
| main meta-analysis | 40-50 vs 60+ | +18.8% | 0.412 | 0.53 [-0.73, 1.78] |
| main meta-analysis | 50-60 vs 60+ | +17.4% | 0.474 | 0.49 [-0.85, 1.82] |
| male | 18+ vs 20-30 | -6.1% | 0.901 | -0.22 [-3.66, 3.23] |
| male | 18+ vs 30-40 | -10.2% | 0.828 | -0.38 [-3.81, 3.05] |
| male | 18+ vs 40-50 | -12.6% | 0.777 | -0.49 [-3.85, 2.88] |
| male | 18+ vs 50-60 | -6.6% | 0.892 | -0.24 [-3.67, 3.19] |
| male | 18+ vs 60+ | +0.4% | 0.993 | 0.01 [-3.40, 3.43] |
| male | 20-30 vs 30-40 | -4.3% | 0.826 | -0.16 [-1.61, 1.28] |
| male | 20-30 vs 40-50 | -6.9% | 0.682 | -0.27 [-1.55, 1.01] |
| male | 20-30 vs 50-60 | -0.5% | 0.980 | -0.02 [-1.47, 1.43] |
| male | 20-30 vs 60+ | +7.0% | 0.744 | 0.23 [-1.17, 1.64] |
| male | 30-40 vs 40-50 | -2.7% | 0.867 | -0.11 [-1.34, 1.13] |
| male | 30-40 vs 50-60 | +4.0% | 0.842 | 0.14 [-1.27, 1.56] |
| male | 30-40 vs 60+ | +11.8% | 0.569 | 0.40 [-0.97, 1.76] |
| male | 40-50 vs 50-60 | +6.9% | 0.694 | 0.25 [-0.99, 1.49] |
| male | 40-50 vs 60+ | +14.9% | 0.407 | 0.50 [-0.69, 1.69] |
| male | 50-60 vs 60+ | +7.5% | 0.718 | 0.25 [-1.12, 1.62] |
| female | 18+ vs 20-30 | -3.0% | 0.659 | -0.09 [-0.49, 0.31] |
| female | 18+ vs 30-40 | +1.7% | 0.840 | 0.05 [-0.43, 0.53] |
| female | 18+ vs 40-50 | -6.4% | 0.261 | -0.20 [-0.55, 0.15] |
| female | 18+ vs 50-60 | -4.9% | 0.399 | -0.15 [-0.51, 0.20] |
| female | 18+ vs 60+ | +20.0% | 0.145 | 0.49 [-0.17, 1.15] |
| female | 20-30 vs 30-40 | +4.8% | 0.615 | 0.14 [-0.40, 0.68] |
| female | 20-30 vs 40-50 | -3.6% | 0.610 | -0.11 [-0.54, 0.32] |
| female | 20-30 vs 50-60 | -2.0% | 0.775 | -0.06 [-0.50, 0.37] |
| female | 20-30 vs 60+ | +23.6% | 0.107 | 0.58 [-0.13, 1.29] |
| female | 30-40 vs 40-50 | -8.0% | 0.333 | -0.25 [-0.76, 0.26] |
| female | 30-40 vs 50-60 | -6.6% | 0.438 | -0.20 [-0.72, 0.31] |
| female | 30-40 vs 60+ | +17.9% | 0.254 | 0.44 [-0.32, 1.20] |
| female | 40-50 vs 50-60 | +1.6% | 0.807 | 0.05 [-0.34, 0.44] |
| female | 40-50 vs 60+ | +28.2% | <0.05 | 0.69 [0.01, 1.37] |
| female | 50-60 vs 60+ | +26.2% | 0.064 | 0.64 [-0.04, 1.33] |
| *Mean difference = Group 1 − Group 2 in component units (mmol/24h, L/24h, or pH); % shift = difference relative to Group 2.* | | | | |

urate_ma_3rd_analysis$wald_tests$sex_within_age_bands

Table 1: **Sex Comparisons Within Age Bands**

| Age Band | % Shift | P (Wald Test) | Mean Diff (95% CI) |
| --- | --- | --- | --- |
| 18+ | +14.3% | 0.802 | 0.42 [-2.87, 3.71] |
| 20-30 | +18.1% | 0.327 | 0.55 [-0.55, 1.65] |
| 30-40 | +29.4% | 0.124 | 0.85 [-0.23, 1.94] |
| 40-50 | +22.4% | 0.077 | 0.71 [-0.08, 1.49] |
| 50-60 | +16.3% | 0.342 | 0.50 [-0.54, 1.55] |
| 60+ | +36.5% | 0.117 | 0.90 [-0.22, 2.02] |
| *Mean difference = Group 1 − Group 2 in component units (mmol/24h, L/24h, or pH); % shift = difference relative to Group 2.* | | | |

#### 6.4 Creatinine

##### 6.4.1 Comparison by Age Band

###### 6.4.1.1 Plot

```
creat_ma_2nd_analysis <- sex_age_subdivision_ma("creat")
## Not enough studies for multivariable meta-regression (k = 3, need > 6). Running univariable models instead.
## Not enough studies for multivariable meta-regression (k = 6, need > 6). Running univariable models instead.
creat_ma_2nd_analysis$combined_plot
## `height` was translated to `width`.
## `height` was translated to `width`.
## `height` was translated to `width`.
```

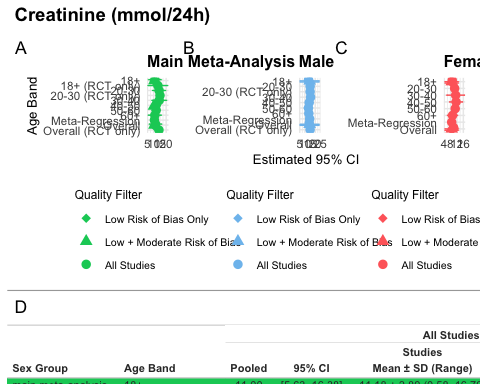

###### 6.4.1.2 Summary Table

creat_ma_2nd_analysis$combined_summary_table

Table 1: **Combined Meta-Analysis Results**

24-Hour Urine Creatinine (mmol/24h)

|  |  | **All Studies** | | | | | | **Low Risk of Bias Only** | | | | | | **Low + Moderate RoB** | | | | | |
| --- | --- | --- | --- | --- | --- | --- | --- | --- | --- | --- | --- | --- | --- | --- | --- | --- | --- | --- | --- |
| Sex Group | Age Band | Pooled | 95% CI | Studies  Mean ± SD (Range) | Studies, n | Q p-value | Egger's p | Pooled | 95% CI | Studies  Mean ± SD (Range) | Studies, n | Q p-value | Egger's p | Pooled | 95% CI | Studies  Mean ± SD (Range) | Studies, n | Q p-value | Egger's p |
| main meta-analysis | 18+ | 11.00 | [5.63, 16.38] | 11.18 ± 2.89 (0.58–16.79) | 48 | <0.001 | <0.001 | 10.49 | [7.50, 13.47] | 10.69 ± 1.81 (8.53–14.59) | 25 | <0.001 | <0.005 | 10.65 | [7.49, 13.81] | 11.03 ± 2.16 (8.53–16.79) | 27 | <0.001 | <0.005 |
| **main meta-analysis** | **18+ (RCT only)** | **13.37** | **[5.46, 21.28]** | **13.86 ± 1.90 (11.16–16.79)** | **6** | **0.17** | **0.54** | **—** | **—** | **—** | **—** | **—** | **—** | **—** | **—** | **—** | **—** | **—** | **—** |
| main meta-analysis | 20-30 | 14.32 | [9.15, 19.50] | 14.40 ± 2.69 (10.56–16.85) | 4 | <0.001 | 0.63 | — | — | — | — | — | — | — | — | — | — | — | — |
| **main meta-analysis** | **20-30 (RCT only)** | **15.04** | **[13.04, 17.04]** | **15.93 ± 1.31 (15.00–16.85)** | **2** | **0.07** | **—** | **—** | **—** | **—** | **—** | **—** | **—** | **—** | **—** | **—** | **—** | **—** | **—** |
| main meta-analysis | 30-40 | 14.43 | [8.59, 20.28] | 14.74 ± 3.26 (11.18–17.58) | 3 | <0.001 | 0.81 | — | — | — | — | — | — | — | — | — | — | — | — |
| main meta-analysis | 40-50 | 12.15 | [8.74, 15.56] | 12.18 ± 1.70 (10.25–15.27) | 6 | <0.001 | 0.26 | — | — | — | — | — | — | 10.25 | [9.15, 11.35] | — (10.25–10.25) | 1 | — | — |
| main meta-analysis | 50-60 | 11.97 | [8.69, 15.25] | 12.00 ± 1.71 (9.85–13.95) | 5 | <0.001 | 0.23 | — | — | — | — | — | — | — | — | — | — | — | — |
| main meta-analysis | 60+ | 10.93 | [5.70, 16.15] | 10.93 ± 2.67 (6.94–14.58) | 8 | <0.001 | <0.05 | — | — | — | — | — | — | — | — | — | — | — | — |
| **main meta-analysis** | **Meta-Regression** | **10.59** | **[8.88, 12.31]** | **—** | **38** | **<0.001** | **0.27** | **—** | **—** | **—** | **—** | **—** | **—** | **—** | **—** | **—** | **—** | **—** | **—** |
| **main meta-analysis** | **Overall** | **11.46** | **[6.16, 16.77]** | **11.61 ± 2.84 (0.58–17.58)** | **74** | **<0.001** | **<0.001** | **10.49** | **[7.50, 13.47]** | **10.69 ± 1.81 (8.53–14.59)** | **25** | **<0.001** | **<0.005** | **10.62** | **[7.55, 13.69]** | **11.00 ± 2.13 (8.53–16.79)** | **28** | **<0.001** | **<0.05** |
| **main meta-analysis** | **Overall (RCT only)** | **14.20** | **[11.40, 16.99]** | **14.38 ± 1.93 (11.16–16.85)** | **8** | **<0.005** | **0.31** | **—** | **—** | **—** | **—** | **—** | **—** | **—** | **—** | **—** | **—** | **—** | **—** |
| male | 18+ | 13.53 | [11.58, 15.47] | 13.65 ± 1.21 (12.56–15.22) | 5 | <0.001 | 0.61 | 14.59 | [5.06, 24.12] | — (14.59–14.59) | 1 | — | — | 14.59 | [5.06, 24.12] | — (14.59–14.59) | 1 | — | — |
| **male** | **18+ (RCT only)** | **—** | **—** | **—** | **—** | **—** | **—** | **—** | **—** | **—** | **—** | **—** | **—** | **—** | **—** | **—** | **—** | **—** | **—** |
| male | 20-30 | 15.09 | [13.04, 17.14] | 15.68 ± 1.02 (15.00–16.85) | 3 | 0.16 | 0.05 | — | — | — | — | — | — | — | — | — | — | — | — |
| **male** | **20-30 (RCT only)** | **15.04** | **[13.04, 17.04]** | **15.93 ± 1.31 (15.00–16.85)** | **2** | **0.07** | **—** | **—** | **—** | **—** | **—** | **—** | **—** | **—** | **—** | **—** | **—** | **—** | **—** |
| male | 30-40 | 15.51 | [12.33, 18.68] | 16.52 ± 1.49 (15.47–17.58) | 2 | 0.32 | — | — | — | — | — | — | — | — | — | — | — | — | — |
| male | 40-50 | 14.15 | [10.68, 17.63] | 13.82 ± 2.05 (12.38–15.27) | 2 | <0.05 | — | — | — | — | — | — | — | — | — | — | — | — | — |
| male | 50-60 | 13.43 | [9.57, 17.30] | 13.64 ± 0.44 (13.33–13.95) | 2 | 0.23 | — | — | — | — | — | — | — | — | — | — | — | — | — |
| male | 60+ | 13.80 | [12.51, 15.08] | 13.77 ± 0.70 (13.36–14.58) | 3 | <0.001 | 0.50 | — | — | — | — | — | — | — | — | — | — | — | — |
| **male** | **Meta-Regression** | **14.59** | **[14.58, 14.59]** | **—** | **3** | **0.12** | **0.70** | **—** | **—** | **—** | **—** | **—** | **—** | **—** | **—** | **—** | **—** | **—** | **—** |
| **male** | **Overall** | **14.23** | **[12.16, 16.29]** | **14.39 ± 1.48 (12.38–17.58)** | **17** | **<0.001** | **0.99** | **14.59** | **[5.06, 24.12]** | **— (14.59–14.59)** | **1** | **—** | **—** | **14.59** | **[5.06, 24.12]** | **— (14.59–14.59)** | **1** | **—** | **—** |
| **male** | **Overall (RCT only)** | **15.04** | **[13.04, 17.04]** | **15.93 ± 1.31 (15.00–16.85)** | **2** | **0.07** | **—** | **—** | **—** | **—** | **—** | **—** | **—** | **—** | **—** | **—** | **—** | **—** | **—** |
| female | 18+ | 9.04 | [7.92, 10.16] | 9.03 ± 0.64 (8.62–10.06) | 8 | <0.001 | 0.55 | 8.70 | [4.31, 13.09] | 8.70 ± 0.00 (8.70–8.70) | 2 | 1.00 | — | 8.70 | [4.31, 13.09] | 8.70 ± 0.00 (8.70–8.70) | 2 | 1.00 | — |
| **female** | **18+ (RCT only)** | **—** | **—** | **—** | **—** | **—** | **—** | **—** | **—** | **—** | **—** | **—** | **—** | **—** | **—** | **—** | **—** | **—** | **—** |
| female | 20-30 | 10.56 | [9.21, 11.91] | — (10.56–10.56) | 1 | — | — | — | — | — | — | — | — | — | — | — | — | — | — |
| **female** | **20-30 (RCT only)** | **—** | **—** | **—** | **—** | **—** | **—** | **—** | **—** | **—** | **—** | **—** | **—** | **—** | **—** | **—** | **—** | **—** | **—** |
| female | 30-40 | 11.18 | [5.69, 16.67] | — (11.18–11.18) | 1 | — | — | — | — | — | — | — | — | — | — | — | — | — | — |
| female | 40-50 | 11.44 | [7.41, 15.47] | 11.73 ± 0.49 (11.39–12.29) | 3 | 0.19 | 0.15 | — | — | — | — | — | — | — | — | — | — | — | — |
| female | 50-60 | 10.90 | [8.78, 13.02] | 10.90 ± 1.12 (9.85–12.07) | 3 | <0.001 | 0.87 | — | — | — | — | — | — | — | — | — | — | — | — |
| female | 60+ | 9.22 | [6.10, 12.34] | 9.23 ± 1.60 (6.94–11.04) | 5 | <0.001 | 0.20 | — | — | — | — | — | — | — | — | — | — | — | — |
| **female** | **Meta-Regression** | **8.70** | **[7.46, 9.94]** | **—** | **6** | **<0.001** | **0.20** | **—** | **—** | **—** | **—** | **—** | **—** | **—** | **—** | **—** | **—** | **—** | **—** |
| **female** | **Overall** | **9.89** | **[7.20, 12.59]** | **9.91 ± 1.40 (6.94–12.29)** | **21** | **<0.001** | **0.18** | **8.70** | **[4.31, 13.09]** | **8.70 ± 0.00 (8.70–8.70)** | **2** | **1.00** | **—** | **8.70** | **[4.31, 13.09]** | **8.70 ± 0.00 (8.70–8.70)** | **2** | **1.00** | **—** |
| **female** | **Overall (RCT only)** | **—** | **—** | **—** | **—** | **—** | **—** | **—** | **—** | **—** | **—** | **—** | **—** | **—** | **—** | **—** | **—** | **—** | **—** |
| *Studies, n = number of studies; Q p-value = Cochran's Q heterogeneity test; Egger's p = linear regression test for funnel plot asymmetry; RoB = Risk of Bias* | | | | | | | | | | | | | | | | | | | |

###### 6.4.1.3 Wald Tests

creat_ma_2nd_analysis$wald_tests$sex_comparison

Table 1: **Sex Comparison (Overall)**

| Comparison | % Shift | P (Wald Test) | Mean Diff (95% CI) |
| --- | --- | --- | --- |
| Male vs Female (Overall) | +43.8% | <0.05 | 4.33 [0.94, 7.73] |
| *Mean difference = Group 1 − Group 2 in component units (mmol/24h, L/24h, or pH); % shift = difference relative to Group 2.* | | | |

creat_ma_2nd_analysis$wald_tests$age_band_comparisons

Table 1: **Age Band Comparisons Within Sex Groups**

| Sex Group | Comparison | % Shift | P (Wald Test) | Mean Diff (95% CI) |
| --- | --- | --- | --- | --- |
| main meta-analysis | 18+ vs 20-30 | -23.2% | 0.383 | -3.32 [-10.78, 4.14] |
| main meta-analysis | 18+ vs 30-40 | -23.8% | 0.397 | -3.43 [-11.37, 4.51] |
| main meta-analysis | 18+ vs 40-50 | -9.5% | 0.724 | -1.15 [-7.52, 5.22] |
| main meta-analysis | 18+ vs 50-60 | -8.1% | 0.763 | -0.97 [-7.27, 5.33] |
| main meta-analysis | 18+ vs 60+ | +0.7% | 0.984 | 0.08 [-7.42, 7.57] |
| main meta-analysis | 20-30 vs 30-40 | -0.8% | 0.978 | -0.11 [-7.92, 7.70] |
| main meta-analysis | 20-30 vs 40-50 | +17.9% | 0.492 | 2.17 [-4.03, 8.37] |
| main meta-analysis | 20-30 vs 50-60 | +19.6% | 0.452 | 2.35 [-3.78, 8.47] |
| main meta-analysis | 20-30 vs 60+ | +31.1% | 0.365 | 3.39 [-3.96, 10.75] |
| main meta-analysis | 30-40 vs 40-50 | +18.8% | 0.509 | 2.28 [-4.49, 9.05] |
| main meta-analysis | 30-40 vs 50-60 | +20.5% | 0.472 | 2.46 [-4.24, 9.16] |
| main meta-analysis | 30-40 vs 60+ | +32.1% | 0.381 | 3.50 [-4.33, 11.34] |
| main meta-analysis | 40-50 vs 50-60 | +1.5% | 0.941 | 0.18 [-4.55, 4.91] |
| main meta-analysis | 40-50 vs 60+ | +11.2% | 0.701 | 1.22 [-5.01, 7.46] |
| main meta-analysis | 50-60 vs 60+ | +9.6% | 0.740 | 1.05 [-5.12, 7.21] |
| male | 18+ vs 20-30 | -10.3% | 0.279 | -1.56 [-4.39, 1.27] |
| male | 18+ vs 30-40 | -12.7% | 0.298 | -1.98 [-5.70, 1.74] |
| male | 18+ vs 40-50 | -4.4% | 0.759 | -0.62 [-4.61, 3.36] |
| male | 18+ vs 50-60 | +0.7% | 0.966 | 0.09 [-4.23, 4.42] |
| male | 18+ vs 60+ | -1.9% | 0.822 | -0.27 [-2.60, 2.07] |
| male | 20-30 vs 30-40 | -2.7% | 0.829 | -0.42 [-4.19, 3.36] |
| male | 20-30 vs 40-50 | +6.6% | 0.649 | 0.94 [-3.10, 4.97] |
| male | 20-30 vs 50-60 | +12.3% | 0.459 | 1.65 [-2.72, 6.03] |
| male | 20-30 vs 60+ | +9.4% | 0.295 | 1.29 [-1.13, 3.71] |
| male | 30-40 vs 40-50 | +9.6% | 0.573 | 1.35 [-3.35, 6.06] |
| male | 30-40 vs 50-60 | +15.4% | 0.417 | 2.07 [-2.93, 7.07] |
| male | 30-40 vs 60+ | +12.4% | 0.328 | 1.71 [-1.71, 5.13] |
| male | 40-50 vs 50-60 | +5.3% | 0.787 | 0.72 [-4.48, 5.92] |
| male | 40-50 vs 60+ | +2.6% | 0.851 | 0.36 [-3.35, 4.06] |
| male | 50-60 vs 60+ | -2.6% | 0.862 | -0.36 [-4.44, 3.71] |
| female | 18+ vs 20-30 | -14.4% | 0.090 | -1.52 [-3.28, 0.24] |
| female | 18+ vs 30-40 | -19.2% | 0.454 | -2.14 [-7.74, 3.46] |
| female | 18+ vs 40-50 | -21.0% | 0.260 | -2.41 [-6.59, 1.78] |
| female | 18+ vs 50-60 | -17.1% | 0.128 | -1.86 [-4.26, 0.53] |
| female | 18+ vs 60+ | -2.0% | 0.915 | -0.18 [-3.50, 3.14] |
| female | 20-30 vs 30-40 | -5.5% | 0.830 | -0.62 [-6.27, 5.03] |
| female | 20-30 vs 40-50 | -7.7% | 0.684 | -0.88 [-5.13, 3.37] |
| female | 20-30 vs 50-60 | -3.1% | 0.790 | -0.34 [-2.85, 2.17] |
| female | 20-30 vs 60+ | +14.5% | 0.440 | 1.34 [-2.06, 4.74] |
| female | 30-40 vs 40-50 | -2.3% | 0.940 | -0.26 [-7.07, 6.54] |
| female | 30-40 vs 50-60 | +2.6% | 0.926 | 0.28 [-5.60, 6.16] |
| female | 30-40 vs 60+ | +21.3% | 0.543 | 1.96 [-4.35, 8.27] |
| female | 40-50 vs 50-60 | +5.0% | 0.815 | 0.54 [-4.01, 5.09] |
| female | 40-50 vs 60+ | +24.1% | 0.392 | 2.22 [-2.87, 7.32] |
| female | 50-60 vs 60+ | +18.2% | 0.382 | 1.68 [-2.09, 5.45] |
| *Mean difference = Group 1 − Group 2 in component units (mmol/24h, L/24h, or pH); % shift = difference relative to Group 2.* | | | | |

creat_ma_2nd_analysis$wald_tests$sex_within_age_bands

Table 1: **Sex Comparisons Within Age Bands**

| Age Band | % Shift | P (Wald Test) | Mean Diff (95% CI) |
| --- | --- | --- | --- |
| 18+ | +49.7% | <0.001 | 4.49 [2.24, 6.74] |
| 20-30 | +42.9% | <0.001 | 4.53 [2.07, 6.98] |
| 30-40 | +38.7% | 0.181 | 4.33 [-2.01, 10.66] |
| 40-50 | +23.7% | 0.318 | 2.71 [-2.61, 8.03] |
| 50-60 | +23.2% | 0.260 | 2.53 [-1.87, 6.94] |
| 60+ | +49.6% | <0.01 | 4.58 [1.20, 7.95] |
| *Mean difference = Group 1 − Group 2 in component units (mmol/24h, L/24h, or pH); % shift = difference relative to Group 2.* | | | |

###### 6.4.1.4 Meta-Regression (plain language)

format_meta_reg_plain("creat", sex_group = "all")

Message

No meta-regression result for creat / all

###### 6.4.1.5 Assay Subgroup Analysis

assay_subgroup_table("creat")

Table 1: **Assay subgroup analysis — Creat**

*Per-assay pooled estimates; categories with k < 3 listed but not pooled.*

| Assay category | Studies, n | Participants | 95% CI | I² | Q p-value | Egger's p | Note |
| --- | --- | --- | --- | --- | --- | --- | --- |
| Unreported | 48 | 6,466 | 11.73 [10.86, 12.60] | 100% | <0.001 | <0.001 |  |
| Autoanalyser | 18 | 2,386 | 10.41 [9.75, 11.07] | 78% | <0.001 | 0.028 |  |
| Jaffe | 8 | 206 | 12.26 [10.38, 14.14] | 99% | <0.001 | 0.251 |  |

###### 6.4.1.6 Preservative Sensitivity (Litholink / acidified samples)

```
preservative_sensitivity_ma("creat")
## # A tibble: 3 × 6
##   group               n_studies n_participants pooled_estimate lower_95 upper_95
##   <chr>                   <int>          <int>           <dbl>    <dbl>    <dbl>
## 1 All studies                74           9058            11.5     10.8     12.1
## 2 Excluding preserva…        73           9048            11.4     10.8     12.1
## 3 Preservative only           1             10            NA       NA       NA
```

###### 6.4.1.7 Creatinine Completeness Sensitivity

creat_completeness_table("creat")

Table 1: **Creatinine-completeness sensitivity**

Message

Not applicable to creatinine itself

##### 6.4.2 Comparison by Age / Sex / race

###### 6.4.2.1 Plot

```
creat_ma_3rd_analysis <- sex_age_race_subdivision_ma("creat",
                                                      urine_ma2,
                                                      overall_bias)
## Not enough studies for multivariable meta-regression (k = 3, need > 6). Running univariable models instead.
## Not enough studies for multivariable meta-regression (k = 6, need > 6). Running univariable models instead.
creat_ma_3rd_analysis$race_plot
## `height` was translated to `width`.
## `height` was translated to `width`.
## `height` was translated to `width`.
```

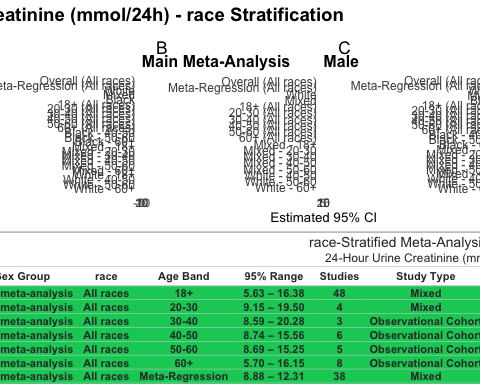

###### 6.4.2.2 Combined MA Results

creat_ma_3rd_analysis$combined_summary_table

Table 1: **Combined Meta-Analysis Results**

24-Hour Urine Creatinine (mmol/24h)

| Sex Group | race | Age Band | Pooled | 95% Range | Studies Mean SD Range | Studies | Study Type | Participants | I² | Cochran's Q | Q (p-value) | Egger's p | Model |
| --- | --- | --- | --- | --- | --- | --- | --- | --- | --- | --- | --- | --- | --- |
| main meta-analysis | All | 18+ | 11.00 | 5.63 – 16.38 | 11.18 ± 2.89 (0.58–16.79) | 48 | Mixed | 4764 | 99.9% | 42505 | <0.005 | <0.001 | Random |
| main meta-analysis | All | 20-30 | 14.32 | 9.15 – 19.50 | 14.40 ± 2.69 (10.56–16.85) | 4 | Mixed | 96 | 99.5% | 599.2 | <0.005 | 0.63 | Random |
| main meta-analysis | All | 30-40 | 14.43 | 8.59 – 20.28 | 14.74 ± 3.26 (11.18–17.58) | 3 | Observational Cohort | 61 | 96% | 49.4 | <0.005 | 0.81 | Random |
| main meta-analysis | All | 40-50 | 12.15 | 8.74 – 15.56 | 12.18 ± 1.70 (10.25–15.27) | 6 | Observational Cohort | 643 | 98.3% | 289.3 | <0.005 | 0.26 | Random |
| main meta-analysis | All | 50-60 | 11.97 | 8.69 – 15.25 | 12.00 ± 1.71 (9.85–13.95) | 5 | Observational Cohort | 2669 | 98.2% | 222.2 | <0.005 | 0.23 | Random |
| main meta-analysis | All | 60+ | 10.93 | 5.70 – 16.15 | 10.93 ± 2.67 (6.94–14.58) | 8 | Observational Cohort | 825 | 99.5% | 1420.8 | <0.005 | <0.05 | Random |
| **main meta-analysis** | **All** | **Meta-Regression** | **10.59** | **8.88 – 12.31** | **—** | **38** | **Mixed** | **3627** | **88.8%** | **165.8** | **<0.005** | **0.27** | **Multivariable** |
| **main meta-analysis** | **All** | **Meta-Regression** | **10.59** | **8.88 – 12.31** | **—** | **38** | **Mixed** | **3627** | **88.8%** | **165.8** | **<0.005** | **0.27** | **Multivariable** |
| **main meta-analysis** | **All** | **Overall** | **11.46** | **6.16 – 16.77** | **11.61 ± 2.84 (0.58–17.58)** | **74** | **Mixed** | **9058** | **99.9%** | **120587.7** | **<0.005** | **<0.001** | **Random** |
| **main meta-analysis** | **All** | **Overall** | **11.46** | **6.16 – 16.77** | **11.61 ± 2.84 (0.58–17.58)** | **74** | **Mixed** | **9058** | **99.9%** | **120587.7** | **<0.005** | **<0.001** | **Random** |
| main meta-analysis | black | 18+ | — | NA – NA | — | NA | NA | NA | — | — | — | — | NA |
| main meta-analysis | black | 20-30 | — | NA – NA | — | NA | NA | NA | — | — | — | — | NA |
| main meta-analysis | black | 30-40 | — | NA – NA | — | NA | NA | NA | — | — | — | — | NA |
| main meta-analysis | black | 40-50 | 12.29 | 7.15 – 17.43 | — (12.29–12.29) | 1 | Observational Cohort | 29 | — | — | — | — | Single study |
| main meta-analysis | black | 50-60 | 12.07 | 6.73 – 17.42 | — (12.07–12.07) | 1 | Observational Cohort | 131 | — | — | — | — | Single study |
| main meta-analysis | black | 60+ | 10.76 | 6.42 – 15.11 | 10.67 ± 0.51 (10.31–11.04) | 2 | Observational Cohort | 144 | 72.4% | 3.6 | 0.06 | — | Fixed |
| **main meta-analysis** | **black** | **Meta-Regression** | **—** | **NA – NA** | **—** | **NA** | **NA** | **NA** | **—** | **—** | **—** | **—** | **NA** |
| **main meta-analysis** | **black** | **Overall** | **11.40** | **9.73 – 13.06** | **11.43 ± 0.92 (10.31–12.29)** | **4** | **Observational Cohort** | **304** | **88.7%** | **26.6** | **<0.005** | **0.89** | **Random** |
| main meta-analysis | mixed | 18+ | 11.11 | 6.80 – 15.41 | 11.34 ± 2.45 (5.91–16.79) | 46 | Mixed | 4183 | 97.5% | 1807.8 | <0.005 | <0.01 | Random |
| main meta-analysis | mixed | 20-30 | 14.32 | 9.15 – 19.50 | 14.40 ± 2.69 (10.56–16.85) | 4 | Mixed | 96 | 99.5% | 599.2 | <0.005 | 0.63 | Random |
| main meta-analysis | mixed | 30-40 | 14.43 | 8.59 – 20.28 | 14.74 ± 3.26 (11.18–17.58) | 3 | Observational Cohort | 61 | 96% | 49.4 | <0.005 | 0.81 | Random |
| main meta-analysis | mixed | 40-50 | 12.33 | 7.24 – 17.42 | 12.34 ± 2.61 (10.25–15.27) | 3 | Observational Cohort | 84 | 99.2% | 265.2 | <0.005 | 0.17 | Random |
| main meta-analysis | mixed | 50-60 | 11.59 | 6.79 – 16.39 | 11.59 ± 2.46 (9.85–13.33) | 2 | Observational Cohort | 50 | 99% | 102.8 | <0.005 | — | Random |
| main meta-analysis | mixed | 60+ | 6.94 | 5.80 – 8.08 | — (6.94–6.94) | 1 | Observational Cohort | 25 | — | — | — | — | Single study |
| **main meta-analysis** | **mixed** | **Meta-Regression** | **—** | **NA – NA** | **—** | **NA** | **NA** | **NA** | **—** | **—** | **—** | **—** | **NA** |
| **main meta-analysis** | **mixed** | **Overall** | **11.52** | **6.64 – 16.40** | **11.71 ± 2.69 (5.91–17.58)** | **59** | **Mixed** | **4499** | **98.9%** | **5155.1** | **<0.005** | **<0.05** | **Random** |
| main meta-analysis | white | 18+ | 7.39 | -11.48 – 26.26 | 7.39 ± 9.63 (0.58–14.20) | 2 | Observational Cohort | 581 | 100% | 2851.6 | <0.005 | — | Random |
| main meta-analysis | white | 20-30 | — | NA – NA | — | NA | NA | NA | — | — | — | — | NA |
| main meta-analysis | white | 30-40 | — | NA – NA | — | NA | NA | NA | — | — | — | — | NA |
| main meta-analysis | white | 40-50 | 11.40 | 6.99 – 15.81 | 11.89 ± 0.69 (11.39–12.38) | 2 | Observational Cohort | 530 | 0% | 0.5 | 0.49 | — | Fixed |
| main meta-analysis | white | 50-60 | 12.33 | 7.96 – 16.69 | 12.36 ± 2.25 (10.77–13.95) | 2 | Observational Cohort | 2488 | 97.8% | 45 | <0.005 | — | Random |
| main meta-analysis | white | 60+ | 11.83 | 6.52 – 17.14 | 11.84 ± 2.72 (8.48–14.58) | 5 | Observational Cohort | 656 | 99.4% | 691.5 | <0.005 | 0.18 | Random |
| **main meta-analysis** | **white** | **Meta-Regression** | **—** | **NA – NA** | **—** | **NA** | **NA** | **NA** | **—** | **—** | **—** | **—** | **NA** |
| **main meta-analysis** | **white** | **Overall** | **11.11** | **3.18 – 19.04** | **11.13 ± 4.03 (0.58–14.58)** | **11** | **Observational Cohort** | **4255** | **100%** | **69897.2** | **<0.005** | **<0.05** | **Random** |
| male | All | 18+ | 13.53 | 11.58 – 15.47 | 13.65 ± 1.21 (12.56–15.22) | 5 | Observational Cohort | 189 | 80.2% | 20.2 | <0.005 | 0.61 | Random |
| male | All | 20-30 | 15.09 | 13.04 – 17.14 | 15.68 ± 1.02 (15.00–16.85) | 3 | Mixed | 71 | 45.5% | 3.7 | 0.16 | 0.05 | Fixed |
| male | All | 30-40 | 15.51 | 12.33 – 18.68 | 16.52 ± 1.49 (15.47–17.58) | 2 | Observational Cohort | 36 | 0% | 1 | 0.32 | — | Fixed |
| male | All | 40-50 | 14.15 | 10.68 – 17.63 | 13.82 ± 2.05 (12.38–15.27) | 2 | Observational Cohort | 31 | 75.1% | 4 | <0.05 | — | Random |
| male | All | 50-60 | 13.43 | 9.57 – 17.30 | 13.64 ± 0.44 (13.33–13.95) | 2 | Observational Cohort | 104 | 31.1% | 1.5 | 0.23 | — | Fixed |
| male | All | 60+ | 13.80 | 12.51 – 15.08 | 13.77 ± 0.70 (13.36–14.58) | 3 | Observational Cohort | 329 | 87% | 15.3 | <0.005 | 0.50 | Random |
| **male** | **All** | **Meta-Regression** | **14.59** | **14.58 – 14.59** | **—** | **3** | **Mixed** | **155** | **0%** | **4.2** | **0.12** | **0.70** | **Univariable** |
| **male** | **All** | **Overall** | **14.23** | **12.16 – 16.29** | **14.39 ± 1.48 (12.38–17.58)** | **17** | **Mixed** | **760** | **91.8%** | **196.2** | **<0.005** | **0.99** | **Random** |
| male | black | 18+ | — | NA – NA | — | NA | NA | NA | — | — | — | — | NA |
| male | black | 20-30 | — | NA – NA | — | NA | NA | NA | — | — | — | — | NA |
| male | black | 30-40 | — | NA – NA | — | NA | NA | NA | — | — | — | — | NA |
| male | black | 40-50 | — | NA – NA | — | NA | NA | NA | — | — | — | — | NA |
| male | black | 50-60 | — | NA – NA | — | NA | NA | NA | — | — | — | — | NA |
| male | black | 60+ | — | NA – NA | — | NA | NA | NA | — | — | — | — | NA |
| **male** | **black** | **Meta-Regression** | **—** | **NA – NA** | **—** | **NA** | **NA** | **NA** | **—** | **—** | **—** | **—** | **NA** |
| **male** | **black** | **Overall** | **—** | **NA – NA** | **—** | **NA** | **NA** | **NA** | **—** | **—** | **—** | **—** | **NA** |
| male | mixed | 18+ | 13.53 | 11.58 – 15.47 | 13.65 ± 1.21 (12.56–15.22) | 5 | Observational Cohort | 189 | 80.2% | 20.2 | <0.005 | 0.61 | Random |
| male | mixed | 20-30 | 15.09 | 13.04 – 17.14 | 15.68 ± 1.02 (15.00–16.85) | 3 | Mixed | 71 | 45.5% | 3.7 | 0.16 | 0.05 | Fixed |
| male | mixed | 30-40 | 15.51 | 12.33 – 18.68 | 16.52 ± 1.49 (15.47–17.58) | 2 | Observational Cohort | 36 | 0% | 1 | 0.32 | — | Fixed |
| male | mixed | 40-50 | 15.27 | 12.31 – 18.23 | — (15.27–15.27) | 1 | Observational Cohort | 25 | — | — | — | — | Single study |
| male | mixed | 50-60 | 13.33 | 11.25 – 15.41 | — (13.33–13.33) | 1 | Observational Cohort | 25 | — | — | — | — | Single study |
| male | mixed | 60+ | — | NA – NA | — | NA | NA | NA | — | — | — | — | NA |
| **male** | **mixed** | **Meta-Regression** | **—** | **NA – NA** | **—** | **NA** | **NA** | **NA** | **—** | **—** | **—** | **—** | **NA** |
| **male** | **mixed** | **Overall** | **14.48** | **12.08 – 16.88** | **14.74 ± 1.58 (12.56–17.58)** | **12** | **Mixed** | **346** | **93.7%** | **173.8** | **<0.005** | **0.73** | **Random** |
| male | white | 18+ | — | NA – NA | — | NA | NA | NA | — | — | — | — | NA |
| male | white | 20-30 | — | NA – NA | — | NA | NA | NA | — | — | — | — | NA |
| male | white | 30-40 | — | NA – NA | — | NA | NA | NA | — | — | — | — | NA |
| male | white | 40-50 | 12.38 | 5.60 – 19.15 | — (12.38–12.38) | 1 | Observational Cohort | 6 | — | — | — | — | Single study |
| male | white | 50-60 | 13.95 | 5.72 – 22.19 | — (13.95–13.95) | 1 | Observational Cohort | 79 | — | — | — | — | Single study |
| male | white | 60+ | 13.80 | 12.51 – 15.08 | 13.77 ± 0.70 (13.36–14.58) | 3 | Observational Cohort | 329 | 87% | 15.3 | <0.005 | 0.50 | Random |
| **male** | **white** | **Meta-Regression** | **—** | **NA – NA** | **—** | **NA** | **NA** | **NA** | **—** | **—** | **—** | **—** | **NA** |
| **male** | **white** | **Overall** | **13.77** | **12.70 – 14.84** | **13.53 ± 0.82 (12.38–14.58)** | **5** | **Observational Cohort** | **414** | **75.8%** | **16.5** | **<0.005** | **0.48** | **Random** |
| female | All | 18+ | 9.04 | 7.92 – 10.16 | 9.03 ± 0.64 (8.62–10.06) | 8 | Observational Cohort | 528 | 82.6% | 40.2 | <0.005 | 0.55 | Random |
| female | All | 20-30 | 10.56 | 9.21 – 11.91 | — (10.56–10.56) | 1 | Observational Cohort | 25 | — | — | — | — | Single study |
| female | All | 30-40 | 11.18 | 5.69 – 16.67 | — (11.18–11.18) | 1 | Observational Cohort | 25 | — | — | — | — | Single study |
| female | All | 40-50 | 11.44 | 7.41 – 15.47 | 11.73 ± 0.49 (11.39–12.29) | 3 | Observational Cohort | 578 | 40.5% | 3.4 | 0.19 | 0.15 | Fixed |
| female | All | 50-60 | 10.90 | 8.78 – 13.02 | 10.90 ± 1.12 (9.85–12.07) | 3 | Observational Cohort | 2565 | 95.2% | 41.5 | <0.005 | 0.87 | Random |
| female | All | 60+ | 9.22 | 6.10 – 12.34 | 9.23 ± 1.60 (6.94–11.04) | 5 | Observational Cohort | 496 | 98.9% | 380.1 | <0.005 | 0.20 | Random |
| **female** | **All** | **Meta-Regression** | **8.70** | **7.46 – 9.94** | **—** | **6** | **Observational Cohort** | **476** | **88.3%** | **37.9** | **<0.005** | **0.20** | **Univariable** |
| **female** | **All** | **Overall** | **9.89** | **7.20 – 12.59** | **9.91 ± 1.40 (6.94–12.29)** | **21** | **Observational Cohort** | **4217** | **98.8%** | **1625.3** | **<0.005** | **0.18** | **Random** |
| female | black | 18+ | — | NA – NA | — | NA | NA | NA | — | — | — | — | NA |
| female | black | 20-30 | — | NA – NA | — | NA | NA | NA | — | — | — | — | NA |
| female | black | 30-40 | — | NA – NA | — | NA | NA | NA | — | — | — | — | NA |
| female | black | 40-50 | 12.29 | 7.15 – 17.43 | — (12.29–12.29) | 1 | Observational Cohort | 29 | — | — | — | — | Single study |
| female | black | 50-60 | 12.07 | 6.73 – 17.42 | — (12.07–12.07) | 1 | Observational Cohort | 131 | — | — | — | — | Single study |
| female | black | 60+ | 10.76 | 6.42 – 15.11 | 10.67 ± 0.51 (10.31–11.04) | 2 | Observational Cohort | 144 | 72.4% | 3.6 | 0.06 | — | Fixed |
| **female** | **black** | **Meta-Regression** | **—** | **NA – NA** | **—** | **NA** | **NA** | **NA** | **—** | **—** | **—** | **—** | **NA** |
| **female** | **black** | **Overall** | **11.40** | **9.73 – 13.06** | **11.43 ± 0.92 (10.31–12.29)** | **4** | **Observational Cohort** | **304** | **88.7%** | **26.6** | **<0.005** | **0.89** | **Random** |
| female | mixed | 18+ | 9.04 | 7.92 – 10.16 | 9.03 ± 0.64 (8.62–10.06) | 8 | Observational Cohort | 528 | 82.6% | 40.2 | <0.005 | 0.55 | Random |
| female | mixed | 20-30 | 10.56 | 9.21 – 11.91 | — (10.56–10.56) | 1 | Observational Cohort | 25 | — | — | — | — | Single study |
| female | mixed | 30-40 | 11.18 | 5.69 – 16.67 | — (11.18–11.18) | 1 | Observational Cohort | 25 | — | — | — | — | Single study |
| female | mixed | 40-50 | 11.50 | 9.66 – 13.34 | — (11.50–11.50) | 1 | Observational Cohort | 25 | — | — | — | — | Single study |
| female | mixed | 50-60 | 9.85 | 7.20 – 12.50 | — (9.85–9.85) | 1 | Observational Cohort | 25 | — | — | — | — | Single study |
| female | mixed | 60+ | 6.94 | 5.80 – 8.08 | — (6.94–6.94) | 1 | Observational Cohort | 25 | — | — | — | — | Single study |
| **female** | **mixed** | **Meta-Regression** | **—** | **NA – NA** | **—** | **NA** | **NA** | **NA** | **—** | **—** | **—** | **—** | **NA** |
| **female** | **mixed** | **Overall** | **9.39** | **6.97 – 11.81** | **9.41 ± 1.26 (6.94–11.50)** | **13** | **Observational Cohort** | **653** | **98.2%** | **684.5** | **<0.005** | **0.29** | **Random** |
| female | white | 18+ | — | NA – NA | — | NA | NA | NA | — | — | — | — | NA |
| female | white | 20-30 | — | NA – NA | — | NA | NA | NA | — | — | — | — | NA |
| female | white | 30-40 | — | NA – NA | — | NA | NA | NA | — | — | — | — | NA |
| female | white | 40-50 | 11.39 | 7.00 – 15.79 | — (11.39–11.39) | 1 | Observational Cohort | 524 | — | — | — | — | Single study |
| female | white | 50-60 | 10.77 | 6.71 – 14.82 | — (10.77–10.77) | 1 | Observational Cohort | 2409 | — | — | — | — | Single study |
| female | white | 60+ | 8.94 | 7.74 – 10.15 | 8.93 ± 0.64 (8.48–9.38) | 2 | Observational Cohort | 327 | 93.2% | 14.7 | <0.005 | — | Random |
| **female** | **white** | **Meta-Regression** | **—** | **NA – NA** | **—** | **NA** | **NA** | **NA** | **—** | **—** | **—** | **—** | **NA** |
| **female** | **white** | **Overall** | **10.01** | **7.45 – 12.58** | **10.01 ± 1.32 (8.48–11.39)** | **4** | **Observational Cohort** | **3260** | **98.9%** | **282.9** | **<0.005** | **0.36** | **Random** |
| *I² represents heterogeneity; Q test assesses between-study variation; Egger's p = linear regression test for funnel plot asymmetry; Meta-Regression shows covariate-adjusted estimates* | | | | | | | | | | | | | |

###### 6.4.2.3 Table of Results subdivided by race / Sex / Age

creat_ma_3rd_analysis$race_summary_table

Table 1: **race-Stratified Meta-Analysis Results**

24-Hour Urine Creatinine (mmol/24h)

| Sex Group | race | Age Band | 95% Range | Studies | Study Type | Participants | I² | Cochran's Q | Q (p-value) | Egger's p | Model |
| --- | --- | --- | --- | --- | --- | --- | --- | --- | --- | --- | --- |
| **main meta-analysis** | **All races** | **18+** | **5.63 – 16.38** | **48** | **Mixed** | **4764** | **99.9%** | **42505** | **<0.005** | **<0.001** | **Random** |
| **main meta-analysis** | **All races** | **20-30** | **9.15 – 19.50** | **4** | **Mixed** | **96** | **99.5%** | **599.2** | **<0.005** | **0.63** | **Random** |
| **main meta-analysis** | **All races** | **30-40** | **8.59 – 20.28** | **3** | **Observational Cohort** | **61** | **96%** | **49.4** | **<0.005** | **0.81** | **Random** |
| **main meta-analysis** | **All races** | **40-50** | **8.74 – 15.56** | **6** | **Observational Cohort** | **643** | **98.3%** | **289.3** | **<0.005** | **0.26** | **Random** |
| **main meta-analysis** | **All races** | **50-60** | **8.69 – 15.25** | **5** | **Observational Cohort** | **2669** | **98.2%** | **222.2** | **<0.005** | **0.23** | **Random** |
| **main meta-analysis** | **All races** | **60+** | **5.70 – 16.15** | **8** | **Observational Cohort** | **825** | **99.5%** | **1420.8** | **<0.005** | **<0.05** | **Random** |
| **main meta-analysis** | **All races** | **Meta-Regression** | **8.88 – 12.31** | **38** | **Mixed** | **3627** | **88.8%** | **165.8** | **<0.005** | **0.27** | **Multivariable** |
| **main meta-analysis** | **All races** | **Meta-Regression** | **8.88 – 12.31** | **38** | **Mixed** | **3627** | **88.8%** | **165.8** | **<0.005** | **0.27** | **Multivariable** |
| **main meta-analysis** | **All races** | **Overall** | **6.16 – 16.77** | **74** | **Mixed** | **9058** | **99.9%** | **120587.7** | **<0.005** | **<0.001** | **Random** |
| **main meta-analysis** | **All races** | **Overall** | **6.16 – 16.77** | **74** | **Mixed** | **9058** | **99.9%** | **120587.7** | **<0.005** | **<0.001** | **Random** |
| main meta-analysis | Black | 18+ | — | — | NA | — | — | — | — | — | — |
| main meta-analysis | Black | 20-30 | — | — | NA | — | — | — | — | — | — |
| main meta-analysis | Black | 30-40 | — | — | NA | — | — | — | — | — | — |
| main meta-analysis | Black | 40-50 | 7.15 – 17.43 | 1 | Observational Cohort | 29 | — | — | — | — | Single study |
| main meta-analysis | Black | 50-60 | 6.73 – 17.42 | 1 | Observational Cohort | 131 | — | — | — | — | Single study |
| main meta-analysis | Black | 60+ | 6.42 – 15.11 | 2 | Observational Cohort | 144 | 72.4% | 3.6 | 0.06 | — | Fixed |
| main meta-analysis | Black | Meta-Regression | — | — | NA | — | — | — | — | — | — |
| **main meta-analysis** | **Black** | **Overall** | **9.73 – 13.06** | **4** | **Observational Cohort** | **304** | **88.7%** | **26.6** | **<0.005** | **0.89** | **Random** |
| main meta-analysis | Mixed | 18+ | 6.80 – 15.41 | 46 | Mixed | 4183 | 97.5% | 1807.8 | <0.005 | <0.01 | Random |
| main meta-analysis | Mixed | 20-30 | 9.15 – 19.50 | 4 | Mixed | 96 | 99.5% | 599.2 | <0.005 | 0.63 | Random |
| main meta-analysis | Mixed | 30-40 | 8.59 – 20.28 | 3 | Observational Cohort | 61 | 96% | 49.4 | <0.005 | 0.81 | Random |
| main meta-analysis | Mixed | 40-50 | 7.24 – 17.42 | 3 | Observational Cohort | 84 | 99.2% | 265.2 | <0.005 | 0.17 | Random |
| main meta-analysis | Mixed | 50-60 | 6.79 – 16.39 | 2 | Observational Cohort | 50 | 99% | 102.8 | <0.005 | — | Random |
| main meta-analysis | Mixed | 60+ | 5.80 – 8.08 | 1 | Observational Cohort | 25 | — | — | — | — | Single study |
| main meta-analysis | Mixed | Meta-Regression | — | — | NA | — | — | — | — | — | — |
| **main meta-analysis** | **Mixed** | **Overall** | **6.64 – 16.40** | **59** | **Mixed** | **4499** | **98.9%** | **5155.1** | **<0.005** | **<0.05** | **Random** |
| main meta-analysis | White | 18+ | -11.48 – 26.26 | 2 | Observational Cohort | 581 | 100% | 2851.6 | <0.005 | — | Random |
| main meta-analysis | White | 20-30 | — | — | NA | — | — | — | — | — | — |
| main meta-analysis | White | 30-40 | — | — | NA | — | — | — | — | — | — |
| main meta-analysis | White | 40-50 | 6.99 – 15.81 | 2 | Observational Cohort | 530 | 0% | 0.5 | 0.49 | — | Fixed |
| main meta-analysis | White | 50-60 | 7.96 – 16.69 | 2 | Observational Cohort | 2488 | 97.8% | 45 | <0.005 | — | Random |
| main meta-analysis | White | 60+ | 6.52 – 17.14 | 5 | Observational Cohort | 656 | 99.4% | 691.5 | <0.005 | 0.18 | Random |
| main meta-analysis | White | Meta-Regression | — | — | NA | — | — | — | — | — | — |
| **main meta-analysis** | **White** | **Overall** | **3.18 – 19.04** | **11** | **Observational Cohort** | **4255** | **100%** | **69897.2** | **<0.005** | **<0.05** | **Random** |
| **male** | **All races** | **18+** | **11.58 – 15.47** | **5** | **Observational Cohort** | **189** | **80.2%** | **20.2** | **<0.005** | **0.61** | **Random** |
| **male** | **All races** | **20-30** | **13.04 – 17.14** | **3** | **Mixed** | **71** | **45.5%** | **3.7** | **0.16** | **0.05** | **Fixed** |
| **male** | **All races** | **30-40** | **12.33 – 18.68** | **2** | **Observational Cohort** | **36** | **0%** | **1** | **0.32** | **—** | **Fixed** |
| **male** | **All races** | **40-50** | **10.68 – 17.63** | **2** | **Observational Cohort** | **31** | **75.1%** | **4** | **<0.05** | **—** | **Random** |
| **male** | **All races** | **50-60** | **9.57 – 17.30** | **2** | **Observational Cohort** | **104** | **31.1%** | **1.5** | **0.23** | **—** | **Fixed** |
| **male** | **All races** | **60+** | **12.51 – 15.08** | **3** | **Observational Cohort** | **329** | **87%** | **15.3** | **<0.005** | **0.50** | **Random** |
| **male** | **All races** | **Meta-Regression** | **14.58 – 14.59** | **3** | **Mixed** | **155** | **0%** | **4.2** | **0.12** | **0.70** | **Univariable** |
| **male** | **All races** | **Overall** | **12.16 – 16.29** | **17** | **Mixed** | **760** | **91.8%** | **196.2** | **<0.005** | **0.99** | **Random** |
| male | Black | 18+ | — | — | NA | — | — | — | — | — | — |
| male | Black | 20-30 | — | — | NA | — | — | — | — | — | — |
| male | Black | 30-40 | — | — | NA | — | — | — | — | — | — |
| male | Black | 40-50 | — | — | NA | — | — | — | — | — | — |
| male | Black | 50-60 | — | — | NA | — | — | — | — | — | — |
| male | Black | 60+ | — | — | NA | — | — | — | — | — | — |
| male | Black | Meta-Regression | — | — | NA | — | — | — | — | — | — |
| **male** | **Black** | **Overall** | **—** | **—** | **NA** | **—** | **—** | **—** | **—** | **—** | **—** |
| male | Mixed | 18+ | 11.58 – 15.47 | 5 | Observational Cohort | 189 | 80.2% | 20.2 | <0.005 | 0.61 | Random |
| male | Mixed | 20-30 | 13.04 – 17.14 | 3 | Mixed | 71 | 45.5% | 3.7 | 0.16 | 0.05 | Fixed |
| male | Mixed | 30-40 | 12.33 – 18.68 | 2 | Observational Cohort | 36 | 0% | 1 | 0.32 | — | Fixed |
| male | Mixed | 40-50 | 12.31 – 18.23 | 1 | Observational Cohort | 25 | — | — | — | — | Single study |
| male | Mixed | 50-60 | 11.25 – 15.41 | 1 | Observational Cohort | 25 | — | — | — | — | Single study |
| male | Mixed | 60+ | — | — | NA | — | — | — | — | — | — |
| male | Mixed | Meta-Regression | — | — | NA | — | — | — | — | — | — |
| **male** | **Mixed** | **Overall** | **12.08 – 16.88** | **12** | **Mixed** | **346** | **93.7%** | **173.8** | **<0.005** | **0.73** | **Random** |
| male | White | 18+ | — | — | NA | — | — | — | — | — | — |
| male | White | 20-30 | — | — | NA | — | — | — | — | — | — |
| male | White | 30-40 | — | — | NA | — | — | — | — | — | — |
| male | White | 40-50 | 5.60 – 19.15 | 1 | Observational Cohort | 6 | — | — | — | — | Single study |
| male | White | 50-60 | 5.72 – 22.19 | 1 | Observational Cohort | 79 | — | — | — | — | Single study |
| male | White | 60+ | 12.51 – 15.08 | 3 | Observational Cohort | 329 | 87% | 15.3 | <0.005 | 0.50 | Random |
| male | White | Meta-Regression | — | — | NA | — | — | — | — | — | — |
| **male** | **White** | **Overall** | **12.70 – 14.84** | **5** | **Observational Cohort** | **414** | **75.8%** | **16.5** | **<0.005** | **0.48** | **Random** |
| **female** | **All races** | **18+** | **7.92 – 10.16** | **8** | **Observational Cohort** | **528** | **82.6%** | **40.2** | **<0.005** | **0.55** | **Random** |
| **female** | **All races** | **20-30** | **9.21 – 11.91** | **1** | **Observational Cohort** | **25** | **—** | **—** | **—** | **—** | **Single study** |
| **female** | **All races** | **30-40** | **5.69 – 16.67** | **1** | **Observational Cohort** | **25** | **—** | **—** | **—** | **—** | **Single study** |
| **female** | **All races** | **40-50** | **7.41 – 15.47** | **3** | **Observational Cohort** | **578** | **40.5%** | **3.4** | **0.19** | **0.15** | **Fixed** |
| **female** | **All races** | **50-60** | **8.78 – 13.02** | **3** | **Observational Cohort** | **2565** | **95.2%** | **41.5** | **<0.005** | **0.87** | **Random** |
| **female** | **All races** | **60+** | **6.10 – 12.34** | **5** | **Observational Cohort** | **496** | **98.9%** | **380.1** | **<0.005** | **0.20** | **Random** |
| **female** | **All races** | **Meta-Regression** | **7.46 – 9.94** | **6** | **Observational Cohort** | **476** | **88.3%** | **37.9** | **<0.005** | **0.20** | **Univariable** |
| **female** | **All races** | **Overall** | **7.20 – 12.59** | **21** | **Observational Cohort** | **4217** | **98.8%** | **1625.3** | **<0.005** | **0.18** | **Random** |
| female | Black | 18+ | — | — | NA | — | — | — | — | — | — |
| female | Black | 20-30 | — | — | NA | — | — | — | — | — | — |
| female | Black | 30-40 | — | — | NA | — | — | — | — | — | — |
| female | Black | 40-50 | 7.15 – 17.43 | 1 | Observational Cohort | 29 | — | — | — | — | Single study |
| female | Black | 50-60 | 6.73 – 17.42 | 1 | Observational Cohort | 131 | — | — | — | — | Single study |
| female | Black | 60+ | 6.42 – 15.11 | 2 | Observational Cohort | 144 | 72.4% | 3.6 | 0.06 | — | Fixed |
| female | Black | Meta-Regression | — | — | NA | — | — | — | — | — | — |
| **female** | **Black** | **Overall** | **9.73 – 13.06** | **4** | **Observational Cohort** | **304** | **88.7%** | **26.6** | **<0.005** | **0.89** | **Random** |
| female | Mixed | 18+ | 7.92 – 10.16 | 8 | Observational Cohort | 528 | 82.6% | 40.2 | <0.005 | 0.55 | Random |
| female | Mixed | 20-30 | 9.21 – 11.91 | 1 | Observational Cohort | 25 | — | — | — | — | Single study |
| female | Mixed | 30-40 | 5.69 – 16.67 | 1 | Observational Cohort | 25 | — | — | — | — | Single study |
| female | Mixed | 40-50 | 9.66 – 13.34 | 1 | Observational Cohort | 25 | — | — | — | — | Single study |
| female | Mixed | 50-60 | 7.20 – 12.50 | 1 | Observational Cohort | 25 | — | — | — | — | Single study |
| female | Mixed | 60+ | 5.80 – 8.08 | 1 | Observational Cohort | 25 | — | — | — | — | Single study |
| female | Mixed | Meta-Regression | — | — | NA | — | — | — | — | — | — |
| **female** | **Mixed** | **Overall** | **6.97 – 11.81** | **13** | **Observational Cohort** | **653** | **98.2%** | **684.5** | **<0.005** | **0.29** | **Random** |
| female | White | 18+ | — | — | NA | — | — | — | — | — | — |
| female | White | 20-30 | — | — | NA | — | — | — | — | — | — |
| female | White | 30-40 | — | — | NA | — | — | — | — | — | — |
| female | White | 40-50 | 7.00 – 15.79 | 1 | Observational Cohort | 524 | — | — | — | — | Single study |
| female | White | 50-60 | 6.71 – 14.82 | 1 | Observational Cohort | 2409 | — | — | — | — | Single study |
| female | White | 60+ | 7.74 – 10.15 | 2 | Observational Cohort | 327 | 93.2% | 14.7 | <0.005 | — | Random |
| female | White | Meta-Regression | — | — | NA | — | — | — | — | — | — |
| **female** | **White** | **Overall** | **7.45 – 12.58** | **4** | **Observational Cohort** | **3260** | **98.9%** | **282.9** | **<0.005** | **0.36** | **Random** |
| *I² represents heterogeneity; Q test assesses between-study variation; Egger's p = linear regression test for funnel plot asymmetry; — indicates insufficient data* | | | | | | | | | | | |

###### 6.4.2.4 Wald Tests

creat_ma_3rd_analysis$wald_tests$sex_comparison

Table 1: **Sex Comparison (Overall)**

| Comparison | % Shift | P (Wald Test) | Mean Diff (95% CI) |
| --- | --- | --- | --- |
| Male vs Female (Overall) | +43.8% | <0.05 | 4.33 [0.94, 7.73] |
| *Mean difference = Group 1 − Group 2 in component units (mmol/24h, L/24h, or pH); % shift = difference relative to Group 2.* | | | |

creat_ma_3rd_analysis$wald_tests$race_comparisons

Table 1: **race Comparisons Within Sex Groups**

| Sex Group | Comparison | % Shift | P (Wald Test) | Mean Diff (95% CI) |
| --- | --- | --- | --- | --- |
| male | black vs mixed | — | — | — |
| male | black vs white | — | — | — |
| male | mixed vs white | +5.1% | 0.599 | 0.71 [-1.92, 3.33] |
| female | black vs mixed | +21.4% | 0.181 | 2.01 [-0.93, 4.94] |
| female | black vs white | +13.8% | 0.376 | 1.38 [-1.68, 4.44] |
| female | mixed vs white | -6.2% | 0.729 | -0.62 [-4.15, 2.90] |
| *Mean difference = Group 1 − Group 2 in component units (mmol/24h, L/24h, or pH); % shift = difference relative to Group 2.* | | | | |

creat_ma_3rd_analysis$wald_tests$age_band_comparisons

Table 1: **Age Band Comparisons Within Sex Groups**

| Sex Group | Comparison | % Shift | P (Wald Test) | Mean Diff (95% CI) |
| --- | --- | --- | --- | --- |
| main meta-analysis | 18+ vs 20-30 | -23.2% | 0.383 | -3.32 [-10.78, 4.14] |
| main meta-analysis | 18+ vs 30-40 | -23.8% | 0.397 | -3.43 [-11.37, 4.51] |
| main meta-analysis | 18+ vs 40-50 | -9.5% | 0.724 | -1.15 [-7.52, 5.22] |
| main meta-analysis | 18+ vs 50-60 | -8.1% | 0.763 | -0.97 [-7.27, 5.33] |
| main meta-analysis | 18+ vs 60+ | +0.7% | 0.984 | 0.08 [-7.42, 7.57] |
| main meta-analysis | 20-30 vs 30-40 | -0.8% | 0.978 | -0.11 [-7.92, 7.70] |
| main meta-analysis | 20-30 vs 40-50 | +17.9% | 0.492 | 2.17 [-4.03, 8.37] |
| main meta-analysis | 20-30 vs 50-60 | +19.6% | 0.452 | 2.35 [-3.78, 8.47] |
| main meta-analysis | 20-30 vs 60+ | +31.1% | 0.365 | 3.39 [-3.96, 10.75] |
| main meta-analysis | 30-40 vs 40-50 | +18.8% | 0.509 | 2.28 [-4.49, 9.05] |
| main meta-analysis | 30-40 vs 50-60 | +20.5% | 0.472 | 2.46 [-4.24, 9.16] |
| main meta-analysis | 30-40 vs 60+ | +32.1% | 0.381 | 3.50 [-4.33, 11.34] |
| main meta-analysis | 40-50 vs 50-60 | +1.5% | 0.941 | 0.18 [-4.55, 4.91] |
| main meta-analysis | 40-50 vs 60+ | +11.2% | 0.701 | 1.22 [-5.01, 7.46] |
| main meta-analysis | 50-60 vs 60+ | +9.6% | 0.740 | 1.05 [-5.12, 7.21] |
| male | 18+ vs 20-30 | -10.3% | 0.279 | -1.56 [-4.39, 1.27] |
| male | 18+ vs 30-40 | -12.7% | 0.298 | -1.98 [-5.70, 1.74] |
| male | 18+ vs 40-50 | -4.4% | 0.759 | -0.62 [-4.61, 3.36] |
| male | 18+ vs 50-60 | +0.7% | 0.966 | 0.09 [-4.23, 4.42] |
| male | 18+ vs 60+ | -1.9% | 0.822 | -0.27 [-2.60, 2.07] |
| male | 20-30 vs 30-40 | -2.7% | 0.829 | -0.42 [-4.19, 3.36] |
| male | 20-30 vs 40-50 | +6.6% | 0.649 | 0.94 [-3.10, 4.97] |
| male | 20-30 vs 50-60 | +12.3% | 0.459 | 1.65 [-2.72, 6.03] |
| male | 20-30 vs 60+ | +9.4% | 0.295 | 1.29 [-1.13, 3.71] |
| male | 30-40 vs 40-50 | +9.6% | 0.573 | 1.35 [-3.35, 6.06] |
| male | 30-40 vs 50-60 | +15.4% | 0.417 | 2.07 [-2.93, 7.07] |
| male | 30-40 vs 60+ | +12.4% | 0.328 | 1.71 [-1.71, 5.13] |
| male | 40-50 vs 50-60 | +5.3% | 0.787 | 0.72 [-4.48, 5.92] |
| male | 40-50 vs 60+ | +2.6% | 0.851 | 0.36 [-3.35, 4.06] |
| male | 50-60 vs 60+ | -2.6% | 0.862 | -0.36 [-4.44, 3.71] |
| female | 18+ vs 20-30 | -14.4% | 0.090 | -1.52 [-3.28, 0.24] |
| female | 18+ vs 30-40 | -19.2% | 0.454 | -2.14 [-7.74, 3.46] |
| female | 18+ vs 40-50 | -21.0% | 0.260 | -2.41 [-6.59, 1.78] |
| female | 18+ vs 50-60 | -17.1% | 0.128 | -1.86 [-4.26, 0.53] |
| female | 18+ vs 60+ | -2.0% | 0.915 | -0.18 [-3.50, 3.14] |
| female | 20-30 vs 30-40 | -5.5% | 0.830 | -0.62 [-6.27, 5.03] |
| female | 20-30 vs 40-50 | -7.7% | 0.684 | -0.88 [-5.13, 3.37] |
| female | 20-30 vs 50-60 | -3.1% | 0.790 | -0.34 [-2.85, 2.17] |
| female | 20-30 vs 60+ | +14.5% | 0.440 | 1.34 [-2.06, 4.74] |
| female | 30-40 vs 40-50 | -2.3% | 0.940 | -0.26 [-7.07, 6.54] |
| female | 30-40 vs 50-60 | +2.6% | 0.926 | 0.28 [-5.60, 6.16] |
| female | 30-40 vs 60+ | +21.3% | 0.543 | 1.96 [-4.35, 8.27] |
| female | 40-50 vs 50-60 | +5.0% | 0.815 | 0.54 [-4.01, 5.09] |
| female | 40-50 vs 60+ | +24.1% | 0.392 | 2.22 [-2.87, 7.32] |
| female | 50-60 vs 60+ | +18.2% | 0.382 | 1.68 [-2.09, 5.45] |
| *Mean difference = Group 1 − Group 2 in component units (mmol/24h, L/24h, or pH); % shift = difference relative to Group 2.* | | | | |

creat_ma_3rd_analysis$wald_tests$sex_within_age_bands

Table 1: **Sex Comparisons Within Age Bands**

| Age Band | % Shift | P (Wald Test) | Mean Diff (95% CI) |
| --- | --- | --- | --- |
| 18+ | +49.7% | <0.001 | 4.49 [2.24, 6.74] |
| 20-30 | +42.9% | <0.001 | 4.53 [2.07, 6.98] |
| 30-40 | +38.7% | 0.181 | 4.33 [-2.01, 10.66] |
| 40-50 | +23.7% | 0.318 | 2.71 [-2.61, 8.03] |
| 50-60 | +23.2% | 0.260 | 2.53 [-1.87, 6.94] |
| 60+ | +49.6% | <0.01 | 4.58 [1.20, 7.95] |
| *Mean difference = Group 1 − Group 2 in component units (mmol/24h, L/24h, or pH); % shift = difference relative to Group 2.* | | | |

#### 6.5 Oxalate

##### 6.5.1 Comparison by Age Band

###### 6.5.1.1 Plot

```
oxalate_ma_2nd_analysis <- sex_age_subdivision_ma("oxalate")
## Not enough studies for multivariable meta-regression (k = 5, need > 6). Running univariable models instead.
oxalate_ma_2nd_analysis$combined_plot
## `height` was translated to `width`.
## `height` was translated to `width`.
## `height` was translated to `width`.
```

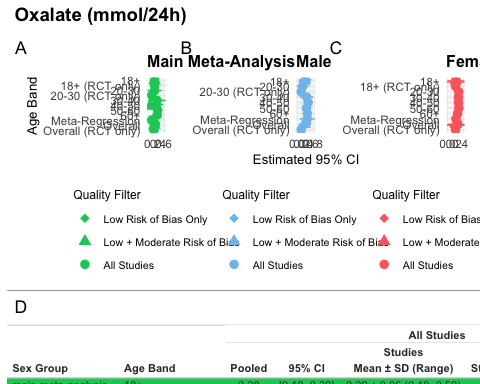

###### 6.5.1.2 Summary Table

oxalate_ma_2nd_analysis$combined_summary_table

Table 1: **Combined Meta-Analysis Results**

24-Hour Urine Oxalate (mmol/24h)

|  |  | **All Studies** | | | | | | **Low Risk of Bias Only** | | | | | | **Low + Moderate RoB** | | | | | |
| --- | --- | --- | --- | --- | --- | --- | --- | --- | --- | --- | --- | --- | --- | --- | --- | --- | --- | --- | --- |
| Sex Group | Age Band | Pooled | 95% CI | Studies  Mean ± SD (Range) | Studies, n | Q p-value | Egger's p | Pooled | 95% CI | Studies  Mean ± SD (Range) | Studies, n | Q p-value | Egger's p | Pooled | 95% CI | Studies  Mean ± SD (Range) | Studies, n | Q p-value | Egger's p |
| main meta-analysis | 18+ | 0.28 | [0.18, 0.39] | 0.29 ± 0.06 (0.18–0.59) | 63 | <0.001 | 0.41 | 0.27 | [0.23, 0.30] | 0.27 ± 0.03 (0.22–0.34) | 28 | <0.001 | 0.38 | 0.28 | [0.20, 0.35] | 0.28 ± 0.04 (0.22–0.41) | 35 | <0.001 | 0.19 |
| **main meta-analysis** | **18+ (RCT only)** | **0.28** | **[0.15, 0.41]** | **0.29 ± 0.07 (0.18–0.37)** | **6** | **<0.001** | **0.28** | **—** | **—** | **—** | **—** | **—** | **—** | **—** | **—** | **—** | **—** | **—** | **—** |
| main meta-analysis | 20-30 | 0.28 | [0.04, 0.51] | 0.28 ± 0.12 (0.17–0.40) | 4 | <0.001 | 0.20 | — | — | — | — | — | — | — | — | — | — | — | — |
| **main meta-analysis** | **20-30 (RCT only)** | **0.18** | **[0.10, 0.26]** | **0.17 ± 0.01 (0.17–0.18)** | **2** | **0.45** | **—** | **—** | **—** | **—** | **—** | **—** | **—** | **—** | **—** | **—** | **—** | **—** | **—** |
| main meta-analysis | 30-40 | 0.36 | [0.07, 0.65] | 0.37 ± 0.16 (0.18–0.60) | 5 | <0.001 | 0.66 | — | — | — | — | — | — | — | — | — | — | — | — |
| main meta-analysis | 40-50 | 0.34 | [0.11, 0.57] | 0.34 ± 0.11 (0.12–0.50) | 7 | <0.001 | 0.07 | — | — | — | — | — | — | 0.12 | [0.10, 0.14] | — (0.12–0.12) | 1 | — | — |
| main meta-analysis | 50-60 | 0.38 | [0.30, 0.47] | 0.39 ± 0.04 (0.34–0.44) | 5 | <0.001 | 0.20 | — | — | — | — | — | — | — | — | — | — | — | — |
| main meta-analysis | 60+ | 0.37 | [0.22, 0.52] | 0.37 ± 0.08 (0.29–0.49) | 8 | <0.001 | 0.33 | — | — | — | — | — | — | — | — | — | — | — | — |
| **main meta-analysis** | **Meta-Regression** | **0.17** | **[0.07, 0.27]** | **—** | **54** | **<0.001** | **0.22** | **—** | **—** | **—** | **—** | **—** | **—** | **—** | **—** | **—** | **—** | **—** | **—** |
| **main meta-analysis** | **Overall** | **0.31** | **[0.15, 0.46]** | **0.31 ± 0.08 (0.12–0.60)** | **92** | **<0.001** | **<0.05** | **0.27** | **[0.23, 0.30]** | **0.27 ± 0.03 (0.22–0.34)** | **28** | **<0.001** | **0.38** | **0.27** | **[0.18, 0.36]** | **0.27 ± 0.05 (0.12–0.41)** | **36** | **<0.001** | **<0.01** |
| **main meta-analysis** | **Overall (RCT only)** | **0.25** | **[0.11, 0.39]** | **0.26 ± 0.08 (0.17–0.37)** | **8** | **<0.001** | **<0.05** | **—** | **—** | **—** | **—** | **—** | **—** | **—** | **—** | **—** | **—** | **—** | **—** |
| male | 18+ | 0.37 | [0.09, 0.65] | 0.37 ± 0.15 (0.26–0.59) | 4 | <0.001 | <0.05 | 0.34 | [0.08, 0.59] | — (0.34–0.34) | 1 | — | — | 0.34 | [0.08, 0.59] | — (0.34–0.34) | 1 | — | — |
| **male** | **18+ (RCT only)** | **—** | **—** | **—** | **—** | **—** | **—** | **—** | **—** | **—** | **—** | **—** | **—** | **—** | **—** | **—** | **—** | **—** | **—** |
| male | 20-30 | 0.25 | [-0.01, 0.51] | 0.25 ± 0.13 (0.17–0.40) | 3 | <0.001 | 0.42 | — | — | — | — | — | — | — | — | — | — | — | — |
| **male** | **20-30 (RCT only)** | **0.18** | **[0.10, 0.26]** | **0.17 ± 0.01 (0.17–0.18)** | **2** | **0.45** | **—** | **—** | **—** | **—** | **—** | **—** | **—** | **—** | **—** | **—** | **—** | **—** | **—** |
| male | 30-40 | 0.43 | [0.15, 0.72] | 0.45 ± 0.16 (0.29–0.60) | 3 | <0.001 | 0.44 | — | — | — | — | — | — | — | — | — | — | — | — |
| male | 40-50 | 0.50 | [0.41, 0.59] | 0.44 ± 0.08 (0.38–0.50) | 2 | 0.06 | — | — | — | — | — | — | — | — | — | — | — | — | — |
| male | 50-60 | 0.43 | [0.30, 0.55] | 0.43 ± 0.01 (0.42–0.44) | 2 | 0.48 | — | — | — | — | — | — | — | — | — | — | — | — | — |
| male | 60+ | 0.46 | [0.40, 0.52] | 0.46 ± 0.03 (0.43–0.49) | 3 | <0.001 | 0.32 | — | — | — | — | — | — | — | — | — | — | — | — |
| **male** | **Meta-Regression** | **0.46** | **[0.13, 0.78]** | **—** | **5** | **<0.001** | **0.70** | **—** | **—** | **—** | **—** | **—** | **—** | **—** | **—** | **—** | **—** | **—** | **—** |
| **male** | **Overall** | **0.39** | **[0.16, 0.62]** | **0.39 ± 0.12 (0.17–0.60)** | **17** | **<0.001** | **0.05** | **0.34** | **[0.08, 0.59]** | **— (0.34–0.34)** | **1** | **—** | **—** | **0.34** | **[0.08, 0.59]** | **— (0.34–0.34)** | **1** | **—** | **—** |
| **male** | **Overall (RCT only)** | **0.18** | **[0.10, 0.26]** | **0.17 ± 0.01 (0.17–0.18)** | **2** | **0.45** | **—** | **—** | **—** | **—** | **—** | **—** | **—** | **—** | **—** | **—** | **—** | **—** | **—** |
| female | 18+ | 0.26 | [0.22, 0.30] | 0.27 ± 0.03 (0.24–0.31) | 7 | <0.01 | 0.10 | 0.26 | [-0.01, 0.53] | 0.26 ± 0.00 (0.26–0.26) | 2 | 1.00 | — | 0.26 | [0.01, 0.51] | 0.26 ± 0.01 (0.25–0.26) | 3 | 0.86 | <0.001 |
| **female** | **18+ (RCT only)** | **0.25** | **[0.14, 0.36]** | **— (0.25–0.25)** | **1** | **—** | **—** | **—** | **—** | **—** | **—** | **—** | **—** | **—** | **—** | **—** | **—** | **—** | **—** |
| female | 20-30 | 0.35 | [0.30, 0.41] | — (0.35–0.35) | 1 | — | — | — | — | — | — | — | — | — | — | — | — | — | — |
| **female** | **20-30 (RCT only)** | **—** | **—** | **—** | **—** | **—** | **—** | **—** | **—** | **—** | **—** | **—** | **—** | **—** | **—** | **—** | **—** | **—** | **—** |
| female | 30-40 | 0.34 | [0.29, 0.39] | — (0.34–0.34) | 1 | — | — | — | — | — | — | — | — | — | — | — | — | — | — |
| female | 40-50 | 0.34 | [0.29, 0.40] | 0.34 ± 0.03 (0.32–0.38) | 3 | <0.001 | 0.86 | — | — | — | — | — | — | — | — | — | — | — | — |
| female | 50-60 | 0.35 | [0.31, 0.39] | 0.36 ± 0.02 (0.34–0.38) | 3 | <0.001 | 0.61 | — | — | — | — | — | — | — | — | — | — | — | — |
| female | 60+ | 0.32 | [0.28, 0.35] | 0.32 ± 0.02 (0.29–0.34) | 5 | <0.001 | 0.20 | — | — | — | — | — | — | — | — | — | — | — | — |
| **female** | **Meta-Regression** | **0.26** | **[0.20, 0.32]** | **—** | **7** | **<0.005** | **0.10** | **—** | **—** | **—** | **—** | **—** | **—** | **—** | **—** | **—** | **—** | **—** | **—** |
| **female** | **Overall** | **0.31** | **[0.23, 0.40]** | **0.31 ± 0.04 (0.24–0.38)** | **20** | **<0.001** | **0.10** | **0.26** | **[-0.01, 0.53]** | **0.26 ± 0.00 (0.26–0.26)** | **2** | **1.00** | **—** | **0.26** | **[0.01, 0.51]** | **0.26 ± 0.01 (0.25–0.26)** | **3** | **0.86** | **<0.001** |
| **female** | **Overall (RCT only)** | **0.25** | **[0.14, 0.36]** | **— (0.25–0.25)** | **1** | **—** | **—** | **—** | **—** | **—** | **—** | **—** | **—** | **—** | **—** | **—** | **—** | **—** | **—** |
| *Studies, n = number of studies; Q p-value = Cochran's Q heterogeneity test; Egger's p = linear regression test for funnel plot asymmetry; RoB = Risk of Bias* | | | | | | | | | | | | | | | | | | | |

###### 6.5.1.3 Wald Tests

oxalate_ma_2nd_analysis$wald_tests$sex_comparison

Table 1: **Sex Comparison (Overall)**

| Comparison | % Shift | P (Wald Test) | Mean Diff (95% CI) |
| --- | --- | --- | --- |
| Male vs Female (Overall) | +24.8% | 0.541 | 0.08 [-0.17, 0.33] |
| *Mean difference = Group 1 − Group 2 in component units (mmol/24h, L/24h, or pH); % shift = difference relative to Group 2.* | | | |

oxalate_ma_2nd_analysis$wald_tests$age_band_comparisons

Table 1: **Age Band Comparisons Within Sex Groups**

| Sex Group | Comparison | % Shift | P (Wald Test) | Mean Diff (95% CI) |
| --- | --- | --- | --- | --- |
| main meta-analysis | 18+ vs 20-30 | +2.5% | 0.958 | 0.01 [-0.25, 0.26] |
| main meta-analysis | 18+ vs 30-40 | -21.6% | 0.617 | -0.08 [-0.39, 0.23] |
| main meta-analysis | 18+ vs 40-50 | -16.8% | 0.652 | -0.06 [-0.31, 0.19] |
| main meta-analysis | 18+ vs 50-60 | -26.2% | 0.139 | -0.10 [-0.23, 0.03] |
| main meta-analysis | 18+ vs 60+ | -23.3% | 0.351 | -0.09 [-0.27, 0.09] |
| main meta-analysis | 20-30 vs 30-40 | -23.5% | 0.653 | -0.09 [-0.46, 0.29] |
| main meta-analysis | 20-30 vs 40-50 | -18.8% | 0.699 | -0.06 [-0.39, 0.26] |
| main meta-analysis | 20-30 vs 50-60 | -28.0% | 0.395 | -0.11 [-0.36, 0.14] |
| main meta-analysis | 20-30 vs 60+ | -25.1% | 0.511 | -0.09 [-0.37, 0.18] |
| main meta-analysis | 30-40 vs 40-50 | +6.2% | 0.910 | 0.02 [-0.35, 0.39] |
| main meta-analysis | 30-40 vs 50-60 | -5.9% | 0.883 | -0.02 [-0.32, 0.28] |
| main meta-analysis | 30-40 vs 60+ | -2.1% | 0.963 | -0.01 [-0.33, 0.32] |
| main meta-analysis | 40-50 vs 50-60 | -11.4% | 0.723 | -0.04 [-0.29, 0.20] |
| main meta-analysis | 40-50 vs 60+ | -7.8% | 0.835 | -0.03 [-0.30, 0.24] |
| main meta-analysis | 50-60 vs 60+ | +4.0% | 0.864 | 0.01 [-0.16, 0.19] |
| male | 18+ vs 20-30 | +47.0% | 0.542 | 0.12 [-0.26, 0.50] |
| male | 18+ vs 30-40 | -14.9% | 0.749 | -0.06 [-0.46, 0.33] |
| male | 18+ vs 40-50 | -25.4% | 0.398 | -0.13 [-0.42, 0.17] |
| male | 18+ vs 50-60 | -13.3% | 0.714 | -0.06 [-0.36, 0.25] |
| male | 18+ vs 60+ | -19.6% | 0.535 | -0.09 [-0.37, 0.19] |
| male | 20-30 vs 30-40 | -42.1% | 0.352 | -0.18 [-0.57, 0.20] |
| male | 20-30 vs 40-50 | -49.3% | 0.081 | -0.24 [-0.52, 0.03] |
| male | 20-30 vs 50-60 | -41.0% | 0.232 | -0.18 [-0.46, 0.11] |
| male | 20-30 vs 60+ | -45.3% | 0.124 | -0.21 [-0.47, 0.06] |
| male | 30-40 vs 40-50 | -12.3% | 0.689 | -0.06 [-0.36, 0.24] |
| male | 30-40 vs 50-60 | +1.9% | 0.959 | 0.01 [-0.30, 0.32] |
| male | 30-40 vs 60+ | -5.4% | 0.866 | -0.02 [-0.32, 0.27] |
| male | 40-50 vs 50-60 | +16.2% | 0.377 | 0.07 [-0.08, 0.22] |
| male | 40-50 vs 60+ | +7.9% | 0.508 | 0.04 [-0.07, 0.14] |
| male | 50-60 vs 60+ | -7.2% | 0.634 | -0.03 [-0.17, 0.10] |
| female | 18+ vs 20-30 | -25.7% | <0.05 | -0.09 [-0.16, -0.02] |
| female | 18+ vs 30-40 | -22.8% | <0.05 | -0.08 [-0.14, -0.02] |
| female | 18+ vs 40-50 | -23.7% | <0.05 | -0.08 [-0.15, -0.01] |
| female | 18+ vs 50-60 | -25.9% | <0.01 | -0.09 [-0.15, -0.03] |
| female | 18+ vs 60+ | -16.8% | 0.061 | -0.05 [-0.11, 0.00] |
| female | 20-30 vs 30-40 | +3.8% | 0.725 | 0.01 [-0.06, 0.09] |
| female | 20-30 vs 40-50 | +2.7% | 0.821 | 0.01 [-0.07, 0.09] |
| female | 20-30 vs 50-60 | -0.4% | 0.970 | -0.00 [-0.07, 0.07] |
| female | 20-30 vs 60+ | +12.0% | 0.276 | 0.04 [-0.03, 0.11] |
| female | 30-40 vs 40-50 | -1.1% | 0.916 | -0.00 [-0.08, 0.07] |
| female | 30-40 vs 50-60 | -4.0% | 0.642 | -0.01 [-0.07, 0.05] |
| female | 30-40 vs 60+ | +7.8% | 0.407 | 0.02 [-0.03, 0.08] |
| female | 40-50 vs 50-60 | -3.0% | 0.763 | -0.01 [-0.08, 0.06] |
| female | 40-50 vs 60+ | +9.0% | 0.401 | 0.03 [-0.04, 0.10] |
| female | 50-60 vs 60+ | +12.4% | 0.162 | 0.04 [-0.02, 0.09] |
| *Mean difference = Group 1 − Group 2 in component units (mmol/24h, L/24h, or pH); % shift = difference relative to Group 2.* | | | | |

oxalate_ma_2nd_analysis$wald_tests$sex_within_age_bands

Table 1: **Sex Comparisons Within Age Bands**

| Age Band | % Shift | P (Wald Test) | Mean Diff (95% CI) |
| --- | --- | --- | --- |
| 18+ | +40.9% | 0.454 | 0.11 [-0.17, 0.39] |
| 20-30 | -28.7% | 0.453 | -0.10 [-0.37, 0.16] |
| 30-40 | +27.9% | 0.520 | 0.09 [-0.19, 0.38] |
| 40-50 | +44.2% | <0.01 | 0.15 [0.05, 0.26] |
| 50-60 | +20.4% | 0.277 | 0.07 [-0.06, 0.20] |
| 60+ | +45.8% | <0.001 | 0.14 [0.08, 0.21] |
| *Mean difference = Group 1 − Group 2 in component units (mmol/24h, L/24h, or pH); % shift = difference relative to Group 2.* | | | |

###### 6.5.1.4 Meta-Regression (plain language)

format_meta_reg_plain("oxalate", sex_group = "all")

Message

No meta-regression result for oxalate / all

###### 6.5.1.5 Assay Subgroup Analysis

assay_subgroup_table("oxalate")

Table 1: **Assay subgroup analysis — Oxalate**

*Per-assay pooled estimates; categories with k < 3 listed but not pooled.*

| Assay category | Studies, n | Participants | 95% CI | I² | Q p-value | Egger's p | Note |
| --- | --- | --- | --- | --- | --- | --- | --- |
| Unreported | 79 | 9,139 | 0.31 [0.30, 0.33] | 98% | <0.001 | 0.954 |  |
| Enzymatic (other) | 6 | 258 | 0.31 [0.25, 0.37] | 99% | <0.001 | 0.931 |  |
| Enzymatic (oxalase) | 2 | 201 | — | — | — | — | k = 2 < min_k = 3 (skipped) |
| HPLC | 2 | 72 | — | — | — | — | k = 2 < min_k = 3 (skipped) |
| Ion chromatography | 2 | 33 | — | — | — | — | k = 2 < min_k = 3 (skipped) |
| Capillary electrophoresis | 1 | 22 | — | — | — | — | k = 1 < min_k = 3 (skipped) |

###### 6.5.1.6 Preservative Sensitivity (Litholink / acidified samples)

```
preservative_sensitivity_ma("oxalate")
## # A tibble: 3 × 6
##   group               n_studies n_participants pooled_estimate lower_95 upper_95
##   <chr>                   <int>          <int>           <dbl>    <dbl>    <dbl>
## 1 All studies                92           9725           0.306    0.289    0.322
## 2 Excluding preserva…        89           9614           0.303    0.286    0.319
## 3 Preservative only           3            111           0.416    0.382    0.449
```

###### 6.5.1.7 Creatinine Completeness Sensitivity

creat_completeness_table("oxalate")

Table 1: **Creatinine-completeness sensitivity — Oxalate**

*Compares the all-studies pooled estimate against subsets defined by whether the study reported 24h urine creatinine. Studies that do not report creatinine cannot verify collection completeness; a substantial shift here flags possible bias from incomplete collections.*

| Subset | Studies, n | Participants | Range (95% CI) | Δ vs all-studies (%) | Flag |
| --- | --- | --- | --- | --- | --- |
| All studies | 92 | 9,725 | 0.31 [0.29, 0.32] | — | — |
| Creatinine reported only | 62 | 7,822 | 0.31 [0.29, 0.33] | +0.1% | OK |
| Creatinine NOT reported | 30 | 1,903 | 0.30 [0.28, 0.33] | -0.3% | OK |

##### 6.5.2 Comparison by Age / Sex / race

###### 6.5.2.1 Plot

```
oxalate_ma_3rd_analysis <- sex_age_race_subdivision_ma("oxalate",
                                                      urine_ma2,
                                                      overall_bias)
## Not enough studies for multivariable meta-regression (k = 5, need > 6). Running univariable models instead.
oxalate_ma_3rd_analysis$race_plot
## `height` was translated to `width`.
## `height` was translated to `width`.
## `height` was translated to `width`.
```

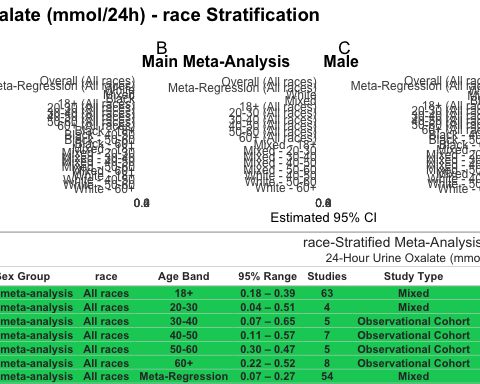

###### 6.5.2.2 Combined MA Results

oxalate_ma_3rd_analysis$combined_summary_table

Table 1: **Combined Meta-Analysis Results**

24-Hour Urine Oxalate (mmol/24h)

| Sex Group | race | Age Band | Pooled | 95% Range | Studies Mean SD Range | Studies | Study Type | Participants | I² | Cochran's Q | Q (p-value) | Egger's p | Model |
| --- | --- | --- | --- | --- | --- | --- | --- | --- | --- | --- | --- | --- | --- |
| main meta-analysis | All | 18+ | 0.28 | 0.18 – 0.39 | 0.29 ± 0.06 (0.18–0.59) | 63 | Mixed | 5285 | 92.5% | 829 | <0.005 | 0.41 | Random |
| main meta-analysis | All | 20-30 | 0.28 | 0.04 – 0.51 | 0.28 ± 0.12 (0.17–0.40) | 4 | Mixed | 96 | 99.7% | 859.6 | <0.005 | 0.20 | Random |
| main meta-analysis | All | 30-40 | 0.36 | 0.07 – 0.65 | 0.37 ± 0.16 (0.18–0.60) | 5 | Observational Cohort | 185 | 99.6% | 1102.3 | <0.005 | 0.66 | Random |
| main meta-analysis | All | 40-50 | 0.34 | 0.11 – 0.57 | 0.34 ± 0.11 (0.12–0.50) | 7 | Observational Cohort | 665 | 99.9% | 6264.2 | <0.005 | 0.07 | Random |
| main meta-analysis | All | 50-60 | 0.38 | 0.30 – 0.47 | 0.39 ± 0.04 (0.34–0.44) | 5 | Observational Cohort | 2669 | 97.5% | 163.1 | <0.005 | 0.20 | Random |
| main meta-analysis | All | 60+ | 0.37 | 0.22 – 0.52 | 0.37 ± 0.08 (0.29–0.49) | 8 | Observational Cohort | 825 | 99.2% | 842.2 | <0.005 | 0.33 | Random |
| **main meta-analysis** | **All** | **Meta-Regression** | **0.17** | **0.07 – 0.27** | **—** | **54** | **Mixed** | **5077** | **95%** | **434.5** | **<0.005** | **0.22** | **Multivariable** |
| **main meta-analysis** | **All** | **Meta-Regression** | **0.17** | **0.07 – 0.27** | **—** | **54** | **Mixed** | **5077** | **95%** | **434.5** | **<0.005** | **0.22** | **Multivariable** |
| **main meta-analysis** | **All** | **Overall** | **0.31** | **0.15 – 0.46** | **0.31 ± 0.08 (0.12–0.60)** | **92** | **Mixed** | **9725** | **99.4%** | **16025.3** | **<0.005** | **<0.05** | **Random** |
| **main meta-analysis** | **All** | **Overall** | **0.31** | **0.15 – 0.46** | **0.31 ± 0.08 (0.12–0.60)** | **92** | **Mixed** | **9725** | **99.4%** | **16025.3** | **<0.005** | **<0.05** | **Random** |
| main meta-analysis | black | 18+ | 0.28 | 0.16 – 0.40 | 0.28 ± 0.00 (0.28–0.28) | 3 | Observational Cohort | 51 | 0% | 0 | 1.00 | — | Fixed |
| main meta-analysis | black | 20-30 | — | NA – NA | — | NA | NA | NA | — | — | — | — | NA |
| main meta-analysis | black | 30-40 | — | NA – NA | — | NA | NA | NA | — | — | — | — | NA |
| main meta-analysis | black | 40-50 | 0.32 | 0.11 – 0.53 | — (0.32–0.32) | 1 | Observational Cohort | 29 | — | — | — | — | Single study |
| main meta-analysis | black | 50-60 | 0.38 | 0.14 – 0.62 | — (0.38–0.38) | 1 | Observational Cohort | 131 | — | — | — | — | Single study |
| main meta-analysis | black | 60+ | 0.34 | 0.12 – 0.56 | 0.34 ± 0.01 (0.33–0.34) | 2 | Observational Cohort | 144 | 0% | 0.2 | 0.69 | — | Fixed |
| **main meta-analysis** | **black** | **Meta-Regression** | **—** | **NA – NA** | **—** | **NA** | **NA** | **NA** | **—** | **—** | **—** | **—** | **NA** |
| **main meta-analysis** | **black** | **Overall** | **0.32** | **0.24 – 0.39** | **0.32 ± 0.04 (0.28–0.38)** | **7** | **Observational Cohort** | **355** | **89.9%** | **59.5** | **<0.005** | **0.18** | **Random** |
| main meta-analysis | mixed | 18+ | 0.28 | 0.17 – 0.40 | 0.29 ± 0.07 (0.18–0.59) | 54 | Mixed | 5069 | 93.5% | 820.2 | <0.005 | 0.54 | Random |
| main meta-analysis | mixed | 20-30 | 0.28 | 0.04 – 0.51 | 0.28 ± 0.12 (0.17–0.40) | 4 | Mixed | 96 | 99.7% | 859.6 | <0.005 | 0.20 | Random |
| main meta-analysis | mixed | 30-40 | 0.36 | 0.07 – 0.65 | 0.37 ± 0.16 (0.18–0.60) | 5 | Observational Cohort | 185 | 99.6% | 1102.3 | <0.005 | 0.66 | Random |
| main meta-analysis | mixed | 40-50 | 0.34 | 0.03 – 0.65 | 0.34 ± 0.16 (0.12–0.50) | 4 | Observational Cohort | 106 | 99.9% | 5297.9 | <0.005 | 0.06 | Random |
| main meta-analysis | mixed | 50-60 | 0.38 | 0.26 – 0.50 | 0.38 ± 0.06 (0.34–0.42) | 2 | Observational Cohort | 50 | 99.2% | 123.7 | <0.005 | — | Random |
| main meta-analysis | mixed | 60+ | 0.30 | 0.27 – 0.33 | — (0.30–0.30) | 1 | Observational Cohort | 25 | — | — | — | — | Single study |
| **main meta-analysis** | **mixed** | **Meta-Regression** | **—** | **NA – NA** | **—** | **NA** | **NA** | **NA** | **—** | **—** | **—** | **—** | **NA** |
| **main meta-analysis** | **mixed** | **Overall** | **0.30** | **0.14 – 0.45** | **0.30 ± 0.09 (0.12–0.60)** | **70** | **Mixed** | **5531** | **99.5%** | **13157.3** | **<0.005** | **0.05** | **Random** |
| main meta-analysis | white | 18+ | 0.28 | 0.17 – 0.40 | 0.29 ± 0.01 (0.28–0.29) | 6 | Observational Cohort | 165 | 0% | 1.1 | 0.95 | <0.001 | Fixed |
| main meta-analysis | white | 20-30 | — | NA – NA | — | NA | NA | NA | — | — | — | — | NA |
| main meta-analysis | white | 30-40 | — | NA – NA | — | NA | NA | NA | — | — | — | — | NA |
| main meta-analysis | white | 40-50 | 0.33 | 0.11 – 0.55 | 0.36 ± 0.04 (0.33–0.38) | 2 | Observational Cohort | 530 | 0% | 0.8 | 0.37 | — | Fixed |
| main meta-analysis | white | 50-60 | 0.39 | 0.27 – 0.51 | 0.39 ± 0.06 (0.35–0.44) | 2 | Observational Cohort | 2488 | 95.9% | 24.6 | <0.005 | — | Random |
| main meta-analysis | white | 60+ | 0.40 | 0.23 – 0.57 | 0.40 ± 0.09 (0.29–0.49) | 5 | Observational Cohort | 656 | 99.1% | 432.4 | <0.005 | 0.68 | Random |
| **main meta-analysis** | **white** | **Meta-Regression** | **—** | **NA – NA** | **—** | **NA** | **NA** | **NA** | **—** | **—** | **—** | **—** | **NA** |
| **main meta-analysis** | **white** | **Overall** | **0.35** | **0.20 – 0.49** | **0.35 ± 0.07 (0.28–0.49)** | **15** | **Observational Cohort** | **3839** | **98.4%** | **856.1** | **<0.005** | **0.80** | **Random** |
| male | All | 18+ | 0.37 | 0.09 – 0.65 | 0.37 ± 0.15 (0.26–0.59) | 4 | Observational Cohort | 182 | 96.9% | 98.1 | <0.005 | <0.05 | Random |
| male | All | 20-30 | 0.25 | -0.01 – 0.51 | 0.25 ± 0.13 (0.17–0.40) | 3 | Mixed | 71 | 99.8% | 816.1 | <0.005 | 0.42 | Random |
| male | All | 30-40 | 0.43 | 0.15 – 0.72 | 0.45 ± 0.16 (0.29–0.60) | 3 | Observational Cohort | 143 | 99.6% | 467.2 | <0.005 | 0.44 | Random |
| male | All | 40-50 | 0.50 | 0.41 – 0.59 | 0.44 ± 0.08 (0.38–0.50) | 2 | Observational Cohort | 31 | 72.2% | 3.6 | 0.06 | — | Fixed |
| male | All | 50-60 | 0.43 | 0.30 – 0.55 | 0.43 ± 0.01 (0.42–0.44) | 2 | Observational Cohort | 104 | 0% | 0.5 | 0.48 | — | Fixed |
| male | All | 60+ | 0.46 | 0.40 – 0.52 | 0.46 ± 0.03 (0.43–0.49) | 3 | Observational Cohort | 329 | 90.2% | 20.3 | <0.005 | 0.32 | Random |
| **male** | **All** | **Meta-Regression** | **0.46** | **0.13 – 0.78** | **—** | **5** | **Mixed** | **299** | **99.8%** | **406.1** | **<0.005** | **0.70** | **Univariable** |
| **male** | **All** | **Overall** | **0.39** | **0.16 – 0.62** | **0.39 ± 0.12 (0.17–0.60)** | **17** | **Mixed** | **860** | **99.5%** | **3373.7** | **<0.005** | **0.05** | **Random** |
| male | black | 18+ | — | NA – NA | — | NA | NA | NA | — | — | — | — | NA |
| male | black | 20-30 | — | NA – NA | — | NA | NA | NA | — | — | — | — | NA |
| male | black | 30-40 | — | NA – NA | — | NA | NA | NA | — | — | — | — | NA |
| male | black | 40-50 | — | NA – NA | — | NA | NA | NA | — | — | — | — | NA |
| male | black | 50-60 | — | NA – NA | — | NA | NA | NA | — | — | — | — | NA |
| male | black | 60+ | — | NA – NA | — | NA | NA | NA | — | — | — | — | NA |
| **male** | **black** | **Meta-Regression** | **—** | **NA – NA** | **—** | **NA** | **NA** | **NA** | **—** | **—** | **—** | **—** | **NA** |
| **male** | **black** | **Overall** | **—** | **NA – NA** | **—** | **NA** | **NA** | **NA** | **—** | **—** | **—** | **—** | **NA** |
| male | mixed | 18+ | 0.37 | 0.09 – 0.65 | 0.37 ± 0.15 (0.26–0.59) | 4 | Observational Cohort | 182 | 96.9% | 98.1 | <0.005 | <0.05 | Random |
| male | mixed | 20-30 | 0.25 | -0.01 – 0.51 | 0.25 ± 0.13 (0.17–0.40) | 3 | Mixed | 71 | 99.8% | 816.1 | <0.005 | 0.42 | Random |
| male | mixed | 30-40 | 0.43 | 0.15 – 0.72 | 0.45 ± 0.16 (0.29–0.60) | 3 | Observational Cohort | 143 | 99.6% | 467.2 | <0.005 | 0.44 | Random |
| male | mixed | 40-50 | 0.50 | 0.42 – 0.58 | — (0.50–0.50) | 1 | Observational Cohort | 25 | — | — | — | — | Single study |
| male | mixed | 50-60 | 0.42 | 0.36 – 0.49 | — (0.42–0.42) | 1 | Observational Cohort | 25 | — | — | — | — | Single study |
| male | mixed | 60+ | — | NA – NA | — | NA | NA | NA | — | — | — | — | NA |
| **male** | **mixed** | **Meta-Regression** | **—** | **NA – NA** | **—** | **NA** | **NA** | **NA** | **—** | **—** | **—** | **—** | **NA** |
| **male** | **mixed** | **Overall** | **0.37** | **0.10 – 0.64** | **0.38 ± 0.14 (0.17–0.60)** | **12** | **Mixed** | **446** | **99.5%** | **2315.5** | **<0.005** | **0.21** | **Random** |
| male | white | 18+ | — | NA – NA | — | NA | NA | NA | — | — | — | — | NA |
| male | white | 20-30 | — | NA – NA | — | NA | NA | NA | — | — | — | — | NA |
| male | white | 30-40 | — | NA – NA | — | NA | NA | NA | — | — | — | — | NA |
| male | white | 40-50 | 0.38 | 0.09 – 0.67 | — (0.38–0.38) | 1 | Observational Cohort | 6 | — | — | — | — | Single study |
| male | white | 50-60 | 0.44 | 0.12 – 0.75 | — (0.44–0.44) | 1 | Observational Cohort | 79 | — | — | — | — | Single study |
| male | white | 60+ | 0.46 | 0.40 – 0.52 | 0.46 ± 0.03 (0.43–0.49) | 3 | Observational Cohort | 329 | 90.2% | 20.3 | <0.005 | 0.32 | Random |
| **male** | **white** | **Meta-Regression** | **—** | **NA – NA** | **—** | **NA** | **NA** | **NA** | **—** | **—** | **—** | **—** | **NA** |
| **male** | **white** | **Overall** | **0.45** | **0.40 – 0.50** | **0.44 ± 0.04 (0.38–0.49)** | **5** | **Observational Cohort** | **414** | **84.7%** | **26.1** | **<0.005** | **0.12** | **Random** |
| female | All | 18+ | 0.26 | 0.22 – 0.30 | 0.27 ± 0.03 (0.24–0.31) | 7 | Mixed | 488 | 64.3% | 16.8 | <0.01 | 0.10 | Random |
| female | All | 20-30 | 0.35 | 0.30 – 0.41 | — (0.35–0.35) | 1 | Observational Cohort | 25 | — | — | — | — | Single study |
| female | All | 30-40 | 0.34 | 0.29 – 0.39 | — (0.34–0.34) | 1 | Observational Cohort | 25 | — | — | — | — | Single study |
| female | All | 40-50 | 0.34 | 0.29 – 0.40 | 0.34 ± 0.03 (0.32–0.38) | 3 | Observational Cohort | 578 | 93.1% | 28.8 | <0.005 | 0.86 | Random |
| female | All | 50-60 | 0.35 | 0.31 – 0.39 | 0.36 ± 0.02 (0.34–0.38) | 3 | Observational Cohort | 2565 | 87.1% | 15.5 | <0.005 | 0.61 | Random |
| female | All | 60+ | 0.32 | 0.28 – 0.35 | 0.32 ± 0.02 (0.29–0.34) | 5 | Observational Cohort | 496 | 82.5% | 22.9 | <0.005 | 0.20 | Random |
| **female** | **All** | **Meta-Regression** | **0.26** | **0.20 – 0.32** | **—** | **7** | **Mixed** | **488** | **80.8%** | **16.5** | **<0.005** | **0.10** | **Multivariable** |
| **female** | **All** | **Overall** | **0.31** | **0.23 – 0.40** | **0.31 ± 0.04 (0.24–0.38)** | **20** | **Mixed** | **4177** | **95.7%** | **439.7** | **<0.005** | **0.10** | **Random** |
| female | black | 18+ | — | NA – NA | — | NA | NA | NA | — | — | — | — | NA |
| female | black | 20-30 | — | NA – NA | — | NA | NA | NA | — | — | — | — | NA |
| female | black | 30-40 | — | NA – NA | — | NA | NA | NA | — | — | — | — | NA |
| female | black | 40-50 | 0.32 | 0.11 – 0.53 | — (0.32–0.32) | 1 | Observational Cohort | 29 | — | — | — | — | Single study |
| female | black | 50-60 | 0.38 | 0.14 – 0.62 | — (0.38–0.38) | 1 | Observational Cohort | 131 | — | — | — | — | Single study |
| female | black | 60+ | 0.34 | 0.12 – 0.56 | 0.34 ± 0.01 (0.33–0.34) | 2 | Observational Cohort | 144 | 0% | 0.2 | 0.69 | — | Fixed |
| **female** | **black** | **Meta-Regression** | **—** | **NA – NA** | **—** | **NA** | **NA** | **NA** | **—** | **—** | **—** | **—** | **NA** |
| **female** | **black** | **Overall** | **0.35** | **0.30 – 0.39** | **0.34 ± 0.03 (0.32–0.38)** | **4** | **Observational Cohort** | **304** | **75.9%** | **12.4** | **<0.01** | **0.31** | **Random** |
| female | mixed | 18+ | 0.26 | 0.22 – 0.30 | 0.27 ± 0.03 (0.24–0.31) | 7 | Mixed | 488 | 64.3% | 16.8 | <0.01 | 0.10 | Random |
| female | mixed | 20-30 | 0.35 | 0.30 – 0.41 | — (0.35–0.35) | 1 | Observational Cohort | 25 | — | — | — | — | Single study |
| female | mixed | 30-40 | 0.34 | 0.29 – 0.39 | — (0.34–0.34) | 1 | Observational Cohort | 25 | — | — | — | — | Single study |
| female | mixed | 40-50 | 0.38 | 0.30 – 0.45 | — (0.38–0.38) | 1 | Observational Cohort | 25 | — | — | — | — | Single study |
| female | mixed | 50-60 | 0.34 | 0.30 – 0.38 | — (0.34–0.34) | 1 | Observational Cohort | 25 | — | — | — | — | Single study |
| female | mixed | 60+ | 0.30 | 0.27 – 0.33 | — (0.30–0.30) | 1 | Observational Cohort | 25 | — | — | — | — | Single study |
| **female** | **mixed** | **Meta-Regression** | **—** | **NA – NA** | **—** | **NA** | **NA** | **NA** | **—** | **—** | **—** | **—** | **NA** |
| **female** | **mixed** | **Overall** | **0.30** | **0.21 – 0.39** | **0.30 ± 0.05 (0.24–0.38)** | **12** | **Mixed** | **613** | **96.6%** | **321.6** | **<0.005** | **0.21** | **Random** |
| female | white | 18+ | — | NA – NA | — | NA | NA | NA | — | — | — | — | NA |
| female | white | 20-30 | — | NA – NA | — | NA | NA | NA | — | — | — | — | NA |
| female | white | 30-40 | — | NA – NA | — | NA | NA | NA | — | — | — | — | NA |
| female | white | 40-50 | 0.33 | 0.11 – 0.55 | — (0.33–0.33) | 1 | Observational Cohort | 524 | — | — | — | — | Single study |
| female | white | 50-60 | 0.35 | 0.09 – 0.61 | — (0.35–0.35) | 1 | Observational Cohort | 2409 | — | — | — | — | Single study |
| female | white | 60+ | 0.31 | 0.27 – 0.35 | 0.31 ± 0.02 (0.29–0.32) | 2 | Observational Cohort | 327 | 83.1% | 5.9 | <0.05 | — | Random |
| **female** | **white** | **Meta-Regression** | **—** | **NA – NA** | **—** | **NA** | **NA** | **NA** | **—** | **—** | **—** | **—** | **NA** |
| **female** | **white** | **Overall** | **0.32** | **0.28 – 0.37** | **0.32 ± 0.02 (0.29–0.35)** | **4** | **Observational Cohort** | **3260** | **93.2%** | **44** | **<0.005** | **<0.05** | **Random** |
| *I² represents heterogeneity; Q test assesses between-study variation; Egger's p = linear regression test for funnel plot asymmetry; Meta-Regression shows covariate-adjusted estimates* | | | | | | | | | | | | | |

###### 6.5.2.3 Table of Results subdivided by race / Sex / Age

oxalate_ma_3rd_analysis$race_summary_table

Table 1: **race-Stratified Meta-Analysis Results**

24-Hour Urine Oxalate (mmol/24h)

| Sex Group | race | Age Band | 95% Range | Studies | Study Type | Participants | I² | Cochran's Q | Q (p-value) | Egger's p | Model |
| --- | --- | --- | --- | --- | --- | --- | --- | --- | --- | --- | --- |
| **main meta-analysis** | **All races** | **18+** | **0.18 – 0.39** | **63** | **Mixed** | **5285** | **92.5%** | **829** | **<0.005** | **0.41** | **Random** |
| **main meta-analysis** | **All races** | **20-30** | **0.04 – 0.51** | **4** | **Mixed** | **96** | **99.7%** | **859.6** | **<0.005** | **0.20** | **Random** |
| **main meta-analysis** | **All races** | **30-40** | **0.07 – 0.65** | **5** | **Observational Cohort** | **185** | **99.6%** | **1102.3** | **<0.005** | **0.66** | **Random** |
| **main meta-analysis** | **All races** | **40-50** | **0.11 – 0.57** | **7** | **Observational Cohort** | **665** | **99.9%** | **6264.2** | **<0.005** | **0.07** | **Random** |
| **main meta-analysis** | **All races** | **50-60** | **0.30 – 0.47** | **5** | **Observational Cohort** | **2669** | **97.5%** | **163.1** | **<0.005** | **0.20** | **Random** |
| **main meta-analysis** | **All races** | **60+** | **0.22 – 0.52** | **8** | **Observational Cohort** | **825** | **99.2%** | **842.2** | **<0.005** | **0.33** | **Random** |
| **main meta-analysis** | **All races** | **Meta-Regression** | **0.07 – 0.27** | **54** | **Mixed** | **5077** | **95%** | **434.5** | **<0.005** | **0.22** | **Multivariable** |
| **main meta-analysis** | **All races** | **Meta-Regression** | **0.07 – 0.27** | **54** | **Mixed** | **5077** | **95%** | **434.5** | **<0.005** | **0.22** | **Multivariable** |
| **main meta-analysis** | **All races** | **Overall** | **0.15 – 0.46** | **92** | **Mixed** | **9725** | **99.4%** | **16025.3** | **<0.005** | **<0.05** | **Random** |
| **main meta-analysis** | **All races** | **Overall** | **0.15 – 0.46** | **92** | **Mixed** | **9725** | **99.4%** | **16025.3** | **<0.005** | **<0.05** | **Random** |
| main meta-analysis | Black | 18+ | 0.16 – 0.40 | 3 | Observational Cohort | 51 | 0% | 0 | 1.00 | — | Fixed |
| main meta-analysis | Black | 20-30 | — | — | NA | — | — | — | — | — | — |
| main meta-analysis | Black | 30-40 | — | — | NA | — | — | — | — | — | — |
| main meta-analysis | Black | 40-50 | 0.11 – 0.53 | 1 | Observational Cohort | 29 | — | — | — | — | Single study |
| main meta-analysis | Black | 50-60 | 0.14 – 0.62 | 1 | Observational Cohort | 131 | — | — | — | — | Single study |
| main meta-analysis | Black | 60+ | 0.12 – 0.56 | 2 | Observational Cohort | 144 | 0% | 0.2 | 0.69 | — | Fixed |
| main meta-analysis | Black | Meta-Regression | — | — | NA | — | — | — | — | — | — |
| **main meta-analysis** | **Black** | **Overall** | **0.24 – 0.39** | **7** | **Observational Cohort** | **355** | **89.9%** | **59.5** | **<0.005** | **0.18** | **Random** |
| main meta-analysis | Mixed | 18+ | 0.17 – 0.40 | 54 | Mixed | 5069 | 93.5% | 820.2 | <0.005 | 0.54 | Random |
| main meta-analysis | Mixed | 20-30 | 0.04 – 0.51 | 4 | Mixed | 96 | 99.7% | 859.6 | <0.005 | 0.20 | Random |
| main meta-analysis | Mixed | 30-40 | 0.07 – 0.65 | 5 | Observational Cohort | 185 | 99.6% | 1102.3 | <0.005 | 0.66 | Random |
| main meta-analysis | Mixed | 40-50 | 0.03 – 0.65 | 4 | Observational Cohort | 106 | 99.9% | 5297.9 | <0.005 | 0.06 | Random |
| main meta-analysis | Mixed | 50-60 | 0.26 – 0.50 | 2 | Observational Cohort | 50 | 99.2% | 123.7 | <0.005 | — | Random |
| main meta-analysis | Mixed | 60+ | 0.27 – 0.33 | 1 | Observational Cohort | 25 | — | — | — | — | Single study |
| main meta-analysis | Mixed | Meta-Regression | — | — | NA | — | — | — | — | — | — |
| **main meta-analysis** | **Mixed** | **Overall** | **0.14 – 0.45** | **70** | **Mixed** | **5531** | **99.5%** | **13157.3** | **<0.005** | **0.05** | **Random** |
| main meta-analysis | White | 18+ | 0.17 – 0.40 | 6 | Observational Cohort | 165 | 0% | 1.1 | 0.95 | <0.001 | Fixed |
| main meta-analysis | White | 20-30 | — | — | NA | — | — | — | — | — | — |
| main meta-analysis | White | 30-40 | — | — | NA | — | — | — | — | — | — |
| main meta-analysis | White | 40-50 | 0.11 – 0.55 | 2 | Observational Cohort | 530 | 0% | 0.8 | 0.37 | — | Fixed |
| main meta-analysis | White | 50-60 | 0.27 – 0.51 | 2 | Observational Cohort | 2488 | 95.9% | 24.6 | <0.005 | — | Random |
| main meta-analysis | White | 60+ | 0.23 – 0.57 | 5 | Observational Cohort | 656 | 99.1% | 432.4 | <0.005 | 0.68 | Random |
| main meta-analysis | White | Meta-Regression | — | — | NA | — | — | — | — | — | — |
| **main meta-analysis** | **White** | **Overall** | **0.20 – 0.49** | **15** | **Observational Cohort** | **3839** | **98.4%** | **856.1** | **<0.005** | **0.80** | **Random** |
| **male** | **All races** | **18+** | **0.09 – 0.65** | **4** | **Observational Cohort** | **182** | **96.9%** | **98.1** | **<0.005** | **<0.05** | **Random** |
| **male** | **All races** | **20-30** | **-0.01 – 0.51** | **3** | **Mixed** | **71** | **99.8%** | **816.1** | **<0.005** | **0.42** | **Random** |
| **male** | **All races** | **30-40** | **0.15 – 0.72** | **3** | **Observational Cohort** | **143** | **99.6%** | **467.2** | **<0.005** | **0.44** | **Random** |
| **male** | **All races** | **40-50** | **0.41 – 0.59** | **2** | **Observational Cohort** | **31** | **72.2%** | **3.6** | **0.06** | **—** | **Fixed** |
| **male** | **All races** | **50-60** | **0.30 – 0.55** | **2** | **Observational Cohort** | **104** | **0%** | **0.5** | **0.48** | **—** | **Fixed** |
| **male** | **All races** | **60+** | **0.40 – 0.52** | **3** | **Observational Cohort** | **329** | **90.2%** | **20.3** | **<0.005** | **0.32** | **Random** |
| **male** | **All races** | **Meta-Regression** | **0.13 – 0.78** | **5** | **Mixed** | **299** | **99.8%** | **406.1** | **<0.005** | **0.70** | **Univariable** |
| **male** | **All races** | **Overall** | **0.16 – 0.62** | **17** | **Mixed** | **860** | **99.5%** | **3373.7** | **<0.005** | **0.05** | **Random** |
| male | Black | 18+ | — | — | NA | — | — | — | — | — | — |
| male | Black | 20-30 | — | — | NA | — | — | — | — | — | — |
| male | Black | 30-40 | — | — | NA | — | — | — | — | — | — |
| male | Black | 40-50 | — | — | NA | — | — | — | — | — | — |
| male | Black | 50-60 | — | — | NA | — | — | — | — | — | — |
| male | Black | 60+ | — | — | NA | — | — | — | — | — | — |
| male | Black | Meta-Regression | — | — | NA | — | — | — | — | — | — |
| **male** | **Black** | **Overall** | **—** | **—** | **NA** | **—** | **—** | **—** | **—** | **—** | **—** |
| male | Mixed | 18+ | 0.09 – 0.65 | 4 | Observational Cohort | 182 | 96.9% | 98.1 | <0.005 | <0.05 | Random |
| male | Mixed | 20-30 | -0.01 – 0.51 | 3 | Mixed | 71 | 99.8% | 816.1 | <0.005 | 0.42 | Random |
| male | Mixed | 30-40 | 0.15 – 0.72 | 3 | Observational Cohort | 143 | 99.6% | 467.2 | <0.005 | 0.44 | Random |
| male | Mixed | 40-50 | 0.42 – 0.58 | 1 | Observational Cohort | 25 | — | — | — | — | Single study |
| male | Mixed | 50-60 | 0.36 – 0.49 | 1 | Observational Cohort | 25 | — | — | — | — | Single study |
| male | Mixed | 60+ | — | — | NA | — | — | — | — | — | — |
| male | Mixed | Meta-Regression | — | — | NA | — | — | — | — | — | — |
| **male** | **Mixed** | **Overall** | **0.10 – 0.64** | **12** | **Mixed** | **446** | **99.5%** | **2315.5** | **<0.005** | **0.21** | **Random** |
| male | White | 18+ | — | — | NA | — | — | — | — | — | — |
| male | White | 20-30 | — | — | NA | — | — | — | — | — | — |
| male | White | 30-40 | — | — | NA | — | — | — | — | — | — |
| male | White | 40-50 | 0.09 – 0.67 | 1 | Observational Cohort | 6 | — | — | — | — | Single study |
| male | White | 50-60 | 0.12 – 0.75 | 1 | Observational Cohort | 79 | — | — | — | — | Single study |
| male | White | 60+ | 0.40 – 0.52 | 3 | Observational Cohort | 329 | 90.2% | 20.3 | <0.005 | 0.32 | Random |
| male | White | Meta-Regression | — | — | NA | — | — | — | — | — | — |
| **male** | **White** | **Overall** | **0.40 – 0.50** | **5** | **Observational Cohort** | **414** | **84.7%** | **26.1** | **<0.005** | **0.12** | **Random** |
| **female** | **All races** | **18+** | **0.22 – 0.30** | **7** | **Mixed** | **488** | **64.3%** | **16.8** | **<0.01** | **0.10** | **Random** |
| **female** | **All races** | **20-30** | **0.30 – 0.41** | **1** | **Observational Cohort** | **25** | **—** | **—** | **—** | **—** | **Single study** |
| **female** | **All races** | **30-40** | **0.29 – 0.39** | **1** | **Observational Cohort** | **25** | **—** | **—** | **—** | **—** | **Single study** |
| **female** | **All races** | **40-50** | **0.29 – 0.40** | **3** | **Observational Cohort** | **578** | **93.1%** | **28.8** | **<0.005** | **0.86** | **Random** |
| **female** | **All races** | **50-60** | **0.31 – 0.39** | **3** | **Observational Cohort** | **2565** | **87.1%** | **15.5** | **<0.005** | **0.61** | **Random** |
| **female** | **All races** | **60+** | **0.28 – 0.35** | **5** | **Observational Cohort** | **496** | **82.5%** | **22.9** | **<0.005** | **0.20** | **Random** |
| **female** | **All races** | **Meta-Regression** | **0.20 – 0.32** | **7** | **Mixed** | **488** | **80.8%** | **16.5** | **<0.005** | **0.10** | **Multivariable** |
| **female** | **All races** | **Overall** | **0.23 – 0.40** | **20** | **Mixed** | **4177** | **95.7%** | **439.7** | **<0.005** | **0.10** | **Random** |
| female | Black | 18+ | — | — | NA | — | — | — | — | — | — |
| female | Black | 20-30 | — | — | NA | — | — | — | — | — | — |
| female | Black | 30-40 | — | — | NA | — | — | — | — | — | — |
| female | Black | 40-50 | 0.11 – 0.53 | 1 | Observational Cohort | 29 | — | — | — | — | Single study |
| female | Black | 50-60 | 0.14 – 0.62 | 1 | Observational Cohort | 131 | — | — | — | — | Single study |
| female | Black | 60+ | 0.12 – 0.56 | 2 | Observational Cohort | 144 | 0% | 0.2 | 0.69 | — | Fixed |
| female | Black | Meta-Regression | — | — | NA | — | — | — | — | — | — |
| **female** | **Black** | **Overall** | **0.30 – 0.39** | **4** | **Observational Cohort** | **304** | **75.9%** | **12.4** | **<0.01** | **0.31** | **Random** |
| female | Mixed | 18+ | 0.22 – 0.30 | 7 | Mixed | 488 | 64.3% | 16.8 | <0.01 | 0.10 | Random |
| female | Mixed | 20-30 | 0.30 – 0.41 | 1 | Observational Cohort | 25 | — | — | — | — | Single study |
| female | Mixed | 30-40 | 0.29 – 0.39 | 1 | Observational Cohort | 25 | — | — | — | — | Single study |
| female | Mixed | 40-50 | 0.30 – 0.45 | 1 | Observational Cohort | 25 | — | — | — | — | Single study |
| female | Mixed | 50-60 | 0.30 – 0.38 | 1 | Observational Cohort | 25 | — | — | — | — | Single study |
| female | Mixed | 60+ | 0.27 – 0.33 | 1 | Observational Cohort | 25 | — | — | — | — | Single study |
| female | Mixed | Meta-Regression | — | — | NA | — | — | — | — | — | — |
| **female** | **Mixed** | **Overall** | **0.21 – 0.39** | **12** | **Mixed** | **613** | **96.6%** | **321.6** | **<0.005** | **0.21** | **Random** |
| female | White | 18+ | — | — | NA | — | — | — | — | — | — |
| female | White | 20-30 | — | — | NA | — | — | — | — | — | — |
| female | White | 30-40 | — | — | NA | — | — | — | — | — | — |
| female | White | 40-50 | 0.11 – 0.55 | 1 | Observational Cohort | 524 | — | — | — | — | Single study |
| female | White | 50-60 | 0.09 – 0.61 | 1 | Observational Cohort | 2409 | — | — | — | — | Single study |
| female | White | 60+ | 0.27 – 0.35 | 2 | Observational Cohort | 327 | 83.1% | 5.9 | <0.05 | — | Random |
| female | White | Meta-Regression | — | — | NA | — | — | — | — | — | — |
| **female** | **White** | **Overall** | **0.28 – 0.37** | **4** | **Observational Cohort** | **3260** | **93.2%** | **44** | **<0.005** | **<0.05** | **Random** |
| *I² represents heterogeneity; Q test assesses between-study variation; Egger's p = linear regression test for funnel plot asymmetry; — indicates insufficient data* | | | | | | | | | | | |

###### 6.5.2.4 Wald Tests

oxalate_ma_3rd_analysis$wald_tests$sex_comparison

Table 1: **Sex Comparison (Overall)**

| Comparison | % Shift | P (Wald Test) | Mean Diff (95% CI) |
| --- | --- | --- | --- |
| Male vs Female (Overall) | +24.8% | 0.541 | 0.08 [-0.17, 0.33] |
| *Mean difference = Group 1 − Group 2 in component units (mmol/24h, L/24h, or pH); % shift = difference relative to Group 2.* | | | |

oxalate_ma_3rd_analysis$wald_tests$race_comparisons

Table 1: **race Comparisons Within Sex Groups**

| Sex Group | Comparison | % Shift | P (Wald Test) | Mean Diff (95% CI) |
| --- | --- | --- | --- | --- |
| male | black vs mixed | — | — | — |
| male | black vs white | — | — | — |
| male | mixed vs white | -18.1% | 0.557 | -0.08 [-0.35, 0.19] |
| female | black vs mixed | +16.4% | 0.350 | 0.05 [-0.05, 0.15] |
| female | black vs white | +7.2% | 0.469 | 0.02 [-0.04, 0.09] |
| female | mixed vs white | -7.9% | 0.620 | -0.03 [-0.13, 0.08] |
| *Mean difference = Group 1 − Group 2 in component units (mmol/24h, L/24h, or pH); % shift = difference relative to Group 2.* | | | | |

oxalate_ma_3rd_analysis$wald_tests$age_band_comparisons

Table 1: **Age Band Comparisons Within Sex Groups**

| Sex Group | Comparison | % Shift | P (Wald Test) | Mean Diff (95% CI) |
| --- | --- | --- | --- | --- |
| main meta-analysis | 18+ vs 20-30 | +2.5% | 0.958 | 0.01 [-0.25, 0.26] |
| main meta-analysis | 18+ vs 30-40 | -21.6% | 0.617 | -0.08 [-0.39, 0.23] |
| main meta-analysis | 18+ vs 40-50 | -16.8% | 0.652 | -0.06 [-0.31, 0.19] |
| main meta-analysis | 18+ vs 50-60 | -26.2% | 0.139 | -0.10 [-0.23, 0.03] |
| main meta-analysis | 18+ vs 60+ | -23.3% | 0.351 | -0.09 [-0.27, 0.09] |
| main meta-analysis | 20-30 vs 30-40 | -23.5% | 0.653 | -0.09 [-0.46, 0.29] |
| main meta-analysis | 20-30 vs 40-50 | -18.8% | 0.699 | -0.06 [-0.39, 0.26] |
| main meta-analysis | 20-30 vs 50-60 | -28.0% | 0.395 | -0.11 [-0.36, 0.14] |
| main meta-analysis | 20-30 vs 60+ | -25.1% | 0.511 | -0.09 [-0.37, 0.18] |
| main meta-analysis | 30-40 vs 40-50 | +6.2% | 0.910 | 0.02 [-0.35, 0.39] |
| main meta-analysis | 30-40 vs 50-60 | -5.9% | 0.883 | -0.02 [-0.32, 0.28] |
| main meta-analysis | 30-40 vs 60+ | -2.1% | 0.963 | -0.01 [-0.33, 0.32] |
| main meta-analysis | 40-50 vs 50-60 | -11.4% | 0.723 | -0.04 [-0.29, 0.20] |
| main meta-analysis | 40-50 vs 60+ | -7.8% | 0.835 | -0.03 [-0.30, 0.24] |
| main meta-analysis | 50-60 vs 60+ | +4.0% | 0.864 | 0.01 [-0.16, 0.19] |
| male | 18+ vs 20-30 | +47.0% | 0.542 | 0.12 [-0.26, 0.50] |
| male | 18+ vs 30-40 | -14.9% | 0.749 | -0.06 [-0.46, 0.33] |
| male | 18+ vs 40-50 | -25.4% | 0.398 | -0.13 [-0.42, 0.17] |
| male | 18+ vs 50-60 | -13.3% | 0.714 | -0.06 [-0.36, 0.25] |
| male | 18+ vs 60+ | -19.6% | 0.535 | -0.09 [-0.37, 0.19] |
| male | 20-30 vs 30-40 | -42.1% | 0.352 | -0.18 [-0.57, 0.20] |
| male | 20-30 vs 40-50 | -49.3% | 0.081 | -0.24 [-0.52, 0.03] |
| male | 20-30 vs 50-60 | -41.0% | 0.232 | -0.18 [-0.46, 0.11] |
| male | 20-30 vs 60+ | -45.3% | 0.124 | -0.21 [-0.47, 0.06] |
| male | 30-40 vs 40-50 | -12.3% | 0.689 | -0.06 [-0.36, 0.24] |
| male | 30-40 vs 50-60 | +1.9% | 0.959 | 0.01 [-0.30, 0.32] |
| male | 30-40 vs 60+ | -5.4% | 0.866 | -0.02 [-0.32, 0.27] |
| male | 40-50 vs 50-60 | +16.2% | 0.377 | 0.07 [-0.08, 0.22] |
| male | 40-50 vs 60+ | +7.9% | 0.508 | 0.04 [-0.07, 0.14] |
| male | 50-60 vs 60+ | -7.2% | 0.634 | -0.03 [-0.17, 0.10] |
| female | 18+ vs 20-30 | -25.7% | <0.05 | -0.09 [-0.16, -0.02] |
| female | 18+ vs 30-40 | -22.8% | <0.05 | -0.08 [-0.14, -0.02] |
| female | 18+ vs 40-50 | -23.7% | <0.05 | -0.08 [-0.15, -0.01] |
| female | 18+ vs 50-60 | -25.9% | <0.01 | -0.09 [-0.15, -0.03] |
| female | 18+ vs 60+ | -16.8% | 0.061 | -0.05 [-0.11, 0.00] |
| female | 20-30 vs 30-40 | +3.8% | 0.725 | 0.01 [-0.06, 0.09] |
| female | 20-30 vs 40-50 | +2.7% | 0.821 | 0.01 [-0.07, 0.09] |
| female | 20-30 vs 50-60 | -0.4% | 0.970 | -0.00 [-0.07, 0.07] |
| female | 20-30 vs 60+ | +12.0% | 0.276 | 0.04 [-0.03, 0.11] |
| female | 30-40 vs 40-50 | -1.1% | 0.916 | -0.00 [-0.08, 0.07] |
| female | 30-40 vs 50-60 | -4.0% | 0.642 | -0.01 [-0.07, 0.05] |
| female | 30-40 vs 60+ | +7.8% | 0.407 | 0.02 [-0.03, 0.08] |
| female | 40-50 vs 50-60 | -3.0% | 0.763 | -0.01 [-0.08, 0.06] |
| female | 40-50 vs 60+ | +9.0% | 0.401 | 0.03 [-0.04, 0.10] |
| female | 50-60 vs 60+ | +12.4% | 0.162 | 0.04 [-0.02, 0.09] |
| *Mean difference = Group 1 − Group 2 in component units (mmol/24h, L/24h, or pH); % shift = difference relative to Group 2.* | | | | |

oxalate_ma_3rd_analysis$wald_tests$sex_within_age_bands

Table 1: **Sex Comparisons Within Age Bands**

| Age Band | % Shift | P (Wald Test) | Mean Diff (95% CI) |
| --- | --- | --- | --- |
| 18+ | +40.9% | 0.454 | 0.11 [-0.17, 0.39] |
| 20-30 | -28.7% | 0.453 | -0.10 [-0.37, 0.16] |
| 30-40 | +27.9% | 0.520 | 0.09 [-0.19, 0.38] |
| 40-50 | +44.2% | <0.01 | 0.15 [0.05, 0.26] |
| 50-60 | +20.4% | 0.277 | 0.07 [-0.06, 0.20] |
| 60+ | +45.8% | <0.001 | 0.14 [0.08, 0.21] |
| *Mean difference = Group 1 − Group 2 in component units (mmol/24h, L/24h, or pH); % shift = difference relative to Group 2.* | | | |

## 6.6 pH

##### 6.6.1 Comparison by Age Band

###### 6.6.1.1 Plot

```
ph_ma_2nd_analysis <- sex_age_subdivision_ma("ph")
## Not enough studies for multivariable meta-regression (k = 4, need > 6). Running univariable models instead.
ph_ma_2nd_analysis$combined_plot
## `height` was translated to `width`.
## `height` was translated to `width`.
## `height` was translated to `width`.
```

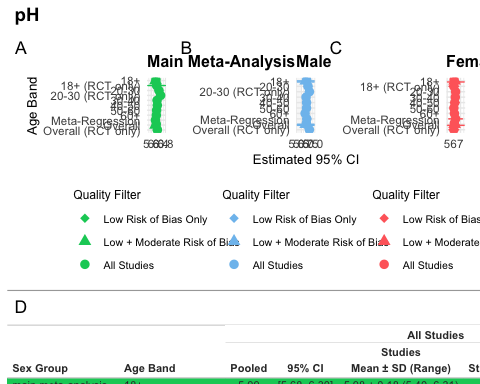

###### 6.6.1.2 Summary Table

ph_ma_2nd_analysis$combined_summary_table

Table 1: **Combined Meta-Analysis Results**

24-Hour Urine pH

|  |  | **All Studies** | | | | | | **Low Risk of Bias Only** | | | | | | **Low + Moderate RoB** | | | | | |
| --- | --- | --- | --- | --- | --- | --- | --- | --- | --- | --- | --- | --- | --- | --- | --- | --- | --- | --- | --- |
| Sex Group | Age Band | Pooled | 95% CI | Studies  Mean ± SD (Range) | Studies, n | Q p-value | Egger's p | Pooled | 95% CI | Studies  Mean ± SD (Range) | Studies, n | Q p-value | Egger's p | Pooled | 95% CI | Studies  Mean ± SD (Range) | Studies, n | Q p-value | Egger's p |
| main meta-analysis | 18+ | 5.99 | [5.68, 6.30] | 5.98 ± 0.18 (5.40–6.31) | 67 | <0.001 | <0.05 | 5.97 | [5.61, 6.32] | 5.97 ± 0.19 (5.45–6.31) | 28 | <0.001 | 0.22 | 5.96 | [5.64, 6.28] | 5.96 ± 0.17 (5.45–6.31) | 35 | <0.001 | 0.14 |
| **main meta-analysis** | **18+ (RCT only)** | **6.04** | **[5.32, 6.77]** | **6.02 ± 0.10 (5.90–6.17)** | **5** | **0.47** | **0.11** | **—** | **—** | **—** | **—** | **—** | **—** | **—** | **—** | **—** | **—** | **—** | **—** |
| main meta-analysis | 20-30 | 6.24 | [5.93, 6.55] | 6.24 ± 0.16 (6.07–6.38) | 3 | <0.001 | 0.83 | — | — | — | — | — | — | — | — | — | — | — | — |
| **main meta-analysis** | **20-30 (RCT only)** | **6.38** | **[6.28, 6.48]** | **— (6.38–6.38)** | **1** | **—** | **—** | **—** | **—** | **—** | **—** | **—** | **—** | **—** | **—** | **—** | **—** | **—** | **—** |
| main meta-analysis | 30-40 | 6.08 | [5.85, 6.32] | 6.08 ± 0.11 (5.91–6.23) | 5 | <0.001 | 0.16 | — | — | — | — | — | — | — | — | — | — | — | — |
| main meta-analysis | 40-50 | 6.11 | [6.01, 6.20] | 6.08 ± 0.07 (6.01–6.18) | 6 | <0.001 | 1.00 | — | — | — | — | — | — | — | — | — | — | — | — |
| main meta-analysis | 50-60 | 6.05 | [5.78, 6.31] | 6.05 ± 0.13 (5.84–6.18) | 6 | <0.001 | 0.83 | — | — | — | — | — | — | — | — | — | — | — | — |
| main meta-analysis | 60+ | 6.02 | [5.75, 6.30] | 6.03 ± 0.15 (5.86–6.22) | 8 | <0.001 | 0.15 | — | — | — | — | — | — | — | — | — | — | — | — |
| **main meta-analysis** | **Meta-Regression** | **5.95** | **[5.63, 6.27]** | **—** | **52** | **<0.001** | **0.35** | **—** | **—** | **—** | **—** | **—** | **—** | **—** | **—** | **—** | **—** | **—** | **—** |
| **main meta-analysis** | **Overall** | **6.02** | **[5.72, 6.32]** | **6.01 ± 0.17 (5.40–6.38)** | **95** | **<0.001** | **0.51** | **5.97** | **[5.61, 6.32]** | **5.97 ± 0.19 (5.45–6.31)** | **28** | **<0.001** | **0.22** | **5.96** | **[5.64, 6.28]** | **5.96 ± 0.17 (5.45–6.31)** | **35** | **<0.001** | **0.14** |
| **main meta-analysis** | **Overall (RCT only)** | **6.11** | **[5.78, 6.44]** | **6.08 ± 0.17 (5.90–6.38)** | **6** | **<0.001** | **<0.001** | **—** | **—** | **—** | **—** | **—** | **—** | **—** | **—** | **—** | **—** | **—** | **—** |
| male | 18+ | 6.08 | [5.85, 6.32] | 6.07 ± 0.14 (5.92–6.21) | 4 | <0.001 | 0.42 | 5.98 | [5.04, 6.92] | — (5.98–5.98) | 1 | — | — | 5.98 | [5.04, 6.92] | — (5.98–5.98) | 1 | — | — |
| **male** | **18+ (RCT only)** | **—** | **—** | **—** | **—** | **—** | **—** | **—** | **—** | **—** | **—** | **—** | **—** | **—** | **—** | **—** | **—** | **—** | **—** |
| male | 20-30 | 6.23 | [5.80, 6.65] | 6.22 ± 0.22 (6.07–6.38) | 2 | <0.001 | — | — | — | — | — | — | — | — | — | — | — | — | — |
| **male** | **20-30 (RCT only)** | **6.38** | **[6.28, 6.48]** | **— (6.38–6.38)** | **1** | **—** | **—** | **—** | **—** | **—** | **—** | **—** | **—** | **—** | **—** | **—** | **—** | **—** | **—** |
| male | 30-40 | 6.02 | [5.80, 6.24] | 6.03 ± 0.10 (5.91–6.10) | 3 | <0.001 | 0.62 | — | — | — | — | — | — | — | — | — | — | — | — |
| male | 40-50 | 6.07 | [5.92, 6.22] | 6.04 ± 0.04 (6.02–6.07) | 2 | 0.75 | — | — | — | — | — | — | — | — | — | — | — | — | — |
| male | 50-60 | 6.06 | [5.77, 6.36] | 6.06 ± 0.16 (5.95–6.17) | 2 | <0.001 | — | — | — | — | — | — | — | — | — | — | — | — | — |
| male | 60+ | 5.91 | [5.83, 6.00] | 5.91 ± 0.05 (5.86–5.95) | 3 | <0.05 | 0.54 | — | — | — | — | — | — | — | — | — | — | — | — |
| **male** | **Meta-Regression** | **6.10** | **[5.67, 6.52]** | **—** | **4** | **<0.001** | **0.40** | **—** | **—** | **—** | **—** | **—** | **—** | **—** | **—** | **—** | **—** | **—** | **—** |
| **male** | **Overall** | **6.05** | **[5.78, 6.32]** | **6.05 ± 0.14 (5.86–6.38)** | **16** | **<0.001** | **0.15** | **5.98** | **[5.04, 6.92]** | **— (5.98–5.98)** | **1** | **—** | **—** | **5.98** | **[5.04, 6.92]** | **— (5.98–5.98)** | **1** | **—** | **—** |
| **male** | **Overall (RCT only)** | **6.38** | **[6.28, 6.48]** | **— (6.38–6.38)** | **1** | **—** | **—** | **—** | **—** | **—** | **—** | **—** | **—** | **—** | **—** | **—** | **—** | **—** | **—** |
| female | 18+ | 6.05 | [5.77, 6.33] | 6.05 ± 0.15 (5.95–6.31) | 9 | <0.001 | 0.48 | 6.31 | [5.02, 7.60] | 6.31 ± 0.00 (6.31–6.31) | 2 | 1.00 | — | 6.12 | [5.79, 6.45] | 6.12 ± 0.18 (5.98–6.31) | 5 | <0.001 | 0.87 |
| **female** | **18+ (RCT only)** | **6.01** | **[5.30, 6.72]** | **— (6.01–6.01)** | **1** | **—** | **—** | **—** | **—** | **—** | **—** | **—** | **—** | **—** | **—** | **—** | **—** | **—** | **—** |
| female | 20-30 | 6.27 | [6.12, 6.42] | — (6.27–6.27) | 1 | — | — | — | — | — | — | — | — | — | — | — | — | — | — |
| **female** | **20-30 (RCT only)** | **—** | **—** | **—** | **—** | **—** | **—** | **—** | **—** | **—** | **—** | **—** | **—** | **—** | **—** | **—** | **—** | **—** | **—** |
| female | 30-40 | 6.23 | [6.03, 6.43] | — (6.23–6.23) | 1 | — | — | — | — | — | — | — | — | — | — | — | — | — | — |
| female | 40-50 | 6.15 | [5.52, 6.79] | 6.11 ± 0.09 (6.01–6.18) | 3 | 0.09 | 0.55 | — | — | — | — | — | — | — | — | — | — | — | — |
| female | 50-60 | 6.02 | [5.68, 6.35] | 6.02 ± 0.17 (5.84–6.18) | 3 | <0.001 | 0.96 | — | — | — | — | — | — | — | — | — | — | — | — |
| female | 60+ | 6.10 | [5.82, 6.37] | 6.10 ± 0.14 (5.89–6.22) | 5 | <0.001 | <0.05 | — | — | — | — | — | — | — | — | — | — | — | — |
| **female** | **Meta-Regression** | **6.27** | **[5.26, 7.28]** | **—** | **9** | **1.00** | **0.48** | **—** | **—** | **—** | **—** | **—** | **—** | **—** | **—** | **—** | **—** | **—** | **—** |
| **female** | **Overall** | **6.09** | **[5.82, 6.35]** | **6.08 ± 0.14 (5.84–6.31)** | **22** | **<0.001** | **0.66** | **6.31** | **[5.02, 7.60]** | **6.31 ± 0.00 (6.31–6.31)** | **2** | **1.00** | **—** | **6.12** | **[5.79, 6.45]** | **6.12 ± 0.18 (5.98–6.31)** | **5** | **<0.001** | **0.87** |
| **female** | **Overall (RCT only)** | **6.01** | **[5.30, 6.72]** | **— (6.01–6.01)** | **1** | **—** | **—** | **—** | **—** | **—** | **—** | **—** | **—** | **—** | **—** | **—** | **—** | **—** | **—** |
| *Studies, n = number of studies; Q p-value = Cochran's Q heterogeneity test; Egger's p = linear regression test for funnel plot asymmetry; RoB = Risk of Bias* | | | | | | | | | | | | | | | | | | | |

###### 6.6.1.3 Wald Tests

ph_ma_2nd_analysis$wald_tests$sex_comparison

Table 1: **Sex Comparison (Overall)**

| Comparison | % Shift | P (Wald Test) | Mean Diff (95% CI) |
| --- | --- | --- | --- |
| Male vs Female (Overall) | -0.6% | 0.857 | -0.03 [-0.42, 0.35] |
| *Mean difference = Group 1 − Group 2 in component units (mmol/24h, L/24h, or pH); % shift = difference relative to Group 2.* | | | |

ph_ma_2nd_analysis$wald_tests$age_band_comparisons

Table 1: **Age Band Comparisons Within Sex Groups**

| Sex Group | Comparison | % Shift | P (Wald Test) | Mean Diff (95% CI) |
| --- | --- | --- | --- | --- |
| main meta-analysis | 18+ vs 20-30 | -4.0% | 0.258 | -0.25 [-0.68, 0.18] |
| main meta-analysis | 18+ vs 30-40 | -1.5% | 0.638 | -0.09 [-0.48, 0.29] |
| main meta-analysis | 18+ vs 40-50 | -1.9% | 0.484 | -0.11 [-0.44, 0.21] |
| main meta-analysis | 18+ vs 50-60 | -0.9% | 0.792 | -0.05 [-0.46, 0.35] |
| main meta-analysis | 18+ vs 60+ | -0.6% | 0.872 | -0.03 [-0.44, 0.38] |
| main meta-analysis | 20-30 vs 30-40 | +2.6% | 0.429 | 0.16 [-0.23, 0.54] |
| main meta-analysis | 20-30 vs 40-50 | +2.2% | 0.410 | 0.13 [-0.19, 0.46] |
| main meta-analysis | 20-30 vs 50-60 | +3.2% | 0.349 | 0.19 [-0.21, 0.60] |
| main meta-analysis | 20-30 vs 60+ | +3.6% | 0.300 | 0.22 [-0.19, 0.62] |
| main meta-analysis | 30-40 vs 40-50 | -0.4% | 0.870 | -0.02 [-0.28, 0.24] |
| main meta-analysis | 30-40 vs 50-60 | +0.6% | 0.834 | 0.04 [-0.32, 0.40] |
| main meta-analysis | 30-40 vs 60+ | +1.0% | 0.747 | 0.06 [-0.30, 0.42] |
| main meta-analysis | 40-50 vs 50-60 | +1.0% | 0.682 | 0.06 [-0.23, 0.35] |
| main meta-analysis | 40-50 vs 60+ | +1.3% | 0.582 | 0.08 [-0.21, 0.37] |
| main meta-analysis | 50-60 vs 60+ | +0.4% | 0.914 | 0.02 [-0.36, 0.40] |
| male | 18+ vs 20-30 | -2.3% | 0.571 | -0.14 [-0.63, 0.35] |
| male | 18+ vs 30-40 | +1.1% | 0.681 | 0.07 [-0.25, 0.39] |
| male | 18+ vs 40-50 | +0.2% | 0.919 | 0.01 [-0.26, 0.29] |
| male | 18+ vs 50-60 | +0.3% | 0.920 | 0.02 [-0.36, 0.40] |
| male | 18+ vs 60+ | +2.9% | 0.179 | 0.17 [-0.08, 0.42] |
| male | 20-30 vs 30-40 | +3.5% | 0.397 | 0.21 [-0.27, 0.69] |
| male | 20-30 vs 40-50 | +2.6% | 0.503 | 0.16 [-0.30, 0.61] |
| male | 20-30 vs 50-60 | +2.6% | 0.546 | 0.16 [-0.36, 0.68] |
| male | 20-30 vs 60+ | +5.3% | 0.162 | 0.31 [-0.13, 0.75] |
| male | 30-40 vs 40-50 | -0.9% | 0.700 | -0.05 [-0.32, 0.21] |
| male | 30-40 vs 50-60 | -0.8% | 0.799 | -0.05 [-0.42, 0.32] |
| male | 30-40 vs 60+ | +1.7% | 0.391 | 0.10 [-0.13, 0.34] |
| male | 40-50 vs 50-60 | +0.1% | 0.978 | 0.00 [-0.33, 0.34] |
| male | 40-50 vs 60+ | +2.6% | 0.081 | 0.16 [-0.02, 0.33] |
| male | 50-60 vs 60+ | +2.6% | 0.335 | 0.15 [-0.16, 0.46] |
| female | 18+ vs 20-30 | -3.5% | 0.182 | -0.22 [-0.54, 0.10] |
| female | 18+ vs 30-40 | -2.9% | 0.309 | -0.18 [-0.52, 0.17] |
| female | 18+ vs 40-50 | -1.7% | 0.774 | -0.10 [-0.80, 0.59] |
| female | 18+ vs 50-60 | +0.6% | 0.876 | 0.04 [-0.41, 0.48] |
| female | 18+ vs 60+ | -0.7% | 0.824 | -0.04 [-0.44, 0.35] |
| female | 20-30 vs 30-40 | +0.6% | 0.751 | 0.04 [-0.21, 0.29] |
| female | 20-30 vs 40-50 | +1.9% | 0.727 | 0.12 [-0.54, 0.77] |
| female | 20-30 vs 50-60 | +4.2% | 0.178 | 0.25 [-0.12, 0.62] |
| female | 20-30 vs 60+ | +2.9% | 0.271 | 0.17 [-0.14, 0.48] |
| female | 30-40 vs 40-50 | +1.2% | 0.822 | 0.08 [-0.59, 0.74] |
| female | 30-40 vs 50-60 | +3.6% | 0.283 | 0.21 [-0.18, 0.60] |
| female | 30-40 vs 60+ | +2.2% | 0.432 | 0.13 [-0.20, 0.47] |
| female | 40-50 vs 50-60 | +2.3% | 0.709 | 0.14 [-0.58, 0.86] |
| female | 40-50 vs 60+ | +0.9% | 0.870 | 0.06 [-0.63, 0.75] |
| female | 50-60 vs 60+ | -1.3% | 0.719 | -0.08 [-0.51, 0.35] |
| *Mean difference = Group 1 − Group 2 in component units (mmol/24h, L/24h, or pH); % shift = difference relative to Group 2.* | | | | |

ph_ma_2nd_analysis$wald_tests$sex_within_age_bands

Table 1: **Sex Comparisons Within Age Bands**

| Age Band | % Shift | P (Wald Test) | Mean Diff (95% CI) |
| --- | --- | --- | --- |
| 18+ | +0.5% | 0.861 | 0.03 [-0.33, 0.40] |
| 20-30 | -0.7% | 0.846 | -0.04 [-0.50, 0.41] |
| 30-40 | -3.4% | 0.156 | -0.21 [-0.51, 0.08] |
| 40-50 | -1.4% | 0.801 | -0.08 [-0.74, 0.57] |
| 50-60 | +0.8% | 0.832 | 0.05 [-0.40, 0.50] |
| 60+ | -3.0% | 0.209 | -0.18 [-0.47, 0.10] |
| *Mean difference = Group 1 − Group 2 in component units (mmol/24h, L/24h, or pH); % shift = difference relative to Group 2.* | | | |

###### 6.6.1.4 Meta-Regression (plain language)

format_meta_reg_plain("ph", sex_group = "all")

Message

No meta-regression result for ph / all

###### 6.6.1.5 Assay Subgroup Analysis

assay_subgroup_table("ph")

Table 1: **Assay subgroup analysis — Ph**

*Per-assay pooled estimates; categories with k < 3 listed but not pooled.*

| Assay category | Studies, n | Participants | 95% CI | I² | Q p-value | Egger's p | Note |
| --- | --- | --- | --- | --- | --- | --- | --- |
| Unreported | 95 | 9,226 | 6.02 [5.99, 6.05] | 96% | <0.001 | 0.512 |  |

###### 6.6.1.6 Preservative Sensitivity (Litholink / acidified samples)

```
preservative_sensitivity_ma("ph")
## # A tibble: 3 × 6
##   group               n_studies n_participants pooled_estimate lower_95 upper_95
##   <chr>                   <int>          <int>           <dbl>    <dbl>    <dbl>
## 1 All studies                95           9226            6.02     5.99     6.05
## 2 Excluding preserva…        95           9226            6.02     5.99     6.05
## 3 Preservative only           0              0           NA       NA       NA
```

###### 6.6.1.7 Creatinine Completeness Sensitivity

creat_completeness_table("ph")

Table 1: **Creatinine-completeness sensitivity — Ph**

*Compares the all-studies pooled estimate against subsets defined by whether the study reported 24h urine creatinine. Studies that do not report creatinine cannot verify collection completeness; a substantial shift here flags possible bias from incomplete collections.*

| Subset | Studies, n | Participants | Range (95% CI) | Δ vs all-studies (%) | Flag |
| --- | --- | --- | --- | --- | --- |
| All studies | 95 | 9,226 | 6.02 [5.99, 6.05] | — | — |
| Creatinine reported only | 61 | 7,809 | 6.02 [5.98, 6.07] | +0.0% | OK |
| Creatinine NOT reported | 34 | 1,417 | 6.01 [5.96, 6.07] | -0.1% | OK |

##### 6.6.2 Comparison by Age / Sex / race

###### 6.6.2.1 Plot

```
ph_ma_3rd_analysis <- sex_age_race_subdivision_ma("ph",
                                                      urine_ma2,
                                                      overall_bias)
## Not enough studies for multivariable meta-regression (k = 4, need > 6). Running univariable models instead.
ph_ma_3rd_analysis$race_plot
## `height` was translated to `width`.
## `height` was translated to `width`.
## `height` was translated to `width`.
```

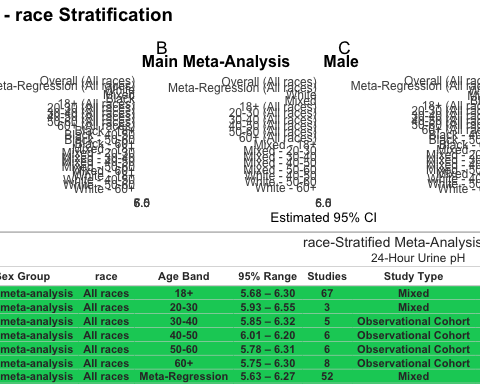

###### 6.6.2.2 Combined MA Results

ph_ma_3rd_analysis$combined_summary_table

Table 1: **Combined Meta-Analysis Results**

24-Hour Urine pH

| Sex Group | race | Age Band | Pooled | 95% Range | Studies Mean SD Range | Studies | Study Type | Participants | I² | Cochran's Q | Q (p-value) | Egger's p | Model |
| --- | --- | --- | --- | --- | --- | --- | --- | --- | --- | --- | --- | --- | --- |
| main meta-analysis | All | 18+ | 5.99 | 5.68 – 6.30 | 5.98 ± 0.18 (5.40–6.31) | 67 | Mixed | 4826 | 85% | 441.1 | <0.005 | <0.05 | Random |
| main meta-analysis | All | 20-30 | 6.24 | 5.93 – 6.55 | 6.24 ± 0.16 (6.07–6.38) | 3 | Mixed | 58 | 98.7% | 155.5 | <0.005 | 0.83 | Random |
| main meta-analysis | All | 30-40 | 6.08 | 5.85 – 6.32 | 6.08 ± 0.11 (5.91–6.23) | 5 | Observational Cohort | 185 | 99% | 397.4 | <0.005 | 0.16 | Random |
| main meta-analysis | All | 40-50 | 6.11 | 6.01 – 6.20 | 6.08 ± 0.07 (6.01–6.18) | 6 | Observational Cohort | 631 | 82.4% | 28.4 | <0.005 | 1.00 | Random |
| main meta-analysis | All | 50-60 | 6.05 | 5.78 – 6.31 | 6.05 ± 0.13 (5.84–6.18) | 6 | Observational Cohort | 2701 | 99.2% | 620 | <0.005 | 0.83 | Random |
| main meta-analysis | All | 60+ | 6.02 | 5.75 – 6.30 | 6.03 ± 0.15 (5.86–6.22) | 8 | Observational Cohort | 825 | 90.9% | 77.1 | <0.005 | 0.15 | Random |
| **main meta-analysis** | **All** | **Meta-Regression** | **5.95** | **5.63 – 6.27** | **—** | **52** | **Mixed** | **4379** | **88.1%** | **324.2** | **<0.005** | **0.35** | **Multivariable** |
| **main meta-analysis** | **All** | **Meta-Regression** | **5.95** | **5.63 – 6.27** | **—** | **52** | **Mixed** | **4379** | **88.1%** | **324.2** | **<0.005** | **0.35** | **Multivariable** |
| **main meta-analysis** | **All** | **Overall** | **6.02** | **5.72 – 6.32** | **6.01 ± 0.17 (5.40–6.38)** | **95** | **Mixed** | **9226** | **96.2%** | **2494.5** | **<0.005** | **0.51** | **Random** |
| **main meta-analysis** | **All** | **Overall** | **6.02** | **5.72 – 6.32** | **6.01 ± 0.17 (5.40–6.38)** | **95** | **Mixed** | **9226** | **96.2%** | **2494.5** | **<0.005** | **0.51** | **Random** |
| main meta-analysis | black | 18+ | 6.20 | 5.47 – 6.93 | 6.20 ± 0.00 (6.20–6.20) | 3 | Observational Cohort | 51 | 0% | 0 | 1.00 | — | Fixed |
| main meta-analysis | black | 20-30 | — | NA – NA | — | NA | NA | NA | — | — | — | — | NA |
| main meta-analysis | black | 30-40 | — | NA – NA | — | NA | NA | NA | — | — | — | — | NA |
| main meta-analysis | black | 40-50 | 6.01 | 5.07 – 6.95 | — (6.01–6.01) | 1 | Observational Cohort | 29 | — | — | — | — | Single study |
| main meta-analysis | black | 50-60 | 6.03 | 4.99 – 7.07 | — (6.03–6.03) | 1 | Observational Cohort | 131 | — | — | — | — | Single study |
| main meta-analysis | black | 60+ | 6.19 | 5.09 – 7.29 | 6.19 ± 0.00 (6.19–6.19) | 2 | Observational Cohort | 144 | 0% | 0 | 1.00 | — | Fixed |
| **main meta-analysis** | **black** | **Meta-Regression** | **—** | **NA – NA** | **—** | **NA** | **NA** | **NA** | **—** | **—** | **—** | **—** | **NA** |
| **main meta-analysis** | **black** | **Overall** | **6.12** | **5.15 – 7.10** | **6.15 ± 0.09 (6.01–6.20)** | **7** | **Observational Cohort** | **355** | **39.5%** | **9.9** | **0.13** | **0.21** | **Fixed** |
| main meta-analysis | mixed | 18+ | 5.97 | 5.65 – 6.29 | 5.97 ± 0.18 (5.40–6.31) | 58 | Mixed | 4610 | 86.7% | 427.8 | <0.005 | <0.01 | Random |
| main meta-analysis | mixed | 20-30 | 6.24 | 5.93 – 6.55 | 6.24 ± 0.16 (6.07–6.38) | 3 | Mixed | 58 | 98.7% | 155.5 | <0.005 | 0.83 | Random |
| main meta-analysis | mixed | 30-40 | 6.08 | 5.85 – 6.32 | 6.08 ± 0.11 (5.91–6.23) | 5 | Observational Cohort | 185 | 99% | 397.4 | <0.005 | 0.16 | Random |
| main meta-analysis | mixed | 40-50 | 6.09 | 6.02 – 6.16 | 6.09 ± 0.04 (6.07–6.14) | 3 | Observational Cohort | 72 | 82% | 11.1 | <0.005 | 0.52 | Random |
| main meta-analysis | mixed | 50-60 | 6.04 | 5.68 – 6.39 | 6.04 ± 0.18 (5.84–6.17) | 3 | Observational Cohort | 82 | 99.1% | 221.5 | <0.005 | 0.62 | Random |
| main meta-analysis | mixed | 60+ | 5.89 | 5.65 – 6.13 | — (5.89–5.89) | 1 | Observational Cohort | 25 | — | — | — | — | Single study |
| **main meta-analysis** | **mixed** | **Meta-Regression** | **—** | **NA – NA** | **—** | **NA** | **NA** | **NA** | **—** | **—** | **—** | **—** | **NA** |
| **main meta-analysis** | **mixed** | **Overall** | **6.00** | **5.68 – 6.33** | **5.99 ± 0.18 (5.40–6.38)** | **73** | **Mixed** | **5032** | **96.6%** | **2104.7** | **<0.005** | **0.44** | **Random** |
| main meta-analysis | white | 18+ | 6.04 | 5.37 – 6.71 | 6.05 ± 0.04 (6.01–6.09) | 6 | Observational Cohort | 165 | 0% | 2.2 | 0.82 | <0.001 | Fixed |
| main meta-analysis | white | 20-30 | — | NA – NA | — | NA | NA | NA | — | — | — | — | NA |
| main meta-analysis | white | 30-40 | — | NA – NA | — | NA | NA | NA | — | — | — | — | NA |
| main meta-analysis | white | 40-50 | 6.18 | 5.24 – 7.12 | 6.10 ± 0.11 (6.02–6.18) | 2 | Observational Cohort | 530 | 0% | 1 | 0.32 | — | Fixed |
| main meta-analysis | white | 50-60 | 6.07 | 5.76 – 6.38 | 6.06 ± 0.16 (5.95–6.18) | 2 | Observational Cohort | 2488 | 95% | 19.8 | <0.005 | — | Random |
| main meta-analysis | white | 60+ | 5.99 | 5.74 – 6.25 | 6.00 ± 0.14 (5.86–6.22) | 5 | Observational Cohort | 656 | 90.9% | 44.2 | <0.005 | 0.55 | Random |
| **main meta-analysis** | **white** | **Meta-Regression** | **—** | **NA – NA** | **—** | **NA** | **NA** | **NA** | **—** | **—** | **—** | **—** | **NA** |
| **main meta-analysis** | **white** | **Overall** | **6.04** | **5.84 – 6.24** | **6.04 ± 0.10 (5.86–6.22)** | **15** | **Observational Cohort** | **3839** | **95%** | **279** | **<0.005** | **0.30** | **Random** |
| male | All | 18+ | 6.08 | 5.85 – 6.32 | 6.07 ± 0.14 (5.92–6.21) | 4 | Observational Cohort | 182 | 84.4% | 19.2 | <0.005 | 0.42 | Random |
| male | All | 20-30 | 6.23 | 5.80 – 6.65 | 6.22 ± 0.22 (6.07–6.38) | 2 | Mixed | 33 | 99.3% | 151 | <0.005 | — | Random |
| male | All | 30-40 | 6.02 | 5.80 – 6.24 | 6.03 ± 0.10 (5.91–6.10) | 3 | Observational Cohort | 143 | 98.7% | 158.2 | <0.005 | 0.62 | Random |
| male | All | 40-50 | 6.07 | 5.92 – 6.22 | 6.04 ± 0.04 (6.02–6.07) | 2 | Observational Cohort | 31 | 0% | 0.1 | 0.75 | — | Fixed |
| male | All | 50-60 | 6.06 | 5.77 – 6.36 | 6.06 ± 0.16 (5.95–6.17) | 2 | Observational Cohort | 104 | 93.9% | 16.3 | <0.005 | — | Random |
| male | All | 60+ | 5.91 | 5.83 – 6.00 | 5.91 ± 0.05 (5.86–5.95) | 3 | Observational Cohort | 329 | 71.1% | 6.9 | <0.05 | 0.54 | Random |
| **male** | **All** | **Meta-Regression** | **6.10** | **5.67 – 6.52** | **—** | **4** | **Mixed** | **261** | **99.1%** | **665.5** | **<0.005** | **0.40** | **Univariable** |
| **male** | **All** | **Overall** | **6.05** | **5.78 – 6.32** | **6.05 ± 0.14 (5.86–6.38)** | **16** | **Mixed** | **822** | **98.5%** | **970.1** | **<0.005** | **0.15** | **Random** |
| male | black | 18+ | — | NA – NA | — | NA | NA | NA | — | — | — | — | NA |
| male | black | 20-30 | — | NA – NA | — | NA | NA | NA | — | — | — | — | NA |
| male | black | 30-40 | — | NA – NA | — | NA | NA | NA | — | — | — | — | NA |
| male | black | 40-50 | — | NA – NA | — | NA | NA | NA | — | — | — | — | NA |
| male | black | 50-60 | — | NA – NA | — | NA | NA | NA | — | — | — | — | NA |
| male | black | 60+ | — | NA – NA | — | NA | NA | NA | — | — | — | — | NA |
| **male** | **black** | **Meta-Regression** | **—** | **NA – NA** | **—** | **NA** | **NA** | **NA** | **—** | **—** | **—** | **—** | **NA** |
| **male** | **black** | **Overall** | **—** | **NA – NA** | **—** | **NA** | **NA** | **NA** | **—** | **—** | **—** | **—** | **NA** |
| male | mixed | 18+ | 6.08 | 5.85 – 6.32 | 6.07 ± 0.14 (5.92–6.21) | 4 | Observational Cohort | 182 | 84.4% | 19.2 | <0.005 | 0.42 | Random |
| male | mixed | 20-30 | 6.23 | 5.80 – 6.65 | 6.22 ± 0.22 (6.07–6.38) | 2 | Mixed | 33 | 99.3% | 151 | <0.005 | — | Random |
| male | mixed | 30-40 | 6.02 | 5.80 – 6.24 | 6.03 ± 0.10 (5.91–6.10) | 3 | Observational Cohort | 143 | 98.7% | 158.2 | <0.005 | 0.62 | Random |
| male | mixed | 40-50 | 6.07 | 5.93 – 6.21 | — (6.07–6.07) | 1 | Observational Cohort | 25 | — | — | — | — | Single study |
| male | mixed | 50-60 | 6.17 | 5.97 – 6.37 | — (6.17–6.17) | 1 | Observational Cohort | 25 | — | — | — | — | Single study |
| male | mixed | 60+ | — | NA – NA | — | NA | NA | NA | — | — | — | — | NA |
| **male** | **mixed** | **Meta-Regression** | **—** | **NA – NA** | **—** | **NA** | **NA** | **NA** | **—** | **—** | **—** | **—** | **NA** |
| **male** | **mixed** | **Overall** | **6.10** | **5.84 – 6.36** | **6.10 ± 0.14 (5.91–6.38)** | **11** | **Mixed** | **408** | **98.9%** | **931** | **<0.005** | **0.07** | **Random** |
| male | white | 18+ | — | NA – NA | — | NA | NA | NA | — | — | — | — | NA |
| male | white | 20-30 | — | NA – NA | — | NA | NA | NA | — | — | — | — | NA |
| male | white | 30-40 | — | NA – NA | — | NA | NA | NA | — | — | — | — | NA |
| male | white | 40-50 | 6.02 | 5.26 – 6.78 | — (6.02–6.02) | 1 | Observational Cohort | 6 | — | — | — | — | Single study |
| male | white | 50-60 | 5.95 | 5.07 – 6.83 | — (5.95–5.95) | 1 | Observational Cohort | 79 | — | — | — | — | Single study |
| male | white | 60+ | 5.91 | 5.83 – 6.00 | 5.91 ± 0.05 (5.86–5.95) | 3 | Observational Cohort | 329 | 71.1% | 6.9 | <0.05 | 0.54 | Random |
| **male** | **white** | **Meta-Regression** | **—** | **NA – NA** | **—** | **NA** | **NA** | **NA** | **—** | **—** | **—** | **—** | **NA** |
| **male** | **white** | **Overall** | **5.94** | **5.45 – 6.42** | **5.94 ± 0.06 (5.86–6.02)** | **5** | **Observational Cohort** | **414** | **45.2%** | **7.3** | **0.12** | **0.71** | **Fixed** |
| female | All | 18+ | 6.05 | 5.77 – 6.33 | 6.05 ± 0.15 (5.95–6.31) | 9 | Mixed | 710 | 86.5% | 59.4 | <0.005 | 0.48 | Random |
| female | All | 20-30 | 6.27 | 6.12 – 6.42 | — (6.27–6.27) | 1 | Observational Cohort | 25 | — | — | — | — | Single study |
| female | All | 30-40 | 6.23 | 6.03 – 6.43 | — (6.23–6.23) | 1 | Observational Cohort | 25 | — | — | — | — | Single study |
| female | All | 40-50 | 6.15 | 5.52 – 6.79 | 6.11 ± 0.09 (6.01–6.18) | 3 | Observational Cohort | 578 | 57.9% | 4.8 | 0.09 | 0.55 | Fixed |
| female | All | 50-60 | 6.02 | 5.68 – 6.35 | 6.02 ± 0.17 (5.84–6.18) | 3 | Observational Cohort | 2565 | 99.6% | 556.2 | <0.005 | 0.96 | Random |
| female | All | 60+ | 6.10 | 5.82 – 6.37 | 6.10 ± 0.14 (5.89–6.22) | 5 | Observational Cohort | 496 | 93.2% | 58.6 | <0.005 | <0.05 | Random |
| **female** | **All** | **Meta-Regression** | **6.27** | **5.26 – 7.28** | **—** | **9** | **Mixed** | **710** | **0%** | **0** | **1.00** | **0.48** | **Multivariable** |
| **female** | **All** | **Overall** | **6.09** | **5.82 – 6.35** | **6.08 ± 0.14 (5.84–6.31)** | **22** | **Mixed** | **4399** | **98%** | **1050.1** | **<0.005** | **0.66** | **Random** |
| female | black | 18+ | — | NA – NA | — | NA | NA | NA | — | — | — | — | NA |
| female | black | 20-30 | — | NA – NA | — | NA | NA | NA | — | — | — | — | NA |
| female | black | 30-40 | — | NA – NA | — | NA | NA | NA | — | — | — | — | NA |
| female | black | 40-50 | 6.01 | 5.07 – 6.95 | — (6.01–6.01) | 1 | Observational Cohort | 29 | — | — | — | — | Single study |
| female | black | 50-60 | 6.03 | 4.99 – 7.07 | — (6.03–6.03) | 1 | Observational Cohort | 131 | — | — | — | — | Single study |
| female | black | 60+ | 6.19 | 5.09 – 7.29 | 6.19 ± 0.00 (6.19–6.19) | 2 | Observational Cohort | 144 | 0% | 0 | 1.00 | — | Fixed |
| **female** | **black** | **Meta-Regression** | **—** | **NA – NA** | **—** | **NA** | **NA** | **NA** | **—** | **—** | **—** | **—** | **NA** |
| **female** | **black** | **Overall** | **6.10** | **5.04 – 7.15** | **6.11 ± 0.10 (6.01–6.19)** | **4** | **Observational Cohort** | **304** | **57.4%** | **7** | **0.07** | **0.74** | **Fixed** |
| female | mixed | 18+ | 6.05 | 5.77 – 6.33 | 6.05 ± 0.15 (5.95–6.31) | 9 | Mixed | 710 | 86.5% | 59.4 | <0.005 | 0.48 | Random |
| female | mixed | 20-30 | 6.27 | 6.12 – 6.42 | — (6.27–6.27) | 1 | Observational Cohort | 25 | — | — | — | — | Single study |
| female | mixed | 30-40 | 6.23 | 6.03 – 6.43 | — (6.23–6.23) | 1 | Observational Cohort | 25 | — | — | — | — | Single study |
| female | mixed | 40-50 | 6.14 | 5.96 – 6.32 | — (6.14–6.14) | 1 | Observational Cohort | 25 | — | — | — | — | Single study |
| female | mixed | 50-60 | 5.84 | 5.74 – 5.94 | — (5.84–5.84) | 1 | Observational Cohort | 25 | — | — | — | — | Single study |
| female | mixed | 60+ | 5.89 | 5.65 – 6.13 | — (5.89–5.89) | 1 | Observational Cohort | 25 | — | — | — | — | Single study |
| **female** | **mixed** | **Meta-Regression** | **—** | **NA – NA** | **—** | **NA** | **NA** | **NA** | **—** | **—** | **—** | **—** | **NA** |
| **female** | **mixed** | **Overall** | **6.06** | **5.75 – 6.37** | **6.06 ± 0.16 (5.84–6.31)** | **14** | **Mixed** | **835** | **98.4%** | **837.6** | **<0.005** | **0.44** | **Random** |
| female | white | 18+ | — | NA – NA | — | NA | NA | NA | — | — | — | — | NA |
| female | white | 20-30 | — | NA – NA | — | NA | NA | NA | — | — | — | — | NA |
| female | white | 30-40 | — | NA – NA | — | NA | NA | NA | — | — | — | — | NA |
| female | white | 40-50 | 6.18 | 5.24 – 7.12 | — (6.18–6.18) | 1 | Observational Cohort | 524 | — | — | — | — | Single study |
| female | white | 50-60 | 6.18 | 5.18 – 7.18 | — (6.18–6.18) | 1 | Observational Cohort | 2409 | — | — | — | — | Single study |
| female | white | 60+ | 6.12 | 5.87 – 6.37 | 6.12 ± 0.13 (6.03–6.22) | 2 | Observational Cohort | 327 | 90.6% | 10.6 | <0.005 | — | Random |
| **female** | **white** | **Meta-Regression** | **—** | **NA – NA** | **—** | **NA** | **NA** | **NA** | **—** | **—** | **—** | **—** | **NA** |
| **female** | **white** | **Overall** | **6.15** | **6.00 – 6.30** | **6.15 ± 0.08 (6.03–6.22)** | **4** | **Observational Cohort** | **3260** | **86.3%** | **21.8** | **<0.005** | **0.60** | **Random** |
| *I² represents heterogeneity; Q test assesses between-study variation; Egger's p = linear regression test for funnel plot asymmetry; Meta-Regression shows covariate-adjusted estimates* | | | | | | | | | | | | | |

###### 6.6.2.3 Table of Results subdivided by race / Sex / Age

ph_ma_3rd_analysis$race_summary_table

Table 1: **race-Stratified Meta-Analysis Results**

24-Hour Urine pH

| Sex Group | race | Age Band | 95% Range | Studies | Study Type | Participants | I² | Cochran's Q | Q (p-value) | Egger's p | Model |
| --- | --- | --- | --- | --- | --- | --- | --- | --- | --- | --- | --- |
| **main meta-analysis** | **All races** | **18+** | **5.68 – 6.30** | **67** | **Mixed** | **4826** | **85%** | **441.1** | **<0.005** | **<0.05** | **Random** |
| **main meta-analysis** | **All races** | **20-30** | **5.93 – 6.55** | **3** | **Mixed** | **58** | **98.7%** | **155.5** | **<0.005** | **0.83** | **Random** |
| **main meta-analysis** | **All races** | **30-40** | **5.85 – 6.32** | **5** | **Observational Cohort** | **185** | **99%** | **397.4** | **<0.005** | **0.16** | **Random** |
| **main meta-analysis** | **All races** | **40-50** | **6.01 – 6.20** | **6** | **Observational Cohort** | **631** | **82.4%** | **28.4** | **<0.005** | **1.00** | **Random** |
| **main meta-analysis** | **All races** | **50-60** | **5.78 – 6.31** | **6** | **Observational Cohort** | **2701** | **99.2%** | **620** | **<0.005** | **0.83** | **Random** |
| **main meta-analysis** | **All races** | **60+** | **5.75 – 6.30** | **8** | **Observational Cohort** | **825** | **90.9%** | **77.1** | **<0.005** | **0.15** | **Random** |
| **main meta-analysis** | **All races** | **Meta-Regression** | **5.63 – 6.27** | **52** | **Mixed** | **4379** | **88.1%** | **324.2** | **<0.005** | **0.35** | **Multivariable** |
| **main meta-analysis** | **All races** | **Meta-Regression** | **5.63 – 6.27** | **52** | **Mixed** | **4379** | **88.1%** | **324.2** | **<0.005** | **0.35** | **Multivariable** |
| **main meta-analysis** | **All races** | **Overall** | **5.72 – 6.32** | **95** | **Mixed** | **9226** | **96.2%** | **2494.5** | **<0.005** | **0.51** | **Random** |
| **main meta-analysis** | **All races** | **Overall** | **5.72 – 6.32** | **95** | **Mixed** | **9226** | **96.2%** | **2494.5** | **<0.005** | **0.51** | **Random** |
| main meta-analysis | Black | 18+ | 5.47 – 6.93 | 3 | Observational Cohort | 51 | 0% | 0 | 1.00 | — | Fixed |
| main meta-analysis | Black | 20-30 | — | — | NA | — | — | — | — | — | — |
| main meta-analysis | Black | 30-40 | — | — | NA | — | — | — | — | — | — |
| main meta-analysis | Black | 40-50 | 5.07 – 6.95 | 1 | Observational Cohort | 29 | — | — | — | — | Single study |
| main meta-analysis | Black | 50-60 | 4.99 – 7.07 | 1 | Observational Cohort | 131 | — | — | — | — | Single study |
| main meta-analysis | Black | 60+ | 5.09 – 7.29 | 2 | Observational Cohort | 144 | 0% | 0 | 1.00 | — | Fixed |
| main meta-analysis | Black | Meta-Regression | — | — | NA | — | — | — | — | — | — |
| **main meta-analysis** | **Black** | **Overall** | **5.15 – 7.10** | **7** | **Observational Cohort** | **355** | **39.5%** | **9.9** | **0.13** | **0.21** | **Fixed** |
| main meta-analysis | Mixed | 18+ | 5.65 – 6.29 | 58 | Mixed | 4610 | 86.7% | 427.8 | <0.005 | <0.01 | Random |
| main meta-analysis | Mixed | 20-30 | 5.93 – 6.55 | 3 | Mixed | 58 | 98.7% | 155.5 | <0.005 | 0.83 | Random |
| main meta-analysis | Mixed | 30-40 | 5.85 – 6.32 | 5 | Observational Cohort | 185 | 99% | 397.4 | <0.005 | 0.16 | Random |
| main meta-analysis | Mixed | 40-50 | 6.02 – 6.16 | 3 | Observational Cohort | 72 | 82% | 11.1 | <0.005 | 0.52 | Random |
| main meta-analysis | Mixed | 50-60 | 5.68 – 6.39 | 3 | Observational Cohort | 82 | 99.1% | 221.5 | <0.005 | 0.62 | Random |
| main meta-analysis | Mixed | 60+ | 5.65 – 6.13 | 1 | Observational Cohort | 25 | — | — | — | — | Single study |
| main meta-analysis | Mixed | Meta-Regression | — | — | NA | — | — | — | — | — | — |
| **main meta-analysis** | **Mixed** | **Overall** | **5.68 – 6.33** | **73** | **Mixed** | **5032** | **96.6%** | **2104.7** | **<0.005** | **0.44** | **Random** |
| main meta-analysis | White | 18+ | 5.37 – 6.71 | 6 | Observational Cohort | 165 | 0% | 2.2 | 0.82 | <0.001 | Fixed |
| main meta-analysis | White | 20-30 | — | — | NA | — | — | — | — | — | — |
| main meta-analysis | White | 30-40 | — | — | NA | — | — | — | — | — | — |
| main meta-analysis | White | 40-50 | 5.24 – 7.12 | 2 | Observational Cohort | 530 | 0% | 1 | 0.32 | — | Fixed |
| main meta-analysis | White | 50-60 | 5.76 – 6.38 | 2 | Observational Cohort | 2488 | 95% | 19.8 | <0.005 | — | Random |
| main meta-analysis | White | 60+ | 5.74 – 6.25 | 5 | Observational Cohort | 656 | 90.9% | 44.2 | <0.005 | 0.55 | Random |
| main meta-analysis | White | Meta-Regression | — | — | NA | — | — | — | — | — | — |
| **main meta-analysis** | **White** | **Overall** | **5.84 – 6.24** | **15** | **Observational Cohort** | **3839** | **95%** | **279** | **<0.005** | **0.30** | **Random** |
| **male** | **All races** | **18+** | **5.85 – 6.32** | **4** | **Observational Cohort** | **182** | **84.4%** | **19.2** | **<0.005** | **0.42** | **Random** |
| **male** | **All races** | **20-30** | **5.80 – 6.65** | **2** | **Mixed** | **33** | **99.3%** | **151** | **<0.005** | **—** | **Random** |
| **male** | **All races** | **30-40** | **5.80 – 6.24** | **3** | **Observational Cohort** | **143** | **98.7%** | **158.2** | **<0.005** | **0.62** | **Random** |
| **male** | **All races** | **40-50** | **5.92 – 6.22** | **2** | **Observational Cohort** | **31** | **0%** | **0.1** | **0.75** | **—** | **Fixed** |
| **male** | **All races** | **50-60** | **5.77 – 6.36** | **2** | **Observational Cohort** | **104** | **93.9%** | **16.3** | **<0.005** | **—** | **Random** |
| **male** | **All races** | **60+** | **5.83 – 6.00** | **3** | **Observational Cohort** | **329** | **71.1%** | **6.9** | **<0.05** | **0.54** | **Random** |
| **male** | **All races** | **Meta-Regression** | **5.67 – 6.52** | **4** | **Mixed** | **261** | **99.1%** | **665.5** | **<0.005** | **0.40** | **Univariable** |
| **male** | **All races** | **Overall** | **5.78 – 6.32** | **16** | **Mixed** | **822** | **98.5%** | **970.1** | **<0.005** | **0.15** | **Random** |
| male | Black | 18+ | — | — | NA | — | — | — | — | — | — |
| male | Black | 20-30 | — | — | NA | — | — | — | — | — | — |
| male | Black | 30-40 | — | — | NA | — | — | — | — | — | — |
| male | Black | 40-50 | — | — | NA | — | — | — | — | — | — |
| male | Black | 50-60 | — | — | NA | — | — | — | — | — | — |
| male | Black | 60+ | — | — | NA | — | — | — | — | — | — |
| male | Black | Meta-Regression | — | — | NA | — | — | — | — | — | — |
| **male** | **Black** | **Overall** | **—** | **—** | **NA** | **—** | **—** | **—** | **—** | **—** | **—** |
| male | Mixed | 18+ | 5.85 – 6.32 | 4 | Observational Cohort | 182 | 84.4% | 19.2 | <0.005 | 0.42 | Random |
| male | Mixed | 20-30 | 5.80 – 6.65 | 2 | Mixed | 33 | 99.3% | 151 | <0.005 | — | Random |
| male | Mixed | 30-40 | 5.80 – 6.24 | 3 | Observational Cohort | 143 | 98.7% | 158.2 | <0.005 | 0.62 | Random |
| male | Mixed | 40-50 | 5.93 – 6.21 | 1 | Observational Cohort | 25 | — | — | — | — | Single study |
| male | Mixed | 50-60 | 5.97 – 6.37 | 1 | Observational Cohort | 25 | — | — | — | — | Single study |
| male | Mixed | 60+ | — | — | NA | — | — | — | — | — | — |
| male | Mixed | Meta-Regression | — | — | NA | — | — | — | — | — | — |
| **male** | **Mixed** | **Overall** | **5.84 – 6.36** | **11** | **Mixed** | **408** | **98.9%** | **931** | **<0.005** | **0.07** | **Random** |
| male | White | 18+ | — | — | NA | — | — | — | — | — | — |
| male | White | 20-30 | — | — | NA | — | — | — | — | — | — |
| male | White | 30-40 | — | — | NA | — | — | — | — | — | — |
| male | White | 40-50 | 5.26 – 6.78 | 1 | Observational Cohort | 6 | — | — | — | — | Single study |
| male | White | 50-60 | 5.07 – 6.83 | 1 | Observational Cohort | 79 | — | — | — | — | Single study |
| male | White | 60+ | 5.83 – 6.00 | 3 | Observational Cohort | 329 | 71.1% | 6.9 | <0.05 | 0.54 | Random |
| male | White | Meta-Regression | — | — | NA | — | — | — | — | — | — |
| **male** | **White** | **Overall** | **5.45 – 6.42** | **5** | **Observational Cohort** | **414** | **45.2%** | **7.3** | **0.12** | **0.71** | **Fixed** |
| **female** | **All races** | **18+** | **5.77 – 6.33** | **9** | **Mixed** | **710** | **86.5%** | **59.4** | **<0.005** | **0.48** | **Random** |
| **female** | **All races** | **20-30** | **6.12 – 6.42** | **1** | **Observational Cohort** | **25** | **—** | **—** | **—** | **—** | **Single study** |
| **female** | **All races** | **30-40** | **6.03 – 6.43** | **1** | **Observational Cohort** | **25** | **—** | **—** | **—** | **—** | **Single study** |
| **female** | **All races** | **40-50** | **5.52 – 6.79** | **3** | **Observational Cohort** | **578** | **57.9%** | **4.8** | **0.09** | **0.55** | **Fixed** |
| **female** | **All races** | **50-60** | **5.68 – 6.35** | **3** | **Observational Cohort** | **2565** | **99.6%** | **556.2** | **<0.005** | **0.96** | **Random** |
| **female** | **All races** | **60+** | **5.82 – 6.37** | **5** | **Observational Cohort** | **496** | **93.2%** | **58.6** | **<0.005** | **<0.05** | **Random** |
| **female** | **All races** | **Meta-Regression** | **5.26 – 7.28** | **9** | **Mixed** | **710** | **0%** | **0** | **1.00** | **0.48** | **Multivariable** |
| **female** | **All races** | **Overall** | **5.82 – 6.35** | **22** | **Mixed** | **4399** | **98%** | **1050.1** | **<0.005** | **0.66** | **Random** |
| female | Black | 18+ | — | — | NA | — | — | — | — | — | — |
| female | Black | 20-30 | — | — | NA | — | — | — | — | — | — |
| female | Black | 30-40 | — | — | NA | — | — | — | — | — | — |
| female | Black | 40-50 | 5.07 – 6.95 | 1 | Observational Cohort | 29 | — | — | — | — | Single study |
| female | Black | 50-60 | 4.99 – 7.07 | 1 | Observational Cohort | 131 | — | — | — | — | Single study |
| female | Black | 60+ | 5.09 – 7.29 | 2 | Observational Cohort | 144 | 0% | 0 | 1.00 | — | Fixed |
| female | Black | Meta-Regression | — | — | NA | — | — | — | — | — | — |
| **female** | **Black** | **Overall** | **5.04 – 7.15** | **4** | **Observational Cohort** | **304** | **57.4%** | **7** | **0.07** | **0.74** | **Fixed** |
| female | Mixed | 18+ | 5.77 – 6.33 | 9 | Mixed | 710 | 86.5% | 59.4 | <0.005 | 0.48 | Random |
| female | Mixed | 20-30 | 6.12 – 6.42 | 1 | Observational Cohort | 25 | — | — | — | — | Single study |
| female | Mixed | 30-40 | 6.03 – 6.43 | 1 | Observational Cohort | 25 | — | — | — | — | Single study |
| female | Mixed | 40-50 | 5.96 – 6.32 | 1 | Observational Cohort | 25 | — | — | — | — | Single study |
| female | Mixed | 50-60 | 5.74 – 5.94 | 1 | Observational Cohort | 25 | — | — | — | — | Single study |
| female | Mixed | 60+ | 5.65 – 6.13 | 1 | Observational Cohort | 25 | — | — | — | — | Single study |
| female | Mixed | Meta-Regression | — | — | NA | — | — | — | — | — | — |
| **female** | **Mixed** | **Overall** | **5.75 – 6.37** | **14** | **Mixed** | **835** | **98.4%** | **837.6** | **<0.005** | **0.44** | **Random** |
| female | White | 18+ | — | — | NA | — | — | — | — | — | — |
| female | White | 20-30 | — | — | NA | — | — | — | — | — | — |
| female | White | 30-40 | — | — | NA | — | — | — | — | — | — |
| female | White | 40-50 | 5.24 – 7.12 | 1 | Observational Cohort | 524 | — | — | — | — | Single study |
| female | White | 50-60 | 5.18 – 7.18 | 1 | Observational Cohort | 2409 | — | — | — | — | Single study |
| female | White | 60+ | 5.87 – 6.37 | 2 | Observational Cohort | 327 | 90.6% | 10.6 | <0.005 | — | Random |
| female | White | Meta-Regression | — | — | NA | — | — | — | — | — | — |
| **female** | **White** | **Overall** | **6.00 – 6.30** | **4** | **Observational Cohort** | **3260** | **86.3%** | **21.8** | **<0.005** | **0.60** | **Random** |
| *I² represents heterogeneity; Q test assesses between-study variation; Egger's p = linear regression test for funnel plot asymmetry; — indicates insufficient data* | | | | | | | | | | | |

###### 6.6.2.4 Wald Tests

ph_ma_3rd_analysis$wald_tests$sex_comparison

Table 1: **Sex Comparison (Overall)**

| Comparison | % Shift | P (Wald Test) | Mean Diff (95% CI) |
| --- | --- | --- | --- |
| Male vs Female (Overall) | -0.6% | 0.857 | -0.03 [-0.42, 0.35] |
| *Mean difference = Group 1 − Group 2 in component units (mmol/24h, L/24h, or pH); % shift = difference relative to Group 2.* | | | |

ph_ma_3rd_analysis$wald_tests$race_comparisons

Table 1: **race Comparisons Within Sex Groups**

| Sex Group | Comparison | % Shift | P (Wald Test) | Mean Diff (95% CI) |
| --- | --- | --- | --- | --- |
| male | black vs mixed | — | — | — |
| male | black vs white | — | — | — |
| male | mixed vs white | +2.8% | 0.550 | 0.17 [-0.38, 0.71] |
| female | black vs mixed | +0.6% | 0.946 | 0.04 [-1.06, 1.14] |
| female | black vs white | -0.9% | 0.921 | -0.05 [-1.12, 1.01] |
| female | mixed vs white | -1.5% | 0.604 | -0.09 [-0.44, 0.25] |
| *Mean difference = Group 1 − Group 2 in component units (mmol/24h, L/24h, or pH); % shift = difference relative to Group 2.* | | | | |

ph_ma_3rd_analysis$wald_tests$age_band_comparisons

Table 1: **Age Band Comparisons Within Sex Groups**

| Sex Group | Comparison | % Shift | P (Wald Test) | Mean Diff (95% CI) |
| --- | --- | --- | --- | --- |
| main meta-analysis | 18+ vs 20-30 | -4.0% | 0.258 | -0.25 [-0.68, 0.18] |
| main meta-analysis | 18+ vs 30-40 | -1.5% | 0.638 | -0.09 [-0.48, 0.29] |
| main meta-analysis | 18+ vs 40-50 | -1.9% | 0.484 | -0.11 [-0.44, 0.21] |
| main meta-analysis | 18+ vs 50-60 | -0.9% | 0.792 | -0.05 [-0.46, 0.35] |
| main meta-analysis | 18+ vs 60+ | -0.6% | 0.872 | -0.03 [-0.44, 0.38] |
| main meta-analysis | 20-30 vs 30-40 | +2.6% | 0.429 | 0.16 [-0.23, 0.54] |
| main meta-analysis | 20-30 vs 40-50 | +2.2% | 0.410 | 0.13 [-0.19, 0.46] |
| main meta-analysis | 20-30 vs 50-60 | +3.2% | 0.349 | 0.19 [-0.21, 0.60] |
| main meta-analysis | 20-30 vs 60+ | +3.6% | 0.300 | 0.22 [-0.19, 0.62] |
| main meta-analysis | 30-40 vs 40-50 | -0.4% | 0.870 | -0.02 [-0.28, 0.24] |
| main meta-analysis | 30-40 vs 50-60 | +0.6% | 0.834 | 0.04 [-0.32, 0.40] |
| main meta-analysis | 30-40 vs 60+ | +1.0% | 0.747 | 0.06 [-0.30, 0.42] |
| main meta-analysis | 40-50 vs 50-60 | +1.0% | 0.682 | 0.06 [-0.23, 0.35] |
| main meta-analysis | 40-50 vs 60+ | +1.3% | 0.582 | 0.08 [-0.21, 0.37] |
| main meta-analysis | 50-60 vs 60+ | +0.4% | 0.914 | 0.02 [-0.36, 0.40] |
| male | 18+ vs 20-30 | -2.3% | 0.571 | -0.14 [-0.63, 0.35] |
| male | 18+ vs 30-40 | +1.1% | 0.681 | 0.07 [-0.25, 0.39] |
| male | 18+ vs 40-50 | +0.2% | 0.919 | 0.01 [-0.26, 0.29] |
| male | 18+ vs 50-60 | +0.3% | 0.920 | 0.02 [-0.36, 0.40] |
| male | 18+ vs 60+ | +2.9% | 0.179 | 0.17 [-0.08, 0.42] |
| male | 20-30 vs 30-40 | +3.5% | 0.397 | 0.21 [-0.27, 0.69] |
| male | 20-30 vs 40-50 | +2.6% | 0.503 | 0.16 [-0.30, 0.61] |
| male | 20-30 vs 50-60 | +2.6% | 0.546 | 0.16 [-0.36, 0.68] |
| male | 20-30 vs 60+ | +5.3% | 0.162 | 0.31 [-0.13, 0.75] |
| male | 30-40 vs 40-50 | -0.9% | 0.700 | -0.05 [-0.32, 0.21] |
| male | 30-40 vs 50-60 | -0.8% | 0.799 | -0.05 [-0.42, 0.32] |
| male | 30-40 vs 60+ | +1.7% | 0.391 | 0.10 [-0.13, 0.34] |
| male | 40-50 vs 50-60 | +0.1% | 0.978 | 0.00 [-0.33, 0.34] |
| male | 40-50 vs 60+ | +2.6% | 0.081 | 0.16 [-0.02, 0.33] |
| male | 50-60 vs 60+ | +2.6% | 0.335 | 0.15 [-0.16, 0.46] |
| female | 18+ vs 20-30 | -3.5% | 0.182 | -0.22 [-0.54, 0.10] |
| female | 18+ vs 30-40 | -2.9% | 0.309 | -0.18 [-0.52, 0.17] |
| female | 18+ vs 40-50 | -1.7% | 0.774 | -0.10 [-0.80, 0.59] |
| female | 18+ vs 50-60 | +0.6% | 0.876 | 0.04 [-0.41, 0.48] |
| female | 18+ vs 60+ | -0.7% | 0.824 | -0.04 [-0.44, 0.35] |
| female | 20-30 vs 30-40 | +0.6% | 0.751 | 0.04 [-0.21, 0.29] |
| female | 20-30 vs 40-50 | +1.9% | 0.727 | 0.12 [-0.54, 0.77] |
| female | 20-30 vs 50-60 | +4.2% | 0.178 | 0.25 [-0.12, 0.62] |
| female | 20-30 vs 60+ | +2.9% | 0.271 | 0.17 [-0.14, 0.48] |
| female | 30-40 vs 40-50 | +1.2% | 0.822 | 0.08 [-0.59, 0.74] |
| female | 30-40 vs 50-60 | +3.6% | 0.283 | 0.21 [-0.18, 0.60] |
| female | 30-40 vs 60+ | +2.2% | 0.432 | 0.13 [-0.20, 0.47] |
| female | 40-50 vs 50-60 | +2.3% | 0.709 | 0.14 [-0.58, 0.86] |
| female | 40-50 vs 60+ | +0.9% | 0.870 | 0.06 [-0.63, 0.75] |
| female | 50-60 vs 60+ | -1.3% | 0.719 | -0.08 [-0.51, 0.35] |
| *Mean difference = Group 1 − Group 2 in component units (mmol/24h, L/24h, or pH); % shift = difference relative to Group 2.* | | | | |

ph_ma_3rd_analysis$wald_tests$sex_within_age_bands

Table 1: **Sex Comparisons Within Age Bands**

| Age Band | % Shift | P (Wald Test) | Mean Diff (95% CI) |
| --- | --- | --- | --- |
| 18+ | +0.5% | 0.861 | 0.03 [-0.33, 0.40] |
| 20-30 | -0.7% | 0.846 | -0.04 [-0.50, 0.41] |
| 30-40 | -3.4% | 0.156 | -0.21 [-0.51, 0.08] |
| 40-50 | -1.4% | 0.801 | -0.08 [-0.74, 0.57] |
| 50-60 | +0.8% | 0.832 | 0.05 [-0.40, 0.50] |
| 60+ | -3.0% | 0.209 | -0.18 [-0.47, 0.10] |
| *Mean difference = Group 1 − Group 2 in component units (mmol/24h, L/24h, or pH); % shift = difference relative to Group 2.* | | | |

#### 6.7 Citrate

##### 6.7.1 Comparison by Age Band

###### 6.7.1.1 Plot

```
citrate_ma_2nd_analysis <- sex_age_subdivision_ma("citrate")
## Not enough studies for multivariable meta-regression (k = 4, need > 6). Running univariable models instead.
citrate_ma_2nd_analysis$combined_plot
## `height` was translated to `width`.
## `height` was translated to `width`.
## `height` was translated to `width`.
```

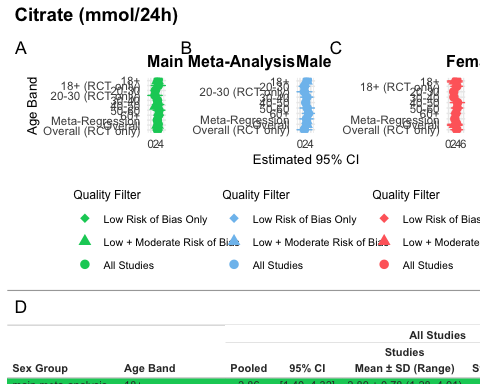

###### 6.7.1.2 Summary Table

citrate_ma_2nd_analysis$combined_summary_table

Table 1: **Combined Meta-Analysis Results**

24-Hour Urine Citrate (mmol/24h)

|  |  | **All Studies** | | | | | | **Low Risk of Bias Only** | | | | | | **Low + Moderate RoB** | | | | | |
| --- | --- | --- | --- | --- | --- | --- | --- | --- | --- | --- | --- | --- | --- | --- | --- | --- | --- | --- | --- |
| Sex Group | Age Band | Pooled | 95% CI | Studies  Mean ± SD (Range) | Studies, n | Q p-value | Egger's p | Pooled | 95% CI | Studies  Mean ± SD (Range) | Studies, n | Q p-value | Egger's p | Pooled | 95% CI | Studies  Mean ± SD (Range) | Studies, n | Q p-value | Egger's p |
| main meta-analysis | 18+ | 2.86 | [1.40, 4.32] | 2.89 ± 0.78 (1.28–4.91) | 67 | <0.001 | <0.005 | 2.47 | [1.51, 3.44] | 2.52 ± 0.57 (1.78–3.99) | 27 | <0.001 | 0.96 | 2.61 | [1.21, 4.00] | 2.65 ± 0.77 (1.28–4.91) | 34 | <0.001 | 0.72 |
| **main meta-analysis** | **18+ (RCT only)** | **2.87** | **[0.19, 5.54]** | **3.00 ± 0.52 (2.26–3.61)** | **7** | **0.07** | **0.35** | **—** | **—** | **—** | **—** | **—** | **—** | **—** | **—** | **—** | **—** | **—** | **—** |
| main meta-analysis | 20-30 | 1.75 | [0.13, 3.37] | 1.75 ± 0.83 (0.64–2.61) | 4 | <0.001 | 0.91 | — | — | — | — | — | — | — | — | — | — | — | — |
| **main meta-analysis** | **20-30 (RCT only)** | **1.62** | **[-1.10, 4.35]** | **1.62 ± 1.39 (0.64–2.61)** | **2** | **<0.001** | **—** | **—** | **—** | **—** | **—** | **—** | **—** | **—** | **—** | **—** | **—** | **—** | **—** |
| main meta-analysis | 30-40 | 2.56 | [1.04, 4.07] | 2.56 ± 0.76 (1.39–3.65) | 6 | <0.001 | 0.36 | — | — | — | — | — | — | — | — | — | — | — | — |
| main meta-analysis | 40-50 | 2.81 | [0.56, 5.07] | 2.83 ± 1.17 (1.68–4.54) | 7 | <0.001 | 0.28 | — | — | — | — | — | — | 1.86 | [1.51, 2.21] | — (1.86–1.86) | 1 | — | — |
| main meta-analysis | 50-60 | 3.13 | [0.90, 5.36] | 3.13 ± 1.14 (1.71–4.15) | 5 | <0.001 | 0.89 | — | — | — | — | — | — | — | — | — | — | — | — |
| main meta-analysis | 60+ | 3.01 | [1.18, 4.83] | 3.02 ± 0.93 (1.33–3.85) | 8 | <0.001 | <0.05 | — | — | — | — | — | — | — | — | — | — | — | — |
| **main meta-analysis** | **Meta-Regression** | **2.52** | **[1.11, 3.93]** | **—** | **51** | **<0.001** | **0.93** | **—** | **—** | **—** | **—** | **—** | **—** | **—** | **—** | **—** | **—** | **—** | **—** |
| **main meta-analysis** | **Overall** | **2.82** | **[1.17, 4.47]** | **2.84 ± 0.86 (0.64–4.91)** | **97** | **<0.001** | **0.15** | **2.47** | **[1.51, 3.44]** | **2.52 ± 0.57 (1.78–3.99)** | **27** | **<0.001** | **0.96** | **2.58** | **[1.19, 3.97]** | **2.62 ± 0.77 (1.28–4.91)** | **35** | **<0.001** | **0.85** |
| **main meta-analysis** | **Overall (RCT only)** | **2.61** | **[0.87, 4.36]** | **2.70 ± 0.90 (0.64–3.61)** | **9** | **<0.001** | **0.13** | **—** | **—** | **—** | **—** | **—** | **—** | **—** | **—** | **—** | **—** | **—** | **—** |
| male | 18+ | 2.71 | [1.63, 3.79] | 2.79 ± 0.66 (2.41–3.56) | 3 | <0.01 | 0.46 | 2.41 | [2.17, 2.65] | — (2.41–2.41) | 1 | — | — | 2.41 | [2.17, 2.65] | — (2.41–2.41) | 1 | — | — |
| **male** | **18+ (RCT only)** | **—** | **—** | **—** | **—** | **—** | **—** | **—** | **—** | **—** | **—** | **—** | **—** | **—** | **—** | **—** | **—** | **—** | **—** |
| male | 20-30 | 1.65 | [-0.28, 3.57] | 1.65 ± 0.99 (0.64–2.61) | 3 | <0.001 | 0.92 | — | — | — | — | — | — | — | — | — | — | — | — |
| **male** | **20-30 (RCT only)** | **1.62** | **[-1.10, 4.35]** | **1.62 ± 1.39 (0.64–2.61)** | **2** | **<0.001** | **—** | **—** | **—** | **—** | **—** | **—** | **—** | **—** | **—** | **—** | **—** | **—** | **—** |
| male | 30-40 | 2.53 | [0.26, 4.79] | 2.53 ± 1.13 (1.39–3.65) | 3 | <0.001 | 0.67 | — | — | — | — | — | — | — | — | — | — | — | — |
| male | 40-50 | 2.12 | [0.76, 3.47] | 2.23 ± 0.78 (1.68–2.78) | 2 | <0.05 | — | — | — | — | — | — | — | — | — | — | — | — | — |
| male | 50-60 | 2.66 | [0.02, 5.30] | 2.67 ± 1.35 (1.71–3.62) | 2 | <0.001 | — | — | — | — | — | — | — | — | — | — | — | — | — |
| male | 60+ | 3.15 | [0.97, 5.33] | 3.16 ± 1.11 (1.87–3.85) | 3 | <0.001 | 0.30 | — | — | — | — | — | — | — | — | — | — | — | — |
| **male** | **Meta-Regression** | **2.41** | **[-0.03, 4.86]** | **—** | **4** | **<0.001** | **0.88** | **—** | **—** | **—** | **—** | **—** | **—** | **—** | **—** | **—** | **—** | **—** | **—** |
| **male** | **Overall** | **2.49** | **[0.59, 4.40]** | **2.51 ± 0.97 (0.64–3.85)** | **16** | **<0.001** | **0.48** | **2.41** | **[2.17, 2.65]** | **— (2.41–2.41)** | **1** | **—** | **—** | **2.41** | **[2.17, 2.65]** | **— (2.41–2.41)** | **1** | **—** | **—** |
| **male** | **Overall (RCT only)** | **1.62** | **[-1.10, 4.35]** | **1.62 ± 1.39 (0.64–2.61)** | **2** | **<0.001** | **—** | **—** | **—** | **—** | **—** | **—** | **—** | **—** | **—** | **—** | **—** | **—** | **—** |
| female | 18+ | 3.29 | [1.98, 4.60] | 3.32 ± 0.70 (2.51–4.22) | 7 | <0.001 | <0.05 | 2.51 | [-0.55, 5.57] | 2.51 ± 0.00 (2.51–2.51) | 2 | 1.00 | — | 2.56 | [-0.46, 5.57] | 2.76 ± 0.44 (2.51–3.27) | 3 | 0.10 | <0.001 |
| **female** | **18+ (RCT only)** | **3.27** | **[0.95, 5.58]** | **— (3.27–3.27)** | **1** | **—** | **—** | **—** | **—** | **—** | **—** | **—** | **—** | **—** | **—** | **—** | **—** | **—** | **—** |
| female | 20-30 | 2.06 | [1.67, 2.45] | — (2.06–2.06) | 1 | — | — | — | — | — | — | — | — | — | — | — | — | — | — |
| **female** | **20-30 (RCT only)** | **—** | **—** | **—** | **—** | **—** | **—** | **—** | **—** | **—** | **—** | **—** | **—** | **—** | **—** | **—** | **—** | **—** | **—** |
| female | 30-40 | 2.14 | [1.67, 2.61] | — (2.14–2.14) | 1 | — | — | — | — | — | — | — | — | — | — | — | — | — | — |
| female | 40-50 | 3.58 | [0.77, 6.39] | 3.60 ± 1.44 (1.94–4.54) | 3 | <0.001 | 0.70 | — | — | — | — | — | — | — | — | — | — | — | — |
| female | 50-60 | 3.44 | [1.19, 5.69] | 3.44 ± 1.15 (2.12–4.15) | 3 | <0.001 | 0.83 | — | — | — | — | — | — | — | — | — | — | — | — |
| female | 60+ | 2.92 | [1.08, 4.76] | 2.93 ± 0.93 (1.33–3.62) | 5 | <0.001 | 0.06 | — | — | — | — | — | — | — | — | — | — | — | — |
| **female** | **Meta-Regression** | **2.51** | **[1.68, 3.34]** | **—** | **7** | **<0.001** | **<0.05** | **—** | **—** | **—** | **—** | **—** | **—** | **—** | **—** | **—** | **—** | **—** | **—** |
| **female** | **Overall** | **3.14** | **[1.33, 4.95]** | **3.16 ± 0.94 (1.33–4.54)** | **20** | **<0.001** | **0.51** | **2.51** | **[-0.55, 5.57]** | **2.51 ± 0.00 (2.51–2.51)** | **2** | **1.00** | **—** | **2.56** | **[-0.46, 5.57]** | **2.76 ± 0.44 (2.51–3.27)** | **3** | **0.10** | **<0.001** |
| **female** | **Overall (RCT only)** | **3.27** | **[0.95, 5.58]** | **— (3.27–3.27)** | **1** | **—** | **—** | **—** | **—** | **—** | **—** | **—** | **—** | **—** | **—** | **—** | **—** | **—** | **—** |
| *Studies, n = number of studies; Q p-value = Cochran's Q heterogeneity test; Egger's p = linear regression test for funnel plot asymmetry; RoB = Risk of Bias* | | | | | | | | | | | | | | | | | | | |

###### 6.7.1.3 Wald Tests

citrate_ma_2nd_analysis$wald_tests$sex_comparison

Table 1: **Sex Comparison (Overall)**

| Comparison | % Shift | P (Wald Test) | Mean Diff (95% CI) |
| --- | --- | --- | --- |
| Male vs Female (Overall) | -20.6% | 0.629 | -0.65 [-3.28, 1.98] |
| *Mean difference = Group 1 − Group 2 in component units (mmol/24h, L/24h, or pH); % shift = difference relative to Group 2.* | | | |

citrate_ma_2nd_analysis$wald_tests$age_band_comparisons

Table 1: **Age Band Comparisons Within Sex Groups**

| Sex Group | Comparison | % Shift | P (Wald Test) | Mean Diff (95% CI) |
| --- | --- | --- | --- | --- |
| main meta-analysis | 18+ vs 20-30 | +63.6% | 0.318 | 1.11 [-1.07, 3.29] |
| main meta-analysis | 18+ vs 30-40 | +12.0% | 0.775 | 0.31 [-1.80, 2.41] |
| main meta-analysis | 18+ vs 40-50 | +1.8% | 0.970 | 0.05 [-2.64, 2.74] |
| main meta-analysis | 18+ vs 50-60 | -8.5% | 0.845 | -0.27 [-2.93, 2.40] |
| main meta-analysis | 18+ vs 60+ | -4.8% | 0.904 | -0.14 [-2.48, 2.20] |
| main meta-analysis | 20-30 vs 30-40 | -31.5% | 0.477 | -0.81 [-3.02, 1.41] |
| main meta-analysis | 20-30 vs 40-50 | -37.8% | 0.454 | -1.06 [-3.84, 1.72] |
| main meta-analysis | 20-30 vs 50-60 | -44.1% | 0.327 | -1.38 [-4.14, 1.38] |
| main meta-analysis | 20-30 vs 60+ | -41.8% | 0.313 | -1.26 [-3.70, 1.19] |
| main meta-analysis | 30-40 vs 40-50 | -9.1% | 0.854 | -0.26 [-2.97, 2.46] |
| main meta-analysis | 30-40 vs 50-60 | -18.3% | 0.678 | -0.57 [-3.27, 2.12] |
| main meta-analysis | 30-40 vs 60+ | -15.0% | 0.710 | -0.45 [-2.83, 1.92] |
| main meta-analysis | 40-50 vs 50-60 | -10.1% | 0.845 | -0.32 [-3.49, 2.86] |
| main meta-analysis | 40-50 vs 60+ | -6.5% | 0.895 | -0.20 [-3.10, 2.71] |
| main meta-analysis | 50-60 vs 60+ | +4.0% | 0.934 | 0.12 [-2.76, 3.01] |
| male | 18+ vs 20-30 | +64.9% | 0.343 | 1.07 [-1.14, 3.28] |
| male | 18+ vs 30-40 | +7.5% | 0.882 | 0.19 [-2.32, 2.69] |
| male | 18+ vs 40-50 | +28.3% | 0.499 | 0.60 [-1.13, 2.33] |
| male | 18+ vs 50-60 | +2.1% | 0.969 | 0.06 [-2.80, 2.91] |
| male | 18+ vs 60+ | -13.8% | 0.727 | -0.43 [-2.87, 2.00] |
| male | 20-30 vs 30-40 | -34.8% | 0.561 | -0.88 [-3.85, 2.09] |
| male | 20-30 vs 40-50 | -22.2% | 0.695 | -0.47 [-2.82, 1.88] |
| male | 20-30 vs 50-60 | -38.1% | 0.543 | -1.01 [-4.28, 2.25] |
| male | 20-30 vs 60+ | -47.7% | 0.312 | -1.50 [-4.41, 1.41] |
| male | 30-40 vs 40-50 | +19.3% | 0.761 | 0.41 [-2.23, 3.04] |
| male | 30-40 vs 50-60 | -5.0% | 0.940 | -0.13 [-3.61, 3.34] |
| male | 30-40 vs 60+ | -19.8% | 0.698 | -0.62 [-3.76, 2.52] |
| male | 40-50 vs 50-60 | -20.4% | 0.720 | -0.54 [-3.51, 2.42] |
| male | 40-50 vs 60+ | -32.8% | 0.431 | -1.03 [-3.60, 1.54] |
| male | 50-60 vs 60+ | -15.5% | 0.780 | -0.49 [-3.91, 2.94] |
| female | 18+ vs 20-30 | +59.6% | 0.078 | 1.23 [-0.14, 2.59] |
| female | 18+ vs 30-40 | +53.6% | 0.106 | 1.15 [-0.24, 2.54] |
| female | 18+ vs 40-50 | -8.1% | 0.855 | -0.29 [-3.39, 2.81] |
| female | 18+ vs 50-60 | -4.4% | 0.909 | -0.15 [-2.75, 2.45] |
| female | 18+ vs 60+ | +12.5% | 0.751 | 0.37 [-1.89, 2.62] |
| female | 20-30 vs 30-40 | -3.7% | 0.798 | -0.08 [-0.69, 0.53] |
| female | 20-30 vs 40-50 | -42.4% | 0.295 | -1.52 [-4.35, 1.32] |
| female | 20-30 vs 50-60 | -40.1% | 0.236 | -1.38 [-3.66, 0.90] |
| female | 20-30 vs 60+ | -29.5% | 0.369 | -0.86 [-2.74, 1.02] |
| female | 30-40 vs 40-50 | -40.2% | 0.323 | -1.44 [-4.29, 1.41] |
| female | 30-40 vs 50-60 | -37.8% | 0.267 | -1.30 [-3.60, 1.00] |
| female | 30-40 vs 60+ | -26.8% | 0.419 | -0.78 [-2.68, 1.12] |
| female | 40-50 vs 50-60 | +4.0% | 0.941 | 0.14 [-3.46, 3.73] |
| female | 40-50 vs 60+ | +22.4% | 0.703 | 0.65 [-2.70, 4.01] |
| female | 50-60 vs 60+ | +17.7% | 0.727 | 0.52 [-2.39, 3.42] |
| *Mean difference = Group 1 − Group 2 in component units (mmol/24h, L/24h, or pH); % shift = difference relative to Group 2.* | | | | |

citrate_ma_2nd_analysis$wald_tests$sex_within_age_bands

Table 1: **Sex Comparisons Within Age Bands**

| Age Band | % Shift | P (Wald Test) | Mean Diff (95% CI) |
| --- | --- | --- | --- |
| 18+ | -17.4% | 0.508 | -0.57 [-2.27, 1.12] |
| 20-30 | -20.1% | 0.679 | -0.41 [-2.38, 1.55] |
| 30-40 | +18.0% | 0.744 | 0.39 [-1.92, 2.69] |
| 40-50 | -40.8% | 0.359 | -1.46 [-4.58, 1.66] |
| 50-60 | -22.7% | 0.659 | -0.78 [-4.25, 2.68] |
| 60+ | +7.7% | 0.877 | 0.23 [-2.63, 3.08] |
| *Mean difference = Group 1 − Group 2 in component units (mmol/24h, L/24h, or pH); % shift = difference relative to Group 2.* | | | |

###### 6.7.1.4 Meta-Regression (plain language)

format_meta_reg_plain("citrate", sex_group = "all")

Message

No meta-regression result for citrate / all

###### 6.7.1.5 Assay Subgroup Analysis

assay_subgroup_table("citrate")

Table 1: **Assay subgroup analysis — Citrate**

*Per-assay pooled estimates; categories with k < 3 listed but not pooled.*

| Assay category | Studies, n | Participants | 95% CI | I² | Q p-value | Egger's p | Note |
| --- | --- | --- | --- | --- | --- | --- | --- |
| Unreported | 64 | 6,294 | 2.90 [2.68, 3.12] | 99% | <0.001 | 0.012 |  |
| HPLC / chromatography | 16 | 2,336 | 2.24 [2.05, 2.42] | 83% | <0.001 | 0.055 |  |
| Citrate lyase / enzymatic | 11 | 471 | 2.75 [2.16, 3.34] | 100% | <0.001 | 0.699 |  |
| Enzymatic (kit) | 7 | 357 | 3.53 [3.03, 4.03] | 97% | <0.001 | <0.001 |  |

###### 6.7.1.6 Preservative Sensitivity (Litholink / acidified samples)

```
preservative_sensitivity_ma("citrate")
## # A tibble: 3 × 6
##   group               n_studies n_participants pooled_estimate lower_95 upper_95
##   <chr>                   <int>          <int>           <dbl>    <dbl>    <dbl>
## 1 All studies                97           9458            2.82     2.64     2.99
## 2 Excluding preserva…        94           9347            2.79     2.62     2.97
## 3 Preservative only           3            111            3.94     3.52     4.37
```

###### 6.7.1.7 Creatinine Completeness Sensitivity

creat_completeness_table("citrate")

Table 1: **Creatinine-completeness sensitivity — Citrate**

*Compares the all-studies pooled estimate against subsets defined by whether the study reported 24h urine creatinine. Studies that do not report creatinine cannot verify collection completeness; a substantial shift here flags possible bias from incomplete collections.*

| Subset | Studies, n | Participants | Range (95% CI) | Δ vs all-studies (%) | Flag |
| --- | --- | --- | --- | --- | --- |
| All studies | 97 | 9,458 | 2.82 [2.64, 2.99] | — | — |
| Creatinine reported only | 65 | 7,916 | 2.73 [2.51, 2.94] | -3.3% | OK |
| Creatinine NOT reported | 32 | 1,542 | 3.01 [2.73, 3.29] | +6.7% | OK |

##### 6.7.2 Comparison by Age / Sex / race

###### 6.7.2.1 Plot

```
citrate_ma_3rd_analysis <- sex_age_race_subdivision_ma("citrate",
                                                      urine_ma2,
                                                      overall_bias)
## Not enough studies for multivariable meta-regression (k = 4, need > 6). Running univariable models instead.
citrate_ma_3rd_analysis$race_plot
## `height` was translated to `width`.
## `height` was translated to `width`.
## `height` was translated to `width`.
```

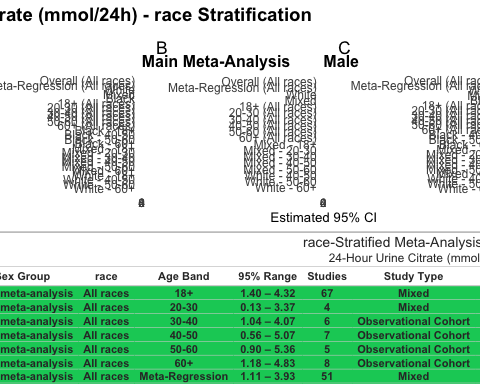

###### 6.7.2.2 Combined MA Results

citrate_ma_3rd_analysis$combined_summary_table

Table 1: **Combined Meta-Analysis Results**

24-Hour Urine Citrate (mmol/24h)

| Sex Group | race | Age Band | Pooled | 95% Range | Studies Mean SD Range | Studies | Study Type | Participants | I² | Cochran's Q | Q (p-value) | Egger's p | Model |
| --- | --- | --- | --- | --- | --- | --- | --- | --- | --- | --- | --- | --- | --- |
| main meta-analysis | All | 18+ | 2.86 | 1.40 – 4.32 | 2.89 ± 0.78 (1.28–4.91) | 67 | Mixed | 4913 | 97.8% | 3042.9 | <0.005 | <0.01 | Random |
| main meta-analysis | All | 20-30 | 1.75 | 0.13 – 3.37 | 1.75 ± 0.83 (0.64–2.61) | 4 | Mixed | 96 | 99.4% | 472.9 | <0.005 | 0.91 | Random |
| main meta-analysis | All | 30-40 | 2.56 | 1.04 – 4.07 | 2.56 ± 0.76 (1.39–3.65) | 6 | Observational Cohort | 290 | 99.8% | 3134.3 | <0.005 | 0.36 | Random |
| main meta-analysis | All | 40-50 | 2.81 | 0.56 – 5.07 | 2.83 ± 1.17 (1.68–4.54) | 7 | Observational Cohort | 665 | 99.6% | 1380.3 | <0.005 | 0.28 | Random |
| main meta-analysis | All | 50-60 | 3.13 | 0.90 – 5.36 | 3.13 ± 1.14 (1.71–4.15) | 5 | Observational Cohort | 2669 | 99.9% | 3028.6 | <0.005 | 0.89 | Random |
| main meta-analysis | All | 60+ | 3.01 | 1.18 – 4.83 | 3.02 ± 0.93 (1.33–3.85) | 8 | Observational Cohort | 825 | 99.4% | 1117.2 | <0.005 | <0.05 | Random |
| **main meta-analysis** | **All** | **Meta-Regression** | **2.52** | **1.11 – 3.93** | **—** | **51** | **Mixed** | **4337** | **96.3%** | **726** | **<0.005** | **0.93** | **Multivariable** |
| **main meta-analysis** | **All** | **Meta-Regression** | **2.52** | **1.11 – 3.93** | **—** | **51** | **Mixed** | **4337** | **96.3%** | **726** | **<0.005** | **0.93** | **Multivariable** |
| **main meta-analysis** | **All** | **Overall** | **2.82** | **1.17 – 4.47** | **2.84 ± 0.86 (0.64–4.91)** | **97** | **Mixed** | **9458** | **99.4%** | **16945** | **<0.005** | **0.15** | **Random** |
| **main meta-analysis** | **All** | **Overall** | **2.82** | **1.17 – 4.47** | **2.84 ± 0.86 (0.64–4.91)** | **97** | **Mixed** | **9458** | **99.4%** | **16945** | **<0.005** | **0.15** | **Random** |
| main meta-analysis | black | 18+ | 3.36 | 0.62 – 6.10 | 3.36 ± 0.00 (3.36–3.36) | 3 | Observational Cohort | 51 | 0% | 0 | 1.00 | — | Fixed |
| main meta-analysis | black | 20-30 | — | NA – NA | — | NA | NA | NA | — | — | — | — | NA |
| main meta-analysis | black | 30-40 | — | NA – NA | — | NA | NA | NA | — | — | — | — | NA |
| main meta-analysis | black | 40-50 | 4.54 | 0.76 – 8.32 | — (4.54–4.54) | 1 | Observational Cohort | 29 | — | — | — | — | Single study |
| main meta-analysis | black | 50-60 | 4.05 | 0.61 – 7.50 | — (4.05–4.05) | 1 | Observational Cohort | 131 | — | — | — | — | Single study |
| main meta-analysis | black | 60+ | 3.42 | 0.05 – 6.78 | 3.32 ± 0.33 (3.08–3.55) | 2 | Observational Cohort | 144 | 53.1% | 2.1 | 0.14 | — | Fixed |
| **main meta-analysis** | **black** | **Meta-Regression** | **—** | **NA – NA** | **—** | **NA** | **NA** | **NA** | **—** | **—** | **—** | **—** | **NA** |
| **main meta-analysis** | **black** | **Overall** | **3.62** | **2.87 – 4.38** | **3.62 ± 0.50 (3.08–4.54)** | **7** | **Observational Cohort** | **355** | **69.8%** | **19.8** | **<0.005** | **0.49** | **Random** |
| main meta-analysis | mixed | 18+ | 2.86 | 1.31 – 4.40 | 2.89 ± 0.82 (1.28–4.91) | 58 | Mixed | 4697 | 98.1% | 2990.1 | <0.005 | <0.01 | Random |
| main meta-analysis | mixed | 20-30 | 1.75 | 0.13 – 3.37 | 1.75 ± 0.83 (0.64–2.61) | 4 | Mixed | 96 | 99.4% | 472.9 | <0.005 | 0.91 | Random |
| main meta-analysis | mixed | 30-40 | 2.56 | 1.04 – 4.07 | 2.56 ± 0.76 (1.39–3.65) | 6 | Observational Cohort | 290 | 99.8% | 3134.3 | <0.005 | 0.36 | Random |
| main meta-analysis | mixed | 40-50 | 2.05 | 1.15 – 2.94 | 2.05 ± 0.46 (1.68–2.72) | 4 | Observational Cohort | 106 | 98.9% | 280.3 | <0.005 | 0.39 | Random |
| main meta-analysis | mixed | 50-60 | 1.91 | 1.35 – 2.48 | 1.92 ± 0.29 (1.71–2.12) | 2 | Observational Cohort | 50 | 97.9% | 47.2 | <0.005 | — | Random |
| main meta-analysis | mixed | 60+ | 1.33 | 0.98 – 1.68 | — (1.33–1.33) | 1 | Observational Cohort | 25 | — | — | — | — | Single study |
| **main meta-analysis** | **mixed** | **Meta-Regression** | **—** | **NA – NA** | **—** | **NA** | **NA** | **NA** | **—** | **—** | **—** | **—** | **NA** |
| **main meta-analysis** | **mixed** | **Overall** | **2.67** | **1.04 – 4.30** | **2.71 ± 0.86 (0.64–4.91)** | **75** | **Mixed** | **5264** | **99.4%** | **11532** | **<0.005** | **0.47** | **Random** |
| main meta-analysis | white | 18+ | 2.61 | 2.07 – 3.15 | 2.72 ± 0.40 (2.36–3.09) | 6 | Observational Cohort | 165 | 55.4% | 11.2 | <0.05 | <0.001 | Random |
| main meta-analysis | white | 20-30 | — | NA – NA | — | NA | NA | NA | — | — | — | — | NA |
| main meta-analysis | white | 30-40 | — | NA – NA | — | NA | NA | NA | — | — | — | — | NA |
| main meta-analysis | white | 40-50 | 3.62 | 1.63 – 5.62 | 3.54 ± 1.08 (2.78–4.31) | 2 | Observational Cohort | 530 | 88.9% | 9 | <0.005 | — | Random |
| main meta-analysis | white | 50-60 | 3.92 | 3.23 – 4.61 | 3.89 ± 0.38 (3.62–4.15) | 2 | Observational Cohort | 2488 | 87.5% | 8 | <0.005 | — | Random |
| main meta-analysis | white | 60+ | 3.22 | 1.62 – 4.83 | 3.23 ± 0.82 (1.87–3.85) | 5 | Observational Cohort | 656 | 99.3% | 558.5 | <0.005 | 0.06 | Random |
| **main meta-analysis** | **white** | **Meta-Regression** | **—** | **NA – NA** | **—** | **NA** | **NA** | **NA** | **—** | **—** | **—** | **—** | **NA** |
| **main meta-analysis** | **white** | **Overall** | **3.17** | **1.73 – 4.61** | **3.16 ± 0.72 (1.87–4.31)** | **15** | **Observational Cohort** | **3839** | **99.4%** | **2409.8** | **<0.005** | **0.67** | **Random** |
| male | All | 18+ | 2.71 | 1.63 – 3.79 | 2.79 ± 0.66 (2.41–3.56) | 3 | Observational Cohort | 145 | 81.1% | 10.6 | <0.01 | 0.46 | Random |
| male | All | 20-30 | 1.65 | -0.28 – 3.57 | 1.65 ± 0.99 (0.64–2.61) | 3 | Mixed | 71 | 99.5% | 375.1 | <0.005 | 0.92 | Random |
| male | All | 30-40 | 2.53 | 0.26 – 4.79 | 2.53 ± 1.13 (1.39–3.65) | 3 | Observational Cohort | 143 | 99.9% | 2639.3 | <0.005 | 0.67 | Random |
| male | All | 40-50 | 2.12 | 0.76 – 3.47 | 2.23 ± 0.78 (1.68–2.78) | 2 | Observational Cohort | 31 | 79% | 4.8 | <0.05 | — | Random |
| male | All | 50-60 | 2.66 | 0.02 – 5.30 | 2.67 ± 1.35 (1.71–3.62) | 2 | Observational Cohort | 104 | 99% | 102.9 | <0.005 | — | Random |
| male | All | 60+ | 3.15 | 0.97 – 5.33 | 3.16 ± 1.11 (1.87–3.85) | 3 | Observational Cohort | 329 | 99.5% | 373.2 | <0.005 | 0.30 | Random |
| **male** | **All** | **Meta-Regression** | **2.41** | **-0.03 – 4.86** | **—** | **4** | **Mixed** | **262** | **100%** | **4336.5** | **<0.005** | **0.88** | **Univariable** |
| **male** | **All** | **Overall** | **2.49** | **0.59 – 4.40** | **2.51 ± 0.97 (0.64–3.85)** | **16** | **Mixed** | **823** | **99.8%** | **7367.9** | **<0.005** | **0.48** | **Random** |
| male | black | 18+ | — | NA – NA | — | NA | NA | NA | — | — | — | — | NA |
| male | black | 20-30 | — | NA – NA | — | NA | NA | NA | — | — | — | — | NA |
| male | black | 30-40 | — | NA – NA | — | NA | NA | NA | — | — | — | — | NA |
| male | black | 40-50 | — | NA – NA | — | NA | NA | NA | — | — | — | — | NA |
| male | black | 50-60 | — | NA – NA | — | NA | NA | NA | — | — | — | — | NA |
| male | black | 60+ | — | NA – NA | — | NA | NA | NA | — | — | — | — | NA |
| **male** | **black** | **Meta-Regression** | **—** | **NA – NA** | **—** | **NA** | **NA** | **NA** | **—** | **—** | **—** | **—** | **NA** |
| **male** | **black** | **Overall** | **—** | **NA – NA** | **—** | **NA** | **NA** | **NA** | **—** | **—** | **—** | **—** | **NA** |
| male | mixed | 18+ | 2.71 | 1.63 – 3.79 | 2.79 ± 0.66 (2.41–3.56) | 3 | Observational Cohort | 145 | 81.1% | 10.6 | <0.01 | 0.46 | Random |
| male | mixed | 20-30 | 1.65 | -0.28 – 3.57 | 1.65 ± 0.99 (0.64–2.61) | 3 | Mixed | 71 | 99.5% | 375.1 | <0.005 | 0.92 | Random |
| male | mixed | 30-40 | 2.53 | 0.26 – 4.79 | 2.53 ± 1.13 (1.39–3.65) | 3 | Observational Cohort | 143 | 99.9% | 2639.3 | <0.005 | 0.67 | Random |
| male | mixed | 40-50 | 1.68 | 1.29 – 2.07 | — (1.68–1.68) | 1 | Observational Cohort | 25 | — | — | — | — | Single study |
| male | mixed | 50-60 | 1.71 | 1.34 – 2.08 | — (1.71–1.71) | 1 | Observational Cohort | 25 | — | — | — | — | Single study |
| male | mixed | 60+ | — | NA – NA | — | NA | NA | NA | — | — | — | — | NA |
| **male** | **mixed** | **Meta-Regression** | **—** | **NA – NA** | **—** | **NA** | **NA** | **NA** | **—** | **—** | **—** | **—** | **NA** |
| **male** | **mixed** | **Overall** | **2.19** | **0.45 – 3.93** | **2.21 ± 0.90 (0.64–3.65)** | **11** | **Mixed** | **409** | **99.9%** | **6884** | **<0.005** | **0.31** | **Random** |
| male | white | 18+ | — | NA – NA | — | NA | NA | NA | — | — | — | — | NA |
| male | white | 20-30 | — | NA – NA | — | NA | NA | NA | — | — | — | — | NA |
| male | white | 30-40 | — | NA – NA | — | NA | NA | NA | — | — | — | — | NA |
| male | white | 40-50 | 2.78 | 0.37 – 5.19 | — (2.78–2.78) | 1 | Observational Cohort | 6 | — | — | — | — | Single study |
| male | white | 50-60 | 3.62 | 0.40 – 6.84 | — (3.62–3.62) | 1 | Observational Cohort | 79 | — | — | — | — | Single study |
| male | white | 60+ | 3.15 | 0.97 – 5.33 | 3.16 ± 1.11 (1.87–3.85) | 3 | Observational Cohort | 329 | 99.5% | 373.2 | <0.005 | 0.30 | Random |
| **male** | **white** | **Meta-Regression** | **—** | **NA – NA** | **—** | **NA** | **NA** | **NA** | **—** | **—** | **—** | **—** | **NA** |
| **male** | **white** | **Overall** | **3.18** | **1.51 – 4.86** | **3.17 ± 0.84 (1.87–3.85)** | **5** | **Observational Cohort** | **414** | **99.1%** | **432.8** | **<0.005** | **0.16** | **Random** |
| female | All | 18+ | 3.29 | 1.98 – 4.60 | 3.32 ± 0.70 (2.51–4.22) | 7 | Mixed | 488 | 92.3% | 78.2 | <0.005 | <0.05 | Random |
| female | All | 20-30 | 2.06 | 1.67 – 2.45 | — (2.06–2.06) | 1 | Observational Cohort | 25 | — | — | — | — | Single study |
| female | All | 30-40 | 2.14 | 1.67 – 2.61 | — (2.14–2.14) | 1 | Observational Cohort | 25 | — | — | — | — | Single study |
| female | All | 40-50 | 3.58 | 0.77 – 6.39 | 3.60 ± 1.44 (1.94–4.54) | 3 | Observational Cohort | 578 | 99.8% | 811.9 | <0.005 | 0.70 | Random |
| female | All | 50-60 | 3.44 | 1.19 – 5.69 | 3.44 ± 1.15 (2.12–4.15) | 3 | Observational Cohort | 2565 | 99.9% | 1375.1 | <0.005 | 0.83 | Random |
| female | All | 60+ | 2.92 | 1.08 – 4.76 | 2.93 ± 0.93 (1.33–3.62) | 5 | Observational Cohort | 496 | 99.4% | 656 | <0.005 | 0.06 | Random |
| **female** | **All** | **Meta-Regression** | **2.51** | **1.68 – 3.34** | **—** | **7** | **Mixed** | **488** | **86.8%** | **20.1** | **<0.005** | **<0.05** | **Multivariable** |
| **female** | **All** | **Overall** | **3.14** | **1.33 – 4.95** | **3.16 ± 0.94 (1.33–4.54)** | **20** | **Mixed** | **4177** | **99.6%** | **5397.1** | **<0.005** | **0.51** | **Random** |
| female | black | 18+ | — | NA – NA | — | NA | NA | NA | — | — | — | — | NA |
| female | black | 20-30 | — | NA – NA | — | NA | NA | NA | — | — | — | — | NA |
| female | black | 30-40 | — | NA – NA | — | NA | NA | NA | — | — | — | — | NA |
| female | black | 40-50 | 4.54 | 0.76 – 8.32 | — (4.54–4.54) | 1 | Observational Cohort | 29 | — | — | — | — | Single study |
| female | black | 50-60 | 4.05 | 0.61 – 7.50 | — (4.05–4.05) | 1 | Observational Cohort | 131 | — | — | — | — | Single study |
| female | black | 60+ | 3.42 | 0.05 – 6.78 | 3.32 ± 0.33 (3.08–3.55) | 2 | Observational Cohort | 144 | 53.1% | 2.1 | 0.14 | — | Fixed |
| **female** | **black** | **Meta-Regression** | **—** | **NA – NA** | **—** | **NA** | **NA** | **NA** | **—** | **—** | **—** | **—** | **NA** |
| **female** | **black** | **Overall** | **3.78** | **2.75 – 4.81** | **3.81 ± 0.63 (3.08–4.54)** | **4** | **Observational Cohort** | **304** | **81.5%** | **16.2** | **<0.005** | **1.00** | **Random** |
| female | mixed | 18+ | 3.29 | 1.98 – 4.60 | 3.32 ± 0.70 (2.51–4.22) | 7 | Mixed | 488 | 92.3% | 78.2 | <0.005 | <0.05 | Random |
| female | mixed | 20-30 | 2.06 | 1.67 – 2.45 | — (2.06–2.06) | 1 | Observational Cohort | 25 | — | — | — | — | Single study |
| female | mixed | 30-40 | 2.14 | 1.67 – 2.61 | — (2.14–2.14) | 1 | Observational Cohort | 25 | — | — | — | — | Single study |
| female | mixed | 40-50 | 1.94 | 1.47 – 2.41 | — (1.94–1.94) | 1 | Observational Cohort | 25 | — | — | — | — | Single study |
| female | mixed | 50-60 | 2.12 | 1.67 – 2.57 | — (2.12–2.12) | 1 | Observational Cohort | 25 | — | — | — | — | Single study |
| female | mixed | 60+ | 1.33 | 0.98 – 1.68 | — (1.33–1.33) | 1 | Observational Cohort | 25 | — | — | — | — | Single study |
| **female** | **mixed** | **Meta-Regression** | **—** | **NA – NA** | **—** | **NA** | **NA** | **NA** | **—** | **—** | **—** | **—** | **NA** |
| **female** | **mixed** | **Overall** | **2.71** | **0.98 – 4.43** | **2.73 ± 0.91 (1.33–4.22)** | **12** | **Mixed** | **613** | **98.4%** | **681.2** | **<0.005** | **<0.01** | **Random** |
| female | white | 18+ | — | NA – NA | — | NA | NA | NA | — | — | — | — | NA |
| female | white | 20-30 | — | NA – NA | — | NA | NA | NA | — | — | — | — | NA |
| female | white | 30-40 | — | NA – NA | — | NA | NA | NA | — | — | — | — | NA |
| female | white | 40-50 | 4.31 | 1.19 – 7.42 | — (4.31–4.31) | 1 | Observational Cohort | 524 | — | — | — | — | Single study |
| female | white | 50-60 | 4.15 | 1.24 – 7.07 | — (4.15–4.15) | 1 | Observational Cohort | 2409 | — | — | — | — | Single study |
| female | white | 60+ | 3.36 | 2.63 – 4.08 | 3.34 ± 0.39 (3.07–3.62) | 2 | Observational Cohort | 327 | 88.4% | 8.6 | <0.005 | — | Random |
| **female** | **white** | **Meta-Regression** | **—** | **NA – NA** | **—** | **NA** | **NA** | **NA** | **—** | **—** | **—** | **—** | **NA** |
| **female** | **white** | **Overall** | **3.80** | **2.73 – 4.87** | **3.79 ± 0.56 (3.07–4.31)** | **4** | **Observational Cohort** | **3260** | **96%** | **75** | **<0.005** | **0.25** | **Random** |
| *I² represents heterogeneity; Q test assesses between-study variation; Egger's p = linear regression test for funnel plot asymmetry; Meta-Regression shows covariate-adjusted estimates* | | | | | | | | | | | | | |

###### 6.7.2.3 Table of Results subdivided by race / Sex / Age

citrate_ma_3rd_analysis$race_summary_table

Table 1: **race-Stratified Meta-Analysis Results**

24-Hour Urine Citrate (mmol/24h)

| Sex Group | race | Age Band | 95% Range | Studies | Study Type | Participants | I² | Cochran's Q | Q (p-value) | Egger's p | Model |
| --- | --- | --- | --- | --- | --- | --- | --- | --- | --- | --- | --- |
| **main meta-analysis** | **All races** | **18+** | **1.40 – 4.32** | **67** | **Mixed** | **4913** | **97.8%** | **3042.9** | **<0.005** | **<0.01** | **Random** |
| **main meta-analysis** | **All races** | **20-30** | **0.13 – 3.37** | **4** | **Mixed** | **96** | **99.4%** | **472.9** | **<0.005** | **0.91** | **Random** |
| **main meta-analysis** | **All races** | **30-40** | **1.04 – 4.07** | **6** | **Observational Cohort** | **290** | **99.8%** | **3134.3** | **<0.005** | **0.36** | **Random** |
| **main meta-analysis** | **All races** | **40-50** | **0.56 – 5.07** | **7** | **Observational Cohort** | **665** | **99.6%** | **1380.3** | **<0.005** | **0.28** | **Random** |
| **main meta-analysis** | **All races** | **50-60** | **0.90 – 5.36** | **5** | **Observational Cohort** | **2669** | **99.9%** | **3028.6** | **<0.005** | **0.89** | **Random** |
| **main meta-analysis** | **All races** | **60+** | **1.18 – 4.83** | **8** | **Observational Cohort** | **825** | **99.4%** | **1117.2** | **<0.005** | **<0.05** | **Random** |
| **main meta-analysis** | **All races** | **Meta-Regression** | **1.11 – 3.93** | **51** | **Mixed** | **4337** | **96.3%** | **726** | **<0.005** | **0.93** | **Multivariable** |
| **main meta-analysis** | **All races** | **Meta-Regression** | **1.11 – 3.93** | **51** | **Mixed** | **4337** | **96.3%** | **726** | **<0.005** | **0.93** | **Multivariable** |
| **main meta-analysis** | **All races** | **Overall** | **1.17 – 4.47** | **97** | **Mixed** | **9458** | **99.4%** | **16945** | **<0.005** | **0.15** | **Random** |
| **main meta-analysis** | **All races** | **Overall** | **1.17 – 4.47** | **97** | **Mixed** | **9458** | **99.4%** | **16945** | **<0.005** | **0.15** | **Random** |
| main meta-analysis | Black | 18+ | 0.62 – 6.10 | 3 | Observational Cohort | 51 | 0% | 0 | 1.00 | — | Fixed |
| main meta-analysis | Black | 20-30 | — | — | NA | — | — | — | — | — | — |
| main meta-analysis | Black | 30-40 | — | — | NA | — | — | — | — | — | — |
| main meta-analysis | Black | 40-50 | 0.76 – 8.32 | 1 | Observational Cohort | 29 | — | — | — | — | Single study |
| main meta-analysis | Black | 50-60 | 0.61 – 7.50 | 1 | Observational Cohort | 131 | — | — | — | — | Single study |
| main meta-analysis | Black | 60+ | 0.05 – 6.78 | 2 | Observational Cohort | 144 | 53.1% | 2.1 | 0.14 | — | Fixed |
| main meta-analysis | Black | Meta-Regression | — | — | NA | — | — | — | — | — | — |
| **main meta-analysis** | **Black** | **Overall** | **2.87 – 4.38** | **7** | **Observational Cohort** | **355** | **69.8%** | **19.8** | **<0.005** | **0.49** | **Random** |
| main meta-analysis | Mixed | 18+ | 1.31 – 4.40 | 58 | Mixed | 4697 | 98.1% | 2990.1 | <0.005 | <0.01 | Random |
| main meta-analysis | Mixed | 20-30 | 0.13 – 3.37 | 4 | Mixed | 96 | 99.4% | 472.9 | <0.005 | 0.91 | Random |
| main meta-analysis | Mixed | 30-40 | 1.04 – 4.07 | 6 | Observational Cohort | 290 | 99.8% | 3134.3 | <0.005 | 0.36 | Random |
| main meta-analysis | Mixed | 40-50 | 1.15 – 2.94 | 4 | Observational Cohort | 106 | 98.9% | 280.3 | <0.005 | 0.39 | Random |
| main meta-analysis | Mixed | 50-60 | 1.35 – 2.48 | 2 | Observational Cohort | 50 | 97.9% | 47.2 | <0.005 | — | Random |
| main meta-analysis | Mixed | 60+ | 0.98 – 1.68 | 1 | Observational Cohort | 25 | — | — | — | — | Single study |
| main meta-analysis | Mixed | Meta-Regression | — | — | NA | — | — | — | — | — | — |
| **main meta-analysis** | **Mixed** | **Overall** | **1.04 – 4.30** | **75** | **Mixed** | **5264** | **99.4%** | **11532** | **<0.005** | **0.47** | **Random** |
| main meta-analysis | White | 18+ | 2.07 – 3.15 | 6 | Observational Cohort | 165 | 55.4% | 11.2 | <0.05 | <0.001 | Random |
| main meta-analysis | White | 20-30 | — | — | NA | — | — | — | — | — | — |
| main meta-analysis | White | 30-40 | — | — | NA | — | — | — | — | — | — |
| main meta-analysis | White | 40-50 | 1.63 – 5.62 | 2 | Observational Cohort | 530 | 88.9% | 9 | <0.005 | — | Random |
| main meta-analysis | White | 50-60 | 3.23 – 4.61 | 2 | Observational Cohort | 2488 | 87.5% | 8 | <0.005 | — | Random |
| main meta-analysis | White | 60+ | 1.62 – 4.83 | 5 | Observational Cohort | 656 | 99.3% | 558.5 | <0.005 | 0.06 | Random |
| main meta-analysis | White | Meta-Regression | — | — | NA | — | — | — | — | — | — |
| **main meta-analysis** | **White** | **Overall** | **1.73 – 4.61** | **15** | **Observational Cohort** | **3839** | **99.4%** | **2409.8** | **<0.005** | **0.67** | **Random** |
| **male** | **All races** | **18+** | **1.63 – 3.79** | **3** | **Observational Cohort** | **145** | **81.1%** | **10.6** | **<0.01** | **0.46** | **Random** |
| **male** | **All races** | **20-30** | **-0.28 – 3.57** | **3** | **Mixed** | **71** | **99.5%** | **375.1** | **<0.005** | **0.92** | **Random** |
| **male** | **All races** | **30-40** | **0.26 – 4.79** | **3** | **Observational Cohort** | **143** | **99.9%** | **2639.3** | **<0.005** | **0.67** | **Random** |
| **male** | **All races** | **40-50** | **0.76 – 3.47** | **2** | **Observational Cohort** | **31** | **79%** | **4.8** | **<0.05** | **—** | **Random** |
| **male** | **All races** | **50-60** | **0.02 – 5.30** | **2** | **Observational Cohort** | **104** | **99%** | **102.9** | **<0.005** | **—** | **Random** |
| **male** | **All races** | **60+** | **0.97 – 5.33** | **3** | **Observational Cohort** | **329** | **99.5%** | **373.2** | **<0.005** | **0.30** | **Random** |
| **male** | **All races** | **Meta-Regression** | **-0.03 – 4.86** | **4** | **Mixed** | **262** | **100%** | **4336.5** | **<0.005** | **0.88** | **Univariable** |
| **male** | **All races** | **Overall** | **0.59 – 4.40** | **16** | **Mixed** | **823** | **99.8%** | **7367.9** | **<0.005** | **0.48** | **Random** |
| male | Black | 18+ | — | — | NA | — | — | — | — | — | — |
| male | Black | 20-30 | — | — | NA | — | — | — | — | — | — |
| male | Black | 30-40 | — | — | NA | — | — | — | — | — | — |
| male | Black | 40-50 | — | — | NA | — | — | — | — | — | — |
| male | Black | 50-60 | — | — | NA | — | — | — | — | — | — |
| male | Black | 60+ | — | — | NA | — | — | — | — | — | — |
| male | Black | Meta-Regression | — | — | NA | — | — | — | — | — | — |
| **male** | **Black** | **Overall** | **—** | **—** | **NA** | **—** | **—** | **—** | **—** | **—** | **—** |
| male | Mixed | 18+ | 1.63 – 3.79 | 3 | Observational Cohort | 145 | 81.1% | 10.6 | <0.01 | 0.46 | Random |
| male | Mixed | 20-30 | -0.28 – 3.57 | 3 | Mixed | 71 | 99.5% | 375.1 | <0.005 | 0.92 | Random |
| male | Mixed | 30-40 | 0.26 – 4.79 | 3 | Observational Cohort | 143 | 99.9% | 2639.3 | <0.005 | 0.67 | Random |
| male | Mixed | 40-50 | 1.29 – 2.07 | 1 | Observational Cohort | 25 | — | — | — | — | Single study |
| male | Mixed | 50-60 | 1.34 – 2.08 | 1 | Observational Cohort | 25 | — | — | — | — | Single study |
| male | Mixed | 60+ | — | — | NA | — | — | — | — | — | — |
| male | Mixed | Meta-Regression | — | — | NA | — | — | — | — | — | — |
| **male** | **Mixed** | **Overall** | **0.45 – 3.93** | **11** | **Mixed** | **409** | **99.9%** | **6884** | **<0.005** | **0.31** | **Random** |
| male | White | 18+ | — | — | NA | — | — | — | — | — | — |
| male | White | 20-30 | — | — | NA | — | — | — | — | — | — |
| male | White | 30-40 | — | — | NA | — | — | — | — | — | — |
| male | White | 40-50 | 0.37 – 5.19 | 1 | Observational Cohort | 6 | — | — | — | — | Single study |
| male | White | 50-60 | 0.40 – 6.84 | 1 | Observational Cohort | 79 | — | — | — | — | Single study |
| male | White | 60+ | 0.97 – 5.33 | 3 | Observational Cohort | 329 | 99.5% | 373.2 | <0.005 | 0.30 | Random |
| male | White | Meta-Regression | — | — | NA | — | — | — | — | — | — |
| **male** | **White** | **Overall** | **1.51 – 4.86** | **5** | **Observational Cohort** | **414** | **99.1%** | **432.8** | **<0.005** | **0.16** | **Random** |
| **female** | **All races** | **18+** | **1.98 – 4.60** | **7** | **Mixed** | **488** | **92.3%** | **78.2** | **<0.005** | **<0.05** | **Random** |
| **female** | **All races** | **20-30** | **1.67 – 2.45** | **1** | **Observational Cohort** | **25** | **—** | **—** | **—** | **—** | **Single study** |
| **female** | **All races** | **30-40** | **1.67 – 2.61** | **1** | **Observational Cohort** | **25** | **—** | **—** | **—** | **—** | **Single study** |
| **female** | **All races** | **40-50** | **0.77 – 6.39** | **3** | **Observational Cohort** | **578** | **99.8%** | **811.9** | **<0.005** | **0.70** | **Random** |
| **female** | **All races** | **50-60** | **1.19 – 5.69** | **3** | **Observational Cohort** | **2565** | **99.9%** | **1375.1** | **<0.005** | **0.83** | **Random** |
| **female** | **All races** | **60+** | **1.08 – 4.76** | **5** | **Observational Cohort** | **496** | **99.4%** | **656** | **<0.005** | **0.06** | **Random** |
| **female** | **All races** | **Meta-Regression** | **1.68 – 3.34** | **7** | **Mixed** | **488** | **86.8%** | **20.1** | **<0.005** | **<0.05** | **Multivariable** |
| **female** | **All races** | **Overall** | **1.33 – 4.95** | **20** | **Mixed** | **4177** | **99.6%** | **5397.1** | **<0.005** | **0.51** | **Random** |
| female | Black | 18+ | — | — | NA | — | — | — | — | — | — |
| female | Black | 20-30 | — | — | NA | — | — | — | — | — | — |
| female | Black | 30-40 | — | — | NA | — | — | — | — | — | — |
| female | Black | 40-50 | 0.76 – 8.32 | 1 | Observational Cohort | 29 | — | — | — | — | Single study |
| female | Black | 50-60 | 0.61 – 7.50 | 1 | Observational Cohort | 131 | — | — | — | — | Single study |
| female | Black | 60+ | 0.05 – 6.78 | 2 | Observational Cohort | 144 | 53.1% | 2.1 | 0.14 | — | Fixed |
| female | Black | Meta-Regression | — | — | NA | — | — | — | — | — | — |
| **female** | **Black** | **Overall** | **2.75 – 4.81** | **4** | **Observational Cohort** | **304** | **81.5%** | **16.2** | **<0.005** | **1.00** | **Random** |
| female | Mixed | 18+ | 1.98 – 4.60 | 7 | Mixed | 488 | 92.3% | 78.2 | <0.005 | <0.05 | Random |
| female | Mixed | 20-30 | 1.67 – 2.45 | 1 | Observational Cohort | 25 | — | — | — | — | Single study |
| female | Mixed | 30-40 | 1.67 – 2.61 | 1 | Observational Cohort | 25 | — | — | — | — | Single study |
| female | Mixed | 40-50 | 1.47 – 2.41 | 1 | Observational Cohort | 25 | — | — | — | — | Single study |
| female | Mixed | 50-60 | 1.67 – 2.57 | 1 | Observational Cohort | 25 | — | — | — | — | Single study |
| female | Mixed | 60+ | 0.98 – 1.68 | 1 | Observational Cohort | 25 | — | — | — | — | Single study |
| female | Mixed | Meta-Regression | — | — | NA | — | — | — | — | — | — |
| **female** | **Mixed** | **Overall** | **0.98 – 4.43** | **12** | **Mixed** | **613** | **98.4%** | **681.2** | **<0.005** | **<0.01** | **Random** |
| female | White | 18+ | — | — | NA | — | — | — | — | — | — |
| female | White | 20-30 | — | — | NA | — | — | — | — | — | — |
| female | White | 30-40 | — | — | NA | — | — | — | — | — | — |
| female | White | 40-50 | 1.19 – 7.42 | 1 | Observational Cohort | 524 | — | — | — | — | Single study |
| female | White | 50-60 | 1.24 – 7.07 | 1 | Observational Cohort | 2409 | — | — | — | — | Single study |
| female | White | 60+ | 2.63 – 4.08 | 2 | Observational Cohort | 327 | 88.4% | 8.6 | <0.005 | — | Random |
| female | White | Meta-Regression | — | — | NA | — | — | — | — | — | — |
| **female** | **White** | **Overall** | **2.73 – 4.87** | **4** | **Observational Cohort** | **3260** | **96%** | **75** | **<0.005** | **0.25** | **Random** |
| *I² represents heterogeneity; Q test assesses between-study variation; Egger's p = linear regression test for funnel plot asymmetry; — indicates insufficient data* | | | | | | | | | | | |

###### 6.7.2.4 Wald Tests

citrate_ma_3rd_analysis$wald_tests$sex_comparison

Table 1: **Sex Comparison (Overall)**

| Comparison | % Shift | P (Wald Test) | Mean Diff (95% CI) |
| --- | --- | --- | --- |
| Male vs Female (Overall) | -20.6% | 0.629 | -0.65 [-3.28, 1.98] |
| *Mean difference = Group 1 − Group 2 in component units (mmol/24h, L/24h, or pH); % shift = difference relative to Group 2.* | | | |

citrate_ma_3rd_analysis$wald_tests$race_comparisons

Table 1: **race Comparisons Within Sex Groups**

| Sex Group | Comparison | % Shift | P (Wald Test) | Mean Diff (95% CI) |
| --- | --- | --- | --- | --- |
| male | black vs mixed | — | — | — |
| male | black vs white | — | — | — |
| male | mixed vs white | -31.3% | 0.418 | -1.00 [-3.41, 1.42] |
| female | black vs mixed | +39.9% | 0.293 | 1.08 [-0.93, 3.09] |
| female | black vs white | -0.5% | 0.981 | -0.02 [-1.50, 1.47] |
| female | mixed vs white | -28.8% | 0.290 | -1.10 [-3.13, 0.93] |
| *Mean difference = Group 1 − Group 2 in component units (mmol/24h, L/24h, or pH); % shift = difference relative to Group 2.* | | | | |

citrate_ma_3rd_analysis$wald_tests$age_band_comparisons

Table 1: **Age Band Comparisons Within Sex Groups**

| Sex Group | Comparison | % Shift | P (Wald Test) | Mean Diff (95% CI) |
| --- | --- | --- | --- | --- |
| main meta-analysis | 18+ vs 20-30 | +63.6% | 0.318 | 1.11 [-1.07, 3.29] |
| main meta-analysis | 18+ vs 30-40 | +12.0% | 0.775 | 0.31 [-1.80, 2.41] |
| main meta-analysis | 18+ vs 40-50 | +1.8% | 0.970 | 0.05 [-2.64, 2.74] |
| main meta-analysis | 18+ vs 50-60 | -8.5% | 0.845 | -0.27 [-2.93, 2.40] |
| main meta-analysis | 18+ vs 60+ | -4.8% | 0.904 | -0.14 [-2.48, 2.20] |
| main meta-analysis | 20-30 vs 30-40 | -31.5% | 0.477 | -0.81 [-3.02, 1.41] |
| main meta-analysis | 20-30 vs 40-50 | -37.8% | 0.454 | -1.06 [-3.84, 1.72] |
| main meta-analysis | 20-30 vs 50-60 | -44.1% | 0.327 | -1.38 [-4.14, 1.38] |
| main meta-analysis | 20-30 vs 60+ | -41.8% | 0.313 | -1.26 [-3.70, 1.19] |
| main meta-analysis | 30-40 vs 40-50 | -9.1% | 0.854 | -0.26 [-2.97, 2.46] |
| main meta-analysis | 30-40 vs 50-60 | -18.3% | 0.678 | -0.57 [-3.27, 2.12] |
| main meta-analysis | 30-40 vs 60+ | -15.0% | 0.710 | -0.45 [-2.83, 1.92] |
| main meta-analysis | 40-50 vs 50-60 | -10.1% | 0.845 | -0.32 [-3.49, 2.86] |
| main meta-analysis | 40-50 vs 60+ | -6.5% | 0.895 | -0.20 [-3.10, 2.71] |
| main meta-analysis | 50-60 vs 60+ | +4.0% | 0.934 | 0.12 [-2.76, 3.01] |
| male | 18+ vs 20-30 | +64.9% | 0.343 | 1.07 [-1.14, 3.28] |
| male | 18+ vs 30-40 | +7.5% | 0.882 | 0.19 [-2.32, 2.69] |
| male | 18+ vs 40-50 | +28.3% | 0.499 | 0.60 [-1.13, 2.33] |
| male | 18+ vs 50-60 | +2.1% | 0.969 | 0.06 [-2.80, 2.91] |
| male | 18+ vs 60+ | -13.8% | 0.727 | -0.43 [-2.87, 2.00] |
| male | 20-30 vs 30-40 | -34.8% | 0.561 | -0.88 [-3.85, 2.09] |
| male | 20-30 vs 40-50 | -22.2% | 0.695 | -0.47 [-2.82, 1.88] |
| male | 20-30 vs 50-60 | -38.1% | 0.543 | -1.01 [-4.28, 2.25] |
| male | 20-30 vs 60+ | -47.7% | 0.312 | -1.50 [-4.41, 1.41] |
| male | 30-40 vs 40-50 | +19.3% | 0.761 | 0.41 [-2.23, 3.04] |
| male | 30-40 vs 50-60 | -5.0% | 0.940 | -0.13 [-3.61, 3.34] |
| male | 30-40 vs 60+ | -19.8% | 0.698 | -0.62 [-3.76, 2.52] |
| male | 40-50 vs 50-60 | -20.4% | 0.720 | -0.54 [-3.51, 2.42] |
| male | 40-50 vs 60+ | -32.8% | 0.431 | -1.03 [-3.60, 1.54] |
| male | 50-60 vs 60+ | -15.5% | 0.780 | -0.49 [-3.91, 2.94] |
| female | 18+ vs 20-30 | +59.6% | 0.078 | 1.23 [-0.14, 2.59] |
| female | 18+ vs 30-40 | +53.6% | 0.106 | 1.15 [-0.24, 2.54] |
| female | 18+ vs 40-50 | -8.1% | 0.855 | -0.29 [-3.39, 2.81] |
| female | 18+ vs 50-60 | -4.4% | 0.909 | -0.15 [-2.75, 2.45] |
| female | 18+ vs 60+ | +12.5% | 0.751 | 0.37 [-1.89, 2.62] |
| female | 20-30 vs 30-40 | -3.7% | 0.798 | -0.08 [-0.69, 0.53] |
| female | 20-30 vs 40-50 | -42.4% | 0.295 | -1.52 [-4.35, 1.32] |
| female | 20-30 vs 50-60 | -40.1% | 0.236 | -1.38 [-3.66, 0.90] |
| female | 20-30 vs 60+ | -29.5% | 0.369 | -0.86 [-2.74, 1.02] |
| female | 30-40 vs 40-50 | -40.2% | 0.323 | -1.44 [-4.29, 1.41] |
| female | 30-40 vs 50-60 | -37.8% | 0.267 | -1.30 [-3.60, 1.00] |
| female | 30-40 vs 60+ | -26.8% | 0.419 | -0.78 [-2.68, 1.12] |
| female | 40-50 vs 50-60 | +4.0% | 0.941 | 0.14 [-3.46, 3.73] |
| female | 40-50 vs 60+ | +22.4% | 0.703 | 0.65 [-2.70, 4.01] |
| female | 50-60 vs 60+ | +17.7% | 0.727 | 0.52 [-2.39, 3.42] |
| *Mean difference = Group 1 − Group 2 in component units (mmol/24h, L/24h, or pH); % shift = difference relative to Group 2.* | | | | |

citrate_ma_3rd_analysis$wald_tests$sex_within_age_bands

Table 1: **Sex Comparisons Within Age Bands**

| Age Band | % Shift | P (Wald Test) | Mean Diff (95% CI) |
| --- | --- | --- | --- |
| 18+ | -17.4% | 0.508 | -0.57 [-2.27, 1.12] |
| 20-30 | -20.1% | 0.679 | -0.41 [-2.38, 1.55] |
| 30-40 | +18.0% | 0.744 | 0.39 [-1.92, 2.69] |
| 40-50 | -40.8% | 0.359 | -1.46 [-4.58, 1.66] |
| 50-60 | -22.7% | 0.659 | -0.78 [-4.25, 2.68] |
| 60+ | +7.7% | 0.877 | 0.23 [-2.63, 3.08] |
| *Mean difference = Group 1 − Group 2 in component units (mmol/24h, L/24h, or pH); % shift = difference relative to Group 2.* | | | |

#### 6.8 Phosphate

##### 6.8.1 Comparison by Age Band

###### 6.8.1.1 Plot

```
phosphate_ma_2nd_analysis <- sex_age_subdivision_ma("phosphate",
                                                    )
## Not enough studies for multivariable meta-regression (k = 4, need > 6). Running univariable models instead.
## Not enough studies for multivariable meta-regression (k = 6, need > 6). Running univariable models instead.
phosphate_ma_2nd_analysis$combined_plot
## `height` was translated to `width`.
## `height` was translated to `width`.
## `height` was translated to `width`.
```

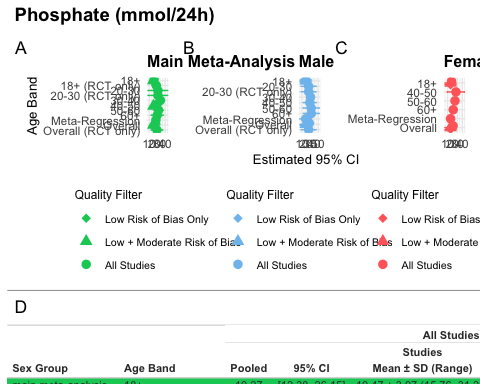

###### 6.8.1.2 Summary Table

phosphate_ma_2nd_analysis$combined_summary_table

Table 1: **Combined Meta-Analysis Results**

24-Hour Urine Phosphate (mmol/24h)

|  |  | **All Studies** | | | | | | **Low Risk of Bias Only** | | | | | | **Low + Moderate RoB** | | | | | |
| --- | --- | --- | --- | --- | --- | --- | --- | --- | --- | --- | --- | --- | --- | --- | --- | --- | --- | --- | --- |
| Sex Group | Age Band | Pooled | 95% CI | Studies  Mean ± SD (Range) | Studies, n | Q p-value | Egger's p | Pooled | 95% CI | Studies  Mean ± SD (Range) | Studies, n | Q p-value | Egger's p | Pooled | 95% CI | Studies  Mean ± SD (Range) | Studies, n | Q p-value | Egger's p |
| main meta-analysis | 18+ | 19.27 | [12.38, 26.15] | 19.47 ± 3.97 (15.76–31.22) | 40 | <0.001 | 0.10 | 17.36 | [15.32, 19.41] | 17.35 ± 1.15 (15.76–19.22) | 25 | <0.001 | 0.67 | 17.81 | [14.87, 20.74] | 18.12 ± 2.60 (15.76–28.80) | 29 | <0.001 | 1.00 |
| **main meta-analysis** | **18+ (RCT only)** | **23.49** | **[15.40, 31.58]** | **23.84 ± 4.43 (18.54–28.80)** | **4** | **<0.001** | **0.11** | **—** | **—** | **—** | **—** | **—** | **—** | **—** | **—** | **—** | **—** | **—** | **—** |
| main meta-analysis | 20-30 | 26.42 | [7.87, 44.96] | 26.37 ± 9.52 (19.64–33.10) | 2 | <0.001 | — | — | — | — | — | — | — | — | — | — | — | — | — |
| **main meta-analysis** | **20-30 (RCT only)** | **26.42** | **[7.87, 44.96]** | **26.37 ± 9.52 (19.64–33.10)** | **2** | **<0.001** | **—** | **—** | **—** | **—** | **—** | **—** | **—** | **—** | **—** | **—** | **—** | **—** | **—** |
| main meta-analysis | 30-40 | 31.20 | [29.24, 33.16] | — (31.20–31.20) | 1 | — | — | — | — | — | — | — | — | — | — | — | — | — | — |
| main meta-analysis | 40-50 | 24.43 | [15.30, 33.56] | 24.86 ± 4.83 (18.08–29.19) | 4 | <0.001 | 0.46 | — | — | — | — | — | — | 18.08 | [15.92, 20.24] | — (18.08–18.08) | 1 | — | — |
| main meta-analysis | 50-60 | 27.61 | [18.38, 36.84] | 27.68 ± 4.85 (23.32–32.91) | 3 | <0.001 | 0.93 | — | — | — | — | — | — | — | — | — | — | — | — |
| main meta-analysis | 60+ | 25.18 | [13.49, 36.88] | 25.19 ± 6.03 (19.13–33.68) | 6 | <0.001 | 0.92 | — | — | — | — | — | — | — | — | — | — | — | — |
| **main meta-analysis** | **Meta-Regression** | **22.63** | **[18.87, 26.40]** | **—** | **40** | **<0.001** | **<0.001** | **—** | **—** | **—** | **—** | **—** | **—** | **—** | **—** | **—** | **—** | **—** | **—** |
| **main meta-analysis** | **Overall** | **21.29** | **[11.16, 31.42]** | **21.37 ± 5.31 (15.76–33.68)** | **56** | **<0.001** | **<0.005** | **17.36** | **[15.32, 19.41]** | **17.35 ± 1.15 (15.76–19.22)** | **25** | **<0.001** | **0.67** | **17.81** | **[14.97, 20.66]** | **18.11 ± 2.56 (15.76–28.80)** | **30** | **<0.001** | **0.89** |
| **main meta-analysis** | **Overall (RCT only)** | **24.67** | **[13.59, 35.75]** | **24.68 ± 5.62 (18.54–33.10)** | **6** | **<0.001** | **0.31** | **—** | **—** | **—** | **—** | **—** | **—** | **—** | **—** | **—** | **—** | **—** | **—** |
| male | 18+ | 19.54 | [5.06, 34.02] | 20.49 ± 2.03 (19.05–21.92) | 2 | 0.10 | — | 19.05 | [4.49, 33.60] | — (19.05–19.05) | 1 | — | — | 19.05 | [4.49, 33.60] | — (19.05–19.05) | 1 | — | — |
| **male** | **18+ (RCT only)** | **—** | **—** | **—** | **—** | **—** | **—** | **—** | **—** | **—** | **—** | **—** | **—** | **—** | **—** | **—** | **—** | **—** | **—** |
| male | 20-30 | 26.42 | [7.87, 44.96] | 26.37 ± 9.52 (19.64–33.10) | 2 | <0.001 | — | — | — | — | — | — | — | — | — | — | — | — | — |
| **male** | **20-30 (RCT only)** | **26.42** | **[7.87, 44.96]** | **26.37 ± 9.52 (19.64–33.10)** | **2** | **<0.001** | **—** | **—** | **—** | **—** | **—** | **—** | **—** | **—** | **—** | **—** | **—** | **—** | **—** |
| male | 30-40 | 31.20 | [29.24, 33.16] | — (31.20–31.20) | 1 | — | — | — | — | — | — | — | — | — | — | — | — | — | — |
| male | 40-50 | 29.19 | [12.85, 45.54] | — (29.19–29.19) | 1 | — | — | — | — | — | — | — | — | — | — | — | — | — | — |
| male | 50-60 | 32.91 | [12.54, 53.27] | — (32.91–32.91) | 1 | — | — | — | — | — | — | — | — | — | — | — | — | — | — |
| male | 60+ | 33.08 | [13.47, 52.68] | 32.59 ± 1.54 (31.50–33.68) | 2 | 0.09 | — | — | — | — | — | — | — | — | — | — | — | — | — |
| **male** | **Meta-Regression** | **19.05** | **[4.56, 33.54]** | **—** | **4** | **<0.001** | **0.32** | **—** | **—** | **—** | **—** | **—** | **—** | **—** | **—** | **—** | **—** | **—** | **—** |
| **male** | **Overall** | **28.04** | **[16.23, 39.86]** | **28.02 ± 6.05 (19.05–33.68)** | **9** | **<0.001** | **0.29** | **19.05** | **[4.49, 33.60]** | **— (19.05–19.05)** | **1** | **—** | **—** | **19.05** | **[4.49, 33.60]** | **— (19.05–19.05)** | **1** | **—** | **—** |
| **male** | **Overall (RCT only)** | **26.42** | **[7.87, 44.96]** | **26.37 ± 9.52 (19.64–33.10)** | **2** | **<0.001** | **—** | **—** | **—** | **—** | **—** | **—** | **—** | **—** | **—** | **—** | **—** | **—** | **—** |
| female | 18+ | 18.02 | [13.18, 22.87] | 18.10 ± 2.64 (15.76–21.47) | 6 | <0.001 | 0.08 | 16.06 | [4.12, 28.00] | 16.05 ± 0.41 (15.76–16.34) | 2 | 0.39 | — | 16.06 | [4.12, 28.00] | 16.05 ± 0.41 (15.76–16.34) | 2 | 0.39 | — |
| **female** | **18+ (RCT only)** | **—** | **—** | **—** | **—** | **—** | **—** | **—** | **—** | **—** | **—** | **—** | **—** | **—** | **—** | **—** | **—** | **—** | **—** |
| female | 20-30 | — | — | — | — | — | — | — | — | — | — | — | — | — | — | — | — | — | — |
| **female** | **20-30 (RCT only)** | **—** | **—** | **—** | **—** | **—** | **—** | **—** | **—** | **—** | **—** | **—** | **—** | **—** | **—** | **—** | **—** | **—** | **—** |
| female | 30-40 | — | — | — | — | — | — | — | — | — | — | — | — | — | — | — | — | — | — |
| female | 40-50 | 27.01 | [9.67, 44.35] | 26.08 ± 1.53 (25.00–27.17) | 2 | 0.13 | — | — | — | — | — | — | — | — | — | — | — | — | — |
| female | 50-60 | 25.13 | [20.41, 29.85] | 25.06 ± 2.46 (23.32–26.81) | 2 | <0.001 | — | — | — | — | — | — | — | — | — | — | — | — | — |
| female | 60+ | 21.56 | [17.35, 25.76] | 21.49 ± 2.24 (19.13–24.31) | 4 | <0.001 | <0.05 | — | — | — | — | — | — | — | — | — | — | — | — |
| **female** | **Meta-Regression** | **16.05** | **[11.20, 20.90]** | **—** | **6** | **<0.001** | **0.08** | **—** | **—** | **—** | **—** | **—** | **—** | **—** | **—** | **—** | **—** | **—** | **—** |
| **female** | **Overall** | **21.21** | **[13.70, 28.71]** | **21.20 ± 3.86 (15.76–27.17)** | **14** | **<0.001** | **<0.05** | **16.06** | **[4.12, 28.00]** | **16.05 ± 0.41 (15.76–16.34)** | **2** | **0.39** | **—** | **16.06** | **[4.12, 28.00]** | **16.05 ± 0.41 (15.76–16.34)** | **2** | **0.39** | **—** |
| **female** | **Overall (RCT only)** | **—** | **—** | **—** | **—** | **—** | **—** | **—** | **—** | **—** | **—** | **—** | **—** | **—** | **—** | **—** | **—** | **—** | **—** |
| *Studies, n = number of studies; Q p-value = Cochran's Q heterogeneity test; Egger's p = linear regression test for funnel plot asymmetry; RoB = Risk of Bias* | | | | | | | | | | | | | | | | | | | |

###### 6.8.1.3 Wald Tests

phosphate_ma_2nd_analysis$wald_tests$sex_comparison

Table 1: **Sex Comparison (Overall)**

| Comparison | % Shift | P (Wald Test) | Mean Diff (95% CI) |
| --- | --- | --- | --- |
| Male vs Female (Overall) | +32.2% | 0.338 | 6.84 [-7.16, 20.83] |
| *Mean difference = Group 1 − Group 2 in component units (mmol/24h, L/24h, or pH); % shift = difference relative to Group 2.* | | | |

phosphate_ma_2nd_analysis$wald_tests$age_band_comparisons

Table 1: **Age Band Comparisons Within Sex Groups**

| Sex Group | Comparison | % Shift | P (Wald Test) | Mean Diff (95% CI) |
| --- | --- | --- | --- | --- |
| main meta-analysis | 18+ vs 20-30 | -27.1% | 0.479 | -7.15 [-26.93, 12.63] |
| main meta-analysis | 18+ vs 30-40 | -38.2% | <0.01 | -11.93 [-19.09, -4.77] |
| main meta-analysis | 18+ vs 40-50 | -21.1% | 0.376 | -5.16 [-16.60, 6.27] |
| main meta-analysis | 18+ vs 50-60 | -30.2% | 0.156 | -8.34 [-19.85, 3.17] |
| main meta-analysis | 18+ vs 60+ | -23.5% | 0.393 | -5.91 [-19.49, 7.66] |
| main meta-analysis | 20-30 vs 30-40 | -15.3% | 0.615 | -4.78 [-23.43, 13.87] |
| main meta-analysis | 20-30 vs 40-50 | +8.1% | 0.851 | 1.99 [-18.68, 22.66] |
| main meta-analysis | 20-30 vs 50-60 | -4.3% | 0.910 | -1.19 [-21.90, 19.52] |
| main meta-analysis | 20-30 vs 60+ | +4.9% | 0.912 | 1.24 [-20.69, 23.16] |
| main meta-analysis | 30-40 vs 40-50 | +27.7% | 0.155 | 6.77 [-2.57, 16.11] |
| main meta-analysis | 30-40 vs 50-60 | +13.0% | 0.456 | 3.59 [-5.84, 13.03] |
| main meta-analysis | 30-40 vs 60+ | +23.9% | 0.320 | 6.02 [-5.84, 17.88] |
| main meta-analysis | 40-50 vs 50-60 | -11.5% | 0.631 | -3.18 [-16.16, 9.81] |
| main meta-analysis | 40-50 vs 60+ | -3.0% | 0.921 | -0.75 [-15.59, 14.09] |
| main meta-analysis | 50-60 vs 60+ | +9.6% | 0.750 | 2.43 [-12.47, 17.32] |
| male | 18+ vs 20-30 | -26.0% | 0.566 | -6.88 [-30.41, 16.65] |
| male | 18+ vs 30-40 | -37.4% | 0.118 | -11.66 [-26.27, 2.95] |
| male | 18+ vs 40-50 | -33.1% | 0.386 | -9.65 [-31.49, 12.18] |
| male | 18+ vs 50-60 | -40.6% | 0.294 | -13.37 [-38.36, 11.62] |
| male | 18+ vs 60+ | -40.9% | 0.276 | -13.54 [-37.91, 10.83] |
| male | 20-30 vs 30-40 | -15.3% | 0.615 | -4.78 [-23.43, 13.87] |
| male | 20-30 vs 40-50 | -9.5% | 0.826 | -2.77 [-27.49, 21.95] |
| male | 20-30 vs 50-60 | -19.7% | 0.644 | -6.49 [-34.03, 21.05] |
| male | 20-30 vs 60+ | -20.1% | 0.629 | -6.66 [-33.64, 20.33] |
| male | 30-40 vs 40-50 | +6.9% | 0.811 | 2.01 [-14.45, 18.47] |
| male | 30-40 vs 50-60 | -5.2% | 0.870 | -1.71 [-22.16, 18.75] |
| male | 30-40 vs 60+ | -5.7% | 0.852 | -1.88 [-21.58, 17.82] |
| male | 40-50 vs 50-60 | -11.3% | 0.780 | -3.72 [-29.83, 22.40] |
| male | 40-50 vs 60+ | -11.7% | 0.766 | -3.88 [-29.41, 21.64] |
| male | 50-60 vs 60+ | -0.5% | 0.991 | -0.17 [-28.43, 28.10] |
| female | 18+ vs 40-50 | -33.3% | 0.328 | -8.98 [-26.99, 9.02] |
| female | 18+ vs 50-60 | -28.3% | <0.05 | -7.11 [-13.87, -0.34] |
| female | 18+ vs 60+ | -16.4% | 0.281 | -3.53 [-9.95, 2.89] |
| female | 40-50 vs 50-60 | +7.5% | 0.838 | 1.88 [-16.09, 19.85] |
| female | 40-50 vs 60+ | +25.3% | 0.549 | 5.45 [-12.39, 23.30] |
| female | 50-60 vs 60+ | +16.6% | 0.267 | 3.58 [-2.74, 9.90] |
| *Mean difference = Group 1 − Group 2 in component units (mmol/24h, L/24h, or pH); % shift = difference relative to Group 2.* | | | | |

phosphate_ma_2nd_analysis$wald_tests$sex_within_age_bands

Table 1: **Sex Comparisons Within Age Bands**

| Age Band | % Shift | P (Wald Test) | Mean Diff (95% CI) |
| --- | --- | --- | --- |
| 18+ | +8.4% | 0.846 | 1.51 [-13.76, 16.78] |
| 40-50 | +8.1% | 0.858 | 2.18 [-21.65, 26.01] |
| 50-60 | +30.9% | 0.466 | 7.77 [-13.13, 28.68] |
| 60+ | +53.4% | 0.260 | 11.52 [-8.53, 31.57] |
| *Mean difference = Group 1 − Group 2 in component units (mmol/24h, L/24h, or pH); % shift = difference relative to Group 2.* | | | |

###### 6.8.1.4 Meta-Regression (plain language)

format_meta_reg_plain("phosphate", sex_group = "all")

Message

No meta-regression result for phosphate / all

###### 6.8.1.5 Assay Subgroup Analysis

assay_subgroup_table("phosphate")

Table 1: **Assay subgroup analysis — Phosphate**

*Per-assay pooled estimates; categories with k < 3 listed but not pooled.*

| Assay category | Studies, n | Participants | 95% CI | I² | Q p-value | Egger's p | Note |
| --- | --- | --- | --- | --- | --- | --- | --- |
| Unreported | 33 | 5,413 | 22.40 [20.58, 24.22] | 99% | <0.001 | 0.796 |  |
| Autoanalyser | 20 | 2,587 | 18.87 [17.01, 20.73] | 100% | <0.001 | <0.01 |  |
| Ammonium molybdate | 3 | 104 | 25.75 [18.38, 33.12] | 98% | <0.001 | 0.217 |  |

###### 6.8.1.6 Preservative Sensitivity (Litholink / acidified samples)

```
preservative_sensitivity_ma("phosphate")
## # A tibble: 3 × 6
##   group               n_studies n_participants pooled_estimate lower_95 upper_95
##   <chr>                   <int>          <int>           <dbl>    <dbl>    <dbl>
## 1 All studies                56           8104            21.3     19.9     22.7
## 2 Excluding preserva…        54           8044            21.1     19.7     22.5
## 3 Preservative only           2             60            27.0     18.2     35.8
```

###### 6.8.1.7 Creatinine Completeness Sensitivity

creat_completeness_table("phosphate")

Table 1: **Creatinine-completeness sensitivity — Phosphate**

*Compares the all-studies pooled estimate against subsets defined by whether the study reported 24h urine creatinine. Studies that do not report creatinine cannot verify collection completeness; a substantial shift here flags possible bias from incomplete collections.*

| Subset | Studies, n | Participants | Range (95% CI) | Δ vs all-studies (%) | Flag |
| --- | --- | --- | --- | --- | --- |
| All studies | 56 | 8,104 | 21.29 [19.90, 22.68] | — | — |
| Creatinine reported only | 49 | 7,524 | 20.62 [19.21, 22.03] | -3.1% | OK |
| Creatinine NOT reported | 7 | 580 | 25.90 [22.12, 29.68] | +21.7% | ⚠ shift > 20% |

##### 6.8.2 Comparison by Age / Sex / race

###### 6.8.2.1 Plot

```
phosphate_ma_3rd_analysis <- sex_age_race_subdivision_ma("phosphate",
                                                      urine_ma2,
                                                      overall_bias)
## Not enough studies for multivariable meta-regression (k = 4, need > 6). Running univariable models instead.
## Not enough studies for multivariable meta-regression (k = 6, need > 6). Running univariable models instead.
phosphate_ma_3rd_analysis$race_plot
## `height` was translated to `width`.
## `height` was translated to `width`.
## `height` was translated to `width`.
```

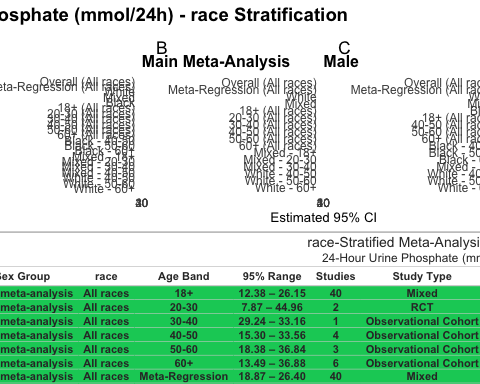

###### 6.8.2.2 Combined MA Results

phosphate_ma_3rd_analysis$combined_summary_table

Table 1: **Combined Meta-Analysis Results**

24-Hour Urine Phosphate (mmol/24h)

| Sex Group | race | Age Band | Pooled | 95% Range | Studies Mean SD Range | Studies | Study Type | Participants | I² | Cochran's Q | Q (p-value) | Egger's p | Model |
| --- | --- | --- | --- | --- | --- | --- | --- | --- | --- | --- | --- | --- | --- |
| main meta-analysis | All | 18+ | 19.27 | 12.38 – 26.15 | 19.47 ± 3.97 (15.76–31.22) | 40 | Mixed | 3964 | 94.1% | 665.5 | <0.005 | 0.10 | Random |
| main meta-analysis | All | 20-30 | 26.42 | 7.87 – 44.96 | 26.37 ± 9.52 (19.64–33.10) | 2 | RCT | 46 | 98.8% | 84.6 | <0.005 | — | Random |
| main meta-analysis | All | 30-40 | 31.20 | 29.24 – 33.16 | — (31.20–31.20) | 1 | Observational Cohort | 107 | — | — | — | — | Single study |
| main meta-analysis | All | 40-50 | 24.43 | 15.30 – 33.56 | 24.86 ± 4.83 (18.08–29.19) | 4 | Observational Cohort | 593 | 99.3% | 459.8 | <0.005 | 0.46 | Random |
| main meta-analysis | All | 50-60 | 27.61 | 18.38 – 36.84 | 27.68 ± 4.85 (23.32–32.91) | 3 | Observational Cohort | 2619 | 96.1% | 50.9 | <0.005 | 0.93 | Random |
| main meta-analysis | All | 60+ | 25.18 | 13.49 – 36.88 | 25.19 ± 6.03 (19.13–33.68) | 6 | Observational Cohort | 775 | 98.3% | 291.7 | <0.005 | 0.92 | Random |
| **main meta-analysis** | **All** | **Meta-Regression** | **22.63** | **18.87 – 26.40** | **—** | **40** | **Mixed** | **3974** | **89.7%** | **183.3** | **<0.005** | **<0.001** | **Multivariable** |
| **main meta-analysis** | **All** | **Meta-Regression** | **22.63** | **18.87 – 26.40** | **—** | **40** | **Mixed** | **3974** | **89.7%** | **183.3** | **<0.005** | **<0.001** | **Multivariable** |
| **main meta-analysis** | **All** | **Overall** | **21.29** | **11.16 – 31.42** | **21.37 ± 5.31 (15.76–33.68)** | **56** | **Mixed** | **8104** | **99.5%** | **11581.6** | **<0.005** | **<0.01** | **Random** |
| **main meta-analysis** | **All** | **Overall** | **21.29** | **11.16 – 31.42** | **21.37 ± 5.31 (15.76–33.68)** | **56** | **Mixed** | **8104** | **99.5%** | **11581.6** | **<0.005** | **<0.01** | **Random** |
| main meta-analysis | black | 18+ | — | NA – NA | — | NA | NA | NA | — | — | — | — | NA |
| main meta-analysis | black | 20-30 | — | NA – NA | — | NA | NA | NA | — | — | — | — | NA |
| main meta-analysis | black | 30-40 | — | NA – NA | — | NA | NA | NA | — | — | — | — | NA |
| main meta-analysis | black | 40-50 | 25.00 | 10.25 – 39.74 | — (25.00–25.00) | 1 | Observational Cohort | 29 | — | — | — | — | Single study |
| main meta-analysis | black | 50-60 | 23.32 | 7.31 – 39.34 | — (23.32–23.32) | 1 | Observational Cohort | 131 | — | — | — | — | Single study |
| main meta-analysis | black | 60+ | 19.84 | 5.85 – 33.83 | 19.78 ± 0.91 (19.13–20.42) | 2 | Observational Cohort | 144 | 14% | 1.2 | 0.28 | — | Fixed |
| **main meta-analysis** | **black** | **Meta-Regression** | **—** | **NA – NA** | **—** | **NA** | **NA** | **NA** | **—** | **—** | **—** | **—** | **NA** |
| **main meta-analysis** | **black** | **Overall** | **21.86** | **17.18 – 26.54** | **21.97 ± 2.67 (19.13–25.00)** | **4** | **Observational Cohort** | **304** | **86.1%** | **21.6** | **<0.005** | **0.77** | **Random** |
| main meta-analysis | mixed | 18+ | 19.27 | 12.38 – 26.15 | 19.47 ± 3.97 (15.76–31.22) | 40 | Mixed | 3964 | 94.1% | 665.5 | <0.005 | 0.10 | Random |
| main meta-analysis | mixed | 20-30 | 26.42 | 7.87 – 44.96 | 26.37 ± 9.52 (19.64–33.10) | 2 | RCT | 46 | 98.8% | 84.6 | <0.005 | — | Random |
| main meta-analysis | mixed | 30-40 | 31.20 | 29.24 – 33.16 | — (31.20–31.20) | 1 | Observational Cohort | 107 | — | — | — | — | Single study |
| main meta-analysis | mixed | 40-50 | 18.08 | 15.92 – 20.24 | — (18.08–18.08) | 1 | Observational Cohort | 34 | — | — | — | — | Single study |
| main meta-analysis | mixed | 50-60 | — | NA – NA | — | NA | NA | NA | — | — | — | — | NA |
| main meta-analysis | mixed | 60+ | — | NA – NA | — | NA | NA | NA | — | — | — | — | NA |
| **main meta-analysis** | **mixed** | **Meta-Regression** | **—** | **NA – NA** | **—** | **NA** | **NA** | **NA** | **—** | **—** | **—** | **—** | **NA** |
| **main meta-analysis** | **mixed** | **Overall** | **19.92** | **11.19 – 28.66** | **20.02 ± 4.64 (15.76–33.10)** | **44** | **Mixed** | **4151** | **99.6%** | **10949.6** | **<0.005** | **<0.01** | **Random** |
| main meta-analysis | white | 18+ | — | NA – NA | — | NA | NA | NA | — | — | — | — | NA |
| main meta-analysis | white | 20-30 | — | NA – NA | — | NA | NA | NA | — | — | — | — | NA |
| main meta-analysis | white | 30-40 | — | NA – NA | — | NA | NA | NA | — | — | — | — | NA |
| main meta-analysis | white | 40-50 | 27.19 | 9.68 – 44.71 | 28.18 ± 1.43 (27.17–29.19) | 2 | Observational Cohort | 530 | 0% | 0.3 | 0.55 | — | Fixed |
| main meta-analysis | white | 50-60 | 29.75 | 21.45 – 38.04 | 29.86 ± 4.31 (26.81–32.91) | 2 | Observational Cohort | 2488 | 96.3% | 26.7 | <0.005 | — | Random |
| main meta-analysis | white | 60+ | 27.88 | 17.06 – 38.70 | 27.89 ± 5.57 (22.08–33.68) | 4 | Observational Cohort | 631 | 98.3% | 179.3 | <0.005 | 0.69 | Random |
| **main meta-analysis** | **white** | **Meta-Regression** | **—** | **NA – NA** | **—** | **NA** | **NA** | **NA** | **—** | **—** | **—** | **—** | **NA** |
| **main meta-analysis** | **white** | **Overall** | **28.37** | **20.27 – 36.47** | **28.46 ± 4.13 (22.08–33.68)** | **8** | **Observational Cohort** | **3649** | **96.6%** | **206.8** | **<0.005** | **0.48** | **Random** |
| male | All | 18+ | 19.54 | 5.06 – 34.02 | 20.49 ± 2.03 (19.05–21.92) | 2 | Observational Cohort | 130 | 64% | 2.8 | 0.10 | — | Fixed |
| male | All | 20-30 | 26.42 | 7.87 – 44.96 | 26.37 ± 9.52 (19.64–33.10) | 2 | RCT | 46 | 98.8% | 84.6 | <0.005 | — | Random |
| male | All | 30-40 | 31.20 | 29.24 – 33.16 | — (31.20–31.20) | 1 | Observational Cohort | 107 | — | — | — | — | Single study |
| male | All | 40-50 | 29.19 | 12.85 – 45.54 | — (29.19–29.19) | 1 | Observational Cohort | 6 | — | — | — | — | Single study |
| male | All | 50-60 | 32.91 | 12.54 – 53.27 | — (32.91–32.91) | 1 | Observational Cohort | 79 | — | — | — | — | Single study |
| male | All | 60+ | 33.08 | 13.47 – 52.68 | 32.59 ± 1.54 (31.50–33.68) | 2 | Observational Cohort | 304 | 65.5% | 2.9 | 0.09 | — | Fixed |
| **male** | **All** | **Meta-Regression** | **19.05** | **4.56 – 33.54** | **—** | **4** | **Mixed** | **262** | **99.4%** | **372** | **<0.005** | **0.32** | **Univariable** |
| **male** | **All** | **Overall** | **28.04** | **16.23 – 39.86** | **28.02 ± 6.05 (19.05–33.68)** | **9** | **Mixed** | **672** | **98.1%** | **424.5** | **<0.005** | **0.29** | **Random** |
| male | black | 18+ | — | NA – NA | — | NA | NA | NA | — | — | — | — | NA |
| male | black | 20-30 | — | NA – NA | — | NA | NA | NA | — | — | — | — | NA |
| male | black | 30-40 | — | NA – NA | — | NA | NA | NA | — | — | — | — | NA |
| male | black | 40-50 | — | NA – NA | — | NA | NA | NA | — | — | — | — | NA |
| male | black | 50-60 | — | NA – NA | — | NA | NA | NA | — | — | — | — | NA |
| male | black | 60+ | — | NA – NA | — | NA | NA | NA | — | — | — | — | NA |
| **male** | **black** | **Meta-Regression** | **—** | **NA – NA** | **—** | **NA** | **NA** | **NA** | **—** | **—** | **—** | **—** | **NA** |
| **male** | **black** | **Overall** | **—** | **NA – NA** | **—** | **NA** | **NA** | **NA** | **—** | **—** | **—** | **—** | **NA** |
| male | mixed | 18+ | 19.54 | 5.06 – 34.02 | 20.49 ± 2.03 (19.05–21.92) | 2 | Observational Cohort | 130 | 64% | 2.8 | 0.10 | — | Fixed |
| male | mixed | 20-30 | 26.42 | 7.87 – 44.96 | 26.37 ± 9.52 (19.64–33.10) | 2 | RCT | 46 | 98.8% | 84.6 | <0.005 | — | Random |
| male | mixed | 30-40 | 31.20 | 29.24 – 33.16 | — (31.20–31.20) | 1 | Observational Cohort | 107 | — | — | — | — | Single study |
| male | mixed | 40-50 | — | NA – NA | — | NA | NA | NA | — | — | — | — | NA |
| male | mixed | 50-60 | — | NA – NA | — | NA | NA | NA | — | — | — | — | NA |
| male | mixed | 60+ | — | NA – NA | — | NA | NA | NA | — | — | — | — | NA |
| **male** | **mixed** | **Meta-Regression** | **—** | **NA – NA** | **—** | **NA** | **NA** | **NA** | **—** | **—** | **—** | **—** | **NA** |
| **male** | **mixed** | **Overall** | **25.05** | **12.04 – 38.07** | **24.98 ± 6.66 (19.05–33.10)** | **5** | **Mixed** | **283** | **99%** | **405.1** | **<0.005** | **0.18** | **Random** |
| male | white | 18+ | — | NA – NA | — | NA | NA | NA | — | — | — | — | NA |
| male | white | 20-30 | — | NA – NA | — | NA | NA | NA | — | — | — | — | NA |
| male | white | 30-40 | — | NA – NA | — | NA | NA | NA | — | — | — | — | NA |
| male | white | 40-50 | 29.19 | 12.85 – 45.54 | — (29.19–29.19) | 1 | Observational Cohort | 6 | — | — | — | — | Single study |
| male | white | 50-60 | 32.91 | 12.54 – 53.27 | — (32.91–32.91) | 1 | Observational Cohort | 79 | — | — | — | — | Single study |
| male | white | 60+ | 33.08 | 13.47 – 52.68 | 32.59 ± 1.54 (31.50–33.68) | 2 | Observational Cohort | 304 | 65.5% | 2.9 | 0.09 | — | Fixed |
| **male** | **white** | **Meta-Regression** | **—** | **NA – NA** | **—** | **NA** | **NA** | **NA** | **—** | **—** | **—** | **—** | **NA** |
| **male** | **white** | **Overall** | **32.96** | **13.27 – 52.64** | **31.82 ± 1.97 (29.19–33.68)** | **4** | **Observational Cohort** | **389** | **27.9%** | **4.2** | **0.24** | **0.19** | **Fixed** |
| female | All | 18+ | 18.02 | 13.18 – 22.87 | 18.10 ± 2.64 (15.76–21.47) | 6 | Observational Cohort | 476 | 90.3% | 51.6 | <0.005 | 0.08 | Random |
| female | All | 20-30 | — | NA – NA | — | NA | NA | NA | — | — | — | — | NA |
| female | All | 30-40 | — | NA – NA | — | NA | NA | NA | — | — | — | — | NA |
| female | All | 40-50 | 27.01 | 9.67 – 44.35 | 26.08 ± 1.53 (25.00–27.17) | 2 | Observational Cohort | 553 | 55.3% | 2.2 | 0.13 | — | Fixed |
| female | All | 50-60 | 25.13 | 20.41 – 29.85 | 25.06 ± 2.46 (23.32–26.81) | 2 | Observational Cohort | 2540 | 95.6% | 22.5 | <0.005 | — | Random |
| female | All | 60+ | 21.56 | 17.35 – 25.76 | 21.49 ± 2.24 (19.13–24.31) | 4 | Observational Cohort | 471 | 91% | 33.3 | <0.005 | <0.05 | Random |
| **female** | **All** | **Meta-Regression** | **16.05** | **11.20 – 20.90** | **—** | **6** | **Observational Cohort** | **476** | **92.5%** | **51.6** | **<0.005** | **0.08** | **Univariable** |
| **female** | **All** | **Overall** | **21.21** | **13.70 – 28.71** | **21.20 ± 3.86 (15.76–27.17)** | **14** | **Observational Cohort** | **4040** | **98.8%** | **1078.8** | **<0.005** | **<0.05** | **Random** |
| female | black | 18+ | — | NA – NA | — | NA | NA | NA | — | — | — | — | NA |
| female | black | 20-30 | — | NA – NA | — | NA | NA | NA | — | — | — | — | NA |
| female | black | 30-40 | — | NA – NA | — | NA | NA | NA | — | — | — | — | NA |
| female | black | 40-50 | 25.00 | 10.25 – 39.74 | — (25.00–25.00) | 1 | Observational Cohort | 29 | — | — | — | — | Single study |
| female | black | 50-60 | 23.32 | 7.31 – 39.34 | — (23.32–23.32) | 1 | Observational Cohort | 131 | — | — | — | — | Single study |
| female | black | 60+ | 19.84 | 5.85 – 33.83 | 19.78 ± 0.91 (19.13–20.42) | 2 | Observational Cohort | 144 | 14% | 1.2 | 0.28 | — | Fixed |
| **female** | **black** | **Meta-Regression** | **—** | **NA – NA** | **—** | **NA** | **NA** | **NA** | **—** | **—** | **—** | **—** | **NA** |
| **female** | **black** | **Overall** | **21.86** | **17.18 – 26.54** | **21.97 ± 2.67 (19.13–25.00)** | **4** | **Observational Cohort** | **304** | **86.1%** | **21.6** | **<0.005** | **0.77** | **Random** |
| female | mixed | 18+ | 18.02 | 13.18 – 22.87 | 18.10 ± 2.64 (15.76–21.47) | 6 | Observational Cohort | 476 | 90.3% | 51.6 | <0.005 | 0.08 | Random |
| female | mixed | 20-30 | — | NA – NA | — | NA | NA | NA | — | — | — | — | NA |
| female | mixed | 30-40 | — | NA – NA | — | NA | NA | NA | — | — | — | — | NA |
| female | mixed | 40-50 | — | NA – NA | — | NA | NA | NA | — | — | — | — | NA |
| female | mixed | 50-60 | — | NA – NA | — | NA | NA | NA | — | — | — | — | NA |
| female | mixed | 60+ | — | NA – NA | — | NA | NA | NA | — | — | — | — | NA |
| **female** | **mixed** | **Meta-Regression** | **—** | **NA – NA** | **—** | **NA** | **NA** | **NA** | **—** | **—** | **—** | **—** | **NA** |
| **female** | **mixed** | **Overall** | **18.02** | **13.18 – 22.87** | **18.10 ± 2.64 (15.76–21.47)** | **6** | **Observational Cohort** | **476** | **90.3%** | **51.6** | **<0.005** | **0.08** | **Random** |
| female | white | 18+ | — | NA – NA | — | NA | NA | NA | — | — | — | — | NA |
| female | white | 20-30 | — | NA – NA | — | NA | NA | NA | — | — | — | — | NA |
| female | white | 30-40 | — | NA – NA | — | NA | NA | NA | — | — | — | — | NA |
| female | white | 40-50 | 27.17 | 9.64 – 44.69 | — (27.17–27.17) | 1 | Observational Cohort | 524 | — | — | — | — | Single study |
| female | white | 50-60 | 26.81 | 10.41 – 43.20 | — (26.81–26.81) | 1 | Observational Cohort | 2409 | — | — | — | — | Single study |
| female | white | 60+ | 23.28 | 20.48 – 26.08 | 23.20 ± 1.58 (22.08–24.31) | 2 | Observational Cohort | 327 | 81.7% | 5.5 | <0.05 | — | Random |
| **female** | **white** | **Meta-Regression** | **—** | **NA – NA** | **—** | **NA** | **NA** | **NA** | **—** | **—** | **—** | **—** | **NA** |
| **female** | **white** | **Overall** | **25.17** | **20.74 – 29.61** | **25.09 ± 2.37 (22.08–27.17)** | **4** | **Observational Cohort** | **3260** | **94.5%** | **54.1** | **<0.005** | **0.19** | **Random** |
| *I² represents heterogeneity; Q test assesses between-study variation; Egger's p = linear regression test for funnel plot asymmetry; Meta-Regression shows covariate-adjusted estimates* | | | | | | | | | | | | | |

###### 6.8.2.3 Table of Results subdivided by race / Sex / Age

phosphate_ma_3rd_analysis$race_summary_table

Table 1: **race-Stratified Meta-Analysis Results**

24-Hour Urine Phosphate (mmol/24h)

| Sex Group | race | Age Band | 95% Range | Studies | Study Type | Participants | I² | Cochran's Q | Q (p-value) | Egger's p | Model |
| --- | --- | --- | --- | --- | --- | --- | --- | --- | --- | --- | --- |
| **main meta-analysis** | **All races** | **18+** | **12.38 – 26.15** | **40** | **Mixed** | **3964** | **94.1%** | **665.5** | **<0.005** | **0.10** | **Random** |
| **main meta-analysis** | **All races** | **20-30** | **7.87 – 44.96** | **2** | **RCT** | **46** | **98.8%** | **84.6** | **<0.005** | **—** | **Random** |
| **main meta-analysis** | **All races** | **30-40** | **29.24 – 33.16** | **1** | **Observational Cohort** | **107** | **—** | **—** | **—** | **—** | **Single study** |
| **main meta-analysis** | **All races** | **40-50** | **15.30 – 33.56** | **4** | **Observational Cohort** | **593** | **99.3%** | **459.8** | **<0.005** | **0.46** | **Random** |
| **main meta-analysis** | **All races** | **50-60** | **18.38 – 36.84** | **3** | **Observational Cohort** | **2619** | **96.1%** | **50.9** | **<0.005** | **0.93** | **Random** |
| **main meta-analysis** | **All races** | **60+** | **13.49 – 36.88** | **6** | **Observational Cohort** | **775** | **98.3%** | **291.7** | **<0.005** | **0.92** | **Random** |
| **main meta-analysis** | **All races** | **Meta-Regression** | **18.87 – 26.40** | **40** | **Mixed** | **3974** | **89.7%** | **183.3** | **<0.005** | **<0.001** | **Multivariable** |
| **main meta-analysis** | **All races** | **Meta-Regression** | **18.87 – 26.40** | **40** | **Mixed** | **3974** | **89.7%** | **183.3** | **<0.005** | **<0.001** | **Multivariable** |
| **main meta-analysis** | **All races** | **Overall** | **11.16 – 31.42** | **56** | **Mixed** | **8104** | **99.5%** | **11581.6** | **<0.005** | **<0.01** | **Random** |
| **main meta-analysis** | **All races** | **Overall** | **11.16 – 31.42** | **56** | **Mixed** | **8104** | **99.5%** | **11581.6** | **<0.005** | **<0.01** | **Random** |
| main meta-analysis | Black | 18+ | — | — | NA | — | — | — | — | — | — |
| main meta-analysis | Black | 20-30 | — | — | NA | — | — | — | — | — | — |
| main meta-analysis | Black | 30-40 | — | — | NA | — | — | — | — | — | — |
| main meta-analysis | Black | 40-50 | 10.25 – 39.74 | 1 | Observational Cohort | 29 | — | — | — | — | Single study |
| main meta-analysis | Black | 50-60 | 7.31 – 39.34 | 1 | Observational Cohort | 131 | — | — | — | — | Single study |
| main meta-analysis | Black | 60+ | 5.85 – 33.83 | 2 | Observational Cohort | 144 | 14% | 1.2 | 0.28 | — | Fixed |
| main meta-analysis | Black | Meta-Regression | — | — | NA | — | — | — | — | — | — |
| **main meta-analysis** | **Black** | **Overall** | **17.18 – 26.54** | **4** | **Observational Cohort** | **304** | **86.1%** | **21.6** | **<0.005** | **0.77** | **Random** |
| main meta-analysis | Mixed | 18+ | 12.38 – 26.15 | 40 | Mixed | 3964 | 94.1% | 665.5 | <0.005 | 0.10 | Random |
| main meta-analysis | Mixed | 20-30 | 7.87 – 44.96 | 2 | RCT | 46 | 98.8% | 84.6 | <0.005 | — | Random |
| main meta-analysis | Mixed | 30-40 | 29.24 – 33.16 | 1 | Observational Cohort | 107 | — | — | — | — | Single study |
| main meta-analysis | Mixed | 40-50 | 15.92 – 20.24 | 1 | Observational Cohort | 34 | — | — | — | — | Single study |
| main meta-analysis | Mixed | 50-60 | — | — | NA | — | — | — | — | — | — |
| main meta-analysis | Mixed | 60+ | — | — | NA | — | — | — | — | — | — |
| main meta-analysis | Mixed | Meta-Regression | — | — | NA | — | — | — | — | — | — |
| **main meta-analysis** | **Mixed** | **Overall** | **11.19 – 28.66** | **44** | **Mixed** | **4151** | **99.6%** | **10949.6** | **<0.005** | **<0.01** | **Random** |
| main meta-analysis | White | 18+ | — | — | NA | — | — | — | — | — | — |
| main meta-analysis | White | 20-30 | — | — | NA | — | — | — | — | — | — |
| main meta-analysis | White | 30-40 | — | — | NA | — | — | — | — | — | — |
| main meta-analysis | White | 40-50 | 9.68 – 44.71 | 2 | Observational Cohort | 530 | 0% | 0.3 | 0.55 | — | Fixed |
| main meta-analysis | White | 50-60 | 21.45 – 38.04 | 2 | Observational Cohort | 2488 | 96.3% | 26.7 | <0.005 | — | Random |
| main meta-analysis | White | 60+ | 17.06 – 38.70 | 4 | Observational Cohort | 631 | 98.3% | 179.3 | <0.005 | 0.69 | Random |
| main meta-analysis | White | Meta-Regression | — | — | NA | — | — | — | — | — | — |
| **main meta-analysis** | **White** | **Overall** | **20.27 – 36.47** | **8** | **Observational Cohort** | **3649** | **96.6%** | **206.8** | **<0.005** | **0.48** | **Random** |
| **male** | **All races** | **18+** | **5.06 – 34.02** | **2** | **Observational Cohort** | **130** | **64%** | **2.8** | **0.10** | **—** | **Fixed** |
| **male** | **All races** | **20-30** | **7.87 – 44.96** | **2** | **RCT** | **46** | **98.8%** | **84.6** | **<0.005** | **—** | **Random** |
| **male** | **All races** | **30-40** | **29.24 – 33.16** | **1** | **Observational Cohort** | **107** | **—** | **—** | **—** | **—** | **Single study** |
| **male** | **All races** | **40-50** | **12.85 – 45.54** | **1** | **Observational Cohort** | **6** | **—** | **—** | **—** | **—** | **Single study** |
| **male** | **All races** | **50-60** | **12.54 – 53.27** | **1** | **Observational Cohort** | **79** | **—** | **—** | **—** | **—** | **Single study** |
| **male** | **All races** | **60+** | **13.47 – 52.68** | **2** | **Observational Cohort** | **304** | **65.5%** | **2.9** | **0.09** | **—** | **Fixed** |
| **male** | **All races** | **Meta-Regression** | **4.56 – 33.54** | **4** | **Mixed** | **262** | **99.4%** | **372** | **<0.005** | **0.32** | **Univariable** |
| **male** | **All races** | **Overall** | **16.23 – 39.86** | **9** | **Mixed** | **672** | **98.1%** | **424.5** | **<0.005** | **0.29** | **Random** |
| male | Black | 18+ | — | — | NA | — | — | — | — | — | — |
| male | Black | 20-30 | — | — | NA | — | — | — | — | — | — |
| male | Black | 30-40 | — | — | NA | — | — | — | — | — | — |
| male | Black | 40-50 | — | — | NA | — | — | — | — | — | — |
| male | Black | 50-60 | — | — | NA | — | — | — | — | — | — |
| male | Black | 60+ | — | — | NA | — | — | — | — | — | — |
| male | Black | Meta-Regression | — | — | NA | — | — | — | — | — | — |
| **male** | **Black** | **Overall** | **—** | **—** | **NA** | **—** | **—** | **—** | **—** | **—** | **—** |
| male | Mixed | 18+ | 5.06 – 34.02 | 2 | Observational Cohort | 130 | 64% | 2.8 | 0.10 | — | Fixed |
| male | Mixed | 20-30 | 7.87 – 44.96 | 2 | RCT | 46 | 98.8% | 84.6 | <0.005 | — | Random |
| male | Mixed | 30-40 | 29.24 – 33.16 | 1 | Observational Cohort | 107 | — | — | — | — | Single study |
| male | Mixed | 40-50 | — | — | NA | — | — | — | — | — | — |
| male | Mixed | 50-60 | — | — | NA | — | — | — | — | — | — |
| male | Mixed | 60+ | — | — | NA | — | — | — | — | — | — |
| male | Mixed | Meta-Regression | — | — | NA | — | — | — | — | — | — |
| **male** | **Mixed** | **Overall** | **12.04 – 38.07** | **5** | **Mixed** | **283** | **99%** | **405.1** | **<0.005** | **0.18** | **Random** |
| male | White | 18+ | — | — | NA | — | — | — | — | — | — |
| male | White | 20-30 | — | — | NA | — | — | — | — | — | — |
| male | White | 30-40 | — | — | NA | — | — | — | — | — | — |
| male | White | 40-50 | 12.85 – 45.54 | 1 | Observational Cohort | 6 | — | — | — | — | Single study |
| male | White | 50-60 | 12.54 – 53.27 | 1 | Observational Cohort | 79 | — | — | — | — | Single study |
| male | White | 60+ | 13.47 – 52.68 | 2 | Observational Cohort | 304 | 65.5% | 2.9 | 0.09 | — | Fixed |
| male | White | Meta-Regression | — | — | NA | — | — | — | — | — | — |
| **male** | **White** | **Overall** | **13.27 – 52.64** | **4** | **Observational Cohort** | **389** | **27.9%** | **4.2** | **0.24** | **0.19** | **Fixed** |
| **female** | **All races** | **18+** | **13.18 – 22.87** | **6** | **Observational Cohort** | **476** | **90.3%** | **51.6** | **<0.005** | **0.08** | **Random** |
| **female** | **All races** | **20-30** | **—** | **—** | **NA** | **—** | **—** | **—** | **—** | **—** | **—** |
| **female** | **All races** | **30-40** | **—** | **—** | **NA** | **—** | **—** | **—** | **—** | **—** | **—** |
| **female** | **All races** | **40-50** | **9.67 – 44.35** | **2** | **Observational Cohort** | **553** | **55.3%** | **2.2** | **0.13** | **—** | **Fixed** |
| **female** | **All races** | **50-60** | **20.41 – 29.85** | **2** | **Observational Cohort** | **2540** | **95.6%** | **22.5** | **<0.005** | **—** | **Random** |
| **female** | **All races** | **60+** | **17.35 – 25.76** | **4** | **Observational Cohort** | **471** | **91%** | **33.3** | **<0.005** | **<0.05** | **Random** |
| **female** | **All races** | **Meta-Regression** | **11.20 – 20.90** | **6** | **Observational Cohort** | **476** | **92.5%** | **51.6** | **<0.005** | **0.08** | **Univariable** |
| **female** | **All races** | **Overall** | **13.70 – 28.71** | **14** | **Observational Cohort** | **4040** | **98.8%** | **1078.8** | **<0.005** | **<0.05** | **Random** |
| female | Black | 18+ | — | — | NA | — | — | — | — | — | — |
| female | Black | 20-30 | — | — | NA | — | — | — | — | — | — |
| female | Black | 30-40 | — | — | NA | — | — | — | — | — | — |
| female | Black | 40-50 | 10.25 – 39.74 | 1 | Observational Cohort | 29 | — | — | — | — | Single study |
| female | Black | 50-60 | 7.31 – 39.34 | 1 | Observational Cohort | 131 | — | — | — | — | Single study |
| female | Black | 60+ | 5.85 – 33.83 | 2 | Observational Cohort | 144 | 14% | 1.2 | 0.28 | — | Fixed |
| female | Black | Meta-Regression | — | — | NA | — | — | — | — | — | — |
| **female** | **Black** | **Overall** | **17.18 – 26.54** | **4** | **Observational Cohort** | **304** | **86.1%** | **21.6** | **<0.005** | **0.77** | **Random** |
| female | Mixed | 18+ | 13.18 – 22.87 | 6 | Observational Cohort | 476 | 90.3% | 51.6 | <0.005 | 0.08 | Random |
| female | Mixed | 20-30 | — | — | NA | — | — | — | — | — | — |
| female | Mixed | 30-40 | — | — | NA | — | — | — | — | — | — |
| female | Mixed | 40-50 | — | — | NA | — | — | — | — | — | — |
| female | Mixed | 50-60 | — | — | NA | — | — | — | — | — | — |
| female | Mixed | 60+ | — | — | NA | — | — | — | — | — | — |
| female | Mixed | Meta-Regression | — | — | NA | — | — | — | — | — | — |
| **female** | **Mixed** | **Overall** | **13.18 – 22.87** | **6** | **Observational Cohort** | **476** | **90.3%** | **51.6** | **<0.005** | **0.08** | **Random** |
| female | White | 18+ | — | — | NA | — | — | — | — | — | — |
| female | White | 20-30 | — | — | NA | — | — | — | — | — | — |
| female | White | 30-40 | — | — | NA | — | — | — | — | — | — |
| female | White | 40-50 | 9.64 – 44.69 | 1 | Observational Cohort | 524 | — | — | — | — | Single study |
| female | White | 50-60 | 10.41 – 43.20 | 1 | Observational Cohort | 2409 | — | — | — | — | Single study |
| female | White | 60+ | 20.48 – 26.08 | 2 | Observational Cohort | 327 | 81.7% | 5.5 | <0.05 | — | Random |
| female | White | Meta-Regression | — | — | NA | — | — | — | — | — | — |
| **female** | **White** | **Overall** | **20.74 – 29.61** | **4** | **Observational Cohort** | **3260** | **94.5%** | **54.1** | **<0.005** | **0.19** | **Random** |
| *I² represents heterogeneity; Q test assesses between-study variation; Egger's p = linear regression test for funnel plot asymmetry; — indicates insufficient data* | | | | | | | | | | | |

###### 6.8.2.4 Wald Tests

phosphate_ma_3rd_analysis$wald_tests$sex_comparison

Table 1: **Sex Comparison (Overall)**

| Comparison | % Shift | P (Wald Test) | Mean Diff (95% CI) |
| --- | --- | --- | --- |
| Male vs Female (Overall) | +32.2% | 0.338 | 6.84 [-7.16, 20.83] |
| *Mean difference = Group 1 − Group 2 in component units (mmol/24h, L/24h, or pH); % shift = difference relative to Group 2.* | | | |

phosphate_ma_3rd_analysis$wald_tests$race_comparisons

Table 1: **race Comparisons Within Sex Groups**

| Sex Group | Comparison | % Shift | P (Wald Test) | Mean Diff (95% CI) |
| --- | --- | --- | --- | --- |
| male | black vs mixed | — | — | — |
| male | black vs white | — | — | — |
| male | mixed vs white | -24.0% | 0.512 | -7.90 [-31.50, 15.69] |
| female | black vs mixed | +21.3% | 0.265 | 3.83 [-2.91, 10.57] |
| female | black vs white | -13.2% | 0.314 | -3.31 [-9.76, 3.13] |
| female | mixed vs white | -28.4% | <0.05 | -7.15 [-13.72, -0.58] |
| *Mean difference = Group 1 − Group 2 in component units (mmol/24h, L/24h, or pH); % shift = difference relative to Group 2.* | | | | |

phosphate_ma_3rd_analysis$wald_tests$age_band_comparisons

Table 1: **Age Band Comparisons Within Sex Groups**

| Sex Group | Comparison | % Shift | P (Wald Test) | Mean Diff (95% CI) |
| --- | --- | --- | --- | --- |
| main meta-analysis | 18+ vs 20-30 | -27.1% | 0.479 | -7.15 [-26.93, 12.63] |
| main meta-analysis | 18+ vs 30-40 | -38.2% | <0.01 | -11.93 [-19.09, -4.77] |
| main meta-analysis | 18+ vs 40-50 | -21.1% | 0.376 | -5.16 [-16.60, 6.27] |
| main meta-analysis | 18+ vs 50-60 | -30.2% | 0.156 | -8.34 [-19.85, 3.17] |
| main meta-analysis | 18+ vs 60+ | -23.5% | 0.393 | -5.91 [-19.49, 7.66] |
| main meta-analysis | 20-30 vs 30-40 | -15.3% | 0.615 | -4.78 [-23.43, 13.87] |
| main meta-analysis | 20-30 vs 40-50 | +8.1% | 0.851 | 1.99 [-18.68, 22.66] |
| main meta-analysis | 20-30 vs 50-60 | -4.3% | 0.910 | -1.19 [-21.90, 19.52] |
| main meta-analysis | 20-30 vs 60+ | +4.9% | 0.912 | 1.24 [-20.69, 23.16] |
| main meta-analysis | 30-40 vs 40-50 | +27.7% | 0.155 | 6.77 [-2.57, 16.11] |
| main meta-analysis | 30-40 vs 50-60 | +13.0% | 0.456 | 3.59 [-5.84, 13.03] |
| main meta-analysis | 30-40 vs 60+ | +23.9% | 0.320 | 6.02 [-5.84, 17.88] |
| main meta-analysis | 40-50 vs 50-60 | -11.5% | 0.631 | -3.18 [-16.16, 9.81] |
| main meta-analysis | 40-50 vs 60+ | -3.0% | 0.921 | -0.75 [-15.59, 14.09] |
| main meta-analysis | 50-60 vs 60+ | +9.6% | 0.750 | 2.43 [-12.47, 17.32] |
| male | 18+ vs 20-30 | -26.0% | 0.566 | -6.88 [-30.41, 16.65] |
| male | 18+ vs 30-40 | -37.4% | 0.118 | -11.66 [-26.27, 2.95] |
| male | 18+ vs 40-50 | -33.1% | 0.386 | -9.65 [-31.49, 12.18] |
| male | 18+ vs 50-60 | -40.6% | 0.294 | -13.37 [-38.36, 11.62] |
| male | 18+ vs 60+ | -40.9% | 0.276 | -13.54 [-37.91, 10.83] |
| male | 20-30 vs 30-40 | -15.3% | 0.615 | -4.78 [-23.43, 13.87] |
| male | 20-30 vs 40-50 | -9.5% | 0.826 | -2.77 [-27.49, 21.95] |
| male | 20-30 vs 50-60 | -19.7% | 0.644 | -6.49 [-34.03, 21.05] |
| male | 20-30 vs 60+ | -20.1% | 0.629 | -6.66 [-33.64, 20.33] |
| male | 30-40 vs 40-50 | +6.9% | 0.811 | 2.01 [-14.45, 18.47] |
| male | 30-40 vs 50-60 | -5.2% | 0.870 | -1.71 [-22.16, 18.75] |
| male | 30-40 vs 60+ | -5.7% | 0.852 | -1.88 [-21.58, 17.82] |
| male | 40-50 vs 50-60 | -11.3% | 0.780 | -3.72 [-29.83, 22.40] |
| male | 40-50 vs 60+ | -11.7% | 0.766 | -3.88 [-29.41, 21.64] |
| male | 50-60 vs 60+ | -0.5% | 0.991 | -0.17 [-28.43, 28.10] |
| female | 18+ vs 20-30 | — | — | — |
| female | 18+ vs 30-40 | — | — | — |
| female | 18+ vs 40-50 | -33.3% | 0.328 | -8.98 [-26.99, 9.02] |
| female | 18+ vs 50-60 | -28.3% | <0.05 | -7.11 [-13.87, -0.34] |
| female | 18+ vs 60+ | -16.4% | 0.281 | -3.53 [-9.95, 2.89] |
| female | 20-30 vs 30-40 | — | — | — |
| female | 20-30 vs 40-50 | — | — | — |
| female | 20-30 vs 50-60 | — | — | — |
| female | 20-30 vs 60+ | — | — | — |
| female | 30-40 vs 40-50 | — | — | — |
| female | 30-40 vs 50-60 | — | — | — |
| female | 30-40 vs 60+ | — | — | — |
| female | 40-50 vs 50-60 | +7.5% | 0.838 | 1.88 [-16.09, 19.85] |
| female | 40-50 vs 60+ | +25.3% | 0.549 | 5.45 [-12.39, 23.30] |
| female | 50-60 vs 60+ | +16.6% | 0.267 | 3.58 [-2.74, 9.90] |
| *Mean difference = Group 1 − Group 2 in component units (mmol/24h, L/24h, or pH); % shift = difference relative to Group 2.* | | | | |

phosphate_ma_3rd_analysis$wald_tests$sex_within_age_bands

Table 1: **Sex Comparisons Within Age Bands**

| Age Band | % Shift | P (Wald Test) | Mean Diff (95% CI) |
| --- | --- | --- | --- |
| 18+ | +8.4% | 0.846 | 1.51 [-13.76, 16.78] |
| 20-30 | — | — | — |
| 30-40 | — | — | — |
| 40-50 | +8.1% | 0.858 | 2.18 [-21.65, 26.01] |
| 50-60 | +30.9% | 0.466 | 7.77 [-13.13, 28.68] |
| 60+ | +53.4% | 0.260 | 11.52 [-8.53, 31.57] |
| *Mean difference = Group 1 − Group 2 in component units (mmol/24h, L/24h, or pH); % shift = difference relative to Group 2.* | | | |

#### 6.9 Sodium

##### 6.9.1 Comparison by Age Band

###### 6.9.1.1 Plot

```
sodium_ma_2nd_analysis <- sex_age_subdivision_ma("sodium")
## Not enough studies for multivariable meta-regression (k = 4, need > 6). Running univariable models instead.
sodium_ma_2nd_analysis$combined_plot
## `height` was translated to `width`.
## `height` was translated to `width`.
## `height` was translated to `width`.
```

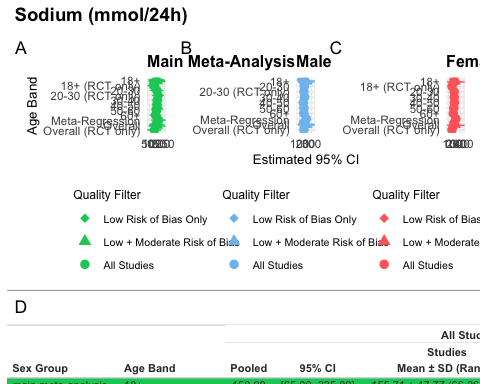

###### 6.9.1.2 Summary Table

sodium_ma_2nd_analysis$combined_summary_table

Table 1: **Combined Meta-Analysis Results**

24-Hour Urine Sodium (mmol/24h)

|  |  | **All Studies** | | | | | | **Low Risk of Bias Only** | | | | | | **Low + Moderate RoB** | | | | | |
| --- | --- | --- | --- | --- | --- | --- | --- | --- | --- | --- | --- | --- | --- | --- | --- | --- | --- | --- | --- |
| Sex Group | Age Band | Pooled | 95% CI | Studies  Mean ± SD (Range) | Studies, n | Q p-value | Egger's p | Pooled | 95% CI | Studies  Mean ± SD (Range) | Studies, n | Q p-value | Egger's p | Pooled | 95% CI | Studies  Mean ± SD (Range) | Studies, n | Q p-value | Egger's p |
| main meta-analysis | 18+ | 150.90 | [65.90, 235.89] | 155.71 ± 47.77 (66.26–237.00) | 66 | <0.001 | 0.07 | 159.68 | [64.47, 254.90] | 163.41 ± 50.17 (72.00–211.73) | 27 | <0.001 | 0.28 | 156.14 | [65.21, 247.08] | 159.69 ± 48.34 (72.00–211.73) | 32 | <0.001 | 0.24 |
| **main meta-analysis** | **18+ (RCT only)** | **117.48** | **[53.59, 181.36]** | **118.52 ± 34.09 (72.00–172.00)** | **7** | **<0.001** | **<0.05** | **—** | **—** | **—** | **—** | **—** | **—** | **—** | **—** | **—** | **—** | **—** | **—** |
| main meta-analysis | 20-30 | 157.00 | [88.79, 225.21] | 156.80 ± 35.18 (107.00–193.00) | 5 | <0.001 | 0.75 | — | — | — | — | — | — | — | — | — | — | — | — |
| **main meta-analysis** | **20-30 (RCT only)** | **170.96** | **[110.19, 231.72]** | **170.15 ± 32.31 (147.30–193.00)** | **2** | **<0.001** | **—** | **—** | **—** | **—** | **—** | **—** | **—** | **—** | **—** | **—** | **—** | **—** | **—** |
| main meta-analysis | 30-40 | 162.56 | [110.21, 214.91] | 162.60 ± 26.98 (131.00–190.20) | 4 | <0.001 | 0.37 | — | — | — | — | — | — | — | — | — | — | — | — |
| main meta-analysis | 40-50 | 157.91 | [102.97, 212.85] | 155.03 ± 4.76 (150.52–161.20) | 6 | 0.06 | 0.48 | — | — | — | — | — | — | 160.60 | [141.39, 179.81] | — (160.60–160.60) | 1 | — | — |
| main meta-analysis | 50-60 | 153.46 | [118.66, 188.26] | 154.48 ± 18.91 (136.30–178.18) | 5 | <0.001 | 0.12 | — | — | — | — | — | — | — | — | — | — | — | — |
| main meta-analysis | 60+ | 142.70 | [81.75, 203.66] | 142.90 ± 31.05 (78.60–175.43) | 8 | <0.001 | 0.11 | — | — | — | — | — | — | — | — | — | — | — | — |
| **main meta-analysis** | **Meta-Regression** | **175.70** | **[98.06, 253.33]** | **—** | **47** | **<0.001** | **<0.05** | **—** | **—** | **—** | **—** | **—** | **—** | **—** | **—** | **—** | **—** | **—** | **—** |
| **main meta-analysis** | **Overall** | **151.03** | **[76.94, 225.12]** | **154.86 ± 42.15 (66.26–237.00)** | **94** | **<0.001** | **0.22** | **159.68** | **[64.47, 254.90]** | **163.41 ± 50.17 (72.00–211.73)** | **27** | **<0.001** | **0.28** | **156.24** | **[66.94, 245.54]** | **159.71 ± 47.58 (72.00–211.73)** | **33** | **<0.001** | **0.88** |
| **main meta-analysis** | **Overall (RCT only)** | **129.77** | **[54.25, 205.30]** | **130.00 ± 38.99 (72.00–193.00)** | **9** | **<0.001** | **0.78** | **—** | **—** | **—** | **—** | **—** | **—** | **—** | **—** | **—** | **—** | **—** | **—** |
| male | 18+ | 139.85 | [55.25, 224.45] | 140.31 ± 43.65 (86.10–211.73) | 7 | <0.001 | 0.90 | 211.73 | [75.45, 348.01] | — (211.73–211.73) | 1 | — | — | 211.73 | [75.45, 348.01] | — (211.73–211.73) | 1 | — | — |
| **male** | **18+ (RCT only)** | **—** | **—** | **—** | **—** | **—** | **—** | **—** | **—** | **—** | **—** | **—** | **—** | **—** | **—** | **—** | **—** | **—** | **—** |
| male | 20-30 | 177.72 | [133.63, 221.80] | 176.23 ± 25.16 (147.30–193.00) | 3 | <0.001 | 0.57 | — | — | — | — | — | — | — | — | — | — | — | — |
| **male** | **20-30 (RCT only)** | **170.96** | **[110.19, 231.72]** | **170.15 ± 32.31 (147.30–193.00)** | **2** | **<0.001** | **—** | **—** | **—** | **—** | **—** | **—** | **—** | **—** | **—** | **—** | **—** | **—** | **—** |
| male | 30-40 | 183.77 | [169.55, 197.99] | 184.60 ± 7.92 (179.00–190.20) | 2 | <0.05 | — | — | — | — | — | — | — | — | — | — | — | — | — |
| male | 40-50 | 161.09 | [132.65, 189.54] | 155.94 ± 7.45 (150.67–161.20) | 2 | 0.68 | — | — | — | — | — | — | — | — | — | — | — | — | — |
| male | 50-60 | 171.71 | [104.40, 239.02] | 174.44 ± 5.29 (170.70–178.18) | 2 | 0.45 | — | — | — | — | — | — | — | — | — | — | — | — | — |
| male | 60+ | 169.33 | [97.42, 241.23] | 171.03 ± 3.97 (167.70–175.43) | 3 | 0.33 | 0.55 | — | — | — | — | — | — | — | — | — | — | — | — |
| **male** | **Meta-Regression** | **211.73** | **[163.04, 260.42]** | **—** | **4** | **<0.001** | **0.65** | **—** | **—** | **—** | **—** | **—** | **—** | **—** | **—** | **—** | **—** | **—** | **—** |
| **male** | **Overall** | **160.98** | **[98.71, 223.25]** | **160.73 ± 31.97 (86.10–211.73)** | **19** | **<0.001** | **0.17** | **211.73** | **[75.45, 348.01]** | **— (211.73–211.73)** | **1** | **—** | **—** | **211.73** | **[75.45, 348.01]** | **— (211.73–211.73)** | **1** | **—** | **—** |
| **male** | **Overall (RCT only)** | **170.96** | **[110.19, 231.72]** | **170.15 ± 32.31 (147.30–193.00)** | **2** | **<0.001** | **—** | **—** | **—** | **—** | **—** | **—** | **—** | **—** | **—** | **—** | **—** | **—** | **—** |
| female | 18+ | 142.05 | [81.95, 202.14] | 142.34 ± 31.66 (90.00–190.17) | 9 | <0.001 | <0.05 | 190.17 | [-32.02, 412.36] | 190.17 ± 0.00 (190.17–190.17) | 2 | 1.00 | — | 156.51 | [43.85, 269.18] | 156.78 ± 57.83 (90.00–190.17) | 3 | <0.001 | <0.001 |
| **female** | **18+ (RCT only)** | **90.00** | **[46.88, 133.12]** | **— (90.00–90.00)** | **1** | **—** | **—** | **—** | **—** | **—** | **—** | **—** | **—** | **—** | **—** | **—** | **—** | **—** | **—** |
| female | 20-30 | 148.30 | [128.90, 167.70] | — (148.30–148.30) | 1 | — | — | — | — | — | — | — | — | — | — | — | — | — | — |
| **female** | **20-30 (RCT only)** | **—** | **—** | **—** | **—** | **—** | **—** | **—** | **—** | **—** | **—** | **—** | **—** | **—** | **—** | **—** | **—** | **—** | **—** |
| female | 30-40 | 150.20 | [122.56, 177.84] | — (150.20–150.20) | 1 | — | — | — | — | — | — | — | — | — | — | — | — | — | — |
| female | 40-50 | 153.44 | [69.76, 237.11] | 152.58 ± 1.89 (150.52–154.22) | 3 | 0.91 | 0.65 | — | — | — | — | — | — | — | — | — | — | — | — |
| female | 50-60 | 137.26 | [38.43, 236.10] | 141.18 ± 6.11 (136.30–148.03) | 3 | 0.08 | <0.05 | — | — | — | — | — | — | — | — | — | — | — | — |
| female | 60+ | 125.60 | [72.44, 178.76] | 126.02 ± 27.01 (78.60–146.46) | 5 | <0.001 | <0.05 | — | — | — | — | — | — | — | — | — | — | — | — |
| **female** | **Meta-Regression** | **190.17** | **[178.07, 202.27]** | **—** | **7** | **0.17** | **<0.01** | **—** | **—** | **—** | **—** | **—** | **—** | **—** | **—** | **—** | **—** | **—** | **—** |
| **female** | **Overall** | **140.13** | **[93.71, 186.55]** | **140.50 ± 24.58 (78.60–190.17)** | **22** | **<0.001** | **0.28** | **190.17** | **[-32.02, 412.36]** | **190.17 ± 0.00 (190.17–190.17)** | **2** | **1.00** | **—** | **156.51** | **[43.85, 269.18]** | **156.78 ± 57.83 (90.00–190.17)** | **3** | **<0.001** | **<0.001** |
| **female** | **Overall (RCT only)** | **90.00** | **[46.88, 133.12]** | **— (90.00–90.00)** | **1** | **—** | **—** | **—** | **—** | **—** | **—** | **—** | **—** | **—** | **—** | **—** | **—** | **—** | **—** |
| *Studies, n = number of studies; Q p-value = Cochran's Q heterogeneity test; Egger's p = linear regression test for funnel plot asymmetry; RoB = Risk of Bias* | | | | | | | | | | | | | | | | | | | |

###### 6.9.1.3 Wald Tests

sodium_ma_2nd_analysis$wald_tests$sex_comparison

Table 1: **Sex Comparison (Overall)**

| Comparison | % Shift | P (Wald Test) | Mean Diff (95% CI) |
| --- | --- | --- | --- |
| Male vs Female (Overall) | +14.9% | 0.599 | 20.85 [-56.82, 98.52] |
| *Mean difference = Group 1 − Group 2 in component units (mmol/24h, L/24h, or pH); % shift = difference relative to Group 2.* | | | |

sodium_ma_2nd_analysis$wald_tests$age_band_comparisons

Table 1: **Age Band Comparisons Within Sex Groups**

| Sex Group | Comparison | % Shift | P (Wald Test) | Mean Diff (95% CI) |
| --- | --- | --- | --- | --- |
| main meta-analysis | 18+ vs 20-30 | -3.9% | 0.913 | -6.10 [-115.08, 102.88] |
| main meta-analysis | 18+ vs 30-40 | -7.2% | 0.819 | -11.66 [-111.48, 88.16] |
| main meta-analysis | 18+ vs 40-50 | -4.4% | 0.892 | -7.02 [-108.22, 94.19] |
| main meta-analysis | 18+ vs 50-60 | -1.7% | 0.956 | -2.56 [-94.41, 89.28] |
| main meta-analysis | 18+ vs 60+ | +5.7% | 0.878 | 8.19 [-96.40, 112.79] |
| main meta-analysis | 20-30 vs 30-40 | -3.4% | 0.899 | -5.56 [-91.54, 80.43] |
| main meta-analysis | 20-30 vs 40-50 | -0.6% | 0.984 | -0.91 [-88.50, 86.67] |
| main meta-analysis | 20-30 vs 50-60 | +2.3% | 0.928 | 3.54 [-73.03, 80.12] |
| main meta-analysis | 20-30 vs 60+ | +10.0% | 0.759 | 14.30 [-77.18, 105.77] |
| main meta-analysis | 30-40 vs 40-50 | +2.9% | 0.905 | 4.64 [-71.25, 80.53] |
| main meta-analysis | 30-40 vs 50-60 | +5.9% | 0.777 | 9.10 [-53.76, 71.96] |
| main meta-analysis | 30-40 vs 60+ | +13.9% | 0.628 | 19.85 [-60.50, 100.20] |
| main meta-analysis | 40-50 vs 50-60 | +2.9% | 0.893 | 4.46 [-60.58, 69.49] |
| main meta-analysis | 40-50 vs 60+ | +10.7% | 0.716 | 15.21 [-66.85, 97.27] |
| main meta-analysis | 50-60 vs 60+ | +7.5% | 0.764 | 10.75 [-59.44, 80.95] |
| male | 18+ vs 20-30 | -21.3% | 0.437 | -37.87 [-133.27, 57.53] |
| male | 18+ vs 30-40 | -23.9% | 0.316 | -43.92 [-129.71, 41.87] |
| male | 18+ vs 40-50 | -13.2% | 0.641 | -21.24 [-110.50, 68.01] |
| male | 18+ vs 50-60 | -18.6% | 0.564 | -31.86 [-139.97, 76.25] |
| male | 18+ vs 60+ | -17.4% | 0.603 | -29.48 [-140.51, 81.55] |
| male | 20-30 vs 30-40 | -3.3% | 0.798 | -6.05 [-52.37, 40.27] |
| male | 20-30 vs 40-50 | +10.3% | 0.534 | 16.63 [-35.84, 69.09] |
| male | 20-30 vs 50-60 | +3.5% | 0.884 | 6.01 [-74.45, 86.47] |
| male | 20-30 vs 60+ | +5.0% | 0.845 | 8.39 [-75.95, 92.74] |
| male | 30-40 vs 40-50 | +14.1% | 0.162 | 22.68 [-9.12, 54.48] |
| male | 30-40 vs 50-60 | +7.0% | 0.731 | 12.06 [-56.73, 80.86] |
| male | 30-40 vs 60+ | +8.5% | 0.699 | 14.44 [-58.85, 87.74] |
| male | 40-50 vs 50-60 | -6.2% | 0.776 | -10.62 [-83.69, 62.46] |
| male | 40-50 vs 60+ | -4.9% | 0.835 | -8.23 [-85.56, 69.09] |
| male | 50-60 vs 60+ | +1.4% | 0.962 | 2.38 [-96.11, 100.87] |
| female | 18+ vs 20-30 | -4.2% | 0.846 | -6.25 [-69.40, 56.89] |
| female | 18+ vs 30-40 | -5.4% | 0.809 | -8.15 [-74.30, 57.99] |
| female | 18+ vs 40-50 | -7.4% | 0.828 | -11.39 [-114.41, 91.62] |
| female | 18+ vs 50-60 | +3.5% | 0.935 | 4.78 [-110.89, 120.45] |
| female | 18+ vs 60+ | +13.1% | 0.688 | 16.44 [-63.79, 96.67] |
| female | 20-30 vs 30-40 | -1.3% | 0.912 | -1.90 [-35.67, 31.87] |
| female | 20-30 vs 40-50 | -3.3% | 0.907 | -5.14 [-91.03, 80.76] |
| female | 20-30 vs 50-60 | +8.0% | 0.830 | 11.04 [-89.68, 111.76] |
| female | 20-30 vs 60+ | +18.1% | 0.432 | 22.70 [-33.90, 79.29] |
| female | 30-40 vs 40-50 | -2.1% | 0.943 | -3.24 [-91.36, 84.88] |
| female | 30-40 vs 50-60 | +9.4% | 0.805 | 12.94 [-89.69, 115.56] |
| female | 30-40 vs 60+ | +19.6% | 0.421 | 24.60 [-35.32, 84.51] |
| female | 40-50 vs 50-60 | +11.8% | 0.807 | 16.18 [-113.32, 145.67] |
| female | 40-50 vs 60+ | +22.2% | 0.582 | 27.83 [-71.30, 126.97] |
| female | 50-60 vs 60+ | +9.3% | 0.839 | 11.66 [-100.56, 123.88] |
| *Mean difference = Group 1 − Group 2 in component units (mmol/24h, L/24h, or pH); % shift = difference relative to Group 2.* | | | | |

sodium_ma_2nd_analysis$wald_tests$sex_within_age_bands

Table 1: **Sex Comparisons Within Age Bands**

| Age Band | % Shift | P (Wald Test) | Mean Diff (95% CI) |
| --- | --- | --- | --- |
| 18+ | -1.5% | 0.967 | -2.20 [-105.97, 101.57] |
| 20-30 | +19.8% | 0.231 | 29.42 [-18.75, 77.59] |
| 30-40 | +22.3% | <0.05 | 33.57 [2.49, 64.65] |
| 40-50 | +5.0% | 0.865 | 7.65 [-80.73, 96.03] |
| 50-60 | +25.1% | 0.572 | 34.44 [-85.13, 154.02] |
| 60+ | +34.8% | 0.338 | 43.72 [-45.70, 133.14] |
| *Mean difference = Group 1 − Group 2 in component units (mmol/24h, L/24h, or pH); % shift = difference relative to Group 2.* | | | |

###### 6.9.1.4 Meta-Regression (plain language)

format_meta_reg_plain("sodium", sex_group = "all")

Message

No meta-regression result for sodium / all

###### 6.9.1.5 Assay Subgroup Analysis

assay_subgroup_table("sodium")

Table 1: **Assay subgroup analysis — Sodium**

*Per-assay pooled estimates; categories with k < 3 listed but not pooled.*

| Assay category | Studies, n | Participants | 95% CI | I² | Q p-value | Egger's p | Note |
| --- | --- | --- | --- | --- | --- | --- | --- |
| Unreported | 63 | 6,173 | 144.72 [135.10, 154.35] | 99% | <0.001 | 0.193 |  |
| Autoanalyser | 20 | 2,510 | 176.96 [161.58, 192.33] | 96% | <0.001 | 0.781 |  |
| Flame photometry / emission | 6 | 311 | 137.65 [97.72, 177.58] | 98% | <0.001 | 0.992 |  |
| Ion-selective electrode | 5 | 126 | 147.64 [136.67, 158.61] | 19% | 0.296 | 0.042 |  |

###### 6.9.1.6 Preservative Sensitivity (Litholink / acidified samples)

```
preservative_sensitivity_ma("sodium")
## # A tibble: 3 × 6
##   group               n_studies n_participants pooled_estimate lower_95 upper_95
##   <chr>                   <int>          <int>           <dbl>    <dbl>    <dbl>
## 1 All studies                94           9120            151.     143.     159.
## 2 Excluding preserva…        91           9009            151.     142.     159.
## 3 Preservative only           3            111            159.     133.     185.
```

###### 6.9.1.7 Creatinine Completeness Sensitivity

creat_completeness_table("sodium")

Table 1: **Creatinine-completeness sensitivity — Sodium**

*Compares the all-studies pooled estimate against subsets defined by whether the study reported 24h urine creatinine. Studies that do not report creatinine cannot verify collection completeness; a substantial shift here flags possible bias from incomplete collections.*

| Subset | Studies, n | Participants | Range (95% CI) | Δ vs all-studies (%) | Flag |
| --- | --- | --- | --- | --- | --- |
| All studies | 94 | 9,120 | 151.03 [142.95, 159.11] | — | — |
| Creatinine reported only | 69 | 8,145 | 149.07 [139.86, 158.29] | -1.3% | OK |
| Creatinine NOT reported | 25 | 975 | 157.52 [140.24, 174.81] | +4.3% | OK |

##### 6.9.2 Comparison by Age / Sex / race

###### 6.9.2.1 Plot

```
sodium_ma_3rd_analysis <- sex_age_race_subdivision_ma("sodium",
                                                      urine_ma2,
                                                      overall_bias)
## Not enough studies for multivariable meta-regression (k = 4, need > 6). Running univariable models instead.
sodium_ma_3rd_analysis$race_plot
## `height` was translated to `width`.
## `height` was translated to `width`.
## `height` was translated to `width`.
```

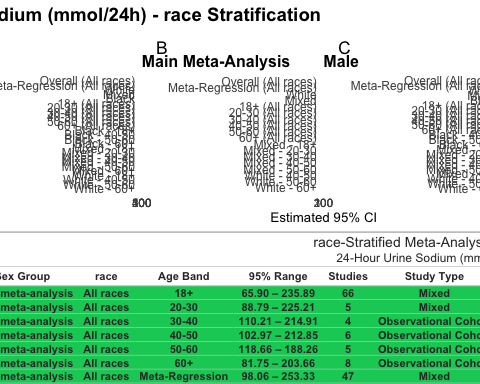

###### 6.9.2.2 Combined MA Results

sodium_ma_3rd_analysis$combined_summary_table

Table 1: **Combined Meta-Analysis Results**

24-Hour Urine Sodium (mmol/24h)

| Sex Group | race | Age Band | Pooled | 95% Range | Studies Mean SD Range | Studies | Study Type | Participants | I² | Cochran's Q | Q (p-value) | Egger's p | Model |
| --- | --- | --- | --- | --- | --- | --- | --- | --- | --- | --- | --- | --- | --- |
| main meta-analysis | All | 18+ | 150.90 | 65.90 – 235.89 | 155.71 ± 47.77 (66.26–237.00) | 66 | Mixed | 4660 | 97.2% | 2335.4 | <0.005 | 0.07 | Random |
| main meta-analysis | All | 20-30 | 157.00 | 88.79 – 225.21 | 156.80 ± 35.18 (107.00–193.00) | 5 | Mixed | 149 | 98.7% | 303 | <0.005 | 0.75 | Random |
| main meta-analysis | All | 30-40 | 162.56 | 110.21 – 214.91 | 162.60 ± 26.98 (131.00–190.20) | 4 | Observational Cohort | 174 | 99% | 303.8 | <0.005 | 0.37 | Random |
| main meta-analysis | All | 40-50 | 157.91 | 102.97 – 212.85 | 155.03 ± 4.76 (150.52–161.20) | 6 | Observational Cohort | 643 | 53.5% | 10.7 | 0.06 | 0.48 | Fixed |
| main meta-analysis | All | 50-60 | 153.46 | 118.66 – 188.26 | 154.48 ± 18.91 (136.30–178.18) | 5 | Observational Cohort | 2669 | 96.1% | 102 | <0.005 | 0.12 | Random |
| main meta-analysis | All | 60+ | 142.70 | 81.75 – 203.66 | 142.90 ± 31.05 (78.60–175.43) | 8 | Observational Cohort | 825 | 99.5% | 1371.2 | <0.005 | 0.11 | Random |
| **main meta-analysis** | **All** | **Meta-Regression** | **175.70** | **98.06 – 253.33** | **—** | **47** | **Mixed** | **3945** | **95.4%** | **857.4** | **<0.005** | **<0.05** | **Multivariable** |
| **main meta-analysis** | **All** | **Meta-Regression** | **175.70** | **98.06 – 253.33** | **—** | **47** | **Mixed** | **3945** | **95.4%** | **857.4** | **<0.005** | **<0.05** | **Multivariable** |
| **main meta-analysis** | **All** | **Overall** | **151.03** | **76.94 – 225.12** | **154.86 ± 42.15 (66.26–237.00)** | **94** | **Mixed** | **9120** | **98.8%** | **7915.6** | **<0.005** | **0.22** | **Random** |
| **main meta-analysis** | **All** | **Overall** | **151.03** | **76.94 – 225.12** | **154.86 ± 42.15 (66.26–237.00)** | **94** | **Mixed** | **9120** | **98.8%** | **7915.6** | **<0.005** | **0.22** | **Random** |
| main meta-analysis | black | 18+ | 237.00 | -17.80 – 491.80 | 237.00 ± 0.00 (237.00–237.00) | 3 | Observational Cohort | 51 | 0% | 0 | 1.00 | — | Fixed |
| main meta-analysis | black | 20-30 | — | NA – NA | — | NA | NA | NA | — | — | — | — | NA |
| main meta-analysis | black | 30-40 | — | NA – NA | — | NA | NA | NA | — | — | — | — | NA |
| main meta-analysis | black | 40-50 | 150.52 | 37.60 – 263.44 | — (150.52–150.52) | 1 | Observational Cohort | 29 | — | — | — | — | Single study |
| main meta-analysis | black | 50-60 | 148.03 | 22.20 – 273.86 | — (148.03–148.03) | 1 | Observational Cohort | 131 | — | — | — | — | Single study |
| main meta-analysis | black | 60+ | 142.55 | 35.40 – 249.70 | 141.83 ± 6.55 (137.20–146.46) | 2 | Observational Cohort | 144 | 0.8% | 1 | 0.32 | — | Fixed |
| **main meta-analysis** | **black** | **Meta-Regression** | **—** | **NA – NA** | **—** | **NA** | **NA** | **NA** | **—** | **—** | **—** | **—** | **NA** |
| **main meta-analysis** | **black** | **Overall** | **175.10** | **96.95 – 253.25** | **184.74 ± 49.05 (137.20–237.00)** | **7** | **Observational Cohort** | **355** | **77.3%** | **26.4** | **<0.005** | **<0.01** | **Random** |
| main meta-analysis | mixed | 18+ | 145.24 | 64.68 – 225.80 | 148.07 ± 43.97 (72.00–211.73) | 55 | Mixed | 4241 | 96.2% | 1427.4 | <0.005 | 0.08 | Random |
| main meta-analysis | mixed | 20-30 | 157.00 | 88.79 – 225.21 | 156.80 ± 35.18 (107.00–193.00) | 5 | Mixed | 149 | 98.7% | 303 | <0.005 | 0.75 | Random |
| main meta-analysis | mixed | 30-40 | 162.56 | 110.21 – 214.91 | 162.60 ± 26.98 (131.00–190.20) | 4 | Observational Cohort | 174 | 99% | 303.8 | <0.005 | 0.37 | Random |
| main meta-analysis | mixed | 40-50 | 158.33 | 150.73 – 165.94 | 158.27 ± 4.57 (153.00–161.20) | 3 | Observational Cohort | 84 | 74.8% | 7.9 | <0.05 | 0.74 | Random |
| main meta-analysis | mixed | 50-60 | 154.82 | 111.56 – 198.08 | 154.95 ± 22.27 (139.20–170.70) | 2 | Observational Cohort | 50 | 98.2% | 55.3 | <0.005 | — | Random |
| main meta-analysis | mixed | 60+ | 78.60 | 63.70 – 93.50 | — (78.60–78.60) | 1 | Observational Cohort | 25 | — | — | — | — | Single study |
| **main meta-analysis** | **mixed** | **Meta-Regression** | **—** | **NA – NA** | **—** | **NA** | **NA** | **NA** | **—** | **—** | **—** | **—** | **NA** |
| **main meta-analysis** | **mixed** | **Overall** | **146.97** | **70.74 – 223.21** | **149.16 ± 41.43 (72.00–211.73)** | **70** | **Mixed** | **4723** | **98.9%** | **6531.2** | **<0.005** | **0.42** | **Random** |
| main meta-analysis | white | 18+ | 171.54 | 68.01 – 275.06 | 179.61 ± 52.96 (66.26–216.00) | 7 | Observational Cohort | 328 | 98% | 301.1 | <0.005 | <0.05 | Random |
| main meta-analysis | white | 20-30 | — | NA – NA | — | NA | NA | NA | — | — | — | — | NA |
| main meta-analysis | white | 30-40 | — | NA – NA | — | NA | NA | NA | — | — | — | — | NA |
| main meta-analysis | white | 40-50 | 154.18 | 30.62 – 277.74 | 152.44 ± 2.51 (150.67–154.22) | 2 | Observational Cohort | 530 | 0% | 0 | 0.89 | — | Fixed |
| main meta-analysis | white | 50-60 | 156.25 | 99.64 – 212.86 | 157.24 ± 29.61 (136.30–178.18) | 2 | Observational Cohort | 2488 | 95.1% | 20.5 | <0.005 | — | Random |
| main meta-analysis | white | 60+ | 156.12 | 117.00 – 195.24 | 156.19 ± 20.53 (133.13–175.43) | 5 | Observational Cohort | 656 | 95.9% | 98.4 | <0.005 | 0.72 | Random |
| **main meta-analysis** | **white** | **Meta-Regression** | **—** | **NA – NA** | **—** | **NA** | **NA** | **NA** | **—** | **—** | **—** | **—** | **NA** |
| **main meta-analysis** | **white** | **Overall** | **158.91** | **94.09 – 223.74** | **166.10 ± 38.03 (66.26–216.00)** | **16** | **Observational Cohort** | **4002** | **98.9%** | **1305.8** | **<0.005** | **0.34** | **Random** |
| male | All | 18+ | 139.85 | 55.25 – 224.45 | 140.31 ± 43.65 (86.10–211.73) | 7 | Observational Cohort | 367 | 98.1% | 315.8 | <0.005 | 0.90 | Random |
| male | All | 20-30 | 177.72 | 133.63 – 221.80 | 176.23 ± 25.16 (147.30–193.00) | 3 | Mixed | 71 | 86% | 14.3 | <0.005 | 0.57 | Random |
| male | All | 30-40 | 183.77 | 169.55 – 197.99 | 184.60 ± 7.92 (179.00–190.20) | 2 | Observational Cohort | 132 | 84% | 6.2 | <0.05 | — | Random |
| male | All | 40-50 | 161.09 | 132.65 – 189.54 | 155.94 ± 7.45 (150.67–161.20) | 2 | Observational Cohort | 31 | 0% | 0.2 | 0.68 | — | Fixed |
| male | All | 50-60 | 171.71 | 104.40 – 239.02 | 174.44 ± 5.29 (170.70–178.18) | 2 | Observational Cohort | 104 | 0% | 0.6 | 0.45 | — | Fixed |
| male | All | 60+ | 169.33 | 97.42 – 241.23 | 171.03 ± 3.97 (167.70–175.43) | 3 | Observational Cohort | 329 | 8.7% | 2.2 | 0.33 | 0.55 | Fixed |
| **male** | **All** | **Meta-Regression** | **211.73** | **163.04 – 260.42** | **—** | **4** | **Mixed** | **262** | **95.5%** | **36.3** | **<0.005** | **0.65** | **Univariable** |
| **male** | **All** | **Overall** | **160.98** | **98.71 – 223.25** | **160.73 ± 31.97 (86.10–211.73)** | **19** | **Mixed** | **1034** | **99.5%** | **3691.7** | **<0.005** | **0.17** | **Random** |
| male | black | 18+ | — | NA – NA | — | NA | NA | NA | — | — | — | — | NA |
| male | black | 20-30 | — | NA – NA | — | NA | NA | NA | — | — | — | — | NA |
| male | black | 30-40 | — | NA – NA | — | NA | NA | NA | — | — | — | — | NA |
| male | black | 40-50 | — | NA – NA | — | NA | NA | NA | — | — | — | — | NA |
| male | black | 50-60 | — | NA – NA | — | NA | NA | NA | — | — | — | — | NA |
| male | black | 60+ | — | NA – NA | — | NA | NA | NA | — | — | — | — | NA |
| **male** | **black** | **Meta-Regression** | **—** | **NA – NA** | **—** | **NA** | **NA** | **NA** | **—** | **—** | **—** | **—** | **NA** |
| **male** | **black** | **Overall** | **—** | **NA – NA** | **—** | **NA** | **NA** | **NA** | **—** | **—** | **—** | **—** | **NA** |
| male | mixed | 18+ | 139.85 | 55.25 – 224.45 | 140.31 ± 43.65 (86.10–211.73) | 7 | Observational Cohort | 367 | 98.1% | 315.8 | <0.005 | 0.90 | Random |
| male | mixed | 20-30 | 177.72 | 133.63 – 221.80 | 176.23 ± 25.16 (147.30–193.00) | 3 | Mixed | 71 | 86% | 14.3 | <0.005 | 0.57 | Random |
| male | mixed | 30-40 | 183.77 | 169.55 – 197.99 | 184.60 ± 7.92 (179.00–190.20) | 2 | Observational Cohort | 132 | 84% | 6.2 | <0.05 | — | Random |
| male | mixed | 40-50 | 161.20 | 135.52 – 186.88 | — (161.20–161.20) | 1 | Observational Cohort | 25 | — | — | — | — | Single study |
| male | mixed | 50-60 | 170.70 | 135.22 – 206.18 | — (170.70–170.70) | 1 | Observational Cohort | 25 | — | — | — | — | Single study |
| male | mixed | 60+ | — | NA – NA | — | NA | NA | NA | — | — | — | — | NA |
| **male** | **mixed** | **Meta-Regression** | **—** | **NA – NA** | **—** | **NA** | **NA** | **NA** | **—** | **—** | **—** | **—** | **NA** |
| **male** | **mixed** | **Overall** | **158.05** | **86.77 – 229.33** | **158.00 ± 36.73 (86.10–211.73)** | **14** | **Mixed** | **620** | **99.6%** | **3516.8** | **<0.005** | **0.30** | **Random** |
| male | white | 18+ | — | NA – NA | — | NA | NA | NA | — | — | — | — | NA |
| male | white | 20-30 | — | NA – NA | — | NA | NA | NA | — | — | — | — | NA |
| male | white | 30-40 | — | NA – NA | — | NA | NA | NA | — | — | — | — | NA |
| male | white | 40-50 | 150.67 | 27.25 – 274.09 | — (150.67–150.67) | 1 | Observational Cohort | 6 | — | — | — | — | Single study |
| male | white | 50-60 | 178.18 | 18.32 – 338.04 | — (178.18–178.18) | 1 | Observational Cohort | 79 | — | — | — | — | Single study |
| male | white | 60+ | 169.33 | 97.42 – 241.23 | 171.03 ± 3.97 (167.70–175.43) | 3 | Observational Cohort | 329 | 8.7% | 2.2 | 0.33 | 0.55 | Fixed |
| **male** | **white** | **Meta-Regression** | **—** | **NA – NA** | **—** | **NA** | **NA** | **NA** | **—** | **—** | **—** | **—** | **NA** |
| **male** | **white** | **Overall** | **169.62** | **91.09 – 248.16** | **168.39 ± 10.75 (150.67–178.18)** | **5** | **Observational Cohort** | **414** | **0%** | **3.6** | **0.46** | **0.66** | **Fixed** |
| female | All | 18+ | 142.05 | 81.95 – 202.14 | 142.34 ± 31.66 (90.00–190.17) | 9 | Mixed | 540 | 93.9% | 132.1 | <0.005 | <0.05 | Random |
| female | All | 20-30 | 148.30 | 128.90 – 167.70 | — (148.30–148.30) | 1 | Observational Cohort | 25 | — | — | — | — | Single study |
| female | All | 30-40 | 150.20 | 122.56 – 177.84 | — (150.20–150.20) | 1 | Observational Cohort | 25 | — | — | — | — | Single study |
| female | All | 40-50 | 153.44 | 69.76 – 237.11 | 152.58 ± 1.89 (150.52–154.22) | 3 | Observational Cohort | 578 | 0% | 0.2 | 0.91 | 0.65 | Fixed |
| female | All | 50-60 | 137.26 | 38.43 – 236.10 | 141.18 ± 6.11 (136.30–148.03) | 3 | Observational Cohort | 2565 | 61.3% | 5.2 | 0.08 | <0.05 | Fixed |
| female | All | 60+ | 125.60 | 72.44 – 178.76 | 126.02 ± 27.01 (78.60–146.46) | 5 | Observational Cohort | 496 | 99% | 387 | <0.005 | <0.05 | Random |
| **female** | **All** | **Meta-Regression** | **190.17** | **178.07 – 202.27** | **—** | **7** | **Mixed** | **488** | **38.4%** | **6.4** | **0.17** | **<0.01** | **Multivariable** |
| **female** | **All** | **Overall** | **140.13** | **93.71 – 186.55** | **140.50 ± 24.58 (78.60–190.17)** | **22** | **Mixed** | **4229** | **98.7%** | **1661** | **<0.005** | **0.28** | **Random** |
| female | black | 18+ | — | NA – NA | — | NA | NA | NA | — | — | — | — | NA |
| female | black | 20-30 | — | NA – NA | — | NA | NA | NA | — | — | — | — | NA |
| female | black | 30-40 | — | NA – NA | — | NA | NA | NA | — | — | — | — | NA |
| female | black | 40-50 | 150.52 | 37.60 – 263.44 | — (150.52–150.52) | 1 | Observational Cohort | 29 | — | — | — | — | Single study |
| female | black | 50-60 | 148.03 | 22.20 – 273.86 | — (148.03–148.03) | 1 | Observational Cohort | 131 | — | — | — | — | Single study |
| female | black | 60+ | 142.55 | 35.40 – 249.70 | 141.83 ± 6.55 (137.20–146.46) | 2 | Observational Cohort | 144 | 0.8% | 1 | 0.32 | — | Fixed |
| **female** | **black** | **Meta-Regression** | **—** | **NA – NA** | **—** | **NA** | **NA** | **NA** | **—** | **—** | **—** | **—** | **NA** |
| **female** | **black** | **Overall** | **145.30** | **30.56 – 260.04** | **145.55 ± 5.81 (137.20–150.52)** | **4** | **Observational Cohort** | **304** | **0%** | **1.8** | **0.60** | **0.98** | **Fixed** |
| female | mixed | 18+ | 142.05 | 81.95 – 202.14 | 142.34 ± 31.66 (90.00–190.17) | 9 | Mixed | 540 | 93.9% | 132.1 | <0.005 | <0.05 | Random |
| female | mixed | 20-30 | 148.30 | 128.90 – 167.70 | — (148.30–148.30) | 1 | Observational Cohort | 25 | — | — | — | — | Single study |
| female | mixed | 30-40 | 150.20 | 122.56 – 177.84 | — (150.20–150.20) | 1 | Observational Cohort | 25 | — | — | — | — | Single study |
| female | mixed | 40-50 | 153.00 | 129.68 – 176.32 | — (153.00–153.00) | 1 | Observational Cohort | 25 | — | — | — | — | Single study |
| female | mixed | 50-60 | 139.20 | 117.64 – 160.76 | — (139.20–139.20) | 1 | Observational Cohort | 25 | — | — | — | — | Single study |
| female | mixed | 60+ | 78.60 | 63.70 – 93.50 | — (78.60–78.60) | 1 | Observational Cohort | 25 | — | — | — | — | Single study |
| **female** | **mixed** | **Meta-Regression** | **—** | **NA – NA** | **—** | **NA** | **NA** | **NA** | **—** | **—** | **—** | **—** | **NA** |
| **female** | **mixed** | **Overall** | **138.97** | **80.41 – 197.53** | **139.31 ± 30.60 (78.60–190.17)** | **14** | **Mixed** | **665** | **99.1%** | **1481.5** | **<0.005** | **0.23** | **Random** |
| female | white | 18+ | — | NA – NA | — | NA | NA | NA | — | — | — | — | NA |
| female | white | 20-30 | — | NA – NA | — | NA | NA | NA | — | — | — | — | NA |
| female | white | 30-40 | — | NA – NA | — | NA | NA | NA | — | — | — | — | NA |
| female | white | 40-50 | 154.22 | 30.66 – 277.78 | — (154.22–154.22) | 1 | Observational Cohort | 524 | — | — | — | — | Single study |
| female | white | 50-60 | 136.30 | 26.70 – 245.90 | — (136.30–136.30) | 1 | Observational Cohort | 2409 | — | — | — | — | Single study |
| female | white | 60+ | 133.55 | 25.93 – 241.17 | 133.93 ± 1.12 (133.13–134.72) | 2 | Observational Cohort | 327 | 0% | 0.1 | 0.82 | — | Fixed |
| **female** | **white** | **Meta-Regression** | **—** | **NA – NA** | **—** | **NA** | **NA** | **NA** | **—** | **—** | **—** | **—** | **NA** |
| **female** | **white** | **Overall** | **139.89** | **121.28 – 158.51** | **139.59 ± 9.84 (133.13–154.22)** | **4** | **Observational Cohort** | **3260** | **92.3%** | **39** | **<0.005** | **0.72** | **Random** |
| *I² represents heterogeneity; Q test assesses between-study variation; Egger's p = linear regression test for funnel plot asymmetry; Meta-Regression shows covariate-adjusted estimates* | | | | | | | | | | | | | |

###### 6.9.2.3 Table of Results subdivided by race / Sex / Age

sodium_ma_3rd_analysis$race_summary_table

Table 1: **race-Stratified Meta-Analysis Results**

24-Hour Urine Sodium (mmol/24h)

| Sex Group | race | Age Band | 95% Range | Studies | Study Type | Participants | I² | Cochran's Q | Q (p-value) | Egger's p | Model |
| --- | --- | --- | --- | --- | --- | --- | --- | --- | --- | --- | --- |
| **main meta-analysis** | **All races** | **18+** | **65.90 – 235.89** | **66** | **Mixed** | **4660** | **97.2%** | **2335.4** | **<0.005** | **0.07** | **Random** |
| **main meta-analysis** | **All races** | **20-30** | **88.79 – 225.21** | **5** | **Mixed** | **149** | **98.7%** | **303** | **<0.005** | **0.75** | **Random** |
| **main meta-analysis** | **All races** | **30-40** | **110.21 – 214.91** | **4** | **Observational Cohort** | **174** | **99%** | **303.8** | **<0.005** | **0.37** | **Random** |
| **main meta-analysis** | **All races** | **40-50** | **102.97 – 212.85** | **6** | **Observational Cohort** | **643** | **53.5%** | **10.7** | **0.06** | **0.48** | **Fixed** |
| **main meta-analysis** | **All races** | **50-60** | **118.66 – 188.26** | **5** | **Observational Cohort** | **2669** | **96.1%** | **102** | **<0.005** | **0.12** | **Random** |
| **main meta-analysis** | **All races** | **60+** | **81.75 – 203.66** | **8** | **Observational Cohort** | **825** | **99.5%** | **1371.2** | **<0.005** | **0.11** | **Random** |
| **main meta-analysis** | **All races** | **Meta-Regression** | **98.06 – 253.33** | **47** | **Mixed** | **3945** | **95.4%** | **857.4** | **<0.005** | **<0.05** | **Multivariable** |
| **main meta-analysis** | **All races** | **Meta-Regression** | **98.06 – 253.33** | **47** | **Mixed** | **3945** | **95.4%** | **857.4** | **<0.005** | **<0.05** | **Multivariable** |
| **main meta-analysis** | **All races** | **Overall** | **76.94 – 225.12** | **94** | **Mixed** | **9120** | **98.8%** | **7915.6** | **<0.005** | **0.22** | **Random** |
| **main meta-analysis** | **All races** | **Overall** | **76.94 – 225.12** | **94** | **Mixed** | **9120** | **98.8%** | **7915.6** | **<0.005** | **0.22** | **Random** |
| main meta-analysis | Black | 18+ | -17.80 – 491.80 | 3 | Observational Cohort | 51 | 0% | 0 | 1.00 | — | Fixed |
| main meta-analysis | Black | 20-30 | — | — | NA | — | — | — | — | — | — |
| main meta-analysis | Black | 30-40 | — | — | NA | — | — | — | — | — | — |
| main meta-analysis | Black | 40-50 | 37.60 – 263.44 | 1 | Observational Cohort | 29 | — | — | — | — | Single study |
| main meta-analysis | Black | 50-60 | 22.20 – 273.86 | 1 | Observational Cohort | 131 | — | — | — | — | Single study |
| main meta-analysis | Black | 60+ | 35.40 – 249.70 | 2 | Observational Cohort | 144 | 0.8% | 1 | 0.32 | — | Fixed |
| main meta-analysis | Black | Meta-Regression | — | — | NA | — | — | — | — | — | — |
| **main meta-analysis** | **Black** | **Overall** | **96.95 – 253.25** | **7** | **Observational Cohort** | **355** | **77.3%** | **26.4** | **<0.005** | **<0.01** | **Random** |
| main meta-analysis | Mixed | 18+ | 64.68 – 225.80 | 55 | Mixed | 4241 | 96.2% | 1427.4 | <0.005 | 0.08 | Random |
| main meta-analysis | Mixed | 20-30 | 88.79 – 225.21 | 5 | Mixed | 149 | 98.7% | 303 | <0.005 | 0.75 | Random |
| main meta-analysis | Mixed | 30-40 | 110.21 – 214.91 | 4 | Observational Cohort | 174 | 99% | 303.8 | <0.005 | 0.37 | Random |
| main meta-analysis | Mixed | 40-50 | 150.73 – 165.94 | 3 | Observational Cohort | 84 | 74.8% | 7.9 | <0.05 | 0.74 | Random |
| main meta-analysis | Mixed | 50-60 | 111.56 – 198.08 | 2 | Observational Cohort | 50 | 98.2% | 55.3 | <0.005 | — | Random |
| main meta-analysis | Mixed | 60+ | 63.70 – 93.50 | 1 | Observational Cohort | 25 | — | — | — | — | Single study |
| main meta-analysis | Mixed | Meta-Regression | — | — | NA | — | — | — | — | — | — |
| **main meta-analysis** | **Mixed** | **Overall** | **70.74 – 223.21** | **70** | **Mixed** | **4723** | **98.9%** | **6531.2** | **<0.005** | **0.42** | **Random** |
| main meta-analysis | White | 18+ | 68.01 – 275.06 | 7 | Observational Cohort | 328 | 98% | 301.1 | <0.005 | <0.05 | Random |
| main meta-analysis | White | 20-30 | — | — | NA | — | — | — | — | — | — |
| main meta-analysis | White | 30-40 | — | — | NA | — | — | — | — | — | — |
| main meta-analysis | White | 40-50 | 30.62 – 277.74 | 2 | Observational Cohort | 530 | 0% | 0 | 0.89 | — | Fixed |
| main meta-analysis | White | 50-60 | 99.64 – 212.86 | 2 | Observational Cohort | 2488 | 95.1% | 20.5 | <0.005 | — | Random |
| main meta-analysis | White | 60+ | 117.00 – 195.24 | 5 | Observational Cohort | 656 | 95.9% | 98.4 | <0.005 | 0.72 | Random |
| main meta-analysis | White | Meta-Regression | — | — | NA | — | — | — | — | — | — |
| **main meta-analysis** | **White** | **Overall** | **94.09 – 223.74** | **16** | **Observational Cohort** | **4002** | **98.9%** | **1305.8** | **<0.005** | **0.34** | **Random** |
| **male** | **All races** | **18+** | **55.25 – 224.45** | **7** | **Observational Cohort** | **367** | **98.1%** | **315.8** | **<0.005** | **0.90** | **Random** |
| **male** | **All races** | **20-30** | **133.63 – 221.80** | **3** | **Mixed** | **71** | **86%** | **14.3** | **<0.005** | **0.57** | **Random** |
| **male** | **All races** | **30-40** | **169.55 – 197.99** | **2** | **Observational Cohort** | **132** | **84%** | **6.2** | **<0.05** | **—** | **Random** |
| **male** | **All races** | **40-50** | **132.65 – 189.54** | **2** | **Observational Cohort** | **31** | **0%** | **0.2** | **0.68** | **—** | **Fixed** |
| **male** | **All races** | **50-60** | **104.40 – 239.02** | **2** | **Observational Cohort** | **104** | **0%** | **0.6** | **0.45** | **—** | **Fixed** |
| **male** | **All races** | **60+** | **97.42 – 241.23** | **3** | **Observational Cohort** | **329** | **8.7%** | **2.2** | **0.33** | **0.55** | **Fixed** |
| **male** | **All races** | **Meta-Regression** | **163.04 – 260.42** | **4** | **Mixed** | **262** | **95.5%** | **36.3** | **<0.005** | **0.65** | **Univariable** |
| **male** | **All races** | **Overall** | **98.71 – 223.25** | **19** | **Mixed** | **1034** | **99.5%** | **3691.7** | **<0.005** | **0.17** | **Random** |
| male | Black | 18+ | — | — | NA | — | — | — | — | — | — |
| male | Black | 20-30 | — | — | NA | — | — | — | — | — | — |
| male | Black | 30-40 | — | — | NA | — | — | — | — | — | — |
| male | Black | 40-50 | — | — | NA | — | — | — | — | — | — |
| male | Black | 50-60 | — | — | NA | — | — | — | — | — | — |
| male | Black | 60+ | — | — | NA | — | — | — | — | — | — |
| male | Black | Meta-Regression | — | — | NA | — | — | — | — | — | — |
| **male** | **Black** | **Overall** | **—** | **—** | **NA** | **—** | **—** | **—** | **—** | **—** | **—** |
| male | Mixed | 18+ | 55.25 – 224.45 | 7 | Observational Cohort | 367 | 98.1% | 315.8 | <0.005 | 0.90 | Random |
| male | Mixed | 20-30 | 133.63 – 221.80 | 3 | Mixed | 71 | 86% | 14.3 | <0.005 | 0.57 | Random |
| male | Mixed | 30-40 | 169.55 – 197.99 | 2 | Observational Cohort | 132 | 84% | 6.2 | <0.05 | — | Random |
| male | Mixed | 40-50 | 135.52 – 186.88 | 1 | Observational Cohort | 25 | — | — | — | — | Single study |
| male | Mixed | 50-60 | 135.22 – 206.18 | 1 | Observational Cohort | 25 | — | — | — | — | Single study |
| male | Mixed | 60+ | — | — | NA | — | — | — | — | — | — |
| male | Mixed | Meta-Regression | — | — | NA | — | — | — | — | — | — |
| **male** | **Mixed** | **Overall** | **86.77 – 229.33** | **14** | **Mixed** | **620** | **99.6%** | **3516.8** | **<0.005** | **0.30** | **Random** |
| male | White | 18+ | — | — | NA | — | — | — | — | — | — |
| male | White | 20-30 | — | — | NA | — | — | — | — | — | — |
| male | White | 30-40 | — | — | NA | — | — | — | — | — | — |
| male | White | 40-50 | 27.25 – 274.09 | 1 | Observational Cohort | 6 | — | — | — | — | Single study |
| male | White | 50-60 | 18.32 – 338.04 | 1 | Observational Cohort | 79 | — | — | — | — | Single study |
| male | White | 60+ | 97.42 – 241.23 | 3 | Observational Cohort | 329 | 8.7% | 2.2 | 0.33 | 0.55 | Fixed |
| male | White | Meta-Regression | — | — | NA | — | — | — | — | — | — |
| **male** | **White** | **Overall** | **91.09 – 248.16** | **5** | **Observational Cohort** | **414** | **0%** | **3.6** | **0.46** | **0.66** | **Fixed** |
| **female** | **All races** | **18+** | **81.95 – 202.14** | **9** | **Mixed** | **540** | **93.9%** | **132.1** | **<0.005** | **<0.05** | **Random** |
| **female** | **All races** | **20-30** | **128.90 – 167.70** | **1** | **Observational Cohort** | **25** | **—** | **—** | **—** | **—** | **Single study** |
| **female** | **All races** | **30-40** | **122.56 – 177.84** | **1** | **Observational Cohort** | **25** | **—** | **—** | **—** | **—** | **Single study** |
| **female** | **All races** | **40-50** | **69.76 – 237.11** | **3** | **Observational Cohort** | **578** | **0%** | **0.2** | **0.91** | **0.65** | **Fixed** |
| **female** | **All races** | **50-60** | **38.43 – 236.10** | **3** | **Observational Cohort** | **2565** | **61.3%** | **5.2** | **0.08** | **<0.05** | **Fixed** |
| **female** | **All races** | **60+** | **72.44 – 178.76** | **5** | **Observational Cohort** | **496** | **99%** | **387** | **<0.005** | **<0.05** | **Random** |
| **female** | **All races** | **Meta-Regression** | **178.07 – 202.27** | **7** | **Mixed** | **488** | **38.4%** | **6.4** | **0.17** | **<0.01** | **Multivariable** |
| **female** | **All races** | **Overall** | **93.71 – 186.55** | **22** | **Mixed** | **4229** | **98.7%** | **1661** | **<0.005** | **0.28** | **Random** |
| female | Black | 18+ | — | — | NA | — | — | — | — | — | — |
| female | Black | 20-30 | — | — | NA | — | — | — | — | — | — |
| female | Black | 30-40 | — | — | NA | — | — | — | — | — | — |
| female | Black | 40-50 | 37.60 – 263.44 | 1 | Observational Cohort | 29 | — | — | — | — | Single study |
| female | Black | 50-60 | 22.20 – 273.86 | 1 | Observational Cohort | 131 | — | — | — | — | Single study |
| female | Black | 60+ | 35.40 – 249.70 | 2 | Observational Cohort | 144 | 0.8% | 1 | 0.32 | — | Fixed |
| female | Black | Meta-Regression | — | — | NA | — | — | — | — | — | — |
| **female** | **Black** | **Overall** | **30.56 – 260.04** | **4** | **Observational Cohort** | **304** | **0%** | **1.8** | **0.60** | **0.98** | **Fixed** |
| female | Mixed | 18+ | 81.95 – 202.14 | 9 | Mixed | 540 | 93.9% | 132.1 | <0.005 | <0.05 | Random |
| female | Mixed | 20-30 | 128.90 – 167.70 | 1 | Observational Cohort | 25 | — | — | — | — | Single study |
| female | Mixed | 30-40 | 122.56 – 177.84 | 1 | Observational Cohort | 25 | — | — | — | — | Single study |
| female | Mixed | 40-50 | 129.68 – 176.32 | 1 | Observational Cohort | 25 | — | — | — | — | Single study |
| female | Mixed | 50-60 | 117.64 – 160.76 | 1 | Observational Cohort | 25 | — | — | — | — | Single study |
| female | Mixed | 60+ | 63.70 – 93.50 | 1 | Observational Cohort | 25 | — | — | — | — | Single study |
| female | Mixed | Meta-Regression | — | — | NA | — | — | — | — | — | — |
| **female** | **Mixed** | **Overall** | **80.41 – 197.53** | **14** | **Mixed** | **665** | **99.1%** | **1481.5** | **<0.005** | **0.23** | **Random** |
| female | White | 18+ | — | — | NA | — | — | — | — | — | — |
| female | White | 20-30 | — | — | NA | — | — | — | — | — | — |
| female | White | 30-40 | — | — | NA | — | — | — | — | — | — |
| female | White | 40-50 | 30.66 – 277.78 | 1 | Observational Cohort | 524 | — | — | — | — | Single study |
| female | White | 50-60 | 26.70 – 245.90 | 1 | Observational Cohort | 2409 | — | — | — | — | Single study |
| female | White | 60+ | 25.93 – 241.17 | 2 | Observational Cohort | 327 | 0% | 0.1 | 0.82 | — | Fixed |
| female | White | Meta-Regression | — | — | NA | — | — | — | — | — | — |
| **female** | **White** | **Overall** | **121.28 – 158.51** | **4** | **Observational Cohort** | **3260** | **92.3%** | **39** | **<0.005** | **0.72** | **Random** |
| *I² represents heterogeneity; Q test assesses between-study variation; Egger's p = linear regression test for funnel plot asymmetry; — indicates insufficient data* | | | | | | | | | | | |

###### 6.9.2.4 Wald Tests

sodium_ma_3rd_analysis$wald_tests$sex_comparison

Table 1: **Sex Comparison (Overall)**

| Comparison | % Shift | P (Wald Test) | Mean Diff (95% CI) |
| --- | --- | --- | --- |
| Male vs Female (Overall) | +14.9% | 0.599 | 20.85 [-56.82, 98.52] |
| *Mean difference = Group 1 − Group 2 in component units (mmol/24h, L/24h, or pH); % shift = difference relative to Group 2.* | | | |

sodium_ma_3rd_analysis$wald_tests$race_comparisons

Table 1: **race Comparisons Within Sex Groups**

| Sex Group | Comparison | % Shift | P (Wald Test) | Mean Diff (95% CI) |
| --- | --- | --- | --- | --- |
| male | black vs mixed | — | — | — |
| male | black vs white | — | — | — |
| male | mixed vs white | -6.8% | 0.831 | -11.57 [-117.64, 94.49] |
| female | black vs mixed | +4.6% | 0.923 | 6.33 [-122.49, 135.15] |
| female | black vs white | +3.9% | 0.927 | 5.41 [-110.84, 121.65] |
| female | mixed vs white | -0.7% | 0.976 | -0.92 [-62.37, 60.52] |
| *Mean difference = Group 1 − Group 2 in component units (mmol/24h, L/24h, or pH); % shift = difference relative to Group 2.* | | | | |

sodium_ma_3rd_analysis$wald_tests$age_band_comparisons

Table 1: **Age Band Comparisons Within Sex Groups**

| Sex Group | Comparison | % Shift | P (Wald Test) | Mean Diff (95% CI) |
| --- | --- | --- | --- | --- |
| main meta-analysis | 18+ vs 20-30 | -3.9% | 0.913 | -6.10 [-115.08, 102.88] |
| main meta-analysis | 18+ vs 30-40 | -7.2% | 0.819 | -11.66 [-111.48, 88.16] |
| main meta-analysis | 18+ vs 40-50 | -4.4% | 0.892 | -7.02 [-108.22, 94.19] |
| main meta-analysis | 18+ vs 50-60 | -1.7% | 0.956 | -2.56 [-94.41, 89.28] |
| main meta-analysis | 18+ vs 60+ | +5.7% | 0.878 | 8.19 [-96.40, 112.79] |
| main meta-analysis | 20-30 vs 30-40 | -3.4% | 0.899 | -5.56 [-91.54, 80.43] |
| main meta-analysis | 20-30 vs 40-50 | -0.6% | 0.984 | -0.91 [-88.50, 86.67] |
| main meta-analysis | 20-30 vs 50-60 | +2.3% | 0.928 | 3.54 [-73.03, 80.12] |
| main meta-analysis | 20-30 vs 60+ | +10.0% | 0.759 | 14.30 [-77.18, 105.77] |
| main meta-analysis | 30-40 vs 40-50 | +2.9% | 0.905 | 4.64 [-71.25, 80.53] |
| main meta-analysis | 30-40 vs 50-60 | +5.9% | 0.777 | 9.10 [-53.76, 71.96] |
| main meta-analysis | 30-40 vs 60+ | +13.9% | 0.628 | 19.85 [-60.50, 100.20] |
| main meta-analysis | 40-50 vs 50-60 | +2.9% | 0.893 | 4.46 [-60.58, 69.49] |
| main meta-analysis | 40-50 vs 60+ | +10.7% | 0.716 | 15.21 [-66.85, 97.27] |
| main meta-analysis | 50-60 vs 60+ | +7.5% | 0.764 | 10.75 [-59.44, 80.95] |
| male | 18+ vs 20-30 | -21.3% | 0.437 | -37.87 [-133.27, 57.53] |
| male | 18+ vs 30-40 | -23.9% | 0.316 | -43.92 [-129.71, 41.87] |
| male | 18+ vs 40-50 | -13.2% | 0.641 | -21.24 [-110.50, 68.01] |
| male | 18+ vs 50-60 | -18.6% | 0.564 | -31.86 [-139.97, 76.25] |
| male | 18+ vs 60+ | -17.4% | 0.603 | -29.48 [-140.51, 81.55] |
| male | 20-30 vs 30-40 | -3.3% | 0.798 | -6.05 [-52.37, 40.27] |
| male | 20-30 vs 40-50 | +10.3% | 0.534 | 16.63 [-35.84, 69.09] |
| male | 20-30 vs 50-60 | +3.5% | 0.884 | 6.01 [-74.45, 86.47] |
| male | 20-30 vs 60+ | +5.0% | 0.845 | 8.39 [-75.95, 92.74] |
| male | 30-40 vs 40-50 | +14.1% | 0.162 | 22.68 [-9.12, 54.48] |
| male | 30-40 vs 50-60 | +7.0% | 0.731 | 12.06 [-56.73, 80.86] |
| male | 30-40 vs 60+ | +8.5% | 0.699 | 14.44 [-58.85, 87.74] |
| male | 40-50 vs 50-60 | -6.2% | 0.776 | -10.62 [-83.69, 62.46] |
| male | 40-50 vs 60+ | -4.9% | 0.835 | -8.23 [-85.56, 69.09] |
| male | 50-60 vs 60+ | +1.4% | 0.962 | 2.38 [-96.11, 100.87] |
| female | 18+ vs 20-30 | -4.2% | 0.846 | -6.25 [-69.40, 56.89] |
| female | 18+ vs 30-40 | -5.4% | 0.809 | -8.15 [-74.30, 57.99] |
| female | 18+ vs 40-50 | -7.4% | 0.828 | -11.39 [-114.41, 91.62] |
| female | 18+ vs 50-60 | +3.5% | 0.935 | 4.78 [-110.89, 120.45] |
| female | 18+ vs 60+ | +13.1% | 0.688 | 16.44 [-63.79, 96.67] |
| female | 20-30 vs 30-40 | -1.3% | 0.912 | -1.90 [-35.67, 31.87] |
| female | 20-30 vs 40-50 | -3.3% | 0.907 | -5.14 [-91.03, 80.76] |
| female | 20-30 vs 50-60 | +8.0% | 0.830 | 11.04 [-89.68, 111.76] |
| female | 20-30 vs 60+ | +18.1% | 0.432 | 22.70 [-33.90, 79.29] |
| female | 30-40 vs 40-50 | -2.1% | 0.943 | -3.24 [-91.36, 84.88] |
| female | 30-40 vs 50-60 | +9.4% | 0.805 | 12.94 [-89.69, 115.56] |
| female | 30-40 vs 60+ | +19.6% | 0.421 | 24.60 [-35.32, 84.51] |
| female | 40-50 vs 50-60 | +11.8% | 0.807 | 16.18 [-113.32, 145.67] |
| female | 40-50 vs 60+ | +22.2% | 0.582 | 27.83 [-71.30, 126.97] |
| female | 50-60 vs 60+ | +9.3% | 0.839 | 11.66 [-100.56, 123.88] |
| *Mean difference = Group 1 − Group 2 in component units (mmol/24h, L/24h, or pH); % shift = difference relative to Group 2.* | | | | |

sodium_ma_3rd_analysis$wald_tests$sex_within_age_bands

Table 1: **Sex Comparisons Within Age Bands**

| Age Band | % Shift | P (Wald Test) | Mean Diff (95% CI) |
| --- | --- | --- | --- |
| 18+ | -1.5% | 0.967 | -2.20 [-105.97, 101.57] |
| 20-30 | +19.8% | 0.231 | 29.42 [-18.75, 77.59] |
| 30-40 | +22.3% | <0.05 | 33.57 [2.49, 64.65] |
| 40-50 | +5.0% | 0.865 | 7.65 [-80.73, 96.03] |
| 50-60 | +25.1% | 0.572 | 34.44 [-85.13, 154.02] |
| 60+ | +34.8% | 0.338 | 43.72 [-45.70, 133.14] |
| *Mean difference = Group 1 − Group 2 in component units (mmol/24h, L/24h, or pH); % shift = difference relative to Group 2.* | | | |

#### 6.10 Potassium

##### 6.10.1 Comparison by Age Band

###### 6.10.1.1 Plot

```
potassium_ma_2nd_analysis <- sex_age_subdivision_ma("potassium")
## Not enough studies for multivariable meta-regression (k = 3, need > 6). Running univariable models instead.
potassium_ma_2nd_analysis$combined_plot
## `height` was translated to `width`.
## `height` was translated to `width`.
## `height` was translated to `width`.
```

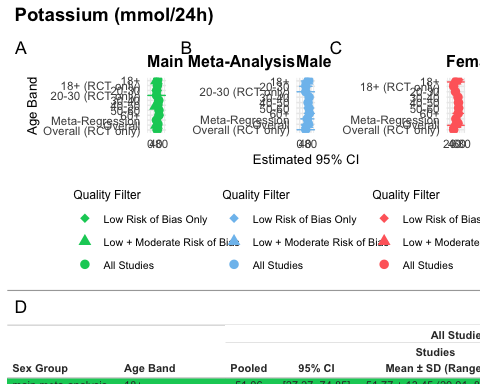

###### 6.10.1.2 Summary Table

potassium_ma_2nd_analysis$combined_summary_table

Table 1: **Combined Meta-Analysis Results**

24-Hour Urine Potassium (mmol/24h)

|  |  | **All Studies** | | | | | | **Low Risk of Bias Only** | | | | | | **Low + Moderate RoB** | | | | | |
| --- | --- | --- | --- | --- | --- | --- | --- | --- | --- | --- | --- | --- | --- | --- | --- | --- | --- | --- | --- |
| Sex Group | Age Band | Pooled | 95% CI | Studies  Mean ± SD (Range) | Studies, n | Q p-value | Egger's p | Pooled | 95% CI | Studies  Mean ± SD (Range) | Studies, n | Q p-value | Egger's p | Pooled | 95% CI | Studies  Mean ± SD (Range) | Studies, n | Q p-value | Egger's p |
| main meta-analysis | 18+ | 51.06 | [27.27, 74.85] | 51.77 ± 13.45 (29.91–86.00) | 66 | <0.001 | 0.05 | 42.93 | [34.79, 51.07] | 42.67 ± 4.99 (32.00–53.90) | 27 | <0.001 | 0.68 | 42.63 | [32.96, 52.30] | 43.25 ± 7.95 (32.00–76.22) | 31 | <0.001 | 0.54 |
| **main meta-analysis** | **18+ (RCT only)** | **48.78** | **[27.85, 69.71]** | **50.27 ± 13.38 (36.00–76.22)** | **7** | **<0.001** | **<0.01** | **—** | **—** | **—** | **—** | **—** | **—** | **—** | **—** | **—** | **—** | **—** | **—** |
| main meta-analysis | 20-30 | 43.97 | [13.16, 74.78] | 44.01 ± 15.77 (21.00–65.50) | 5 | <0.001 | 0.67 | — | — | — | — | — | — | — | — | — | — | — | — |
| **main meta-analysis** | **20-30 (RCT only)** | **43.23** | **[-18.37, 104.83]** | **43.25 ± 31.47 (21.00–65.50)** | **2** | **<0.001** | **—** | **—** | **—** | **—** | **—** | **—** | **—** | **—** | **—** | **—** | **—** | **—** | **—** |
| main meta-analysis | 30-40 | 58.86 | [45.04, 72.67] | 59.00 ± 7.31 (53.66–67.33) | 3 | <0.001 | 0.28 | — | — | — | — | — | — | — | — | — | — | — | — |
| main meta-analysis | 40-50 | 53.04 | [34.38, 71.70] | 53.84 ± 9.53 (36.10–61.50) | 6 | <0.001 | 0.24 | — | — | — | — | — | — | 36.10 | [31.59, 40.61] | — (36.10–36.10) | 1 | — | — |
| main meta-analysis | 50-60 | 59.95 | [38.46, 81.43] | 60.19 ± 11.41 (49.33–77.76) | 5 | <0.001 | 0.53 | — | — | — | — | — | — | — | — | — | — | — | — |
| main meta-analysis | 60+ | 60.02 | [31.86, 88.18] | 60.13 ± 14.47 (37.07–79.80) | 8 | <0.001 | <0.01 | — | — | — | — | — | — | — | — | — | — | — | — |
| **main meta-analysis** | **Meta-Regression** | **42.40** | **[25.05, 59.75]** | **—** | **45** | **<0.001** | **<0.005** | **—** | **—** | **—** | **—** | **—** | **—** | **—** | **—** | **—** | **—** | **—** | **—** |
| **main meta-analysis** | **Overall** | **52.41** | **[27.72, 77.11]** | **52.89 ± 13.43 (21.00–86.00)** | **93** | **<0.001** | **0.74** | **42.93** | **[34.79, 51.07]** | **42.67 ± 4.99 (32.00–53.90)** | **27** | **<0.001** | **0.68** | **42.38** | **[32.53, 52.22]** | **43.03 ± 7.93 (32.00–76.22)** | **32** | **<0.001** | **<0.01** |
| **main meta-analysis** | **Overall (RCT only)** | **47.95** | **[17.67, 78.24]** | **48.71 ± 16.36 (21.00–76.22)** | **9** | **<0.001** | **0.15** | **—** | **—** | **—** | **—** | **—** | **—** | **—** | **—** | **—** | **—** | **—** | **—** |
| male | 18+ | 62.35 | [31.90, 92.81] | 63.26 ± 16.64 (41.00–83.68) | 7 | <0.001 | 0.31 | 53.90 | [11.58, 96.22] | — (53.90–53.90) | 1 | — | — | 53.90 | [11.58, 96.22] | — (53.90–53.90) | 1 | — | — |
| **male** | **18+ (RCT only)** | **—** | **—** | **—** | **—** | **—** | **—** | **—** | **—** | **—** | **—** | **—** | **—** | **—** | **—** | **—** | **—** | **—** | **—** |
| male | 20-30 | 43.90 | [0.34, 87.46] | 43.93 ± 22.28 (21.00–65.50) | 3 | <0.001 | 0.72 | — | — | — | — | — | — | — | — | — | — | — | — |
| **male** | **20-30 (RCT only)** | **43.23** | **[-18.37, 104.83]** | **43.25 ± 31.47 (21.00–65.50)** | **2** | **<0.001** | **—** | **—** | **—** | **—** | **—** | **—** | **—** | **—** | **—** | **—** | **—** | **—** | **—** |
| male | 30-40 | 67.33 | [48.32, 86.34] | — (67.33–67.33) | 1 | — | — | — | — | — | — | — | — | — | — | — | — | — | — |
| male | 40-50 | 60.04 | [48.40, 71.69] | 60.77 ± 1.04 (60.03–61.50) | 2 | 0.89 | — | — | — | — | — | — | — | — | — | — | — | — | — |
| male | 50-60 | 65.85 | [33.71, 97.98] | 66.05 ± 16.56 (54.34–77.76) | 2 | <0.001 | — | — | — | — | — | — | — | — | — | — | — | — | — |
| male | 60+ | 69.05 | [34.75, 103.35] | 69.16 ± 17.52 (48.94–79.80) | 3 | <0.001 | 0.24 | — | — | — | — | — | — | — | — | — | — | — | — |
| **male** | **Meta-Regression** | **53.90** | **[8.71, 99.09]** | **—** | **3** | **<0.001** | **0.28** | **—** | **—** | **—** | **—** | **—** | **—** | **—** | **—** | **—** | **—** | **—** | **—** |
| **male** | **Overall** | **60.85** | **[28.71, 92.99]** | **61.28 ± 16.70 (21.00–83.68)** | **18** | **<0.001** | **0.53** | **53.90** | **[11.58, 96.22]** | **— (53.90–53.90)** | **1** | **—** | **—** | **53.90** | **[11.58, 96.22]** | **— (53.90–53.90)** | **1** | **—** | **—** |
| **male** | **Overall (RCT only)** | **43.23** | **[-18.37, 104.83]** | **43.25 ± 31.47 (21.00–65.50)** | **2** | **<0.001** | **—** | **—** | **—** | **—** | **—** | **—** | **—** | **—** | **—** | **—** | **—** | **—** | **—** |
| female | 18+ | 52.77 | [30.13, 75.41] | 53.53 ± 12.48 (36.00–71.10) | 9 | <0.001 | <0.005 | 44.79 | [8.90, 80.68] | 44.79 ± 0.00 (44.79–44.79) | 2 | 1.00 | — | 41.86 | [32.32, 51.40] | 41.86 ± 5.07 (36.00–44.79) | 3 | <0.001 | <0.001 |
| **female** | **18+ (RCT only)** | **36.00** | **[26.20, 45.80]** | **— (36.00–36.00)** | **1** | **—** | **—** | **—** | **—** | **—** | **—** | **—** | **—** | **—** | **—** | **—** | **—** | **—** | **—** |
| female | 20-30 | 43.26 | [33.46, 53.06] | — (43.26–43.26) | 1 | — | — | — | — | — | — | — | — | — | — | — | — | — | — |
| **female** | **20-30 (RCT only)** | **—** | **—** | **—** | **—** | **—** | **—** | **—** | **—** | **—** | **—** | **—** | **—** | **—** | **—** | **—** | **—** | **—** | **—** |
| female | 30-40 | 53.66 | [42.98, 64.34] | — (53.66–53.66) | 1 | — | — | — | — | — | — | — | — | — | — | — | — | — | — |
| female | 40-50 | 55.67 | [48.15, 63.19] | 55.13 ± 4.33 (51.24–59.80) | 3 | <0.001 | 0.85 | — | — | — | — | — | — | — | — | — | — | — | — |
| female | 50-60 | 56.32 | [40.40, 72.25] | 56.28 ± 8.11 (49.33–65.19) | 3 | <0.001 | 0.43 | — | — | — | — | — | — | — | — | — | — | — | — |
| female | 60+ | 54.61 | [33.54, 75.68] | 54.71 ± 10.71 (37.07–64.05) | 5 | <0.001 | <0.05 | — | — | — | — | — | — | — | — | — | — | — | — |
| **female** | **Meta-Regression** | **44.79** | **[33.24, 56.34]** | **—** | **7** | **<0.001** | **<0.05** | **—** | **—** | **—** | **—** | **—** | **—** | **—** | **—** | **—** | **—** | **—** | **—** |
| **female** | **Overall** | **53.49** | **[35.51, 71.47]** | **53.93 ± 9.79 (36.00–71.10)** | **22** | **<0.001** | **0.91** | **44.79** | **[8.90, 80.68]** | **44.79 ± 0.00 (44.79–44.79)** | **2** | **1.00** | **—** | **41.86** | **[32.32, 51.40]** | **41.86 ± 5.07 (36.00–44.79)** | **3** | **<0.001** | **<0.001** |
| **female** | **Overall (RCT only)** | **36.00** | **[26.20, 45.80]** | **— (36.00–36.00)** | **1** | **—** | **—** | **—** | **—** | **—** | **—** | **—** | **—** | **—** | **—** | **—** | **—** | **—** | **—** |
| *Studies, n = number of studies; Q p-value = Cochran's Q heterogeneity test; Egger's p = linear regression test for funnel plot asymmetry; RoB = Risk of Bias* | | | | | | | | | | | | | | | | | | | |

###### 6.10.1.3 Wald Tests

potassium_ma_2nd_analysis$wald_tests$sex_comparison

Table 1: **Sex Comparison (Overall)**

| Comparison | % Shift | P (Wald Test) | Mean Diff (95% CI) |
| --- | --- | --- | --- |
| Male vs Female (Overall) | +13.8% | 0.695 | 7.36 [-29.47, 44.18] |
| *Mean difference = Group 1 − Group 2 in component units (mmol/24h, L/24h, or pH); % shift = difference relative to Group 2.* | | | |

potassium_ma_2nd_analysis$wald_tests$age_band_comparisons

Table 1: **Age Band Comparisons Within Sex Groups**

| Sex Group | Comparison | % Shift | P (Wald Test) | Mean Diff (95% CI) |
| --- | --- | --- | --- | --- |
| main meta-analysis | 18+ vs 20-30 | +16.1% | 0.721 | 7.09 [-31.84, 46.02] |
| main meta-analysis | 18+ vs 30-40 | -13.2% | 0.579 | -7.79 [-35.30, 19.72] |
| main meta-analysis | 18+ vs 40-50 | -3.7% | 0.898 | -1.98 [-32.21, 28.26] |
| main meta-analysis | 18+ vs 50-60 | -14.8% | 0.587 | -8.89 [-40.94, 23.17] |
| main meta-analysis | 18+ vs 60+ | -14.9% | 0.634 | -8.96 [-45.82, 27.91] |
| main meta-analysis | 20-30 vs 30-40 | -25.3% | 0.388 | -14.88 [-48.66, 18.89] |
| main meta-analysis | 20-30 vs 40-50 | -17.1% | 0.622 | -9.07 [-45.09, 26.95] |
| main meta-analysis | 20-30 vs 50-60 | -26.7% | 0.404 | -15.98 [-53.54, 21.59] |
| main meta-analysis | 20-30 vs 60+ | -26.7% | 0.451 | -16.05 [-57.79, 25.69] |
| main meta-analysis | 30-40 vs 40-50 | +11.0% | 0.623 | 5.82 [-17.40, 29.03] |
| main meta-analysis | 30-40 vs 50-60 | -1.8% | 0.933 | -1.09 [-26.64, 24.45] |
| main meta-analysis | 30-40 vs 60+ | -1.9% | 0.942 | -1.17 [-32.53, 30.20] |
| main meta-analysis | 40-50 vs 50-60 | -11.5% | 0.634 | -6.91 [-35.36, 21.55] |
| main meta-analysis | 40-50 vs 60+ | -11.6% | 0.685 | -6.98 [-40.76, 26.80] |
| main meta-analysis | 50-60 vs 60+ | -0.1% | 0.997 | -0.07 [-35.49, 35.35] |
| male | 18+ vs 20-30 | +42.0% | 0.496 | 18.45 [-34.70, 71.60] |
| male | 18+ vs 30-40 | -7.4% | 0.786 | -4.98 [-40.88, 30.93] |
| male | 18+ vs 40-50 | +3.8% | 0.890 | 2.31 [-30.30, 34.92] |
| male | 18+ vs 50-60 | -5.3% | 0.877 | -3.49 [-47.77, 40.78] |
| male | 18+ vs 60+ | -9.7% | 0.775 | -6.70 [-52.57, 39.18] |
| male | 20-30 vs 30-40 | -34.8% | 0.334 | -23.43 [-70.96, 24.10] |
| male | 20-30 vs 40-50 | -26.9% | 0.483 | -16.14 [-61.23, 28.94] |
| male | 20-30 vs 50-60 | -33.3% | 0.427 | -21.95 [-76.07, 32.18] |
| male | 20-30 vs 60+ | -36.4% | 0.374 | -25.15 [-80.59, 30.29] |
| male | 30-40 vs 40-50 | +12.1% | 0.522 | 7.29 [-15.01, 29.58] |
| male | 30-40 vs 50-60 | +2.3% | 0.938 | 1.48 [-35.85, 38.82] |
| male | 30-40 vs 60+ | -2.5% | 0.931 | -1.72 [-40.94, 37.50] |
| male | 40-50 vs 50-60 | -8.8% | 0.739 | -5.80 [-39.98, 28.38] |
| male | 40-50 vs 60+ | -13.0% | 0.626 | -9.01 [-45.23, 27.22] |
| male | 50-60 vs 60+ | -4.6% | 0.894 | -3.20 [-50.21, 43.80] |
| female | 18+ vs 20-30 | +22.0% | 0.450 | 9.51 [-15.16, 34.18] |
| female | 18+ vs 30-40 | -1.7% | 0.945 | -0.89 [-25.92, 24.15] |
| female | 18+ vs 40-50 | -5.2% | 0.812 | -2.90 [-26.76, 20.96] |
| female | 18+ vs 50-60 | -6.3% | 0.802 | -3.55 [-31.23, 24.13] |
| female | 18+ vs 60+ | -3.4% | 0.907 | -1.84 [-32.76, 29.09] |
| female | 20-30 vs 30-40 | -19.4% | 0.160 | -10.40 [-24.90, 4.10] |
| female | 20-30 vs 40-50 | -22.3% | <0.05 | -12.41 [-24.77, -0.06] |
| female | 20-30 vs 50-60 | -23.2% | 0.171 | -13.06 [-31.76, 5.64] |
| female | 20-30 vs 60+ | -20.8% | 0.338 | -11.35 [-34.59, 11.89] |
| female | 30-40 vs 40-50 | -3.6% | 0.763 | -2.01 [-15.08, 11.06] |
| female | 30-40 vs 50-60 | -4.7% | 0.786 | -2.66 [-21.84, 16.51] |
| female | 30-40 vs 60+ | -1.7% | 0.937 | -0.95 [-24.57, 22.67] |
| female | 40-50 vs 50-60 | -1.2% | 0.942 | -0.65 [-18.26, 16.96] |
| female | 40-50 vs 60+ | +1.9% | 0.926 | 1.06 [-21.31, 23.43] |
| female | 50-60 vs 60+ | +3.1% | 0.899 | 1.71 [-24.70, 28.12] |
| *Mean difference = Group 1 − Group 2 in component units (mmol/24h, L/24h, or pH); % shift = difference relative to Group 2.* | | | | |

potassium_ma_2nd_analysis$wald_tests$sex_within_age_bands

Table 1: **Sex Comparisons Within Age Bands**

| Age Band | % Shift | P (Wald Test) | Mean Diff (95% CI) |
| --- | --- | --- | --- |
| 18+ | +18.2% | 0.621 | 9.58 [-28.37, 47.53] |
| 20-30 | +1.5% | 0.978 | 0.64 [-44.01, 45.29] |
| 30-40 | +25.5% | 0.219 | 13.67 [-8.14, 35.48] |
| 40-50 | +7.9% | 0.536 | 4.37 [-9.49, 18.24] |
| 50-60 | +16.9% | 0.603 | 9.52 [-26.34, 45.39] |
| 60+ | +26.4% | 0.482 | 14.44 [-25.82, 54.70] |
| *Mean difference = Group 1 − Group 2 in component units (mmol/24h, L/24h, or pH); % shift = difference relative to Group 2.* | | | |

###### 6.10.1.4 Meta-Regression (plain language)

format_meta_reg_plain("potassium", sex_group = "all")

Message

No meta-regression result for potassium / all

###### 6.10.1.5 Assay Subgroup Analysis

assay_subgroup_table("potassium")

Table 1: **Assay subgroup analysis — Potassium**

*Per-assay pooled estimates; categories with k < 3 listed but not pooled.*

| Assay category | Studies, n | Participants | 95% CI | I² | Q p-value | Egger's p | Note |
| --- | --- | --- | --- | --- | --- | --- | --- |
| Unreported | 63 | 6,578 | 54.40 [51.28, 57.53] | 99% | <0.001 | 0.717 |  |
| Autoanalyser | 19 | 2,403 | 43.02 [39.83, 46.22] | 97% | <0.001 | 0.649 |  |
| Flame photometry / emission | 6 | 311 | 51.39 [42.24, 60.54] | 97% | <0.001 | 0.319 |  |
| Ion-selective electrode | 5 | 126 | 67.94 [49.13, 86.74] | 97% | <0.001 | <0.001 |  |

###### 6.10.1.6 Preservative Sensitivity (Litholink / acidified samples)

```
preservative_sensitivity_ma("potassium")
## # A tibble: 3 × 6
##   group               n_studies n_participants pooled_estimate lower_95 upper_95
##   <chr>                   <int>          <int>           <dbl>    <dbl>    <dbl>
## 1 All studies                93           9418            52.4     49.8     55.1
## 2 Excluding preserva…        92           9408            52.5     49.8     55.2
## 3 Preservative only           1             10            NA       NA       NA
```

###### 6.10.1.7 Creatinine Completeness Sensitivity

creat_completeness_table("potassium")

Table 1: **Creatinine-completeness sensitivity — Potassium**

*Compares the all-studies pooled estimate against subsets defined by whether the study reported 24h urine creatinine. Studies that do not report creatinine cannot verify collection completeness; a substantial shift here flags possible bias from incomplete collections.*

| Subset | Studies, n | Participants | Range (95% CI) | Δ vs all-studies (%) | Flag |
| --- | --- | --- | --- | --- | --- |
| All studies | 93 | 9,418 | 52.41 [49.76, 55.07] | — | — |
| Creatinine reported only | 69 | 8,551 | 52.09 [48.85, 55.32] | -0.6% | OK |
| Creatinine NOT reported | 24 | 867 | 53.46 [49.01, 57.91] | +2.0% | OK |

##### 6.10.2 Comparison by Age / Sex / race

###### 6.10.2.1 Plot

```
potassium_ma_3rd_analysis <- sex_age_race_subdivision_ma("potassium",
                                                      urine_ma2,
                                                      overall_bias)
## Not enough studies for multivariable meta-regression (k = 3, need > 6). Running univariable models instead.
potassium_ma_3rd_analysis$race_plot
## `height` was translated to `width`.
## `height` was translated to `width`.
## `height` was translated to `width`.
```

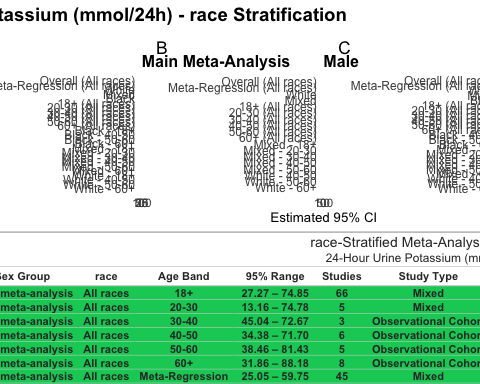

###### 6.10.2.2 Combined MA Results

potassium_ma_3rd_analysis$combined_summary_table

Table 1: **Combined Meta-Analysis Results**

24-Hour Urine Potassium (mmol/24h)

| Sex Group | race | Age Band | Pooled | 95% Range | Studies Mean SD Range | Studies | Study Type | Participants | I² | Cochran's Q | Q (p-value) | Egger's p | Model |
| --- | --- | --- | --- | --- | --- | --- | --- | --- | --- | --- | --- | --- | --- |
| main meta-analysis | All | 18+ | 51.06 | 27.27 – 74.85 | 51.77 ± 13.45 (29.91–86.00) | 66 | Mixed | 5065 | 97.3% | 2437.2 | <0.005 | 0.05 | Random |
| main meta-analysis | All | 20-30 | 43.97 | 13.16 – 74.78 | 44.01 ± 15.77 (21.00–65.50) | 5 | Mixed | 149 | 99.2% | 518.6 | <0.005 | 0.67 | Random |
| main meta-analysis | All | 30-40 | 58.86 | 45.04 – 72.67 | 59.00 ± 7.31 (53.66–67.33) | 3 | Observational Cohort | 67 | 94.8% | 38.2 | <0.005 | 0.28 | Random |
| main meta-analysis | All | 40-50 | 53.04 | 34.38 – 71.70 | 53.84 ± 9.53 (36.10–61.50) | 6 | Observational Cohort | 643 | 99.6% | 1170.7 | <0.005 | 0.24 | Random |
| main meta-analysis | All | 50-60 | 59.95 | 38.46 – 81.43 | 60.19 ± 11.41 (49.33–77.76) | 5 | Observational Cohort | 2669 | 98.9% | 374 | <0.005 | 0.53 | Random |
| main meta-analysis | All | 60+ | 60.02 | 31.86 – 88.18 | 60.13 ± 14.47 (37.07–79.80) | 8 | Observational Cohort | 825 | 99.4% | 1147.5 | <0.005 | <0.01 | Random |
| **main meta-analysis** | **All** | **Meta-Regression** | **42.40** | **25.05 – 59.75** | **—** | **45** | **Mixed** | **3787** | **95.6%** | **548.7** | **<0.005** | **<0.01** | **Multivariable** |
| **main meta-analysis** | **All** | **Meta-Regression** | **42.40** | **25.05 – 59.75** | **—** | **45** | **Mixed** | **3787** | **95.6%** | **548.7** | **<0.005** | **<0.01** | **Multivariable** |
| **main meta-analysis** | **All** | **Overall** | **52.41** | **27.72 – 77.11** | **52.89 ± 13.43 (21.00–86.00)** | **93** | **Mixed** | **9418** | **98.8%** | **7644.4** | **<0.005** | **0.74** | **Random** |
| **main meta-analysis** | **All** | **Overall** | **52.41** | **27.72 – 77.11** | **52.89 ± 13.43 (21.00–86.00)** | **93** | **Mixed** | **9418** | **98.8%** | **7644.4** | **<0.005** | **0.74** | **Random** |
| main meta-analysis | black | 18+ | 50.00 | -4.88 – 104.88 | 50.00 ± 0.00 (50.00–50.00) | 3 | Observational Cohort | 51 | 0% | 0 | 1.00 | — | Fixed |
| main meta-analysis | black | 20-30 | — | NA – NA | — | NA | NA | NA | — | — | — | — | NA |
| main meta-analysis | black | 30-40 | — | NA – NA | — | NA | NA | NA | — | — | — | — | NA |
| main meta-analysis | black | 40-50 | 51.24 | 18.45 – 84.03 | — (51.24–51.24) | 1 | Observational Cohort | 29 | — | — | — | — | Single study |
| main meta-analysis | black | 50-60 | 54.31 | 18.15 – 90.47 | — (54.31–54.31) | 1 | Observational Cohort | 131 | — | — | — | — | Single study |
| main meta-analysis | black | 60+ | 54.84 | 20.68 – 88.99 | 55.01 ± 1.09 (54.24–55.78) | 2 | Observational Cohort | 144 | 0% | 0.3 | 0.61 | — | Fixed |
| **main meta-analysis** | **black** | **Meta-Regression** | **—** | **NA – NA** | **—** | **NA** | **NA** | **NA** | **—** | **—** | **—** | **—** | **NA** |
| **main meta-analysis** | **black** | **Overall** | **53.97** | **17.54 – 90.40** | **52.22 ± 2.48 (50.00–55.78)** | **7** | **Observational Cohort** | **355** | **0%** | **2.5** | **0.87** | **0.05** | **Fixed** |
| main meta-analysis | mixed | 18+ | 50.98 | 26.44 – 75.52 | 51.74 ± 14.12 (32.00–86.00) | 55 | Mixed | 4268 | 96.9% | 1759.6 | <0.005 | <0.05 | Random |
| main meta-analysis | mixed | 20-30 | 43.97 | 13.16 – 74.78 | 44.01 ± 15.77 (21.00–65.50) | 5 | Mixed | 149 | 99.2% | 518.6 | <0.005 | 0.67 | Random |
| main meta-analysis | mixed | 30-40 | 58.86 | 45.04 – 72.67 | 59.00 ± 7.31 (53.66–67.33) | 3 | Observational Cohort | 67 | 94.8% | 38.2 | <0.005 | 0.28 | Random |
| main meta-analysis | mixed | 40-50 | 50.14 | 25.66 – 74.62 | 50.16 ± 12.50 (36.10–60.03) | 3 | Observational Cohort | 84 | 99.7% | 775.3 | <0.005 | 0.13 | Random |
| main meta-analysis | mixed | 50-60 | 51.84 | 45.11 – 58.58 | 51.84 ± 3.54 (49.33–54.34) | 2 | Observational Cohort | 50 | 94.1% | 16.9 | <0.005 | — | Random |
| main meta-analysis | mixed | 60+ | 37.07 | 32.60 – 41.54 | — (37.07–37.07) | 1 | Observational Cohort | 25 | — | — | — | — | Single study |
| **main meta-analysis** | **mixed** | **Meta-Regression** | **—** | **NA – NA** | **—** | **NA** | **NA** | **NA** | **—** | **—** | **—** | **—** | **NA** |
| **main meta-analysis** | **mixed** | **Overall** | **50.58** | **26.31 – 74.84** | **51.22 ± 13.75 (21.00–86.00)** | **69** | **Mixed** | **4643** | **98.7%** | **5093.6** | **<0.005** | **0.34** | **Random** |
| main meta-analysis | white | 18+ | 52.35 | 27.62 – 77.07 | 52.68 ± 11.81 (29.91–73.50) | 8 | Observational Cohort | 746 | 98.8% | 570.9 | <0.005 | 0.39 | Random |
| main meta-analysis | white | 20-30 | — | NA – NA | — | NA | NA | NA | — | — | — | — | NA |
| main meta-analysis | white | 30-40 | — | NA – NA | — | NA | NA | NA | — | — | — | — | NA |
| main meta-analysis | white | 40-50 | 59.81 | 21.88 – 97.74 | 60.65 ± 1.20 (59.80–61.50) | 2 | Observational Cohort | 530 | 0% | 0 | 0.88 | — | Fixed |
| main meta-analysis | white | 50-60 | 71.08 | 54.24 – 87.91 | 71.47 ± 8.89 (65.19–77.76) | 2 | Observational Cohort | 2488 | 93.4% | 15.2 | <0.005 | — | Random |
| main meta-analysis | white | 60+ | 66.66 | 41.72 – 91.60 | 66.79 ± 12.82 (48.94–79.80) | 5 | Observational Cohort | 656 | 98.9% | 375.9 | <0.005 | 0.05 | Random |
| **main meta-analysis** | **white** | **Meta-Regression** | **—** | **NA – NA** | **—** | **NA** | **NA** | **NA** | **—** | **—** | **—** | **—** | **NA** |
| **main meta-analysis** | **white** | **Overall** | **60.18** | **34.47 – 85.89** | **59.98 ± 12.84 (29.91–79.80)** | **17** | **Observational Cohort** | **4420** | **99%** | **1596.3** | **<0.005** | **0.98** | **Random** |
| male | All | 18+ | 62.35 | 31.90 – 92.81 | 63.26 ± 16.64 (41.00–83.68) | 7 | Observational Cohort | 367 | 94.6% | 110.8 | <0.005 | 0.31 | Random |
| male | All | 20-30 | 43.90 | 0.34 – 87.46 | 43.93 ± 22.28 (21.00–65.50) | 3 | Mixed | 71 | 99.6% | 508.4 | <0.005 | 0.72 | Random |
| male | All | 30-40 | 67.33 | 48.32 – 86.34 | — (67.33–67.33) | 1 | Observational Cohort | 25 | — | — | — | — | Single study |
| male | All | 40-50 | 60.04 | 48.40 – 71.69 | 60.77 ± 1.04 (60.03–61.50) | 2 | Observational Cohort | 31 | 0% | 0 | 0.89 | — | Fixed |
| male | All | 50-60 | 65.85 | 33.71 – 97.98 | 66.05 ± 16.56 (54.34–77.76) | 2 | Observational Cohort | 104 | 98% | 50.2 | <0.005 | — | Random |
| male | All | 60+ | 69.05 | 34.75 – 103.35 | 69.16 ± 17.52 (48.94–79.80) | 3 | Observational Cohort | 329 | 99.4% | 331.8 | <0.005 | 0.24 | Random |
| **male** | **All** | **Meta-Regression** | **53.90** | **8.71 – 99.09** | **—** | **3** | **Mixed** | **155** | **99.5%** | **516** | **<0.005** | **0.28** | **Univariable** |
| **male** | **All** | **Overall** | **60.85** | **28.71 – 92.99** | **61.28 ± 16.70 (21.00–83.68)** | **18** | **Mixed** | **927** | **98.9%** | **1526.4** | **<0.005** | **0.53** | **Random** |
| male | black | 18+ | — | NA – NA | — | NA | NA | NA | — | — | — | — | NA |
| male | black | 20-30 | — | NA – NA | — | NA | NA | NA | — | — | — | — | NA |
| male | black | 30-40 | — | NA – NA | — | NA | NA | NA | — | — | — | — | NA |
| male | black | 40-50 | — | NA – NA | — | NA | NA | NA | — | — | — | — | NA |
| male | black | 50-60 | — | NA – NA | — | NA | NA | NA | — | — | — | — | NA |
| male | black | 60+ | — | NA – NA | — | NA | NA | NA | — | — | — | — | NA |
| **male** | **black** | **Meta-Regression** | **—** | **NA – NA** | **—** | **NA** | **NA** | **NA** | **—** | **—** | **—** | **—** | **NA** |
| **male** | **black** | **Overall** | **—** | **NA – NA** | **—** | **NA** | **NA** | **NA** | **—** | **—** | **—** | **—** | **NA** |
| male | mixed | 18+ | 62.35 | 31.90 – 92.81 | 63.26 ± 16.64 (41.00–83.68) | 7 | Observational Cohort | 367 | 94.6% | 110.8 | <0.005 | 0.31 | Random |
| male | mixed | 20-30 | 43.90 | 0.34 – 87.46 | 43.93 ± 22.28 (21.00–65.50) | 3 | Mixed | 71 | 99.6% | 508.4 | <0.005 | 0.72 | Random |
| male | mixed | 30-40 | 67.33 | 48.32 – 86.34 | — (67.33–67.33) | 1 | Observational Cohort | 25 | — | — | — | — | Single study |
| male | mixed | 40-50 | 60.03 | 49.52 – 70.54 | — (60.03–60.03) | 1 | Observational Cohort | 25 | — | — | — | — | Single study |
| male | mixed | 50-60 | 54.34 | 46.17 – 62.51 | — (54.34–54.34) | 1 | Observational Cohort | 25 | — | — | — | — | Single study |
| male | mixed | 60+ | — | NA – NA | — | NA | NA | NA | — | — | — | — | NA |
| **male** | **mixed** | **Meta-Regression** | **—** | **NA – NA** | **—** | **NA** | **NA** | **NA** | **—** | **—** | **—** | **—** | **NA** |
| **male** | **mixed** | **Overall** | **57.60** | **25.39 – 89.81** | **58.18 ± 17.19 (21.00–83.68)** | **13** | **Mixed** | **513** | **98.9%** | **1136.4** | **<0.005** | **0.94** | **Random** |
| male | white | 18+ | — | NA – NA | — | NA | NA | NA | — | — | — | — | NA |
| male | white | 20-30 | — | NA – NA | — | NA | NA | NA | — | — | — | — | NA |
| male | white | 30-40 | — | NA – NA | — | NA | NA | NA | — | — | — | — | NA |
| male | white | 40-50 | 61.50 | 8.56 – 114.44 | — (61.50–61.50) | 1 | Observational Cohort | 6 | — | — | — | — | Single study |
| male | white | 50-60 | 77.76 | 22.02 – 133.50 | — (77.76–77.76) | 1 | Observational Cohort | 79 | — | — | — | — | Single study |
| male | white | 60+ | 69.05 | 34.75 – 103.35 | 69.16 ± 17.52 (48.94–79.80) | 3 | Observational Cohort | 329 | 99.4% | 331.8 | <0.005 | 0.24 | Random |
| **male** | **white** | **Meta-Regression** | **—** | **NA – NA** | **—** | **NA** | **NA** | **NA** | **—** | **—** | **—** | **—** | **NA** |
| **male** | **white** | **Overall** | **69.82** | **42.69 – 96.94** | **69.35 ± 13.66 (48.94–79.80)** | **5** | **Observational Cohort** | **414** | **99%** | **382.2** | **<0.005** | **0.17** | **Random** |
| female | All | 18+ | 52.77 | 30.13 – 75.41 | 53.53 ± 12.48 (36.00–71.10) | 9 | Mixed | 540 | 94.2% | 138 | <0.005 | <0.01 | Random |
| female | All | 20-30 | 43.26 | 33.46 – 53.06 | — (43.26–43.26) | 1 | Observational Cohort | 25 | — | — | — | — | Single study |
| female | All | 30-40 | 53.66 | 42.98 – 64.34 | — (53.66–53.66) | 1 | Observational Cohort | 25 | — | — | — | — | Single study |
| female | All | 40-50 | 55.67 | 48.15 – 63.19 | 55.13 ± 4.33 (51.24–59.80) | 3 | Observational Cohort | 578 | 92.4% | 26.2 | <0.005 | 0.85 | Random |
| female | All | 50-60 | 56.32 | 40.40 – 72.25 | 56.28 ± 8.11 (49.33–65.19) | 3 | Observational Cohort | 2565 | 99.3% | 279.7 | <0.005 | 0.43 | Random |
| female | All | 60+ | 54.61 | 33.54 – 75.68 | 54.71 ± 10.71 (37.07–64.05) | 5 | Observational Cohort | 496 | 99.2% | 526.3 | <0.005 | <0.05 | Random |
| **female** | **All** | **Meta-Regression** | **44.79** | **33.24 – 56.34** | **—** | **7** | **Mixed** | **488** | **89.5%** | **26.7** | **<0.005** | **<0.05** | **Multivariable** |
| **female** | **All** | **Overall** | **53.49** | **35.51 – 71.47** | **53.93 ± 9.79 (36.00–71.10)** | **22** | **Mixed** | **4229** | **99.2%** | **2516.4** | **<0.005** | **0.91** | **Random** |
| female | black | 18+ | — | NA – NA | — | NA | NA | NA | — | — | — | — | NA |
| female | black | 20-30 | — | NA – NA | — | NA | NA | NA | — | — | — | — | NA |
| female | black | 30-40 | — | NA – NA | — | NA | NA | NA | — | — | — | — | NA |
| female | black | 40-50 | 51.24 | 18.45 – 84.03 | — (51.24–51.24) | 1 | Observational Cohort | 29 | — | — | — | — | Single study |
| female | black | 50-60 | 54.31 | 18.15 – 90.47 | — (54.31–54.31) | 1 | Observational Cohort | 131 | — | — | — | — | Single study |
| female | black | 60+ | 54.84 | 20.68 – 88.99 | 55.01 ± 1.09 (54.24–55.78) | 2 | Observational Cohort | 144 | 0% | 0.3 | 0.61 | — | Fixed |
| **female** | **black** | **Meta-Regression** | **—** | **NA – NA** | **—** | **NA** | **NA** | **NA** | **—** | **—** | **—** | **—** | **NA** |
| **female** | **black** | **Overall** | **54.24** | **19.41 – 89.07** | **53.89 ± 1.91 (51.24–55.78)** | **4** | **Observational Cohort** | **304** | **0%** | **1.4** | **0.71** | **0.52** | **Fixed** |
| female | mixed | 18+ | 52.77 | 30.13 – 75.41 | 53.53 ± 12.48 (36.00–71.10) | 9 | Mixed | 540 | 94.2% | 138 | <0.005 | <0.01 | Random |
| female | mixed | 20-30 | 43.26 | 33.46 – 53.06 | — (43.26–43.26) | 1 | Observational Cohort | 25 | — | — | — | — | Single study |
| female | mixed | 30-40 | 53.66 | 42.98 – 64.34 | — (53.66–53.66) | 1 | Observational Cohort | 25 | — | — | — | — | Single study |
| female | mixed | 40-50 | 54.36 | 46.97 – 61.75 | — (54.36–54.36) | 1 | Observational Cohort | 25 | — | — | — | — | Single study |
| female | mixed | 50-60 | 49.33 | 40.61 – 58.05 | — (49.33–49.33) | 1 | Observational Cohort | 25 | — | — | — | — | Single study |
| female | mixed | 60+ | 37.07 | 32.60 – 41.54 | — (37.07–37.07) | 1 | Observational Cohort | 25 | — | — | — | — | Single study |
| **female** | **mixed** | **Meta-Regression** | **—** | **NA – NA** | **—** | **NA** | **NA** | **NA** | **—** | **—** | **—** | **—** | **NA** |
| **female** | **mixed** | **Overall** | **50.65** | **31.32 – 69.98** | **51.39 ± 11.01 (36.00–71.10)** | **14** | **Mixed** | **665** | **98.1%** | **679** | **<0.005** | **<0.05** | **Random** |
| female | white | 18+ | — | NA – NA | — | NA | NA | NA | — | — | — | — | NA |
| female | white | 20-30 | — | NA – NA | — | NA | NA | NA | — | — | — | — | NA |
| female | white | 30-40 | — | NA – NA | — | NA | NA | NA | — | — | — | — | NA |
| female | white | 40-50 | 59.80 | 21.97 – 97.63 | — (59.80–59.80) | 1 | Observational Cohort | 524 | — | — | — | — | Single study |
| female | white | 50-60 | 65.19 | 23.72 – 106.66 | — (65.19–65.19) | 1 | Observational Cohort | 2409 | — | — | — | — | Single study |
| female | white | 60+ | 63.59 | 21.56 – 105.61 | 63.22 ± 1.17 (62.40–64.05) | 2 | Observational Cohort | 327 | 0% | 0.4 | 0.53 | — | Fixed |
| **female** | **white** | **Meta-Regression** | **—** | **NA – NA** | **—** | **NA** | **NA** | **NA** | **—** | **—** | **—** | **—** | **NA** |
| **female** | **white** | **Overall** | **62.92** | **58.22 – 67.61** | **62.86 ± 2.34 (59.80–65.19)** | **4** | **Observational Cohort** | **3260** | **90.9%** | **32.9** | **<0.005** | **0.50** | **Random** |
| *I² represents heterogeneity; Q test assesses between-study variation; Egger's p = linear regression test for funnel plot asymmetry; Meta-Regression shows covariate-adjusted estimates* | | | | | | | | | | | | | |

###### 6.10.2.3 Table of Results subdivided by race / Sex / Age

potassium_ma_3rd_analysis$race_summary_table

Table 1: **race-Stratified Meta-Analysis Results**

24-Hour Urine Potassium (mmol/24h)

| Sex Group | race | Age Band | 95% Range | Studies | Study Type | Participants | I² | Cochran's Q | Q (p-value) | Egger's p | Model |
| --- | --- | --- | --- | --- | --- | --- | --- | --- | --- | --- | --- |
| **main meta-analysis** | **All races** | **18+** | **27.27 – 74.85** | **66** | **Mixed** | **5065** | **97.3%** | **2437.2** | **<0.005** | **0.05** | **Random** |
| **main meta-analysis** | **All races** | **20-30** | **13.16 – 74.78** | **5** | **Mixed** | **149** | **99.2%** | **518.6** | **<0.005** | **0.67** | **Random** |
| **main meta-analysis** | **All races** | **30-40** | **45.04 – 72.67** | **3** | **Observational Cohort** | **67** | **94.8%** | **38.2** | **<0.005** | **0.28** | **Random** |
| **main meta-analysis** | **All races** | **40-50** | **34.38 – 71.70** | **6** | **Observational Cohort** | **643** | **99.6%** | **1170.7** | **<0.005** | **0.24** | **Random** |
| **main meta-analysis** | **All races** | **50-60** | **38.46 – 81.43** | **5** | **Observational Cohort** | **2669** | **98.9%** | **374** | **<0.005** | **0.53** | **Random** |
| **main meta-analysis** | **All races** | **60+** | **31.86 – 88.18** | **8** | **Observational Cohort** | **825** | **99.4%** | **1147.5** | **<0.005** | **<0.01** | **Random** |
| **main meta-analysis** | **All races** | **Meta-Regression** | **25.05 – 59.75** | **45** | **Mixed** | **3787** | **95.6%** | **548.7** | **<0.005** | **<0.01** | **Multivariable** |
| **main meta-analysis** | **All races** | **Meta-Regression** | **25.05 – 59.75** | **45** | **Mixed** | **3787** | **95.6%** | **548.7** | **<0.005** | **<0.01** | **Multivariable** |
| **main meta-analysis** | **All races** | **Overall** | **27.72 – 77.11** | **93** | **Mixed** | **9418** | **98.8%** | **7644.4** | **<0.005** | **0.74** | **Random** |
| **main meta-analysis** | **All races** | **Overall** | **27.72 – 77.11** | **93** | **Mixed** | **9418** | **98.8%** | **7644.4** | **<0.005** | **0.74** | **Random** |
| main meta-analysis | Black | 18+ | -4.88 – 104.88 | 3 | Observational Cohort | 51 | 0% | 0 | 1.00 | — | Fixed |
| main meta-analysis | Black | 20-30 | — | — | NA | — | — | — | — | — | — |
| main meta-analysis | Black | 30-40 | — | — | NA | — | — | — | — | — | — |
| main meta-analysis | Black | 40-50 | 18.45 – 84.03 | 1 | Observational Cohort | 29 | — | — | — | — | Single study |
| main meta-analysis | Black | 50-60 | 18.15 – 90.47 | 1 | Observational Cohort | 131 | — | — | — | — | Single study |
| main meta-analysis | Black | 60+ | 20.68 – 88.99 | 2 | Observational Cohort | 144 | 0% | 0.3 | 0.61 | — | Fixed |
| main meta-analysis | Black | Meta-Regression | — | — | NA | — | — | — | — | — | — |
| **main meta-analysis** | **Black** | **Overall** | **17.54 – 90.40** | **7** | **Observational Cohort** | **355** | **0%** | **2.5** | **0.87** | **0.05** | **Fixed** |
| main meta-analysis | Mixed | 18+ | 26.44 – 75.52 | 55 | Mixed | 4268 | 96.9% | 1759.6 | <0.005 | <0.05 | Random |
| main meta-analysis | Mixed | 20-30 | 13.16 – 74.78 | 5 | Mixed | 149 | 99.2% | 518.6 | <0.005 | 0.67 | Random |
| main meta-analysis | Mixed | 30-40 | 45.04 – 72.67 | 3 | Observational Cohort | 67 | 94.8% | 38.2 | <0.005 | 0.28 | Random |
| main meta-analysis | Mixed | 40-50 | 25.66 – 74.62 | 3 | Observational Cohort | 84 | 99.7% | 775.3 | <0.005 | 0.13 | Random |
| main meta-analysis | Mixed | 50-60 | 45.11 – 58.58 | 2 | Observational Cohort | 50 | 94.1% | 16.9 | <0.005 | — | Random |
| main meta-analysis | Mixed | 60+ | 32.60 – 41.54 | 1 | Observational Cohort | 25 | — | — | — | — | Single study |
| main meta-analysis | Mixed | Meta-Regression | — | — | NA | — | — | — | — | — | — |
| **main meta-analysis** | **Mixed** | **Overall** | **26.31 – 74.84** | **69** | **Mixed** | **4643** | **98.7%** | **5093.6** | **<0.005** | **0.34** | **Random** |
| main meta-analysis | White | 18+ | 27.62 – 77.07 | 8 | Observational Cohort | 746 | 98.8% | 570.9 | <0.005 | 0.39 | Random |
| main meta-analysis | White | 20-30 | — | — | NA | — | — | — | — | — | — |
| main meta-analysis | White | 30-40 | — | — | NA | — | — | — | — | — | — |
| main meta-analysis | White | 40-50 | 21.88 – 97.74 | 2 | Observational Cohort | 530 | 0% | 0 | 0.88 | — | Fixed |
| main meta-analysis | White | 50-60 | 54.24 – 87.91 | 2 | Observational Cohort | 2488 | 93.4% | 15.2 | <0.005 | — | Random |
| main meta-analysis | White | 60+ | 41.72 – 91.60 | 5 | Observational Cohort | 656 | 98.9% | 375.9 | <0.005 | 0.05 | Random |
| main meta-analysis | White | Meta-Regression | — | — | NA | — | — | — | — | — | — |
| **main meta-analysis** | **White** | **Overall** | **34.47 – 85.89** | **17** | **Observational Cohort** | **4420** | **99%** | **1596.3** | **<0.005** | **0.98** | **Random** |
| **male** | **All races** | **18+** | **31.90 – 92.81** | **7** | **Observational Cohort** | **367** | **94.6%** | **110.8** | **<0.005** | **0.31** | **Random** |
| **male** | **All races** | **20-30** | **0.34 – 87.46** | **3** | **Mixed** | **71** | **99.6%** | **508.4** | **<0.005** | **0.72** | **Random** |
| **male** | **All races** | **30-40** | **48.32 – 86.34** | **1** | **Observational Cohort** | **25** | **—** | **—** | **—** | **—** | **Single study** |
| **male** | **All races** | **40-50** | **48.40 – 71.69** | **2** | **Observational Cohort** | **31** | **0%** | **0** | **0.89** | **—** | **Fixed** |
| **male** | **All races** | **50-60** | **33.71 – 97.98** | **2** | **Observational Cohort** | **104** | **98%** | **50.2** | **<0.005** | **—** | **Random** |
| **male** | **All races** | **60+** | **34.75 – 103.35** | **3** | **Observational Cohort** | **329** | **99.4%** | **331.8** | **<0.005** | **0.24** | **Random** |
| **male** | **All races** | **Meta-Regression** | **8.71 – 99.09** | **3** | **Mixed** | **155** | **99.5%** | **516** | **<0.005** | **0.28** | **Univariable** |
| **male** | **All races** | **Overall** | **28.71 – 92.99** | **18** | **Mixed** | **927** | **98.9%** | **1526.4** | **<0.005** | **0.53** | **Random** |
| male | Black | 18+ | — | — | NA | — | — | — | — | — | — |
| male | Black | 20-30 | — | — | NA | — | — | — | — | — | — |
| male | Black | 30-40 | — | — | NA | — | — | — | — | — | — |
| male | Black | 40-50 | — | — | NA | — | — | — | — | — | — |
| male | Black | 50-60 | — | — | NA | — | — | — | — | — | — |
| male | Black | 60+ | — | — | NA | — | — | — | — | — | — |
| male | Black | Meta-Regression | — | — | NA | — | — | — | — | — | — |
| **male** | **Black** | **Overall** | **—** | **—** | **NA** | **—** | **—** | **—** | **—** | **—** | **—** |
| male | Mixed | 18+ | 31.90 – 92.81 | 7 | Observational Cohort | 367 | 94.6% | 110.8 | <0.005 | 0.31 | Random |
| male | Mixed | 20-30 | 0.34 – 87.46 | 3 | Mixed | 71 | 99.6% | 508.4 | <0.005 | 0.72 | Random |
| male | Mixed | 30-40 | 48.32 – 86.34 | 1 | Observational Cohort | 25 | — | — | — | — | Single study |
| male | Mixed | 40-50 | 49.52 – 70.54 | 1 | Observational Cohort | 25 | — | — | — | — | Single study |
| male | Mixed | 50-60 | 46.17 – 62.51 | 1 | Observational Cohort | 25 | — | — | — | — | Single study |
| male | Mixed | 60+ | — | — | NA | — | — | — | — | — | — |
| male | Mixed | Meta-Regression | — | — | NA | — | — | — | — | — | — |
| **male** | **Mixed** | **Overall** | **25.39 – 89.81** | **13** | **Mixed** | **513** | **98.9%** | **1136.4** | **<0.005** | **0.94** | **Random** |
| male | White | 18+ | — | — | NA | — | — | — | — | — | — |
| male | White | 20-30 | — | — | NA | — | — | — | — | — | — |
| male | White | 30-40 | — | — | NA | — | — | — | — | — | — |
| male | White | 40-50 | 8.56 – 114.44 | 1 | Observational Cohort | 6 | — | — | — | — | Single study |
| male | White | 50-60 | 22.02 – 133.50 | 1 | Observational Cohort | 79 | — | — | — | — | Single study |
| male | White | 60+ | 34.75 – 103.35 | 3 | Observational Cohort | 329 | 99.4% | 331.8 | <0.005 | 0.24 | Random |
| male | White | Meta-Regression | — | — | NA | — | — | — | — | — | — |
| **male** | **White** | **Overall** | **42.69 – 96.94** | **5** | **Observational Cohort** | **414** | **99%** | **382.2** | **<0.005** | **0.17** | **Random** |
| **female** | **All races** | **18+** | **30.13 – 75.41** | **9** | **Mixed** | **540** | **94.2%** | **138** | **<0.005** | **<0.01** | **Random** |
| **female** | **All races** | **20-30** | **33.46 – 53.06** | **1** | **Observational Cohort** | **25** | **—** | **—** | **—** | **—** | **Single study** |
| **female** | **All races** | **30-40** | **42.98 – 64.34** | **1** | **Observational Cohort** | **25** | **—** | **—** | **—** | **—** | **Single study** |
| **female** | **All races** | **40-50** | **48.15 – 63.19** | **3** | **Observational Cohort** | **578** | **92.4%** | **26.2** | **<0.005** | **0.85** | **Random** |
| **female** | **All races** | **50-60** | **40.40 – 72.25** | **3** | **Observational Cohort** | **2565** | **99.3%** | **279.7** | **<0.005** | **0.43** | **Random** |
| **female** | **All races** | **60+** | **33.54 – 75.68** | **5** | **Observational Cohort** | **496** | **99.2%** | **526.3** | **<0.005** | **<0.05** | **Random** |
| **female** | **All races** | **Meta-Regression** | **33.24 – 56.34** | **7** | **Mixed** | **488** | **89.5%** | **26.7** | **<0.005** | **<0.05** | **Multivariable** |
| **female** | **All races** | **Overall** | **35.51 – 71.47** | **22** | **Mixed** | **4229** | **99.2%** | **2516.4** | **<0.005** | **0.91** | **Random** |
| female | Black | 18+ | — | — | NA | — | — | — | — | — | — |
| female | Black | 20-30 | — | — | NA | — | — | — | — | — | — |
| female | Black | 30-40 | — | — | NA | — | — | — | — | — | — |
| female | Black | 40-50 | 18.45 – 84.03 | 1 | Observational Cohort | 29 | — | — | — | — | Single study |
| female | Black | 50-60 | 18.15 – 90.47 | 1 | Observational Cohort | 131 | — | — | — | — | Single study |
| female | Black | 60+ | 20.68 – 88.99 | 2 | Observational Cohort | 144 | 0% | 0.3 | 0.61 | — | Fixed |
| female | Black | Meta-Regression | — | — | NA | — | — | — | — | — | — |
| **female** | **Black** | **Overall** | **19.41 – 89.07** | **4** | **Observational Cohort** | **304** | **0%** | **1.4** | **0.71** | **0.52** | **Fixed** |
| female | Mixed | 18+ | 30.13 – 75.41 | 9 | Mixed | 540 | 94.2% | 138 | <0.005 | <0.01 | Random |
| female | Mixed | 20-30 | 33.46 – 53.06 | 1 | Observational Cohort | 25 | — | — | — | — | Single study |
| female | Mixed | 30-40 | 42.98 – 64.34 | 1 | Observational Cohort | 25 | — | — | — | — | Single study |
| female | Mixed | 40-50 | 46.97 – 61.75 | 1 | Observational Cohort | 25 | — | — | — | — | Single study |
| female | Mixed | 50-60 | 40.61 – 58.05 | 1 | Observational Cohort | 25 | — | — | — | — | Single study |
| female | Mixed | 60+ | 32.60 – 41.54 | 1 | Observational Cohort | 25 | — | — | — | — | Single study |
| female | Mixed | Meta-Regression | — | — | NA | — | — | — | — | — | — |
| **female** | **Mixed** | **Overall** | **31.32 – 69.98** | **14** | **Mixed** | **665** | **98.1%** | **679** | **<0.005** | **<0.05** | **Random** |
| female | White | 18+ | — | — | NA | — | — | — | — | — | — |
| female | White | 20-30 | — | — | NA | — | — | — | — | — | — |
| female | White | 30-40 | — | — | NA | — | — | — | — | — | — |
| female | White | 40-50 | 21.97 – 97.63 | 1 | Observational Cohort | 524 | — | — | — | — | Single study |
| female | White | 50-60 | 23.72 – 106.66 | 1 | Observational Cohort | 2409 | — | — | — | — | Single study |
| female | White | 60+ | 21.56 – 105.61 | 2 | Observational Cohort | 327 | 0% | 0.4 | 0.53 | — | Fixed |
| female | White | Meta-Regression | — | — | NA | — | — | — | — | — | — |
| **female** | **White** | **Overall** | **58.22 – 67.61** | **4** | **Observational Cohort** | **3260** | **90.9%** | **32.9** | **<0.005** | **0.50** | **Random** |
| *I² represents heterogeneity; Q test assesses between-study variation; Egger's p = linear regression test for funnel plot asymmetry; — indicates insufficient data* | | | | | | | | | | | |

###### 6.10.2.4 Wald Tests

potassium_ma_3rd_analysis$wald_tests$sex_comparison

Table 1: **Sex Comparison (Overall)**

| Comparison | % Shift | P (Wald Test) | Mean Diff (95% CI) |
| --- | --- | --- | --- |
| Male vs Female (Overall) | +13.8% | 0.695 | 7.36 [-29.47, 44.18] |
| *Mean difference = Group 1 − Group 2 in component units (mmol/24h, L/24h, or pH); % shift = difference relative to Group 2.* | | | |

potassium_ma_3rd_analysis$wald_tests$race_comparisons

Table 1: **race Comparisons Within Sex Groups**

| Sex Group | Comparison | % Shift | P (Wald Test) | Mean Diff (95% CI) |
| --- | --- | --- | --- | --- |
| male | black vs mixed | — | — | — |
| male | black vs white | — | — | — |
| male | mixed vs white | -17.5% | 0.570 | -12.22 [-54.32, 29.89] |
| female | black vs mixed | +7.1% | 0.860 | 3.59 [-36.25, 43.42] |
| female | black vs white | -13.8% | 0.628 | -8.68 [-43.82, 26.47] |
| female | mixed vs white | -19.5% | 0.227 | -12.27 [-32.16, 7.62] |
| *Mean difference = Group 1 − Group 2 in component units (mmol/24h, L/24h, or pH); % shift = difference relative to Group 2.* | | | | |

potassium_ma_3rd_analysis$wald_tests$age_band_comparisons

Table 1: **Age Band Comparisons Within Sex Groups**

| Sex Group | Comparison | % Shift | P (Wald Test) | Mean Diff (95% CI) |
| --- | --- | --- | --- | --- |
| main meta-analysis | 18+ vs 20-30 | +16.1% | 0.721 | 7.09 [-31.84, 46.02] |
| main meta-analysis | 18+ vs 30-40 | -13.2% | 0.579 | -7.79 [-35.30, 19.72] |
| main meta-analysis | 18+ vs 40-50 | -3.7% | 0.898 | -1.98 [-32.21, 28.26] |
| main meta-analysis | 18+ vs 50-60 | -14.8% | 0.587 | -8.89 [-40.94, 23.17] |
| main meta-analysis | 18+ vs 60+ | -14.9% | 0.634 | -8.96 [-45.82, 27.91] |
| main meta-analysis | 20-30 vs 30-40 | -25.3% | 0.388 | -14.88 [-48.66, 18.89] |
| main meta-analysis | 20-30 vs 40-50 | -17.1% | 0.622 | -9.07 [-45.09, 26.95] |
| main meta-analysis | 20-30 vs 50-60 | -26.7% | 0.404 | -15.98 [-53.54, 21.59] |
| main meta-analysis | 20-30 vs 60+ | -26.7% | 0.451 | -16.05 [-57.79, 25.69] |
| main meta-analysis | 30-40 vs 40-50 | +11.0% | 0.623 | 5.82 [-17.40, 29.03] |
| main meta-analysis | 30-40 vs 50-60 | -1.8% | 0.933 | -1.09 [-26.64, 24.45] |
| main meta-analysis | 30-40 vs 60+ | -1.9% | 0.942 | -1.17 [-32.53, 30.20] |
| main meta-analysis | 40-50 vs 50-60 | -11.5% | 0.634 | -6.91 [-35.36, 21.55] |
| main meta-analysis | 40-50 vs 60+ | -11.6% | 0.685 | -6.98 [-40.76, 26.80] |
| main meta-analysis | 50-60 vs 60+ | -0.1% | 0.997 | -0.07 [-35.49, 35.35] |
| male | 18+ vs 20-30 | +42.0% | 0.496 | 18.45 [-34.70, 71.60] |
| male | 18+ vs 30-40 | -7.4% | 0.786 | -4.98 [-40.88, 30.93] |
| male | 18+ vs 40-50 | +3.8% | 0.890 | 2.31 [-30.30, 34.92] |
| male | 18+ vs 50-60 | -5.3% | 0.877 | -3.49 [-47.77, 40.78] |
| male | 18+ vs 60+ | -9.7% | 0.775 | -6.70 [-52.57, 39.18] |
| male | 20-30 vs 30-40 | -34.8% | 0.334 | -23.43 [-70.96, 24.10] |
| male | 20-30 vs 40-50 | -26.9% | 0.483 | -16.14 [-61.23, 28.94] |
| male | 20-30 vs 50-60 | -33.3% | 0.427 | -21.95 [-76.07, 32.18] |
| male | 20-30 vs 60+ | -36.4% | 0.374 | -25.15 [-80.59, 30.29] |
| male | 30-40 vs 40-50 | +12.1% | 0.522 | 7.29 [-15.01, 29.58] |
| male | 30-40 vs 50-60 | +2.3% | 0.938 | 1.48 [-35.85, 38.82] |
| male | 30-40 vs 60+ | -2.5% | 0.931 | -1.72 [-40.94, 37.50] |
| male | 40-50 vs 50-60 | -8.8% | 0.739 | -5.80 [-39.98, 28.38] |
| male | 40-50 vs 60+ | -13.0% | 0.626 | -9.01 [-45.23, 27.22] |
| male | 50-60 vs 60+ | -4.6% | 0.894 | -3.20 [-50.21, 43.80] |
| female | 18+ vs 20-30 | +22.0% | 0.450 | 9.51 [-15.16, 34.18] |
| female | 18+ vs 30-40 | -1.7% | 0.945 | -0.89 [-25.92, 24.15] |
| female | 18+ vs 40-50 | -5.2% | 0.812 | -2.90 [-26.76, 20.96] |
| female | 18+ vs 50-60 | -6.3% | 0.802 | -3.55 [-31.23, 24.13] |
| female | 18+ vs 60+ | -3.4% | 0.907 | -1.84 [-32.76, 29.09] |
| female | 20-30 vs 30-40 | -19.4% | 0.160 | -10.40 [-24.90, 4.10] |
| female | 20-30 vs 40-50 | -22.3% | <0.05 | -12.41 [-24.77, -0.06] |
| female | 20-30 vs 50-60 | -23.2% | 0.171 | -13.06 [-31.76, 5.64] |
| female | 20-30 vs 60+ | -20.8% | 0.338 | -11.35 [-34.59, 11.89] |
| female | 30-40 vs 40-50 | -3.6% | 0.763 | -2.01 [-15.08, 11.06] |
| female | 30-40 vs 50-60 | -4.7% | 0.786 | -2.66 [-21.84, 16.51] |
| female | 30-40 vs 60+ | -1.7% | 0.937 | -0.95 [-24.57, 22.67] |
| female | 40-50 vs 50-60 | -1.2% | 0.942 | -0.65 [-18.26, 16.96] |
| female | 40-50 vs 60+ | +1.9% | 0.926 | 1.06 [-21.31, 23.43] |
| female | 50-60 vs 60+ | +3.1% | 0.899 | 1.71 [-24.70, 28.12] |
| *Mean difference = Group 1 − Group 2 in component units (mmol/24h, L/24h, or pH); % shift = difference relative to Group 2.* | | | | |

potassium_ma_3rd_analysis$wald_tests$sex_within_age_bands

Table 1: **Sex Comparisons Within Age Bands**

| Age Band | % Shift | P (Wald Test) | Mean Diff (95% CI) |
| --- | --- | --- | --- |
| 18+ | +18.2% | 0.621 | 9.58 [-28.37, 47.53] |
| 20-30 | +1.5% | 0.978 | 0.64 [-44.01, 45.29] |
| 30-40 | +25.5% | 0.219 | 13.67 [-8.14, 35.48] |
| 40-50 | +7.9% | 0.536 | 4.37 [-9.49, 18.24] |
| 50-60 | +16.9% | 0.603 | 9.52 [-26.34, 45.39] |
| 60+ | +26.4% | 0.482 | 14.44 [-25.82, 54.70] |
| *Mean difference = Group 1 − Group 2 in component units (mmol/24h, L/24h, or pH); % shift = difference relative to Group 2.* | | | |

#### 6.11 Magnesium

##### 6.11.1 Comparison by Age Band

###### 6.11.1.1 Plot

```
magnesium_ma_2nd_analysis <- sex_age_subdivision_ma("magnesium")
## Not enough studies for multivariable meta-regression (k = 4, need > 6). Running univariable models instead.
magnesium_ma_2nd_analysis$combined_plot
## `height` was translated to `width`.
## `height` was translated to `width`.
## `height` was translated to `width`.
```

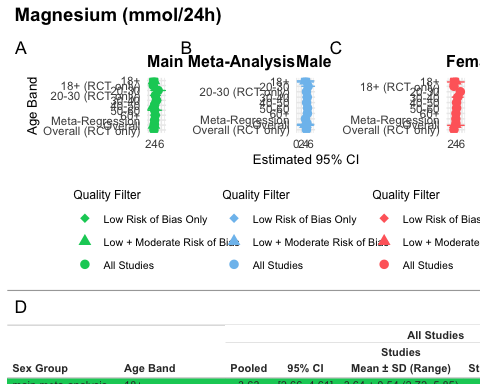

###### 6.11.1.2 Summary Table

magnesium_ma_2nd_analysis$combined_summary_table

Table 1: **Combined Meta-Analysis Results**

24-Hour Urine Magnesium (mmol/24h)

|  |  | **All Studies** | | | | | | **Low Risk of Bias Only** | | | | | | **Low + Moderate RoB** | | | | | |
| --- | --- | --- | --- | --- | --- | --- | --- | --- | --- | --- | --- | --- | --- | --- | --- | --- | --- | --- | --- |
| Sex Group | Age Band | Pooled | 95% CI | Studies  Mean ± SD (Range) | Studies, n | Q p-value | Egger's p | Pooled | 95% CI | Studies  Mean ± SD (Range) | Studies, n | Q p-value | Egger's p | Pooled | 95% CI | Studies  Mean ± SD (Range) | Studies, n | Q p-value | Egger's p |
| main meta-analysis | 18+ | 3.63 | [2.66, 4.61] | 3.64 ± 0.54 (2.72–5.85) | 59 | <0.001 | 0.98 | 3.73 | [3.16, 4.30] | 3.74 ± 0.39 (2.92–4.48) | 28 | <0.001 | 0.71 | 3.70 | [3.04, 4.35] | 3.72 ± 0.42 (2.83–4.48) | 33 | <0.001 | 0.60 |
| **main meta-analysis** | **18+ (RCT only)** | **3.50** | **[2.85, 4.15]** | **3.58 ± 0.49 (2.88–4.30)** | **7** | **<0.05** | **0.11** | **—** | **—** | **—** | **—** | **—** | **—** | **—** | **—** | **—** | **—** | **—** | **—** |
| main meta-analysis | 20-30 | 5.01 | [2.80, 7.21] | 4.86 ± 1.03 (3.60–6.29) | 4 | <0.001 | 0.14 | — | — | — | — | — | — | — | — | — | — | — | — |
| **main meta-analysis** | **20-30 (RCT only)** | **4.16** | **[2.67, 5.65]** | **4.15 ± 0.78 (3.60–4.70)** | **2** | **<0.001** | **—** | **—** | **—** | **—** | **—** | **—** | **—** | **—** | **—** | **—** | **—** | **—** | **—** |
| main meta-analysis | 30-40 | 4.51 | [3.20, 5.82] | 4.44 ± 0.78 (3.33–5.08) | 4 | <0.001 | 0.56 | — | — | — | — | — | — | — | — | — | — | — | — |
| main meta-analysis | 40-50 | 3.94 | [3.29, 4.59] | 3.85 ± 0.39 (3.36–4.28) | 6 | <0.001 | 0.67 | — | — | — | — | — | — | 3.54 | [3.11, 3.97] | — (3.54–3.54) | 1 | — | — |
| main meta-analysis | 50-60 | 3.93 | [1.83, 6.02] | 3.93 ± 1.07 (1.90–4.92) | 6 | <0.001 | 0.91 | — | — | — | — | — | — | — | — | — | — | — | — |
| main meta-analysis | 60+ | 4.11 | [2.76, 5.46] | 4.12 ± 0.69 (2.98–5.07) | 8 | <0.001 | <0.05 | — | — | — | — | — | — | — | — | — | — | — | — |
| **main meta-analysis** | **Meta-Regression** | **3.92** | **[3.06, 4.78]** | **—** | **49** | **<0.001** | **<0.001** | **—** | **—** | **—** | **—** | **—** | **—** | **—** | **—** | **—** | **—** | **—** | **—** |
| **main meta-analysis** | **Overall** | **3.83** | **[2.47, 5.19]** | **3.82 ± 0.70 (1.90–6.29)** | **87** | **<0.001** | **0.36** | **3.73** | **[3.16, 4.30]** | **3.74 ± 0.39 (2.92–4.48)** | **28** | **<0.001** | **0.71** | **3.69** | **[3.05, 4.33]** | **3.71 ± 0.42 (2.83–4.48)** | **34** | **<0.001** | **0.77** |
| **main meta-analysis** | **Overall (RCT only)** | **3.70** | **[2.66, 4.75]** | **3.71 ± 0.56 (2.88–4.70)** | **9** | **<0.001** | **0.14** | **—** | **—** | **—** | **—** | **—** | **—** | **—** | **—** | **—** | **—** | **—** | **—** |
| male | 18+ | 3.77 | [3.22, 4.33] | 3.79 ± 0.31 (3.59–4.25) | 4 | <0.001 | 0.18 | 3.75 | [0.23, 7.26] | — (3.75–3.75) | 1 | — | — | 3.75 | [0.23, 7.26] | — (3.75–3.75) | 1 | — | — |
| **male** | **18+ (RCT only)** | **—** | **—** | **—** | **—** | **—** | **—** | **—** | **—** | **—** | **—** | **—** | **—** | **—** | **—** | **—** | **—** | **—** | **—** |
| male | 20-30 | 4.87 | [2.24, 7.50] | 4.86 ± 1.35 (3.60–6.29) | 3 | <0.001 | 0.34 | — | — | — | — | — | — | — | — | — | — | — | — |
| **male** | **20-30 (RCT only)** | **4.16** | **[2.67, 5.65]** | **4.15 ± 0.78 (3.60–4.70)** | **2** | **<0.001** | **—** | **—** | **—** | **—** | **—** | **—** | **—** | **—** | **—** | **—** | **—** | **—** | **—** |
| male | 30-40 | 4.49 | [2.79, 6.19] | 4.43 ± 0.95 (3.33–5.08) | 3 | <0.001 | 0.82 | — | — | — | — | — | — | — | — | — | — | — | — |
| male | 40-50 | 4.22 | [3.55, 4.89] | 3.80 ± 0.62 (3.36–4.23) | 2 | 0.13 | — | — | — | — | — | — | — | — | — | — | — | — | — |
| male | 50-60 | 4.61 | [3.17, 6.05] | 4.74 ± 0.25 (4.56–4.92) | 2 | 0.08 | — | — | — | — | — | — | — | — | — | — | — | — | — |
| male | 60+ | 4.57 | [3.06, 6.07] | 4.58 ± 0.77 (3.69–5.07) | 3 | <0.001 | 0.30 | — | — | — | — | — | — | — | — | — | — | — | — |
| **male** | **Meta-Regression** | **3.75** | **[2.50, 4.99]** | **—** | **4** | **<0.001** | **0.10** | **—** | **—** | **—** | **—** | **—** | **—** | **—** | **—** | **—** | **—** | **—** | **—** |
| **male** | **Overall** | **4.38** | **[2.85, 5.90]** | **4.34 ± 0.81 (3.33–6.29)** | **17** | **<0.001** | **0.58** | **3.75** | **[0.23, 7.26]** | **— (3.75–3.75)** | **1** | **—** | **—** | **3.75** | **[0.23, 7.26]** | **— (3.75–3.75)** | **1** | **—** | **—** |
| **male** | **Overall (RCT only)** | **4.16** | **[2.67, 5.65]** | **4.15 ± 0.78 (3.60–4.70)** | **2** | **<0.001** | **—** | **—** | **—** | **—** | **—** | **—** | **—** | **—** | **—** | **—** | **—** | **—** | **—** |
| female | 18+ | 3.31 | [2.43, 4.19] | 3.31 ± 0.46 (2.72–3.80) | 7 | <0.001 | 0.91 | 3.80 | [0.68, 6.92] | 3.80 ± 0.00 (3.80–3.80) | 2 | 1.00 | — | 3.60 | [2.93, 4.26] | 3.58 ± 0.39 (3.13–3.80) | 3 | <0.01 | <0.001 |
| **female** | **18+ (RCT only)** | **3.13** | **[1.84, 4.42]** | **— (3.13–3.13)** | **1** | **—** | **—** | **—** | **—** | **—** | **—** | **—** | **—** | **—** | **—** | **—** | **—** | **—** | **—** |
| female | 20-30 | 5.41 | [4.31, 6.51] | — (5.41–5.41) | 1 | — | — | — | — | — | — | — | — | — | — | — | — | — | — |
| **female** | **20-30 (RCT only)** | **—** | **—** | **—** | **—** | **—** | **—** | **—** | **—** | **—** | **—** | **—** | **—** | **—** | **—** | **—** | **—** | **—** | **—** |
| female | 30-40 | 4.47 | [2.55, 6.39] | — (4.47–4.47) | 1 | — | — | — | — | — | — | — | — | — | — | — | — | — | — |
| female | 40-50 | 4.05 | [3.54, 4.56] | 3.99 ± 0.34 (3.62–4.28) | 3 | <0.005 | 0.68 | — | — | — | — | — | — | — | — | — | — | — | — |
| female | 50-60 | 4.08 | [3.44, 4.71] | 4.07 ± 0.32 (3.83–4.43) | 3 | <0.001 | 0.61 | — | — | — | — | — | — | — | — | — | — | — | — |
| female | 60+ | 3.83 | [2.76, 4.90] | 3.84 ± 0.54 (2.98–4.38) | 5 | <0.001 | 0.09 | — | — | — | — | — | — | — | — | — | — | — | — |
| **female** | **Meta-Regression** | **3.80** | **[3.10, 4.50]** | **—** | **7** | **<0.001** | **0.91** | **—** | **—** | **—** | **—** | **—** | **—** | **—** | **—** | **—** | **—** | **—** | **—** |
| **female** | **Overall** | **3.82** | **[2.57, 5.08]** | **3.82 ± 0.65 (2.72–5.41)** | **20** | **<0.001** | **0.94** | **3.80** | **[0.68, 6.92]** | **3.80 ± 0.00 (3.80–3.80)** | **2** | **1.00** | **—** | **3.60** | **[2.93, 4.26]** | **3.58 ± 0.39 (3.13–3.80)** | **3** | **<0.01** | **<0.001** |
| **female** | **Overall (RCT only)** | **3.13** | **[1.84, 4.42]** | **— (3.13–3.13)** | **1** | **—** | **—** | **—** | **—** | **—** | **—** | **—** | **—** | **—** | **—** | **—** | **—** | **—** | **—** |
| *Studies, n = number of studies; Q p-value = Cochran's Q heterogeneity test; Egger's p = linear regression test for funnel plot asymmetry; RoB = Risk of Bias* | | | | | | | | | | | | | | | | | | | |

###### 6.11.1.3 Wald Tests

magnesium_ma_2nd_analysis$wald_tests$sex_comparison

Table 1: **Sex Comparison (Overall)**

| Comparison | % Shift | P (Wald Test) | Mean Diff (95% CI) |
| --- | --- | --- | --- |
| Male vs Female (Overall) | +14.4% | 0.585 | 0.55 [-1.42, 2.52] |
| *Mean difference = Group 1 − Group 2 in component units (mmol/24h, L/24h, or pH); % shift = difference relative to Group 2.* | | | |

magnesium_ma_2nd_analysis$wald_tests$age_band_comparisons

Table 1: **Age Band Comparisons Within Sex Groups**

| Sex Group | Comparison | % Shift | P (Wald Test) | Mean Diff (95% CI) |
| --- | --- | --- | --- | --- |
| main meta-analysis | 18+ vs 20-30 | -27.5% | 0.263 | -1.38 [-3.79, 1.03] |
| main meta-analysis | 18+ vs 30-40 | -19.5% | 0.290 | -0.88 [-2.51, 0.75] |
| main meta-analysis | 18+ vs 40-50 | -7.8% | 0.608 | -0.31 [-1.48, 0.86] |
| main meta-analysis | 18+ vs 50-60 | -7.5% | 0.803 | -0.29 [-2.61, 2.02] |
| main meta-analysis | 18+ vs 60+ | -11.7% | 0.571 | -0.48 [-2.14, 1.18] |
| main meta-analysis | 20-30 vs 30-40 | +11.0% | 0.705 | 0.49 [-2.07, 3.06] |
| main meta-analysis | 20-30 vs 40-50 | +27.2% | 0.362 | 1.07 [-1.23, 3.37] |
| main meta-analysis | 20-30 vs 50-60 | +27.5% | 0.486 | 1.08 [-1.96, 4.12] |
| main meta-analysis | 20-30 vs 60+ | +21.8% | 0.497 | 0.90 [-1.69, 3.48] |
| main meta-analysis | 30-40 vs 40-50 | +14.6% | 0.441 | 0.57 [-0.89, 2.03] |
| main meta-analysis | 30-40 vs 50-60 | +14.9% | 0.642 | 0.59 [-1.89, 3.06] |
| main meta-analysis | 30-40 vs 60+ | +9.7% | 0.676 | 0.40 [-1.48, 2.28] |
| main meta-analysis | 40-50 vs 50-60 | +0.3% | 0.992 | 0.01 [-2.18, 2.21] |
| main meta-analysis | 40-50 vs 60+ | -4.2% | 0.820 | -0.17 [-1.67, 1.32] |
| main meta-analysis | 50-60 vs 60+ | -4.5% | 0.884 | -0.19 [-2.68, 2.31] |
| male | 18+ vs 20-30 | -22.5% | 0.424 | -1.10 [-3.79, 1.59] |
| male | 18+ vs 30-40 | -16.0% | 0.432 | -0.72 [-2.51, 1.07] |
| male | 18+ vs 40-50 | -10.6% | 0.315 | -0.45 [-1.32, 0.43] |
| male | 18+ vs 50-60 | -18.2% | 0.287 | -0.84 [-2.38, 0.71] |
| male | 18+ vs 60+ | -17.4% | 0.331 | -0.80 [-2.40, 0.81] |
| male | 20-30 vs 30-40 | +8.4% | 0.813 | 0.38 [-2.76, 3.51] |
| male | 20-30 vs 40-50 | +15.4% | 0.639 | 0.65 [-2.07, 3.37] |
| male | 20-30 vs 50-60 | +5.6% | 0.866 | 0.26 [-2.74, 3.26] |
| male | 20-30 vs 60+ | +6.6% | 0.846 | 0.30 [-2.73, 3.33] |
| male | 30-40 vs 40-50 | +6.4% | 0.772 | 0.27 [-1.56, 2.10] |
| male | 30-40 vs 50-60 | -2.6% | 0.915 | -0.12 [-2.35, 2.11] |
| male | 30-40 vs 60+ | -1.7% | 0.947 | -0.08 [-2.35, 2.19] |
| male | 40-50 vs 50-60 | -8.5% | 0.629 | -0.39 [-1.98, 1.20] |
| male | 40-50 vs 60+ | -7.6% | 0.679 | -0.35 [-2.00, 1.30] |
| male | 50-60 vs 60+ | +1.0% | 0.967 | 0.04 [-2.04, 2.13] |
| female | 18+ vs 20-30 | -38.9% | <0.01 | -2.10 [-3.51, -0.69] |
| female | 18+ vs 30-40 | -26.0% | 0.281 | -1.16 [-3.28, 0.95] |
| female | 18+ vs 40-50 | -18.4% | 0.152 | -0.74 [-1.76, 0.27] |
| female | 18+ vs 50-60 | -18.8% | 0.165 | -0.77 [-1.85, 0.32] |
| female | 18+ vs 60+ | -13.7% | 0.458 | -0.52 [-1.91, 0.86] |
| female | 20-30 vs 30-40 | +21.0% | 0.405 | 0.94 [-1.27, 3.15] |
| female | 20-30 vs 40-50 | +33.5% | <0.05 | 1.36 [0.15, 2.57] |
| female | 20-30 vs 50-60 | +32.7% | <0.05 | 1.33 [0.07, 2.60] |
| female | 20-30 vs 60+ | +41.2% | <0.05 | 1.58 [0.04, 3.11] |
| female | 30-40 vs 40-50 | +10.3% | 0.680 | 0.42 [-1.57, 2.41] |
| female | 30-40 vs 50-60 | +9.7% | 0.702 | 0.39 [-1.63, 2.42] |
| female | 30-40 vs 60+ | +16.6% | 0.570 | 0.64 [-1.56, 2.84] |
| female | 40-50 vs 50-60 | -0.6% | 0.954 | -0.02 [-0.84, 0.79] |
| female | 40-50 vs 60+ | +5.7% | 0.717 | 0.22 [-0.97, 1.41] |
| female | 50-60 vs 60+ | +6.4% | 0.701 | 0.24 [-1.00, 1.49] |
| *Mean difference = Group 1 − Group 2 in component units (mmol/24h, L/24h, or pH); % shift = difference relative to Group 2.* | | | | |

magnesium_ma_2nd_analysis$wald_tests$sex_within_age_bands

Table 1: **Sex Comparisons Within Age Bands**

| Age Band | % Shift | P (Wald Test) | Mean Diff (95% CI) |
| --- | --- | --- | --- |
| 18+ | +14.1% | 0.381 | 0.47 [-0.58, 1.51] |
| 20-30 | -10.0% | 0.711 | -0.54 [-3.39, 2.31] |
| 30-40 | +0.5% | 0.987 | 0.02 [-2.54, 2.59] |
| 40-50 | +4.1% | 0.696 | 0.17 [-0.68, 1.01] |
| 50-60 | +13.2% | 0.504 | 0.54 [-1.04, 2.11] |
| 60+ | +19.2% | 0.435 | 0.74 [-1.11, 2.58] |
| *Mean difference = Group 1 − Group 2 in component units (mmol/24h, L/24h, or pH); % shift = difference relative to Group 2.* | | | |

###### 6.11.1.4 Meta-Regression (plain language)

format_meta_reg_plain("magnesium", sex_group = "all")

Message

No meta-regression result for magnesium / all

###### 6.11.1.5 Assay Subgroup Analysis

assay_subgroup_table("magnesium")

Table 1: **Assay subgroup analysis — Magnesium**

*Per-assay pooled estimates; categories with k < 3 listed but not pooled.*

| Assay category | Studies, n | Participants | 95% CI | I² | Q p-value | Egger's p | Note |
| --- | --- | --- | --- | --- | --- | --- | --- |
| Unreported | 67 | 8,602 | 3.85 [3.67, 4.03] | 98% | <0.001 | 0.568 |  |
| Atomic absorption | 15 | 420 | 3.77 [3.39, 4.15] | 93% | <0.001 | 0.477 |  |
| Colourimetric (other) | 3 | 287 | 3.59 [3.57, 3.62] | 0% | 0.674 | <0.001 |  |
| Autoanalyser | 1 | 107 | — | — | — | — | k = 1 < min_k = 3 (skipped) |
| ICP / ICP-MS | 1 | 22 | — | — | — | — | k = 1 < min_k = 3 (skipped) |
| Xylidyl blue | 1 | 75 | — | — | — | — | k = 1 < min_k = 3 (skipped) |

###### 6.11.1.6 Preservative Sensitivity (Litholink / acidified samples)

```
preservative_sensitivity_ma("magnesium")
## # A tibble: 3 × 6
##   group               n_studies n_participants pooled_estimate lower_95 upper_95
##   <chr>                   <int>          <int>           <dbl>    <dbl>    <dbl>
## 1 All studies                87           9513            3.83     3.67     3.98
## 2 Excluding preserva…        85           9452            3.82     3.66     3.98
## 3 Preservative only           2             61            4.15     3.65     4.66
```

###### 6.11.1.7 Creatinine Completeness Sensitivity

creat_completeness_table("magnesium")

Table 1: **Creatinine-completeness sensitivity — Magnesium**

*Compares the all-studies pooled estimate against subsets defined by whether the study reported 24h urine creatinine. Studies that do not report creatinine cannot verify collection completeness; a substantial shift here flags possible bias from incomplete collections.*

| Subset | Studies, n | Participants | Range (95% CI) | Δ vs all-studies (%) | Flag |
| --- | --- | --- | --- | --- | --- |
| All studies | 87 | 9,513 | 3.83 [3.67, 3.98] | — | — |
| Creatinine reported only | 61 | 7,801 | 3.91 [3.74, 4.08] | +2.1% | OK |
| Creatinine NOT reported | 26 | 1,712 | 3.63 [3.32, 3.95] | -5.1% | OK |

##### 6.11.2 Comparison by Age / Sex / race

###### 6.11.2.1 Plot

```
magnesium_ma_3rd_analysis <- sex_age_race_subdivision_ma("magnesium",
                                                      urine_ma2,
                                                      overall_bias)
## Not enough studies for multivariable meta-regression (k = 4, need > 6). Running univariable models instead.
magnesium_ma_3rd_analysis$race_plot
## `height` was translated to `width`.
## `height` was translated to `width`.
## `height` was translated to `width`.
```

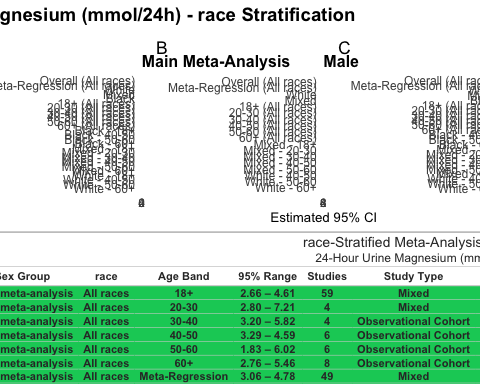

###### 6.11.2.2 Combined MA Results

magnesium_ma_3rd_analysis$combined_summary_table

Table 1: **Combined Meta-Analysis Results**

24-Hour Urine Magnesium (mmol/24h)

| Sex Group | race | Age Band | Pooled | 95% Range | Studies Mean SD Range | Studies | Study Type | Participants | I² | Cochran's Q | Q (p-value) | Egger's p | Model |
| --- | --- | --- | --- | --- | --- | --- | --- | --- | --- | --- | --- | --- | --- |
| main meta-analysis | All | 18+ | 3.63 | 2.66 – 4.61 | 3.64 ± 0.54 (2.72–5.85) | 59 | Mixed | 5075 | 92.8% | 805.9 | <0.005 | 0.98 | Random |
| main meta-analysis | All | 20-30 | 5.01 | 2.80 – 7.21 | 4.86 ± 1.03 (3.60–6.29) | 4 | Mixed | 101 | 98.5% | 195.2 | <0.005 | 0.14 | Random |
| main meta-analysis | All | 30-40 | 4.51 | 3.20 – 5.82 | 4.44 ± 0.78 (3.33–5.08) | 4 | Observational Cohort | 168 | 88.8% | 26.8 | <0.005 | 0.56 | Random |
| main meta-analysis | All | 40-50 | 3.94 | 3.29 – 4.59 | 3.85 ± 0.39 (3.36–4.28) | 6 | Observational Cohort | 643 | 97% | 169.1 | <0.005 | 0.67 | Random |
| main meta-analysis | All | 50-60 | 3.93 | 1.83 – 6.02 | 3.93 ± 1.07 (1.90–4.92) | 6 | Observational Cohort | 2701 | 99.7% | 1773.1 | <0.005 | 0.91 | Random |
| main meta-analysis | All | 60+ | 4.11 | 2.76 – 5.46 | 4.12 ± 0.69 (2.98–5.07) | 8 | Observational Cohort | 825 | 98.7% | 539.2 | <0.005 | <0.05 | Random |
| **main meta-analysis** | **All** | **Meta-Regression** | **3.92** | **3.06 – 4.78** | **—** | **49** | **Mixed** | **4724** | **88.7%** | **306** | **<0.005** | **<0.001** | **Multivariable** |
| **main meta-analysis** | **All** | **Meta-Regression** | **3.92** | **3.06 – 4.78** | **—** | **49** | **Mixed** | **4724** | **88.7%** | **306** | **<0.005** | **<0.001** | **Multivariable** |
| **main meta-analysis** | **All** | **Overall** | **3.83** | **2.47 – 5.19** | **3.82 ± 0.70 (1.90–6.29)** | **87** | **Mixed** | **9513** | **99.1%** | **9600.7** | **<0.005** | **0.36** | **Random** |
| **main meta-analysis** | **All** | **Overall** | **3.83** | **2.47 – 5.19** | **3.82 ± 0.70 (1.90–6.29)** | **87** | **Mixed** | **9513** | **99.1%** | **9600.7** | **<0.005** | **0.36** | **Random** |
| main meta-analysis | black | 18+ | 3.47 | 0.49 – 6.45 | 3.47 ± 0.00 (3.47–3.47) | 3 | Observational Cohort | 51 | 0% | 0 | 1.00 | — | Fixed |
| main meta-analysis | black | 20-30 | — | NA – NA | — | NA | NA | NA | — | — | — | — | NA |
| main meta-analysis | black | 30-40 | — | NA – NA | — | NA | NA | NA | — | — | — | — | NA |
| main meta-analysis | black | 40-50 | 3.62 | 1.22 – 6.01 | — (3.62–3.62) | 1 | Observational Cohort | 29 | — | — | — | — | Single study |
| main meta-analysis | black | 50-60 | 3.94 | 1.12 – 6.77 | — (3.94–3.94) | 1 | Observational Cohort | 131 | — | — | — | — | Single study |
| main meta-analysis | black | 60+ | 3.82 | 0.64 – 7.00 | 3.83 ± 0.07 (3.79–3.88) | 2 | Observational Cohort | 144 | 0% | 0.1 | 0.74 | — | Fixed |
| **main meta-analysis** | **black** | **Meta-Regression** | **—** | **NA – NA** | **—** | **NA** | **NA** | **NA** | **—** | **—** | **—** | **—** | **NA** |
| **main meta-analysis** | **black** | **Overall** | **3.80** | **0.87 – 6.72** | **3.66 ± 0.21 (3.47–3.94)** | **7** | **Observational Cohort** | **355** | **0%** | **4.5** | **0.61** | **<0.01** | **Fixed** |
| main meta-analysis | mixed | 18+ | 3.68 | 2.64 – 4.71 | 3.69 ± 0.57 (2.72–5.85) | 49 | Mixed | 4819 | 93.9% | 793.1 | <0.005 | 0.73 | Random |
| main meta-analysis | mixed | 20-30 | 5.01 | 2.80 – 7.21 | 4.86 ± 1.03 (3.60–6.29) | 4 | Mixed | 101 | 98.5% | 195.2 | <0.005 | 0.14 | Random |
| main meta-analysis | mixed | 30-40 | 4.51 | 3.20 – 5.82 | 4.44 ± 0.78 (3.33–5.08) | 4 | Observational Cohort | 168 | 88.8% | 26.8 | <0.005 | 0.56 | Random |
| main meta-analysis | mixed | 40-50 | 3.95 | 3.24 – 4.65 | 3.95 ± 0.36 (3.54–4.23) | 3 | Observational Cohort | 84 | 98.4% | 125.7 | <0.005 | 0.18 | Random |
| main meta-analysis | mixed | 50-60 | 3.43 | 0.74 – 6.12 | 3.43 ± 1.37 (1.90–4.56) | 3 | Observational Cohort | 82 | 99.8% | 1063.5 | <0.005 | 0.69 | Random |
| main meta-analysis | mixed | 60+ | 2.98 | 2.63 – 3.33 | — (2.98–2.98) | 1 | Observational Cohort | 25 | — | — | — | — | Single study |
| **main meta-analysis** | **mixed** | **Meta-Regression** | **—** | **NA – NA** | **—** | **NA** | **NA** | **NA** | **—** | **—** | **—** | **—** | **NA** |
| **main meta-analysis** | **mixed** | **Overall** | **3.81** | **2.37 – 5.25** | **3.81 ± 0.74 (1.90–6.29)** | **64** | **Mixed** | **5279** | **99.3%** | **9166.9** | **<0.005** | **0.37** | **Random** |
| main meta-analysis | white | 18+ | 3.42 | 0.07 – 6.78 | 3.42 ± 0.02 (3.41–3.44) | 6 | Observational Cohort | 165 | 0% | 0 | 1.00 | <0.001 | Fixed |
| main meta-analysis | white | 20-30 | — | NA – NA | — | NA | NA | NA | — | — | — | — | NA |
| main meta-analysis | white | 30-40 | — | NA – NA | — | NA | NA | NA | — | — | — | — | NA |
| main meta-analysis | white | 40-50 | 4.27 | 1.34 – 7.21 | 3.82 ± 0.65 (3.36–4.28) | 2 | Observational Cohort | 530 | 60.5% | 2.5 | 0.11 | — | Fixed |
| main meta-analysis | white | 50-60 | 4.64 | 4.02 – 5.26 | 4.68 ± 0.34 (4.43–4.92) | 2 | Observational Cohort | 2488 | 84.3% | 6.4 | <0.05 | — | Random |
| main meta-analysis | white | 60+ | 4.45 | 3.35 – 5.55 | 4.46 ± 0.57 (3.69–5.07) | 5 | Observational Cohort | 656 | 97% | 131.2 | <0.005 | 0.13 | Random |
| **main meta-analysis** | **white** | **Meta-Regression** | **—** | **NA – NA** | **—** | **NA** | **NA** | **NA** | **—** | **—** | **—** | **—** | **NA** |
| **main meta-analysis** | **white** | **Overall** | **4.06** | **2.89 – 5.23** | **3.99 ± 0.65 (3.36–5.07)** | **15** | **Observational Cohort** | **3839** | **93.4%** | **212.9** | **<0.005** | **0.31** | **Random** |
| male | All | 18+ | 3.77 | 3.22 – 4.33 | 3.79 ± 0.31 (3.59–4.25) | 4 | Observational Cohort | 302 | 86.4% | 22.1 | <0.005 | 0.18 | Random |
| male | All | 20-30 | 4.87 | 2.24 – 7.50 | 4.86 ± 1.35 (3.60–6.29) | 3 | Mixed | 71 | 99% | 195 | <0.005 | 0.34 | Random |
| male | All | 30-40 | 4.49 | 2.79 – 6.19 | 4.43 ± 0.95 (3.33–5.08) | 3 | Observational Cohort | 143 | 91.1% | 22.5 | <0.005 | 0.82 | Random |
| male | All | 40-50 | 4.22 | 3.55 – 4.89 | 3.80 ± 0.62 (3.36–4.23) | 2 | Observational Cohort | 31 | 55.5% | 2.2 | 0.13 | — | Fixed |
| male | All | 50-60 | 4.61 | 3.17 – 6.05 | 4.74 ± 0.25 (4.56–4.92) | 2 | Observational Cohort | 104 | 67.4% | 3.1 | 0.08 | — | Fixed |
| male | All | 60+ | 4.57 | 3.06 – 6.07 | 4.58 ± 0.77 (3.69–5.07) | 3 | Observational Cohort | 329 | 98.4% | 125.1 | <0.005 | 0.30 | Random |
| **male** | **All** | **Meta-Regression** | **3.75** | **2.50 – 4.99** | **—** | **4** | **Mixed** | **262** | **97.1%** | **86** | **<0.005** | **0.10** | **Univariable** |
| **male** | **All** | **Overall** | **4.38** | **2.85 – 5.90** | **4.34 ± 0.81 (3.33–6.29)** | **17** | **Mixed** | **980** | **99.7%** | **5622.9** | **<0.005** | **0.58** | **Random** |
| male | black | 18+ | — | NA – NA | — | NA | NA | NA | — | — | — | — | NA |
| male | black | 20-30 | — | NA – NA | — | NA | NA | NA | — | — | — | — | NA |
| male | black | 30-40 | — | NA – NA | — | NA | NA | NA | — | — | — | — | NA |
| male | black | 40-50 | — | NA – NA | — | NA | NA | NA | — | — | — | — | NA |
| male | black | 50-60 | — | NA – NA | — | NA | NA | NA | — | — | — | — | NA |
| male | black | 60+ | — | NA – NA | — | NA | NA | NA | — | — | — | — | NA |
| **male** | **black** | **Meta-Regression** | **—** | **NA – NA** | **—** | **NA** | **NA** | **NA** | **—** | **—** | **—** | **—** | **NA** |
| **male** | **black** | **Overall** | **—** | **NA – NA** | **—** | **NA** | **NA** | **NA** | **—** | **—** | **—** | **—** | **NA** |
| male | mixed | 18+ | 3.77 | 3.22 – 4.33 | 3.79 ± 0.31 (3.59–4.25) | 4 | Observational Cohort | 302 | 86.4% | 22.1 | <0.005 | 0.18 | Random |
| male | mixed | 20-30 | 4.87 | 2.24 – 7.50 | 4.86 ± 1.35 (3.60–6.29) | 3 | Mixed | 71 | 99% | 195 | <0.005 | 0.34 | Random |
| male | mixed | 30-40 | 4.49 | 2.79 – 6.19 | 4.43 ± 0.95 (3.33–5.08) | 3 | Observational Cohort | 143 | 91.1% | 22.5 | <0.005 | 0.82 | Random |
| male | mixed | 40-50 | 4.23 | 3.62 – 4.84 | — (4.23–4.23) | 1 | Observational Cohort | 25 | — | — | — | — | Single study |
| male | mixed | 50-60 | 4.56 | 3.80 – 5.32 | — (4.56–4.56) | 1 | Observational Cohort | 25 | — | — | — | — | Single study |
| male | mixed | 60+ | — | NA – NA | — | NA | NA | NA | — | — | — | — | NA |
| **male** | **mixed** | **Meta-Regression** | **—** | **NA – NA** | **—** | **NA** | **NA** | **NA** | **—** | **—** | **—** | **—** | **NA** |
| **male** | **mixed** | **Overall** | **4.34** | **2.71 – 5.96** | **4.32 ± 0.85 (3.33–6.29)** | **12** | **Mixed** | **566** | **99.8%** | **5478.1** | **<0.005** | **0.67** | **Random** |
| male | white | 18+ | — | NA – NA | — | NA | NA | NA | — | — | — | — | NA |
| male | white | 20-30 | — | NA – NA | — | NA | NA | NA | — | — | — | — | NA |
| male | white | 30-40 | — | NA – NA | — | NA | NA | NA | — | — | — | — | NA |
| male | white | 40-50 | 3.36 | 0.59 – 6.13 | — (3.36–3.36) | 1 | Observational Cohort | 6 | — | — | — | — | Single study |
| male | white | 50-60 | 4.92 | 1.63 – 8.21 | — (4.92–4.92) | 1 | Observational Cohort | 79 | — | — | — | — | Single study |
| male | white | 60+ | 4.57 | 3.06 – 6.07 | 4.58 ± 0.77 (3.69–5.07) | 3 | Observational Cohort | 329 | 98.4% | 125.1 | <0.005 | 0.30 | Random |
| **male** | **white** | **Meta-Regression** | **—** | **NA – NA** | **—** | **NA** | **NA** | **NA** | **—** | **—** | **—** | **—** | **NA** |
| **male** | **white** | **Overall** | **4.48** | **3.08 – 5.87** | **4.40 ± 0.81 (3.36–5.07)** | **5** | **Observational Cohort** | **414** | **97.2%** | **144.5** | **<0.005** | **0.34** | **Random** |
| female | All | 18+ | 3.31 | 2.43 – 4.19 | 3.31 ± 0.46 (2.72–3.80) | 7 | Mixed | 488 | 91.9% | 74.4 | <0.005 | 0.91 | Random |
| female | All | 20-30 | 5.41 | 4.31 – 6.51 | — (5.41–5.41) | 1 | Observational Cohort | 25 | — | — | — | — | Single study |
| female | All | 30-40 | 4.47 | 2.55 – 6.39 | — (4.47–4.47) | 1 | Observational Cohort | 25 | — | — | — | — | Single study |
| female | All | 40-50 | 4.05 | 3.54 – 4.56 | 3.99 ± 0.34 (3.62–4.28) | 3 | Observational Cohort | 578 | 82.7% | 11.6 | <0.005 | 0.68 | Random |
| female | All | 50-60 | 4.08 | 3.44 – 4.71 | 4.07 ± 0.32 (3.83–4.43) | 3 | Observational Cohort | 2565 | 98.1% | 106.4 | <0.005 | 0.61 | Random |
| female | All | 60+ | 3.83 | 2.76 – 4.90 | 3.84 ± 0.54 (2.98–4.38) | 5 | Observational Cohort | 496 | 98.2% | 224.5 | <0.005 | 0.09 | Random |
| **female** | **All** | **Meta-Regression** | **3.80** | **3.10 – 4.50** | **—** | **7** | **Mixed** | **488** | **86.3%** | **26.2** | **<0.005** | **0.91** | **Multivariable** |
| **female** | **All** | **Overall** | **3.82** | **2.57 – 5.08** | **3.82 ± 0.65 (2.72–5.41)** | **20** | **Mixed** | **4177** | **98.6%** | **1362** | **<0.005** | **0.94** | **Random** |
| female | black | 18+ | — | NA – NA | — | NA | NA | NA | — | — | — | — | NA |
| female | black | 20-30 | — | NA – NA | — | NA | NA | NA | — | — | — | — | NA |
| female | black | 30-40 | — | NA – NA | — | NA | NA | NA | — | — | — | — | NA |
| female | black | 40-50 | 3.62 | 1.22 – 6.01 | — (3.62–3.62) | 1 | Observational Cohort | 29 | — | — | — | — | Single study |
| female | black | 50-60 | 3.94 | 1.12 – 6.77 | — (3.94–3.94) | 1 | Observational Cohort | 131 | — | — | — | — | Single study |
| female | black | 60+ | 3.82 | 0.64 – 7.00 | 3.83 ± 0.07 (3.79–3.88) | 2 | Observational Cohort | 144 | 0% | 0.1 | 0.74 | — | Fixed |
| **female** | **black** | **Meta-Regression** | **—** | **NA – NA** | **—** | **NA** | **NA** | **NA** | **—** | **—** | **—** | **—** | **NA** |
| **female** | **black** | **Overall** | **3.85** | **0.93 – 6.76** | **3.81 ± 0.14 (3.62–3.94)** | **4** | **Observational Cohort** | **304** | **0%** | **1.8** | **0.62** | **0.29** | **Fixed** |
| female | mixed | 18+ | 3.31 | 2.43 – 4.19 | 3.31 ± 0.46 (2.72–3.80) | 7 | Mixed | 488 | 91.9% | 74.4 | <0.005 | 0.91 | Random |
| female | mixed | 20-30 | 5.41 | 4.31 – 6.51 | — (5.41–5.41) | 1 | Observational Cohort | 25 | — | — | — | — | Single study |
| female | mixed | 30-40 | 4.47 | 2.55 – 6.39 | — (4.47–4.47) | 1 | Observational Cohort | 25 | — | — | — | — | Single study |
| female | mixed | 40-50 | 4.08 | 3.59 – 4.57 | — (4.08–4.08) | 1 | Observational Cohort | 25 | — | — | — | — | Single study |
| female | mixed | 50-60 | 3.83 | 3.34 – 4.32 | — (3.83–3.83) | 1 | Observational Cohort | 25 | — | — | — | — | Single study |
| female | mixed | 60+ | 2.98 | 2.63 – 3.33 | — (2.98–2.98) | 1 | Observational Cohort | 25 | — | — | — | — | Single study |
| **female** | **mixed** | **Meta-Regression** | **—** | **NA – NA** | **—** | **NA** | **NA** | **NA** | **—** | **—** | **—** | **—** | **NA** |
| **female** | **mixed** | **Overall** | **3.66** | **2.17 – 5.15** | **3.66 ± 0.77 (2.72–5.41)** | **12** | **Mixed** | **613** | **98.6%** | **785.3** | **<0.005** | **0.50** | **Random** |
| female | white | 18+ | — | NA – NA | — | NA | NA | NA | — | — | — | — | NA |
| female | white | 20-30 | — | NA – NA | — | NA | NA | NA | — | — | — | — | NA |
| female | white | 30-40 | — | NA – NA | — | NA | NA | NA | — | — | — | — | NA |
| female | white | 40-50 | 4.28 | 1.35 – 7.22 | — (4.28–4.28) | 1 | Observational Cohort | 524 | — | — | — | — | Single study |
| female | white | 50-60 | 4.43 | 1.22 – 7.64 | — (4.43–4.43) | 1 | Observational Cohort | 2409 | — | — | — | — | Single study |
| female | white | 60+ | 4.33 | 1.22 – 7.43 | 4.28 ± 0.15 (4.18–4.38) | 2 | Observational Cohort | 327 | 10.6% | 1.1 | 0.29 | — | Fixed |
| **female** | **white** | **Meta-Regression** | **—** | **NA – NA** | **—** | **NA** | **NA** | **NA** | **—** | **—** | **—** | **—** | **NA** |
| **female** | **white** | **Overall** | **4.39** | **1.24 – 7.55** | **4.32 ± 0.11 (4.18–4.43)** | **4** | **Observational Cohort** | **3260** | **49.6%** | **5.9** | **0.11** | **0.22** | **Fixed** |
| *I² represents heterogeneity; Q test assesses between-study variation; Egger's p = linear regression test for funnel plot asymmetry; Meta-Regression shows covariate-adjusted estimates* | | | | | | | | | | | | | |

###### 6.11.2.3 Table of Results subdivided by race / Sex / Age

magnesium_ma_3rd_analysis$race_summary_table

Table 1: **race-Stratified Meta-Analysis Results**

24-Hour Urine Magnesium (mmol/24h)

| Sex Group | race | Age Band | 95% Range | Studies | Study Type | Participants | I² | Cochran's Q | Q (p-value) | Egger's p | Model |
| --- | --- | --- | --- | --- | --- | --- | --- | --- | --- | --- | --- |
| **main meta-analysis** | **All races** | **18+** | **2.66 – 4.61** | **59** | **Mixed** | **5075** | **92.8%** | **805.9** | **<0.005** | **0.98** | **Random** |
| **main meta-analysis** | **All races** | **20-30** | **2.80 – 7.21** | **4** | **Mixed** | **101** | **98.5%** | **195.2** | **<0.005** | **0.14** | **Random** |
| **main meta-analysis** | **All races** | **30-40** | **3.20 – 5.82** | **4** | **Observational Cohort** | **168** | **88.8%** | **26.8** | **<0.005** | **0.56** | **Random** |
| **main meta-analysis** | **All races** | **40-50** | **3.29 – 4.59** | **6** | **Observational Cohort** | **643** | **97%** | **169.1** | **<0.005** | **0.67** | **Random** |
| **main meta-analysis** | **All races** | **50-60** | **1.83 – 6.02** | **6** | **Observational Cohort** | **2701** | **99.7%** | **1773.1** | **<0.005** | **0.91** | **Random** |
| **main meta-analysis** | **All races** | **60+** | **2.76 – 5.46** | **8** | **Observational Cohort** | **825** | **98.7%** | **539.2** | **<0.005** | **<0.05** | **Random** |
| **main meta-analysis** | **All races** | **Meta-Regression** | **3.06 – 4.78** | **49** | **Mixed** | **4724** | **88.7%** | **306** | **<0.005** | **<0.001** | **Multivariable** |
| **main meta-analysis** | **All races** | **Meta-Regression** | **3.06 – 4.78** | **49** | **Mixed** | **4724** | **88.7%** | **306** | **<0.005** | **<0.001** | **Multivariable** |
| **main meta-analysis** | **All races** | **Overall** | **2.47 – 5.19** | **87** | **Mixed** | **9513** | **99.1%** | **9600.7** | **<0.005** | **0.36** | **Random** |
| **main meta-analysis** | **All races** | **Overall** | **2.47 – 5.19** | **87** | **Mixed** | **9513** | **99.1%** | **9600.7** | **<0.005** | **0.36** | **Random** |
| main meta-analysis | Black | 18+ | 0.49 – 6.45 | 3 | Observational Cohort | 51 | 0% | 0 | 1.00 | — | Fixed |
| main meta-analysis | Black | 20-30 | — | — | NA | — | — | — | — | — | — |
| main meta-analysis | Black | 30-40 | — | — | NA | — | — | — | — | — | — |
| main meta-analysis | Black | 40-50 | 1.22 – 6.01 | 1 | Observational Cohort | 29 | — | — | — | — | Single study |
| main meta-analysis | Black | 50-60 | 1.12 – 6.77 | 1 | Observational Cohort | 131 | — | — | — | — | Single study |
| main meta-analysis | Black | 60+ | 0.64 – 7.00 | 2 | Observational Cohort | 144 | 0% | 0.1 | 0.74 | — | Fixed |
| main meta-analysis | Black | Meta-Regression | — | — | NA | — | — | — | — | — | — |
| **main meta-analysis** | **Black** | **Overall** | **0.87 – 6.72** | **7** | **Observational Cohort** | **355** | **0%** | **4.5** | **0.61** | **<0.01** | **Fixed** |
| main meta-analysis | Mixed | 18+ | 2.64 – 4.71 | 49 | Mixed | 4819 | 93.9% | 793.1 | <0.005 | 0.73 | Random |
| main meta-analysis | Mixed | 20-30 | 2.80 – 7.21 | 4 | Mixed | 101 | 98.5% | 195.2 | <0.005 | 0.14 | Random |
| main meta-analysis | Mixed | 30-40 | 3.20 – 5.82 | 4 | Observational Cohort | 168 | 88.8% | 26.8 | <0.005 | 0.56 | Random |
| main meta-analysis | Mixed | 40-50 | 3.24 – 4.65 | 3 | Observational Cohort | 84 | 98.4% | 125.7 | <0.005 | 0.18 | Random |
| main meta-analysis | Mixed | 50-60 | 0.74 – 6.12 | 3 | Observational Cohort | 82 | 99.8% | 1063.5 | <0.005 | 0.69 | Random |
| main meta-analysis | Mixed | 60+ | 2.63 – 3.33 | 1 | Observational Cohort | 25 | — | — | — | — | Single study |
| main meta-analysis | Mixed | Meta-Regression | — | — | NA | — | — | — | — | — | — |
| **main meta-analysis** | **Mixed** | **Overall** | **2.37 – 5.25** | **64** | **Mixed** | **5279** | **99.3%** | **9166.9** | **<0.005** | **0.37** | **Random** |
| main meta-analysis | White | 18+ | 0.07 – 6.78 | 6 | Observational Cohort | 165 | 0% | 0 | 1.00 | <0.001 | Fixed |
| main meta-analysis | White | 20-30 | — | — | NA | — | — | — | — | — | — |
| main meta-analysis | White | 30-40 | — | — | NA | — | — | — | — | — | — |
| main meta-analysis | White | 40-50 | 1.34 – 7.21 | 2 | Observational Cohort | 530 | 60.5% | 2.5 | 0.11 | — | Fixed |
| main meta-analysis | White | 50-60 | 4.02 – 5.26 | 2 | Observational Cohort | 2488 | 84.3% | 6.4 | <0.05 | — | Random |
| main meta-analysis | White | 60+ | 3.35 – 5.55 | 5 | Observational Cohort | 656 | 97% | 131.2 | <0.005 | 0.13 | Random |
| main meta-analysis | White | Meta-Regression | — | — | NA | — | — | — | — | — | — |
| **main meta-analysis** | **White** | **Overall** | **2.89 – 5.23** | **15** | **Observational Cohort** | **3839** | **93.4%** | **212.9** | **<0.005** | **0.31** | **Random** |
| **male** | **All races** | **18+** | **3.22 – 4.33** | **4** | **Observational Cohort** | **302** | **86.4%** | **22.1** | **<0.005** | **0.18** | **Random** |
| **male** | **All races** | **20-30** | **2.24 – 7.50** | **3** | **Mixed** | **71** | **99%** | **195** | **<0.005** | **0.34** | **Random** |
| **male** | **All races** | **30-40** | **2.79 – 6.19** | **3** | **Observational Cohort** | **143** | **91.1%** | **22.5** | **<0.005** | **0.82** | **Random** |
| **male** | **All races** | **40-50** | **3.55 – 4.89** | **2** | **Observational Cohort** | **31** | **55.5%** | **2.2** | **0.13** | **—** | **Fixed** |
| **male** | **All races** | **50-60** | **3.17 – 6.05** | **2** | **Observational Cohort** | **104** | **67.4%** | **3.1** | **0.08** | **—** | **Fixed** |
| **male** | **All races** | **60+** | **3.06 – 6.07** | **3** | **Observational Cohort** | **329** | **98.4%** | **125.1** | **<0.005** | **0.30** | **Random** |
| **male** | **All races** | **Meta-Regression** | **2.50 – 4.99** | **4** | **Mixed** | **262** | **97.1%** | **86** | **<0.005** | **0.10** | **Univariable** |
| **male** | **All races** | **Overall** | **2.85 – 5.90** | **17** | **Mixed** | **980** | **99.7%** | **5622.9** | **<0.005** | **0.58** | **Random** |
| male | Black | 18+ | — | — | NA | — | — | — | — | — | — |
| male | Black | 20-30 | — | — | NA | — | — | — | — | — | — |
| male | Black | 30-40 | — | — | NA | — | — | — | — | — | — |
| male | Black | 40-50 | — | — | NA | — | — | — | — | — | — |
| male | Black | 50-60 | — | — | NA | — | — | — | — | — | — |
| male | Black | 60+ | — | — | NA | — | — | — | — | — | — |
| male | Black | Meta-Regression | — | — | NA | — | — | — | — | — | — |
| **male** | **Black** | **Overall** | **—** | **—** | **NA** | **—** | **—** | **—** | **—** | **—** | **—** |
| male | Mixed | 18+ | 3.22 – 4.33 | 4 | Observational Cohort | 302 | 86.4% | 22.1 | <0.005 | 0.18 | Random |
| male | Mixed | 20-30 | 2.24 – 7.50 | 3 | Mixed | 71 | 99% | 195 | <0.005 | 0.34 | Random |
| male | Mixed | 30-40 | 2.79 – 6.19 | 3 | Observational Cohort | 143 | 91.1% | 22.5 | <0.005 | 0.82 | Random |
| male | Mixed | 40-50 | 3.62 – 4.84 | 1 | Observational Cohort | 25 | — | — | — | — | Single study |
| male | Mixed | 50-60 | 3.80 – 5.32 | 1 | Observational Cohort | 25 | — | — | — | — | Single study |
| male | Mixed | 60+ | — | — | NA | — | — | — | — | — | — |
| male | Mixed | Meta-Regression | — | — | NA | — | — | — | — | — | — |
| **male** | **Mixed** | **Overall** | **2.71 – 5.96** | **12** | **Mixed** | **566** | **99.8%** | **5478.1** | **<0.005** | **0.67** | **Random** |
| male | White | 18+ | — | — | NA | — | — | — | — | — | — |
| male | White | 20-30 | — | — | NA | — | — | — | — | — | — |
| male | White | 30-40 | — | — | NA | — | — | — | — | — | — |
| male | White | 40-50 | 0.59 – 6.13 | 1 | Observational Cohort | 6 | — | — | — | — | Single study |
| male | White | 50-60 | 1.63 – 8.21 | 1 | Observational Cohort | 79 | — | — | — | — | Single study |
| male | White | 60+ | 3.06 – 6.07 | 3 | Observational Cohort | 329 | 98.4% | 125.1 | <0.005 | 0.30 | Random |
| male | White | Meta-Regression | — | — | NA | — | — | — | — | — | — |
| **male** | **White** | **Overall** | **3.08 – 5.87** | **5** | **Observational Cohort** | **414** | **97.2%** | **144.5** | **<0.005** | **0.34** | **Random** |
| **female** | **All races** | **18+** | **2.43 – 4.19** | **7** | **Mixed** | **488** | **91.9%** | **74.4** | **<0.005** | **0.91** | **Random** |
| **female** | **All races** | **20-30** | **4.31 – 6.51** | **1** | **Observational Cohort** | **25** | **—** | **—** | **—** | **—** | **Single study** |
| **female** | **All races** | **30-40** | **2.55 – 6.39** | **1** | **Observational Cohort** | **25** | **—** | **—** | **—** | **—** | **Single study** |
| **female** | **All races** | **40-50** | **3.54 – 4.56** | **3** | **Observational Cohort** | **578** | **82.7%** | **11.6** | **<0.005** | **0.68** | **Random** |
| **female** | **All races** | **50-60** | **3.44 – 4.71** | **3** | **Observational Cohort** | **2565** | **98.1%** | **106.4** | **<0.005** | **0.61** | **Random** |
| **female** | **All races** | **60+** | **2.76 – 4.90** | **5** | **Observational Cohort** | **496** | **98.2%** | **224.5** | **<0.005** | **0.09** | **Random** |
| **female** | **All races** | **Meta-Regression** | **3.10 – 4.50** | **7** | **Mixed** | **488** | **86.3%** | **26.2** | **<0.005** | **0.91** | **Multivariable** |
| **female** | **All races** | **Overall** | **2.57 – 5.08** | **20** | **Mixed** | **4177** | **98.6%** | **1362** | **<0.005** | **0.94** | **Random** |
| female | Black | 18+ | — | — | NA | — | — | — | — | — | — |
| female | Black | 20-30 | — | — | NA | — | — | — | — | — | — |
| female | Black | 30-40 | — | — | NA | — | — | — | — | — | — |
| female | Black | 40-50 | 1.22 – 6.01 | 1 | Observational Cohort | 29 | — | — | — | — | Single study |
| female | Black | 50-60 | 1.12 – 6.77 | 1 | Observational Cohort | 131 | — | — | — | — | Single study |
| female | Black | 60+ | 0.64 – 7.00 | 2 | Observational Cohort | 144 | 0% | 0.1 | 0.74 | — | Fixed |
| female | Black | Meta-Regression | — | — | NA | — | — | — | — | — | — |
| **female** | **Black** | **Overall** | **0.93 – 6.76** | **4** | **Observational Cohort** | **304** | **0%** | **1.8** | **0.62** | **0.29** | **Fixed** |
| female | Mixed | 18+ | 2.43 – 4.19 | 7 | Mixed | 488 | 91.9% | 74.4 | <0.005 | 0.91 | Random |
| female | Mixed | 20-30 | 4.31 – 6.51 | 1 | Observational Cohort | 25 | — | — | — | — | Single study |
| female | Mixed | 30-40 | 2.55 – 6.39 | 1 | Observational Cohort | 25 | — | — | — | — | Single study |
| female | Mixed | 40-50 | 3.59 – 4.57 | 1 | Observational Cohort | 25 | — | — | — | — | Single study |
| female | Mixed | 50-60 | 3.34 – 4.32 | 1 | Observational Cohort | 25 | — | — | — | — | Single study |
| female | Mixed | 60+ | 2.63 – 3.33 | 1 | Observational Cohort | 25 | — | — | — | — | Single study |
| female | Mixed | Meta-Regression | — | — | NA | — | — | — | — | — | — |
| **female** | **Mixed** | **Overall** | **2.17 – 5.15** | **12** | **Mixed** | **613** | **98.6%** | **785.3** | **<0.005** | **0.50** | **Random** |
| female | White | 18+ | — | — | NA | — | — | — | — | — | — |
| female | White | 20-30 | — | — | NA | — | — | — | — | — | — |
| female | White | 30-40 | — | — | NA | — | — | — | — | — | — |
| female | White | 40-50 | 1.35 – 7.22 | 1 | Observational Cohort | 524 | — | — | — | — | Single study |
| female | White | 50-60 | 1.22 – 7.64 | 1 | Observational Cohort | 2409 | — | — | — | — | Single study |
| female | White | 60+ | 1.22 – 7.43 | 2 | Observational Cohort | 327 | 10.6% | 1.1 | 0.29 | — | Fixed |
| female | White | Meta-Regression | — | — | NA | — | — | — | — | — | — |
| **female** | **White** | **Overall** | **1.24 – 7.55** | **4** | **Observational Cohort** | **3260** | **49.6%** | **5.9** | **0.11** | **0.22** | **Fixed** |
| *I² represents heterogeneity; Q test assesses between-study variation; Egger's p = linear regression test for funnel plot asymmetry; — indicates insufficient data* | | | | | | | | | | | |

###### 6.11.2.4 Wald Tests

magnesium_ma_3rd_analysis$wald_tests$sex_comparison

Table 1: **Sex Comparison (Overall)**

| Comparison | % Shift | P (Wald Test) | Mean Diff (95% CI) |
| --- | --- | --- | --- |
| Male vs Female (Overall) | +14.4% | 0.585 | 0.55 [-1.42, 2.52] |
| *Mean difference = Group 1 − Group 2 in component units (mmol/24h, L/24h, or pH); % shift = difference relative to Group 2.* | | | |

magnesium_ma_3rd_analysis$wald_tests$race_comparisons

Table 1: **race Comparisons Within Sex Groups**

| Sex Group | Comparison | % Shift | P (Wald Test) | Mean Diff (95% CI) |
| --- | --- | --- | --- | --- |
| male | black vs mixed | — | — | — |
| male | black vs white | — | — | — |
| male | mixed vs white | -3.1% | 0.898 | -0.14 [-2.28, 2.00] |
| female | black vs mixed | +5.1% | 0.910 | 0.19 [-3.09, 3.46] |
| female | black vs white | -12.4% | 0.803 | -0.55 [-4.84, 3.75] |
| female | mixed vs white | -16.7% | 0.680 | -0.73 [-4.22, 2.75] |
| *Mean difference = Group 1 − Group 2 in component units (mmol/24h, L/24h, or pH); % shift = difference relative to Group 2.* | | | | |

magnesium_ma_3rd_analysis$wald_tests$age_band_comparisons

Table 1: **Age Band Comparisons Within Sex Groups**

| Sex Group | Comparison | % Shift | P (Wald Test) | Mean Diff (95% CI) |
| --- | --- | --- | --- | --- |
| main meta-analysis | 18+ vs 20-30 | -27.5% | 0.263 | -1.38 [-3.79, 1.03] |
| main meta-analysis | 18+ vs 30-40 | -19.5% | 0.290 | -0.88 [-2.51, 0.75] |
| main meta-analysis | 18+ vs 40-50 | -7.8% | 0.608 | -0.31 [-1.48, 0.86] |
| main meta-analysis | 18+ vs 50-60 | -7.5% | 0.803 | -0.29 [-2.61, 2.02] |
| main meta-analysis | 18+ vs 60+ | -11.7% | 0.571 | -0.48 [-2.14, 1.18] |
| main meta-analysis | 20-30 vs 30-40 | +11.0% | 0.705 | 0.49 [-2.07, 3.06] |
| main meta-analysis | 20-30 vs 40-50 | +27.2% | 0.362 | 1.07 [-1.23, 3.37] |
| main meta-analysis | 20-30 vs 50-60 | +27.5% | 0.486 | 1.08 [-1.96, 4.12] |
| main meta-analysis | 20-30 vs 60+ | +21.8% | 0.497 | 0.90 [-1.69, 3.48] |
| main meta-analysis | 30-40 vs 40-50 | +14.6% | 0.441 | 0.57 [-0.89, 2.03] |
| main meta-analysis | 30-40 vs 50-60 | +14.9% | 0.642 | 0.59 [-1.89, 3.06] |
| main meta-analysis | 30-40 vs 60+ | +9.7% | 0.676 | 0.40 [-1.48, 2.28] |
| main meta-analysis | 40-50 vs 50-60 | +0.3% | 0.992 | 0.01 [-2.18, 2.21] |
| main meta-analysis | 40-50 vs 60+ | -4.2% | 0.820 | -0.17 [-1.67, 1.32] |
| main meta-analysis | 50-60 vs 60+ | -4.5% | 0.884 | -0.19 [-2.68, 2.31] |
| male | 18+ vs 20-30 | -22.5% | 0.424 | -1.10 [-3.79, 1.59] |
| male | 18+ vs 30-40 | -16.0% | 0.432 | -0.72 [-2.51, 1.07] |
| male | 18+ vs 40-50 | -10.6% | 0.315 | -0.45 [-1.32, 0.43] |
| male | 18+ vs 50-60 | -18.2% | 0.287 | -0.84 [-2.38, 0.71] |
| male | 18+ vs 60+ | -17.4% | 0.331 | -0.80 [-2.40, 0.81] |
| male | 20-30 vs 30-40 | +8.4% | 0.813 | 0.38 [-2.76, 3.51] |
| male | 20-30 vs 40-50 | +15.4% | 0.639 | 0.65 [-2.07, 3.37] |
| male | 20-30 vs 50-60 | +5.6% | 0.866 | 0.26 [-2.74, 3.26] |
| male | 20-30 vs 60+ | +6.6% | 0.846 | 0.30 [-2.73, 3.33] |
| male | 30-40 vs 40-50 | +6.4% | 0.772 | 0.27 [-1.56, 2.10] |
| male | 30-40 vs 50-60 | -2.6% | 0.915 | -0.12 [-2.35, 2.11] |
| male | 30-40 vs 60+ | -1.7% | 0.947 | -0.08 [-2.35, 2.19] |
| male | 40-50 vs 50-60 | -8.5% | 0.629 | -0.39 [-1.98, 1.20] |
| male | 40-50 vs 60+ | -7.6% | 0.679 | -0.35 [-2.00, 1.30] |
| male | 50-60 vs 60+ | +1.0% | 0.967 | 0.04 [-2.04, 2.13] |
| female | 18+ vs 20-30 | -38.9% | <0.01 | -2.10 [-3.51, -0.69] |
| female | 18+ vs 30-40 | -26.0% | 0.281 | -1.16 [-3.28, 0.95] |
| female | 18+ vs 40-50 | -18.4% | 0.152 | -0.74 [-1.76, 0.27] |
| female | 18+ vs 50-60 | -18.8% | 0.165 | -0.77 [-1.85, 0.32] |
| female | 18+ vs 60+ | -13.7% | 0.458 | -0.52 [-1.91, 0.86] |
| female | 20-30 vs 30-40 | +21.0% | 0.405 | 0.94 [-1.27, 3.15] |
| female | 20-30 vs 40-50 | +33.5% | <0.05 | 1.36 [0.15, 2.57] |
| female | 20-30 vs 50-60 | +32.7% | <0.05 | 1.33 [0.07, 2.60] |
| female | 20-30 vs 60+ | +41.2% | <0.05 | 1.58 [0.04, 3.11] |
| female | 30-40 vs 40-50 | +10.3% | 0.680 | 0.42 [-1.57, 2.41] |
| female | 30-40 vs 50-60 | +9.7% | 0.702 | 0.39 [-1.63, 2.42] |
| female | 30-40 vs 60+ | +16.6% | 0.570 | 0.64 [-1.56, 2.84] |
| female | 40-50 vs 50-60 | -0.6% | 0.954 | -0.02 [-0.84, 0.79] |
| female | 40-50 vs 60+ | +5.7% | 0.717 | 0.22 [-0.97, 1.41] |
| female | 50-60 vs 60+ | +6.4% | 0.701 | 0.24 [-1.00, 1.49] |
| *Mean difference = Group 1 − Group 2 in component units (mmol/24h, L/24h, or pH); % shift = difference relative to Group 2.* | | | | |

magnesium_ma_3rd_analysis$wald_tests$sex_within_age_bands

Table 1: **Sex Comparisons Within Age Bands**

| Age Band | % Shift | P (Wald Test) | Mean Diff (95% CI) |
| --- | --- | --- | --- |
| 18+ | +14.1% | 0.381 | 0.47 [-0.58, 1.51] |
| 20-30 | -10.0% | 0.711 | -0.54 [-3.39, 2.31] |
| 30-40 | +0.5% | 0.987 | 0.02 [-2.54, 2.59] |
| 40-50 | +4.1% | 0.696 | 0.17 [-0.68, 1.01] |
| 50-60 | +13.2% | 0.504 | 0.54 [-1.04, 2.11] |
| 60+ | +19.2% | 0.435 | 0.74 [-1.11, 2.58] |
| *Mean difference = Group 1 − Group 2 in component units (mmol/24h, L/24h, or pH); % shift = difference relative to Group 2.* | | | |

#### 6.12 Ammonium

##### 6.12.1 Comparison by Age Band

###### 6.12.1.1 Plot

```
ammonium_ma_2nd_analysis <- sex_age_subdivision_ma("ammonium")
## Meta-regression failed for male: No studies to combine in meta-analysis.
## Not enough studies for multivariable meta-regression (k = 5, need > 6). Running univariable models instead.
ammonium_ma_2nd_analysis$combined_plot
## `height` was translated to `width`.
## `height` was translated to `width`.
## `height` was translated to `width`.
```

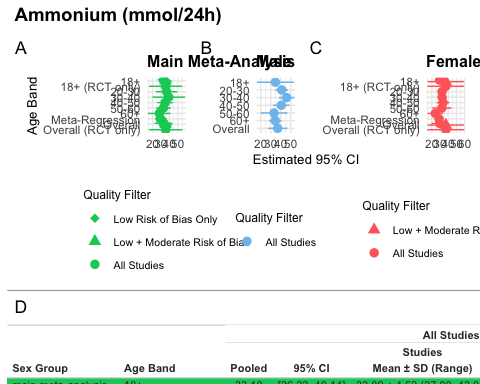

###### 6.12.1.2 Summary Table

ammonium_ma_2nd_analysis$combined_summary_table

Table 1: **Combined Meta-Analysis Results**

24-Hour Urine Ammonium (mmol/24h)

|  |  | **All Studies** | | | | | | **Low Risk of Bias Only** | | | | | | **Low + Moderate RoB** | | | | | |
| --- | --- | --- | --- | --- | --- | --- | --- | --- | --- | --- | --- | --- | --- | --- | --- | --- | --- | --- | --- |
| Sex Group | Age Band | Pooled | 95% CI | Studies  Mean ± SD (Range) | Studies, n | Q p-value | Egger's p | Pooled | 95% CI | Studies  Mean ± SD (Range) | Studies, n | Q p-value | Egger's p | Pooled | 95% CI | Studies  Mean ± SD (Range) | Studies, n | Q p-value | Egger's p |
| main meta-analysis | 18+ | 33.18 | [26.22, 40.14] | 33.89 ± 4.52 (27.00–43.00) | 23 | <0.001 | <0.05 | 35.10 | [27.74, 42.45] | 35.04 ± 3.85 (28.00–39.00) | 7 | <0.001 | 0.71 | 35.29 | [28.35, 42.23] | 35.29 ± 3.63 (28.00–39.00) | 8 | <0.001 | 0.63 |
| **main meta-analysis** | **18+ (RCT only)** | **35.57** | **[16.72, 54.42]** | **35.75 ± 2.08 (33.35–37.00)** | **3** | **0.54** | **0.68** | **—** | **—** | **—** | **—** | **—** | **—** | **—** | **—** | **—** | **—** | **—** | **—** |
| main meta-analysis | 20-30 | 36.68 | [24.73, 48.62] | 36.69 ± 6.12 (32.36–41.01) | 2 | <0.001 | — | — | — | — | — | — | — | — | — | — | — | — | — |
| main meta-analysis | 30-40 | 39.13 | [20.54, 57.72] | 39.14 ± 9.50 (32.42–45.86) | 2 | <0.001 | — | — | — | — | — | — | — | — | — | — | — | — | — |
| main meta-analysis | 40-50 | 34.96 | [26.39, 43.52] | 34.47 ± 4.40 (30.21–40.69) | 5 | <0.001 | 0.66 | — | — | — | — | — | — | — | — | — | — | — | — |
| main meta-analysis | 50-60 | 32.88 | [24.84, 40.91] | 32.94 ± 4.15 (28.63–38.71) | 5 | <0.001 | 0.21 | — | — | — | — | — | — | — | — | — | — | — | — |
| main meta-analysis | 60+ | 28.10 | [15.53, 40.67] | 28.12 ± 6.45 (20.50–37.63) | 8 | <0.001 | <0.05 | — | — | — | — | — | — | — | — | — | — | — | — |
| **main meta-analysis** | **Meta-Regression** | **29.27** | **[23.61, 34.93]** | **—** | **16** | **<0.001** | **0.15** | **—** | **—** | **—** | **—** | **—** | **—** | **—** | **—** | **—** | **—** | **—** | **—** |
| **main meta-analysis** | **Overall** | **32.92** | **[22.48, 43.35]** | **33.18 ± 5.51 (20.50–45.86)** | **45** | **<0.001** | **0.08** | **35.10** | **[27.74, 42.45]** | **35.04 ± 3.85 (28.00–39.00)** | **7** | **<0.001** | **0.71** | **35.29** | **[28.35, 42.23]** | **35.29 ± 3.63 (28.00–39.00)** | **8** | **<0.001** | **0.63** |
| **main meta-analysis** | **Overall (RCT only)** | **35.57** | **[16.72, 54.42]** | **35.75 ± 2.08 (33.35–37.00)** | **3** | **0.54** | **0.68** | **—** | **—** | **—** | **—** | **—** | **—** | **—** | **—** | **—** | **—** | **—** | **—** |
| male | 18+ | 34.70 | [17.26, 52.14] | — (34.70–34.70) | 1 | — | — | — | — | — | — | — | — | — | — | — | — | — | — |
| **male** | **18+ (RCT only)** | **—** | **—** | **—** | **—** | **—** | **—** | **—** | **—** | **—** | **—** | **—** | **—** | **—** | **—** | **—** | **—** | **—** | **—** |
| male | 20-30 | 41.01 | [35.40, 46.62] | — (41.01–41.01) | 1 | — | — | — | — | — | — | — | — | — | — | — | — | — | — |
| male | 30-40 | 45.86 | [38.94, 52.78] | — (45.86–45.86) | 1 | — | — | — | — | — | — | — | — | — | — | — | — | — | — |
| male | 40-50 | 40.52 | [33.21, 47.83] | 35.84 ± 6.85 (31.00–40.69) | 2 | 0.06 | — | — | — | — | — | — | — | — | — | — | — | — | — |
| male | 50-60 | 33.34 | [20.82, 45.86] | 34.01 ± 1.32 (33.08–34.94) | 2 | 0.30 | — | — | — | — | — | — | — | — | — | — | — | — | — |
| male | 60+ | 34.58 | [29.00, 40.16] | 34.46 ± 3.07 (31.50–37.63) | 3 | <0.001 | 0.46 | — | — | — | — | — | — | — | — | — | — | — | — |
| **male** | **Meta-Regression** | **—** | **—** | **—** | **—** | **—** | **—** | **—** | **—** | **—** | **—** | **—** | **—** | **—** | **—** | **—** | **—** | **—** | **—** |
| **male** | **Overall** | **36.87** | **[27.96, 45.78]** | **36.47 ± 4.77 (31.00–45.86)** | **10** | **<0.001** | **0.29** | **—** | **—** | **—** | **—** | **—** | **—** | **—** | **—** | **—** | **—** | **—** | **—** |
| **male** | **Overall (RCT only)** | **—** | **—** | **—** | **—** | **—** | **—** | **—** | **—** | **—** | **—** | **—** | **—** | **—** | **—** | **—** | **—** | **—** | **—** |
| female | 18+ | 29.50 | [25.47, 33.53] | 30.20 ± 4.09 (27.00–37.00) | 5 | <0.05 | 0.37 | — | — | — | — | — | — | 37.00 | [15.44, 58.56] | — (37.00–37.00) | 1 | — | — |
| **female** | **18+ (RCT only)** | **37.00** | **[15.44, 58.56]** | **— (37.00–37.00)** | **1** | **—** | **—** | **—** | **—** | **—** | **—** | **—** | **—** | **—** | **—** | **—** | **—** | **—** | **—** |
| female | 20-30 | 32.36 | [27.93, 36.79] | — (32.36–32.36) | 1 | — | — | — | — | — | — | — | — | — | — | — | — | — | — |
| female | 30-40 | 32.42 | [27.87, 36.97] | — (32.42–32.42) | 1 | — | — | — | — | — | — | — | — | — | — | — | — | — | — |
| female | 40-50 | 33.55 | [26.50, 40.60] | 33.55 ± 3.48 (30.21–37.15) | 3 | <0.001 | 0.73 | — | — | — | — | — | — | — | — | — | — | — | — |
| female | 50-60 | 32.21 | [21.30, 43.13] | 32.22 ± 5.63 (28.63–38.71) | 3 | <0.001 | 0.57 | — | — | — | — | — | — | — | — | — | — | — | — |
| female | 60+ | 24.30 | [15.65, 32.95] | 24.32 ± 4.46 (20.50–30.74) | 5 | <0.001 | 0.19 | — | — | — | — | — | — | — | — | — | — | — | — |
| **female** | **Meta-Regression** | **28.76** | **[24.73, 32.79]** | **—** | **5** | **<0.05** | **0.37** | **—** | **—** | **—** | **—** | **—** | **—** | **—** | **—** | **—** | **—** | **—** | **—** |
| **female** | **Overall** | **29.60** | **[19.68, 39.52]** | **29.71 ± 5.19 (20.50–38.71)** | **18** | **<0.001** | **0.66** | **—** | **—** | **—** | **—** | **—** | **—** | **37.00** | **[15.44, 58.56]** | **— (37.00–37.00)** | **1** | **—** | **—** |
| **female** | **Overall (RCT only)** | **37.00** | **[15.44, 58.56]** | **— (37.00–37.00)** | **1** | **—** | **—** | **—** | **—** | **—** | **—** | **—** | **—** | **—** | **—** | **—** | **—** | **—** | **—** |
| *Studies, n = number of studies; Q p-value = Cochran's Q heterogeneity test; Egger's p = linear regression test for funnel plot asymmetry; RoB = Risk of Bias* | | | | | | | | | | | | | | | | | | | |

###### 6.12.1.3 Wald Tests

ammonium_ma_2nd_analysis$wald_tests$sex_comparison

Table 1: **Sex Comparison (Overall)**

| Comparison | % Shift | P (Wald Test) | Mean Diff (95% CI) |
| --- | --- | --- | --- |
| Male vs Female (Overall) | +24.6% | 0.285 | 7.27 [-6.07, 20.61] |
| *Mean difference = Group 1 − Group 2 in component units (mmol/24h, L/24h, or pH); % shift = difference relative to Group 2.* | | | |

ammonium_ma_2nd_analysis$wald_tests$age_band_comparisons

Table 1: **Age Band Comparisons Within Sex Groups**

| Sex Group | Comparison | % Shift | P (Wald Test) | Mean Diff (95% CI) |
| --- | --- | --- | --- | --- |
| main meta-analysis | 18+ vs 20-30 | -9.5% | 0.620 | -3.50 [-17.32, 10.33] |
| main meta-analysis | 18+ vs 30-40 | -15.2% | 0.557 | -5.95 [-25.80, 13.90] |
| main meta-analysis | 18+ vs 40-50 | -5.1% | 0.752 | -1.78 [-12.81, 9.26] |
| main meta-analysis | 18+ vs 50-60 | +0.9% | 0.955 | 0.31 [-10.32, 10.94] |
| main meta-analysis | 18+ vs 60+ | +18.1% | 0.488 | 5.08 [-9.29, 19.45] |
| main meta-analysis | 20-30 vs 30-40 | -6.3% | 0.828 | -2.45 [-24.55, 19.65] |
| main meta-analysis | 20-30 vs 40-50 | +4.9% | 0.818 | 1.72 [-12.98, 16.42] |
| main meta-analysis | 20-30 vs 50-60 | +11.6% | 0.605 | 3.80 [-10.59, 18.20] |
| main meta-analysis | 20-30 vs 60+ | +30.5% | 0.332 | 8.58 [-8.77, 25.92] |
| main meta-analysis | 30-40 vs 40-50 | +11.9% | 0.689 | 4.17 [-16.29, 24.64] |
| main meta-analysis | 30-40 vs 50-60 | +19.0% | 0.545 | 6.25 [-14.00, 26.51] |
| main meta-analysis | 30-40 vs 60+ | +39.2% | 0.336 | 11.03 [-11.41, 33.47] |
| main meta-analysis | 40-50 vs 50-60 | +6.3% | 0.728 | 2.08 [-9.66, 13.82] |
| main meta-analysis | 40-50 vs 60+ | +24.4% | 0.377 | 6.85 [-8.36, 22.06] |
| main meta-analysis | 50-60 vs 60+ | +17.0% | 0.531 | 4.77 [-10.15, 19.69] |
| male | 18+ vs 20-30 | -15.4% | 0.500 | -6.31 [-24.63, 12.01] |
| male | 18+ vs 30-40 | -24.3% | 0.244 | -11.16 [-29.93, 7.61] |
| male | 18+ vs 40-50 | -14.4% | 0.547 | -5.82 [-24.73, 13.10] |
| male | 18+ vs 50-60 | +4.1% | 0.901 | 1.36 [-20.12, 22.83] |
| male | 18+ vs 60+ | +0.3% | 0.990 | 0.12 [-18.20, 18.43] |
| male | 20-30 vs 30-40 | -10.6% | 0.286 | -4.85 [-13.75, 4.05] |
| male | 20-30 vs 40-50 | +1.2% | 0.917 | 0.49 [-8.72, 9.70] |
| male | 20-30 vs 50-60 | +23.0% | 0.273 | 7.67 [-6.05, 21.39] |
| male | 20-30 vs 60+ | +18.6% | 0.111 | 6.43 [-1.48, 14.34] |
| male | 30-40 vs 40-50 | +13.2% | 0.298 | 5.34 [-4.72, 15.41] |
| male | 30-40 vs 50-60 | +37.5% | 0.086 | 12.52 [-1.79, 26.82] |
| male | 30-40 vs 60+ | +32.6% | <0.05 | 11.28 [2.39, 20.17] |
| male | 40-50 vs 50-60 | +21.5% | 0.332 | 7.18 [-7.32, 21.68] |
| male | 40-50 vs 60+ | +17.2% | 0.206 | 5.94 [-3.26, 15.14] |
| male | 50-60 vs 60+ | -3.6% | 0.860 | -1.24 [-14.95, 12.47] |
| female | 18+ vs 20-30 | -8.8% | 0.349 | -2.86 [-8.85, 3.13] |
| female | 18+ vs 30-40 | -9.0% | 0.346 | -2.92 [-9.00, 3.16] |
| female | 18+ vs 40-50 | -12.1% | 0.329 | -4.05 [-12.17, 4.07] |
| female | 18+ vs 50-60 | -8.4% | 0.648 | -2.71 [-14.35, 8.92] |
| female | 18+ vs 60+ | +21.4% | 0.286 | 5.20 [-4.34, 14.74] |
| female | 20-30 vs 30-40 | -0.2% | 0.985 | -0.06 [-6.41, 6.29] |
| female | 20-30 vs 40-50 | -3.5% | 0.780 | -1.19 [-9.52, 7.14] |
| female | 20-30 vs 50-60 | +0.5% | 0.980 | 0.15 [-11.63, 11.93] |
| female | 20-30 vs 60+ | +33.2% | 0.104 | 8.06 [-1.66, 17.78] |
| female | 30-40 vs 40-50 | -3.4% | 0.792 | -1.13 [-9.52, 7.26] |
| female | 30-40 vs 50-60 | +0.6% | 0.972 | 0.21 [-11.62, 12.03] |
| female | 30-40 vs 60+ | +33.4% | 0.104 | 8.12 [-1.66, 17.89] |
| female | 40-50 vs 50-60 | +4.2% | 0.840 | 1.34 [-11.66, 14.33] |
| female | 40-50 vs 60+ | +38.1% | 0.104 | 9.25 [-1.91, 20.41] |
| female | 50-60 vs 60+ | +32.5% | 0.266 | 7.91 [-6.02, 21.84] |
| *Mean difference = Group 1 − Group 2 in component units (mmol/24h, L/24h, or pH); % shift = difference relative to Group 2.* | | | | |

ammonium_ma_2nd_analysis$wald_tests$sex_within_age_bands

Table 1: **Sex Comparisons Within Age Bands**

| Age Band | % Shift | P (Wald Test) | Mean Diff (95% CI) |
| --- | --- | --- | --- |
| 18+ | +17.6% | 0.569 | 5.20 [-12.70, 23.10] |
| 20-30 | +26.7% | <0.05 | 8.65 [1.51, 15.79] |
| 30-40 | +41.5% | <0.01 | 13.44 [5.16, 21.72] |
| 40-50 | +20.8% | 0.179 | 6.97 [-3.19, 17.13] |
| 50-60 | +3.5% | 0.894 | 1.13 [-15.48, 17.74] |
| 60+ | +42.3% | 0.050 | 10.28 [-0.02, 20.57] |
| *Mean difference = Group 1 − Group 2 in component units (mmol/24h, L/24h, or pH); % shift = difference relative to Group 2.* | | | |

###### 6.12.1.4 Meta-Regression (plain language)

format_meta_reg_plain("ammonium", sex_group = "all")

Message

No meta-regression result for ammonium / all

###### 6.12.1.5 Assay Subgroup Analysis

assay_subgroup_table("ammonium")

Table 1: **Assay subgroup analysis — Ammonium**

*Per-assay pooled estimates; categories with k < 3 listed but not pooled.*

| Assay category | Studies, n | Participants | 95% CI | I² | Q p-value | Egger's p | Note |
| --- | --- | --- | --- | --- | --- | --- | --- |
| Unreported | 36 | 4,481 | 32.78 [30.80, 34.76] | 99% | <0.001 | 0.121 |  |
| Glutamate dehydrogenase | 6 | 242 | 32.58 [29.95, 35.20] | 57% | 0.040 | 0.505 |  |
| Autoanalyser | 2 | 106 | — | — | — | — | k = 2 < min_k = 3 (skipped) |
| Enzymatic | 1 | 12 | — | — | — | — | k = 1 < min_k = 3 (skipped) |

###### 6.12.1.6 Preservative Sensitivity (Litholink / acidified samples)

```
preservative_sensitivity_ma("ammonium")
## # A tibble: 3 × 6
##   group               n_studies n_participants pooled_estimate lower_95 upper_95
##   <chr>                   <int>          <int>           <dbl>    <dbl>    <dbl>
## 1 All studies                45           4841            32.9     31.3     34.6
## 2 Excluding preserva…        45           4841            32.9     31.3     34.6
## 3 Preservative only           0              0            NA       NA       NA
```

###### 6.12.1.7 Creatinine Completeness Sensitivity

creat_completeness_table("ammonium")

Table 1: **Creatinine-completeness sensitivity — Ammonium**

*Compares the all-studies pooled estimate against subsets defined by whether the study reported 24h urine creatinine. Studies that do not report creatinine cannot verify collection completeness; a substantial shift here flags possible bias from incomplete collections.*

| Subset | Studies, n | Participants | Range (95% CI) | Δ vs all-studies (%) | Flag |
| --- | --- | --- | --- | --- | --- |
| All studies | 45 | 4,841 | 32.92 [31.26, 34.57] | — | — |
| Creatinine reported only | 35 | 4,519 | 32.89 [30.93, 34.85] | -0.1% | OK |
| Creatinine NOT reported | 10 | 322 | 32.54 [29.99, 35.08] | -1.2% | OK |

##### 6.12.2 Comparison by Age / Sex / race

###### 6.12.2.1 Plot

```
ammonium_ma_3rd_analysis <- sex_age_race_subdivision_ma("ammonium",
                                                      urine_ma2,
                                                      overall_bias)
## Meta-regression failed for male: No studies to combine in meta-analysis.
## Not enough studies for multivariable meta-regression (k = 5, need > 6). Running univariable models instead.
ammonium_ma_3rd_analysis$race_plot
## `height` was translated to `width`.
## `height` was translated to `width`.
## `height` was translated to `width`.
```

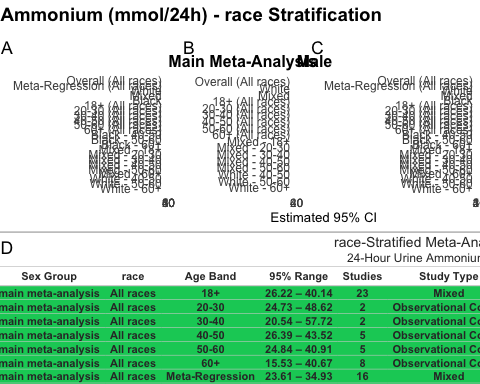

###### 6.12.2.2 Combined MA Results

ammonium_ma_3rd_analysis$combined_summary_table

Table 1: **Combined Meta-Analysis Results**

24-Hour Urine Ammonium (mmol/24h)

| Sex Group | race | Age Band | Pooled | 95% Range | Studies Mean SD Range | Studies | Study Type | Participants | I² | Cochran's Q | Q (p-value) | Egger's p | Model |
| --- | --- | --- | --- | --- | --- | --- | --- | --- | --- | --- | --- | --- | --- |
| main meta-analysis | All | 18+ | 33.18 | 26.22 – 40.14 | 33.89 ± 4.52 (27.00–43.00) | 23 | Mixed | 638 | 76.3% | 92.6 | <0.005 | <0.05 | Random |
| main meta-analysis | All | 20-30 | 36.68 | 24.73 – 48.62 | 36.69 ± 6.12 (32.36–41.01) | 2 | Observational Cohort | 50 | 99.3% | 140.8 | <0.005 | — | Random |
| main meta-analysis | All | 30-40 | 39.13 | 20.54 – 57.72 | 39.14 ± 9.50 (32.42–45.86) | 2 | Observational Cohort | 50 | 99.6% | 253.1 | <0.005 | — | Random |
| main meta-analysis | All | 40-50 | 34.96 | 26.39 – 43.52 | 34.47 ± 4.40 (30.21–40.69) | 5 | Observational Cohort | 609 | 98% | 203.9 | <0.005 | 0.66 | Random |
| main meta-analysis | All | 50-60 | 32.88 | 24.84 – 40.91 | 32.94 ± 4.15 (28.63–38.71) | 5 | Observational Cohort | 2669 | 97.6% | 164.1 | <0.005 | 0.21 | Random |
| main meta-analysis | All | 60+ | 28.10 | 15.53 – 40.67 | 28.12 ± 6.45 (20.50–37.63) | 8 | Observational Cohort | 825 | 99% | 686.2 | <0.005 | <0.05 | Random |
| **main meta-analysis** | **All** | **Meta-Regression** | **29.27** | **23.61 – 34.93** | **—** | **16** | **Mixed** | **349** | **68.5%** | **39.1** | **<0.005** | **0.15** | **Multivariable** |
| **main meta-analysis** | **All** | **Meta-Regression** | **29.27** | **23.61 – 34.93** | **—** | **16** | **Mixed** | **349** | **68.5%** | **39.1** | **<0.005** | **0.15** | **Multivariable** |
| **main meta-analysis** | **All** | **Overall** | **32.92** | **22.48 – 43.35** | **33.18 ± 5.51 (20.50–45.86)** | **45** | **Mixed** | **4841** | **98.4%** | **2767.6** | **<0.005** | **0.08** | **Random** |
| **main meta-analysis** | **All** | **Overall** | **32.92** | **22.48 – 43.35** | **33.18 ± 5.51 (20.50–45.86)** | **45** | **Mixed** | **4841** | **98.4%** | **2767.6** | **<0.005** | **0.08** | **Random** |
| main meta-analysis | black | 18+ | — | NA – NA | — | NA | NA | NA | — | — | — | — | NA |
| main meta-analysis | black | 20-30 | — | NA – NA | — | NA | NA | NA | — | — | — | — | NA |
| main meta-analysis | black | 30-40 | — | NA – NA | — | NA | NA | NA | — | — | — | — | NA |
| main meta-analysis | black | 40-50 | 33.29 | 10.63 – 55.95 | — (33.29–33.29) | 1 | Observational Cohort | 29 | — | — | — | — | Single study |
| main meta-analysis | black | 50-60 | 28.63 | 8.87 – 48.39 | — (28.63–28.63) | 1 | Observational Cohort | 131 | — | — | — | — | Single study |
| main meta-analysis | black | 60+ | 20.50 | 13.56 – 27.44 | 20.50 ± 0.00 (20.50–20.50) | 2 | Observational Cohort | 144 | 0% | 0 | 1.00 | — | Fixed |
| **main meta-analysis** | **black** | **Meta-Regression** | **—** | **NA – NA** | **—** | **NA** | **NA** | **NA** | **—** | **—** | **—** | **—** | **NA** |
| **main meta-analysis** | **black** | **Overall** | **25.51** | **13.60 – 37.43** | **25.73 ± 6.33 (20.50–33.29)** | **4** | **Observational Cohort** | **304** | **97.2%** | **107.2** | **<0.005** | **0.11** | **Random** |
| main meta-analysis | mixed | 18+ | 33.18 | 26.22 – 40.14 | 33.89 ± 4.52 (27.00–43.00) | 23 | Mixed | 638 | 76.3% | 92.6 | <0.005 | <0.05 | Random |
| main meta-analysis | mixed | 20-30 | 36.68 | 24.73 – 48.62 | 36.69 ± 6.12 (32.36–41.01) | 2 | Observational Cohort | 50 | 99.3% | 140.8 | <0.005 | — | Random |
| main meta-analysis | mixed | 30-40 | 39.13 | 20.54 – 57.72 | 39.14 ± 9.50 (32.42–45.86) | 2 | Observational Cohort | 50 | 99.6% | 253.1 | <0.005 | — | Random |
| main meta-analysis | mixed | 40-50 | 38.92 | 34.19 – 43.65 | 38.92 ± 2.50 (37.15–40.69) | 2 | Observational Cohort | 50 | 93% | 14.2 | <0.005 | — | Random |
| main meta-analysis | mixed | 50-60 | 35.88 | 28.21 – 43.55 | 35.89 ± 3.98 (33.08–38.71) | 2 | Observational Cohort | 50 | 96.6% | 29.4 | <0.005 | — | Random |
| main meta-analysis | mixed | 60+ | 30.74 | 22.86 – 38.62 | — (30.74–30.74) | 1 | Observational Cohort | 25 | — | — | — | — | Single study |
| **main meta-analysis** | **mixed** | **Meta-Regression** | **—** | **NA – NA** | **—** | **NA** | **NA** | **NA** | **—** | **—** | **—** | **—** | **NA** |
| **main meta-analysis** | **mixed** | **Overall** | **34.51** | **25.72 – 43.30** | **34.73 ± 4.81 (27.00–45.86)** | **32** | **Mixed** | **863** | **95.4%** | **681** | **<0.005** | **0.51** | **Random** |
| main meta-analysis | white | 18+ | — | NA – NA | — | NA | NA | NA | — | — | — | — | NA |
| main meta-analysis | white | 20-30 | — | NA – NA | — | NA | NA | NA | — | — | — | — | NA |
| main meta-analysis | white | 30-40 | — | NA – NA | — | NA | NA | NA | — | — | — | — | NA |
| main meta-analysis | white | 40-50 | 30.22 | 11.42 – 49.01 | 30.61 ± 0.56 (30.21–31.00) | 2 | Observational Cohort | 530 | 0% | 0 | 0.88 | — | Fixed |
| main meta-analysis | white | 50-60 | 31.89 | 24.47 – 39.31 | 32.13 ± 3.97 (29.33–34.94) | 2 | Observational Cohort | 2488 | 91% | 11.2 | <0.005 | — | Random |
| main meta-analysis | white | 60+ | 30.68 | 19.43 – 41.93 | 30.65 ± 5.83 (22.91–37.63) | 5 | Observational Cohort | 656 | 97.4% | 155.5 | <0.005 | 0.63 | Random |
| **main meta-analysis** | **white** | **Meta-Regression** | **—** | **NA – NA** | **—** | **NA** | **NA** | **NA** | **—** | **—** | **—** | **—** | **NA** |
| **main meta-analysis** | **white** | **Overall** | **30.95** | **22.31 – 39.58** | **30.97 ± 4.41 (22.91–37.63)** | **9** | **Observational Cohort** | **3674** | **95.7%** | **186.9** | **<0.005** | **0.45** | **Random** |
| male | All | 18+ | 34.70 | 17.26 – 52.14 | — (34.70–34.70) | 1 | Observational Cohort | 21 | — | — | — | — | Single study |
| male | All | 20-30 | 41.01 | 35.40 – 46.62 | — (41.01–41.01) | 1 | Observational Cohort | 25 | — | — | — | — | Single study |
| male | All | 30-40 | 45.86 | 38.94 – 52.78 | — (45.86–45.86) | 1 | Observational Cohort | 25 | — | — | — | — | Single study |
| male | All | 40-50 | 40.52 | 33.21 – 47.83 | 35.84 ± 6.85 (31.00–40.69) | 2 | Observational Cohort | 31 | 72.5% | 3.6 | 0.06 | — | Fixed |
| male | All | 50-60 | 33.34 | 20.82 – 45.86 | 34.01 ± 1.32 (33.08–34.94) | 2 | Observational Cohort | 104 | 6.4% | 1.1 | 0.30 | — | Fixed |
| male | All | 60+ | 34.58 | 29.00 – 40.16 | 34.46 ± 3.07 (31.50–37.63) | 3 | Observational Cohort | 329 | 88.8% | 17.8 | <0.005 | 0.46 | Random |
| **male** | **All** | **Meta-Regression** | **—** | **NA – NA** | **—** | **NA** | **NA** | **NA** | **—** | **—** | **—** | **—** | **NA** |
| **male** | **All** | **Overall** | **36.87** | **27.96 – 45.78** | **36.47 ± 4.77 (31.00–45.86)** | **10** | **Observational Cohort** | **535** | **96.6%** | **267.5** | **<0.005** | **0.29** | **Random** |
| male | black | 18+ | — | NA – NA | — | NA | NA | NA | — | — | — | — | NA |
| male | black | 20-30 | — | NA – NA | — | NA | NA | NA | — | — | — | — | NA |
| male | black | 30-40 | — | NA – NA | — | NA | NA | NA | — | — | — | — | NA |
| male | black | 40-50 | — | NA – NA | — | NA | NA | NA | — | — | — | — | NA |
| male | black | 50-60 | — | NA – NA | — | NA | NA | NA | — | — | — | — | NA |
| male | black | 60+ | — | NA – NA | — | NA | NA | NA | — | — | — | — | NA |
| **male** | **black** | **Meta-Regression** | **—** | **NA – NA** | **—** | **NA** | **NA** | **NA** | **—** | **—** | **—** | **—** | **NA** |
| **male** | **black** | **Overall** | **—** | **NA – NA** | **—** | **NA** | **NA** | **NA** | **—** | **—** | **—** | **—** | **NA** |
| male | mixed | 18+ | 34.70 | 17.26 – 52.14 | — (34.70–34.70) | 1 | Observational Cohort | 21 | — | — | — | — | Single study |
| male | mixed | 20-30 | 41.01 | 35.40 – 46.62 | — (41.01–41.01) | 1 | Observational Cohort | 25 | — | — | — | — | Single study |
| male | mixed | 30-40 | 45.86 | 38.94 – 52.78 | — (45.86–45.86) | 1 | Observational Cohort | 25 | — | — | — | — | Single study |
| male | mixed | 40-50 | 40.69 | 34.07 – 47.31 | — (40.69–40.69) | 1 | Observational Cohort | 25 | — | — | — | — | Single study |
| male | mixed | 50-60 | 33.08 | 26.46 – 39.70 | — (33.08–33.08) | 1 | Observational Cohort | 25 | — | — | — | — | Single study |
| male | mixed | 60+ | — | NA – NA | — | NA | NA | NA | — | — | — | — | NA |
| **male** | **mixed** | **Meta-Regression** | **—** | **NA – NA** | **—** | **NA** | **NA** | **NA** | **—** | **—** | **—** | **—** | **NA** |
| **male** | **mixed** | **Overall** | **39.17** | **29.19 – 49.15** | **39.07 ± 5.18 (33.08–45.86)** | **5** | **Observational Cohort** | **121** | **97.8%** | **185.3** | **<0.005** | **0.76** | **Random** |
| male | white | 18+ | — | NA – NA | — | NA | NA | NA | — | — | — | — | NA |
| male | white | 20-30 | — | NA – NA | — | NA | NA | NA | — | — | — | — | NA |
| male | white | 30-40 | — | NA – NA | — | NA | NA | NA | — | — | — | — | NA |
| male | white | 40-50 | 31.00 | 6.83 – 55.17 | — (31.00–31.00) | 1 | Observational Cohort | 6 | — | — | — | — | Single study |
| male | white | 50-60 | 34.94 | 5.89 – 63.99 | — (34.94–34.94) | 1 | Observational Cohort | 79 | — | — | — | — | Single study |
| male | white | 60+ | 34.58 | 29.00 – 40.16 | 34.46 ± 3.07 (31.50–37.63) | 3 | Observational Cohort | 329 | 88.8% | 17.8 | <0.005 | 0.46 | Random |
| **male** | **white** | **Meta-Regression** | **—** | **NA – NA** | **—** | **NA** | **NA** | **NA** | **—** | **—** | **—** | **—** | **NA** |
| **male** | **white** | **Overall** | **34.51** | **30.08 – 38.94** | **33.86 ± 2.71 (31.00–37.63)** | **5** | **Observational Cohort** | **414** | **78.5%** | **18.6** | **<0.005** | **0.44** | **Random** |
| female | All | 18+ | 29.50 | 25.47 – 33.53 | 30.20 ± 4.09 (27.00–37.00) | 5 | Mixed | 168 | 59% | 9.8 | <0.05 | 0.37 | Random |
| female | All | 20-30 | 32.36 | 27.93 – 36.79 | — (32.36–32.36) | 1 | Observational Cohort | 25 | — | — | — | — | Single study |
| female | All | 30-40 | 32.42 | 27.87 – 36.97 | — (32.42–32.42) | 1 | Observational Cohort | 25 | — | — | — | — | Single study |
| female | All | 40-50 | 33.55 | 26.50 – 40.60 | 33.55 ± 3.48 (30.21–37.15) | 3 | Observational Cohort | 578 | 97.5% | 80.5 | <0.005 | 0.73 | Random |
| female | All | 50-60 | 32.21 | 21.30 – 43.13 | 32.22 ± 5.63 (28.63–38.71) | 3 | Observational Cohort | 2565 | 98.5% | 134.8 | <0.005 | 0.57 | Random |
| female | All | 60+ | 24.30 | 15.65 – 32.95 | 24.32 ± 4.46 (20.50–30.74) | 5 | Observational Cohort | 496 | 97.9% | 186.4 | <0.005 | 0.19 | Random |
| **female** | **All** | **Meta-Regression** | **28.76** | **24.73 – 32.79** | **—** | **5** | **Mixed** | **168** | **58%** | **9.8** | **<0.05** | **0.37** | **Univariable** |
| **female** | **All** | **Overall** | **29.60** | **19.68 – 39.52** | **29.71 ± 5.19 (20.50–38.71)** | **18** | **Mixed** | **3857** | **98.7%** | **1303.8** | **<0.005** | **0.66** | **Random** |
| female | black | 18+ | — | NA – NA | — | NA | NA | NA | — | — | — | — | NA |
| female | black | 20-30 | — | NA – NA | — | NA | NA | NA | — | — | — | — | NA |
| female | black | 30-40 | — | NA – NA | — | NA | NA | NA | — | — | — | — | NA |
| female | black | 40-50 | 33.29 | 10.63 – 55.95 | — (33.29–33.29) | 1 | Observational Cohort | 29 | — | — | — | — | Single study |
| female | black | 50-60 | 28.63 | 8.87 – 48.39 | — (28.63–28.63) | 1 | Observational Cohort | 131 | — | — | — | — | Single study |
| female | black | 60+ | 20.50 | 13.56 – 27.44 | 20.50 ± 0.00 (20.50–20.50) | 2 | Observational Cohort | 144 | 0% | 0 | 1.00 | — | Fixed |
| **female** | **black** | **Meta-Regression** | **—** | **NA – NA** | **—** | **NA** | **NA** | **NA** | **—** | **—** | **—** | **—** | **NA** |
| **female** | **black** | **Overall** | **25.51** | **13.60 – 37.43** | **25.73 ± 6.33 (20.50–33.29)** | **4** | **Observational Cohort** | **304** | **97.2%** | **107.2** | **<0.005** | **0.11** | **Random** |
| female | mixed | 18+ | 29.50 | 25.47 – 33.53 | 30.20 ± 4.09 (27.00–37.00) | 5 | Mixed | 168 | 59% | 9.8 | <0.05 | 0.37 | Random |
| female | mixed | 20-30 | 32.36 | 27.93 – 36.79 | — (32.36–32.36) | 1 | Observational Cohort | 25 | — | — | — | — | Single study |
| female | mixed | 30-40 | 32.42 | 27.87 – 36.97 | — (32.42–32.42) | 1 | Observational Cohort | 25 | — | — | — | — | Single study |
| female | mixed | 40-50 | 37.15 | 30.76 – 43.54 | — (37.15–37.15) | 1 | Observational Cohort | 25 | — | — | — | — | Single study |
| female | mixed | 50-60 | 38.71 | 30.99 – 46.43 | — (38.71–38.71) | 1 | Observational Cohort | 25 | — | — | — | — | Single study |
| female | mixed | 60+ | 30.74 | 22.86 – 38.62 | — (30.74–30.74) | 1 | Observational Cohort | 25 | — | — | — | — | Single study |
| **female** | **mixed** | **Meta-Regression** | **—** | **NA – NA** | **—** | **NA** | **NA** | **NA** | **—** | **—** | **—** | **—** | **NA** |
| **female** | **mixed** | **Overall** | **32.25** | **24.75 – 39.75** | **32.24 ± 4.16 (27.00–38.71)** | **10** | **Mixed** | **293** | **93.4%** | **137.3** | **<0.005** | **0.57** | **Random** |
| female | white | 18+ | — | NA – NA | — | NA | NA | NA | — | — | — | — | NA |
| female | white | 20-30 | — | NA – NA | — | NA | NA | NA | — | — | — | — | NA |
| female | white | 30-40 | — | NA – NA | — | NA | NA | NA | — | — | — | — | NA |
| female | white | 40-50 | 30.21 | 11.45 – 48.97 | — (30.21–30.21) | 1 | Observational Cohort | 524 | — | — | — | — | Single study |
| female | white | 50-60 | 29.33 | 9.67 – 48.99 | — (29.33–29.33) | 1 | Observational Cohort | 2409 | — | — | — | — | Single study |
| female | white | 60+ | 25.05 | 19.78 – 30.32 | 24.94 ± 2.87 (22.91–26.97) | 2 | Observational Cohort | 327 | 87.7% | 8.1 | <0.005 | — | Random |
| **female** | **white** | **Meta-Regression** | **—** | **NA – NA** | **—** | **NA** | **NA** | **NA** | **—** | **—** | **—** | **—** | **NA** |
| **female** | **white** | **Overall** | **27.51** | **21.53 – 33.48** | **27.36 ± 3.26 (22.91–30.21)** | **4** | **Observational Cohort** | **3260** | **92.8%** | **41.9** | **<0.005** | **0.26** | **Random** |
| *I² represents heterogeneity; Q test assesses between-study variation; Egger's p = linear regression test for funnel plot asymmetry; Meta-Regression shows covariate-adjusted estimates* | | | | | | | | | | | | | |

###### 6.12.2.3 Table of Results subdivided by race / Sex / Age

ammonium_ma_3rd_analysis$race_summary_table

Table 1: **race-Stratified Meta-Analysis Results**

24-Hour Urine Ammonium (mmol/24h)

| Sex Group | race | Age Band | 95% Range | Studies | Study Type | Participants | I² | Cochran's Q | Q (p-value) | Egger's p | Model |
| --- | --- | --- | --- | --- | --- | --- | --- | --- | --- | --- | --- |
| **main meta-analysis** | **All races** | **18+** | **26.22 – 40.14** | **23** | **Mixed** | **638** | **76.3%** | **92.6** | **<0.005** | **<0.05** | **Random** |
| **main meta-analysis** | **All races** | **20-30** | **24.73 – 48.62** | **2** | **Observational Cohort** | **50** | **99.3%** | **140.8** | **<0.005** | **—** | **Random** |
| **main meta-analysis** | **All races** | **30-40** | **20.54 – 57.72** | **2** | **Observational Cohort** | **50** | **99.6%** | **253.1** | **<0.005** | **—** | **Random** |
| **main meta-analysis** | **All races** | **40-50** | **26.39 – 43.52** | **5** | **Observational Cohort** | **609** | **98%** | **203.9** | **<0.005** | **0.66** | **Random** |
| **main meta-analysis** | **All races** | **50-60** | **24.84 – 40.91** | **5** | **Observational Cohort** | **2669** | **97.6%** | **164.1** | **<0.005** | **0.21** | **Random** |
| **main meta-analysis** | **All races** | **60+** | **15.53 – 40.67** | **8** | **Observational Cohort** | **825** | **99%** | **686.2** | **<0.005** | **<0.05** | **Random** |
| **main meta-analysis** | **All races** | **Meta-Regression** | **23.61 – 34.93** | **16** | **Mixed** | **349** | **68.5%** | **39.1** | **<0.005** | **0.15** | **Multivariable** |
| **main meta-analysis** | **All races** | **Meta-Regression** | **23.61 – 34.93** | **16** | **Mixed** | **349** | **68.5%** | **39.1** | **<0.005** | **0.15** | **Multivariable** |
| **main meta-analysis** | **All races** | **Overall** | **22.48 – 43.35** | **45** | **Mixed** | **4841** | **98.4%** | **2767.6** | **<0.005** | **0.08** | **Random** |
| **main meta-analysis** | **All races** | **Overall** | **22.48 – 43.35** | **45** | **Mixed** | **4841** | **98.4%** | **2767.6** | **<0.005** | **0.08** | **Random** |
| main meta-analysis | Black | 18+ | — | — | NA | — | — | — | — | — | — |
| main meta-analysis | Black | 20-30 | — | — | NA | — | — | — | — | — | — |
| main meta-analysis | Black | 30-40 | — | — | NA | — | — | — | — | — | — |
| main meta-analysis | Black | 40-50 | 10.63 – 55.95 | 1 | Observational Cohort | 29 | — | — | — | — | Single study |
| main meta-analysis | Black | 50-60 | 8.87 – 48.39 | 1 | Observational Cohort | 131 | — | — | — | — | Single study |
| main meta-analysis | Black | 60+ | 13.56 – 27.44 | 2 | Observational Cohort | 144 | 0% | 0 | 1.00 | — | Fixed |
| main meta-analysis | Black | Meta-Regression | — | — | NA | — | — | — | — | — | — |
| **main meta-analysis** | **Black** | **Overall** | **13.60 – 37.43** | **4** | **Observational Cohort** | **304** | **97.2%** | **107.2** | **<0.005** | **0.11** | **Random** |
| main meta-analysis | Mixed | 18+ | 26.22 – 40.14 | 23 | Mixed | 638 | 76.3% | 92.6 | <0.005 | <0.05 | Random |
| main meta-analysis | Mixed | 20-30 | 24.73 – 48.62 | 2 | Observational Cohort | 50 | 99.3% | 140.8 | <0.005 | — | Random |
| main meta-analysis | Mixed | 30-40 | 20.54 – 57.72 | 2 | Observational Cohort | 50 | 99.6% | 253.1 | <0.005 | — | Random |
| main meta-analysis | Mixed | 40-50 | 34.19 – 43.65 | 2 | Observational Cohort | 50 | 93% | 14.2 | <0.005 | — | Random |
| main meta-analysis | Mixed | 50-60 | 28.21 – 43.55 | 2 | Observational Cohort | 50 | 96.6% | 29.4 | <0.005 | — | Random |
| main meta-analysis | Mixed | 60+ | 22.86 – 38.62 | 1 | Observational Cohort | 25 | — | — | — | — | Single study |
| main meta-analysis | Mixed | Meta-Regression | — | — | NA | — | — | — | — | — | — |
| **main meta-analysis** | **Mixed** | **Overall** | **25.72 – 43.30** | **32** | **Mixed** | **863** | **95.4%** | **681** | **<0.005** | **0.51** | **Random** |
| main meta-analysis | White | 18+ | — | — | NA | — | — | — | — | — | — |
| main meta-analysis | White | 20-30 | — | — | NA | — | — | — | — | — | — |
| main meta-analysis | White | 30-40 | — | — | NA | — | — | — | — | — | — |
| main meta-analysis | White | 40-50 | 11.42 – 49.01 | 2 | Observational Cohort | 530 | 0% | 0 | 0.88 | — | Fixed |
| main meta-analysis | White | 50-60 | 24.47 – 39.31 | 2 | Observational Cohort | 2488 | 91% | 11.2 | <0.005 | — | Random |
| main meta-analysis | White | 60+ | 19.43 – 41.93 | 5 | Observational Cohort | 656 | 97.4% | 155.5 | <0.005 | 0.63 | Random |
| main meta-analysis | White | Meta-Regression | — | — | NA | — | — | — | — | — | — |
| **main meta-analysis** | **White** | **Overall** | **22.31 – 39.58** | **9** | **Observational Cohort** | **3674** | **95.7%** | **186.9** | **<0.005** | **0.45** | **Random** |
| **male** | **All races** | **18+** | **17.26 – 52.14** | **1** | **Observational Cohort** | **21** | **—** | **—** | **—** | **—** | **Single study** |
| **male** | **All races** | **20-30** | **35.40 – 46.62** | **1** | **Observational Cohort** | **25** | **—** | **—** | **—** | **—** | **Single study** |
| **male** | **All races** | **30-40** | **38.94 – 52.78** | **1** | **Observational Cohort** | **25** | **—** | **—** | **—** | **—** | **Single study** |
| **male** | **All races** | **40-50** | **33.21 – 47.83** | **2** | **Observational Cohort** | **31** | **72.5%** | **3.6** | **0.06** | **—** | **Fixed** |
| **male** | **All races** | **50-60** | **20.82 – 45.86** | **2** | **Observational Cohort** | **104** | **6.4%** | **1.1** | **0.30** | **—** | **Fixed** |
| **male** | **All races** | **60+** | **29.00 – 40.16** | **3** | **Observational Cohort** | **329** | **88.8%** | **17.8** | **<0.005** | **0.46** | **Random** |
| **male** | **All races** | **Meta-Regression** | **—** | **—** | **NA** | **—** | **—** | **—** | **—** | **—** | **—** |
| **male** | **All races** | **Overall** | **27.96 – 45.78** | **10** | **Observational Cohort** | **535** | **96.6%** | **267.5** | **<0.005** | **0.29** | **Random** |
| male | Black | 18+ | — | — | NA | — | — | — | — | — | — |
| male | Black | 20-30 | — | — | NA | — | — | — | — | — | — |
| male | Black | 30-40 | — | — | NA | — | — | — | — | — | — |
| male | Black | 40-50 | — | — | NA | — | — | — | — | — | — |
| male | Black | 50-60 | — | — | NA | — | — | — | — | — | — |
| male | Black | 60+ | — | — | NA | — | — | — | — | — | — |
| male | Black | Meta-Regression | — | — | NA | — | — | — | — | — | — |
| **male** | **Black** | **Overall** | **—** | **—** | **NA** | **—** | **—** | **—** | **—** | **—** | **—** |
| male | Mixed | 18+ | 17.26 – 52.14 | 1 | Observational Cohort | 21 | — | — | — | — | Single study |
| male | Mixed | 20-30 | 35.40 – 46.62 | 1 | Observational Cohort | 25 | — | — | — | — | Single study |
| male | Mixed | 30-40 | 38.94 – 52.78 | 1 | Observational Cohort | 25 | — | — | — | — | Single study |
| male | Mixed | 40-50 | 34.07 – 47.31 | 1 | Observational Cohort | 25 | — | — | — | — | Single study |
| male | Mixed | 50-60 | 26.46 – 39.70 | 1 | Observational Cohort | 25 | — | — | — | — | Single study |
| male | Mixed | 60+ | — | — | NA | — | — | — | — | — | — |
| male | Mixed | Meta-Regression | — | — | NA | — | — | — | — | — | — |
| **male** | **Mixed** | **Overall** | **29.19 – 49.15** | **5** | **Observational Cohort** | **121** | **97.8%** | **185.3** | **<0.005** | **0.76** | **Random** |
| male | White | 18+ | — | — | NA | — | — | — | — | — | — |
| male | White | 20-30 | — | — | NA | — | — | — | — | — | — |
| male | White | 30-40 | — | — | NA | — | — | — | — | — | — |
| male | White | 40-50 | 6.83 – 55.17 | 1 | Observational Cohort | 6 | — | — | — | — | Single study |
| male | White | 50-60 | 5.89 – 63.99 | 1 | Observational Cohort | 79 | — | — | — | — | Single study |
| male | White | 60+ | 29.00 – 40.16 | 3 | Observational Cohort | 329 | 88.8% | 17.8 | <0.005 | 0.46 | Random |
| male | White | Meta-Regression | — | — | NA | — | — | — | — | — | — |
| **male** | **White** | **Overall** | **30.08 – 38.94** | **5** | **Observational Cohort** | **414** | **78.5%** | **18.6** | **<0.005** | **0.44** | **Random** |
| **female** | **All races** | **18+** | **25.47 – 33.53** | **5** | **Mixed** | **168** | **59%** | **9.8** | **<0.05** | **0.37** | **Random** |
| **female** | **All races** | **20-30** | **27.93 – 36.79** | **1** | **Observational Cohort** | **25** | **—** | **—** | **—** | **—** | **Single study** |
| **female** | **All races** | **30-40** | **27.87 – 36.97** | **1** | **Observational Cohort** | **25** | **—** | **—** | **—** | **—** | **Single study** |
| **female** | **All races** | **40-50** | **26.50 – 40.60** | **3** | **Observational Cohort** | **578** | **97.5%** | **80.5** | **<0.005** | **0.73** | **Random** |
| **female** | **All races** | **50-60** | **21.30 – 43.13** | **3** | **Observational Cohort** | **2565** | **98.5%** | **134.8** | **<0.005** | **0.57** | **Random** |
| **female** | **All races** | **60+** | **15.65 – 32.95** | **5** | **Observational Cohort** | **496** | **97.9%** | **186.4** | **<0.005** | **0.19** | **Random** |
| **female** | **All races** | **Meta-Regression** | **24.73 – 32.79** | **5** | **Mixed** | **168** | **58%** | **9.8** | **<0.05** | **0.37** | **Univariable** |
| **female** | **All races** | **Overall** | **19.68 – 39.52** | **18** | **Mixed** | **3857** | **98.7%** | **1303.8** | **<0.005** | **0.66** | **Random** |
| female | Black | 18+ | — | — | NA | — | — | — | — | — | — |
| female | Black | 20-30 | — | — | NA | — | — | — | — | — | — |
| female | Black | 30-40 | — | — | NA | — | — | — | — | — | — |
| female | Black | 40-50 | 10.63 – 55.95 | 1 | Observational Cohort | 29 | — | — | — | — | Single study |
| female | Black | 50-60 | 8.87 – 48.39 | 1 | Observational Cohort | 131 | — | — | — | — | Single study |
| female | Black | 60+ | 13.56 – 27.44 | 2 | Observational Cohort | 144 | 0% | 0 | 1.00 | — | Fixed |
| female | Black | Meta-Regression | — | — | NA | — | — | — | — | — | — |
| **female** | **Black** | **Overall** | **13.60 – 37.43** | **4** | **Observational Cohort** | **304** | **97.2%** | **107.2** | **<0.005** | **0.11** | **Random** |
| female | Mixed | 18+ | 25.47 – 33.53 | 5 | Mixed | 168 | 59% | 9.8 | <0.05 | 0.37 | Random |
| female | Mixed | 20-30 | 27.93 – 36.79 | 1 | Observational Cohort | 25 | — | — | — | — | Single study |
| female | Mixed | 30-40 | 27.87 – 36.97 | 1 | Observational Cohort | 25 | — | — | — | — | Single study |
| female | Mixed | 40-50 | 30.76 – 43.54 | 1 | Observational Cohort | 25 | — | — | — | — | Single study |
| female | Mixed | 50-60 | 30.99 – 46.43 | 1 | Observational Cohort | 25 | — | — | — | — | Single study |
| female | Mixed | 60+ | 22.86 – 38.62 | 1 | Observational Cohort | 25 | — | — | — | — | Single study |
| female | Mixed | Meta-Regression | — | — | NA | — | — | — | — | — | — |
| **female** | **Mixed** | **Overall** | **24.75 – 39.75** | **10** | **Mixed** | **293** | **93.4%** | **137.3** | **<0.005** | **0.57** | **Random** |
| female | White | 18+ | — | — | NA | — | — | — | — | — | — |
| female | White | 20-30 | — | — | NA | — | — | — | — | — | — |
| female | White | 30-40 | — | — | NA | — | — | — | — | — | — |
| female | White | 40-50 | 11.45 – 48.97 | 1 | Observational Cohort | 524 | — | — | — | — | Single study |
| female | White | 50-60 | 9.67 – 48.99 | 1 | Observational Cohort | 2409 | — | — | — | — | Single study |
| female | White | 60+ | 19.78 – 30.32 | 2 | Observational Cohort | 327 | 87.7% | 8.1 | <0.005 | — | Random |
| female | White | Meta-Regression | — | — | NA | — | — | — | — | — | — |
| **female** | **White** | **Overall** | **21.53 – 33.48** | **4** | **Observational Cohort** | **3260** | **92.8%** | **41.9** | **<0.005** | **0.26** | **Random** |
| *I² represents heterogeneity; Q test assesses between-study variation; Egger's p = linear regression test for funnel plot asymmetry; — indicates insufficient data* | | | | | | | | | | | |

###### 6.12.2.4 Wald Tests

ammonium_ma_3rd_analysis$wald_tests$sex_comparison

Table 1: **Sex Comparison (Overall)**

| Comparison | % Shift | P (Wald Test) | Mean Diff (95% CI) |
| --- | --- | --- | --- |
| Male vs Female (Overall) | +24.6% | 0.285 | 7.27 [-6.07, 20.61] |
| *Mean difference = Group 1 − Group 2 in component units (mmol/24h, L/24h, or pH); % shift = difference relative to Group 2.* | | | |

ammonium_ma_3rd_analysis$wald_tests$race_comparisons

Table 1: **race Comparisons Within Sex Groups**

| Sex Group | Comparison | % Shift | P (Wald Test) | Mean Diff (95% CI) |
| --- | --- | --- | --- | --- |
| male | black vs mixed | — | — | — |
| male | black vs white | — | — | — |
| male | mixed vs white | +13.5% | 0.403 | 4.65 [-6.26, 15.57] |
| female | black vs mixed | -20.9% | 0.348 | -6.74 [-20.82, 7.34] |
| female | black vs white | -7.2% | 0.769 | -1.99 [-15.32, 11.34] |
| female | mixed vs white | +17.2% | 0.332 | 4.74 [-4.84, 14.33] |
| *Mean difference = Group 1 − Group 2 in component units (mmol/24h, L/24h, or pH); % shift = difference relative to Group 2.* | | | | |

ammonium_ma_3rd_analysis$wald_tests$age_band_comparisons

Table 1: **Age Band Comparisons Within Sex Groups**

| Sex Group | Comparison | % Shift | P (Wald Test) | Mean Diff (95% CI) |
| --- | --- | --- | --- | --- |
| main meta-analysis | 18+ vs 20-30 | -9.5% | 0.620 | -3.50 [-17.32, 10.33] |
| main meta-analysis | 18+ vs 30-40 | -15.2% | 0.557 | -5.95 [-25.80, 13.90] |
| main meta-analysis | 18+ vs 40-50 | -5.1% | 0.752 | -1.78 [-12.81, 9.26] |
| main meta-analysis | 18+ vs 50-60 | +0.9% | 0.955 | 0.31 [-10.32, 10.94] |
| main meta-analysis | 18+ vs 60+ | +18.1% | 0.488 | 5.08 [-9.29, 19.45] |
| main meta-analysis | 20-30 vs 30-40 | -6.3% | 0.828 | -2.45 [-24.55, 19.65] |
| main meta-analysis | 20-30 vs 40-50 | +4.9% | 0.818 | 1.72 [-12.98, 16.42] |
| main meta-analysis | 20-30 vs 50-60 | +11.6% | 0.605 | 3.80 [-10.59, 18.20] |
| main meta-analysis | 20-30 vs 60+ | +30.5% | 0.332 | 8.58 [-8.77, 25.92] |
| main meta-analysis | 30-40 vs 40-50 | +11.9% | 0.689 | 4.17 [-16.29, 24.64] |
| main meta-analysis | 30-40 vs 50-60 | +19.0% | 0.545 | 6.25 [-14.00, 26.51] |
| main meta-analysis | 30-40 vs 60+ | +39.2% | 0.336 | 11.03 [-11.41, 33.47] |
| main meta-analysis | 40-50 vs 50-60 | +6.3% | 0.728 | 2.08 [-9.66, 13.82] |
| main meta-analysis | 40-50 vs 60+ | +24.4% | 0.377 | 6.85 [-8.36, 22.06] |
| main meta-analysis | 50-60 vs 60+ | +17.0% | 0.531 | 4.77 [-10.15, 19.69] |
| male | 18+ vs 20-30 | -15.4% | 0.500 | -6.31 [-24.63, 12.01] |
| male | 18+ vs 30-40 | -24.3% | 0.244 | -11.16 [-29.93, 7.61] |
| male | 18+ vs 40-50 | -14.4% | 0.547 | -5.82 [-24.73, 13.10] |
| male | 18+ vs 50-60 | +4.1% | 0.901 | 1.36 [-20.12, 22.83] |
| male | 18+ vs 60+ | +0.3% | 0.990 | 0.12 [-18.20, 18.43] |
| male | 20-30 vs 30-40 | -10.6% | 0.286 | -4.85 [-13.75, 4.05] |
| male | 20-30 vs 40-50 | +1.2% | 0.917 | 0.49 [-8.72, 9.70] |
| male | 20-30 vs 50-60 | +23.0% | 0.273 | 7.67 [-6.05, 21.39] |
| male | 20-30 vs 60+ | +18.6% | 0.111 | 6.43 [-1.48, 14.34] |
| male | 30-40 vs 40-50 | +13.2% | 0.298 | 5.34 [-4.72, 15.41] |
| male | 30-40 vs 50-60 | +37.5% | 0.086 | 12.52 [-1.79, 26.82] |
| male | 30-40 vs 60+ | +32.6% | <0.05 | 11.28 [2.39, 20.17] |
| male | 40-50 vs 50-60 | +21.5% | 0.332 | 7.18 [-7.32, 21.68] |
| male | 40-50 vs 60+ | +17.2% | 0.206 | 5.94 [-3.26, 15.14] |
| male | 50-60 vs 60+ | -3.6% | 0.860 | -1.24 [-14.95, 12.47] |
| female | 18+ vs 20-30 | -8.8% | 0.349 | -2.86 [-8.85, 3.13] |
| female | 18+ vs 30-40 | -9.0% | 0.346 | -2.92 [-9.00, 3.16] |
| female | 18+ vs 40-50 | -12.1% | 0.329 | -4.05 [-12.17, 4.07] |
| female | 18+ vs 50-60 | -8.4% | 0.648 | -2.71 [-14.35, 8.92] |
| female | 18+ vs 60+ | +21.4% | 0.286 | 5.20 [-4.34, 14.74] |
| female | 20-30 vs 30-40 | -0.2% | 0.985 | -0.06 [-6.41, 6.29] |
| female | 20-30 vs 40-50 | -3.5% | 0.780 | -1.19 [-9.52, 7.14] |
| female | 20-30 vs 50-60 | +0.5% | 0.980 | 0.15 [-11.63, 11.93] |
| female | 20-30 vs 60+ | +33.2% | 0.104 | 8.06 [-1.66, 17.78] |
| female | 30-40 vs 40-50 | -3.4% | 0.792 | -1.13 [-9.52, 7.26] |
| female | 30-40 vs 50-60 | +0.6% | 0.972 | 0.21 [-11.62, 12.03] |
| female | 30-40 vs 60+ | +33.4% | 0.104 | 8.12 [-1.66, 17.89] |
| female | 40-50 vs 50-60 | +4.2% | 0.840 | 1.34 [-11.66, 14.33] |
| female | 40-50 vs 60+ | +38.1% | 0.104 | 9.25 [-1.91, 20.41] |
| female | 50-60 vs 60+ | +32.5% | 0.266 | 7.91 [-6.02, 21.84] |
| *Mean difference = Group 1 − Group 2 in component units (mmol/24h, L/24h, or pH); % shift = difference relative to Group 2.* | | | | |

ammonium_ma_3rd_analysis$wald_tests$sex_within_age_bands

Table 1: **Sex Comparisons Within Age Bands**

| Age Band | % Shift | P (Wald Test) | Mean Diff (95% CI) |
| --- | --- | --- | --- |
| 18+ | +17.6% | 0.569 | 5.20 [-12.70, 23.10] |
| 20-30 | +26.7% | <0.05 | 8.65 [1.51, 15.79] |
| 30-40 | +41.5% | <0.01 | 13.44 [5.16, 21.72] |
| 40-50 | +20.8% | 0.179 | 6.97 [-3.19, 17.13] |
| 50-60 | +3.5% | 0.894 | 1.13 [-15.48, 17.74] |
| 60+ | +42.3% | 0.050 | 10.28 [-0.02, 20.57] |
| *Mean difference = Group 1 − Group 2 in component units (mmol/24h, L/24h, or pH); % shift = difference relative to Group 2.* | | | |

#### 6.13 Chloride

##### 6.13.1 Comparison by Age Band

###### 6.13.1.1 Plot

```
chloride_ma_2nd_analysis <- sex_age_subdivision_ma("chloride")
## Not enough studies for multivariable meta-regression (k = 3, need > 6). Running univariable models instead.
## Not enough studies for multivariable meta-regression (k = 6, need > 6). Running univariable models instead.
chloride_ma_2nd_analysis$combined_plot
## `height` was translated to `width`.
## `height` was translated to `width`.
## `height` was translated to `width`.
```

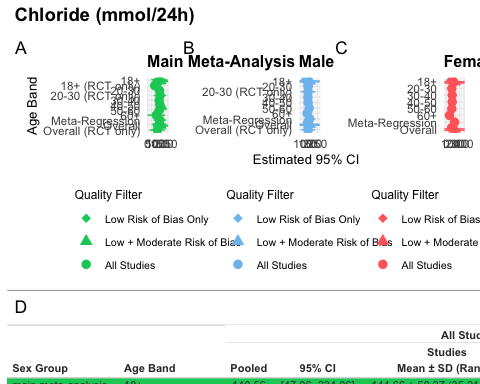

###### 6.13.1.2 Summary Table

chloride_ma_2nd_analysis$combined_summary_table

Table 1: **Combined Meta-Analysis Results**

24-Hour Urine Chloride (mmol/24h)

|  |  | **All Studies** | | | | | | **Low Risk of Bias Only** | | | | | | **Low + Moderate RoB** | | | | | |
| --- | --- | --- | --- | --- | --- | --- | --- | --- | --- | --- | --- | --- | --- | --- | --- | --- | --- | --- | --- |
| Sex Group | Age Band | Pooled | 95% CI | Studies  Mean ± SD (Range) | Studies, n | Q p-value | Egger's p | Pooled | 95% CI | Studies  Mean ± SD (Range) | Studies, n | Q p-value | Egger's p | Pooled | 95% CI | Studies  Mean ± SD (Range) | Studies, n | Q p-value | Egger's p |
| main meta-analysis | 18+ | 140.56 | [47.06, 234.06] | 144.66 ± 50.27 (35.21–205.68) | 38 | <0.001 | <0.005 | 155.89 | [65.76, 246.02] | 160.55 ± 47.64 (69.80–205.68) | 26 | <0.001 | 0.22 | 155.89 | [65.76, 246.02] | 160.55 ± 47.64 (69.80–205.68) | 26 | <0.001 | 0.22 |
| **main meta-analysis** | **18+ (RCT only)** | **87.07** | **[37.82, 136.32]** | **89.00 ± 27.15 (69.80–108.20)** | **2** | **<0.01** | **—** | **—** | **—** | **—** | **—** | **—** | **—** | **—** | **—** | **—** | **—** | **—** | **—** |
| main meta-analysis | 20-30 | 145.11 | [98.93, 191.28] | 146.72 ± 25.64 (123.90–178.40) | 4 | <0.001 | 0.82 | — | — | — | — | — | — | — | — | — | — | — | — |
| **main meta-analysis** | **20-30 (RCT only)** | **151.87** | **[84.06, 219.68]** | **153.20 ± 35.64 (128.00–178.40)** | **2** | **<0.001** | **—** | **—** | **—** | **—** | **—** | **—** | **—** | **—** | **—** | **—** | **—** | **—** | **—** |
| main meta-analysis | 30-40 | 141.30 | [117.78, 164.82] | 141.20 ± 12.16 (132.60–149.80) | 2 | <0.001 | — | — | — | — | — | — | — | — | — | — | — | — | — |
| main meta-analysis | 40-50 | 149.67 | [119.14, 180.20] | 149.63 ± 15.68 (133.10–164.30) | 3 | <0.001 | 0.44 | — | — | — | — | — | — | 164.30 | [145.68, 182.92] | — (164.30–164.30) | 1 | — | — |
| main meta-analysis | 50-60 | 143.15 | [90.42, 195.88] | 143.20 ± 27.01 (124.10–162.30) | 2 | <0.001 | — | — | — | — | — | — | — | — | — | — | — | — | — |
| main meta-analysis | 60+ | 106.49 | [6.48, 206.50] | 106.50 ± 51.05 (70.40–142.60) | 2 | <0.001 | — | — | — | — | — | — | — | — | — | — | — | — | — |
| **main meta-analysis** | **Meta-Regression** | **150.95** | **[83.96, 217.95]** | **—** | **36** | **<0.001** | **0.80** | **—** | **—** | **—** | **—** | **—** | **—** | **—** | **—** | **—** | **—** | **—** | **—** |
| **main meta-analysis** | **Overall** | **139.96** | **[56.59, 223.32]** | **143.42 ± 45.26 (35.21–205.68)** | **51** | **<0.001** | **0.53** | **155.89** | **[65.76, 246.02]** | **160.55 ± 47.64 (69.80–205.68)** | **26** | **<0.001** | **0.22** | **156.15** | **[67.95, 244.35]** | **160.69 ± 46.73 (69.80–205.68)** | **27** | **<0.001** | **0.73** |
| **main meta-analysis** | **Overall (RCT only)** | **120.59** | **[34.11, 207.07]** | **121.10 ± 45.20 (69.80–178.40)** | **4** | **<0.001** | **0.85** | **—** | **—** | **—** | **—** | **—** | **—** | **—** | **—** | **—** | **—** | **—** | **—** |
| male | 18+ | 132.07 | [83.90, 180.23] | 130.91 ± 24.70 (118.33–167.96) | 4 | <0.001 | 0.87 | 167.96 | [65.96, 269.96] | — (167.96–167.96) | 1 | — | — | 167.96 | [65.96, 269.96] | — (167.96–167.96) | 1 | — | — |
| **male** | **18+ (RCT only)** | **—** | **—** | **—** | **—** | **—** | **—** | **—** | **—** | **—** | **—** | **—** | **—** | **—** | **—** | **—** | **—** | **—** | **—** |
| male | 20-30 | 152.70 | [106.30, 199.10] | 154.33 ± 25.28 (128.00–178.40) | 3 | <0.001 | 0.95 | — | — | — | — | — | — | — | — | — | — | — | — |
| **male** | **20-30 (RCT only)** | **151.87** | **[84.06, 219.68]** | **153.20 ± 35.64 (128.00–178.40)** | **2** | **<0.001** | **—** | **—** | **—** | **—** | **—** | **—** | **—** | **—** | **—** | **—** | **—** | **—** | **—** |
| male | 30-40 | 149.80 | [135.10, 164.50] | — (149.80–149.80) | 1 | — | — | — | — | — | — | — | — | — | — | — | — | — | — |
| male | 40-50 | 151.50 | [128.96, 174.04] | — (151.50–151.50) | 1 | — | — | — | — | — | — | — | — | — | — | — | — | — | — |
| male | 50-60 | 162.30 | [134.86, 189.74] | — (162.30–162.30) | 1 | — | — | — | — | — | — | — | — | — | — | — | — | — | — |
| male | 60+ | 142.60 | [124.37, 160.83] | — (142.60–142.60) | 1 | — | — | — | — | — | — | — | — | — | — | — | — | — | — |
| **male** | **Meta-Regression** | **167.96** | **[117.00, 218.92]** | **—** | **3** | **<0.001** | **0.36** | **—** | **—** | **—** | **—** | **—** | **—** | **—** | **—** | **—** | **—** | **—** | **—** |
| **male** | **Overall** | **145.43** | **[109.16, 181.70]** | **144.80 ± 21.29 (118.33–178.40)** | **11** | **<0.001** | **0.77** | **167.96** | **[65.96, 269.96]** | **— (167.96–167.96)** | **1** | **—** | **—** | **167.96** | **[65.96, 269.96]** | **— (167.96–167.96)** | **1** | **—** | **—** |
| **male** | **Overall (RCT only)** | **151.87** | **[84.06, 219.68]** | **153.20 ± 35.64 (128.00–178.40)** | **2** | **<0.001** | **—** | **—** | **—** | **—** | **—** | **—** | **—** | **—** | **—** | **—** | **—** | **—** | **—** |
| female | 18+ | 125.31 | [8.69, 241.92] | 125.70 ± 59.62 (35.21–185.58) | 8 | <0.001 | <0.001 | 185.58 | [-36.80, 407.96] | 185.58 ± 0.00 (185.58–185.58) | 2 | 1.00 | — | 185.58 | [-36.80, 407.96] | 185.58 ± 0.00 (185.58–185.58) | 2 | 1.00 | — |
| **female** | **18+ (RCT only)** | **—** | **—** | **—** | **—** | **—** | **—** | **—** | **—** | **—** | **—** | **—** | **—** | **—** | **—** | **—** | **—** | **—** | **—** |
| female | 20-30 | 123.90 | [105.28, 142.52] | — (123.90–123.90) | 1 | — | — | — | — | — | — | — | — | — | — | — | — | — | — |
| **female** | **20-30 (RCT only)** | **—** | **—** | **—** | **—** | **—** | **—** | **—** | **—** | **—** | **—** | **—** | **—** | **—** | **—** | **—** | **—** | **—** | **—** |
| female | 30-40 | 132.60 | [109.28, 155.92] | — (132.60–132.60) | 1 | — | — | — | — | — | — | — | — | — | — | — | — | — | — |
| female | 40-50 | 133.10 | [111.93, 154.27] | — (133.10–133.10) | 1 | — | — | — | — | — | — | — | — | — | — | — | — | — | — |
| female | 50-60 | 124.10 | [104.70, 143.50] | — (124.10–124.10) | 1 | — | — | — | — | — | — | — | — | — | — | — | — | — | — |
| female | 60+ | 70.40 | [55.99, 84.81] | — (70.40–70.40) | 1 | — | — | — | — | — | — | — | — | — | — | — | — | — | — |
| **female** | **Meta-Regression** | **185.58** | **[140.39, 230.77]** | **—** | **6** | **<0.001** | **<0.05** | **—** | **—** | **—** | **—** | **—** | **—** | **—** | **—** | **—** | **—** | **—** | **—** |
| **female** | **Overall** | **121.87** | **[28.04, 215.70]** | **122.28 ± 48.22 (35.21–185.58)** | **13** | **<0.001** | **0.42** | **185.58** | **[-36.80, 407.96]** | **185.58 ± 0.00 (185.58–185.58)** | **2** | **1.00** | **—** | **185.58** | **[-36.80, 407.96]** | **185.58 ± 0.00 (185.58–185.58)** | **2** | **1.00** | **—** |
| **female** | **Overall (RCT only)** | **—** | **—** | **—** | **—** | **—** | **—** | **—** | **—** | **—** | **—** | **—** | **—** | **—** | **—** | **—** | **—** | **—** | **—** |
| *Studies, n = number of studies; Q p-value = Cochran's Q heterogeneity test; Egger's p = linear regression test for funnel plot asymmetry; RoB = Risk of Bias* | | | | | | | | | | | | | | | | | | | |

###### 6.13.1.3 Wald Tests

chloride_ma_2nd_analysis$wald_tests$sex_comparison

Table 1: **Sex Comparison (Overall)**

| Comparison | % Shift | P (Wald Test) | Mean Diff (95% CI) |
| --- | --- | --- | --- |
| Male vs Female (Overall) | +19.3% | 0.646 | 23.56 [-77.04, 124.16] |
| *Mean difference = Group 1 − Group 2 in component units (mmol/24h, L/24h, or pH); % shift = difference relative to Group 2.* | | | |

chloride_ma_2nd_analysis$wald_tests$age_band_comparisons

Table 1: **Age Band Comparisons Within Sex Groups**

| Sex Group | Comparison | % Shift | P (Wald Test) | Mean Diff (95% CI) |
| --- | --- | --- | --- | --- |
| main meta-analysis | 18+ vs 20-30 | -3.1% | 0.932 | -4.55 [-108.82, 99.73] |
| main meta-analysis | 18+ vs 30-40 | -0.5% | 0.988 | -0.74 [-97.15, 95.67] |
| main meta-analysis | 18+ vs 40-50 | -6.1% | 0.856 | -9.11 [-107.46, 89.25] |
| main meta-analysis | 18+ vs 50-60 | -1.8% | 0.962 | -2.59 [-109.93, 104.76] |
| main meta-analysis | 18+ vs 60+ | +32.0% | 0.626 | 34.07 [-102.84, 170.98] |
| main meta-analysis | 20-30 vs 30-40 | +2.7% | 0.885 | 3.81 [-48.01, 55.63] |
| main meta-analysis | 20-30 vs 40-50 | -3.0% | 0.872 | -4.56 [-59.92, 50.79] |
| main meta-analysis | 20-30 vs 50-60 | +1.4% | 0.956 | 1.96 [-68.13, 72.05] |
| main meta-analysis | 20-30 vs 60+ | +36.3% | 0.492 | 38.62 [-71.54, 148.77] |
| main meta-analysis | 30-40 vs 40-50 | -5.6% | 0.670 | -8.37 [-46.91, 30.16] |
| main meta-analysis | 30-40 vs 50-60 | -1.3% | 0.950 | -1.85 [-59.58, 55.89] |
| main meta-analysis | 30-40 vs 60+ | +32.7% | 0.507 | 34.81 [-67.93, 137.55] |
| main meta-analysis | 40-50 vs 50-60 | +4.6% | 0.834 | 6.52 [-54.41, 67.45] |
| main meta-analysis | 40-50 vs 60+ | +40.5% | 0.418 | 43.18 [-61.38, 147.75] |
| main meta-analysis | 50-60 vs 60+ | +34.4% | 0.525 | 36.66 [-76.40, 149.72] |
| male | 18+ vs 20-30 | -13.5% | 0.545 | -20.63 [-87.51, 46.25] |
| male | 18+ vs 30-40 | -11.8% | 0.490 | -17.73 [-68.09, 32.63] |
| male | 18+ vs 40-50 | -12.8% | 0.474 | -19.43 [-72.61, 33.75] |
| male | 18+ vs 50-60 | -18.6% | 0.285 | -30.23 [-85.66, 25.20] |
| male | 18+ vs 60+ | -7.4% | 0.689 | -10.53 [-62.03, 40.97] |
| male | 20-30 vs 30-40 | +1.9% | 0.907 | 2.90 [-45.77, 51.57] |
| male | 20-30 vs 40-50 | +0.8% | 0.964 | 1.20 [-50.38, 52.78] |
| male | 20-30 vs 50-60 | -5.9% | 0.727 | -9.60 [-63.50, 44.30] |
| male | 20-30 vs 60+ | +7.1% | 0.691 | 10.10 [-39.75, 59.95] |
| male | 30-40 vs 40-50 | -1.1% | 0.901 | -1.70 [-28.61, 25.21] |
| male | 30-40 vs 50-60 | -7.7% | 0.431 | -12.50 [-43.63, 18.63] |
| male | 30-40 vs 60+ | +5.0% | 0.547 | 7.20 [-16.22, 30.62] |
| male | 40-50 vs 50-60 | -6.7% | 0.551 | -10.80 [-46.31, 24.71] |
| male | 40-50 vs 60+ | +6.2% | 0.547 | 8.90 [-20.09, 37.89] |
| male | 50-60 vs 60+ | +13.8% | 0.241 | 19.70 [-13.24, 52.64] |
| female | 18+ vs 20-30 | +1.1% | 0.981 | 1.41 [-116.68, 119.50] |
| female | 18+ vs 30-40 | -5.5% | 0.904 | -7.29 [-126.22, 111.63] |
| female | 18+ vs 40-50 | -5.9% | 0.897 | -7.79 [-126.31, 110.73] |
| female | 18+ vs 50-60 | +1.0% | 0.984 | 1.21 [-117.01, 119.43] |
| female | 18+ vs 60+ | +78.0% | 0.360 | 54.91 [-62.59, 172.41] |
| female | 20-30 vs 30-40 | -6.6% | 0.568 | -8.70 [-38.54, 21.14] |
| female | 20-30 vs 40-50 | -6.9% | 0.522 | -9.20 [-37.39, 18.99] |
| female | 20-30 vs 50-60 | -0.2% | 0.988 | -0.20 [-27.09, 26.69] |
| female | 20-30 vs 60+ | +76.0% | <0.001 | 53.50 [29.96, 77.04] |
| female | 30-40 vs 40-50 | -0.4% | 0.975 | -0.50 [-32.00, 31.00] |
| female | 30-40 vs 50-60 | +6.8% | 0.583 | 8.50 [-21.84, 38.84] |
| female | 30-40 vs 60+ | +88.4% | <0.001 | 62.20 [34.79, 89.61] |
| female | 40-50 vs 50-60 | +7.3% | 0.539 | 9.00 [-19.72, 37.72] |
| female | 40-50 vs 60+ | +89.1% | <0.001 | 62.70 [37.09, 88.31] |
| female | 50-60 vs 60+ | +76.3% | <0.001 | 53.70 [29.53, 77.87] |
| *Mean difference = Group 1 − Group 2 in component units (mmol/24h, L/24h, or pH); % shift = difference relative to Group 2.* | | | | |

chloride_ma_2nd_analysis$wald_tests$sex_within_age_bands

Table 1: **Sex Comparisons Within Age Bands**

| Age Band | % Shift | P (Wald Test) | Mean Diff (95% CI) |
| --- | --- | --- | --- |
| 18+ | +5.4% | 0.916 | 6.76 [-119.41, 132.93] |
| 20-30 | +23.2% | 0.259 | 28.80 [-21.19, 78.79] |
| 30-40 | +13.0% | 0.221 | 17.20 [-10.37, 44.77] |
| 40-50 | +13.8% | 0.243 | 18.40 [-12.52, 49.32] |
| 50-60 | +30.8% | <0.05 | 38.20 [4.59, 71.81] |
| 60+ | +102.6% | <0.001 | 72.20 [48.97, 95.43] |
| *Mean difference = Group 1 − Group 2 in component units (mmol/24h, L/24h, or pH); % shift = difference relative to Group 2.* | | | |

###### 6.13.1.4 Meta-Regression (plain language)

format_meta_reg_plain("chloride", sex_group = "all")

Message

No meta-regression result for chloride / all

###### 6.13.1.5 Assay Subgroup Analysis

assay_subgroup_table("chloride")

Table 1: **Assay subgroup analysis — Chloride**

*Per-assay pooled estimates; categories with k < 3 listed but not pooled.*

| Assay category | Studies, n | Participants | 95% CI | I² | Q p-value | Egger's p | Note |
| --- | --- | --- | --- | --- | --- | --- | --- |
| Unreported | 44 | 3,337 | 146.12 [134.36, 157.88] | 99% | <0.001 | 0.269 |  |
| Ion-selective electrode | 5 | 104 | 86.23 [44.15, 128.32] | 99% | <0.001 | 0.822 |  |
| Autoanalyser | 1 | 38 | — | — | — | — | k = 1 < min_k = 3 (skipped) |
| Coulometric / silver | 1 | 55 | — | — | — | — | k = 1 < min_k = 3 (skipped) |

###### 6.13.1.6 Preservative Sensitivity (Litholink / acidified samples)

```
preservative_sensitivity_ma("chloride")
## # A tibble: 3 × 6
##   group               n_studies n_participants pooled_estimate lower_95 upper_95
##   <chr>                   <int>          <int>           <dbl>    <dbl>    <dbl>
## 1 All studies                51           3534            140.     128.     152.
## 2 Excluding preserva…        51           3534            140.     128.     152.
## 3 Preservative only           0              0             NA       NA       NA
```

###### 6.13.1.7 Creatinine Completeness Sensitivity

creat_completeness_table("chloride")

Table 1: **Creatinine-completeness sensitivity — Chloride**

*Compares the all-studies pooled estimate against subsets defined by whether the study reported 24h urine creatinine. Studies that do not report creatinine cannot verify collection completeness; a substantial shift here flags possible bias from incomplete collections.*

| Subset | Studies, n | Participants | Range (95% CI) | Δ vs all-studies (%) | Flag |
| --- | --- | --- | --- | --- | --- |
| All studies | 51 | 3,534 | 139.96 [127.80, 152.11] | — | — |
| Creatinine reported only | 48 | 3,493 | 142.33 [129.69, 154.96] | +1.7% | OK |
| Creatinine NOT reported | 3 | 41 | 104.46 [82.73, 126.20] | -25.4% | ⚠ shift > 20% |

##### 6.13.2 Comparison by Age / Sex / race

###### 6.13.2.1 Plot

```
chloride_ma_3rd_analysis <- sex_age_race_subdivision_ma("chloride",
                                                      urine_ma2,
                                                      overall_bias)
## Not enough studies for multivariable meta-regression (k = 3, need > 6). Running univariable models instead.
## Not enough studies for multivariable meta-regression (k = 6, need > 6). Running univariable models instead.
chloride_ma_3rd_analysis$race_plot
## `height` was translated to `width`.
## `height` was translated to `width`.
## `height` was translated to `width`.
```

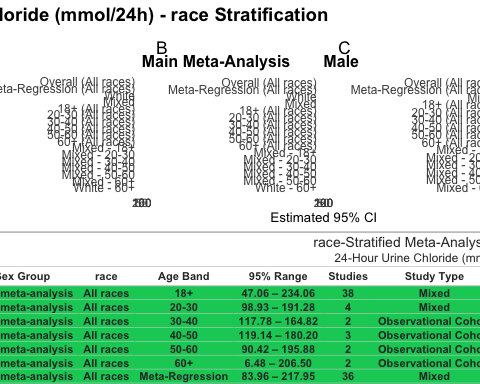

###### 6.13.2.2 Combined MA Results

chloride_ma_3rd_analysis$combined_summary_table

Table 1: **Combined Meta-Analysis Results**

24-Hour Urine Chloride (mmol/24h)

| Sex Group | race | Age Band | Pooled | 95% Range | Studies Mean SD Range | Studies | Study Type | Participants | I² | Cochran's Q | Q (p-value) | Egger's p | Model |
| --- | --- | --- | --- | --- | --- | --- | --- | --- | --- | --- | --- | --- | --- |
| main meta-analysis | All | 18+ | 140.56 | 47.06 – 234.06 | 144.66 ± 50.27 (35.21–205.68) | 38 | Mixed | 3204 | 98.4% | 2274.4 | <0.005 | <0.01 | Random |
| main meta-analysis | All | 20-30 | 145.11 | 98.93 – 191.28 | 146.72 ± 25.64 (123.90–178.40) | 4 | Mixed | 96 | 98.3% | 176.8 | <0.005 | 0.82 | Random |
| main meta-analysis | All | 30-40 | 141.30 | 117.78 – 164.82 | 141.20 ± 12.16 (132.60–149.80) | 2 | Observational Cohort | 50 | 97.3% | 37.4 | <0.005 | — | Random |
| main meta-analysis | All | 40-50 | 149.67 | 119.14 – 180.20 | 149.63 ± 15.68 (133.10–164.30) | 3 | Observational Cohort | 84 | 98.5% | 133.3 | <0.005 | 0.44 | Random |
| main meta-analysis | All | 50-60 | 143.15 | 90.42 – 195.88 | 143.20 ± 27.01 (124.10–162.30) | 2 | Observational Cohort | 50 | 99.2% | 124.1 | <0.005 | — | Random |
| main meta-analysis | All | 60+ | 106.49 | 6.48 – 206.50 | 106.50 ± 51.05 (70.40–142.60) | 2 | Observational Cohort | 50 | 99.9% | 927.5 | <0.005 | — | Random |
| **main meta-analysis** | **All** | **Meta-Regression** | **150.95** | **83.96 – 217.95** | **—** | **36** | **Mixed** | **3173** | **94.2%** | **553.3** | **<0.005** | **0.80** | **Multivariable** |
| **main meta-analysis** | **All** | **Meta-Regression** | **150.95** | **83.96 – 217.95** | **—** | **36** | **Mixed** | **3173** | **94.2%** | **553.3** | **<0.005** | **0.80** | **Multivariable** |
| **main meta-analysis** | **All** | **Overall** | **139.96** | **56.59 – 223.32** | **143.42 ± 45.26 (35.21–205.68)** | **51** | **Mixed** | **3534** | **99%** | **5197.7** | **<0.005** | **0.53** | **Random** |
| **main meta-analysis** | **All** | **Overall** | **139.96** | **56.59 – 223.32** | **143.42 ± 45.26 (35.21–205.68)** | **51** | **Mixed** | **3534** | **99%** | **5197.7** | **<0.005** | **0.53** | **Random** |
| main meta-analysis | black | 18+ | — | NA – NA | — | NA | NA | NA | — | — | — | — | NA |
| main meta-analysis | black | 20-30 | — | NA – NA | — | NA | NA | NA | — | — | — | — | NA |
| main meta-analysis | black | 30-40 | — | NA – NA | — | NA | NA | NA | — | — | — | — | NA |
| main meta-analysis | black | 40-50 | — | NA – NA | — | NA | NA | NA | — | — | — | — | NA |
| main meta-analysis | black | 50-60 | — | NA – NA | — | NA | NA | NA | — | — | — | — | NA |
| main meta-analysis | black | 60+ | — | NA – NA | — | NA | NA | NA | — | — | — | — | NA |
| **main meta-analysis** | **black** | **Meta-Regression** | **—** | **NA – NA** | **—** | **NA** | **NA** | **NA** | **—** | **—** | **—** | **—** | **NA** |
| **main meta-analysis** | **black** | **Overall** | **—** | **NA – NA** | **—** | **NA** | **NA** | **NA** | **—** | **—** | **—** | **—** | **NA** |
| main meta-analysis | mixed | 18+ | 140.56 | 47.06 – 234.06 | 144.66 ± 50.27 (35.21–205.68) | 38 | Mixed | 3204 | 98.4% | 2274.4 | <0.005 | <0.01 | Random |
| main meta-analysis | mixed | 20-30 | 145.11 | 98.93 – 191.28 | 146.72 ± 25.64 (123.90–178.40) | 4 | Mixed | 96 | 98.3% | 176.8 | <0.005 | 0.82 | Random |
| main meta-analysis | mixed | 30-40 | 141.30 | 117.78 – 164.82 | 141.20 ± 12.16 (132.60–149.80) | 2 | Observational Cohort | 50 | 97.3% | 37.4 | <0.005 | — | Random |
| main meta-analysis | mixed | 40-50 | 149.67 | 119.14 – 180.20 | 149.63 ± 15.68 (133.10–164.30) | 3 | Observational Cohort | 84 | 98.5% | 133.3 | <0.005 | 0.44 | Random |
| main meta-analysis | mixed | 50-60 | 143.15 | 90.42 – 195.88 | 143.20 ± 27.01 (124.10–162.30) | 2 | Observational Cohort | 50 | 99.2% | 124.1 | <0.005 | — | Random |
| main meta-analysis | mixed | 60+ | 70.40 | 55.99 – 84.81 | — (70.40–70.40) | 1 | Observational Cohort | 25 | — | — | — | — | Single study |
| **main meta-analysis** | **mixed** | **Meta-Regression** | **—** | **NA – NA** | **—** | **NA** | **NA** | **NA** | **—** | **—** | **—** | **—** | **NA** |
| **main meta-analysis** | **mixed** | **Overall** | **139.94** | **55.62 – 224.25** | **143.44 ± 45.71 (35.21–205.68)** | **50** | **Mixed** | **3509** | **99%** | **5148.3** | **<0.005** | **0.48** | **Random** |
| main meta-analysis | white | 18+ | — | NA – NA | — | NA | NA | NA | — | — | — | — | NA |
| main meta-analysis | white | 20-30 | — | NA – NA | — | NA | NA | NA | — | — | — | — | NA |
| main meta-analysis | white | 30-40 | — | NA – NA | — | NA | NA | NA | — | — | — | — | NA |
| main meta-analysis | white | 40-50 | — | NA – NA | — | NA | NA | NA | — | — | — | — | NA |
| main meta-analysis | white | 50-60 | — | NA – NA | — | NA | NA | NA | — | — | — | — | NA |
| main meta-analysis | white | 60+ | 142.60 | 124.37 – 160.83 | — (142.60–142.60) | 1 | Observational Cohort | 25 | — | — | — | — | Single study |
| **main meta-analysis** | **white** | **Meta-Regression** | **—** | **NA – NA** | **—** | **NA** | **NA** | **NA** | **—** | **—** | **—** | **—** | **NA** |
| **main meta-analysis** | **white** | **Overall** | **142.60** | **124.37 – 160.83** | **— (142.60–142.60)** | **1** | **Observational Cohort** | **25** | **—** | **—** | **—** | **—** | **Single study** |
| male | All | 18+ | 132.07 | 83.90 – 180.23 | 130.91 ± 24.70 (118.33–167.96) | 4 | Observational Cohort | 168 | 95.8% | 71.6 | <0.005 | 0.87 | Random |
| male | All | 20-30 | 152.70 | 106.30 – 199.10 | 154.33 ± 25.28 (128.00–178.40) | 3 | Mixed | 71 | 97.8% | 90.8 | <0.005 | 0.95 | Random |
| male | All | 30-40 | 149.80 | 135.10 – 164.50 | — (149.80–149.80) | 1 | Observational Cohort | 25 | — | — | — | — | Single study |
| male | All | 40-50 | 151.50 | 128.96 – 174.04 | — (151.50–151.50) | 1 | Observational Cohort | 25 | — | — | — | — | Single study |
| male | All | 50-60 | 162.30 | 134.86 – 189.74 | — (162.30–162.30) | 1 | Observational Cohort | 25 | — | — | — | — | Single study |
| male | All | 60+ | 142.60 | 124.37 – 160.83 | — (142.60–142.60) | 1 | Observational Cohort | 25 | — | — | — | — | Single study |
| **male** | **All** | **Meta-Regression** | **167.96** | **117.00 – 218.92** | **—** | **3** | **Mixed** | **155** | **96.3%** | **64** | **<0.005** | **0.36** | **Univariable** |
| **male** | **All** | **Overall** | **145.43** | **109.16 – 181.70** | **144.80 ± 21.29 (118.33–178.40)** | **11** | **Mixed** | **339** | **95.9%** | **241.8** | **<0.005** | **0.77** | **Random** |
| male | black | 18+ | — | NA – NA | — | NA | NA | NA | — | — | — | — | NA |
| male | black | 20-30 | — | NA – NA | — | NA | NA | NA | — | — | — | — | NA |
| male | black | 30-40 | — | NA – NA | — | NA | NA | NA | — | — | — | — | NA |
| male | black | 40-50 | — | NA – NA | — | NA | NA | NA | — | — | — | — | NA |
| male | black | 50-60 | — | NA – NA | — | NA | NA | NA | — | — | — | — | NA |
| male | black | 60+ | — | NA – NA | — | NA | NA | NA | — | — | — | — | NA |
| **male** | **black** | **Meta-Regression** | **—** | **NA – NA** | **—** | **NA** | **NA** | **NA** | **—** | **—** | **—** | **—** | **NA** |
| **male** | **black** | **Overall** | **—** | **NA – NA** | **—** | **NA** | **NA** | **NA** | **—** | **—** | **—** | **—** | **NA** |
| male | mixed | 18+ | 132.07 | 83.90 – 180.23 | 130.91 ± 24.70 (118.33–167.96) | 4 | Observational Cohort | 168 | 95.8% | 71.6 | <0.005 | 0.87 | Random |
| male | mixed | 20-30 | 152.70 | 106.30 – 199.10 | 154.33 ± 25.28 (128.00–178.40) | 3 | Mixed | 71 | 97.8% | 90.8 | <0.005 | 0.95 | Random |
| male | mixed | 30-40 | 149.80 | 135.10 – 164.50 | — (149.80–149.80) | 1 | Observational Cohort | 25 | — | — | — | — | Single study |
| male | mixed | 40-50 | 151.50 | 128.96 – 174.04 | — (151.50–151.50) | 1 | Observational Cohort | 25 | — | — | — | — | Single study |
| male | mixed | 50-60 | 162.30 | 134.86 – 189.74 | — (162.30–162.30) | 1 | Observational Cohort | 25 | — | — | — | — | Single study |
| male | mixed | 60+ | — | NA – NA | — | NA | NA | NA | — | — | — | — | NA |
| **male** | **mixed** | **Meta-Regression** | **—** | **NA – NA** | **—** | **NA** | **NA** | **NA** | **—** | **—** | **—** | **—** | **NA** |
| **male** | **mixed** | **Overall** | **145.68** | **106.81 – 184.54** | **145.02 ± 22.43 (118.33–178.40)** | **10** | **Mixed** | **314** | **96.2%** | **234.7** | **<0.005** | **0.73** | **Random** |
| male | white | 18+ | — | NA – NA | — | NA | NA | NA | — | — | — | — | NA |
| male | white | 20-30 | — | NA – NA | — | NA | NA | NA | — | — | — | — | NA |
| male | white | 30-40 | — | NA – NA | — | NA | NA | NA | — | — | — | — | NA |
| male | white | 40-50 | — | NA – NA | — | NA | NA | NA | — | — | — | — | NA |
| male | white | 50-60 | — | NA – NA | — | NA | NA | NA | — | — | — | — | NA |
| male | white | 60+ | 142.60 | 124.37 – 160.83 | — (142.60–142.60) | 1 | Observational Cohort | 25 | — | — | — | — | Single study |
| **male** | **white** | **Meta-Regression** | **—** | **NA – NA** | **—** | **NA** | **NA** | **NA** | **—** | **—** | **—** | **—** | **NA** |
| **male** | **white** | **Overall** | **142.60** | **124.37 – 160.83** | **— (142.60–142.60)** | **1** | **Observational Cohort** | **25** | **—** | **—** | **—** | **—** | **Single study** |
| female | All | 18+ | 125.31 | 8.69 – 241.92 | 125.70 ± 59.62 (35.21–185.58) | 8 | Observational Cohort | 528 | 99.2% | 833.3 | <0.005 | <0.001 | Random |
| female | All | 20-30 | 123.90 | 105.28 – 142.52 | — (123.90–123.90) | 1 | Observational Cohort | 25 | — | — | — | — | Single study |
| female | All | 30-40 | 132.60 | 109.28 – 155.92 | — (132.60–132.60) | 1 | Observational Cohort | 25 | — | — | — | — | Single study |
| female | All | 40-50 | 133.10 | 111.93 – 154.27 | — (133.10–133.10) | 1 | Observational Cohort | 25 | — | — | — | — | Single study |
| female | All | 50-60 | 124.10 | 104.70 – 143.50 | — (124.10–124.10) | 1 | Observational Cohort | 25 | — | — | — | — | Single study |
| female | All | 60+ | 70.40 | 55.99 – 84.81 | — (70.40–70.40) | 1 | Observational Cohort | 25 | — | — | — | — | Single study |
| **female** | **All** | **Meta-Regression** | **185.58** | **140.39 – 230.77** | **—** | **6** | **Observational Cohort** | **476** | **88.7%** | **41.6** | **<0.005** | **<0.05** | **Univariable** |
| **female** | **All** | **Overall** | **121.87** | **28.04 – 215.70** | **122.28 ± 48.22 (35.21–185.58)** | **13** | **Observational Cohort** | **653** | **99.4%** | **2057.3** | **<0.005** | **0.42** | **Random** |
| female | black | 18+ | — | NA – NA | — | NA | NA | NA | — | — | — | — | NA |
| female | black | 20-30 | — | NA – NA | — | NA | NA | NA | — | — | — | — | NA |
| female | black | 30-40 | — | NA – NA | — | NA | NA | NA | — | — | — | — | NA |
| female | black | 40-50 | — | NA – NA | — | NA | NA | NA | — | — | — | — | NA |
| female | black | 50-60 | — | NA – NA | — | NA | NA | NA | — | — | — | — | NA |
| female | black | 60+ | — | NA – NA | — | NA | NA | NA | — | — | — | — | NA |
| **female** | **black** | **Meta-Regression** | **—** | **NA – NA** | **—** | **NA** | **NA** | **NA** | **—** | **—** | **—** | **—** | **NA** |
| **female** | **black** | **Overall** | **—** | **NA – NA** | **—** | **NA** | **NA** | **NA** | **—** | **—** | **—** | **—** | **NA** |
| female | mixed | 18+ | 125.31 | 8.69 – 241.92 | 125.70 ± 59.62 (35.21–185.58) | 8 | Observational Cohort | 528 | 99.2% | 833.3 | <0.005 | <0.001 | Random |
| female | mixed | 20-30 | 123.90 | 105.28 – 142.52 | — (123.90–123.90) | 1 | Observational Cohort | 25 | — | — | — | — | Single study |
| female | mixed | 30-40 | 132.60 | 109.28 – 155.92 | — (132.60–132.60) | 1 | Observational Cohort | 25 | — | — | — | — | Single study |
| female | mixed | 40-50 | 133.10 | 111.93 – 154.27 | — (133.10–133.10) | 1 | Observational Cohort | 25 | — | — | — | — | Single study |
| female | mixed | 50-60 | 124.10 | 104.70 – 143.50 | — (124.10–124.10) | 1 | Observational Cohort | 25 | — | — | — | — | Single study |
| female | mixed | 60+ | 70.40 | 55.99 – 84.81 | — (70.40–70.40) | 1 | Observational Cohort | 25 | — | — | — | — | Single study |
| **female** | **mixed** | **Meta-Regression** | **—** | **NA – NA** | **—** | **NA** | **NA** | **NA** | **—** | **—** | **—** | **—** | **NA** |
| **female** | **mixed** | **Overall** | **121.87** | **28.04 – 215.70** | **122.28 ± 48.22 (35.21–185.58)** | **13** | **Observational Cohort** | **653** | **99.4%** | **2057.3** | **<0.005** | **0.42** | **Random** |
| female | white | 18+ | — | NA – NA | — | NA | NA | NA | — | — | — | — | NA |
| female | white | 20-30 | — | NA – NA | — | NA | NA | NA | — | — | — | — | NA |
| female | white | 30-40 | — | NA – NA | — | NA | NA | NA | — | — | — | — | NA |
| female | white | 40-50 | — | NA – NA | — | NA | NA | NA | — | — | — | — | NA |
| female | white | 50-60 | — | NA – NA | — | NA | NA | NA | — | — | — | — | NA |
| female | white | 60+ | — | NA – NA | — | NA | NA | NA | — | — | — | — | NA |
| **female** | **white** | **Meta-Regression** | **—** | **NA – NA** | **—** | **NA** | **NA** | **NA** | **—** | **—** | **—** | **—** | **NA** |
| **female** | **white** | **Overall** | **—** | **NA – NA** | **—** | **NA** | **NA** | **NA** | **—** | **—** | **—** | **—** | **NA** |
| *I² represents heterogeneity; Q test assesses between-study variation; Egger's p = linear regression test for funnel plot asymmetry; Meta-Regression shows covariate-adjusted estimates* | | | | | | | | | | | | | |

###### 6.13.2.3 Table of Results subdivided by race / Sex / Age

chloride_ma_3rd_analysis$race_summary_table

Table 1: **race-Stratified Meta-Analysis Results**

24-Hour Urine Chloride (mmol/24h)

| Sex Group | race | Age Band | 95% Range | Studies | Study Type | Participants | I² | Cochran's Q | Q (p-value) | Egger's p | Model |
| --- | --- | --- | --- | --- | --- | --- | --- | --- | --- | --- | --- |
| **main meta-analysis** | **All races** | **18+** | **47.06 – 234.06** | **38** | **Mixed** | **3204** | **98.4%** | **2274.4** | **<0.005** | **<0.01** | **Random** |
| **main meta-analysis** | **All races** | **20-30** | **98.93 – 191.28** | **4** | **Mixed** | **96** | **98.3%** | **176.8** | **<0.005** | **0.82** | **Random** |
| **main meta-analysis** | **All races** | **30-40** | **117.78 – 164.82** | **2** | **Observational Cohort** | **50** | **97.3%** | **37.4** | **<0.005** | **—** | **Random** |
| **main meta-analysis** | **All races** | **40-50** | **119.14 – 180.20** | **3** | **Observational Cohort** | **84** | **98.5%** | **133.3** | **<0.005** | **0.44** | **Random** |
| **main meta-analysis** | **All races** | **50-60** | **90.42 – 195.88** | **2** | **Observational Cohort** | **50** | **99.2%** | **124.1** | **<0.005** | **—** | **Random** |
| **main meta-analysis** | **All races** | **60+** | **6.48 – 206.50** | **2** | **Observational Cohort** | **50** | **99.9%** | **927.5** | **<0.005** | **—** | **Random** |
| **main meta-analysis** | **All races** | **Meta-Regression** | **83.96 – 217.95** | **36** | **Mixed** | **3173** | **94.2%** | **553.3** | **<0.005** | **0.80** | **Multivariable** |
| **main meta-analysis** | **All races** | **Meta-Regression** | **83.96 – 217.95** | **36** | **Mixed** | **3173** | **94.2%** | **553.3** | **<0.005** | **0.80** | **Multivariable** |
| **main meta-analysis** | **All races** | **Overall** | **56.59 – 223.32** | **51** | **Mixed** | **3534** | **99%** | **5197.7** | **<0.005** | **0.53** | **Random** |
| **main meta-analysis** | **All races** | **Overall** | **56.59 – 223.32** | **51** | **Mixed** | **3534** | **99%** | **5197.7** | **<0.005** | **0.53** | **Random** |
| main meta-analysis | Black | 18+ | — | — | NA | — | — | — | — | — | — |
| main meta-analysis | Black | 20-30 | — | — | NA | — | — | — | — | — | — |
| main meta-analysis | Black | 30-40 | — | — | NA | — | — | — | — | — | — |
| main meta-analysis | Black | 40-50 | — | — | NA | — | — | — | — | — | — |
| main meta-analysis | Black | 50-60 | — | — | NA | — | — | — | — | — | — |
| main meta-analysis | Black | 60+ | — | — | NA | — | — | — | — | — | — |
| main meta-analysis | Black | Meta-Regression | — | — | NA | — | — | — | — | — | — |
| **main meta-analysis** | **Black** | **Overall** | **—** | **—** | **NA** | **—** | **—** | **—** | **—** | **—** | **—** |
| main meta-analysis | Mixed | 18+ | 47.06 – 234.06 | 38 | Mixed | 3204 | 98.4% | 2274.4 | <0.005 | <0.01 | Random |
| main meta-analysis | Mixed | 20-30 | 98.93 – 191.28 | 4 | Mixed | 96 | 98.3% | 176.8 | <0.005 | 0.82 | Random |
| main meta-analysis | Mixed | 30-40 | 117.78 – 164.82 | 2 | Observational Cohort | 50 | 97.3% | 37.4 | <0.005 | — | Random |
| main meta-analysis | Mixed | 40-50 | 119.14 – 180.20 | 3 | Observational Cohort | 84 | 98.5% | 133.3 | <0.005 | 0.44 | Random |
| main meta-analysis | Mixed | 50-60 | 90.42 – 195.88 | 2 | Observational Cohort | 50 | 99.2% | 124.1 | <0.005 | — | Random |
| main meta-analysis | Mixed | 60+ | 55.99 – 84.81 | 1 | Observational Cohort | 25 | — | — | — | — | Single study |
| main meta-analysis | Mixed | Meta-Regression | — | — | NA | — | — | — | — | — | — |
| **main meta-analysis** | **Mixed** | **Overall** | **55.62 – 224.25** | **50** | **Mixed** | **3509** | **99%** | **5148.3** | **<0.005** | **0.48** | **Random** |
| main meta-analysis | White | 18+ | — | — | NA | — | — | — | — | — | — |
| main meta-analysis | White | 20-30 | — | — | NA | — | — | — | — | — | — |
| main meta-analysis | White | 30-40 | — | — | NA | — | — | — | — | — | — |
| main meta-analysis | White | 40-50 | — | — | NA | — | — | — | — | — | — |
| main meta-analysis | White | 50-60 | — | — | NA | — | — | — | — | — | — |
| main meta-analysis | White | 60+ | 124.37 – 160.83 | 1 | Observational Cohort | 25 | — | — | — | — | Single study |
| main meta-analysis | White | Meta-Regression | — | — | NA | — | — | — | — | — | — |
| **main meta-analysis** | **White** | **Overall** | **124.37 – 160.83** | **1** | **Observational Cohort** | **25** | **—** | **—** | **—** | **—** | **Single study** |
| **male** | **All races** | **18+** | **83.90 – 180.23** | **4** | **Observational Cohort** | **168** | **95.8%** | **71.6** | **<0.005** | **0.87** | **Random** |
| **male** | **All races** | **20-30** | **106.30 – 199.10** | **3** | **Mixed** | **71** | **97.8%** | **90.8** | **<0.005** | **0.95** | **Random** |
| **male** | **All races** | **30-40** | **135.10 – 164.50** | **1** | **Observational Cohort** | **25** | **—** | **—** | **—** | **—** | **Single study** |
| **male** | **All races** | **40-50** | **128.96 – 174.04** | **1** | **Observational Cohort** | **25** | **—** | **—** | **—** | **—** | **Single study** |
| **male** | **All races** | **50-60** | **134.86 – 189.74** | **1** | **Observational Cohort** | **25** | **—** | **—** | **—** | **—** | **Single study** |
| **male** | **All races** | **60+** | **124.37 – 160.83** | **1** | **Observational Cohort** | **25** | **—** | **—** | **—** | **—** | **Single study** |
| **male** | **All races** | **Meta-Regression** | **117.00 – 218.92** | **3** | **Mixed** | **155** | **96.3%** | **64** | **<0.005** | **0.36** | **Univariable** |
| **male** | **All races** | **Overall** | **109.16 – 181.70** | **11** | **Mixed** | **339** | **95.9%** | **241.8** | **<0.005** | **0.77** | **Random** |
| male | Black | 18+ | — | — | NA | — | — | — | — | — | — |
| male | Black | 20-30 | — | — | NA | — | — | — | — | — | — |
| male | Black | 30-40 | — | — | NA | — | — | — | — | — | — |
| male | Black | 40-50 | — | — | NA | — | — | — | — | — | — |
| male | Black | 50-60 | — | — | NA | — | — | — | — | — | — |
| male | Black | 60+ | — | — | NA | — | — | — | — | — | — |
| male | Black | Meta-Regression | — | — | NA | — | — | — | — | — | — |
| **male** | **Black** | **Overall** | **—** | **—** | **NA** | **—** | **—** | **—** | **—** | **—** | **—** |
| male | Mixed | 18+ | 83.90 – 180.23 | 4 | Observational Cohort | 168 | 95.8% | 71.6 | <0.005 | 0.87 | Random |
| male | Mixed | 20-30 | 106.30 – 199.10 | 3 | Mixed | 71 | 97.8% | 90.8 | <0.005 | 0.95 | Random |
| male | Mixed | 30-40 | 135.10 – 164.50 | 1 | Observational Cohort | 25 | — | — | — | — | Single study |
| male | Mixed | 40-50 | 128.96 – 174.04 | 1 | Observational Cohort | 25 | — | — | — | — | Single study |
| male | Mixed | 50-60 | 134.86 – 189.74 | 1 | Observational Cohort | 25 | — | — | — | — | Single study |
| male | Mixed | 60+ | — | — | NA | — | — | — | — | — | — |
| male | Mixed | Meta-Regression | — | — | NA | — | — | — | — | — | — |
| **male** | **Mixed** | **Overall** | **106.81 – 184.54** | **10** | **Mixed** | **314** | **96.2%** | **234.7** | **<0.005** | **0.73** | **Random** |
| male | White | 18+ | — | — | NA | — | — | — | — | — | — |
| male | White | 20-30 | — | — | NA | — | — | — | — | — | — |
| male | White | 30-40 | — | — | NA | — | — | — | — | — | — |
| male | White | 40-50 | — | — | NA | — | — | — | — | — | — |
| male | White | 50-60 | — | — | NA | — | — | — | — | — | — |
| male | White | 60+ | 124.37 – 160.83 | 1 | Observational Cohort | 25 | — | — | — | — | Single study |
| male | White | Meta-Regression | — | — | NA | — | — | — | — | — | — |
| **male** | **White** | **Overall** | **124.37 – 160.83** | **1** | **Observational Cohort** | **25** | **—** | **—** | **—** | **—** | **Single study** |
| **female** | **All races** | **18+** | **8.69 – 241.92** | **8** | **Observational Cohort** | **528** | **99.2%** | **833.3** | **<0.005** | **<0.001** | **Random** |
| **female** | **All races** | **20-30** | **105.28 – 142.52** | **1** | **Observational Cohort** | **25** | **—** | **—** | **—** | **—** | **Single study** |
| **female** | **All races** | **30-40** | **109.28 – 155.92** | **1** | **Observational Cohort** | **25** | **—** | **—** | **—** | **—** | **Single study** |
| **female** | **All races** | **40-50** | **111.93 – 154.27** | **1** | **Observational Cohort** | **25** | **—** | **—** | **—** | **—** | **Single study** |
| **female** | **All races** | **50-60** | **104.70 – 143.50** | **1** | **Observational Cohort** | **25** | **—** | **—** | **—** | **—** | **Single study** |
| **female** | **All races** | **60+** | **55.99 – 84.81** | **1** | **Observational Cohort** | **25** | **—** | **—** | **—** | **—** | **Single study** |
| **female** | **All races** | **Meta-Regression** | **140.39 – 230.77** | **6** | **Observational Cohort** | **476** | **88.7%** | **41.6** | **<0.005** | **<0.05** | **Univariable** |
| **female** | **All races** | **Overall** | **28.04 – 215.70** | **13** | **Observational Cohort** | **653** | **99.4%** | **2057.3** | **<0.005** | **0.42** | **Random** |
| female | Black | 18+ | — | — | NA | — | — | — | — | — | — |
| female | Black | 20-30 | — | — | NA | — | — | — | — | — | — |
| female | Black | 30-40 | — | — | NA | — | — | — | — | — | — |
| female | Black | 40-50 | — | — | NA | — | — | — | — | — | — |
| female | Black | 50-60 | — | — | NA | — | — | — | — | — | — |
| female | Black | 60+ | — | — | NA | — | — | — | — | — | — |
| female | Black | Meta-Regression | — | — | NA | — | — | — | — | — | — |
| **female** | **Black** | **Overall** | **—** | **—** | **NA** | **—** | **—** | **—** | **—** | **—** | **—** |
| female | Mixed | 18+ | 8.69 – 241.92 | 8 | Observational Cohort | 528 | 99.2% | 833.3 | <0.005 | <0.001 | Random |
| female | Mixed | 20-30 | 105.28 – 142.52 | 1 | Observational Cohort | 25 | — | — | — | — | Single study |
| female | Mixed | 30-40 | 109.28 – 155.92 | 1 | Observational Cohort | 25 | — | — | — | — | Single study |
| female | Mixed | 40-50 | 111.93 – 154.27 | 1 | Observational Cohort | 25 | — | — | — | — | Single study |
| female | Mixed | 50-60 | 104.70 – 143.50 | 1 | Observational Cohort | 25 | — | — | — | — | Single study |
| female | Mixed | 60+ | 55.99 – 84.81 | 1 | Observational Cohort | 25 | — | — | — | — | Single study |
| female | Mixed | Meta-Regression | — | — | NA | — | — | — | — | — | — |
| **female** | **Mixed** | **Overall** | **28.04 – 215.70** | **13** | **Observational Cohort** | **653** | **99.4%** | **2057.3** | **<0.005** | **0.42** | **Random** |
| female | White | 18+ | — | — | NA | — | — | — | — | — | — |
| female | White | 20-30 | — | — | NA | — | — | — | — | — | — |
| female | White | 30-40 | — | — | NA | — | — | — | — | — | — |
| female | White | 40-50 | — | — | NA | — | — | — | — | — | — |
| female | White | 50-60 | — | — | NA | — | — | — | — | — | — |
| female | White | 60+ | — | — | NA | — | — | — | — | — | — |
| female | White | Meta-Regression | — | — | NA | — | — | — | — | — | — |
| **female** | **White** | **Overall** | **—** | **—** | **NA** | **—** | **—** | **—** | **—** | **—** | **—** |
| *I² represents heterogeneity; Q test assesses between-study variation; Egger's p = linear regression test for funnel plot asymmetry; — indicates insufficient data* | | | | | | | | | | | |

###### 6.13.2.4 Wald Tests

chloride_ma_3rd_analysis$wald_tests$sex_comparison

Table 1: **Sex Comparison (Overall)**

| Comparison | % Shift | P (Wald Test) | Mean Diff (95% CI) |
| --- | --- | --- | --- |
| Male vs Female (Overall) | +19.3% | 0.646 | 23.56 [-77.04, 124.16] |
| *Mean difference = Group 1 − Group 2 in component units (mmol/24h, L/24h, or pH); % shift = difference relative to Group 2.* | | | |

chloride_ma_3rd_analysis$wald_tests$race_comparisons

Table 1: **race Comparisons Within Sex Groups**

| Sex Group | Comparison | % Shift | P (Wald Test) | Mean Diff (95% CI) |
| --- | --- | --- | --- | --- |
| male | black vs mixed | — | — | — |
| male | black vs white | — | — | — |
| male | mixed vs white | +2.2% | 0.888 | 3.08 [-39.85, 46.01] |
| female | black vs mixed | — | — | — |
| female | black vs white | — | — | — |
| female | mixed vs white | — | — | — |
| *Mean difference = Group 1 − Group 2 in component units (mmol/24h, L/24h, or pH); % shift = difference relative to Group 2.* | | | | |

chloride_ma_3rd_analysis$wald_tests$age_band_comparisons

Table 1: **Age Band Comparisons Within Sex Groups**

| Sex Group | Comparison | % Shift | P (Wald Test) | Mean Diff (95% CI) |
| --- | --- | --- | --- | --- |
| main meta-analysis | 18+ vs 20-30 | -3.1% | 0.932 | -4.55 [-108.82, 99.73] |
| main meta-analysis | 18+ vs 30-40 | -0.5% | 0.988 | -0.74 [-97.15, 95.67] |
| main meta-analysis | 18+ vs 40-50 | -6.1% | 0.856 | -9.11 [-107.46, 89.25] |
| main meta-analysis | 18+ vs 50-60 | -1.8% | 0.962 | -2.59 [-109.93, 104.76] |
| main meta-analysis | 18+ vs 60+ | +32.0% | 0.626 | 34.07 [-102.84, 170.98] |
| main meta-analysis | 20-30 vs 30-40 | +2.7% | 0.885 | 3.81 [-48.01, 55.63] |
| main meta-analysis | 20-30 vs 40-50 | -3.0% | 0.872 | -4.56 [-59.92, 50.79] |
| main meta-analysis | 20-30 vs 50-60 | +1.4% | 0.956 | 1.96 [-68.13, 72.05] |
| main meta-analysis | 20-30 vs 60+ | +36.3% | 0.492 | 38.62 [-71.54, 148.77] |
| main meta-analysis | 30-40 vs 40-50 | -5.6% | 0.670 | -8.37 [-46.91, 30.16] |
| main meta-analysis | 30-40 vs 50-60 | -1.3% | 0.950 | -1.85 [-59.58, 55.89] |
| main meta-analysis | 30-40 vs 60+ | +32.7% | 0.507 | 34.81 [-67.93, 137.55] |
| main meta-analysis | 40-50 vs 50-60 | +4.6% | 0.834 | 6.52 [-54.41, 67.45] |
| main meta-analysis | 40-50 vs 60+ | +40.5% | 0.418 | 43.18 [-61.38, 147.75] |
| main meta-analysis | 50-60 vs 60+ | +34.4% | 0.525 | 36.66 [-76.40, 149.72] |
| male | 18+ vs 20-30 | -13.5% | 0.545 | -20.63 [-87.51, 46.25] |
| male | 18+ vs 30-40 | -11.8% | 0.490 | -17.73 [-68.09, 32.63] |
| male | 18+ vs 40-50 | -12.8% | 0.474 | -19.43 [-72.61, 33.75] |
| male | 18+ vs 50-60 | -18.6% | 0.285 | -30.23 [-85.66, 25.20] |
| male | 18+ vs 60+ | -7.4% | 0.689 | -10.53 [-62.03, 40.97] |
| male | 20-30 vs 30-40 | +1.9% | 0.907 | 2.90 [-45.77, 51.57] |
| male | 20-30 vs 40-50 | +0.8% | 0.964 | 1.20 [-50.38, 52.78] |
| male | 20-30 vs 50-60 | -5.9% | 0.727 | -9.60 [-63.50, 44.30] |
| male | 20-30 vs 60+ | +7.1% | 0.691 | 10.10 [-39.75, 59.95] |
| male | 30-40 vs 40-50 | -1.1% | 0.901 | -1.70 [-28.61, 25.21] |
| male | 30-40 vs 50-60 | -7.7% | 0.431 | -12.50 [-43.63, 18.63] |
| male | 30-40 vs 60+ | +5.0% | 0.547 | 7.20 [-16.22, 30.62] |
| male | 40-50 vs 50-60 | -6.7% | 0.551 | -10.80 [-46.31, 24.71] |
| male | 40-50 vs 60+ | +6.2% | 0.547 | 8.90 [-20.09, 37.89] |
| male | 50-60 vs 60+ | +13.8% | 0.241 | 19.70 [-13.24, 52.64] |
| female | 18+ vs 20-30 | +1.1% | 0.981 | 1.41 [-116.68, 119.50] |
| female | 18+ vs 30-40 | -5.5% | 0.904 | -7.29 [-126.22, 111.63] |
| female | 18+ vs 40-50 | -5.9% | 0.897 | -7.79 [-126.31, 110.73] |
| female | 18+ vs 50-60 | +1.0% | 0.984 | 1.21 [-117.01, 119.43] |
| female | 18+ vs 60+ | +78.0% | 0.360 | 54.91 [-62.59, 172.41] |
| female | 20-30 vs 30-40 | -6.6% | 0.568 | -8.70 [-38.54, 21.14] |
| female | 20-30 vs 40-50 | -6.9% | 0.522 | -9.20 [-37.39, 18.99] |
| female | 20-30 vs 50-60 | -0.2% | 0.988 | -0.20 [-27.09, 26.69] |
| female | 20-30 vs 60+ | +76.0% | <0.001 | 53.50 [29.96, 77.04] |
| female | 30-40 vs 40-50 | -0.4% | 0.975 | -0.50 [-32.00, 31.00] |
| female | 30-40 vs 50-60 | +6.8% | 0.583 | 8.50 [-21.84, 38.84] |
| female | 30-40 vs 60+ | +88.4% | <0.001 | 62.20 [34.79, 89.61] |
| female | 40-50 vs 50-60 | +7.3% | 0.539 | 9.00 [-19.72, 37.72] |
| female | 40-50 vs 60+ | +89.1% | <0.001 | 62.70 [37.09, 88.31] |
| female | 50-60 vs 60+ | +76.3% | <0.001 | 53.70 [29.53, 77.87] |
| *Mean difference = Group 1 − Group 2 in component units (mmol/24h, L/24h, or pH); % shift = difference relative to Group 2.* | | | | |

chloride_ma_3rd_analysis$wald_tests$sex_within_age_bands

Table 1: **Sex Comparisons Within Age Bands**

| Age Band | % Shift | P (Wald Test) | Mean Diff (95% CI) |
| --- | --- | --- | --- |
| 18+ | +5.4% | 0.916 | 6.76 [-119.41, 132.93] |
| 20-30 | +23.2% | 0.259 | 28.80 [-21.19, 78.79] |
| 30-40 | +13.0% | 0.221 | 17.20 [-10.37, 44.77] |
| 40-50 | +13.8% | 0.243 | 18.40 [-12.52, 49.32] |
| 50-60 | +30.8% | <0.05 | 38.20 [4.59, 71.81] |
| 60+ | +102.6% | <0.001 | 72.20 [48.97, 95.43] |
| *Mean difference = Group 1 − Group 2 in component units (mmol/24h, L/24h, or pH); % shift = difference relative to Group 2.* | | | |

#### 6.14 Sulphate

##### 6.14.1 Comparison by Age Band

###### 6.14.1.1 Plot

```
sulphate_ma_2nd_analysis <- sex_age_subdivision_ma("sulphate")
## Not enough studies for multivariable meta-regression (k = 1, need > 6). Running univariable models instead.
## Not enough studies for multivariable meta-regression (k = 4, need > 6). Running univariable models instead.
sulphate_ma_2nd_analysis$combined_plot
## `height` was translated to `width`.
## `height` was translated to `width`.
## `height` was translated to `width`.
```

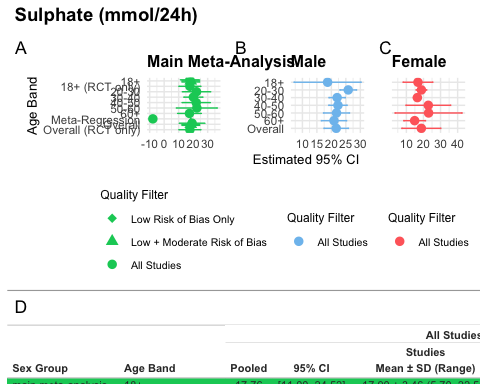

###### 6.14.1.2 Summary Table

sulphate_ma_2nd_analysis$combined_summary_table

Table 1: **Combined Meta-Analysis Results**

24-Hour Urine Sulphate (mmol/24h)

|  |  | **All Studies** | | | | | | **Low Risk of Bias Only** | | | | | | **Low + Moderate RoB** | | | | | |
| --- | --- | --- | --- | --- | --- | --- | --- | --- | --- | --- | --- | --- | --- | --- | --- | --- | --- | --- | --- |
| Sex Group | Age Band | Pooled | 95% CI | Studies  Mean ± SD (Range) | Studies, n | Q p-value | Egger's p | Pooled | 95% CI | Studies  Mean ± SD (Range) | Studies, n | Q p-value | Egger's p | Pooled | 95% CI | Studies  Mean ± SD (Range) | Studies, n | Q p-value | Egger's p |
| main meta-analysis | 18+ | 17.76 | [11.00, 24.52] | 17.90 ± 3.46 (5.70–22.50) | 19 | <0.001 | <0.001 | 18.36 | [11.98, 24.74] | 18.46 ± 0.69 (17.30–19.00) | 5 | 0.54 | 0.46 | 18.36 | [11.98, 24.74] | 18.46 ± 0.69 (17.30–19.00) | 5 | 0.54 | 0.46 |
| **main meta-analysis** | **18+ (RCT only)** | **17.76** | **[10.15, 25.36]** | **18.38 ± 1.08 (17.30–19.45)** | **3** | **0.45** | **0.38** | **—** | **—** | **—** | **—** | **—** | **—** | **—** | **—** | **—** | **—** | **—** | **—** |
| main meta-analysis | 20-30 | 22.31 | [12.57, 32.05] | 22.31 ± 4.98 (18.79–25.83) | 2 | <0.001 | — | — | — | — | — | — | — | — | — | — | — | — | — |
| main meta-analysis | 30-40 | 20.13 | [13.55, 26.71] | 20.13 ± 3.36 (16.43–23.00) | 3 | <0.001 | 0.32 | — | — | — | — | — | — | — | — | — | — | — | — |
| main meta-analysis | 40-50 | 22.10 | [12.37, 31.83] | 21.98 ± 5.00 (17.93–30.45) | 5 | <0.001 | 0.96 | — | — | — | — | — | — | — | — | — | — | — | — |
| main meta-analysis | 50-60 | 22.56 | [8.44, 36.67] | 22.56 ± 7.20 (17.18–34.53) | 5 | <0.001 | 0.96 | — | — | — | — | — | — | — | — | — | — | — | — |
| main meta-analysis | 60+ | 17.46 | [9.31, 25.61] | 17.47 ± 4.17 (10.59–23.27) | 7 | <0.001 | 0.12 | — | — | — | — | — | — | — | — | — | — | — | — |
| **main meta-analysis** | **Meta-Regression** | **-7.80** | **[-9.49, -6.10]** | **—** | **14** | **0.06** | **0.14** | **—** | **—** | **—** | **—** | **—** | **—** | **—** | **—** | **—** | **—** | **—** | **—** |
| **main meta-analysis** | **Overall** | **19.26** | **[10.19, 28.33]** | **19.27 ± 4.59 (5.70–34.53)** | **41** | **<0.001** | **0.41** | **18.36** | **[11.98, 24.74]** | **18.46 ± 0.69 (17.30–19.00)** | **5** | **0.54** | **0.46** | **18.36** | **[11.98, 24.74]** | **18.46 ± 0.69 (17.30–19.00)** | **5** | **0.54** | **0.46** |
| **main meta-analysis** | **Overall (RCT only)** | **17.76** | **[10.15, 25.36]** | **18.38 ± 1.08 (17.30–19.45)** | **3** | **0.45** | **0.38** | **—** | **—** | **—** | **—** | **—** | **—** | **—** | **—** | **—** | **—** | **—** | **—** |
| male | 18+ | 18.70 | [7.14, 30.26] | — (18.70–18.70) | 1 | — | — | — | — | — | — | — | — | — | — | — | — | — | — |
| **male** | **18+ (RCT only)** | **—** | **—** | **—** | **—** | **—** | **—** | **—** | **—** | **—** | **—** | **—** | **—** | **—** | **—** | **—** | **—** | **—** | **—** |
| male | 20-30 | 25.83 | [22.97, 28.69] | — (25.83–25.83) | 1 | — | — | — | — | — | — | — | — | — | — | — | — | — | — |
| male | 30-40 | 21.99 | [19.18, 24.80] | 21.98 ± 1.44 (20.96–23.00) | 2 | <0.001 | — | — | — | — | — | — | — | — | — | — | — | — | — |
| male | 40-50 | 22.28 | [19.12, 25.44] | 20.75 ± 2.23 (19.17–22.33) | 2 | 0.17 | — | — | — | — | — | — | — | — | — | — | — | — | — |
| male | 50-60 | 21.78 | [17.31, 26.26] | 21.91 ± 2.40 (20.22–23.61) | 2 | <0.005 | — | — | — | — | — | — | — | — | — | — | — | — | — |
| male | 60+ | 20.90 | [16.56, 25.25] | 20.94 ± 2.24 (18.81–23.27) | 3 | <0.001 | 0.38 | — | — | — | — | — | — | — | — | — | — | — | — |
| **male** | **Meta-Regression** | **—** | **—** | **—** | **—** | **—** | **—** | **—** | **—** | **—** | **—** | **—** | **—** | **—** | **—** | **—** | **—** | **—** | **—** |
| **male** | **Overall** | **21.67** | **[17.45, 25.89]** | **21.51 ± 2.29 (18.70–25.83)** | **11** | **<0.001** | **0.33** | **—** | **—** | **—** | **—** | **—** | **—** | **—** | **—** | **—** | **—** | **—** | **—** |
| **male** | **Overall (RCT only)** | **—** | **—** | **—** | **—** | **—** | **—** | **—** | **—** | **—** | **—** | **—** | **—** | **—** | **—** | **—** | **—** | **—** | **—** |
| female | 18+ | 16.78 | [8.12, 25.44] | 17.00 ± 1.15 (16.00–18.00) | 4 | 0.05 | <0.001 | — | — | — | — | — | — | — | — | — | — | — | — |
| **female** | **18+ (RCT only)** | **—** | **—** | **—** | **—** | **—** | **—** | **—** | **—** | **—** | **—** | **—** | **—** | **—** | **—** | **—** | **—** | **—** | **—** |
| female | 20-30 | 18.79 | [15.69, 21.89] | — (18.79–18.79) | 1 | — | — | — | — | — | — | — | — | — | — | — | — | — | — |
| female | 30-40 | 16.43 | [14.16, 18.70] | — (16.43–16.43) | 1 | — | — | — | — | — | — | — | — | — | — | — | — | — | — |
| female | 40-50 | 22.83 | [9.76, 35.89] | 22.80 ± 6.71 (17.93–30.45) | 3 | <0.001 | 0.92 | — | — | — | — | — | — | — | — | — | — | — | — |
| female | 50-60 | 22.99 | [3.39, 42.58] | 22.98 ± 10.00 (17.18–34.53) | 3 | <0.001 | 0.97 | — | — | — | — | — | — | — | — | — | — | — | — |
| female | 60+ | 14.84 | [8.51, 21.17] | 14.87 ± 3.23 (10.59–18.24) | 4 | <0.001 | 0.10 | — | — | — | — | — | — | — | — | — | — | — | — |
| **female** | **Meta-Regression** | **—** | **—** | **—** | **—** | **—** | **—** | **—** | **—** | **—** | **—** | **—** | **—** | **—** | **—** | **—** | **—** | **—** | **—** |
| **female** | **Overall** | **18.76** | **[7.37, 30.14]** | **18.75 ± 5.81 (10.59–34.53)** | **16** | **<0.001** | **0.24** | **—** | **—** | **—** | **—** | **—** | **—** | **—** | **—** | **—** | **—** | **—** | **—** |
| **female** | **Overall (RCT only)** | **—** | **—** | **—** | **—** | **—** | **—** | **—** | **—** | **—** | **—** | **—** | **—** | **—** | **—** | **—** | **—** | **—** | **—** |
| *Studies, n = number of studies; Q p-value = Cochran's Q heterogeneity test; Egger's p = linear regression test for funnel plot asymmetry; RoB = Risk of Bias* | | | | | | | | | | | | | | | | | | | |

###### 6.14.1.3 Wald Tests

sulphate_ma_2nd_analysis$wald_tests$sex_comparison

Table 1: **Sex Comparison (Overall)**

| Comparison | % Shift | P (Wald Test) | Mean Diff (95% CI) |
| --- | --- | --- | --- |
| Male vs Female (Overall) | +15.5% | 0.638 | 2.91 [-9.22, 15.05] |
| *Mean difference = Group 1 − Group 2 in component units (mmol/24h, L/24h, or pH); % shift = difference relative to Group 2.* | | | |

sulphate_ma_2nd_analysis$wald_tests$age_band_comparisons

Table 1: **Age Band Comparisons Within Sex Groups**

| Sex Group | Comparison | % Shift | P (Wald Test) | Mean Diff (95% CI) |
| --- | --- | --- | --- | --- |
| main meta-analysis | 18+ vs 20-30 | -20.4% | 0.452 | -4.55 [-16.40, 7.30] |
| main meta-analysis | 18+ vs 30-40 | -11.8% | 0.622 | -2.37 [-11.81, 7.06] |
| main meta-analysis | 18+ vs 40-50 | -19.6% | 0.473 | -4.34 [-16.19, 7.50] |
| main meta-analysis | 18+ vs 50-60 | -21.3% | 0.548 | -4.80 [-20.45, 10.85] |
| main meta-analysis | 18+ vs 60+ | +1.7% | 0.955 | 0.30 [-10.28, 10.89] |
| main meta-analysis | 20-30 vs 30-40 | +10.8% | 0.717 | 2.18 [-9.58, 13.93] |
| main meta-analysis | 20-30 vs 40-50 | +1.0% | 0.976 | 0.21 [-13.56, 13.98] |
| main meta-analysis | 20-30 vs 50-60 | -1.1% | 0.978 | -0.25 [-17.39, 16.90] |
| main meta-analysis | 20-30 vs 60+ | +27.8% | 0.454 | 4.86 [-7.84, 17.55] |
| main meta-analysis | 30-40 vs 40-50 | -8.9% | 0.743 | -1.97 [-13.71, 9.78] |
| main meta-analysis | 30-40 vs 50-60 | -10.7% | 0.760 | -2.42 [-18.00, 13.15] |
| main meta-analysis | 30-40 vs 60+ | +15.3% | 0.616 | 2.68 [-7.80, 13.15] |
| main meta-analysis | 40-50 vs 50-60 | -2.0% | 0.958 | -0.46 [-17.60, 16.69] |
| main meta-analysis | 40-50 vs 60+ | +26.6% | 0.473 | 4.65 [-8.05, 17.34] |
| main meta-analysis | 50-60 vs 60+ | +29.2% | 0.540 | 5.10 [-11.20, 21.40] |
| male | 18+ vs 20-30 | -27.6% | 0.241 | -7.13 [-19.04, 4.78] |
| male | 18+ vs 30-40 | -15.0% | 0.588 | -3.29 [-15.19, 8.61] |
| male | 18+ vs 40-50 | -16.1% | 0.558 | -3.58 [-15.57, 8.41] |
| male | 18+ vs 50-60 | -14.2% | 0.626 | -3.08 [-15.48, 9.32] |
| male | 18+ vs 60+ | -10.5% | 0.727 | -2.20 [-14.56, 10.15] |
| male | 20-30 vs 30-40 | +17.5% | 0.061 | 3.84 [-0.17, 7.85] |
| male | 20-30 vs 40-50 | +15.9% | 0.103 | 3.55 [-0.71, 7.81] |
| male | 20-30 vs 50-60 | +18.6% | 0.135 | 4.05 [-1.26, 9.36] |
| male | 20-30 vs 60+ | +23.6% | 0.063 | 4.93 [-0.28, 10.13] |
| male | 30-40 vs 40-50 | -1.3% | 0.894 | -0.29 [-4.52, 3.94] |
| male | 30-40 vs 50-60 | +1.0% | 0.938 | 0.21 [-5.07, 5.49] |
| male | 30-40 vs 60+ | +5.2% | 0.680 | 1.09 [-4.08, 6.26] |
| male | 40-50 vs 50-60 | +2.3% | 0.859 | 0.50 [-4.98, 5.98] |
| male | 40-50 vs 60+ | +6.6% | 0.616 | 1.38 [-4.00, 6.75] |
| male | 50-60 vs 60+ | +4.2% | 0.782 | 0.88 [-5.36, 7.11] |
| female | 18+ vs 20-30 | -10.7% | 0.668 | -2.01 [-11.20, 7.19] |
| female | 18+ vs 30-40 | +2.1% | 0.939 | 0.35 [-8.60, 9.30] |
| female | 18+ vs 40-50 | -26.5% | 0.450 | -6.04 [-21.72, 9.63] |
| female | 18+ vs 50-60 | -27.0% | 0.570 | -6.21 [-27.63, 15.21] |
| female | 18+ vs 60+ | +13.1% | 0.723 | 1.94 [-8.78, 12.67] |
| female | 20-30 vs 30-40 | +14.4% | 0.229 | 2.36 [-1.48, 6.20] |
| female | 20-30 vs 40-50 | -17.7% | 0.556 | -4.04 [-17.46, 9.39] |
| female | 20-30 vs 50-60 | -18.3% | 0.678 | -4.20 [-24.03, 15.64] |
| female | 20-30 vs 60+ | +26.6% | 0.272 | 3.95 [-3.09, 11.00] |
| female | 30-40 vs 40-50 | -28.0% | 0.345 | -6.40 [-19.66, 6.86] |
| female | 30-40 vs 50-60 | -28.5% | 0.515 | -6.56 [-26.28, 13.17] |
| female | 30-40 vs 60+ | +10.7% | 0.643 | 1.59 [-5.13, 8.32] |
| female | 40-50 vs 50-60 | -0.7% | 0.989 | -0.16 [-23.71, 23.39] |
| female | 40-50 vs 60+ | +53.8% | 0.281 | 7.99 [-6.53, 22.50] |
| female | 50-60 vs 60+ | +54.9% | 0.438 | 8.15 [-12.44, 28.74] |
| *Mean difference = Group 1 − Group 2 in component units (mmol/24h, L/24h, or pH); % shift = difference relative to Group 2.* | | | | |

sulphate_ma_2nd_analysis$wald_tests$sex_within_age_bands

Table 1: **Sex Comparisons Within Age Bands**

| Age Band | % Shift | P (Wald Test) | Mean Diff (95% CI) |
| --- | --- | --- | --- |
| 18+ | +11.4% | 0.795 | 1.92 [-12.53, 16.37] |
| 20-30 | +37.5% | <0.01 | 7.04 [2.82, 11.26] |
| 30-40 | +33.8% | <0.01 | 5.56 [1.95, 9.17] |
| 40-50 | -2.4% | 0.937 | -0.55 [-13.99, 12.90] |
| 50-60 | -5.2% | 0.906 | -1.20 [-21.30, 18.89] |
| 60+ | +40.9% | 0.121 | 6.07 [-1.61, 13.74] |
| *Mean difference = Group 1 − Group 2 in component units (mmol/24h, L/24h, or pH); % shift = difference relative to Group 2.* | | | |

###### 6.14.1.4 Meta-Regression (plain language)

format_meta_reg_plain("sulphate", sex_group = "all")

Message

No meta-regression result for sulphate / all

###### 6.14.1.5 Assay Subgroup Analysis

assay_subgroup_table("sulphate")

Table 1: **Assay subgroup analysis — Sulphate**

*Per-assay pooled estimates; categories with k < 3 listed but not pooled.*

| Assay category | Studies, n | Participants | 95% CI | I² | Q p-value | Egger's p | Note |
| --- | --- | --- | --- | --- | --- | --- | --- |
| Unreported | 32 | 4,402 | 19.50 [17.92, 21.09] | 100% | <0.001 | 0.243 |  |
| Ion chromatography | 6 | 242 | 17.59 [12.53, 22.65] | 99% | <0.001 | <0.01 |  |
| Autoanalyser | 1 | 12 | — | — | — | — | k = 1 < min_k = 3 (skipped) |
| HPLC | 1 | 107 | — | — | — | — | k = 1 < min_k = 3 (skipped) |
| Turbidometric | 1 | 94 | — | — | — | — | k = 1 < min_k = 3 (skipped) |

###### 6.14.1.6 Preservative Sensitivity (Litholink / acidified samples)

```
preservative_sensitivity_ma("sulphate")
## # A tibble: 3 × 6
##   group               n_studies n_participants pooled_estimate lower_95 upper_95
##   <chr>                   <int>          <int>           <dbl>    <dbl>    <dbl>
## 1 All studies                41           4857            19.3     17.8     20.7
## 2 Excluding preserva…        41           4857            19.3     17.8     20.7
## 3 Preservative only           0              0            NA       NA       NA
```

###### 6.14.1.7 Creatinine Completeness Sensitivity

creat_completeness_table("sulphate")

Table 1: **Creatinine-completeness sensitivity — Sulphate**

*Compares the all-studies pooled estimate against subsets defined by whether the study reported 24h urine creatinine. Studies that do not report creatinine cannot verify collection completeness; a substantial shift here flags possible bias from incomplete collections.*

| Subset | Studies, n | Participants | Range (95% CI) | Δ vs all-studies (%) | Flag |
| --- | --- | --- | --- | --- | --- |
| All studies | 41 | 4,857 | 19.26 [17.82, 20.71] | — | — |
| Creatinine reported only | 34 | 4,470 | 19.11 [17.42, 20.79] | -0.8% | OK |
| Creatinine NOT reported | 7 | 387 | 20.02 [18.03, 22.01] | +3.9% | OK |

##### 6.14.2 Comparison by Age / Sex / race

###### 6.14.2.1 Plot

```
sulphate_ma_3rd_analysis <- sex_age_race_subdivision_ma("sulphate",
                                                      urine_ma2,
                                                      overall_bias)
## Not enough studies for multivariable meta-regression (k = 1, need > 6). Running univariable models instead.
## Not enough studies for multivariable meta-regression (k = 4, need > 6). Running univariable models instead.
sulphate_ma_3rd_analysis$race_plot
## `height` was translated to `width`.
## `height` was translated to `width`.
## `height` was translated to `width`.
```

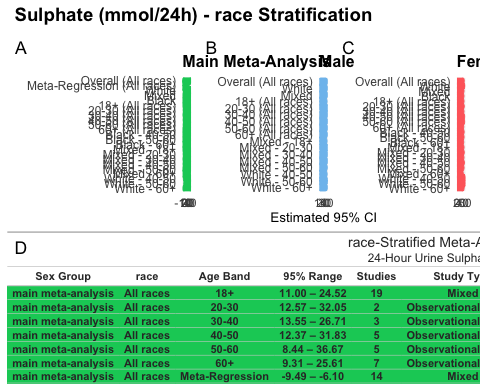

###### 6.14.2.2 Combined MA Results

sulphate_ma_3rd_analysis$combined_summary_table

Table 1: **Combined Meta-Analysis Results**

24-Hour Urine Sulphate (mmol/24h)

| Sex Group | race | Age Band | Pooled | 95% Range | Studies Mean SD Range | Studies | Study Type | Participants | I² | Cochran's Q | Q (p-value) | Egger's p | Model |
| --- | --- | --- | --- | --- | --- | --- | --- | --- | --- | --- | --- | --- | --- |
| main meta-analysis | All | 18+ | 17.76 | 11.00 – 24.52 | 17.90 ± 3.46 (5.70–22.50) | 19 | Mixed | 598 | 98.8% | 1520.8 | <0.005 | <0.001 | Random |
| main meta-analysis | All | 20-30 | 22.31 | 12.57 – 32.05 | 22.31 ± 4.98 (18.79–25.83) | 2 | Observational Cohort | 50 | 99.6% | 267.7 | <0.005 | — | Random |
| main meta-analysis | All | 30-40 | 20.13 | 13.55 – 26.71 | 20.13 ± 3.36 (16.43–23.00) | 3 | Observational Cohort | 157 | 99.7% | 781.8 | <0.005 | 0.32 | Random |
| main meta-analysis | All | 40-50 | 22.10 | 12.37 – 31.83 | 21.98 ± 5.00 (17.93–30.45) | 5 | Observational Cohort | 609 | 98.2% | 217 | <0.005 | 0.96 | Random |
| main meta-analysis | All | 50-60 | 22.56 | 8.44 – 36.67 | 22.56 ± 7.20 (17.18–34.53) | 5 | Observational Cohort | 2669 | 99.9% | 2759.8 | <0.005 | 0.96 | Random |
| main meta-analysis | All | 60+ | 17.46 | 9.31 – 25.61 | 17.47 ± 4.17 (10.59–23.27) | 7 | Observational Cohort | 774 | 99.6% | 1533.5 | <0.005 | 0.12 | Random |
| **main meta-analysis** | **All** | **Meta-Regression** | **-7.80** | **-9.49 – -6.10** | **—** | **14** | **Mixed** | **424** | **48.2%** | **16.5** | **0.06** | **0.14** | **Multivariable** |
| **main meta-analysis** | **All** | **Meta-Regression** | **-7.80** | **-9.49 – -6.10** | **—** | **14** | **Mixed** | **424** | **48.2%** | **16.5** | **0.06** | **0.14** | **Multivariable** |
| **main meta-analysis** | **All** | **Overall** | **19.26** | **10.19 – 28.33** | **19.27 ± 4.59 (5.70–34.53)** | **41** | **Mixed** | **4857** | **99.7%** | **14494.6** | **<0.005** | **0.41** | **Random** |
| **main meta-analysis** | **All** | **Overall** | **19.26** | **10.19 – 28.33** | **19.27 ± 4.59 (5.70–34.53)** | **41** | **Mixed** | **4857** | **99.7%** | **14494.6** | **<0.005** | **0.41** | **Random** |
| main meta-analysis | black | 18+ | — | NA – NA | — | NA | NA | NA | — | — | — | — | NA |
| main meta-analysis | black | 20-30 | — | NA – NA | — | NA | NA | NA | — | — | — | — | NA |
| main meta-analysis | black | 30-40 | — | NA – NA | — | NA | NA | NA | — | — | — | — | NA |
| main meta-analysis | black | 40-50 | 17.93 | 5.88 – 29.98 | — (17.93–17.93) | 1 | Observational Cohort | 29 | — | — | — | — | Single study |
| main meta-analysis | black | 50-60 | 17.24 | 5.09 – 29.39 | — (17.24–17.24) | 1 | Observational Cohort | 131 | — | — | — | — | Single study |
| main meta-analysis | black | 60+ | 14.57 | 3.26 – 25.88 | — (14.57–14.57) | 1 | Observational Cohort | 93 | — | — | — | — | Single study |
| **main meta-analysis** | **black** | **Meta-Regression** | **—** | **NA – NA** | **—** | **NA** | **NA** | **NA** | **—** | **—** | **—** | **—** | **NA** |
| **main meta-analysis** | **black** | **Overall** | **16.47** | **13.30 – 19.63** | **16.58 ± 1.77 (14.57–17.93)** | **3** | **Observational Cohort** | **253** | **85.1%** | **13.4** | **<0.005** | **0.81** | **Random** |
| main meta-analysis | mixed | 18+ | 17.76 | 11.00 – 24.52 | 17.90 ± 3.46 (5.70–22.50) | 19 | Mixed | 598 | 98.8% | 1520.8 | <0.005 | <0.001 | Random |
| main meta-analysis | mixed | 20-30 | 22.31 | 12.57 – 32.05 | 22.31 ± 4.98 (18.79–25.83) | 2 | Observational Cohort | 50 | 99.6% | 267.7 | <0.005 | — | Random |
| main meta-analysis | mixed | 30-40 | 20.13 | 13.55 – 26.71 | 20.13 ± 3.36 (16.43–23.00) | 3 | Observational Cohort | 157 | 99.7% | 781.8 | <0.005 | 0.32 | Random |
| main meta-analysis | mixed | 40-50 | 21.19 | 18.06 – 24.33 | 21.17 ± 1.64 (20.01–22.33) | 2 | Observational Cohort | 50 | 95% | 20.2 | <0.005 | — | Random |
| main meta-analysis | mixed | 50-60 | 18.69 | 14.51 – 22.88 | 18.70 ± 2.15 (17.18–20.22) | 2 | Observational Cohort | 50 | 98.5% | 66.6 | <0.005 | — | Random |
| main meta-analysis | mixed | 60+ | 10.59 | 9.22 – 11.96 | — (10.59–10.59) | 1 | Observational Cohort | 25 | — | — | — | — | Single study |
| **main meta-analysis** | **mixed** | **Meta-Regression** | **—** | **NA – NA** | **—** | **NA** | **NA** | **NA** | **—** | **—** | **—** | **—** | **NA** |
| **main meta-analysis** | **mixed** | **Overall** | **18.42** | **11.04 – 25.79** | **18.46 ± 3.74 (5.70–25.83)** | **29** | **Mixed** | **930** | **99.7%** | **10936.6** | **<0.005** | **0.26** | **Random** |
| main meta-analysis | white | 18+ | — | NA – NA | — | NA | NA | NA | — | — | — | — | NA |
| main meta-analysis | white | 20-30 | — | NA – NA | — | NA | NA | NA | — | — | — | — | NA |
| main meta-analysis | white | 30-40 | — | NA – NA | — | NA | NA | NA | — | — | — | — | NA |
| main meta-analysis | white | 40-50 | 25.03 | 9.75 – 40.31 | 24.81 ± 7.98 (19.17–30.45) | 2 | Observational Cohort | 530 | 95.5% | 22.4 | <0.005 | — | Random |
| main meta-analysis | white | 50-60 | 29.11 | 14.04 – 44.18 | 29.07 ± 7.72 (23.61–34.53) | 2 | Observational Cohort | 2488 | 99.1% | 113 | <0.005 | — | Random |
| main meta-analysis | white | 60+ | 19.43 | 14.27 – 24.59 | 19.42 ± 2.72 (16.07–23.27) | 5 | Observational Cohort | 656 | 95.4% | 87.5 | <0.005 | 0.74 | Random |
| **main meta-analysis** | **white** | **Meta-Regression** | **—** | **NA – NA** | **—** | **NA** | **NA** | **NA** | **—** | **—** | **—** | **—** | **NA** |
| **main meta-analysis** | **white** | **Overall** | **22.82** | **10.88 – 34.76** | **22.76 ± 6.09 (16.07–34.53)** | **9** | **Observational Cohort** | **3674** | **99.7%** | **2517.7** | **<0.005** | **0.77** | **Random** |
| male | All | 18+ | 18.70 | 7.14 – 30.26 | — (18.70–18.70) | 1 | Observational Cohort | 21 | — | — | — | — | Single study |
| male | All | 20-30 | 25.83 | 22.97 – 28.69 | — (25.83–25.83) | 1 | Observational Cohort | 25 | — | — | — | — | Single study |
| male | All | 30-40 | 21.99 | 19.18 – 24.80 | 21.98 ± 1.44 (20.96–23.00) | 2 | Observational Cohort | 132 | 98.7% | 76 | <0.005 | — | Random |
| male | All | 40-50 | 22.28 | 19.12 – 25.44 | 20.75 ± 2.23 (19.17–22.33) | 2 | Observational Cohort | 31 | 46.1% | 1.9 | 0.17 | — | Fixed |
| male | All | 50-60 | 21.78 | 17.31 – 26.26 | 21.91 ± 2.40 (20.22–23.61) | 2 | Observational Cohort | 104 | 90.7% | 10.7 | <0.005 | — | Random |
| male | All | 60+ | 20.90 | 16.56 – 25.25 | 20.94 ± 2.24 (18.81–23.27) | 3 | Observational Cohort | 329 | 96.7% | 61.4 | <0.005 | 0.38 | Random |
| **male** | **All** | **Meta-Regression** | **—** | **NA – NA** | **—** | **NA** | **NA** | **NA** | **—** | **—** | **—** | **—** | **NA** |
| **male** | **All** | **Overall** | **21.67** | **17.45 – 25.89** | **21.51 ± 2.29 (18.70–25.83)** | **11** | **Observational Cohort** | **642** | **98.3%** | **588.1** | **<0.005** | **0.33** | **Random** |
| male | black | 18+ | — | NA – NA | — | NA | NA | NA | — | — | — | — | NA |
| male | black | 20-30 | — | NA – NA | — | NA | NA | NA | — | — | — | — | NA |
| male | black | 30-40 | — | NA – NA | — | NA | NA | NA | — | — | — | — | NA |
| male | black | 40-50 | — | NA – NA | — | NA | NA | NA | — | — | — | — | NA |
| male | black | 50-60 | — | NA – NA | — | NA | NA | NA | — | — | — | — | NA |
| male | black | 60+ | — | NA – NA | — | NA | NA | NA | — | — | — | — | NA |
| **male** | **black** | **Meta-Regression** | **—** | **NA – NA** | **—** | **NA** | **NA** | **NA** | **—** | **—** | **—** | **—** | **NA** |
| **male** | **black** | **Overall** | **—** | **NA – NA** | **—** | **NA** | **NA** | **NA** | **—** | **—** | **—** | **—** | **NA** |
| male | mixed | 18+ | 18.70 | 7.14 – 30.26 | — (18.70–18.70) | 1 | Observational Cohort | 21 | — | — | — | — | Single study |
| male | mixed | 20-30 | 25.83 | 22.97 – 28.69 | — (25.83–25.83) | 1 | Observational Cohort | 25 | — | — | — | — | Single study |
| male | mixed | 30-40 | 21.99 | 19.18 – 24.80 | 21.98 ± 1.44 (20.96–23.00) | 2 | Observational Cohort | 132 | 98.7% | 76 | <0.005 | — | Random |
| male | mixed | 40-50 | 22.33 | 19.47 – 25.19 | — (22.33–22.33) | 1 | Observational Cohort | 25 | — | — | — | — | Single study |
| male | mixed | 50-60 | 20.22 | 17.32 – 23.12 | — (20.22–20.22) | 1 | Observational Cohort | 25 | — | — | — | — | Single study |
| male | mixed | 60+ | — | NA – NA | — | NA | NA | NA | — | — | — | — | NA |
| **male** | **mixed** | **Meta-Regression** | **—** | **NA – NA** | **—** | **NA** | **NA** | **NA** | **—** | **—** | **—** | **—** | **NA** |
| **male** | **mixed** | **Overall** | **21.96** | **17.39 – 26.54** | **21.84 ± 2.48 (18.70–25.83)** | **6** | **Observational Cohort** | **228** | **98.2%** | **272.5** | **<0.005** | **0.52** | **Random** |
| male | white | 18+ | — | NA – NA | — | NA | NA | NA | — | — | — | — | NA |
| male | white | 20-30 | — | NA – NA | — | NA | NA | NA | — | — | — | — | NA |
| male | white | 30-40 | — | NA – NA | — | NA | NA | NA | — | — | — | — | NA |
| male | white | 40-50 | 19.17 | 8.12 – 30.22 | — (19.17–19.17) | 1 | Observational Cohort | 6 | — | — | — | — | Single study |
| male | white | 50-60 | 23.61 | 6.32 – 40.90 | — (23.61–23.61) | 1 | Observational Cohort | 79 | — | — | — | — | Single study |
| male | white | 60+ | 20.90 | 16.56 – 25.25 | 20.94 ± 2.24 (18.81–23.27) | 3 | Observational Cohort | 329 | 96.7% | 61.4 | <0.005 | 0.38 | Random |
| **male** | **white** | **Meta-Regression** | **—** | **NA – NA** | **—** | **NA** | **NA** | **NA** | **—** | **—** | **—** | **—** | **NA** |
| **male** | **white** | **Overall** | **21.25** | **17.14 – 25.36** | **21.12 ± 2.24 (18.81–23.61)** | **5** | **Observational Cohort** | **414** | **94.8%** | **77.5** | **<0.005** | **0.24** | **Random** |
| female | All | 18+ | 16.78 | 8.12 – 25.44 | 17.00 ± 1.15 (16.00–18.00) | 4 | Observational Cohort | 156 | 60.6% | 7.6 | 0.05 | <0.001 | Fixed |
| female | All | 20-30 | 18.79 | 15.69 – 21.89 | — (18.79–18.79) | 1 | Observational Cohort | 25 | — | — | — | — | Single study |
| female | All | 30-40 | 16.43 | 14.16 – 18.70 | — (16.43–16.43) | 1 | Observational Cohort | 25 | — | — | — | — | Single study |
| female | All | 40-50 | 22.83 | 9.76 – 35.89 | 22.80 ± 6.71 (17.93–30.45) | 3 | Observational Cohort | 578 | 99.1% | 213.3 | <0.005 | 0.92 | Random |
| female | All | 50-60 | 22.99 | 3.39 – 42.58 | 22.98 ± 10.00 (17.18–34.53) | 3 | Observational Cohort | 2565 | 99.9% | 2645.8 | <0.005 | 0.97 | Random |
| female | All | 60+ | 14.84 | 8.51 – 21.17 | 14.87 ± 3.23 (10.59–18.24) | 4 | Observational Cohort | 445 | 99% | 292.3 | <0.005 | 0.10 | Random |
| **female** | **All** | **Meta-Regression** | **—** | **NA – NA** | **—** | **NA** | **NA** | **NA** | **—** | **—** | **—** | **—** | **NA** |
| **female** | **All** | **Overall** | **18.76** | **7.37 – 30.14** | **18.75 ± 5.81 (10.59–34.53)** | **16** | **Observational Cohort** | **3794** | **99.8%** | **7067** | **<0.005** | **0.24** | **Random** |
| female | black | 18+ | — | NA – NA | — | NA | NA | NA | — | — | — | — | NA |
| female | black | 20-30 | — | NA – NA | — | NA | NA | NA | — | — | — | — | NA |
| female | black | 30-40 | — | NA – NA | — | NA | NA | NA | — | — | — | — | NA |
| female | black | 40-50 | 17.93 | 5.88 – 29.98 | — (17.93–17.93) | 1 | Observational Cohort | 29 | — | — | — | — | Single study |
| female | black | 50-60 | 17.24 | 5.09 – 29.39 | — (17.24–17.24) | 1 | Observational Cohort | 131 | — | — | — | — | Single study |
| female | black | 60+ | 14.57 | 3.26 – 25.88 | — (14.57–14.57) | 1 | Observational Cohort | 93 | — | — | — | — | Single study |
| **female** | **black** | **Meta-Regression** | **—** | **NA – NA** | **—** | **NA** | **NA** | **NA** | **—** | **—** | **—** | **—** | **NA** |
| **female** | **black** | **Overall** | **16.47** | **13.30 – 19.63** | **16.58 ± 1.77 (14.57–17.93)** | **3** | **Observational Cohort** | **253** | **85.1%** | **13.4** | **<0.005** | **0.81** | **Random** |
| female | mixed | 18+ | 16.78 | 8.12 – 25.44 | 17.00 ± 1.15 (16.00–18.00) | 4 | Observational Cohort | 156 | 60.6% | 7.6 | 0.05 | <0.001 | Fixed |
| female | mixed | 20-30 | 18.79 | 15.69 – 21.89 | — (18.79–18.79) | 1 | Observational Cohort | 25 | — | — | — | — | Single study |
| female | mixed | 30-40 | 16.43 | 14.16 – 18.70 | — (16.43–16.43) | 1 | Observational Cohort | 25 | — | — | — | — | Single study |
| female | mixed | 40-50 | 20.01 | 15.84 – 24.18 | — (20.01–20.01) | 1 | Observational Cohort | 25 | — | — | — | — | Single study |
| female | mixed | 50-60 | 17.18 | 14.97 – 19.39 | — (17.18–17.18) | 1 | Observational Cohort | 25 | — | — | — | — | Single study |
| female | mixed | 60+ | 10.59 | 9.22 – 11.96 | — (10.59–10.59) | 1 | Observational Cohort | 25 | — | — | — | — | Single study |
| **female** | **mixed** | **Meta-Regression** | **—** | **NA – NA** | **—** | **NA** | **NA** | **NA** | **—** | **—** | **—** | **—** | **NA** |
| **female** | **mixed** | **Overall** | **16.75** | **11.48 – 22.03** | **16.78 ± 2.68 (10.59–20.01)** | **9** | **Observational Cohort** | **281** | **99.4%** | **1387.2** | **<0.005** | **0.07** | **Random** |
| female | white | 18+ | — | NA – NA | — | NA | NA | NA | — | — | — | — | NA |
| female | white | 20-30 | — | NA – NA | — | NA | NA | NA | — | — | — | — | NA |
| female | white | 30-40 | — | NA – NA | — | NA | NA | NA | — | — | — | — | NA |
| female | white | 40-50 | 30.45 | 2.64 – 58.26 | — (30.45–30.45) | 1 | Observational Cohort | 524 | — | — | — | — | Single study |
| female | white | 50-60 | 34.53 | 9.05 – 60.01 | — (34.53–34.53) | 1 | Observational Cohort | 2409 | — | — | — | — | Single study |
| female | white | 60+ | 17.22 | 14.46 – 19.97 | 17.16 ± 1.53 (16.07–18.24) | 2 | Observational Cohort | 327 | 83.8% | 6.2 | <0.05 | — | Random |
| **female** | **white** | **Meta-Regression** | **—** | **NA – NA** | **—** | **NA** | **NA** | **NA** | **—** | **—** | **—** | **—** | **NA** |
| **female** | **white** | **Overall** | **24.83** | **7.12 – 42.55** | **24.82 ± 9.05 (16.07–34.53)** | **4** | **Observational Cohort** | **3260** | **99.8%** | **1225.1** | **<0.005** | **0.20** | **Random** |
| *I² represents heterogeneity; Q test assesses between-study variation; Egger's p = linear regression test for funnel plot asymmetry; Meta-Regression shows covariate-adjusted estimates* | | | | | | | | | | | | | |

###### 6.14.2.3 Table of Results subdivided by race / Sex / Age

sulphate_ma_3rd_analysis$race_summary_table

Table 1: **race-Stratified Meta-Analysis Results**

24-Hour Urine Sulphate (mmol/24h)

| Sex Group | race | Age Band | 95% Range | Studies | Study Type | Participants | I² | Cochran's Q | Q (p-value) | Egger's p | Model |
| --- | --- | --- | --- | --- | --- | --- | --- | --- | --- | --- | --- |
| **main meta-analysis** | **All races** | **18+** | **11.00 – 24.52** | **19** | **Mixed** | **598** | **98.8%** | **1520.8** | **<0.005** | **<0.001** | **Random** |
| **main meta-analysis** | **All races** | **20-30** | **12.57 – 32.05** | **2** | **Observational Cohort** | **50** | **99.6%** | **267.7** | **<0.005** | **—** | **Random** |
| **main meta-analysis** | **All races** | **30-40** | **13.55 – 26.71** | **3** | **Observational Cohort** | **157** | **99.7%** | **781.8** | **<0.005** | **0.32** | **Random** |
| **main meta-analysis** | **All races** | **40-50** | **12.37 – 31.83** | **5** | **Observational Cohort** | **609** | **98.2%** | **217** | **<0.005** | **0.96** | **Random** |
| **main meta-analysis** | **All races** | **50-60** | **8.44 – 36.67** | **5** | **Observational Cohort** | **2669** | **99.9%** | **2759.8** | **<0.005** | **0.96** | **Random** |
| **main meta-analysis** | **All races** | **60+** | **9.31 – 25.61** | **7** | **Observational Cohort** | **774** | **99.6%** | **1533.5** | **<0.005** | **0.12** | **Random** |
| **main meta-analysis** | **All races** | **Meta-Regression** | **-9.49 – -6.10** | **14** | **Mixed** | **424** | **48.2%** | **16.5** | **0.06** | **0.14** | **Multivariable** |
| **main meta-analysis** | **All races** | **Meta-Regression** | **-9.49 – -6.10** | **14** | **Mixed** | **424** | **48.2%** | **16.5** | **0.06** | **0.14** | **Multivariable** |
| **main meta-analysis** | **All races** | **Overall** | **10.19 – 28.33** | **41** | **Mixed** | **4857** | **99.7%** | **14494.6** | **<0.005** | **0.41** | **Random** |
| **main meta-analysis** | **All races** | **Overall** | **10.19 – 28.33** | **41** | **Mixed** | **4857** | **99.7%** | **14494.6** | **<0.005** | **0.41** | **Random** |
| main meta-analysis | Black | 18+ | — | — | NA | — | — | — | — | — | — |
| main meta-analysis | Black | 20-30 | — | — | NA | — | — | — | — | — | — |
| main meta-analysis | Black | 30-40 | — | — | NA | — | — | — | — | — | — |
| main meta-analysis | Black | 40-50 | 5.88 – 29.98 | 1 | Observational Cohort | 29 | — | — | — | — | Single study |
| main meta-analysis | Black | 50-60 | 5.09 – 29.39 | 1 | Observational Cohort | 131 | — | — | — | — | Single study |
| main meta-analysis | Black | 60+ | 3.26 – 25.88 | 1 | Observational Cohort | 93 | — | — | — | — | Single study |
| main meta-analysis | Black | Meta-Regression | — | — | NA | — | — | — | — | — | — |
| **main meta-analysis** | **Black** | **Overall** | **13.30 – 19.63** | **3** | **Observational Cohort** | **253** | **85.1%** | **13.4** | **<0.005** | **0.81** | **Random** |
| main meta-analysis | Mixed | 18+ | 11.00 – 24.52 | 19 | Mixed | 598 | 98.8% | 1520.8 | <0.005 | <0.001 | Random |
| main meta-analysis | Mixed | 20-30 | 12.57 – 32.05 | 2 | Observational Cohort | 50 | 99.6% | 267.7 | <0.005 | — | Random |
| main meta-analysis | Mixed | 30-40 | 13.55 – 26.71 | 3 | Observational Cohort | 157 | 99.7% | 781.8 | <0.005 | 0.32 | Random |
| main meta-analysis | Mixed | 40-50 | 18.06 – 24.33 | 2 | Observational Cohort | 50 | 95% | 20.2 | <0.005 | — | Random |
| main meta-analysis | Mixed | 50-60 | 14.51 – 22.88 | 2 | Observational Cohort | 50 | 98.5% | 66.6 | <0.005 | — | Random |
| main meta-analysis | Mixed | 60+ | 9.22 – 11.96 | 1 | Observational Cohort | 25 | — | — | — | — | Single study |
| main meta-analysis | Mixed | Meta-Regression | — | — | NA | — | — | — | — | — | — |
| **main meta-analysis** | **Mixed** | **Overall** | **11.04 – 25.79** | **29** | **Mixed** | **930** | **99.7%** | **10936.6** | **<0.005** | **0.26** | **Random** |
| main meta-analysis | White | 18+ | — | — | NA | — | — | — | — | — | — |
| main meta-analysis | White | 20-30 | — | — | NA | — | — | — | — | — | — |
| main meta-analysis | White | 30-40 | — | — | NA | — | — | — | — | — | — |
| main meta-analysis | White | 40-50 | 9.75 – 40.31 | 2 | Observational Cohort | 530 | 95.5% | 22.4 | <0.005 | — | Random |
| main meta-analysis | White | 50-60 | 14.04 – 44.18 | 2 | Observational Cohort | 2488 | 99.1% | 113 | <0.005 | — | Random |
| main meta-analysis | White | 60+ | 14.27 – 24.59 | 5 | Observational Cohort | 656 | 95.4% | 87.5 | <0.005 | 0.74 | Random |
| main meta-analysis | White | Meta-Regression | — | — | NA | — | — | — | — | — | — |
| **main meta-analysis** | **White** | **Overall** | **10.88 – 34.76** | **9** | **Observational Cohort** | **3674** | **99.7%** | **2517.7** | **<0.005** | **0.77** | **Random** |
| **male** | **All races** | **18+** | **7.14 – 30.26** | **1** | **Observational Cohort** | **21** | **—** | **—** | **—** | **—** | **Single study** |
| **male** | **All races** | **20-30** | **22.97 – 28.69** | **1** | **Observational Cohort** | **25** | **—** | **—** | **—** | **—** | **Single study** |
| **male** | **All races** | **30-40** | **19.18 – 24.80** | **2** | **Observational Cohort** | **132** | **98.7%** | **76** | **<0.005** | **—** | **Random** |
| **male** | **All races** | **40-50** | **19.12 – 25.44** | **2** | **Observational Cohort** | **31** | **46.1%** | **1.9** | **0.17** | **—** | **Fixed** |
| **male** | **All races** | **50-60** | **17.31 – 26.26** | **2** | **Observational Cohort** | **104** | **90.7%** | **10.7** | **<0.005** | **—** | **Random** |
| **male** | **All races** | **60+** | **16.56 – 25.25** | **3** | **Observational Cohort** | **329** | **96.7%** | **61.4** | **<0.005** | **0.38** | **Random** |
| **male** | **All races** | **Meta-Regression** | **—** | **—** | **NA** | **—** | **—** | **—** | **—** | **—** | **—** |
| **male** | **All races** | **Overall** | **17.45 – 25.89** | **11** | **Observational Cohort** | **642** | **98.3%** | **588.1** | **<0.005** | **0.33** | **Random** |
| male | Black | 18+ | — | — | NA | — | — | — | — | — | — |
| male | Black | 20-30 | — | — | NA | — | — | — | — | — | — |
| male | Black | 30-40 | — | — | NA | — | — | — | — | — | — |
| male | Black | 40-50 | — | — | NA | — | — | — | — | — | — |
| male | Black | 50-60 | — | — | NA | — | — | — | — | — | — |
| male | Black | 60+ | — | — | NA | — | — | — | — | — | — |
| male | Black | Meta-Regression | — | — | NA | — | — | — | — | — | — |
| **male** | **Black** | **Overall** | **—** | **—** | **NA** | **—** | **—** | **—** | **—** | **—** | **—** |
| male | Mixed | 18+ | 7.14 – 30.26 | 1 | Observational Cohort | 21 | — | — | — | — | Single study |
| male | Mixed | 20-30 | 22.97 – 28.69 | 1 | Observational Cohort | 25 | — | — | — | — | Single study |
| male | Mixed | 30-40 | 19.18 – 24.80 | 2 | Observational Cohort | 132 | 98.7% | 76 | <0.005 | — | Random |
| male | Mixed | 40-50 | 19.47 – 25.19 | 1 | Observational Cohort | 25 | — | — | — | — | Single study |
| male | Mixed | 50-60 | 17.32 – 23.12 | 1 | Observational Cohort | 25 | — | — | — | — | Single study |
| male | Mixed | 60+ | — | — | NA | — | — | — | — | — | — |
| male | Mixed | Meta-Regression | — | — | NA | — | — | — | — | — | — |
| **male** | **Mixed** | **Overall** | **17.39 – 26.54** | **6** | **Observational Cohort** | **228** | **98.2%** | **272.5** | **<0.005** | **0.52** | **Random** |
| male | White | 18+ | — | — | NA | — | — | — | — | — | — |
| male | White | 20-30 | — | — | NA | — | — | — | — | — | — |
| male | White | 30-40 | — | — | NA | — | — | — | — | — | — |
| male | White | 40-50 | 8.12 – 30.22 | 1 | Observational Cohort | 6 | — | — | — | — | Single study |
| male | White | 50-60 | 6.32 – 40.90 | 1 | Observational Cohort | 79 | — | — | — | — | Single study |
| male | White | 60+ | 16.56 – 25.25 | 3 | Observational Cohort | 329 | 96.7% | 61.4 | <0.005 | 0.38 | Random |
| male | White | Meta-Regression | — | — | NA | — | — | — | — | — | — |
| **male** | **White** | **Overall** | **17.14 – 25.36** | **5** | **Observational Cohort** | **414** | **94.8%** | **77.5** | **<0.005** | **0.24** | **Random** |
| **female** | **All races** | **18+** | **8.12 – 25.44** | **4** | **Observational Cohort** | **156** | **60.6%** | **7.6** | **0.05** | **<0.001** | **Fixed** |
| **female** | **All races** | **20-30** | **15.69 – 21.89** | **1** | **Observational Cohort** | **25** | **—** | **—** | **—** | **—** | **Single study** |
| **female** | **All races** | **30-40** | **14.16 – 18.70** | **1** | **Observational Cohort** | **25** | **—** | **—** | **—** | **—** | **Single study** |
| **female** | **All races** | **40-50** | **9.76 – 35.89** | **3** | **Observational Cohort** | **578** | **99.1%** | **213.3** | **<0.005** | **0.92** | **Random** |
| **female** | **All races** | **50-60** | **3.39 – 42.58** | **3** | **Observational Cohort** | **2565** | **99.9%** | **2645.8** | **<0.005** | **0.97** | **Random** |
| **female** | **All races** | **60+** | **8.51 – 21.17** | **4** | **Observational Cohort** | **445** | **99%** | **292.3** | **<0.005** | **0.10** | **Random** |
| **female** | **All races** | **Meta-Regression** | **—** | **—** | **NA** | **—** | **—** | **—** | **—** | **—** | **—** |
| **female** | **All races** | **Overall** | **7.37 – 30.14** | **16** | **Observational Cohort** | **3794** | **99.8%** | **7067** | **<0.005** | **0.24** | **Random** |
| female | Black | 18+ | — | — | NA | — | — | — | — | — | — |
| female | Black | 20-30 | — | — | NA | — | — | — | — | — | — |
| female | Black | 30-40 | — | — | NA | — | — | — | — | — | — |
| female | Black | 40-50 | 5.88 – 29.98 | 1 | Observational Cohort | 29 | — | — | — | — | Single study |
| female | Black | 50-60 | 5.09 – 29.39 | 1 | Observational Cohort | 131 | — | — | — | — | Single study |
| female | Black | 60+ | 3.26 – 25.88 | 1 | Observational Cohort | 93 | — | — | — | — | Single study |
| female | Black | Meta-Regression | — | — | NA | — | — | — | — | — | — |
| **female** | **Black** | **Overall** | **13.30 – 19.63** | **3** | **Observational Cohort** | **253** | **85.1%** | **13.4** | **<0.005** | **0.81** | **Random** |
| female | Mixed | 18+ | 8.12 – 25.44 | 4 | Observational Cohort | 156 | 60.6% | 7.6 | 0.05 | <0.001 | Fixed |
| female | Mixed | 20-30 | 15.69 – 21.89 | 1 | Observational Cohort | 25 | — | — | — | — | Single study |
| female | Mixed | 30-40 | 14.16 – 18.70 | 1 | Observational Cohort | 25 | — | — | — | — | Single study |
| female | Mixed | 40-50 | 15.84 – 24.18 | 1 | Observational Cohort | 25 | — | — | — | — | Single study |
| female | Mixed | 50-60 | 14.97 – 19.39 | 1 | Observational Cohort | 25 | — | — | — | — | Single study |
| female | Mixed | 60+ | 9.22 – 11.96 | 1 | Observational Cohort | 25 | — | — | — | — | Single study |
| female | Mixed | Meta-Regression | — | — | NA | — | — | — | — | — | — |
| **female** | **Mixed** | **Overall** | **11.48 – 22.03** | **9** | **Observational Cohort** | **281** | **99.4%** | **1387.2** | **<0.005** | **0.07** | **Random** |
| female | White | 18+ | — | — | NA | — | — | — | — | — | — |
| female | White | 20-30 | — | — | NA | — | — | — | — | — | — |
| female | White | 30-40 | — | — | NA | — | — | — | — | — | — |
| female | White | 40-50 | 2.64 – 58.26 | 1 | Observational Cohort | 524 | — | — | — | — | Single study |
| female | White | 50-60 | 9.05 – 60.01 | 1 | Observational Cohort | 2409 | — | — | — | — | Single study |
| female | White | 60+ | 14.46 – 19.97 | 2 | Observational Cohort | 327 | 83.8% | 6.2 | <0.05 | — | Random |
| female | White | Meta-Regression | — | — | NA | — | — | — | — | — | — |
| **female** | **White** | **Overall** | **7.12 – 42.55** | **4** | **Observational Cohort** | **3260** | **99.8%** | **1225.1** | **<0.005** | **0.20** | **Random** |
| *I² represents heterogeneity; Q test assesses between-study variation; Egger's p = linear regression test for funnel plot asymmetry; — indicates insufficient data* | | | | | | | | | | | |

###### 6.14.2.4 Wald Tests

sulphate_ma_3rd_analysis$wald_tests$sex_comparison

Table 1: **Sex Comparison (Overall)**

| Comparison | % Shift | P (Wald Test) | Mean Diff (95% CI) |
| --- | --- | --- | --- |
| Male vs Female (Overall) | +15.5% | 0.638 | 2.91 [-9.22, 15.05] |
| *Mean difference = Group 1 − Group 2 in component units (mmol/24h, L/24h, or pH); % shift = difference relative to Group 2.* | | | |

sulphate_ma_3rd_analysis$wald_tests$race_comparisons

Table 1: **race Comparisons Within Sex Groups**

| Sex Group | Comparison | % Shift | P (Wald Test) | Mean Diff (95% CI) |
| --- | --- | --- | --- | --- |
| male | black vs mixed | — | — | — |
| male | black vs white | — | — | — |
| male | mixed vs white | +3.4% | 0.820 | 0.71 [-5.44, 6.86] |
| female | black vs mixed | -1.7% | 0.927 | -0.29 [-6.44, 5.87] |
| female | black vs white | -33.7% | 0.362 | -8.36 [-26.36, 9.63] |
| female | mixed vs white | -32.5% | 0.392 | -8.08 [-26.56, 10.41] |
| *Mean difference = Group 1 − Group 2 in component units (mmol/24h, L/24h, or pH); % shift = difference relative to Group 2.* | | | | |

sulphate_ma_3rd_analysis$wald_tests$age_band_comparisons

Table 1: **Age Band Comparisons Within Sex Groups**

| Sex Group | Comparison | % Shift | P (Wald Test) | Mean Diff (95% CI) |
| --- | --- | --- | --- | --- |
| main meta-analysis | 18+ vs 20-30 | -20.4% | 0.452 | -4.55 [-16.40, 7.30] |
| main meta-analysis | 18+ vs 30-40 | -11.8% | 0.622 | -2.37 [-11.81, 7.06] |
| main meta-analysis | 18+ vs 40-50 | -19.6% | 0.473 | -4.34 [-16.19, 7.50] |
| main meta-analysis | 18+ vs 50-60 | -21.3% | 0.548 | -4.80 [-20.45, 10.85] |
| main meta-analysis | 18+ vs 60+ | +1.7% | 0.955 | 0.30 [-10.28, 10.89] |
| main meta-analysis | 20-30 vs 30-40 | +10.8% | 0.717 | 2.18 [-9.58, 13.93] |
| main meta-analysis | 20-30 vs 40-50 | +1.0% | 0.976 | 0.21 [-13.56, 13.98] |
| main meta-analysis | 20-30 vs 50-60 | -1.1% | 0.978 | -0.25 [-17.39, 16.90] |
| main meta-analysis | 20-30 vs 60+ | +27.8% | 0.454 | 4.86 [-7.84, 17.55] |
| main meta-analysis | 30-40 vs 40-50 | -8.9% | 0.743 | -1.97 [-13.71, 9.78] |
| main meta-analysis | 30-40 vs 50-60 | -10.7% | 0.760 | -2.42 [-18.00, 13.15] |
| main meta-analysis | 30-40 vs 60+ | +15.3% | 0.616 | 2.68 [-7.80, 13.15] |
| main meta-analysis | 40-50 vs 50-60 | -2.0% | 0.958 | -0.46 [-17.60, 16.69] |
| main meta-analysis | 40-50 vs 60+ | +26.6% | 0.473 | 4.65 [-8.05, 17.34] |
| main meta-analysis | 50-60 vs 60+ | +29.2% | 0.540 | 5.10 [-11.20, 21.40] |
| male | 18+ vs 20-30 | -27.6% | 0.241 | -7.13 [-19.04, 4.78] |
| male | 18+ vs 30-40 | -15.0% | 0.588 | -3.29 [-15.19, 8.61] |
| male | 18+ vs 40-50 | -16.1% | 0.558 | -3.58 [-15.57, 8.41] |
| male | 18+ vs 50-60 | -14.2% | 0.626 | -3.08 [-15.48, 9.32] |
| male | 18+ vs 60+ | -10.5% | 0.727 | -2.20 [-14.56, 10.15] |
| male | 20-30 vs 30-40 | +17.5% | 0.061 | 3.84 [-0.17, 7.85] |
| male | 20-30 vs 40-50 | +15.9% | 0.103 | 3.55 [-0.71, 7.81] |
| male | 20-30 vs 50-60 | +18.6% | 0.135 | 4.05 [-1.26, 9.36] |
| male | 20-30 vs 60+ | +23.6% | 0.063 | 4.93 [-0.28, 10.13] |
| male | 30-40 vs 40-50 | -1.3% | 0.894 | -0.29 [-4.52, 3.94] |
| male | 30-40 vs 50-60 | +1.0% | 0.938 | 0.21 [-5.07, 5.49] |
| male | 30-40 vs 60+ | +5.2% | 0.680 | 1.09 [-4.08, 6.26] |
| male | 40-50 vs 50-60 | +2.3% | 0.859 | 0.50 [-4.98, 5.98] |
| male | 40-50 vs 60+ | +6.6% | 0.616 | 1.38 [-4.00, 6.75] |
| male | 50-60 vs 60+ | +4.2% | 0.782 | 0.88 [-5.36, 7.11] |
| female | 18+ vs 20-30 | -10.7% | 0.668 | -2.01 [-11.20, 7.19] |
| female | 18+ vs 30-40 | +2.1% | 0.939 | 0.35 [-8.60, 9.30] |
| female | 18+ vs 40-50 | -26.5% | 0.450 | -6.04 [-21.72, 9.63] |
| female | 18+ vs 50-60 | -27.0% | 0.570 | -6.21 [-27.63, 15.21] |
| female | 18+ vs 60+ | +13.1% | 0.723 | 1.94 [-8.78, 12.67] |
| female | 20-30 vs 30-40 | +14.4% | 0.229 | 2.36 [-1.48, 6.20] |
| female | 20-30 vs 40-50 | -17.7% | 0.556 | -4.04 [-17.46, 9.39] |
| female | 20-30 vs 50-60 | -18.3% | 0.678 | -4.20 [-24.03, 15.64] |
| female | 20-30 vs 60+ | +26.6% | 0.272 | 3.95 [-3.09, 11.00] |
| female | 30-40 vs 40-50 | -28.0% | 0.345 | -6.40 [-19.66, 6.86] |
| female | 30-40 vs 50-60 | -28.5% | 0.515 | -6.56 [-26.28, 13.17] |
| female | 30-40 vs 60+ | +10.7% | 0.643 | 1.59 [-5.13, 8.32] |
| female | 40-50 vs 50-60 | -0.7% | 0.989 | -0.16 [-23.71, 23.39] |
| female | 40-50 vs 60+ | +53.8% | 0.281 | 7.99 [-6.53, 22.50] |
| female | 50-60 vs 60+ | +54.9% | 0.438 | 8.15 [-12.44, 28.74] |
| *Mean difference = Group 1 − Group 2 in component units (mmol/24h, L/24h, or pH); % shift = difference relative to Group 2.* | | | | |

sulphate_ma_3rd_analysis$wald_tests$sex_within_age_bands

Table 1: **Sex Comparisons Within Age Bands**

| Age Band | % Shift | P (Wald Test) | Mean Diff (95% CI) |
| --- | --- | --- | --- |
| 18+ | +11.4% | 0.795 | 1.92 [-12.53, 16.37] |
| 20-30 | +37.5% | <0.01 | 7.04 [2.82, 11.26] |
| 30-40 | +33.8% | <0.01 | 5.56 [1.95, 9.17] |
| 40-50 | -2.4% | 0.937 | -0.55 [-13.99, 12.90] |
| 50-60 | -5.2% | 0.906 | -1.20 [-21.30, 18.89] |
| 60+ | +40.9% | 0.121 | 6.07 [-1.61, 13.74] |
| *Mean difference = Group 1 − Group 2 in component units (mmol/24h, L/24h, or pH); % shift = difference relative to Group 2.* | | | |

#### 6.15 Urea

##### 6.15.1 Comparison by Age Band

###### 6.15.1.1 Plot

```
urea_ma_2nd_analysis <- sex_age_subdivision_ma("urea")
## Not enough studies for multivariable meta-regression (k = 3, need > 6). Running univariable models instead.
## Not enough studies for multivariable meta-regression (k = 4, need > 6). Running univariable models instead.
urea_ma_2nd_analysis$combined_plot
## `height` was translated to `width`.
## `height` was translated to `width`.
## `height` was translated to `width`.
```

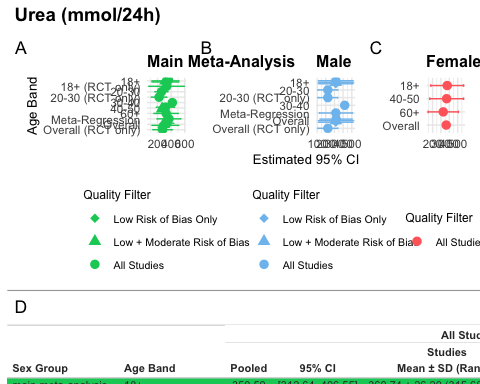

###### 6.15.1.2 Summary Table

urea_ma_2nd_analysis$combined_summary_table

Table 1: **Combined Meta-Analysis Results**

24-Hour Urine Urea (mmol/24h)

|  |  | **All Studies** | | | | | | **Low Risk of Bias Only** | | | | | | **Low + Moderate RoB** | | | | | |
| --- | --- | --- | --- | --- | --- | --- | --- | --- | --- | --- | --- | --- | --- | --- | --- | --- | --- | --- | --- |
| Sex Group | Age Band | Pooled | 95% CI | Studies  Mean ± SD (Range) | Studies, n | Q p-value | Egger's p | Pooled | 95% CI | Studies  Mean ± SD (Range) | Studies, n | Q p-value | Egger's p | Pooled | 95% CI | Studies  Mean ± SD (Range) | Studies, n | Q p-value | Egger's p |
| main meta-analysis | 18+ | 359.59 | [312.64, 406.55] | 360.74 ± 26.29 (315.68–382.95) | 7 | <0.001 | 0.22 | 315.68 | [111.72, 519.65] | — (315.68–315.68) | 1 | — | — | 315.68 | [111.72, 519.65] | — (315.68–315.68) | 1 | — | — |
| **main meta-analysis** | **18+ (RCT only)** | **333.00** | **[71.93, 594.07]** | **— (333.00–333.00)** | **1** | **—** | **—** | **—** | **—** | **—** | **—** | **—** | **—** | **—** | **—** | **—** | **—** | **—** | **—** |
| main meta-analysis | 20-30 | 257.68 | [160.94, 354.41] | 259.53 ± 51.58 (223.06–296.00) | 2 | <0.001 | — | — | — | — | — | — | — | — | — | — | — | — | — |
| **main meta-analysis** | **20-30 (RCT only)** | **223.06** | **[108.95, 337.17]** | **— (223.06–223.06)** | **1** | **—** | **—** | **—** | **—** | **—** | **—** | **—** | **—** | **—** | **—** | **—** | **—** | **—** | **—** |
| main meta-analysis | 30-40 | 418.00 | [388.60, 447.40] | — (418.00–418.00) | 1 | — | — | — | — | — | — | — | — | — | — | — | — | — | — |
| main meta-analysis | 40-50 | 317.60 | [179.12, 456.08] | 317.59 ± 70.69 (267.60–367.57) | 2 | <0.001 | — | — | — | — | — | — | — | 267.60 | [237.42, 297.78] | — (267.60–267.60) | 1 | — | — |
| main meta-analysis | 60+ | 326.77 | [146.26, 507.29] | — (326.77–326.77) | 1 | — | — | — | — | — | — | — | — | — | — | — | — | — | — |
| **main meta-analysis** | **Meta-Regression** | **367.84** | **[192.12, 543.57]** | **—** | **9** | **0.78** | **0.38** | **—** | **—** | **—** | **—** | **—** | **—** | **—** | **—** | **—** | **—** | **—** | **—** |
| **main meta-analysis** | **Overall** | **339.45** | **[233.61, 445.30]** | **340.32 ± 54.03 (223.06–418.00)** | **13** | **<0.001** | **0.28** | **315.68** | **[111.72, 519.65]** | **— (315.68–315.68)** | **1** | **—** | **—** | **290.68** | **[225.59, 355.77]** | **291.64 ± 34.00 (267.60–315.68)** | **2** | **<0.001** | **—** |
| **main meta-analysis** | **Overall (RCT only)** | **274.40** | **[128.32, 420.49]** | **278.03 ± 77.74 (223.06–333.00)** | **2** | **<0.001** | **—** | **—** | **—** | **—** | **—** | **—** | **—** | **—** | **—** | **—** | **—** | **—** | **—** |
| male | 18+ | 315.68 | [111.72, 519.65] | — (315.68–315.68) | 1 | — | — | 315.68 | [111.72, 519.65] | — (315.68–315.68) | 1 | — | — | 315.68 | [111.72, 519.65] | — (315.68–315.68) | 1 | — | — |
| **male** | **18+ (RCT only)** | **—** | **—** | **—** | **—** | **—** | **—** | **—** | **—** | **—** | **—** | **—** | **—** | **—** | **—** | **—** | **—** | **—** | **—** |
| male | 20-30 | 223.06 | [108.95, 337.17] | — (223.06–223.06) | 1 | — | — | — | — | — | — | — | — | — | — | — | — | — | — |
| **male** | **20-30 (RCT only)** | **223.06** | **[108.95, 337.17]** | **— (223.06–223.06)** | **1** | **—** | **—** | **—** | **—** | **—** | **—** | **—** | **—** | **—** | **—** | **—** | **—** | **—** | **—** |
| male | 30-40 | 418.00 | [388.60, 447.40] | — (418.00–418.00) | 1 | — | — | — | — | — | — | — | — | — | — | — | — | — | — |
| male | 40-50 | — | — | — | — | — | — | — | — | — | — | — | — | — | — | — | — | — | — |
| male | 60+ | — | — | — | — | — | — | — | — | — | — | — | — | — | — | — | — | — | — |
| **male** | **Meta-Regression** | **315.68** | **[124.81, 506.55]** | **—** | **3** | **<0.001** | **0.21** | **—** | **—** | **—** | **—** | **—** | **—** | **—** | **—** | **—** | **—** | **—** | **—** |
| **male** | **Overall** | **319.22** | **[128.35, 510.09]** | **318.91 ± 97.51 (223.06–418.00)** | **3** | **<0.001** | **0.21** | **315.68** | **[111.72, 519.65]** | **— (315.68–315.68)** | **1** | **—** | **—** | **315.68** | **[111.72, 519.65]** | **— (315.68–315.68)** | **1** | **—** | **—** |
| **male** | **Overall (RCT only)** | **223.06** | **[108.95, 337.17]** | **— (223.06–223.06)** | **1** | **—** | **—** | **—** | **—** | **—** | **—** | **—** | **—** | **—** | **—** | **—** | **—** | **—** | **—** |
| female | 18+ | 374.63 | [178.82, 570.43] | 374.63 ± 9.61 (366.30–382.95) | 4 | 0.78 | — | — | — | — | — | — | — | — | — | — | — | — | — |
| **female** | **18+ (RCT only)** | **—** | **—** | **—** | **—** | **—** | **—** | **—** | **—** | **—** | **—** | **—** | **—** | **—** | **—** | **—** | **—** | **—** | **—** |
| female | 20-30 | — | — | — | — | — | — | — | — | — | — | — | — | — | — | — | — | — | — |
| **female** | **20-30 (RCT only)** | **—** | **—** | **—** | **—** | **—** | **—** | **—** | **—** | **—** | **—** | **—** | **—** | **—** | **—** | **—** | **—** | **—** | **—** |
| female | 30-40 | — | — | — | — | — | — | — | — | — | — | — | — | — | — | — | — | — | — |
| female | 40-50 | 367.57 | [165.66, 569.48] | — (367.57–367.57) | 1 | — | — | — | — | — | — | — | — | — | — | — | — | — | — |
| female | 60+ | 326.77 | [146.26, 507.29] | — (326.77–326.77) | 1 | — | — | — | — | — | — | — | — | — | — | — | — | — | — |
| **female** | **Meta-Regression** | **—** | **—** | **—** | **—** | **—** | **—** | **—** | **—** | **—** | **—** | **—** | **—** | **—** | **—** | **—** | **—** | **—** | **—** |
| **female** | **Overall** | **362.62** | **[322.03, 403.21]** | **365.47 ± 20.56 (326.77–382.95)** | **6** | **<0.001** | **0.98** | **—** | **—** | **—** | **—** | **—** | **—** | **—** | **—** | **—** | **—** | **—** | **—** |
| **female** | **Overall (RCT only)** | **—** | **—** | **—** | **—** | **—** | **—** | **—** | **—** | **—** | **—** | **—** | **—** | **—** | **—** | **—** | **—** | **—** | **—** |
| *Studies, n = number of studies; Q p-value = Cochran's Q heterogeneity test; Egger's p = linear regression test for funnel plot asymmetry; RoB = Risk of Bias* | | | | | | | | | | | | | | | | | | | |

###### 6.15.1.3 Wald Tests

urea_ma_2nd_analysis$wald_tests$sex_comparison

Table 1: **Sex Comparison (Overall)**

| Comparison | % Shift | P (Wald Test) | Mean Diff (95% CI) |
| --- | --- | --- | --- |
| Male vs Female (Overall) | -12.0% | 0.663 | -43.40 [-238.54, 151.73] |
| *Mean difference = Group 1 − Group 2 in component units (mmol/24h, L/24h, or pH); % shift = difference relative to Group 2.* | | | |

urea_ma_2nd_analysis$wald_tests$age_band_comparisons

Table 1: **Age Band Comparisons Within Sex Groups**

| Sex Group | Comparison | % Shift | P (Wald Test) | Mean Diff (95% CI) |
| --- | --- | --- | --- | --- |
| main meta-analysis | 18+ vs 20-30 | +39.6% | 0.063 | 101.92 [-5.61, 209.44] |
| main meta-analysis | 18+ vs 30-40 | -14.0% | <0.05 | -58.41 [-113.80, -3.01] |
| main meta-analysis | 18+ vs 40-50 | +13.2% | 0.573 | 41.99 [-104.23, 188.21] |
| main meta-analysis | 18+ vs 60+ | +10.0% | 0.730 | 32.82 [-153.70, 219.34] |
| main meta-analysis | 20-30 vs 30-40 | -38.4% | <0.01 | -160.32 [-261.43, -59.22] |
| main meta-analysis | 20-30 vs 40-50 | -18.9% | 0.487 | -59.92 [-228.84, 109.00] |
| main meta-analysis | 20-30 vs 60+ | -21.1% | 0.508 | -69.09 [-273.90, 135.71] |
| main meta-analysis | 30-40 vs 40-50 | +31.6% | 0.165 | 100.40 [-41.16, 241.96] |
| main meta-analysis | 30-40 vs 60+ | +27.9% | 0.328 | 91.23 [-91.67, 274.12] |
| main meta-analysis | 40-50 vs 60+ | -2.8% | 0.937 | -9.17 [-236.68, 218.34] |
| male | 18+ vs 20-30 | +41.5% | 0.437 | 92.62 [-141.09, 326.34] |
| male | 18+ vs 30-40 | -24.5% | 0.330 | -102.32 [-308.39, 103.76] |
| male | 20-30 vs 30-40 | -46.6% | <0.01 | -194.94 [-312.78, -77.10] |
| female | 18+ vs 40-50 | +1.9% | 0.961 | 7.05 [-274.21, 288.31] |
| female | 18+ vs 60+ | +14.6% | 0.725 | 47.85 [-218.47, 314.17] |
| female | 40-50 vs 60+ | +12.5% | 0.768 | 40.80 [-230.04, 311.64] |
| *Mean difference = Group 1 − Group 2 in component units (mmol/24h, L/24h, or pH); % shift = difference relative to Group 2.* | | | | |

urea_ma_2nd_analysis$wald_tests$sex_within_age_bands

Table 1: **Sex Comparisons Within Age Bands**

| Age Band | % Shift | P (Wald Test) | Mean Diff (95% CI) |
| --- | --- | --- | --- |
| 18+ | -15.7% | 0.683 | -58.94 [-341.68, 223.80] |
| *Mean difference = Group 1 − Group 2 in component units (mmol/24h, L/24h, or pH); % shift = difference relative to Group 2.* | | | |

###### 6.15.1.4 Meta-Regression (plain language)

format_meta_reg_plain("urea", sex_group = "all")

Message

No meta-regression result for urea / all

###### 6.15.1.5 Assay Subgroup Analysis

assay_subgroup_table("urea")

Table 1: **Assay subgroup analysis — Urea**

*Per-assay pooled estimates; categories with k < 3 listed but not pooled.*

| Assay category | Studies, n | Participants | 95% CI | I² | Q p-value | Egger's p | Note |
| --- | --- | --- | --- | --- | --- | --- | --- |
| Unreported | 10 | 3,711 | 342.83 [316.76, 368.89] | 99% | <0.001 | 0.931 |  |
| Autoanalyser | 2 | 145 | — | — | — | — | k = 2 < min_k = 3 (skipped) |
| Enzymatic / ultraviolet | 1 | 20 | — | — | — | — | k = 1 < min_k = 3 (skipped) |

###### 6.15.1.6 Preservative Sensitivity (Litholink / acidified samples)

```
preservative_sensitivity_ma("urea")
## # A tibble: 3 × 6
##   group               n_studies n_participants pooled_estimate lower_95 upper_95
##   <chr>                   <int>          <int>           <dbl>    <dbl>    <dbl>
## 1 All studies                13           3876            339.     309.     370.
## 2 Excluding preserva…        13           3876            339.     309.     370.
## 3 Preservative only           0              0             NA       NA       NA
```

###### 6.15.1.7 Creatinine Completeness Sensitivity

creat_completeness_table("urea")

Table 1: **Creatinine-completeness sensitivity — Urea**

*Compares the all-studies pooled estimate against subsets defined by whether the study reported 24h urine creatinine. Studies that do not report creatinine cannot verify collection completeness; a substantial shift here flags possible bias from incomplete collections.*

| Subset | Studies, n | Participants | Range (95% CI) | Δ vs all-studies (%) | Flag |
| --- | --- | --- | --- | --- | --- |
| All studies | 13 | 3,876 | 339.45 [308.88, 370.02] | — | — |
| Creatinine reported only | 9 | 365 | 332.84 [294.00, 371.67] | -1.9% | OK |
| Creatinine NOT reported | 4 | 3,511 | 353.84 [302.83, 404.84] | +4.2% | OK |

##### 6.15.2 Comparison by Age / Sex / race

###### 6.15.2.1 Plot

```
urea_ma_3rd_analysis <- sex_age_race_subdivision_ma("urea",
                                                      urine_ma2,
                                                      overall_bias)
## Not enough studies for multivariable meta-regression (k = 3, need > 6). Running univariable models instead.
## Not enough studies for multivariable meta-regression (k = 4, need > 6). Running univariable models instead.
urea_ma_3rd_analysis$race_plot
## `height` was translated to `width`.
## `height` was translated to `width`.
## `height` was translated to `width`.
```

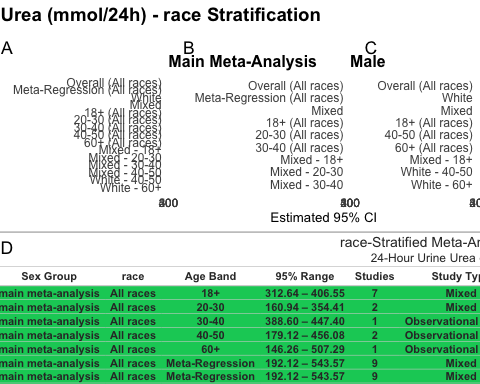

###### 6.15.2.2 Combined MA Results

urea_ma_3rd_analysis$combined_summary_table

Table 1: **Combined Meta-Analysis Results**

24-Hour Urine Urea (mmol/24h)

| Sex Group | race | Age Band | Pooled | 95% Range | Studies Mean SD Range | Studies | Study Type | Participants | I² | Cochran's Q | Q (p-value) | Egger's p | Model |
| --- | --- | --- | --- | --- | --- | --- | --- | --- | --- | --- | --- | --- | --- |
| main meta-analysis | All | 18+ | 359.59 | 312.64 – 406.55 | 360.74 ± 26.29 (315.68–382.95) | 7 | Mixed | 293 | 74.3% | 23.4 | <0.005 | 0.22 | Random |
| main meta-analysis | All | 20-30 | 257.68 | 160.94 – 354.41 | 259.53 ± 51.58 (223.06–296.00) | 2 | Mixed | 91 | 91.6% | 11.9 | <0.005 | — | Random |
| main meta-analysis | All | 30-40 | 418.00 | 388.60 – 447.40 | — (418.00–418.00) | 1 | Observational Cohort | 107 | — | — | — | — | Single study |
| main meta-analysis | All | 40-50 | 317.60 | 179.12 – 456.08 | 317.59 ± 70.69 (267.60–367.57) | 2 | Observational Cohort | 2601 | 99.9% | 899.6 | <0.005 | — | Random |
| main meta-analysis | All | 60+ | 326.77 | 146.26 – 507.29 | — (326.77–326.77) | 1 | Observational Cohort | 784 | — | — | — | — | Single study |
| **main meta-analysis** | **All** | **Meta-Regression** | **367.84** | **192.12 – 543.57** | **—** | **9** | **Mixed** | **464** | **0%** | **1.1** | **0.78** | **0.38** | **Multivariable** |
| **main meta-analysis** | **All** | **Meta-Regression** | **367.84** | **192.12 – 543.57** | **—** | **9** | **Mixed** | **464** | **0%** | **1.1** | **0.78** | **0.38** | **Multivariable** |
| **main meta-analysis** | **All** | **Overall** | **339.45** | **233.61 – 445.30** | **340.32 ± 54.03 (223.06–418.00)** | **13** | **Mixed** | **3876** | **99.6%** | **3061.6** | **<0.005** | **0.28** | **Random** |
| **main meta-analysis** | **All** | **Overall** | **339.45** | **233.61 – 445.30** | **340.32 ± 54.03 (223.06–418.00)** | **13** | **Mixed** | **3876** | **99.6%** | **3061.6** | **<0.005** | **0.28** | **Random** |
| main meta-analysis | black | 18+ | — | NA – NA | — | NA | NA | NA | — | — | — | — | NA |
| main meta-analysis | black | 20-30 | — | NA – NA | — | NA | NA | NA | — | — | — | — | NA |
| main meta-analysis | black | 30-40 | — | NA – NA | — | NA | NA | NA | — | — | — | — | NA |
| main meta-analysis | black | 40-50 | — | NA – NA | — | NA | NA | NA | — | — | — | — | NA |
| main meta-analysis | black | 60+ | — | NA – NA | — | NA | NA | NA | — | — | — | — | NA |
| **main meta-analysis** | **black** | **Meta-Regression** | **—** | **NA – NA** | **—** | **NA** | **NA** | **NA** | **—** | **—** | **—** | **—** | **NA** |
| **main meta-analysis** | **black** | **Overall** | **—** | **NA – NA** | **—** | **NA** | **NA** | **NA** | **—** | **—** | **—** | **—** | **NA** |
| main meta-analysis | mixed | 18+ | 359.59 | 312.64 – 406.55 | 360.74 ± 26.29 (315.68–382.95) | 7 | Mixed | 293 | 74.3% | 23.4 | <0.005 | 0.22 | Random |
| main meta-analysis | mixed | 20-30 | 257.68 | 160.94 – 354.41 | 259.53 ± 51.58 (223.06–296.00) | 2 | Mixed | 91 | 91.6% | 11.9 | <0.005 | — | Random |
| main meta-analysis | mixed | 30-40 | 418.00 | 388.60 – 447.40 | — (418.00–418.00) | 1 | Observational Cohort | 107 | — | — | — | — | Single study |
| main meta-analysis | mixed | 40-50 | 267.60 | 237.42 – 297.78 | — (267.60–267.60) | 1 | Observational Cohort | 34 | — | — | — | — | Single study |
| main meta-analysis | mixed | 60+ | — | NA – NA | — | NA | NA | NA | — | — | — | — | NA |
| **main meta-analysis** | **mixed** | **Meta-Regression** | **—** | **NA – NA** | **—** | **NA** | **NA** | **NA** | **—** | **—** | **—** | **—** | **NA** |
| **main meta-analysis** | **mixed** | **Overall** | **338.06** | **223.14 – 452.98** | **339.08 ± 58.38 (223.06–418.00)** | **11** | **Mixed** | **525** | **99.6%** | **2838.3** | **<0.005** | **0.34** | **Random** |
| main meta-analysis | white | 18+ | — | NA – NA | — | NA | NA | NA | — | — | — | — | NA |
| main meta-analysis | white | 20-30 | — | NA – NA | — | NA | NA | NA | — | — | — | — | NA |
| main meta-analysis | white | 30-40 | — | NA – NA | — | NA | NA | NA | — | — | — | — | NA |
| main meta-analysis | white | 40-50 | 367.57 | 165.66 – 569.48 | — (367.57–367.57) | 1 | Observational Cohort | 2567 | — | — | — | — | Single study |
| main meta-analysis | white | 60+ | 326.77 | 146.26 – 507.29 | — (326.77–326.77) | 1 | Observational Cohort | 784 | — | — | — | — | Single study |
| **main meta-analysis** | **white** | **Meta-Regression** | **—** | **NA – NA** | **—** | **NA** | **NA** | **NA** | **—** | **—** | **—** | **—** | **NA** |
| **main meta-analysis** | **white** | **Overall** | **347.25** | **290.96 – 403.54** | **347.17 ± 28.85 (326.77–367.57)** | **2** | **Observational Cohort** | **3351** | **99.1%** | **111.3** | **<0.005** | **—** | **Random** |
| male | All | 18+ | 315.68 | 111.72 – 519.65 | — (315.68–315.68) | 1 | Observational Cohort | 109 | — | — | — | — | Single study |
| male | All | 20-30 | 223.06 | 108.95 – 337.17 | — (223.06–223.06) | 1 | RCT | 38 | — | — | — | — | Single study |
| male | All | 30-40 | 418.00 | 388.60 – 447.40 | — (418.00–418.00) | 1 | Observational Cohort | 107 | — | — | — | — | Single study |
| male | All | 40-50 | — | NA – NA | — | NA | NA | NA | — | — | — | — | NA |
| male | All | 60+ | — | NA – NA | — | NA | NA | NA | — | — | — | — | NA |
| **male** | **All** | **Meta-Regression** | **315.68** | **124.81 – 506.55** | **—** | **3** | **Mixed** | **254** | **99.5%** | **510.6** | **<0.005** | **0.21** | **Univariable** |
| **male** | **All** | **Overall** | **319.22** | **128.35 – 510.09** | **318.91 ± 97.51 (223.06–418.00)** | **3** | **Mixed** | **254** | **99.6%** | **510.6** | **<0.005** | **0.21** | **Random** |
| male | black | 18+ | — | NA – NA | — | NA | NA | NA | — | — | — | — | NA |
| male | black | 20-30 | — | NA – NA | — | NA | NA | NA | — | — | — | — | NA |
| male | black | 30-40 | — | NA – NA | — | NA | NA | NA | — | — | — | — | NA |
| male | black | 40-50 | — | NA – NA | — | NA | NA | NA | — | — | — | — | NA |
| male | black | 60+ | — | NA – NA | — | NA | NA | NA | — | — | — | — | NA |
| **male** | **black** | **Meta-Regression** | **—** | **NA – NA** | **—** | **NA** | **NA** | **NA** | **—** | **—** | **—** | **—** | **NA** |
| **male** | **black** | **Overall** | **—** | **NA – NA** | **—** | **NA** | **NA** | **NA** | **—** | **—** | **—** | **—** | **NA** |
| male | mixed | 18+ | 315.68 | 111.72 – 519.65 | — (315.68–315.68) | 1 | Observational Cohort | 109 | — | — | — | — | Single study |
| male | mixed | 20-30 | 223.06 | 108.95 – 337.17 | — (223.06–223.06) | 1 | RCT | 38 | — | — | — | — | Single study |
| male | mixed | 30-40 | 418.00 | 388.60 – 447.40 | — (418.00–418.00) | 1 | Observational Cohort | 107 | — | — | — | — | Single study |
| male | mixed | 40-50 | — | NA – NA | — | NA | NA | NA | — | — | — | — | NA |
| male | mixed | 60+ | — | NA – NA | — | NA | NA | NA | — | — | — | — | NA |
| **male** | **mixed** | **Meta-Regression** | **—** | **NA – NA** | **—** | **NA** | **NA** | **NA** | **—** | **—** | **—** | **—** | **NA** |
| **male** | **mixed** | **Overall** | **319.22** | **128.35 – 510.09** | **318.91 ± 97.51 (223.06–418.00)** | **3** | **Mixed** | **254** | **99.6%** | **510.6** | **<0.005** | **0.21** | **Random** |
| male | white | 18+ | — | NA – NA | — | NA | NA | NA | — | — | — | — | NA |
| male | white | 20-30 | — | NA – NA | — | NA | NA | NA | — | — | — | — | NA |
| male | white | 30-40 | — | NA – NA | — | NA | NA | NA | — | — | — | — | NA |
| male | white | 40-50 | — | NA – NA | — | NA | NA | NA | — | — | — | — | NA |
| male | white | 60+ | — | NA – NA | — | NA | NA | NA | — | — | — | — | NA |
| **male** | **white** | **Meta-Regression** | **—** | **NA – NA** | **—** | **NA** | **NA** | **NA** | **—** | **—** | **—** | **—** | **NA** |
| **male** | **white** | **Overall** | **—** | **NA – NA** | **—** | **NA** | **NA** | **NA** | **—** | **—** | **—** | **—** | **NA** |
| female | All | 18+ | 374.63 | 178.82 – 570.43 | 374.63 ± 9.61 (366.30–382.95) | 4 | Observational Cohort | 156 | 0% | 1.1 | 0.78 | — | Fixed |
| female | All | 20-30 | — | NA – NA | — | NA | NA | NA | — | — | — | — | NA |
| female | All | 30-40 | — | NA – NA | — | NA | NA | NA | — | — | — | — | NA |
| female | All | 40-50 | 367.57 | 165.66 – 569.48 | — (367.57–367.57) | 1 | Observational Cohort | 2567 | — | — | — | — | Single study |
| female | All | 60+ | 326.77 | 146.26 – 507.29 | — (326.77–326.77) | 1 | Observational Cohort | 784 | — | — | — | — | Single study |
| **female** | **All** | **Meta-Regression** | **—** | **NA – NA** | **—** | **NA** | **NA** | **NA** | **—** | **—** | **—** | **—** | **NA** |
| **female** | **All** | **Overall** | **362.62** | **322.03 – 403.21** | **365.47 ± 20.56 (326.77–382.95)** | **6** | **Observational Cohort** | **3507** | **95.7%** | **117.4** | **<0.005** | **0.98** | **Random** |
| female | black | 18+ | — | NA – NA | — | NA | NA | NA | — | — | — | — | NA |
| female | black | 20-30 | — | NA – NA | — | NA | NA | NA | — | — | — | — | NA |
| female | black | 30-40 | — | NA – NA | — | NA | NA | NA | — | — | — | — | NA |
| female | black | 40-50 | — | NA – NA | — | NA | NA | NA | — | — | — | — | NA |
| female | black | 60+ | — | NA – NA | — | NA | NA | NA | — | — | — | — | NA |
| **female** | **black** | **Meta-Regression** | **—** | **NA – NA** | **—** | **NA** | **NA** | **NA** | **—** | **—** | **—** | **—** | **NA** |
| **female** | **black** | **Overall** | **—** | **NA – NA** | **—** | **NA** | **NA** | **NA** | **—** | **—** | **—** | **—** | **NA** |
| female | mixed | 18+ | 374.63 | 178.82 – 570.43 | 374.63 ± 9.61 (366.30–382.95) | 4 | Observational Cohort | 156 | 0% | 1.1 | 0.78 | — | Fixed |
| female | mixed | 20-30 | — | NA – NA | — | NA | NA | NA | — | — | — | — | NA |
| female | mixed | 30-40 | — | NA – NA | — | NA | NA | NA | — | — | — | — | NA |
| female | mixed | 40-50 | — | NA – NA | — | NA | NA | NA | — | — | — | — | NA |
| female | mixed | 60+ | — | NA – NA | — | NA | NA | NA | — | — | — | — | NA |
| **female** | **mixed** | **Meta-Regression** | **—** | **NA – NA** | **—** | **NA** | **NA** | **NA** | **—** | **—** | **—** | **—** | **NA** |
| **female** | **mixed** | **Overall** | **374.63** | **178.82 – 570.43** | **374.63 ± 9.61 (366.30–382.95)** | **4** | **Observational Cohort** | **156** | **0%** | **1.1** | **0.78** | **—** | **Fixed** |
| female | white | 18+ | — | NA – NA | — | NA | NA | NA | — | — | — | — | NA |
| female | white | 20-30 | — | NA – NA | — | NA | NA | NA | — | — | — | — | NA |
| female | white | 30-40 | — | NA – NA | — | NA | NA | NA | — | — | — | — | NA |
| female | white | 40-50 | 367.57 | 165.66 – 569.48 | — (367.57–367.57) | 1 | Observational Cohort | 2567 | — | — | — | — | Single study |
| female | white | 60+ | 326.77 | 146.26 – 507.29 | — (326.77–326.77) | 1 | Observational Cohort | 784 | — | — | — | — | Single study |
| **female** | **white** | **Meta-Regression** | **—** | **NA – NA** | **—** | **NA** | **NA** | **NA** | **—** | **—** | **—** | **—** | **NA** |
| **female** | **white** | **Overall** | **347.25** | **290.96 – 403.54** | **347.17 ± 28.85 (326.77–367.57)** | **2** | **Observational Cohort** | **3351** | **99.1%** | **111.3** | **<0.005** | **—** | **Random** |
| *I² represents heterogeneity; Q test assesses between-study variation; Egger's p = linear regression test for funnel plot asymmetry; Meta-Regression shows covariate-adjusted estimates* | | | | | | | | | | | | | |

###### 6.15.2.3 Table of Results subdivided by race / Sex / Age

urea_ma_3rd_analysis$race_summary_table

Table 1: **race-Stratified Meta-Analysis Results**

24-Hour Urine Urea (mmol/24h)

| Sex Group | race | Age Band | 95% Range | Studies | Study Type | Participants | I² | Cochran's Q | Q (p-value) | Egger's p | Model |
| --- | --- | --- | --- | --- | --- | --- | --- | --- | --- | --- | --- |
| **main meta-analysis** | **All races** | **18+** | **312.64 – 406.55** | **7** | **Mixed** | **293** | **74.3%** | **23.4** | **<0.005** | **0.22** | **Random** |
| **main meta-analysis** | **All races** | **20-30** | **160.94 – 354.41** | **2** | **Mixed** | **91** | **91.6%** | **11.9** | **<0.005** | **—** | **Random** |
| **main meta-analysis** | **All races** | **30-40** | **388.60 – 447.40** | **1** | **Observational Cohort** | **107** | **—** | **—** | **—** | **—** | **Single study** |
| **main meta-analysis** | **All races** | **40-50** | **179.12 – 456.08** | **2** | **Observational Cohort** | **2601** | **99.9%** | **899.6** | **<0.005** | **—** | **Random** |
| **main meta-analysis** | **All races** | **60+** | **146.26 – 507.29** | **1** | **Observational Cohort** | **784** | **—** | **—** | **—** | **—** | **Single study** |
| **main meta-analysis** | **All races** | **Meta-Regression** | **192.12 – 543.57** | **9** | **Mixed** | **464** | **0%** | **1.1** | **0.78** | **0.38** | **Multivariable** |
| **main meta-analysis** | **All races** | **Meta-Regression** | **192.12 – 543.57** | **9** | **Mixed** | **464** | **0%** | **1.1** | **0.78** | **0.38** | **Multivariable** |
| **main meta-analysis** | **All races** | **Overall** | **233.61 – 445.30** | **13** | **Mixed** | **3876** | **99.6%** | **3061.6** | **<0.005** | **0.28** | **Random** |
| **main meta-analysis** | **All races** | **Overall** | **233.61 – 445.30** | **13** | **Mixed** | **3876** | **99.6%** | **3061.6** | **<0.005** | **0.28** | **Random** |
| main meta-analysis | Black | 18+ | — | — | NA | — | — | — | — | — | — |
| main meta-analysis | Black | 20-30 | — | — | NA | — | — | — | — | — | — |
| main meta-analysis | Black | 30-40 | — | — | NA | — | — | — | — | — | — |
| main meta-analysis | Black | 40-50 | — | — | NA | — | — | — | — | — | — |
| main meta-analysis | Black | 60+ | — | — | NA | — | — | — | — | — | — |
| main meta-analysis | Black | Meta-Regression | — | — | NA | — | — | — | — | — | — |
| **main meta-analysis** | **Black** | **Overall** | **—** | **—** | **NA** | **—** | **—** | **—** | **—** | **—** | **—** |
| main meta-analysis | Mixed | 18+ | 312.64 – 406.55 | 7 | Mixed | 293 | 74.3% | 23.4 | <0.005 | 0.22 | Random |
| main meta-analysis | Mixed | 20-30 | 160.94 – 354.41 | 2 | Mixed | 91 | 91.6% | 11.9 | <0.005 | — | Random |
| main meta-analysis | Mixed | 30-40 | 388.60 – 447.40 | 1 | Observational Cohort | 107 | — | — | — | — | Single study |
| main meta-analysis | Mixed | 40-50 | 237.42 – 297.78 | 1 | Observational Cohort | 34 | — | — | — | — | Single study |
| main meta-analysis | Mixed | 60+ | — | — | NA | — | — | — | — | — | — |
| main meta-analysis | Mixed | Meta-Regression | — | — | NA | — | — | — | — | — | — |
| **main meta-analysis** | **Mixed** | **Overall** | **223.14 – 452.98** | **11** | **Mixed** | **525** | **99.6%** | **2838.3** | **<0.005** | **0.34** | **Random** |
| main meta-analysis | White | 18+ | — | — | NA | — | — | — | — | — | — |
| main meta-analysis | White | 20-30 | — | — | NA | — | — | — | — | — | — |
| main meta-analysis | White | 30-40 | — | — | NA | — | — | — | — | — | — |
| main meta-analysis | White | 40-50 | 165.66 – 569.48 | 1 | Observational Cohort | 2567 | — | — | — | — | Single study |
| main meta-analysis | White | 60+ | 146.26 – 507.29 | 1 | Observational Cohort | 784 | — | — | — | — | Single study |
| main meta-analysis | White | Meta-Regression | — | — | NA | — | — | — | — | — | — |
| **main meta-analysis** | **White** | **Overall** | **290.96 – 403.54** | **2** | **Observational Cohort** | **3351** | **99.1%** | **111.3** | **<0.005** | **—** | **Random** |
| **male** | **All races** | **18+** | **111.72 – 519.65** | **1** | **Observational Cohort** | **109** | **—** | **—** | **—** | **—** | **Single study** |
| **male** | **All races** | **20-30** | **108.95 – 337.17** | **1** | **RCT** | **38** | **—** | **—** | **—** | **—** | **Single study** |
| **male** | **All races** | **30-40** | **388.60 – 447.40** | **1** | **Observational Cohort** | **107** | **—** | **—** | **—** | **—** | **Single study** |
| **male** | **All races** | **40-50** | **—** | **—** | **NA** | **—** | **—** | **—** | **—** | **—** | **—** |
| **male** | **All races** | **60+** | **—** | **—** | **NA** | **—** | **—** | **—** | **—** | **—** | **—** |
| **male** | **All races** | **Meta-Regression** | **124.81 – 506.55** | **3** | **Mixed** | **254** | **99.5%** | **510.6** | **<0.005** | **0.21** | **Univariable** |
| **male** | **All races** | **Overall** | **128.35 – 510.09** | **3** | **Mixed** | **254** | **99.6%** | **510.6** | **<0.005** | **0.21** | **Random** |
| male | Black | 18+ | — | — | NA | — | — | — | — | — | — |
| male | Black | 20-30 | — | — | NA | — | — | — | — | — | — |
| male | Black | 30-40 | — | — | NA | — | — | — | — | — | — |
| male | Black | 40-50 | — | — | NA | — | — | — | — | — | — |
| male | Black | 60+ | — | — | NA | — | — | — | — | — | — |
| male | Black | Meta-Regression | — | — | NA | — | — | — | — | — | — |
| **male** | **Black** | **Overall** | **—** | **—** | **NA** | **—** | **—** | **—** | **—** | **—** | **—** |
| male | Mixed | 18+ | 111.72 – 519.65 | 1 | Observational Cohort | 109 | — | — | — | — | Single study |
| male | Mixed | 20-30 | 108.95 – 337.17 | 1 | RCT | 38 | — | — | — | — | Single study |
| male | Mixed | 30-40 | 388.60 – 447.40 | 1 | Observational Cohort | 107 | — | — | — | — | Single study |
| male | Mixed | 40-50 | — | — | NA | — | — | — | — | — | — |
| male | Mixed | 60+ | — | — | NA | — | — | — | — | — | — |
| male | Mixed | Meta-Regression | — | — | NA | — | — | — | — | — | — |
| **male** | **Mixed** | **Overall** | **128.35 – 510.09** | **3** | **Mixed** | **254** | **99.6%** | **510.6** | **<0.005** | **0.21** | **Random** |
| male | White | 18+ | — | — | NA | — | — | — | — | — | — |
| male | White | 20-30 | — | — | NA | — | — | — | — | — | — |
| male | White | 30-40 | — | — | NA | — | — | — | — | — | — |
| male | White | 40-50 | — | — | NA | — | — | — | — | — | — |
| male | White | 60+ | — | — | NA | — | — | — | — | — | — |
| male | White | Meta-Regression | — | — | NA | — | — | — | — | — | — |
| **male** | **White** | **Overall** | **—** | **—** | **NA** | **—** | **—** | **—** | **—** | **—** | **—** |
| **female** | **All races** | **18+** | **178.82 – 570.43** | **4** | **Observational Cohort** | **156** | **0%** | **1.1** | **0.78** | **—** | **Fixed** |
| **female** | **All races** | **20-30** | **—** | **—** | **NA** | **—** | **—** | **—** | **—** | **—** | **—** |
| **female** | **All races** | **30-40** | **—** | **—** | **NA** | **—** | **—** | **—** | **—** | **—** | **—** |
| **female** | **All races** | **40-50** | **165.66 – 569.48** | **1** | **Observational Cohort** | **2567** | **—** | **—** | **—** | **—** | **Single study** |
| **female** | **All races** | **60+** | **146.26 – 507.29** | **1** | **Observational Cohort** | **784** | **—** | **—** | **—** | **—** | **Single study** |
| **female** | **All races** | **Meta-Regression** | **—** | **—** | **NA** | **—** | **—** | **—** | **—** | **—** | **—** |
| **female** | **All races** | **Overall** | **322.03 – 403.21** | **6** | **Observational Cohort** | **3507** | **95.7%** | **117.4** | **<0.005** | **0.98** | **Random** |
| female | Black | 18+ | — | — | NA | — | — | — | — | — | — |
| female | Black | 20-30 | — | — | NA | — | — | — | — | — | — |
| female | Black | 30-40 | — | — | NA | — | — | — | — | — | — |
| female | Black | 40-50 | — | — | NA | — | — | — | — | — | — |
| female | Black | 60+ | — | — | NA | — | — | — | — | — | — |
| female | Black | Meta-Regression | — | — | NA | — | — | — | — | — | — |
| **female** | **Black** | **Overall** | **—** | **—** | **NA** | **—** | **—** | **—** | **—** | **—** | **—** |
| female | Mixed | 18+ | 178.82 – 570.43 | 4 | Observational Cohort | 156 | 0% | 1.1 | 0.78 | — | Fixed |
| female | Mixed | 20-30 | — | — | NA | — | — | — | — | — | — |
| female | Mixed | 30-40 | — | — | NA | — | — | — | — | — | — |
| female | Mixed | 40-50 | — | — | NA | — | — | — | — | — | — |
| female | Mixed | 60+ | — | — | NA | — | — | — | — | — | — |
| female | Mixed | Meta-Regression | — | — | NA | — | — | — | — | — | — |
| **female** | **Mixed** | **Overall** | **178.82 – 570.43** | **4** | **Observational Cohort** | **156** | **0%** | **1.1** | **0.78** | **—** | **Fixed** |
| female | White | 18+ | — | — | NA | — | — | — | — | — | — |
| female | White | 20-30 | — | — | NA | — | — | — | — | — | — |
| female | White | 30-40 | — | — | NA | — | — | — | — | — | — |
| female | White | 40-50 | 165.66 – 569.48 | 1 | Observational Cohort | 2567 | — | — | — | — | Single study |
| female | White | 60+ | 146.26 – 507.29 | 1 | Observational Cohort | 784 | — | — | — | — | Single study |
| female | White | Meta-Regression | — | — | NA | — | — | — | — | — | — |
| **female** | **White** | **Overall** | **290.96 – 403.54** | **2** | **Observational Cohort** | **3351** | **99.1%** | **111.3** | **<0.005** | **—** | **Random** |
| *I² represents heterogeneity; Q test assesses between-study variation; Egger's p = linear regression test for funnel plot asymmetry; — indicates insufficient data* | | | | | | | | | | | |

###### 6.15.2.4 Wald Tests

urea_ma_3rd_analysis$wald_tests$sex_comparison

Table 1: **Sex Comparison (Overall)**

| Comparison | % Shift | P (Wald Test) | Mean Diff (95% CI) |
| --- | --- | --- | --- |
| Male vs Female (Overall) | -12.0% | 0.663 | -43.40 [-238.54, 151.73] |
| *Mean difference = Group 1 − Group 2 in component units (mmol/24h, L/24h, or pH); % shift = difference relative to Group 2.* | | | |

urea_ma_3rd_analysis$wald_tests$race_comparisons

Table 1: **race Comparisons Within Sex Groups**

| Sex Group | Comparison | % Shift | P (Wald Test) | Mean Diff (95% CI) |
| --- | --- | --- | --- | --- |
| male | black vs mixed | — | — | — |
| male | black vs white | — | — | — |
| male | mixed vs white | — | — | — |
| female | black vs mixed | — | — | — |
| female | black vs white | — | — | — |
| female | mixed vs white | +7.9% | 0.792 | 27.37 [-176.36, 231.11] |
| *Mean difference = Group 1 − Group 2 in component units (mmol/24h, L/24h, or pH); % shift = difference relative to Group 2.* | | | | |

urea_ma_3rd_analysis$wald_tests$age_band_comparisons

Table 1: **Age Band Comparisons Within Sex Groups**

| Sex Group | Comparison | % Shift | P (Wald Test) | Mean Diff (95% CI) |
| --- | --- | --- | --- | --- |
| main meta-analysis | 18+ vs 20-30 | +39.6% | 0.063 | 101.92 [-5.61, 209.44] |
| main meta-analysis | 18+ vs 30-40 | -14.0% | <0.05 | -58.41 [-113.80, -3.01] |
| main meta-analysis | 18+ vs 40-50 | +13.2% | 0.573 | 41.99 [-104.23, 188.21] |
| main meta-analysis | 18+ vs 60+ | +10.0% | 0.730 | 32.82 [-153.70, 219.34] |
| main meta-analysis | 20-30 vs 30-40 | -38.4% | <0.01 | -160.32 [-261.43, -59.22] |
| main meta-analysis | 20-30 vs 40-50 | -18.9% | 0.487 | -59.92 [-228.84, 109.00] |
| main meta-analysis | 20-30 vs 60+ | -21.1% | 0.508 | -69.09 [-273.90, 135.71] |
| main meta-analysis | 30-40 vs 40-50 | +31.6% | 0.165 | 100.40 [-41.16, 241.96] |
| main meta-analysis | 30-40 vs 60+ | +27.9% | 0.328 | 91.23 [-91.67, 274.12] |
| main meta-analysis | 40-50 vs 60+ | -2.8% | 0.937 | -9.17 [-236.68, 218.34] |
| male | 18+ vs 20-30 | +41.5% | 0.437 | 92.62 [-141.09, 326.34] |
| male | 18+ vs 30-40 | -24.5% | 0.330 | -102.32 [-308.39, 103.76] |
| male | 18+ vs 40-50 | — | — | — |
| male | 18+ vs 60+ | — | — | — |
| male | 20-30 vs 30-40 | -46.6% | <0.01 | -194.94 [-312.78, -77.10] |
| male | 20-30 vs 40-50 | — | — | — |
| male | 20-30 vs 60+ | — | — | — |
| male | 30-40 vs 40-50 | — | — | — |
| male | 30-40 vs 60+ | — | — | — |
| male | 40-50 vs 60+ | — | — | — |
| female | 18+ vs 20-30 | — | — | — |
| female | 18+ vs 30-40 | — | — | — |
| female | 18+ vs 40-50 | +1.9% | 0.961 | 7.05 [-274.21, 288.31] |
| female | 18+ vs 60+ | +14.6% | 0.725 | 47.85 [-218.47, 314.17] |
| female | 20-30 vs 30-40 | — | — | — |
| female | 20-30 vs 40-50 | — | — | — |
| female | 20-30 vs 60+ | — | — | — |
| female | 30-40 vs 40-50 | — | — | — |
| female | 30-40 vs 60+ | — | — | — |
| female | 40-50 vs 60+ | +12.5% | 0.768 | 40.80 [-230.04, 311.64] |
| *Mean difference = Group 1 − Group 2 in component units (mmol/24h, L/24h, or pH); % shift = difference relative to Group 2.* | | | | |

urea_ma_3rd_analysis$wald_tests$sex_within_age_bands

Table 1: **Sex Comparisons Within Age Bands**

| Age Band | % Shift | P (Wald Test) | Mean Diff (95% CI) |
| --- | --- | --- | --- |
| 18+ | -15.7% | 0.683 | -58.94 [-341.68, 223.80] |
| 20-30 | — | — | — |
| 30-40 | — | — | — |
| 40-50 | — | — | — |
| 60+ | — | — | — |
| *Mean difference = Group 1 − Group 2 in component units (mmol/24h, L/24h, or pH); % shift = difference relative to Group 2.* | | | |

### 7 Bias Analysis

#### 7.1 Observational Studies

```
bias_observational_table <- bias_observational %>%
  rename_with(~str_remove(., "_low_moderate_high"), everything()) %>%
  arrange(study) %>%
  rename(
    Study = study,
    `Study Participation` = study_participation,
    `Study Attrition` = study_attrition,
    `Prognostic Factor Measurement` = prognostic_factor_measurement,
    `Outcomes Measurement` = outcomes_measurement,
    `Study Confounding` = study_confounding,
    `Statistical Analysis` = statistical_analysis,
    Overall = overall
  )

bias_observational_table %>%
  gt() %>%
  tab_header(
    title = "Risk of Bias Assessment for Observational Studies"
  ) %>%
  # Color coding function
  data_color(
    columns = c(`Study Participation`, `Study Attrition`, `Prognostic Factor Measurement`,
                `Outcomes Measurement`, `Study Confounding`, `Statistical Analysis`, Overall),
    fn = function(x) {
      case_when(
        x == "Low" ~ "#90EE90",
        x == "Moderate" ~ "#FFD700",
        x == "High" ~ "#FF6B6B",
        TRUE ~ "#FFFFFF"
      )
    }
  ) %>%
  tab_style(
    style = cell_text(weight = "bold"),
    locations = cells_column_labels()
  ) %>%
  tab_style(
    style = cell_text(weight = "bold"),
    locations = cells_body(columns = Overall)
  ) %>%
  cols_align(
    align = "center",
    columns = c(`Study Participation`, `Study Attrition`, `Prognostic Factor Measurement`,
                `Outcomes Measurement`, `Study Confounding`, `Statistical Analysis`, Overall)
  ) %>%
  cols_align(
    align = "left",
    columns = Study
  ) %>%
  opt_row_striping()
```

Table 1: Risk of Bias Assessment for Observational Studies

| Study | Study Participation | Study Attrition | Prognostic Factor Measurement | Outcomes Measurement | Study Confounding | Statistical Analysis | Overall |
| --- | --- | --- | --- | --- | --- | --- | --- |
| Abate et al. 2004 | Moderate | High | Low | Low | Moderate | Low | **High** |
| Arora et al. 2017 | High | High | Moderate | Moderate | High | High | **High** |
| Arrabal-Polo et al. 2012 | Moderate | High | Low | Low | Moderate | Moderate | **High** |
| Arrabal-Polo et al. 2015 | Low | Low | Low | Low | Moderate | Moderate | **Moderate** |
| Batailee et al. 1991 | Moderate | High | Moderate | Low | Moderate | Low | **High** |
| Buchholz et al. 1996 | High | High | Low | Low | High | Moderate | **High** |
| Cameron et al 2006 | Low | Low | Low | Low | Low | Low | **Low** |
| Christie et al. 1996 | Moderate | Low | Low | Moderate | High | Moderate | **High** |
| Ciacci et al. 2008 | Moderate | High | Moderate | Low | High | Moderate | **High** |
| Curhan et al. 2001 | Low | Low | Low | Moderate | Moderate | Low | **Moderate** |
| Defoor et al. 2005 | Moderate | Moderate | Low | Low | High | High | **High** |
| Deng et al. 2017 | Low | Low | Low | Low | Low | Low | **Low** |
| Doenyas-Barak et al. 2017 | High | High | Moderate | Moderate | High | Moderate | **High** |
| Doizi et al. 2016 | Low | Low | Low | Low | Low | Low | **Low** |
| Duffey et al. 2011 | Low | Moderate | Low | Low | Moderate | Low | **Moderate** |
| Elkoushy et al. 2014 | Low | Moderate | Low | Low | Moderate | Moderate | **Moderate** |
| Erwin et al. 1994 | Low | High | Low | Low | High | Low | **High** |
| Ferraro et al. 2023 | Low | Low | Low | Low | Low | Low | **Low** |
| Ferraro et al. 2024 | Low | Low | Low | Low | Low | Low | **Low** |
| Goldschmied et al. 1975 | Moderate | Low | Low | Low | Low | High | **High** |
| Goodzari et al. 2012 | Moderate | High | Moderate | Moderate | High | High | **High** |
| Grases et al. 2015 | Moderate | High | Moderate | Moderate | High | High | **High** |
| HPFS et al. 40-50 | Low | Low | Low | Low | Moderate | Low | **Low** |
| HPFS et al. 50-60 | Low | Low | Low | Low | Moderate | Low | **Low** |
| HPFS et al. 60-70 | Low | Low | Low | Low | Moderate | Low | **Low** |
| HPFS et al. 70-80 | Low | Low | Low | Low | Moderate | Low | **Low** |
| Hess et al. 1999 | Moderate | High | Moderate | Moderate | High | Low | **High** |
| Hong et al. 2012 | Moderate | Low | Moderate | Moderate | Moderate | Low | **Moderate** |
| Hunag et al. 2003 | Low | High | Low | Low | High | Low | **High** |
| Kinder et al. 2002 | Moderate | Moderate | Low | Low | Low | Low | **Moderate** |
| Kuo et al. 2003 | Moderate | Low | Low | Low | Moderate | Low | **Moderate** |
| Lemann Jr et al. 1996 | Moderate | Low | Low | Low | Moderate | Low | **Moderate** |
| Ma et al. 2014 | Low | Low | Low | Low | Low | Moderate | **Low** |
| Maalouf et al. 2010 | Low | Low | Low | Low | Low | Low | **Low** |
| Mai et al. 2019 | Low | Low | Low | Low | Low | Low | **Low** |
| Marya et al. 1979 | High | High | High | High | High | High | **High** |
| Matin et al. 2000 | Moderate | Low | Moderate | Moderate | Low | Low | **Moderate** |
| Michelacci et al. 1989 | High | High | Low | Low | Moderate | Moderate | **High** |
| Mikaye et al. 2001 | High | Low | Low | Low | High | High | **High** |
| NHS et al. 40-50 | Low | Low | Low | Low | Moderate | Low | **Low** |
| NHS et al. 40-50 | Low | Low | Low | Low | Moderate | Low | **Low** |
| NHS et al. 50-60 | Low | Low | Low | Low | Moderate | Low | **Low** |
| NHS et al. 50-60 | Low | Low | Low | Low | Moderate | Low | **Low** |
| NHS et al. 60-70 | Low | Low | Low | Low | Moderate | Low | **Low** |
| NHS et al. 60-70 | Low | Low | Low | Low | Moderate | Low | **Low** |
| NHS et al. 70-80 | Low | Low | Low | Low | Moderate | Low | **Low** |
| NHS et al. 70-80 | Low | Low | Low | Low | Moderate | Low | **Low** |
| Nouvenne et al. 2013 | High | Low | Low | Low | Low | Low | **High** |
| Ogawa et al. 2003 | High | Low | Low | Low | High | Low | **High** |
| Pais et al. 2007 | High | High | Low | Low | High | Moderate | **High** |
| Parvin et al. 2011 | Low | Moderate | Low | Low | Low | Low | **Low** |
| Perez-Brayfield et al. 2002 | Low | Moderate | Low | Low | Moderate | Moderate | **Moderate** |
| Perinpam et al. 2015 | Low | Moderate | Low | Low | Low | Low | **Low** |
| Pigna et al. 2013 | Moderate | High | Low | Low | Moderate | Moderate | **High** |
| Prochaska et al. 2018 | Low | Low | Low | Low | Low | Low | **Low** |
| Roders et al. 2009 | Low | High | Low | Low | High | Moderate | **High** |
| Rodgers et al. 1997 | High | High | High | High | High | High | **High** |
| Semins et al. 2010 | Low | Low | Low | Low | Low | Low | **Low** |
| Shah et al. 2005 | Moderate | High | Low | Moderate | Low | Low | **High** |
| Sorensen et al. 2011 | Low | Moderate | Low | Low | Moderate | Low | **Moderate** |
| Stern et al. 1980 | High | High | Moderate | Moderate | Low | Moderate | **High** |
| Stitchantrakul et al. 2007 | Moderate | Low | Low | Low | Moderate | Moderate | **Moderate** |
| Taylor et al. 2006 | Low | Low | Low | Low | Low | Low | **Low** |
| Terris et al. 2001 | Moderate | Low | Low | Low | High | Moderate | **High** |
| Thongprayoon et al. 2022 | Low | Moderate | Low | Low | Moderate | Low | **Low** |
| Welshman et al 1976 | Low | Low | Moderate | Low | Moderate | Low | **Moderate** |
| Welshmanet al. 1975 | High | High | Moderate | Moderate | High | Moderate | **High** |
| Williams et al. 2001 | Moderate | High | Low | Moderate | Moderate | Low | **Moderate** |

#### 7.2 RCTs

```
bias_rcts_table <- bias_rcts %>%
  mutate(
    risk_of_bias_in_selection_of_reported_result_low_some_concern_high = case_when(
      risk_of_bias_in_selection_of_reported_result_low_some_concern_high == "Low" ~ "Low",
      risk_of_bias_in_selection_of_reported_result_low_some_concern_high == "Some Concern" ~ "Moderate",
      TRUE ~ "Low"
    )
  ) %>%
  rename_with(~str_remove(., "_low_some_concern_high"), everything()) %>%
  arrange(study) %>%
  rename(
    Study = study,
    `Randomisation` = risk_of_bias_from_randomising_process,
    `Deviations from Intended Intervention Effect - Assignment` = risk_of_bias_due_to_deviations_from_the_intended_interventions_effect_of_assingment_to_intervention,
    `Deviations from Intended Intervention Effect - Adherence` = risk_of_bias_due_to_deviations_from_the_intended_interventions_effect_of_adhering_to_intervention,

    `Missing Outcomes` = missing_outcome_data,
    `Measurement of Outcome` = risk_of_bias_in_measurement_of_outcome,
    `Selection of Reported Result` = risk_of_bias_in_selection_of_reported_result,
    Overall = overall
  ) %>%
  mutate(across(-Study, ~str_replace_all(., "Some concern", "Moderate")),
         across(-Study, ~str_replace_all(., "Low ", "Low")),
         across(-Study, ~str_replace_all(., "Some Concern", "Moderate")))

bias_rcts_table %>%
  gt() %>%
  tab_header(
    title = "Risk of Bias Assessment for Randomised Controlled Studies"
  ) %>%
  # Color coding function
  data_color(
    columns = c(`Randomisation`, `Deviations from Intended Intervention Effect - Assignment`, `Deviations from Intended Intervention Effect - Adherence`,
                `Missing Outcomes`, `Measurement of Outcome`, `Selection of Reported Result`, Overall),
    fn = function(x) {
      case_when(
        x == "Low" ~ "#90EE90",
        x == "Moderate" ~ "#FFD700",
        x == "High" ~ "#FF6B6B",
        TRUE ~ "#FFFFFF"
      )
    }
  ) %>%
  tab_style(
    style = cell_text(weight = "bold"),
    locations = cells_column_labels()
  ) %>%
  tab_style(
    style = cell_text(weight = "bold"),
    locations = cells_body(columns = Overall)
  ) %>%
  cols_align(
    align = "center",
    columns = c(`Randomisation`, `Deviations from Intended Intervention Effect - Assignment`, `Deviations from Intended Intervention Effect - Adherence`,
                `Missing Outcomes`, `Measurement of Outcome`, `Selection of Reported Result`, Overall)
  ) %>%
  cols_align(
    align = "left",
    columns = Study
  ) %>%
  opt_row_striping()
```

Table 1: Risk of Bias Assessment for Randomised Controlled Studies

| Study | Randomisation | Deviations from Intended Intervention Effect - Assignment | Deviations from Intended Intervention Effect - Adherence | Missing Outcomes | Measurement of Outcome | Selection of Reported Result | Overall |
| --- | --- | --- | --- | --- | --- | --- | --- |
| Allie et al. 2003 | High | Moderate | Moderate | Low | Moderate | Low | **High** |
| Baxmann et al. 2003 | High | Moderate | Moderate | Low | Moderate | Low | **High** |
| Chai et al. 2004 | Moderate | Moderate | Moderate | Low | Moderate | Low | **Moderate** |
| Gettman et al. | Moderate | Moderate | Moderate | Low | Moderate | Low | **Moderate** |
| Herrel et al. 2012 | Moderate | Low | Low | Low | Moderate | Low | **Moderate** |
| Holmes et al 2001 | Low | Low | Moderate | Low | Moderate | Low | **Moderate** |
| Lange et al. 2014 | Moderate | Moderate | Moderate | Low | Moderate | Low | **Moderate** |
| Lewandowski et al. 2004 | Moderate | Moderate | Moderate | Low | Moderate | Low | **Moderate** |
| Matsumoto et al. 2006 | High | Moderate | Moderate | Low | Moderate | Low | **High** |
| Mirazazadeh et al. 2011 | High | Moderate | Moderate | Low | High | Low | **High** |
| Rodgers et al. 2002 | Low | Low | Moderate | Low | Moderate | Low | **Moderate** |
| Stitchantrakul et al. 2004 | High | Moderate | Moderate | High | Moderate | Low | **High** |
| Sweeney et al. 2009 | High | Moderate | Moderate | Low | Moderate | Low | **High** |
| Traxer et al. 2003 | Low | Low | Low | Low | Low | Low | **Low** |
| Wabner et al. 1992 | Moderate | Moderate | Moderate | Low | Moderate | Moderate | **Moderate** |

#### 7.3 Overall Bias

```
overall_bias %>%
  mutate(
    Study = study_name,
    Overall = factor(overall, levels = c("Low", "Moderate", "High"))
  ) %>%
  select(Study, Overall) %>%
  arrange(Study) %>%
  arrange(Overall) %>%
  gt() %>%
  tab_header(
    title = md("**Overall Risk of Bias Assessment**"),
    subtitle = "Summary across all studies"
  ) %>%
  data_color(
    columns = Overall,
    fn = function(x) {
      case_when(
        x == "Low" ~ "#90EE90",
        x == "Moderate" ~ "#FFD700",
        x == "High" ~ "#FF6B6B",
        TRUE ~ "#FFFFFF"
      )
    }
  ) %>%
  # Styling
  tab_style(
    style = cell_text(weight = "bold", align = "center"),
    locations = cells_column_labels()
  ) %>%
  tab_style(
    style = cell_text(weight = "bold", align = "center"),
    locations = cells_body(columns = Overall)
  ) %>%
  cols_align(
    align = "left",
    columns = Study
  ) %>%
  cols_align(
    align = "center",
    columns = Overall
  ) %>%
  cols_width(
    Study ~ px(300),
    Overall ~ px(150)
  ) %>%
  opt_row_striping() %>%
  tab_options(
    table.font.size = 14,
    heading.title.font.size = 18,
    heading.subtitle.font.size = 14,
    column_labels.font.weight = "bold",
    table.border.top.style = "solid",
    table.border.top.width = px(3),
    table.border.top.color = "#323232",
    column_labels.border.bottom.style = "solid",
    column_labels.border.bottom.width = px(2),
    column_labels.border.bottom.color = "#323232"
  )
```

Table 1: **Overall Risk of Bias Assessment**

Summary across all studies

| Study | Overall |
| --- | --- |
| Cameron et al 2006 | **Low** |
| Deng et al. 2017 | **Low** |
| Doizi et al. 2016 | **Low** |
| Ferraro et al. 2023 | **Low** |
| Ferraro et al. 2024 | **Low** |
| HPFS et al. 40-50 | **Low** |
| HPFS et al. 50-60 | **Low** |
| HPFS et al. 60-70 | **Low** |
| HPFS et al. 70-80 | **Low** |
| Ma et al. 2014 | **Low** |
| Maalouf et al. 2010 | **Low** |
| Mai et al. 2019 | **Low** |
| NHS et al. 40-50 | **Low** |
| NHS et al. 40-50 | **Low** |
| NHS et al. 50-60 | **Low** |
| NHS et al. 50-60 | **Low** |
| NHS et al. 60-70 | **Low** |
| NHS et al. 60-70 | **Low** |
| NHS et al. 70-80 | **Low** |
| NHS et al. 70-80 | **Low** |
| Parvin et al. 2011 | **Low** |
| Perinpam et al. 2015 | **Low** |
| Prochaska et al. 2018 | **Low** |
| Semins et al. 2010 | **Low** |
| Taylor et al. 2006 | **Low** |
| Thongprayoon et al. 2022 | **Low** |
| Traxer et al. 2003 | **Low** |
| Arrabal-Polo et al. 2015 | **Moderate** |
| Chai et al. 2004 | **Moderate** |
| Curhan et al. 2001 | **Moderate** |
| Duffey et al. 2011 | **Moderate** |
| Elkoushy et al. 2014 | **Moderate** |
| Gettman et al. | **Moderate** |
| Herrel et al. 2012 | **Moderate** |
| Holmes et al 2001 | **Moderate** |
| Hong et al. 2012 | **Moderate** |
| Kinder et al. 2002 | **Moderate** |
| Kuo et al. 2003 | **Moderate** |
| Lange et al. 2014 | **Moderate** |
| Lemann Jr et al. 1996 | **Moderate** |
| Lewandowski et al. 2004 | **Moderate** |
| Matin et al. 2000 | **Moderate** |
| Perez-Brayfield et al. 2002 | **Moderate** |
| Rodgers et al. 2002 | **Moderate** |
| Sorensen et al. 2011 | **Moderate** |
| Stitchantrakul et al. 2007 | **Moderate** |
| Wabner et al. 1992 | **Moderate** |
| Welshman et al 1976 | **Moderate** |
| Williams et al. 2001 | **Moderate** |
| Abate et al. 2004 | **High** |
| Allie et al. 2003 | **High** |
| Arora et al. 2017 | **High** |
| Arrabal-Polo et al. 2012 | **High** |
| Batailee et al. 1991 | **High** |
| Baxmann et al. 2003 | **High** |
| Buchholz et al. 1996 | **High** |
| Christie et al. 1996 | **High** |
| Ciacci et al. 2008 | **High** |
| Defoor et al. 2005 | **High** |
| Doenyas-Barak et al. 2017 | **High** |
| Erwin et al. 1994 | **High** |
| Goldschmied et al. 1975 | **High** |
| Goodzari et al. 2012 | **High** |
| Grases et al. 2015 | **High** |
| Hess et al. 1999 | **High** |
| Hunag et al. 2003 | **High** |
| Marya et al. 1979 | **High** |
| Matsumoto et al. 2006 | **High** |
| Michelacci et al. 1989 | **High** |
| Mikaye et al. 2001 | **High** |
| Mirazazadeh et al. 2011 | **High** |
| Nouvenne et al. 2013 | **High** |
| Ogawa et al. 2003 | **High** |
| Pais et al. 2007 | **High** |
| Pigna et al. 2013 | **High** |
| Roders et al. 2009 | **High** |
| Rodgers et al. 1997 | **High** |
| Shah et al. 2005 | **High** |
| Stern et al. 1980 | **High** |
| Stitchantrakul et al. 2004 | **High** |
| Sweeney et al. 2009 | **High** |
| Terris et al. 2001 | **High** |
| Welshmanet al. 1975 | **High** |

### 8 GRADE

#### 8.1 Function to Derive GRADE rating

```
calculate_grade <- function(study_design,
                           risk_of_bias,
                           inconsistency,
                           indirectness,
                           imprecision,
                           publication_bias,
                           large_effect = FALSE,
                           dose_response = FALSE,
                           confounders_reduce_effect = FALSE) {

  # Starting point based on study design
  if (study_design == "RCT") {
    certainty <- 4  # High
  } else if (study_design == "Observational" | study_design == "Mixed") {
    certainty <- 2  # Low
  } else {
    certainty <- 1  # Very Low
  }

  # Downgrade for limitations
  if (risk_of_bias == "Serious") certainty <- certainty - 1
  if (risk_of_bias == "Very serious") certainty <- certainty - 2

  if (inconsistency == "Serious") certainty <- certainty - 1
  if (inconsistency == "Very serious") certainty <- certainty - 2

  if (indirectness == "Serious") certainty <- certainty - 1
  if (indirectness == "Very serious") certainty <- certainty - 2

  if (imprecision == "Serious") certainty <- certainty - 1
  if (imprecision == "Very serious") certainty <- certainty - 2

  if (publication_bias == "Strongly suspected") certainty <- certainty - 1

  # Upgrade for observational studies
  if (study_design == "Observational" | study_design == "Mixed") {
    if (large_effect) certainty <- certainty + 1
    if (dose_response) certainty <- certainty + 1
    if (confounders_reduce_effect) certainty <- certainty + 1
  }

  # Ensure certainty stays within bounds
  certainty <- max(1, min(4, certainty))

  # Convert to GRADE rating
  grade_rating <- case_when(
    certainty == 4 ~ "High",
    certainty == 3 ~ "Moderate",
    certainty == 2 ~ "Low",
    certainty == 1 ~ "Very Low"
  )

  return(list(
    certainty_score = certainty,
    grade_rating = grade_rating
  ))
}

# Function to create GRADE quality symbols
grade_symbol <- function(grade_rating) {
  symbols <- case_when(
    grade_rating == "High" | grade_rating == 4 ~ "⊕⊕⊕⊕",
    grade_rating == "Moderate" | grade_rating == 3 ~ "⊕⊕⊕◯",
    grade_rating == "Low" | grade_rating == 2 ~ "⊕⊕◯◯",
    grade_rating == "Very Low" | grade_rating == 1 ~ "⊕◯◯◯",
    TRUE ~ "◯◯◯◯"
  )
  return(symbols)
}

# Helper for the GRADE `confounders_reduce_effect` upgrade.
# Returns TRUE when EITHER:
#   (a) meta-regression R² > 25% (substantial heterogeneity explained), OR
#   (b) meta-regression pooled estimate differs from overall pooled by ≥ 5%
#       (regression-adjusted result is meaningfully different from unadjusted)
# `ma_results` is the standard *_ma_2nd_analysis$ma_results data frame.
mr_reduces_effect <- function(ma_results,
                              R2_threshold      = 0.25,
                              shift_threshold   = 0.05) {
  ov <- ma_results %>%
    dplyr::filter(sex_group == "main meta-analysis",
                  age_band == "Overall",
                  quality_filter == "All Studies")
  mr <- ma_results %>%
    dplyr::filter(sex_group == "main meta-analysis",
                  age_band == "Meta-Regression",
                  quality_filter == "All Studies")

  if (!nrow(ov) || !nrow(mr)) return(FALSE)

  # Rule (a): high R² alone is sufficient
  R2 <- if ("R2" %in% names(mr)) mr$R2[1] else NA_real_
  if (!is.na(R2) && R2 > R2_threshold) return(TRUE)

  # Rule (b): MR pooled meaningfully different from overall pooled
  ov_est <- ov$pooled_estimate[1]
  mr_est <- mr$pooled_estimate[1]
  if (is.na(ov_est) || is.na(mr_est) || ov_est == 0) return(FALSE)
  rel_diff <- abs(mr_est - ov_est) / abs(ov_est)
  rel_diff >= shift_threshold
}
```

### 9 Finalised Results

#### 9.1 Functions

##### 9.1.1 Function to get final result

```
get_final_ma_results <- function(ma_results, sex_specific = FALSE, meta_regression = FALSE, low_rob = FALSE) {
  # Helper: apply strict filter; if 0 rows, fall back to relaxed filter that
  # drops the quality_filter constraint (handles cases where Meta-Regression
  # rows lack a quality_filter value, e.g. NA from bind_rows column propagation).
  strict_then_relaxed <- function(strict, relaxed) {
    res <- strict
    if (nrow(res) == 0) res <- relaxed
    res
  }

  if (sex_specific) {
    strict_then_relaxed(
      strict  = ma_results %>% filter(age_band == "Overall", sex_group != "main meta-analysis",
                                      quality_filter == "All Studies"),
      relaxed = ma_results %>% filter(age_band == "Overall", sex_group != "main meta-analysis")
    )
  } else if (meta_regression) {
    res <- strict_then_relaxed(
      strict  = ma_results %>% filter(age_band == "Meta-Regression", sex_group == "main meta-analysis",
                                      quality_filter == "All Studies"),
      relaxed = ma_results %>% filter(age_band == "Meta-Regression", sex_group == "main meta-analysis")
    )
    # Final safety net: if meta-regression row genuinely missing (helper
    # silently failed upstream), fall back to overall pooled so the chunk
    # produces output rather than crashing on a 0-row cbind.
    if (nrow(res) == 0) {
      message("get_final_ma_results: no Meta-Regression row available; falling back to Overall pooled")
      res <- ma_results %>% filter(sex_group == "main meta-analysis", age_band == "Overall",
                                   quality_filter == "All Studies")
    }
    res
  } else if (low_rob) {
    ma_results %>% filter(age_band == "Overall", sex_group == "main meta-analysis",
                          quality_filter == "Low Risk of Bias Only")
  } else {
    strict_then_relaxed(
      strict  = ma_results %>% filter(sex_group == "main meta-analysis", age_band == "Overall",
                                      quality_filter == "All Studies"),
      relaxed = ma_results %>% filter(sex_group == "main meta-analysis", age_band == "Overall")
    )
  }
}
```

#### 9.2 Recommendations Summary

This summary applies the decision framework defined earlier (see *Flow Diagram detailing Decision Making*) to each urine component using the per-component meta-analysis objects (*_ma_2nd_analysis and *_ma_3rd_analysis). It is placed here, at the top of the Finalised Results, so the reader can see **which outcome was selected for which component and why** before reading the per-component conclusions below.

```
# Pull together rows from the *_ma_2nd_analysis$ma_results outputs
all_ma_long <- purrr::map_dfr(analysis_components, function(comp) {
  obj_name <- paste0(comp, "_ma_2nd_analysis")
  if (!exists(obj_name)) return(tibble::tibble())
  obj <- get(obj_name)
  if (is.null(obj$ma_results)) return(tibble::tibble())
  res <- obj$ma_results
  if (!"urine_component" %in% names(res)) res$urine_component <- comp
  if (!"quality_filter" %in% names(res))  res$quality_filter  <- "All Studies"
  res
})

# Guard: if no per-component objects are loaded yet, render a placeholder
if (nrow(all_ma_long) == 0) {
  gt::gt(tibble::tibble(
    Message = "No *_ma_2nd_analysis objects found — re-run all per-component sections first."
  ))
} else {
  all_ma_long <- all_ma_long %>%
    dplyr::mutate(age_band  = tolower(as.character(age_band)),
                  sex_group = tolower(as.character(sex_group)))

  # Build race-stratified long table from *_ma_3rd_analysis (for Q4)
  all_ma_race_long <- purrr::map_dfr(analysis_components, function(comp) {
    obj_name <- paste0(comp, "_ma_3rd_analysis")
    if (!exists(obj_name)) return(tibble::tibble())
    obj <- get(obj_name)
    if (is.null(obj$ma_results)) return(tibble::tibble())
    res <- obj$ma_results
    if (!"urine_component" %in% names(res)) res$urine_component <- comp
    res
  })

  # Build long-format meta-regression coefficient table from the global
  # `meta_reg_full_results` cache populated by meta_regression(). Used by
  # Bridge 1 (women_prop_c coefficient) and Bridge 4 (R² for Q6).
  all_meta_reg_long <- if (exists("meta_reg_full_results") &&
                            length(meta_reg_full_results) > 0) {
    purrr::map_dfr(names(meta_reg_full_results), function(key) {
      tbl <- meta_reg_full_results[[key]]
      if (is.null(tbl) || !nrow(tbl)) return(tibble::tibble())
      parts <- strsplit(key, "__", fixed = TRUE)[[1]]
      tbl %>% dplyr::mutate(urine_component = parts[1],
                             sex_group       = parts[2])
    })
  } else NULL

  # Apply the integrated framework
  recommendations <- purrr::map_dfr(analysis_components, function(comp) {
    tryCatch(
      recommend_analysis(comp,
                         ma_long       = all_ma_long,
                         ma_race_long  = all_ma_race_long,
                         data          = urine_ma2,
                         meta_reg_long = all_meta_reg_long),
      error = function(e) tibble::tibble(component = comp,
                                          outcome  = "ERROR",
                                          branch   = NA_character_,
                                          reason   = e$message)
    )
  })

  recommendations %>%
    dplyr::mutate(component = stringr::str_to_title(component),
                  R2 = ifelse(is.na(R2), "—", sprintf("%.1f%%", R2 * 100)),
                  k  = ifelse(is.na(k), "—", as.character(k)),
                  n  = ifelse(is.na(n), "—", format(n, big.mark = ","))) %>%
    dplyr::rowwise() %>%
    dplyr::mutate(
      `Sex p (women_prop / overall / MR / LRoB)` =
        paste(format_p(p_women_prop %||% NA_real_),
              format_p(p_sex_overall),
              format_p(p_sex_metareg),
              format_p(p_sex_lowrob), sep = " / "),
      `Age p (min)`   = format_p(p_age),
      `race p (min)`  = format_p(p_race),
      `Assay p (min)` = format_p(p_assay),
      `Egger's p`     = format_p(eggers_p %||% NA_real_),
      # Quality-gate display: Cochran's Q normalised by df, MR R² for chosen subset
      `Q/df (chosen / overall)` = paste0(
        ifelse(is.na(qc_q_ratio_chosen),  "—", sprintf("%.1f", qc_q_ratio_chosen)),
        " / ",
        ifelse(is.na(qc_q_ratio_overall), "—", sprintf("%.1f", qc_q_ratio_overall))
      ),
      `Q/df ratio (chosen÷overall)` = ifelse(is.na(qc_q_ratio_rel), "—",
                                              sprintf("%.2f", qc_q_ratio_rel)),
      `Q p (chosen)` = format_p(qc_q_pval_chosen),
      `MR R² (chosen)` = ifelse(is.na(qc_mr_R2_chosen), "—",
                                 sprintf("%.0f%%", qc_mr_R2_chosen * 100)),
      `QC warnings` = ifelse(qc_warnings == "" | is.na(qc_warnings), "—", qc_warnings)
    ) %>%
    dplyr::ungroup() %>%
    dplyr::select(Component = component,
                  k, n, `R²` = R2,
                  `Sex p (women_prop / overall / MR / LRoB)`,
                  `Age p (min)`,
                  `race p (min)`,
                  `Assay p (min)`,
                  `Egger's p`,
                  `Q/df (chosen / overall)`,
                  `Q/df ratio (chosen÷overall)`,
                  `Q p (chosen)`,
                  `MR R² (chosen)`,
                  `Recommended Analysis` = outcome,
                  Branch  = branch,
                  Rationale = reason,
                  `QC warnings`) %>%
    gt::gt() %>%
    gt::tab_header(
      title    = gt::md("**Decision-Framework Recommendations per Urine Component**"),
      subtitle = gt::md(sprintf(
        "Q1 data → Q2 sex → Q3 age → Q4 race → Q5 assay → Q6 meta-reg (R²) → Q6b meta-reg (Egger's) → Q6c meta-reg (no stratum effect + valid range) → Q7 low-RoB → Q8 overall pooled. Thresholds: k≥%d, n≥%d, R²≥%.0f%% (Q6) or ≥%.0f%% (Q6c) & k≥%d, Wald p<%.2f, Egger p<%.2f, consistency ≤%.0f%%.",
        THRESHOLDS$k_min, THRESHOLDS$n_min,
        THRESHOLDS$R2_min * 100, THRESHOLDS$R2_q6c_min * 100, THRESHOLDS$R2_k_min, THRESHOLDS$wald_alpha,
        THRESHOLDS$eggers_alpha,
        THRESHOLDS$consistency_pct * 100))
    ) %>%
    gt::data_color(
      columns = `Recommended Analysis`,
      fn = function(x) {
        dplyr::case_when(
          x == "Sex-stratified"             ~ "#cce5ff",
          x == "Age-stratified"             ~ "#cff4fc",
          x == "race-stratified"            ~ "#d1e7dd",
          x == "Assay/country-stratified"   ~ "#cfe2ff",
          x == "Meta-regression"            ~ "#d4edda",
          x == "Low risk-of-bias overall"   ~ "#e2d4f0",
          x == "Overall pooled (caveats)"   ~ "#f8d7da",
          x == "Overall pooled"             ~ "#ffe5d0",
          TRUE                               ~ "#f0f0f0"
        )
      }
    ) %>%
    gt::tab_options(table.font.size = 10, data_row.padding = gt::px(4))
}
```

Table 1: **Decision-Framework Recommendations per Urine Component**

Q1 data → Q2 sex → Q3 age → Q4 race → Q5 assay → Q6 meta-reg (R²) → Q6b meta-reg (Egger's) → Q6c meta-reg (no stratum effect + valid range) → Q7 low-RoB → Q8 overall pooled. Thresholds: k≥5, n≥500, R²≥10% (Q6) or ≥10% (Q6c) & k≥20, Wald p<0.05, Egger p<0.10, consistency ≤20%.

| Component | k | n | R² | Sex p (women_prop / overall / MR / LRoB) | Age p (min) | race p (min) | Assay p (min) | Egger's p | Q/df (chosen / overall) | Q/df ratio (chosen÷overall) | Q p (chosen) | MR R² (chosen) | Recommended Analysis | Branch | Rationale | QC warnings |
| --- | --- | --- | --- | --- | --- | --- | --- | --- | --- | --- | --- | --- | --- | --- | --- | --- |
| Volume | 89 | 9,144 | 7.6% | 0.438 / 0.937 / — / 0.970 | 0.840 | 0.762 | — | 0.811 | 104.1 / 104.1 | 1.00 | <0.001 | 8% | Overall pooled | 8 — Default pooled | No branch criteria met; default to overall pooled with heterogeneity caveat | — |
| Ph | 95 | 9,226 | 11.6% | 0.279 / 0.857 / 0.753 / 0.687 | 0.484 | 0.835 | — | 0.512 | 6.4 / 26.5 | 0.24 | <0.001 | 12% | Meta-regression | 6 — Meta-regression effective | R² = 11.6% ≥ 10% with k = 95 ≥ 20 | — |
| Calcium | 100 | 10,284 | 29.4% | 0.075 / 0.675 / 0.601 / 0.911 | 0.409 | 0.974 | 0.535 | 0.093 | 47.3 / 214.0 | 0.22 | <0.001 | 29% | Meta-regression | 6 — Meta-regression effective | R² = 29.4% ≥ 10% with k = 100 ≥ 20 | — |
| Oxalate | 92 | 9,725 | 38.3% | 0.012 / 0.541 / 0.238 / 0.687 | — | — | — | 0.020 | 117.0 / 176.1 | 0.66 | <0.001 | 0% | Sex-stratified | 2 — Robust sex difference | women_prop coef p=0.012 in main meta-regression | — |
| Urate | 93 | 9,544 | 5.9% | 0.042 / 0.436 / 0.461 / 0.592 | — | — | — | 0.443 | 69.8 / 40.9 | 1.71 | <0.001 | 51% | Sex-stratified | 2 — Robust sex difference | women_prop coef p=0.042 in main meta-regression | Chosen Q/df (69.8) is 171% of overall Q/df (40.9) — exceeds limit of 150% |
| Citrate | 97 | 9,458 | 17.5% | 0.363 / 0.629 / 0.941 / 0.951 | 0.845 | 0.654 | <0.001 | 0.148 | — / 176.5 | — | — | 18% | Assay/country-stratified | 5 — Robust assay/country difference | Assay min Wald p = <0.001 (rel shift 44.8%) across 3 named-category pair(s); 2/3 significant after 20% shift filter | — |
| Creat | 74 | 9,058 | 82.0% | <0.001 / 0.012 / <0.001 / 0.272 | — | — | — | <0.001 | 46.8 / 1651.9 | 0.03 | <0.001 | — | Sex-stratified | 2 — Robust sex difference | women_prop coef p=<0.001 in main meta-regression; sex p<0.05 in 2 of 3 tiers (overall=0.012, MR=<0.001, LRoB=0.272) | — |
| Phosphate | 56 | 8,104 | 76.8% | 0.067 / 0.338 / 0.701 / 0.756 | 0.393 | 0.165 | 0.076 | 0.001 | 4.7 / 210.6 | 0.02 | <0.001 | 77% | Meta-regression | 6 — Meta-regression effective | R² = 76.8% ≥ 10% with k = 56 ≥ 20 | — |
| Sodium | 94 | 9,120 | 11.5% | 0.657 / 0.599 / 0.400 / 0.871 | 0.716 | 0.815 | 0.002 | 0.219 | 18.6 / 85.1 | 0.22 | <0.001 | 11% | Meta-regression | 6 — Meta-regression effective | R² = 11.5% ≥ 10% with k = 94 ≥ 20 | — |
| Potassium | 93 | 9,418 | 22.2% | 0.205 / 0.695 / 0.702 / 0.748 | 0.587 | 0.594 | 0.010 | 0.744 | — / 83.1 | — | — | 22% | Assay/country-stratified | 5 — Robust assay/country difference | Assay min Wald p = 0.010 (rel shift 44.9%) across 3 named-category pair(s); 1/3 significant after 20% shift filter | — |
| Magnesium | 87 | 9,513 | 17.3% | 0.421 / 0.585 / 0.940 / 0.982 | 0.571 | 0.793 | 0.363 | 0.359 | 6.4 / 111.6 | 0.06 | <0.001 | 17% | Meta-regression | 6 — Meta-regression effective | R² = 17.3% ≥ 10% with k = 87 ≥ 20 | — |
| Ammonium | 45 | 4,841 | 45.8% | 0.035 / 0.285 / — / — | — | — | — | 0.081 | 53.2 / 62.9 | 0.85 | <0.001 | — | Sex-stratified | 2 — Robust sex difference | women_prop coef p=0.035 in main meta-regression | — |
| Chloride | 51 | 3,534 | 29.7% | 0.059 / 0.646 / 0.612 / 0.888 | — | — | — | 0.529 | 15.8 / 104.0 | 0.15 | <0.001 | 30% | Meta-regression | 6 — Meta-regression effective | R² = 29.7% ≥ 10% with k = 51 ≥ 20 | — |
| Sulphate | 41 | 4,857 | 95.1% | <0.001 / 0.638 / — / — | — | — | — | 0.415 | 265.0 / 362.4 | 0.73 | <0.001 | — | Sex-stratified | 2 — Robust sex difference | women_prop coef p=<0.001 in main meta-regression | — |
| Urea | 13 | 3,876 | 100.0% | 0.110 / 0.663 / — / — | — | — | — | 0.278 | 255.1 / 255.1 | 1.00 | <0.001 | 100% | Overall pooled | 8 — Default pooled | Meta-regression range 192.12–543.57 valid but k = 13 < 20 (model saturated); default to overall pooled | — |

##### 9.2.1 Framework Recommendation Helper

```
# Render a one-row "Framework Recommendation" callout for a single component.
# Pulls from the global `recommendations` tibble built above.
framework_callout <- function(component_code) {
  if (!exists("recommendations") || !inherits(recommendations, "data.frame") ||
      nrow(recommendations) == 0) {
    return(gt::gt(tibble::tibble(
      Message = "Framework not yet computed — see Recommendations Summary."
    )))
  }
  rec <- recommendations %>%
    dplyr::filter(tolower(component) == tolower(component_code))
  if (nrow(rec) == 0) {
    return(gt::gt(tibble::tibble(
      Message = sprintf("No framework recommendation for %s", component_code)
    )))
  }
  rec %>%
    dplyr::transmute(
      Component   = component,
      Recommended = outcome,
      Branch      = branch,
      Reason      = reason,
      k = ifelse(is.na(k), "—", as.character(k)),
      n = ifelse(is.na(n), "—", format(n, big.mark = ",")),
      `R²` = ifelse(is.na(R2), "—", sprintf("%.1f%%", R2 * 100)),
      `Sex p (women_prop / overall / MR / LRoB)` =
        paste(format_p(p_women_prop %||% NA_real_),
              format_p(p_sex_overall),
              format_p(p_sex_metareg),
              format_p(p_sex_lowrob), sep = " / "),
      `Age p (min)`   = format_p(p_age),
      `race p (min)`  = format_p(p_race),
      `Assay p (min)` = format_p(p_assay),
      `Egger's p`     = format_p(eggers_p %||% NA_real_),
      `Q/df (chosen / overall)` = paste0(
        ifelse(is.na(qc_q_ratio_chosen),  "—", sprintf("%.1f", qc_q_ratio_chosen)),
        " / ",
        ifelse(is.na(qc_q_ratio_overall), "—", sprintf("%.1f", qc_q_ratio_overall))
      ),
      `Q/df ratio (chosen÷overall)` = ifelse(is.na(qc_q_ratio_rel), "—",
                                              sprintf("%.2f", qc_q_ratio_rel)),
      `Q p (chosen)` = format_p(qc_q_pval_chosen),
      `MR R² (chosen)` = ifelse(is.na(qc_mr_R2_chosen), "—",
                                 sprintf("%.0f%%", qc_mr_R2_chosen * 100)),
      `QC warnings` = ifelse(qc_warnings == "" | is.na(qc_warnings),
                             "—", qc_warnings)
    ) %>%
    gt::gt() %>%
    gt::tab_header(
      title    = gt::md(sprintf("**Framework Recommendation — %s**", rec$component)),
      subtitle = gt::md(sprintf("Branch %s", rec$branch))
    ) %>%
    gt::data_color(
      columns = Recommended,
      fn = function(x) {
        dplyr::case_when(
          x == "Sex-stratified"             ~ "#cce5ff",
          x == "Age-stratified"             ~ "#cff4fc",
          x == "race-stratified"            ~ "#d1e7dd",
          x == "Assay/country-stratified"   ~ "#cfe2ff",
          x == "Meta-regression"            ~ "#d4edda",
          x == "Low risk-of-bias overall"   ~ "#e2d4f0",
          x == "Overall pooled (caveats)"   ~ "#f8d7da",
          x == "Overall pooled"             ~ "#ffe5d0",
          TRUE                               ~ "#f0f0f0"
        )
      }
    ) %>%
    gt::cols_align(align = "center", columns = -c(Component, Reason)) %>%
    gt::tab_options(table.font.size = 11, data_row.padding = gt::px(4))
}
```

#### 9.3 Volume

##### 9.3.1 Framework Recommendation

framework_callout("volume")

Table 1: **Framework Recommendation — volume**

Branch 8 — Default pooled

| Component | Recommended | Branch | Reason | k | n | R² | Sex p (women_prop / overall / MR / LRoB) | Age p (min) | race p (min) | Assay p (min) | Egger's p | Q/df (chosen / overall) | Q/df ratio (chosen÷overall) | Q p (chosen) | MR R² (chosen) | QC warnings |
| --- | --- | --- | --- | --- | --- | --- | --- | --- | --- | --- | --- | --- | --- | --- | --- | --- |
| volume | Overall pooled | 8 — Default pooled | No branch criteria met; default to overall pooled with heterogeneity caveat | 89 | 9,144 | 7.6% | 0.438 / 0.937 / — / 0.970 | 0.840 | 0.762 | — | 0.811 | 104.1 / 104.1 | 1.00 | <0.001 | 8% | — |

##### 9.3.2 Comment

For 24-hour urine volume, the meta-analysis included k = 89 subgroup-level records and n = 9,144 participants. Heterogeneity was extreme throughout (I² ≈ 99%, Cochran’s Q p < 0.001) and the dataset comfortably met the framework’s data-sufficiency floor (k ≥ 5, n ≥ 500).

Stratum tests were uniformly non-significant. Sex differences were not significant on meta-regression (p = 0.438) or in the overall sex Wald test (p = 0.937). Age comparisons across powered bands gave a minimum pairwise Wald p of 0.840. race was severely underpowered (minimum Wald p = 0.762), with several strata being single studies of implausibly wide range; the one true pairwise between-race contrast that did reach significance was Black women (1.40 – 1.75 L/24h, n = 304, I² = 60%) vs White women (1.76 – 2.05 L/24h, n = 3,260, I² = 78%), while a further apparent contrast between the all-race pooled group (0.94 – 2.40, n = 4,177) and White women is a comparison between a pooled group and one of its constituent strata and should not be interpreted as a true between-group difference. Assay/country stratification was not tested for volume.

Meta-regression accounted for only R² = 7.6% of heterogeneity (k = 89), which is below the framework’s 10% threshold, so the meta-regression branch did not fire. Egger’s test was not significant (p = 0.811), ruling out the small-study-bias override; and the no-stratum-effect override was not triggered because the meta-regression-adjusted result did not differ materially from the unadjusted overall.

The low risk-of-bias overall sub-analysis was consistent with the main result, as were the RCT-only sub-analyses (the latter were limited by small sample sizes but agreed on the ~1 L lower bound). No sensitivity analysis suggested a more conservative or more informative estimate.

The framework therefore defaulted to the unadjusted overall pooled meta-analysis. The recommended expected range is **1.0 – 2.5 L/24h** (Overall, All Studies, n = 9,144). The dominant caveat is the extreme between-study heterogeneity, which is largely unexplained by any tested moderator; a women-only meta-regression shows a marked decrease in the lower limit (from 0.94 L/24h in the main MA to 0.55 L/24h), suggesting that female-only datasets may have systematically different lower bounds.

##### 9.3.3 Plot

```
volume_ma_2nd_analysis$main_meta_analysis$plot %+%
  subset(volume_ma_2nd_analysis$main_meta_analysis$plot$data,
         age_band %in% ("Overall"))
## `height` was translated to `width`.
```

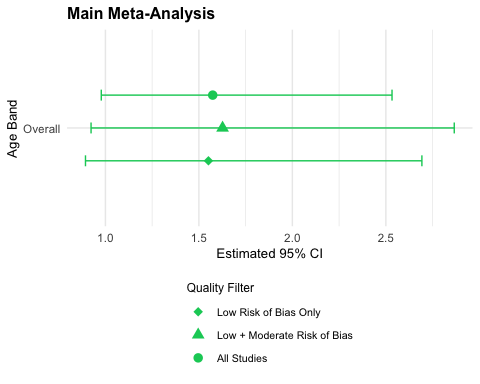

##### 9.3.4 Results Table

```
volume_ma_2nd_analysis$ma_results %>%
  filter(
    sex_group == "main meta-analysis",
    age_band %in% ("Overall")
  ) %>%
  mutate(
    pooled_estimate = round(pooled_estimate, 2),
    pooled_sd       = round(pooled_sd, 2),
    I2              = round(I2, 2),
    Q               = round(Q, 2),
    Q_pval = case_when(
      Q_pval < 0.001 ~ "<0.001",
      Q_pval < 0.01  ~ "<0.01",
      Q_pval < 0.05  ~ "<0.05",
      TRUE           ~ as.character(round(Q_pval, 2))
    ),
    ci_95 = paste0(
      round(lower_95, 1), " – ", round(upper_95, 1)),
    Quality = quality_filter
  ) %>%
  select(
    Quality,
    total_participants,
    pooled_estimate,
    pooled_sd,
    ci_95,
    I2,
    Q,
    Q_pval
  ) %>%
  gt() %>%
  cols_label(
    Quality           = "Study Quality",
    total_participants  = "Patients, n",
    pooled_estimate = "Pooled Estimate",
    pooled_sd       = "SD",
    ci_95           = "95% CI",
    I2              = html("I<sup>2</sup> (%)"),
    Q               = "Q statistic",
    Q_pval          = "Q p-value"
  ) %>%
  cols_align(
    align = "center",
    columns = c(total_participants, pooled_estimate, pooled_sd, ci_95, I2, Q, Q_pval)
  ) %>%
  tab_header(
    title = "Meta-analysis Results by Age Band",
    subtitle = "Random-effects meta-analysis"
  ) %>%
  tab_source_note(
    source_note = md(
      "*I² quantifies heterogeneity; Q p-value tests heterogeneity across studies.*"
    )
  )
```

Table 1: Meta-analysis Results by Age Band

Random-effects meta-analysis

| Study Quality | Patients, n | Pooled Estimate | SD | 95% CI | I2 (%) | Q statistic | Q p-value |
| --- | --- | --- | --- | --- | --- | --- | --- |
| All Studies | 9144 | 1.57 | 0.24 | 1 – 2.5 | 0.99 | 9158.78 | <0.001 |
| Low + Moderate Risk of Bias | 3648 | 1.63 | 0.29 | 0.9 – 2.9 | 0.99 | 2591.19 | <0.001 |
| Low Risk of Bias Only | 3288 | 1.55 | 0.28 | 0.9 – 2.7 | 0.98 | 1057.54 | <0.001 |
| *I² quantifies heterogeneity; Q p-value tests heterogeneity across studies.* | | | | | | | |

##### 9.3.5 Bias analysis

```
volume_rob <- urine_ma2 %>%
  mutate(
    women_prop = dplyr::case_when(
          male_female == "female" ~ 1,
          male_female == "male" ~ 0,
          male_female == "mixed" & !is.na(female_n) ~ female_n / total_n,
          male_female == "mixed" & is.na(female_n) ~ 0.5,
          TRUE ~ NA_real_
        )
  ) %>%
  select(study_name, age_band, study_type, volume_sd_m_l, women_prop, race, total_n) %>%
  drop_na(volume_sd_m_l, women_prop) %>%
  left_join(
    overall_bias, by = "study_name"
  ) %>%
  select(study_name, overall, study_type) %>%
  mutate(
    overall = case_when(
      is.na(overall) ~ "Moderate",
      TRUE ~ overall
    )
  ) %>%
  distinct(study_name, .keep_all = TRUE) %>%
  arrange(overall) %>%
  gt() %>%
  cols_label(
    study_name = "Study",
    overall    = "Risk of Bias",
    study_type = "Study Type"
  ) %>%
  data_color(
    columns = overall,
    fn = function(x) {
      case_when(
        x == "Low"      ~ "#90EE90",
        x == "Moderate" ~ "#FFD700",
        x == "High"     ~ "#FF6B6B",
        TRUE            ~ "#FFFFFF"
      )
    }
  ) %>%
  opt_all_caps() %>%
  cols_align(
    align = "center",
    overall
  ) %>%
  tab_header(
    title = "Risk of Bias by Study and Age Band for 24 Hour Urine Volume"
  )

volume_rob
```

Table 1: Risk of Bias by Study and Age Band for 24 Hour Urine Volume

| Study | Risk of Bias | Study Type |
| --- | --- | --- |
| Arora et al. 2017 | High | Observational Cohort |
| Grases et al. 2015 | High | Observational Cohort |
| Nouvenne et al. 2013 | High | Observational Cohort |
| Arrabal-Polo et al. 2012 | High | Observational Cohort |
| Sweeney et al. 2009 | High | RCT |
| Matsumoto et al. 2006 | High | RCT |
| Abate et al. 2004 | High | Observational Cohort |
| Stitchantrakul et al. 2004 | High | RCT |
| Baxmann et al. 2003 | High | RCT |
| Hess et al. 1999 | High | Observational Cohort |
| Rodgers et al. 1997 | High | Observational Cohort |
| Christie et al. 1996 | High | Observational Cohort |
| Erwin et al. 1994 | High | Observational Cohort |
| Stern et al. 1980 | High | Observational Cohort |
| Thongprayoon et al. 2022 | Low | Observational Cohort |
| Deng et al. 2017 | Low | Observational Cohort |
| Parvin et al. 2011 | Low | Observational Cohort |
| Maalouf et al. 2010 | Low | Observational Cohort |
| Traxer et al. 2003 | Low | RCT |
| Mai et al. 2019 | Low | Observational Cohort |
| Doizi et al. 2016 | Low | Observational Cohort |
| Elkoushy et al. 2014 | Moderate | Observational Cohort |
| Lange et al. 2014 | Moderate | RCT |
| Pigna et al. 2014 | Moderate | Observational Cohort |
| Goodarzi et al. 2012 | Moderate | Observational Cohort |
| Herrel et al. 2012 | Moderate | RCT |
| Hong et al. 2012 | Moderate | Observational Cohort |
| Sorensen et al. 2012 | Moderate | Observational Cohort |
| Duffey et al. 2011 | Moderate | Observational Cohort |
| Stitchantrakul et al. 2007 | Moderate | Observational Cohort |
| Cameron et al. 2006 | Moderate | Observational Cohort |
| Gettman et al. 2005 | Moderate | Observational Cohort |
| Kuo et al. 2003 | Moderate | Observational Cohort |
| Holmes et al. 2001 | Moderate | Observational Cohort |
| Miyake et al. 2001 | Moderate | Observational Cohort |
| Lemann et al. 1996 | Moderate | Observational Cohort |
| Wabner et al. 1992 | Moderate | RCT |
| Matin and Streem, 2000 | Moderate | Observational Cohort |
| Baric et al. 2019 | Moderate | Observational Cohort |
| Rodgers et al. 2015 | Moderate | Observational Cohort |
| Hesse et al. 1986 | Moderate | Observational Cohort |
| Athanasatou et al. 2018 | Moderate | Observational Cohort |
| HPFS et al. 40-50 | Moderate | Observational Cohort |
| HPFS et al. 50-60 | Moderate | Observational Cohort |
| HPFS et al. 60-70 | Moderate | Observational Cohort |
| HPFS et al. 70-80 | Moderate | Observational Cohort |
| NHS et al. 40-50 | Moderate | Observational Cohort |
| NHS et al. 50-60 | Moderate | Observational Cohort |
| NHS et al. 60-70 | Moderate | Observational Cohort |
| NHS et al. 70-80 | Moderate | Observational Cohort |

##### 9.3.6 24 Hour Urine Volume Suggested Expected Range inc. GRADE

```
volume_grade<- calculate_grade(
  study_design= "RCT", #RCT results seem comparable with overall analysis
  risk_of_bias = "Not Serious", #Low risk of bias sub analysis consistent with main result
  inconsistency = "Moderate", #Deviation across analyses esp on upper limit
  indirectness = "Not Serious", #Populations examined are appropriate, assay type not applicable in this scenario
  imprecision = "Not Serious", #++sample size - precision ok
  publication_bias = "Suspected", #Large cochran's Q + I2
  large_effect = TRUE,
  dose_response = FALSE, # Not applicable
  confounders_reduce_effect = mr_reduces_effect(volume_ma_2nd_analysis$ma_results)
)

volume_grade <- cbind(
  sex = c("Overall"),
  grade = volume_grade$grade_rating
) %>% as_tibble() %>%
  mutate(
    grade_rating = grade_symbol(grade)
  )

volume_final_results <- get_final_ma_results(volume_ma_2nd_analysis$ma_results, sex_specific = FALSE, meta_regression = FALSE, low_rob = FALSE) %>%
  mutate(
    pooled_estimate = round(pooled_estimate, 2),
    pooled_sd       = round(pooled_sd, 2),
    Q               = round(Q, 2),
    Q_pval = case_when(
      Q_pval < 0.001 ~ "<0.001",
      Q_pval < 0.01  ~ "<0.01",
      Q_pval < 0.05  ~ "<0.05",
      TRUE           ~ sprintf("%.2f", Q_pval)
    ),
    ci_95 = paste0(
      round(lower_95, 1), " – ", round(upper_95, 1)
    )
  ) %>%
  select(
    age_band,
    total_participants,
    ci_95,
    Q,
    Q_pval
  ) %>%
  cbind(
    volume_grade) %>%
  mutate(grade_rating = paste0(grade, " ", grade_rating)) %>%
  select(-c(sex,grade))

volume_final_results %>%
  gt() %>%
  cols_label(
    age_band       = "Analysis",
    total_participants  = "Patients, n",
    ci_95           = "95% CI",
    Q               = "Q statistic",
    Q_pval          = "Q p-value",
    grade_rating    = "GRADE Rating"
  ) %>%
  cols_align(
    align = "center",
    -age_band
  ) %>%
  tab_header(
    title = "Finalised Meta-Analysis Results for Volume",
    subtitle = "Subdivided by Age Band"
  ) %>%
  tab_source_note(
    source_note = md(
      "*Q p-value = Cochran's Q heterogeneity test; RoB = Risk of Bias*"
    )
  )
```

Table 1: Finalised Meta-Analysis Results for Volume

Subdivided by Age Band

| Analysis | Patients, n | 95% CI | Q statistic | Q p-value | GRADE Rating |
| --- | --- | --- | --- | --- | --- |
| Overall | 9144 | 1 – 2.5 | 9158.78 | <0.001 | High ⊕⊕⊕⊕ |
| *Q p-value = Cochran's Q heterogeneity test; RoB = Risk of Bias* | | | | | |

```
volume_final_results <- volume_final_results %>%
  mutate(
    urine_component = "volume"
  )
```

#### 9.4 Calcium

##### 9.4.1 Framework Recommendation

framework_callout("calcium")

Table 1: **Framework Recommendation — calcium**

Branch 6 — Meta-regression effective

| Component | Recommended | Branch | Reason | k | n | R² | Sex p (women_prop / overall / MR / LRoB) | Age p (min) | race p (min) | Assay p (min) | Egger's p | Q/df (chosen / overall) | Q/df ratio (chosen÷overall) | Q p (chosen) | MR R² (chosen) | QC warnings |
| --- | --- | --- | --- | --- | --- | --- | --- | --- | --- | --- | --- | --- | --- | --- | --- | --- |
| calcium | Meta-regression | 6 — Meta-regression effective | R² = 29.4% ≥ 10% with k = 100 ≥ 20 | 100 | 10,284 | 29.4% | 0.075 / 0.675 / 0.601 / 0.911 | 0.409 | 0.974 | 0.535 | 0.093 | 47.3 / 214.0 | 0.22 | <0.001 | 29% | — |

##### 9.4.2 Comment

For 24-hour urine calcium, the meta-analysis included k = 100 subgroup-level records and n = 10,284 participants — the largest dataset of any analyte. Heterogeneity was extreme throughout (I² > 99%, Cochran’s Q p < 0.001).

Sex differences on meta-regression approached but did not reach significance (p = 0.075; overall sex Wald p = 0.675). Age comparisons across powered bands gave a minimum pairwise Wald p of 0.409. race was severely underpowered (minimum Wald p = 0.974), with most strata being single studies and some producing negative values. The assay/country stratum test was not significant (minimum Wald p = 0.535). Several stratified analyses, notably the RCT subgroup, showed tighter intervals than the main result but had substantially fewer patients and are unlikely to be representative of the wider population.

Meta-regression explained R² = 29.4% of heterogeneity with k = 100, well above the framework’s 10% threshold, so the meta-regression branch was satisfied. Egger’s test was borderline (p = 0.093), just above the 0.10 small-study-bias threshold; the small-study-bias override was therefore not invoked but the suggestion of small-study bias is noted.

The low risk-of-bias overall sub-analysis was consistent with the main meta-analysis, and the RCT-only sub-analysis (with substantially smaller n) gave a tighter interval that sat within the meta-regression range. Sensitivity analyses supported the meta-regression choice.

The framework selected the meta-regression result. The recommended expected range is **1.9 – 6.2 mmol/24h** (Meta-regression, n = 5,418). Substantial residual heterogeneity remains even after meta-regression adjustment.

##### 9.4.3 Plot

```
calcium_ma_2nd_analysis$main_meta_analysis$plot %+%
  subset(calcium_ma_2nd_analysis$main_meta_analysis$plot$data,
         age_band %in% ("Meta-Regression"))
## `height` was translated to `width`.
```

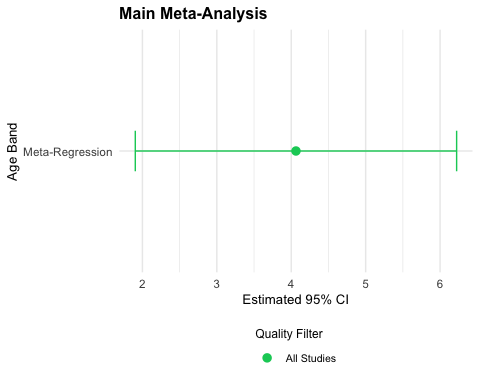

##### 9.4.4 Results Table

```
calcium_ma_2nd_analysis$ma_results %>%
  filter(
    age_band %in% c("Meta-Regression")
  ) %>% filter(
    sex_group == "main meta-analysis"
  ) %>%
  mutate(
    pooled_estimate = round(pooled_estimate, 2),
    pooled_sd       = round(pooled_sd, 2),
    I2              = round((I2 * 100), 0),
    Q               = round(Q, 2),
    Q_pval = case_when(
      Q_pval < 0.001 ~ "<0.001",
      Q_pval < 0.01  ~ "<0.01",
      Q_pval < 0.05  ~ "<0.05",
      TRUE           ~ as.character(round(Q_pval, 2))
    ),
    ci_95 = paste0(
      round(lower_95, 1), " – ", round(upper_95, 1)
    ),
    sex_group = str_to_title(age_band)
  ) %>%
  select(
    sex_group,
    total_participants,
    pooled_estimate,
    pooled_sd,
    ci_95,
    I2,
    Q,
    Q_pval
  ) %>%
  gt() %>%
  cols_label(
    sex_group = "Analysis",
    total_participants  = "Patients, n",
    pooled_estimate = "Pooled Estimate",
    pooled_sd       = "SD",
    ci_95           = "95% CI",
    I2              = html("I<sup>2</sup> (%)"),
    Q               = "Q statistic",
    Q_pval          = "Q p-value"
  ) %>%
  cols_align(
    align = "center",
    columns = c(total_participants, pooled_estimate, pooled_sd, ci_95, I2, Q, Q_pval)
  ) %>%
  tab_header(
    title = "Calcium Meta-analysis Results by Age Band",
    subtitle = "Random-effects meta-analysis"
  ) %>%
  tab_source_note(
    source_note = md(
      "*I² quantifies heterogeneity; Q p-value tests heterogeneity across studies.*"
    )
  )
```

Table 1: Calcium Meta-analysis Results by Age Band

Random-effects meta-analysis

| Analysis | Patients, n | Pooled Estimate | SD | 95% CI | I2 (%) | Q statistic | Q p-value |
| --- | --- | --- | --- | --- | --- | --- | --- |
| Meta-Regression | 5418 | 4.06 | 1.1 | 1.9 – 6.2 | 98 | 2744.79 | <0.001 |
| *I² quantifies heterogeneity; Q p-value tests heterogeneity across studies.* | | | | | | | |

##### 9.4.5 Bias analysis

```
calcium_rob <- urine_ma2 %>%
  drop_na(calcium_sd_mmol_24h) %>%
  left_join(
    overall_bias, by = "study_name"
  ) %>%
  select(study_name, overall, study_type) %>%
  mutate(
    overall = case_when(
      is.na(overall) ~ "Moderate",
      TRUE ~ overall
    )
  ) %>%
  distinct(study_name, .keep_all = TRUE) %>%
  arrange(overall) %>%
  gt() %>%
  cols_label(
    study_name = "Study",
    overall    = "Risk of Bias",
    study_type = "Study Type"
  ) %>%
  data_color(
    columns = overall,
    fn = function(x) {
      case_when(
        x == "Low"      ~ "#90EE90",
        x == "Moderate" ~ "#FFD700",
        x == "High"     ~ "#FF6B6B",
        TRUE            ~ "#FFFFFF"
      )
    }
  ) %>%
  opt_all_caps() %>%
  cols_align(
    align = "center",
    overall
  ) %>%
  tab_header(
    title = "Risk of Bias by Study and Age Band for 24 Hour Urine Calcium"
  )

calcium_rob
```

Table 1: Risk of Bias by Study and Age Band for 24 Hour Urine Calcium

| Study | Risk of Bias | Study Type |
| --- | --- | --- |
| Arora et al. 2017 | High | Observational Cohort |
| Grases et al. 2015 | High | Observational Cohort |
| Nouvenne et al. 2013 | High | Observational Cohort |
| Arrabal-Polo et al. 2012 | High | Observational Cohort |
| Sweeney et al. 2009 | High | RCT |
| Matsumoto et al. 2006 | High | RCT |
| Stitchantrakul et al. 2004 | High | RCT |
| Allie et al. 2003 | High | RCT |
| Baxmann et al. 2003 | High | RCT |
| Ogawa et al. 2003 | High | Observational Cohort |
| Hess et al. 1999 | High | Observational Cohort |
| Rodgers et al. 1997 | High | Observational Cohort |
| Buchholz et al. 1996 | High | Observational Cohort |
| Christie et al. 1996 | High | Observational Cohort |
| Erwin et al. 1994 | High | Observational Cohort |
| Thongprayoon et al. 2022 | Low | Observational Cohort |
| Deng et al. 2017 | Low | Observational Cohort |
| Parvin et al. 2011 | Low | Observational Cohort |
| Maalouf et al. 2010 | Low | Observational Cohort |
| Traxer et al. 2003 | Low | RCT |
| Mai et al. 2019 | Low | Observational Cohort |
| Doizi et al. 2016 | Low | Observational Cohort |
| Perinpam et al. 2015 | Low | Observational Cohort |
| Elkoushy et al. 2014 | Moderate | Observational Cohort |
| Lange et al. 2014 | Moderate | RCT |
| Pigna et al. 2014 | Moderate | Observational Cohort |
| Herrel et al. 2012 | Moderate | RCT |
| Hong et al. 2012 | Moderate | Observational Cohort |
| Sorensen et al. 2012 | Moderate | Observational Cohort |
| Duffey et al. 2011 | Moderate | Observational Cohort |
| Rodgers et al. 2009 | Moderate | Observational Cohort |
| Stitchantrakul et al. 2007 | Moderate | Observational Cohort |
| Gettman et al. 2005 | Moderate | Observational Cohort |
| Kuo et al. 2003 | Moderate | Observational Cohort |
| Miyake et al. 2001 | Moderate | Observational Cohort |
| Lemann et al. 1996 | Moderate | Observational Cohort |
| Wabner et al. 1992 | Moderate | RCT |
| Welshman et al. 1975 | Moderate | Observational Cohort |
| Arrabal-Polo et al. 2015 | Moderate | Observational Cohort |
| Kinder et al. 2002 | Moderate | Observational Cohort |
| Matin and Streem, 2000 | Moderate | Observational Cohort |
| Rodgers et al. 2015 | Moderate | Observational Cohort |
| Hesse et al. 1986 | Moderate | Observational Cohort |
| Cao et al. 2023 | Moderate | Observational Cohort |
| HPFS et al. 40-50 | Moderate | Observational Cohort |
| HPFS et al. 50-60 | Moderate | Observational Cohort |
| HPFS et al. 60-70 | Moderate | Observational Cohort |
| HPFS et al. 70-80 | Moderate | Observational Cohort |
| NHS et al. 40-50 | Moderate | Observational Cohort |
| NHS et al. 50-60 | Moderate | Observational Cohort |
| NHS et al. 60-70 | Moderate | Observational Cohort |
| NHS et al. 70-80 | Moderate | Observational Cohort |

##### 9.4.6 24 Hour Urine Calcium Suggested Expected Range inc. GRADE

```
calcium_grade <- calculate_grade(
  study_design= "Mixed", # Main results are from both Observational + RCTs
  risk_of_bias = "Moderate", # from bias analysis - low ROB subanalysis widens range
  inconsistency = "Serious", # upper limits range from ~3.7 -> 6.7
  indirectness = "Not Serious", # Large spread of studies likely to be representative
  imprecision = "Not Serious", # High precision
  publication_bias = "Strongly suspected", # Large cochran's Q values + I2
  large_effect = TRUE,
  dose_response = FALSE,
  confounders_reduce_effect = mr_reduces_effect(calcium_ma_2nd_analysis$ma_results)
)

calcium_grade <- cbind(
  sex = c("Overall"),
  grade = calcium_grade$grade_rating
) %>% as_tibble() %>%
  mutate(
    grade_rating = grade_symbol(grade)
  )

calcium_final_results <- get_final_ma_results(calcium_ma_2nd_analysis$ma_results, sex_specific = FALSE, meta_regression = TRUE) %>%
  mutate(
    pooled_estimate = round(pooled_estimate, 2),
    pooled_sd       = round(pooled_sd, 2),
    Q               = round(Q, 2),
    Q_pval = case_when(
      Q_pval < 0.001 ~ "<0.001",
      Q_pval < 0.01  ~ "<0.01",
      Q_pval < 0.05  ~ "<0.05",
      TRUE           ~ sprintf("%.2f", Q_pval)
    ),
    ci_95 = paste0(
      round(lower_95, 1), " – ", round(upper_95, 1)
    )
  ) %>%
  select(
    age_band,
    total_participants,
    ci_95,
    Q,
    Q_pval
  ) %>%
  cbind(
    calcium_grade) %>%
  mutate(grade_rating = paste0(grade, " ", grade_rating)) %>%
  select(-c(sex,grade))

calcium_final_results %>%
  gt() %>%
  cols_label(
    age_band        = "Analysis",
    total_participants  = "Patients, n",
    ci_95           = "95% CI",
    Q               = "Q statistic",
    Q_pval          = "Q p-value",
    grade_rating    = "GRADE Rating"
  ) %>%
  cols_align(
    align = "center"
  ) %>%
  tab_header(
    title = "Finalised Meta-Analysis Results for Calcium",
    subtitle = "Subdivided by Age Band"
  ) %>%
  tab_source_note(
    source_note = md(
      "*Q p-value = Cochran's Q heterogeneity test; RoB = Risk of Bias*"
    )
  )
```

Table 1: Finalised Meta-Analysis Results for Calcium

Subdivided by Age Band

| Analysis | Patients, n | 95% CI | Q statistic | Q p-value | GRADE Rating |
| --- | --- | --- | --- | --- | --- |
| Meta-Regression | 5418 | 1.9 – 6.2 | 2744.79 | <0.001 | Low ⊕⊕◯◯ |
| *Q p-value = Cochran's Q heterogeneity test; RoB = Risk of Bias* | | | | | |

```
calcium_final_results <- calcium_final_results %>%
  mutate(
    urine_component = "calcium"
  )
```

#### 9.5 Urate

##### 9.5.1 Framework Recommendation

framework_callout("urate")

Table 1: **Framework Recommendation — urate**

Branch 2 — Robust sex difference

| Component | Recommended | Branch | Reason | k | n | R² | Sex p (women_prop / overall / MR / LRoB) | Age p (min) | race p (min) | Assay p (min) | Egger's p | Q/df (chosen / overall) | Q/df ratio (chosen÷overall) | Q p (chosen) | MR R² (chosen) | QC warnings |
| --- | --- | --- | --- | --- | --- | --- | --- | --- | --- | --- | --- | --- | --- | --- | --- | --- |
| urate | Sex-stratified | 2 — Robust sex difference | women_prop coef p=0.042 in main meta-regression | 93 | 9,544 | 5.9% | 0.042 / 0.436 / 0.461 / 0.592 | — | — | — | 0.443 | 69.8 / 40.9 | 1.71 | <0.001 | 51% | Chosen Q/df (69.8) is 171% of overall Q/df (40.9) — exceeds limit of 150% |

##### 9.5.2 Comment

For 24-hour urine urate, the meta-analysis included k = 93 subgroup-level records and n = 9,544 participants. Most ranges lay between approximately 2 – 5 mmol/24h with high between-study heterogeneity (I² > 95%, Cochran’s Q p < 0.001), only modestly narrowed by meta-regression.

Significant sex differences were detected on meta-regression (p = 0.042) with a coefficient of −0.77 mmol/24h for women. The overall sex Wald test was non-significant (p = 0.436) but underpowered. Age comparisons were not testable across powered bands (no powered pairs available). race was severely underpowered (no powered pairs) with mostly single studies and wide ranges. Assay/country stratification was not tested for urate.

Meta-regression on the overall dataset explained very little heterogeneity (R² = 5.9%, k = 93), below the framework’s 10% threshold, and Egger’s test was non-significant (p = 0.443) — so the meta-regression branch and the small-study-bias override would not have been triggered in any case. Because the sex stratum test is evaluated first in the framework, meta-regression was bypassed regardless.

The RCT-only sub-analyses produced results within the respective sex-stratified ranges; the wider spread of the latter is consistent with their being more representative of the wider population. The low risk-of-bias sub-analysis did not suggest a more conservative estimate.

The framework selected the sex-stratified main results. The recommended expected ranges are **2.0 – 5.1 mmol/24h (male, n = 860)** and **2.3 – 3.5 mmol/24h (female, n = 4,177)**. The male cohort is substantially smaller than the female cohort, so the male estimate should be interpreted with caution; an automatic QC warning flagged that the chosen Q/df ratio (69.8) exceeded the framework’s 150% consistency limit relative to the overall Q/df (40.9), reflecting that the sex-stratified analysis is more heterogeneous per degree of freedom than the unadjusted overall.

##### 9.5.3 Plot

```
male_plot <- urate_ma_2nd_analysis$male$plot %+%
  subset(urate_ma_2nd_analysis$male$plot$data, age_band == "Overall") +
  scale_x_continuous(limits = c(-1,6))

female_plot <- urate_ma_2nd_analysis$female$plot %+%
  subset(urate_ma_2nd_analysis$female$plot$data, age_band == "Overall") +
  scale_x_continuous(limits = c(-1,6))

urate_final_plot <- (male_plot / female_plot) + plot_layout(axes = "collect")

urate_final_plot
## `height` was translated to `width`.
## `height` was translated to `width`.
```

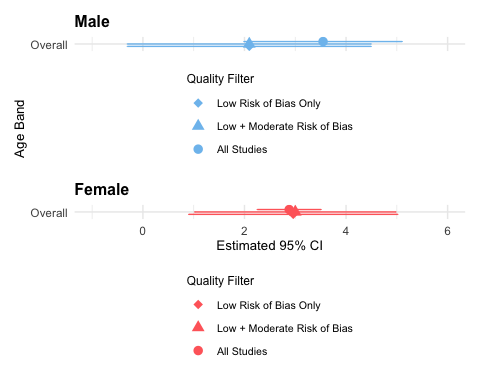

##### 9.5.4 Results Table

```
urate_ma_2nd_analysis$ma_results %>%
  filter(
    age_band %in% c("Overall"),
    quality_filter == "All Studies"
  ) %>% filter(
    sex_group != "main meta-analysis"
  ) %>%
  mutate(
    pooled_estimate = round(pooled_estimate, 2),
    pooled_sd       = round(pooled_sd, 2),
    I2              = round((I2 * 100), 0),
    Q               = round(Q, 2),
    Q_pval = case_when(
      Q_pval < 0.001 ~ "<0.001",
      Q_pval < 0.01  ~ "<0.01",
      Q_pval < 0.05  ~ "<0.05",
      TRUE           ~ as.character(round(Q_pval, 2))
    ),
    ci_95 = paste0(
      round(lower_95, 1), " – ", round(upper_95, 1)
    ),
    sex_group = str_to_title(sex_group)
  ) %>%
  select(
    sex_group,
    total_participants,
    pooled_estimate,
    pooled_sd,
    ci_95,
    I2,
    Q,
    Q_pval
  ) %>%
  gt() %>%
  cols_label(
    sex_group          = "Sex",
    total_participants  = "Patients, n",
    pooled_estimate = "Pooled Estimate",
    pooled_sd       = "SD",
    ci_95           = "95% CI",
    I2              = html("I<sup>2</sup> (%)"),
    Q               = "Q statistic",
    Q_pval          = "Q p-value"
  ) %>%
  cols_align(
    align = "center",
    columns = c(total_participants, pooled_estimate, pooled_sd, ci_95, I2, Q, Q_pval)
  ) %>%
  tab_header(
    title = "Urate Meta-analysis Results by Age Band",
    subtitle = "Random-effects meta-analysis"
  ) %>%
  tab_source_note(
    source_note = md(
      "*I² quantifies heterogeneity; Q p-value tests heterogeneity across studies.*"
    )
  )
```

Table 1: Urate Meta-analysis Results by Age Band

Random-effects meta-analysis

| Sex | Patients, n | Pooled Estimate | SD | 95% CI | I2 (%) | Q statistic | Q p-value |
| --- | --- | --- | --- | --- | --- | --- | --- |
| Male | 860 | 3.55 | 0.79 | 2 – 5.1 | 99 | 1118.46 | <0.001 |
| Female | 4177 | 2.88 | 0.32 | 2.3 – 3.5 | 99 | 1324.26 | <0.001 |
| *I² quantifies heterogeneity; Q p-value tests heterogeneity across studies.* | | | | | | | |

##### 9.5.5 Bias analysis

```
urate_rob <- urine_ma2 %>%
  drop_na(urate_sd_mmol_24h) %>%
  left_join(
    overall_bias, by = "study_name"
  ) %>%
  select(study_name, overall, study_type, male_female) %>%
  filter(male_female != "mixed") %>%
  mutate(
    overall = case_when(
      is.na(overall) ~ "Moderate",
      TRUE ~ overall
    ),
    sex_group = str_to_title(male_female)
  ) %>%
  distinct(study_name, .keep_all = TRUE) %>%
  arrange(overall) %>%
  group_by(sex_group) %>%
  gt() %>%
  cols_label(
    study_name = "Study",
    overall    = "Risk of Bias",
    study_type = "Study Type"
  ) %>%
  data_color(
    columns = overall,
    fn = function(x) {
      case_when(
        x == "Low"      ~ "#90EE90",
        x == "Moderate" ~ "#FFD700",
        x == "High"     ~ "#FF6B6B",
        TRUE            ~ "#FFFFFF"
      )
    }
  ) %>%
  opt_all_caps() %>%
  cols_align(
    align = "center",
    overall
  ) %>%
  tab_header(
    title = "Risk of Bias by Study and Age Band for 24 Hour Urine Urate"
  )

urate_rob
```

Table 1: Risk of Bias by Study and Age Band for 24 Hour Urine Urate

| Study | Risk of Bias | Study Type | male_female |
| --- | --- | --- | --- |
| Female | | | |
| Nouvenne et al. 2013 | High | Observational Cohort | female |
| Mai et al. 2019 | Low | Observational Cohort | female |
| Wabner et al. 1992 | Moderate | RCT | female |
| Hesse et al. 1986 | Moderate | Observational Cohort | female |
| NHS et al. 40-50 | Moderate | Observational Cohort | female |
| NHS et al. 50-60 | Moderate | Observational Cohort | female |
| NHS et al. 60-70 | Moderate | Observational Cohort | female |
| NHS et al. 70-80 | Moderate | Observational Cohort | female |
| Male | | | |
| Stitchantrakul et al. 2004 | High | RCT | male |
| Allie et al. 2003 | High | RCT | male |
| Hess et al. 1999 | High | Observational Cohort | male |
| Buchholz et al. 1996 | High | Observational Cohort | male |
| Parvin et al. 2011 | Low | Observational Cohort | male |
| Pigna et al. 2014 | Moderate | Observational Cohort | male |
| Miyake et al. 2001 | Moderate | Observational Cohort | male |
| Rodgers et al. 2015 | Moderate | Observational Cohort | male |
| HPFS et al. 40-50 | Moderate | Observational Cohort | male |
| HPFS et al. 50-60 | Moderate | Observational Cohort | male |
| HPFS et al. 60-70 | Moderate | Observational Cohort | male |
| HPFS et al. 70-80 | Moderate | Observational Cohort | male |

##### 9.5.6 24 Hour Urine Urate Suggested Expected Range inc. GRADE

```
urate_male_grade <- calculate_grade(
  study_design= "Mixed", # RCTs + Observational cohorts
  risk_of_bias = "Serious", # See bias analysis
  inconsistency = "Not Serious", # similar ranges throughout analyses
  indirectness = "Not Serious", # Large cohorts
  imprecision = "Not Serious", # Large cohorts with precise metrics
  publication_bias = "Strongly suspected", # Large I2 / Cochran's Q
  large_effect = TRUE, # Effect seems similar throughout sub-analyses
  dose_response = FALSE,
  confounders_reduce_effect = mr_reduces_effect(urate_ma_2nd_analysis$ma_results)
)

urate_female_grade <- calculate_grade(
  study_design= "Mixed", # RCTs + Observational cohorts
  risk_of_bias = "Moderate", # See bias analysis
  inconsistency = "Serious", # Wider variation in values than in men
  indirectness = "Not Serious", # Large cohorts
  imprecision = "Serious", # Wider variation in values than in men
  publication_bias = "Strongly Suspected", # Large I2 / Cochran's Q
  large_effect = FALSE, # Effect varies throughout sub-analyses
  dose_response = FALSE,
  confounders_reduce_effect = mr_reduces_effect(urate_ma_2nd_analysis$ma_results)
)

urate_grade <- cbind(
  sex = c("Male", "Female"),
  grade = c(urate_male_grade$grade_rating,
            urate_female_grade$grade_rating)
) %>% as_tibble() %>%
  mutate(
    grade_rating = grade_symbol(grade)
  )

urate_final_results <- get_final_ma_results(urate_ma_2nd_analysis$ma_results, sex_specific = TRUE) %>%
  mutate(
    pooled_estimate = round(pooled_estimate, 2),
    pooled_sd       = round(pooled_sd, 2),
    Q               = round(Q, 2),
    Q_pval = case_when(
      Q_pval < 0.001 ~ "<0.001",
      Q_pval < 0.01  ~ "<0.01",
      Q_pval < 0.05  ~ "<0.05",
      TRUE           ~ sprintf("%.2f", Q_pval)
    ),
    ci_95 = paste0(
      round(lower_95, 1), " – ", round(upper_95, 1)
    ),
    age_band = str_to_title(sex_group)
  ) %>%
  select(
    age_band,
    total_participants,
    ci_95,
    Q,
    Q_pval
  ) %>%
  cbind(
    urate_grade) %>%
  mutate(grade_rating = paste0(grade, " ", grade_rating)) %>%
  select(-c(sex, grade))

urate_final_results %>%
  gt() %>%
  cols_label(
    age_band       = "Analysis",
    total_participants  = "Patients, n",
    ci_95           = "95% CI",
    Q               = "Q statistic",
    Q_pval          = "Q p-value",
    grade_rating    = "GRADE Rating"
  ) %>%
  cols_align(
    align = "center"
  ) %>%
  tab_header(
    title = "Finalised Meta-Analysis Results for Urate",
    subtitle = "Subdivided by Age Band"
  ) %>%
  tab_source_note(
    source_note = md(
      "*Q p-value = Cochran's Q heterogeneity test; RoB = Risk of Bias*"
    )
  )
```

Table 1: Finalised Meta-Analysis Results for Urate

Subdivided by Age Band

| Analysis | Patients, n | 95% CI | Q statistic | Q p-value | GRADE Rating |
| --- | --- | --- | --- | --- | --- |
| Male | 860 | 2 – 5.1 | 1118.46 | <0.001 | Very Low ⊕◯◯◯ |
| Female | 4177 | 2.3 – 3.5 | 1324.26 | <0.001 | Very Low ⊕◯◯◯ |
| *Q p-value = Cochran's Q heterogeneity test; RoB = Risk of Bias* | | | | | |

```
urate_final_results <- urate_final_results %>%
  mutate(
    urine_component = "urate"
  )
```

#### 9.6 Creatinine

##### 9.6.1 Framework Recommendation

framework_callout("creat")

Table 1: **Framework Recommendation — creat**

Branch 2 — Robust sex difference

| Component | Recommended | Branch | Reason | k | n | R² | Sex p (women_prop / overall / MR / LRoB) | Age p (min) | race p (min) | Assay p (min) | Egger's p | Q/df (chosen / overall) | Q/df ratio (chosen÷overall) | Q p (chosen) | MR R² (chosen) | QC warnings |
| --- | --- | --- | --- | --- | --- | --- | --- | --- | --- | --- | --- | --- | --- | --- | --- | --- |
| creat | Sex-stratified | 2 — Robust sex difference | women_prop coef p=<0.001 in main meta-regression; sex p<0.05 in 2 of 3 tiers (overall=0.012, MR=<0.001, LRoB=0.272) | 74 | 9,058 | 82.0% | <0.001 / 0.012 / <0.001 / 0.272 | — | — | — | <0.001 | 46.8 / 1651.9 | 0.03 | <0.001 | — | — |

##### 9.6.2 Comment

For 24-hour urine creatinine, the meta-analysis included k = 74 subgroup-level records and n = 9,058 participants. Heterogeneity was substantial in all analyses (I² > 95%, Cochran’s Q p < 0.001).

Highly significant sex differences were detected on meta-regression (p < 0.001), and the overall sex Wald test also reached significance (p = 0.012). Age comparisons across powered bands were not testable (no powered pairs available). race stratification was severely underpowered with mostly single studies (no powered pairs). Assay/country stratification was not tested for creatinine.

Meta-regression explained an exceptionally high R² = 82.0% of heterogeneity with k = 74, well above the framework’s 10% threshold. Egger’s test was highly significant (p < 0.001), indicating strong small-study bias. However, because the sex stratum test is evaluated first in the framework, the framework proceeded directly to sex-stratified rather than meta-regression.

Age stratification dramatically reduces the number of eligible studies and participants, reducing power; the low risk-of-bias sub-analysis is consistent with the sex-stratified result and does not suggest a more informative or conservative estimate.

The framework selected the sex-stratified main results. The recommended expected ranges are **12.2 – 16.3 mmol/24h (male, n = 760)** and **7.2 – 12.6 mmol/24h (female, n = 4,217)**. The male cohort is substantially smaller than the female cohort, so the male estimate should be interpreted with caution. The very high meta-regression R² (82%) combined with significant Egger’s small-study bias suggests that future analyses should treat the meta-regression result as a useful sanity check alongside the sex-stratified ranges.

##### 9.6.3 Plot

```
creatinine_final_plot <- (creat_ma_2nd_analysis$male$plot %+%
  subset(creat_ma_2nd_analysis$main_meta_analysis$plot$data,
         age_band %in% ("Overall"))) / (creat_ma_2nd_analysis$female$plot %+%
  subset(creat_ma_2nd_analysis$main_meta_analysis$plot$data,
         age_band %in% ("Overall"))) + plot_layout(axes = "collect")

creatinine_final_plot
## `height` was translated to `width`.
## `height` was translated to `width`.
```

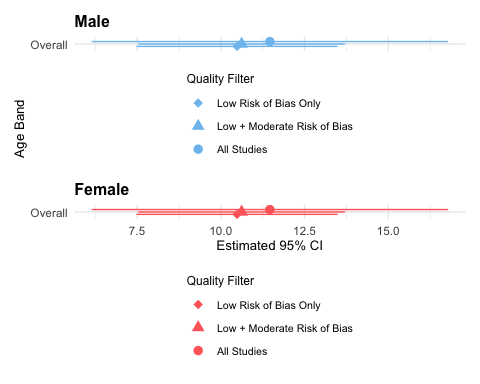

##### 9.6.4 Results Table

```
creat_ma_2nd_analysis$ma_results %>%
  filter(
    age_band %in% c("Overall"),
    quality_filter == "All Studies"
  ) %>% filter(
    sex_group != "main meta-analysis"
  ) %>%
  mutate(
    pooled_estimate = round(pooled_estimate, 2),
    pooled_sd       = round(pooled_sd, 2),
    I2              = round((I2 * 100), 0),
    Q               = round(Q, 2),
    Q_pval = case_when(
      Q_pval < 0.001 ~ "<0.001",
      Q_pval < 0.01  ~ "<0.01",
      Q_pval < 0.05  ~ "<0.05",
      TRUE           ~ as.character(round(Q_pval, 2))
    ),
    ci_95 = paste0(
      round(lower_95, 1), " – ", round(upper_95, 1)
    ),
    sex_group = str_to_title(sex_group)
  ) %>%
  select(
    sex_group,
    total_participants,
    pooled_estimate,
    pooled_sd,
    ci_95,
    I2,
    Q,
    Q_pval
  ) %>%
  gt() %>%
  cols_label(
    sex_group          = "Sex",
    total_participants  = "Patients, n",
    pooled_estimate = "Pooled Estimate",
    pooled_sd       = "SD",
    ci_95           = "95% CI",
    I2              = html("I<sup>2</sup> (%)"),
    Q               = "Q statistic",
    Q_pval          = "Q p-value"
  ) %>%
  cols_align(
    align = "center",
    columns = c(total_participants, pooled_estimate, pooled_sd, ci_95, I2, Q, Q_pval)
  ) %>%
  tab_header(
    title = "Creatinine Meta-analysis Results by Age Band",
    subtitle = "Random-effects meta-analysis"
  ) %>%
  tab_source_note(
    source_note = md(
      "*I² quantifies heterogeneity; Q p-value tests heterogeneity across studies.*"
    )
  )
```

Table 1: Creatinine Meta-analysis Results by Age Band

Random-effects meta-analysis

| Sex | Patients, n | Pooled Estimate | SD | 95% CI | I2 (%) | Q statistic | Q p-value |
| --- | --- | --- | --- | --- | --- | --- | --- |
| Male | 760 | 14.23 | 1.05 | 12.2 – 16.3 | 92 | 196.17 | <0.001 |
| Female | 4217 | 9.89 | 1.37 | 7.2 – 12.6 | 99 | 1625.32 | <0.001 |
| *I² quantifies heterogeneity; Q p-value tests heterogeneity across studies.* | | | | | | | |

##### 9.6.5 Bias analysis

```
creatinine_rob <- urine_ma2 %>%
  drop_na(creatinine_sd_mmol_24h) %>%
  left_join(
    overall_bias, by = "study_name"
  ) %>%
  select(study_name, overall, study_type, male_female) %>%
  filter(male_female != "mixed") %>%
  mutate(
    overall = case_when(
      is.na(overall) ~ "Moderate",
      TRUE ~ overall
    ),
    sex_group = str_to_title(male_female)
  ) %>%
  distinct(study_name, .keep_all = TRUE) %>%
  arrange(overall) %>%
  group_by(sex_group) %>%
  gt() %>%
  cols_label(
    study_name = "Study",
    overall    = "Risk of Bias",
    study_type = "Study Type"
  ) %>%
  data_color(
    columns = overall,
    fn = function(x) {
      case_when(
        x == "Low"      ~ "#90EE90",
        x == "Moderate" ~ "#FFD700",
        x == "High"     ~ "#FF6B6B",
        TRUE            ~ "#FFFFFF"
      )
    }
  ) %>%
  opt_all_caps() %>%
  cols_align(
    align = "center",
    overall
  ) %>%
  tab_header(
    title = "Risk of Bias by Study and Age Band for 24 Hour Urine Creatinine"
  )

creatinine_rob
```

Table 1: Risk of Bias by Study and Age Band for 24 Hour Urine Creatinine

| Study | Risk of Bias | Study Type | male_female |
| --- | --- | --- | --- |
| Female | | | |
| Nouvenne et al. 2013 | High | Observational Cohort | female |
| Mai et al. 2019 | Low | Observational Cohort | female |
| Hesse et al. 1986 | Moderate | Observational Cohort | female |
| NHS et al. 40-50 | Moderate | Observational Cohort | female |
| NHS et al. 50-60 | Moderate | Observational Cohort | female |
| NHS et al. 60-70 | Moderate | Observational Cohort | female |
| NHS et al. 70-80 | Moderate | Observational Cohort | female |
| Male | | | |
| Stitchantrakul et al. 2004 | High | RCT | male |
| Allie et al. 2003 | High | RCT | male |
| Parvin et al. 2011 | Low | Observational Cohort | male |
| Doenyas-Barak et al. 2015 | Moderate | Observational Cohort | male |
| Pigna et al. 2014 | Moderate | Observational Cohort | male |
| Miyake et al. 2001 | Moderate | Observational Cohort | male |
| Rodgers et al. 2015 | Moderate | Observational Cohort | male |
| HPFS et al. 40-50 | Moderate | Observational Cohort | male |
| HPFS et al. 50-60 | Moderate | Observational Cohort | male |
| HPFS et al. 60-70 | Moderate | Observational Cohort | male |
| HPFS et al. 70-80 | Moderate | Observational Cohort | male |

##### 9.6.6 24 Hour Urine Creatinine Suggested Expected Range inc. GRADE

```
creatinine_male_grade <- calculate_grade(
  study_design= "Mixed",
  risk_of_bias = "Serious", # Most studies at serious risk of bias
  inconsistency = "Serious",
  indirectness = "Moderate",
  imprecision = "Serious",
  publication_bias = "Strongly suspected", # Significant Cochran's Q
  large_effect = TRUE,
  dose_response = FALSE,
  confounders_reduce_effect = mr_reduces_effect(creat_ma_2nd_analysis$ma_results)
)

creatinine_female_grade <- calculate_grade(
  study_design= "Mixed",
  risk_of_bias = "Moderate", # Low ROB analysis comparable to overall female result
  inconsistency = "Serious",
  indirectness = "Moderate",
  imprecision = "Serious",
  publication_bias = "Suspected", # Significant Cochran's Q
  large_effect = TRUE,
  dose_response = FALSE,
  confounders_reduce_effect = mr_reduces_effect(creat_ma_2nd_analysis$ma_results)
)

creatinine_grade <- cbind(
  sex = c("Male", "Female"),
  grade = c(creatinine_male_grade$grade_rating,
            creatinine_female_grade$grade_rating)
) %>% as_tibble() %>%
  mutate(
    grade_rating = grade_symbol(grade)
  )

creatinine_final_results <- get_final_ma_results(creat_ma_2nd_analysis$ma_results, sex_specific = TRUE) %>%
  mutate(
    pooled_estimate = round(pooled_estimate, 2),
    pooled_sd       = round(pooled_sd, 2),
    Q               = round(Q, 2),
    Q_pval = case_when(
      Q_pval < 0.001 ~ "<0.001",
      Q_pval < 0.01  ~ "<0.01",
      Q_pval < 0.05  ~ "<0.05",
      TRUE           ~ sprintf("%.2f", Q_pval)
    ),
    ci_95 = paste0(
      round(lower_95, 1), " – ", round(upper_95, 1)
    ),
    age_band = str_to_title(sex_group)
  ) %>%
  select(
    age_band,
    total_participants,
    ci_95,
    Q,
    Q_pval
  ) %>%
  cbind(
    creatinine_grade) %>%
  mutate(grade_rating = paste0(grade, " ", grade_rating)) %>%
  select(-c(sex, grade))

creatinine_final_results %>%
  gt() %>%
  cols_label(
    age_band       = "Analysis",
    total_participants  = "Patients, n",
    ci_95           = "95% CI",
    Q               = "Q statistic",
    Q_pval          = "Q p-value",
    grade_rating    = "GRADE Rating"
  ) %>%
  cols_align(
    align = "center"
  ) %>%
  tab_header(
    title = "Finalised Meta-Analysis Results for creatinine",
    subtitle = "Subdivided by Age Band"
  ) %>%
  tab_source_note(
    source_note = md(
      "*Q p-value = Cochran's Q heterogeneity test; RoB = Risk of Bias*"
    )
  )
```

Table 1: Finalised Meta-Analysis Results for creatinine

Subdivided by Age Band

| Analysis | Patients, n | 95% CI | Q statistic | Q p-value | GRADE Rating |
| --- | --- | --- | --- | --- | --- |
| Male | 760 | 12.2 – 16.3 | 196.17 | <0.001 | Very Low ⊕◯◯◯ |
| Female | 4217 | 7.2 – 12.6 | 1625.32 | <0.001 | Low ⊕⊕◯◯ |
| *Q p-value = Cochran's Q heterogeneity test; RoB = Risk of Bias* | | | | | |

```
creatinine_final_results <- creatinine_final_results %>%
  mutate(
    urine_component = "creatinine"
  )
```

#### 9.7 Oxalate

##### 9.7.1 Framework Recommendation

framework_callout("oxalate")

Table 1: **Framework Recommendation — oxalate**

Branch 2 — Robust sex difference

| Component | Recommended | Branch | Reason | k | n | R² | Sex p (women_prop / overall / MR / LRoB) | Age p (min) | race p (min) | Assay p (min) | Egger's p | Q/df (chosen / overall) | Q/df ratio (chosen÷overall) | Q p (chosen) | MR R² (chosen) | QC warnings |
| --- | --- | --- | --- | --- | --- | --- | --- | --- | --- | --- | --- | --- | --- | --- | --- | --- |
| oxalate | Sex-stratified | 2 — Robust sex difference | women_prop coef p=0.012 in main meta-regression | 92 | 9,725 | 38.3% | 0.012 / 0.541 / 0.238 / 0.687 | — | — | — | 0.020 | 117.0 / 176.1 | 0.66 | <0.001 | 0% | — |

##### 9.7.2 Comment

For 24-hour urine oxalate, the meta-analysis included k = 92 subgroup-level records and n = 9,725 participants. Heterogeneity was high across most strata (I² > 95%, Cochran’s Q p < 0.001), save for some small subgroups limited to a small number of studies.

Significant sex differences were detected in the univariable meta-regression (estimate = −0.15 mmol/24h per unit increase in proportion-female, 95% CI: −0.22 to −0.07, p < 0.001) and remained significant (p = 0.012) in the multivariable framework meta-regression. The overall sex Wald test was non-significant (p = 0.541), reflecting the imprecise stratum-level comparison. Age comparisons across powered bands were not testable (no powered pairs). race stratification was severely underpowered (no powered pairs). Assay/country stratification was not tested.

Meta-regression explained R² = 38.3% of heterogeneity with k = 92, well above the framework’s 10% threshold; Egger’s test was significant (p = 0.020), indicating small-study bias. Both the meta-regression and the small-study-bias override would therefore have been satisfied — but because the sex stratum test is evaluated first, meta-regression was bypassed.

The low risk-of-bias sub-analysis produced sex-stratified ranges close to the main analysis; the RCT-only sub-analysis was consistent. Sample size showed no significant effect in the meta-regression. Sensitivity analyses therefore supported the sex-stratified choice.

The framework selected the sex-stratified main meta-analysis. The recommended expected ranges are **0.2 – 0.6 mmol/24h (male, n = 860)** and **0.2 – 0.4 mmol/24h (female, n = 4,177)**. The male cohort is substantially smaller than the female cohort. Reviewer DF flagged that enzymatic and ion-chromatography oxalate assays may give systematically different results; the current dataset uses country as a proxy for assay and was unable to detect a significant assay/country effect, but residual assay variability is a plausible contributor to the remaining heterogeneity within each sex.

##### 9.7.3 Plot

```
oxalate_final_plot <- (oxalate_ma_2nd_analysis$male$plot %+%
  subset(oxalate_ma_2nd_analysis$main_meta_analysis$plot$data,
         age_band %in% ("Overall"))) / (oxalate_ma_2nd_analysis$female$plot %+%
  subset(oxalate_ma_2nd_analysis$main_meta_analysis$plot$data,
         age_band %in% ("Overall"))) + plot_layout(axes = "collect")

oxalate_final_plot
## `height` was translated to `width`.
## `height` was translated to `width`.
```

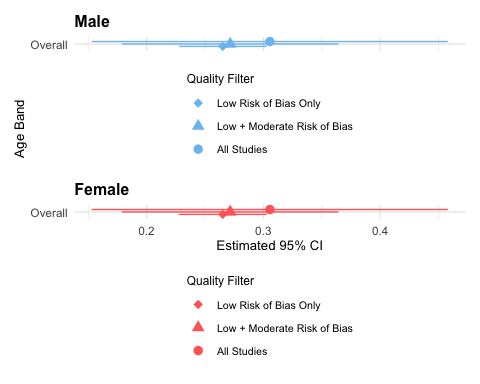

##### 9.7.4 Results Table

```
oxalate_ma_2nd_analysis$ma_results %>%
  filter(
    age_band %in% c("Overall"),
    quality_filter == "All Studies"
  ) %>% filter(
    sex_group != "main meta-analysis"
  ) %>%
  mutate(
    pooled_estimate = round(pooled_estimate, 2),
    pooled_sd       = round(pooled_sd, 2),
    I2              = round((I2 * 100), 0),
    Q               = round(Q, 2),
    Q_pval = case_when(
      Q_pval < 0.001 ~ "<0.001",
      Q_pval < 0.01  ~ "<0.01",
      Q_pval < 0.05  ~ "<0.05",
      TRUE           ~ as.character(round(Q_pval, 2))
    ),
    ci_95 = paste0(
      round(lower_95, 1), " – ", round(upper_95, 1)
    ),
    sex_group = str_to_title(sex_group)
  ) %>%
  select(
    sex_group,
    total_participants,
    pooled_estimate,
    pooled_sd,
    ci_95,
    I2,
    Q,
    Q_pval
  ) %>%
  gt() %>%
  cols_label(
    sex_group          = "Analysis",
    total_participants  = "Patients, n",
    pooled_estimate = "Pooled Estimate",
    pooled_sd       = "SD",
    ci_95           = "95% CI",
    I2              = html("I<sup>2</sup> (%)"),
    Q               = "Q statistic",
    Q_pval          = "Q p-value"
  ) %>%
  cols_align(
    align = "center",
    columns = c(total_participants, pooled_estimate, pooled_sd, ci_95, I2, Q, Q_pval)
  ) %>%
  tab_header(
    title = "Oxalate Meta-analysis Results by Age Band",
    subtitle = "Random-effects meta-analysis"
  ) %>%
  tab_source_note(
    source_note = md(
      "*I² quantifies heterogeneity; Q p-value tests heterogeneity across studies.*"
    )
  )
```

Table 1: Oxalate Meta-analysis Results by Age Band

Random-effects meta-analysis

| Analysis | Patients, n | Pooled Estimate | SD | 95% CI | I2 (%) | Q statistic | Q p-value |
| --- | --- | --- | --- | --- | --- | --- | --- |
| Male | 860 | 0.39 | 0.12 | 0.2 – 0.6 | 100 | 3373.74 | <0.001 |
| Female | 4177 | 0.31 | 0.04 | 0.2 – 0.4 | 96 | 439.70 | <0.001 |
| *I² quantifies heterogeneity; Q p-value tests heterogeneity across studies.* | | | | | | | |

##### 9.7.5 Bias analysis

```
oxalate_rob <- urine_ma2 %>%
  drop_na(oxalate_sd_mmol_24h) %>%
  left_join(
    overall_bias, by = "study_name"
  ) %>%
  select(study_name, overall, study_type, male_female) %>%
  filter(male_female != "mixed") %>%
  mutate(
    overall = case_when(
      is.na(overall) ~ "Moderate",
      TRUE ~ overall
    ),
    sex_group = str_to_title(male_female)
  ) %>%
  distinct(study_name, .keep_all = TRUE) %>%
  arrange(overall) %>%
  group_by(sex_group) %>%
  gt() %>%
  cols_label(
    study_name = "Study",
    overall    = "Risk of Bias",
    study_type = "Study Type"
  ) %>%
  data_color(
    columns = overall,
    fn = function(x) {
      case_when(
        x == "Low"      ~ "#90EE90",
        x == "Moderate" ~ "#FFD700",
        x == "High"     ~ "#FF6B6B",
        TRUE            ~ "#FFFFFF"
      )
    }
  ) %>%
  opt_all_caps() %>%
  cols_align(
    align = "center",
    overall
  ) %>%
  tab_header(
    title = "Risk of Bias by Study and Age Band for 24 Hour Urine Oxalate"
  )

oxalate_rob
```

Table 1: Risk of Bias by Study and Age Band for 24 Hour Urine Oxalate

| Study | Risk of Bias | Study Type | male_female |
| --- | --- | --- | --- |
| Female | | | |
| Nouvenne et al. 2013 | High | Observational Cohort | female |
| Mai et al. 2019 | Low | Observational Cohort | female |
| Wabner et al. 1992 | Moderate | RCT | female |
| Hesse et al. 1986 | Moderate | Observational Cohort | female |
| NHS et al. 40-50 | Moderate | Observational Cohort | female |
| NHS et al. 50-60 | Moderate | Observational Cohort | female |
| NHS et al. 60-70 | Moderate | Observational Cohort | female |
| NHS et al. 70-80 | Moderate | Observational Cohort | female |
| Male | | | |
| Stitchantrakul et al. 2004 | High | RCT | male |
| Allie et al. 2003 | High | RCT | male |
| Hess et al. 1999 | High | Observational Cohort | male |
| Buchholz et al. 1996 | High | Observational Cohort | male |
| Parvin et al. 2011 | Low | Observational Cohort | male |
| Pigna et al. 2014 | Moderate | Observational Cohort | male |
| Miyake et al. 2001 | Moderate | Observational Cohort | male |
| Rodgers et al. 2015 | Moderate | Observational Cohort | male |
| HPFS et al. 40-50 | Moderate | Observational Cohort | male |
| HPFS et al. 50-60 | Moderate | Observational Cohort | male |
| HPFS et al. 60-70 | Moderate | Observational Cohort | male |
| HPFS et al. 70-80 | Moderate | Observational Cohort | male |

##### 9.7.6 24 Hour Urine Oxalate Suggested Expected Range inc. GRADE

```
oxalate_grade_female <- calculate_grade(
  study_design= "Mixed", # Overall result consistent with RCT subanalysis
  risk_of_bias = "Serious", # RoB subanalyses
  inconsistency = "Serious", # Consistent across subanalyses
  indirectness = "Moderate",
  imprecision = "Not Serious", # Consistent across subanalyses
  publication_bias = "Strongly suspected", # Significant Cochran's Q
  large_effect = TRUE,
  dose_response = FALSE,
  confounders_reduce_effect = mr_reduces_effect(oxalate_ma_2nd_analysis$ma_results)
)

oxalate_grade_male <- calculate_grade(
  study_design= "Mixed", # Overall result consistent with RCT subanalysis
  risk_of_bias = "Serious", # Overall result consistent with Low ROB subanalysis
  inconsistency = "Serious", # Consistent across subanalyses
  indirectness = "Moderate",
  imprecision = "Not Serious", # Consistent across subanalyses
  publication_bias = "Strongly suspected", # Significant Cochran's Q
  large_effect = TRUE,
  dose_response = FALSE,
  confounders_reduce_effect = mr_reduces_effect(oxalate_ma_2nd_analysis$ma_results)
)

oxalate_grade <- cbind(
  sex = c("Male", "Female"),
  grade = c(oxalate_grade_male$grade_rating,
            oxalate_grade_female$grade_rating)
) %>% as_tibble() %>%
  mutate(
    grade_rating = grade_symbol(grade)
  )

oxalate_final_results <- get_final_ma_results(oxalate_ma_2nd_analysis$ma_results, sex_specific = TRUE, meta_regression = FALSE, low_rob = FALSE) %>%
  mutate(
    pooled_estimate = round(pooled_estimate, 2),
    pooled_sd       = round(pooled_sd, 2),
    Q               = round(Q, 2),
    Q_pval = case_when(
      Q_pval < 0.001 ~ "<0.001",
      Q_pval < 0.01  ~ "<0.01",
      Q_pval < 0.05  ~ "<0.05",
      TRUE           ~ sprintf("%.2f", Q_pval)
    ),
    ci_95 = paste0(
      round(lower_95, 1), " – ", round(upper_95, 1)
    ),
    age_band = str_to_title(sex_group)
  ) %>%
  select(
    age_band,
    total_participants,
    ci_95,
    Q,
    Q_pval
  ) %>%
  cbind(
    oxalate_grade) %>%
  mutate(grade_rating = paste0(grade, " ", grade_rating)) %>%
  select(-c(sex, grade))

oxalate_final_results %>%
  gt() %>%
  cols_label(
    age_band       = "Sex",
    total_participants  = "Patients, n",
    ci_95           = "95% CI",
    Q               = "Q statistic",
    Q_pval          = "Q p-value",
    grade_rating    = "GRADE Rating"
  ) %>%
  cols_align(
    align = "center"
  ) %>%
  tab_header(
    title = "Finalised Meta-Analysis Results for Oxalate",
    subtitle = "Subdivided by Age Band"
  ) %>%
  tab_source_note(
    source_note = md(
      "*Q p-value = Cochran's Q heterogeneity test; RoB = Risk of Bias*"
    )
  )
```

Table 1: Finalised Meta-Analysis Results for Oxalate

Subdivided by Age Band

| Sex | Patients, n | 95% CI | Q statistic | Q p-value | GRADE Rating |
| --- | --- | --- | --- | --- | --- |
| Male | 860 | 0.2 – 0.6 | 3373.74 | <0.001 | Very Low ⊕◯◯◯ |
| Female | 4177 | 0.2 – 0.4 | 439.70 | <0.001 | Very Low ⊕◯◯◯ |
| *Q p-value = Cochran's Q heterogeneity test; RoB = Risk of Bias* | | | | | |

```
oxalate_final_results <- oxalate_final_results %>%
  mutate(
    urine_component = "oxalate"
  )
```

## 9.8 pH

##### 9.8.1 Framework Recommendation

framework_callout("ph")

Table 1: **Framework Recommendation — ph**

Branch 6 — Meta-regression effective

| Component | Recommended | Branch | Reason | k | n | R² | Sex p (women_prop / overall / MR / LRoB) | Age p (min) | race p (min) | Assay p (min) | Egger's p | Q/df (chosen / overall) | Q/df ratio (chosen÷overall) | Q p (chosen) | MR R² (chosen) | QC warnings |
| --- | --- | --- | --- | --- | --- | --- | --- | --- | --- | --- | --- | --- | --- | --- | --- | --- |
| ph | Meta-regression | 6 — Meta-regression effective | R² = 11.6% ≥ 10% with k = 95 ≥ 20 | 95 | 9,226 | 11.6% | 0.279 / 0.857 / 0.753 / 0.687 | 0.484 | 0.835 | — | 0.512 | 6.4 / 26.5 | 0.24 | <0.001 | 12% | — |

##### 9.8.2 Comment

For 24-hour urine pH, the meta-analysis included k = 95 subgroup-level records and n = 9,226 participants. Although there was substantial heterogeneity across most analyses (I² > 95%, Cochran’s Q p < 0.001), the absolute spread of values was relatively tight.

Stratum tests were uniformly non-significant. Sex differences were not significant on meta-regression (p = 0.279; overall sex Wald p = 0.857). Age comparisons across powered bands gave a minimum pairwise Wald p of 0.484, with little absolute difference between bands. race was underpowered (minimum Wald p = 0.835) with few studies. The assay/country stratum test was not applicable for pH.

Meta-regression explained R² = 11.6% of heterogeneity with k = 95, just above the framework’s 10% threshold — the meta-regression branch was satisfied. Egger’s test was not significant (p = 0.512), so the small-study-bias override would not have been invoked but was not needed. Reassuringly, the meta-regression-adjusted estimate coincides closely with the unadjusted overall pooled estimate.

The low risk-of-bias overall and RCT-only sub-analyses were consistent with the main meta-analysis, and the absolute pH range was stable across all sensitivity analyses.

The framework selected the meta-regression result. The recommended expected range is **5.7 – 6.3** (pH is dimensionless, n = 9,226). For this analyte the meta-regression and unadjusted overall pooled estimates are numerically equivalent.

##### 9.8.3 Plot

```
ph_ma_2nd_analysis$main_meta_analysis$plot %+%
  subset(ph_ma_2nd_analysis$main_meta_analysis$plot$data,
         age_band %in% ("Meta-Regression"))
## `height` was translated to `width`.
```

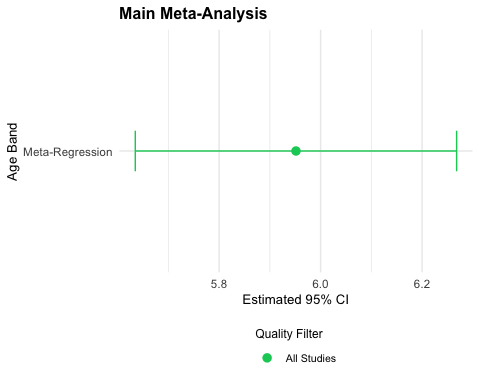

##### 9.8.4 Results Table

```
ph_ma_2nd_analysis$ma_results %>%
  filter(
    age_band %in% c("Meta-Regression")
  ) %>% filter(
    sex_group == "main meta-analysis"
  ) %>%
  mutate(
    pooled_estimate = round(pooled_estimate, 2),
    pooled_sd       = round(pooled_sd, 2),
    I2              = round((I2 * 100), 0),
    Q               = round(Q, 2),
    Q_pval = case_when(
      Q_pval < 0.001 ~ "<0.001",
      Q_pval < 0.01  ~ "<0.01",
      Q_pval < 0.05  ~ "<0.05",
      TRUE           ~ as.character(round(Q_pval, 2))
    ),
    ci_95 = paste0(
      round(lower_95, 1), " – ", round(upper_95, 1)
    )
  ) %>%
  select(
    age_band,
    total_participants,
    pooled_estimate,
    pooled_sd,
    ci_95,
    I2,
    Q,
    Q_pval
  ) %>%
  gt() %>%
  cols_label(
    age_band = "Analysis",
    total_participants  = "Patients, n",
    pooled_estimate = "Pooled Estimate",
    pooled_sd       = "SD",
    ci_95           = "95% CI",
    I2              = html("I<sup>2</sup> (%)"),
    Q               = "Q statistic",
    Q_pval          = "Q p-value"
  ) %>%
  cols_align(
    align = "center",
    columns = c(total_participants, pooled_estimate, pooled_sd, ci_95, I2, Q, Q_pval)
  ) %>%
  tab_header(
    title = "pH Meta-analysis Results by Age Band",
    subtitle = "Meta-Regression"
  ) %>%
  tab_source_note(
    source_note = md(
      "*I² quantifies heterogeneity; Q p-value tests heterogeneity across studies.*"
    )
  )
```

Table 1: pH Meta-analysis Results by Age Band

Meta-Regression

| Analysis | Patients, n | Pooled Estimate | SD | 95% CI | I2 (%) | Q statistic | Q p-value |
| --- | --- | --- | --- | --- | --- | --- | --- |
| Meta-Regression | 4379 | 5.95 | 0.16 | 5.6 – 6.3 | 88 | 324.21 | <0.001 |
| *I² quantifies heterogeneity; Q p-value tests heterogeneity across studies.* | | | | | | | |

##### 9.8.5 Bias analysis

```
ph_rob <- urine_ma2 %>%
  drop_na(ph_sd) %>%
  left_join(
    overall_bias, by = "study_name"
  ) %>%
  select(study_name, overall, study_type) %>%
  mutate(
    overall = case_when(
      is.na(overall) ~ "Moderate",
      TRUE ~ overall
    )
  ) %>%
  distinct(study_name, .keep_all = TRUE) %>%
  arrange(overall) %>%
  gt() %>%
  cols_label(
    study_name = "Study",
    overall    = "Risk of Bias",
    study_type = "Study Type"
  ) %>%
  data_color(
    columns = overall,
    fn = function(x) {
      case_when(
        x == "Low"      ~ "#90EE90",
        x == "Moderate" ~ "#FFD700",
        x == "High"     ~ "#FF6B6B",
        TRUE            ~ "#FFFFFF"
      )
    }
  ) %>%
  opt_all_caps() %>%
  cols_align(
    align = "center",
    overall
  ) %>%
  tab_header(
    title = "Risk of Bias by Study and Age Band for 24 Hour Urine ph"
  )

ph_rob
```

Table 1: Risk of Bias by Study and Age Band for 24 Hour Urine ph

| Study | Risk of Bias | Study Type |
| --- | --- | --- |
| Arora et al. 2017 | High | Observational Cohort |
| Nouvenne et al. 2013 | High | Observational Cohort |
| Arrabal-Polo et al. 2012 | High | Observational Cohort |
| Sweeney et al. 2009 | High | RCT |
| Matsumoto et al. 2006 | High | RCT |
| Defoor et al. 2005 | High | Observational Cohort |
| Abate et al. 2004 | High | Observational Cohort |
| Allie et al. 2003 | High | RCT |
| Hess et al. 1999 | High | Observational Cohort |
| Buchholz et al. 1996 | High | Observational Cohort |
| Christie et al. 1996 | High | Observational Cohort |
| Erwin et al. 1994 | High | Observational Cohort |
| Stern et al. 1980 | High | Observational Cohort |
| Marya et al. 1979 | High | Observational Cohort |
| Thongprayoon et al. 2022 | Low | Observational Cohort |
| Deng et al. 2017 | Low | Observational Cohort |
| Parvin et al. 2011 | Low | Observational Cohort |
| Maalouf et al. 2010 | Low | Observational Cohort |
| Traxer et al. 2003 | Low | RCT |
| Mai et al. 2019 | Low | Observational Cohort |
| Doizi et al. 2016 | Low | Observational Cohort |
| Ma et al. 2014 | Low | Observational Cohort |
| Pigna et al. 2014 | Moderate | Observational Cohort |
| Goodarzi et al. 2012 | Moderate | Observational Cohort |
| Herrel et al. 2012 | Moderate | RCT |
| Hong et al. 2012 | Moderate | Observational Cohort |
| Sorensen et al. 2012 | Moderate | Observational Cohort |
| Duffey et al. 2011 | Moderate | Observational Cohort |
| Rodgers et al. 2009 | Moderate | Observational Cohort |
| Cameron et al. 2006 | Moderate | Observational Cohort |
| Gettman et al. 2005 | Moderate | Observational Cohort |
| Huang et al. 2003 | Moderate | Observational Cohort |
| Kuo et al. 2003 | Moderate | Observational Cohort |
| Miyake et al. 2001 | Moderate | Observational Cohort |
| Lemann et al. 1996 | Moderate | Observational Cohort |
| Wabner et al. 1992 | Moderate | RCT |
| Kinder et al. 2002 | Moderate | Observational Cohort |
| Rodgers et al. 2015 | Moderate | Observational Cohort |
| Hesse et al. 1986 | Moderate | Observational Cohort |
| HPFS et al. 40-50 | Moderate | Observational Cohort |
| HPFS et al. 50-60 | Moderate | Observational Cohort |
| HPFS et al. 60-70 | Moderate | Observational Cohort |
| HPFS et al. 70-80 | Moderate | Observational Cohort |
| NHS et al. 40-50 | Moderate | Observational Cohort |
| NHS et al. 50-60 | Moderate | Observational Cohort |
| NHS et al. 60-70 | Moderate | Observational Cohort |
| NHS et al. 70-80 | Moderate | Observational Cohort |

##### 9.8.6 24 Hour Urine pH Suggested Expected Range inc. GRADE

```
ph_grade <- calculate_grade(
  study_design= "RCT", # Correlation with RCTs
  risk_of_bias = "Not Serious", # Very similar results between main result + low ROB subanalysis
  inconsistency = "Low", # Strongly consistent throughout studies
  indirectness = "Moderate",
  imprecision = "Moderate",
  publication_bias = "Strongly suspected", #significant cochran's Q + I2 high
  large_effect = TRUE,
  dose_response = FALSE,
  confounders_reduce_effect = mr_reduces_effect(ph_ma_2nd_analysis$ma_results)
)

ph_grade <- cbind(
  sex = c("Overall"),
  grade = ph_grade$grade_rating
) %>% as_tibble() %>%
  mutate(
    grade_rating = grade_symbol(grade)
  )

ph_final_results <- get_final_ma_results(ph_ma_2nd_analysis$ma_results, sex_specific = FALSE, meta_regression = TRUE, low_rob = FALSE) %>%
  mutate(
    pooled_estimate = round(pooled_estimate, 2),
    pooled_sd       = round(pooled_sd, 2),
    Q               = round(Q, 2),
    Q_pval = case_when(
      Q_pval < 0.001 ~ "<0.001",
      Q_pval < 0.01  ~ "<0.01",
      Q_pval < 0.05  ~ "<0.05",
      TRUE           ~ sprintf("%.2f", Q_pval)
    ),
    ci_95 = paste0(
      round(lower_95, 1), " – ", round(upper_95, 1)
    )
  ) %>%
  select(
    age_band,
    total_participants,
    ci_95,
    Q,
    Q_pval
  ) %>%
  cbind(
    ph_grade) %>%
  mutate(grade_rating = paste0(grade, " ", grade_rating)) %>%
  select(-c(sex, grade))

ph_final_results %>%
  gt() %>%
  cols_label(
    age_band       = "Sex",
    total_participants  = "Patients, n",
    ci_95           = "95% CI",
    Q               = "Q statistic",
    Q_pval          = "Q p-value",
    grade_rating    = "GRADE Rating"
  ) %>%
  cols_align(
    align = "center"
  ) %>%
  tab_header(
    title = "Finalised Meta-Analysis Results for pH",
    subtitle = "Subdivided by Age Band"
  ) %>%
  tab_source_note(
    source_note = md(
      "*Q p-value = Cochran's Q heterogeneity test; RoB = Risk of Bias*"
    )
  )
```

Table 1: Finalised Meta-Analysis Results for pH

Subdivided by Age Band

| Sex | Patients, n | 95% CI | Q statistic | Q p-value | GRADE Rating |
| --- | --- | --- | --- | --- | --- |
| Meta-Regression | 4379 | 5.6 – 6.3 | 324.21 | <0.001 | Moderate ⊕⊕⊕◯ |
| *Q p-value = Cochran's Q heterogeneity test; RoB = Risk of Bias* | | | | | |

```
ph_final_results <- ph_final_results %>%
  mutate(
    urine_component = "pH"
  )
```

#### 9.9 Citrate

##### 9.9.1 Framework Recommendation

framework_callout("citrate")

Table 1: **Framework Recommendation — citrate**

Branch 5 — Robust assay/country difference

| Component | Recommended | Branch | Reason | k | n | R² | Sex p (women_prop / overall / MR / LRoB) | Age p (min) | race p (min) | Assay p (min) | Egger's p | Q/df (chosen / overall) | Q/df ratio (chosen÷overall) | Q p (chosen) | MR R² (chosen) | QC warnings |
| --- | --- | --- | --- | --- | --- | --- | --- | --- | --- | --- | --- | --- | --- | --- | --- | --- |
| citrate | Assay/country-stratified | 5 — Robust assay/country difference | Assay min Wald p = <0.001 (rel shift 44.8%) across 3 named-category pair(s); 2/3 significant after 20% shift filter | 97 | 9,458 | 17.5% | 0.363 / 0.629 / 0.941 / 0.951 | 0.845 | 0.654 | <0.001 | 0.148 | — / 176.5 | — | — | 18% | — |

##### 9.9.2 Comment

For 24-hour urine citrate, the meta-analysis included k = 97 subgroup-level records and n = 9,458 participants. Heterogeneity was substantial across all analyses (I² > 95%, Cochran’s Q p < 0.001), with notable instability in the lower limit.

Sex differences on meta-regression were not significant (p = 0.363; overall sex Wald p = 0.629). Age comparisons across powered bands gave a minimum pairwise Wald p of 0.845. race was underpowered (minimum Wald p = 0.654). A strong between-assay/country effect was identified: the minimum pairwise Wald p between named assay/country categories was < 0.001, with 2 of 3 named pairs significant after the 20% relative-shift filter and a relative shift of 44.8% between categories.

Meta-regression explained R² = 17.5% of heterogeneity with k = 97 — above the framework’s 10% threshold — but because the assay/country stratum test fires first, the meta-regression branch was not reached. Egger’s test was not significant (p = 0.148).

The low risk-of-bias sub-analysis was consistent with the main result; the RCT-only sub-analysis gave a slightly tighter range.

The framework selected the assay/country-stratified branch. However, because clinicians do not generally know which assay method or laboratory produced an individual citrate result, assay-specific reference ranges are not clinically actionable. The displayed result is therefore the overall pooled estimate with an explicit caveat that citrate values vary materially by assay method. The recommended expected range is **1.2 – 4.5 mmol/24h** (Overall, with assay-variability caveat, n = 9,458). The lower limit is unstable across sensitivity analyses, and the wide assay-related variation is a substantive limitation for clinical use of a single pooled citrate range.

##### 9.9.3 Plot

```
citrate_final_plot <- citrate_ma_2nd_analysis$main_meta_analysis$plot %+%
  subset(citrate_ma_2nd_analysis$main_meta_analysis$plot$data, age_band == "Overall" & quality_filter == "All Studies") +
  scale_x_continuous(limits = c(0,5.5))

citrate_final_plot
## `height` was translated to `width`.
```

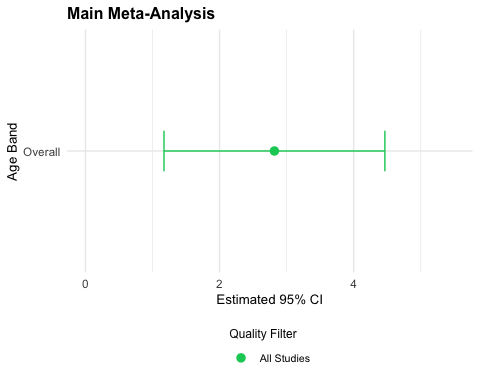

##### 9.9.4 Results Table

```
citrate_ma_2nd_analysis$ma_results %>%
  filter(
    age_band %in% c("Overall")
  ) %>% filter(
    sex_group == "main meta-analysis"
  ) %>% filter(
    quality_filter == "All Studies"
  ) %>%
  mutate(
    pooled_estimate = round(pooled_estimate, 2),
    pooled_sd       = round(pooled_sd, 2),
    I2              = round((I2 * 100), 0),
    Q               = round(Q, 2),
    Q_pval = case_when(
      Q_pval < 0.001 ~ "<0.001",
      Q_pval < 0.01  ~ "<0.01",
      Q_pval < 0.05  ~ "<0.05",
      TRUE           ~ as.character(round(Q_pval, 2))
    ),
    ci_95 = paste0(
      round(lower_95, 1), " – ", round(upper_95, 1)
    ),
    sex_group = "Overall"
  ) %>%
  select(
    sex_group,
    total_participants,
    pooled_estimate,
    pooled_sd,
    ci_95,
    I2,
    Q,
    Q_pval
  ) %>%
  gt() %>%
  cols_label(
    sex_group          = "Analysis",
    total_participants  = "Patients, n",
    pooled_estimate = "Pooled Estimate",
    pooled_sd       = "SD",
    ci_95           = "95% CI",
    I2              = html("I<sup>2</sup> (%)"),
    Q               = "Q statistic",
    Q_pval          = "Q p-value"
  ) %>%
  cols_align(
    align = "center",
    columns = c(total_participants, pooled_estimate, pooled_sd, ci_95, I2, Q, Q_pval)
  ) %>%
  tab_header(
    title = "Citrate Meta-analysis Results by Age Band",
    subtitle = "Random-effects meta-analysis"
  ) %>%
  tab_source_note(
    source_note = md(
      "*I² quantifies heterogeneity; Q p-value tests heterogeneity across studies.*"
    )
  )
```

Table 1: Citrate Meta-analysis Results by Age Band

Random-effects meta-analysis

| Analysis | Patients, n | Pooled Estimate | SD | 95% CI | I2 (%) | Q statistic | Q p-value |
| --- | --- | --- | --- | --- | --- | --- | --- |
| Overall | 9458 | 2.82 | 0.84 | 1.2 – 4.5 | 99 | 16945 | <0.001 |
| *I² quantifies heterogeneity; Q p-value tests heterogeneity across studies.* | | | | | | | |

##### 9.9.5 Bias analysis

```
citrate_rob <- urine_ma2 %>%
  drop_na(citrate_sd_mmol_24h) %>%
  left_join(
    overall_bias, by = "study_name"
  ) %>%
  select(study_name, overall, study_type) %>%
  mutate(
    overall = case_when(
      is.na(overall) ~ "Moderate",
      TRUE ~ overall
    )
  ) %>%
  distinct(study_name, .keep_all = TRUE) %>%
  arrange(overall) %>%
  gt() %>%
  cols_label(
    study_name = "Study",
    overall    = "Risk of Bias",
    study_type = "Study Type"
  ) %>%
  data_color(
    columns = overall,
    fn = function(x) {
      case_when(
        x == "Low"      ~ "#90EE90",
        x == "Moderate" ~ "#FFD700",
        x == "High"     ~ "#FF6B6B",
        TRUE            ~ "#FFFFFF"
      )
    }
  ) %>%
  opt_all_caps() %>%
  cols_align(
    align = "center",
    overall
  ) %>%
  tab_header(
    title = "Risk of Bias by Study and Age Band for 24 Hour Urine Citrate"
  )

citrate_rob
```

Table 1: Risk of Bias by Study and Age Band for 24 Hour Urine Citrate

| Study | Risk of Bias | Study Type |
| --- | --- | --- |
| Arora et al. 2017 | High | Observational Cohort |
| Grases et al. 2015 | High | Observational Cohort |
| Nouvenne et al. 2013 | High | Observational Cohort |
| Sweeney et al. 2009 | High | RCT |
| Ciacci et al. 2008 | High | Observational Cohort |
| Matsumoto et al. 2006 | High | RCT |
| Abate et al. 2004 | High | Observational Cohort |
| Stitchantrakul et al. 2004 | High | RCT |
| Allie et al. 2003 | High | RCT |
| Baxmann et al. 2003 | High | RCT |
| Ogawa et al. 2003 | High | Observational Cohort |
| Hess et al. 1999 | High | Observational Cohort |
| Rodgers et al. 1997 | High | Observational Cohort |
| Erwin et al. 1994 | High | Observational Cohort |
| Thongprayoon et al. 2022 | Low | Observational Cohort |
| Deng et al. 2017 | Low | Observational Cohort |
| Parvin et al. 2011 | Low | Observational Cohort |
| Maalouf et al. 2010 | Low | Observational Cohort |
| Traxer et al. 2003 | Low | RCT |
| Mai et al. 2019 | Low | Observational Cohort |
| Doizi et al. 2016 | Low | Observational Cohort |
| Elkoushy et al. 2014 | Moderate | Observational Cohort |
| Lange et al. 2014 | Moderate | RCT |
| Pigna et al. 2014 | Moderate | Observational Cohort |
| Goodarzi et al. 2012 | Moderate | Observational Cohort |
| Herrel et al. 2012 | Moderate | RCT |
| Hong et al. 2012 | Moderate | Observational Cohort |
| Sorensen et al. 2012 | Moderate | Observational Cohort |
| Duffey et al. 2011 | Moderate | Observational Cohort |
| Rodgers et al. 2009 | Moderate | Observational Cohort |
| Stitchantrakul et al. 2007 | Moderate | Observational Cohort |
| Cameron et al. 2006 | Moderate | Observational Cohort |
| Gettman et al. 2005 | Moderate | Observational Cohort |
| Miyake et al. 2001 | Moderate | Observational Cohort |
| Lemann et al. 1996 | Moderate | Observational Cohort |
| Wabner et al. 1992 | Moderate | RCT |
| Welshman et al. 1976 | Moderate | Observational Cohort |
| Arrabal-Polo et al. 2015 | Moderate | Observational Cohort |
| Shah, 2005 | Moderate | Observational Cohort |
| Matin and Streem, 2000 | Moderate | Observational Cohort |
| Rodgers et al. 2015 | Moderate | Observational Cohort |
| Hesse et al. 1986 | Moderate | Observational Cohort |
| HPFS et al. 40-50 | Moderate | Observational Cohort |
| HPFS et al. 50-60 | Moderate | Observational Cohort |
| HPFS et al. 60-70 | Moderate | Observational Cohort |
| HPFS et al. 70-80 | Moderate | Observational Cohort |
| NHS et al. 40-50 | Moderate | Observational Cohort |
| NHS et al. 50-60 | Moderate | Observational Cohort |
| NHS et al. 60-70 | Moderate | Observational Cohort |
| NHS et al. 70-80 | Moderate | Observational Cohort |

##### 9.9.6 24 Hour Urine Citrate Suggested Expected Range inc. GRADE

```
citrate_grade <- calculate_grade(
  study_design= "Mixed", # Mixed overall Result
  risk_of_bias = "Serious", #Low RoB sub-analysis used for estimation
  inconsistency = "Serious", # Marked variation in values
  indirectness = "Serious", # Assay variability detected
  imprecision = "Serious", # Large number of patients in each analysis
  publication_bias = "Suspected", # Significant Cochran's Q
  large_effect = TRUE,
  dose_response = FALSE,
  confounders_reduce_effect = mr_reduces_effect(citrate_ma_2nd_analysis$ma_results)
)

citrate_grade <- citrate_grade %>%
  as_tibble() %>%
  mutate(
    sex = c("Overall"),
    grade = grade_rating,
    grade_rating = grade_symbol(grade))

citrate_final_results <- get_final_ma_results(citrate_ma_2nd_analysis$ma_results, sex_specific = FALSE, meta_regression = FALSE, low_rob = FALSE) %>%
  mutate(
    pooled_estimate = round(pooled_estimate, 2),
    pooled_sd       = round(pooled_sd, 2),
    Q               = round(Q, 2),
    Q_pval = case_when(
      Q_pval < 0.001 ~ "<0.001",
      Q_pval < 0.01  ~ "<0.01",
      Q_pval < 0.05  ~ "<0.05",
      TRUE           ~ sprintf("%.2f", Q_pval)
    ),
    ci_95 = paste0(
      round(lower_95, 1), " – ", round(upper_95, 1)
    )
  ) %>%
  select(
    age_band,
    total_participants,
    ci_95,
    Q,
    Q_pval
  ) %>%
  cbind(
    citrate_grade) %>%
  mutate(grade_rating = paste0(grade, " ", grade_rating)) %>%
  select(-c(sex,grade, certainty_score))

citrate_final_results %>%
  gt() %>%
  cols_label(
    age_band       = "Analysis",
    total_participants  = "Patients, n",
    ci_95           = "95% CI",
    Q               = "Q statistic",
    Q_pval          = "Q p-value",
    grade_rating    = "GRADE Rating"
  ) %>%
  cols_align(
    align = "center"
  ) %>%
  tab_header(
    title = "Finalised Meta-Analysis Results for Citrate",
    subtitle = "Subdivided by Age Band"
  ) %>%
  tab_source_note(
    source_note = md(
      "*Q p-value = Cochran's Q heterogeneity test; RoB = Risk of Bias*"
    )
  )
```

Table 1: Finalised Meta-Analysis Results for Citrate

Subdivided by Age Band

| Analysis | Patients, n | 95% CI | Q statistic | Q p-value | GRADE Rating |
| --- | --- | --- | --- | --- | --- |
| Overall | 9458 | 1.2 – 4.5 | 16945 | <0.001 | Very Low ⊕◯◯◯ |
| *Q p-value = Cochran's Q heterogeneity test; RoB = Risk of Bias* | | | | | |

```
citrate_final_results <- citrate_final_results %>%
  mutate(
    urine_component = "citrate"
  )
```

#### 9.10 Phosphate

##### 9.10.1 Framework Recommendation

framework_callout("phosphate")

Table 1: **Framework Recommendation — phosphate**

Branch 6 — Meta-regression effective

| Component | Recommended | Branch | Reason | k | n | R² | Sex p (women_prop / overall / MR / LRoB) | Age p (min) | race p (min) | Assay p (min) | Egger's p | Q/df (chosen / overall) | Q/df ratio (chosen÷overall) | Q p (chosen) | MR R² (chosen) | QC warnings |
| --- | --- | --- | --- | --- | --- | --- | --- | --- | --- | --- | --- | --- | --- | --- | --- | --- |
| phosphate | Meta-regression | 6 — Meta-regression effective | R² = 76.8% ≥ 10% with k = 56 ≥ 20 | 56 | 8,104 | 76.8% | 0.067 / 0.338 / 0.701 / 0.756 | 0.393 | 0.165 | 0.076 | 0.001 | 4.7 / 210.6 | 0.02 | <0.001 | 77% | — |

##### 9.10.2 Comment

For 24-hour urine phosphate, the meta-analysis included k = 56 subgroup-level records and n = 8,104 participants. Heterogeneity was substantial across all analyses (I² > 95%, Cochran’s Q p < 0.001), and the upper limit was notably unstable across sensitivity analyses.

Sex differences on meta-regression approached but did not reach significance (p = 0.067; overall sex Wald p = 0.338). Age comparisons across powered bands gave a minimum pairwise Wald p of 0.393. race was severely underpowered (minimum Wald p = 0.165), with race sub-analyses often limited to single studies or no data at all. The assay/country stratum test approached significance (minimum Wald p = 0.076) but did not pass the threshold. Significant amounts of missing data limited the stratum analyses.

Meta-regression explained R² = 76.8% of heterogeneity with k = 56, well above the framework’s 10% threshold, so the meta-regression branch was satisfied. Egger’s test was highly significant (p = 0.001), indicating strong small-study bias; the small-study-bias override would also have selected meta-regression independently. The high R² combined with modest k carries a residual saturation/over-fit concern that the reader should bear in mind.

The low risk-of-bias and RCT-only sub-analyses were broadly consistent with the meta-regression result, but the upper limit was less stable; the meta-regression-adjusted upper limit (~26) is tighter than the unadjusted main analysis.

The framework selected the meta-regression result. The recommended expected range is **19 – 26 mmol/24h** (Meta-regression, n = 3,974). The dominant caveats are (i) the very high meta-regression R² combined with k = 56, which raises a saturation risk; (ii) the unstable upper limit across sensitivity analyses; and (iii) the borderline assay/country effect, suggesting citrate-style assay variability may also apply here.

##### 9.10.3 Plot

```
phosphate_final_plot <- phosphate_ma_2nd_analysis$main_meta_analysis$plot %+%
  subset(phosphate_ma_2nd_analysis$main_meta_analysis$plot$data,
         quality_filter == "All Studies" & age_band == "Meta-Regression") +
  scale_x_continuous(limits = c(0, 40))

phosphate_final_plot
## `height` was translated to `width`.
```

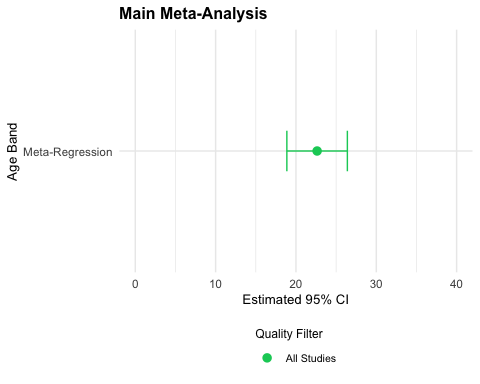

##### 9.10.4 Results Table

```
phosphate_ma_2nd_analysis$ma_results %>%
  filter(
    age_band == "Meta-Regression"
  ) %>% filter(
    sex_group == "main meta-analysis"
  ) %>%
  filter(quality_filter == "All Studies") %>%
  mutate(
    pooled_estimate = round(pooled_estimate, 2),
    pooled_sd       = round(pooled_sd, 2),
    I2              = round((I2 * 100), 0),
    Q               = round(Q, 2),
    Q_pval = case_when(
      Q_pval < 0.001 ~ "<0.001",
      Q_pval < 0.01  ~ "<0.01",
      Q_pval < 0.05  ~ "<0.05",
      TRUE           ~ as.character(round(Q_pval, 2))
    ),
    ci_95 = paste0(
      round(lower_95, 0), " – ", round(upper_95, 0)
    ),
    sex_group = str_to_title(age_band)
  ) %>%
  select(
    sex_group,
    total_participants,
    pooled_estimate,
    pooled_sd,
    ci_95,
    I2,
    Q,
    Q_pval
  ) %>%
  gt() %>%
  cols_label(
    sex_group          = "Analysis",
    total_participants  = "Patients, n",
    pooled_estimate = "Pooled Estimate",
    pooled_sd       = "SD",
    ci_95           = "95% CI",
    I2              = html("I<sup>2</sup> (%)"),
    Q               = "Q statistic",
    Q_pval          = "Q p-value"
  ) %>%
  cols_align(
    align = "center",
    columns = c(total_participants, pooled_estimate, pooled_sd, ci_95, I2, Q, Q_pval)
  ) %>%
  tab_header(
    title = "Phosphate Meta-analysis Results by Age Band",
    subtitle = "Random-effects meta-analysis"
  ) %>%
  tab_source_note(
    source_note = md(
      "*I² quantifies heterogeneity; Q p-value tests heterogeneity across studies.*"
    )
  )
```

Table 1: Phosphate Meta-analysis Results by Age Band

Random-effects meta-analysis

| Analysis | Patients, n | Pooled Estimate | SD | 95% CI | I2 (%) | Q statistic | Q p-value |
| --- | --- | --- | --- | --- | --- | --- | --- |
| Meta-Regression | 3974 | 22.63 | 1.92 | 19 – 26 | 90 | 183.33 | <0.001 |
| *I² quantifies heterogeneity; Q p-value tests heterogeneity across studies.* | | | | | | | |

##### 9.10.5 Bias analysis

```
phosphate_rob <- urine_ma2 %>%
  drop_na(phosphate_sd_mmol_24h) %>%
  left_join(
    overall_bias, by = "study_name"
  ) %>%
  select(study_name, overall, study_type) %>%
  mutate(
    overall = case_when(
      is.na(overall) ~ "Moderate",
      TRUE ~ overall
    )
  ) %>%
  distinct(study_name, .keep_all = TRUE) %>%
  arrange(overall) %>%
  gt() %>%
  cols_label(
    study_name = "Study",
    overall    = "Risk of Bias",
    study_type = "Study Type"
  ) %>%
  data_color(
    columns = overall,
    fn = function(x) {
      case_when(
        x == "Low"      ~ "#90EE90",
        x == "Moderate" ~ "#FFD700",
        x == "High"     ~ "#FF6B6B",
        TRUE            ~ "#FFFFFF"
      )
    }
  ) %>%
  opt_all_caps() %>%
  cols_align(
    align = "center",
    overall
  ) %>%
  tab_header(
    title = "Risk of Bias by Study and Age Band for 24 Hour Urine Phosphate"
  )

phosphate_rob
```

Table 1: Risk of Bias by Study and Age Band for 24 Hour Urine Phosphate

| Study | Risk of Bias | Study Type |
| --- | --- | --- |
| Arora et al. 2017 | High | Observational Cohort |
| Grases et al. 2015 | High | Observational Cohort |
| Nouvenne et al. 2013 | High | Observational Cohort |
| Matsumoto et al. 2006 | High | RCT |
| Stitchantrakul et al. 2004 | High | RCT |
| Allie et al. 2003 | High | RCT |
| Hess et al. 1999 | High | Observational Cohort |
| Thongprayoon et al. 2022 | Low | Observational Cohort |
| Deng et al. 2017 | Low | Observational Cohort |
| Parvin et al. 2011 | Low | Observational Cohort |
| Maalouf et al. 2010 | Low | Observational Cohort |
| Traxer et al. 2003 | Low | RCT |
| Mai et al. 2019 | Low | Observational Cohort |
| Elkoushy et al. 2014 | Moderate | Observational Cohort |
| Lange et al. 2014 | Moderate | RCT |
| Pigna et al. 2014 | Moderate | Observational Cohort |
| Herrel et al. 2012 | Moderate | RCT |
| Hong et al. 2012 | Moderate | Observational Cohort |
| Sorensen et al. 2012 | Moderate | Observational Cohort |
| Stitchantrakul et al. 2007 | Moderate | Observational Cohort |
| Gettman et al. 2005 | Moderate | Observational Cohort |
| Lemann et al. 1996 | Moderate | Observational Cohort |
| HPFS et al. 40-50 | Moderate | Observational Cohort |
| HPFS et al. 50-60 | Moderate | Observational Cohort |
| HPFS et al. 60-70 | Moderate | Observational Cohort |
| HPFS et al. 70-80 | Moderate | Observational Cohort |
| NHS et al. 40-50 | Moderate | Observational Cohort |
| NHS et al. 50-60 | Moderate | Observational Cohort |
| NHS et al. 60-70 | Moderate | Observational Cohort |
| NHS et al. 70-80 | Moderate | Observational Cohort |

##### 9.10.6 24 Hour Urine Phosphate Suggested Expected Range inc. GRADE

```
phosphate_grade <- calculate_grade(
  study_design= "RCT", # Overall result similar to RCT
  risk_of_bias = "Moderate", # Studies with serious risk of bias included
  inconsistency = "Serious", # marked variation between sensitivity analyses
  indirectness = "Moderate",
  imprecision = "Serious", # marked variation between sensitivity analyses suggests imprecision
  publication_bias = "Suspected", # Cochran's Q significant
  large_effect = TRUE,
  dose_response = FALSE,
  confounders_reduce_effect = mr_reduces_effect(phosphate_ma_2nd_analysis$ma_results)
)

phosphate_grade <- cbind(
  sex = c("Overall"),
  grade = phosphate_grade$grade_rating
) %>% as_tibble() %>%
  mutate(
    grade_rating = grade_symbol(grade)
  )

phosphate_final_results <- get_final_ma_results(phosphate_ma_2nd_analysis$ma_results, sex_specific = FALSE, meta_regression = TRUE, low_rob = FALSE) %>%
  mutate(
    pooled_estimate = round(pooled_estimate, 2),
    pooled_sd       = round(pooled_sd, 2),
    Q               = round(Q, 2),
    Q_pval = case_when(
      Q_pval < 0.001 ~ "<0.001",
      Q_pval < 0.01  ~ "<0.01",
      Q_pval < 0.05  ~ "<0.05",
      TRUE           ~ sprintf("%.2f", Q_pval)
    ),
    ci_95 = paste0(
      round(lower_95, 0), " – ", round(upper_95, 0)
    )
    # age_band stays as "Meta-Regression" (set by get_final_ma_results)
  ) %>%
  select(
    age_band,
    total_participants,
    ci_95,
    Q,
    Q_pval
  ) %>%
  cbind(
    phosphate_grade) %>%
  mutate(grade_rating = paste0(grade, " ", grade_rating)) %>%
  select(-c(sex, grade))

phosphate_final_results %>%
  gt() %>%
  cols_label(
    age_band       = "Sex",
    total_participants  = "Patients, n",
    ci_95           = "95% CI",
    Q               = "Q statistic",
    Q_pval          = "Q p-value",
    grade_rating    = "GRADE Rating"
  ) %>%
  cols_align(
    align = "center"
  ) %>%
  tab_header(
    title = "Finalised Meta-Analysis Results for phosphate",
    subtitle = "Subdivided by Age Band"
  ) %>%
  tab_source_note(
    source_note = md(
      "*Q p-value = Cochran's Q heterogeneity test; RoB = Risk of Bias*"
    )
  )
```

Table 1: Finalised Meta-Analysis Results for phosphate

Subdivided by Age Band

| Sex | Patients, n | 95% CI | Q statistic | Q p-value | GRADE Rating |
| --- | --- | --- | --- | --- | --- |
| Meta-Regression | 3974 | 19 – 26 | 183.33 | <0.001 | Low ⊕⊕◯◯ |
| *Q p-value = Cochran's Q heterogeneity test; RoB = Risk of Bias* | | | | | |

```
phosphate_final_results <- phosphate_final_results %>%
  mutate(
    urine_component = "phosphate"
  )
```

#### 9.11 Sodium

##### 9.11.1 Framework Recommendation

framework_callout("sodium")

Table 1: **Framework Recommendation — sodium**

Branch 6 — Meta-regression effective

| Component | Recommended | Branch | Reason | k | n | R² | Sex p (women_prop / overall / MR / LRoB) | Age p (min) | race p (min) | Assay p (min) | Egger's p | Q/df (chosen / overall) | Q/df ratio (chosen÷overall) | Q p (chosen) | MR R² (chosen) | QC warnings |
| --- | --- | --- | --- | --- | --- | --- | --- | --- | --- | --- | --- | --- | --- | --- | --- | --- |
| sodium | Meta-regression | 6 — Meta-regression effective | R² = 11.5% ≥ 10% with k = 94 ≥ 20 | 94 | 9,120 | 11.5% | 0.657 / 0.599 / 0.400 / 0.871 | 0.716 | 0.815 | 0.002 | 0.219 | 18.6 / 85.1 | 0.22 | <0.001 | 11% | — |

##### 9.11.2 Comment

For 24-hour urine sodium, the meta-analysis included k = 94 subgroup-level records and n = 9,120 participants. Heterogeneity was substantial across all analyses (I² > 95%, Cochran’s Q p < 0.001).

Sex differences on meta-regression were not significant (p = 0.657; overall sex Wald p = 0.599) — only the 30 – 40 age band demonstrated a significant sex difference, with all other age-banded sex tests non-significant. Age comparisons across powered bands gave a minimum pairwise Wald p of 0.716. race was underpowered (minimum Wald p = 0.815). The assay/country stratum test reached significance (minimum Wald p = 0.002), but the relative shift between categories did not pass the 20% magnitude floor required by the assay branch, so the assay branch did not fire.

Meta-regression explained R² = 11.5% of heterogeneity with k = 94, just above the framework’s 10% threshold — the meta-regression branch was satisfied. Egger’s test was not significant (p = 0.219). Additionally, the meta-regression-adjusted pooled estimate differed from the unadjusted overall by ~15% (above the 5% margin used by the no-stratum-effect override), confirming that moderator adjustment meaningfully changes the result and further supporting the meta-regression result.

Reductions in heterogeneity in specific scenarios (e.g. males aged 40 – 50) are likely artefacts of underpowered analyses. Sex-specific ranges are at high risk of bias from the imbalanced subgroups. The low risk-of-bias sub-analysis was consistent with the meta-regression result.

The framework selected the meta-regression result. The recommended expected range is **98 – 253 mmol/24h** (Meta-regression, n = 3,945). Sodium intake is highly variable culturally and dietarily, which is consistent with the residual heterogeneity even after meta-regression adjustment; the wide expected range reflects this real biological variability.

##### 9.11.3 Plot

```
sodium_final_plot <- sodium_ma_2nd_analysis$main_meta_analysis$plot %+%
  subset(sodium_ma_2nd_analysis$main_meta_analysis$plot$data, age_band == "Meta-Regression" & quality_filter == "All Studies") +
  scale_x_continuous(limits = c(50,270))

sodium_final_plot
## `height` was translated to `width`.
```

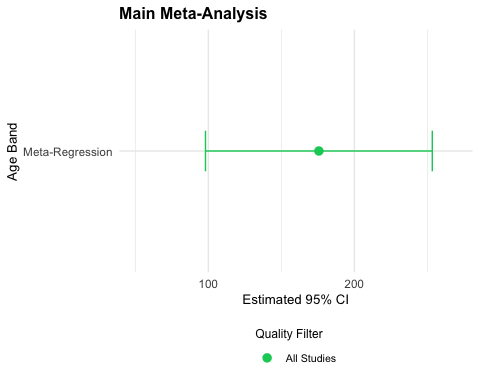

##### 9.11.4 Results Table

```
sodium_ma_2nd_analysis$ma_results %>%
  filter(
    age_band %in% c("Meta-Regression")
  ) %>% filter(
    sex_group == "main meta-analysis"
  ) %>% filter(
    quality_filter == "All Studies"
  ) %>%
  mutate(
    pooled_estimate = round(pooled_estimate, 2),
    pooled_sd       = round(pooled_sd, 2),
    I2              = round((I2 * 100), 0),
    Q               = round(Q, 2),
    Q_pval = case_when(
      Q_pval < 0.001 ~ "<0.001",
      Q_pval < 0.01  ~ "<0.01",
      Q_pval < 0.05  ~ "<0.05",
      TRUE           ~ as.character(round(Q_pval, 2))
    ),
    ci_95 = paste0(
      round(lower_95, 0), " – ", round(upper_95, 0)
    ),
    sex_group = str_to_title(age_band)
  ) %>%
  select(
    sex_group,
    total_participants,
    pooled_estimate,
    pooled_sd,
    ci_95,
    I2,
    Q,
    Q_pval
  ) %>%
  gt() %>%
  cols_label(
    sex_group          = "Sex",
    total_participants  = "Patients, n",
    pooled_estimate = "Pooled Estimate",
    pooled_sd       = "SD",
    ci_95           = "95% CI",
    I2              = html("I<sup>2</sup> (%)"),
    Q               = "Q statistic",
    Q_pval          = "Q p-value"
  ) %>%
  cols_align(
    align = "center",
    columns = c(total_participants, pooled_estimate, pooled_sd, ci_95, I2, Q, Q_pval)
  ) %>%
  tab_header(
    title = "Sodium Meta-analysis Results by Age Band",
    subtitle = "Random-effects meta-analysis"
  ) %>%
  tab_source_note(
    source_note = md(
      "*I² quantifies heterogeneity; Q p-value tests heterogeneity across studies.*"
    )
  )
```

Table 1: Sodium Meta-analysis Results by Age Band

Random-effects meta-analysis

| Sex | Patients, n | Pooled Estimate | SD | 95% CI | I2 (%) | Q statistic | Q p-value |
| --- | --- | --- | --- | --- | --- | --- | --- |
| Meta-Regression | 3945 | 175.7 | 39.61 | 98 – 253 | 95 | 857.39 | <0.001 |
| *I² quantifies heterogeneity; Q p-value tests heterogeneity across studies.* | | | | | | | |

##### 9.11.5 Bias analysis

```
sodium_rob <- urine_ma2 %>%
  drop_na(sodium_sd_mmol_24h) %>%
  left_join(
    overall_bias, by = "study_name"
  ) %>%
  select(study_name, overall, study_type) %>%
  mutate(
    overall = case_when(
      is.na(overall) ~ "Moderate",
      TRUE ~ overall
    )
  ) %>%
  distinct(study_name, .keep_all = TRUE) %>%
  arrange(overall) %>%
  gt() %>%
  cols_label(
    study_name = "Study",
    overall    = "Risk of Bias",
    study_type = "Study Type"
  ) %>%
  data_color(
    columns = overall,
    fn = function(x) {
      case_when(
        x == "Low"      ~ "#90EE90",
        x == "Moderate" ~ "#FFD700",
        x == "High"     ~ "#FF6B6B",
        TRUE            ~ "#FFFFFF"
      )
    }
  ) %>%
  opt_all_caps() %>%
  cols_align(
    align = "center",
    overall
  ) %>%
  tab_header(
    title = "Risk of Bias by Study and Age Band for 24 Hour Urine Sodium"
  )

sodium_rob
```

Table 1: Risk of Bias by Study and Age Band for 24 Hour Urine Sodium

| Study | Risk of Bias | Study Type |
| --- | --- | --- |
| Arora et al. 2017 | High | Observational Cohort |
| Nouvenne et al. 2013 | High | Observational Cohort |
| Arrabal-Polo et al. 2012 | High | Observational Cohort |
| Sweeney et al. 2009 | High | RCT |
| Matsumoto et al. 2006 | High | RCT |
| Abate et al. 2004 | High | Observational Cohort |
| Stitchantrakul et al. 2004 | High | RCT |
| Allie et al. 2003 | High | RCT |
| Baxmann et al. 2003 | High | RCT |
| Hess et al. 1999 | High | Observational Cohort |
| Christie et al. 1996 | High | Observational Cohort |
| Thongprayoon et al. 2022 | Low | Observational Cohort |
| Deng et al. 2017 | Low | Observational Cohort |
| Parvin et al. 2011 | Low | Observational Cohort |
| Maalouf et al. 2010 | Low | Observational Cohort |
| Traxer et al. 2003 | Low | RCT |
| Mai et al. 2019 | Low | Observational Cohort |
| Doizi et al. 2016 | Low | Observational Cohort |
| Doenyas-Barak et al. 2015 | Moderate | Observational Cohort |
| Lange et al. 2014 | Moderate | RCT |
| Pigna et al. 2014 | Moderate | Observational Cohort |
| Herrel et al. 2012 | Moderate | RCT |
| Hong et al. 2012 | Moderate | Observational Cohort |
| Sorensen et al. 2012 | Moderate | Observational Cohort |
| Duffey et al. 2011 | Moderate | Observational Cohort |
| Rodgers et al. 2009 | Moderate | Observational Cohort |
| Stitchantrakul et al. 2007 | Moderate | Observational Cohort |
| Gettman et al. 2005 | Moderate | Observational Cohort |
| Lemann et al. 1996 | Moderate | Observational Cohort |
| Wabner et al. 1992 | Moderate | RCT |
| Welshman et al. 1975 | Moderate | Observational Cohort |
| Matin and Streem, 2000 | Moderate | Observational Cohort |
| Baric et al. 2019 | Moderate | Observational Cohort |
| Bingham et al. 1988 | Moderate | Observational Cohort |
| Rodgers et al. 2015 | Moderate | Observational Cohort |
| Hesse et al. 1986 | Moderate | Observational Cohort |
| Athanasatou et al. 2018 | Moderate | Observational Cohort |
| Cao et al. 2023 | Moderate | Observational Cohort |
| HPFS et al. 40-50 | Moderate | Observational Cohort |
| HPFS et al. 50-60 | Moderate | Observational Cohort |
| HPFS et al. 60-70 | Moderate | Observational Cohort |
| HPFS et al. 70-80 | Moderate | Observational Cohort |
| NHS et al. 40-50 | Moderate | Observational Cohort |
| NHS et al. 50-60 | Moderate | Observational Cohort |
| NHS et al. 60-70 | Moderate | Observational Cohort |
| NHS et al. 70-80 | Moderate | Observational Cohort |

##### 9.11.6 24 Hour Urine Sodium Suggested Expected Range inc. GRADE

```
sodium_grade <- calculate_grade(
  study_design= "Mixed", # Mix of Observational + RCTs
  risk_of_bias = "Not Serious", # Low ROB subanalysis used
  inconsistency = "Serious", # Inconsistency across analyses
  indirectness = "Moderate",
  imprecision = "Not Serious", # Good level of precision
  publication_bias = "Suspected", # Significant cochran's Q
  large_effect = TRUE,
  dose_response = FALSE,
  confounders_reduce_effect = mr_reduces_effect(sodium_ma_2nd_analysis$ma_results)
)

sodium_grade <- sodium_grade %>%
  as_tibble() %>%
  mutate(
    sex = "Overall",
    grade = grade_rating,
    grade_rating = grade_symbol(grade)
  )

sodium_final_results <- get_final_ma_results(sodium_ma_2nd_analysis$ma_results, sex_specific = FALSE, low_rob = FALSE, meta_regression = TRUE) %>%
  mutate(
    pooled_estimate = round(pooled_estimate, 2),
    pooled_sd       = round(pooled_sd, 2),
    Q               = round(Q, 2),
    Q_pval = case_when(
      Q_pval < 0.001 ~ "<0.001",
      Q_pval < 0.01  ~ "<0.01",
      Q_pval < 0.05  ~ "<0.05",
      TRUE           ~ sprintf("%.2f", Q_pval)
    ),
    ci_95 = paste0(
      round(lower_95, 0), " – ", round(upper_95, 0)
    )
  ) %>%
  select(
    age_band,
    total_participants,
    ci_95,
    Q,
    Q_pval
  ) %>%
  cbind(
    sodium_grade) %>%
  mutate(grade_rating = paste0(grade, " ", grade_rating)) %>%
  select(-c(sex, certainty_score, grade))

sodium_final_results %>%
  gt() %>%
  cols_label(
    age_band       = "Sex",
    total_participants  = "Patients, n",
    ci_95           = "95% CI",
    Q               = "Q statistic",
    Q_pval          = "Q p-value",
    grade_rating    = "GRADE Rating"
  ) %>%
  cols_align(
    align = "center"
  ) %>%
  tab_header(
    title = "Finalised Meta-Analysis Results for Sodium",
    subtitle = "Subdivided by Age Band"
  ) %>%
  tab_source_note(
    source_note = md(
      "*Q p-value = Cochran's Q heterogeneity test; RoB = Risk of Bias*"
    )
  )
```

Table 1: Finalised Meta-Analysis Results for Sodium

Subdivided by Age Band

| Sex | Patients, n | 95% CI | Q statistic | Q p-value | GRADE Rating |
| --- | --- | --- | --- | --- | --- |
| Meta-Regression | 3945 | 98 – 253 | 857.39 | <0.001 | Moderate ⊕⊕⊕◯ |
| *Q p-value = Cochran's Q heterogeneity test; RoB = Risk of Bias* | | | | | |

```
sodium_final_results <- sodium_final_results %>%
  mutate(
    urine_component = "sodium"
  )
```

#### 9.12 Potassium

##### 9.12.1 Framework Recommendation

framework_callout("potassium")

Table 1: **Framework Recommendation — potassium**

Branch 5 — Robust assay/country difference

| Component | Recommended | Branch | Reason | k | n | R² | Sex p (women_prop / overall / MR / LRoB) | Age p (min) | race p (min) | Assay p (min) | Egger's p | Q/df (chosen / overall) | Q/df ratio (chosen÷overall) | Q p (chosen) | MR R² (chosen) | QC warnings |
| --- | --- | --- | --- | --- | --- | --- | --- | --- | --- | --- | --- | --- | --- | --- | --- | --- |
| potassium | Assay/country-stratified | 5 — Robust assay/country difference | Assay min Wald p = 0.010 (rel shift 44.9%) across 3 named-category pair(s); 1/3 significant after 20% shift filter | 93 | 9,418 | 22.2% | 0.205 / 0.695 / 0.702 / 0.748 | 0.587 | 0.594 | 0.010 | 0.744 | — / 83.1 | — | — | 22% | — |

##### 9.12.2 Comment

For 24-hour urine potassium, the meta-analysis included k = 93 subgroup-level records and n = 9,418 participants. Heterogeneity was high throughout (I² > 95%, Cochran’s Q p < 0.001), with substantial numbers of missing strata.

Sex differences on meta-regression were not significant (p = 0.205; overall sex Wald p = 0.695). Age comparisons across powered bands gave a minimum pairwise Wald p of 0.587. race was underpowered (minimum Wald p = 0.594) and mainly limited to single studies. A significant between-assay/country effect was identified: minimum pairwise Wald p = 0.010, with 1 of 3 named pairs significant after the 20% relative-shift filter and a relative shift of 44.9% — sufficient to trigger the assay/country branch.

Meta-regression explained R² = 22.2% of heterogeneity with k = 93, comfortably above the framework’s 10% threshold; Egger’s test was not significant (p = 0.744). Under the meta-regression branch in isolation, meta-regression would have been a defensible choice. However, because the assay/country stratum test fires before meta-regression in the framework, the framework proceeded to the assay branch.

The low risk-of-bias and RCT-only sub-analyses were consistent with the main result. Meta-regression slightly reduced intervals and I² but substantial heterogeneity remained, suggesting study-level covariates only partially explain the variability.

The framework selected the assay/country-stratified branch. As with citrate, assay-specific reference ranges are not clinically actionable because clinicians do not generally know which assay method or laboratory produced an individual potassium result. The displayed result is therefore the overall pooled estimate with an explicit assay-variability caveat. The recommended expected range is **28 – 77 mmol/24h** (Overall, with assay-variability caveat, n = 9,418). The wide range reflects substantial cultural and dietary variability in potassium intake; assay variability adds further uncertainty.

##### 9.12.3 Plot

```
potassium_ma_2nd_analysis$main_meta_analysis$plot %+%
  subset(potassium_ma_2nd_analysis$main_meta_analysis$plot$data,
         age_band %in% ("Overall") & quality_filter == "All Studies")
## `height` was translated to `width`.
```

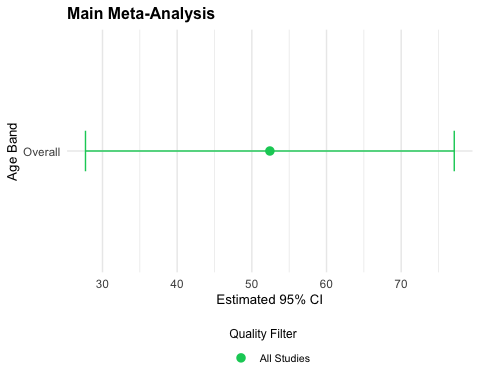

##### 9.12.4 Results Table

```
potassium_ma_2nd_analysis$ma_results %>%
  filter(
    sex_group == "main meta-analysis",
    age_band %in% ("Overall"),
    quality_filter == "All Studies"
  ) %>%
  mutate(
    pooled_estimate = round(pooled_estimate, 2),
    pooled_sd       = round(pooled_sd, 2),
    I2              = round((I2 * 100), 0),
    Q               = round(Q, 2),
    Q_pval = case_when(
      Q_pval < 0.001 ~ "<0.001",
      Q_pval < 0.01  ~ "<0.01",
      Q_pval < 0.05  ~ "<0.05",
      TRUE           ~ as.character(round(Q_pval, 2))
    ),
    ci_95 = paste0(
      round(lower_95, 0), " – ", round(upper_95, 0)
    ),
    sex_group = str_to_title(sex_group)
  ) %>%
  select(
    sex_group,
    total_participants,
    pooled_estimate,
    pooled_sd,
    ci_95,
    I2,
    Q,
    Q_pval
  ) %>%
  gt() %>%
  cols_label(
    sex_group          = "Analysis",
    total_participants  = "Patients, n",
    pooled_estimate = "Pooled Estimate",
    pooled_sd       = "SD",
    ci_95           = "95% CI",
    I2              = html("I<sup>2</sup> (%)"),
    Q               = "Q statistic",
    Q_pval          = "Q p-value"
  ) %>%
  cols_align(
    align = "center",
    columns = c(total_participants, pooled_estimate, pooled_sd, ci_95, I2, Q, Q_pval)
  ) %>%
  tab_header(
    title = "Potassium Meta-analysis Results",
    subtitle = "Random Effects Meta-analysis"
  ) %>%
  tab_source_note(
    source_note = md(
      "*I² quantifies heterogeneity; Q p-value tests heterogeneity across studies.*"
    )
  )
```

Table 1: Potassium Meta-analysis Results

Random Effects Meta-analysis

| Analysis | Patients, n | Pooled Estimate | SD | 95% CI | I2 (%) | Q statistic | Q p-value |
| --- | --- | --- | --- | --- | --- | --- | --- |
| Main Meta-Analysis | 9418 | 52.41 | 12.6 | 28 – 77 | 99 | 7644.37 | <0.001 |
| *I² quantifies heterogeneity; Q p-value tests heterogeneity across studies.* | | | | | | | |

##### 9.12.5 Bias analysis

```
potassium_rob <- urine_ma2 %>%
  drop_na(potassium_sd_mmol_24h) %>%
  left_join(
    overall_bias, by = "study_name"
  ) %>%
  select(study_name, overall, study_type) %>%
  mutate(
    overall = case_when(
      is.na(overall) ~ "Moderate",
      TRUE ~ overall
    )
  ) %>%
  distinct(study_name, .keep_all = TRUE) %>%
  arrange(overall) %>%
  gt() %>%
  cols_label(
    study_name = "Study",
    overall    = "Risk of Bias",
    study_type = "Study Type"
  ) %>%
  data_color(
    columns = overall,
    fn = function(x) {
      case_when(
        x == "Low"      ~ "#90EE90",
        x == "Moderate" ~ "#FFD700",
        x == "High"     ~ "#FF6B6B",
        TRUE            ~ "#FFFFFF"
      )
    }
  ) %>%
  opt_all_caps() %>%
  cols_align(
    align = "center",
    overall
  ) %>%
  tab_header(
    title = "Risk of Bias by Study and Age Band for 24 Hour Urine potassium"
  )

potassium_rob
```

Table 1: Risk of Bias by Study and Age Band for 24 Hour Urine potassium

| Study | Risk of Bias | Study Type |
| --- | --- | --- |
| Arora et al. 2017 | High | Observational Cohort |
| Nouvenne et al. 2013 | High | Observational Cohort |
| Arrabal-Polo et al. 2012 | High | Observational Cohort |
| Sweeney et al. 2009 | High | RCT |
| Matsumoto et al. 2006 | High | RCT |
| Abate et al. 2004 | High | Observational Cohort |
| Stitchantrakul et al. 2004 | High | RCT |
| Allie et al. 2003 | High | RCT |
| Baxmann et al. 2003 | High | RCT |
| Christie et al. 1996 | High | Observational Cohort |
| Thongprayoon et al. 2022 | Low | Observational Cohort |
| Deng et al. 2017 | Low | Observational Cohort |
| Parvin et al. 2011 | Low | Observational Cohort |
| Maalouf et al. 2010 | Low | Observational Cohort |
| Traxer et al. 2003 | Low | RCT |
| Mai et al. 2019 | Low | Observational Cohort |
| Doizi et al. 2016 | Low | Observational Cohort |
| Doenyas-Barak et al. 2015 | Moderate | Observational Cohort |
| Lange et al. 2014 | Moderate | RCT |
| Pigna et al. 2014 | Moderate | Observational Cohort |
| Herrel et al. 2012 | Moderate | RCT |
| Hong et al. 2012 | Moderate | Observational Cohort |
| Rodgers et al. 2009 | Moderate | Observational Cohort |
| Stitchantrakul et al. 2007 | Moderate | Observational Cohort |
| Cameron et al. 2006 | Moderate | Observational Cohort |
| Lemann et al. 1996 | Moderate | Observational Cohort |
| Wabner et al. 1992 | Moderate | RCT |
| Welshman et al. 1975 | Moderate | Observational Cohort |
| Baric et al. 2019 | Moderate | Observational Cohort |
| Bingham et al. 1988 | Moderate | Observational Cohort |
| Rodgers et al. 2015 | Moderate | Observational Cohort |
| Rodríguez-Rodríguez et al. 2015 | Moderate | Observational Cohort |
| Hesse et al. 1986 | Moderate | Observational Cohort |
| Athanasatou et al. 2018 | Moderate | Observational Cohort |
| HPFS et al. 40-50 | Moderate | Observational Cohort |
| HPFS et al. 50-60 | Moderate | Observational Cohort |
| HPFS et al. 60-70 | Moderate | Observational Cohort |
| HPFS et al. 70-80 | Moderate | Observational Cohort |
| NHS et al. 40-50 | Moderate | Observational Cohort |
| NHS et al. 50-60 | Moderate | Observational Cohort |
| NHS et al. 60-70 | Moderate | Observational Cohort |
| NHS et al. 70-80 | Moderate | Observational Cohort |

##### 9.12.6 24 Hour Urine Potassium Suggested Expected Range inc. GRADE

```
potassium_grade <- calculate_grade(
  study_design= "Mixed",
  risk_of_bias = "Moderate", # Low + Low/Moderate ROB subanalyses aren't dissimilar to MR result
  inconsistency = "Serious",
  indirectness = "Serious",
  imprecision = "Serious", # assay differences detected
  publication_bias = "Strongly suspected", # Significant Cochran's Q
  large_effect = TRUE,
  dose_response = FALSE,
  confounders_reduce_effect = mr_reduces_effect(potassium_ma_2nd_analysis$ma_results)
)

potassium_grade <- cbind(
  sex = c("Overall"),
  grade = potassium_grade$grade_rating
) %>% as_tibble() %>%
  mutate(
    grade_rating = grade_symbol(grade)
  )

potassium_final_results <- get_final_ma_results(potassium_ma_2nd_analysis$ma_results, sex_specific = FALSE, meta_regression = FALSE, low_rob = FALSE) %>%
  mutate(
    pooled_estimate = round(pooled_estimate, 2),
    pooled_sd       = round(pooled_sd, 2),
    Q               = round(Q, 2),
    Q_pval = case_when(
      Q_pval < 0.001 ~ "<0.001",
      Q_pval < 0.01  ~ "<0.01",
      Q_pval < 0.05  ~ "<0.05",
      TRUE           ~ sprintf("%.2f", Q_pval)
    ),
    ci_95 = paste0(
      round(lower_95, 0), " – ", round(upper_95, 0)
    )
  ) %>%
  select(
    age_band,
    total_participants,
    ci_95,
    Q,
    Q_pval
  ) %>%
  cbind(
    potassium_grade) %>%
  mutate(grade_rating = paste0(grade, " ", grade_rating)) %>%
  select(-c(sex, grade))

potassium_final_results %>%
  gt() %>%
  cols_label(
    age_band       = "Analysis",
    total_participants  = "Patients, n",
    ci_95           = "95% CI",
    Q               = "Q statistic",
    Q_pval          = "Q p-value",
    grade_rating    = "GRADE Rating"
  ) %>%
  cols_align(
    align = "center"
  ) %>%
  tab_header(
    title = "Finalised Meta-Analysis Results for Potassium",
    subtitle = "Subdivided by Age Band"
  ) %>%
  tab_source_note(
    source_note = md(
      "*Q p-value = Cochran's Q heterogeneity test; RoB = Risk of Bias*"
    )
  )
```

Table 1: Finalised Meta-Analysis Results for Potassium

Subdivided by Age Band

| Analysis | Patients, n | 95% CI | Q statistic | Q p-value | GRADE Rating |
| --- | --- | --- | --- | --- | --- |
| Overall | 9418 | 28 – 77 | 7644.37 | <0.001 | Very Low ⊕◯◯◯ |
| *Q p-value = Cochran's Q heterogeneity test; RoB = Risk of Bias* | | | | | |

```
potassium_final_results <- potassium_final_results %>%
  mutate(
    urine_component = "potassium"
  )
```

#### 9.13 Magnesium

##### 9.13.1 Framework Recommendation

framework_callout("magnesium")

Table 1: **Framework Recommendation — magnesium**

Branch 6 — Meta-regression effective

| Component | Recommended | Branch | Reason | k | n | R² | Sex p (women_prop / overall / MR / LRoB) | Age p (min) | race p (min) | Assay p (min) | Egger's p | Q/df (chosen / overall) | Q/df ratio (chosen÷overall) | Q p (chosen) | MR R² (chosen) | QC warnings |
| --- | --- | --- | --- | --- | --- | --- | --- | --- | --- | --- | --- | --- | --- | --- | --- | --- |
| magnesium | Meta-regression | 6 — Meta-regression effective | R² = 17.3% ≥ 10% with k = 87 ≥ 20 | 87 | 9,513 | 17.3% | 0.421 / 0.585 / 0.940 / 0.982 | 0.571 | 0.793 | 0.363 | 0.359 | 6.4 / 111.6 | 0.06 | <0.001 | 17% | — |

##### 9.13.2 Comment

For 24-hour urine magnesium, the meta-analysis included k = 87 subgroup-level records and n = 9,513 participants. Heterogeneity was high throughout (I² > 95%, Cochran’s Q p < 0.001), with substantial — albeit non-significant — variation between strata.

Sex differences on meta-regression were not significant (p = 0.421; overall sex Wald p = 0.585). Age comparisons across powered bands gave a minimum pairwise Wald p of 0.571 with no clear age-related fluctuation. race was underpowered (minimum Wald p = 0.793). The assay/country stratum test was not significant (minimum Wald p = 0.363).

Meta-regression explained R² = 17.3% of heterogeneity with k = 87, above the framework’s 10% threshold — under the framework algorithm the meta-regression branch is satisfied. Egger’s test was not significant (p = 0.359).

The low risk-of-bias overall sub-analysis was consistent with the RCT-only sub-analysis and aligned with the meta-regression result.

The framework selected the meta-regression result. The recommended expected range is **3.1 – 4.8 mmol/24h** (Meta-Regression, n = 4,724).

##### 9.13.3 Plot

```
magnesium_ma_2nd_analysis$main_meta_analysis$plot %+%
  subset(magnesium_ma_2nd_analysis$main_meta_analysis$plot$data,
         age_band %in% ("Meta-Regression"))
## `height` was translated to `width`.
```

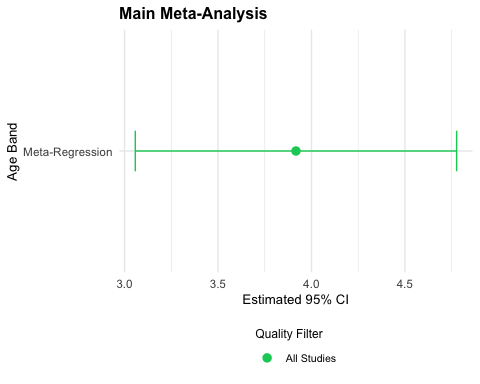

##### 9.13.4 Results Table

```
magnesium_ma_2nd_analysis$ma_results %>%
  filter(
    sex_group == "main meta-analysis",
    age_band %in% ("Meta-Regression"),
    quality_filter == "All Studies"
  ) %>%
  mutate(
    pooled_estimate = round(pooled_estimate, 2),
    pooled_sd       = round(pooled_sd, 2),
    I2              = round((I2 * 100), 0),
    Q               = round(Q, 2),
    Q_pval = case_when(
      Q_pval < 0.001 ~ "<0.001",
      Q_pval < 0.01  ~ "<0.01",
      Q_pval < 0.05  ~ "<0.05",
      TRUE           ~ as.character(round(Q_pval, 2))
    ),
    ci_95 = paste0(
      round(lower_95, 1), " – ", round(upper_95, 1)
    ),
    sex_group = str_to_title(age_band)
  ) %>%
  select(
    sex_group,
    total_participants,
    pooled_estimate,
    pooled_sd,
    ci_95,
    I2,
    Q,
    Q_pval
  ) %>%
  gt() %>%
  cols_label(
    sex_group          = "Analysis",
    total_participants  = "Patients, n",
    pooled_estimate = "Pooled Estimate",
    pooled_sd       = "SD",
    ci_95           = "95% CI",
    I2              = html("I<sup>2</sup> (%)"),
    Q               = "Q statistic",
    Q_pval          = "Q p-value"
  ) %>%
  cols_align(
    align = "center",
    columns = c(total_participants, pooled_estimate, pooled_sd, ci_95, I2, Q, Q_pval)
  ) %>%
  tab_header(
    title = "Magnesium Meta-analysis Results",
    subtitle = "Meta-Regression"
  ) %>%
  tab_source_note(
    source_note = md(
      "*I² quantifies heterogeneity; Q p-value tests heterogeneity across studies.*"
    )
  )
```

Table 1: Magnesium Meta-analysis Results

Meta-Regression

| Analysis | Patients, n | Pooled Estimate | SD | 95% CI | I2 (%) | Q statistic | Q p-value |
| --- | --- | --- | --- | --- | --- | --- | --- |
| Meta-Regression | 4724 | 3.92 | 0.44 | 3.1 – 4.8 | 89 | 306 | <0.001 |
| *I² quantifies heterogeneity; Q p-value tests heterogeneity across studies.* | | | | | | | |

##### 9.13.5 Bias analysis

```
magnesium_rob <- urine_ma2 %>%
  drop_na(magnesium_sd_mmol_24h) %>%
  left_join(
    overall_bias, by = "study_name"
  ) %>%
  select(study_name, overall, study_type) %>%
  mutate(
    overall = case_when(
      is.na(overall) ~ "Moderate",
      TRUE ~ overall
    )
  ) %>%
  distinct(study_name, .keep_all = TRUE) %>%
  arrange(overall) %>%
  gt() %>%
  cols_label(
    study_name = "Study",
    overall    = "Risk of Bias",
    study_type = "Study Type"
  ) %>%
  data_color(
    columns = overall,
    fn = function(x) {
      case_when(
        x == "Low"      ~ "#90EE90",
        x == "Moderate" ~ "#FFD700",
        x == "High"     ~ "#FF6B6B",
        TRUE            ~ "#FFFFFF"
      )
    }
  ) %>%
  opt_all_caps() %>%
  cols_align(
    align = "center",
    overall
  ) %>%
  tab_header(
    title = "Risk of Bias by Study and Age Band for 24 Hour Urine Magnesium"
  )

magnesium_rob
```

Table 1: Risk of Bias by Study and Age Band for 24 Hour Urine Magnesium

| Study | Risk of Bias | Study Type |
| --- | --- | --- |
| Arora et al. 2017 | High | Observational Cohort |
| Grases et al. 2015 | High | Observational Cohort |
| Nouvenne et al. 2013 | High | Observational Cohort |
| Arrabal-Polo et al. 2012 | High | Observational Cohort |
| Sweeney et al. 2009 | High | RCT |
| Matsumoto et al. 2006 | High | RCT |
| Stitchantrakul et al. 2004 | High | RCT |
| Allie et al. 2003 | High | RCT |
| Baxmann et al. 2003 | High | RCT |
| Ogawa et al. 2003 | High | Observational Cohort |
| Terris et al. 2001 | High | Observational Cohort |
| Hess et al. 1999 | High | Observational Cohort |
| Rodgers et al. 1997 | High | Observational Cohort |
| Thongprayoon et al. 2022 | Low | Observational Cohort |
| Deng et al. 2017 | Low | Observational Cohort |
| Parvin et al. 2011 | Low | Observational Cohort |
| Maalouf et al. 2010 | Low | Observational Cohort |
| Traxer et al. 2003 | Low | RCT |
| Mai et al. 2019 | Low | Observational Cohort |
| Doizi et al. 2016 | Low | Observational Cohort |
| Perinpam et al. 2015 | Low | Observational Cohort |
| Lange et al. 2014 | Moderate | RCT |
| Herrel et al. 2012 | Moderate | RCT |
| Hong et al. 2012 | Moderate | Observational Cohort |
| Duffey et al. 2011 | Moderate | Observational Cohort |
| Rodgers et al. 2009 | Moderate | Observational Cohort |
| Stitchantrakul et al. 2007 | Moderate | Observational Cohort |
| Huang et al. 2003 | Moderate | Observational Cohort |
| Miyake et al. 2001 | Moderate | Observational Cohort |
| Lemann et al. 1996 | Moderate | Observational Cohort |
| Wabner et al. 1992 | Moderate | RCT |
| Welshman et al. 1975 | Moderate | Observational Cohort |
| Rodgers et al. 2015 | Moderate | Observational Cohort |
| Hesse et al. 1986 | Moderate | Observational Cohort |
| Cao et al. 2023 | Moderate | Observational Cohort |
| HPFS et al. 40-50 | Moderate | Observational Cohort |
| HPFS et al. 50-60 | Moderate | Observational Cohort |
| HPFS et al. 60-70 | Moderate | Observational Cohort |
| HPFS et al. 70-80 | Moderate | Observational Cohort |
| NHS et al. 40-50 | Moderate | Observational Cohort |
| NHS et al. 50-60 | Moderate | Observational Cohort |
| NHS et al. 60-70 | Moderate | Observational Cohort |
| NHS et al. 70-80 | Moderate | Observational Cohort |

##### 9.13.6 24 Hour Urine Magnesium Suggested Expected Range inc. GRADE

```
magnesium_grade <- calculate_grade(
  study_design= "RCT", #RCTs have similar ranges to MR result
  risk_of_bias = "Moderate", #consistent results + replicated in Low RoB subanalyses
  inconsistency = "Moderate",
  indirectness = "Moderate",
  imprecision = "Moderate",
  publication_bias = "Strongly suspected", #significant Q
  large_effect = TRUE,
  dose_response = FALSE,
  confounders_reduce_effect = mr_reduces_effect(magnesium_ma_2nd_analysis$ma_results)
)

magnesium_grade <- cbind(
  sex = c("Overall"),
  grade = magnesium_grade$grade_rating
) %>% as_tibble() %>%
  mutate(
    grade_rating = grade_symbol(grade)
  )

magnesium_final_results <- get_final_ma_results(magnesium_ma_2nd_analysis$ma_results, sex_specific = FALSE, meta_regression = TRUE, low_rob = FALSE) %>%
  mutate(
    pooled_estimate = round(pooled_estimate, 2),
    pooled_sd       = round(pooled_sd, 2),
    Q               = round(Q, 2),
    Q_pval = case_when(
      Q_pval < 0.001 ~ "<0.001",
      Q_pval < 0.01  ~ "<0.01",
      Q_pval < 0.05  ~ "<0.05",
      TRUE           ~ sprintf("%.2f", Q_pval)
    ),
    ci_95 = paste0(
      round(lower_95, 1), " – ", round(upper_95, 1)
    )
  ) %>%
  select(
    age_band,
    total_participants,
    ci_95,
    Q,
    Q_pval
  ) %>%
  cbind(
    magnesium_grade) %>%
  mutate(grade_rating = paste0(grade, " ", grade_rating)) %>%
  select(-c(sex, grade))

magnesium_final_results %>%
  gt() %>%
  cols_label(
    age_band       = "Sex",
    total_participants  = "Patients, n",
    ci_95           = "95% CI",
    Q               = "Q statistic",
    Q_pval          = "Q p-value",
    grade_rating    = "GRADE Rating"
  ) %>%
  cols_align(
    align = "center"
  ) %>%
  tab_header(
    title = "Finalised Meta-Analysis Results for Magnesium",
    subtitle = "Subdivided by Age Band"
  ) %>%
  tab_source_note(
    source_note = md(
      "*Q p-value = Cochran's Q heterogeneity test; RoB = Risk of Bias*"
    )
  )
```

Table 1: Finalised Meta-Analysis Results for Magnesium

Subdivided by Age Band

| Sex | Patients, n | 95% CI | Q statistic | Q p-value | GRADE Rating |
| --- | --- | --- | --- | --- | --- |
| Meta-Regression | 4724 | 3.1 – 4.8 | 306 | <0.001 | Moderate ⊕⊕⊕◯ |
| *Q p-value = Cochran's Q heterogeneity test; RoB = Risk of Bias* | | | | | |

```
magnesium_final_results <- magnesium_final_results %>%
  mutate(
    urine_component = "magnesium"
  )
```

#### 9.14 Ammonium

##### 9.14.1 Framework Recommendation

framework_callout("ammonium")

Table 1: **Framework Recommendation — ammonium**

Branch 2 — Robust sex difference

| Component | Recommended | Branch | Reason | k | n | R² | Sex p (women_prop / overall / MR / LRoB) | Age p (min) | race p (min) | Assay p (min) | Egger's p | Q/df (chosen / overall) | Q/df ratio (chosen÷overall) | Q p (chosen) | MR R² (chosen) | QC warnings |
| --- | --- | --- | --- | --- | --- | --- | --- | --- | --- | --- | --- | --- | --- | --- | --- | --- |
| ammonium | Sex-stratified | 2 — Robust sex difference | women_prop coef p=0.035 in main meta-regression | 45 | 4,841 | 45.8% | 0.035 / 0.285 / — / — | — | — | — | 0.081 | 53.2 / 62.9 | 0.85 | <0.001 | — | — |

##### 9.14.2 Comment

For 24-hour urine ammonium, the meta-analysis included k = 45 subgroup-level records and n = 4,841 participants. Heterogeneity remained high across analyses (I² > 90%, Cochran’s Q p < 0.001), but with reasonable consistency across strata.

Significant sex differences were detected on meta-regression (p = 0.035). The overall sex Wald test was non-significant (p = 0.285). Age comparisons across powered bands were not testable (no powered pairs available); a significant 30 – 40 vs 60+ band-level comparison was detected but only in men, where the sample size is too small to support a stand-alone meta-regression, so that result is not interpretable. race was severely underpowered (no powered pairs). Assay/country stratification was not tested.

Meta-regression explained R² = 45.8% of heterogeneity with k = 45, well above the framework’s 10% threshold. Egger’s test approached but did not reach significance (p = 0.081). Because the sex stratum test is evaluated first in the framework, meta-regression was not reached as the preferred analysis.

Meta-regression on the overall dataset reduced both interval width and I², suggesting at least partial adjustment for between-study variation; the RCT-only and low risk-of-bias sub-analyses were broadly consistent with the sex-stratified main results. An age-related decline is suggested in some sub-analyses but they are underpowered.

The framework selected the sex-stratified main results. The recommended expected ranges are **28 – 46 mmol/24h (male, n = 535)** and **20 – 40 mmol/24h (female, n = 3,857)**. The male cohort is substantially smaller than the female cohort, so the male estimate should be interpreted with caution. No meta-regression is available for the male sub-cohort, which limits the ability to adjust for moderators in men.

##### 9.14.3 Plot

```
ammonium_male_plot <- ammonium_ma_2nd_analysis$male$plot %+%
  subset(ammonium_ma_2nd_analysis$male$plot$data, age_band == "Overall")

ammonium_female_plot <- ammonium_ma_2nd_analysis$female$plot %+%
  subset(ammonium_ma_2nd_analysis$female$plot$data, age_band == "Overall")

ammonium_final_plot <- (ammonium_male_plot / ammonium_female_plot) + plot_layout(axes = "collect")

ammonium_final_plot
## `height` was translated to `width`.
## `height` was translated to `width`.
```

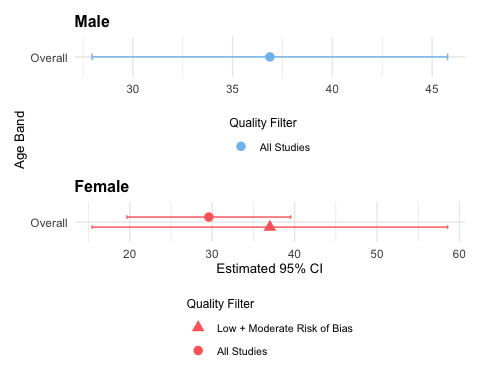

##### 9.14.4 Results Table

```
ammonium_ma_2nd_analysis$ma_results %>%
  filter(
    age_band %in% c("Overall"),
    sex_group != "main meta-analysis",
    quality_filter == "All Studies"
  ) %>%
  mutate(
    pooled_estimate = round(pooled_estimate, 2),
    pooled_sd       = round(pooled_sd, 2),
    I2              = round((I2 * 100), 0),
    Q               = round(Q, 2),
    Q_pval = case_when(
      Q_pval < 0.001 ~ "<0.001",
      Q_pval < 0.01  ~ "<0.01",
      Q_pval < 0.05  ~ "<0.05",
      TRUE           ~ as.character(round(Q_pval, 2))
    ),
    ci_95 = paste0(
      round(lower_95, 0), " – ", round(upper_95, 0)
    ),
    sex_group = str_to_title(sex_group)
  ) %>%
  select(
    sex_group,
    total_participants,
    pooled_estimate,
    pooled_sd,
    ci_95,
    I2,
    Q,
    Q_pval
  ) %>%
  gt() %>%
  cols_label(
    sex_group          = "Sex",
    total_participants  = "Patients, n",
    pooled_estimate = "Pooled Estimate",
    pooled_sd       = "SD",
    ci_95           = "95% CI",
    I2              = html("I<sup>2</sup> (%)"),
    Q               = "Q statistic",
    Q_pval          = "Q p-value"
  ) %>%
  cols_align(
    align = "center",
    columns = c(total_participants, pooled_estimate, pooled_sd, ci_95, I2, Q, Q_pval)
  ) %>%
  tab_header(
    title = "Ammonium Meta-Analysis Results",
    subtitle = "Meta-Regression"
  ) %>%
  tab_source_note(
    source_note = md(
      "*I² quantifies heterogeneity; Q p-value tests heterogeneity across studies.*"
    )
  )
```

Table 1: Ammonium Meta-Analysis Results

Meta-Regression

| Sex | Patients, n | Pooled Estimate | SD | 95% CI | I2 (%) | Q statistic | Q p-value |
| --- | --- | --- | --- | --- | --- | --- | --- |
| Male | 535 | 36.87 | 4.55 | 28 – 46 | 97 | 267.48 | <0.001 |
| Female | 3857 | 29.60 | 5.06 | 20 – 40 | 99 | 1303.78 | <0.001 |
| *I² quantifies heterogeneity; Q p-value tests heterogeneity across studies.* | | | | | | | |

##### 9.14.5 Bias analysis

```
ammonium_rob <- urine_ma2 %>%
  drop_na(ammonium_sd_mmol_24h) %>%
  left_join(
    overall_bias, by = "study_name"
  ) %>%
  select(study_name, overall, study_type) %>%
  mutate(
    overall = case_when(
      is.na(overall) ~ "Moderate",
      TRUE ~ overall
    )
  ) %>%
  distinct(study_name, .keep_all = TRUE) %>%
  arrange(overall) %>%
  gt() %>%
  cols_label(
    study_name = "Study",
    overall    = "Risk of Bias",
    study_type = "Study Type"
  ) %>%
  data_color(
    columns = overall,
    fn = function(x) {
      case_when(
        x == "Low"      ~ "#90EE90",
        x == "Moderate" ~ "#FFD700",
        x == "High"     ~ "#FF6B6B",
        TRUE            ~ "#FFFFFF"
      )
    }
  ) %>%
  opt_all_caps() %>%
  cols_align(
    align = "center",
    overall
  ) %>%
  tab_header(
    title = "Risk of Bias by Study and Age Band for 24 Hour Urine Ammonium"
  )

ammonium_rob
```

Table 1: Risk of Bias by Study and Age Band for 24 Hour Urine Ammonium

| Study | Risk of Bias | Study Type |
| --- | --- | --- |
| Arora et al. 2017 | High | Observational Cohort |
| Nouvenne et al. 2013 | High | Observational Cohort |
| Sweeney et al. 2009 | High | RCT |
| Abate et al. 2004 | High | Observational Cohort |
| Stern et al. 1980 | High | Observational Cohort |
| Maalouf et al. 2010 | Low | Observational Cohort |
| Traxer et al. 2003 | Low | RCT |
| Doizi et al. 2016 | Low | Observational Cohort |
| Pigna et al. 2014 | Moderate | Observational Cohort |
| Cameron et al. 2006 | Moderate | Observational Cohort |
| Lemann et al. 1996 | Moderate | Observational Cohort |
| Wabner et al. 1992 | Moderate | RCT |
| Bingham et al. 1988 | Moderate | Observational Cohort |
| Hesse et al. 1986 | Moderate | Observational Cohort |
| HPFS et al. 40-50 | Moderate | Observational Cohort |
| HPFS et al. 50-60 | Moderate | Observational Cohort |
| HPFS et al. 60-70 | Moderate | Observational Cohort |
| HPFS et al. 70-80 | Moderate | Observational Cohort |
| NHS et al. 40-50 | Moderate | Observational Cohort |
| NHS et al. 50-60 | Moderate | Observational Cohort |
| NHS et al. 60-70 | Moderate | Observational Cohort |
| NHS et al. 70-80 | Moderate | Observational Cohort |

##### 9.14.6 24 Hour Urine ammonium Suggested Expected Range inc. GRADE

```
ammonium_grade <- calculate_grade(
  study_design= "Mixed", # RCTs + Cohorts
  risk_of_bias = "Low", #Low RoB result being used
  inconsistency = "Moderate",
  indirectness = "Moderate",
  imprecision = "Moderate",
  publication_bias = "Strongly suspected", #significant Q
  large_effect = TRUE,
  dose_response = FALSE,
  confounders_reduce_effect = mr_reduces_effect(ammonium_ma_2nd_analysis$ma_results)
)

ammonium_grade <- cbind(
  sex = c("Overall"),
  grade = ammonium_grade$grade_rating
) %>% as_tibble() %>%
  mutate(
    grade_rating = grade_symbol(grade)
  )

ammonium_final_results <- get_final_ma_results(ammonium_ma_2nd_analysis$ma_results, sex_specific = TRUE, low_rob = FALSE) %>%
  mutate(
    pooled_estimate = round(pooled_estimate, 2),
    pooled_sd       = round(pooled_sd, 2),
    Q               = round(Q, 2),
    Q_pval = case_when(
      Q_pval < 0.001 ~ "<0.001",
      Q_pval < 0.01  ~ "<0.01",
      Q_pval < 0.05  ~ "<0.05",
      TRUE           ~ sprintf("%.2f", Q_pval)
    ),
    ci_95 = paste0(
      round(lower_95, 0), " – ", round(upper_95, 0)
    ),
    age_band = str_to_title(sex_group)
  ) %>%
  select(
    age_band,
    total_participants,
    ci_95,
    Q,
    Q_pval
  ) %>%
  cbind(
    ammonium_grade) %>%
  mutate(grade_rating = paste0(grade, " ", grade_rating)) %>%
  select(-c(sex, grade))

ammonium_final_results %>%
  gt() %>%
  cols_label(
    age_band       = "Analysis",
    total_participants  = "Patients, n",
    ci_95           = "95% CI",
    Q               = "Q statistic",
    Q_pval          = "Q p-value",
    grade_rating    = "GRADE Rating"
  ) %>%
  cols_align(
    align = "center"
  ) %>%
  tab_header(
    title = "Finalised Meta-Analysis Results for Ammonium",
    subtitle = "Subdivided by Age Band"
  ) %>%
  tab_source_note(
    source_note = md(
      "*Q p-value = Cochran's Q heterogeneity test; RoB = Risk of Bias*"
    )
  )
```

Table 1: Finalised Meta-Analysis Results for Ammonium

Subdivided by Age Band

| Analysis | Patients, n | 95% CI | Q statistic | Q p-value | GRADE Rating |
| --- | --- | --- | --- | --- | --- |
| Male | 535 | 28 – 46 | 267.48 | <0.001 | Moderate ⊕⊕⊕◯ |
| Female | 3857 | 20 – 40 | 1303.78 | <0.001 | Moderate ⊕⊕⊕◯ |
| *Q p-value = Cochran's Q heterogeneity test; RoB = Risk of Bias* | | | | | |

```
ammonium_final_results <- ammonium_final_results %>%
  mutate(
    urine_component = "ammonium"
  )
```

#### 9.15 Chloride

##### 9.15.1 Framework Recommendation

framework_callout("chloride")

Table 1: **Framework Recommendation — chloride**

Branch 6 — Meta-regression effective

| Component | Recommended | Branch | Reason | k | n | R² | Sex p (women_prop / overall / MR / LRoB) | Age p (min) | race p (min) | Assay p (min) | Egger's p | Q/df (chosen / overall) | Q/df ratio (chosen÷overall) | Q p (chosen) | MR R² (chosen) | QC warnings |
| --- | --- | --- | --- | --- | --- | --- | --- | --- | --- | --- | --- | --- | --- | --- | --- | --- |
| chloride | Meta-regression | 6 — Meta-regression effective | R² = 29.7% ≥ 10% with k = 51 ≥ 20 | 51 | 3,534 | 29.7% | 0.059 / 0.646 / 0.612 / 0.888 | — | — | — | 0.529 | 15.8 / 104.0 | 0.15 | <0.001 | 30% | — |

##### 9.15.2 Comment

For 24-hour urine chloride, the meta-analysis included k = 51 subgroup-level records and n = 3,534 participants. Heterogeneity was substantial throughout (I² > 95%, Cochran’s Q p < 0.001), with minimal data available for age and race stratifications.

Sex differences on meta-regression approached but did not reach significance (p = 0.059; overall sex Wald p = 0.646). Age comparisons were not testable across powered bands (no powered pairs). race was severely underpowered (no powered pairs). Assay/country stratification was not tested.

Meta-regression explained R² = 29.7% of heterogeneity with k = 51 — well above the framework’s 10% threshold — so the meta-regression branch was satisfied. Egger’s test was not significant (p = 0.529). Meta-regression also reduced interval width and I² in both the male and female sub-analyses.

The low risk-of-bias and RCT-only sub-analyses were consistent with the meta-regression result; other stratification analyses remained underpowered.

The framework selected the meta-regression result. The recommended expected range is **84 – 218 mmol/24h** (Meta-regression, n = 3,173). The dominant caveat is the very limited data for stratified analyses, with no powered age or race comparisons available. The wide range reflects the substantial dietary variability of chloride intake.

##### 9.15.3 Plot

```
chloride_ma_2nd_analysis$main_meta_analysis$plot %+%
  subset(chloride_ma_2nd_analysis$main_meta_analysis$plot$data,
         age_band %in% ("Meta-Regression") & quality_filter == "All Studies")
## `height` was translated to `width`.
```

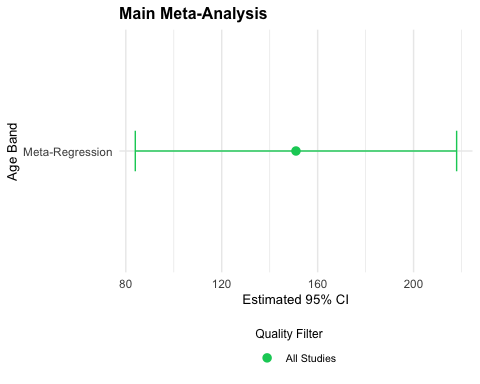

##### 9.15.4 Results Table

```
chloride_ma_2nd_analysis$ma_results %>%
  filter(
    sex_group == "main meta-analysis",
    age_band %in% ("Meta-Regression"),
    quality_filter == "All Studies"
  ) %>%
  mutate(
    pooled_estimate = round(pooled_estimate, 2),
    pooled_sd       = round(pooled_sd, 2),
    I2              = round((I2 * 100), 0),
    Q               = round(Q, 2),
    Q_pval = case_when(
      Q_pval < 0.001 ~ "<0.001",
      Q_pval < 0.01  ~ "<0.01",
      Q_pval < 0.05  ~ "<0.05",
      TRUE           ~ as.character(round(Q_pval, 2))
    ),
    ci_95 = paste0(
      round(lower_95, 0), " – ", round(upper_95, 0)
    ),
    sex_group = "Meta-Regression"
  ) %>%
  select(
    sex_group,
    total_participants,
    pooled_estimate,
    pooled_sd,
    ci_95,
    I2,
    Q,
    Q_pval
  ) %>%
  gt() %>%
  cols_label(
    sex_group          = "Analysis",
    total_participants  = "Patients, n",
    pooled_estimate = "Pooled Estimate",
    pooled_sd       = "SD",
    ci_95           = "95% CI",
    I2              = html("I<sup>2</sup> (%)"),
    Q               = "Q statistic",
    Q_pval          = "Q p-value"
  ) %>%
  cols_align(
    align = "center",
    columns = c(total_participants, pooled_estimate, pooled_sd, ci_95, I2, Q, Q_pval)
  ) %>%
  tab_header(
    title = "Chloride Meta-analysis Results",
    subtitle = "Meta-Regression"
  ) %>%
  tab_source_note(
    source_note = md(
      "*I² quantifies heterogeneity; Q p-value tests heterogeneity across studies.*"
    )
  )
```

Table 1: Chloride Meta-analysis Results

Meta-Regression

| Analysis | Patients, n | Pooled Estimate | SD | 95% CI | I2 (%) | Q statistic | Q p-value |
| --- | --- | --- | --- | --- | --- | --- | --- |
| Meta-Regression | 3173 | 150.95 | 34.18 | 84 – 218 | 94 | 553.32 | <0.001 |
| *I² quantifies heterogeneity; Q p-value tests heterogeneity across studies.* | | | | | | | |

##### 9.15.5 Bias analysis

```
chloride_rob <- urine_ma2 %>%
  drop_na(chloride_sd_mmol_24h) %>%
  left_join(
    overall_bias, by = "study_name"
  ) %>%
  select(study_name, overall, study_type) %>%
  mutate(
    overall = case_when(
      is.na(overall) ~ "Moderate",
      TRUE ~ overall
    )
  ) %>%
  distinct(study_name, .keep_all = TRUE) %>%
  arrange(overall) %>%
  gt() %>%
  cols_label(
    study_name = "Study",
    overall    = "Risk of Bias",
    study_type = "Study Type"
  ) %>%
  data_color(
    columns = overall,
    fn = function(x) {
      case_when(
        x == "Low"      ~ "#90EE90",
        x == "Moderate" ~ "#FFD700",
        x == "High"     ~ "#FF6B6B",
        TRUE            ~ "#FFFFFF"
      )
    }
  ) %>%
  opt_all_caps() %>%
  cols_align(
    align = "center",
    overall
  ) %>%
  tab_header(
    title = "Risk of Bias by Study and Age Band for 24 Hour Urine chloride"
  )

chloride_rob
```

Table 1: Risk of Bias by Study and Age Band for 24 Hour Urine chloride

| Study | Risk of Bias | Study Type |
| --- | --- | --- |
| Arora et al. 2017 | High | Observational Cohort |
| Nouvenne et al. 2013 | High | Observational Cohort |
| Sweeney et al. 2009 | High | RCT |
| Abate et al. 2004 | High | Observational Cohort |
| Stitchantrakul et al. 2004 | High | RCT |
| Allie et al. 2003 | High | RCT |
| Deng et al. 2017 | Low | Observational Cohort |
| Parvin et al. 2011 | Low | Observational Cohort |
| Maalouf et al. 2010 | Low | Observational Cohort |
| Traxer et al. 2003 | Low | RCT |
| Mai et al. 2019 | Low | Observational Cohort |
| Doizi et al. 2016 | Low | Observational Cohort |
| Doenyas-Barak et al. 2015 | Moderate | Observational Cohort |
| Stitchantrakul et al. 2007 | Moderate | Observational Cohort |
| Rodgers et al. 2015 | Moderate | Observational Cohort |
| Hesse et al. 1986 | Moderate | Observational Cohort |

##### 9.15.6 24 Hour Urine chloride Suggested Expected Range inc. GRADE

```
chloride_grade <- calculate_grade(
  study_design= "Mixed", # Mixture of RCTs and Cohorts
  risk_of_bias = "Moderate", #consistent with low RoB analyses
  inconsistency = "Moderate",
  indirectness = "Moderate",
  imprecision = "Moderate",
  publication_bias = "Strongly suspected", #significant Q
  large_effect = TRUE,
  dose_response = FALSE,
  confounders_reduce_effect = mr_reduces_effect(chloride_ma_2nd_analysis$ma_results)
)

chloride_grade <- cbind(
  sex = c("Overall"),
  grade = chloride_grade$grade_rating
) %>% as_tibble() %>%
  mutate(
    grade_rating = grade_symbol(grade)
  )

chloride_final_results <- get_final_ma_results(chloride_ma_2nd_analysis$ma_results, sex_specific = FALSE, meta_regression = TRUE, low_rob = FALSE) %>%
  mutate(
    pooled_estimate = round(pooled_estimate, 2),
    pooled_sd       = round(pooled_sd, 2),
    Q               = round(Q, 2),
    Q_pval = case_when(
      Q_pval < 0.001 ~ "<0.001",
      Q_pval < 0.01  ~ "<0.01",
      Q_pval < 0.05  ~ "<0.05",
      TRUE           ~ sprintf("%.2f", Q_pval)
    ),
    ci_95 = paste0(
      round(lower_95, 0), " – ", round(upper_95, 0)
    )
  ) %>%
  select(
    age_band,
    total_participants,
    ci_95,
    Q,
    Q_pval
  ) %>%
  cbind(
    chloride_grade) %>%
  mutate(grade_rating = paste0(grade, " ", grade_rating)) %>%
  select(-c(sex, grade))

chloride_final_results %>%
  gt() %>%
  cols_label(
    age_band       = "Analysis",
    total_participants  = "Patients, n",
    ci_95           = "95% CI",
    Q               = "Q statistic",
    Q_pval          = "Q p-value",
    grade_rating    = "GRADE Rating"
  ) %>%
  cols_align(
    align = "center"
  ) %>%
  tab_header(
    title = "Finalised Meta-Analysis Results for Chloride",
    subtitle = "Subdivided by Age Band"
  ) %>%
  tab_source_note(
    source_note = md(
      "*Q p-value = Cochran's Q heterogeneity test; RoB = Risk of Bias*"
    )
  )
```

Table 1: Finalised Meta-Analysis Results for Chloride

Subdivided by Age Band

| Analysis | Patients, n | 95% CI | Q statistic | Q p-value | GRADE Rating |
| --- | --- | --- | --- | --- | --- |
| Meta-Regression | 3173 | 84 – 218 | 553.32 | <0.001 | Moderate ⊕⊕⊕◯ |
| *Q p-value = Cochran's Q heterogeneity test; RoB = Risk of Bias* | | | | | |

```
chloride_final_results <- chloride_final_results %>%
  mutate(
    urine_component = "chloride"
  )
```

#### 9.16 Sulphate

##### 9.16.1 Framework Recommendation

framework_callout("sulphate")

Table 1: **Framework Recommendation — sulphate**

Branch 2 — Robust sex difference

| Component | Recommended | Branch | Reason | k | n | R² | Sex p (women_prop / overall / MR / LRoB) | Age p (min) | race p (min) | Assay p (min) | Egger's p | Q/df (chosen / overall) | Q/df ratio (chosen÷overall) | Q p (chosen) | MR R² (chosen) | QC warnings |
| --- | --- | --- | --- | --- | --- | --- | --- | --- | --- | --- | --- | --- | --- | --- | --- | --- |
| sulphate | Sex-stratified | 2 — Robust sex difference | women_prop coef p=<0.001 in main meta-regression | 41 | 4,857 | 95.1% | <0.001 / 0.638 / — / — | — | — | — | 0.415 | 265.0 / 362.4 | 0.73 | <0.001 | — | — |

##### 9.16.2 Comment

For 24-hour urine sulphate, the meta-analysis included k = 41 subgroup-level records and n = 4,857 participants — the second-smallest dataset after urea. Heterogeneity was high throughout (I² > 95%, Cochran’s Q p < 0.001), with marked variation between men and women.

Highly significant sex differences were detected on meta-regression (p < 0.001). The overall sex Wald test was non-significant (p = 0.638) but underpowered. Age comparisons across powered bands were not testable (no powered pairs available). race was severely underpowered (no powered pairs). Assay/country stratification was not tested.

Meta-regression explained R² = 95.1% of heterogeneity with k = 41 — formally above the 10% threshold, but the very high R² combined with modest k raises a saturation/over-fit concern. Egger’s test was not significant (p = 0.415). Because the sex stratum test is evaluated first in the framework, the meta-regression branch was bypassed regardless.

The low risk-of-bias sub-analysis was underpowered but produced ranges within the sex-stratified main results; the RCT-only sub-analysis was similarly consistent.

The framework selected the sex-stratified main results. The recommended expected ranges are **17 – 26 mmol/24h (male, n = 642)** and **7 – 30 mmol/24h (female, n = 3,794)**. The male cohort is substantially smaller than the female cohort, so the male estimate should be interpreted with caution. The high meta-regression R² combined with modest k means that the meta-regression result would not be a safe alternative even though it formally passes the framework threshold.

##### 9.16.3 Plot

```
sulphate_male_plot <- sulphate_ma_2nd_analysis$male$plot %+%
  subset(sulphate_ma_2nd_analysis$male$plot$data, age_band == "Overall")

sulphate_female_plot <- sulphate_ma_2nd_analysis$female$plot %+%
  subset(sulphate_ma_2nd_analysis$female$plot$data, age_band == "Overall")

sulphate_final_plot <- (sulphate_male_plot / sulphate_female_plot) + plot_layout(axes = "collect")

sulphate_final_plot
## `height` was translated to `width`.
## `height` was translated to `width`.
```

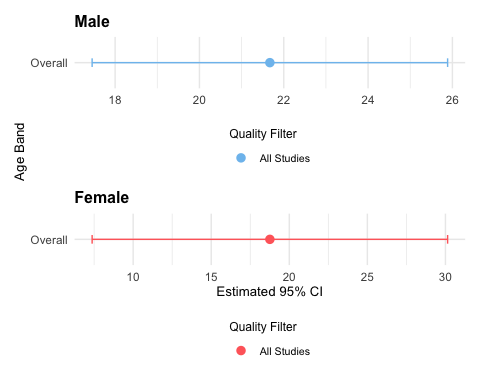

##### 9.16.4 Results Table

```
sulphate_ma_2nd_analysis$ma_results %>%
  filter(
    age_band %in% c("Overall"),
    sex_group != "main meta-analysis",
    quality_filter == "All Studies"
  ) %>%
  mutate(
    pooled_estimate = round(pooled_estimate, 2),
    pooled_sd       = round(pooled_sd, 2),
    I2              = round((I2 * 100), 0),
    Q               = round(Q, 2),
    Q_pval = case_when(
      Q_pval < 0.001 ~ "<0.001",
      Q_pval < 0.01  ~ "<0.01",
      Q_pval < 0.05  ~ "<0.05",
      TRUE           ~ as.character(round(Q_pval, 2))
    ),
    ci_95 = paste0(
      round(lower_95, 0), " – ", round(upper_95, 0)
    ),
    sex_group = str_to_title(sex_group)
  ) %>%
  select(
    sex_group,
    total_participants,
    pooled_estimate,
    pooled_sd,
    ci_95,
    I2,
    Q,
    Q_pval
  ) %>%
  gt() %>%
  cols_label(
    sex_group          = "Sex",
    total_participants  = "Patients, n",
    pooled_estimate = "Pooled Estimate",
    pooled_sd       = "SD",
    ci_95           = "95% CI",
    I2              = html("I<sup>2</sup> (%)"),
    Q               = "Q statistic",
    Q_pval          = "Q p-value"
  ) %>%
  cols_align(
    align = "center",
    columns = c(total_participants, pooled_estimate, pooled_sd, ci_95, I2, Q, Q_pval)
  ) %>%
  tab_header(
    title = "Sulphate Meta-analysis Results",
    subtitle = "Meta-Analysis"
  ) %>%
  tab_source_note(
    source_note = md(
      "*I² quantifies heterogeneity; Q p-value tests heterogeneity across studies.*"
    )
  )
```

Table 1: Sulphate Meta-analysis Results

Meta-Analysis

| Sex | Patients, n | Pooled Estimate | SD | 95% CI | I2 (%) | Q statistic | Q p-value |
| --- | --- | --- | --- | --- | --- | --- | --- |
| Male | 642 | 21.67 | 2.15 | 17 – 26 | 98 | 588.12 | <0.001 |
| Female | 3794 | 18.76 | 5.81 | 7 – 30 | 100 | 7067.01 | <0.001 |
| *I² quantifies heterogeneity; Q p-value tests heterogeneity across studies.* | | | | | | | |

##### 9.16.5 Bias analysis

```
sulphate_rob <- urine_ma2 %>%
  drop_na(sulphate_sd_mmol_24h) %>%
  left_join(
    overall_bias, by = "study_name"
  ) %>%
  select(study_name, overall, study_type) %>%
  mutate(
    overall = case_when(
      is.na(overall) ~ "Moderate",
      TRUE ~ overall
    )
  ) %>%
  distinct(study_name, .keep_all = TRUE) %>%
  arrange(overall) %>%
  gt() %>%
  cols_label(
    study_name = "Study",
    overall    = "Risk of Bias",
    study_type = "Study Type"
  ) %>%
  data_color(
    columns = overall,
    fn = function(x) {
      case_when(
        x == "Low"      ~ "#90EE90",
        x == "Moderate" ~ "#FFD700",
        x == "High"     ~ "#FF6B6B",
        TRUE            ~ "#FFFFFF"
      )
    }
  ) %>%
  opt_all_caps() %>%
  cols_align(
    align = "center",
    overall
  ) %>%
  tab_header(
    title = "Risk of Bias by Study and Age Band for 24 Hour Urine Sulphate"
  )

sulphate_rob
```

Table 1: Risk of Bias by Study and Age Band for 24 Hour Urine Sulphate

| Study | Risk of Bias | Study Type |
| --- | --- | --- |
| Arora et al. 2017 | High | Observational Cohort |
| Nouvenne et al. 2013 | High | Observational Cohort |
| Sweeney et al. 2009 | High | RCT |
| Matsumoto et al. 2006 | High | RCT |
| Abate et al. 2004 | High | Observational Cohort |
| Hess et al. 1999 | High | Observational Cohort |
| Maalouf et al. 2010 | Low | Observational Cohort |
| Traxer et al. 2003 | Low | RCT |
| Pigna et al. 2014 | Moderate | Observational Cohort |
| Cameron et al. 2006 | Moderate | Observational Cohort |
| Lemann et al. 1996 | Moderate | Observational Cohort |
| Hesse et al. 1986 | Moderate | Observational Cohort |
| HPFS et al. 40-50 | Moderate | Observational Cohort |
| HPFS et al. 50-60 | Moderate | Observational Cohort |
| HPFS et al. 60-70 | Moderate | Observational Cohort |
| HPFS et al. 70-80 | Moderate | Observational Cohort |
| NHS et al. 40-50 | Moderate | Observational Cohort |
| NHS et al. 50-60 | Moderate | Observational Cohort |
| NHS et al. 60-70 | Moderate | Observational Cohort |
| NHS et al. 70-80 | Moderate | Observational Cohort |

##### 9.16.6 24 Hour Urine sulphate Suggested Expected Range inc. GRADE

```
sulphate_grade <- calculate_grade(
  study_design= "RCT", #Consistent with RCT results
  risk_of_bias = "Moderate", # Overall result consistent with RoB sub-analyses
  inconsistency = "Strongly Suspected", # Power limited
  indirectness = "Moderate",
  imprecision = "Moderate",
  publication_bias = "Strongly suspected", #significant Q
  large_effect = TRUE,
  dose_response = FALSE,
  confounders_reduce_effect = mr_reduces_effect(sulphate_ma_2nd_analysis$ma_results)
)

sulphate_grade <- cbind(
  sex = c("Overall"),
  grade = sulphate_grade$grade_rating
) %>% as_tibble() %>%
  mutate(
    grade_rating = grade_symbol(grade)
  )

sulphate_final_results <- get_final_ma_results(sulphate_ma_2nd_analysis$ma_results, sex_specific = TRUE) %>%
  mutate(
    pooled_estimate = round(pooled_estimate, 2),
    pooled_sd       = round(pooled_sd, 2),
    Q               = round(Q, 2),
    Q_pval = case_when(
      Q_pval < 0.001 ~ "<0.001",
      Q_pval < 0.01  ~ "<0.01",
      Q_pval < 0.05  ~ "<0.05",
      TRUE           ~ sprintf("%.2f", Q_pval)
    ),
    ci_95 = paste0(
      round(lower_95, 0), " – ", round(upper_95, 0)
    ),
    age_band = str_to_title(sex_group)
  ) %>%
  select(
    age_band,
    total_participants,
    ci_95,
    Q,
    Q_pval
  ) %>%
  cbind(
    sulphate_grade) %>%
  mutate(grade_rating = paste0(grade, " ", grade_rating)) %>%
  select(-c(sex, grade))

sulphate_final_results %>%
  gt() %>%
  cols_label(
    age_band       = "Analysis",
    total_participants  = "Patients, n",
    ci_95           = "95% CI",
    Q               = "Q statistic",
    Q_pval          = "Q p-value",
    grade_rating    = "GRADE Rating"
  ) %>%
  cols_align(
    align = "center"
  ) %>%
  tab_header(
    title = "Finalised Meta-Analysis Results for Sulphate",
    subtitle = "Subdivided by Age Band"
  ) %>%
  tab_source_note(
    source_note = md(
      "*Q p-value = Cochran's Q heterogeneity test; RoB = Risk of Bias*"
    )
  )
```

Table 1: Finalised Meta-Analysis Results for Sulphate

Subdivided by Age Band

| Analysis | Patients, n | 95% CI | Q statistic | Q p-value | GRADE Rating |
| --- | --- | --- | --- | --- | --- |
| Male | 642 | 17 – 26 | 588.12 | <0.001 | Moderate ⊕⊕⊕◯ |
| Female | 3794 | 7 – 30 | 7067.01 | <0.001 | Moderate ⊕⊕⊕◯ |
| *Q p-value = Cochran's Q heterogeneity test; RoB = Risk of Bias* | | | | | |

```
sulphate_final_results <- sulphate_final_results %>%
  mutate(
    urine_component = "sulphate"
  )
```

#### 9.17 Urea

##### 9.17.1 Framework Recommendation

framework_callout("urea")

Table 1: **Framework Recommendation — urea**

Branch 8 — Default pooled

| Component | Recommended | Branch | Reason | k | n | R² | Sex p (women_prop / overall / MR / LRoB) | Age p (min) | race p (min) | Assay p (min) | Egger's p | Q/df (chosen / overall) | Q/df ratio (chosen÷overall) | Q p (chosen) | MR R² (chosen) | QC warnings |
| --- | --- | --- | --- | --- | --- | --- | --- | --- | --- | --- | --- | --- | --- | --- | --- | --- |
| urea | Overall pooled | 8 — Default pooled | Meta-regression range 192.12–543.57 valid but k = 13 < 20 (model saturated); default to overall pooled | 13 | 3,876 | 100.0% | 0.110 / 0.663 / — / — | — | — | — | 0.278 | 255.1 / 255.1 | 1.00 | <0.001 | 100% | — |

##### 9.17.2 Comment

For 24-hour urine urea, the meta-analysis included k = 13 subgroup-level records and n = 3,876 participants. The dataset was substantially augmented by the inclusion of summarised Nurses’ Health Study II (premenopausal and postmenopausal) cohorts kindly provided by Curhan et al., taking k from 11 to 13 and n from 525 to 3,876. Heterogeneity was high (I² > 95%, Cochran’s Q p < 0.001); the data-sufficiency floor (k ≥ 5, n ≥ 500) is now comfortably met.

Sex differences on meta-regression were not significant (p = 0.110; overall sex Wald p = 0.663). Age, race and assay/country stratum tests were not testable — no powered pairs were available because the dataset remains the smallest of any analyte and the strata are sparse.

Meta-regression returned R² = 100%, which indicates a saturated/over-fit model rather than meaningful explanation of heterogeneity, given k = 13 < 20 (the framework’s sanity floor on k for meta-regression preference). Egger’s test was not significant (p = 0.278), so the small-study-bias override would not fire either.

The low risk-of-bias and RCT-only sub-analyses are too sparse to provide a more informative or conservative estimate.

The framework therefore defaulted to the unadjusted overall pooled meta-analysis. The recommended expected range is **234 – 445 mmol/24h** (Overall, All Studies, n = 3,876). The dominant caveat is the small number of contributing study cohorts (k = 13), even after the NHS II augmentation. Future analyses may benefit further from additional contemporary cohort data; the saturated meta-regression model could become interpretable once k ≥ 20.

##### 9.17.3 Plot

```
urea_ma_2nd_analysis$main_meta_analysis$plot %+%
  subset(urea_ma_2nd_analysis$main_meta_analysis$plot$data,
         age_band %in% ("Overall") & quality_filter == "All Studies")
## `height` was translated to `width`.
```

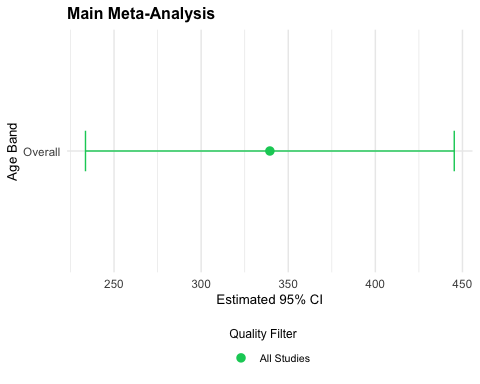

##### 9.17.4 Results Table

```
urea_ma_2nd_analysis$ma_results %>%
  filter(
    sex_group == "main meta-analysis",
    age_band %in% ("Overall"),
    quality_filter == "All Studies"
  ) %>%
  mutate(
    pooled_estimate = round(pooled_estimate, 2),
    pooled_sd       = round(pooled_sd, 2),
    I2              = round((I2 * 100), 0),
    Q               = round(Q, 2),
    Q_pval = case_when(
      Q_pval < 0.001 ~ "<0.001",
      Q_pval < 0.01  ~ "<0.01",
      Q_pval < 0.05  ~ "<0.05",
      TRUE           ~ as.character(round(Q_pval, 2))
    ),
    ci_95 = paste0(
      round(lower_95, 0), " – ", round(upper_95, 0)
    ),
    sex_group = "Meta-Regression"
  ) %>%
  select(
    sex_group,
    total_participants,
    pooled_estimate,
    pooled_sd,
    ci_95,
    I2,
    Q,
    Q_pval
  ) %>%
  gt() %>%
  cols_label(
    sex_group          = "Analysis",
    total_participants  = "Patients, n",
    pooled_estimate = "Pooled Estimate",
    pooled_sd       = "SD",
    ci_95           = "95% CI",
    I2              = html("I<sup>2</sup> (%)"),
    Q               = "Q statistic",
    Q_pval          = "Q p-value"
  ) %>%
  cols_align(
    align = "center",
    columns = c(total_participants, pooled_estimate, pooled_sd, ci_95, I2, Q, Q_pval)
  ) %>%
  tab_header(
    title = "Urea Meta-analysis Results",
    subtitle = "Meta-Regression"
  ) %>%
  tab_source_note(
    source_note = md(
      "*I² quantifies heterogeneity; Q p-value tests heterogeneity across studies.*"
    )
  )
```

Table 1: Urea Meta-analysis Results

Meta-Regression

| Analysis | Patients, n | Pooled Estimate | SD | 95% CI | I2 (%) | Q statistic | Q p-value |
| --- | --- | --- | --- | --- | --- | --- | --- |
| Meta-Regression | 3876 | 339.45 | 54 | 234 – 445 | 100 | 3061.65 | <0.001 |
| *I² quantifies heterogeneity; Q p-value tests heterogeneity across studies.* | | | | | | | |

##### 9.17.5 Bias analysis

```
urea_rob <- urine_ma2 %>%
  drop_na(urea_sd_mmol_24h) %>%
  left_join(
    overall_bias, by = "study_name"
  ) %>%
  select(study_name, overall, study_type) %>%
  mutate(
    overall = case_when(
      is.na(overall) ~ "Moderate",
      TRUE ~ overall
    )
  ) %>%
  distinct(study_name, .keep_all = TRUE) %>%
  arrange(overall) %>%
  gt() %>%
  cols_label(
    study_name = "Study",
    overall    = "Risk of Bias",
    study_type = "Study Type"
  ) %>%
  data_color(
    columns = overall,
    fn = function(x) {
      case_when(
        x == "Low"      ~ "#90EE90",
        x == "Moderate" ~ "#FFD700",
        x == "High"     ~ "#FF6B6B",
        TRUE            ~ "#FFFFFF"
      )
    }
  ) %>%
  opt_all_caps() %>%
  cols_align(
    align = "center",
    overall
  ) %>%
  tab_header(
    title = "Risk of Bias by Study and Age Band for 24 Hour Urine Urea"
  )

urea_rob
```

Table 1: Risk of Bias by Study and Age Band for 24 Hour Urine Urea

| Study | Risk of Bias | Study Type |
| --- | --- | --- |
| Nouvenne et al. 2013 | High | Observational Cohort |
| Stitchantrakul et al. 2004 | High | RCT |
| Baxmann et al. 2003 | High | RCT |
| Hess et al. 1999 | High | Observational Cohort |
| Parvin et al. 2011 | Low | Observational Cohort |
| Stitchantrakul et al. 2007 | Moderate | Observational Cohort |
| Baric et al. 2019 | Moderate | Observational Cohort |
| Bingham et al. 1988 | Moderate | Observational Cohort |
| NHS II et al. premenopausal | Moderate | Observational Cohort |
| NHS II et al. postmenopausal | Moderate | Observational Cohort |

##### 9.17.6 24 Hour Urine urea Suggested Expected Range inc. GRADE

```
urea_grade <- calculate_grade(
  study_design= "Mixed", # RCT result different
  risk_of_bias = "Very Serious", #Minimal studies with lower RoB
  inconsistency = "Serious",
  indirectness = "Moderate",
  imprecision = "Moderate",
  publication_bias = "Strongly suspected", #significant Q
  large_effect = TRUE,
  dose_response = FALSE,
  confounders_reduce_effect = mr_reduces_effect(urea_ma_2nd_analysis$ma_results)
)

urea_grade <- cbind(
  sex = c("Overall"),
  grade = urea_grade$grade_rating
) %>% as_tibble() %>%
  mutate(
    grade_rating = grade_symbol(grade)
  )

urea_final_results <- get_final_ma_results(urea_ma_2nd_analysis$ma_results, sex_specific = FALSE) %>%
  mutate(
    pooled_estimate = round(pooled_estimate, 2),
    pooled_sd       = round(pooled_sd, 2),
    Q               = round(Q, 2),
    Q_pval = case_when(
      Q_pval < 0.001 ~ "<0.001",
      Q_pval < 0.01  ~ "<0.01",
      Q_pval < 0.05  ~ "<0.05",
      TRUE           ~ sprintf("%.2f", Q_pval)
    ),
    ci_95 = paste0(
      round(lower_95, 0), " – ", round(upper_95, 0)
    )
  ) %>%
  select(
    age_band,
    total_participants,
    ci_95,
    Q,
    Q_pval
  ) %>%
  cbind(
    urea_grade) %>%
  mutate(grade_rating = paste0(grade, " ", grade_rating)) %>%
  select(-c(sex, grade))

urea_final_results %>%
  gt() %>%
  cols_label(
    age_band       = "Analysis",
    total_participants  = "Patients, n",
    ci_95           = "95% CI",
    Q               = "Q statistic",
    Q_pval          = "Q p-value",
    grade_rating    = "GRADE Rating"
  ) %>%
  cols_align(
    align = "center"
  ) %>%
  tab_header(
    title = "Finalised Meta-Analysis Results for Urea",
    subtitle = "Subdivided by Age Band"
  ) %>%
  tab_source_note(
    source_note = md(
      "*Q p-value = Cochran's Q heterogeneity test; RoB = Risk of Bias*"
    )
  )
```

Table 1: Finalised Meta-Analysis Results for Urea

Subdivided by Age Band

| Analysis | Patients, n | 95% CI | Q statistic | Q p-value | GRADE Rating |
| --- | --- | --- | --- | --- | --- |
| Overall | 3876 | 234 – 445 | 3061.65 | <0.001 | Low ⊕⊕◯◯ |
| *Q p-value = Cochran's Q heterogeneity test; RoB = Risk of Bias* | | | | | |

```
urea_final_results <- urea_final_results %>%
  mutate(
    urine_component = "urea"
  )
```

### 10 Final Table of All Results

#### 10.1 All MA Results

```
final_table <- rbind(volume_final_results,
                     calcium_final_results,
                     urate_final_results,
                     creatinine_final_results,
                     oxalate_final_results,
                     ph_final_results,
                     citrate_final_results,
                     phosphate_final_results,
                     sodium_final_results,
                     potassium_final_results,
                     magnesium_final_results,
                     ammonium_final_results,
                     chloride_final_results,
                     sulphate_final_results,
                     urea_final_results
                     )

component_cols <- c(
  "Ammonium"   = "#E2ECE9",
  "Calcium"    = "#FDE2E4",
  "Chloride"   = "#E0F2FE",
  "Citrate"    = "#FFF1C1",
  "Creatinine" = "#E2F0CB",
  "Magnesium"  = "#F1C0E8",
  "Oxalate"    = "#CDE7F0",
  "Ph"         = "#F8F9FA",
  "Phosphate"  = "#FFE5EC",
  "Potassium"  = "#D0F4DE",
  "Sodium"     = "#D3F8E2",
  "Sulphate"   = "#E4C1F9",
  "Urate"      = "#FFD6A5",
  "Urea"       = "#FAD2E1",
  "Volume"     = "#EADCF8"
)

final_gt <- final_table %>%
  mutate(urine_component = case_when(
    urine_component == "ph" ~ "pH",
    urine_component == "creat" ~ "Creatinine",
    TRUE ~ str_to_title(urine_component)
  )) %>%
  group_by(urine_component) %>%
  gt() %>%
  cols_label(
    age_band       = "Meta-Analysis Type",
    total_participants  = "Patients, n",
    ci_95           = "95% CI",
    Q               = "Q statistic",
    Q_pval          = "Q p-value",
    grade_rating    = "GRADE Rating"
  ) %>%
  cols_align(
    align = "center",
    -age_band
  ) %>%
  tab_header(
    title = "Finalised 24 Hour Urine Expected Range Suggestions",
    subtitle = "Recommendations based on meta-analyses with most robust evidence"
  ) %>%
  tab_source_note(
    source_note = md(
      "*Q p-value = Cochran's Q heterogeneity test; RoB = Risk of Bias*"
    )
  ) %>%
  opt_row_striping() %>%
  tab_options(
    row.striping.background_color = "lightgrey"
  )

row_groups <- final_gt[["_row_groups"]]

for (comp in names(component_cols)) {

  matching <- row_groups[str_detect(row_groups, comp)]

  final_gt <- final_gt %>%
    tab_style(
      style = list(
        cell_fill(color = component_cols[comp]),
        cell_text(weight = "bold")
      ),
      locations = cells_row_groups(groups = matching)
    )
}

final_gt
```

Table 1: Finalised 24 Hour Urine Expected Range Suggestions

Recommendations based on meta-analyses with most robust evidence

| Meta-Analysis Type | Patients, n | 95% CI | Q statistic | Q p-value | GRADE Rating |
| --- | --- | --- | --- | --- | --- |
| **Volume** | | | | | |
| Overall | 9144 | 1 – 2.5 | 9158.78 | <0.001 | High ⊕⊕⊕⊕ |
| **Calcium** | | | | | |
| Meta-Regression | 5418 | 1.9 – 6.2 | 2744.79 | <0.001 | Low ⊕⊕◯◯ |
| **Urate** | | | | | |
| Male | 860 | 2 – 5.1 | 1118.46 | <0.001 | Very Low ⊕◯◯◯ |
| Female | 4177 | 2.3 – 3.5 | 1324.26 | <0.001 | Very Low ⊕◯◯◯ |
| **Creatinine** | | | | | |
| Male | 760 | 12.2 – 16.3 | 196.17 | <0.001 | Very Low ⊕◯◯◯ |
| Female | 4217 | 7.2 – 12.6 | 1625.32 | <0.001 | Low ⊕⊕◯◯ |
| **Oxalate** | | | | | |
| Male | 860 | 0.2 – 0.6 | 3373.74 | <0.001 | Very Low ⊕◯◯◯ |
| Female | 4177 | 0.2 – 0.4 | 439.70 | <0.001 | Very Low ⊕◯◯◯ |
| **Ph** | | | | | |
| Meta-Regression | 4379 | 5.6 – 6.3 | 324.21 | <0.001 | Moderate ⊕⊕⊕◯ |
| **Citrate** | | | | | |
| Overall | 9458 | 1.2 – 4.5 | 16945.00 | <0.001 | Very Low ⊕◯◯◯ |
| **Phosphate** | | | | | |
| Meta-Regression | 3974 | 19 – 26 | 183.33 | <0.001 | Low ⊕⊕◯◯ |
| **Sodium** | | | | | |
| Meta-Regression | 3945 | 98 – 253 | 857.39 | <0.001 | Moderate ⊕⊕⊕◯ |
| **Potassium** | | | | | |
| Overall | 9418 | 28 – 77 | 7644.37 | <0.001 | Very Low ⊕◯◯◯ |
| **Magnesium** | | | | | |
| Meta-Regression | 4724 | 3.1 – 4.8 | 306.00 | <0.001 | Moderate ⊕⊕⊕◯ |
| **Ammonium** | | | | | |
| Male | 535 | 28 – 46 | 267.48 | <0.001 | Moderate ⊕⊕⊕◯ |
| Female | 3857 | 20 – 40 | 1303.78 | <0.001 | Moderate ⊕⊕⊕◯ |
| **Chloride** | | | | | |
| Meta-Regression | 3173 | 84 – 218 | 553.32 | <0.001 | Moderate ⊕⊕⊕◯ |
| **Sulphate** | | | | | |
| Male | 642 | 17 – 26 | 588.12 | <0.001 | Moderate ⊕⊕⊕◯ |
| Female | 3794 | 7 – 30 | 7067.01 | <0.001 | Moderate ⊕⊕⊕◯ |
| **Urea** | | | | | |
| Overall | 3876 | 234 – 445 | 3061.65 | <0.001 | Low ⊕⊕◯◯ |
| *Q p-value = Cochran's Q heterogeneity test; RoB = Risk of Bias* | | | | | |

```
# Persist framework-aligned final_table for the Shiny app
if (!dir.exists("shiny_app_data")) dir.create("shiny_app_data", showWarnings = FALSE)
saveRDS(final_table, "shiny_app_data/final_table.rds")
message("Saved framework-aligned final_table.rds (", nrow(final_table), " rows)")
## Saved framework-aligned final_table.rds (20 rows)
# Also persist the current guideline reference ranges + bias tables for the app
saveRDS(current_ref_range, "shiny_app_data/current_ref_range.rds")
saveRDS(bias_observational, "shiny_app_data/bias_observational.rds")
saveRDS(bias_rcts,          "shiny_app_data/bias_rcts.rds")
message("Saved current_ref_range.rds (", nrow(current_ref_range),
        " rows), bias_observational.rds, bias_rcts.rds")
## Saved current_ref_range.rds (125 rows), bias_observational.rds, bias_rcts.rds
# Pre-compute creatinine-completeness sensitivity for every component and
# save for the Shiny app's Results Explorer tab. Skips "creat" itself
# (would be circular). Each row carries the component name + the standard
# creat_completeness_ma() output (group, n_studies, n_participants, pooled,
# CI). The app will compute Δ vs all-studies on the fly per component.
creat_completeness_results <- purrr::map_dfr(
  c("volume", "ph", "calcium", "oxalate", "urate", "citrate",
    "phosphate", "sodium", "potassium", "magnesium", "ammonium",
    "chloride", "sulphate", "urea"),
  function(comp) {
    res <- tryCatch(creat_completeness_ma(comp, data = urine_ma2),
                    error = function(e) NULL)
    if (is.null(res) || !nrow(res)) return(tibble::tibble())
    res %>% dplyr::mutate(urine_component = comp, .before = 1)
  }
)

if (!dir.exists("shiny_app_data")) dir.create("shiny_app_data", showWarnings = FALSE)
saveRDS(creat_completeness_results, "shiny_app_data/creat_completeness.rds")
message("Saved creat_completeness.rds (",
        dplyr::n_distinct(creat_completeness_results$urine_component),
        " components, ", nrow(creat_completeness_results), " rows)")
## Saved creat_completeness.rds (14 components, 42 rows)
```

#### 10.2 Comparison to Current Expected Range

```
summary_table_separated <- summary_table_data %>%
  mutate(
    sex_group = case_when(
      str_detect(urine_component, regex("women only", ignore_case = TRUE)) ~ "Women",
      str_detect(urine_component, regex("men only", ignore_case = TRUE)) ~ "Men",
      str_detect(urine_component, regex("overall", ignore_case = TRUE)) ~ "Overall",
      TRUE ~ "Overall"
    ),
    urine_component = urine_component %>%
      str_remove(regex("\\s*-\\s*(men only|women only|overall)", ignore_case = TRUE)) %>%
      str_trim()
  ) %>%
  arrange(urine_component)

proposed_lookup <- final_table %>%
  mutate(
    urine_component = str_to_title(urine_component),
    sex_group = case_when(age_band == "Male" ~ "Men",
                          age_band == "Female" ~ "Women",
                          age_band == "Main Meta-Analysis" ~ "Overall",
                          age_band == "Meta-Regression" ~ "Overall",
                          TRUE ~ NA_character_),
    `Expected Range` = ci_95,
    "Expected Range GRADE Rating" = grade_rating,
    join_key = str_to_lower(urine_component),
    .keep = "unused"
  ) %>%
  dplyr::select(urine_component,
                sex_group,
                `Expected Range`,
                `Expected Range GRADE Rating`,
                join_key) %>%
  mutate(
    sex_group = case_when(
      is.na(sex_group) ~ "Overall",

      TRUE ~ sex_group
    )
  )

gt_tbl <- summary_table_separated %>%
  mutate(join_key = str_to_lower(str_remove(urine_component, "\\s*\\(.*\\)"))) %>%
  left_join(
    proposed_lookup %>% select(join_key, sex_group, `Expected Range`, `Expected Range GRADE Rating`),
    by = c("join_key", "sex_group")
  ) %>%
  select(-join_key) %>%
  bind_rows(
    proposed_lookup %>%
      filter(!join_key %in% c("ph")) %>%
      anti_join(
        summary_table_separated %>%
          mutate(join_key = str_to_lower(str_remove(urine_component, "\\s*\\(.*\\)"))),
        by = c("join_key", "sex_group")
      ) %>%
      mutate(urine_component = case_when(
        join_key == "volume" ~ "Volume (L / 24hrs)",
        join_key == "ph" ~ "Ph",
        TRUE ~ paste0(str_to_title(join_key), " (Mmol / 24hrs)")
      )) %>%
      select(-join_key)
  ) %>%
  arrange(factor(sex_group, levels = c("Overall", "Men", "Women")), urine_component) %>%
  group_by(urine_component) %>%
  gt(groupname_col = "urine_component") %>%
  cols_move_to_start(columns = sex_group) %>%
  tab_style(
    style = list(cell_fill(color = "#FFF3CD"), cell_text(weight = "bold")),
    locations = list(
      cells_body(columns = `Expected Range`),
      cells_column_labels(columns = `Expected Range`)
    )
  ) %>%
  fmt_missing(columns = everything(), missing_text = "-") %>%
  tab_style(style = cell_text(weight = "bold"),
            locations = cells_body(columns = sex_group)) %>%
  cols_label(sex_group = "Sex", urine_component = "Urine Component") %>%
  cols_align("center",
             c(EAU, NDH, NHS, Litholink, `Mayo Clinic`,`Expected Range`, `Expected Range GRADE Rating`)) %>%
  tab_style(style = list(cell_text(weight = "bold")),
            locations = cells_column_labels(everything())) %>%
  opt_row_striping() %>%
  tab_options(table.font.size = 12,
              row.striping.background_color = "lightgrey")

# Apply row group colours
row_groups <- gt_tbl[["_row_groups"]]
for (comp in names(component_cols)) {
  matching <- row_groups[str_detect(row_groups, comp)]
  if (length(matching) > 0) {
    gt_tbl <- gt_tbl %>%
      tab_style(
        style = list(cell_fill(color = component_cols[comp]), cell_text(weight = "bold")),
        locations = cells_row_groups(groups = matching)
      )
  }
}

gt_tbl
```

| Sex | EAU | NHS | Mayo Clinic | Litholink | NDH | Expected Range | Expected Range GRADE Rating |
| --- | --- | --- | --- | --- | --- | --- | --- |
| **Ammonium (Mmol / 24hrs)** | | | | | | | |
| **Overall** | < 50 | - | 15 - 56 | 18 - 56 | - | **-** | - |
| **Men** | - | - | - | - | - | **28 – 46** | Moderate ⊕⊕⊕◯ |
| **Women** | - | - | - | - | - | **20 – 40** | Moderate ⊕⊕⊕◯ |
| **Calcium (Mmol / 24hrs)** | | | | | | | |
| **Overall** | < 8 | 2.5 - 7.5 | < 5 | - | 2 - 7.4 | **1.9 – 6.2** | Low ⊕⊕◯◯ |
| **Men** | - | - | - | 1 - 6.2 | - | **-** | - |
| **Women** | - | - | - | 0.7 - 5 | - | **-** | - |
| **Chloride (Mmol / 24hrs)** | | | | | | | |
| **Overall** | - | 110 - 250 | 34 - 286 | 50 - 120 | 110 - 250 | **84 – 218** | Moderate ⊕⊕⊕◯ |
| **Citrate (Mmol / 24hrs)** | | | | | | | |
| **Overall** | - | 0.6 - 4.8 | 1.3 - 6.2 | - | - | **1.2 – 4.5** | Very Low ⊕◯◯◯ |
| **Men** | > 1.7 | - | - | > 2.1 | - | **-** | - |
| **Women** | > 1.9 | - | - | > 2.8 | - | **-** | - |
| **Magnesium (Mmol / 24hrs)** | | | | | | | |
| **Overall** | > 3 | 2.4 - 6.5 | 2.1 - 4.3 | 2.6 - 7.1 | - | **3.1 – 4.8** | Moderate ⊕⊕⊕◯ |
| **Oxalate (Mmol / 24hrs)** | | | | | | | |
| **Overall** | < 0.5 | 0.1 - 0.5 | 0.1 - 0.5 | - | - | **-** | - |
| **Men** | - | - | - | < 0.5 | - | **0.2 – 0.6** | Very Low ⊕◯◯◯ |
| **Women** | - | - | - | < 0.4 | - | **0.2 – 0.4** | Very Low ⊕◯◯◯ |
| **Ph** | | | | | | | |
| **Overall** | 5.5 - 7 | - | 4.5 - 8 | 5.7 - 6.3 | 4.5 - 8 | **5.6 – 6.3** | Moderate ⊕⊕⊕◯ |
| **Phosphate (Mmol / 24hrs)** | | | | | | | |
| **Overall** | < 35 | 15 - 50 | 7.3 - 58 | 19.4 - 38.7 | 12.9 - 42 | **19 – 26** | Low ⊕⊕◯◯ |
| **Potassium (Mmol / 24hrs)** | | | | | | | |
| **Overall** | - | 25 - 125 | 16 - 105 | 40 - 100 | 25 - 125 | **28 – 77** | Very Low ⊕◯◯◯ |
| **Sodium (Mmol / 24hrs)** | | | | | | | |
| **Overall** | - | 40 - 250 | 22 - 328 | 50 - 120 | 40 - 220 | **98 – 253** | Moderate ⊕⊕⊕◯ |
| **Sulphate (Mmol / 24hrs)** | | | | | | | |
| **Overall** | - | - | 7 - 47 | 12.5 - 28 | - | **-** | - |
| **Men** | - | - | - | - | - | **17 – 26** | Moderate ⊕⊕⊕◯ |
| **Women** | - | - | - | - | - | **7 – 30** | Moderate ⊕⊕⊕◯ |
| **Urate (Mmol / 24hrs)** | | | | | | | |
| **Overall** | - | 1.5 - 4.5 | - | - | 1.5 - 4.5 | **-** | - |
| **Men** | < 5 | - | 1.2 - 5.9 | 2.7 - 4.8 | - | **2 – 5.1** | Very Low ⊕◯◯◯ |
| **Women** | < 4 | - | 1.5 - 4.5 | 2.2 - 4.5 | - | **2.3 – 3.5** | Very Low ⊕◯◯◯ |
| **Urea (Mmol / 24hrs)** | | | | | | | |
| **Overall** | - | 250 - 570 | 124.9 - 749.7 | 249.9 - 535.5 | 250 - 4.5 | **234 – 445** | Low ⊕⊕◯◯ |
| **Volume (L / 24hrs)** | | | | | | | |
| **Overall** | > 0.5 | < 3 | - | > 2.5 | - | **1 – 2.5** | High ⊕⊕⊕⊕ |
| **Creatinine (Mmol / 24hrs)** | | | | | | | |
| **Men** | 13 - 18 | 9 - 19 | 8.2 - 26.1 | - | 7.1 - 17.7 | **12.2 – 16.3** | Very Low ⊕◯◯◯ |
| **Women** | 7 - 13 | 6 - 13 | 5.3 - 15.8 | - | 5.3 - 15.9 | **7.2 – 12.6** | Low ⊕⊕◯◯ |
